# Mapping risks of hospital-recorded health conditions in people with eczema

**DOI:** 10.1101/2025.03.24.25324470

**Authors:** Julian Matthewman, Anna Schultze, Krishnan Bhaskaran, Amanda Roberts, Spiros Denaxas, Kathryn E Mansfield, Sinéad M Langan

## Abstract

Atopic eczema may be associated with multiple health conditions. Here, we systematically explored risks across the full health spectrum based on the International Classification of Diseases, assessing associations between eczema and 2,058 ICD-10 codes, 1,593 phecodes, and 201 Global Burden of Disease codes.

In English primary care electronic health records (1997−2023) we identified cohorts of people with eczema (up to 3 million) and compared to matched comparators (individuals without eczema (by age, sex, general practice) (up to 14 million). In up to 25 years of follow-up, we captured outcomes recorded during hospital admissions.

People with eczema had higher rates of several outcomes across multiple organ systems. Among those followed up from eczema diagnosis in childhood, atopic/allergic conditions and infections accounted for most excess diagnoses. Besides skin and atopic/allergic outcomes, lymphomas, coeliac disease, inflammatory bowel conditions, and specific eye diseases had particularly large relative risk increases.

## Introduction

Eczema, also referred to as atopic eczema or atopic dermatitis, is common and associated with the subsequent occurrence and co-occurrence of other atopic diseases such as asthma and allergies. Associations with subsequent non-atopic diseases are less clear.^1,2^ Our previous study offered evidence on a range of outcomes – recorded in primary care – chosen for their inclusion in guidelines or previous research.^1,3^ Knowledge of risks across the full health spectrum faced by people with eczema is however still limited, as is evidence on numerous individual disease outcomes, in particular those better captured in a hospital setting.

Diagnoses in UK hospital records are structured using the International Classification of Diseases (ICD) Version 10; a hierarchical terminology of which the 2,058 codes of the third level (“category-level”) correspond to diseases (e.g., asthma, atopic dermatitis, scabies, impetigo, psoriasis, etc…), symptoms, or health-related occurrences. ICD-10 codes can also be mapped to other disease classification systems, including phecodes (covering 1,593 diseases)^4^ and Global Burden of Disease codes (covering 201 diseases).^5^

Here we used the hierarchical nature of ICD-10 codes to systematically explore associations between eczema and outcomes recorded as part of hospital admissions, with the aim of learning about risks across the full health spectrum.

## Results

### Descriptive Statistics

From English primary care data from the Clinical Research Practice Datalink (CPRD) Aurum linked to Hospital Episode Statistics (HES) Admitted Patient Care (APC) data from April 1st 1997 to March 31st 2023 (N= 35,968,445), we identified 3,513,875 individuals who met the eczema definition (at least one record of an eczema diagnostic code and at least two records for eczema therapies in primary care) and were eligible for matching. We matched them with up to five eligible individuals who had no record of eczema at the diagnosis date of the exposed people. Without restrictions on age (any-age cohort) this resulted in a cohort of 17,180,124 (with and without eczema); 7,478,587 when including only individuals who met the eczema definition before their 18^th^ birthday (<18 cohort), 12,772,961 for a cohort of individuals at least 18 years of age (18+ cohort), and 7,075,624 for a cohort of individuals at least 40 years of age (40+ cohort) (**Figure 1**). For analyses of each specific outcome, individuals who already had a hospital record for that outcome of interest at index date were excluded (e.g., when assessing the association with asthma, 1,545,003 [9%] who already had a hospital record for asthma were excluded; for stroke 113,085 [0.7%] were excluded). We assessed the presence of primary-care-recorded comorbidities at index date, for some of which there were considerable differences between groups (e.g., previous asthma 12% in unexposed versus 21% in exposed) (**Table 1, Supplementary Table 1**).

**Figure 1:**
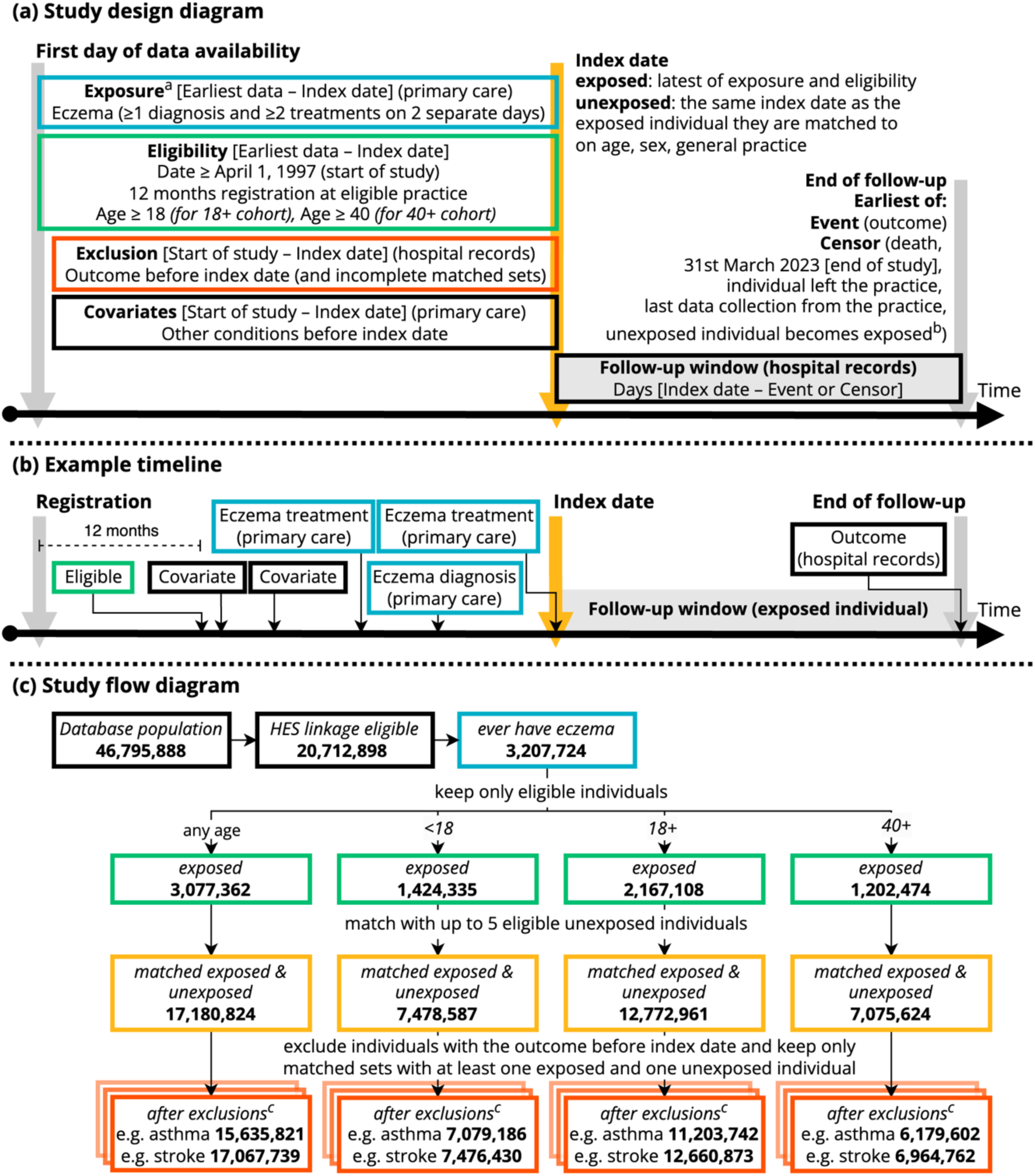
Study design and flow diagram. (a) Study design diagram, (b) example timeline, and (c) Study flow diagram, colour-coded by step. ^a^ Treatments include emollients, topical glucocorticoids, topical calcineurin inhibitors, systemic immunosuppressants (azathioprine, methotrexate, ciclosporin, mycophenolate), and oral glucocorticoids. ^b^ Unexposed individuals are censored on the day they meet the eczema diagnostic algorithm themselves, and can then be re-matched, this time as exposed individuals. ^c^Example outcomes are J45 “Asthma” and I63 “Cerebral infarction”.

**Table 1:**
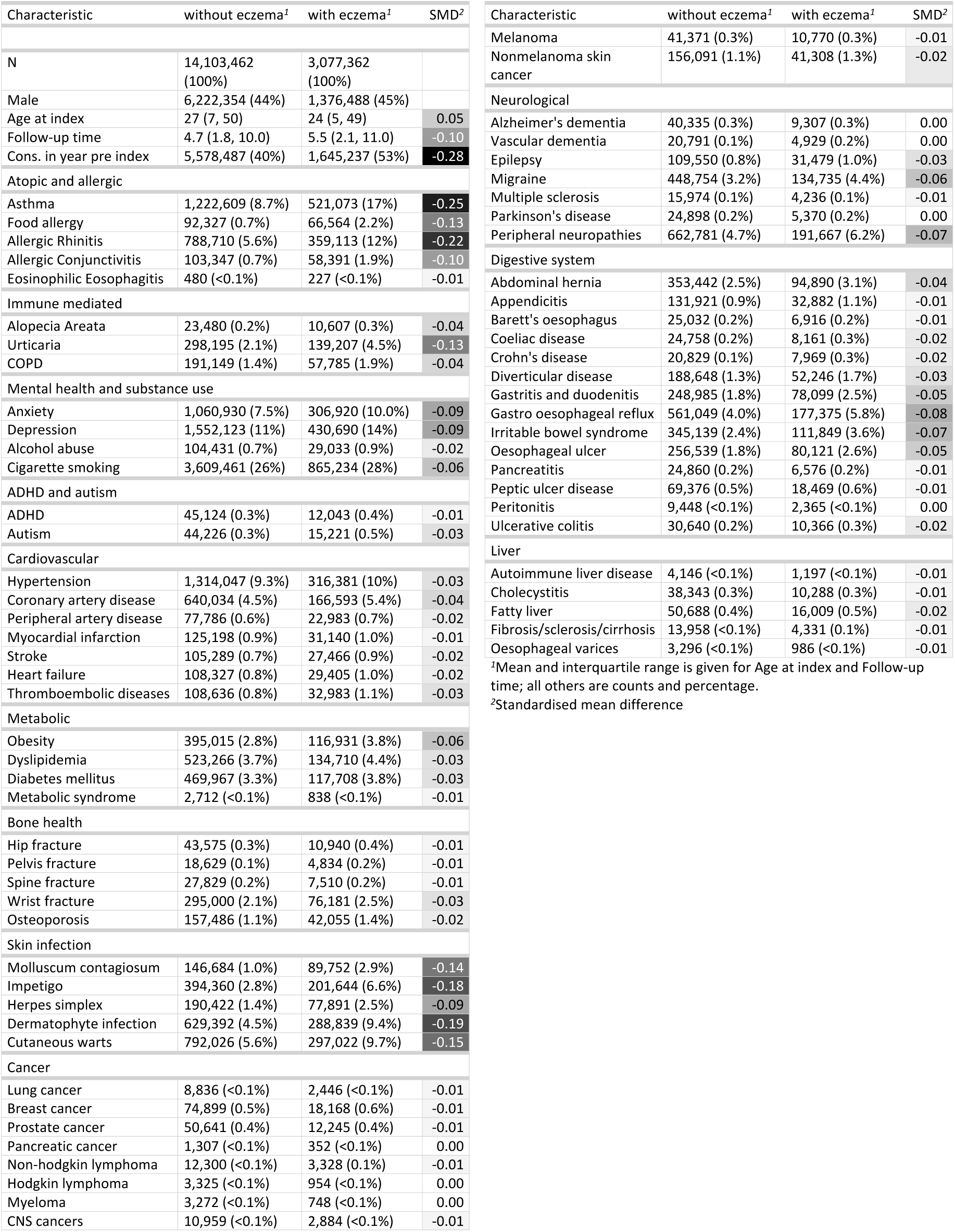
Baseline characteristics recorded in primary care (any-age cohort) Demographic and clinical characteristics at baseline (index date). All clinical characteristics are assessed in primary care.

For each outcome, individuals were followed up using their hospital admission records until they had the outcome of interest (event) recorded in any diagnostic position (i.e., primary or secondary reasons for admission or medical history), or were censored (died, left the GP practice, last data collection from practice, or the 31^st^ March 2023). Median follow-up time was 5.5 (interquartile range 2.1, 11.0) years in the any-age cohort, 4.8 (1.9, 9.9) for the 18+ cohort, 6.2 (2.7, 11.3) for the 40+ cohort, and 5.2 (2.0, 10.8) for the <18 cohort (in the unexposed 4.7 [1.8, 10.0], 4.4 [1.8, 9.3], 5.8 [2.5, 10.8], and 3.9 [1.5, 9.1], respectively) (**Table 1, Supplementary Table 1**).

### Associations between eczema and subsequent hospital-recorded diagnoses

For each outcome of interest, we estimated rate differences, hazard ratios and p-values from Cox regression stratified on matched set. We provide these results from crude and comorbidity-adjusted models (adjusted for 71 baseline conditions assessed in primary care from the following disease categories: Atopic and allergic, Immune mediated, Mental health and substance use, attention deficit hyperactivity disorder and autism, Cardiovascular, Metabolic, Bone health, Skin infection, Cancer, Neurological, Digestive system, Liver), by age cohort (any-age, <18, 18+, 40+), and from sensitivity analyses (1. Including only individuals that had been hospitalised at least one year before index date; 2. excluding non-consulters, i.e., individuals who did not have any of four common records [9N11.00 - Seen in GP’s surgery, 22A..00 - Body weight, 4….00 - Laboratory test, 246..00 - O/E - blood pressure reading] in the year prior to index date). In **Table 2** we show examples of synthesising all available information for selected outcomes.

**Table 2:**
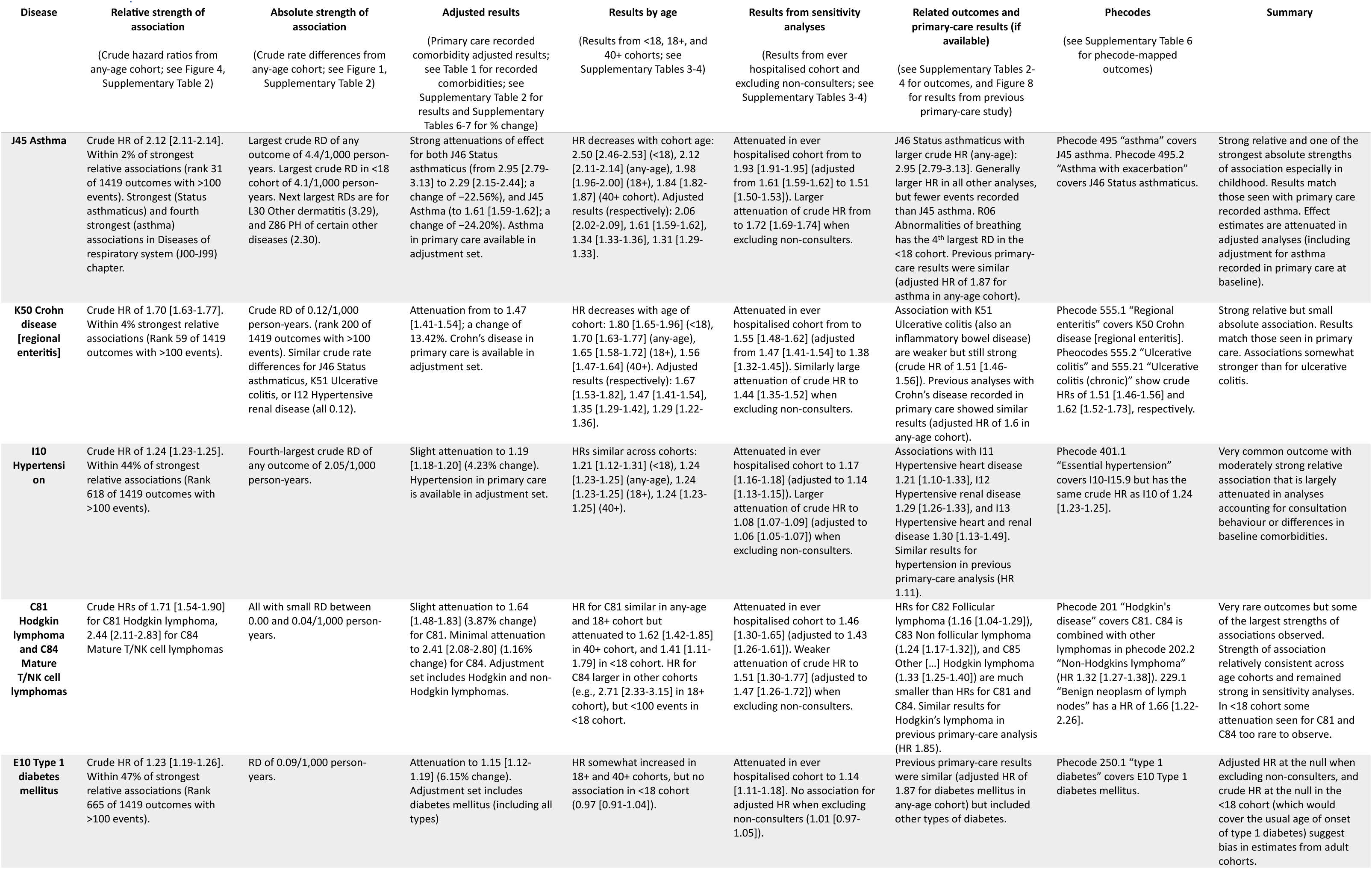
Example associations. Examples demonstrating the range of available results and how these can be used to examine individual associations.

| Disease | Relative strength of association | Absolute strength of association | Adjusted results | Results by age | Results from sensitivity analyses | Related outcomes and primary-care results (if available) | Phecodes | Summary |
| --- | --- | --- | --- | --- | --- | --- | --- | --- |
|  | (Crude hazard ratios from any-age cohort; see Figure 4, Supplementary Table 2) | (Crude rate differences from any-age cohort; see Figure 1, Supplementary Table 2) | (Primary care recorded comorbidity adjusted results; see Table 1 for recorded comorbidities; see Supplementary Table 2 for results and Supplementary Tables 6-7 for % change) | (Results from <18, 18+, and 40+ cohorts; see Supplementary Tables 3-4) | (Results from ever hospitalised cohort and excluding non-consulters; see Supplementary Tables 3-4) | (see Supplementary Tables 2-4 for outcomes, and Figure 8 for results from previous primary-care study) | (see Supplementary Table 6 for phecode-mapped outcomes) |  |
| **J45 Asthma** | Crude HR of 2.12 [2.11-2.14]. Within 2% of strongest relative associations (rank 31 of 1419 outcomes with >100 events). Strongest (Status asthmaticus) and fourth strongest (asthma) associations in Diseases of respiratory system (J00-J99) chapter. | Largest crude RD of any outcome of 4.4/1,000 person-years. Largest crude RD in <18 cohort of 4.1/1,000 person-years. Next largest RDs are for L30 Other dermatitis (3.29), and Z86 PH of certain other diseases (2.30). | Strong attenuations of effect for both J46 Status asthmaticus (from 2.95 [2.79-3.13] to 2.29 [2.15-2.44]; a change of -22.56%), and J45 Asthma (to 1.61 [1.59-1.62]; a change of -24.20%). Asthma in primary care available in adjustment set. | HR decreases with cohort age: 2.50 [2.46-2.53] (<18), 2.12 [2.11-2.14] (any-age), 1.98 [1.96-2.00] (18+), 1.84 [1.82-1.87] (40+ cohort). Adjusted results (respectively): 2.06 [2.02-2.09], 1.61 [1.59-1.62], 1.34 [1.33-1.36], 1.31 [1.29-1.33]. | Attenuated in ever hospitalised cohort from to 1.93 [1.91-1.95] (adjusted from 1.61 [1.59-1.62] to 1.51 [1.50-1.53]). Larger attenuation of crude HR from to 1.72 [1.69-1.74] when excluding non-consulters. | J46 Status asthmaticus with larger crude HR (any-age): 2.95 [2.79-3.13]. Generally larger HR in all other analyses, but fewer events recorded than J45 asthma. R06 Abnormalities of breathing has the 4<sup>th</sup> largest RD in the <18 cohort. Previous primary-care results were similar (adjusted HR of 1.87 for asthma in any-age cohort). | Phecode 495 “asthma” covers J45 asthma. Phecode 495.2 “Asthma with exacerbation” covers J46 Status asthmaticus. | Strong relative and one of the strongest absolute strengths of association especially in childhood. Results match those seen with primary care recorded asthma. Effect estimates are attenuated in adjusted analyses (including adjustment for asthma recorded in primary care at baseline). |
| **K50 Crohn disease [regional enteritis]** | Crude HR of 1.70 [1.63-1.77]. Within 4% strongest relative associations (Rank 59 of 1419 outcomes with >100 events). | Crude RD of 0.12/1,000 person-years. (rank 200 of 1419 outcomes with >100 events). Similar crude rate differences for J46 Status asthmaticus, K51 Ulcerative colitis, or I12 Hypertensive renal disease (all 0.12). | Attenuation from to 1.47 [1.41-1.54]; a change of 13.42%. Crohn’s disease in primary care is available in adjustment set. | HR decreases with age of cohort: 1.80 [1.65-1.96] (<18), 1.70 [1.63-1.77] (any-age), 1.65 [1.58-1.72] (18+), 1.56 [1.47-1.64] (40+). Adjusted results (respectively): 1.67 [1.53-1.82], 1.47 [1.41-1.54], 1.35 [1.29-1.42], 1.29 [1.22-1.36]. | Attenuated in ever hospitalised cohort from to 1.55 [1.48-1.62] (adjusted from 1.47 [1.41-1.54] to 1.38 [1.32-1.45]). Similarly large attenuation of crude HR to 1.44 [1.35-1.52] when excluding non-consulters. | Association with K51 Ulcerative colitis (also an inflammatory bowel disease) are weaker but still strong (crude HR of 1.51 [1.46-1.56]). Previous analyses with Crohn’s disease recorded in primary care showed similar results (adjusted HR of 1.6 in any-age cohort). | Phecode 555.1 “Regional enteritis” covers K50 Crohn disease [regional enteritis]. Phecodes 555.2 “Ulcerative colitis” and 555.21 “Ulcerative colitis (chronic)” show crude HRs of 1.51 [1.46-1.56] and 1.62 [1.52-1.73], respectively. | Strong relative but small absolute association. Results match those seen in primary care. Associations somewhat stronger than for ulcerative colitis. |
| **I10 Hypertension** | Crude HR of 1.24 [1.23-1.25]. Within 44% of strongest relative associations (Rank 618 of 1419 outcomes with >100 events). | Fourth-largest crude RD of any outcome of 2.05/1,000 person-years. | Slight attenuation to 1.19 [1.18-1.20] (4.23% change). Hypertension in primary care is available in adjustment set. | HRs similar across cohorts: 1.21 [1.12-1.31] (<18), 1.24 [1.23-1.25] (any-age), 1.24 [1.23-1.25] (18+), 1.24 [1.23-1.25] (40+). | Attenuated in ever hospitalised cohort to 1.17 [1.16-1.18] (adjusted to 1.14 [1.13-1.15]). Larger attenuation of crude HR to 1.08 [1.07-1.09] (adjusted to 1.06 [1.05-1.07]) when excluding non-consulters. | Associations with I11 Hypertensive heart disease 1.21 [1.10-1.33], I12 Hypertensive renal disease 1.29 [1.26-1.33], and I13 Hypertensive heart and renal disease 1.30 [1.13-1.49]. Similar results for hypertension in previous primary-care analysis (HR 1.11). | Phecode 401.1 “Essential hypertension” covers I10-I15.9 but has the same crude HR as I10 of 1.24 [1.23-1.25]. | Very common outcome with moderately strong relative association that is largely attenuated in analyses accounting for consultation behaviour or differences in baseline comorbidities. |
| **C81 Hodgkin lymphoma and C84 Mature T/NK cell lymphomas** | Crude HRs of 1.71 [1.54-1.90] for C81 Hodgkin lymphoma, 2.44 [2.11-2.83] for C84 Mature T/NK cell lymphomas | All with small RD between 0.00 and 0.04/1,000 person-years. | Slight attenuation to 1.64 [1.48-1.83] (3.87% change) for C81. Minimal attenuation to 2.41 [2.08-2.80] (1.16% change) for C84. Adjustment set includes Hodgkin and non-Hodgkin lymphomas. | HR for C81 similar in any-age and 18+ cohort but attenuated to 1.62 [1.42-1.85] in 40+ cohort, and 1.41 [1.11-1.79] in <18 cohort. HR for C84 larger in other cohorts (e.g., 2.71 [2.33-3.15] in 18+ cohort), but <100 events in <18 cohort. | Attenuated in ever hospitalised cohort to 1.46 [1.30-1.65] (adjusted to 1.43 [1.26-1.61]). Weaker attenuation of crude HR to 1.51 [1.30-1.77] (adjusted to 1.47 [1.26-1.72]) when excluding non-consulters. | HRs for C82 Follicular lymphoma (1.16 [1.04-1.29]), C83 Non follicular lymphoma (1.24 [1.17-1.32]), and C85 Other [...] Hodgkin lymphoma (1.33 [1.25-1.40]) are much smaller than HRs for C81 and C84. Similar results for Hodgkin’s lymphoma in previous primary-care analysis (HR 1.85). | Phecode 201 “Hodgkin’s disease” covers C81. C84 is combined with other lymphomas in phecode 202.2 “Non-Hodgkins lymphoma” (HR 1.32 [1.27-1.38]). 229.1 “Benign neoplasm of lymph nodes” has a HR of 1.66 [1.22-2.26]. | Very rare outcomes but some of the largest strengths of associations observed. Strength of association relatively consistent across age cohorts and remained strong in sensitivity analyses. In <18 cohort some attenuation seen for C81 and C84 too rare to observe. |
| **E10 Type 1 diabetes mellitus** | Crude HR of 1.23 [1.19-1.26]. Within 47% of strongest relative associations (Rank 665 of 1419 outcomes with >100 events). | RD of 0.09/1,000 person-years. | Attenuation to 1.15 [1.12-1.19] (6.15% change). Adjustment set includes diabetes mellitus (including all types) | HR somewhat increased in 18+ and 40+ cohorts, but no association in <18 cohort (0.97 [0.91-1.04]). | Attenuated in ever hospitalised cohort to 1.14 [1.11-1.18]. No association for adjusted HR when excluding non-consulters (1.01 [0.97-1.05]). | Previous primary-care results were similar (adjusted HR of 1.87 for diabetes mellitus in any-age cohort) but included other types of diabetes. | Phecode 250.1 “type 1 diabetes” covers E10 Type 1 diabetes mellitus. | Adjusted HR at the null when excluding non-consulters, and crude HR at the null in the <18 cohort (which would cover the usual age of onset of type 1 diabetes) suggest bias in estimates from adult cohorts. |

#### p-values

Of the 2,058 tested associations between eczema and outcomes based on ICD-10 codes (crude results from Cox regression), 44% (any-age), 11% (<18), 44% (18+), and 40% (40+) had p-values below the Bonferroni-corrected 1% significance level (0.01/2,058 = 0.000005) (**Figure 2**). The percentage of significant outcomes decreased to 36% (any-age), 8% (<18), 30% (18+), and 29% (40+) when adjusting for comorbidities at baseline, and 21% (any-age), 2% (<18), 17% (18+), and 17% (40+) when also excluding non-consulters.

**Figure 2:**
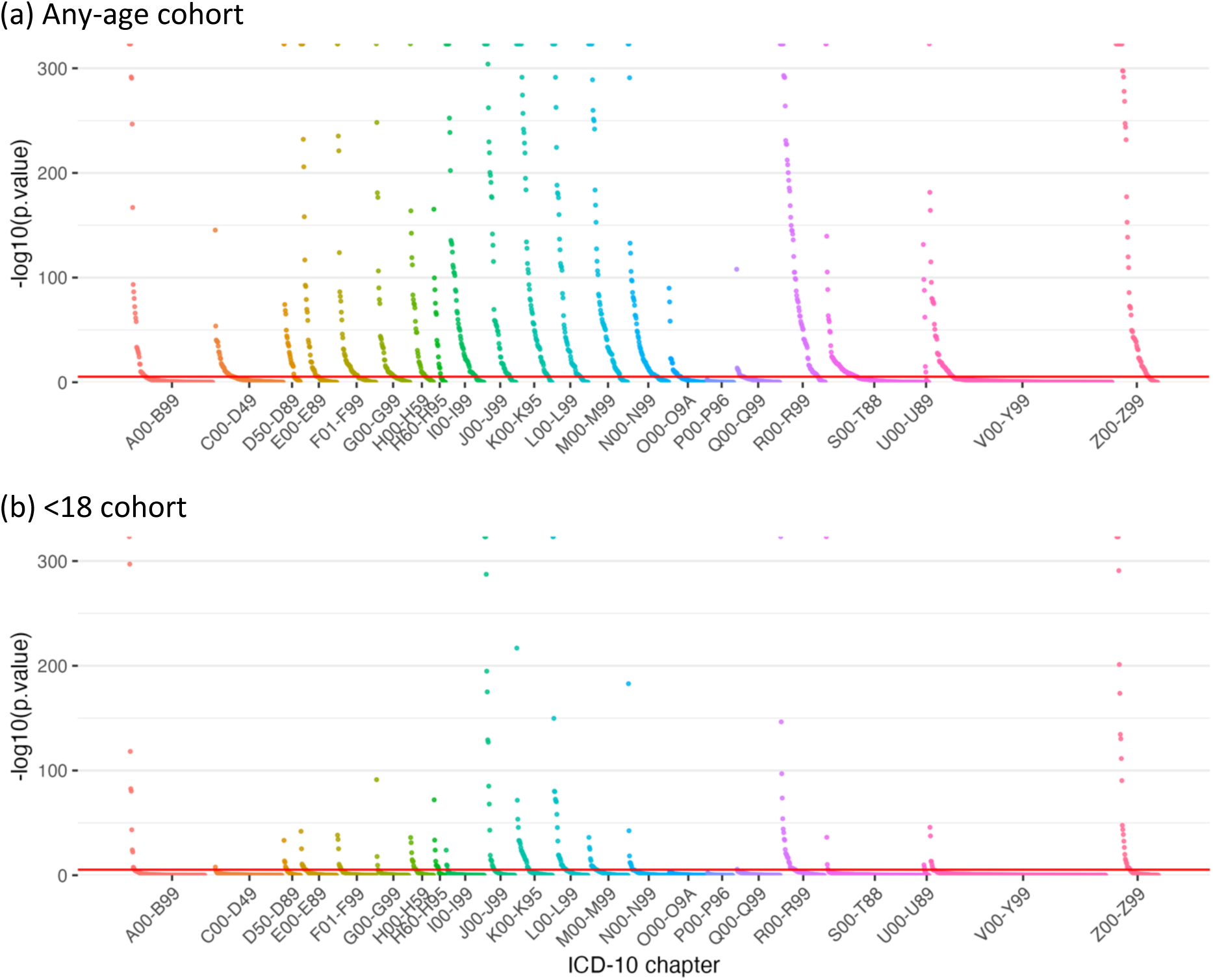
p-values. Plot of p-values from crude Cox-regression for each ICD-10 category from (a) the any-age cohort, (b) the <18 cohort. The red horizontal line shows the Bonferroni-corrected significance level with 2,056 outcomes (0.01/2058).

#### Excess rate

Some of the largest excess rates of recorded outcomes in people with eczema were seen in Chapters R (clinical signs and symptoms) (e.g., rate difference per 1,000 person-years [RD] for Abnormalities of breathing: 1.22 in any-age cohort; 1.66 in the <18 cohort), and Z (health status) (e.g., RD for Personal history of risk factors: 1.34 in any-age cohort; 1.48 in <18 cohort). Together, outcomes in chapters R and Z made up more than a quarter of the total excess burden (calculated as the sum of the excess rate in people with eczema for each outcome) in both the any-age and <18 cohorts. L30 Other dermatitis had a large excess rate in all cohorts (3.29 in any-age cohort; 3.96 in <18 cohort) (**Figure 3**).

**Figure 3:**
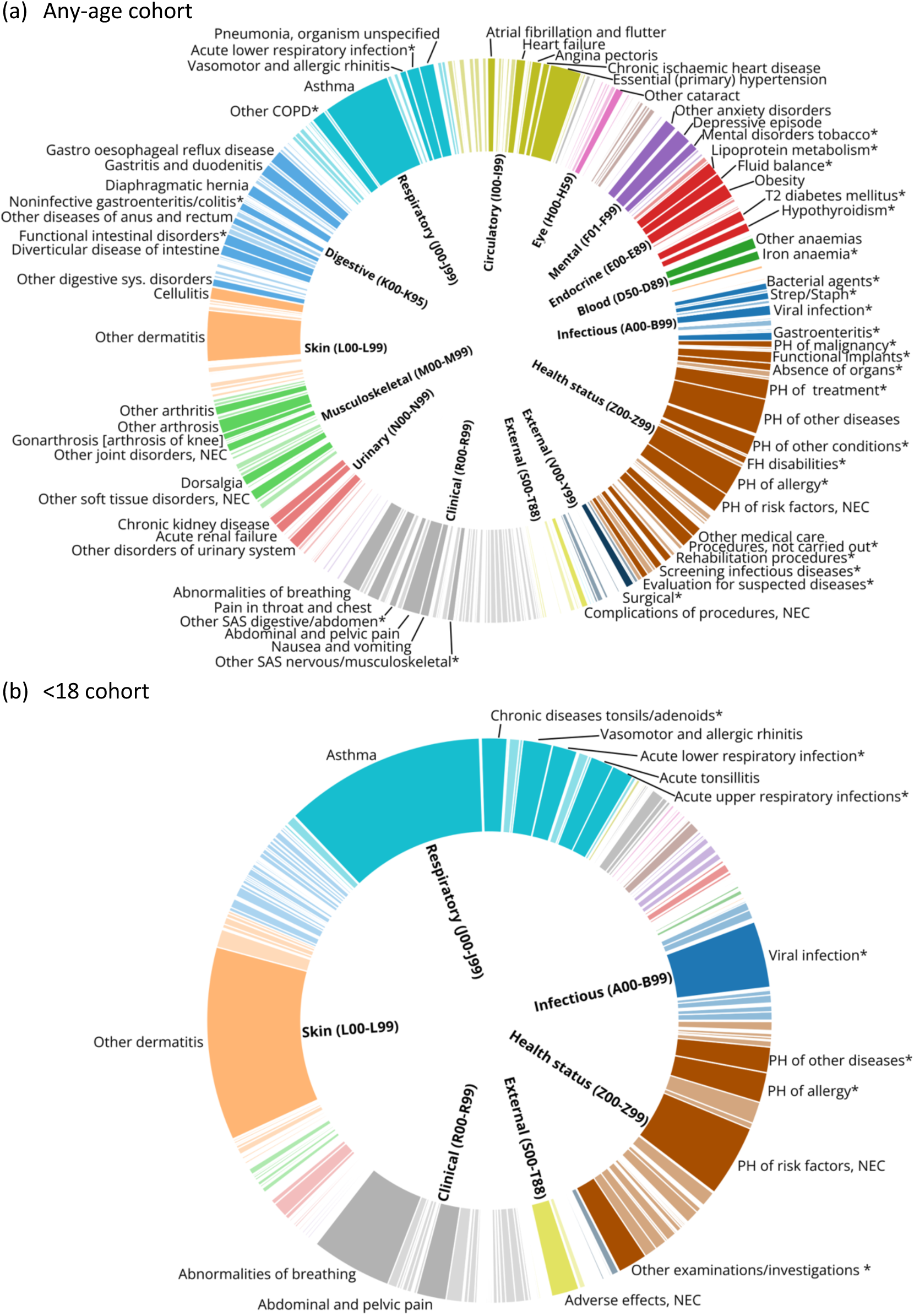
Total sum of excess rates. Excess rate (segment width) as the proportion of the sum of all excess rates (width of bars) from crude results from (a) the any-age cohort, (b) the <18 cohort. Each outcome is defined as a 3-character ICD-10 code and its descendants. Labels are only shown for outcomes with the largest individual excess rate estimates that cumulatively make up 50% of the total excess rate. Outcomes with an excess rate in the control group rather than the eczema group are not shown. Outcome labels are abbreviated. Abbreviations: PH = Personal history, SAS = signs and symptoms, OAU = Other and unspecified, NEC = not elsewhere classified

When excluding chapters L (skin), Q (congenital conditions), and R-Z (symptoms and other health related occurrences), diagnoses in Chapters J (respiratory) and A-B (infections) accounted for more than half of the total excess burden in the <18 cohort. In the any-age cohort, the total excess disease burden was more evenly distributed across chapters. The largest rate differences in the any-age cohort were for J45 Asthma (4.41), I10 Essential hypertension (2.05), F32 Depressive episode (1.05), and J18 Pneumonia, organism unspecified (0.97). For the <18 cohort, it was J45 Asthma (4.13), B34 Viral infection of unspecified site (1.36), J30 Vasomotor and allergic rhinitis (0.60), J35 Chronic diseases of tonsils and adenoids (0.53), and J22 Unspecified acute lower respiratory infection (0.51) (**Figure 4**; **Supplementary Table 2**).

**Figure 4:**
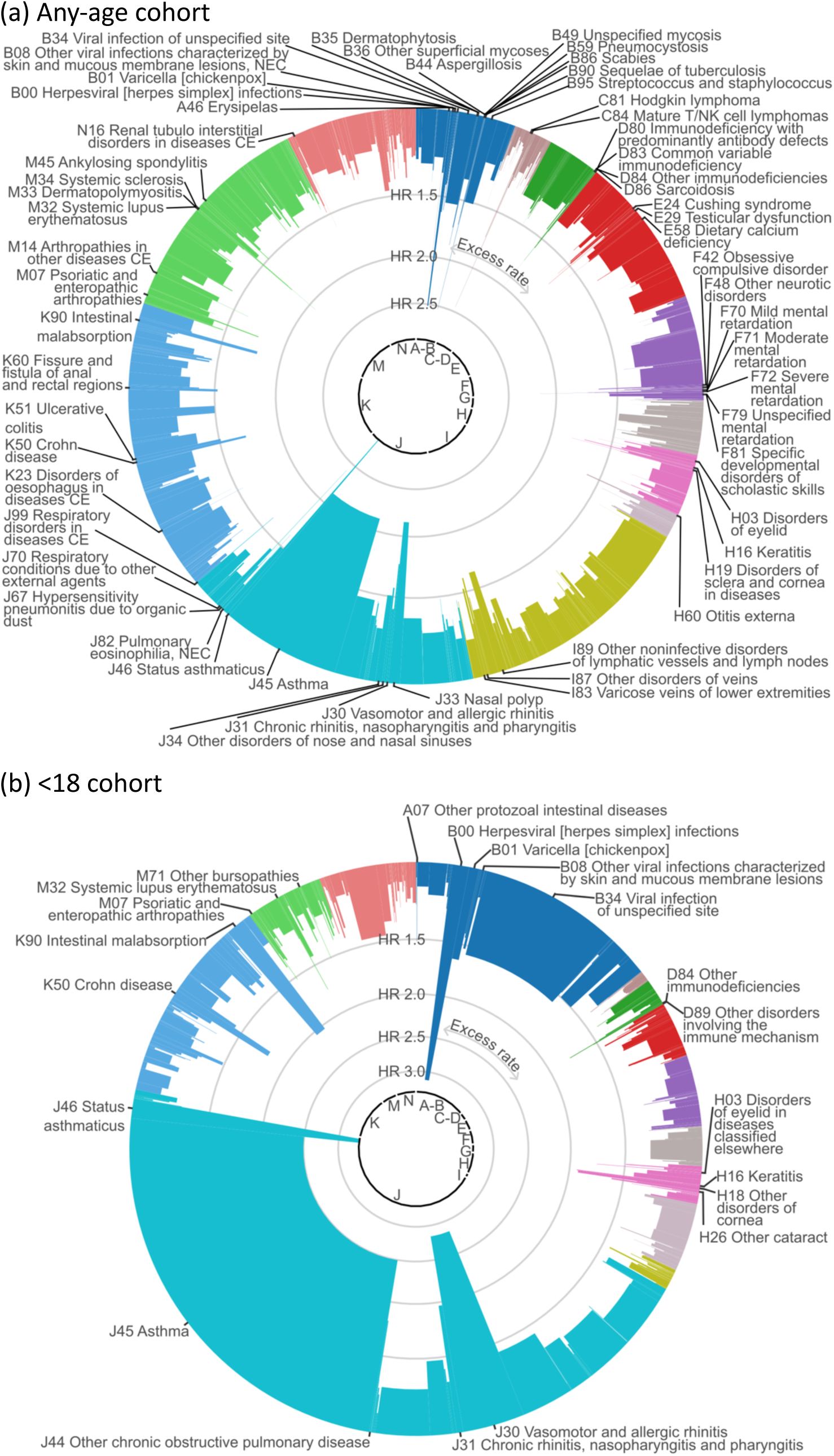
Hazard ratios and excess rate for all outcomes (excluding chapters L, Q-Z) Hazard ratios (height of bars) and excess rate as the proportion of the sum of all excess rates (width of bars) from crude results from (a) the any-age cohort, (b) the <18 cohort. Excludes chapters L & Q-Z. Labels are shown if HR > 1.5. Outcomes with an excess rate in the control group rather than the eczema group are not shown.

##### Hazard ratios

Excluding Chapters L and Q-Z and outcomes with <1,000 events in the eczema group, in the any-age cohort hazard ratios were largest for J46 Status asthmaticus (2.95 [2.79-3.13]), B00 Herpesviral [herpes simplex] infections (2.52 [2.38-2.67]), J30 Vasomotor and allergic rhinitis (2.14 [2.08-2.20]), J45 Asthma (2.12 [2.11-2.14]), M32 Systemic lupus erythematosus (1.79 [1.65-1.95]), K90 Intestinal malabsorption (1.78 [1.72-1.85]), M07 Psoriatic and enteropathic arthropathies (1.75 [1.64-1.87]), J31 Chronic rhinitis, nasopharyngitis and pharyngitis (1.75 [1.66-1.86]), B35 Dermatophytosis (1.72 [1.58-1.86]), and H16 Keratitis (1.71 [1.57-1.86]). In **Figure 4** we show hazard ratios alongside excess rate. For each chapter, the 5 outcomes with the largest crude HRs are shown in **Figure 5**. Crude and adjusted HRs for all outcomes are shown in **Supplementary Table 2-3**.

**Figure 5:**
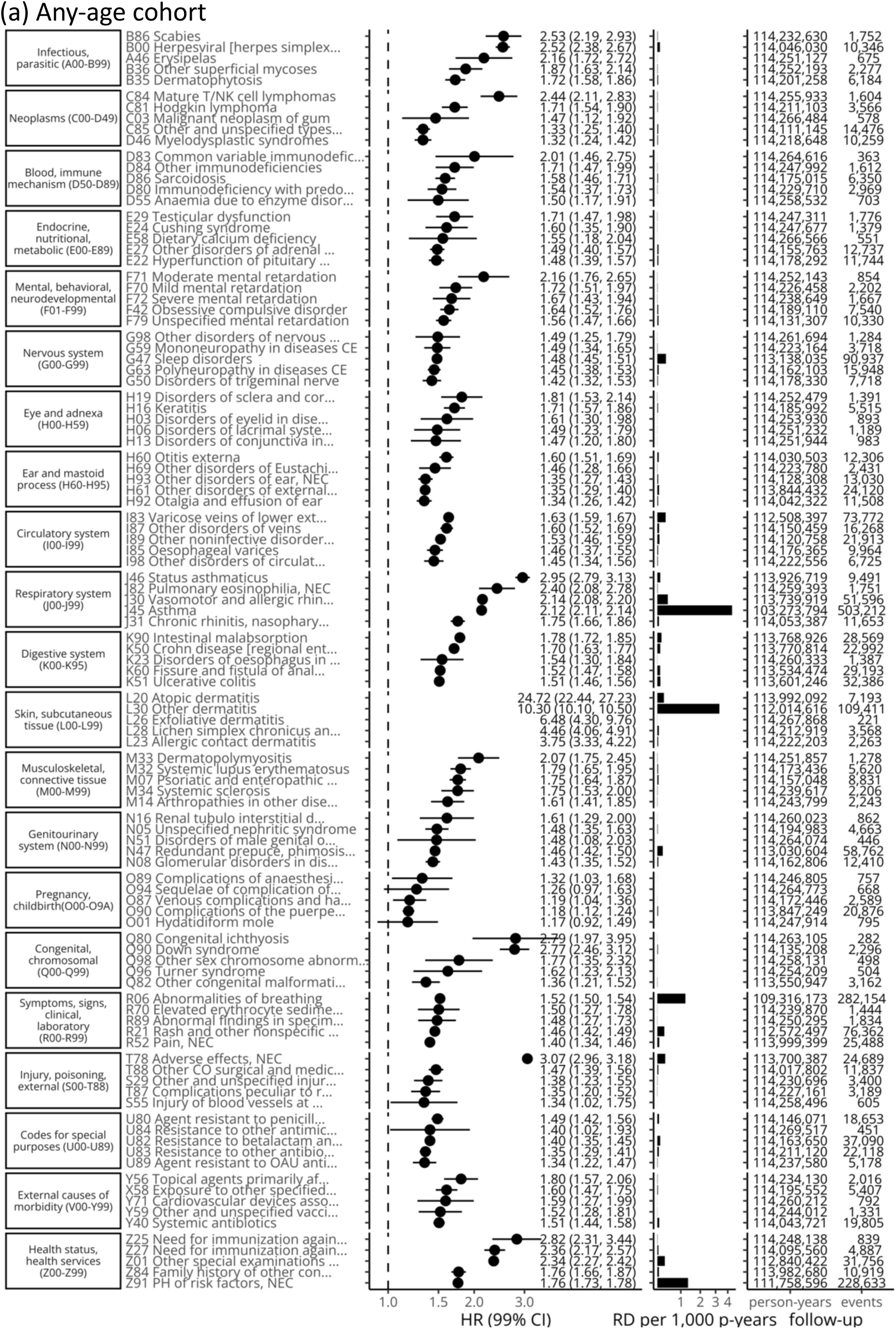
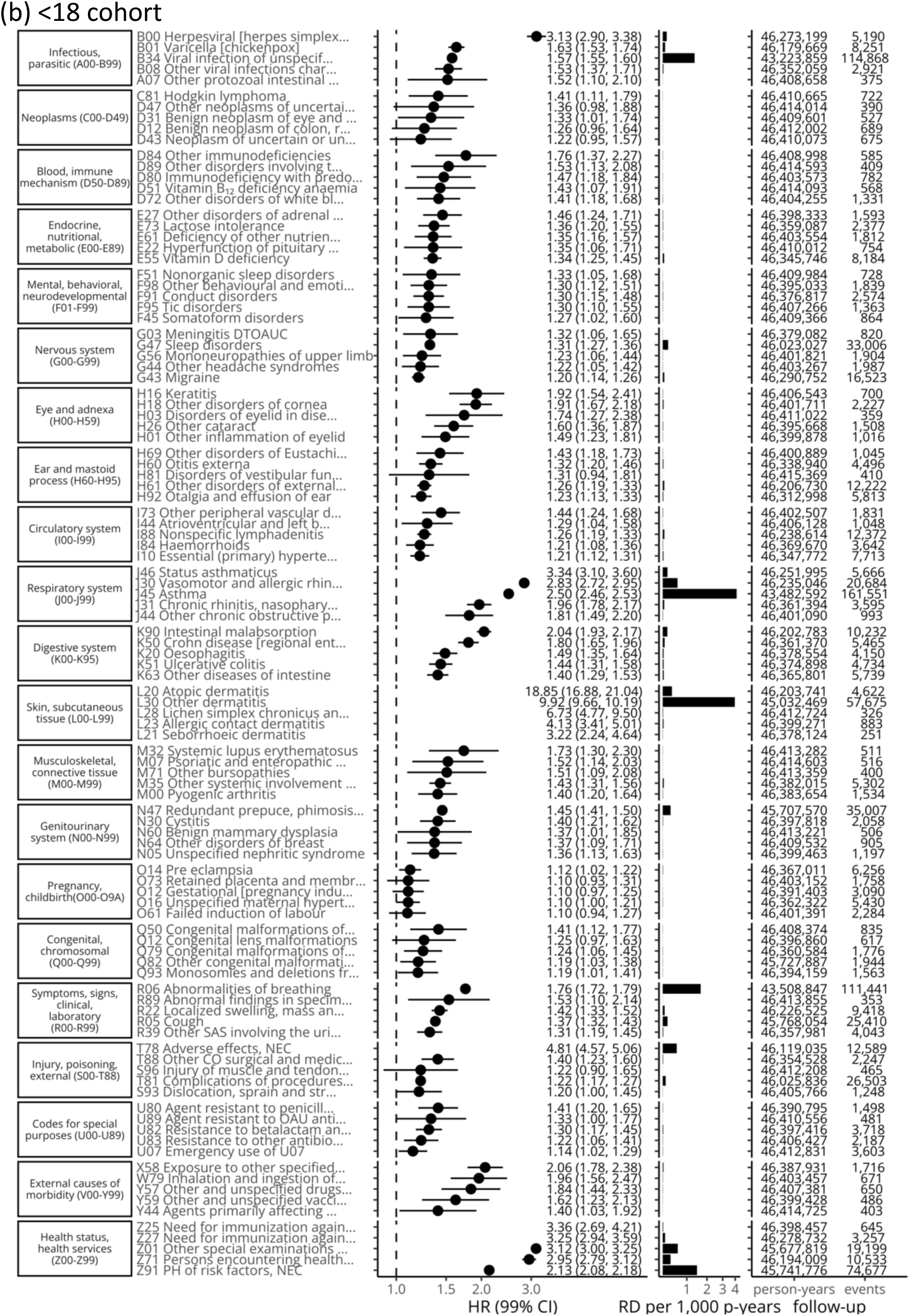
Five strongest associations between eczema and outcomes for each ICD-10 chapter (with >100 events) Hazard ratios (HR) with 99% confidence intervals (99%CI) (represented by vertical bars) from Cox regression and crude rate difference per 1,000 person-years (RD per 1,000 p-years) from (a) the any-age cohort, (b) the <18 cohort.

Hazard ratios from adjusted models are shown in **Supplementary Table 2-3**. There were substantial differences between crude and adjusted hazard ratios for some outcomes (e.g., asthma: crude HR 1.78 [1.74-1.82], adjusted HR 1.51 [1.46-1.56], change −15.12%) but not for others (e.g., essential [primary] hypertension: crude HR 1.14 [1.12-1.16], adjusted HR 1.14 [1.12-1.16], change −0.06%)

#### Sensitivity analyses

In **Supplementary Tables 3** we provide results from all age cohorts. In **Supplementary Tables 3-4** we provide results from sensitivity analyses (the any-age cohort including only individuals that had been hospitalised at least one year before index date, and from all cohorts excluding non-consulters). In both sensitivity analyses, hazard ratios were generally attenuated.

Mappings to phecodes and GBDcodes

In **Figure 6** and **Supplementary Table 5** we show results for outcomes mapped to 201 Global Burden of Disease (GBD) codes, which are ordered hierarchically (e.g., A. Malignant neoplasms: HR 1.09 [1.08-1.10], 108,133 events; 22. Lymphomas, multiple myeloma: HR 1.29 [1.15-1.34], 7,749 events; and a. Hodgkin lymphoma: HR 1.71 [1.54-1.90], 969 events). In **Figure 7** and **Supplementary Table 6** we show results mapped to 1,593 phecodes, which have a different hierarchical structure (e.g., 201 Hodgkin’s disease: HR 1.71 [1.54-1.9], 969 events; 202 Cancer of other lymphoid, histiocytic tissue: HR 1.33 [1.26-1.4], 3,821 events; 202.2 Non-Hodgkins lymphoma: HR 1.32 [1.27-1.38], 4,824 events; 202.21 Nodular lymphoma: HR 1.16 [1.04-1.29], 790 events; 202.24 Large cell lymphoma: HR 1.31 [1.22-1.42], 1,690 events). In **Table 3** we show the 6 strongest associations per phecode category.

**Figure 6:**
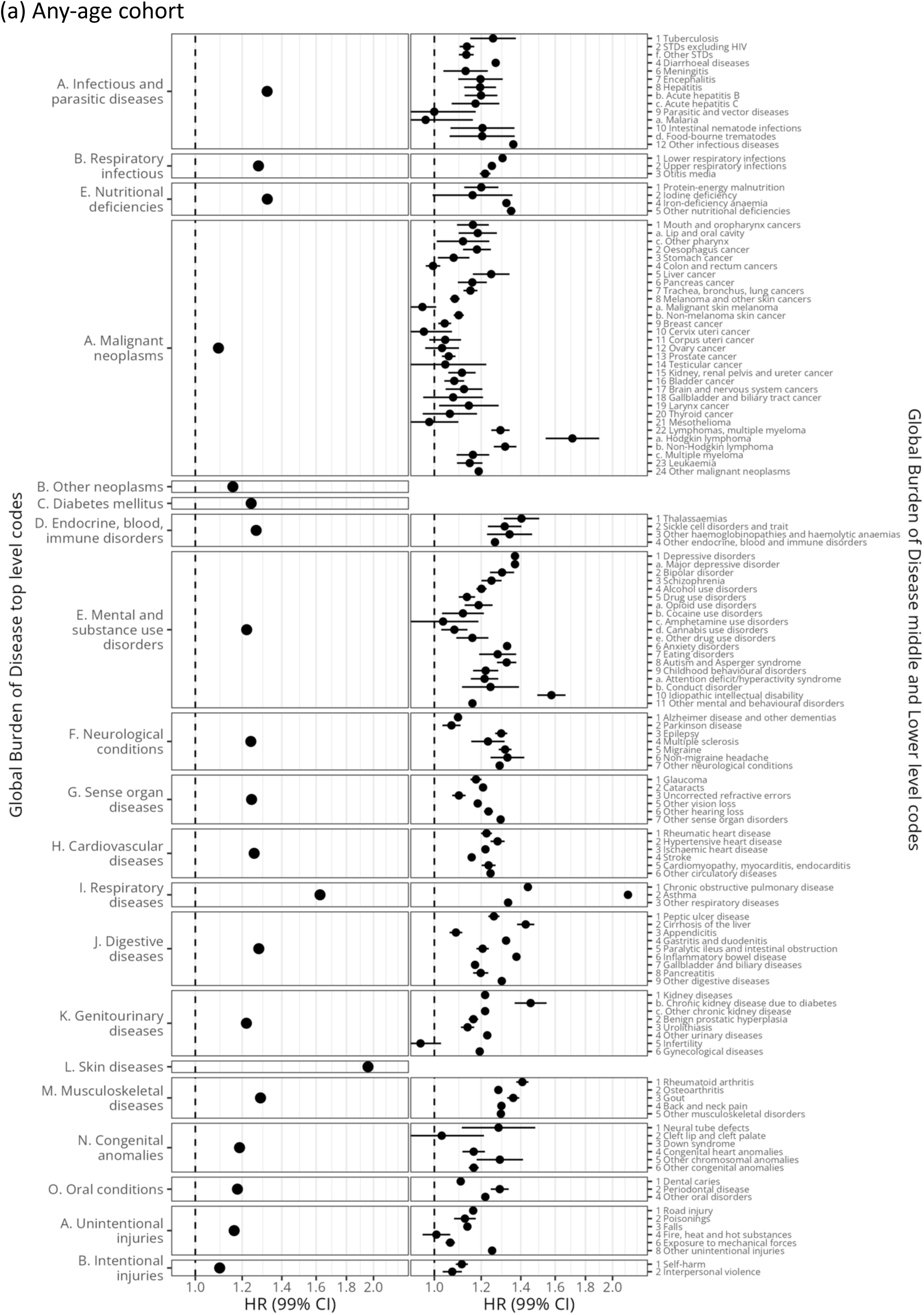
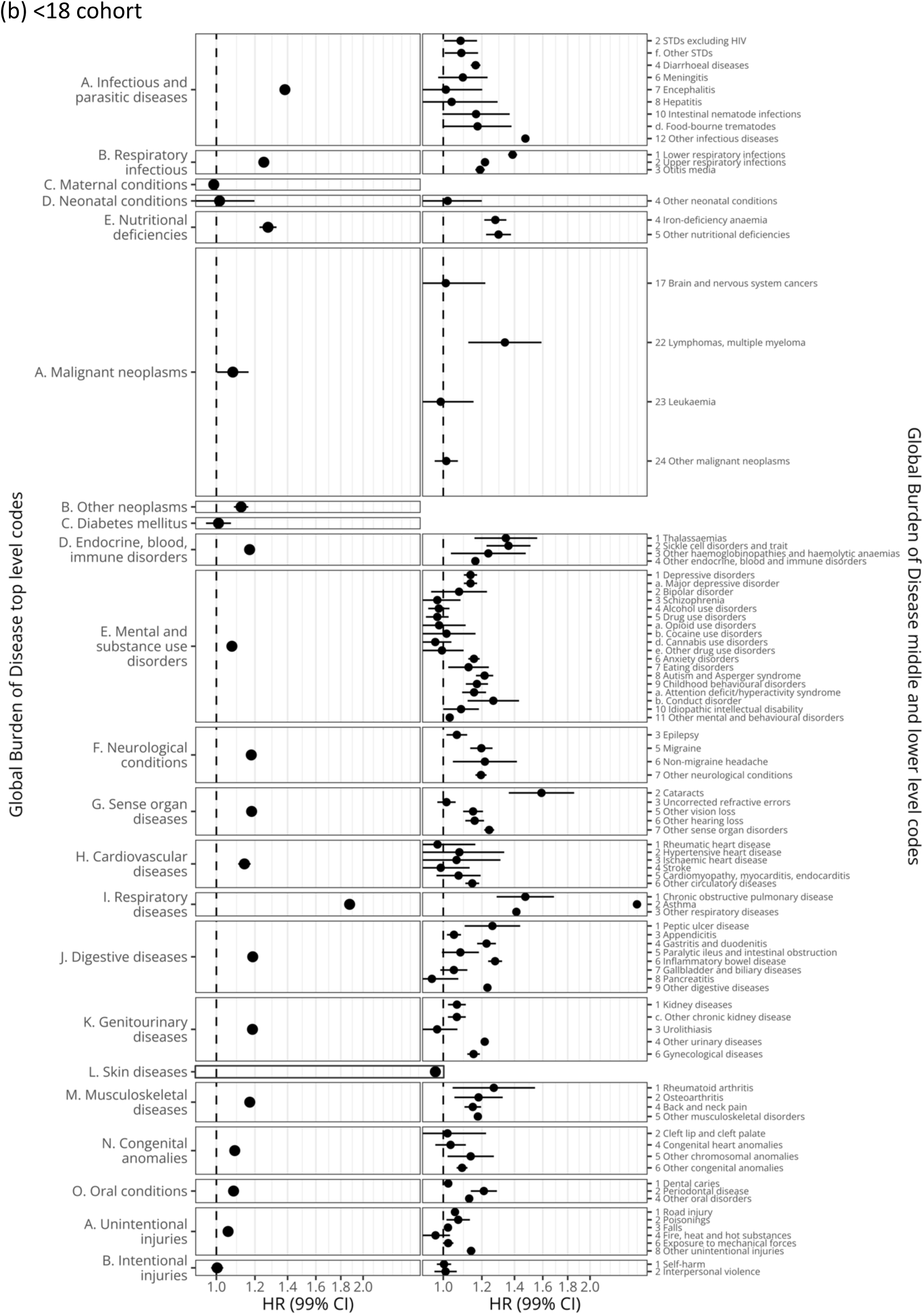
Global Burden of Disease code-mapped results. Hazard ratios (HR) with 99% confidence intervals (99%CI) (represented by vertical bars) from Cox regression from (a) the any-age cohort, (b) the <18 cohort. Excludes the following outcomes with 200 or fewer events in the exposed in the any-age cohort: Infectious and parasitic diseases (a. Syphilis, b. Chlamydia, c. Gonorrhoea, d. Trichomoniasis, e. Genital herpes, 3 HIV/AIDS, b. HIV resulting in other diseases, 5 Childhood-cluster diseases, a. Whooping cough, b. Diphtheria, c. Measles, d. Tetanus, a. Acute hepatitis A, d. Acute hepatitis E, b. Trypanosomiasis, c. Chagas disease, d. Schistosomiasis, e. Leishmaniasis, f. Lymphatic filariasis, g. Onchocerciasis, h. Cysticercosis, i. Echinococcosis, k. Trachoma, l. Yellow fever, m. Rabies, a. Ascariasis, b. Trichuriasis, c. Hookworm disease, 11 Leprosy), Neonatal conditions (1 Preterm birth complications, 2 Birth asphyxia and birth trauma, 3 Neonatal sepsis and infections), Nutritional deficiencies (3 Vitamin A deficiency), Malignant neoplasms (b. Nasopharynx), Genitourinary diseases (a. Acute glomerulonephritis), Sudden infant death syndrome (P. Sudden infant death syndrome), Unintentional injuries (5 Drowning, 7 Natural disasters), Intentional injuries (3 Collective violence and legal intervention). Outcomes that are included in (a) but not (b) have 200 or fewer events in the <18 cohort.

**Figure 7:**
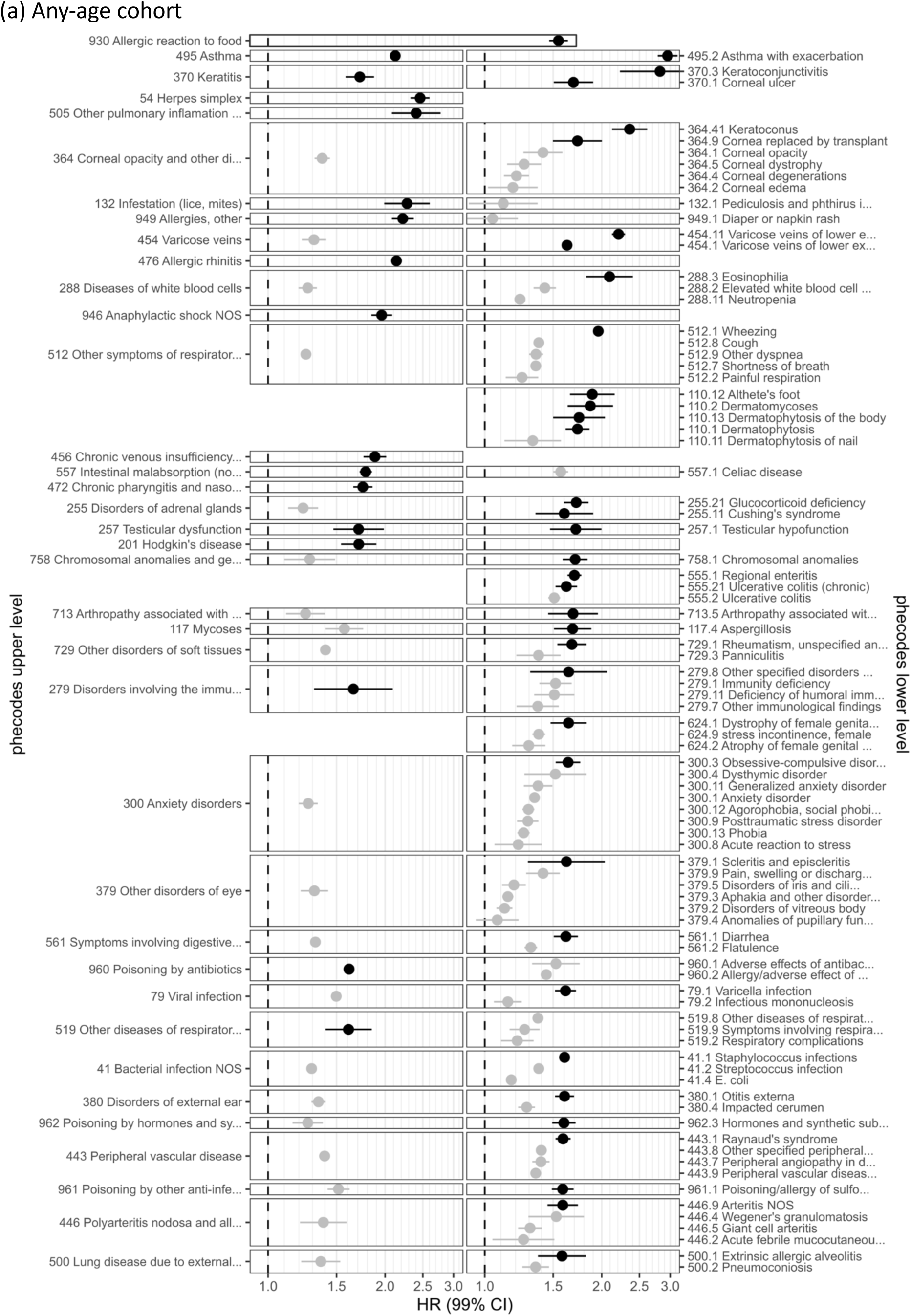
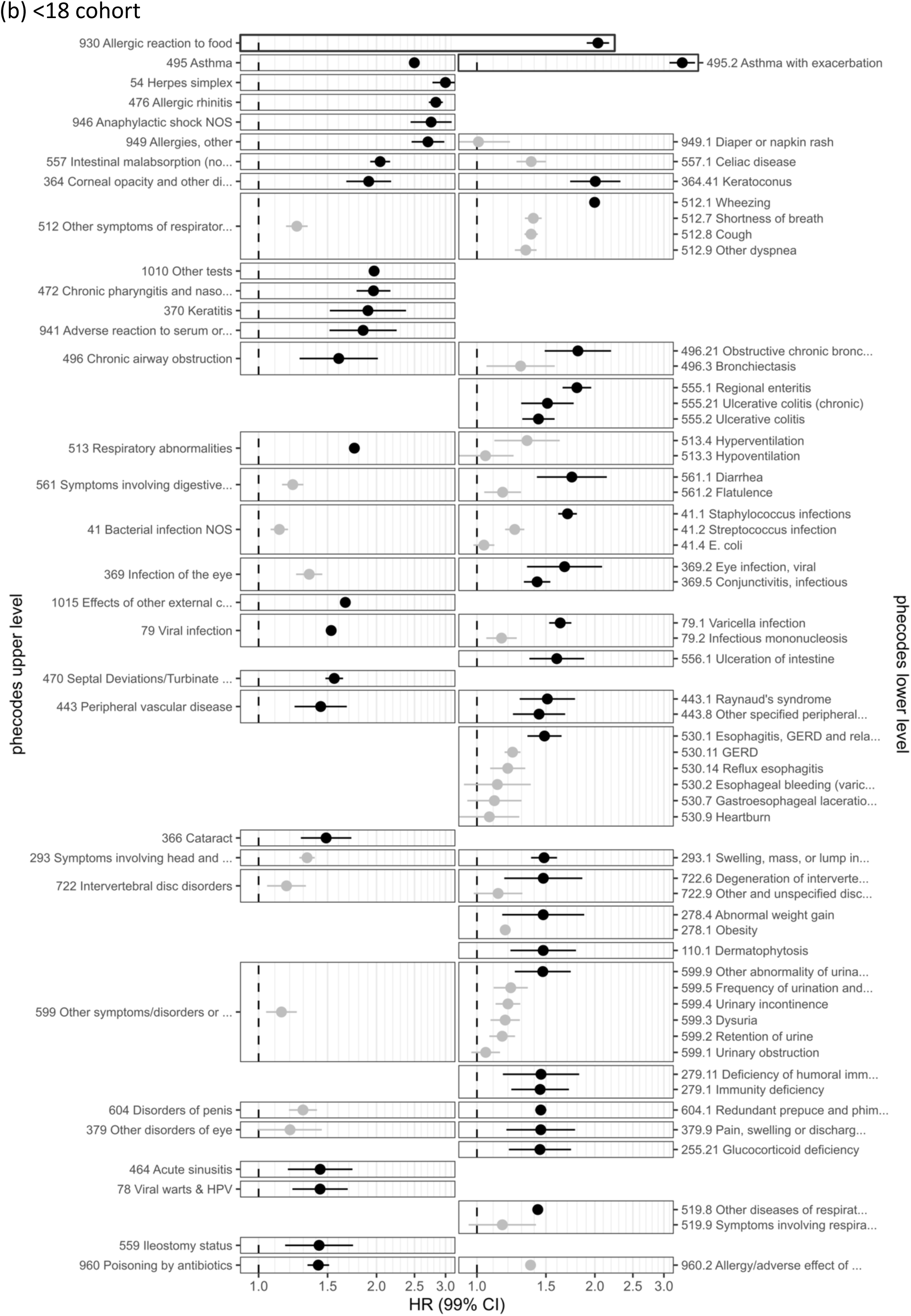
phecode-mapped results (40 largest HRs and related codes) Hazard ratios (HR) with 99% confidence intervals (99%CI) (represented by vertical bars) from Cox regression from (a) the any-age cohort, (b) the <18 cohort. The 40 phecodes with the largest hazard ratios are shown (in black), together with all codes that have the same parent code. Excludes outcomes with 200 or fewer events in the exposed. (a) Truncated phecode labels: 132.1 Pediculosis and phthirus infestation, 279 Disorders involving the immune mechanism, 279.11 Deficiency of humoral immunity, 279.8 Other specified disorders involving the immune mechanism, 288.2 Elevated white blood cell count, 300.12 Agorophobia, social phobia, and panic disorder, 300.3 Obsessive-compulsive disorders, 364 Corneal opacity and other disorders of cornea, 379.3 Aphakia and other disorders of lens, 379.4 Anomalies of pupillary function, 379.5 Disorders of iris and ciliary body, 379.9 Pain, swelling or discharge of eye, 443.7 Peripheral angiopathy in diseases classified elsewhere, 443.8 Other specified peripheral vascular diseases, 443.9 Peripheral vascular disease, unspecified, 446 Polyarteritis nodosa and allied conditions, 446.2 Acute febrile mucocutaneous lymph node syndrome (Kawasaki disease), 454.1 Varicose veins of lower extremity, 454.11 Varicose veins of lower extremity, symptomtic, 456 Chronic venous insufficiency [CVI], 472 Chronic pharyngitis and nasopharyngitis, 500 Lung disease due to external agents, 505 Other pulmonary inflamation or edema, 512 Other symptoms of respiratory system, 519 Other diseases of respiratory system, not elsewhere classified, 519.8 Other diseases of respiratory system, NEC, 519.9 Symptoms involving respiratory system and other chest symptoms, 557 Intestinal malabsorption (non-celiac), 561 Symptoms involving digestive system, 624.1 Dystrophy of female genital tract, 624.2 Atrophy of female genital tract, 713 Arthropathy associated with other disorders classified elsewhere, 713.5 Arthropathy associated with neurological disorders, 729.1 Rheumatism, unspecified and fibrositis, 758 Chromosomal anomalies and genetic disorders, 960.1 Adverse effects of antibacterials (not penicillins), 960.2 Allergy/adverse effect of penicillin, 961 Poisoning by other anti-infectives, 961.1 Poisoning/allergy of sulfonamides, 962 Poisoning by hormones and synthetic substitutes, 962.3 Hormones and synthetic substitutes causing adverse effects in therapeutic use. (b) Truncated phecode labels: 1015 Effects of other external causes, 279.11 Deficiency of humoral immunity, 293 Symptoms involving head and neck, 293.1 Swelling, mass, or lump in head and neck [Space-occupying lesion, intracranial NOS], 364 Corneal opacity and other disorders of cornea, 379.9 Pain, swelling or discharge of eye, 443.8 Other specified peripheral vascular diseases, 470 Septal Deviations/Turbinate Hypertrophy, 472 Chronic pharyngitis and nasopharyngitis, 496.21 Obstructive chronic bronchitis, 512 Other symptoms of respiratory system, 519.8 Other diseases of respiratory system, NEC, 519.9 Symptoms involving respiratory system and other chest symptoms, 530.1 Esophagitis, GERD and related diseases, 530.2 Esophageal bleeding (varices/hemorrhage), 530.7 Gastroesophageal laceration-hemorrhage syndrome, 557 Intestinal malabsorption (non-celiac), 561 Symptoms involving digestive system, 599 Other symptoms/disorders or the urinary system, 599.5 Frequency of urination and polyuria, 599.9 Other abnormality of urination, 604.1 Redundant prepuce and phimosis/BXO, 722.6 Degeneration of intervertebral disc, 722.9 Other and unspecified disc disorder, 941 Adverse reaction to serum or vaccine, 960.2 Allergy/adverse effect of penicillin.

**Table 3:** Phecode-mapped results (6 strongest associations per category, >500 events) Crude hazard ratios (HR) with 99% confidence intervals (99%CI) from Cox regression and number of events from the any-age cohort. Cells are shaded by strength of association from 1 and below (white) to 2 and above (black).

| phecode | phenotype | crude HR(99%CI) | events |
| --- | --- | --- | --- |
| circulatory system |  |  |  |
| 454.11 | Varicose veins of lower ext... | 2.21 [2.12-2.3] | 8,205 |
| 456 | Chronic venous insufficienc... | 1.88 [1.76-2.01] | 2,673 |
| 454.1 | Varicose veins of lower ext... | 1.63 [1.59-1.67] | 19,256 |
| 443.1 | Raynaud's syndrome | 1.59 [1.52-1.66] | 5,376 |
| 446.9 | Arteritis NOS | 1.59 [1.45-1.74] | 1,278 |
| 450 | Noninfectious disorders of ... | 1.52 [1.46-1.59] | 5,602 |
| congenital anomalies |  |  |  |
| 758.1 | Chromosomal anomalies | 1.71 [1.59-1.84] | 2,051 |
| 756.3 | Congenital anomalies of mus... | 1.43 [1.28-1.61] | 797 |
| 757 | Congenital anomalies of the... | 1.36 [1.22-1.51] | 857 |
| 752.11 | Spina bifida | 1.3 [1.17-1.44] | 964 |
| 756.5 | Congenital osteodystrophies | 1.27 [1.18-1.37] | 1,865 |
| 755.1 | Congenital deformities of feet | 1.26 [1.15-1.37] | 1,329 |
| dermatologic |  |  |  |
| 690 | Erythematousquamous dermatosis | 27.4 [23.4-32.2] | 2,347 |
| 939 | Atopic/contact dermatitis d... | 7.81 [7.68-7.95] | 84,723 |
| 690.1 | Seborrheic dermatitis | 3.29 [2.87-3.77] | 776 |
| 695.7 | Prurigo and Lichen | 3 [2.81-3.19] | 3,243 |
| 695.22 | Pemphigus and pemphigoid | 2.81 [2.55-3.09] | 1,545 |
| 696.41 | Psoriasis vulgaris | 2.3 [2.23-2.37] | 12,400 |
| digestive |  |  |  |
| 557 | Intestinal malabsorption (n... | 1.78 [1.72-1.85] | 8,443 |
| 555.1 | Regional enteritis | 1.7 [1.63-1.77] | 6,324 |
| 555.21 | Ulcerative colitis (chronic) | 1.62 [1.52-1.73] | 2,605 |
| 561.1 | Diarrhea | 1.62 [1.51-1.74] | 2,038 |
| 557.1 | Celiac disease | 1.57 [1.5-1.64] | 5,403 |
| 556.1 | Ulceration of intestine | 1.54 [1.45-1.63] | 2,973 |
| endocrine/metabolic |  |  |  |
| 255.21 | Glucocorticoid deficiency | 1.72 [1.6-1.85] | 2,070 |
| 250.14 | Type 1 diabetes with neurol... | 1.52 [1.36-1.71] | 762 |
| 279.1 | Immunity deficiency | 1.52 [1.38-1.67] | 1,161 |
| 253.7 | Other disorders of neurohyp... | 1.52 [1.42-1.63] | 2,343 |
| 279.11 | Deficiency of humoral immunity | 1.51 [1.34-1.7] | 744 |
| 250.12 | Type 1 diabetes with renal ... | 1.51 [1.32-1.73] | 527 |
| genitourinary |  |  |  |
| 624.1 | Dystrophy of female genital... | 1.64 [1.48-1.83] | 927 |
| 580.12 | Non-proliferative glomerulo... | 1.54 [1.35-1.75] | 595 |
| 580.32 | Nephritis and nephropathy w... | 1.47 [1.36-1.59] | 1,569 |
| 604.1 | Redundant prepuce and phimo... | 1.46 [1.42-1.5] | 16,264 |
| 601.4 | Balanoposthitis | 1.45 [1.35-1.56] | 1,874 |
| 592.13 | Chronic interstitial cystitis | 1.45 [1.28-1.64] | 657 |
| hematopoietic |  |  |  |
| 288.3 | Eosinophilia | 2.09 [1.82-2.4] | 636 |
| 283.1 | Autoimmune hemolytic anemias | 1.57 [1.36-1.81] | 517 |
| 281.9 | Deficiency anemias | 1.48 [1.32-1.66] | 835 |
| 283 | Acquired hemolytic anemias | 1.44 [1.28-1.62] | 745 |
| 288.2 | Elevated white blood cell c... | 1.43 [1.34-1.53] | 2,421 |
| 285.2 | Anemia of chronic disease | 1.43 [1.34-1.52] | 2,731 |
| infectious diseases |  |  |  |
| 54 | Herpes simplex | 2.46 [2.33-2.6] | 3,948 |
| 132 | Infestation (lice, mites) | 2.28 [1.99-2.61] | 669 |
| 110.12 | Althete's foot | 1.89 [1.66-2.16] | 648 |
| 110.2 | Dermatomycoses | 1.87 [1.63-2.14] | 651 |
| 110.1 | Dermatophytosis | 1.73 [1.61-1.86] | 2,264 |
| 117.4 | Aspergillosis | 1.68 [1.51-1.88] | 857 |
| injuries & poisonings |  |  |  |
| 930 | Allergic reaction to food | 5.58 [5.28-5.9] | 6,437 |
| 949 | Allergies, other | 2.22 [2.08-2.37] | 2,834 |
| 946 | Anaphylactic shock NOS | 1.96 [1.84-2.08] | 2,962 |
| 960 | Poisoning by antibiotics | 1.62 [1.58-1.65] | 24,400 |
| 962.3 | Hormones and synthetic subs... | 1.6 [1.49-1.71] | 2,226 |
| 961.1 | Poisoning/allergy of sulfon... | 1.59 [1.49-1.69] | 2,515 |
| mental disorders |  |  |  |
| 300.3 | Obsessive-compulsive disorders | 1.64 [1.52-1.76] | 2,087 |
| 315.3 | Mental retardation | 1.58 [1.49-1.67] | 3,630 |
| 315.1 | Learning disorder | 1.56 [1.5-1.62] | 8,341 |
| 302 | Sexual and gender identity ... | 1.49 [1.33-1.68] | 707 |
| 317.11 | Alcoholic liver damage | 1.44 [1.38-1.51] | 5,266 |
| 303.4 | Somatoform disorder | 1.44 [1.34-1.55] | 1,793 |
| musculoskeletal |  |  |  |
| 729.1 | Rheumatism, unspecified and... | 1.68 [1.54-1.83] | 1,338 |
| 726.3 | Bursitis | 1.51 [1.41-1.62] | 2,124 |
| 715.2 | Ankylosing spondylitis | 1.5 [1.39-1.62] | 1,810 |
| 711.1 | Pyogenic arthritis | 1.48 [1.39-1.59] | 2,261 |
| 716 | Other arthropathies | 1.48 [1.31-1.68] | 648 |
| 714 | Rheumatoid arthritis and ot... | 1.46 [1.36-1.56] | 2,226 |
| neoplasms |  |  |  |
| 201 | Hodgkin's disease | 1.71 [1.54-1.9] | 969 |
| 217.1 | Nevus, non-neoplastic | 1.34 [1.24-1.45] | 1,597 |
| 202 | Cancer of other lymphoid, h... | 1.33 [1.26-1.4] | 3,821 |
| 202.2 | Non-Hodgkins lymphoma | 1.32 [1.27-1.38] | 4,824 |
| 204.21 | Myeloid leukemia, acute | 1.32 [1.21-1.44] | 1,349 |
| 202.24 | Large cell lymphoma | 1.31 [1.22-1.42] | 1,690 |
| neurological |  |  |  |
| 327.3 | Sleep apnea | 1.51 [1.48-1.55] | 20,679 |
| 327 | Sleep disorders | 1.48 [1.45-1.51] | 23,995 |
| 338.2 | Chronic pain | 1.46 [1.37-1.56] | 2,496 |
| 345.12 | Partial epilepsy | 1.46 [1.33-1.59] | 1,394 |
| 352.1 | Trigeminal nerve disorders ... | 1.42 [1.32-1.53] | 1,837 |
| 338.1 | Acute pain | 1.4 [1.3-1.51] | 1,800 |
| pregnancy complications |  |  |  |
| 649 | Other conditions or status ... | 1.21 [1.17-1.25] | 9,473 |
| 668 | Complications of the admini... | 1.2 [1.06-1.37] | 626 |
| 674 | Other complications of the ... | 1.19 [1.14-1.25] | 5,402 |
| 656 | Other perinatal conditions ... | 1.18 [1.14-1.22] | 9,997 |
| 646 | Other complications of preg... | 1.18 [1.16-1.2] | 31,167 |
| 636 | Early or threatened labor; ... | 1.16 [1.12-1.21] | 6,347 |
| respiratory |  |  |  |
| 495.2 | Asthma with exacerbation | 2.95 [2.79-3.13] | 3,968 |
| 505 | Other pulmonary inflammation... | 2.4 [2.08-2.78] | 600 |
| 476 | Allergic rhinitis | 2.14 [2.08-2.2] | 17,750 |
| 495 | Asthma | 2.12 [2.11-2.14] | 165,066 |
| 512.1 | Wheezing | 1.96 [1.92-2] | 29,127 |
| 472 | Chronic pharyngitis and nas... | 1.75 [1.66-1.86] | 3,383 |
| sense organs |  |  |  |
| 364.41 | Keratoconus | 2.36 [2.13-2.62] | 1,203 |
| 364.9 | Cornea replaced by transplant | 1.73 [1.5-2] | 507 |
| 370 | Keratitis | 1.72 [1.58-1.87] | 1,569 |
| 370.1 | Corneal ulcer | 1.69 [1.51-1.9] | 802 |
| 380.1 | Otitis externa | 1.6 [1.52-1.7] | 3,397 |
| 369.2 | Eye infection, viral | 1.54 [1.39-1.71] | 985 |
| symptoms |  |  |  |
| 798.1 | Chronic fatigue syndrome | 1.49 [1.4-1.6] | 2,283 |
| 771.1 | Swelling of limb | 1.4 [1.37-1.43] | 21,683 |
| 770 | Myalgia and myositis unspec... | 1.37 [1.3-1.44] | 4,383 |
| 773 | Pain in limb | 1.37 [1.34-1.4] | 25,521 |
| 771 | Musculoskeletal symptoms re... | 1.36 [1.28-1.44] | 2,936 |
| 761 | Cervicalgia | 1.34 [1.3-1.39] | 7,994 |

Comparison with results with primary care outcomes

We previously assessed associations with 71 outcomes recorded in primary care in the same cohort (without requiring eligibility for HES linkage). In **Figure 8** we compare our results against these previous results (for the 69 outcomes we were able to identify a corresponding ICD-10 code or phecode), using the same adjustment set. Results were generally similar (previous vs this study e.g., food allergy HR [99%CI] 4.03 [3.95-4.11] vs 930 Allergic reaction to food 4.37 [4.11-4.63]; Diverticular disease 1.17 [1.16-1.18] vs K57 Diverticular disease of intestine 1.17 [1.16-1.19]; Prostate cancer 1.01 [0.99-1.03] vs C61 Malignant neoplasm of prostate 1.04 [1.01-1.06]).

**Figure 8:**
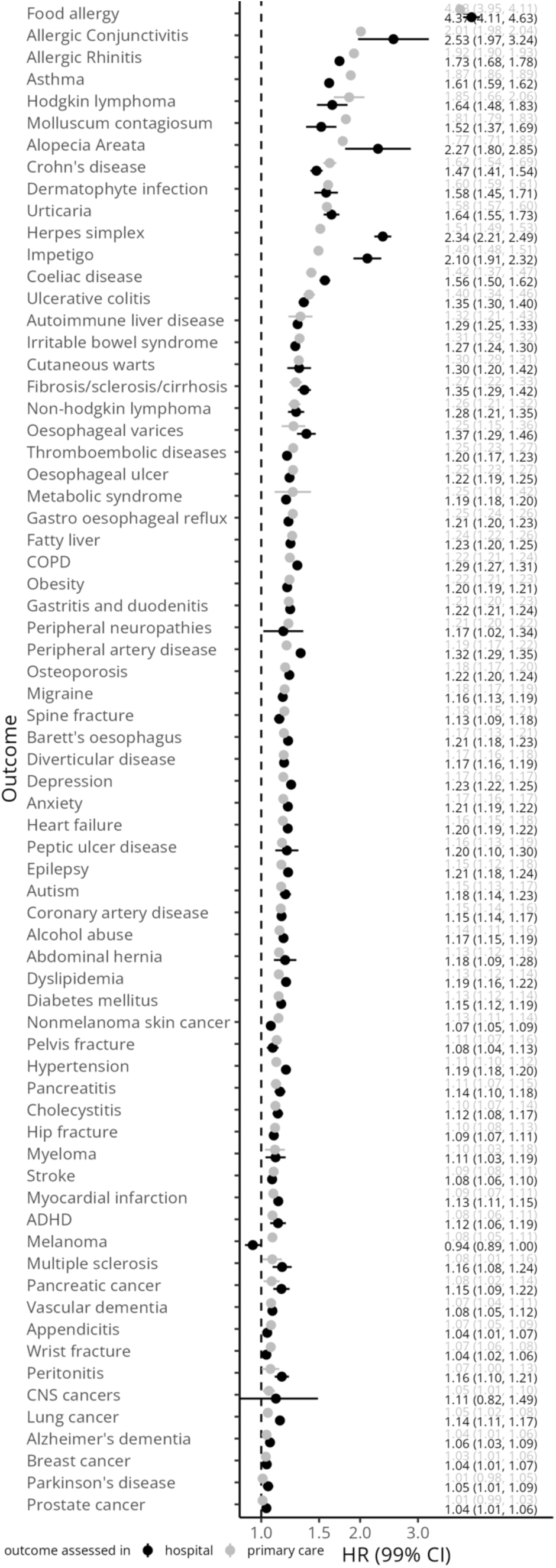
Comparison with previous study with primary-care outcomes. Hazard ratios from “Cohort studies on 71 outcomes among people with atopic eczema in UK primary care data” ranked by main adjusted hazard ratio from the any-age cohort, compared to corresponding outcomes defined either as category-level ICD-10 code or phecode and their ranking by main adjusted hazard ratio from the any-age cohort. No corresponding outcomes were available for eosinophilic oesophagitis and cigarette smoking. Food allergy=930 Allergic reaction to food; Allergic Conjunctivitis=370.3 Keratoconjunctivitis; Allergic Rhinitis=476 Allergic rhinitis; Asthma=J45 Asthma; Hodgkin lymphoma=C81 Hodgkin lymphoma; Molluscum contagiosum=B08 Other viral infections chara…; Alopecia Areata=L63 Alopecia areata; Crohn’s disease=K50 Crohn disease [regional ente…; Dermatophyte infection=B35 Dermatophytosis; Urticaria=L50 Urticaria; Herpes simplex=B00 Herpesviral [herpes simplex]…; Impetigo=L01 Impetigo; Coeliac disease=K90 Intestinal malabsorption; Ulcerative colitis=K51 Ulcerative colitis; Autoimmune liver disease=571.5 Other chronic nonalcoholic…; Irritable bowel syndrome=K58 Irritable bowel syndrome; Cutaneous warts=B07 Viral warts; Fibrosis/sclerosis/cirrhosis=K74 Fibrosis and cirrhosis of liver; Non-Hodgkin lymphoma=C85 Other and unspecified types …; Oesophageal varices=I85 Oesophageal varices; Thromboembolic diseases=I80 Phlebitis and thrombophlebitis; Oesophageal ulcer=K20 Oesophagitis; Metabolic syndrome=E11 Type 2 diabetes mellitus; Gastro oesophageal reflux=K21 Gastro oesophageal reflux di…; Fatty liver=K76 Other diseases of liver; COPD=J44 Other chronic obstructive pu…; Obesity=E66 Obesity; Gastritis and duodenitis=K29 Gastritis and duodenitis; Peripheral neuropathies=356 Hereditary and idiopathic peripheral neuropathy; Peripheral artery disease=I73 Other peripheral vascular di…; Osteoporosis=M81 Osteoporosis without patholo…; Migraine=G43 Migraine; Spine fracture=805 Fracture of vertebral column…; Barett’s oesophagus=K22 Other diseases of oesophagus; Diverticular disease=K57 Diverticular disease of inte…; Depression=296.2 Depression; Anxiety=300.1 Anxiety disorder; Heart failure=I50 Heart failure; Peptic ulcer disease=K27 Peptic ulcer, site unspecified; Epilepsy=G40 Epilepsy; Autism=313.3 Autism; Coronary artery disease=411.4 Coronary atherosclerosis; Alcohol abuse=317.1 Alcoholism; Abdominal hernia=K46 Unspecified abdominal hernia; Dyslipidemia=272.1 Hyperlipidemia; Diabetes mellitus=E10 Type 1 diabetes mellitus; Nonmelanoma skin cancer=C44 Other malignant neoplasms of…; Pelvis fracture=802 Fracture of pelvis; Hypertension=I10 Essential (primary) hyperten…; Pancreatitis=K85 Acute pancreatitis; Cholecystitis=K81 Cholecystitis; Hip fracture=S72 Fracture of femur; Myeloma=C90 Multiple myeloma and maligna…; Stroke=I63 Cerebral infarction; Myocardial infarction=I21 Acute myocardial infarction; ADHD=313.1 Attention deficit hyperact…; Melanoma=C43 Malignant melanoma of skin; Multiple sclerosis=G35 Multiple sclerosis; Pancreatic cancer=C25 Malignant neoplasm of pancreas; Vascular dementia=F01 Vascular dementia; Appendicitis=K35 Acute appendicitis; Wrist fracture=S52 Fracture of forearm; Peritonitis=K65 Peritonitis; CNS cancers=C72 Malignant neoplasm of spinal…; Lung cancer=C34 Malignant neoplasm of bronch…; Alzheimer’s dementia=G30 Alzheimer disease; Breast cancer=C50 Malignant neoplasm of breast; Parkinson’s disease=332 Parkinson’s disease; Prostate cancer=C61 Malignant neoplasm of prostate.

## Discussion

### Summary of most important findings

Besides asthma, allergies and skin infections, our results indicate that the absolute burden of other outcomes after childhood eczema was likely to be small. In contrast, adults with new or pre-existing eczema were more likely to be diagnosed with numerous conditions when admitted to hospitals. While much of this excess in outcomes may be explained by factors including misclassification of eczema, or confounding, our study identifies outcomes that may warrant further investigation.

For example, of comorbidities currently not recognised in guidelines,^1^ certain lymphomas, coeliac disease, inflammatory bowel conditions, and certain diseases of the eye demonstrated particularly strong associations with eczema. However, strength of association is only one of several results provided for each outcome in this study; we recommend considering the full range of results available for each outcome to draw conclusions (we demonstrated this for example outcomes in **Table 2**).

### Strengths and limitations

The use of routinely collected electronic health records (EHR) data comes with several strengths. Large sample size enabled follow-up of even rare outcomes. CPRD Aurum has been found to be representative of the general population of England in terms of age, sex, geographical spread and deprivation (albeit it may not necessarily be generalisable to other settings).^6^ Using primary care data to define the exposure of eczema we were likely able to avoid some of the selection biases that can occur when studying hospital-based populations.^7^

The use of ICD-10 coded hospital records allowed us to systematically and comprehensively study all hospital-recorded diseases; a previously underutilised strength of electronic health records. Ascertaining outcomes in hospital records may also make observed differences less likely to be due to ascertainment bias (e.g., people with eczema may consult their GP more frequently because of their eczema but are unlikely to be admitted to hospital more frequently).

The use of different ICD-10 mappings is another strength of this study. Firstly, the use of GBD codes gives a more compact overview across diseases, and results will be comparable to those from the Global Burden of Disease Studies. Secondly, the use of phecodes complements the use of category-level ICD-10 codes. While many diseases of interest correspond well to category-level ICD-10 codes, for some, phecodes offer a more clinically useful definition. However, the use of unmapped category-level ICD-10 codes has the advantage of giving a complete picture of all hospital records, as well as outcomes being non-overlapping.

Previous hypothesis-free studies exploring eczema outcomes employed study designs not accounting for temporality of associations and low granularity of outcomes (e.g., on ICD-10-chapter level).^8^ Our study offers high granularity of outcomes and study design that could be employed in hypothesis-testing studies. The evidence created through this approach is a more useful starting point for future studies, allowing these to focus on outcome-specific considerations.

Our results were generally consistent with those from our previous primary-care study both for serious conditions likely to be diagnosed in hospital, as well as conditions likely to be managed in primary care, suggesting these diagnoses are recorded during hospital admissions, even if they are not a primary reason for admission.

This study has limitations. Results may be biased by individuals already having the outcome of interest before it is captured in hospital records. We excluded individuals who already had a record for the outcome under analysis before index date and performed sensitivity analyses requiring previous hospital admission. These measures are however unlikely to account for all instances of individuals already having the relevant outcome before they develop eczema. For some outcomes, adjustment for comorbidities recorded in primary care at baseline can partially account for outcomes pre-dating eczema (e.g., a large attenuation is seen for asthma in comorbidity adjusted [including for asthma] results).

A limitation of this large-scale study is a lack of outcome-specific confounding adjustment strategies. While our covariate set of primary care assessed comorbidities was chosen based on the existing literature to include the most relevant comorbidities of eczema, there may still be residual confounding for any outcomes.

Information bias, including misclassification of exposure and outcome as well as their timings needs to be considered, and can vary for each individual outcome. For example, outcomes that can present with a rash might be initially misdiagnosed as eczema until they get the correct diagnosis; a likely explanation for many other skin disease outcomes including psoriasis, but also other outcomes presenting with rash such as systemic lupus erythematosus and certain lymphomas.^9^ Outcome status and timing may also be misclassified, both through factors relating to disease progression (e.g., a long lead time before diagnosis), as well as factors relating to the use of hospital admission data to capture outcomes (e.g., outcomes not requiring hospital admission only being recorded when individuals are admitted to hospital for a different reason).

### Interpretation of findings

#### Comparisons between cohorts

An important finding of our study was the stark difference between the <18 cohort and the other cohorts. The <18 cohort had a much lower proportion of significant outcomes (11% <18 vs 44% any-age) going down to 2% when also adjusting for comorbidities and excluding non-consulters. Several factors may explain these differences. Firstly, follow-up extending a maximum of 25 years may be too short to observe outcomes occurring most commonly in older age.

Secondly, the <18 cohort consists mainly of incident eczema rather than incident and prevalent eczema (as do the other cohorts), which may better account for temporality of associations. For example, despite excluding individuals with hospital records for the outcome before index date there is a strong association with Down’s syndrome (a congenital condition that is guaranteed to have occurred before eczema and is likely to increase the risk of eczema)^10^ in all cohorts except the <18 cohort where there was no association.

Thirdly, there may be increased confidence in the diagnosis of atopic eczema in childhood, as opposed to other forms of dermatitis (irritant, contact, varicose eczema, etc.). An example of this may be the very strong association with I83 “Varicose veins […]” in the any-age cohort, as opposed to no association in the <18 cohort: the any-age cohort may include individuals that have varicose eczema rather than atopic eczema. However, other outcomes that are likely to be strongly associated due to misclassified eczema remain strongly associated in the <18 cohort, including psoriasis (2.04 [1.84-2.26]) and systemic lupus erythematosus (1.73 [1.30-2.30]).

Eczema often most commonly occurs first in childhood and often improves in adulthood but can also persist or occur at older ages.^11^ All cohorts include a mix of individuals whose eczema has gotten better or even disappeared over time, as well as individuals who continue to have eczema throughout their life.

Chapters including respiratory, eye, and digestive system conditions stand out in that the outcomes with the largest HRs were larger in the <18 than the any-age cohort. While unsurprising for atopic and allergic outcomes (asthma, allergic conjunctivitis/rhinitis, allergies), other outcomes including cataract, intestinal malabsorption (which includes coeliac disease), and inflammatory bowel conditions (Crohn’s disease and ulcerative colitis) have large HRs. We highlighted inflammatory bowel conditions as outcomes of particular interest in our previous primary-care study.^3^

Generally, results from the 18+ and 40+ cohorts were similar to those from the any-age cohort, however, where differences occur, they may suggest differences in outcome risk by age. For example, atopic and allergic outcomes (food allergy, asthma, allergic rhinitis) and viral infections (herpes simplex, varicella) had much smaller hazard ratios in the 40+ cohort than the any-age cohort, which seems plausible given the usual ages of onset for these conditions.

#### Discussion of findings by chapter

The following discussion of results from all chapters references the adjusted results from the <18 cohort. Several infections were associated with eczema(A-B). A tendency towards cutaneous infections is a diagnostic feature of eczema,^12^ of which we saw herpes simplex infections more strongly associated than varicella, streptococcal and staphylococcal infections. We also found evidence for an association with herpes zoster. There was little evidence for an increased risk of several other non-skin infections (e.g., gastroenteritis, intestinal infections, candidiasis, sepsis). Cancers (C) were generally not found to be associated with eczema, except for Hodgkin lymphoma. Reverse causality may be an explanation for this finding,^9^ however previous studies have not found evidence for this hypothesis.^13,14^ Of immune-related and blood disorders (D), there was a relatively strong association for the rare outcome D84 Other immunodeficiencies. Relatively strong evidence was found for an association with iron deficiency anaemia, the most common of all outcomes in this chapter.

Of endocrine disorders (E), disorders of the adrenal and pituitary gland were associated, as were nutritional deficiencies, most clearly for the more commonly recorded Vitamin D deficiency. While type 2 diabetes mellitus was associated, type 1 was not.

Sleep disorders were the most clearly associated mental and behavioural disorder (F). For the commonly recorded anxiety and depression there was evidence of only a small increase in risk.

Nervous system disorders (G) were generally not associated with eczema. Several diseases of the eye (H) were associated. Those likely related to the well-characterised increased risk of atopic and allergic conjunctivitis include Conjunctivitis, Keratitis, corneal disorders, and disorders of the eyelid. A clear signal was also seen for keratoconus and cataracts.

There was little evidence for diseases of the heart and circulatory system (I).

The largest relative and absolute increases in risk were seen for respiratory diseases (J), especially asthma, allergic rhinitis, and related conditions. Associations were also seen for pneumonia and upper respiratory tract infections.

Coeliac disease and Crohn’s disease were the most strongly associated gastrointestinal conditions (K). There was also evidence for ulcerative colitis, oesophagitis, and fissures, abscesses and haemorrhoids of the anal and rectal region. The existing evidence on the risk of inflammatory bowel diseases from large cohort studies is conflicting.^15,16^

Of skin diseases (L) not indicating atopic eczema itself, itch, or other dermatitis (e.g., contact dermatitis, seborrhoeic dermatitis), there were strong associations with vitiligo, psoriasis, skin infections, and several others. These observed associations are likely due to eczema being a differential diagnosis (e.g., an individual may have eczema recorded in primary care before receiving the actual diagnosis of psoriasis).^17^ This likely also applies to the strong observed association with Systemic lupus erythematosus, recorded as a musculoskeletal/connective tissue condition (M). Other associated conditions include different forms of arthritis (possibly related to psoriasis) and osteoporosis.

There was some evidence for genitourinary conditions (N), in particular phimosis and cystitis.

### Implications

Our findings are generally reassuring and seem plausible judging from clinical practice: in a cohort of people with childhood eczema, besides the already well-known outcomes including atopic and allergic conditions and infections, there is little evidence for considerable increases in risks for most other diseases. Some other strongly associated outcomes have potential alternative explanations relating to eczema misdiagnosis (e.g., lymphomas). As in our previous study in primary care, inflammatory bowel diseases, in particular Crohn’s disease, emerge as outcomes of interest. Eye diseases including allergic conjunctivitis, but also as cataract and keratoconus, showed some of the strongest associations. There is currently limited awareness for the ocular complications of eczema,^18^ despite being listed in the commonly used diagnostic features of atopic dermatitis by Hanifin & Rajka,^12^ possibly due to a lack of evidence from epidemiological studies.

We may consider results from the <18 cohort as most accurate given the clearer diagnosis of eczema in childhood, as well as the avoidance of time-related biases (through the use of a cohort with primarily incident eczema). However, given a maximum follow-up time of 25 years, it is also useful to examine cohorts of adults, including both incident and prevalent eczema to assess risks across the life course. Despite larger potential for bias, results from these adult cohorts suggest increased rates of hospital-recorded diagnoses across multiple organ systems. To know if these outcomes could be prevented, e.g., through better treatment of eczema, we need further research considering factors such as eczema severity, treatments, and outcome-specific considerations.

## Conclusion

In conclusion, our study to our knowledge is the first to offer both a big-picture view of risks faced by people with eczema across the entire disease spectrum, as well as allowing detailed inspection of each individual association. Results will be useful to inform guidelines and clinical practice. Incorporating all diseases and allowing for comparisons make our findings a hypothesis-generating tool for future studies on any disease outcome.

## Methods

### Ethics

The study was approved by the London School of Hygiene & Tropical Medicine Research Ethics Committee (Reference number: 29781). This study is based on data from the CPRD obtained under license from the U.K. Medicines and Healthcare products Regulatory Agency. The data are provided by patients and collected by the National Health Service (NHS) as part of their care and support. The study was approved by the Independent Scientific Advisory Committee (Protocol reference number: 23_002665). Individual patient consent is not required or possible since CPRD provides anonymised data. Consent is given by the GP practices that contribute data to CPRD. Individual patient consent is implied. However, patients are offered the right to opt out from the use of their pseudonymised data.

### Study population

We used an algorithm to identify individuals with eczema (at least one record of an eczema diagnosis code and at least two prescription records for eczema therapies [emollients, topical glucocorticoids, topical calcineurin inhibitors, oral glucocorticoids or systemic immunosuppressants] on two separate days). An analogous algorithm has been previously validated in UK primary care data and was found to have a positive predictive value of 86%.^19^ We included individuals in the eczema cohort from the latest of: (1) Date they met the eczema definition; (2) One year after practice registration (to allow us to reliably capture baseline health status); (3) Study start (April 1, 1997); and (4) 18^th^ (18+ cohort) or 40^th^ birthday (40+ cohort), or no age limitation (any-age cohort). For the 18+ and 40+ cohorts, meeting the eczema definition could occur before individuals became eligible (i.e., individuals with both new and existing eczema were included, a recommended approach for relapsing conditions like eczema to better assess longer-term effects of an exposure).^20^

Eczema exposed individuals were matched (without replacement) to up to 5 unexposed individuals with at least 1-year prior registration, on age (2-year calliper), sex, and general practice in calendar date order. The index date for comparators was set to the index date of the exposed individual they were matched to. Comparators were censored on the day they met the eczema definition themselves and were then included in the eczema cohort.

Individuals were followed until the date of specific outcome (depending on outcome under investigation), or until they were censored (death, left practice, or for comparators, when they met the eczema definition). For each outcome-specific analysis, individuals who had the outcome before their index date were excluded (**Figure 1**).

To address consultation bias or differential health seeking behaviour, individuals who did not have any of four common records (9N11.00 - Seen in GP’s surgery, 22A..00 - Body weight, 4….00 - Laboratory test, 246..00 - O/E - blood pressure reading) in the year prior to index date were excluded as was done in our previous study.^3^

### Outcomes

We assessed outcomes recorded in any diagnostic position (i.e., primary or any other reason for admission or medical history) in Hospital Episode Statistics (HES) Admitted Patient Care (APC) data. We defined outcomes based on: 1) 3-character ICD-10 codes (including all descendant codes) (2,058 outcomes); 2) phecodes (a high-throughput phenotyping used to define clinically meaningful diseases and conditions)^21^ (1,593 outcomes);^4^ and 3) Global Burden of Disease codes at different levels of granularity from a technical paper on the methods for the Global Burden of Disease studies (201 outcomes).^5^

### Statistical analysis

We used Cox proportional hazards regression, stratifying on matched set, to estimate hazard ratios (HRs) and 99% confidence intervals (99%CI) for the association of eczema with each outcome. For each analysis, we estimated crude HRs (implicitly adjusted through matching on age, sex and general practice, and calendar time, as comparators entered the cohort on the same day as exposed individuals). To account for baseline differences in comorbidities and to allow a comparison with adjusted-results from our previous primary care study, we also estimated comorbidity-adjusted HRs (additionally adjusted for history of 71 conditions assessed in primary care from the following disease categories: Atopic and allergic, Immune mediated, Mental health and substance use, attention deficit hyperactivity disorder and autism, Cardiovascular, Metabolic, Bone health, Skin infection, Cancer, Neurological, Digestive system, Liver), as was done in our previous study.^3^

Since comparators were matched, we estimated rates of outcomes among patients without eczema as the rate in people with eczema multiplied by the inverse of the corresponding hazard ratio.

To account for multiple testing, we reported wider 99% instead of usual 95% confidence intervals and defined a Bonferroni corrected significance threshold (with 2,058 ICD-10 categories considered, estimates are considered significant under Bonferroni correction with a p-value less than 0.01/2,058 = 0.000005).

### Pipeline

For all 2,058 outcomes based on 3-character ICD-10 codes, we ran analyses for all five cohorts (any-age, <18, 18+, 40+, ever hospitalised), two models (crude and comorbidity adjusted), and with and without exclusion of non-consulters (sensitivity analyses), resulting in a total of 41,160 combinations. For all 1,593 phecodes and 201 Global Burden of Disease (GBD) codes, we ran crude and adjusted analyses for all five cohorts (15,930 for phecodes, 2,010 combinations for GBD codes). We used R version 4.3.1 and organised the research pipeline using the targets R package, ensuring reproducibility of the computationally expensive pipeline.^22^

## Data availability

Data supporting the findings of this study are available in the article and its Supplementary information. The data underlying this article is provided by the UK CPRD electronic health record database, which is only accessible to researchers with protocols approved by the CPRD’s independent scientific advisory committee. Data access may incur a cost, and further details can be found here: https://www.cprd.com/data-access.

## Funding and acknowledgements

This work uses data provided by patients and collected by the NHS as part of their care and support.

This work was funded by a Wellcome Trust Senior Research Fellowship in Clinical Science (205039/Z/16/Z) awarded to Sinéad M. Langan. Krishnan Bhaskaran is funded by a Wellcome Senior Research Fellowship (220283/Z/20/Z). The views expressed in this publication are those of the author(s) and not necessarily those of the NIHR, NHS or the UK Department of Health and Social Care.

## Author contributions

JM contributed to Conceptualization, Data curation, Formal Analysis, Funding acquisition, Investigation, Methodology, Project administration, Resources, Software, Supervision, Validation, Visualization, Writing – original draft, and Writing – review & editing.

SML contributed to Methodology, Supervision, Validation, and Writing – review & editing. JM, AS, KB, AR, SD, KEM, and SML contributed to Methodology and Writing – review & editing.

## References

1. Davis, D. M. R. et al. AAD Guidelines: awareness of comorbidities associated with atopic dermatitis in adults. Journal of the American Academy of Dermatology S0190962222000809 (2022) doi:10.1016/j.jaad.2022.01.009.

2. Langan, S. M., Irvine, A. D. & Weidinger, S. Atopic dermatitis. The Lancet 396, 345–360 (2020).

3. Matthewman, J. et al. Cohort studies on 71 outcomes among people with atopic eczema in UK primary care data. Nat Commun 15, 9573 (2024).

4. Wu, P. et al. Mapping ICD-10 and ICD-10-CM Codes to Phecodes: Workflow Development and Initial Evaluation. JMIR Med Inform 7, e14325 (2019).

5. WHO. WHO Methods and Data Sources for Global Burden of Disease Estimates 2000-2019. https://www.who.int/docs/default-source/gho-documents/global-health-estimates/ghe2019_daly-methods.pdf (2020).

6. Wolf, A. et al. Data resource profile: Clinical Practice Research Datalink (CPRD) Aurum. International Journal of Epidemiology 48, 1740–1740g (2019).

7. Phelan, M., Bhavsar, N. A. & Goldstein, B. A. Illustrating Informed Presence Bias in Electronic Health Records Data: How Patient Interactions with a Health System Can Impact Inference. eGEMs 5, 22 (2017).

8. Mulick, A. R. et al. Novel multimorbidity clusters in people with eczema and asthma: a population-based cluster analysis. Sci Rep 12, 21866 (2022).

9. Rubenstein, M. & Duvic, M. Cutaneous manifestations of Hodgkin’s disease. International Journal of Dermatology 45, 251–256 (2006).

10. Lam, M., Lu, J. D., Elhadad, L., Sibbald, C. & Alhusayen, R. Common Dermatologic Disorders in Down Syndrome: Systematic Review. JMIR Dermatol 5, e33391 (2022).

11. Abuabara, K., Yu, A. M., Okhovat, J.-P., Allen, I. E. & Langan, S. M. The prevalence of atopic dermatitis beyond childhood: A systematic review and meta-analysis of longitudinal studies. Allergy 73, 696–704 (2018).

12. Hanifin, J. M. & Rajka, G. Diagnostic Features of Atopic Dermatitis. Acta Dermato-Venereologica 60, 44–47 (1980).

13. Mansfield, K. E. et al. Association Between Atopic Eczema and Cancer in England and Denmark. JAMA Dermatol 156, 1086 (2020).

14. Rafiq, M. et al. Allergic disease, corticosteroid use, and risk of Hodgkin lymphoma: A United Kingdom nationwide case-control study. Journal of Allergy and Clinical Immunology 145, 868–876 (2020).

15. Schneeweiss, M. C. et al. Occurrence of inflammatory bowel disease in patients with chronic inflammatory skin diseases: a cohort study*. British Journal of Dermatology 187, 692–703 (2022).

16. Chiesa Fuxench, Z. C., et al. Risk of Inflammatory Bowel Disease in Patients With Atopic Dermatitis. JAMA Dermatology 159, 1085–1092 (2023).

17. Differential diagnosis | Diagnosis | Psoriasis | CKS | NICE. https://cks.nice.org.uk/topics/psoriasis/diagnosis/differential-diagnosis/.

18. Thyssen, J., et al. Management of Ocular Manifestations of Atopic Dermatitis: A Consensus Meeting Using a Modified Delphi Process. Acta Derm Venereol 100, adv00264 (2020).

19. Abuabara, K. et al. Development and Validation of an Algorithm to Accurately Identify Atopic Eczema Patients in Primary Care Electronic Health Records from the UK. Journal of Investigative Dermatology 137, 1655–1662 (2017).

20. Vandenbroucke, J. & Pearce, N. Point: Incident Exposures, Prevalent Exposures, and Causal Inference: Does Limiting Studies to Persons Who Are Followed From First Exposure Onward Damage Epidemiology? American Journal of Epidemiology 182, 826– 833 (2015).

21. Denaxas, S. spiros/phemap. https://github.com/spiros/phemap (2025).

22. Landau, W. The targets R package: a dynamic Make-like function-oriented pipeline toolkit for reproducibility and high-performance computing. JOSS 6, 2959 (2021).

