## Supplementary Table 1 for "Mapping risks of hospital-recorded health conditions in people with eczema"

| Supplementary Table 1: Baseline characteristics | | | |
|---|---|---|---|
| Characteristic | without eczema[1] | with eczema[1] | SMD[2] |
| any age - | | | |
| N | 14,103,462 (100%) | 3,077,362 (100%) | NA |
| Male | 6,222,354 (44%) | 1,376,488 (45%) | NA |
| Age at index | 27 (7, 50) | 24 (5, 49) | 0.05 |
| Follow-up time | 4.7 (1.8, 10.0) | 5.5 (2.1, 11.0) | -0.10 |
| Cons. in year pre index | 5,578,487 (40%) | 1,645,237 (53%) | -0.28 |
| any age - Atopic and allergic | | | |
| Asthma | 1,222,609 (8.7%) | 521,073 (17%) | -0.25 |
| Food allergy | 92,327 (0.7%) | 66,564 (2.2%) | -0.13 |
| Allergic Rhinitis | 788,710 (5.6%) | 359,113 (12%) | -0.22 |
| Allergic Conjunctivitis | 103,347 (0.7%) | 58,391 (1.9%) | -0.10 |
| Eosinophilic Eosophagitis | 480 (<0.1%) | 227 (<0.1%) | -0.01 |
| any age - Immune mediated | | | |
| Alopecia Areata | 23,480 (0.2%) | 10,607 (0.3%) | -0.04 |
| Urticaria | 298,195 (2.1%) | 139,207 (4.5%) | -0.13 |
| COPD | 191,149 (1.4%) | 57,785 (1.9%) | -0.04 |
| any age - Mental health and substance use | | | |
| Anxiety | 1,060,930 (7.5%) | 306,920 (10.0%) | -0.09 |
| Depression | 1,552,123 (11%) | 430,690 (14%) | -0.09 |
| Alcohol abuse | 104,431 (0.7%) | 29,033 (0.9%) | -0.02 |
| Cigarette smoking | 3,609,461 (26%) | 865,234 (28%) | -0.06 |
| any age - ADHD and autism | | | |
| ADHD | 45,124 (0.3%) | 12,043 (0.4%) | -0.01 |
| Autism | 44,226 (0.3%) | 15,221 (0.5%) | -0.03 |
| any age - Cardiovascular | | | |
| Hypertension | 1,314,047 (9.3%) | 316,381 (10%) | -0.03 |
| Coronary artery disease | 640,034 (4.5%) | 166,593 (5.4%) | -0.04 |
| Peripheral artery disease | 77,786 (0.6%) | 22,983 (0.7%) | -0.02 |
| Myocardial infarction | 125,198 (0.9%) | 31,140 (1.0%) | -0.01 |
| Stroke | 105,289 (0.7%) | 27,466 (0.9%) | -0.02 |
| Heart failure | 108,327 (0.8%) | 29,405 (1.0%) | -0.02 |
| Thromboembolic diseases | 108,636 (0.8%) | 32,983 (1.1%) | -0.03 |
| any age - Metabolic | | | |
| Obesity | 395,015 (2.8%) | 116,931 (3.8%) | -0.06 |
| Dyslipidemia | 523,266 (3.7%) | 134,710 (4.4%) | -0.03 |
| Diabetes mellitus | 469,967 (3.3%) | 117,708 (3.8%) | -0.03 |
| Metabolic syndrome | 2,712 (<0.1%) | 838 (<0.1%) | -0.01 |
| any age - Bone health | | | |
| Hip fracture | 43,575 (0.3%) | 10,940 (0.4%) | -0.01 |

[1] Mean and interquartile range is given for Age at index and Follow-up time; all others are counts and percentage.

[2] Standardised mean difference

| Supplementary Table 1: Baseline characteristics | | | |
|---|---|---|---|
| Characteristic | without eczema[1] | with eczema[1] | SMD[2] |
| Pelvis fracture | 18,629 (0.1%) | 4,834 (0.2%) | -0.01 |
| Spine fracture | 27,829 (0.2%) | 7,510 (0.2%) | -0.01 |
| Wrist fracture | 295,000 (2.1%) | 76,181 (2.5%) | -0.03 |
| Osteoporosis | 157,486 (1.1%) | 42,055 (1.4%) | -0.02 |
| any age - Skin infection | | | |
| Molluscum contagiosum | 146,684 (1.0%) | 89,752 (2.9%) | -0.14 |
| Impetigo | 394,360 (2.8%) | 201,644 (6.6%) | -0.18 |
| Herpes simplex | 190,422 (1.4%) | 77,891 (2.5%) | -0.09 |
| Dermatophyte infection | 629,392 (4.5%) | 288,839 (9.4%) | -0.19 |
| Cutaneous warts | 792,026 (5.6%) | 297,022 (9.7%) | -0.15 |
| any age - Cancer | | | |
| Lung cancer | 8,836 (<0.1%) | 2,446 (<0.1%) | -0.01 |
| Breast cancer | 74,899 (0.5%) | 18,168 (0.6%) | -0.01 |
| Prostate cancer | 50,641 (0.4%) | 12,245 (0.4%) | -0.01 |
| Pancreatic cancer | 1,307 (<0.1%) | 352 (<0.1%) | 0.00 |
| Non-hodgkin lymphoma | 12,300 (<0.1%) | 3,328 (0.1%) | -0.01 |
| Hodgkin lymphoma | 3,325 (<0.1%) | 954 (<0.1%) | 0.00 |
| Myeloma | 3,272 (<0.1%) | 748 (<0.1%) | 0.00 |
| CNS cancers | 10,959 (<0.1%) | 2,884 (<0.1%) | -0.01 |
| Melanoma | 41,371 (0.3%) | 10,770 (0.3%) | -0.01 |
| Nonmelanoma skin cancer | 156,091 (1.1%) | 41,308 (1.3%) | -0.02 |
| any age - Neurological | | | |
| Alzheimer's dementia | 40,335 (0.3%) | 9,307 (0.3%) | 0.00 |
| Vascular dementia | 20,791 (0.1%) | 4,929 (0.2%) | 0.00 |
| Epilepsy | 109,550 (0.8%) | 31,479 (1.0%) | -0.03 |
| Migraine | 448,754 (3.2%) | 134,735 (4.4%) | -0.06 |
| Multiple sclerosis | 15,974 (0.1%) | 4,236 (0.1%) | -0.01 |
| Parkinson's disease | 24,898 (0.2%) | 5,370 (0.2%) | 0.00 |
| Peripheral neuropathies | 662,781 (4.7%) | 191,667 (6.2%) | -0.07 |
| any age - Digestive system | | | |
| Abdominal hernia | 353,442 (2.5%) | 94,890 (3.1%) | -0.04 |
| Appendicitis | 131,921 (0.9%) | 32,882 (1.1%) | -0.01 |
| Barett's oesophagus | 25,032 (0.2%) | 6,916 (0.2%) | -0.01 |
| Coeliac disease | 24,758 (0.2%) | 8,161 (0.3%) | -0.02 |
| Crohn's disease | 20,829 (0.1%) | 7,969 (0.3%) | -0.02 |
| Diverticular disease | 188,648 (1.3%) | 52,246 (1.7%) | -0.03 |
| Gastritis and duodenitis | 248,985 (1.8%) | 78,099 (2.5%) | -0.05 |
| Gastro oesophageal reflux | 561,049 (4.0%) | 177,375 (5.8%) | -0.08 |
| Irritable bowel syndrome | 345,139 (2.4%) | 111,849 (3.6%) | -0.07 |
| Oesophageal ulcer | 256,539 (1.8%) | 80,121 (2.6%) | -0.05 |
| Pancreatitis | 24,860 (0.2%) | 6,576 (0.2%) | -0.01 |
| Peptic ulcer disease | 69,376 (0.5%) | 18,469 (0.6%) | -0.01 |

[1] Mean and interquartile range is given for Age at index and Follow-up time; all others are counts and percentage.

[2] Standardised mean difference

| Supplementary Table 1: Baseline characteristics | | | |
|---|---|---|---|
| Characteristic | without eczema[1] | with eczema[1] | SMD[2] |
| Peritonitis | 9,448 (<0.1%) | 2,365 (<0.1%) | 0.00 |
| Ulcerative colitis | 30,640 (0.2%) | 10,366 (0.3%) | -0.02 |
| any age - Liver | | | |
| Autoimmune liver disease | 4,146 (<0.1%) | 1,197 (<0.1%) | -0.01 |
| Cholecystitis | 38,343 (0.3%) | 10,288 (0.3%) | -0.01 |
| Fatty liver | 50,688 (0.4%) | 16,009 (0.5%) | -0.02 |
| Fibrosis/sclerosis/cirrhosis | 13,958 (<0.1%) | 4,331 (0.1%) | -0.01 |
| Oesophageal varices | 3,296 (<0.1%) | 986 (<0.1%) | -0.01 |
| 18+ - | | | |
| N | 10,605,853 (100%) | 2,167,108 (100%) | NA |
| Male | 4,431,643 (42%) | 903,026 (42%) | NA |
| Age at index | 37 (23, 58) | 37 (23, 59) | -0.02 |
| Follow-up time | 4.4 (1.8, 9.3) | 4.8 (1.9, 9.9) | -0.06 |
| Cons. in year pre index | 4,893,589 (46%) | 1,398,020 (65%) | -0.38 |
| 18+ - Atopic and allergic | | | |
| Asthma | 1,234,248 (12%) | 495,151 (23%) | -0.30 |
| Food allergy | 85,028 (0.8%) | 46,079 (2.1%) | -0.11 |
| Allergic Rhinitis | 875,117 (8.3%) | 368,215 (17%) | -0.27 |
| Allergic Conjunctivitis | 120,141 (1.1%) | 59,572 (2.7%) | -0.12 |
| Eosinophilic Eosophagitis | 555 (<0.1%) | 256 (<0.1%) | -0.01 |
| 18+ - Immune mediated | | | |
| Alopecia Areata | 25,818 (0.2%) | 11,205 (0.5%) | -0.04 |
| Urticaria | 315,375 (3.0%) | 128,031 (5.9%) | -0.14 |
| COPD | 192,222 (1.8%) | 57,946 (2.7%) | -0.06 |
| 18+ - Mental health and substance use | | | |
| Anxiety | 1,122,564 (11%) | 322,655 (15%) | -0.13 |
| Depression | 1,612,428 (15%) | 449,111 (21%) | -0.14 |
| Alcohol abuse | 104,670 (1.0%) | 29,123 (1.3%) | -0.03 |
| Cigarette smoking | 3,709,250 (35%) | 893,121 (41%) | -0.13 |
| 18+ - ADHD and autism | | | |
| ADHD | 51,678 (0.5%) | 12,103 (0.6%) | -0.01 |
| Autism | 49,653 (0.5%) | 13,462 (0.6%) | -0.02 |
| 18+ - Cardiovascular | | | |
| Hypertension | 1,314,149 (12%) | 316,170 (15%) | -0.06 |
| Coronary artery disease | 639,324 (6.0%) | 166,602 (7.7%) | -0.07 |
| Peripheral artery disease | 79,447 (0.7%) | 23,539 (1.1%) | -0.04 |
| Myocardial infarction | 125,009 (1.2%) | 31,139 (1.4%) | -0.02 |
| Stroke | 104,897 (1.0%) | 27,346 (1.3%) | -0.03 |
| Heart failure | 107,091 (1.0%) | 29,277 (1.4%) | -0.03 |
| Thromboembolic diseases | 108,806 (1.0%) | 33,180 (1.5%) | -0.04 |

[1] Mean and interquartile range is given for Age at index and Follow-up time; all others are counts and percentage.

[2] Standardised mean difference

Supplementary Table 1: Baseline characteristics

| Characteristic | without eczema[1] | with eczema[1] | SMD[2] |
|---|---|---|---|
| 18+ - Metabolic | | | |
| Obesity | 402,981 (3.8%) | 118,834 (5.5%) | -0.08 |
| Dyslipidemia | 523,680 (4.9%) | 134,784 (6.2%) | -0.06 |
| Diabetes mellitus | 472,368 (4.5%) | 117,877 (5.4%) | -0.05 |
| Metabolic syndrome | 2,726 (<0.1%) | 846 (<0.1%) | -0.01 |
| 18+ - Bone health | | | |
| Hip fracture | 43,828 (0.4%) | 10,970 (0.5%) | -0.01 |
| Pelvis fracture | 19,317 (0.2%) | 5,005 (0.2%) | -0.01 |
| Spine fracture | 28,833 (0.3%) | 7,729 (0.4%) | -0.02 |
| Wrist fracture | 338,711 (3.2%) | 83,507 (3.9%) | -0.04 |
| Osteoporosis | 158,047 (1.5%) | 42,185 (1.9%) | -0.04 |
| 18+ - Skin infection | | | |
| Molluscum contagiosum | 194,304 (1.8%) | 83,829 (3.9%) | -0.12 |
| Impetigo | 432,957 (4.1%) | 178,705 (8.2%) | -0.17 |
| Herpes simplex | 202,523 (1.9%) | 77,197 (3.6%) | -0.10 |
| Dermatophyte infection | 662,551 (6.2%) | 277,474 (13%) | -0.22 |
| Cutaneous warts | 981,375 (9.3%) | 322,054 (15%) | -0.17 |
| 18+ - Cancer | | | |
| Lung cancer | 8,678 (<0.1%) | 2,436 (0.1%) | -0.01 |
| Breast cancer | 74,747 (0.7%) | 18,178 (0.8%) | -0.02 |
| Prostate cancer | 50,873 (0.5%) | 12,243 (0.6%) | -0.01 |
| Pancreatic cancer | 1,285 (<0.1%) | 354 (<0.1%) | 0.00 |
| Non-hodgkin lymphoma | 12,455 (0.1%) | 3,333 (0.2%) | -0.01 |
| Hodgkin lymphoma | 3,456 (<0.1%) | 1,010 (<0.1%) | -0.01 |
| Myeloma | 3,352 (<0.1%) | 748 (<0.1%) | 0.00 |
| CNS cancers | 10,896 (0.1%) | 2,877 (0.1%) | -0.01 |
| Melanoma | 41,166 (0.4%) | 10,719 (0.5%) | -0.02 |
| Nonmelanoma skin cancer | 156,122 (1.5%) | 41,315 (1.9%) | -0.03 |
| 18+ - Neurological | | | |
| Alzheimer's dementia | 40,145 (0.4%) | 9,326 (0.4%) | -0.01 |
| Vascular dementia | 20,756 (0.2%) | 4,922 (0.2%) | -0.01 |
| Epilepsy | 108,905 (1.0%) | 30,926 (1.4%) | -0.04 |
| Migraine | 493,450 (4.7%) | 146,192 (6.7%) | -0.09 |
| Multiple sclerosis | 15,922 (0.2%) | 4,245 (0.2%) | -0.01 |
| Parkinson's disease | 24,920 (0.2%) | 5,372 (0.2%) | 0.00 |
| Peripheral neuropathies | 664,777 (6.3%) | 191,806 (8.9%) | -0.10 |
| 18+ - Digestive system | | | |
| Abdominal hernia | 286,265 (2.7%) | 72,461 (3.3%) | -0.04 |
| Appendicitis | 145,789 (1.4%) | 35,705 (1.6%) | -0.02 |
| Barett's oesophagus | 24,999 (0.2%) | 6,939 (0.3%) | -0.02 |
| Coeliac disease | 25,285 (0.2%) | 8,101 (0.4%) | -0.02 |

[1] Mean and interquartile range is given for Age at index and Follow-up time; all others are counts and percentage.

[2] Standardised mean difference

| Supplementary Table 1: Baseline characteristics | | | |
|---|---|---|---|
| Characteristic | without eczema[1] | with eczema[1] | SMD[2] |
| Crohn's disease | 22,397 (0.2%) | 8,416 (0.4%) | -0.03 |
| Diverticular disease | 188,506 (1.8%) | 52,228 (2.4%) | -0.04 |
| Gastritis and duodenitis | 254,816 (2.4%) | 78,445 (3.6%) | -0.07 |
| Gastro oesophageal reflux | 415,368 (3.9%) | 126,694 (5.8%) | -0.09 |
| Irritable bowel syndrome | 356,169 (3.4%) | 115,391 (5.3%) | -0.10 |
| Oesophageal ulcer | 242,547 (2.3%) | 74,675 (3.4%) | -0.07 |
| Pancreatitis | 25,028 (0.2%) | 6,648 (0.3%) | -0.01 |
| Peptic ulcer disease | 69,170 (0.7%) | 18,504 (0.9%) | -0.02 |
| Peritonitis | 9,671 (<0.1%) | 2,405 (0.1%) | -0.01 |
| Ulcerative colitis | 31,321 (0.3%) | 10,573 (0.5%) | -0.03 |
| 18+ - Liver | | | |
| Autoimmune liver disease | 4,267 (<0.1%) | 1,219 (<0.1%) | -0.01 |
| Cholecystitis | 38,448 (0.4%) | 10,335 (0.5%) | -0.02 |
| Fatty liver | 51,022 (0.5%) | 16,154 (0.7%) | -0.03 |
| Fibrosis/sclerosis/cirrhosis | 13,955 (0.1%) | 4,318 (0.2%) | -0.02 |
| Oesophageal varices | 3,317 (<0.1%) | 984 (<0.1%) | -0.01 |
| 40+ - | | | |
| N | 5,873,150 (100%) | 1,202,474 (100%) | NA |
| Male | 2,487,358 (42%) | 509,194 (42%) | NA |
| Age at index | 55 (43, 69) | 56 (43, 70) | -0.03 |
| Follow-up time | 5.8 (2.5, 10.8) | 6.2 (2.7, 11.3) | -0.05 |
| Cons. in year pre index | 3,207,342 (55%) | 843,989 (70%) | -0.33 |
| 40+ - Atopic and allergic | | | |
| Asthma | 615,931 (10%) | 225,131 (19%) | -0.23 |
| Food allergy | 28,900 (0.5%) | 11,175 (0.9%) | -0.05 |
| Allergic Rhinitis | 399,891 (6.8%) | 153,553 (13%) | -0.20 |
| Allergic Conjunctivitis | 60,001 (1.0%) | 29,110 (2.4%) | -0.11 |
| Eosinophilic Eosophagitis | 249 (<0.1%) | 102 (<0.1%) | -0.01 |
| 40+ - Immune mediated | | | |
| Alopecia Areata | 13,648 (0.2%) | 5,695 (0.5%) | -0.04 |
| Urticaria | 153,364 (2.6%) | 64,373 (5.4%) | -0.14 |
| COPD | 190,152 (3.2%) | 56,912 (4.7%) | -0.08 |
| 40+ - Mental health and substance use | | | |
| Anxiety | 735,641 (13%) | 204,416 (17%) | -0.13 |
| Depression | 1,126,879 (19%) | 304,057 (25%) | -0.15 |
| Alcohol abuse | 90,659 (1.5%) | 25,208 (2.1%) | -0.04 |
| Cigarette smoking | 2,407,939 (41%) | 564,196 (47%) | -0.12 |
| 40+ - ADHD and autism | | | |
| ADHD | 2,331 (<0.1%) | 648 (<0.1%) | -0.01 |
| Autism | 3,354 (<0.1%) | 1,312 (0.1%) | -0.02 |
| 40+ - Cardiovascular | | | |

[1] Mean and interquartile range is given for Age at index and Follow-up time; all others are counts and percentage.

[2] Standardised mean difference

### Supplementary Table 1: Baseline characteristics

| Characteristic | without eczema[1] | with eczema[1] | SMD[2] |
|---|---|---|---|
| Hypertension | 1,311,220 (22%) | 315,565 (26%) | -0.09 |
| Coronary artery disease | 645,785 (11%) | 168,509 (14%) | -0.09 |
| Peripheral artery disease | 69,165 (1.2%) | 20,173 (1.7%) | -0.04 |
| Myocardial infarction | 123,373 (2.1%) | 30,779 (2.6%) | -0.03 |
| Stroke | 102,507 (1.7%) | 26,766 (2.2%) | -0.03 |
| Heart failure | 106,574 (1.8%) | 29,024 (2.4%) | -0.04 |
| Thromboembolic diseases | 102,151 (1.7%) | 31,091 (2.6%) | -0.06 |
| 40+ - Metabolic | | | |
| Obesity | 344,910 (5.9%) | 98,421 (8.2%) | -0.09 |
| Dyslipidemia | 517,580 (8.8%) | 133,075 (11%) | -0.08 |
| Diabetes mellitus | 453,376 (7.7%) | 113,130 (9.4%) | -0.06 |
| Metabolic syndrome | 2,609 (<0.1%) | 806 (<0.1%) | -0.01 |
| 40+ - Bone health | | | |
| Hip fracture | 42,271 (0.7%) | 10,581 (0.9%) | -0.02 |
| Pelvis fracture | 15,797 (0.3%) | 4,054 (0.3%) | -0.01 |
| Spine fracture | 23,834 (0.4%) | 6,443 (0.5%) | -0.02 |
| Wrist fracture | 147,902 (2.5%) | 35,427 (2.9%) | -0.03 |
| Osteoporosis | 154,605 (2.6%) | 40,971 (3.4%) | -0.05 |
| 40+ - Skin infection | | | |
| Molluscum contagiosum | 7,980 (0.1%) | 2,920 (0.2%) | -0.02 |
| Impetigo | 84,427 (1.4%) | 36,361 (3.0%) | -0.11 |
| Herpes simplex | 114,192 (1.9%) | 41,360 (3.4%) | -0.09 |
| Dermatophyte infection | 423,613 (7.2%) | 169,264 (14%) | -0.22 |
| Cutaneous warts | 390,155 (6.6%) | 122,114 (10%) | -0.13 |
| 40+ - Cancer | | | |
| Lung cancer | 8,722 (0.1%) | 2,421 (0.2%) | -0.01 |
| Breast cancer | 75,400 (1.3%) | 18,189 (1.5%) | -0.02 |
| Prostate cancer | 50,537 (0.9%) | 12,128 (1.0%) | -0.02 |
| Pancreatic cancer | 1,249 (<0.1%) | 348 (<0.1%) | 0.00 |
| Non-hodgkin lymphoma | 11,594 (0.2%) | 3,134 (0.3%) | -0.01 |
| Hodgkin lymphoma | 2,226 (<0.1%) | 687 (<0.1%) | -0.01 |
| Myeloma | 3,365 (<0.1%) | 744 (<0.1%) | 0.00 |
| CNS cancers | 9,322 (0.2%) | 2,408 (0.2%) | -0.01 |
| Melanoma | 39,683 (0.7%) | 10,136 (0.8%) | -0.02 |
| Nonmelanoma skin cancer | 156,596 (2.7%) | 41,296 (3.4%) | -0.04 |
| 40+ - Neurological | | | |
| Alzheimer's dementia | 40,453 (0.7%) | 9,330 (0.8%) | -0.01 |
| Vascular dementia | 20,670 (0.4%) | 4,940 (0.4%) | -0.01 |
| Epilepsy | 67,580 (1.2%) | 19,627 (1.6%) | -0.04 |
| Migraine | 279,686 (4.8%) | 79,192 (6.6%) | -0.08 |
| Multiple sclerosis | 14,963 (0.3%) | 3,907 (0.3%) | -0.01 |

[1] Mean and interquartile range is given for Age at index and Follow-up time; all others are counts and percentage.

[2] Standardised mean difference

### Supplementary Table 1: Baseline characteristics

| Characteristic | without eczema[1] | with eczema[1] | SMD[2] |
|---|---|---|---|
| Parkinson's disease | 24,805 (0.4%) | 5,369 (0.4%) | 0.00 |
| Peripheral neuropathies | 633,963 (11%) | 180,408 (15%) | -0.13 |
| 40+ - Digestive system | | | |
| Abdominal hernia | 238,468 (4.1%) | 59,439 (4.9%) | -0.04 |
| Appendicitis | 61,707 (1.1%) | 14,256 (1.2%) | -0.01 |
| Barett's oesophagus | 24,748 (0.4%) | 6,863 (0.6%) | -0.02 |
| Coeliac disease | 15,929 (0.3%) | 4,961 (0.4%) | -0.02 |
| Crohn's disease | 15,546 (0.3%) | 5,242 (0.4%) | -0.03 |
| Diverticular disease | 189,288 (3.2%) | 52,332 (4.4%) | -0.06 |
| Gastritis and duodenitis | 202,446 (3.4%) | 60,761 (5.1%) | -0.08 |
| Gastro oesophageal reflux | 350,610 (6.0%) | 105,448 (8.8%) | -0.11 |
| Irritable bowel syndrome | 256,647 (4.4%) | 80,667 (6.7%) | -0.10 |
| Oesophageal ulcer | 225,636 (3.8%) | 68,437 (5.7%) | -0.09 |
| Pancreatitis | 22,724 (0.4%) | 5,927 (0.5%) | -0.02 |
| Peptic ulcer disease | 66,563 (1.1%) | 17,664 (1.5%) | -0.03 |
| Peritonitis | 7,813 (0.1%) | 1,901 (0.2%) | -0.01 |
| Ulcerative colitis | 25,794 (0.4%) | 8,212 (0.7%) | -0.03 |
| 40+ - Liver | | | |
| Autoimmune liver disease | 3,778 (<0.1%) | 1,058 (<0.1%) | -0.01 |
| Cholecystitis | 36,644 (0.6%) | 9,648 (0.8%) | -0.02 |
| Fatty liver | 49,350 (0.8%) | 15,243 (1.3%) | -0.04 |
| Fibrosis/sclerosis/cirrhosis | 13,794 (0.2%) | 4,244 (0.4%) | -0.02 |
| Oesophageal varices | 3,252 (<0.1%) | 953 (<0.1%) | -0.01 |
| <18 - | | | |
| N | 6,054,252 (100%) | 1,424,335 (100%) | NA |
| Male | 2,974,532 (49%) | 711,754 (50%) | NA |
| Age at index | 4 (1, 13) | 4 (2, 12) | 0.05 |
| Follow-up time | 3.9 (1.5, 9.1) | 5.2 (2.0, 10.8) | -0.16 |
| Cons. in year pre index | 1,464,135 (24%) | 487,852 (34%) | -0.22 |
| <18 - Atopic and allergic | | | |
| Asthma | 386,222 (6.4%) | 192,274 (13%) | -0.24 |
| Food allergy | 46,431 (0.8%) | 44,241 (3.1%) | -0.17 |
| Allergic Rhinitis | 232,553 (3.8%) | 132,302 (9.3%) | -0.22 |
| Allergic Conjunctivitis | 33,224 (0.5%) | 22,837 (1.6%) | -0.10 |
| <18 - Immune mediated | | | |
| Alopecia Areata | 5,888 (<0.1%) | 2,898 (0.2%) | -0.03 |
| Urticaria | 113,247 (1.9%) | 59,243 (4.2%) | -0.13 |
| COPD | 1,942 (<0.1%) | 876 (<0.1%) | -0.01 |
| <18 - Mental health and substance use | | | |
| Anxiety | 161,974 (2.7%) | 47,560 (3.3%) | -0.04 |
| Depression | 177,780 (2.9%) | 47,255 (3.3%) | -0.02 |

[1] Mean and interquartile range is given for Age at index and Follow-up time; all others are counts and percentage.

[2] Standardised mean difference

### Supplementary Table 1: Baseline characteristics

| Characteristic | without eczema[1] | with eczema[1] | SMD[2] |
|---|---|---|---|
| Alcohol abuse | 4,429 (<0.1%) | 979 (<0.1%) | 0.00 |
| Cigarette smoking | 468,067 (7.7%) | 121,591 (8.5%) | -0.03 |
| <18 - ADHD and autism | | | |
| ADHD | 34,156 (0.6%) | 8,734 (0.6%) | -0.01 |
| Autism | 35,302 (0.6%) | 11,363 (0.8%) | -0.03 |
| <18 - Cardiovascular | | | |
| Hypertension | 5,594 (<0.1%) | 1,423 (<0.1%) | 0.00 |
| Coronary artery disease | 667 (<0.1%) | 255 (<0.1%) | -0.01 |
| Peripheral artery disease | 4,651 (<0.1%) | 1,670 (0.1%) | -0.01 |
| Myocardial infarction | 691 (<0.1%) | 183 (<0.1%) | 0.00 |
| Stroke | 1,957 (<0.1%) | 532 (<0.1%) | 0.00 |
| Heart failure | 1,267 (<0.1%) | 340 (<0.1%) | 0.00 |
| Thromboembolic diseases | 3,173 (<0.1%) | 910 (<0.1%) | 0.00 |
| <18 - Metabolic | | | |
| Obesity | 28,013 (0.5%) | 9,493 (0.7%) | -0.03 |
| Dyslipidemia | 4,389 (<0.1%) | 1,142 (<0.1%) | 0.00 |
| Diabetes mellitus | 11,467 (0.2%) | 2,676 (0.2%) | 0.00 |
| <18 - Bone health | | | |
| Hip fracture | 900 (<0.1%) | 223 (<0.1%) | 0.00 |
| Pelvis fracture | 1,200 (<0.1%) | 359 (<0.1%) | 0.00 |
| Spine fracture | 1,639 (<0.1%) | 420 (<0.1%) | 0.00 |
| Wrist fracture | 82,959 (1.4%) | 23,700 (1.7%) | -0.02 |
| Osteoporosis | 1,933 (<0.1%) | 514 (<0.1%) | 0.00 |
| <18 - Skin infection | | | |
| Molluscum contagiosum | 119,971 (2.0%) | 78,159 (5.5%) | -0.19 |
| Impetigo | 249,959 (4.1%) | 137,124 (9.6%) | -0.22 |
| Herpes simplex | 47,178 (0.8%) | 23,848 (1.7%) | -0.08 |
| Dermatophyte infection | 148,086 (2.4%) | 86,252 (6.1%) | -0.18 |
| Cutaneous warts | 285,312 (4.7%) | 130,243 (9.1%) | -0.18 |
| <18 - Cancer | | | |
| Non-hodgkin lymphoma | 411 (<0.1%) | 137 (<0.1%) | 0.00 |
| Hodgkin lymphoma | 388 (<0.1%) | 107 (<0.1%) | 0.00 |
| CNS cancers | 1,093 (<0.1%) | 270 (<0.1%) | 0.00 |
| Melanoma | 938 (<0.1%) | 446 (<0.1%) | -0.01 |
| <18 - Digestive system | | | |
| Abdominal hernia | 107,056 (1.8%) | 32,820 (2.3%) | -0.04 |
| Appendicitis | 33,004 (0.5%) | 8,863 (0.6%) | -0.01 |
| Coeliac disease | 5,922 (<0.1%) | 2,061 (0.1%) | -0.01 |
| Crohn's disease | 2,249 (<0.1%) | 977 (<0.1%) | -0.01 |
| Gastritis and duodenitis | 27,950 (0.5%) | 10,173 (0.7%) | -0.03 |
| Gastro oesophageal reflux | 193,570 (3.2%) | 64,001 (4.5%) | -0.07 |

[1] Mean and interquartile range is given for Age at index and Follow-up time; all others are counts and percentage.

[2] Standardised mean difference

Supplementary Table 1: Baseline characteristics

| Characteristic | without eczema[1] | with eczema[1] | SMD[2] |
|---|---|---|---|
| Irritable bowel syndrome | 33,226 (0.5%) | 11,078 (0.8%) | -0.03 |
| Oesophageal ulcer | 26,166 (0.4%) | 9,473 (0.7%) | -0.03 |
| Pancreatitis | 912 (<0.1%) | 238 (<0.1%) | 0.00 |
| Peptic ulcer disease | 961 (<0.1%) | 290 (<0.1%) | 0.00 |
| Peritonitis | 908 (<0.1%) | 247 (<0.1%) | 0.00 |
| Ulcerative colitis | 2,109 (<0.1%) | 764 (<0.1%) | -0.01 |
| <18 - Liver | | | |
| Cholecystitis | 1,003 (<0.1%) | 284 (<0.1%) | 0.00 |
| Fatty liver | 1,776 (<0.1%) | 482 (<0.1%) | 0.00 |
| <18 - Neurological | | | |
| Epilepsy | 25,838 (0.4%) | 6,723 (0.5%) | -0.01 |
| Migraine | 81,867 (1.4%) | 26,094 (1.8%) | -0.04 |
| Peripheral neuropathies | 22,993 (0.4%) | 6,640 (0.5%) | -0.01 |
| hospitalised - | | | |
| N | 10,571,189 (100%) | 2,435,001 (100%) | NA |
| Male | 4,534,880 (43%) | 1,067,196 (44%) | NA |
| Age at index | 30 (6, 55) | 26 (5, 53) | 0.07 |
| Follow-up time | 4.7 (1.9, 9.4) | 5.2 (2.1, 10.1) | -0.08 |
| Cons. in year pre index | 4,910,747 (46%) | 1,360,996 (56%) | -0.19 |
| hospitalised - Atopic and allergic | | | |
| Asthma | 1,084,680 (10%) | 437,131 (18%) | -0.22 |
| Food allergy | 77,587 (0.7%) | 56,828 (2.3%) | -0.13 |
| Allergic Rhinitis | 694,223 (6.6%) | 299,414 (12%) | -0.20 |
| Allergic Conjunctivitis | 97,505 (0.9%) | 52,351 (2.1%) | -0.10 |
| Eosinophilic Eosophagitis | 700 (<0.1%) | 264 (<0.1%) | 0.00 |
| hospitalised - Immune mediated | | | |
| Alopecia Areata | 21,715 (0.2%) | 9,105 (0.4%) | -0.03 |
| Urticaria | 283,121 (2.7%) | 125,424 (5.2%) | -0.13 |
| COPD | 207,102 (2.0%) | 60,286 (2.5%) | -0.04 |
| hospitalised - Mental health and substance use | | | |
| Anxiety | 1,032,394 (9.8%) | 281,310 (12%) | -0.06 |
| Depression | 1,549,101 (15%) | 401,815 (17%) | -0.05 |
| Alcohol abuse | 113,440 (1.1%) | 28,362 (1.2%) | -0.01 |
| Cigarette smoking | 3,193,677 (30%) | 756,990 (31%) | -0.02 |
| hospitalised - ADHD and autism | | | |
| ADHD | 39,177 (0.4%) | 10,278 (0.4%) | -0.01 |
| Autism | 39,629 (0.4%) | 13,118 (0.5%) | -0.02 |
| hospitalised - Cardiovascular | | | |
| Hypertension | 1,313,426 (12%) | 311,061 (13%) | -0.01 |
| Coronary artery disease | 690,211 (6.5%) | 172,480 (7.1%) | -0.02 |

[1] Mean and interquartile range is given for Age at index and Follow-up time; all others are counts and percentage.

[2] Standardised mean difference

### Supplementary Table 1: Baseline characteristics

| Characteristic | without eczema[1] | with eczema[1] | SMD[2] |
|---|---|---|---|
| Peripheral artery disease | 83,654 (0.8%) | 23,354 (1.0%) | -0.02 |
| Myocardial infarction | 147,185 (1.4%) | 33,959 (1.4%) | 0.00 |
| Stroke | 120,087 (1.1%) | 30,027 (1.2%) | -0.01 |
| Heart failure | 123,385 (1.2%) | 31,916 (1.3%) | -0.01 |
| Thromboembolic diseases | 123,365 (1.2%) | 35,211 (1.4%) | -0.02 |
| hospitalised - Metabolic | | | |
| Obesity | 401,323 (3.8%) | 114,185 (4.7%) | -0.04 |
| Dyslipidemia | 536,131 (5.1%) | 134,222 (5.5%) | -0.02 |
| Diabetes mellitus | 483,228 (4.6%) | 116,987 (4.8%) | -0.01 |
| Metabolic syndrome | 2,952 (<0.1%) | 867 (<0.1%) | 0.00 |
| hospitalised - Bone health | | | |
| Hip fracture | 52,147 (0.5%) | 12,377 (0.5%) | 0.00 |
| Pelvis fracture | 21,182 (0.2%) | 5,127 (0.2%) | 0.00 |
| Spine fracture | 31,905 (0.3%) | 7,970 (0.3%) | 0.00 |
| Wrist fracture | 293,825 (2.8%) | 69,994 (2.9%) | -0.01 |
| Osteoporosis | 169,971 (1.6%) | 44,606 (1.8%) | -0.02 |
| hospitalised - Skin infection | | | |
| Molluscum contagiosum | 120,247 (1.1%) | 70,284 (2.9%) | -0.12 |
| Impetigo | 349,957 (3.3%) | 168,333 (6.9%) | -0.16 |
| Herpes simplex | 185,290 (1.8%) | 71,661 (2.9%) | -0.08 |
| Dermatophyte infection | 600,933 (5.7%) | 256,233 (11%) | -0.18 |
| Cutaneous warts | 731,181 (6.9%) | 255,819 (11%) | -0.13 |
| hospitalised - Cancer | | | |
| Lung cancer | 11,011 (0.1%) | 2,931 (0.1%) | 0.00 |
| Breast cancer | 91,299 (0.9%) | 20,295 (0.8%) | 0.00 |
| Prostate cancer | 60,101 (0.6%) | 13,798 (0.6%) | 0.00 |
| Pancreatic cancer | 1,624 (<0.1%) | 443 (<0.1%) | 0.00 |
| Non-hodgkin lymphoma | 15,368 (0.1%) | 3,914 (0.2%) | 0.00 |
| Hodgkin lymphoma | 4,018 (<0.1%) | 1,142 (<0.1%) | 0.00 |
| Myeloma | 4,449 (<0.1%) | 946 (<0.1%) | 0.00 |
| CNS cancers | 13,089 (0.1%) | 3,135 (0.1%) | 0.00 |
| Melanoma | 47,025 (0.4%) | 11,468 (0.5%) | 0.00 |
| Nonmelanoma skin cancer | 176,412 (1.7%) | 45,059 (1.9%) | -0.01 |
| hospitalised - Neurological | | | |
| Alzheimer's dementia | 40,947 (0.4%) | 9,603 (0.4%) | 0.00 |
| Vascular dementia | 21,458 (0.2%) | 5,198 (0.2%) | 0.00 |
| Epilepsy | 113,365 (1.1%) | 30,053 (1.2%) | -0.02 |
| Migraine | 434,659 (4.1%) | 122,685 (5.0%) | -0.04 |
| Multiple sclerosis | 17,794 (0.2%) | 4,269 (0.2%) | 0.00 |
| Parkinson's disease | 26,332 (0.2%) | 5,628 (0.2%) | 0.00 |
| Peripheral neuropathies | 703,573 (6.7%) | 193,371 (7.9%) | -0.05 |

[1] Mean and interquartile range is given for Age at index and Follow-up time; all others are counts and percentage.

[2] Standardised mean difference

| Supplementary Table 1: Baseline characteristics | | | |
|---|---|---|---|
| Characteristic | without eczema[1] | with eczema[1] | SMD[2] |
| hospitalised - Digestive system | | | |
| Abdominal hernia | 389,682 (3.7%) | 96,133 (3.9%) | -0.01 |
| Appendicitis | 156,542 (1.5%) | 34,630 (1.4%) | 0.00 |
| Barett's oesophagus | 30,187 (0.3%) | 7,750 (0.3%) | -0.01 |
| Coeliac disease | 27,021 (0.3%) | 8,413 (0.3%) | -0.02 |
| Crohn's disease | 25,284 (0.2%) | 8,472 (0.3%) | -0.02 |
| Diverticular disease | 217,706 (2.1%) | 57,586 (2.4%) | -0.02 |
| Gastritis and duodenitis | 276,680 (2.6%) | 79,448 (3.3%) | -0.04 |
| Gastro oesophageal reflux | 590,227 (5.6%) | 176,166 (7.2%) | -0.07 |
| Irritable bowel syndrome | 351,337 (3.3%) | 105,751 (4.3%) | -0.05 |
| Oesophageal ulcer | 286,158 (2.7%) | 82,937 (3.4%) | -0.04 |
| Pancreatitis | 30,459 (0.3%) | 7,257 (0.3%) | 0.00 |
| Peptic ulcer disease | 79,157 (0.7%) | 19,471 (0.8%) | -0.01 |
| Peritonitis | 11,593 (0.1%) | 2,576 (0.1%) | 0.00 |
| Ulcerative colitis | 36,561 (0.3%) | 10,972 (0.5%) | -0.02 |
| hospitalised - Liver | | | |
| Autoimmune liver disease | 4,843 (<0.1%) | 1,282 (<0.1%) | 0.00 |
| Cholecystitis | 46,403 (0.4%) | 11,282 (0.5%) | 0.00 |
| Fatty liver | 56,315 (0.5%) | 16,509 (0.7%) | -0.02 |
| Fibrosis/sclerosis/cirrhosis | 16,818 (0.2%) | 4,685 (0.2%) | -0.01 |
| Oesophageal varices | 4,173 (<0.1%) | 1,101 (<0.1%) | 0.00 |

[1] Mean and interquartile range is given for Age at index and Follow-up time; all others are counts and percentage.

[2] Standardised mean difference
