## Supplementary Table 2 for "Mapping risks of hospital-recorded health conditions in people with eczema"

Supplementary Table 2: Rates, events, and rate differences from all cohorts

| Outcome | events (overall) | person-years | Crude Rate (per 1,000 person-years) | | | Rate difference (per 1,000 person-years) | | | Hazard ratio (99% confidence interval)[1] | |
|---|---|---|---|---|---|---|---|---|---|---|
| | | | exposed | unexposed (observed) | unexposed (estimated) | observed | crude estimated[2] | adjusted estimated[2] | minimally-adjusted | comorbidity-adjusted |
| any age - A00-B99 | | | | | | | | | | |
| A02 | 1,720 | 114,221,040 | 0.02 | 0.01 | 0.01 | 0.00 | 0.00 | 0.00 | 1.23 [1.05-1.44] | 1.15 [0.98-1.36] |
| A04 | 51,460 | 113,666,050 | 0.51 | 0.44 | 0.41 | 0.08 | 0.11 | 0.09 | 1.27 [1.23-1.31]* | 1.21 [1.17-1.24]* |
| A05 | 520 | 114,263,960 | 0.01 | 0.00 | 0.00 | 0.00 | 0.00 | 0.00 | 1.26 [0.93-1.69] | 1.20 [0.89-1.63] |
| A07 | 970 | 114,252,240 | 0.01 | 0.01 | 0.01 | 0.00 | 0.00 | 0.00 | 1.21 [0.98-1.50] | 1.14 [0.92-1.42] |
| A08 | 62,940 | 112,513,630 | 0.68 | 0.53 | 0.57 | 0.15 | 0.11 | 0.09 | 1.20 [1.17-1.23]* | 1.15 [1.12-1.18]* |
| A09 | 223,490 | 112,582,510 | 2.38 | 1.89 | 1.84 | 0.49 | 0.54 | 0.40 | 1.29 [1.28-1.31]* | 1.20 [1.18-1.22]* |
| A15 | 1,320 | 114,244,380 | 0.01 | 0.01 | 0.01 | 0.00 | 0.00 | 0.00 | 1.09 [0.90-1.32] | 1.06 [0.87-1.29] |
| A16 | 2,620 | 114,216,350 | 0.03 | 0.02 | 0.02 | 0.01 | 0.01 | 0.01 | 1.33 [1.17-1.51]* | 1.28 [1.12-1.46]* |
| A18 | 1,360 | 114,233,310 | 0.01 | 0.01 | 0.01 | 0.00 | 0.00 | 0.00 | 1.19 [0.99-1.42] | 1.14 [0.95-1.38] |
| A31 | 1,100 | 114,260,560 | 0.01 | 0.01 | 0.01 | 0.00 | 0.00 | 0.00 | 1.41 [1.16-1.71]* | 1.26 [1.03-1.54] |
| A38 | 2,000 | 114,245,630 | 0.02 | 0.02 | 0.02 | 0.00 | 0.00 | 0.00 | 1.08 [0.93-1.25] | 1.06 [0.91-1.22] |
| A39 | 2,380 | 114,167,390 | 0.02 | 0.02 | 0.02 | 0.00 | 0.00 | 0.00 | 1.06 [0.92-1.21] | 1.03 [0.90-1.18] |
| A40 | 10,380 | 114,180,090 | 0.11 | 0.09 | 0.08 | 0.02 | 0.03 | 0.02 | 1.33 [1.25-1.43]* | 1.28 [1.20-1.37]* |
| A41 | 195,330 | 113,361,790 | 1.96 | 1.67 | 1.56 | 0.29 | 0.39 | 0.34 | 1.25 [1.23-1.27]* | 1.21 [1.19-1.23]* |
| A46 | 680 | 114,251,130 | 0.01 | 0.01 | 0.00 | 0.00 | 0.01 | 0.01 | 2.16 [1.72-2.72]* | 2.11 [1.67-2.66]* |
| A48 | 1,020 | 114,255,470 | 0.01 | 0.01 | 0.01 | 0.00 | 0.00 | 0.00 | 1.21 [0.98-1.49] | 1.22 [0.99-1.51] |
| A49 | 28,210 | 113,966,200 | 0.30 | 0.24 | 0.22 | 0.06 | 0.08 | 0.07 | 1.34 [1.29-1.40]* | 1.28 [1.23-1.34]* |
| A63 | 940 | 114,178,580 | 0.01 | 0.01 | 0.01 | 0.00 | 0.00 | 0.00 | 1.49 [1.21-1.83]* | 1.35 [1.09-1.67]* |
| A69 | 750 | 114,260,870 | 0.01 | 0.01 | 0.01 | 0.00 | 0.00 | 0.00 | 1.25 [0.99-1.59] | 1.15 [0.90-1.47] |
| A86 | 1,500 | 114,250,770 | 0.01 | 0.01 | 0.01 | 0.00 | 0.00 | 0.00 | 1.01 [0.84-1.21] | 0.98 [0.81-1.17] |
| A87 | 3,700 | 114,154,320 | 0.04 | 0.03 | 0.03 | 0.01 | 0.01 | 0.01 | 1.31 [1.17-1.45]* | 1.20 [1.08-1.34]* |
| B00 | 10,350 | 114,046,030 | 0.18 | 0.07 | 0.07 | 0.11 | 0.11 | 0.10 | 2.52 [2.38-2.67]* | 2.34 [2.21-2.49]* |
| B01 | 9,210 | 114,001,710 | 0.12 | 0.07 | 0.08 | 0.05 | 0.05 | 0.04 | 1.61 [1.52-1.72]* | 1.56 [1.46-1.66]* |
| B02 | 14,520 | 114,138,900 | 0.16 | 0.12 | 0.11 | 0.04 | 0.05 | 0.04 | 1.42 [1.34-1.50]* | 1.34 [1.26-1.42]* |
| B07 | 6,380 | 114,112,930 | 0.07 | 0.05 | 0.05 | 0.02 | 0.02 | 0.02 | 1.41 [1.30-1.53]* | 1.30 [1.20-1.42]* |
| B08 | 3,300 | 114,191,180 | 0.04 | 0.03 | 0.03 | 0.02 | 0.02 | 0.01 | 1.60 [1.44-1.77]* | 1.52 [1.37-1.69]* |
| B09 | 1,460 | 114,221,410 | 0.02 | 0.01 | 0.01 | 0.00 | 0.00 | 0.00 | 1.19 [1.01-1.41] | 1.18 [0.99-1.40] |
| B15 | 800 | 114,254,390 | 0.01 | 0.01 | 0.01 | 0.00 | 0.00 | 0.00 | 1.10 [0.86-1.39] | 1.06 [0.83-1.36] |
| B16 | 1,380 | 114,232,470 | 0.01 | 0.01 | 0.01 | 0.00 | 0.00 | 0.00 | 1.34 [1.12-1.59]* | 1.31 [1.09-1.56]* |
| B17 | 2,140 | 114,224,270 | 0.02 | 0.02 | 0.02 | 0.00 | 0.01 | 0.01 | 1.39 [1.21-1.60]* | 1.33 [1.15-1.53]* |
| B18 | 8,820 | 114,110,750 | 0.08 | 0.08 | 0.07 | 0.01 | 0.01 | 0.01 | 1.19 [1.11-1.28]* | 1.16 [1.08-1.25]* |
| B19 | 560 | 114,260,820 | 0.01 | 0.00 | 0.00 | 0.00 | 0.00 | 0.00 | 1.16 [0.87-1.54] | 1.07 [0.80-1.43] |
| B25 | 1,720 | 114,247,760 | 0.02 | 0.01 | 0.01 | 0.00 | 0.01 | 0.00 | 1.41 [1.21-1.64]* | 1.35 [1.15-1.58]* |
| B26 | 640 | 114,257,510 | 0.01 | 0.01 | 0.01 | 0.00 | 0.00 | 0.00 | 1.20 [0.92-1.56] | 1.21 [0.92-1.58] |
| B27 | 6,690 | 114,156,630 | 0.07 | 0.06 | 0.06 | 0.02 | 0.01 | 0.01 | 1.15 [1.06-1.24]* | 1.10 [1.01-1.19] |
| B30 | 650 | 114,257,500 | 0.01 | 0.01 | 0.01 | 0.00 | 0.00 | 0.00 | 1.26 [0.98-1.62] | 1.16 [0.89-1.50] |
| B33 | 470 | 114,259,950 | 0.00 | 0.00 | 0.00 | 0.00 | 0.00 | 0.00 | 1.24 [0.93-1.66] | 1.22 [0.91-1.64] |
| B34 | 138,970 | 110,561,040 | 1.89 | 1.11 | 1.23 | 0.78 | 0.66 | 0.61 | 1.54 [1.52-1.57]* | 1.48 [1.45-1.50]* |
| B35 | 6,180 | 114,201,260 | 0.08 | 0.05 | 0.05 | 0.03 | 0.03 | 0.03 | 1.72 [1.58-1.86]* | 1.58 [1.45-1.71]* |
| B36 | 2,280 | 114,252,190 | 0.03 | 0.02 | 0.02 | 0.01 | 0.01 | 0.01 | 1.87 [1.63-2.14]* | 1.72 [1.50-1.97]* |
| B37 | 81,580 | 113,457,400 | 0.87 | 0.68 | 0.64 | 0.18 | 0.23 | 0.17 | 1.36 [1.33-1.39]* | 1.25 [1.22-1.28]* |
| B44 | 3,180 | 114,242,930 | 0.04 | 0.03 | 0.02 | 0.01 | 0.02 | 0.01 | 1.69 [1.51-1.88]* | 1.40 [1.24-1.57]* |
| B49 | 1,890 | 114,256,280 | 0.02 | 0.02 | 0.01 | 0.01 | 0.01 | 0.01 | 1.63 [1.41-1.88]* | 1.54 [1.33-1.79]* |
| B50 | 1,050 | 114,239,870 | 0.01 | 0.01 | 0.01 | 0.00 | 0.00 | 0.00 | 0.95 [0.76-1.19] | 0.97 [0.77-1.22] |
| B59 | 940 | 114,264,600 | 0.01 | 0.01 | 0.01 | 0.00 | 0.00 | 0.00 | 1.53 [1.24-1.88]* | 1.44 [1.16-1.78]* |
| B80 | 2,610 | 114,229,070 | 0.03 | 0.02 | 0.02 | 0.01 | 0.00 | 0.00 | 1.21 [1.06-1.37]* | 1.14 [1.00-1.30] |
| B85 | 1,080 | 114,251,810 | 0.01 | 0.01 | 0.01 | 0.00 | 0.00 | 0.00 | 1.12 [0.91-1.37] | 1.11 [0.90-1.36] |
| B86 | 1,750 | 114,232,630 | 0.03 | 0.01 | 0.01 | 0.02 | 0.02 | 0.02 | 2.53 [2.19-2.93]* | 2.49 [2.15-2.89]* |
| B90 | 740 | 114,262,250 | 0.01 | 0.01 | 0.01 | 0.00 | 0.00 | 0.00 | 1.60 [1.25-2.03]* | 1.46 [1.14-1.87]* |
| B91 | 1,180 | 114,252,480 | 0.01 | 0.01 | 0.01 | 0.00 | 0.00 | 0.00 | 1.30 [1.07-1.58]* | 1.30 [1.06-1.58] |
| B94 | 2,030 | 114,246,300 | 0.02 | 0.02 | 0.02 | 0.00 | 0.01 | 0.00 | 1.32 [1.14-1.52]* | 1.19 [1.02-1.39] |
| B95 | 118,310 | 112,611,290 | 1.37 | 0.97 | 0.91 | 0.40 | 0.46 | 0.42 | 1.51 [1.48-1.54]* | 1.44 [1.42-1.47]* |
| B96 | 235,480 | 111,557,930 | 2.38 | 2.05 | 1.94 | 0.34 | 0.44 | 0.35 | 1.23 [1.21-1.24]* | 1.17 [1.16-1.19]* |
| B97 | 64,700 | 113,345,550 | 0.72 | 0.54 | 0.54 | 0.18 | 0.17 | 0.14 | 1.32 [1.29-1.35]* | 1.23 [1.20-1.27]* |
| B98 | 23,060 | 114,109,250 | 0.22 | 0.20 | 0.19 | 0.03 | 0.04 | 0.03 | 1.20 [1.14-1.25]* | 1.14 [1.09-1.19]* |
| B99 | 6,520 | 114,237,370 | 0.07 | 0.06 | 0.05 | 0.01 | 0.01 | 0.01 | 1.24 [1.14-1.36]* | 1.20 [1.10-1.31]* |
| any age - C00-D49 | | | | | | | | | | |
| C00 | 780 | 114,260,080 | 0.01 | 0.01 | 0.01 | 0.00 | 0.00 | 0.00 | 1.04 [0.81-1.35] | 1.03 [0.80-1.33] |
| C01 | 1,640 | 114,258,730 | 0.01 | 0.01 | 0.01 | 0.00 | 0.00 | 0.00 | 1.08 [0.91-1.28] | 1.08 [0.90-1.28] |
| C02 | 2,530 | 114,243,900 | 0.02 | 0.02 | 0.02 | 0.00 | 0.01 | 0.01 | 1.28 [1.13-1.46]* | 1.27 [1.11-1.45]* |
| C03 | 580 | 114,266,480 | 0.01 | 0.00 | 0.00 | 0.00 | 0.00 | 0.00 | 1.47 [1.12-1.92]* | 1.46 [1.11-1.91]* |
| C04 | 750 | 114,261,650 | 0.01 | 0.01 | 0.01 | 0.00 | 0.00 | 0.00 | 1.13 [0.88-1.45] | 1.14 [0.89-1.47] |
| C05 | 820 | 114,262,400 | 0.01 | 0.01 | 0.01 | 0.00 | 0.00 | 0.00 | 1.28 [1.01-1.62] | 1.29 [1.02-1.64] |
| C06 | 1,170 | 114,260,580 | 0.01 | 0.01 | 0.01 | 0.00 | 0.00 | 0.00 | 1.25 [1.03-1.52] | 1.24 [1.01-1.51] |
| C07 | 1,180 | 114,257,890 | 0.01 | 0.01 | 0.01 | 0.00 | 0.00 | 0.00 | 1.03 [0.84-1.27] | 1.02 [0.83-1.25] |
| C09 | 2,110 | 114,249,450 | 0.02 | 0.02 | 0.02 | 0.00 | 0.00 | 0.00 | 1.07 [0.92-1.25] | 1.04 [0.89-1.22] |
| C10 | 1,070 | 114,264,510 | 0.01 | 0.01 | 0.01 | 0.00 | 0.00 | 0.00 | 1.11 [0.90-1.37] | 1.09 [0.88-1.35] |
| C11 | 560 | 114,265,080 | 0.01 | 0.00 | 0.00 | 0.00 | 0.00 | 0.00 | 1.19 [0.90-1.58] | 1.19 [0.89-1.58] |
| C12 | 580 | 114,267,020 | 0.01 | 0.00 | 0.00 | 0.00 | 0.00 | 0.00 | 1.24 [0.94-1.63] | 1.25 [0.94-1.65] |
| C13 | 650 | 114,268,040 | 0.01 | 0.01 | 0.01 | 0.00 | 0.00 | 0.00 | 1.16 [0.89-1.52] | 1.20 [0.91-1.57] |
| C14 | 1,210 | 114,264,160 | 0.01 | 0.01 | 0.01 | 0.00 | 0.00 | 0.00 | 1.10 [0.90-1.34] | 1.08 [0.88-1.33] |
| C15 | 16,200 | 114,213,050 | 0.15 | 0.14 | 0.13 | 0.01 | 0.02 | 0.02 | 1.18 [1.12-1.25]* | 1.17 [1.11-1.24]* |
| C16 | 13,250 | 114,215,520 | 0.11 | 0.12 | 0.11 | 0.00 | 0.01 | 0.01 | 1.08 [1.01-1.15] | 1.07 [1.01-1.14] |
| C17 | 2,680 | 114,255,610 | 0.02 | 0.02 | 0.02 | 0.00 | 0.00 | 0.00 | 1.11 [0.98-1.27] | 1.10 [0.96-1.26] |
| C18 | 45,750 | 113,878,370 | 0.38 | 0.41 | 0.37 | −0.03 | 0.01 | 0.00 | 1.01 [0.98-1.05] | 1.00 [0.97-1.04] |
| C19 | 9,480 | 114,197,600 | 0.07 | 0.09 | 0.08 | −0.01 | 0.00 | 0.00 | 0.95 [0.88-1.02] | 0.95 [0.88-1.02] |
| C20 | 19,640 | 114,069,270 | 0.15 | 0.18 | 0.16 | −0.02 | −0.01 | 0.00 | 0.97 [0.92-1.02] | 0.98 [0.93-1.03] |
| C21 | 2,440 | 114,245,640 | 0.02 | 0.02 | 0.02 | 0.00 | 0.00 | 0.00 | 1.08 [0.94-1.24] | 1.07 [0.93-1.23] |
| C22 | 9,010 | 114,248,510 | 0.09 | 0.08 | 0.07 | 0.01 | 0.02 | 0.02 | 1.25 [1.16-1.34]* | 1.21 [1.13-1.31]* |
| C23 | 1,320 | 114,268,220 | 0.01 | 0.01 | 0.01 | 0.00 | 0.00 | 0.00 | 1.07 [0.88-1.31] | 1.07 [0.88-1.30] |
| C24 | 2,450 | 114,261,970 | 0.02 | 0.02 | 0.02 | 0.00 | 0.00 | 0.00 | 1.09 [0.94-1.25] | 1.08 [0.94-1.25] |
| C25 | 15,190 | 114,243,700 | 0.14 | 0.13 | 0.12 | 0.01 | 0.02 | 0.02 | 1.16 [1.10-1.23]* | 1.15 [1.09-1.22]* |
| C26 | 3,390 | 114,256,370 | 0.03 | 0.03 | 0.03 | 0.00 | 0.00 | 0.00 | 1.13 [1.00-1.28] | 1.10 [0.98-1.25] |
| C30 | 570 | 114,264,750 | 0.00 | 0.01 | 0.00 | 0.00 | 0.00 | 0.00 | 1.07 [0.80-1.43] | 1.06 [0.79-1.42] |
| C32 | 3,540 | 114,230,960 | 0.03 | 0.03 | 0.03 | 0.00 | 0.00 | 0.00 | 1.14 [1.02-1.28] | 1.16 [1.03-1.30] |
| C34 | 65,460 | 114,089,990 | 0.60 | 0.57 | 0.52 | 0.03 | 0.08 | 0.07 | 1.15 [1.12-1.18]* | 1.14 [1.11-1.17]* |
| C38 | 880 | 114,266,870 | 0.01 | 0.01 | 0.01 | 0.00 | 0.00 | 0.00 | 1.07 [0.85-1.35] | 1.08 [0.85-1.37] |
| C40 | 690 | 114,261,970 | 0.01 | 0.01 | 0.01 | 0.00 | 0.00 | 0.00 | 1.05 [0.80-1.37] | 1.02 [0.78-1.34] |
| C41 | 1,890 | 114,254,810 | 0.02 | 0.02 | 0.01 | 0.00 | 0.00 | 0.00 | 1.25 [1.07-1.46]* | 1.24 [1.06-1.46]* |
| C43 | 18,240 | 114,055,240 | 0.14 | 0.16 | 0.15 | −0.02 | −0.01 | −0.01 | 0.95 [0.90-1.01] | 0.94 [0.89-1.00] |
| C44 | 129,100 | 112,713,740 | 1.16 | 1.14 | 1.05 | 0.01 | 0.10 | 0.07 | 1.10 [1.08-1.12]* | 1.07 [1.05-1.09]* |
| C45 | 4,100 | 114,262,400 | 0.03 | 0.04 | 0.03 | 0.00 | 0.00 | 0.00 | 0.98 [0.88-1.10] | 0.99 [0.89-1.11] |
| C48 | 2,300 | 114,260,420 | 0.02 | 0.02 | 0.02 | 0.00 | 0.00 | 0.00 | 1.05 [0.90-1.21] | 1.03 [0.89-1.20] |
| C49 | 4,240 | 114,229,950 | 0.04 | 0.04 | 0.03 | 0.00 | 0.00 | 0.00 | 1.08 [0.97-1.20] | 1.06 [0.95-1.18] |
| C50 | 77,280 | 113,050,660 | 0.66 | 0.69 | 0.63 | −0.03 | 0.03 | 0.02 | 1.04 [1.02-1.07]* | 1.04 [1.01-1.07]* |
| C51 | 2,060 | 114,247,150 | 0.02 | 0.02 | 0.02 | 0.00 | 0.00 | 0.00 | 1.20 [1.04-1.39] | 1.18 [1.02-1.38] |
| C52 | 650 | 114,266,240 | 0.01 | 0.01 | 0.01 | 0.00 | 0.00 | 0.00 | 0.98 [0.74-1.29] | 1.00 [0.76-1.33] |
| C53 | 4,290 | 114,206,450 | 0.03 | 0.04 | 0.04 | 0.00 | 0.00 | 0.00 | 0.96 [0.86-1.07] | 0.97 [0.87-1.09] |

| | | | | | | | | | | |
|---|---|---|---|---|---|---|---|---|---|---|
| C54 | 12,680 | 114,108,260 | 0.11 | 0.11 | 0.10 | 0.00 | 0.00 | 0.01 | 1.04 [0.98-1.11] | 1.06 [0.99-1.12] |
| C55 | 1,740 | 114,256,580 | 0.01 | 0.02 | 0.01 | 0.00 | 0.00 | 0.00 | 1.06 [0.90-1.25] | 1.05 [0.88-1.24] |
| C56 | 11,460 | 114,171,690 | 0.10 | 0.10 | 0.09 | −0.01 | 0.00 | 0.00 | 1.03 [0.97-1.10] | 1.04 [0.97-1.11] |
| C57 | 1,270 | 114,264,240 | 0.01 | 0.01 | 0.01 | 0.00 | 0.00 | 0.00 | 1.18 [0.97-1.42] | 1.20 [0.99-1.46] |
| C60 | 980 | 114,258,200 | 0.01 | 0.01 | 0.01 | 0.00 | 0.00 | 0.00 | 1.19 [0.95-1.48] | 1.20 [0.96-1.50] |
| C61 | 72,980 | 113,592,100 | 0.63 | 0.65 | 0.59 | −0.02 | 0.03 | 0.02 | 1.06 [1.03-1.09]* | 1.04 [1.01-1.06]* |
| C62 | 2,020 | 114,222,460 | 0.02 | 0.02 | 0.02 | 0.00 | 0.00 | 0.00 | 1.04 [0.89-1.22] | 1.02 [0.87-1.20] |
| C64 | 15,190 | 114,147,330 | 0.13 | 0.13 | 0.12 | 0.00 | 0.01 | 0.01 | 1.11 [1.05-1.18]* | 1.09 [1.03-1.15]* |
| C65 | 1,340 | 114,260,100 | 0.01 | 0.01 | 0.01 | 0.00 | 0.00 | 0.00 | 1.22 [1.01-1.47] | 1.24 [1.02-1.50] |
| C66 | 1,870 | 114,257,520 | 0.02 | 0.02 | 0.01 | 0.00 | 0.00 | 0.00 | 1.10 [0.93-1.29] | 1.09 [0.93-1.29] |
| C67 | 33,740 | 113,869,260 | 0.29 | 0.30 | 0.27 | 0.00 | 0.02 | 0.02 | 1.08 [1.04-1.12]* | 1.07 [1.03-1.12]* |
| C68 | 720 | 114,264,870 | 0.01 | 0.01 | 0.01 | 0.00 | 0.00 | 0.00 | 1.03 [0.79-1.35] | 1.02 [0.78-1.33] |
| C69 | 1,400 | 114,245,660 | 0.01 | 0.01 | 0.01 | 0.00 | 0.00 | 0.00 | 0.98 [0.81-1.18] | 0.96 [0.79-1.16] |
| C71 | 8,490 | 114,216,660 | 0.08 | 0.07 | 0.07 | 0.00 | 0.01 | 0.01 | 1.12 [1.04-1.21]* | 1.11 [1.03-1.19]* |
| C72 | 540 | 114,263,370 | 0.01 | 0.00 | 0.00 | 0.00 | 0.00 | 0.00 | 1.20 [0.90-1.59] | 1.11 [0.82-1.49] |
| C73 | 4,310 | 114,206,790 | 0.04 | 0.04 | 0.04 | 0.00 | 0.00 | 0.00 | 1.06 [0.96-1.18] | 1.04 [0.93-1.16] |
| C74 | 690 | 114,262,330 | 0.01 | 0.01 | 0.01 | 0.00 | 0.00 | 0.00 | 1.20 [0.93-1.55] | 1.19 [0.92-1.54] |
| C75 | 720 | 114,263,440 | 0.01 | 0.01 | 0.01 | 0.00 | 0.00 | 0.00 | 1.07 [0.83-1.39] | 1.04 [0.80-1.36] |
| C76 | 2,400 | 114,252,700 | 0.02 | 0.02 | 0.02 | 0.00 | 0.00 | 0.00 | 1.10 [0.96-1.27] | 1.11 [0.96-1.28] |
| C77 | 75,990 | 113,728,310 | 0.65 | 0.67 | 0.61 | −0.02 | 0.04 | 0.04 | 1.06 [1.03-1.09]* | 1.06 [1.04-1.09]* |
| C78 | 111,980 | 114,049,850 | 0.96 | 0.99 | 0.90 | −0.03 | 0.06 | 0.06 | 1.06 [1.04-1.09]* | 1.07 [1.04-1.09]* |
| C79 | 94,520 | 114,075,420 | 0.82 | 0.83 | 0.76 | −0.02 | 0.06 | 0.06 | 1.08 [1.05-1.10]* | 1.08 [1.06-1.11]* |
| C80 | 33,220 | 114,186,460 | 0.29 | 0.29 | 0.26 | 0.00 | 0.03 | 0.03 | 1.10 [1.06-1.15]* | 1.10 [1.06-1.14]* |
| C81 | 3,570 | 114,211,100 | 0.04 | 0.03 | 0.03 | 0.02 | 0.02 | 0.02 | 1.71 [1.54-1.90]* | 1.64 [1.48-1.83]* |
| C82 | 4,030 | 114,221,070 | 0.04 | 0.04 | 0.03 | 0.00 | 0.00 | 0.00 | 1.16 [1.04-1.29]* | 1.10 [0.99-1.23] |
| C83 | 11,500 | 114,181,920 | 0.11 | 0.10 | 0.09 | 0.01 | 0.02 | 0.02 | 1.24 [1.17-1.32]* | 1.21 [1.13-1.29]* |
| C84 | 1,600 | 114,255,930 | 0.02 | 0.01 | 0.01 | 0.01 | 0.01 | 0.01 | 2.44 [2.11-2.83]* | 2.41 [2.08-2.80]* |
| C85 | 14,480 | 114,111,140 | 0.14 | 0.12 | 0.11 | 0.02 | 0.04 | 0.03 | 1.33 [1.25-1.40]* | 1.28 [1.21-1.35]* |
| C88 | 1,900 | 114,257,780 | 0.02 | 0.02 | 0.01 | 0.00 | 0.00 | 0.00 | 1.28 [1.10-1.50]* | 1.26 [1.08-1.48]* |
| C90 | 10,050 | 114,204,890 | 0.09 | 0.09 | 0.08 | 0.00 | 0.01 | 0.01 | 1.13 [1.06-1.22]* | 1.11 [1.03-1.19]* |
| C91 | 10,940 | 114,159,750 | 0.10 | 0.10 | 0.09 | 0.00 | 0.01 | 0.01 | 1.07 [1.00-1.15] | 1.05 [0.99-1.13] |
| C92 | 8,090 | 114,221,140 | 0.08 | 0.07 | 0.06 | 0.01 | 0.02 | 0.02 | 1.28 [1.18-1.38]* | 1.25 [1.16-1.35]* |
| C93 | 1,040 | 114,268,110 | 0.01 | 0.01 | 0.01 | 0.00 | 0.00 | 0.00 | 1.17 [0.94-1.46] | 1.19 [0.95-1.48] |
| C94 | 490 | 114,268,490 | 0.00 | 0.00 | 0.00 | 0.00 | 0.00 | 0.00 | 1.25 [0.92-1.70] | 1.24 [0.91-1.70] |
| C95 | 1,800 | 114,258,090 | 0.02 | 0.02 | 0.01 | 0.00 | 0.00 | 0.00 | 1.22 [1.04-1.43] | 1.19 [1.02-1.40] |
| C97 | 8,550 | 114,245,010 | 0.07 | 0.08 | 0.07 | 0.00 | 0.00 | 0.00 | 1.06 [0.98-1.15] | 1.04 [0.96-1.12] |
| D00 | 940 | 114,261,430 | 0.01 | 0.01 | 0.01 | 0.00 | 0.00 | 0.00 | 1.09 [0.87-1.36] | 1.05 [0.84-1.32] |
| D01 | 950 | 114,259,340 | 0.01 | 0.01 | 0.01 | 0.00 | 0.00 | 0.00 | 1.10 [0.88-1.38] | 1.04 [0.82-1.30] |
| D02 | 940 | 114,261,100 | 0.01 | 0.01 | 0.01 | 0.00 | 0.00 | 0.00 | 1.27 [1.02-1.57] | 1.23 [0.99-1.54] |
| D03 | 6,960 | 114,184,000 | 0.06 | 0.06 | 0.06 | 0.00 | 0.00 | 0.00 | 1.06 [0.97-1.15] | 1.02 [0.94-1.11] |
| D04 | 14,400 | 114,128,590 | 0.14 | 0.12 | 0.11 | 0.02 | 0.03 | 0.02 | 1.25 [1.18-1.33]* | 1.19 [1.12-1.27]* |
| D05 | 12,660 | 114,042,010 | 0.10 | 0.11 | 0.10 | −0.01 | 0.00 | 0.00 | 1.01 [0.95-1.07] | 0.99 [0.93-1.06] |
| D06 | 10,930 | 113,899,090 | 0.09 | 0.10 | 0.09 | 0.00 | 0.00 | 0.00 | 1.05 [0.98-1.12] | 1.03 [0.96-1.11] |
| D07 | 3,930 | 114,205,110 | 0.04 | 0.03 | 0.03 | 0.00 | 0.01 | 0.00 | 1.20 [1.08-1.33]* | 1.14 [1.02-1.28] |
| D09 | 3,550 | 114,234,110 | 0.03 | 0.03 | 0.03 | 0.00 | 0.00 | 0.00 | 1.12 [1.00-1.26] | 1.11 [0.98-1.25] |
| D10 | 4,090 | 114,181,840 | 0.04 | 0.03 | 0.03 | 0.01 | 0.01 | 0.01 | 1.27 [1.15-1.41]* | 1.21 [1.09-1.35]* |
| D11 | 4,380 | 114,187,160 | 0.04 | 0.04 | 0.04 | 0.00 | 0.00 | 0.00 | 1.04 [0.94-1.16] | 1.04 [0.93-1.16] |
| D12 | 123,720 | 112,693,960 | 1.18 | 1.08 | 0.98 | 0.11 | 0.21 | 0.15 | 1.21 [1.19-1.24]* | 1.14 [1.12-1.16]* |
| D13 | 8,410 | 114,058,360 | 0.08 | 0.07 | 0.07 | 0.01 | 0.01 | 0.01 | 1.22 [1.13-1.31]* | 1.12 [1.04-1.21]* |
| D14 | 2,760 | 114,213,450 | 0.03 | 0.02 | 0.02 | 0.00 | 0.01 | 0.01 | 1.31 [1.15-1.48]* | 1.23 [1.08-1.40]* |
| D15 | 1,060 | 114,258,150 | 0.01 | 0.01 | 0.01 | 0.00 | 0.00 | 0.00 | 1.15 [0.93-1.42] | 1.14 [0.92-1.41] |
| D16 | 4,960 | 114,176,090 | 0.05 | 0.04 | 0.04 | 0.01 | 0.01 | 0.01 | 1.18 [1.07-1.29]* | 1.14 [1.03-1.25]* |
| D17 | 39,130 | 113,238,760 | 0.36 | 0.34 | 0.31 | 0.02 | 0.05 | 0.03 | 1.16 [1.12-1.20]* | 1.10 [1.07-1.14]* |
| D18 | 17,970 | 113,893,460 | 0.17 | 0.15 | 0.15 | 0.02 | 0.02 | 0.02 | 1.17 [1.11-1.23]* | 1.11 [1.05-1.17]* |
| D21 | 4,320 | 114,179,870 | 0.04 | 0.04 | 0.03 | 0.01 | 0.01 | 0.01 | 1.28 [1.16-1.42]* | 1.23 [1.11-1.36]* |
| D22 | 40,170 | 112,843,030 | 0.38 | 0.35 | 0.33 | 0.03 | 0.05 | 0.03 | 1.14 [1.10-1.18]* | 1.09 [1.05-1.13]* |
| D23 | 26,120 | 113,493,750 | 0.26 | 0.22 | 0.21 | 0.04 | 0.05 | 0.04 | 1.23 [1.18-1.28]* | 1.17 [1.13-1.22]* |
| D24 | 13,600 | 113,927,600 | 0.12 | 0.12 | 0.11 | 0.00 | 0.01 | 0.00 | 1.05 [0.99-1.11] | 1.01 [0.95-1.07] |
| D25 | 64,900 | 112,701,380 | 0.58 | 0.58 | 0.54 | 0.00 | 0.04 | 0.02 | 1.07 [1.04-1.10]* | 1.04 [1.01-1.07]* |
| D26 | 2,140 | 114,210,440 | 0.02 | 0.02 | 0.02 | 0.00 | 0.00 | 0.00 | 1.12 [0.96-1.29] | 1.09 [0.94-1.27] |
| D27 | 15,320 | 113,980,960 | 0.13 | 0.13 | 0.13 | 0.00 | 0.00 | 0.00 | 1.04 [0.98-1.10] | 1.02 [0.96-1.08] |
| D28 | 1,610 | 114,229,980 | 0.02 | 0.01 | 0.01 | 0.00 | 0.00 | 0.00 | 1.25 [1.06-1.47]* | 1.19 [1.00-1.41] |
| D29 | 1,270 | 114,248,200 | 0.01 | 0.01 | 0.01 | 0.00 | 0.00 | 0.00 | 1.23 [1.02-1.49] | 1.19 [0.98-1.45] |
| D30 | 2,350 | 114,239,560 | 0.02 | 0.02 | 0.02 | 0.00 | 0.00 | 0.00 | 1.11 [0.96-1.28] | 1.08 [0.93-1.25] |
| D31 | 2,530 | 114,234,340 | 0.03 | 0.02 | 0.02 | 0.00 | 0.01 | 0.01 | 1.28 [1.12-1.46]* | 1.25 [1.09-1.43]* |
| D32 | 8,570 | 114,191,750 | 0.08 | 0.07 | 0.07 | 0.00 | 0.01 | 0.01 | 1.13 [1.05-1.22]* | 1.09 [1.01-1.18] |
| D33 | 3,400 | 114,221,500 | 0.03 | 0.03 | 0.03 | 0.00 | 0.00 | 0.00 | 1.13 [1.00-1.27] | 1.05 [0.93-1.18] |
| D34 | 2,270 | 114,226,280 | 0.02 | 0.02 | 0.02 | 0.00 | 0.00 | 0.00 | 1.07 [0.93-1.24] | 1.03 [0.88-1.19] |
| D35 | 12,650 | 114,117,610 | 0.11 | 0.11 | 0.10 | 0.00 | 0.01 | 0.01 | 1.13 [1.06-1.20]* | 1.08 [1.02-1.15] |
| D36 | 6,150 | 114,135,610 | 0.06 | 0.05 | 0.05 | 0.01 | 0.01 | 0.01 | 1.22 [1.12-1.33]* | 1.14 [1.04-1.24]* |
| D37 | 16,960 | 114,098,180 | 0.15 | 0.15 | 0.14 | 0.00 | 0.01 | 0.01 | 1.10 [1.04-1.16]* | 1.07 [1.01-1.13] |
| D38 | 4,000 | 114,241,040 | 0.04 | 0.03 | 0.03 | 0.00 | 0.01 | 0.01 | 1.22 [1.10-1.36]* | 1.20 [1.08-1.34]* |
| D39 | 2,470 | 114,228,430 | 0.02 | 0.02 | 0.02 | 0.00 | 0.00 | 0.00 | 1.06 [0.92-1.22] | 1.06 [0.92-1.22] |
| D40 | 830 | 114,255,000 | 0.01 | 0.01 | 0.01 | 0.00 | 0.00 | 0.00 | 1.12 [0.88-1.43] | 1.11 [0.87-1.43] |
| D41 | 11,860 | 114,134,340 | 0.10 | 0.10 | 0.10 | 0.00 | 0.01 | 0.01 | 1.09 [1.02-1.16]* | 1.07 [1.00-1.15] |
| D43 | 4,130 | 114,235,150 | 0.04 | 0.04 | 0.03 | 0.00 | 0.00 | 0.00 | 1.13 [1.02-1.26] | 1.11 [0.99-1.23] |
| D44 | 2,470 | 114,232,440 | 0.02 | 0.02 | 0.02 | 0.00 | 0.00 | 0.00 | 1.17 [1.02-1.34] | 1.10 [0.96-1.27] |
| D45 | 4,420 | 114,199,650 | 0.04 | 0.04 | 0.03 | 0.00 | 0.00 | 0.00 | 1.10 [0.99-1.23] | 1.09 [0.98-1.21] |
| D46 | 10,260 | 114,218,650 | 0.10 | 0.09 | 0.08 | 0.02 | 0.03 | 0.02 | 1.32 [1.24-1.42]* | 1.29 [1.20-1.38]* |
| D47 | 16,650 | 114,171,230 | 0.17 | 0.14 | 0.13 | 0.03 | 0.03 | 0.03 | 1.26 [1.20-1.33]* | 1.20 [1.13-1.26]* |
| D48 | 8,690 | 114,113,920 | 0.08 | 0.08 | 0.07 | 0.00 | 0.01 | 0.00 | 1.09 [1.01-1.17] | 1.05 [0.97-1.13] |
| any age - D50-D89 | | | | | | | | | | |
| D50 | 231,860 | 112,083,610 | 2.46 | 1.97 | 1.86 | 0.49 | 0.60 | 0.49 | 1.32 [1.31-1.34]* | 1.25 [1.23-1.27]* |
| D51 | 20,450 | 114,061,640 | 0.21 | 0.17 | 0.16 | 0.04 | 0.06 | 0.05 | 1.38 [1.32-1.44]* | 1.29 [1.23-1.35]* |
| D52 | 15,540 | 114,194,900 | 0.16 | 0.13 | 0.12 | 0.03 | 0.03 | 0.03 | 1.28 [1.21-1.35]* | 1.23 [1.17-1.31]* |
| D53 | 4,190 | 114,241,410 | 0.05 | 0.03 | 0.03 | 0.01 | 0.01 | 0.01 | 1.42 [1.28-1.58]* | 1.34 [1.20-1.49]* |
| D55 | 700 | 114,258,530 | 0.01 | 0.01 | 0.01 | 0.00 | 0.00 | 0.00 | 1.50 [1.17-1.91]* | 1.47 [1.14-1.89]* |
| D56 | 8,860 | 114,122,140 | 0.10 | 0.07 | 0.07 | 0.03 | 0.03 | 0.03 | 1.40 [1.31-1.50]* | 1.37 [1.27-1.47]* |
| D57 | 10,180 | 114,052,440 | 0.12 | 0.08 | 0.09 | 0.03 | 0.03 | 0.03 | 1.31 [1.23-1.40]* | 1.28 [1.20-1.37]* |
| D58 | 2,470 | 114,231,070 | 0.02 | 0.02 | 0.02 | 0.00 | 0.00 | 0.00 | 1.21 [1.05-1.38]* | 1.19 [1.04-1.37] |
| D59 | 3,080 | 114,232,240 | 0.03 | 0.03 | 0.02 | 0.01 | 0.01 | 0.01 | 1.44 [1.28-1.62]* | 1.42 [1.26-1.59]* |
| D61 | 14,570 | 114,194,810 | 0.15 | 0.12 | 0.11 | 0.03 | 0.04 | 0.03 | 1.31 [1.24-1.39]* | 1.27 [1.20-1.35]* |
| D62 | 7,010 | 114,145,740 | 0.07 | 0.06 | 0.06 | 0.00 | 0.01 | 0.01 | 1.13 [1.04-1.23]* | 1.12 [1.03-1.22]* |
| D63 | 22,310 | 114,142,120 | 0.22 | 0.19 | 0.17 | 0.03 | 0.05 | 0.04 | 1.28 [1.23-1.34]* | 1.24 [1.19-1.30]* |
| D64 | 308,600 | 111,309,680 | 3.23 | 2.66 | 2.50 | 0.57 | 0.74 | 0.63 | 1.30 [1.28-1.31]* | 1.24 [1.23-1.26]* |
| D65 | 1,060 | 114,264,970 | 0.01 | 0.01 | 0.01 | 0.00 | 0.00 | 0.00 | 1.19 [0.96-1.46] | 1.17 [0.94-1.44] |
| D66 | 1,140 | 114,244,450 | 0.01 | 0.01 | 0.01 | 0.00 | 0.00 | 0.00 | 1.15 [0.94-1.40] | 1.11 [0.90-1.36] |
| D68 | 17,780 | 114,016,940 | 0.18 | 0.15 | 0.14 | 0.03 | 0.04 | 0.03 | 1.29 [1.22-1.35]* | 1.19 [1.13-1.25]* |
| D69 | 53,400 | 113,746,430 | 0.54 | 0.45 | 0.44 | 0.09 | 0.10 | 0.09 | 1.23 [1.20-1.27]* | 1.20 [1.16-1.23]* |
| D70 | 45,520 | 113,893,820 | 0.45 | 0.39 | 0.36 | 0.06 | 0.08 | 0.08 | 1.23 [1.19-1.27]* | 1.20 [1.16-1.24]* |
| D72 | 9,910 | 114,193,710 | 0.11 | 0.08 | 0.08 | 0.03 | 0.03 | 0.03 | 1.43 [1.34-1.53]* | 1.31 [1.22-1.40]* |
| D73 | 10,330 | 114,177,740 | 0.10 | 0.09 | 0.08 | 0.01 | 0.02 | 0.02 | 1.25 [1.17-1.34]* | 1.20 [1.12-1.28]* |
| D75 | 12,720 | 114,139,920 | 0.12 | 0.11 | 0.10 | 0.01 | 0.02 | 0.02 | 1.21 [1.14-1.29]* | 1.17 [1.10-1.24]* |
| D76 | 940 | 114,250,730 | 0.01 | 0.01 | 0.01 | 0.00 | 0.00 | 0.00 | 1.27 [1.02-1.57] | 1.20 [0.97-1.50] |
| D80 | 2,970 | 114,229,710 | 0.04 | 0.02 | 0.02 | 0.01 | 0.01 | 0.01 | 1.54 [1.37-1.73]* | 1.36 [1.20-1.53]* |
| D82 | 790 | 114,256,460 | 0.01 | 0.01 | 0.01 | 0.00 | 0.00 | 0.00 | 1.31 [1.04-1.65] | 1.19 [0.94-1.51] |
| D83 | 360 | 114,264,620 | 0.01 | 0.00 | 0.00 | 0.00 | 0.00 | 0.00 | 2.01 [1.46-2.75]* | 1.76 [1.27-2.43]* |
| D84 | 1,610 | 114,247,990 | 0.02 | 0.01 | 0.01 | 0.01 | 0.01 | 0.01 | 1.71 [1.47-1.99]* | 1.43 [1.22-1.69]* |

| | | | | | | | | | | |
|---|---|---|---|---|---|---|---|---|---|---|
| D86 | 6,350 | 114,175,010 | 0.07 | 0.05 | 0.05 | 0.02 | 0.03 | 0.02 | 1.58 [1.46-1.71]* | 1.47 [1.35-1.59]* |
| D89 | 5,210 | 114,223,090 | 0.06 | 0.04 | 0.04 | 0.02 | 0.02 | 0.01 | 1.43 [1.31-1.57]* | 1.33 [1.21-1.46]* |
| any age - E00-E89 | | | | | | | | | | |
| E02 | 1,930 | 114,261,430 | 0.02 | 0.02 | 0.02 | 0.00 | 0.00 | 0.00 | 1.14 [0.97-1.35] | 1.12 [0.95-1.33] |
| E03 | 222,190 | 111,509,480 | 2.35 | 1.91 | 1.76 | 0.45 | 0.60 | 0.51 | 1.34 [1.32-1.36]* | 1.28 [1.26-1.30]* |
| E04 | 23,040 | 113,951,780 | 0.21 | 0.20 | 0.19 | 0.01 | 0.03 | 0.02 | 1.15 [1.10-1.20]* | 1.09 [1.04-1.14]* |
| E05 | 37,470 | 113,814,430 | 0.38 | 0.32 | 0.29 | 0.06 | 0.09 | 0.07 | 1.29 [1.25-1.34]* | 1.23 [1.18-1.27]* |
| E06 | 4,130 | 114,212,330 | 0.04 | 0.04 | 0.03 | 0.01 | 0.01 | 0.01 | 1.22 [1.10-1.35]* | 1.17 [1.05-1.30]* |
| E07 | 6,400 | 114,196,100 | 0.06 | 0.05 | 0.05 | 0.01 | 0.01 | 0.01 | 1.26 [1.16-1.37]* | 1.17 [1.07-1.27]* |
| E10 | 48,670 | 113,231,490 | 0.49 | 0.42 | 0.40 | 0.07 | 0.09 | 0.06 | 1.23 [1.19-1.26]* | 1.15 [1.12-1.19]* |
| E11 | 344,180 | 109,746,160 | 3.53 | 3.04 | 2.81 | 0.49 | 0.72 | 0.56 | 1.26 [1.24-1.27]* | 1.19 [1.18-1.20]* |
| E13 | 4,480 | 114,235,960 | 0.05 | 0.04 | 0.03 | 0.01 | 0.01 | 0.01 | 1.41 [1.28-1.55]* | 1.25 [1.13-1.39]* |
| E14 | 48,270 | 113,678,920 | 0.51 | 0.40 | 0.37 | 0.10 | 0.14 | 0.10 | 1.37 [1.33-1.41]* | 1.25 [1.22-1.29]* |
| E16 | 57,430 | 113,875,050 | 0.58 | 0.49 | 0.46 | 0.09 | 0.12 | 0.09 | 1.25 [1.22-1.29]* | 1.18 [1.14-1.21]* |
| E20 | 1,540 | 114,255,280 | 0.02 | 0.01 | 0.01 | 0.00 | 0.01 | 0.00 | 1.44 [1.21-1.70]* | 1.26 [1.06-1.50]* |
| E21 | 17,410 | 114,133,900 | 0.16 | 0.15 | 0.14 | 0.01 | 0.02 | 0.02 | 1.17 [1.11-1.23]* | 1.12 [1.06-1.18]* |
| E22 | 11,740 | 114,178,290 | 0.13 | 0.10 | 0.09 | 0.04 | 0.04 | 0.04 | 1.48 [1.39-1.57]* | 1.41 [1.32-1.50]* |
| E23 | 6,800 | 114,166,710 | 0.08 | 0.06 | 0.05 | 0.02 | 0.02 | 0.02 | 1.39 [1.28-1.50]* | 1.29 [1.19-1.40]* |
| E24 | 1,380 | 114,247,680 | 0.02 | 0.01 | 0.01 | 0.01 | 0.01 | 0.00 | 1.60 [1.35-1.90]* | 1.32 [1.10-1.58]* |
| E26 | 810 | 114,257,610 | 0.01 | 0.01 | 0.01 | 0.00 | 0.00 | 0.00 | 1.21 [0.95-1.54] | 1.13 [0.88-1.44] |
| E27 | 12,740 | 114,155,760 | 0.14 | 0.10 | 0.10 | 0.04 | 0.05 | 0.04 | 1.49 [1.40-1.57]* | 1.34 [1.26-1.42]* |
| E28 | 31,970 | 113,839,200 | 0.33 | 0.27 | 0.27 | 0.06 | 0.06 | 0.03 | 1.21 [1.17-1.26]* | 1.11 [1.07-1.15]* |
| E29 | 1,780 | 114,247,310 | 0.02 | 0.01 | 0.01 | 0.01 | 0.01 | 0.01 | 1.71 [1.47-1.98]* | 1.50 [1.28-1.75]* |
| E30 | 3,170 | 114,225,040 | 0.04 | 0.03 | 0.03 | 0.01 | 0.01 | 0.01 | 1.24 [1.11-1.39]* | 1.19 [1.06-1.34]* |
| E34 | 6,000 | 114,180,060 | 0.07 | 0.05 | 0.06 | 0.02 | 0.01 | 0.01 | 1.23 [1.13-1.34]* | 1.14 [1.05-1.24]* |
| E41 | 650 | 114,268,540 | 0.01 | 0.01 | 0.01 | 0.00 | 0.00 | 0.00 | 1.07 [0.80-1.42] | 1.10 [0.82-1.47] |
| E43 | 750 | 114,267,340 | 0.01 | 0.01 | 0.01 | 0.00 | 0.00 | 0.00 | 1.20 [0.94-1.54] | 1.18 [0.92-1.53] |
| E46 | 9,660 | 114,225,920 | 0.09 | 0.08 | 0.08 | 0.01 | 0.02 | 0.01 | 1.22 [1.13-1.31]* | 1.19 [1.11-1.28]* |
| E51 | 930 | 114,261,790 | 0.01 | 0.01 | 0.01 | 0.00 | 0.00 | 0.00 | 1.47 [1.19-1.82]* | 1.39 [1.12-1.73]* |
| E53 | 54,380 | 113,982,430 | 0.56 | 0.46 | 0.43 | 0.10 | 0.13 | 0.11 | 1.30 [1.27-1.34]* | 1.23 [1.20-1.27]* |
| E55 | 74,830 | 113,919,860 | 0.83 | 0.62 | 0.60 | 0.21 | 0.22 | 0.17 | 1.37 [1.34-1.40]* | 1.26 [1.23-1.29]* |
| E56 | 780 | 114,264,910 | 0.01 | 0.01 | 0.01 | 0.00 | 0.00 | 0.00 | 1.27 [1.00-1.62] | 1.19 [0.93-1.53] |
| E58 | 550 | 114,266,570 | 0.01 | 0.00 | 0.00 | 0.00 | 0.00 | 0.00 | 1.55 [1.18-2.04]* | 1.48 [1.11-1.95]* |
| E61 | 11,560 | 114,197,480 | 0.13 | 0.09 | 0.09 | 0.03 | 0.04 | 0.03 | 1.38 [1.30-1.47]* | 1.26 [1.18-1.34]* |
| E63 | 710 | 114,265,240 | 0.01 | 0.01 | 0.01 | 0.00 | 0.00 | 0.00 | 1.30 [1.01-1.67] | 1.24 [0.96-1.60] |
| E65 | 480 | 114,251,420 | 0.01 | 0.00 | 0.00 | 0.00 | 0.00 | 0.00 | 1.45 [1.08-1.95] | 1.33 [0.98-1.80] |
| E66 | 389,760 | 110,758,960 | 4.14 | 3.37 | 3.17 | 0.77 | 0.97 | 0.69 | 1.30 [1.29-1.32]* | 1.20 [1.19-1.21]* |
| E70 | 510 | 114,255,830 | 0.01 | 0.00 | 0.00 | 0.00 | 0.00 | 0.00 | 1.35 [1.02-1.79] | 1.30 [0.98-1.74] |
| E72 | 880 | 114,255,440 | 0.01 | 0.01 | 0.01 | 0.00 | 0.00 | 0.00 | 1.17 [0.93-1.46] | 1.09 [0.87-1.37] |
| E73 | 4,380 | 114,193,970 | 0.05 | 0.04 | 0.04 | 0.02 | 0.02 | 0.01 | 1.41 [1.28-1.55]* | 1.22 [1.11-1.35]* |
| E74 | 1,520 | 114,247,230 | 0.02 | 0.01 | 0.01 | 0.00 | 0.00 | 0.00 | 1.35 [1.14-1.60]* | 1.24 [1.05-1.48] |
| E75 | 650 | 114,260,440 | 0.01 | 0.01 | 0.01 | 0.00 | 0.00 | 0.00 | 1.20 [0.93-1.55] | 1.17 [0.90-1.52] |
| E78 | 448,700 | 108,519,310 | 4.62 | 4.02 | 3.76 | 0.61 | 0.87 | 0.68 | 1.23 [1.22-1.24]* | 1.17 [1.16-1.18]* |
| E79 | 610 | 114,264,990 | 0.01 | 0.01 | 0.00 | 0.00 | 0.00 | 0.00 | 1.32 [1.01-1.72] | 1.25 [0.95-1.65] |
| E80 | 6,170 | 114,191,470 | 0.06 | 0.05 | 0.05 | 0.01 | 0.01 | 0.01 | 1.17 [1.07-1.28]* | 1.09 [1.00-1.20] |
| E83 | 83,760 | 113,857,720 | 0.82 | 0.72 | 0.67 | 0.10 | 0.14 | 0.11 | 1.21 [1.18-1.24]* | 1.16 [1.13-1.19]* |
| E84 | 720 | 114,234,860 | 0.01 | 0.01 | 0.01 | 0.00 | 0.00 | 0.00 | 1.23 [0.96-1.58] | 1.11 [0.86-1.43] |
| E85 | 3,060 | 114,252,710 | 0.03 | 0.03 | 0.02 | 0.00 | 0.01 | 0.00 | 1.25 [1.10-1.41]* | 1.20 [1.06-1.36]* |
| E86 | 222,050 | 113,007,960 | 2.19 | 1.91 | 1.84 | 0.28 | 0.36 | 0.31 | 1.20 [1.18-1.21]* | 1.16 [1.14-1.18]* |
| E87 | 330,700 | 112,670,290 | 3.40 | 2.82 | 2.65 | 0.58 | 0.75 | 0.64 | 1.28 [1.27-1.30]* | 1.23 [1.22-1.25]* |
| E88 | 16,000 | 114,181,140 | 0.16 | 0.13 | 0.13 | 0.03 | 0.04 | 0.03 | 1.29 [1.23-1.36]* | 1.23 [1.17-1.30]* |
| E89 | 21,020 | 114,047,640 | 0.21 | 0.18 | 0.17 | 0.03 | 0.04 | 0.03 | 1.25 [1.19-1.31]* | 1.17 [1.12-1.23]* |
| any age - F01-F99 | | | | | | | | | | |
| F00 | 51,390 | 114,115,300 | 0.46 | 0.45 | 0.44 | 0.02 | 0.03 | 0.03 | 1.06 [1.03-1.10]* | 1.06 [1.03-1.10]* |
| F01 | 58,730 | 114,084,850 | 0.54 | 0.51 | 0.49 | 0.03 | 0.05 | 0.04 | 1.10 [1.07-1.13]* | 1.08 [1.05-1.12]* |
| F02 | 10,210 | 114,238,310 | 0.09 | 0.09 | 0.09 | 0.00 | 0.00 | 0.00 | 1.01 [0.94-1.09] | 1.02 [0.95-1.10] |
| F03 | 141,060 | 113,801,810 | 1.29 | 1.23 | 1.20 | 0.06 | 0.09 | 0.08 | 1.07 [1.05-1.09]* | 1.07 [1.05-1.09]* |
| F05 | 129,730 | 113,952,600 | 1.25 | 1.11 | 1.07 | 0.14 | 0.18 | 0.16 | 1.17 [1.15-1.20]* | 1.14 [1.12-1.17]* |
| F06 | 20,420 | 114,173,640 | 0.19 | 0.18 | 0.17 | 0.02 | 0.03 | 0.02 | 1.16 [1.10-1.22]* | 1.13 [1.07-1.19]* |
| F07 | 1,390 | 114,248,780 | 0.01 | 0.01 | 0.01 | 0.00 | 0.00 | 0.00 | 1.28 [1.07-1.53]* | 1.18 [0.99-1.42] |
| F10 | 111,920 | 112,396,930 | 1.08 | 0.98 | 0.90 | 0.10 | 0.18 | 0.16 | 1.20 [1.18-1.23]* | 1.17 [1.15-1.19]* |
| F11 | 8,350 | 114,034,480 | 0.08 | 0.07 | 0.07 | 0.00 | 0.01 | 0.01 | 1.17 [1.08-1.26]* | 1.08 [1.00-1.17] |
| F12 | 18,400 | 114,061,480 | 0.17 | 0.16 | 0.16 | 0.01 | 0.01 | 0.00 | 1.08 [1.03-1.14]* | 1.01 [0.96-1.07] |
| F13 | 1,870 | 114,240,750 | 0.02 | 0.02 | 0.02 | 0.00 | 0.00 | 0.00 | 1.26 [1.08-1.47]* | 1.14 [0.97-1.33] |
| F14 | 7,220 | 114,174,230 | 0.07 | 0.06 | 0.06 | 0.00 | 0.01 | 0.00 | 1.12 [1.03-1.21]* | 1.04 [0.95-1.13] |
| F15 | 2,620 | 114,221,740 | 0.02 | 0.02 | 0.02 | 0.00 | 0.00 | 0.00 | 1.03 [0.90-1.19] | 0.97 [0.85-1.12] |
| F16 | 800 | 114,251,510 | 0.01 | 0.01 | 0.01 | 0.00 | 0.00 | 0.00 | 0.95 [0.73-1.22] | 0.91 [0.70-1.19] |
| F17 | 533,320 | 109,366,710 | 5.14 | 4.81 | 4.55 | 0.32 | 0.58 | 0.51 | 1.13 [1.12-1.14]* | 1.11 [1.10-1.12]* |
| F19 | 7,920 | 114,137,290 | 0.07 | 0.07 | 0.07 | 0.00 | 0.00 | 0.00 | 1.07 [0.99-1.16] | 1.00 [0.92-1.09] |
| F20 | 15,750 | 113,872,320 | 0.16 | 0.13 | 0.12 | 0.02 | 0.03 | 0.03 | 1.28 [1.21-1.34]* | 1.26 [1.19-1.33]* |
| F22 | 7,080 | 114,184,140 | 0.07 | 0.06 | 0.06 | 0.01 | 0.02 | 0.01 | 1.29 [1.19-1.40]* | 1.23 [1.14-1.34]* |
| F23 | 4,440 | 114,180,250 | 0.04 | 0.04 | 0.04 | 0.00 | 0.01 | 0.00 | 1.16 [1.05-1.29]* | 1.13 [1.02-1.26] |
| F25 | 3,270 | 114,184,600 | 0.03 | 0.03 | 0.03 | 0.01 | 0.01 | 0.01 | 1.36 [1.21-1.52]* | 1.32 [1.17-1.49]* |
| F29 | 8,190 | 114,172,850 | 0.08 | 0.07 | 0.07 | 0.01 | 0.01 | 0.01 | 1.22 [1.13-1.32]* | 1.16 [1.07-1.25]* |
| F30 | 2,140 | 114,212,070 | 0.02 | 0.02 | 0.02 | 0.00 | 0.01 | 0.00 | 1.34 [1.16-1.54]* | 1.28 [1.11-1.48]* |
| F31 | 19,010 | 113,948,710 | 0.20 | 0.16 | 0.15 | 0.04 | 0.05 | 0.03 | 1.31 [1.24-1.37]* | 1.19 [1.13-1.25]* |
| F32 | 352,560 | 110,591,970 | 3.90 | 3.02 | 2.84 | 0.88 | 1.05 | 0.74 | 1.37 [1.35-1.38]* | 1.23 [1.22-1.25]* |
| F33 | 10,900 | 114,015,750 | 0.11 | 0.09 | 0.08 | 0.02 | 0.03 | 0.02 | 1.31 [1.23-1.39]* | 1.18 [1.10-1.26]* |
| F34 | 1,830 | 114,242,170 | 0.02 | 0.02 | 0.01 | 0.01 | 0.01 | 0.00 | 1.44 [1.24-1.68]* | 1.25 [1.06-1.46]* |
| F38 | 880 | 114,261,580 | 0.01 | 0.01 | 0.01 | 0.00 | 0.00 | 0.00 | 1.30 [1.04-1.64] | 1.19 [0.94-1.51] |
| F39 | 1,550 | 114,252,080 | 0.02 | 0.01 | 0.01 | 0.00 | 0.00 | 0.00 | 1.33 [1.12-1.57]* | 1.20 [1.00-1.42] |
| F40 | 34,110 | 113,960,660 | 0.36 | 0.28 | 0.29 | 0.08 | 0.07 | 0.05 | 1.26 [1.21-1.30]* | 1.16 [1.12-1.21]* |
| F41 | 301,930 | 112,056,780 | 3.29 | 2.55 | 2.45 | 0.73 | 0.83 | 0.56 | 1.34 [1.32-1.36]* | 1.20 [1.19-1.22]* |
| F42 | 7,540 | 114,189,110 | 0.10 | 0.06 | 0.06 | 0.04 | 0.04 | 0.03 | 1.64 [1.52-1.76]* | 1.43 [1.32-1.54]* |
| F43 | 17,280 | 114,012,180 | 0.18 | 0.15 | 0.14 | 0.03 | 0.04 | 0.02 | 1.26 [1.20-1.33]* | 1.11 [1.05-1.17]* |
| F44 | 4,140 | 114,219,250 | 0.05 | 0.03 | 0.03 | 0.01 | 0.01 | 0.01 | 1.40 [1.26-1.55]* | 1.21 [1.09-1.34]* |
| F45 | 4,960 | 114,184,440 | 0.06 | 0.04 | 0.04 | 0.01 | 0.02 | 0.01 | 1.45 [1.32-1.59]* | 1.28 [1.16-1.41]* |
| F48 | 1,360 | 114,251,430 | 0.02 | 0.01 | 0.01 | 0.00 | 0.01 | 0.00 | 1.55 [1.30-1.84]* | 1.35 [1.12-1.62]* |
| F50 | 8,450 | 114,155,800 | 0.09 | 0.07 | 0.07 | 0.02 | 0.02 | 0.01 | 1.28 [1.19-1.37]* | 1.17 [1.08-1.26]* |
| F51 | 1,780 | 114,250,060 | 0.02 | 0.01 | 0.01 | 0.01 | 0.01 | 0.00 | 1.43 [1.23-1.67]* | 1.30 [1.11-1.52]* |
| F52 | 2,710 | 114,223,540 | 0.03 | 0.02 | 0.02 | 0.01 | 0.01 | 0.01 | 1.49 [1.32-1.69]* | 1.35 [1.19-1.53]* |
| F53 | 2,660 | 114,222,820 | 0.03 | 0.02 | 0.02 | 0.01 | 0.01 | 0.01 | 1.35 [1.19-1.53]* | 1.24 [1.08-1.41]* |
| F60 | 14,190 | 114,029,770 | 0.15 | 0.12 | 0.12 | 0.04 | 0.04 | 0.02 | 1.34 [1.27-1.42]* | 1.15 [1.09-1.22]* |
| F61 | 540 | 114,260,920 | 0.01 | 0.00 | 0.00 | 0.00 | 0.00 | 0.00 | 1.13 [0.85-1.51] | 0.99 [0.73-1.35] |
| F69 | 1,170 | 114,247,550 | 0.01 | 0.01 | 0.01 | 0.00 | 0.00 | 0.00 | 1.47 [1.22-1.77]* | 1.27 [1.05-1.55] |
| F70 | 2,200 | 114,226,460 | 0.03 | 0.02 | 0.02 | 0.01 | 0.01 | 0.01 | 1.72 [1.51-1.97]* | 1.54 [1.34-1.76]* |
| F71 | 850 | 114,252,140 | 0.01 | 0.01 | 0.01 | 0.01 | 0.01 | 0.01 | 2.16 [1.76-2.65]* | 1.93 [1.56-2.39]* |
| F72 | 1,670 | 114,238,650 | 0.02 | 0.01 | 0.01 | 0.01 | 0.01 | 0.01 | 1.67 [1.43-1.94]* | 1.51 [1.29-1.76]* |
| F79 | 10,330 | 114,131,310 | 0.13 | 0.08 | 0.08 | 0.05 | 0.05 | 0.04 | 1.56 [1.47-1.66]* | 1.44 [1.35-1.54]* |
| F80 | 11,880 | 114,137,020 | 0.13 | 0.10 | 0.11 | 0.03 | 0.02 | 0.02 | 1.22 [1.15-1.29]* | 1.16 [1.10-1.24]* |
| F81 | 30,440 | 113,772,470 | 0.38 | 0.24 | 0.25 | 0.14 | 0.14 | 0.12 | 1.56 [1.50-1.61]* | 1.44 [1.39-1.50]* |
| F82 | 1,680 | 114,245,650 | 0.02 | 0.01 | 0.02 | 0.00 | 0.00 | 0.00 | 1.19 [1.02-1.39] | 1.11 [0.95-1.31] |
| F83 | 1,530 | 114,253,820 | 0.01 | 0.01 | 0.02 | 0.00 | 0.00 | 0.00 | 0.95 [0.80-1.13] | 0.89 [0.74-1.06] |
| F84 | 28,810 | 113,981,400 | 0.36 | 0.23 | 0.27 | 0.13 | 0.09 | 0.06 | 1.32 [1.28-1.38]* | 1.18 [1.14-1.23]* |
| F89 | 7,340 | 114,209,390 | 0.08 | 0.06 | 0.07 | 0.01 | 0.00 | 0.00 | 1.02 [0.95-1.10] | 0.94 [0.87-1.02] |
| F90 | 14,870 | 114,132,570 | 0.18 | 0.12 | 0.15 | 0.06 | 0.03 | 0.02 | 1.22 [1.15-1.28]* | 1.12 [1.06-1.19]* |
| F91 | 3,210 | 114,211,000 | 0.04 | 0.03 | 0.03 | 0.01 | 0.01 | 0.01 | 1.27 [1.13-1.42]* | 1.19 [1.06-1.34]* |

| | | | | | | | | | | |
|---|---|---|---|---|---|---|---|---|---|---|
| F94 | 610 | 114,265,240 | 0.01 | 0.00 | 0.01 | 0.00 | 0.00 | 0.00 | 1.22 [0.94-1.58] | 1.15 [0.88-1.51] |
| F95 | 1,710 | 114,253,610 | 0.02 | 0.01 | 0.02 | 0.01 | 0.01 | 0.00 | 1.38 [1.18-1.61]* | 1.23 [1.05-1.45] |
| F98 | 2,190 | 114,238,850 | 0.03 | 0.02 | 0.02 | 0.01 | 0.01 | 0.00 | 1.32 [1.15-1.52]* | 1.22 [1.06-1.41]* |
| F99 | 5,910 | 114,147,030 | 0.06 | 0.05 | 0.05 | 0.01 | 0.01 | 0.01 | 1.24 [1.14-1.36]* | 1.16 [1.06-1.27]* |
| any age - G00-G99 | | | | | | | | | | |
| G00 | 1,720 | 114,207,870 | 0.02 | 0.01 | 0.01 | 0.00 | 0.00 | 0.00 | 1.12 [0.95-1.31] | 1.07 [0.90-1.26] |
| G01 | 910 | 114,232,140 | 0.01 | 0.01 | 0.01 | 0.00 | 0.00 | 0.00 | 1.11 [0.89-1.39] | 1.06 [0.84-1.33] |
| G02 | 780 | 114,245,840 | 0.01 | 0.01 | 0.01 | 0.00 | 0.00 | 0.00 | 1.08 [0.85-1.39] | 0.98 [0.76-1.27] |
| G03 | 2,130 | 114,211,870 | 0.02 | 0.02 | 0.02 | 0.00 | 0.00 | 0.00 | 1.24 [1.07-1.43]* | 1.15 [1.00-1.34] |
| G04 | 4,690 | 114,217,390 | 0.05 | 0.04 | 0.04 | 0.01 | 0.01 | 0.01 | 1.25 [1.13-1.38]* | 1.19 [1.08-1.32]* |
| G05 | 1,210 | 114,254,830 | 0.01 | 0.01 | 0.01 | 0.00 | 0.00 | 0.00 | 1.42 [1.17-1.71]* | 1.37 [1.14-1.66]* |
| G06 | 2,080 | 114,243,350 | 0.02 | 0.02 | 0.02 | 0.00 | 0.00 | 0.00 | 1.25 [1.08-1.45]* | 1.20 [1.03-1.40] |
| G08 | 1,160 | 114,256,880 | 0.01 | 0.01 | 0.01 | 0.00 | 0.00 | 0.00 | 1.16 [0.95-1.42] | 1.12 [0.92-1.38] |
| G09 | 1,310 | 114,246,960 | 0.01 | 0.01 | 0.01 | 0.00 | 0.00 | 0.00 | 1.36 [1.14-1.63]* | 1.28 [1.06-1.54]* |
| G10 | 600 | 114,262,480 | 0.01 | 0.01 | 0.00 | 0.00 | 0.00 | 0.00 | 1.40 [1.07-1.84] | 1.42 [1.08-1.87] |
| G11 | 2,160 | 114,242,600 | 0.02 | 0.02 | 0.02 | 0.00 | 0.00 | 0.00 | 1.17 [1.01-1.36] | 1.13 [0.97-1.31] |
| G12 | 4,500 | 114,245,650 | 0.04 | 0.04 | 0.04 | 0.00 | 0.00 | 0.00 | 1.08 [0.97-1.20] | 1.04 [0.93-1.15] |
| G20 | 39,140 | 114,013,930 | 0.34 | 0.34 | 0.32 | 0.00 | 0.02 | 0.02 | 1.06 [1.02-1.09]* | 1.05 [1.01-1.09] |
| G21 | 3,400 | 114,252,110 | 0.04 | 0.03 | 0.03 | 0.01 | 0.01 | 0.01 | 1.34 [1.19-1.50]* | 1.31 [1.17-1.48]* |
| G23 | 2,110 | 114,261,620 | 0.02 | 0.02 | 0.02 | 0.00 | 0.00 | 0.00 | 1.17 [1.00-1.36] | 1.14 [0.97-1.33] |
| G24 | 4,860 | 114,205,480 | 0.05 | 0.04 | 0.04 | 0.01 | 0.01 | 0.00 | 1.14 [1.03-1.25]* | 1.09 [0.98-1.20] |
| G25 | 22,100 | 114,103,630 | 0.22 | 0.19 | 0.18 | 0.04 | 0.05 | 0.03 | 1.28 [1.23-1.34]* | 1.16 [1.11-1.22]* |
| G30 | 68,860 | 114,028,030 | 0.62 | 0.60 | 0.58 | 0.02 | 0.04 | 0.04 | 1.07 [1.04-1.10]* | 1.06 [1.03-1.09]* |
| G31 | 49,740 | 114,035,670 | 0.47 | 0.43 | 0.40 | 0.04 | 0.07 | 0.06 | 1.17 [1.13-1.21]* | 1.14 [1.10-1.18]* |
| G32 | 790 | 114,263,120 | 0.01 | 0.01 | 0.01 | 0.00 | 0.00 | 0.00 | 1.39 [1.10-1.76]* | 1.30 [1.02-1.66] |
| G35 | 10,520 | 114,020,700 | 0.10 | 0.09 | 0.08 | 0.01 | 0.02 | 0.01 | 1.23 [1.15-1.31]* | 1.16 [1.08-1.24]* |
| G37 | 2,930 | 114,209,940 | 0.03 | 0.02 | 0.02 | 0.00 | 0.01 | 0.01 | 1.28 [1.14-1.45]* | 1.22 [1.07-1.38]* |
| G40 | 75,530 | 112,792,190 | 0.80 | 0.64 | 0.61 | 0.16 | 0.19 | 0.14 | 1.31 [1.28-1.34]* | 1.21 [1.18-1.24]* |
| G41 | 5,770 | 114,183,590 | 0.06 | 0.05 | 0.05 | 0.01 | 0.01 | 0.01 | 1.20 [1.10-1.31]* | 1.13 [1.04-1.24]* |
| G43 | 70,130 | 113,469,880 | 0.75 | 0.59 | 0.57 | 0.17 | 0.18 | 0.11 | 1.32 [1.28-1.35]* | 1.16 [1.13-1.19]* |
| G44 | 10,050 | 114,123,630 | 0.11 | 0.08 | 0.08 | 0.02 | 0.03 | 0.02 | 1.33 [1.24-1.42]* | 1.20 [1.12-1.28]* |
| G45 | 47,400 | 113,709,290 | 0.45 | 0.41 | 0.38 | 0.04 | 0.07 | 0.05 | 1.18 [1.15-1.22]* | 1.14 [1.10-1.17]* |
| G46 | 3,110 | 114,250,730 | 0.03 | 0.03 | 0.02 | 0.00 | 0.01 | 0.00 | 1.22 [1.07-1.38]* | 1.19 [1.04-1.35]* |
| G47 | 90,940 | 113,138,040 | 1.09 | 0.74 | 0.74 | 0.35 | 0.35 | 0.27 | 1.48 [1.45-1.51]* | 1.32 [1.30-1.35]* |
| G50 | 7,720 | 114,178,330 | 0.08 | 0.06 | 0.06 | 0.02 | 0.02 | 0.02 | 1.42 [1.32-1.53]* | 1.29 [1.20-1.39]* |
| G51 | 17,200 | 114,044,900 | 0.17 | 0.15 | 0.14 | 0.02 | 0.03 | 0.02 | 1.21 [1.15-1.28]* | 1.15 [1.09-1.21]* |
| G52 | 1,270 | 114,255,190 | 0.01 | 0.01 | 0.01 | 0.00 | 0.00 | 0.00 | 1.23 [1.02-1.49] | 1.17 [0.97-1.42] |
| G53 | 2,700 | 114,248,480 | 0.03 | 0.02 | 0.02 | 0.01 | 0.01 | 0.01 | 1.33 [1.17-1.52]* | 1.26 [1.10-1.44]* |
| G54 | 4,900 | 114,205,020 | 0.05 | 0.04 | 0.04 | 0.01 | 0.01 | 0.01 | 1.33 [1.21-1.46]* | 1.21 [1.10-1.33]* |
| G55 | 55,280 | 113,576,570 | 0.55 | 0.47 | 0.43 | 0.07 | 0.12 | 0.08 | 1.27 [1.24-1.31]* | 1.17 [1.14-1.21]* |
| G56 | 82,090 | 112,484,540 | 0.85 | 0.70 | 0.63 | 0.15 | 0.22 | 0.16 | 1.35 [1.32-1.38]* | 1.24 [1.21-1.27]* |
| G57 | 9,610 | 114,099,740 | 0.10 | 0.08 | 0.07 | 0.02 | 0.03 | 0.02 | 1.39 [1.30-1.48]* | 1.28 [1.19-1.37]* |
| G58 | 4,940 | 114,196,590 | 0.05 | 0.04 | 0.04 | 0.01 | 0.01 | 0.01 | 1.32 [1.20-1.45]* | 1.20 [1.09-1.32]* |
| G59 | 3,720 | 114,223,160 | 0.04 | 0.03 | 0.03 | 0.01 | 0.01 | 0.01 | 1.49 [1.34-1.65]* | 1.31 [1.18-1.47]* |
| G60 | 2,360 | 114,238,510 | 0.02 | 0.02 | 0.02 | 0.00 | 0.01 | 0.00 | 1.28 [1.11-1.46]* | 1.17 [1.02-1.34] |
| G61 | 2,940 | 114,221,830 | 0.03 | 0.03 | 0.02 | 0.00 | 0.01 | 0.00 | 1.25 [1.11-1.41]* | 1.21 [1.07-1.37]* |
| G62 | 30,530 | 114,039,910 | 0.32 | 0.26 | 0.24 | 0.06 | 0.08 | 0.06 | 1.35 [1.30-1.40]* | 1.23 [1.18-1.28]* |
| G63 | 15,950 | 114,162,100 | 0.18 | 0.13 | 0.12 | 0.04 | 0.05 | 0.04 | 1.45 [1.38-1.53]* | 1.30 [1.23-1.37]* |
| G70 | 2,910 | 114,230,380 | 0.03 | 0.03 | 0.02 | 0.00 | 0.00 | 0.00 | 1.12 [0.99-1.28] | 1.07 [0.94-1.22] |
| G71 | 2,590 | 114,224,610 | 0.03 | 0.02 | 0.02 | 0.00 | 0.00 | 0.00 | 1.18 [1.04-1.35] | 1.15 [1.00-1.31] |
| G72 | 3,460 | 114,233,280 | 0.04 | 0.03 | 0.03 | 0.01 | 0.01 | 0.01 | 1.41 [1.26-1.58]* | 1.32 [1.18-1.48]* |
| G80 | 8,700 | 114,046,970 | 0.08 | 0.08 | 0.08 | 0.00 | 0.00 | 0.00 | 1.01 [0.94-1.09] | 0.96 [0.89-1.04] |
| G81 | 56,170 | 113,743,260 | 0.53 | 0.48 | 0.45 | 0.05 | 0.08 | 0.07 | 1.17 [1.14-1.20]* | 1.14 [1.11-1.18]* |
| G82 | 6,180 | 114,160,580 | 0.06 | 0.05 | 0.05 | 0.00 | 0.01 | 0.01 | 1.15 [1.05-1.25]* | 1.10 [1.01-1.20] |
| G83 | 8,700 | 114,178,200 | 0.08 | 0.07 | 0.07 | 0.01 | 0.01 | 0.01 | 1.21 [1.12-1.30]* | 1.15 [1.06-1.23]* |
| G90 | 4,610 | 114,223,450 | 0.05 | 0.04 | 0.04 | 0.01 | 0.01 | 0.01 | 1.26 [1.15-1.40]* | 1.17 [1.05-1.29]* |
| G91 | 11,190 | 114,128,260 | 0.11 | 0.10 | 0.09 | 0.01 | 0.02 | 0.01 | 1.17 [1.10-1.25]* | 1.15 [1.08-1.23]* |
| G93 | 45,820 | 113,861,400 | 0.46 | 0.39 | 0.37 | 0.08 | 0.10 | 0.07 | 1.26 [1.22-1.30]* | 1.17 [1.13-1.21]* |
| G94 | 1,690 | 114,257,180 | 0.02 | 0.01 | 0.01 | 0.00 | 0.00 | 0.00 | 1.18 [1.00-1.39] | 1.17 [0.99-1.38] |
| G95 | 12,040 | 114,156,690 | 0.11 | 0.10 | 0.10 | 0.01 | 0.02 | 0.01 | 1.19 [1.12-1.27]* | 1.14 [1.07-1.21]* |
| G96 | 3,850 | 114,217,310 | 0.04 | 0.03 | 0.03 | 0.01 | 0.01 | 0.01 | 1.30 [1.17-1.44]* | 1.20 [1.07-1.34]* |
| G97 | 3,740 | 114,211,560 | 0.04 | 0.03 | 0.03 | 0.01 | 0.01 | 0.00 | 1.22 [1.09-1.36]* | 1.11 [0.99-1.24] |
| G98 | 1,280 | 114,261,690 | 0.01 | 0.01 | 0.01 | 0.00 | 0.00 | 0.00 | 1.49 [1.25-1.79]* | 1.33 [1.10-1.60]* |
| G99 | 18,300 | 114,091,180 | 0.18 | 0.16 | 0.14 | 0.02 | 0.04 | 0.03 | 1.26 [1.20-1.33]* | 1.18 [1.12-1.24]* |
| any age - H00-H59 | | | | | | | | | | |
| H00 | 17,570 | 113,819,440 | 0.18 | 0.15 | 0.15 | 0.04 | 0.04 | 0.03 | 1.25 [1.19-1.31]* | 1.20 [1.14-1.27]* |
| H01 | 11,540 | 114,128,780 | 0.13 | 0.10 | 0.09 | 0.03 | 0.04 | 0.03 | 1.42 [1.33-1.51]* | 1.33 [1.25-1.42]* |
| H02 | 60,580 | 113,228,740 | 0.62 | 0.51 | 0.48 | 0.11 | 0.15 | 0.12 | 1.31 [1.27-1.34]* | 1.24 [1.21-1.28]* |
| H03 | 890 | 114,253,930 | 0.01 | 0.01 | 0.01 | 0.00 | 0.00 | 0.00 | 1.61 [1.30-1.98]* | 1.50 [1.20-1.86]* |
| H04 | 30,700 | 113,743,620 | 0.33 | 0.25 | 0.24 | 0.08 | 0.10 | 0.08 | 1.40 [1.35-1.45]* | 1.30 [1.25-1.35]* |
| H05 | 11,070 | 114,051,510 | 0.13 | 0.09 | 0.09 | 0.04 | 0.04 | 0.03 | 1.40 [1.32-1.49]* | 1.36 [1.28-1.44]* |
| H06 | 1,190 | 114,251,230 | 0.01 | 0.01 | 0.01 | 0.00 | 0.00 | 0.00 | 1.49 [1.23-1.79]* | 1.43 [1.18-1.73]* |
| H10 | 17,430 | 113,936,020 | 0.20 | 0.14 | 0.14 | 0.06 | 0.06 | 0.05 | 1.43 [1.36-1.50]* | 1.34 [1.27-1.41]* |
| H11 | 12,680 | 114,076,650 | 0.13 | 0.11 | 0.10 | 0.02 | 0.02 | 0.02 | 1.24 [1.16-1.31]* | 1.19 [1.12-1.26]* |
| H13 | 980 | 114,251,940 | 0.01 | 0.01 | 0.01 | 0.00 | 0.00 | 0.00 | 1.47 [1.20-1.80]* | 1.37 [1.11-1.69]* |
| H15 | 1,150 | 114,253,710 | 0.01 | 0.01 | 0.01 | 0.00 | 0.00 | 0.00 | 1.41 [1.17-1.71]* | 1.31 [1.08-1.60]* |
| H16 | 5,520 | 114,185,990 | 0.07 | 0.04 | 0.04 | 0.03 | 0.03 | 0.03 | 1.71 [1.57-1.86]* | 1.60 [1.47-1.75]* |
| H17 | 3,180 | 114,231,140 | 0.04 | 0.03 | 0.03 | 0.01 | 0.01 | 0.01 | 1.41 [1.26-1.59]* | 1.34 [1.19-1.51]* |
| H18 | 21,950 | 114,067,940 | 0.24 | 0.18 | 0.17 | 0.06 | 0.07 | 0.06 | 1.38 [1.32-1.44]* | 1.31 [1.25-1.37]* |
| H19 | 1,390 | 114,252,480 | 0.02 | 0.01 | 0.01 | 0.01 | 0.01 | 0.01 | 1.81 [1.53-2.14]* | 1.60 [1.34-1.90]* |
| H20 | 5,960 | 114,181,370 | 0.07 | 0.05 | 0.05 | 0.02 | 0.02 | 0.02 | 1.43 [1.32-1.56]* | 1.38 [1.27-1.50]* |
| H21 | 9,290 | 114,178,110 | 0.09 | 0.08 | 0.07 | 0.01 | 0.01 | 0.01 | 1.18 [1.10-1.27]* | 1.14 [1.06-1.23]* |
| H22 | 440 | 114,265,760 | 0.01 | 0.00 | 0.00 | 0.00 | 0.00 | 0.00 | 1.46 [1.07-1.99] | 1.40 [1.02-1.91] |
| H25 | 220,680 | 111,840,070 | 2.14 | 1.93 | 1.83 | 0.21 | 0.31 | 0.25 | 1.17 [1.15-1.19]* | 1.13 [1.12-1.15]* |
| H26 | 311,990 | 110,390,570 | 3.15 | 2.75 | 2.61 | 0.40 | 0.55 | 0.45 | 1.21 [1.19-1.23]* | 1.17 [1.15-1.18]* |
| H27 | 4,460 | 114,201,180 | 0.04 | 0.04 | 0.04 | 0.00 | 0.01 | 0.01 | 1.19 [1.08-1.32]* | 1.16 [1.05-1.29]* |
| H28 | 1,630 | 114,251,990 | 0.02 | 0.01 | 0.01 | 0.00 | 0.00 | 0.00 | 1.32 [1.12-1.55]* | 1.24 [1.05-1.47] |
| H30 | 530 | 114,261,530 | 0.01 | 0.00 | 0.00 | 0.00 | 0.00 | 0.00 | 1.33 [1.00-1.76] | 1.26 [0.94-1.68] |
| H31 | 5,310 | 114,220,520 | 0.05 | 0.05 | 0.04 | 0.00 | 0.00 | 0.00 | 1.11 [1.01-1.22] | 1.09 [0.99-1.20] |
| H33 | 21,100 | 113,879,380 | 0.18 | 0.19 | 0.17 | −0.01 | 0.01 | 0.01 | 1.05 [1.00-1.10] | 1.03 [0.98-1.08] |
| H34 | 12,590 | 114,145,760 | 0.12 | 0.11 | 0.10 | 0.01 | 0.02 | 0.01 | 1.17 [1.10-1.24]* | 1.14 [1.07-1.21]* |
| H35 | 115,730 | 113,214,060 | 1.10 | 1.00 | 0.94 | 0.10 | 0.16 | 0.13 | 1.17 [1.14-1.19]* | 1.13 [1.11-1.16]* |
| H36 | 45,990 | 113,813,430 | 0.45 | 0.39 | 0.36 | 0.06 | 0.09 | 0.07 | 1.25 [1.21-1.29]* | 1.18 [1.14-1.22]* |
| H40 | 106,080 | 113,192,320 | 1.01 | 0.92 | 0.86 | 0.09 | 0.15 | 0.13 | 1.18 [1.15-1.20]* | 1.14 [1.12-1.17]* |
| H43 | 21,680 | 114,020,430 | 0.20 | 0.19 | 0.17 | 0.01 | 0.02 | 0.02 | 1.13 [1.07-1.18]* | 1.08 [1.03-1.14]* |
| H44 | 7,060 | 114,161,440 | 0.07 | 0.06 | 0.06 | 0.01 | 0.02 | 0.01 | 1.30 [1.20-1.40]* | 1.24 [1.14-1.34]* |
| H46 | 2,150 | 114,239,310 | 0.02 | 0.02 | 0.02 | 0.01 | 0.01 | 0.01 | 1.37 [1.19-1.58]* | 1.33 [1.15-1.54]* |
| H47 | 9,550 | 114,165,530 | 0.10 | 0.08 | 0.08 | 0.01 | 0.01 | 0.01 | 1.18 [1.10-1.26]* | 1.13 [1.06-1.22]* |
| H49 | 6,050 | 114,196,800 | 0.05 | 0.05 | 0.05 | 0.00 | 0.00 | 0.00 | 1.06 [0.97-1.16] | 1.01 [0.92-1.11] |
| H50 | 28,450 | 113,669,630 | 0.28 | 0.24 | 0.26 | 0.04 | 0.02 | 0.01 | 1.07 [1.03-1.12]* | 1.04 [1.00-1.08] |
| H51 | 1,120 | 114,254,270 | 0.01 | 0.01 | 0.01 | 0.00 | 0.00 | 0.00 | 1.20 [0.98-1.46] | 1.12 [0.91-1.38] |
| H52 | 37,710 | 113,946,940 | 0.35 | 0.33 | 0.31 | 0.02 | 0.04 | 0.03 | 1.13 [1.09-1.17]* | 1.10 [1.06-1.14]* |
| H53 | 75,260 | 113,343,430 | 0.76 | 0.64 | 0.62 | 0.12 | 0.15 | 0.10 | 1.24 [1.21-1.27]* | 1.16 [1.13-1.18]* |
| H54 | 53,750 | 113,902,700 | 0.52 | 0.46 | 0.44 | 0.06 | 0.09 | 0.07 | 1.20 [1.16-1.24]* | 1.16 [1.12-1.20]* |
| H55 | 5,860 | 114,198,220 | 0.06 | 0.05 | 0.05 | 0.01 | 0.01 | 0.00 | 1.12 [1.03-1.23] | 1.07 [0.98-1.17] |
| H57 | 12,420 | 114,120,420 | 0.13 | 0.10 | 0.10 | 0.03 | 0.03 | 0.02 | 1.27 [1.20-1.35]* | 1.19 [1.12-1.27]* |
| H58 | 2,390 | 114,248,580 | 0.02 | 0.02 | 0.02 | 0.00 | 0.00 | 0.00 | 1.23 [1.07-1.42]* | 1.18 [1.02-1.36] |

| | | | | | | | | | | |
|---|---|---|---|---|---|---|---|---|---|---|
| H59 | 4,180 | 114,207,230 | 0.04 | 0.04 | 0.03 | 0.00 | 0.01 | 0.00 | 1.17 [1.05-1.30]* | 1.14 [1.03-1.27] |
| any age - H60-H95 | | | | | | | | | | |
| H60 | 12,310 | 114,030,500 | 0.15 | 0.10 | 0.10 | 0.06 | 0.06 | 0.05 | 1.60 [1.51-1.69]* | 1.50 [1.42-1.59]* |
| H61 | 24,120 | 113,844,430 | 0.27 | 0.20 | 0.20 | 0.08 | 0.07 | 0.06 | 1.35 [1.29-1.40]* | 1.30 [1.24-1.35]* |
| H65 | 68,730 | 112,511,020 | 0.77 | 0.57 | 0.64 | 0.20 | 0.13 | 0.11 | 1.21 [1.18-1.24]* | 1.17 [1.14-1.20]* |
| H66 | 33,480 | 113,341,490 | 0.38 | 0.28 | 0.30 | 0.10 | 0.08 | 0.07 | 1.26 [1.22-1.31]* | 1.21 [1.17-1.26]* |
| H69 | 2,430 | 114,223,780 | 0.03 | 0.02 | 0.02 | 0.01 | 0.01 | 0.01 | 1.46 [1.28-1.66]* | 1.36 [1.19-1.56]* |
| H70 | 3,140 | 114,215,150 | 0.03 | 0.03 | 0.03 | 0.01 | 0.00 | 0.00 | 1.18 [1.05-1.33]* | 1.13 [1.00-1.28] |
| H71 | 7,220 | 114,112,790 | 0.07 | 0.06 | 0.06 | 0.01 | 0.01 | 0.01 | 1.10 [1.02-1.19] | 1.08 [0.99-1.17] |
| H72 | 14,500 | 113,917,460 | 0.15 | 0.12 | 0.12 | 0.03 | 0.03 | 0.03 | 1.24 [1.18-1.31]* | 1.19 [1.13-1.26]* |
| H73 | 10,030 | 114,058,810 | 0.11 | 0.08 | 0.09 | 0.03 | 0.02 | 0.02 | 1.22 [1.14-1.30]* | 1.17 [1.10-1.25]* |
| H74 | 7,430 | 114,104,090 | 0.08 | 0.06 | 0.06 | 0.01 | 0.01 | 0.01 | 1.19 [1.10-1.29]* | 1.15 [1.06-1.24]* |
| H80 | 1,500 | 114,234,030 | 0.01 | 0.01 | 0.01 | 0.00 | 0.00 | 0.00 | 1.09 [0.91-1.30] | 1.07 [0.90-1.29] |
| H81 | 22,090 | 114,069,080 | 0.22 | 0.19 | 0.18 | 0.03 | 0.04 | 0.03 | 1.26 [1.20-1.32]* | 1.17 [1.12-1.23]* |
| H83 | 9,460 | 114,127,220 | 0.09 | 0.08 | 0.07 | 0.01 | 0.02 | 0.01 | 1.26 [1.18-1.35]* | 1.17 [1.09-1.25]* |
| H90 | 29,430 | 113,934,360 | 0.32 | 0.24 | 0.25 | 0.07 | 0.07 | 0.06 | 1.29 [1.24-1.34]* | 1.22 [1.17-1.27]* |
| H91 | 140,390 | 113,276,850 | 1.41 | 1.20 | 1.15 | 0.21 | 0.26 | 0.21 | 1.23 [1.20-1.25]* | 1.17 [1.15-1.20]* |
| H92 | 11,510 | 114,042,320 | 0.13 | 0.09 | 0.10 | 0.04 | 0.03 | 0.03 | 1.34 [1.26-1.42]* | 1.25 [1.18-1.33]* |
| H93 | 13,030 | 114,128,310 | 0.14 | 0.11 | 0.10 | 0.03 | 0.04 | 0.03 | 1.35 [1.27-1.43]* | 1.26 [1.19-1.34]* |
| H95 | 1,510 | 114,235,370 | 0.02 | 0.01 | 0.01 | 0.00 | 0.00 | 0.00 | 1.27 [1.07-1.50]* | 1.24 [1.05-1.48] |
| any age - I00-I99 | | | | | | | | | | |
| I05 | 7,130 | 114,185,600 | 0.07 | 0.06 | 0.06 | 0.01 | 0.02 | 0.01 | 1.30 [1.20-1.40]* | 1.22 [1.12-1.32]* |
| I06 | 690 | 114,257,890 | 0.01 | 0.01 | 0.01 | 0.00 | 0.00 | 0.00 | 1.34 [1.05-1.73] | 1.26 [0.98-1.63] |
| I07 | 18,210 | 114,186,150 | 0.19 | 0.15 | 0.14 | 0.03 | 0.04 | 0.04 | 1.31 [1.24-1.38]* | 1.25 [1.19-1.32]* |
| I08 | 93,060 | 113,808,890 | 0.91 | 0.80 | 0.75 | 0.11 | 0.16 | 0.13 | 1.21 [1.19-1.24]* | 1.17 [1.14-1.20]* |
| I09 | 1,060 | 114,258,150 | 0.01 | 0.01 | 0.01 | 0.00 | 0.00 | 0.00 | 1.42 [1.17-1.74]* | 1.36 [1.11-1.67]* |
| I10 | 923,670 | 100,710,140 | 10.51 | 8.85 | 8.46 | 1.66 | 2.05 | 1.67 | 1.24 [1.23-1.25]* | 1.19 [1.18-1.20]* |
| I11 | 5,330 | 114,209,680 | 0.05 | 0.05 | 0.04 | 0.00 | 0.01 | 0.01 | 1.21 [1.10-1.33]* | 1.15 [1.04-1.27]* |
| I12 | 52,680 | 113,792,280 | 0.52 | 0.45 | 0.40 | 0.07 | 0.12 | 0.10 | 1.29 [1.26-1.33]* | 1.25 [1.21-1.29]* |
| I13 | 2,620 | 114,262,390 | 0.03 | 0.02 | 0.02 | 0.00 | 0.01 | 0.00 | 1.30 [1.13-1.49]* | 1.22 [1.06-1.40]* |
| I15 | 3,980 | 114,220,120 | 0.04 | 0.03 | 0.03 | 0.01 | 0.01 | 0.01 | 1.28 [1.15-1.42]* | 1.22 [1.09-1.36]* |
| I20 | 203,010 | 110,384,760 | 2.10 | 1.78 | 1.64 | 0.32 | 0.45 | 0.34 | 1.28 [1.26-1.29]* | 1.20 [1.18-1.21]* |
| I21 | 138,620 | 112,614,840 | 1.29 | 1.22 | 1.12 | 0.08 | 0.17 | 0.15 | 1.16 [1.13-1.18]* | 1.13 [1.11-1.15]* |
| I22 | 12,550 | 114,094,910 | 0.12 | 0.11 | 0.10 | 0.02 | 0.03 | 0.02 | 1.28 [1.20-1.36]* | 1.22 [1.15-1.30]* |
| I23 | 790 | 114,259,200 | 0.01 | 0.01 | 0.01 | 0.00 | 0.00 | 0.00 | 1.24 [0.98-1.57] | 1.22 [0.95-1.55] |
| I24 | 39,460 | 113,982,260 | 0.39 | 0.34 | 0.31 | 0.05 | 0.07 | 0.06 | 1.24 [1.20-1.28]* | 1.18 [1.13-1.22]* |
| I25 | 348,810 | 109,192,490 | 3.58 | 3.10 | 2.93 | 0.47 | 0.64 | 0.50 | 1.22 [1.21-1.23]* | 1.16 [1.15-1.18]* |
| I26 | 71,350 | 113,645,530 | 0.65 | 0.62 | 0.57 | 0.02 | 0.07 | 0.05 | 1.13 [1.10-1.16]* | 1.08 [1.05-1.11]* |
| I27 | 43,020 | 114,049,030 | 0.44 | 0.36 | 0.33 | 0.08 | 0.11 | 0.09 | 1.32 [1.28-1.37]* | 1.25 [1.21-1.29]* |
| I28 | 1,310 | 114,257,620 | 0.01 | 0.01 | 0.01 | 0.00 | 0.00 | 0.00 | 1.16 [0.96-1.41] | 1.07 [0.88-1.30] |
| I30 | 2,240 | 114,228,230 | 0.02 | 0.02 | 0.02 | 0.00 | 0.00 | 0.00 | 1.24 [1.07-1.42]* | 1.19 [1.03-1.37] |
| I31 | 27,080 | 114,041,680 | 0.26 | 0.23 | 0.21 | 0.03 | 0.05 | 0.04 | 1.22 [1.17-1.28]* | 1.17 [1.12-1.22]* |
| I33 | 4,440 | 114,230,360 | 0.05 | 0.04 | 0.03 | 0.01 | 0.01 | 0.01 | 1.35 [1.23-1.50]* | 1.30 [1.17-1.44]* |
| I34 | 60,900 | 113,768,400 | 0.58 | 0.52 | 0.49 | 0.05 | 0.09 | 0.07 | 1.18 [1.14-1.21]* | 1.13 [1.10-1.16]* |
| I35 | 88,920 | 113,628,130 | 0.87 | 0.76 | 0.71 | 0.11 | 0.16 | 0.13 | 1.23 [1.20-1.26]* | 1.17 [1.15-1.20]* |
| I36 | 6,560 | 114,238,880 | 0.06 | 0.06 | 0.05 | 0.01 | 0.01 | 0.01 | 1.23 [1.12-1.34]* | 1.16 [1.06-1.26]* |
| I37 | 7,200 | 114,227,670 | 0.07 | 0.06 | 0.06 | 0.01 | 0.01 | 0.01 | 1.22 [1.12-1.32]* | 1.15 [1.06-1.25]* |
| I38 | 9,950 | 114,156,420 | 0.10 | 0.08 | 0.08 | 0.02 | 0.02 | 0.02 | 1.29 [1.21-1.38]* | 1.23 [1.15-1.32]* |
| I40 | 750 | 114,261,620 | 0.01 | 0.01 | 0.01 | 0.00 | 0.00 | 0.00 | 1.29 [1.01-1.64] | 1.23 [0.96-1.59] |
| I42 | 22,830 | 114,017,610 | 0.22 | 0.20 | 0.18 | 0.02 | 0.04 | 0.03 | 1.21 [1.16-1.27]* | 1.15 [1.10-1.20]* |
| I43 | 960 | 114,266,280 | 0.01 | 0.01 | 0.01 | 0.00 | 0.00 | 0.00 | 0.98 [0.78-1.25] | 0.94 [0.74-1.20] |
| I44 | 139,340 | 113,350,830 | 1.36 | 1.20 | 1.13 | 0.16 | 0.23 | 0.18 | 1.20 [1.18-1.22]* | 1.16 [1.13-1.18]* |
| I45 | 87,760 | 113,679,790 | 0.85 | 0.75 | 0.71 | 0.10 | 0.14 | 0.11 | 1.20 [1.18-1.23]* | 1.15 [1.12-1.18]* |
| I46 | 45,670 | 114,131,180 | 0.44 | 0.39 | 0.36 | 0.05 | 0.08 | 0.07 | 1.23 [1.19-1.27]* | 1.19 [1.15-1.23]* |
| I47 | 51,050 | 113,582,230 | 0.50 | 0.44 | 0.41 | 0.06 | 0.09 | 0.07 | 1.22 [1.19-1.26]* | 1.15 [1.12-1.19]* |
| I48 | 387,240 | 111,079,210 | 3.79 | 3.41 | 3.26 | 0.37 | 0.53 | 0.42 | 1.16 [1.15-1.18]* | 1.13 [1.11-1.14]* |
| I49 | 60,360 | 113,711,580 | 0.58 | 0.52 | 0.49 | 0.06 | 0.09 | 0.06 | 1.18 [1.15-1.22]* | 1.12 [1.09-1.15]* |
| I50 | 294,080 | 112,545,450 | 2.99 | 2.52 | 2.37 | 0.47 | 0.62 | 0.51 | 1.26 [1.25-1.28]* | 1.20 [1.19-1.22]* |
| I51 | 164,050 | 113,262,330 | 1.63 | 1.41 | 1.31 | 0.22 | 0.32 | 0.25 | 1.25 [1.23-1.27]* | 1.18 [1.16-1.21]* |
| I60 | 9,660 | 114,144,600 | 0.09 | 0.08 | 0.08 | 0.00 | 0.01 | 0.01 | 1.16 [1.08-1.24]* | 1.15 [1.07-1.24]* |
| I61 | 24,080 | 114,113,730 | 0.22 | 0.21 | 0.19 | 0.02 | 0.03 | 0.03 | 1.17 [1.12-1.23]* | 1.16 [1.11-1.22]* |
| I62 | 13,830 | 114,182,210 | 0.13 | 0.12 | 0.11 | 0.01 | 0.02 | 0.02 | 1.16 [1.09-1.23]* | 1.14 [1.07-1.21]* |
| I63 | 113,000 | 113,453,940 | 1.01 | 0.99 | 0.93 | 0.02 | 0.08 | 0.07 | 1.09 [1.07-1.11]* | 1.08 [1.06-1.10]* |
| I64 | 29,380 | 113,926,560 | 0.27 | 0.26 | 0.23 | 0.01 | 0.03 | 0.03 | 1.15 [1.10-1.19]* | 1.13 [1.08-1.18]* |
| I65 | 27,300 | 114,004,160 | 0.26 | 0.24 | 0.22 | 0.02 | 0.04 | 0.03 | 1.17 [1.13-1.22]* | 1.14 [1.10-1.19]* |
| I66 | 1,780 | 114,255,750 | 0.02 | 0.02 | 0.01 | 0.00 | 0.00 | 0.00 | 1.15 [0.97-1.36] | 1.11 [0.94-1.32] |
| I67 | 170,160 | 113,461,500 | 1.63 | 1.47 | 1.38 | 0.17 | 0.25 | 0.21 | 1.18 [1.16-1.20]* | 1.15 [1.13-1.17]* |
| I68 | 1,000 | 114,267,090 | 0.01 | 0.01 | 0.01 | 0.00 | 0.00 | 0.00 | 1.22 [0.99-1.52] | 1.20 [0.96-1.49] |
| I69 | 50,450 | 113,820,340 | 0.48 | 0.44 | 0.40 | 0.04 | 0.07 | 0.06 | 1.18 [1.15-1.22]* | 1.15 [1.12-1.19]* |
| I70 | 42,380 | 113,957,290 | 0.42 | 0.36 | 0.33 | 0.06 | 0.09 | 0.07 | 1.26 [1.21-1.30]* | 1.21 [1.17-1.26]* |
| I71 | 51,970 | 113,903,530 | 0.48 | 0.45 | 0.42 | 0.04 | 0.07 | 0.06 | 1.17 [1.13-1.20]* | 1.15 [1.12-1.19]* |
| I72 | 12,450 | 114,157,250 | 0.12 | 0.11 | 0.10 | 0.01 | 0.02 | 0.01 | 1.17 [1.10-1.25]* | 1.14 [1.07-1.21]* |
| I73 | 93,220 | 113,349,500 | 1.00 | 0.78 | 0.72 | 0.22 | 0.28 | 0.24 | 1.40 [1.37-1.43]* | 1.32 [1.29-1.35]* |
| I74 | 24,020 | 114,036,070 | 0.22 | 0.21 | 0.19 | 0.02 | 0.03 | 0.03 | 1.18 [1.13-1.23]* | 1.14 [1.09-1.19]* |
| I77 | 31,750 | 113,922,830 | 0.32 | 0.27 | 0.25 | 0.06 | 0.08 | 0.07 | 1.32 [1.27-1.37]* | 1.26 [1.21-1.31]* |
| I78 | 8,230 | 114,163,730 | 0.08 | 0.07 | 0.06 | 0.01 | 0.02 | 0.02 | 1.30 [1.21-1.40]* | 1.24 [1.15-1.34]* |
| I79 | 3,130 | 114,240,520 | 0.03 | 0.03 | 0.02 | 0.01 | 0.01 | 0.01 | 1.34 [1.19-1.51]* | 1.24 [1.10-1.40]* |
| I80 | 64,000 | 113,400,140 | 0.63 | 0.55 | 0.50 | 0.08 | 0.13 | 0.10 | 1.25 [1.22-1.29]* | 1.20 [1.17-1.23]* |
| I81 | 3,600 | 114,253,110 | 0.03 | 0.03 | 0.03 | 0.00 | 0.00 | 0.00 | 1.14 [1.01-1.28] | 1.08 [0.96-1.22] |
| I82 | 8,300 | 114,204,010 | 0.08 | 0.07 | 0.07 | 0.01 | 0.01 | 0.01 | 1.21 [1.12-1.30]* | 1.15 [1.07-1.25]* |
| I83 | 73,770 | 112,508,400 | 0.89 | 0.60 | 0.55 | 0.29 | 0.34 | 0.32 | 1.63 [1.59-1.67]* | 1.57 [1.54-1.61]* |
| I84 | 115,820 | 111,349,500 | 1.22 | 1.00 | 0.89 | 0.22 | 0.33 | 0.26 | 1.37 [1.34-1.39]* | 1.28 [1.25-1.30]* |
| I85 | 9,960 | 114,176,370 | 0.11 | 0.08 | 0.07 | 0.03 | 0.03 | 0.03 | 1.46 [1.37-1.55]* | 1.37 [1.29-1.46]* |
| I86 | 8,510 | 114,148,180 | 0.09 | 0.07 | 0.07 | 0.02 | 0.02 | 0.02 | 1.31 [1.22-1.41]* | 1.22 [1.14-1.32]* |
| I87 | 16,270 | 114,150,460 | 0.19 | 0.13 | 0.12 | 0.06 | 0.07 | 0.07 | 1.60 [1.52-1.69]* | 1.52 [1.44-1.60]* |
| I88 | 14,270 | 114,038,230 | 0.17 | 0.11 | 0.13 | 0.06 | 0.04 | 0.03 | 1.28 [1.22-1.36]* | 1.25 [1.18-1.32]* |
| I89 | 21,910 | 114,120,760 | 0.25 | 0.18 | 0.16 | 0.07 | 0.09 | 0.08 | 1.53 [1.46-1.59]* | 1.43 [1.37-1.50]* |
| I95 | 207,390 | 113,135,460 | 2.03 | 1.79 | 1.69 | 0.24 | 0.34 | 0.27 | 1.20 [1.18-1.22]* | 1.16 [1.14-1.17]* |
| I97 | 2,640 | 114,215,960 | 0.03 | 0.02 | 0.02 | 0.00 | 0.01 | 0.01 | 1.33 [1.17-1.51]* | 1.26 [1.10-1.43]* |
| I98 | 6,720 | 114,222,560 | 0.07 | 0.06 | 0.05 | 0.02 | 0.02 | 0.02 | 1.45 [1.34-1.56]* | 1.34 [1.24-1.45]* |
| I99 | 3,840 | 114,232,630 | 0.04 | 0.03 | 0.03 | 0.00 | 0.01 | 0.01 | 1.25 [1.11-1.39]* | 1.20 [1.07-1.34]* |
| any age - J00-J99 | | | | | | | | | | |
| J00 | 13,540 | 113,939,120 | 0.16 | 0.11 | 0.11 | 0.05 | 0.05 | 0.04 | 1.41 [1.33-1.48]* | 1.33 [1.26-1.40]* |
| J01 | 3,390 | 114,210,530 | 0.04 | 0.03 | 0.03 | 0.01 | 0.01 | 0.01 | 1.46 [1.31-1.63]* | 1.31 [1.17-1.47]* |
| J02 | 25,070 | 113,831,290 | 0.28 | 0.21 | 0.21 | 0.08 | 0.07 | 0.06 | 1.32 [1.27-1.38]* | 1.24 [1.19-1.29]* |
| J03 | 130,520 | 111,894,800 | 1.49 | 1.09 | 1.22 | 0.40 | 0.27 | 0.22 | 1.22 [1.20-1.24]* | 1.17 [1.15-1.19]* |
| J04 | 3,040 | 114,214,990 | 0.03 | 0.03 | 0.02 | 0.01 | 0.01 | 0.01 | 1.34 [1.19-1.51]* | 1.23 [1.09-1.39]* |
| J05 | 22,750 | 113,550,870 | 0.22 | 0.20 | 0.22 | 0.03 | 0.00 | 0.00 | 1.02 [0.97-1.06] | 0.99 [0.95-1.04] |
| J06 | 86,740 | 110,834,210 | 1.03 | 0.72 | 0.80 | 0.31 | 0.24 | 0.20 | 1.30 [1.27-1.33]* | 1.24 [1.21-1.27]* |
| J10 | 20,020 | 114,149,530 | 0.22 | 0.16 | 0.16 | 0.05 | 0.06 | 0.04 | 1.35 [1.28-1.41]* | 1.22 [1.16-1.28]* |
| J11 | 4,260 | 114,202,770 | 0.05 | 0.04 | 0.03 | 0.01 | 0.01 | 0.01 | 1.35 [1.22-1.49]* | 1.24 [1.12-1.38]* |
| J12 | 31,080 | 114,194,250 | 0.32 | 0.26 | 0.25 | 0.06 | 0.07 | 0.05 | 1.28 [1.23-1.33]* | 1.19 [1.14-1.24]* |
| J13 | 6,920 | 114,195,510 | 0.07 | 0.06 | 0.05 | 0.01 | 0.02 | 0.01 | 1.32 [1.22-1.43]* | 1.21 [1.12-1.32]* |
| J14 | 2,900 | 114,244,570 | 0.03 | 0.02 | 0.02 | 0.01 | 0.01 | 0.00 | 1.33 [1.17-1.50]* | 1.17 [1.03-1.33] |
| J15 | 15,170 | 114,125,420 | 0.16 | 0.13 | 0.12 | 0.04 | 0.04 | 0.03 | 1.38 [1.31-1.45]* | 1.26 [1.19-1.33]* |
| J17 | 2,990 | 114,245,460 | 0.03 | 0.02 | 0.02 | 0.01 | 0.01 | 0.01 | 1.49 [1.32-1.67]* | 1.38 [1.23-1.56]* |

| | | | | | | | | | | |
|---|---|---|---|---|---|---|---|---|---|---|
| J18 | 438,130 | 111,330,990 | 4.54 | 3.79 | 3.57 | 0.75 | 0.97 | 0.78 | 1.27 [1.26-1.28]* | 1.21 [1.20-1.22]* |
| J20 | 5,960 | 114,163,670 | 0.06 | 0.05 | 0.05 | 0.01 | 0.01 | 0.01 | 1.29 [1.18-1.40]* | 1.18 [1.08-1.29]* |
| J21 | 5,040 | 112,463,840 | 0.05 | 0.04 | 0.05 | 0.01 | 0.01 | 0.00 | 1.14 [1.05-1.25]* | 1.07 [0.97-1.17] |
| J22 | 285,020 | 110,816,300 | 3.19 | 2.43 | 2.34 | 0.76 | 0.85 | 0.68 | 1.36 [1.35-1.38]* | 1.27 [1.26-1.29]* |
| J30 | 51,600 | 113,739,920 | 0.81 | 0.37 | 0.38 | 0.45 | 0.43 | 0.34 | 2.14 [2.08-2.20]* | 1.73 [1.68-1.78]* |
| J31 | 11,650 | 114,053,390 | 0.15 | 0.09 | 0.09 | 0.06 | 0.07 | 0.05 | 1.75 [1.66-1.86]* | 1.44 [1.35-1.53]* |
| J32 | 27,210 | 113,745,000 | 0.30 | 0.22 | 0.21 | 0.08 | 0.09 | 0.06 | 1.42 [1.36-1.47]* | 1.22 [1.18-1.28]* |
| J33 | 17,910 | 113,746,650 | 0.20 | 0.15 | 0.13 | 0.06 | 0.07 | 0.05 | 1.56 [1.49-1.64]* | 1.29 [1.23-1.36]* |
| J34 | 63,770 | 112,468,740 | 0.76 | 0.52 | 0.50 | 0.24 | 0.25 | 0.20 | 1.51 [1.47-1.54]* | 1.35 [1.31-1.38]* |
| J35 | 101,640 | 111,518,930 | 1.21 | 0.84 | 0.94 | 0.36 | 0.27 | 0.23 | 1.28 [1.26-1.31]* | 1.23 [1.21-1.26]* |
| J36 | 11,690 | 114,030,650 | 0.11 | 0.10 | 0.10 | 0.01 | 0.01 | 0.01 | 1.14 [1.07-1.22]* | 1.11 [1.04-1.18]* |
| J37 | 610 | 114,251,570 | 0.01 | 0.01 | 0.00 | 0.00 | 0.00 | 0.00 | 1.39 [1.07-1.81] | 1.24 [0.95-1.63] |
| J38 | 20,980 | 113,877,360 | 0.21 | 0.18 | 0.16 | 0.04 | 0.05 | 0.04 | 1.30 [1.24-1.36]* | 1.21 [1.15-1.27]* |
| J39 | 11,180 | 114,102,740 | 0.12 | 0.09 | 0.09 | 0.03 | 0.03 | 0.02 | 1.33 [1.25-1.42]* | 1.23 [1.15-1.31]* |
| J40 | 11,910 | 114,110,590 | 0.13 | 0.10 | 0.09 | 0.03 | 0.04 | 0.03 | 1.44 [1.36-1.53]* | 1.29 [1.21-1.37]* |
| J41 | 580 | 114,263,520 | 0.01 | 0.00 | 0.00 | 0.00 | 0.00 | 0.00 | 1.44 [1.09-1.89]* | 1.32 [0.99-1.74] |
| J42 | 4,820 | 114,204,800 | 0.05 | 0.04 | 0.04 | 0.01 | 0.02 | 0.01 | 1.45 [1.32-1.60]* | 1.28 [1.16-1.41]* |
| J43 | 65,960 | 113,834,960 | 0.69 | 0.55 | 0.51 | 0.13 | 0.18 | 0.14 | 1.35 [1.32-1.39]* | 1.25 [1.22-1.29]* |
| J44 | 262,890 | 111,926,660 | 2.94 | 2.21 | 2.02 | 0.73 | 0.92 | 0.66 | 1.46 [1.44-1.48]* | 1.29 [1.27-1.31]* |
| J45 | 503,210 | 103,273,790 | 8.34 | 4.05 | 3.93 | 4.29 | 4.41 | 3.16 | 2.12 [2.11-2.14]* | 1.61 [1.59-1.62]* |
| J46 | 9,490 | 113,926,720 | 0.18 | 0.06 | 0.06 | 0.12 | 0.12 | 0.10 | 2.95 [2.79-3.13]* | 2.29 [2.15-2.44]* |
| J47 | 48,030 | 113,922,430 | 0.54 | 0.39 | 0.36 | 0.15 | 0.18 | 0.12 | 1.49 [1.45-1.54]* | 1.29 [1.25-1.33]* |
| J61 | 6,510 | 114,232,750 | 0.07 | 0.05 | 0.05 | 0.01 | 0.02 | 0.02 | 1.39 [1.28-1.52]* | 1.32 [1.21-1.43]* |
| J67 | 1,930 | 114,256,010 | 0.02 | 0.02 | 0.01 | 0.01 | 0.01 | 0.01 | 1.61 [1.39-1.86]* | 1.44 [1.24-1.67]* |
| J69 | 64,700 | 114,106,290 | 0.62 | 0.55 | 0.53 | 0.06 | 0.09 | 0.08 | 1.17 [1.14-1.20]* | 1.15 [1.12-1.18]* |
| J70 | 1,310 | 114,263,830 | 0.01 | 0.01 | 0.01 | 0.00 | 0.01 | 0.00 | 1.52 [1.27-1.82]* | 1.42 [1.18-1.70]* |
| J80 | 5,040 | 114,235,320 | 0.05 | 0.04 | 0.04 | 0.01 | 0.01 | 0.01 | 1.37 [1.25-1.51]* | 1.31 [1.19-1.44]* |
| J81 | 22,390 | 114,120,810 | 0.22 | 0.19 | 0.17 | 0.03 | 0.05 | 0.05 | 1.30 [1.24-1.36]* | 1.25 [1.20-1.31]* |
| J82 | 1,750 | 114,259,390 | 0.03 | 0.01 | 0.01 | 0.01 | 0.02 | 0.01 | 2.40 [2.08-2.78]* | 1.72 [1.47-2.02]* |
| J84 | 41,200 | 114,083,230 | 0.45 | 0.34 | 0.31 | 0.11 | 0.14 | 0.12 | 1.44 [1.39-1.49]* | 1.36 [1.31-1.41]* |
| J85 | 2,310 | 114,251,300 | 0.02 | 0.02 | 0.02 | 0.00 | 0.01 | 0.00 | 1.31 [1.14-1.51]* | 1.21 [1.05-1.39]* |
| J86 | 8,030 | 114,183,450 | 0.08 | 0.07 | 0.06 | 0.01 | 0.01 | 0.01 | 1.20 [1.12-1.30]* | 1.17 [1.08-1.26]* |
| J90 | 210,860 | 113,227,020 | 2.03 | 1.82 | 1.71 | 0.21 | 0.33 | 0.28 | 1.19 [1.17-1.21]* | 1.16 [1.14-1.18]* |
| J91 | 5,070 | 114,250,760 | 0.05 | 0.04 | 0.04 | 0.01 | 0.01 | 0.01 | 1.24 [1.12-1.37]* | 1.21 [1.09-1.33]* |
| J92 | 23,580 | 114,154,380 | 0.23 | 0.20 | 0.18 | 0.03 | 0.05 | 0.04 | 1.26 [1.20-1.31]* | 1.22 [1.16-1.27]* |
| J93 | 20,490 | 114,050,810 | 0.19 | 0.18 | 0.17 | 0.01 | 0.02 | 0.02 | 1.15 [1.09-1.20]* | 1.12 [1.06-1.17]* |
| J94 | 7,920 | 114,221,710 | 0.07 | 0.07 | 0.06 | 0.01 | 0.01 | 0.01 | 1.18 [1.09-1.28]* | 1.15 [1.07-1.25]* |
| J95 | 6,590 | 114,177,850 | 0.06 | 0.06 | 0.05 | 0.01 | 0.01 | 0.01 | 1.21 [1.11-1.31]* | 1.17 [1.08-1.27]* |
| J96 | 134,510 | 113,785,820 | 1.38 | 1.14 | 1.05 | 0.24 | 0.33 | 0.26 | 1.32 [1.30-1.34]* | 1.23 [1.21-1.25]* |
| J98 | 129,100 | 113,382,360 | 1.29 | 1.10 | 1.03 | 0.19 | 0.26 | 0.19 | 1.25 [1.23-1.28]* | 1.17 [1.15-1.20]* |
| J99 | 3,890 | 114,238,600 | 0.05 | 0.03 | 0.03 | 0.02 | 0.02 | 0.01 | 1.60 [1.45-1.77]* | 1.39 [1.25-1.54]* |
| any age - K00-K95 | | | | | | | | | | |
| K00 | 26,680 | 113,812,440 | 0.30 | 0.22 | 0.26 | 0.08 | 0.04 | 0.03 | 1.16 [1.12-1.21]* | 1.12 [1.08-1.17]* |
| K01 | 64,810 | 112,598,380 | 0.69 | 0.55 | 0.57 | 0.14 | 0.12 | 0.09 | 1.21 [1.18-1.25]* | 1.16 [1.13-1.19]* |
| K02 | 191,290 | 110,815,100 | 2.00 | 1.66 | 1.80 | 0.34 | 0.19 | 0.13 | 1.11 [1.09-1.13]* | 1.07 [1.05-1.09]* |
| K03 | 4,670 | 114,208,490 | 0.05 | 0.04 | 0.04 | 0.01 | 0.01 | 0.01 | 1.17 [1.07-1.29]* | 1.12 [1.01-1.23] |
| K04 | 41,400 | 113,490,440 | 0.41 | 0.35 | 0.36 | 0.06 | 0.05 | 0.03 | 1.14 [1.10-1.18]* | 1.09 [1.05-1.13]* |
| K05 | 36,920 | 113,595,030 | 0.39 | 0.31 | 0.30 | 0.08 | 0.09 | 0.07 | 1.29 [1.25-1.34]* | 1.22 [1.17-1.26]* |
| K06 | 4,860 | 114,196,150 | 0.05 | 0.04 | 0.04 | 0.01 | 0.01 | 0.01 | 1.38 [1.26-1.52]* | 1.31 [1.19-1.44]* |
| K07 | 38,970 | 113,505,900 | 0.42 | 0.32 | 0.36 | 0.10 | 0.07 | 0.05 | 1.19 [1.15-1.23]* | 1.14 [1.10-1.18]* |
| K08 | 57,980 | 113,263,340 | 0.59 | 0.49 | 0.49 | 0.09 | 0.09 | 0.07 | 1.19 [1.15-1.22]* | 1.13 [1.10-1.16]* |
| K09 | 4,320 | 114,162,120 | 0.04 | 0.04 | 0.04 | 0.01 | 0.01 | 0.01 | 1.26 [1.14-1.40]* | 1.24 [1.12-1.38]* |
| K10 | 5,530 | 114,199,020 | 0.06 | 0.05 | 0.05 | 0.01 | 0.01 | 0.01 | 1.25 [1.14-1.36]* | 1.15 [1.05-1.26]* |
| K11 | 13,230 | 114,035,250 | 0.14 | 0.11 | 0.11 | 0.03 | 0.03 | 0.03 | 1.31 [1.24-1.39]* | 1.24 [1.17-1.32]* |
| K12 | 20,820 | 114,033,530 | 0.22 | 0.17 | 0.17 | 0.05 | 0.05 | 0.04 | 1.29 [1.24-1.35]* | 1.24 [1.19-1.30]* |
| K13 | 27,380 | 113,790,920 | 0.30 | 0.23 | 0.22 | 0.07 | 0.08 | 0.07 | 1.37 [1.31-1.42]* | 1.30 [1.25-1.35]* |
| K14 | 13,280 | 114,066,920 | 0.14 | 0.11 | 0.10 | 0.03 | 0.04 | 0.03 | 1.39 [1.31-1.47]* | 1.28 [1.21-1.36]* |
| K20 | 81,790 | 112,805,980 | 0.85 | 0.70 | 0.64 | 0.15 | 0.21 | 0.15 | 1.33 [1.30-1.36]* | 1.22 [1.19-1.25]* |
| K21 | 315,740 | 109,902,060 | 3.44 | 2.74 | 2.54 | 0.70 | 0.90 | 0.60 | 1.36 [1.34-1.37]* | 1.21 [1.20-1.23]* |
| K22 | 120,180 | 112,636,160 | 1.22 | 1.03 | 0.94 | 0.19 | 0.28 | 0.21 | 1.29 [1.27-1.32]* | 1.21 [1.18-1.23]* |
| K23 | 1,390 | 114,260,330 | 0.02 | 0.01 | 0.01 | 0.00 | 0.01 | 0.00 | 1.54 [1.30-1.84]* | 1.38 [1.16-1.65]* |
| K25 | 59,100 | 113,430,030 | 0.60 | 0.50 | 0.46 | 0.09 | 0.14 | 0.11 | 1.29 [1.26-1.33]* | 1.22 [1.18-1.25]* |
| K26 | 42,600 | 113,641,690 | 0.40 | 0.37 | 0.34 | 0.04 | 0.07 | 0.06 | 1.19 [1.15-1.23]* | 1.16 [1.12-1.20]* |
| K27 | 7,230 | 114,168,840 | 0.07 | 0.06 | 0.06 | 0.01 | 0.02 | 0.01 | 1.29 [1.20-1.40]* | 1.20 [1.10-1.30]* |
| K28 | 1,040 | 114,258,320 | 0.01 | 0.01 | 0.01 | 0.00 | 0.00 | 0.00 | 1.47 [1.20-1.79]* | 1.24 [1.00-1.52] |
| K29 | 306,110 | 109,363,150 | 3.30 | 2.68 | 2.50 | 0.63 | 0.80 | 0.61 | 1.32 [1.31-1.34]* | 1.22 [1.21-1.24]* |
| K30 | 73,680 | 112,429,780 | 0.78 | 0.63 | 0.57 | 0.15 | 0.21 | 0.15 | 1.36 [1.33-1.39]* | 1.24 [1.21-1.27]* |
| K31 | 102,340 | 113,237,500 | 1.04 | 0.87 | 0.80 | 0.17 | 0.24 | 0.16 | 1.30 [1.27-1.33]* | 1.18 [1.16-1.21]* |
| K35 | 64,400 | 112,976,120 | 0.62 | 0.56 | 0.57 | 0.06 | 0.04 | 0.03 | 1.08 [1.05-1.11]* | 1.04 [1.01-1.07]* |
| K36 | 1,370 | 114,244,690 | 0.01 | 0.01 | 0.01 | 0.00 | 0.00 | 0.00 | 1.17 [0.98-1.40] | 1.15 [0.96-1.38] |
| K37 | 12,120 | 114,036,730 | 0.12 | 0.10 | 0.11 | 0.02 | 0.01 | 0.01 | 1.14 [1.07-1.21]* | 1.07 [1.01-1.14] |
| K38 | 8,010 | 114,148,810 | 0.08 | 0.07 | 0.07 | 0.01 | 0.01 | 0.01 | 1.15 [1.07-1.24]* | 1.10 [1.01-1.18] |
| K40 | 121,060 | 111,468,670 | 1.11 | 1.08 | 1.01 | 0.03 | 0.10 | 0.06 | 1.10 [1.07-1.12]* | 1.06 [1.04-1.08]* |
| K41 | 8,350 | 114,134,000 | 0.08 | 0.07 | 0.07 | 0.01 | 0.01 | 0.01 | 1.19 [1.10-1.28]* | 1.13 [1.05-1.22]* |
| K42 | 54,130 | 113,303,900 | 0.55 | 0.46 | 0.43 | 0.09 | 0.12 | 0.08 | 1.29 [1.25-1.32]* | 1.17 [1.14-1.21]* |
| K43 | 41,540 | 113,607,970 | 0.41 | 0.35 | 0.33 | 0.06 | 0.08 | 0.06 | 1.26 [1.22-1.30]* | 1.16 [1.12-1.20]* |
| K44 | 295,680 | 109,862,970 | 3.12 | 2.59 | 2.39 | 0.53 | 0.73 | 0.51 | 1.31 [1.29-1.32]* | 1.20 [1.18-1.21]* |
| K45 | 1,930 | 114,247,050 | 0.02 | 0.02 | 0.02 | 0.00 | 0.00 | 0.00 | 1.22 [1.04-1.42] | 1.12 [0.95-1.31] |
| K46 | 7,960 | 114,208,020 | 0.08 | 0.07 | 0.06 | 0.01 | 0.02 | 0.01 | 1.31 [1.21-1.41]* | 1.18 [1.09-1.28]* |
| K50 | 22,990 | 113,770,810 | 0.29 | 0.18 | 0.17 | 0.11 | 0.12 | 0.09 | 1.70 [1.63-1.77]* | 1.47 [1.41-1.54]* |
| K51 | 32,390 | 113,601,250 | 0.37 | 0.27 | 0.24 | 0.10 | 0.12 | 0.10 | 1.51 [1.46-1.56]* | 1.35 [1.30-1.40]* |
| K52 | 179,030 | 109,897,870 | 1.95 | 1.55 | 1.44 | 0.39 | 0.51 | 0.41 | 1.35 [1.33-1.38]* | 1.27 [1.25-1.29]* |
| K55 | 25,930 | 114,088,570 | 0.25 | 0.22 | 0.20 | 0.03 | 0.05 | 0.04 | 1.25 [1.20-1.30]* | 1.19 [1.14-1.24]* |
| K56 | 80,780 | 113,496,030 | 0.78 | 0.69 | 0.65 | 0.09 | 0.13 | 0.10 | 1.21 [1.18-1.24]* | 1.15 [1.12-1.17]* |
| K57 | 337,850 | 110,472,750 | 3.42 | 2.97 | 2.73 | 0.45 | 0.69 | 0.51 | 1.25 [1.24-1.27]* | 1.17 [1.16-1.19]* |
| K58 | 96,360 | 113,076,990 | 1.12 | 0.79 | 0.74 | 0.33 | 0.37 | 0.24 | 1.50 [1.47-1.53]* | 1.27 [1.24-1.30]* |
| K59 | 332,310 | 111,007,730 | 3.53 | 2.87 | 2.77 | 0.66 | 0.76 | 0.58 | 1.27 [1.26-1.29]* | 1.20 [1.18-1.21]* |
| K60 | 29,190 | 113,534,470 | 0.34 | 0.24 | 0.22 | 0.10 | 0.11 | 0.10 | 1.52 [1.47-1.58]* | 1.41 [1.36-1.46]* |
| K61 | 18,450 | 113,826,890 | 0.20 | 0.15 | 0.14 | 0.04 | 0.06 | 0.05 | 1.39 [1.33-1.46]* | 1.32 [1.26-1.39]* |
| K62 | 177,520 | 110,848,840 | 1.87 | 1.54 | 1.40 | 0.33 | 0.47 | 0.37 | 1.33 [1.31-1.35]* | 1.25 [1.23-1.27]* |
| K63 | 178,680 | 112,477,140 | 1.79 | 1.54 | 1.40 | 0.25 | 0.39 | 0.28 | 1.27 [1.25-1.30]* | 1.19 [1.17-1.21]* |
| K64 | 126,200 | 113,775,740 | 1.30 | 1.06 | 0.99 | 0.24 | 0.31 | 0.23 | 1.31 [1.29-1.34]* | 1.21 [1.19-1.23]* |
| K65 | 19,400 | 114,105,660 | 0.18 | 0.17 | 0.15 | 0.02 | 0.03 | 0.02 | 1.19 [1.13-1.25]* | 1.16 [1.10-1.21]* |
| K66 | 68,500 | 113,521,050 | 0.66 | 0.59 | 0.55 | 0.07 | 0.11 | 0.07 | 1.19 [1.16-1.22]* | 1.12 [1.09-1.15]* |
| K70 | 22,450 | 114,021,850 | 0.24 | 0.19 | 0.17 | 0.05 | 0.07 | 0.06 | 1.44 [1.38-1.51]* | 1.37 [1.31-1.43]* |
| K71 | 1,450 | 114,253,650 | 0.02 | 0.01 | 0.01 | 0.00 | 0.00 | 0.00 | 1.37 [1.15-1.62]* | 1.29 [1.08-1.54]* |
| K72 | 14,090 | 114,212,050 | 0.15 | 0.12 | 0.11 | 0.03 | 0.04 | 0.03 | 1.36 [1.28-1.43]* | 1.28 [1.21-1.36]* |
| K73 | 1,380 | 114,241,240 | 0.02 | 0.01 | 0.01 | 0.00 | 0.00 | 0.00 | 1.48 [1.25-1.75]* | 1.39 [1.16-1.65]* |
| K74 | 20,900 | 114,113,730 | 0.23 | 0.17 | 0.16 | 0.06 | 0.07 | 0.06 | 1.46 [1.39-1.52]* | 1.35 [1.29-1.42]* |
| K75 | 13,910 | 114,143,540 | 0.15 | 0.11 | 0.11 | 0.04 | 0.04 | 0.04 | 1.40 [1.33-1.48]* | 1.30 [1.23-1.38]* |
| K76 | 100,850 | 113,575,820 | 1.05 | 0.85 | 0.79 | 0.20 | 0.26 | 0.19 | 1.33 [1.31-1.36]* | 1.23 [1.20-1.25]* |
| K80 | 191,490 | 111,433,250 | 1.83 | 1.69 | 1.57 | 0.14 | 0.26 | 0.17 | 1.16 [1.14-1.18]* | 1.10 [1.09-1.12]* |
| K81 | 35,060 | 113,724,020 | 0.33 | 0.30 | 0.28 | 0.03 | 0.05 | 0.04 | 1.19 [1.15-1.23]* | 1.12 [1.08-1.17]* |
| K82 | 26,820 | 113,922,940 | 0.25 | 0.23 | 0.21 | 0.02 | 0.04 | 0.03 | 1.18 [1.13-1.23]* | 1.11 [1.07-1.16]* |
| K83 | 50,290 | 113,808,320 | 0.47 | 0.44 | 0.41 | 0.03 | 0.06 | 0.05 | 1.16 [1.12-1.19]* | 1.11 [1.08-1.15]* |
| K85 | 33,650 | 113,797,680 | 0.32 | 0.29 | 0.27 | 0.03 | 0.05 | 0.04 | 1.20 [1.16-1.25]* | 1.14 [1.10-1.18]* |
| K86 | 28,040 | 114,026,110 | 0.27 | 0.24 | 0.22 | 0.03 | 0.05 | 0.03 | 1.21 [1.17-1.27]* | 1.15 [1.10-1.19]* |
| K90 | 28,570 | 113,768,930 | 0.39 | 0.22 | 0.22 | 0.17 | 0.17 | 0.14 | 1.78 [1.72-1.85]* | 1.56 [1.50-1.62]* |

| | | | | | | | | | | |
|---|---|---|---|---|---|---|---|---|---|---|
| K91 | 25,800 | 113,951,270 | 0.26 | 0.22 | 0.20 | 0.04 | 0.06 | 0.04 | 1.29 [1.24-1.35]* | 1.20 [1.15-1.25]* |
| K92 | 224,610 | 112,058,810 | 2.31 | 1.93 | 1.80 | 0.38 | 0.51 | 0.40 | 1.28 [1.27-1.30]* | 1.21 [1.19-1.23]* |
| any age - L00-L99 | | | | | | | | | | |
| L00 | 720 | 114,229,250 | 0.01 | 0.01 | 0.01 | 0.00 | 0.00 | 0.00 | 1.54 [1.22-1.93]* | 1.54 [1.22-1.93]* |
| L01 | 3,670 | 114,151,010 | 0.06 | 0.03 | 0.03 | 0.04 | 0.03 | 0.03 | 2.28 [2.08-2.51]* | 2.10 [1.91-2.32]* |
| L02 | 50,310 | 113,294,260 | 0.55 | 0.42 | 0.40 | 0.13 | 0.15 | 0.13 | 1.38 [1.34-1.42]* | 1.30 [1.26-1.34]* |
| L03 | 175,050 | 112,175,720 | 2.12 | 1.43 | 1.32 | 0.70 | 0.80 | 0.73 | 1.61 [1.58-1.63]* | 1.53 [1.50-1.55]* |
| L04 | 5,080 | 114,164,240 | 0.05 | 0.04 | 0.05 | 0.01 | 0.01 | 0.01 | 1.21 [1.11-1.33]* | 1.19 [1.08-1.30]* |
| L05 | 17,880 | 113,734,920 | 0.17 | 0.15 | 0.15 | 0.02 | 0.02 | 0.02 | 1.15 [1.09-1.21]* | 1.11 [1.05-1.17]* |
| L08 | 47,270 | 113,790,560 | 0.55 | 0.38 | 0.36 | 0.17 | 0.19 | 0.18 | 1.54 [1.50-1.59]* | 1.47 [1.43-1.52]* |
| L10 | 560 | 114,264,510 | 0.01 | 0.00 | 0.00 | 0.00 | 0.00 | 0.00 | 2.28 [1.75-2.95]* | 2.11 [1.62-2.76]* |
| L11 | 460 | 114,264,280 | 0.01 | 0.00 | 0.00 | 0.00 | 0.00 | 0.00 | 1.67 [1.24-2.25]* | 1.57 [1.16-2.13]* |
| L12 | 3,810 | 114,246,770 | 0.06 | 0.03 | 0.02 | 0.04 | 0.04 | 0.04 | 2.86 [2.59-3.16]* | 2.75 [2.48-3.04]* |
| L13 | 680 | 114,260,200 | 0.01 | 0.00 | 0.00 | 0.01 | 0.01 | 0.01 | 3.14 [2.51-3.92]* | 2.84 [2.26-3.58]* |
| L20 | 7,190 | 113,992,090 | 0.28 | 0.01 | 0.01 | 0.27 | 0.27 | 0.27 | 24.72 [22.44-27.23]* | 21.74 [19.72-23.97]* |
| L21 | 1,860 | 114,212,560 | 0.04 | 0.01 | 0.01 | 0.02 | 0.02 | 0.02 | 3.29 [2.87-3.77]* | 3.09 [2.69-3.56]* |
| L22 | 2,060 | 114,160,750 | 0.02 | 0.02 | 0.02 | 0.00 | 0.00 | 0.00 | 1.05 [0.90-1.22] | 1.01 [0.87-1.18] |
| L23 | 2,260 | 114,222,200 | 0.05 | 0.01 | 0.01 | 0.04 | 0.04 | 0.03 | 3.75 [3.33-4.22]* | 3.33 [2.94-3.76]* |
| L24 | 10,730 | 114,250,180 | 0.12 | 0.09 | 0.08 | 0.03 | 0.04 | 0.03 | 1.45 [1.35-1.55]* | 1.39 [1.30-1.49]* |
| L25 | 2,100 | 114,239,310 | 0.04 | 0.01 | 0.01 | 0.03 | 0.03 | 0.03 | 2.99 [2.63-3.39]* | 2.84 [2.49-3.23]* |
| L26 | 220 | 114,267,870 | 0.01 | 0.00 | 0.00 | 0.00 | 0.00 | 0.00 | 6.48 [4.30-9.76]* | 6.06 [3.99-9.21]* |
| L27 | 10,870 | 114,129,140 | 0.14 | 0.08 | 0.08 | 0.06 | 0.06 | 0.05 | 1.75 [1.65-1.86]* | 1.61 [1.51-1.71]* |
| L28 | 3,570 | 114,212,920 | 0.08 | 0.02 | 0.02 | 0.06 | 0.06 | 0.06 | 4.46 [4.06-4.91]* | 4.14 [3.75-4.56]* |
| L29 | 17,360 | 113,985,790 | 0.22 | 0.14 | 0.13 | 0.09 | 0.09 | 0.08 | 1.72 [1.64-1.80]* | 1.60 [1.52-1.68]* |
| L30 | 109,410 | 112,014,620 | 3.64 | 0.35 | 0.35 | 3.29 | 3.29 | 3.26 | 10.30 [10.10-10.50]* | 9.48 [9.30-9.67]* |
| L40 | 42,900 | 113,784,710 | 0.64 | 0.31 | 0.29 | 0.33 | 0.35 | 0.34 | 2.23 [2.17-2.30]* | 2.13 [2.07-2.20]* |
| L42 | 730 | 114,262,580 | 0.01 | 0.01 | 0.01 | 0.00 | 0.00 | 0.00 | 1.54 [1.22-1.95]* | 1.44 [1.13-1.83]* |
| L43 | 4,550 | 114,196,140 | 0.07 | 0.03 | 0.03 | 0.03 | 0.04 | 0.03 | 2.18 [1.99-2.38]* | 2.03 [1.86-2.23]* |
| L44 | 260 | 114,265,110 | 0.00 | 0.00 | 0.00 | 0.00 | 0.00 | 0.00 | 3.12 [2.21-4.41]* | 2.92 [2.05-4.16]* |
| L50 | 12,880 | 114,038,850 | 0.19 | 0.09 | 0.10 | 0.10 | 0.09 | 0.07 | 1.93 [1.83-2.03]* | 1.64 [1.55-1.73]* |
| L51 | 3,180 | 114,198,300 | 0.03 | 0.03 | 0.03 | 0.01 | 0.01 | 0.01 | 1.28 [1.14-1.43]* | 1.21 [1.07-1.35]* |
| L52 | 1,260 | 114,245,010 | 0.02 | 0.01 | 0.01 | 0.00 | 0.00 | 0.00 | 1.49 [1.25-1.79]* | 1.38 [1.15-1.66]* |
| L53 | 15,850 | 114,084,140 | 0.20 | 0.12 | 0.12 | 0.08 | 0.09 | 0.08 | 1.75 [1.66-1.84]* | 1.66 [1.58-1.75]* |
| L55 | 540 | 114,260,680 | 0.01 | 0.00 | 0.00 | 0.00 | 0.00 | 0.00 | 1.15 [0.86-1.53] | 1.10 [0.82-1.48] |
| L56 | 620 | 114,260,860 | 0.01 | 0.00 | 0.00 | 0.01 | 0.01 | 0.01 | 2.80 [2.22-3.53]* | 2.65 [2.09-3.36]* |
| L57 | 34,630 | 113,880,160 | 0.36 | 0.29 | 0.27 | 0.06 | 0.09 | 0.07 | 1.32 [1.27-1.37]* | 1.25 [1.21-1.30]* |
| L59 | 520 | 114,264,400 | 0.01 | 0.00 | 0.00 | 0.00 | 0.00 | 0.00 | 1.42 [1.07-1.89] | 1.38 [1.02-1.85] |
| L60 | 14,740 | 113,848,160 | 0.17 | 0.12 | 0.12 | 0.05 | 0.05 | 0.04 | 1.41 [1.34-1.49]* | 1.30 [1.23-1.38]* |
| L63 | 720 | 114,260,800 | 0.01 | 0.00 | 0.00 | 0.01 | 0.01 | 0.01 | 2.87 [2.32-3.56]* | 2.27 [1.80-2.85]* |
| L65 | 2,500 | 114,241,560 | 0.04 | 0.02 | 0.02 | 0.02 | 0.02 | 0.02 | 1.99 [1.76-2.25]* | 1.75 [1.54-1.99]* |
| L66 | 500 | 114,262,820 | 0.01 | 0.00 | 0.00 | 0.00 | 0.00 | 0.00 | 1.73 [1.31-2.27]* | 1.62 [1.22-2.14]* |
| L68 | 910 | 114,251,310 | 0.01 | 0.01 | 0.01 | 0.00 | 0.00 | 0.00 | 1.25 [1.00-1.56] | 1.19 [0.95-1.49] |
| L70 | 3,340 | 114,232,290 | 0.04 | 0.03 | 0.03 | 0.01 | 0.01 | 0.01 | 1.45 [1.30-1.62]* | 1.32 [1.18-1.49]* |
| L71 | 3,530 | 114,234,320 | 0.04 | 0.03 | 0.03 | 0.01 | 0.01 | 0.01 | 1.55 [1.39-1.72]* | 1.44 [1.29-1.61]* |
| L72 | 53,830 | 112,748,340 | 0.50 | 0.47 | 0.43 | 0.03 | 0.07 | 0.05 | 1.16 [1.13-1.19]* | 1.11 [1.08-1.14]* |
| L73 | 6,820 | 114,132,620 | 0.08 | 0.05 | 0.05 | 0.03 | 0.03 | 0.02 | 1.57 [1.46-1.70]* | 1.44 [1.33-1.56]* |
| L80 | 1,580 | 114,253,190 | 0.02 | 0.01 | 0.01 | 0.01 | 0.01 | 0.01 | 1.97 [1.69-2.29]* | 1.80 [1.54-2.11]* |
| L81 | 8,340 | 114,122,460 | 0.09 | 0.07 | 0.07 | 0.02 | 0.02 | 0.02 | 1.29 [1.20-1.38]* | 1.23 [1.14-1.32]* |
| L82 | 23,390 | 113,887,380 | 0.22 | 0.20 | 0.18 | 0.02 | 0.04 | 0.03 | 1.22 [1.17-1.28]* | 1.16 [1.11-1.21]* |
| L83 | 1,600 | 114,246,470 | 0.02 | 0.01 | 0.01 | 0.01 | 0.01 | 0.01 | 1.67 [1.43-1.96]* | 1.56 [1.33-1.84]* |
| L84 | 2,570 | 114,227,230 | 0.03 | 0.02 | 0.02 | 0.01 | 0.01 | 0.01 | 1.49 [1.31-1.69]* | 1.39 [1.22-1.58]* |
| L85 | 9,870 | 114,147,190 | 0.12 | 0.08 | 0.07 | 0.04 | 0.04 | 0.04 | 1.62 [1.52-1.73]* | 1.54 [1.44-1.65]* |
| L88 | 570 | 114,263,170 | 0.01 | 0.00 | 0.00 | 0.00 | 0.00 | 0.00 | 1.56 [1.20-2.04]* | 1.37 [1.04-1.80] |
| L89 | 121,870 | 113,961,150 | 1.20 | 1.04 | 0.98 | 0.16 | 0.22 | 0.20 | 1.22 [1.20-1.25]* | 1.20 [1.17-1.22]* |
| L90 | 25,210 | 113,726,160 | 0.26 | 0.21 | 0.20 | 0.04 | 0.06 | 0.05 | 1.29 [1.24-1.34]* | 1.22 [1.17-1.27]* |
| L91 | 9,360 | 114,010,680 | 0.10 | 0.08 | 0.07 | 0.02 | 0.03 | 0.02 | 1.35 [1.26-1.44]* | 1.27 [1.19-1.36]* |
| L92 | 7,520 | 114,106,240 | 0.08 | 0.06 | 0.06 | 0.02 | 0.02 | 0.01 | 1.30 [1.20-1.40]* | 1.21 [1.12-1.31]* |
| L93 | 2,690 | 114,232,710 | 0.04 | 0.02 | 0.02 | 0.02 | 0.02 | 0.01 | 1.86 [1.66-2.10]* | 1.67 [1.48-1.88]* |
| L94 | 2,020 | 114,241,050 | 0.02 | 0.02 | 0.02 | 0.01 | 0.01 | 0.01 | 1.55 [1.35-1.78]* | 1.45 [1.26-1.68]* |
| L95 | 2,440 | 114,239,660 | 0.03 | 0.02 | 0.02 | 0.01 | 0.01 | 0.01 | 1.61 [1.42-1.83]* | 1.50 [1.31-1.71]* |
| L97 | 86,530 | 113,764,860 | 0.99 | 0.71 | 0.64 | 0.28 | 0.35 | 0.33 | 1.54 [1.51-1.58]* | 1.49 [1.46-1.53]* |
| L98 | 73,890 | 113,100,030 | 0.80 | 0.62 | 0.57 | 0.18 | 0.22 | 0.20 | 1.39 [1.36-1.42]* | 1.32 [1.29-1.36]* |
| any age - M00-M99 | | | | | | | | | | |
| M00 | 9,110 | 114,140,710 | 0.10 | 0.07 | 0.07 | 0.03 | 0.03 | 0.03 | 1.48 [1.39-1.59]* | 1.41 [1.32-1.51]* |
| M02 | 2,370 | 114,233,040 | 0.03 | 0.02 | 0.02 | 0.01 | 0.00 | 0.00 | 1.22 [1.07-1.40]* | 1.18 [1.03-1.35] |
| M05 | 10,180 | 114,141,640 | 0.11 | 0.08 | 0.08 | 0.02 | 0.03 | 0.03 | 1.40 [1.32-1.50]* | 1.35 [1.27-1.44]* |
| M06 | 74,500 | 113,334,180 | 0.80 | 0.62 | 0.57 | 0.18 | 0.23 | 0.19 | 1.40 [1.37-1.44]* | 1.31 [1.28-1.34]* |
| M07 | 8,830 | 114,157,050 | 0.11 | 0.07 | 0.06 | 0.04 | 0.05 | 0.04 | 1.75 [1.64-1.87]* | 1.62 [1.52-1.74]* |
| M08 | 3,230 | 114,204,690 | 0.04 | 0.03 | 0.03 | 0.01 | 0.01 | 0.01 | 1.33 [1.19-1.48]* | 1.28 [1.14-1.43]* |
| M10 | 84,410 | 113,647,810 | 0.88 | 0.71 | 0.65 | 0.17 | 0.23 | 0.20 | 1.36 [1.33-1.39]* | 1.29 [1.26-1.32]* |
| M11 | 9,460 | 114,187,290 | 0.10 | 0.08 | 0.08 | 0.02 | 0.02 | 0.02 | 1.29 [1.20-1.39]* | 1.21 [1.12-1.30]* |
| M12 | 2,050 | 114,239,420 | 0.02 | 0.02 | 0.02 | 0.00 | 0.01 | 0.00 | 1.40 [1.21-1.61]* | 1.27 [1.10-1.48]* |
| M13 | 213,590 | 112,162,460 | 2.26 | 1.82 | 1.68 | 0.44 | 0.57 | 0.44 | 1.34 [1.32-1.36]* | 1.24 [1.22-1.26]* |
| M14 | 2,240 | 114,243,800 | 0.03 | 0.02 | 0.02 | 0.01 | 0.01 | 0.01 | 1.61 [1.41-1.85]* | 1.46 [1.28-1.68]* |
| M15 | 78,160 | 113,535,800 | 0.83 | 0.66 | 0.60 | 0.17 | 0.23 | 0.17 | 1.38 [1.34-1.41]* | 1.26 [1.23-1.29]* |
| M16 | 131,070 | 112,378,840 | 1.20 | 1.16 | 1.06 | 0.04 | 0.13 | 0.10 | 1.13 [1.10-1.15]* | 1.09 [1.07-1.11]* |
| M17 | 211,420 | 110,949,980 | 2.13 | 1.85 | 1.70 | 0.28 | 0.43 | 0.32 | 1.25 [1.23-1.27]* | 1.18 [1.16-1.20]* |
| M18 | 11,530 | 114,116,800 | 0.12 | 0.10 | 0.09 | 0.03 | 0.04 | 0.03 | 1.43 [1.35-1.52]* | 1.27 [1.19-1.35]* |
| M19 | 330,800 | 111,418,910 | 3.55 | 2.83 | 2.62 | 0.72 | 0.93 | 0.71 | 1.36 [1.34-1.37]* | 1.25 [1.24-1.27]* |
| M20 | 50,220 | 113,125,210 | 0.51 | 0.43 | 0.39 | 0.08 | 0.12 | 0.09 | 1.30 [1.26-1.34]* | 1.22 [1.18-1.26]* |
| M21 | 30,160 | 113,891,470 | 0.30 | 0.26 | 0.25 | 0.04 | 0.05 | 0.03 | 1.19 [1.15-1.24]* | 1.12 [1.07-1.16]* |
| M22 | 9,680 | 114,002,890 | 0.10 | 0.08 | 0.08 | 0.02 | 0.02 | 0.02 | 1.32 [1.24-1.41]* | 1.20 [1.12-1.29]* |
| M23 | 98,050 | 111,891,390 | 0.95 | 0.86 | 0.78 | 0.09 | 0.17 | 0.12 | 1.22 [1.19-1.25]* | 1.14 [1.12-1.17]* |
| M24 | 39,650 | 113,571,400 | 0.40 | 0.34 | 0.32 | 0.06 | 0.08 | 0.06 | 1.26 [1.22-1.30]* | 1.17 [1.13-1.21]* |
| M25 | 225,360 | 111,218,950 | 2.38 | 1.94 | 1.84 | 0.44 | 0.54 | 0.40 | 1.29 [1.27-1.31]* | 1.20 [1.18-1.22]* |
| M30 | 2,040 | 114,229,470 | 0.02 | 0.02 | 0.02 | 0.01 | 0.01 | 0.01 | 1.39 [1.21-1.59]* | 1.28 [1.11-1.48]* |
| M31 | 11,970 | 114,148,760 | 0.12 | 0.10 | 0.09 | 0.02 | 0.03 | 0.03 | 1.34 [1.26-1.42]* | 1.26 [1.18-1.34]* |
| M32 | 5,620 | 114,173,440 | 0.07 | 0.04 | 0.04 | 0.03 | 0.03 | 0.03 | 1.79 [1.65-1.95]* | 1.63 [1.49-1.77]* |
| M33 | 1,280 | 114,251,860 | 0.02 | 0.01 | 0.01 | 0.01 | 0.01 | 0.01 | 2.07 [1.75-2.45]* | 2.00 [1.69-2.38]* |
| M34 | 2,210 | 114,239,620 | 0.03 | 0.02 | 0.02 | 0.01 | 0.01 | 0.01 | 1.75 [1.53-2.00]* | 1.62 [1.41-1.85]* |
| M35 | 57,660 | 113,762,580 | 0.64 | 0.47 | 0.45 | 0.17 | 0.20 | 0.16 | 1.44 [1.40-1.48]* | 1.33 [1.29-1.36]* |
| M36 | 540 | 114,261,490 | 0.01 | 0.00 | 0.00 | 0.00 | 0.00 | 0.00 | 1.43 [1.07-1.89] | 1.30 [0.97-1.74] |
| M40 | 13,790 | 114,197,230 | 0.14 | 0.12 | 0.11 | 0.02 | 0.03 | 0.02 | 1.25 [1.18-1.33]* | 1.18 [1.11-1.25]* |
| M41 | 39,770 | 113,936,200 | 0.41 | 0.34 | 0.33 | 0.07 | 0.08 | 0.06 | 1.24 [1.20-1.28]* | 1.16 [1.12-1.20]* |
| M42 | 810 | 114,261,550 | 0.01 | 0.01 | 0.01 | 0.00 | 0.00 | 0.00 | 1.13 [0.89-1.44] | 0.98 [0.76-1.26] |
| M43 | 31,610 | 113,918,390 | 0.32 | 0.27 | 0.25 | 0.06 | 0.07 | 0.05 | 1.29 [1.24-1.34]* | 1.18 [1.14-1.23]* |
| M45 | 7,300 | 114,164,290 | 0.08 | 0.06 | 0.05 | 0.02 | 0.03 | 0.02 | 1.50 [1.40-1.62]* | 1.39 [1.29-1.50]* |
| M46 | 24,800 | 114,021,180 | 0.27 | 0.21 | 0.19 | 0.06 | 0.08 | 0.06 | 1.42 [1.36-1.48]* | 1.28 [1.22-1.33]* |
| M47 | 140,600 | 112,837,020 | 1.48 | 1.19 | 1.09 | 0.29 | 0.39 | 0.28 | 1.35 [1.33-1.38]* | 1.23 [1.21-1.26]* |
| M48 | 82,960 | 113,519,610 | 0.84 | 0.71 | 0.65 | 0.13 | 0.19 | 0.13 | 1.29 [1.26-1.32]* | 1.19 [1.16-1.22]* |
| M49 | 3,000 | 114,256,140 | 0.03 | 0.03 | 0.02 | 0.00 | 0.00 | 0.00 | 1.08 [0.95-1.23] | 1.06 [0.93-1.21] |
| M50 | 21,770 | 114,024,370 | 0.22 | 0.18 | 0.17 | 0.04 | 0.06 | 0.04 | 1.33 [1.27-1.40]* | 1.22 [1.17-1.28]* |
| M51 | 105,880 | 112,836,700 | 1.08 | 0.91 | 0.83 | 0.17 | 0.25 | 0.17 | 1.30 [1.28-1.33]* | 1.19 [1.16-1.21]* |
| M53 | 6,730 | 114,163,050 | 0.07 | 0.06 | 0.05 | 0.01 | 0.01 | 0.01 | 1.26 [1.16-1.36]* | 1.12 [1.03-1.22]* |
| M54 | 267,020 | 110,400,930 | 2.82 | 2.32 | 2.17 | 0.50 | 0.65 | 0.45 | 1.30 [1.28-1.32]* | 1.19 [1.17-1.21]* |

| | | | | | | | | | | |
|---|---|---|---|---|---|---|---|---|---|---|
| M60 | 5,520 | 114,203,100 | 0.06 | 0.04 | 0.04 | 0.02 | 0.02 | 0.02 | 1.45 [1.33-1.58]* | 1.38 [1.27-1.51]* |
| M62 | 23,300 | 114,043,900 | 0.23 | 0.20 | 0.19 | 0.03 | 0.04 | 0.03 | 1.22 [1.17-1.28]* | 1.16 [1.11-1.22]* |
| M65 | 46,990 | 113,425,640 | 0.50 | 0.39 | 0.37 | 0.10 | 0.13 | 0.10 | 1.36 [1.32-1.40]* | 1.25 [1.21-1.29]* |
| M66 | 4,650 | 114,190,050 | 0.05 | 0.04 | 0.04 | 0.01 | 0.01 | 0.01 | 1.29 [1.17-1.42]* | 1.21 [1.10-1.34]* |
| M67 | 34,180 | 113,501,310 | 0.35 | 0.29 | 0.28 | 0.06 | 0.07 | 0.05 | 1.27 [1.22-1.31]* | 1.19 [1.14-1.23]* |
| M70 | 18,030 | 113,983,870 | 0.20 | 0.15 | 0.14 | 0.05 | 0.06 | 0.05 | 1.44 [1.37-1.51]* | 1.31 [1.25-1.38]* |
| M71 | 11,430 | 114,126,080 | 0.12 | 0.10 | 0.09 | 0.02 | 0.03 | 0.02 | 1.34 [1.26-1.42]* | 1.24 [1.16-1.32]* |
| M72 | 25,940 | 113,780,260 | 0.23 | 0.23 | 0.21 | 0.00 | 0.02 | 0.02 | 1.12 [1.07-1.17]* | 1.08 [1.03-1.13]* |
| M75 | 72,830 | 112,974,880 | 0.74 | 0.62 | 0.56 | 0.12 | 0.18 | 0.13 | 1.32 [1.28-1.35]* | 1.20 [1.17-1.23]* |
| M76 | 6,040 | 114,192,140 | 0.07 | 0.05 | 0.05 | 0.02 | 0.02 | 0.01 | 1.42 [1.31-1.54]* | 1.28 [1.18-1.40]* |
| M77 | 14,150 | 114,001,050 | 0.16 | 0.12 | 0.11 | 0.04 | 0.05 | 0.04 | 1.47 [1.40-1.56]* | 1.33 [1.26-1.40]* |
| M79 | 223,590 | 111,199,670 | 2.48 | 1.90 | 1.78 | 0.58 | 0.70 | 0.53 | 1.39 [1.37-1.41]* | 1.27 [1.25-1.29]* |
| M80 | 26,260 | 114,072,310 | 0.27 | 0.22 | 0.21 | 0.04 | 0.06 | 0.05 | 1.28 [1.23-1.34]* | 1.22 [1.16-1.27]* |
| M81 | 160,120 | 113,090,090 | 1.66 | 1.36 | 1.27 | 0.30 | 0.39 | 0.30 | 1.31 [1.28-1.33]* | 1.22 [1.20-1.24]* |
| M83 | 1,020 | 114,256,300 | 0.01 | 0.01 | 0.01 | 0.00 | 0.00 | 0.00 | 1.34 [1.09-1.65]* | 1.22 [0.98-1.51] |
| M84 | 24,110 | 113,895,630 | 0.24 | 0.21 | 0.19 | 0.03 | 0.04 | 0.03 | 1.23 [1.17-1.28]* | 1.16 [1.11-1.21]* |
| M85 | 35,720 | 114,049,690 | 0.37 | 0.30 | 0.28 | 0.07 | 0.09 | 0.06 | 1.31 [1.27-1.36]* | 1.20 [1.16-1.25]* |
| M86 | 16,520 | 114,088,300 | 0.18 | 0.14 | 0.13 | 0.04 | 0.05 | 0.04 | 1.37 [1.30-1.44]* | 1.31 [1.25-1.38]* |
| M87 | 7,040 | 114,168,920 | 0.07 | 0.06 | 0.05 | 0.01 | 0.02 | 0.01 | 1.28 [1.18-1.39]* | 1.21 [1.11-1.31]* |
| M88 | 4,420 | 114,196,890 | 0.04 | 0.04 | 0.03 | 0.00 | 0.01 | 0.00 | 1.17 [1.05-1.30]* | 1.13 [1.01-1.26] |
| M89 | 18,810 | 113,941,420 | 0.19 | 0.16 | 0.15 | 0.03 | 0.04 | 0.03 | 1.29 [1.23-1.35]* | 1.20 [1.14-1.26]* |
| M90 | 7,280 | 114,244,340 | 0.06 | 0.06 | 0.06 | 0.00 | 0.00 | 0.00 | 1.06 [0.97-1.15] | 1.06 [0.97-1.15] |
| M91 | 1,780 | 114,230,500 | 0.02 | 0.01 | 0.02 | 0.00 | 0.00 | 0.00 | 1.19 [1.02-1.39] | 1.16 [0.99-1.36] |
| M92 | 2,640 | 114,227,410 | 0.03 | 0.02 | 0.02 | 0.01 | 0.01 | 0.01 | 1.45 [1.28-1.64]* | 1.31 [1.16-1.50]* |
| M93 | 5,140 | 114,166,960 | 0.05 | 0.04 | 0.04 | 0.01 | 0.01 | 0.01 | 1.31 [1.20-1.44]* | 1.22 [1.11-1.34]* |
| M94 | 13,420 | 114,082,290 | 0.15 | 0.11 | 0.11 | 0.04 | 0.04 | 0.03 | 1.41 [1.33-1.49]* | 1.26 [1.19-1.34]* |
| M95 | 3,480 | 114,166,320 | 0.04 | 0.03 | 0.03 | 0.01 | 0.01 | 0.01 | 1.36 [1.21-1.51]* | 1.27 [1.13-1.42]* |
| M96 | 8,910 | 114,184,610 | 0.09 | 0.07 | 0.07 | 0.02 | 0.02 | 0.02 | 1.28 [1.19-1.38]* | 1.20 [1.12-1.30]* |
| M99 | 2,560 | 114,248,490 | 0.03 | 0.02 | 0.02 | 0.00 | 0.01 | 0.00 | 1.34 [1.17-1.53]* | 1.20 [1.05-1.38]* |
| any age - N00-N99 | | | | | | | | | | |
| N00 | 590 | 114,260,870 | 0.01 | 0.00 | 0.00 | 0.00 | 0.00 | 0.00 | 1.32 [1.01-1.73] | 1.30 [0.98-1.71] |
| N02 | 7,120 | 114,131,150 | 0.07 | 0.06 | 0.06 | 0.01 | 0.02 | 0.02 | 1.32 [1.22-1.43]* | 1.27 [1.17-1.37]* |
| N03 | 27,140 | 114,059,330 | 0.28 | 0.23 | 0.21 | 0.05 | 0.07 | 0.06 | 1.34 [1.28-1.39]* | 1.27 [1.22-1.32]* |
| N04 | 4,800 | 114,193,950 | 0.05 | 0.04 | 0.04 | 0.01 | 0.01 | 0.01 | 1.34 [1.22-1.48]* | 1.28 [1.16-1.41]* |
| N05 | 4,660 | 114,194,980 | 0.05 | 0.04 | 0.04 | 0.02 | 0.02 | 0.02 | 1.48 [1.35-1.63]* | 1.42 [1.29-1.56]* |
| N08 | 12,410 | 114,162,810 | 0.13 | 0.10 | 0.09 | 0.03 | 0.04 | 0.03 | 1.43 [1.35-1.52]* | 1.35 [1.27-1.43]* |
| N10 | 5,850 | 114,176,950 | 0.06 | 0.05 | 0.05 | 0.01 | 0.01 | 0.01 | 1.17 [1.07-1.28]* | 1.10 [1.01-1.21] |
| N11 | 2,670 | 114,219,050 | 0.03 | 0.02 | 0.02 | 0.00 | 0.01 | 0.00 | 1.25 [1.10-1.42]* | 1.20 [1.05-1.37]* |
| N12 | 32,490 | 113,816,420 | 0.32 | 0.28 | 0.27 | 0.04 | 0.05 | 0.03 | 1.18 [1.13-1.22]* | 1.10 [1.06-1.15]* |
| N13 | 68,690 | 113,613,290 | 0.60 | 0.61 | 0.57 | −0.01 | 0.03 | 0.03 | 1.06 [1.03-1.09]* | 1.05 [1.02-1.08]* |
| N14 | 960 | 114,264,240 | 0.01 | 0.01 | 0.01 | 0.00 | 0.00 | 0.00 | 1.34 [1.08-1.66]* | 1.29 [1.03-1.61] |
| N15 | 1,770 | 114,250,340 | 0.02 | 0.02 | 0.01 | 0.00 | 0.00 | 0.00 | 1.20 [1.02-1.41] | 1.13 [0.96-1.33] |
| N16 | 860 | 114,260,020 | 0.01 | 0.01 | 0.01 | 0.00 | 0.00 | 0.00 | 1.61 [1.29-2.00]* | 1.53 [1.22-1.92]* |
| N17 | 358,760 | 113,040,440 | 3.54 | 3.09 | 2.89 | 0.46 | 0.65 | 0.56 | 1.23 [1.21-1.24]* | 1.19 [1.17-1.20]* |
| N18 | 300,050 | 112,886,960 | 3.01 | 2.57 | 2.39 | 0.43 | 0.62 | 0.51 | 1.26 [1.24-1.28]* | 1.21 [1.19-1.22]* |
| N19 | 44,610 | 113,877,480 | 0.45 | 0.38 | 0.34 | 0.07 | 0.10 | 0.09 | 1.30 [1.26-1.34]* | 1.24 [1.20-1.28]* |
| N20 | 54,980 | 113,371,610 | 0.50 | 0.48 | 0.44 | 0.02 | 0.06 | 0.04 | 1.15 [1.11-1.18]* | 1.10 [1.06-1.13]* |
| N21 | 13,520 | 114,098,760 | 0.11 | 0.12 | 0.11 | −0.01 | 0.00 | 0.00 | 1.02 [0.96-1.08] | 1.00 [0.94-1.07] |
| N23 | 12,720 | 113,886,210 | 0.12 | 0.11 | 0.10 | 0.01 | 0.02 | 0.01 | 1.17 [1.11-1.25]* | 1.08 [1.02-1.15] |
| N25 | 2,660 | 114,238,490 | 0.03 | 0.02 | 0.02 | 0.00 | 0.01 | 0.01 | 1.31 [1.15-1.48]* | 1.25 [1.10-1.43]* |
| N26 | 5,200 | 114,238,570 | 0.05 | 0.04 | 0.04 | 0.01 | 0.01 | 0.01 | 1.20 [1.09-1.32]* | 1.14 [1.03-1.26]* |
| N27 | 2,920 | 114,237,040 | 0.03 | 0.02 | 0.02 | 0.01 | 0.01 | 0.01 | 1.30 [1.15-1.47]* | 1.27 [1.12-1.44]* |
| N28 | 73,690 | 113,670,770 | 0.70 | 0.64 | 0.59 | 0.07 | 0.11 | 0.08 | 1.18 [1.15-1.22]* | 1.13 [1.11-1.16]* |
| N29 | 680 | 114,259,590 | 0.01 | 0.01 | 0.01 | 0.00 | 0.00 | 0.00 | 1.32 [1.02-1.71] | 1.30 [1.00-1.68] |
| N30 | 33,960 | 113,705,020 | 0.34 | 0.29 | 0.27 | 0.05 | 0.07 | 0.06 | 1.27 [1.22-1.32]* | 1.20 [1.16-1.25]* |
| N31 | 10,280 | 114,042,850 | 0.11 | 0.09 | 0.08 | 0.02 | 0.03 | 0.02 | 1.35 [1.26-1.43]* | 1.24 [1.16-1.32]* |
| N32 | 99,370 | 112,750,230 | 0.98 | 0.86 | 0.79 | 0.12 | 0.18 | 0.14 | 1.23 [1.20-1.26]* | 1.17 [1.14-1.19]* |
| N34 | 1,740 | 114,230,940 | 0.02 | 0.01 | 0.01 | 0.00 | 0.00 | 0.00 | 1.35 [1.16-1.58]* | 1.30 [1.11-1.53]* |
| N35 | 34,270 | 113,608,330 | 0.35 | 0.29 | 0.27 | 0.06 | 0.08 | 0.07 | 1.31 [1.27-1.36]* | 1.25 [1.20-1.30]* |
| N36 | 9,740 | 114,098,100 | 0.10 | 0.08 | 0.08 | 0.01 | 0.02 | 0.01 | 1.23 [1.15-1.32]* | 1.17 [1.10-1.26]* |
| N39 | 448,510 | 109,115,830 | 4.58 | 4.00 | 3.82 | 0.59 | 0.76 | 0.61 | 1.20 [1.19-1.21]* | 1.15 [1.14-1.17]* |
| N40 | 144,020 | 112,509,190 | 1.39 | 1.25 | 1.19 | 0.13 | 0.20 | 0.15 | 1.16 [1.14-1.19]* | 1.12 [1.10-1.14]* |
| N41 | 10,590 | 114,084,470 | 0.09 | 0.09 | 0.08 | 0.00 | 0.01 | 0.01 | 1.15 [1.08-1.23]* | 1.10 [1.02-1.17]* |
| N42 | 13,750 | 114,059,490 | 0.12 | 0.12 | 0.11 | 0.00 | 0.01 | 0.01 | 1.11 [1.05-1.18]* | 1.07 [1.00-1.13] |
| N43 | 21,610 | 113,825,610 | 0.21 | 0.19 | 0.19 | 0.02 | 0.02 | 0.02 | 1.13 [1.08-1.18]* | 1.10 [1.05-1.15]* |
| N44 | 7,350 | 114,153,960 | 0.08 | 0.06 | 0.07 | 0.03 | 0.01 | 0.01 | 1.18 [1.09-1.27]* | 1.16 [1.07-1.25]* |
| N45 | 15,500 | 114,008,990 | 0.15 | 0.13 | 0.13 | 0.02 | 0.03 | 0.02 | 1.21 [1.15-1.28]* | 1.15 [1.09-1.22]* |
| N47 | 58,760 | 113,030,600 | 0.71 | 0.47 | 0.49 | 0.24 | 0.22 | 0.20 | 1.46 [1.42-1.50]* | 1.40 [1.37-1.44]* |
| N48 | 30,710 | 113,710,890 | 0.35 | 0.25 | 0.25 | 0.10 | 0.10 | 0.09 | 1.40 [1.35-1.46]* | 1.34 [1.29-1.40]* |
| N49 | 3,590 | 114,215,000 | 0.04 | 0.03 | 0.03 | 0.01 | 0.01 | 0.01 | 1.35 [1.21-1.50]* | 1.29 [1.16-1.45]* |
| N50 | 33,020 | 113,634,280 | 0.34 | 0.28 | 0.28 | 0.06 | 0.06 | 0.05 | 1.22 [1.17-1.26]* | 1.16 [1.12-1.21]* |
| N51 | 450 | 114,264,070 | 0.00 | 0.00 | 0.00 | 0.00 | 0.00 | 0.00 | 1.48 [1.08-2.03] | 1.37 [0.99-1.90] |
| N60 | 10,790 | 113,963,480 | 0.10 | 0.09 | 0.09 | 0.01 | 0.01 | 0.01 | 1.17 [1.10-1.25]* | 1.13 [1.06-1.21]* |
| N61 | 7,260 | 114,106,170 | 0.08 | 0.06 | 0.06 | 0.02 | 0.02 | 0.02 | 1.34 [1.24-1.44]* | 1.28 [1.18-1.38]* |
| N62 | 6,270 | 114,065,990 | 0.06 | 0.05 | 0.05 | 0.01 | 0.01 | 0.01 | 1.28 [1.18-1.39]* | 1.21 [1.11-1.32]* |
| N63 | 11,490 | 113,955,840 | 0.11 | 0.10 | 0.09 | 0.01 | 0.01 | 0.01 | 1.16 [1.09-1.23]* | 1.11 [1.04-1.18]* |
| N64 | 11,000 | 114,038,650 | 0.11 | 0.09 | 0.09 | 0.01 | 0.02 | 0.02 | 1.25 [1.18-1.34]* | 1.19 [1.12-1.27]* |
| N70 | 7,890 | 114,082,010 | 0.07 | 0.07 | 0.07 | 0.00 | 0.00 | 0.00 | 1.07 [0.99-1.15] | 1.03 [0.95-1.11] |
| N71 | 3,790 | 114,181,890 | 0.04 | 0.03 | 0.03 | 0.00 | 0.01 | 0.00 | 1.17 [1.05-1.31]* | 1.08 [0.97-1.21] |
| N72 | 7,510 | 113,992,030 | 0.07 | 0.07 | 0.06 | 0.00 | 0.01 | 0.00 | 1.09 [1.01-1.18] | 1.04 [0.96-1.13] |
| N73 | 42,730 | 113,241,550 | 0.40 | 0.37 | 0.35 | 0.03 | 0.05 | 0.03 | 1.14 [1.10-1.17]* | 1.07 [1.04-1.11]* |
| N75 | 7,440 | 114,060,100 | 0.07 | 0.07 | 0.06 | 0.00 | 0.00 | 0.00 | 1.06 [0.97-1.15] | 1.04 [0.95-1.12] |
| N76 | 11,480 | 113,999,530 | 0.13 | 0.09 | 0.09 | 0.03 | 0.04 | 0.03 | 1.39 [1.31-1.47]* | 1.31 [1.23-1.39]* |
| N77 | 14,070 | 114,011,200 | 0.15 | 0.12 | 0.11 | 0.03 | 0.03 | 0.02 | 1.28 [1.21-1.36]* | 1.19 [1.12-1.26]* |
| N80 | 42,360 | 113,185,470 | 0.40 | 0.37 | 0.35 | 0.03 | 0.05 | 0.02 | 1.13 [1.09-1.17]* | 1.06 [1.02-1.10]* |
| N81 | 68,030 | 112,718,500 | 0.66 | 0.59 | 0.54 | 0.07 | 0.11 | 0.08 | 1.21 [1.18-1.24]* | 1.14 [1.12-1.17]* |
| N82 | 3,230 | 114,224,420 | 0.03 | 0.03 | 0.03 | 0.01 | 0.01 | 0.01 | 1.32 [1.18-1.49]* | 1.24 [1.11-1.40]* |
| N83 | 70,540 | 112,757,740 | 0.67 | 0.61 | 0.59 | 0.06 | 0.08 | 0.04 | 1.13 [1.10-1.16]* | 1.07 [1.04-1.10]* |
| N84 | 56,920 | 112,868,180 | 0.52 | 0.50 | 0.46 | 0.02 | 0.06 | 0.05 | 1.12 [1.09-1.16]* | 1.10 [1.07-1.13]* |
| N85 | 45,220 | 113,342,110 | 0.43 | 0.39 | 0.37 | 0.04 | 0.06 | 0.05 | 1.17 [1.14-1.21]* | 1.13 [1.09-1.16]* |
| N86 | 13,580 | 113,875,230 | 0.13 | 0.12 | 0.11 | 0.01 | 0.02 | 0.02 | 1.19 [1.12-1.26]* | 1.13 [1.07-1.20]* |
| N87 | 16,590 | 113,540,450 | 0.15 | 0.15 | 0.13 | 0.00 | 0.01 | 0.01 | 1.10 [1.05-1.17]* | 1.06 [1.01-1.12] |
| N88 | 13,420 | 113,961,110 | 0.13 | 0.12 | 0.11 | 0.01 | 0.02 | 0.02 | 1.21 [1.14-1.28]* | 1.16 [1.10-1.23]* |
| N89 | 23,000 | 113,670,890 | 0.24 | 0.19 | 0.18 | 0.04 | 0.05 | 0.04 | 1.29 [1.23-1.34]* | 1.21 [1.16-1.27]* |
| N90 | 21,160 | 113,798,310 | 0.23 | 0.18 | 0.16 | 0.05 | 0.06 | 0.05 | 1.38 [1.32-1.45]* | 1.31 [1.25-1.37]* |
| N91 | 2,610 | 114,203,250 | 0.03 | 0.02 | 0.02 | 0.01 | 0.01 | 0.00 | 1.29 [1.13-1.47]* | 1.22 [1.06-1.39]* |
| N92 | 92,940 | 111,576,480 | 0.93 | 0.81 | 0.77 | 0.12 | 0.16 | 0.11 | 1.21 [1.18-1.23]* | 1.13 [1.11-1.16]* |
| N93 | 43,170 | 113,028,540 | 0.42 | 0.37 | 0.35 | 0.05 | 0.07 | 0.05 | 1.20 [1.16-1.24]* | 1.14 [1.10-1.17]* |
| N94 | 35,030 | 113,126,440 | 0.36 | 0.30 | 0.28 | 0.06 | 0.07 | 0.05 | 1.26 [1.21-1.30]* | 1.16 [1.12-1.20]* |
| N95 | 48,570 | 113,029,600 | 0.48 | 0.42 | 0.38 | 0.06 | 0.10 | 0.08 | 1.25 [1.21-1.29]* | 1.20 [1.16-1.24]* |
| N96 | 680 | 114,255,380 | 0.01 | 0.01 | 0.01 | 0.00 | 0.00 | 0.00 | 1.08 [0.83-1.41] | 1.07 [0.81-1.41] |
| N97 | 7,800 | 113,712,940 | 0.06 | 0.07 | 0.06 | −0.01 | 0.00 | 0.00 | 0.93 [0.86-1.01] | 0.92 [0.85-1.00] |
| N98 | 950 | 114,241,510 | 0.01 | 0.01 | 0.01 | 0.00 | 0.00 | 0.00 | 0.80 [0.63-1.03] | 0.76 [0.59-0.98] |
| N99 | 12,220 | 114,038,560 | 0.12 | 0.10 | 0.10 | 0.01 | 0.02 | 0.02 | 1.22 [1.15-1.30]* | 1.17 [1.10-1.25]* |
| any age - O00-O9A | | | | | | | | | | |
| O00 | 13,390 | 113,900,020 | 0.12 | 0.12 | 0.11 | 0.00 | 0.00 | 0.00 | 1.04 [0.98-1.11] | 1.02 [0.95-1.08] |
| O01 | 800 | 114,247,910 | 0.01 | 0.01 | 0.01 | 0.00 | 0.00 | 0.00 | 1.17 [0.92-1.49] | 1.19 [0.92-1.53] |

| | | | | | | | | | | |
|---|---|---|---|---|---|---|---|---|---|---|
| O02 | 30,480 | 113,193,690 | 0.28 | 0.27 | 0.25 | 0.01 | 0.02 | 0.02 | 1.10 [1.05-1.14]* | 1.07 [1.02-1.11]* |
| O03 | 46,050 | 112,760,510 | 0.41 | 0.41 | 0.39 | 0.01 | 0.02 | 0.01 | 1.06 [1.02-1.09]* | 1.04 [1.00-1.07] |
| O04 | 59,590 | 112,296,900 | 0.54 | 0.53 | 0.53 | 0.01 | 0.01 | 0.00 | 1.02 [1.00-1.05] | 1.01 [0.98-1.04] |
| O08 | 4,820 | 114,155,580 | 0.04 | 0.04 | 0.04 | 0.00 | 0.00 | 0.00 | 1.05 [0.95-1.16] | 1.01 [0.91-1.12] |
| O10 | 4,880 | 114,160,380 | 0.05 | 0.04 | 0.04 | 0.00 | 0.01 | 0.00 | 1.13 [1.02-1.25] | 1.10 [0.99-1.21] |
| O11 | 700 | 114,254,240 | 0.01 | 0.01 | 0.01 | 0.00 | 0.00 | 0.00 | 1.17 [0.90-1.51] | 1.12 [0.86-1.45] |
| O12 | 10,150 | 113,910,560 | 0.10 | 0.09 | 0.08 | 0.01 | 0.01 | 0.01 | 1.13 [1.06-1.21]* | 1.08 [1.01-1.16] |
| O13 | 28,060 | 113,330,620 | 0.26 | 0.25 | 0.24 | 0.01 | 0.02 | 0.01 | 1.08 [1.04-1.13]* | 1.06 [1.01-1.11]* |
| O14 | 20,420 | 113,620,810 | 0.19 | 0.18 | 0.17 | 0.01 | 0.02 | 0.01 | 1.10 [1.05-1.16]* | 1.07 [1.02-1.13]* |
| O15 | 810 | 114,244,590 | 0.01 | 0.01 | 0.01 | 0.00 | 0.00 | 0.00 | 1.10 [0.86-1.41] | 1.08 [0.84-1.39] |
| O16 | 22,710 | 113,298,600 | 0.21 | 0.20 | 0.19 | 0.01 | 0.02 | 0.02 | 1.12 [1.07-1.17]* | 1.09 [1.04-1.14]* |
| O20 | 40,380 | 112,914,650 | 0.37 | 0.35 | 0.35 | 0.02 | 0.02 | 0.01 | 1.06 [1.03-1.10]* | 1.03 [0.99-1.07] |
| O21 | 28,640 | 113,456,490 | 0.27 | 0.25 | 0.25 | 0.03 | 0.03 | 0.01 | 1.11 [1.06-1.15]* | 1.04 [1.00-1.09] |
| O22 | 4,560 | 114,161,090 | 0.04 | 0.04 | 0.04 | 0.00 | 0.00 | 0.00 | 1.11 [1.00-1.23] | 1.09 [0.98-1.21] |
| O23 | 31,230 | 113,319,110 | 0.31 | 0.27 | 0.26 | 0.04 | 0.04 | 0.03 | 1.16 [1.12-1.20]* | 1.11 [1.07-1.15]* |
| O24 | 37,970 | 113,620,160 | 0.35 | 0.33 | 0.33 | 0.02 | 0.02 | 0.02 | 1.07 [1.03-1.11]* | 1.05 [1.01-1.09]* |
| O26 | 116,280 | 110,304,620 | 1.19 | 1.02 | 1.03 | 0.17 | 0.16 | 0.11 | 1.16 [1.13-1.18]* | 1.10 [1.08-1.13]* |
| O28 | 11,630 | 114,067,930 | 0.11 | 0.10 | 0.10 | 0.01 | 0.01 | 0.00 | 1.06 [0.99-1.13] | 1.01 [0.95-1.08] |
| O30 | 14,430 | 113,890,610 | 0.12 | 0.13 | 0.12 | −0.01 | 0.00 | 0.00 | 1.00 [0.94-1.06] | 0.98 [0.92-1.04] |
| O31 | 810 | 114,249,620 | 0.01 | 0.01 | 0.01 | 0.00 | 0.00 | 0.00 | 1.05 [0.82-1.34] | 1.01 [0.78-1.30] |
| O32 | 57,040 | 112,728,690 | 0.51 | 0.51 | 0.49 | 0.00 | 0.02 | 0.01 | 1.03 [1.00-1.06] | 1.02 [0.99-1.05] |
| O33 | 1,350 | 114,201,270 | 0.01 | 0.01 | 0.01 | 0.00 | 0.00 | 0.00 | 1.15 [0.95-1.39] | 1.13 [0.93-1.37] |
| O34 | 93,950 | 112,009,270 | 0.87 | 0.83 | 0.80 | 0.04 | 0.07 | 0.06 | 1.09 [1.06-1.11]* | 1.07 [1.05-1.10]* |
| O35 | 14,160 | 113,953,850 | 0.12 | 0.12 | 0.12 | 0.00 | 0.00 | 0.00 | 1.02 [0.96-1.08] | 1.00 [0.94-1.06] |
| O36 | 154,890 | 110,226,410 | 1.57 | 1.37 | 1.41 | 0.20 | 0.16 | 0.12 | 1.12 [1.10-1.14]* | 1.08 [1.06-1.10]* |
| O40 | 12,970 | 114,033,840 | 0.12 | 0.11 | 0.11 | 0.01 | 0.01 | 0.01 | 1.09 [1.03-1.16]* | 1.05 [0.99-1.12] |
| O41 | 17,530 | 113,721,110 | 0.16 | 0.15 | 0.15 | 0.01 | 0.01 | 0.01 | 1.10 [1.04-1.16]* | 1.08 [1.02-1.14]* |
| O42 | 93,460 | 111,714,650 | 0.85 | 0.83 | 0.83 | 0.02 | 0.03 | 0.02 | 1.03 [1.01-1.06]* | 1.03 [1.01-1.05] |
| O43 | 12,900 | 113,868,720 | 0.11 | 0.11 | 0.11 | 0.00 | 0.00 | 0.00 | 1.03 [0.97-1.10] | 1.02 [0.96-1.09] |
| O44 | 8,740 | 114,036,220 | 0.08 | 0.08 | 0.07 | 0.00 | 0.00 | 0.00 | 1.03 [0.95-1.11] | 0.99 [0.91-1.07] |
| O45 | 3,840 | 114,160,950 | 0.03 | 0.03 | 0.03 | 0.00 | 0.00 | 0.00 | 1.02 [0.91-1.14] | 1.00 [0.89-1.12] |
| O46 | 43,420 | 112,887,170 | 0.41 | 0.38 | 0.37 | 0.03 | 0.04 | 0.03 | 1.12 [1.09-1.16]* | 1.09 [1.05-1.13]* |
| O47 | 30,950 | 112,334,650 | 0.29 | 0.27 | 0.25 | 0.02 | 0.04 | 0.03 | 1.16 [1.12-1.21]* | 1.12 [1.07-1.16]* |
| O48 | 57,620 | 112,150,800 | 0.50 | 0.52 | 0.50 | −0.01 | 0.01 | 0.01 | 1.02 [0.99-1.05] | 1.02 [0.99-1.05] |
| O60 | 48,180 | 112,889,580 | 0.44 | 0.42 | 0.41 | 0.02 | 0.03 | 0.02 | 1.08 [1.04-1.11]* | 1.05 [1.01-1.08]* |
| O61 | 9,070 | 113,980,620 | 0.08 | 0.08 | 0.08 | 0.00 | 0.01 | 0.00 | 1.08 [1.01-1.17] | 1.05 [0.97-1.13] |
| O62 | 36,590 | 113,332,360 | 0.33 | 0.32 | 0.31 | 0.01 | 0.02 | 0.02 | 1.05 [1.01-1.09]* | 1.05 [1.01-1.09] |
| O63 | 83,060 | 111,318,330 | 0.74 | 0.75 | 0.73 | 0.00 | 0.01 | 0.01 | 1.02 [0.99-1.04] | 1.01 [0.98-1.03] |
| O64 | 16,270 | 113,596,780 | 0.14 | 0.14 | 0.14 | −0.01 | 0.00 | 0.00 | 1.02 [0.97-1.08] | 1.02 [0.96-1.08] |
| O65 | 1,190 | 114,198,580 | 0.01 | 0.01 | 0.01 | 0.00 | 0.00 | 0.00 | 0.98 [0.80-1.21] | 0.99 [0.80-1.22] |
| O66 | 15,620 | 113,743,600 | 0.14 | 0.14 | 0.13 | 0.00 | 0.00 | 0.00 | 1.01 [0.96-1.07] | 1.00 [0.94-1.06] |
| O67 | 4,160 | 114,121,330 | 0.04 | 0.04 | 0.03 | 0.00 | 0.00 | 0.00 | 1.02 [0.91-1.13] | 1.01 [0.90-1.13] |
| O68 | 173,530 | 108,628,640 | 1.69 | 1.57 | 1.62 | 0.12 | 0.07 | 0.06 | 1.04 [1.03-1.06]* | 1.03 [1.02-1.05]* |
| O69 | 16,010 | 113,458,940 | 0.14 | 0.14 | 0.13 | 0.00 | 0.01 | 0.01 | 1.09 [1.03-1.15]* | 1.08 [1.02-1.14]* |
| O70 | 232,730 | 106,299,640 | 2.34 | 2.15 | 2.26 | 0.19 | 0.08 | 0.08 | 1.03 [1.02-1.05]* | 1.04 [1.02-1.05]* |
| O71 | 14,160 | 113,835,660 | 0.12 | 0.12 | 0.12 | 0.00 | 0.00 | 0.00 | 1.01 [0.96-1.08] | 1.01 [0.95-1.07] |
| O72 | 117,020 | 111,597,950 | 1.09 | 1.04 | 1.04 | 0.06 | 0.06 | 0.05 | 1.05 [1.03-1.08]* | 1.05 [1.02-1.07]* |
| O73 | 8,220 | 113,964,730 | 0.07 | 0.07 | 0.07 | 0.00 | 0.01 | 0.01 | 1.10 [1.02-1.19] | 1.09 [1.01-1.18] |
| O74 | 2,230 | 114,210,570 | 0.02 | 0.02 | 0.02 | 0.00 | 0.00 | 0.00 | 1.16 [1.00-1.34] | 1.12 [0.96-1.30] |
| O75 | 58,240 | 112,705,400 | 0.53 | 0.51 | 0.51 | 0.01 | 0.02 | 0.02 | 1.04 [1.01-1.07]* | 1.04 [1.01-1.07] |
| O80 | 62,900 | 111,215,660 | 0.58 | 0.56 | 0.54 | 0.02 | 0.04 | 0.05 | 1.08 [1.05-1.11]* | 1.09 [1.06-1.12]* |
| O81 | 990 | 114,158,510 | 0.01 | 0.01 | 0.01 | 0.00 | 0.00 | 0.00 | 1.07 [0.86-1.34] | 1.06 [0.84-1.33] |
| O82 | 9,060 | 113,774,780 | 0.08 | 0.08 | 0.07 | 0.00 | 0.01 | 0.01 | 1.10 [1.03-1.19]* | 1.08 [1.00-1.16] |
| O85 | 4,840 | 114,198,180 | 0.05 | 0.04 | 0.04 | 0.00 | 0.00 | 0.00 | 1.07 [0.97-1.19] | 1.03 [0.93-1.15] |
| O86 | 17,780 | 113,820,400 | 0.17 | 0.15 | 0.15 | 0.02 | 0.02 | 0.02 | 1.15 [1.09-1.21]* | 1.12 [1.06-1.18]* |
| O87 | 2,590 | 114,172,450 | 0.02 | 0.02 | 0.02 | 0.00 | 0.00 | 0.00 | 1.19 [1.04-1.36] | 1.17 [1.02-1.34] |
| O88 | 950 | 114,251,140 | 0.01 | 0.01 | 0.01 | 0.00 | 0.00 | 0.00 | 1.15 [0.92-1.44] | 1.07 [0.85-1.35] |
| O89 | 760 | 114,246,810 | 0.01 | 0.01 | 0.01 | 0.00 | 0.00 | 0.00 | 1.32 [1.03-1.68] | 1.25 [0.97-1.62] |
| O90 | 20,880 | 113,847,250 | 0.20 | 0.18 | 0.17 | 0.03 | 0.03 | 0.02 | 1.18 [1.12-1.24]* | 1.14 [1.08-1.19]* |
| O91 | 2,130 | 114,226,930 | 0.02 | 0.02 | 0.02 | 0.00 | 0.00 | 0.00 | 1.02 [0.87-1.18] | 0.99 [0.84-1.16] |
| O92 | 3,220 | 114,176,540 | 0.03 | 0.03 | 0.03 | 0.00 | 0.00 | 0.00 | 1.11 [0.98-1.25] | 1.07 [0.94-1.21] |
| O94 | 670 | 114,264,770 | 0.01 | 0.01 | 0.01 | 0.00 | 0.00 | 0.00 | 1.26 [0.97-1.63] | 1.14 [0.86-1.50] |
| O98 | 7,740 | 114,083,070 | 0.08 | 0.07 | 0.07 | 0.01 | 0.01 | 0.01 | 1.15 [1.07-1.25]* | 1.11 [1.02-1.20] |
| O99 | 127,350 | 110,852,290 | 1.30 | 1.11 | 1.12 | 0.19 | 0.18 | 0.13 | 1.16 [1.14-1.19]* | 1.11 [1.09-1.13]* |
| any age - P00-P96 | | | | | | | | | | |
| P83 | 1,660 | 113,235,600 | 0.02 | 0.01 | 0.02 | 0.00 | 0.00 | 0.00 | 1.02 [0.87-1.20] | 1.02 [0.86-1.20] |
| any age - Q00-Q99 | | | | | | | | | | |
| Q02 | 2,060 | 114,217,950 | 0.02 | 0.02 | 0.02 | 0.00 | 0.00 | 0.00 | 1.10 [0.96-1.27] | 1.05 [0.91-1.21] |
| Q03 | 850 | 114,229,970 | 0.01 | 0.01 | 0.01 | 0.00 | 0.00 | 0.00 | 1.20 [0.96-1.50] | 1.15 [0.91-1.45] |
| Q04 | 2,220 | 114,196,740 | 0.02 | 0.02 | 0.02 | 0.00 | 0.00 | 0.00 | 1.08 [0.94-1.25] | 1.01 [0.87-1.17] |
| Q05 | 2,140 | 114,210,590 | 0.02 | 0.02 | 0.02 | 0.00 | 0.00 | 0.00 | 1.29 [1.12-1.49]* | 1.18 [1.02-1.37] |
| Q06 | 650 | 114,258,070 | 0.01 | 0.01 | 0.01 | 0.00 | 0.00 | 0.00 | 0.91 [0.70-1.20] | 0.88 [0.66-1.16] |
| Q07 | 2,530 | 114,236,140 | 0.03 | 0.02 | 0.02 | 0.01 | 0.00 | 0.00 | 1.20 [1.05-1.36]* | 1.09 [0.95-1.24] |
| Q10 | 2,600 | 114,203,990 | 0.03 | 0.02 | 0.02 | 0.01 | 0.00 | 0.00 | 1.19 [1.05-1.35]* | 1.18 [1.04-1.34] |
| Q12 | 1,410 | 114,232,500 | 0.01 | 0.01 | 0.01 | 0.00 | 0.00 | 0.00 | 1.22 [1.02-1.46] | 1.17 [0.98-1.41] |
| Q13 | 510 | 114,254,330 | 0.01 | 0.00 | 0.00 | 0.00 | 0.00 | 0.00 | 1.33 [1.00-1.77] | 1.21 [0.90-1.62] |
| Q14 | 680 | 114,258,230 | 0.01 | 0.01 | 0.01 | 0.00 | 0.00 | 0.00 | 1.11 [0.86-1.44] | 1.07 [0.82-1.39] |
| Q16 | 430 | 114,260,970 | 0.00 | 0.00 | 0.00 | 0.00 | 0.00 | 0.00 | 1.28 [0.93-1.75] | 1.24 [0.90-1.70] |
| Q17 | 6,290 | 114,016,810 | 0.06 | 0.05 | 0.06 | 0.01 | 0.00 | 0.00 | 1.08 [0.99-1.17] | 1.06 [0.97-1.15] |
| Q18 | 3,470 | 114,172,350 | 0.04 | 0.03 | 0.03 | 0.01 | 0.01 | 0.00 | 1.16 [1.04-1.30]* | 1.13 [1.01-1.27] |
| Q20 | 660 | 114,225,600 | 0.01 | 0.01 | 0.01 | 0.00 | 0.00 | 0.00 | 1.01 [0.77-1.32] | 0.98 [0.75-1.30] |
| Q21 | 10,710 | 113,839,690 | 0.11 | 0.09 | 0.09 | 0.01 | 0.01 | 0.01 | 1.16 [1.09-1.24]* | 1.10 [1.03-1.18]* |
| Q22 | 1,360 | 114,205,230 | 0.01 | 0.01 | 0.01 | 0.00 | 0.00 | 0.00 | 0.96 [0.79-1.15] | 0.96 [0.79-1.17] |
| Q23 | 4,780 | 114,181,960 | 0.05 | 0.04 | 0.04 | 0.00 | 0.01 | 0.00 | 1.15 [1.04-1.26]* | 1.11 [1.01-1.23] |
| Q24 | 3,670 | 114,156,270 | 0.04 | 0.03 | 0.03 | 0.01 | 0.01 | 0.00 | 1.17 [1.05-1.31]* | 1.10 [0.99-1.23] |
| Q25 | 3,950 | 113,944,950 | 0.04 | 0.03 | 0.04 | 0.00 | 0.00 | 0.00 | 1.06 [0.95-1.18] | 1.03 [0.92-1.14] |
| Q26 | 640 | 114,251,980 | 0.01 | 0.01 | 0.01 | 0.00 | 0.00 | 0.00 | 1.11 [0.85-1.44] | 1.04 [0.80-1.37] |
| Q27 | 3,180 | 114,209,260 | 0.03 | 0.03 | 0.03 | 0.00 | 0.00 | 0.00 | 1.08 [0.96-1.22] | 1.04 [0.92-1.18] |
| Q28 | 1,700 | 114,237,990 | 0.02 | 0.01 | 0.01 | 0.00 | 0.00 | 0.00 | 1.32 [1.12-1.54]* | 1.27 [1.08-1.49]* |
| Q31 | 1,360 | 114,183,290 | 0.01 | 0.01 | 0.01 | 0.00 | 0.00 | 0.00 | 1.03 [0.86-1.24] | 0.94 [0.78-1.14] |
| Q32 | 590 | 114,251,740 | 0.01 | 0.01 | 0.01 | 0.00 | 0.00 | 0.00 | 0.96 [0.73-1.27] | 0.85 [0.64-1.13] |
| Q33 | 540 | 114,235,040 | 0.01 | 0.00 | 0.00 | 0.00 | 0.00 | 0.00 | 1.05 [0.79-1.41] | 0.93 [0.69-1.26] |
| Q35 | 1,500 | 114,200,270 | 0.02 | 0.01 | 0.01 | 0.00 | 0.00 | 0.00 | 1.11 [0.94-1.32] | 1.06 [0.89-1.26] |
| Q38 | 10,400 | 113,612,180 | 0.11 | 0.09 | 0.10 | 0.02 | 0.01 | 0.01 | 1.14 [1.07-1.21]* | 1.11 [1.04-1.18]* |
| Q39 | 1,690 | 114,236,620 | 0.02 | 0.01 | 0.01 | 0.00 | 0.00 | 0.00 | 1.27 [1.08-1.49]* | 1.17 [0.99-1.39] |
| Q40 | 1,070 | 114,173,750 | 0.01 | 0.01 | 0.01 | 0.00 | 0.00 | 0.00 | 1.24 [1.01-1.52] | 1.11 [0.90-1.38] |
| Q43 | 4,350 | 114,155,360 | 0.04 | 0.04 | 0.04 | 0.01 | 0.01 | 0.00 | 1.17 [1.06-1.29]* | 1.10 [0.99-1.22] |
| Q44 | 1,380 | 114,244,540 | 0.01 | 0.01 | 0.01 | 0.00 | 0.00 | 0.00 | 1.15 [0.95-1.38] | 1.09 [0.90-1.31] |
| Q45 | 570 | 114,261,670 | 0.01 | 0.00 | 0.00 | 0.00 | 0.00 | 0.00 | 1.34 [1.02-1.76] | 1.24 [0.93-1.64] |
| Q50 | 3,050 | 114,197,920 | 0.03 | 0.03 | 0.02 | 0.01 | 0.01 | 0.01 | 1.25 [1.11-1.41]* | 1.19 [1.06-1.35]* |
| Q51 | 3,040 | 114,210,700 | 0.03 | 0.03 | 0.03 | 0.00 | 0.00 | 0.00 | 1.06 [0.94-1.21] | 1.01 [0.89-1.15] |
| Q52 | 1,600 | 114,220,550 | 0.02 | 0.01 | 0.01 | 0.00 | 0.00 | 0.00 | 1.21 [1.03-1.42] | 1.12 [0.94-1.32] |
| Q53 | 13,720 | 113,693,310 | 0.14 | 0.12 | 0.14 | 0.02 | 0.00 | 0.00 | 1.00 [0.94-1.06] | 0.98 [0.92-1.04] |
| Q54 | 4,020 | 114,021,240 | 0.04 | 0.03 | 0.04 | 0.01 | 0.00 | 0.00 | 1.11 [1.00-1.23] | 1.11 [0.99-1.23] |
| Q55 | 8,670 | 114,071,870 | 0.10 | 0.07 | 0.09 | 0.03 | 0.01 | 0.01 | 1.13 [1.06-1.22]* | 1.11 [1.03-1.19]* |
| Q60 | 2,940 | 114,218,010 | 0.03 | 0.03 | 0.02 | 0.00 | 0.00 | 0.00 | 1.17 [1.03-1.33] | 1.12 [0.99-1.28] |
| Q61 | 7,280 | 114,141,410 | 0.07 | 0.06 | 0.06 | 0.01 | 0.01 | 0.01 | 1.16 [1.07-1.26]* | 1.11 [1.03-1.21] |

| | | | | | | | | | | |
|---|---|---|---|---|---|---|---|---|---|---|
| Q62 | 1,720 | 114,068,190 | 0.02 | 0.01 | 0.02 | 0.00 | 0.00 | 0.00 | 1.00 [0.85-1.18] | 0.98 [0.83-1.16] |
| Q63 | 5,700 | 114,135,950 | 0.06 | 0.05 | 0.05 | 0.01 | 0.01 | 0.00 | 1.16 [1.06-1.27]* | 1.09 [0.99-1.19] |
| Q64 | 1,260 | 114,213,790 | 0.01 | 0.01 | 0.01 | 0.00 | 0.00 | 0.00 | 1.15 [0.95-1.38] | 1.09 [0.90-1.32] |
| Q65 | 4,710 | 114,080,300 | 0.04 | 0.04 | 0.04 | 0.00 | 0.00 | 0.00 | 1.04 [0.94-1.15] | 0.99 [0.89-1.09] |
| Q66 | 5,520 | 113,410,300 | 0.06 | 0.05 | 0.05 | 0.02 | 0.01 | 0.01 | 1.26 [1.15-1.37]* | 1.19 [1.09-1.30]* |
| Q67 | 3,570 | 114,163,700 | 0.04 | 0.03 | 0.03 | 0.01 | 0.01 | 0.00 | 1.20 [1.07-1.33]* | 1.14 [1.02-1.28] |
| Q68 | 1,240 | 114,224,690 | 0.01 | 0.01 | 0.01 | 0.00 | 0.00 | 0.00 | 1.21 [1.00-1.46] | 1.16 [0.96-1.40] |
| Q69 | 630 | 114,162,420 | 0.01 | 0.01 | 0.01 | 0.00 | 0.00 | 0.00 | 0.90 [0.69-1.18] | 0.88 [0.67-1.15] |
| Q70 | 600 | 114,213,280 | 0.01 | 0.01 | 0.01 | 0.00 | 0.00 | 0.00 | 0.87 [0.65-1.16] | 0.87 [0.65-1.16] |
| Q72 | 540 | 114,253,520 | 0.01 | 0.00 | 0.00 | 0.00 | 0.00 | 0.00 | 1.18 [0.88-1.57] | 1.15 [0.86-1.54] |
| Q74 | 3,740 | 114,182,900 | 0.04 | 0.03 | 0.03 | 0.01 | 0.01 | 0.01 | 1.21 [1.09-1.35]* | 1.17 [1.05-1.31]* |
| Q75 | 1,990 | 114,181,440 | 0.02 | 0.02 | 0.02 | 0.00 | 0.00 | 0.00 | 1.03 [0.89-1.19] | 0.99 [0.85-1.15] |
| Q76 | 2,910 | 114,204,820 | 0.03 | 0.02 | 0.03 | 0.00 | 0.00 | 0.00 | 1.15 [1.02-1.31] | 1.06 [0.94-1.21] |
| Q78 | 1,860 | 114,236,790 | 0.02 | 0.02 | 0.02 | 0.00 | 0.00 | 0.00 | 1.22 [1.05-1.42] | 1.15 [0.99-1.35] |
| Q79 | 3,700 | 114,186,580 | 0.04 | 0.03 | 0.03 | 0.01 | 0.01 | 0.00 | 1.35 [1.22-1.51]* | 1.13 [1.01-1.26] |
| Q80 | 280 | 114,263,110 | 0.01 | 0.00 | 0.00 | 0.00 | 0.00 | 0.00 | 2.79 [1.97-3.95]* | 2.58 [1.81-3.68]* |
| Q82 | 3,160 | 113,550,950 | 0.04 | 0.03 | 0.03 | 0.01 | 0.01 | 0.01 | 1.36 [1.21-1.52]* | 1.28 [1.14-1.44]* |
| Q83 | 1,700 | 114,203,600 | 0.02 | 0.01 | 0.01 | 0.00 | 0.00 | 0.00 | 1.13 [0.96-1.33] | 1.06 [0.90-1.26] |
| Q85 | 2,610 | 114,216,440 | 0.02 | 0.02 | 0.02 | 0.00 | 0.00 | 0.00 | 1.02 [0.89-1.16] | 0.97 [0.85-1.12] |
| Q87 | 3,680 | 114,172,710 | 0.04 | 0.03 | 0.03 | 0.01 | 0.01 | 0.01 | 1.23 [1.10-1.37]* | 1.15 [1.03-1.29] |
| Q89 | 3,360 | 114,191,290 | 0.03 | 0.03 | 0.03 | 0.00 | 0.00 | 0.00 | 1.09 [0.97-1.22] | 1.06 [0.94-1.19] |
| Q90 | 2,300 | 114,135,210 | 0.04 | 0.02 | 0.01 | 0.02 | 0.02 | 0.02 | 2.77 [2.46-3.12]* | 2.68 [2.37-3.02]* |
| Q92 | 650 | 114,261,790 | 0.01 | 0.01 | 0.01 | 0.00 | 0.00 | 0.00 | 1.12 [0.87-1.45] | 1.07 [0.83-1.40] |
| Q93 | 1,770 | 114,242,640 | 0.02 | 0.01 | 0.02 | 0.01 | 0.00 | 0.00 | 1.27 [1.09-1.47]* | 1.17 [1.00-1.36] |
| Q96 | 500 | 114,254,210 | 0.01 | 0.00 | 0.00 | 0.00 | 0.00 | 0.00 | 1.62 [1.23-2.13]* | 1.65 [1.24-2.19]* |
| Q98 | 500 | 114,258,130 | 0.01 | 0.00 | 0.00 | 0.00 | 0.00 | 0.00 | 1.77 [1.35-2.32]* | 1.60 [1.21-2.11]* |
| Q99 | 2,120 | 114,235,370 | 0.02 | 0.02 | 0.02 | 0.01 | 0.00 | 0.00 | 1.16 [1.00-1.33] | 1.08 [0.94-1.25] |
| any age - R00-R99 | | | | | | | | | | |
| R00 | 206,340 | 112,193,240 | 2.07 | 1.79 | 1.71 | 0.28 | 0.36 | 0.26 | 1.21 [1.19-1.23]* | 1.14 [1.13-1.16]* |
| R01 | 33,350 | 113,688,640 | 0.34 | 0.28 | 0.28 | 0.06 | 0.06 | 0.04 | 1.21 [1.17-1.26]* | 1.15 [1.11-1.19]* |
| R02 | 30,180 | 114,087,600 | 0.29 | 0.26 | 0.24 | 0.04 | 0.05 | 0.05 | 1.22 [1.17-1.27]* | 1.19 [1.14-1.24]* |
| R03 | 71,720 | 113,546,050 | 0.69 | 0.62 | 0.58 | 0.08 | 0.11 | 0.09 | 1.18 [1.15-1.22]* | 1.14 [1.11-1.17]* |
| R04 | 80,700 | 113,133,360 | 0.85 | 0.68 | 0.64 | 0.17 | 0.21 | 0.17 | 1.33 [1.30-1.36]* | 1.25 [1.22-1.28]* |
| R05 | 85,010 | 113,011,960 | 0.95 | 0.70 | 0.69 | 0.25 | 0.26 | 0.20 | 1.38 [1.35-1.41]* | 1.27 [1.24-1.30]* |
| R06 | 282,150 | 109,316,170 | 3.56 | 2.35 | 2.35 | 1.22 | 1.22 | 1.07 | 1.52 [1.50-1.54]* | 1.43 [1.41-1.44]* |
| R07 | 341,950 | 107,976,700 | 3.67 | 3.05 | 2.84 | 0.62 | 0.83 | 0.60 | 1.29 [1.28-1.31]* | 1.20 [1.18-1.21]* |
| R09 | 30,610 | 113,956,370 | 0.32 | 0.26 | 0.24 | 0.06 | 0.07 | 0.06 | 1.31 [1.26-1.36]* | 1.22 [1.17-1.26]* |
| R10 | 498,030 | 103,240,900 | 5.74 | 4.60 | 4.52 | 1.13 | 1.21 | 0.88 | 1.27 [1.26-1.28]* | 1.18 [1.17-1.19]* |
| R11 | 292,230 | 109,627,550 | 3.11 | 2.56 | 2.53 | 0.56 | 0.58 | 0.42 | 1.23 [1.22-1.25]* | 1.16 [1.14-1.17]* |
| R12 | 18,070 | 113,951,110 | 0.19 | 0.15 | 0.14 | 0.04 | 0.05 | 0.03 | 1.35 [1.29-1.42]* | 1.22 [1.16-1.28]* |
| R13 | 133,640 | 113,119,120 | 1.39 | 1.13 | 1.06 | 0.25 | 0.33 | 0.25 | 1.31 [1.29-1.34]* | 1.22 [1.19-1.24]* |
| R14 | 31,190 | 113,859,920 | 0.32 | 0.26 | 0.25 | 0.06 | 0.08 | 0.05 | 1.31 [1.27-1.36]* | 1.20 [1.16-1.25]* |
| R15 | 56,780 | 113,978,110 | 0.55 | 0.48 | 0.46 | 0.07 | 0.09 | 0.08 | 1.20 [1.16-1.24]* | 1.16 [1.13-1.20]* |
| R16 | 21,970 | 114,065,630 | 0.24 | 0.18 | 0.17 | 0.05 | 0.07 | 0.05 | 1.38 [1.32-1.45]* | 1.30 [1.24-1.36]* |
| R17 | 18,030 | 114,036,950 | 0.17 | 0.15 | 0.14 | 0.02 | 0.03 | 0.03 | 1.22 [1.16-1.28]* | 1.19 [1.13-1.25]* |
| R18 | 45,510 | 114,016,960 | 0.43 | 0.39 | 0.36 | 0.04 | 0.07 | 0.06 | 1.20 [1.17-1.24]* | 1.17 [1.14-1.21]* |
| R19 | 172,930 | 112,074,130 | 1.83 | 1.47 | 1.37 | 0.36 | 0.46 | 0.35 | 1.34 [1.31-1.36]* | 1.23 [1.21-1.25]* |
| R20 | 47,880 | 113,608,950 | 0.49 | 0.41 | 0.38 | 0.08 | 0.10 | 0.07 | 1.27 [1.23-1.30]* | 1.16 [1.12-1.20]* |
| R21 | 76,360 | 112,572,500 | 0.92 | 0.62 | 0.63 | 0.30 | 0.29 | 0.26 | 1.46 [1.42-1.49]* | 1.39 [1.35-1.42]* |
| R22 | 37,270 | 113,599,990 | 0.41 | 0.31 | 0.30 | 0.10 | 0.11 | 0.09 | 1.38 [1.34-1.43]* | 1.29 [1.25-1.34]* |
| R23 | 30,470 | 113,695,690 | 0.33 | 0.25 | 0.26 | 0.07 | 0.06 | 0.05 | 1.25 [1.20-1.29]* | 1.19 [1.14-1.24]* |
| R25 | 28,180 | 113,750,170 | 0.29 | 0.24 | 0.23 | 0.05 | 0.05 | 0.04 | 1.24 [1.19-1.29]* | 1.15 [1.10-1.20]* |
| R26 | 208,060 | 113,406,160 | 2.07 | 1.78 | 1.71 | 0.29 | 0.36 | 0.29 | 1.21 [1.19-1.23]* | 1.16 [1.15-1.18]* |
| R27 | 8,710 | 114,174,010 | 0.09 | 0.07 | 0.07 | 0.01 | 0.01 | 0.01 | 1.18 [1.10-1.27]* | 1.12 [1.04-1.21]* |
| R29 | 273,190 | 112,756,550 | 2.69 | 2.36 | 2.26 | 0.33 | 0.43 | 0.34 | 1.19 [1.18-1.21]* | 1.15 [1.13-1.16]* |
| R30 | 22,530 | 113,905,650 | 0.24 | 0.19 | 0.18 | 0.06 | 0.06 | 0.05 | 1.35 [1.29-1.41]* | 1.24 [1.19-1.30]* |
| R31 | 149,230 | 111,674,990 | 1.45 | 1.31 | 1.21 | 0.14 | 0.24 | 0.19 | 1.19 [1.17-1.22]* | 1.15 [1.13-1.17]* |
| R32 | 109,890 | 113,571,950 | 1.07 | 0.94 | 0.90 | 0.13 | 0.17 | 0.14 | 1.19 [1.17-1.22]* | 1.15 [1.13-1.18]* |
| R33 | 206,830 | 112,713,930 | 1.99 | 1.80 | 1.70 | 0.19 | 0.28 | 0.23 | 1.17 [1.15-1.19]* | 1.13 [1.11-1.15]* |
| R34 | 5,610 | 114,236,720 | 0.06 | 0.05 | 0.05 | 0.01 | 0.01 | 0.01 | 1.21 [1.11-1.33]* | 1.15 [1.05-1.27]* |
| R35 | 34,910 | 113,516,350 | 0.37 | 0.29 | 0.27 | 0.07 | 0.09 | 0.08 | 1.35 [1.30-1.40]* | 1.26 [1.21-1.30]* |
| R36 | 760 | 114,259,380 | 0.01 | 0.01 | 0.01 | 0.00 | 0.00 | 0.00 | 1.28 [1.00-1.63] | 1.25 [0.97-1.60] |
| R39 | 40,110 | 113,494,870 | 0.42 | 0.34 | 0.31 | 0.08 | 0.11 | 0.08 | 1.35 [1.30-1.39]* | 1.25 [1.21-1.29]* |
| R40 | 33,190 | 114,004,880 | 0.33 | 0.28 | 0.28 | 0.05 | 0.06 | 0.04 | 1.21 [1.17-1.26]* | 1.15 [1.11-1.20]* |
| R41 | 199,520 | 113,248,680 | 1.93 | 1.72 | 1.64 | 0.21 | 0.29 | 0.24 | 1.17 [1.16-1.19]* | 1.14 [1.12-1.16]* |
| R42 | 111,950 | 113,041,220 | 1.12 | 0.96 | 0.91 | 0.17 | 0.21 | 0.15 | 1.24 [1.21-1.26]* | 1.15 [1.13-1.18]* |
| R43 | 1,500 | 114,257,410 | 0.02 | 0.01 | 0.01 | 0.00 | 0.00 | 0.00 | 1.35 [1.14-1.61]* | 1.23 [1.03-1.47] |
| R44 | 29,420 | 114,078,790 | 0.29 | 0.25 | 0.24 | 0.04 | 0.05 | 0.04 | 1.21 [1.16-1.26]* | 1.15 [1.11-1.20]* |
| R45 | 67,540 | 113,855,600 | 0.69 | 0.57 | 0.58 | 0.12 | 0.11 | 0.08 | 1.19 [1.16-1.22]* | 1.13 [1.10-1.16]* |
| R46 | 19,220 | 114,159,970 | 0.18 | 0.17 | 0.16 | 0.02 | 0.02 | 0.02 | 1.13 [1.08-1.19]* | 1.11 [1.06-1.17]* |
| R47 | 55,830 | 113,859,260 | 0.53 | 0.48 | 0.46 | 0.05 | 0.08 | 0.06 | 1.17 [1.13-1.20]* | 1.13 [1.09-1.16]* |
| R48 | 2,280 | 114,252,380 | 0.03 | 0.02 | 0.02 | 0.01 | 0.01 | 0.00 | 1.29 [1.12-1.48]* | 1.15 [1.00-1.33] |
| R49 | 11,260 | 114,060,520 | 0.12 | 0.09 | 0.09 | 0.02 | 0.03 | 0.02 | 1.35 [1.27-1.44]* | 1.23 [1.16-1.32]* |
| R50 | 125,160 | 112,045,030 | 1.33 | 1.07 | 1.09 | 0.27 | 0.24 | 0.19 | 1.22 [1.20-1.25]* | 1.17 [1.15-1.19]* |
| R51 | 164,460 | 111,561,010 | 1.77 | 1.40 | 1.38 | 0.37 | 0.39 | 0.27 | 1.28 [1.26-1.30]* | 1.18 [1.16-1.20]* |
| R52 | 25,490 | 113,999,400 | 0.28 | 0.21 | 0.20 | 0.06 | 0.08 | 0.06 | 1.40 [1.34-1.46]* | 1.26 [1.21-1.31]* |
| R53 | 106,190 | 113,406,850 | 1.08 | 0.90 | 0.87 | 0.17 | 0.21 | 0.16 | 1.24 [1.21-1.27]* | 1.18 [1.15-1.20]* |
| R54 | 128,060 | 113,818,500 | 1.22 | 1.10 | 1.05 | 0.12 | 0.18 | 0.15 | 1.17 [1.14-1.19]* | 1.14 [1.12-1.17]* |
| R55 | 204,250 | 111,935,580 | 2.02 | 1.78 | 1.69 | 0.24 | 0.33 | 0.26 | 1.20 [1.18-1.21]* | 1.15 [1.13-1.17]* |
| R56 | 96,350 | 112,250,170 | 0.96 | 0.83 | 0.84 | 0.13 | 0.12 | 0.09 | 1.14 [1.11-1.16]* | 1.10 [1.08-1.13]* |
| R57 | 17,300 | 114,219,960 | 0.16 | 0.15 | 0.14 | 0.02 | 0.03 | 0.02 | 1.18 [1.12-1.25]* | 1.15 [1.09-1.22]* |
| R58 | 2,750 | 114,246,900 | 0.03 | 0.02 | 0.02 | 0.01 | 0.01 | 0.01 | 1.35 [1.19-1.53]* | 1.30 [1.14-1.48]* |
| R59 | 59,160 | 113,578,790 | 0.64 | 0.49 | 0.47 | 0.15 | 0.17 | 0.15 | 1.35 [1.32-1.39]* | 1.31 [1.28-1.35]* |
| R60 | 92,340 | 113,763,250 | 0.96 | 0.78 | 0.72 | 0.18 | 0.24 | 0.20 | 1.33 [1.30-1.36]* | 1.26 [1.23-1.29]* |
| R61 | 10,700 | 114,111,200 | 0.11 | 0.09 | 0.08 | 0.03 | 0.03 | 0.02 | 1.36 [1.27-1.45]* | 1.24 [1.16-1.32]* |
| R62 | 16,720 | 113,655,560 | 0.17 | 0.14 | 0.16 | 0.03 | 0.01 | 0.00 | 1.07 [1.01-1.12] | 1.02 [0.97-1.07] |
| R63 | 200,090 | 112,055,920 | 2.05 | 1.72 | 1.65 | 0.32 | 0.40 | 0.31 | 1.24 [1.22-1.26]* | 1.18 [1.16-1.20]* |
| R64 | 5,220 | 114,260,690 | 0.05 | 0.05 | 0.04 | 0.00 | 0.01 | 0.01 | 1.16 [1.05-1.28]* | 1.17 [1.06-1.30]* |
| R65 | 11,950 | 114,240,850 | 0.12 | 0.10 | 0.09 | 0.02 | 0.02 | 0.02 | 1.23 [1.16-1.31]* | 1.18 [1.10-1.26]* |
| R68 | 19,650 | 113,923,150 | 0.19 | 0.17 | 0.16 | 0.02 | 0.03 | 0.03 | 1.21 [1.15-1.27]* | 1.19 [1.13-1.25]* |
| R69 | 169,740 | 106,597,460 | 1.78 | 1.55 | 1.43 | 0.24 | 0.35 | 0.29 | 1.25 [1.23-1.27]* | 1.19 [1.18-1.22]* |
| R70 | 1,440 | 114,239,870 | 0.02 | 0.01 | 0.01 | 0.00 | 0.01 | 0.00 | 1.50 [1.27-1.78]* | 1.43 [1.20-1.70]* |
| R71 | 1,190 | 114,257,880 | 0.01 | 0.01 | 0.01 | 0.00 | 0.00 | 0.00 | 1.22 [1.00-1.49] | 1.18 [0.97-1.45] |
| R72 | 6,570 | 114,210,350 | 0.06 | 0.06 | 0.05 | 0.01 | 0.01 | 0.01 | 1.23 [1.13-1.34]* | 1.18 [1.09-1.29]* |
| R73 | 24,160 | 114,027,320 | 0.25 | 0.20 | 0.19 | 0.05 | 0.06 | 0.04 | 1.31 [1.26-1.37]* | 1.21 [1.16-1.27]* |
| R74 | 10,960 | 114,178,830 | 0.11 | 0.09 | 0.09 | 0.01 | 0.02 | 0.01 | 1.21 [1.14-1.30]* | 1.15 [1.07-1.23]* |
| R76 | 3,080 | 114,226,880 | 0.03 | 0.03 | 0.02 | 0.01 | 0.01 | 0.01 | 1.38 [1.23-1.56]* | 1.31 [1.16-1.47]* |
| R77 | 6,710 | 114,229,350 | 0.07 | 0.06 | 0.05 | 0.01 | 0.01 | 0.01 | 1.26 [1.16-1.37]* | 1.18 [1.09-1.29]* |
| R78 | 900 | 114,255,170 | 0.01 | 0.01 | 0.01 | 0.00 | 0.00 | 0.00 | 1.19 [0.95-1.49] | 1.15 [0.91-1.45] |
| R79 | 129,520 | 113,225,980 | 1.29 | 1.11 | 1.04 | 0.18 | 0.25 | 0.19 | 1.24 [1.22-1.26]* | 1.18 [1.15-1.20]* |
| R80 | 5,510 | 114,174,500 | 0.06 | 0.05 | 0.04 | 0.01 | 0.01 | 0.01 | 1.33 [1.22-1.45]* | 1.28 [1.17-1.40]* |
| R81 | 1,600 | 114,177,010 | 0.02 | 0.01 | 0.01 | 0.00 | 0.00 | 0.00 | 1.20 [1.02-1.42] | 1.15 [0.97-1.36] |
| R82 | 11,310 | 114,135,950 | 0.11 | 0.10 | 0.09 | 0.01 | 0.02 | 0.01 | 1.20 [1.13-1.28]* | 1.15 [1.07-1.22]* |
| R83 | 490 | 114,263,100 | 0.00 | 0.00 | 0.00 | 0.00 | 0.00 | 0.00 | 1.20 [0.89-1.62] | 1.14 [0.84-1.56] |
| R84 | 3,660 | 114,244,720 | 0.04 | 0.03 | 0.03 | 0.01 | 0.01 | 0.01 | 1.29 [1.15-1.44]* | 1.16 [1.03-1.30] |
| R85 | 1,960 | 114,222,400 | 0.02 | 0.02 | 0.02 | 0.00 | 0.01 | 0.00 | 1.32 [1.14-1.53]* | 1.21 [1.04-1.40] |
| R87 | 9,950 | 113,723,210 | 0.09 | 0.09 | 0.08 | 0.00 | 0.01 | 0.01 | 1.16 [1.08-1.24]* | 1.12 [1.05-1.20]* |
| R89 | 1,830 | 114,250,300 | 0.02 | 0.01 | 0.01 | 0.01 | 0.01 | 0.01 | 1.48 [1.27-1.73]* | 1.41 [1.21-1.65]* |

| | | | | | | | | | | |
|---|---|---|---|---|---|---|---|---|---|---|
| R90 | 6,170 | 114,209,920 | 0.06 | 0.05 | 0.05 | 0.01 | 0.01 | 0.01 | 1.21 [1.11-1.32]* | 1.16 [1.06-1.27]* |
| R91 | 63,380 | 113,817,410 | 0.63 | 0.54 | 0.50 | 0.09 | 0.13 | 0.11 | 1.26 [1.22-1.29]* | 1.20 [1.17-1.24]* |
| R92 | 590 | 114,263,450 | 0.00 | 0.01 | 0.00 | 0.00 | 0.00 | 0.00 | 1.09 [0.81-1.46] | 1.06 [0.78-1.43] |
| R93 | 64,880 | 113,674,380 | 0.63 | 0.56 | 0.52 | 0.07 | 0.11 | 0.09 | 1.21 [1.18-1.25]* | 1.16 [1.13-1.19]* |
| R94 | 114,510 | 113,305,880 | 1.16 | 0.98 | 0.92 | 0.18 | 0.24 | 0.20 | 1.26 [1.24-1.29]* | 1.21 [1.18-1.23]* |
| any age - S00-T88 | | | | | | | | | | |
| S00 | 126,580 | 112,940,920 | 1.23 | 1.10 | 1.08 | 0.13 | 0.14 | 0.11 | 1.13 [1.11-1.15]* | 1.10 [1.08-1.12]* |
| S01 | 135,300 | 112,669,540 | 1.32 | 1.17 | 1.16 | 0.14 | 0.15 | 0.12 | 1.13 [1.11-1.15]* | 1.10 [1.08-1.12]* |
| S02 | 52,400 | 113,241,270 | 0.50 | 0.45 | 0.45 | 0.04 | 0.05 | 0.04 | 1.11 [1.07-1.14]* | 1.08 [1.04-1.11]* |
| S03 | 2,210 | 114,240,290 | 0.02 | 0.02 | 0.02 | 0.00 | 0.00 | 0.00 | 1.08 [0.94-1.25] | 1.04 [0.90-1.21] |
| S05 | 11,420 | 114,107,500 | 0.11 | 0.10 | 0.09 | 0.01 | 0.01 | 0.01 | 1.14 [1.06-1.21]* | 1.11 [1.03-1.18]* |
| S06 | 36,390 | 113,869,270 | 0.34 | 0.31 | 0.31 | 0.03 | 0.03 | 0.03 | 1.11 [1.07-1.15]* | 1.09 [1.05-1.13]* |
| S09 | 72,790 | 112,786,440 | 0.72 | 0.63 | 0.63 | 0.10 | 0.10 | 0.07 | 1.16 [1.13-1.19]* | 1.11 [1.08-1.14]* |
| S10 | 3,900 | 114,213,540 | 0.04 | 0.03 | 0.03 | 0.00 | 0.00 | 0.00 | 1.05 [0.94-1.17] | 1.00 [0.89-1.12] |
| S11 | 2,080 | 114,239,200 | 0.02 | 0.02 | 0.02 | 0.00 | 0.00 | 0.00 | 1.19 [1.03-1.38] | 1.13 [0.97-1.31] |
| S12 | 8,700 | 114,194,430 | 0.08 | 0.07 | 0.07 | 0.01 | 0.01 | 0.01 | 1.18 [1.09-1.27]* | 1.15 [1.06-1.24]* |
| S13 | 2,320 | 114,217,790 | 0.02 | 0.02 | 0.02 | 0.00 | 0.00 | 0.00 | 1.20 [1.04-1.38] | 1.12 [0.97-1.30] |
| S14 | 1,700 | 114,246,740 | 0.02 | 0.01 | 0.01 | 0.00 | 0.00 | 0.00 | 1.07 [0.90-1.27] | 1.07 [0.90-1.27] |
| S19 | 3,000 | 114,209,090 | 0.03 | 0.03 | 0.02 | 0.00 | 0.01 | 0.00 | 1.24 [1.10-1.41]* | 1.15 [1.02-1.30] |
| S20 | 13,120 | 114,136,550 | 0.13 | 0.11 | 0.11 | 0.02 | 0.02 | 0.01 | 1.18 [1.11-1.25]* | 1.12 [1.05-1.19]* |
| S21 | 2,880 | 114,227,160 | 0.03 | 0.02 | 0.02 | 0.00 | 0.00 | 0.00 | 1.11 [0.97-1.26] | 1.06 [0.93-1.21] |
| S22 | 42,790 | 113,855,770 | 0.41 | 0.37 | 0.34 | 0.05 | 0.07 | 0.06 | 1.21 [1.17-1.25]* | 1.15 [1.12-1.19]* |
| S27 | 13,690 | 114,108,850 | 0.12 | 0.12 | 0.11 | 0.00 | 0.01 | 0.01 | 1.11 [1.05-1.18]* | 1.08 [1.02-1.15] |
| S29 | 3,400 | 114,230,700 | 0.04 | 0.03 | 0.03 | 0.01 | 0.01 | 0.01 | 1.38 [1.23-1.55]* | 1.27 [1.13-1.43]* |
| S30 | 25,390 | 113,987,990 | 0.25 | 0.22 | 0.21 | 0.03 | 0.04 | 0.03 | 1.18 [1.13-1.23]* | 1.12 [1.07-1.17]* |
| S31 | 8,010 | 114,130,960 | 0.08 | 0.07 | 0.07 | 0.01 | 0.01 | 0.01 | 1.11 [1.03-1.19]* | 1.07 [0.99-1.16] |
| S32 | 47,250 | 113,876,780 | 0.44 | 0.41 | 0.39 | 0.03 | 0.05 | 0.04 | 1.13 [1.09-1.16]* | 1.09 [1.05-1.13]* |
| S33 | 600 | 114,260,130 | 0.01 | 0.01 | 0.01 | 0.00 | 0.00 | 0.00 | 1.08 [0.81-1.45] | 1.02 [0.76-1.37] |
| S35 | 590 | 114,264,450 | 0.01 | 0.01 | 0.01 | 0.00 | 0.00 | 0.00 | 1.08 [0.82-1.43] | 1.09 [0.82-1.45] |
| S36 | 5,840 | 114,189,370 | 0.05 | 0.05 | 0.05 | 0.00 | 0.01 | 0.00 | 1.11 [1.01-1.22] | 1.09 [1.00-1.20] |
| S37 | 4,310 | 114,219,690 | 0.04 | 0.04 | 0.04 | 0.00 | 0.00 | 0.00 | 1.00 [0.90-1.12] | 0.98 [0.87-1.09] |
| S39 | 10,090 | 114,106,080 | 0.10 | 0.09 | 0.08 | 0.01 | 0.01 | 0.01 | 1.17 [1.09-1.25]* | 1.09 [1.02-1.17] |
| S40 | 19,220 | 114,125,830 | 0.18 | 0.16 | 0.16 | 0.02 | 0.02 | 0.01 | 1.14 [1.08-1.20]* | 1.09 [1.03-1.15]* |
| S41 | 7,810 | 114,195,570 | 0.08 | 0.07 | 0.06 | 0.01 | 0.01 | 0.01 | 1.22 [1.13-1.32]* | 1.16 [1.07-1.26]* |
| S42 | 68,860 | 113,381,550 | 0.66 | 0.60 | 0.59 | 0.06 | 0.06 | 0.05 | 1.11 [1.08-1.14]* | 1.08 [1.06-1.11]* |
| S43 | 9,780 | 114,140,440 | 0.09 | 0.09 | 0.08 | 0.00 | 0.01 | 0.01 | 1.10 [1.02-1.18]* | 1.07 [0.99-1.15] |
| S44 | 550 | 114,261,770 | 0.01 | 0.00 | 0.00 | 0.00 | 0.00 | 0.00 | 1.24 [0.93-1.66] | 1.21 [0.90-1.63] |
| S46 | 4,000 | 114,229,020 | 0.04 | 0.03 | 0.03 | 0.00 | 0.00 | 0.00 | 1.10 [0.99-1.23] | 1.05 [0.94-1.18] |
| S49 | 5,700 | 114,215,910 | 0.06 | 0.05 | 0.05 | 0.01 | 0.01 | 0.01 | 1.20 [1.09-1.31]* | 1.12 [1.02-1.23] |
| S50 | 21,230 | 114,116,770 | 0.21 | 0.18 | 0.18 | 0.03 | 0.03 | 0.02 | 1.16 [1.10-1.22]* | 1.11 [1.05-1.16]* |
| S51 | 30,580 | 114,007,980 | 0.30 | 0.26 | 0.26 | 0.04 | 0.04 | 0.02 | 1.15 [1.11-1.20]* | 1.09 [1.04-1.14]* |
| S52 | 114,140 | 112,223,550 | 1.09 | 1.00 | 1.03 | 0.09 | 0.06 | 0.04 | 1.06 [1.04-1.08]* | 1.04 [1.02-1.06]* |
| S53 | 3,250 | 114,209,060 | 0.03 | 0.03 | 0.03 | 0.00 | 0.00 | 0.00 | 0.98 [0.87-1.11] | 0.95 [0.84-1.08] |
| S54 | 1,370 | 114,239,230 | 0.01 | 0.01 | 0.01 | 0.00 | 0.00 | 0.00 | 1.13 [0.94-1.35] | 1.09 [0.90-1.32] |
| S55 | 600 | 114,258,500 | 0.01 | 0.01 | 0.00 | 0.00 | 0.00 | 0.00 | 1.34 [1.02-1.75] | 1.33 [1.01-1.76] |
| S56 | 4,720 | 114,180,870 | 0.04 | 0.04 | 0.04 | 0.00 | 0.00 | 0.00 | 1.00 [0.90-1.11] | 0.98 [0.88-1.09] |
| S59 | 2,430 | 114,243,250 | 0.02 | 0.02 | 0.02 | 0.00 | 0.00 | 0.00 | 1.24 [1.08-1.43]* | 1.17 [1.01-1.35] |
| S60 | 17,040 | 114,106,760 | 0.16 | 0.15 | 0.14 | 0.02 | 0.02 | 0.01 | 1.14 [1.08-1.20]* | 1.10 [1.04-1.16]* |
| S61 | 72,940 | 113,057,610 | 0.67 | 0.64 | 0.64 | 0.03 | 0.03 | 0.01 | 1.05 [1.02-1.08]* | 1.02 [0.99-1.04] |
| S62 | 51,530 | 113,409,820 | 0.47 | 0.45 | 0.45 | 0.02 | 0.02 | 0.01 | 1.05 [1.02-1.08]* | 1.02 [0.98-1.05] |
| S63 | 7,660 | 114,139,110 | 0.07 | 0.07 | 0.06 | 0.01 | 0.01 | 0.01 | 1.19 [1.11-1.29]* | 1.14 [1.06-1.24]* |
| S64 | 10,010 | 114,049,320 | 0.08 | 0.09 | 0.08 | 0.00 | 0.00 | 0.00 | 1.00 [0.93-1.07] | 0.98 [0.91-1.06] |
| S65 | 2,720 | 114,215,310 | 0.02 | 0.02 | 0.02 | 0.00 | 0.00 | 0.00 | 1.02 [0.89-1.17] | 1.03 [0.90-1.18] |
| S66 | 18,380 | 113,853,050 | 0.16 | 0.16 | 0.15 | 0.00 | 0.00 | 0.00 | 1.03 [0.98-1.09] | 1.01 [0.95-1.06] |
| S67 | 2,790 | 114,200,260 | 0.02 | 0.02 | 0.03 | 0.00 | 0.00 | 0.00 | 0.87 [0.76-1.00] | 0.86 [0.75-0.98] |
| S68 | 6,380 | 114,130,470 | 0.05 | 0.06 | 0.06 | 0.00 | 0.00 | 0.00 | 0.98 [0.90-1.07] | 0.98 [0.90-1.08] |
| S69 | 6,770 | 114,177,830 | 0.07 | 0.06 | 0.06 | 0.01 | 0.01 | 0.01 | 1.21 [1.11-1.32]* | 1.15 [1.05-1.25]* |
| S70 | 18,860 | 114,128,460 | 0.18 | 0.16 | 0.16 | 0.02 | 0.02 | 0.02 | 1.16 [1.10-1.22]* | 1.11 [1.05-1.17]* |
| S71 | 4,800 | 114,201,600 | 0.05 | 0.04 | 0.04 | 0.01 | 0.00 | 0.00 | 1.07 [0.97-1.18] | 1.02 [0.92-1.13] |
| S72 | 128,350 | 113,226,460 | 1.19 | 1.12 | 1.08 | 0.07 | 0.11 | 0.10 | 1.10 [1.08-1.12]* | 1.09 [1.07-1.11]* |
| S73 | 1,830 | 114,242,960 | 0.02 | 0.02 | 0.02 | 0.00 | 0.00 | 0.00 | 1.09 [0.93-1.28] | 1.04 [0.88-1.22] |
| S76 | 3,980 | 114,221,130 | 0.04 | 0.03 | 0.03 | 0.00 | 0.00 | 0.00 | 1.14 [1.02-1.27] | 1.07 [0.96-1.20] |
| S79 | 9,520 | 114,195,470 | 0.09 | 0.08 | 0.08 | 0.01 | 0.02 | 0.02 | 1.25 [1.16-1.34]* | 1.20 [1.11-1.29]* |
| S80 | 36,840 | 113,968,360 | 0.35 | 0.32 | 0.31 | 0.04 | 0.04 | 0.02 | 1.14 [1.10-1.18]* | 1.08 [1.04-1.12]* |
| S81 | 28,110 | 113,996,560 | 0.28 | 0.24 | 0.23 | 0.04 | 0.05 | 0.03 | 1.20 [1.15-1.26]* | 1.12 [1.07-1.17]* |
| S82 | 81,330 | 112,712,560 | 0.75 | 0.72 | 0.69 | 0.03 | 0.06 | 0.04 | 1.09 [1.06-1.11]* | 1.05 [1.03-1.08]* |
| S83 | 10,720 | 114,056,350 | 0.10 | 0.09 | 0.09 | 0.01 | 0.01 | 0.01 | 1.15 [1.08-1.23]* | 1.08 [1.01-1.16] |
| S86 | 5,340 | 114,144,630 | 0.05 | 0.05 | 0.04 | 0.00 | 0.01 | 0.00 | 1.14 [1.04-1.25]* | 1.08 [0.98-1.19] |
| S89 | 7,640 | 114,181,540 | 0.08 | 0.06 | 0.06 | 0.01 | 0.01 | 0.01 | 1.23 [1.14-1.33]* | 1.15 [1.07-1.25]* |
| S90 | 9,660 | 114,170,210 | 0.09 | 0.08 | 0.08 | 0.01 | 0.01 | 0.00 | 1.11 [1.04-1.20]* | 1.06 [0.98-1.14] |
| S91 | 10,680 | 114,107,390 | 0.11 | 0.09 | 0.09 | 0.02 | 0.01 | 0.01 | 1.12 [1.05-1.19]* | 1.07 [1.00-1.15] |
| S92 | 16,520 | 113,997,170 | 0.15 | 0.14 | 0.14 | 0.01 | 0.02 | 0.01 | 1.13 [1.07-1.19]* | 1.08 [1.02-1.14]* |
| S93 | 5,220 | 114,193,160 | 0.05 | 0.04 | 0.04 | 0.01 | 0.01 | 0.01 | 1.19 [1.09-1.31]* | 1.12 [1.01-1.23] |
| S96 | 1,280 | 114,247,900 | 0.01 | 0.01 | 0.01 | 0.00 | 0.00 | 0.00 | 1.14 [0.95-1.38] | 1.06 [0.87-1.29] |
| S99 | 3,990 | 114,220,780 | 0.04 | 0.03 | 0.03 | 0.00 | 0.00 | 0.00 | 1.15 [1.03-1.28] | 1.06 [0.94-1.18] |
| T00 | 8,680 | 114,200,910 | 0.08 | 0.07 | 0.07 | 0.01 | 0.01 | 0.01 | 1.12 [1.04-1.21]* | 1.07 [0.99-1.16] |
| T01 | 4,150 | 114,237,960 | 0.04 | 0.04 | 0.04 | 0.01 | 0.00 | 0.00 | 1.12 [1.00-1.25] | 1.05 [0.94-1.18] |
| T02 | 2,800 | 114,236,060 | 0.03 | 0.02 | 0.02 | 0.00 | 0.00 | 0.00 | 1.17 [1.03-1.33] | 1.15 [1.01-1.32] |
| T09 | 1,400 | 114,248,940 | 0.01 | 0.01 | 0.01 | 0.00 | 0.00 | 0.00 | 1.22 [1.02-1.47] | 1.14 [0.95-1.38] |
| T11 | 5,760 | 114,220,790 | 0.06 | 0.05 | 0.05 | 0.01 | 0.01 | 0.01 | 1.19 [1.08-1.31]* | 1.12 [1.01-1.23] |
| T13 | 3,680 | 114,228,430 | 0.03 | 0.03 | 0.03 | 0.00 | 0.00 | 0.00 | 1.17 [1.04-1.31] | 1.10 [0.97-1.24] |
| T14 | 3,670 | 114,165,040 | 0.03 | 0.03 | 0.03 | 0.00 | 0.00 | 0.00 | 1.16 [1.03-1.29] | 1.09 [0.97-1.22] |
| T15 | 880 | 114,249,860 | 0.01 | 0.01 | 0.01 | 0.00 | 0.00 | 0.00 | 1.05 [0.83-1.32] | 1.03 [0.82-1.30] |
| T16 | 8,910 | 114,111,310 | 0.09 | 0.07 | 0.09 | 0.02 | 0.00 | 0.00 | 1.05 [0.98-1.12] | 1.03 [0.96-1.11] |
| T17 | 13,540 | 114,041,250 | 0.13 | 0.11 | 0.12 | 0.02 | 0.02 | 0.01 | 1.14 [1.07-1.20]* | 1.08 [1.02-1.15]* |
| T18 | 10,690 | 114,086,140 | 0.11 | 0.09 | 0.09 | 0.02 | 0.01 | 0.01 | 1.16 [1.09-1.24]* | 1.09 [1.02-1.16] |
| T19 | 1,020 | 114,252,480 | 0.01 | 0.01 | 0.01 | 0.00 | 0.00 | 0.00 | 1.19 [0.96-1.46] | 1.11 [0.89-1.37] |
| T20 | 4,680 | 114,165,740 | 0.04 | 0.04 | 0.04 | 0.00 | 0.00 | 0.00 | 0.99 [0.89-1.09] | 0.98 [0.88-1.08] |
| T21 | 6,670 | 114,135,910 | 0.06 | 0.06 | 0.06 | 0.00 | 0.00 | 0.00 | 1.00 [0.92-1.08] | 0.99 [0.91-1.08] |
| T22 | 5,570 | 114,153,870 | 0.05 | 0.05 | 0.05 | 0.00 | 0.00 | 0.00 | 1.01 [0.92-1.11] | 1.00 [0.91-1.09] |
| T23 | 5,650 | 114,150,400 | 0.05 | 0.05 | 0.05 | 0.00 | 0.00 | 0.00 | 1.01 [0.93-1.11] | 1.00 [0.91-1.09] |
| T24 | 4,870 | 114,182,010 | 0.04 | 0.04 | 0.05 | 0.00 | 0.00 | 0.00 | 0.92 [0.83-1.02] | 0.92 [0.83-1.02] |
| T25 | 2,490 | 114,222,310 | 0.02 | 0.02 | 0.02 | 0.00 | 0.00 | 0.00 | 0.88 [0.77-1.02] | 0.85 [0.74-0.98] |
| T29 | 1,080 | 114,248,500 | 0.01 | 0.01 | 0.01 | 0.00 | 0.00 | 0.00 | 0.93 [0.75-1.15] | 0.90 [0.73-1.12] |
| T31 | 15,310 | 113,970,280 | 0.14 | 0.13 | 0.14 | 0.01 | 0.00 | −0.01 | 0.97 [0.92-1.02] | 0.95 [0.90-1.01] |
| T36 | 3,430 | 114,188,880 | 0.04 | 0.03 | 0.03 | 0.01 | 0.01 | 0.01 | 1.26 [1.13-1.41]* | 1.17 [1.04-1.32]* |
| T37 | 1,290 | 114,241,450 | 0.01 | 0.01 | 0.01 | 0.00 | 0.00 | 0.00 | 1.30 [1.08-1.56]* | 1.21 [1.00-1.46] |
| T38 | 5,380 | 114,184,200 | 0.06 | 0.04 | 0.04 | 0.01 | 0.01 | 0.01 | 1.26 [1.15-1.37]* | 1.14 [1.04-1.25]* |
| T39 | 64,110 | 112,832,120 | 0.63 | 0.55 | 0.57 | 0.08 | 0.06 | 0.03 | 1.10 [1.07-1.13]* | 1.05 [1.02-1.08]* |
| T40 | 27,960 | 113,812,350 | 0.27 | 0.24 | 0.23 | 0.03 | 0.04 | 0.02 | 1.18 [1.13-1.22]* | 1.08 [1.03-1.12]* |
| T41 | 730 | 114,260,920 | 0.01 | 0.01 | 0.01 | 0.00 | 0.00 | 0.00 | 1.18 [0.92-1.50] | 1.06 [0.82-1.37] |
| T42 | 20,040 | 113,769,620 | 0.19 | 0.17 | 0.17 | 0.02 | 0.03 | 0.01 | 1.17 [1.12-1.23]* | 1.08 [1.02-1.13]* |
| T43 | 32,120 | 113,520,310 | 0.32 | 0.27 | 0.27 | 0.04 | 0.04 | 0.03 | 1.16 [1.12-1.21]* | 1.09 [1.05-1.13]* |
| T44 | 6,900 | 114,140,510 | 0.07 | 0.06 | 0.06 | 0.01 | 0.01 | 0.01 | 1.16 [1.07-1.26]* | 1.10 [1.01-1.19] |
| T45 | 9,170 | 114,112,260 | 0.10 | 0.08 | 0.08 | 0.02 | 0.02 | 0.01 | 1.26 [1.18-1.35]* | 1.15 [1.08-1.24]* |
| T46 | 6,220 | 114,185,630 | 0.06 | 0.05 | 0.05 | 0.01 | 0.01 | 0.01 | 1.19 [1.10-1.30]* | 1.11 [1.02-1.22] |
| T47 | 3,580 | 114,212,760 | 0.04 | 0.03 | 0.03 | 0.01 | 0.00 | 0.00 | 1.15 [1.03-1.29] | 1.04 [0.92-1.17] |
| T48 | 1,910 | 114,224,170 | 0.02 | 0.02 | 0.02 | 0.00 | 0.00 | 0.00 | 1.20 [1.04-1.40] | 1.08 [0.93-1.26] |

| | | | | | | | | | | |
|---|---|---|---|---|---|---|---|---|---|---|
| T49 | 1,610 | 114,226,920 | 0.02 | 0.01 | 0.01 | 0.00 | 0.00 | 0.00 | 1.02 [0.87-1.21] | 0.98 [0.82-1.16] |
| T50 | 8,420 | 114,035,030 | 0.08 | 0.07 | 0.07 | 0.01 | 0.01 | 0.01 | 1.18 [1.09-1.27]* | 1.08 [1.00-1.17] |
| T51 | 26,670 | 113,639,370 | 0.25 | 0.23 | 0.22 | 0.02 | 0.03 | 0.02 | 1.14 [1.10-1.19]* | 1.07 [1.03-1.12]* |
| T52 | 1,610 | 114,229,250 | 0.01 | 0.01 | 0.02 | 0.00 | 0.00 | 0.00 | 0.87 [0.74-1.04] | 0.85 [0.72-1.02] |
| T54 | 1,630 | 114,242,270 | 0.02 | 0.01 | 0.02 | 0.00 | 0.00 | 0.00 | 0.98 [0.82-1.16] | 0.95 [0.80-1.14] |
| T56 | 720 | 114,255,770 | 0.01 | 0.01 | 0.01 | 0.00 | 0.00 | 0.00 | 1.17 [0.91-1.50] | 1.10 [0.85-1.42] |
| T58 | 600 | 114,254,090 | 0.01 | 0.01 | 0.00 | 0.00 | 0.00 | 0.00 | 1.08 [0.81-1.43] | 1.07 [0.80-1.43] |
| T59 | 2,130 | 114,223,220 | 0.02 | 0.02 | 0.02 | 0.00 | 0.00 | 0.00 | 1.05 [0.90-1.23] | 1.02 [0.87-1.19] |
| T62 | 780 | 114,249,880 | 0.01 | 0.01 | 0.01 | 0.00 | 0.00 | 0.00 | 1.24 [0.98-1.56] | 1.17 [0.92-1.48] |
| T63 | 1,010 | 114,246,240 | 0.01 | 0.01 | 0.01 | 0.00 | 0.00 | 0.00 | 1.10 [0.89-1.37] | 1.02 [0.82-1.27] |
| T65 | 1,640 | 114,233,320 | 0.02 | 0.02 | 0.01 | 0.00 | 0.00 | 0.00 | 1.05 [0.89-1.24] | 1.00 [0.84-1.18] |
| T68 | 9,510 | 114,232,630 | 0.09 | 0.08 | 0.08 | 0.00 | 0.01 | 0.01 | 1.10 [1.02-1.19] | 1.12 [1.04-1.21]* |
| T71 | 870 | 114,257,950 | 0.01 | 0.01 | 0.01 | 0.00 | 0.00 | 0.00 | 0.96 [0.76-1.22] | 0.94 [0.74-1.21] |
| T74 | 1,580 | 114,238,740 | 0.01 | 0.01 | 0.02 | 0.00 | 0.00 | 0.00 | 0.91 [0.76-1.08] | 0.86 [0.72-1.03] |
| T75 | 1,080 | 114,241,720 | 0.01 | 0.01 | 0.01 | 0.00 | 0.00 | 0.00 | 1.09 [0.89-1.34] | 1.02 [0.83-1.26] |
| T78 | 24,690 | 113,700,390 | 0.48 | 0.15 | 0.16 | 0.33 | 0.33 | 0.29 | 3.07 [2.96-3.18]* | 2.54 [2.45-2.64]* |
| T79 | 12,940 | 114,111,220 | 0.12 | 0.11 | 0.11 | 0.00 | 0.01 | 0.01 | 1.09 [1.03-1.17]* | 1.07 [1.00-1.14] |
| T80 | 5,910 | 114,184,060 | 0.06 | 0.05 | 0.05 | 0.01 | 0.02 | 0.01 | 1.33 [1.23-1.45]* | 1.27 [1.16-1.39]* |
| T81 | 180,710 | 111,293,390 | 1.89 | 1.56 | 1.46 | 0.33 | 0.43 | 0.34 | 1.29 [1.27-1.31]* | 1.22 [1.20-1.24]* |
| T82 | 42,800 | 113,763,580 | 0.44 | 0.36 | 0.34 | 0.08 | 0.10 | 0.09 | 1.31 [1.27-1.35]* | 1.25 [1.21-1.29]* |
| T83 | 42,140 | 113,964,420 | 0.39 | 0.37 | 0.34 | 0.02 | 0.05 | 0.04 | 1.14 [1.10-1.18]* | 1.11 [1.07-1.15]* |
| T84 | 56,750 | 113,346,570 | 0.58 | 0.48 | 0.44 | 0.10 | 0.14 | 0.11 | 1.31 [1.28-1.35]* | 1.24 [1.20-1.27]* |
| T85 | 38,000 | 113,776,720 | 0.39 | 0.32 | 0.30 | 0.07 | 0.09 | 0.07 | 1.30 [1.26-1.35]* | 1.23 [1.19-1.28]* |
| T86 | 5,280 | 114,190,880 | 0.05 | 0.04 | 0.04 | 0.01 | 0.01 | 0.01 | 1.33 [1.22-1.46]* | 1.29 [1.18-1.41]* |
| T87 | 3,190 | 114,227,160 | 0.03 | 0.03 | 0.02 | 0.01 | 0.01 | 0.01 | 1.35 [1.20-1.52]* | 1.25 [1.11-1.41]* |
| T88 | 11,840 | 114,017,800 | 0.13 | 0.10 | 0.09 | 0.04 | 0.04 | 0.03 | 1.47 [1.39-1.56]* | 1.33 [1.25-1.41]* |
| T90 | 9,870 | 114,056,180 | 0.10 | 0.08 | 0.08 | 0.02 | 0.02 | 0.02 | 1.29 [1.21-1.38]* | 1.23 [1.15-1.31]* |
| T91 | 4,200 | 114,187,190 | 0.04 | 0.04 | 0.03 | 0.01 | 0.01 | 0.01 | 1.30 [1.17-1.44]* | 1.19 [1.07-1.32]* |
| T92 | 7,940 | 114,081,110 | 0.07 | 0.07 | 0.06 | 0.01 | 0.01 | 0.01 | 1.17 [1.09-1.26]* | 1.12 [1.04-1.21]* |
| T93 | 10,340 | 114,041,320 | 0.10 | 0.09 | 0.08 | 0.01 | 0.02 | 0.01 | 1.22 [1.15-1.31]* | 1.15 [1.07-1.23]* |
| T94 | 900 | 114,253,490 | 0.01 | 0.01 | 0.01 | 0.00 | 0.00 | 0.00 | 1.26 [1.01-1.57] | 1.21 [0.96-1.52] |
| T95 | 1,110 | 114,243,120 | 0.01 | 0.01 | 0.01 | 0.00 | 0.00 | 0.00 | 0.97 [0.79-1.19] | 0.96 [0.78-1.18] |
| T98 | 1,600 | 114,242,620 | 0.02 | 0.01 | 0.01 | 0.00 | 0.00 | 0.00 | 1.19 [1.01-1.41] | 1.10 [0.92-1.30] |
| any age - U00-U89 | | | | | | | | | | |
| U07 | 52,700 | 114,218,800 | 0.54 | 0.44 | 0.43 | 0.10 | 0.11 | 0.08 | 1.27 [1.23-1.31]* | 1.18 [1.15-1.22]* |
| U80 | 18,650 | 114,146,070 | 0.21 | 0.15 | 0.14 | 0.06 | 0.07 | 0.06 | 1.49 [1.42-1.56]* | 1.40 [1.33-1.47]* |
| U81 | 1,080 | 114,265,170 | 0.01 | 0.01 | 0.01 | 0.00 | 0.00 | 0.00 | 1.19 [0.96-1.46] | 1.14 [0.92-1.41] |
| U82 | 37,090 | 114,163,650 | 0.41 | 0.30 | 0.29 | 0.10 | 0.12 | 0.10 | 1.40 [1.35-1.45]* | 1.32 [1.27-1.37]* |
| U83 | 22,120 | 114,211,120 | 0.24 | 0.18 | 0.18 | 0.06 | 0.06 | 0.05 | 1.35 [1.29-1.41]* | 1.28 [1.22-1.34]* |
| U84 | 450 | 114,269,520 | 0.00 | 0.00 | 0.00 | 0.00 | 0.00 | 0.00 | 1.40 [1.02-1.93] | 1.35 [0.97-1.87] |
| U88 | 4,540 | 114,243,610 | 0.05 | 0.04 | 0.04 | 0.01 | 0.01 | 0.01 | 1.29 [1.16-1.42]* | 1.22 [1.10-1.35]* |
| U89 | 5,180 | 114,237,580 | 0.06 | 0.04 | 0.04 | 0.01 | 0.01 | 0.01 | 1.34 [1.22-1.47]* | 1.26 [1.14-1.38]* |
| any age - V00-Y99 | | | | | | | | | | |
| V01 | 690 | 114,258,600 | 0.01 | 0.01 | 0.01 | 0.00 | 0.00 | 0.00 | 0.99 [0.76-1.29] | 0.96 [0.73-1.26] |
| V02 | 600 | 114,257,400 | 0.01 | 0.01 | 0.01 | 0.00 | 0.00 | 0.00 | 0.99 [0.74-1.32] | 0.99 [0.74-1.33] |
| V03 | 11,280 | 114,025,790 | 0.10 | 0.10 | 0.10 | 0.00 | 0.00 | 0.00 | 1.02 [0.96-1.09] | 1.00 [0.94-1.07] |
| V04 | 700 | 114,254,370 | 0.01 | 0.01 | 0.01 | 0.00 | 0.00 | 0.00 | 1.32 [1.03-1.70] | 1.27 [0.98-1.64] |
| V09 | 2,480 | 114,235,660 | 0.03 | 0.02 | 0.02 | 0.00 | 0.00 | 0.00 | 1.21 [1.05-1.38]* | 1.13 [0.98-1.30] |
| V11 | 620 | 114,261,740 | 0.01 | 0.01 | 0.01 | 0.00 | 0.00 | 0.00 | 1.00 [0.76-1.31] | 0.95 [0.71-1.26] |
| V13 | 3,230 | 114,208,290 | 0.03 | 0.03 | 0.03 | 0.00 | 0.00 | 0.00 | 0.91 [0.80-1.03] | 0.91 [0.80-1.03] |
| V17 | 1,250 | 114,250,880 | 0.01 | 0.01 | 0.01 | 0.00 | 0.00 | 0.00 | 0.98 [0.80-1.19] | 0.97 [0.79-1.18] |
| V18 | 19,430 | 113,897,890 | 0.18 | 0.17 | 0.18 | 0.01 | 0.00 | 0.00 | 1.02 [0.97-1.07] | 1.00 [0.95-1.05] |
| V19 | 2,920 | 114,218,500 | 0.03 | 0.02 | 0.03 | 0.00 | 0.00 | 0.00 | 1.07 [0.95-1.22] | 1.05 [0.92-1.19] |
| V23 | 4,020 | 114,183,990 | 0.03 | 0.04 | 0.03 | 0.00 | 0.00 | 0.00 | 0.96 [0.86-1.08] | 0.96 [0.86-1.08] |
| V27 | 980 | 114,249,490 | 0.01 | 0.01 | 0.01 | 0.00 | 0.00 | 0.00 | 0.87 [0.69-1.10] | 0.82 [0.64-1.04] |
| V28 | 5,530 | 114,158,440 | 0.05 | 0.05 | 0.05 | 0.00 | 0.00 | 0.00 | 1.02 [0.92-1.12] | 0.99 [0.90-1.09] |
| V29 | 1,380 | 114,234,020 | 0.01 | 0.01 | 0.01 | 0.00 | 0.00 | 0.00 | 1.05 [0.87-1.26] | 1.01 [0.83-1.22] |
| V43 | 9,210 | 114,052,090 | 0.09 | 0.08 | 0.07 | 0.01 | 0.01 | 0.01 | 1.14 [1.06-1.22]* | 1.09 [1.01-1.17] |
| V44 | 850 | 114,249,730 | 0.01 | 0.01 | 0.01 | 0.00 | 0.00 | 0.00 | 1.19 [0.95-1.50] | 1.11 [0.87-1.40] |
| V47 | 4,250 | 114,183,920 | 0.04 | 0.04 | 0.04 | 0.00 | 0.01 | 0.00 | 1.16 [1.04-1.28]* | 1.11 [1.00-1.24] |
| V48 | 5,190 | 114,189,050 | 0.05 | 0.04 | 0.04 | 0.00 | 0.01 | 0.00 | 1.14 [1.03-1.25]* | 1.09 [0.99-1.21] |
| V49 | 3,980 | 114,164,690 | 0.04 | 0.03 | 0.03 | 0.00 | 0.00 | 0.00 | 1.11 [0.99-1.23] | 1.08 [0.96-1.20] |
| V78 | 1,940 | 114,248,810 | 0.02 | 0.02 | 0.02 | 0.00 | 0.00 | 0.00 | 1.14 [0.98-1.34] | 1.09 [0.93-1.29] |
| V79 | 660 | 114,260,040 | 0.01 | 0.01 | 0.01 | 0.00 | 0.00 | 0.00 | 1.08 [0.83-1.42] | 1.03 [0.78-1.35] |
| V80 | 5,990 | 114,148,660 | 0.05 | 0.05 | 0.05 | 0.00 | 0.00 | 0.00 | 0.95 [0.86-1.04] | 0.93 [0.85-1.02] |
| V82 | 480 | 114,264,050 | 0.01 | 0.00 | 0.00 | 0.00 | 0.00 | 0.00 | 1.42 [1.05-1.92] | 1.42 [1.05-1.93] |
| V86 | 1,430 | 114,252,040 | 0.01 | 0.01 | 0.01 | 0.00 | 0.00 | 0.00 | 1.08 [0.90-1.29] | 1.02 [0.85-1.23] |
| V89 | 1,790 | 114,193,790 | 0.02 | 0.02 | 0.01 | 0.00 | 0.00 | 0.00 | 1.20 [1.02-1.40] | 1.12 [0.95-1.32] |
| W00 | 7,880 | 114,139,160 | 0.07 | 0.07 | 0.06 | 0.00 | 0.01 | 0.00 | 1.09 [1.01-1.18] | 1.07 [0.99-1.16] |
| W01 | 163,090 | 112,375,980 | 1.57 | 1.42 | 1.38 | 0.15 | 0.19 | 0.15 | 1.14 [1.12-1.16]* | 1.10 [1.08-1.12]* |
| W02 | 8,990 | 114,090,990 | 0.09 | 0.08 | 0.09 | 0.01 | 0.00 | 0.00 | 1.01 [0.94-1.09] | 0.99 [0.92-1.07] |
| W03 | 12,540 | 114,034,150 | 0.12 | 0.11 | 0.12 | 0.01 | 0.01 | 0.00 | 1.05 [0.99-1.11] | 1.01 [0.95-1.08] |
| W04 | 1,980 | 114,152,760 | 0.02 | 0.02 | 0.02 | 0.00 | 0.00 | 0.00 | 1.01 [0.87-1.18] | 1.00 [0.86-1.17] |
| W05 | 3,280 | 114,243,210 | 0.03 | 0.03 | 0.03 | 0.01 | 0.01 | 0.01 | 1.24 [1.10-1.39]* | 1.19 [1.05-1.34]* |
| W06 | 38,680 | 113,959,670 | 0.38 | 0.33 | 0.32 | 0.05 | 0.05 | 0.04 | 1.17 [1.12-1.21]* | 1.13 [1.09-1.17]* |
| W07 | 23,040 | 114,022,370 | 0.22 | 0.20 | 0.20 | 0.03 | 0.03 | 0.02 | 1.14 [1.09-1.19]* | 1.11 [1.06-1.16]* |
| W08 | 7,900 | 114,138,180 | 0.08 | 0.07 | 0.07 | 0.01 | 0.01 | 0.00 | 1.09 [1.00-1.17] | 1.06 [0.98-1.15] |
| W09 | 23,160 | 113,882,380 | 0.24 | 0.19 | 0.23 | 0.05 | 0.01 | 0.01 | 1.05 [1.01-1.10] | 1.04 [0.99-1.08] |
| W10 | 66,850 | 113,331,210 | 0.63 | 0.58 | 0.55 | 0.05 | 0.08 | 0.06 | 1.15 [1.12-1.18]* | 1.11 [1.08-1.14]* |
| W11 | 9,730 | 114,073,200 | 0.08 | 0.09 | 0.08 | 0.00 | 0.00 | 0.00 | 1.06 [0.98-1.13] | 1.02 [0.94-1.09] |
| W12 | 590 | 114,256,110 | 0.01 | 0.00 | 0.00 | 0.00 | 0.00 | 0.00 | 1.19 [0.90-1.57] | 1.16 [0.88-1.55] |
| W13 | 8,370 | 114,075,130 | 0.08 | 0.07 | 0.08 | 0.00 | 0.00 | 0.00 | 0.99 [0.92-1.07] | 0.97 [0.90-1.05] |
| W14 | 2,960 | 114,202,700 | 0.03 | 0.03 | 0.03 | 0.00 | 0.00 | 0.00 | 0.99 [0.87-1.12] | 0.99 [0.87-1.12] |
| W17 | 15,790 | 113,943,730 | 0.15 | 0.14 | 0.14 | 0.01 | 0.01 | 0.00 | 1.05 [0.99-1.11] | 1.02 [0.97-1.08] |
| W18 | 108,010 | 113,415,770 | 1.07 | 0.93 | 0.90 | 0.14 | 0.17 | 0.14 | 1.19 [1.16-1.22]* | 1.15 [1.12-1.17]* |
| W19 | 243,800 | 112,102,860 | 2.40 | 2.12 | 2.04 | 0.28 | 0.36 | 0.29 | 1.17 [1.16-1.19]* | 1.14 [1.12-1.16]* |
| W20 | 7,300 | 114,125,080 | 0.07 | 0.06 | 0.06 | 0.00 | 0.00 | 0.00 | 1.03 [0.95-1.11] | 0.99 [0.91-1.07] |
| W21 | 6,690 | 114,143,180 | 0.07 | 0.06 | 0.06 | 0.01 | 0.01 | 0.01 | 1.11 [1.03-1.21] | 1.08 [0.99-1.18] |
| W22 | 35,960 | 113,708,720 | 0.35 | 0.31 | 0.31 | 0.04 | 0.04 | 0.02 | 1.12 [1.08-1.16]* | 1.08 [1.04-1.12]* |
| W23 | 23,440 | 113,856,700 | 0.22 | 0.20 | 0.21 | 0.02 | 0.00 | 0.00 | 1.02 [0.98-1.07] | 1.00 [0.96-1.05] |
| W25 | 11,920 | 113,960,230 | 0.11 | 0.10 | 0.10 | 0.00 | 0.00 | 0.00 | 1.02 [0.96-1.09] | 1.00 [0.94-1.07] |
| W26 | 10,520 | 114,105,850 | 0.10 | 0.09 | 0.09 | 0.01 | 0.01 | 0.00 | 1.06 [0.99-1.13] | 1.02 [0.95-1.09] |
| W27 | 4,890 | 114,161,880 | 0.04 | 0.04 | 0.04 | 0.00 | 0.00 | 0.00 | 0.99 [0.89-1.10] | 0.95 [0.86-1.06] |
| W28 | 670 | 114,256,060 | 0.00 | 0.01 | 0.01 | 0.00 | 0.00 | 0.00 | 0.85 [0.63-1.13] | 0.83 [0.62-1.11] |
| W29 | 5,040 | 114,182,900 | 0.04 | 0.04 | 0.04 | 0.00 | 0.00 | 0.00 | 0.99 [0.89-1.09] | 0.98 [0.88-1.08] |
| W31 | 4,680 | 114,163,100 | 0.04 | 0.04 | 0.04 | 0.00 | 0.00 | 0.00 | 1.02 [0.92-1.13] | 1.02 [0.92-1.13] |
| W34 | 790 | 114,249,910 | 0.01 | 0.01 | 0.01 | 0.00 | 0.00 | 0.00 | 1.06 [0.83-1.35] | 1.03 [0.80-1.33] |
| W44 | 25,020 | 113,834,870 | 0.25 | 0.21 | 0.23 | 0.04 | 0.02 | 0.01 | 1.10 [1.06-1.15]* | 1.06 [1.02-1.11]* |
| W45 | 8,300 | 114,089,460 | 0.08 | 0.07 | 0.07 | 0.01 | 0.01 | 0.00 | 1.07 [0.99-1.15] | 1.03 [0.96-1.12] |
| W46 | 990 | 114,264,200 | 0.01 | 0.01 | 0.01 | 0.00 | 0.00 | 0.00 | 1.08 [0.86-1.34] | 0.99 [0.78-1.24] |
| W49 | 2,110 | 114,217,390 | 0.02 | 0.02 | 0.02 | 0.00 | 0.00 | 0.00 | 1.19 [1.03-1.38] | 1.18 [1.02-1.37] |
| W50 | 11,980 | 114,046,680 | 0.12 | 0.10 | 0.11 | 0.02 | 0.01 | 0.01 | 1.11 [1.05-1.18]* | 1.08 [1.01-1.15] |
| W51 | 9,270 | 114,083,900 | 0.09 | 0.08 | 0.09 | 0.01 | 0.01 | 0.00 | 1.06 [0.99-1.14] | 1.03 [0.96-1.10] |
| W54 | 12,010 | 114,086,980 | 0.11 | 0.11 | 0.11 | 0.00 | 0.00 | −0.01 | 0.98 [0.92-1.05] | 0.95 [0.89-1.01] |
| W55 | 5,310 | 114,191,360 | 0.04 | 0.05 | 0.05 | 0.00 | 0.00 | 0.00 | 0.98 [0.89-1.08] | 0.92 [0.83-1.02] |
| W57 | 6,480 | 114,173,710 | 0.07 | 0.05 | 0.05 | 0.02 | 0.02 | 0.02 | 1.40 [1.29-1.52]* | 1.31 [1.21-1.43]* |
| W60 | 590 | 114,261,330 | 0.01 | 0.00 | 0.00 | 0.00 | 0.00 | 0.00 | 1.35 [1.04-1.76] | 1.27 [0.97-1.67] |

| | | | | | | | | | | |
|---|---|---|---|---|---|---|---|---|---|---|
| W64 | 1,330 | 114,257,110 | 0.01 | 0.01 | 0.01 | 0.00 | 0.00 | 0.00 | 1.04 [0.86-1.26] | 1.03 [0.85-1.25] |
| W78 | 3,720 | 114,257,790 | 0.03 | 0.03 | 0.03 | 0.00 | 0.00 | 0.00 | 1.10 [0.97-1.23] | 1.07 [0.95-1.21] |
| W79 | 5,690 | 114,228,490 | 0.06 | 0.05 | 0.05 | 0.01 | 0.01 | 0.01 | 1.27 [1.16-1.39]* | 1.21 [1.10-1.32]* |
| W80 | 2,320 | 114,256,690 | 0.02 | 0.02 | 0.02 | 0.00 | 0.00 | 0.00 | 1.18 [1.02-1.37] | 1.12 [0.96-1.30] |
| W84 | 2,160 | 114,264,860 | 0.02 | 0.02 | 0.02 | 0.00 | 0.00 | 0.00 | 1.06 [0.90-1.24] | 1.03 [0.88-1.21] |
| X04 | 700 | 114,255,340 | 0.01 | 0.01 | 0.01 | 0.00 | 0.00 | 0.00 | 1.02 [0.78-1.33] | 0.99 [0.75-1.29] |
| X08 | 660 | 114,257,940 | 0.01 | 0.01 | 0.01 | 0.00 | 0.00 | 0.00 | 1.04 [0.79-1.37] | 1.04 [0.79-1.37] |
| X09 | 930 | 114,250,620 | 0.01 | 0.01 | 0.01 | 0.00 | 0.00 | 0.00 | 1.01 [0.80-1.27] | 0.98 [0.78-1.25] |
| X10 | 4,420 | 114,173,720 | 0.04 | 0.04 | 0.04 | 0.00 | 0.00 | 0.00 | 1.00 [0.91-1.11] | 0.99 [0.89-1.10] |
| X11 | 1,400 | 114,237,020 | 0.01 | 0.01 | 0.01 | 0.00 | 0.00 | 0.00 | 0.97 [0.81-1.17] | 0.94 [0.78-1.14] |
| X12 | 2,670 | 114,206,380 | 0.02 | 0.02 | 0.03 | 0.00 | 0.00 | 0.00 | 0.94 [0.82-1.08] | 0.92 [0.81-1.06] |
| X15 | 1,860 | 114,234,120 | 0.02 | 0.02 | 0.02 | 0.00 | 0.00 | 0.00 | 1.04 [0.89-1.22] | 1.03 [0.88-1.21] |
| X16 | 1,320 | 114,249,860 | 0.01 | 0.01 | 0.01 | 0.00 | 0.00 | 0.00 | 0.99 [0.82-1.21] | 0.96 [0.79-1.17] |
| X19 | 860 | 114,255,330 | 0.01 | 0.01 | 0.01 | 0.00 | 0.00 | 0.00 | 1.12 [0.89-1.41] | 1.07 [0.85-1.36] |
| X23 | 1,330 | 114,246,880 | 0.01 | 0.01 | 0.01 | 0.00 | 0.00 | 0.00 | 1.17 [0.97-1.41] | 1.09 [0.90-1.32] |
| X31 | 1,320 | 114,263,320 | 0.01 | 0.01 | 0.01 | 0.00 | 0.00 | 0.00 | 1.05 [0.85-1.29] | 1.06 [0.86-1.31] |
| X40 | 15,820 | 113,938,010 | 0.15 | 0.14 | 0.14 | 0.02 | 0.01 | 0.00 | 1.07 [1.02-1.13]* | 1.03 [0.97-1.09] |
| X41 | 9,200 | 114,055,650 | 0.09 | 0.08 | 0.08 | 0.01 | 0.01 | 0.01 | 1.15 [1.08-1.24]* | 1.08 [1.01-1.16] |
| X42 | 8,270 | 114,166,080 | 0.08 | 0.07 | 0.07 | 0.01 | 0.01 | 0.01 | 1.22 [1.13-1.31]* | 1.11 [1.03-1.20]* |
| X43 | 1,940 | 114,235,500 | 0.02 | 0.02 | 0.02 | 0.00 | 0.00 | 0.00 | 1.10 [0.94-1.29] | 1.05 [0.90-1.24] |
| X44 | 12,970 | 114,020,420 | 0.13 | 0.11 | 0.11 | 0.02 | 0.02 | 0.01 | 1.17 [1.10-1.24]* | 1.09 [1.03-1.16]* |
| X45 | 3,580 | 114,181,050 | 0.03 | 0.03 | 0.03 | 0.00 | 0.00 | 0.00 | 1.12 [1.00-1.26] | 1.07 [0.95-1.21] |
| X46 | 1,100 | 114,239,960 | 0.01 | 0.01 | 0.01 | 0.00 | 0.00 | 0.00 | 0.91 [0.73-1.12] | 0.88 [0.71-1.09] |
| X47 | 1,300 | 114,244,730 | 0.01 | 0.01 | 0.01 | 0.00 | 0.00 | 0.00 | 1.11 [0.92-1.34] | 1.08 [0.89-1.31] |
| X49 | 5,210 | 114,144,650 | 0.06 | 0.04 | 0.04 | 0.02 | 0.02 | 0.01 | 1.35 [1.24-1.47]* | 1.27 [1.16-1.38]* |
| X50 | 18,770 | 113,982,060 | 0.17 | 0.16 | 0.16 | 0.01 | 0.02 | 0.01 | 1.11 [1.06-1.17]* | 1.05 [1.00-1.10] |
| X51 | 1,240 | 114,250,550 | 0.01 | 0.01 | 0.01 | 0.00 | 0.00 | 0.00 | 1.19 [0.98-1.44] | 1.15 [0.95-1.40] |
| X58 | 5,410 | 114,195,550 | 0.07 | 0.04 | 0.04 | 0.02 | 0.03 | 0.02 | 1.60 [1.47-1.75]* | 1.44 [1.32-1.58]* |
| X59 | 77,870 | 112,918,590 | 0.78 | 0.67 | 0.65 | 0.11 | 0.13 | 0.09 | 1.19 [1.16-1.22]* | 1.14 [1.11-1.17]* |
| X60 | 50,280 | 113,147,240 | 0.50 | 0.43 | 0.45 | 0.07 | 0.05 | 0.03 | 1.11 [1.08-1.15]* | 1.06 [1.02-1.09]* |
| X61 | 34,660 | 113,448,130 | 0.34 | 0.30 | 0.29 | 0.04 | 0.05 | 0.03 | 1.16 [1.12-1.21]* | 1.08 [1.04-1.13]* |
| X62 | 19,650 | 113,924,460 | 0.19 | 0.17 | 0.17 | 0.02 | 0.03 | 0.01 | 1.16 [1.10-1.22]* | 1.05 [1.00-1.11] |
| X63 | 4,860 | 114,180,910 | 0.05 | 0.04 | 0.04 | 0.01 | 0.01 | 0.00 | 1.17 [1.06-1.28]* | 1.10 [1.00-1.21] |
| X64 | 17,510 | 113,879,610 | 0.18 | 0.15 | 0.15 | 0.03 | 0.03 | 0.02 | 1.20 [1.14-1.26]* | 1.10 [1.05-1.16]* |
| X65 | 23,830 | 113,735,250 | 0.23 | 0.21 | 0.20 | 0.02 | 0.03 | 0.02 | 1.15 [1.10-1.21]* | 1.08 [1.03-1.13]* |
| X69 | 1,820 | 114,237,990 | 0.02 | 0.02 | 0.02 | 0.00 | 0.00 | 0.00 | 1.07 [0.91-1.26] | 1.02 [0.86-1.20] |
| X70 | 1,380 | 114,253,350 | 0.01 | 0.01 | 0.01 | 0.00 | 0.00 | 0.00 | 0.97 [0.81-1.17] | 0.96 [0.79-1.17] |
| X78 | 12,040 | 114,090,340 | 0.12 | 0.10 | 0.11 | 0.02 | 0.01 | 0.00 | 1.10 [1.03-1.17]* | 1.02 [0.96-1.09] |
| X79 | 470 | 114,265,450 | 0.00 | 0.00 | 0.00 | 0.00 | 0.00 | 0.00 | 1.23 [0.90-1.67] | 1.08 [0.78-1.50] |
| X83 | 1,030 | 114,259,270 | 0.01 | 0.01 | 0.01 | 0.00 | 0.00 | 0.00 | 1.03 [0.83-1.28] | 0.95 [0.76-1.18] |
| X84 | 1,540 | 114,247,360 | 0.01 | 0.01 | 0.01 | 0.00 | 0.00 | 0.00 | 1.04 [0.87-1.24] | 0.94 [0.78-1.13] |
| X99 | 5,250 | 114,156,100 | 0.05 | 0.05 | 0.05 | 0.00 | 0.00 | 0.00 | 1.06 [0.96-1.16] | 1.04 [0.94-1.14] |
| Y00 | 2,670 | 114,197,940 | 0.02 | 0.02 | 0.02 | 0.00 | 0.00 | 0.00 | 1.08 [0.94-1.23] | 1.03 [0.89-1.18] |
| Y04 | 23,200 | 113,652,980 | 0.21 | 0.20 | 0.20 | 0.01 | 0.01 | 0.01 | 1.07 [1.02-1.12]* | 1.03 [0.98-1.08] |
| Y07 | 1,630 | 114,234,220 | 0.01 | 0.01 | 0.02 | 0.00 | 0.00 | 0.00 | 0.92 [0.77-1.09] | 0.88 [0.74-1.05] |
| Y08 | 1,070 | 114,245,730 | 0.01 | 0.01 | 0.01 | 0.00 | 0.00 | 0.00 | 0.92 [0.74-1.15] | 0.90 [0.71-1.12] |
| Y09 | 3,180 | 114,165,910 | 0.03 | 0.03 | 0.03 | 0.00 | 0.00 | 0.00 | 1.09 [0.96-1.23] | 1.05 [0.92-1.19] |
| Y28 | 800 | 114,250,470 | 0.01 | 0.01 | 0.01 | 0.00 | 0.00 | 0.00 | 1.04 [0.81-1.33] | 0.96 [0.75-1.24] |
| Y34 | 700 | 114,247,750 | 0.01 | 0.01 | 0.01 | 0.00 | 0.00 | 0.00 | 1.12 [0.86-1.44] | 1.03 [0.79-1.34] |
| Y40 | 19,800 | 114,043,720 | 0.23 | 0.16 | 0.15 | 0.07 | 0.08 | 0.06 | 1.51 [1.44-1.58]* | 1.39 [1.32-1.45]* |
| Y41 | 4,020 | 114,224,040 | 0.04 | 0.03 | 0.03 | 0.01 | 0.01 | 0.01 | 1.39 [1.26-1.55]* | 1.26 [1.14-1.41]* |
| Y42 | 15,560 | 114,115,160 | 0.17 | 0.13 | 0.12 | 0.05 | 0.05 | 0.04 | 1.45 [1.38-1.53]* | 1.29 [1.22-1.36]* |
| Y43 | 33,250 | 114,029,700 | 0.32 | 0.28 | 0.27 | 0.04 | 0.06 | 0.05 | 1.22 [1.17-1.26]* | 1.18 [1.14-1.23]* |
| Y44 | 10,940 | 114,181,470 | 0.11 | 0.09 | 0.09 | 0.02 | 0.02 | 0.02 | 1.27 [1.18-1.35]* | 1.20 [1.13-1.29]* |
| Y45 | 25,310 | 114,023,280 | 0.27 | 0.21 | 0.19 | 0.05 | 0.07 | 0.06 | 1.36 [1.31-1.42]* | 1.26 [1.21-1.32]* |
| Y46 | 3,660 | 114,227,620 | 0.04 | 0.03 | 0.03 | 0.01 | 0.01 | 0.01 | 1.40 [1.25-1.56]* | 1.28 [1.14-1.43]* |
| Y47 | 1,900 | 114,250,440 | 0.02 | 0.02 | 0.02 | 0.00 | 0.00 | 0.00 | 1.27 [1.09-1.49]* | 1.17 [1.00-1.38] |
| Y48 | 2,200 | 114,242,900 | 0.02 | 0.02 | 0.02 | 0.00 | 0.01 | 0.00 | 1.32 [1.14-1.52]* | 1.20 [1.04-1.39] |
| Y49 | 6,150 | 114,208,920 | 0.06 | 0.05 | 0.05 | 0.01 | 0.02 | 0.01 | 1.35 [1.24-1.47]* | 1.29 [1.18-1.41]* |
| Y51 | 9,440 | 114,185,110 | 0.09 | 0.08 | 0.07 | 0.01 | 0.02 | 0.01 | 1.22 [1.13-1.31]* | 1.18 [1.10-1.27]* |
| Y52 | 22,410 | 114,101,370 | 0.23 | 0.19 | 0.17 | 0.04 | 0.06 | 0.05 | 1.32 [1.26-1.39]* | 1.26 [1.20-1.32]* |
| Y53 | 4,290 | 114,239,850 | 0.05 | 0.04 | 0.03 | 0.01 | 0.01 | 0.01 | 1.38 [1.25-1.53]* | 1.25 [1.13-1.39]* |
| Y54 | 17,710 | 114,167,870 | 0.19 | 0.15 | 0.14 | 0.04 | 0.05 | 0.04 | 1.35 [1.28-1.42]* | 1.27 [1.21-1.34]* |
| Y55 | 2,550 | 114,245,460 | 0.03 | 0.02 | 0.02 | 0.01 | 0.01 | 0.01 | 1.46 [1.28-1.66]* | 1.28 [1.12-1.46]* |
| Y56 | 2,020 | 114,234,130 | 0.03 | 0.02 | 0.02 | 0.01 | 0.01 | 0.01 | 1.80 [1.57-2.06]* | 1.62 [1.41-1.87]* |
| Y57 | 10,280 | 114,181,770 | 0.11 | 0.08 | 0.08 | 0.03 | 0.03 | 0.03 | 1.43 [1.34-1.53]* | 1.31 [1.22-1.40]* |
| Y59 | 1,330 | 114,244,010 | 0.02 | 0.01 | 0.01 | 0.01 | 0.01 | 0.00 | 1.52 [1.28-1.81]* | 1.40 [1.17-1.68]* |
| Y60 | 12,830 | 114,156,750 | 0.13 | 0.11 | 0.10 | 0.02 | 0.02 | 0.02 | 1.24 [1.17-1.32]* | 1.18 [1.11-1.25]* |
| Y65 | 1,810 | 114,249,730 | 0.02 | 0.02 | 0.01 | 0.00 | 0.00 | 0.00 | 1.20 [1.02-1.41] | 1.14 [0.97-1.34] |
| Y71 | 790 | 114,260,210 | 0.01 | 0.01 | 0.01 | 0.00 | 0.00 | 0.00 | 1.59 [1.27-1.99]* | 1.51 [1.20-1.90]* |
| Y73 | 780 | 114,261,840 | 0.01 | 0.01 | 0.01 | 0.00 | 0.00 | 0.00 | 0.97 [0.75-1.25] | 0.93 [0.71-1.21] |
| Y76 | 450 | 114,259,720 | 0.00 | 0.00 | 0.00 | 0.00 | 0.00 | 0.00 | 1.35 [1.00-1.82] | 1.32 [0.97-1.79] |
| Y77 | 840 | 114,262,560 | 0.01 | 0.01 | 0.01 | 0.00 | 0.00 | 0.00 | 1.01 [0.78-1.30] | 0.98 [0.76-1.27] |
| Y79 | 1,820 | 114,241,430 | 0.02 | 0.02 | 0.01 | 0.00 | 0.00 | 0.00 | 1.20 [1.02-1.41] | 1.15 [0.97-1.35] |
| Y82 | 680 | 114,259,610 | 0.01 | 0.01 | 0.01 | 0.00 | 0.00 | 0.00 | 1.31 [1.01-1.68] | 1.24 [0.96-1.61] |
| Y83 | 237,780 | 110,687,950 | 2.48 | 2.07 | 1.94 | 0.42 | 0.54 | 0.43 | 1.28 [1.26-1.30]* | 1.21 [1.19-1.22]* |
| Y84 | 51,560 | 113,838,910 | 0.51 | 0.44 | 0.41 | 0.07 | 0.10 | 0.08 | 1.24 [1.21-1.28]* | 1.19 [1.16-1.23]* |
| Y85 | 5,600 | 114,156,010 | 0.06 | 0.05 | 0.04 | 0.01 | 0.01 | 0.01 | 1.30 [1.19-1.42]* | 1.23 [1.12-1.34]* |
| Y86 | 10,780 | 114,034,430 | 0.10 | 0.09 | 0.09 | 0.01 | 0.01 | 0.01 | 1.16 [1.09-1.24]* | 1.11 [1.03-1.18]* |
| Y87 | 1,770 | 114,230,000 | 0.02 | 0.01 | 0.01 | 0.00 | 0.00 | 0.00 | 1.23 [1.05-1.44]* | 1.15 [0.97-1.35] |
| Y88 | 1,940 | 114,239,820 | 0.02 | 0.02 | 0.02 | 0.00 | 0.00 | 0.00 | 1.17 [1.00-1.37] | 1.07 [0.91-1.26] |
| Y89 | 640 | 114,257,270 | 0.01 | 0.01 | 0.01 | 0.00 | 0.00 | 0.00 | 1.26 [0.96-1.64] | 1.21 [0.92-1.59] |
| Y90 | 1,140 | 114,256,100 | 0.01 | 0.01 | 0.01 | 0.00 | 0.00 | 0.00 | 1.13 [0.93-1.39] | 1.10 [0.90-1.36] |
| Y91 | 2,290 | 114,222,180 | 0.02 | 0.02 | 0.02 | 0.00 | 0.00 | 0.00 | 1.15 [1.00-1.33] | 1.12 [0.96-1.29] |
| Y95 | 117,620 | 113,985,260 | 1.17 | 1.00 | 0.94 | 0.18 | 0.23 | 0.19 | 1.25 [1.22-1.27]* | 1.19 [1.16-1.21]* |
| any age - Z00-Z99 | | | | | | | | | | |
| Z00 | 5,350 | 114,100,500 | 0.05 | 0.05 | 0.04 | 0.00 | 0.01 | 0.01 | 1.19 [1.08-1.30]* | 1.12 [1.02-1.23] |
| Z01 | 31,760 | 112,840,420 | 0.54 | 0.22 | 0.23 | 0.32 | 0.31 | 0.26 | 2.34 [2.27-2.42]* | 1.96 [1.90-2.03]* |
| Z02 | 19,320 | 114,072,870 | 0.18 | 0.17 | 0.15 | 0.01 | 0.02 | 0.02 | 1.16 [1.10-1.22]* | 1.11 [1.05-1.17]* |
| Z03 | 156,740 | 110,281,390 | 1.73 | 1.35 | 1.29 | 0.38 | 0.44 | 0.34 | 1.34 [1.32-1.36]* | 1.25 [1.23-1.27]* |
| Z04 | 29,550 | 113,115,220 | 0.30 | 0.25 | 0.24 | 0.05 | 0.05 | 0.04 | 1.22 [1.17-1.27]* | 1.16 [1.11-1.21]* |
| Z08 | 44,890 | 113,470,140 | 0.39 | 0.40 | 0.36 | −0.01 | 0.03 | 0.02 | 1.08 [1.05-1.12]* | 1.07 [1.03-1.10]* |
| Z09 | 87,880 | 112,560,570 | 0.90 | 0.75 | 0.70 | 0.14 | 0.20 | 0.14 | 1.28 [1.25-1.31]* | 1.19 [1.17-1.22]* |
| Z11 | 272,790 | 113,709,140 | 2.77 | 2.31 | 2.27 | 0.46 | 0.50 | 0.36 | 1.22 [1.20-1.23]* | 1.15 [1.13-1.16]* |
| Z12 | 63,210 | 113,641,280 | 0.57 | 0.55 | 0.50 | 0.02 | 0.08 | 0.06 | 1.16 [1.13-1.19]* | 1.12 [1.09-1.15]* |
| Z13 | 175,920 | 111,670,540 | 1.85 | 1.51 | 1.44 | 0.34 | 0.42 | 0.33 | 1.29 [1.27-1.31]* | 1.22 [1.19-1.24]* |
| Z20 | 9,680 | 114,195,190 | 0.10 | 0.08 | 0.08 | 0.02 | 0.02 | 0.02 | 1.25 [1.17-1.34]* | 1.19 [1.10-1.28]* |
| Z22 | 81,500 | 113,387,010 | 0.90 | 0.68 | 0.64 | 0.22 | 0.25 | 0.22 | 1.40 [1.36-1.43]* | 1.32 [1.29-1.35]* |
| Z24 | 1,760 | 114,153,520 | 0.02 | 0.02 | 0.02 | 0.00 | 0.00 | 0.00 | 1.07 [0.91-1.26] | 1.07 [0.90-1.27] |
| Z25 | 840 | 114,248,140 | 0.02 | 0.01 | 0.01 | 0.01 | 0.01 | 0.01 | 2.82 [2.31-3.44]* | 2.27 [1.84-2.80]* |
| Z27 | 4,890 | 114,095,560 | 0.08 | 0.03 | 0.03 | 0.04 | 0.04 | 0.04 | 2.36 [2.17-2.57]* | 2.10 [1.92-2.29]* |
| Z29 | 45,890 | 113,064,430 | 0.47 | 0.39 | 0.37 | 0.08 | 0.09 | 0.07 | 1.25 [1.21-1.29]* | 1.19 [1.15-1.23]* |
| Z30 | 53,370 | 111,703,510 | 0.50 | 0.47 | 0.43 | 0.02 | 0.07 | 0.05 | 1.16 [1.13-1.20]* | 1.12 [1.09-1.15]* |
| Z31 | 910 | 114,157,150 | 0.01 | 0.01 | 0.01 | 0.00 | 0.00 | 0.00 | 1.02 [0.81-1.28] | 0.97 [0.77-1.23] |
| Z32 | 3,850 | 114,094,760 | 0.04 | 0.03 | 0.03 | 0.00 | 0.01 | 0.01 | 1.23 [1.10-1.37]* | 1.17 [1.05-1.31]* |
| Z33 | 35,520 | 113,139,030 | 0.36 | 0.30 | 0.30 | 0.06 | 0.07 | 0.05 | 1.23 [1.18-1.27]* | 1.14 [1.10-1.19]* |
| Z34 | 97,020 | 111,476,280 | 0.93 | 0.85 | 0.86 | 0.08 | 0.08 | 0.06 | 1.09 [1.07-1.11]* | 1.06 [1.04-1.09]* |
| Z35 | 86,100 | 112,684,920 | 0.80 | 0.75 | 0.75 | 0.05 | 0.05 | 0.03 | 1.07 [1.04-1.09]* | 1.04 [1.02-1.07]* |

| | | | | | | | | | | |
|---|---|---|---|---|---|---|---|---|---|---|
| Z36 | 18,140 | 113,067,420 | 0.17 | 0.16 | 0.15 | 0.01 | 0.02 | 0.02 | 1.12 [1.07-1.18]* | 1.10 [1.04-1.16]* |
| Z37 | 337,140 | 99,896,670 | 3.96 | 3.23 | 3.69 | 0.73 | 0.27 | 0.23 | 1.07 [1.06-1.09]* | 1.06 [1.05-1.08]* |
| Z38 | 1,010 | 88,256,730 | 0.01 | 0.01 | 0.01 | 0.00 | 0.00 | 0.00 | 1.09 [0.88-1.35] | 1.09 [0.88-1.36] |
| Z39 | 29,460 | 113,210,160 | 0.27 | 0.26 | 0.25 | 0.01 | 0.02 | 0.02 | 1.08 [1.04-1.12]* | 1.07 [1.03-1.12]* |
| Z40 | 5,720 | 114,190,700 | 0.05 | 0.05 | 0.05 | 0.00 | 0.01 | 0.00 | 1.12 [1.02-1.22] | 1.10 [1.00-1.20] |
| Z41 | 6,560 | 114,032,850 | 0.07 | 0.05 | 0.05 | 0.02 | 0.02 | 0.02 | 1.34 [1.24-1.45]* | 1.29 [1.19-1.40]* |
| Z42 | 15,180 | 113,966,600 | 0.14 | 0.13 | 0.12 | 0.01 | 0.02 | 0.02 | 1.18 [1.12-1.25]* | 1.16 [1.09-1.22]* |
| Z43 | 22,450 | 113,877,910 | 0.21 | 0.19 | 0.18 | 0.01 | 0.03 | 0.02 | 1.15 [1.10-1.20]* | 1.11 [1.06-1.16]* |
| Z44 | 490 | 114,254,290 | 0.00 | 0.00 | 0.00 | 0.00 | 0.00 | 0.00 | 1.32 [0.98-1.79] | 1.28 [0.93-1.74] |
| Z45 | 46,900 | 113,671,940 | 0.45 | 0.40 | 0.38 | 0.05 | 0.07 | 0.06 | 1.20 [1.16-1.24]* | 1.14 [1.11-1.18]* |
| Z46 | 57,850 | 113,377,840 | 0.53 | 0.51 | 0.46 | 0.02 | 0.07 | 0.05 | 1.14 [1.11-1.17]* | 1.11 [1.08-1.14]* |
| Z47 | 46,750 | 113,063,620 | 0.46 | 0.40 | 0.41 | 0.06 | 0.05 | 0.03 | 1.12 [1.08-1.15]* | 1.07 [1.04-1.11]* |
| Z48 | 17,260 | 113,816,190 | 0.17 | 0.15 | 0.14 | 0.02 | 0.03 | 0.02 | 1.20 [1.14-1.26]* | 1.15 [1.10-1.22]* |
| Z49 | 8,820 | 114,166,210 | 0.09 | 0.07 | 0.07 | 0.02 | 0.03 | 0.03 | 1.44 [1.34-1.54]* | 1.40 [1.31-1.50]* |
| Z50 | 331,390 | 112,224,960 | 3.26 | 2.88 | 2.75 | 0.38 | 0.51 | 0.41 | 1.19 [1.17-1.20]* | 1.14 [1.13-1.16]* |
| Z51 | 394,130 | 111,027,540 | 3.91 | 3.46 | 3.22 | 0.45 | 0.69 | 0.56 | 1.21 [1.20-1.23]* | 1.17 [1.16-1.18]* |
| Z52 | 2,200 | 114,227,270 | 0.02 | 0.02 | 0.02 | 0.00 | 0.00 | 0.00 | 1.00 [0.86-1.17] | 1.01 [0.86-1.18] |
| Z53 | 342,090 | 108,273,730 | 3.73 | 3.02 | 2.84 | 0.71 | 0.89 | 0.70 | 1.31 [1.30-1.33]* | 1.23 [1.22-1.25]* |
| Z54 | 2,890 | 114,195,540 | 0.03 | 0.02 | 0.02 | 0.01 | 0.01 | 0.01 | 1.40 [1.24-1.58]* | 1.36 [1.20-1.54]* |
| Z55 | 1,590 | 114,250,160 | 0.02 | 0.01 | 0.01 | 0.01 | 0.00 | 0.00 | 1.33 [1.13-1.56]* | 1.25 [1.06-1.48]* |
| Z56 | 4,120 | 114,215,600 | 0.04 | 0.04 | 0.03 | 0.00 | 0.01 | 0.00 | 1.17 [1.06-1.31]* | 1.09 [0.98-1.22] |
| Z57 | 11,960 | 114,199,590 | 0.12 | 0.10 | 0.09 | 0.02 | 0.03 | 0.02 | 1.27 [1.20-1.35]* | 1.21 [1.14-1.29]* |
| Z58 | 9,050 | 114,194,770 | 0.10 | 0.07 | 0.07 | 0.02 | 0.02 | 0.02 | 1.32 [1.23-1.41]* | 1.22 [1.14-1.31]* |
| Z59 | 9,280 | 114,159,310 | 0.08 | 0.08 | 0.08 | 0.00 | 0.01 | 0.00 | 1.09 [1.01-1.17] | 1.05 [0.97-1.13] |
| Z60 | 106,020 | 113,087,560 | 1.01 | 0.92 | 0.85 | 0.08 | 0.15 | 0.12 | 1.18 [1.15-1.21]* | 1.14 [1.11-1.16]* |
| Z61 | 4,780 | 114,202,950 | 0.05 | 0.04 | 0.04 | 0.01 | 0.00 | 0.00 | 1.06 [0.96-1.17] | 1.00 [0.90-1.11] |
| Z62 | 1,090 | 114,246,470 | 0.01 | 0.01 | 0.01 | 0.00 | 0.00 | 0.00 | 1.01 [0.82-1.23] | 0.93 [0.76-1.15] |
| Z63 | 35,170 | 113,816,780 | 0.34 | 0.30 | 0.31 | 0.04 | 0.04 | 0.02 | 1.12 [1.08-1.16]* | 1.05 [1.01-1.09] |
| Z64 | 900 | 114,229,890 | 0.01 | 0.01 | 0.01 | 0.00 | 0.00 | 0.00 | 0.99 [0.78-1.25] | 0.94 [0.74-1.20] |
| Z65 | 2,850 | 114,228,600 | 0.03 | 0.02 | 0.03 | 0.00 | 0.00 | 0.00 | 1.09 [0.96-1.24] | 1.04 [0.91-1.18] |
| Z71 | 35,110 | 113,762,180 | 0.45 | 0.28 | 0.27 | 0.17 | 0.18 | 0.15 | 1.65 [1.59-1.70]* | 1.49 [1.44-1.55]* |
| Z72 | 229,660 | 108,372,280 | 2.26 | 2.09 | 1.88 | 0.17 | 0.37 | 0.32 | 1.20 [1.18-1.22]* | 1.17 [1.15-1.18]* |
| Z73 | 54,930 | 114,010,380 | 0.53 | 0.47 | 0.45 | 0.06 | 0.08 | 0.06 | 1.18 [1.14-1.22]* | 1.13 [1.10-1.17]* |
| Z74 | 42,210 | 114,016,830 | 0.40 | 0.36 | 0.34 | 0.03 | 0.06 | 0.05 | 1.18 [1.14-1.22]* | 1.16 [1.12-1.20]* |
| Z75 | 83,140 | 113,864,080 | 0.78 | 0.72 | 0.69 | 0.06 | 0.09 | 0.08 | 1.13 [1.10-1.16]* | 1.11 [1.08-1.14]* |
| Z76 | 6,650 | 113,221,920 | 0.06 | 0.06 | 0.05 | 0.01 | 0.01 | 0.01 | 1.20 [1.10-1.30]* | 1.18 [1.08-1.28]* |
| Z80 | 90,850 | 113,023,020 | 0.85 | 0.79 | 0.73 | 0.06 | 0.12 | 0.09 | 1.17 [1.14-1.20]* | 1.11 [1.09-1.14]* |
| Z81 | 14,740 | 114,014,230 | 0.16 | 0.12 | 0.13 | 0.03 | 0.02 | 0.01 | 1.18 [1.11-1.24]* | 1.11 [1.05-1.17]* |
| Z82 | 194,870 | 110,845,770 | 2.08 | 1.68 | 1.60 | 0.40 | 0.48 | 0.38 | 1.30 [1.28-1.32]* | 1.23 [1.21-1.24]* |
| Z83 | 75,220 | 112,474,520 | 0.79 | 0.64 | 0.64 | 0.16 | 0.15 | 0.12 | 1.23 [1.21-1.27]* | 1.17 [1.14-1.20]* |
| Z84 | 10,920 | 113,982,080 | 0.15 | 0.09 | 0.09 | 0.07 | 0.07 | 0.06 | 1.76 [1.66-1.87]* | 1.69 [1.59-1.80]* |
| Z85 | 321,340 | 110,790,740 | 3.10 | 2.85 | 2.66 | 0.25 | 0.44 | 0.36 | 1.17 [1.15-1.18]* | 1.13 [1.12-1.15]* |
| Z86 | 1,016,710 | 103,940,520 | 11.18 | 9.44 | 8.88 | 1.74 | 2.30 | 1.81 | 1.26 [1.25-1.27]* | 1.19 [1.19-1.20]* |
| Z87 | 537,640 | 107,644,600 | 5.91 | 4.77 | 4.59 | 1.14 | 1.33 | 0.98 | 1.29 [1.28-1.30]* | 1.20 [1.19-1.21]* |
| Z88 | 527,060 | 107,275,920 | 6.33 | 4.57 | 4.32 | 1.75 | 2.01 | 1.56 | 1.47 [1.45-1.48]* | 1.33 [1.31-1.34]* |
| Z89 | 15,080 | 114,138,110 | 0.15 | 0.13 | 0.12 | 0.02 | 0.03 | 0.03 | 1.28 [1.21-1.35]* | 1.23 [1.16-1.30]* |
| Z90 | 278,340 | 111,791,120 | 2.77 | 2.42 | 2.26 | 0.34 | 0.50 | 0.36 | 1.22 [1.21-1.24]* | 1.15 [1.13-1.16]* |
| Z91 | 228,630 | 111,758,600 | 3.12 | 1.79 | 1.78 | 1.33 | 1.34 | 1.09 | 1.76 [1.73-1.78]* | 1.54 [1.52-1.56]* |
| Z92 | 656,400 | 108,686,690 | 6.78 | 5.86 | 5.48 | 0.92 | 1.30 | 1.01 | 1.24 [1.23-1.25]* | 1.17 [1.16-1.18]* |
| Z93 | 65,200 | 113,637,850 | 0.62 | 0.56 | 0.52 | 0.06 | 0.10 | 0.08 | 1.19 [1.16-1.23]* | 1.15 [1.12-1.18]* |
| Z94 | 13,060 | 114,062,060 | 0.14 | 0.11 | 0.10 | 0.03 | 0.04 | 0.03 | 1.36 [1.28-1.44]* | 1.31 [1.23-1.38]* |
| Z95 | 213,710 | 111,781,730 | 2.12 | 1.86 | 1.73 | 0.26 | 0.39 | 0.31 | 1.22 [1.21-1.24]* | 1.17 [1.15-1.19]* |
| Z96 | 415,170 | 110,481,860 | 4.18 | 3.66 | 3.43 | 0.52 | 0.74 | 0.59 | 1.22 [1.20-1.23]* | 1.17 [1.15-1.18]* |
| Z97 | 41,380 | 113,915,180 | 0.42 | 0.35 | 0.33 | 0.07 | 0.09 | 0.07 | 1.26 [1.22-1.31]* | 1.18 [1.14-1.23]* |
| Z98 | 55,760 | 113,621,780 | 0.57 | 0.47 | 0.44 | 0.09 | 0.13 | 0.09 | 1.29 [1.26-1.33]* | 1.18 [1.15-1.22]* |
| Z99 | 68,690 | 113,775,710 | 0.74 | 0.57 | 0.53 | 0.17 | 0.21 | 0.16 | 1.39 [1.36-1.43]* | 1.28 [1.25-1.31]* |
| 18+ - A00-B99 | | | | | | | | | | |
| A02 | 1,240 | 80,137,040 | 0.02 | 0.01 | 0.02 | 0.00 | 0.00 | 0.00 | 1.17 [0.97-1.42] | 1.04 [0.86-1.27] |
| A04 | 49,160 | 79,601,460 | 0.76 | 0.59 | 0.59 | 0.17 | 0.17 | 0.11 | 1.28 [1.24-1.32]* | 1.18 [1.14-1.22]* |
| A05 | 460 | 80,166,580 | 0.01 | 0.01 | 0.01 | 0.00 | 0.00 | 0.00 | 1.31 [0.96-1.80] | 1.23 [0.89-1.70] |
| A07 | 660 | 80,159,900 | 0.01 | 0.01 | 0.01 | 0.00 | 0.00 | 0.00 | 1.19 [0.91-1.56] | 1.04 [0.79-1.38] |
| A08 | 25,100 | 79,670,080 | 0.40 | 0.30 | 0.30 | 0.10 | 0.10 | 0.06 | 1.31 [1.26-1.37]* | 1.17 [1.12-1.22]* |
| A09 | 196,020 | 78,925,500 | 3.18 | 2.33 | 2.41 | 0.85 | 0.78 | 0.46 | 1.32 [1.30-1.34]* | 1.17 [1.15-1.19]* |
| A15 | 1,240 | 80,147,010 | 0.02 | 0.02 | 0.02 | 0.00 | 0.00 | 0.00 | 1.13 [0.93-1.37] | 1.08 [0.88-1.32] |
| A16 | 2,320 | 80,121,820 | 0.04 | 0.03 | 0.03 | 0.01 | 0.01 | 0.01 | 1.41 [1.23-1.61]* | 1.30 [1.14-1.50]* |
| A18 | 1,200 | 80,137,380 | 0.02 | 0.01 | 0.01 | 0.00 | 0.00 | 0.00 | 1.35 [1.11-1.63]* | 1.31 [1.08-1.60]* |
| A31 | 910 | 80,164,630 | 0.02 | 0.01 | 0.01 | 0.01 | 0.01 | 0.00 | 1.63 [1.32-2.01]* | 1.39 [1.12-1.73]* |
| A39 | 580 | 80,128,400 | 0.01 | 0.01 | 0.01 | 0.00 | 0.00 | 0.00 | 1.10 [0.82-1.46] | 1.06 [0.79-1.42] |
| A40 | 9,550 | 80,104,470 | 0.15 | 0.11 | 0.11 | 0.04 | 0.04 | 0.03 | 1.35 [1.26-1.45]* | 1.27 [1.18-1.36]* |
| A41 | 189,650 | 79,399,180 | 2.93 | 2.27 | 2.33 | 0.65 | 0.60 | 0.43 | 1.26 [1.24-1.28]* | 1.17 [1.15-1.19]* |
| A46 | 660 | 80,154,330 | 0.01 | 0.01 | 0.01 | 0.01 | 0.01 | 0.01 | 2.10 [1.67-2.65]* | 1.95 [1.54-2.48]* |
| A48 | 840 | 80,160,380 | 0.01 | 0.01 | 0.01 | 0.00 | 0.00 | 0.00 | 1.26 [1.00-1.59] | 1.20 [0.95-1.52] |
| A49 | 25,660 | 79,924,900 | 0.41 | 0.30 | 0.30 | 0.11 | 0.11 | 0.08 | 1.36 [1.31-1.42]* | 1.25 [1.20-1.31]* |
| A63 | 880 | 80,081,650 | 0.01 | 0.01 | 0.01 | 0.00 | 0.00 | 0.00 | 1.47 [1.19-1.81]* | 1.31 [1.06-1.63] |
| A69 | 620 | 80,164,210 | 0.01 | 0.01 | 0.01 | 0.00 | 0.00 | 0.00 | 1.23 [0.94-1.61] | 1.06 [0.80-1.41] |
| A86 | 1,230 | 80,157,210 | 0.02 | 0.01 | 0.02 | 0.00 | 0.00 | 0.00 | 1.09 [0.89-1.33] | 1.01 [0.82-1.23] |
| A87 | 3,120 | 80,087,040 | 0.05 | 0.04 | 0.04 | 0.02 | 0.01 | 0.01 | 1.40 [1.25-1.58]* | 1.22 [1.08-1.38]* |
| B00 | 5,800 | 80,050,970 | 0.13 | 0.06 | 0.06 | 0.07 | 0.06 | 0.06 | 2.06 [1.91-2.24]* | 1.80 [1.66-1.96]* |
| B01 | 1,110 | 80,096,880 | 0.02 | 0.01 | 0.01 | 0.01 | 0.01 | 0.00 | 1.39 [1.14-1.68]* | 1.27 [1.04-1.55] |
| B02 | 13,430 | 80,050,650 | 0.22 | 0.16 | 0.16 | 0.07 | 0.06 | 0.05 | 1.41 [1.33-1.49]* | 1.29 [1.21-1.37]* |
| B07 | 5,300 | 80,021,600 | 0.09 | 0.06 | 0.06 | 0.03 | 0.02 | 0.02 | 1.40 [1.28-1.53]* | 1.25 [1.14-1.37]* |
| B08 | 490 | 80,145,560 | 0.01 | 0.01 | 0.01 | 0.00 | 0.00 | 0.00 | 1.95 [1.48-2.56]* | 1.70 [1.27-2.27]* |
| B15 | 650 | 80,157,960 | 0.01 | 0.01 | 0.01 | 0.00 | 0.00 | 0.00 | 1.24 [0.95-1.60] | 1.17 [0.89-1.53] |
| B16 | 1,350 | 80,135,350 | 0.02 | 0.02 | 0.02 | 0.01 | 0.01 | 0.01 | 1.39 [1.17-1.66]* | 1.37 [1.14-1.64]* |
| B17 | 2,030 | 80,126,740 | 0.03 | 0.02 | 0.02 | 0.01 | 0.01 | 0.01 | 1.41 [1.22-1.62]* | 1.26 [1.08-1.46]* |
| B18 | 8,690 | 80,015,250 | 0.13 | 0.10 | 0.10 | 0.02 | 0.02 | 0.01 | 1.23 [1.14-1.32]* | 1.13 [1.05-1.22]* |
| B25 | 1,540 | 80,154,650 | 0.03 | 0.02 | 0.02 | 0.01 | 0.01 | 0.01 | 1.40 [1.19-1.65]* | 1.27 [1.07-1.50]* |
| B27 | 3,910 | 80,064,570 | 0.06 | 0.05 | 0.05 | 0.01 | 0.01 | 0.00 | 1.11 [1.00-1.24] | 1.02 [0.91-1.14] |
| B34 | 29,660 | 79,096,870 | 0.48 | 0.35 | 0.36 | 0.12 | 0.12 | 0.06 | 1.33 [1.28-1.38]* | 1.15 [1.11-1.20]* |
| B35 | 5,530 | 80,113,490 | 0.11 | 0.06 | 0.06 | 0.05 | 0.05 | 0.04 | 1.77 [1.62-1.93]* | 1.56 [1.43-1.71]* |
| B36 | 2,070 | 80,158,400 | 0.04 | 0.02 | 0.02 | 0.02 | 0.02 | 0.02 | 1.86 [1.62-2.14]* | 1.66 [1.44-1.92]* |
| B37 | 77,500 | 79,516,330 | 1.27 | 0.91 | 0.93 | 0.36 | 0.34 | 0.21 | 1.37 [1.34-1.40]* | 1.20 [1.17-1.23]* |
| B44 | 2,860 | 80,146,770 | 0.06 | 0.03 | 0.03 | 0.03 | 0.03 | 0.02 | 1.84 [1.64-2.06]* | 1.47 [1.30-1.66]* |
| B49 | 1,700 | 80,160,430 | 0.03 | 0.02 | 0.02 | 0.01 | 0.01 | 0.01 | 1.53 [1.30-1.78]* | 1.40 [1.19-1.65]* |
| B50 | 800 | 80,144,800 | 0.01 | 0.01 | 0.01 | 0.00 | 0.00 | 0.00 | 0.98 [0.76-1.26] | 1.05 [0.81-1.37] |
| B59 | 910 | 80,167,300 | 0.02 | 0.01 | 0.01 | 0.01 | 0.01 | 0.00 | 1.55 [1.26-1.91]* | 1.44 [1.16-1.78]* |
| B80 | 1,230 | 80,140,030 | 0.02 | 0.01 | 0.01 | 0.00 | 0.00 | 0.00 | 1.24 [1.03-1.50] | 1.13 [0.93-1.37] |
| B85 | 560 | 80,161,670 | 0.01 | 0.01 | 0.01 | 0.00 | 0.00 | 0.00 | 1.19 [0.89-1.59] | 1.09 [0.81-1.47] |
| B86 | 1,480 | 80,146,040 | 0.04 | 0.01 | 0.01 | 0.02 | 0.02 | 0.02 | 2.59 [2.22-3.04]* | 2.43 [2.07-2.86]* |
| B90 | 730 | 80,164,240 | 0.01 | 0.01 | 0.01 | 0.00 | 0.00 | 0.00 | 1.43 [1.12-1.82]* | 1.29 [1.00-1.65] |
| B91 | 1,190 | 80,154,460 | 0.02 | 0.01 | 0.01 | 0.00 | 0.00 | 0.00 | 1.29 [1.06-1.57] | 1.27 [1.04-1.56] |
| B94 | 1,580 | 80,153,800 | 0.02 | 0.02 | 0.02 | 0.01 | 0.01 | 0.00 | 1.31 [1.11-1.55]* | 1.14 [0.96-1.36] |
| B95 | 104,630 | 78,835,720 | 1.83 | 1.22 | 1.22 | 0.62 | 0.62 | 0.51 | 1.50 [1.47-1.54]* | 1.39 [1.36-1.42]* |
| B96 | 220,280 | 77,863,530 | 3.44 | 2.70 | 2.78 | 0.75 | 0.66 | 0.43 | 1.24 [1.22-1.26]* | 1.14 [1.13-1.16]* |
| B97 | 42,520 | 79,817,470 | 0.68 | 0.50 | 0.53 | 0.18 | 0.15 | 0.09 | 1.29 [1.25-1.34]* | 1.15 [1.11-1.19]* |
| B98 | 22,620 | 80,013,200 | 0.34 | 0.27 | 0.28 | 0.07 | 0.06 | 0.04 | 1.22 [1.16-1.27]* | 1.13 [1.08-1.18]* |
| B99 | 6,210 | 80,145,070 | 0.09 | 0.07 | 0.07 | 0.02 | 0.02 | 0.01 | 1.26 [1.16-1.38]* | 1.17 [1.07-1.28]* |

| | | | | | | | | | | |
|---|---|---|---|---|---|---|---|---|---|---|
| C00 | 790 | 80,162,340 | 0.01 | 0.01 | 0.01 | 0.00 | 0.00 | 0.00 | 1.08 [0.84-1.40] | 1.05 [0.81-1.36] |
| C01 | 1,610 | 80,161,150 | 0.02 | 0.02 | 0.02 | 0.00 | 0.00 | 0.00 | 1.09 [0.92-1.30] | 1.05 [0.88-1.26] |
| C02 | 2,540 | 80,146,300 | 0.04 | 0.03 | 0.03 | 0.01 | 0.01 | 0.01 | 1.31 [1.15-1.50]* | 1.24 [1.08-1.42]* |
| C03 | 590 | 80,168,690 | 0.01 | 0.01 | 0.01 | 0.00 | 0.00 | 0.00 | 1.38 [1.06-1.81] | 1.38 [1.05-1.82] |
| C04 | 780 | 80,163,560 | 0.01 | 0.01 | 0.01 | 0.00 | 0.00 | 0.00 | 1.15 [0.90-1.48] | 1.08 [0.84-1.39] |
| C05 | 810 | 80,164,400 | 0.01 | 0.01 | 0.01 | 0.00 | 0.00 | 0.00 | 1.32 [1.05-1.67] | 1.29 [1.01-1.63] |
| C06 | 1,160 | 80,162,860 | 0.02 | 0.01 | 0.01 | 0.00 | 0.00 | 0.00 | 1.24 [1.02-1.52] | 1.18 [0.96-1.44] |
| C07 | 1,160 | 80,159,990 | 0.01 | 0.01 | 0.01 | 0.00 | 0.00 | 0.00 | 1.05 [0.85-1.29] | 1.00 [0.80-1.24] |
| C09 | 2,070 | 80,152,020 | 0.03 | 0.03 | 0.02 | 0.00 | 0.00 | 0.00 | 1.12 [0.97-1.31] | 1.06 [0.91-1.24] |
| C10 | 1,060 | 80,166,870 | 0.01 | 0.01 | 0.01 | 0.00 | 0.00 | 0.00 | 1.18 [0.95-1.45] | 1.10 [0.88-1.36] |
| C11 | 530 | 80,167,510 | 0.01 | 0.01 | 0.01 | 0.00 | 0.00 | 0.00 | 1.20 [0.90-1.60] | 1.17 [0.87-1.56] |
| C12 | 570 | 80,169,190 | 0.01 | 0.01 | 0.01 | 0.00 | 0.00 | 0.00 | 1.29 [0.98-1.70] | 1.24 [0.93-1.65] |
| C13 | 630 | 80,170,290 | 0.01 | 0.01 | 0.01 | 0.00 | 0.00 | 0.00 | 1.25 [0.95-1.63] | 1.24 [0.94-1.63] |
| C14 | 1,180 | 80,166,460 | 0.02 | 0.01 | 0.01 | 0.00 | 0.00 | 0.00 | 1.12 [0.91-1.36] | 1.06 [0.87-1.31] |
| C15 | 16,370 | 80,115,170 | 0.23 | 0.20 | 0.20 | 0.03 | 0.03 | 0.02 | 1.15 [1.09-1.22]* | 1.12 [1.06-1.18]* |
| C16 | 13,260 | 80,117,230 | 0.18 | 0.16 | 0.17 | 0.01 | 0.01 | 0.01 | 1.07 [1.00-1.13] | 1.04 [0.98-1.11] |
| C17 | 2,700 | 80,157,710 | 0.04 | 0.03 | 0.03 | 0.00 | 0.00 | 0.00 | 1.15 [1.01-1.32] | 1.09 [0.95-1.25] |
| C18 | 45,800 | 79,778,300 | 0.58 | 0.57 | 0.58 | 0.01 | 0.01 | 0.00 | 1.01 [0.98-1.05] | 1.00 [0.97-1.03] |
| C19 | 9,430 | 80,099,230 | 0.11 | 0.12 | 0.12 | 0.00 | 0.00 | 0.00 | 0.96 [0.89-1.04] | 0.96 [0.89-1.04] |
| C20 | 19,540 | 79,971,740 | 0.24 | 0.25 | 0.25 | −0.01 | −0.01 | −0.01 | 0.97 [0.92-1.02] | 0.98 [0.93-1.03] |
| C21 | 2,390 | 80,148,280 | 0.03 | 0.03 | 0.03 | 0.00 | 0.00 | 0.00 | 1.08 [0.94-1.25] | 1.05 [0.91-1.21] |
| C22 | 9,020 | 80,151,390 | 0.14 | 0.11 | 0.11 | 0.03 | 0.03 | 0.02 | 1.25 [1.16-1.34]* | 1.18 [1.10-1.27]* |
| C23 | 1,320 | 80,170,360 | 0.02 | 0.02 | 0.02 | 0.00 | 0.00 | 0.00 | 1.06 [0.87-1.29] | 1.04 [0.85-1.26] |
| C24 | 2,450 | 80,164,240 | 0.03 | 0.03 | 0.03 | 0.00 | 0.00 | 0.00 | 1.07 [0.93-1.24] | 1.04 [0.90-1.20] |
| C25 | 15,100 | 80,146,000 | 0.21 | 0.18 | 0.19 | 0.03 | 0.03 | 0.02 | 1.14 [1.08-1.21]* | 1.10 [1.04-1.17]* |
| C26 | 3,400 | 80,158,470 | 0.05 | 0.04 | 0.04 | 0.01 | 0.00 | 0.00 | 1.10 [0.97-1.24] | 1.05 [0.93-1.19] |
| C30 | 540 | 80,167,150 | 0.01 | 0.01 | 0.01 | 0.00 | 0.00 | 0.00 | 1.12 [0.84-1.51] | 1.09 [0.81-1.46] |
| C32 | 3,480 | 80,133,450 | 0.05 | 0.04 | 0.04 | 0.01 | 0.01 | 0.01 | 1.17 [1.04-1.32]* | 1.14 [1.01-1.28] |
| C34 | 65,420 | 79,991,990 | 0.92 | 0.80 | 0.80 | 0.13 | 0.12 | 0.07 | 1.15 [1.12-1.18]* | 1.09 [1.06-1.12]* |
| C38 | 910 | 80,169,620 | 0.01 | 0.01 | 0.01 | 0.00 | 0.00 | 0.00 | 0.97 [0.76-1.22] | 0.95 [0.74-1.20] |
| C41 | 1,710 | 80,158,210 | 0.02 | 0.02 | 0.02 | 0.00 | 0.00 | 0.00 | 1.23 [1.04-1.45] | 1.19 [1.00-1.40] |
| C43 | 18,260 | 79,957,940 | 0.22 | 0.23 | 0.23 | −0.01 | −0.01 | −0.01 | 0.97 [0.92-1.02] | 0.95 [0.90-1.01] |
| C44 | 128,910 | 78,616,050 | 1.80 | 1.61 | 1.62 | 0.19 | 0.17 | 0.11 | 1.11 [1.08-1.13]* | 1.07 [1.04-1.09]* |
| C45 | 4,100 | 80,164,600 | 0.05 | 0.05 | 0.05 | 0.00 | 0.00 | 0.00 | 1.00 [0.89-1.12] | 1.01 [0.90-1.13] |
| C48 | 2,250 | 80,163,070 | 0.03 | 0.03 | 0.03 | 0.00 | 0.00 | 0.00 | 1.07 [0.93-1.24] | 1.07 [0.92-1.24] |
| C49 | 3,900 | 80,134,780 | 0.05 | 0.05 | 0.05 | 0.01 | 0.01 | 0.01 | 1.13 [1.01-1.26] | 1.10 [0.99-1.24] |
| C50 | 77,330 | 78,953,740 | 1.02 | 0.97 | 0.99 | 0.05 | 0.04 | 0.03 | 1.04 [1.01-1.06]* | 1.03 [1.00-1.05] |
| C51 | 2,070 | 80,149,550 | 0.03 | 0.02 | 0.02 | 0.01 | 0.01 | 0.01 | 1.24 [1.07-1.44]* | 1.22 [1.05-1.42]* |
| C52 | 660 | 80,168,560 | 0.01 | 0.01 | 0.01 | 0.00 | 0.00 | 0.00 | 0.90 [0.68-1.19] | 0.90 [0.68-1.19] |
| C53 | 4,300 | 80,108,340 | 0.05 | 0.05 | 0.06 | 0.00 | 0.00 | 0.00 | 0.96 [0.86-1.07] | 0.97 [0.87-1.08] |
| C54 | 12,830 | 80,010,230 | 0.17 | 0.16 | 0.16 | 0.01 | 0.01 | 0.01 | 1.04 [0.98-1.11] | 1.06 [1.00-1.13] |
| C55 | 1,790 | 80,158,940 | 0.02 | 0.02 | 0.02 | 0.00 | 0.00 | 0.00 | 1.02 [0.86-1.20] | 1.01 [0.85-1.19] |
| C56 | 11,460 | 80,073,480 | 0.15 | 0.14 | 0.14 | 0.00 | 0.00 | 0.00 | 1.01 [0.95-1.08] | 1.02 [0.95-1.09] |
| C57 | 1,270 | 80,166,510 | 0.02 | 0.02 | 0.02 | 0.00 | 0.00 | 0.00 | 1.19 [0.99-1.44] | 1.22 [1.00-1.47] |
| C60 | 930 | 80,160,630 | 0.01 | 0.01 | 0.01 | 0.00 | 0.00 | 0.00 | 1.30 [1.04-1.62] | 1.29 [1.03-1.62] |
| C61 | 72,820 | 79,495,220 | 0.97 | 0.90 | 0.92 | 0.07 | 0.05 | 0.03 | 1.06 [1.03-1.09]* | 1.03 [1.00-1.06] |
| C62 | 1,940 | 80,125,010 | 0.02 | 0.02 | 0.02 | 0.00 | 0.00 | 0.00 | 1.02 [0.87-1.20] | 0.99 [0.84-1.17] |
| C64 | 14,940 | 80,054,640 | 0.20 | 0.18 | 0.18 | 0.02 | 0.02 | 0.01 | 1.10 [1.04-1.17]* | 1.05 [0.99-1.11] |
| C65 | 1,350 | 80,162,050 | 0.02 | 0.02 | 0.02 | 0.00 | 0.00 | 0.00 | 1.22 [1.01-1.47] | 1.18 [0.98-1.43] |
| C66 | 1,880 | 80,159,820 | 0.03 | 0.02 | 0.02 | 0.00 | 0.00 | 0.00 | 1.09 [0.92-1.28] | 1.05 [0.89-1.24] |
| C67 | 33,640 | 79,771,300 | 0.45 | 0.41 | 0.42 | 0.04 | 0.04 | 0.03 | 1.09 [1.05-1.13]* | 1.06 [1.02-1.10]* |
| C68 | 750 | 80,167,240 | 0.01 | 0.01 | 0.01 | 0.00 | 0.00 | 0.00 | 1.05 [0.81-1.37] | 1.04 [0.80-1.36] |
| C69 | 1,350 | 80,152,360 | 0.02 | 0.02 | 0.02 | 0.00 | 0.00 | 0.00 | 0.97 [0.80-1.17] | 0.95 [0.78-1.16] |
| C71 | 7,480 | 80,126,100 | 0.10 | 0.09 | 0.09 | 0.01 | 0.01 | 0.01 | 1.15 [1.06-1.24]* | 1.12 [1.03-1.21]* |
| C73 | 4,260 | 80,108,450 | 0.06 | 0.05 | 0.05 | 0.00 | 0.00 | 0.00 | 1.06 [0.95-1.18] | 1.02 [0.92-1.14] |
| C74 | 510 | 80,167,620 | 0.01 | 0.01 | 0.01 | 0.00 | 0.00 | 0.00 | 1.13 [0.83-1.52] | 1.06 [0.77-1.44] |
| C75 | 680 | 80,165,830 | 0.01 | 0.01 | 0.01 | 0.00 | 0.00 | 0.00 | 1.00 [0.76-1.32] | 0.96 [0.73-1.27] |
| C76 | 2,220 | 80,156,940 | 0.03 | 0.03 | 0.03 | 0.00 | 0.00 | 0.00 | 1.11 [0.95-1.28] | 1.09 [0.94-1.27] |
| C77 | 75,680 | 79,632,510 | 1.00 | 0.94 | 0.95 | 0.06 | 0.05 | 0.04 | 1.05 [1.03-1.08]* | 1.04 [1.01-1.07]* |
| C78 | 111,800 | 79,951,960 | 1.47 | 1.38 | 1.40 | 0.09 | 0.08 | 0.05 | 1.05 [1.03-1.08]* | 1.04 [1.01-1.06]* |
| C79 | 93,930 | 79,978,990 | 1.25 | 1.16 | 1.17 | 0.10 | 0.09 | 0.06 | 1.07 [1.05-1.10]* | 1.05 [1.03-1.08]* |
| C80 | 33,240 | 80,089,150 | 0.45 | 0.41 | 0.41 | 0.04 | 0.04 | 0.02 | 1.09 [1.05-1.13]* | 1.06 [1.02-1.10]* |
| C81 | 3,220 | 80,113,110 | 0.06 | 0.04 | 0.04 | 0.03 | 0.03 | 0.02 | 1.72 [1.54-1.91]* | 1.65 [1.48-1.85]* |
| C82 | 4,000 | 80,123,180 | 0.06 | 0.05 | 0.05 | 0.01 | 0.01 | 0.00 | 1.10 [0.99-1.23] | 1.04 [0.93-1.16] |
| C83 | 11,180 | 80,086,150 | 0.17 | 0.13 | 0.13 | 0.03 | 0.03 | 0.03 | 1.26 [1.18-1.34]* | 1.22 [1.14-1.30]* |
| C84 | 1,490 | 80,159,180 | 0.04 | 0.01 | 0.01 | 0.02 | 0.02 | 0.02 | 2.71 [2.33-3.15]* | 2.64 [2.26-3.08]* |
| C85 | 14,210 | 80,014,610 | 0.22 | 0.17 | 0.17 | 0.05 | 0.05 | 0.04 | 1.31 [1.24-1.39]* | 1.26 [1.19-1.33]* |
| C88 | 1,900 | 80,159,950 | 0.03 | 0.02 | 0.02 | 0.01 | 0.01 | 0.00 | 1.23 [1.06-1.44]* | 1.21 [1.03-1.42] |
| C90 | 9,990 | 80,107,150 | 0.14 | 0.12 | 0.12 | 0.02 | 0.02 | 0.02 | 1.15 [1.07-1.23]* | 1.13 [1.05-1.21]* |
| C91 | 9,460 | 80,077,760 | 0.13 | 0.12 | 0.11 | 0.01 | 0.01 | 0.01 | 1.12 [1.04-1.21]* | 1.08 [1.01-1.17] |
| C92 | 7,770 | 80,126,140 | 0.12 | 0.09 | 0.09 | 0.03 | 0.03 | 0.02 | 1.29 [1.20-1.39]* | 1.26 [1.16-1.36]* |
| C93 | 1,050 | 80,170,360 | 0.02 | 0.01 | 0.01 | 0.00 | 0.00 | 0.00 | 1.18 [0.95-1.47] | 1.14 [0.91-1.42] |
| C94 | 460 | 80,171,030 | 0.01 | 0.01 | 0.01 | 0.00 | 0.00 | 0.00 | 1.31 [0.96-1.80] | 1.27 [0.92-1.74] |
| C95 | 1,540 | 80,162,350 | 0.02 | 0.02 | 0.02 | 0.01 | 0.01 | 0.01 | 1.34 [1.13-1.60]* | 1.27 [1.06-1.52]* |
| C97 | 8,460 | 80,147,380 | 0.11 | 0.10 | 0.10 | 0.01 | 0.01 | 0.00 | 1.08 [1.00-1.17] | 1.04 [0.96-1.13] |
| D00 | 960 | 80,163,610 | 0.01 | 0.01 | 0.01 | 0.00 | 0.00 | 0.00 | 1.05 [0.84-1.32] | 0.99 [0.79-1.24] |
| D01 | 950 | 80,161,630 | 0.01 | 0.01 | 0.01 | 0.00 | 0.00 | 0.00 | 1.11 [0.89-1.40] | 1.02 [0.81-1.29] |
| D02 | 970 | 80,163,240 | 0.01 | 0.01 | 0.01 | 0.00 | 0.00 | 0.00 | 1.20 [0.97-1.49] | 1.13 [0.91-1.41] |
| D03 | 6,900 | 80,086,960 | 0.09 | 0.09 | 0.08 | 0.01 | 0.01 | 0.00 | 1.08 [0.99-1.18] | 1.04 [0.95-1.13] |
| D04 | 14,350 | 80,031,760 | 0.22 | 0.17 | 0.17 | 0.05 | 0.05 | 0.03 | 1.26 [1.19-1.34]* | 1.18 [1.11-1.25]* |
| D05 | 12,600 | 79,943,710 | 0.16 | 0.16 | 0.16 | 0.00 | 0.00 | 0.00 | 1.01 [0.95-1.07] | 1.00 [0.93-1.06] |
| D06 | 10,880 | 79,802,200 | 0.15 | 0.13 | 0.14 | 0.01 | 0.01 | 0.00 | 1.06 [0.99-1.13] | 1.00 [0.93-1.07] |
| D07 | 3,830 | 80,108,230 | 0.06 | 0.05 | 0.04 | 0.01 | 0.01 | 0.01 | 1.28 [1.15-1.42]* | 1.20 [1.07-1.34]* |
| D09 | 3,570 | 80,136,310 | 0.05 | 0.04 | 0.04 | 0.01 | 0.01 | 0.00 | 1.13 [1.00-1.27] | 1.08 [0.96-1.21] |
| D10 | 3,550 | 80,087,610 | 0.06 | 0.04 | 0.04 | 0.01 | 0.01 | 0.01 | 1.33 [1.20-1.49]* | 1.19 [1.06-1.33]* |
| D11 | 4,320 | 80,089,110 | 0.06 | 0.05 | 0.05 | 0.00 | 0.00 | 0.00 | 1.04 [0.94-1.16] | 1.00 [0.90-1.12] |
| D12 | 123,630 | 78,593,830 | 1.84 | 1.52 | 1.52 | 0.32 | 0.32 | 0.18 | 1.21 [1.19-1.24]* | 1.11 [1.09-1.13]* |
| D13 | 8,450 | 79,961,050 | 0.12 | 0.10 | 0.10 | 0.02 | 0.02 | 0.01 | 1.21 [1.12-1.30]* | 1.09 [1.01-1.17] |
| D14 | 2,610 | 80,117,590 | 0.04 | 0.03 | 0.03 | 0.01 | 0.01 | 0.01 | 1.30 [1.14-1.48]* | 1.16 [1.02-1.33] |
| D15 | 1,000 | 80,161,660 | 0.01 | 0.01 | 0.01 | 0.00 | 0.00 | 0.00 | 1.15 [0.92-1.43] | 1.11 [0.89-1.38] |
| D16 | 2,990 | 80,084,290 | 0.05 | 0.04 | 0.04 | 0.01 | 0.01 | 0.01 | 1.27 [1.13-1.43]* | 1.18 [1.04-1.33] |
| D17 | 38,020 | 79,151,530 | 0.54 | 0.47 | 0.46 | 0.07 | 0.07 | 0.04 | 1.16 [1.12-1.20]* | 1.08 [1.05-1.12]* |
| D18 | 15,290 | 79,904,850 | 0.22 | 0.19 | 0.19 | 0.03 | 0.03 | 0.01 | 1.16 [1.10-1.23]* | 1.07 [1.01-1.13] |
| D21 | 3,990 | 80,086,100 | 0.06 | 0.05 | 0.05 | 0.01 | 0.02 | 0.01 | 1.34 [1.21-1.49]* | 1.24 [1.11-1.38]* |
| D22 | 33,250 | 79,176,850 | 0.47 | 0.41 | 0.41 | 0.06 | 0.06 | 0.04 | 1.15 [1.11-1.20]* | 1.09 [1.05-1.13]* |
| D23 | 21,150 | 79,455,400 | 0.32 | 0.26 | 0.25 | 0.06 | 0.06 | 0.05 | 1.26 [1.20-1.32]* | 1.17 [1.12-1.23]* |
| D24 | 13,030 | 79,826,070 | 0.17 | 0.16 | 0.17 | 0.01 | 0.01 | 0.00 | 1.04 [0.98-1.11] | 0.99 [0.93-1.06] |
| D25 | 64,730 | 78,603,730 | 0.90 | 0.81 | 0.83 | 0.09 | 0.07 | 0.04 | 1.08 [1.05-1.11]* | 1.05 [1.02-1.08]* |
| D26 | 2,190 | 80,112,140 | 0.03 | 0.03 | 0.03 | 0.00 | 0.00 | 0.00 | 1.10 [0.95-1.27] | 1.08 [0.93-1.25] |
| D27 | 15,010 | 79,883,220 | 0.20 | 0.19 | 0.19 | 0.01 | 0.01 | 0.00 | 1.06 [1.00-1.12] | 1.01 [0.96-1.07] |
| D28 | 1,560 | 80,133,260 | 0.02 | 0.02 | 0.02 | 0.00 | 0.00 | 0.00 | 1.26 [1.06-1.49]* | 1.19 [1.00-1.41] |
| D29 | 1,170 | 80,151,300 | 0.02 | 0.01 | 0.01 | 0.00 | 0.00 | 0.00 | 1.21 [0.99-1.49] | 1.16 [0.95-1.43] |
| D30 | 2,350 | 80,142,180 | 0.03 | 0.03 | 0.03 | 0.00 | 0.00 | 0.00 | 1.08 [0.94-1.25] | 1.02 [0.88-1.18] |
| D31 | 2,120 | 80,140,740 | 0.03 | 0.03 | 0.02 | 0.01 | 0.01 | 0.01 | 1.30 [1.12-1.50]* | 1.23 [1.06-1.43]* |

| | | | | | | | | | | |
|---|---|---|---|---|---|---|---|---|---|---|
| D32 | 8,640 | 80,093,100 | 0.12 | 0.11 | 0.11 | 0.01 | 0.01 | 0.00 | 1.09 [1.01-1.18] | 1.03 [0.95-1.11] |
| D33 | 3,180 | 80,126,400 | 0.04 | 0.04 | 0.04 | 0.00 | 0.00 | 0.00 | 1.08 [0.96-1.22] | 0.98 [0.87-1.11] |
| D34 | 2,270 | 80,128,250 | 0.03 | 0.03 | 0.03 | 0.00 | 0.00 | 0.00 | 1.04 [0.90-1.21] | 0.98 [0.84-1.14] |
| D35 | 12,550 | 80,020,830 | 0.18 | 0.15 | 0.16 | 0.02 | 0.02 | 0.01 | 1.13 [1.06-1.20]* | 1.05 [0.99-1.12] |
| D36 | 5,530 | 80,044,060 | 0.08 | 0.07 | 0.07 | 0.01 | 0.01 | 0.01 | 1.23 [1.12-1.34]* | 1.13 [1.03-1.24] |
| D37 | 16,780 | 80,001,960 | 0.23 | 0.21 | 0.21 | 0.02 | 0.02 | 0.01 | 1.10 [1.05-1.16]* | 1.06 [1.00-1.12] |
| D38 | 3,970 | 80,143,440 | 0.06 | 0.05 | 0.05 | 0.01 | 0.01 | 0.01 | 1.21 [1.09-1.35]* | 1.15 [1.03-1.28] |
| D39 | 2,360 | 80,130,930 | 0.03 | 0.03 | 0.03 | 0.00 | 0.00 | 0.00 | 1.02 [0.88-1.18] | 1.02 [0.88-1.18] |
| D40 | 790 | 80,157,400 | 0.01 | 0.01 | 0.01 | 0.00 | 0.00 | 0.00 | 1.13 [0.88-1.45] | 1.11 [0.86-1.43] |
| D41 | 11,780 | 80,037,680 | 0.16 | 0.14 | 0.15 | 0.02 | 0.01 | 0.01 | 1.09 [1.02-1.17]* | 1.06 [0.99-1.13] |
| D43 | 3,580 | 80,141,700 | 0.05 | 0.04 | 0.04 | 0.01 | 0.01 | 0.00 | 1.11 [0.99-1.25] | 1.07 [0.95-1.21] |
| D44 | 2,300 | 80,135,570 | 0.03 | 0.03 | 0.03 | 0.00 | 0.00 | 0.00 | 1.16 [1.01-1.34] | 1.07 [0.92-1.24] |
| D45 | 4,400 | 80,102,540 | 0.06 | 0.05 | 0.05 | 0.01 | 0.01 | 0.00 | 1.13 [1.01-1.25] | 1.06 [0.95-1.18] |
| D46 | 10,110 | 80,122,030 | 0.16 | 0.12 | 0.12 | 0.04 | 0.04 | 0.04 | 1.34 [1.26-1.44]* | 1.28 [1.19-1.37]* |
| D47 | 16,380 | 80,076,130 | 0.26 | 0.19 | 0.20 | 0.06 | 0.05 | 0.04 | 1.27 [1.20-1.34]* | 1.18 [1.11-1.24]* |
| D48 | 8,020 | 80,024,330 | 0.11 | 0.10 | 0.10 | 0.01 | 0.01 | 0.01 | 1.12 [1.03-1.21]* | 1.06 [0.98-1.15] |
| 18+ - D50-D89 | | | | | | | | | | |
| D50 | 223,670 | 78,080,690 | 3.67 | 2.69 | 2.78 | 0.98 | 0.89 | 0.62 | 1.32 [1.30-1.34]* | 1.20 [1.18-1.22]* |
| D51 | 20,560 | 79,963,700 | 0.33 | 0.24 | 0.24 | 0.09 | 0.09 | 0.06 | 1.35 [1.29-1.42]* | 1.23 [1.17-1.29]* |
| D52 | 15,390 | 80,097,780 | 0.24 | 0.18 | 0.18 | 0.06 | 0.06 | 0.04 | 1.31 [1.24-1.38]* | 1.22 [1.15-1.29]* |
| D53 | 4,040 | 80,145,140 | 0.07 | 0.05 | 0.05 | 0.02 | 0.02 | 0.02 | 1.43 [1.28-1.59]* | 1.30 [1.16-1.45]* |
| D55 | 430 | 80,166,800 | 0.01 | 0.00 | 0.00 | 0.00 | 0.00 | 0.00 | 1.61 [1.18-2.18]* | 1.50 [1.09-2.06] |
| D56 | 7,920 | 80,035,810 | 0.13 | 0.09 | 0.09 | 0.04 | 0.04 | 0.04 | 1.42 [1.32-1.53]* | 1.37 [1.27-1.48]* |
| D57 | 7,490 | 79,996,600 | 0.12 | 0.09 | 0.09 | 0.04 | 0.03 | 0.03 | 1.37 [1.27-1.47]* | 1.32 [1.22-1.43]* |
| D58 | 2,020 | 80,142,250 | 0.03 | 0.02 | 0.02 | 0.01 | 0.01 | 0.00 | 1.24 [1.07-1.44]* | 1.19 [1.02-1.39] |
| D59 | 2,600 | 80,141,520 | 0.05 | 0.03 | 0.03 | 0.02 | 0.01 | 0.01 | 1.47 [1.30-1.67]* | 1.43 [1.25-1.63]* |
| D61 | 13,840 | 80,103,750 | 0.22 | 0.16 | 0.16 | 0.06 | 0.05 | 0.04 | 1.33 [1.25-1.41]* | 1.26 [1.19-1.34]* |
| D62 | 6,890 | 80,048,390 | 0.10 | 0.08 | 0.09 | 0.02 | 0.01 | 0.01 | 1.13 [1.04-1.23]* | 1.10 [1.01-1.20] |
| D63 | 21,970 | 80,047,120 | 0.34 | 0.26 | 0.26 | 0.08 | 0.07 | 0.06 | 1.29 [1.23-1.35]* | 1.21 [1.15-1.27]* |
| D64 | 302,270 | 77,299,520 | 4.93 | 3.69 | 3.79 | 1.24 | 1.14 | 0.85 | 1.30 [1.29-1.32]* | 1.21 [1.19-1.22]* |
| D65 | 900 | 80,168,020 | 0.01 | 0.01 | 0.01 | 0.00 | 0.00 | 0.00 | 1.25 [1.00-1.57] | 1.21 [0.96-1.51] |
| D66 | 880 | 80,152,510 | 0.01 | 0.01 | 0.01 | 0.00 | 0.00 | 0.00 | 1.13 [0.90-1.43] | 1.03 [0.81-1.31] |
| D68 | 16,230 | 79,935,350 | 0.25 | 0.19 | 0.19 | 0.06 | 0.06 | 0.04 | 1.32 [1.25-1.39]* | 1.18 [1.11-1.24]* |
| D69 | 43,460 | 79,771,080 | 0.67 | 0.52 | 0.52 | 0.15 | 0.14 | 0.11 | 1.27 [1.23-1.31]* | 1.20 [1.16-1.24]* |
| D70 | 41,210 | 79,850,660 | 0.62 | 0.49 | 0.50 | 0.12 | 0.12 | 0.10 | 1.24 [1.20-1.28]* | 1.19 [1.15-1.23]* |
| D72 | 9,080 | 80,104,200 | 0.15 | 0.10 | 0.11 | 0.05 | 0.05 | 0.03 | 1.44 [1.35-1.55]* | 1.27 [1.18-1.37]* |
| D73 | 9,910 | 80,083,030 | 0.15 | 0.12 | 0.12 | 0.03 | 0.03 | 0.02 | 1.24 [1.16-1.32]* | 1.15 [1.07-1.23]* |
| D75 | 12,550 | 80,046,470 | 0.19 | 0.15 | 0.15 | 0.04 | 0.03 | 0.02 | 1.22 [1.15-1.30]* | 1.12 [1.05-1.19]* |
| D76 | 640 | 80,157,350 | 0.01 | 0.01 | 0.01 | 0.00 | 0.00 | 0.00 | 1.37 [1.06-1.78] | 1.26 [0.96-1.65] |
| D80 | 2,340 | 80,139,550 | 0.04 | 0.03 | 0.03 | 0.02 | 0.02 | 0.01 | 1.56 [1.36-1.77]* | 1.32 [1.15-1.52]* |
| D82 | 490 | 80,163,810 | 0.01 | 0.01 | 0.01 | 0.00 | 0.00 | 0.00 | 1.51 [1.12-2.02]* | 1.33 [0.98-1.80] |
| D83 | 330 | 80,167,000 | 0.01 | 0.00 | 0.00 | 0.00 | 0.00 | 0.00 | 2.05 [1.48-2.85]* | 1.75 [1.24-2.46]* |
| D84 | 1,100 | 80,156,070 | 0.02 | 0.01 | 0.01 | 0.01 | 0.01 | 0.01 | 1.70 [1.41-2.05]* | 1.35 [1.10-1.65]* |
| D86 | 6,290 | 80,076,740 | 0.11 | 0.07 | 0.07 | 0.04 | 0.04 | 0.03 | 1.56 [1.44-1.69]* | 1.41 [1.30-1.54]* |
| D89 | 4,940 | 80,127,480 | 0.08 | 0.06 | 0.06 | 0.03 | 0.02 | 0.02 | 1.42 [1.29-1.56]* | 1.29 [1.17-1.42]* |
| 18+ - E00-E89 | | | | | | | | | | |
| E02 | 1,850 | 80,164,040 | 0.03 | 0.02 | 0.02 | 0.01 | 0.00 | 0.00 | 1.20 [1.02-1.41] | 1.16 [0.98-1.37] |
| E03 | 220,270 | 77,442,500 | 3.63 | 2.67 | 2.72 | 0.96 | 0.91 | 0.71 | 1.34 [1.32-1.36]* | 1.24 [1.23-1.26]* |
| E04 | 22,870 | 79,854,010 | 0.32 | 0.28 | 0.29 | 0.04 | 0.04 | 0.02 | 1.13 [1.08-1.18]* | 1.05 [1.01-1.10] |
| E05 | 36,880 | 79,718,180 | 0.58 | 0.44 | 0.45 | 0.14 | 0.13 | 0.09 | 1.29 [1.24-1.33]* | 1.18 [1.14-1.22]* |
| E06 | 3,960 | 80,115,030 | 0.06 | 0.05 | 0.05 | 0.01 | 0.01 | 0.01 | 1.22 [1.10-1.36]* | 1.13 [1.01-1.26] |
| E07 | 6,230 | 80,099,420 | 0.10 | 0.07 | 0.07 | 0.02 | 0.02 | 0.01 | 1.29 [1.19-1.41]* | 1.17 [1.08-1.28]* |
| E10 | 39,890 | 79,193,410 | 0.61 | 0.48 | 0.47 | 0.13 | 0.14 | 0.07 | 1.30 [1.25-1.34]* | 1.12 [1.09-1.16]* |
| E11 | 343,850 | 75,646,640 | 5.54 | 4.33 | 4.41 | 1.21 | 1.14 | 0.61 | 1.26 [1.24-1.27]* | 1.12 [1.11-1.14]* |
| E13 | 4,340 | 80,138,520 | 0.07 | 0.05 | 0.05 | 0.02 | 0.02 | 0.01 | 1.39 [1.26-1.54]* | 1.18 [1.06-1.31]* |
| E14 | 47,840 | 79,581,090 | 0.78 | 0.56 | 0.57 | 0.22 | 0.22 | 0.12 | 1.38 [1.34-1.42]* | 1.19 [1.15-1.23]* |
| E16 | 51,520 | 79,856,800 | 0.80 | 0.61 | 0.63 | 0.19 | 0.17 | 0.08 | 1.27 [1.24-1.31]* | 1.11 [1.08-1.14]* |
| E20 | 1,410 | 80,158,600 | 0.02 | 0.02 | 0.02 | 0.01 | 0.01 | 0.01 | 1.49 [1.25-1.77]* | 1.30 [1.09-1.56]* |
| E21 | 17,310 | 80,037,460 | 0.25 | 0.21 | 0.21 | 0.04 | 0.04 | 0.02 | 1.17 [1.11-1.23]* | 1.08 [1.02-1.14]* |
| E22 | 11,350 | 80,084,240 | 0.20 | 0.13 | 0.13 | 0.07 | 0.06 | 0.05 | 1.47 [1.38-1.56]* | 1.37 [1.28-1.46]* |
| E23 | 5,200 | 80,080,540 | 0.09 | 0.06 | 0.06 | 0.03 | 0.03 | 0.02 | 1.43 [1.30-1.56]* | 1.27 [1.15-1.39]* |
| E24 | 1,300 | 80,150,800 | 0.02 | 0.01 | 0.02 | 0.01 | 0.01 | 0.00 | 1.46 [1.22-1.74]* | 1.12 [0.93-1.36] |
| E26 | 770 | 80,160,640 | 0.01 | 0.01 | 0.01 | 0.00 | 0.00 | 0.00 | 1.16 [0.91-1.49] | 1.04 [0.80-1.34] |
| E27 | 11,690 | 80,067,770 | 0.20 | 0.13 | 0.13 | 0.06 | 0.06 | 0.04 | 1.48 [1.39-1.57]* | 1.28 [1.20-1.36]* |
| E28 | 31,030 | 79,740,110 | 0.49 | 0.37 | 0.39 | 0.12 | 0.10 | 0.04 | 1.24 [1.20-1.29]* | 1.08 [1.04-1.12]* |
| E29 | 1,700 | 80,150,470 | 0.03 | 0.02 | 0.02 | 0.01 | 0.01 | 0.01 | 1.76 [1.51-2.05]* | 1.42 [1.21-1.67]* |
| E34 | 1,770 | 80,110,540 | 0.03 | 0.02 | 0.02 | 0.01 | 0.01 | 0.00 | 1.31 [1.12-1.53]* | 1.13 [0.96-1.34] |
| E41 | 630 | 80,170,980 | 0.01 | 0.01 | 0.01 | 0.00 | 0.00 | 0.00 | 1.05 [0.78-1.41] | 1.06 [0.78-1.42] |
| E43 | 760 | 80,169,640 | 0.01 | 0.01 | 0.01 | 0.00 | 0.00 | 0.00 | 1.09 [0.84-1.40] | 1.00 [0.77-1.30] |
| E46 | 9,480 | 80,130,200 | 0.14 | 0.11 | 0.12 | 0.02 | 0.02 | 0.01 | 1.18 [1.10-1.27]* | 1.10 [1.02-1.18] |
| E51 | 940 | 80,163,950 | 0.02 | 0.01 | 0.01 | 0.00 | 0.00 | 0.00 | 1.41 [1.14-1.74]* | 1.23 [0.99-1.54] |
| E53 | 54,100 | 79,884,220 | 0.86 | 0.64 | 0.65 | 0.22 | 0.20 | 0.13 | 1.31 [1.27-1.35]* | 1.18 [1.15-1.22]* |
| E55 | 70,300 | 79,861,030 | 1.17 | 0.82 | 0.85 | 0.36 | 0.32 | 0.21 | 1.38 [1.34-1.42]* | 1.22 [1.19-1.25]* |
| E56 | 680 | 80,168,120 | 0.01 | 0.01 | 0.01 | 0.00 | 0.00 | 0.00 | 1.29 [1.00-1.67] | 1.14 [0.87-1.48] |
| E58 | 520 | 80,169,190 | 0.01 | 0.01 | 0.01 | 0.00 | 0.00 | 0.00 | 1.41 [1.06-1.88] | 1.25 [0.93-1.68] |
| E61 | 10,600 | 80,106,180 | 0.18 | 0.12 | 0.13 | 0.05 | 0.05 | 0.03 | 1.39 [1.30-1.48]* | 1.21 [1.14-1.30]* |
| E63 | 650 | 80,168,850 | 0.01 | 0.01 | 0.01 | 0.00 | 0.00 | 0.00 | 1.17 [0.89-1.52] | 1.08 [0.82-1.41] |
| E65 | 440 | 80,153,630 | 0.01 | 0.01 | 0.00 | 0.00 | 0.00 | 0.00 | 1.54 [1.14-2.08]* | 1.39 [1.02-1.90] |
| E66 | 382,520 | 76,679,140 | 6.33 | 4.70 | 4.80 | 1.63 | 1.53 | 0.85 | 1.32 [1.30-1.33]* | 1.15 [1.14-1.17]* |
| E72 | 610 | 80,162,480 | 0.01 | 0.01 | 0.01 | 0.00 | 0.00 | 0.00 | 1.39 [1.07-1.81] | 1.24 [0.94-1.62] |
| E73 | 2,470 | 80,146,600 | 0.04 | 0.03 | 0.03 | 0.01 | 0.01 | 0.00 | 1.38 [1.21-1.57]* | 1.11 [0.96-1.27] |
| E74 | 1,140 | 80,155,760 | 0.02 | 0.01 | 0.01 | 0.00 | 0.00 | 0.00 | 1.34 [1.10-1.63]* | 1.22 [1.00-1.49] |
| E78 | 448,200 | 74,422,520 | 7.31 | 5.74 | 5.94 | 1.57 | 1.37 | 0.76 | 1.23 [1.22-1.24]* | 1.12 [1.10-1.13]* |
| E79 | 590 | 80,167,340 | 0.01 | 0.01 | 0.01 | 0.00 | 0.00 | 0.00 | 1.35 [1.02-1.77] | 1.21 [0.91-1.60] |
| E80 | 5,590 | 80,108,470 | 0.08 | 0.07 | 0.07 | 0.01 | 0.01 | 0.01 | 1.21 [1.11-1.33]* | 1.11 [1.01-1.22] |
| E83 | 81,730 | 79,797,030 | 1.23 | 0.98 | 1.01 | 0.25 | 0.22 | 0.13 | 1.22 [1.19-1.25]* | 1.12 [1.09-1.15]* |
| E85 | 2,950 | 80,156,370 | 0.04 | 0.04 | 0.04 | 0.01 | 0.01 | 0.01 | 1.22 [1.07-1.38]* | 1.14 [1.00-1.29] |
| E86 | 207,200 | 79,213,790 | 3.12 | 2.51 | 2.62 | 0.62 | 0.50 | 0.33 | 1.19 [1.17-1.21]* | 1.12 [1.10-1.14]* |
| E87 | 323,540 | 78,737,560 | 5.16 | 3.88 | 4.01 | 1.27 | 1.14 | 0.80 | 1.29 [1.27-1.30]* | 1.18 [1.17-1.20]* |
| E88 | 14,800 | 80,099,900 | 0.23 | 0.17 | 0.18 | 0.06 | 0.06 | 0.04 | 1.32 [1.24-1.39]* | 1.21 [1.14-1.28]* |
| E89 | 20,760 | 79,950,920 | 0.32 | 0.25 | 0.25 | 0.07 | 0.07 | 0.04 | 1.26 [1.20-1.32]* | 1.14 [1.09-1.20]* |
| 18+ - F01-F99 | | | | | | | | | | |
| F00 | 51,300 | 80,017,420 | 0.72 | 0.62 | 0.67 | 0.09 | 0.04 | 0.03 | 1.06 [1.03-1.10]* | 1.04 [1.01-1.08] |
| F01 | 58,730 | 79,987,690 | 0.83 | 0.71 | 0.76 | 0.12 | 0.07 | 0.04 | 1.10 [1.06-1.13]* | 1.05 [1.02-1.08]* |
| F02 | 10,280 | 80,140,290 | 0.13 | 0.13 | 0.13 | 0.01 | 0.00 | 0.00 | 1.01 [0.94-1.09] | 0.99 [0.92-1.07] |
| F03 | 141,440 | 79,704,490 | 1.99 | 1.73 | 1.87 | 0.27 | 0.12 | 0.08 | 1.06 [1.04-1.09]* | 1.04 [1.02-1.06]* |
| F05 | 129,540 | 79,856,300 | 1.93 | 1.56 | 1.65 | 0.38 | 0.28 | 0.18 | 1.17 [1.15-1.19]* | 1.10 [1.08-1.13]* |
| F06 | 20,330 | 80,075,900 | 0.30 | 0.24 | 0.26 | 0.05 | 0.04 | 0.03 | 1.16 [1.10-1.22]* | 1.10 [1.04-1.16]* |
| F07 | 1,240 | 80,151,110 | 0.02 | 0.02 | 0.02 | 0.01 | 0.00 | 0.00 | 1.29 [1.07-1.55]* | 1.16 [0.96-1.41] |
| F10 | 105,170 | 78,294,640 | 1.58 | 1.29 | 1.29 | 0.29 | 0.29 | 0.16 | 1.23 [1.20-1.25]* | 1.11 [1.09-1.14]* |
| F11 | 8,220 | 79,936,370 | 0.12 | 0.10 | 0.10 | 0.02 | 0.02 | −0.01 | 1.18 [1.10-1.28]* | 0.95 [0.88-1.03] |
| F12 | 16,910 | 79,960,920 | 0.24 | 0.21 | 0.21 | 0.03 | 0.02 | −0.01 | 1.12 [1.06-1.18]* | 0.95 [0.90-1.00] |
| F13 | 1,820 | 80,143,720 | 0.03 | 0.02 | 0.02 | 0.01 | 0.01 | 0.00 | 1.24 [1.06-1.45]* | 0.98 [0.83-1.15] |
| F14 | 7,070 | 80,076,110 | 0.10 | 0.09 | 0.09 | 0.01 | 0.01 | 0.00 | 1.16 [1.07-1.26]* | 0.97 [0.89-1.05] |
| F15 | 2,380 | 80,123,500 | 0.03 | 0.03 | 0.03 | 0.00 | 0.00 | 0.00 | 1.04 [0.90-1.20] | 0.93 [0.80-1.08] |

| | | | | | | | | | | |
|---|---|---|---|---|---|---|---|---|---|---|
| F16 | 710 | 80,153,180 | 0.01 | 0.01 | 0.01 | 0.00 | 0.00 | 0.00 | 0.87 [0.66-1.15] | 0.78 [0.59-1.03] |
| F17 | 522,180 | 75,245,470 | 7.89 | 6.73 | 6.86 | 1.16 | 1.03 | 0.16 | 1.15 [1.14-1.16]* | 1.02 [1.01-1.03]* |
| F19 | 7,530 | 80,039,390 | 0.10 | 0.09 | 0.09 | 0.01 | 0.01 | −0.01 | 1.08 [1.00-1.17] | 0.89 [0.82-0.97]* |
| F20 | 15,640 | 79,774,600 | 0.24 | 0.19 | 0.19 | 0.05 | 0.05 | 0.04 | 1.27 [1.20-1.34]* | 1.17 [1.11-1.24]* |
| F22 | 6,980 | 80,086,630 | 0.11 | 0.08 | 0.08 | 0.02 | 0.02 | 0.02 | 1.28 [1.18-1.39]* | 1.18 [1.08-1.28]* |
| F23 | 4,240 | 80,081,270 | 0.06 | 0.05 | 0.05 | 0.01 | 0.01 | 0.00 | 1.12 [1.01-1.24] | 1.05 [0.94-1.17] |
| F25 | 3,290 | 80,086,970 | 0.05 | 0.04 | 0.04 | 0.01 | 0.01 | 0.01 | 1.34 [1.20-1.50]* | 1.19 [1.06-1.34]* |
| F29 | 7,820 | 80,074,980 | 0.12 | 0.09 | 0.09 | 0.03 | 0.02 | 0.01 | 1.26 [1.17-1.36]* | 1.11 [1.03-1.20]* |
| F30 | 2,050 | 80,114,130 | 0.03 | 0.02 | 0.02 | 0.01 | 0.01 | 0.01 | 1.33 [1.15-1.54]* | 1.21 [1.05-1.41] |
| F31 | 18,770 | 79,851,170 | 0.30 | 0.22 | 0.23 | 0.08 | 0.07 | 0.02 | 1.32 [1.26-1.39]* | 1.08 [1.03-1.14]* |
| F32 | 344,020 | 76,481,710 | 5.91 | 4.19 | 4.29 | 1.72 | 1.63 | 0.68 | 1.38 [1.36-1.40]* | 1.13 [1.12-1.14]* |
| F33 | 10,780 | 79,917,480 | 0.17 | 0.13 | 0.13 | 0.04 | 0.04 | 0.01 | 1.30 [1.22-1.39]* | 1.07 [1.00-1.14] |
| F34 | 1,720 | 80,144,770 | 0.03 | 0.02 | 0.02 | 0.01 | 0.01 | 0.00 | 1.47 [1.26-1.72]* | 1.18 [1.00-1.39] |
| F38 | 810 | 80,163,640 | 0.01 | 0.01 | 0.01 | 0.00 | 0.00 | 0.00 | 1.27 [1.00-1.62] | 1.09 [0.85-1.40] |
| F39 | 1,360 | 80,154,190 | 0.02 | 0.02 | 0.02 | 0.01 | 0.01 | 0.00 | 1.35 [1.13-1.61]* | 1.10 [0.92-1.33] |
| F40 | 29,180 | 79,880,880 | 0.46 | 0.34 | 0.36 | 0.12 | 0.11 | 0.05 | 1.30 [1.25-1.35]* | 1.13 [1.09-1.18]* |
| F41 | 289,410 | 77,972,800 | 4.84 | 3.47 | 3.58 | 1.37 | 1.25 | 0.50 | 1.35 [1.33-1.37]* | 1.11 [1.10-1.13]* |
| F42 | 6,670 | 80,091,320 | 0.13 | 0.07 | 0.08 | 0.05 | 0.05 | 0.03 | 1.67 [1.54-1.80]* | 1.33 [1.22-1.44]* |
| F43 | 16,300 | 79,913,940 | 0.25 | 0.19 | 0.20 | 0.06 | 0.06 | 0.00 | 1.28 [1.21-1.35]* | 1.01 [0.95-1.06] |
| F44 | 3,700 | 80,122,590 | 0.06 | 0.04 | 0.04 | 0.02 | 0.02 | 0.01 | 1.43 [1.28-1.59]* | 1.11 [1.00-1.25] |
| F45 | 4,560 | 80,088,620 | 0.08 | 0.05 | 0.05 | 0.02 | 0.02 | 0.01 | 1.40 [1.27-1.54]* | 1.15 [1.04-1.27]* |
| F48 | 1,250 | 80,153,460 | 0.02 | 0.01 | 0.01 | 0.01 | 0.01 | 0.00 | 1.56 [1.30-1.87]* | 1.27 [1.05-1.54] |
| F50 | 5,670 | 80,058,330 | 0.09 | 0.07 | 0.07 | 0.02 | 0.02 | 0.01 | 1.29 [1.18-1.41]* | 1.08 [0.99-1.18] |
| F51 | 1,170 | 80,156,970 | 0.02 | 0.01 | 0.01 | 0.01 | 0.01 | 0.01 | 1.57 [1.30-1.89]* | 1.32 [1.09-1.60]* |
| F52 | 2,680 | 80,126,510 | 0.04 | 0.03 | 0.03 | 0.01 | 0.01 | 0.01 | 1.48 [1.30-1.67]* | 1.25 [1.10-1.42]* |
| F53 | 2,640 | 80,124,710 | 0.04 | 0.03 | 0.03 | 0.01 | 0.01 | 0.01 | 1.34 [1.18-1.52]* | 1.14 [0.99-1.30] |
| F60 | 13,830 | 79,931,610 | 0.23 | 0.16 | 0.17 | 0.07 | 0.06 | 0.01 | 1.39 [1.31-1.47]* | 1.05 [0.99-1.11] |
| F61 | 520 | 80,163,070 | 0.01 | 0.01 | 0.01 | 0.00 | 0.00 | 0.00 | 1.17 [0.88-1.56] | 0.97 [0.71-1.32] |
| F69 | 910 | 80,151,420 | 0.02 | 0.01 | 0.01 | 0.01 | 0.01 | 0.00 | 1.62 [1.31-1.99]* | 1.27 [1.01-1.58] |
| F70 | 1,780 | 80,130,620 | 0.04 | 0.02 | 0.02 | 0.02 | 0.02 | 0.01 | 1.87 [1.62-2.16]* | 1.65 [1.42-1.92]* |
| F71 | 680 | 80,155,010 | 0.02 | 0.01 | 0.01 | 0.01 | 0.01 | 0.01 | 2.40 [1.92-3.00]* | 2.23 [1.77-2.82]* |
| F72 | 1,100 | 80,144,110 | 0.03 | 0.01 | 0.01 | 0.01 | 0.01 | 0.01 | 2.15 [1.80-2.57]* | 2.09 [1.74-2.52]* |
| F79 | 6,820 | 80,056,830 | 0.14 | 0.07 | 0.07 | 0.07 | 0.07 | 0.06 | 1.89 [1.76-2.04]* | 1.74 [1.61-1.88]* |
| F80 | 2,960 | 80,124,610 | 0.05 | 0.03 | 0.03 | 0.01 | 0.01 | 0.01 | 1.38 [1.22-1.56]* | 1.29 [1.13-1.46]* |
| F81 | 18,290 | 79,755,950 | 0.38 | 0.20 | 0.20 | 0.18 | 0.18 | 0.17 | 1.92 [1.83-2.00]* | 1.79 [1.71-1.88]* |
| F84 | 8,200 | 79,985,810 | 0.15 | 0.09 | 0.10 | 0.06 | 0.05 | 0.04 | 1.54 [1.44-1.65]* | 1.34 [1.24-1.44]* |
| F89 | 650 | 80,159,130 | 0.01 | 0.01 | 0.01 | 0.00 | 0.00 | 0.00 | 1.53 [1.20-1.96]* | 1.33 [1.02-1.73] |
| F90 | 4,690 | 80,072,810 | 0.07 | 0.06 | 0.06 | 0.02 | 0.01 | 0.01 | 1.23 [1.12-1.35]* | 1.09 [0.99-1.21] |
| F91 | 770 | 80,132,720 | 0.01 | 0.01 | 0.01 | 0.00 | 0.00 | 0.00 | 1.19 [0.93-1.52] | 1.08 [0.84-1.39] |
| F95 | 550 | 80,160,130 | 0.01 | 0.01 | 0.01 | 0.00 | 0.00 | 0.00 | 1.52 [1.16-1.98]* | 1.23 [0.93-1.64] |
| F98 | 580 | 80,152,410 | 0.01 | 0.01 | 0.01 | 0.00 | 0.00 | 0.00 | 1.26 [0.95-1.66] | 1.08 [0.81-1.44] |
| F99 | 5,420 | 80,049,540 | 0.08 | 0.06 | 0.07 | 0.02 | 0.02 | 0.01 | 1.27 [1.16-1.39]* | 1.08 [0.99-1.19] |
| 18+ - G00-G99 | | | | | | | | | | |
| G00 | 1,300 | 80,145,200 | 0.02 | 0.02 | 0.02 | 0.00 | 0.00 | 0.00 | 1.20 [1.00-1.44] | 1.12 [0.92-1.35] |
| G02 | 620 | 80,163,070 | 0.01 | 0.01 | 0.01 | 0.00 | 0.00 | 0.00 | 1.19 [0.90-1.56] | 1.04 [0.78-1.39] |
| G03 | 1,540 | 80,144,640 | 0.02 | 0.02 | 0.02 | 0.00 | 0.00 | 0.00 | 1.17 [0.98-1.39] | 1.03 [0.86-1.24] |
| G04 | 3,780 | 80,130,350 | 0.06 | 0.04 | 0.05 | 0.01 | 0.01 | 0.01 | 1.28 [1.15-1.43]* | 1.17 [1.04-1.30]* |
| G05 | 960 | 80,161,550 | 0.02 | 0.01 | 0.01 | 0.01 | 0.01 | 0.01 | 1.55 [1.26-1.91]* | 1.46 [1.18-1.81]* |
| G06 | 1,790 | 80,149,230 | 0.03 | 0.02 | 0.02 | 0.00 | 0.00 | 0.00 | 1.20 [1.02-1.41] | 1.14 [0.97-1.34] |
| G08 | 910 | 80,161,160 | 0.01 | 0.01 | 0.01 | 0.00 | 0.00 | 0.00 | 1.32 [1.06-1.65] | 1.26 [1.00-1.58] |
| G09 | 960 | 80,154,610 | 0.02 | 0.01 | 0.01 | 0.01 | 0.00 | 0.00 | 1.38 [1.12-1.70]* | 1.27 [1.03-1.57] |
| G10 | 620 | 80,164,800 | 0.01 | 0.01 | 0.01 | 0.00 | 0.00 | 0.00 | 1.33 [1.02-1.74] | 1.39 [1.05-1.83] |
| G11 | 1,820 | 80,147,570 | 0.03 | 0.02 | 0.02 | 0.00 | 0.01 | 0.00 | 1.25 [1.07-1.46]* | 1.16 [0.99-1.36] |
| G12 | 4,240 | 80,151,580 | 0.06 | 0.05 | 0.05 | 0.01 | 0.01 | 0.00 | 1.12 [1.00-1.24] | 1.07 [0.96-1.20] |
| G20 | 39,030 | 79,917,280 | 0.53 | 0.48 | 0.50 | 0.04 | 0.03 | 0.01 | 1.05 [1.01-1.09]* | 1.02 [0.99-1.06] |
| G21 | 3,390 | 80,154,550 | 0.05 | 0.04 | 0.04 | 0.01 | 0.01 | 0.01 | 1.27 [1.13-1.43]* | 1.20 [1.06-1.35]* |
| G23 | 2,110 | 80,164,420 | 0.03 | 0.03 | 0.03 | 0.00 | 0.00 | 0.00 | 1.12 [0.96-1.30] | 1.07 [0.92-1.25] |
| G24 | 3,890 | 80,116,180 | 0.06 | 0.05 | 0.05 | 0.01 | 0.01 | 0.01 | 1.20 [1.08-1.34]* | 1.10 [0.98-1.23] |
| G25 | 20,980 | 80,033,280 | 0.33 | 0.25 | 0.25 | 0.08 | 0.08 | 0.04 | 1.30 [1.24-1.36]* | 1.12 [1.07-1.17]* |
| G30 | 68,840 | 79,929,490 | 0.96 | 0.84 | 0.90 | 0.12 | 0.07 | 0.05 | 1.07 [1.04-1.10]* | 1.05 [1.02-1.08]* |
| G31 | 49,290 | 79,945,050 | 0.72 | 0.59 | 0.61 | 0.12 | 0.11 | 0.08 | 1.18 [1.14-1.22]* | 1.12 [1.08-1.16]* |
| G32 | 800 | 80,165,300 | 0.01 | 0.01 | 0.01 | 0.00 | 0.00 | 0.00 | 1.29 [1.02-1.63] | 1.12 [0.88-1.43] |
| G35 | 10,540 | 79,921,820 | 0.16 | 0.13 | 0.13 | 0.03 | 0.03 | 0.01 | 1.22 [1.15-1.30]* | 1.08 [1.01-1.15] |
| G37 | 2,670 | 80,113,840 | 0.04 | 0.03 | 0.03 | 0.01 | 0.01 | 0.01 | 1.30 [1.14-1.47]* | 1.16 [1.02-1.33] |
| G40 | 62,420 | 78,853,860 | 1.01 | 0.74 | 0.74 | 0.27 | 0.27 | 0.16 | 1.36 [1.32-1.40]* | 1.19 [1.15-1.22]* |
| G41 | 3,840 | 80,113,080 | 0.06 | 0.05 | 0.05 | 0.02 | 0.02 | 0.01 | 1.33 [1.20-1.48]* | 1.20 [1.07-1.34]* |
| G43 | 64,020 | 79,382,920 | 1.04 | 0.76 | 0.78 | 0.28 | 0.25 | 0.08 | 1.32 [1.29-1.36]* | 1.08 [1.05-1.11]* |
| G44 | 9,430 | 80,026,910 | 0.15 | 0.11 | 0.11 | 0.04 | 0.04 | 0.02 | 1.31 [1.23-1.40]* | 1.12 [1.04-1.20]* |
| G45 | 47,120 | 79,613,830 | 0.69 | 0.57 | 0.58 | 0.12 | 0.11 | 0.06 | 1.18 [1.15-1.22]* | 1.10 [1.07-1.14]* |
| G46 | 3,070 | 80,152,960 | 0.05 | 0.04 | 0.04 | 0.01 | 0.01 | 0.00 | 1.18 [1.04-1.34]* | 1.12 [0.98-1.27] |
| G47 | 60,510 | 79,340,920 | 1.09 | 0.69 | 0.69 | 0.40 | 0.40 | 0.24 | 1.58 [1.54-1.62]* | 1.28 [1.24-1.31]* |
| G50 | 7,680 | 80,081,670 | 0.13 | 0.09 | 0.09 | 0.04 | 0.04 | 0.02 | 1.43 [1.33-1.55]* | 1.22 [1.13-1.32]* |
| G51 | 15,090 | 79,965,510 | 0.23 | 0.18 | 0.18 | 0.05 | 0.04 | 0.03 | 1.24 [1.17-1.31]* | 1.13 [1.06-1.19]* |
| G52 | 1,160 | 80,158,070 | 0.02 | 0.01 | 0.01 | 0.00 | 0.00 | 0.00 | 1.32 [1.08-1.60]* | 1.18 [0.97-1.45] |
| G53 | 2,650 | 80,151,010 | 0.04 | 0.03 | 0.03 | 0.01 | 0.01 | 0.01 | 1.32 [1.15-1.51]* | 1.21 [1.06-1.39]* |
| G54 | 4,970 | 80,108,040 | 0.08 | 0.06 | 0.06 | 0.02 | 0.02 | 0.01 | 1.29 [1.17-1.42]* | 1.10 [1.00-1.22] |
| G55 | 54,890 | 79,479,170 | 0.84 | 0.66 | 0.66 | 0.18 | 0.18 | 0.08 | 1.27 [1.23-1.31]* | 1.11 [1.07-1.14]* |
| G56 | 82,240 | 78,384,650 | 1.32 | 0.99 | 0.99 | 0.33 | 0.33 | 0.20 | 1.34 [1.31-1.37]* | 1.18 [1.15-1.21]* |
| G57 | 9,510 | 80,000,380 | 0.15 | 0.11 | 0.11 | 0.04 | 0.04 | 0.03 | 1.40 [1.31-1.50]* | 1.25 [1.17-1.34]* |
| G58 | 4,760 | 80,100,780 | 0.08 | 0.06 | 0.05 | 0.02 | 0.02 | 0.01 | 1.40 [1.27-1.54]* | 1.21 [1.09-1.33]* |
| G59 | 3,740 | 80,125,150 | 0.06 | 0.04 | 0.04 | 0.02 | 0.02 | 0.01 | 1.47 [1.33-1.64]* | 1.18 [1.05-1.32]* |
| G60 | 2,180 | 80,141,610 | 0.03 | 0.03 | 0.03 | 0.01 | 0.01 | 0.00 | 1.26 [1.10-1.46]* | 1.12 [0.97-1.30] |
| G61 | 2,730 | 80,125,460 | 0.04 | 0.03 | 0.03 | 0.01 | 0.01 | 0.01 | 1.26 [1.11-1.43]* | 1.19 [1.05-1.36]* |
| G62 | 30,200 | 79,943,570 | 0.48 | 0.36 | 0.36 | 0.13 | 0.13 | 0.07 | 1.36 [1.30-1.41]* | 1.16 [1.12-1.21]* |
| G63 | 15,880 | 80,064,770 | 0.27 | 0.18 | 0.19 | 0.09 | 0.08 | 0.04 | 1.44 [1.37-1.52]* | 1.16 [1.10-1.23]* |
| G70 | 2,660 | 80,136,020 | 0.04 | 0.03 | 0.03 | 0.01 | 0.01 | 0.00 | 1.17 [1.02-1.33] | 1.09 [0.95-1.25] |
| G71 | 1,790 | 80,135,240 | 0.03 | 0.02 | 0.02 | 0.01 | 0.00 | 0.00 | 1.22 [1.04-1.43] | 1.16 [0.98-1.36] |
| G72 | 3,180 | 80,138,570 | 0.05 | 0.04 | 0.04 | 0.02 | 0.02 | 0.01 | 1.42 [1.27-1.60]* | 1.28 [1.13-1.44]* |
| G80 | 3,500 | 80,023,650 | 0.06 | 0.04 | 0.04 | 0.02 | 0.02 | 0.01 | 1.39 [1.25-1.55]* | 1.33 [1.19-1.49]* |
| G81 | 53,970 | 79,669,480 | 0.79 | 0.65 | 0.67 | 0.14 | 0.13 | 0.08 | 1.19 [1.15-1.23]* | 1.12 [1.08-1.15]* |
| G82 | 5,380 | 80,071,480 | 0.08 | 0.06 | 0.06 | 0.01 | 0.01 | 0.01 | 1.22 [1.11-1.34]* | 1.14 [1.04-1.25]* |
| G83 | 8,340 | 80,086,550 | 0.12 | 0.10 | 0.10 | 0.02 | 0.02 | 0.01 | 1.20 [1.12-1.30]* | 1.09 [1.01-1.17] |
| G90 | 4,420 | 80,127,690 | 0.07 | 0.05 | 0.05 | 0.01 | 0.01 | 0.00 | 1.23 [1.11-1.36]* | 1.07 [0.97-1.19] |
| G91 | 9,700 | 80,064,700 | 0.14 | 0.12 | 0.12 | 0.03 | 0.03 | 0.02 | 1.21 [1.13-1.30]* | 1.16 [1.08-1.25]* |
| G93 | 40,710 | 79,815,670 | 0.63 | 0.48 | 0.49 | 0.14 | 0.13 | 0.07 | 1.27 [1.23-1.31]* | 1.12 [1.08-1.16]* |
| G94 | 1,330 | 80,162,300 | 0.02 | 0.02 | 0.02 | 0.00 | 0.00 | 0.00 | 1.25 [1.04-1.50] | 1.22 [1.01-1.47] |
| G95 | 11,430 | 80,062,640 | 0.17 | 0.14 | 0.14 | 0.03 | 0.03 | 0.02 | 1.22 [1.14-1.29]* | 1.13 [1.06-1.21]* |
| G96 | 3,430 | 80,124,140 | 0.05 | 0.04 | 0.04 | 0.01 | 0.01 | 0.01 | 1.35 [1.20-1.50]* | 1.18 [1.05-1.32]* |
| G97 | 3,220 | 80,116,420 | 0.05 | 0.04 | 0.04 | 0.01 | 0.01 | 0.00 | 1.26 [1.12-1.42]* | 1.09 [0.97-1.23] |
| G98 | 1,160 | 80,164,560 | 0.02 | 0.01 | 0.01 | 0.01 | 0.01 | 0.00 | 1.46 [1.20-1.76]* | 1.18 [0.97-1.45] |
| G99 | 18,070 | 79,994,670 | 0.27 | 0.22 | 0.21 | 0.06 | 0.06 | 0.03 | 1.28 [1.21-1.34]* | 1.13 [1.08-1.19]* |
| 18+ - H00-H59 | | | | | | | | | | |
| H00 | 11,560 | 79,772,400 | 0.17 | 0.14 | 0.14 | 0.03 | 0.04 | 0.03 | 1.27 [1.19-1.35]* | 1.22 [1.14-1.29]* |
| H01 | 10,800 | 80,042,890 | 0.18 | 0.13 | 0.13 | 0.05 | 0.05 | 0.04 | 1.39 [1.30-1.48]* | 1.29 [1.21-1.38]* |
| H02 | 57,230 | 79,164,860 | 0.90 | 0.68 | 0.69 | 0.22 | 0.22 | 0.16 | 1.32 [1.28-1.35]* | 1.22 [1.19-1.26]* |
| H03 | 590 | 80,158,970 | 0.01 | 0.01 | 0.01 | 0.00 | 0.00 | 0.00 | 1.41 [1.07-1.84] | 1.25 [0.95-1.65] |

| | | | | | | | | | | |
|---|---|---|---|---|---|---|---|---|---|---|
| H04 | 26,580 | 79,731,600 | 0.45 | 0.31 | 0.31 | 0.14 | 0.14 | 0.11 | 1.44 [1.38-1.50]* | 1.32 [1.27-1.37]* |
| H05 | 4,880 | 80,053,780 | 0.08 | 0.06 | 0.06 | 0.03 | 0.02 | 0.02 | 1.43 [1.30-1.57]* | 1.31 [1.19-1.45]* |
| H06 | 1,180 | 80,153,280 | 0.02 | 0.01 | 0.01 | 0.01 | 0.01 | 0.00 | 1.45 [1.21-1.75]* | 1.28 [1.05-1.55] |
| H10 | 11,670 | 80,047,420 | 0.20 | 0.14 | 0.14 | 0.06 | 0.06 | 0.04 | 1.43 [1.34-1.52]* | 1.29 [1.21-1.38]* |
| H11 | 11,910 | 79,990,360 | 0.18 | 0.14 | 0.14 | 0.04 | 0.03 | 0.03 | 1.24 [1.16-1.32]* | 1.18 [1.11-1.26]* |
| H13 | 660 | 80,161,870 | 0.01 | 0.01 | 0.01 | 0.00 | 0.00 | 0.00 | 1.59 [1.24-2.03]* | 1.50 [1.16-1.93]* |
| H15 | 1,070 | 80,156,610 | 0.02 | 0.01 | 0.01 | 0.01 | 0.01 | 0.00 | 1.45 [1.19-1.77]* | 1.28 [1.05-1.58] |
| H16 | 5,130 | 80,092,560 | 0.09 | 0.06 | 0.06 | 0.04 | 0.04 | 0.03 | 1.64 [1.50-1.80]* | 1.52 [1.39-1.67]* |
| H17 | 3,010 | 80,135,600 | 0.05 | 0.03 | 0.04 | 0.02 | 0.01 | 0.01 | 1.40 [1.24-1.57]* | 1.30 [1.15-1.48]* |
| H18 | 21,360 | 79,971,630 | 0.35 | 0.25 | 0.26 | 0.10 | 0.09 | 0.08 | 1.36 [1.30-1.43]* | 1.27 [1.22-1.34]* |
| H19 | 1,300 | 80,155,570 | 0.03 | 0.01 | 0.01 | 0.01 | 0.01 | 0.01 | 1.85 [1.55-2.19]* | 1.59 [1.33-1.90]* |
| H20 | 5,410 | 80,087,020 | 0.09 | 0.06 | 0.06 | 0.03 | 0.02 | 0.02 | 1.38 [1.26-1.50]* | 1.29 [1.18-1.41]* |
| H21 | 9,050 | 80,083,880 | 0.13 | 0.11 | 0.11 | 0.02 | 0.02 | 0.01 | 1.18 [1.09-1.26]* | 1.12 [1.04-1.21]* |
| H22 | 420 | 80,168,300 | 0.01 | 0.00 | 0.01 | 0.00 | 0.00 | 0.00 | 1.45 [1.06-1.98] | 1.28 [0.92-1.77] |
| H25 | 220,590 | 77,744,420 | 3.34 | 2.73 | 2.85 | 0.61 | 0.49 | 0.35 | 1.17 [1.16-1.19]* | 1.12 [1.10-1.13]* |
| H26 | 310,740 | 76,301,420 | 4.93 | 3.89 | 4.08 | 1.04 | 0.85 | 0.62 | 1.21 [1.19-1.22]* | 1.14 [1.13-1.16]* |
| H27 | 4,110 | 80,108,110 | 0.06 | 0.05 | 0.05 | 0.01 | 0.01 | 0.01 | 1.22 [1.09-1.35]* | 1.16 [1.04-1.29]* |
| H28 | 1,610 | 80,154,350 | 0.03 | 0.02 | 0.02 | 0.01 | 0.01 | 0.00 | 1.35 [1.14-1.59]* | 1.22 [1.03-1.44] |
| H30 | 490 | 80,164,030 | 0.01 | 0.01 | 0.01 | 0.00 | 0.00 | 0.00 | 1.29 [0.96-1.74] | 1.17 [0.87-1.59] |
| H31 | 5,180 | 80,123,870 | 0.07 | 0.06 | 0.07 | 0.01 | 0.01 | 0.01 | 1.13 [1.02-1.24] | 1.09 [0.98-1.20] |
| H33 | 20,280 | 79,786,420 | 0.27 | 0.25 | 0.25 | 0.02 | 0.02 | 0.01 | 1.08 [1.02-1.13]* | 1.05 [1.00-1.11] |
| H34 | 12,600 | 80,048,470 | 0.18 | 0.15 | 0.16 | 0.03 | 0.03 | 0.02 | 1.17 [1.10-1.24]* | 1.11 [1.04-1.18]* |
| H35 | 114,660 | 79,168,620 | 1.69 | 1.40 | 1.44 | 0.30 | 0.25 | 0.18 | 1.17 [1.15-1.20]* | 1.12 [1.10-1.15]* |
| H36 | 45,930 | 79,714,600 | 0.70 | 0.55 | 0.56 | 0.15 | 0.14 | 0.06 | 1.25 [1.21-1.29]* | 1.09 [1.06-1.13]* |
| H40 | 105,600 | 79,099,740 | 1.55 | 1.29 | 1.32 | 0.26 | 0.23 | 0.18 | 1.18 [1.15-1.20]* | 1.13 [1.11-1.16]* |
| H43 | 21,350 | 79,925,070 | 0.30 | 0.26 | 0.27 | 0.04 | 0.03 | 0.02 | 1.13 [1.07-1.18]* | 1.07 [1.01-1.12] |
| H44 | 6,590 | 80,085,460 | 0.10 | 0.08 | 0.08 | 0.02 | 0.02 | 0.02 | 1.32 [1.22-1.43]* | 1.25 [1.15-1.36]* |
| H46 | 1,960 | 80,142,220 | 0.03 | 0.02 | 0.02 | 0.01 | 0.01 | 0.01 | 1.41 [1.22-1.63]* | 1.27 [1.09-1.48]* |
| H47 | 7,460 | 80,079,790 | 0.11 | 0.09 | 0.09 | 0.02 | 0.02 | 0.01 | 1.23 [1.13-1.33]* | 1.13 [1.04-1.23]* |
| H49 | 5,000 | 80,107,370 | 0.07 | 0.06 | 0.06 | 0.01 | 0.01 | 0.00 | 1.10 [1.00-1.21] | 1.02 [0.92-1.13] |
| H50 | 8,680 | 79,816,600 | 0.13 | 0.10 | 0.10 | 0.03 | 0.03 | 0.02 | 1.27 [1.18-1.37]* | 1.19 [1.11-1.28]* |
| H51 | 800 | 80,160,280 | 0.01 | 0.01 | 0.01 | 0.00 | 0.00 | 0.00 | 1.25 [0.98-1.59] | 1.12 [0.88-1.44] |
| H52 | 34,000 | 79,881,490 | 0.48 | 0.41 | 0.42 | 0.07 | 0.06 | 0.04 | 1.13 [1.09-1.17]* | 1.09 [1.05-1.13]* |
| H53 | 66,430 | 79,290,290 | 1.02 | 0.80 | 0.82 | 0.22 | 0.20 | 0.10 | 1.25 [1.22-1.28]* | 1.11 [1.08-1.14]* |
| H54 | 50,990 | 79,833,400 | 0.76 | 0.61 | 0.63 | 0.15 | 0.14 | 0.10 | 1.22 [1.18-1.25]* | 1.14 [1.11-1.18]* |
| H55 | 4,360 | 80,122,850 | 0.06 | 0.05 | 0.05 | 0.01 | 0.01 | 0.01 | 1.19 [1.07-1.32]* | 1.09 [0.98-1.21] |
| H57 | 10,630 | 80,050,920 | 0.16 | 0.13 | 0.13 | 0.04 | 0.03 | 0.02 | 1.27 [1.19-1.36]* | 1.15 [1.07-1.23]* |
| H58 | 2,220 | 80,152,120 | 0.03 | 0.03 | 0.03 | 0.01 | 0.01 | 0.00 | 1.26 [1.09-1.46]* | 1.15 [1.00-1.34] |
| H59 | 4,090 | 80,110,430 | 0.06 | 0.05 | 0.05 | 0.01 | 0.01 | 0.01 | 1.16 [1.05-1.30]* | 1.13 [1.01-1.26] |
| 18+ - H60-H95 | | | | | | | | | | |
| H60 | 8,970 | 79,975,150 | 0.17 | 0.10 | 0.10 | 0.07 | 0.07 | 0.06 | 1.71 [1.60-1.83]* | 1.54 [1.43-1.64]* |
| H61 | 12,720 | 79,877,880 | 0.22 | 0.15 | 0.15 | 0.07 | 0.07 | 0.06 | 1.45 [1.37-1.54]* | 1.35 [1.27-1.43]* |
| H65 | 7,060 | 79,326,880 | 0.11 | 0.08 | 0.08 | 0.03 | 0.03 | 0.02 | 1.35 [1.25-1.46]* | 1.22 [1.12-1.32]* |
| H66 | 8,700 | 79,756,590 | 0.15 | 0.10 | 0.10 | 0.05 | 0.05 | 0.04 | 1.53 [1.43-1.64]* | 1.39 [1.29-1.49]* |
| H69 | 1,670 | 80,133,000 | 0.03 | 0.02 | 0.02 | 0.01 | 0.01 | 0.01 | 1.49 [1.27-1.74]* | 1.28 [1.09-1.51]* |
| H70 | 1,660 | 80,138,850 | 0.03 | 0.02 | 0.02 | 0.01 | 0.01 | 0.00 | 1.38 [1.17-1.63]* | 1.23 [1.04-1.46] |
| H71 | 4,440 | 80,029,630 | 0.06 | 0.05 | 0.05 | 0.01 | 0.01 | 0.00 | 1.12 [1.01-1.24] | 1.08 [0.98-1.21] |
| H72 | 7,880 | 79,869,510 | 0.12 | 0.09 | 0.10 | 0.02 | 0.02 | 0.02 | 1.25 [1.16-1.35]* | 1.18 [1.09-1.27]* |
| H73 | 3,390 | 80,021,070 | 0.05 | 0.04 | 0.04 | 0.01 | 0.01 | 0.01 | 1.32 [1.18-1.48]* | 1.23 [1.09-1.38]* |
| H74 | 4,210 | 80,029,740 | 0.06 | 0.05 | 0.05 | 0.01 | 0.01 | 0.01 | 1.24 [1.12-1.37]* | 1.16 [1.04-1.29]* |
| H80 | 1,440 | 80,136,440 | 0.02 | 0.02 | 0.02 | 0.00 | 0.00 | 0.00 | 1.12 [0.93-1.34] | 1.09 [0.91-1.32] |
| H81 | 21,900 | 79,971,160 | 0.34 | 0.26 | 0.27 | 0.08 | 0.07 | 0.04 | 1.26 [1.20-1.32]* | 1.14 [1.09-1.20]* |
| H83 | 8,840 | 80,033,850 | 0.14 | 0.11 | 0.11 | 0.03 | 0.03 | 0.02 | 1.28 [1.19-1.38]* | 1.16 [1.08-1.25]* |
| H90 | 19,960 | 79,934,330 | 0.33 | 0.23 | 0.24 | 0.09 | 0.09 | 0.06 | 1.36 [1.30-1.43]* | 1.23 [1.17-1.29]* |
| H91 | 128,680 | 79,322,380 | 1.98 | 1.55 | 1.60 | 0.43 | 0.38 | 0.28 | 1.24 [1.21-1.26]* | 1.16 [1.14-1.19]* |
| H92 | 6,490 | 80,000,300 | 0.11 | 0.07 | 0.07 | 0.04 | 0.04 | 0.03 | 1.49 [1.37-1.61]* | 1.31 [1.20-1.42]* |
| H93 | 12,120 | 80,039,050 | 0.20 | 0.14 | 0.14 | 0.05 | 0.05 | 0.04 | 1.37 [1.29-1.46]* | 1.23 [1.16-1.31]* |
| H95 | 970 | 80,140,400 | 0.02 | 0.01 | 0.01 | 0.00 | 0.00 | 0.00 | 1.36 [1.10-1.67]* | 1.27 [1.03-1.58] |
| 18+ - I00-I99 | | | | | | | | | | |
| I05 | 7,070 | 80,086,800 | 0.11 | 0.08 | 0.09 | 0.03 | 0.02 | 0.02 | 1.27 [1.17-1.37]* | 1.17 [1.07-1.27]* |
| I06 | 720 | 80,160,050 | 0.01 | 0.01 | 0.01 | 0.00 | 0.00 | 0.00 | 1.25 [0.97-1.60] | 1.14 [0.89-1.48] |
| I07 | 17,970 | 80,091,840 | 0.28 | 0.21 | 0.22 | 0.07 | 0.07 | 0.05 | 1.30 [1.23-1.37]* | 1.20 [1.14-1.26]* |
| I08 | 92,780 | 79,713,320 | 1.40 | 1.11 | 1.15 | 0.29 | 0.25 | 0.17 | 1.22 [1.19-1.25]* | 1.13 [1.11-1.16]* |
| I09 | 1,040 | 80,160,100 | 0.02 | 0.01 | 0.01 | 0.01 | 0.01 | 0.00 | 1.43 [1.17-1.74]* | 1.35 [1.10-1.65]* |
| I10 | 920,510 | 66,639,780 | 17.21 | 13.07 | 13.86 | 4.15 | 3.35 | 1.94 | 1.24 [1.23-1.25]* | 1.13 [1.12-1.14]* |
| I11 | 5,240 | 80,112,440 | 0.08 | 0.06 | 0.06 | 0.01 | 0.01 | 0.01 | 1.21 [1.10-1.33]* | 1.08 [0.98-1.19] |
| I12 | 52,310 | 79,698,950 | 0.81 | 0.62 | 0.62 | 0.18 | 0.19 | 0.13 | 1.30 [1.27-1.34]* | 1.19 [1.15-1.23]* |
| I13 | 2,600 | 80,164,800 | 0.04 | 0.03 | 0.03 | 0.01 | 0.01 | 0.01 | 1.29 [1.12-1.48]* | 1.15 [1.00-1.33] |
| I15 | 3,370 | 80,128,940 | 0.05 | 0.04 | 0.04 | 0.01 | 0.01 | 0.01 | 1.31 [1.17-1.47]* | 1.17 [1.04-1.32] |
| I20 | 203,410 | 76,281,770 | 3.29 | 2.53 | 2.58 | 0.76 | 0.71 | 0.40 | 1.27 [1.26-1.29]* | 1.14 [1.12-1.15]* |
| I21 | 138,650 | 78,518,590 | 2.01 | 1.71 | 1.74 | 0.30 | 0.27 | 0.16 | 1.15 [1.13-1.18]* | 1.08 [1.06-1.11]* |
| I22 | 12,540 | 79,997,550 | 0.19 | 0.15 | 0.15 | 0.04 | 0.04 | 0.03 | 1.28 [1.21-1.36]* | 1.16 [1.09-1.23]* |
| I23 | 790 | 80,161,780 | 0.01 | 0.01 | 0.01 | 0.00 | 0.00 | 0.00 | 1.23 [0.97-1.56] | 1.15 [0.90-1.47] |
| I24 | 39,370 | 79,883,590 | 0.60 | 0.47 | 0.48 | 0.13 | 0.12 | 0.07 | 1.25 [1.21-1.29]* | 1.13 [1.09-1.17]* |
| I25 | 348,390 | 75,094,330 | 5.63 | 4.42 | 4.61 | 1.21 | 1.02 | 0.57 | 1.22 [1.21-1.24]* | 1.11 [1.10-1.13]* |
| I26 | 71,410 | 79,547,270 | 1.00 | 0.88 | 0.89 | 0.12 | 0.11 | 0.05 | 1.12 [1.09-1.15]* | 1.05 [1.02-1.08]* |
| I27 | 42,540 | 79,972,120 | 0.68 | 0.50 | 0.51 | 0.18 | 0.16 | 0.11 | 1.32 [1.28-1.37]* | 1.19 [1.15-1.23]* |
| I28 | 1,120 | 80,161,690 | 0.02 | 0.01 | 0.01 | 0.00 | 0.00 | 0.00 | 1.19 [0.97-1.46] | 1.06 [0.86-1.31] |
| I30 | 2,140 | 80,131,130 | 0.03 | 0.03 | 0.03 | 0.01 | 0.01 | 0.00 | 1.20 [1.04-1.39] | 1.13 [0.98-1.31] |
| I31 | 26,240 | 79,955,950 | 0.39 | 0.32 | 0.32 | 0.07 | 0.07 | 0.04 | 1.23 [1.18-1.28]* | 1.13 [1.08-1.18]* |
| I33 | 4,320 | 80,134,200 | 0.07 | 0.05 | 0.05 | 0.02 | 0.02 | 0.01 | 1.36 [1.23-1.51]* | 1.26 [1.13-1.40]* |
| I34 | 60,300 | 79,676,220 | 0.89 | 0.73 | 0.74 | 0.16 | 0.14 | 0.09 | 1.19 [1.16-1.22]* | 1.11 [1.08-1.14]* |
| I35 | 88,160 | 79,535,630 | 1.34 | 1.06 | 1.09 | 0.28 | 0.25 | 0.16 | 1.23 [1.20-1.26]* | 1.14 [1.11-1.17]* |
| I36 | 6,350 | 80,142,560 | 0.10 | 0.08 | 0.08 | 0.02 | 0.02 | 0.01 | 1.21 [1.11-1.32]* | 1.12 [1.02-1.22] |
| I37 | 6,690 | 80,134,830 | 0.10 | 0.08 | 0.08 | 0.02 | 0.02 | 0.01 | 1.26 [1.15-1.37]* | 1.17 [1.07-1.27]* |
| I38 | 9,390 | 80,079,940 | 0.15 | 0.11 | 0.11 | 0.04 | 0.03 | 0.02 | 1.30 [1.21-1.40]* | 1.20 [1.11-1.29]* |
| I40 | 710 | 80,164,370 | 0.01 | 0.01 | 0.01 | 0.00 | 0.00 | 0.00 | 1.23 [0.96-1.58] | 1.19 [0.92-1.54] |
| I42 | 22,020 | 79,937,940 | 0.33 | 0.26 | 0.27 | 0.06 | 0.06 | 0.03 | 1.23 [1.18-1.29]* | 1.12 [1.07-1.17]* |
| I43 | 920 | 80,168,950 | 0.01 | 0.01 | 0.01 | 0.00 | 0.00 | 0.00 | 1.07 [0.85-1.36] | 0.99 [0.78-1.27] |
| I44 | 139,120 | 79,258,740 | 2.09 | 1.68 | 1.74 | 0.41 | 0.36 | 0.23 | 1.20 [1.18-1.23]* | 1.13 [1.10-1.15]* |
| I45 | 86,440 | 79,590,720 | 1.30 | 1.04 | 1.08 | 0.26 | 0.22 | 0.14 | 1.21 [1.18-1.24]* | 1.12 [1.09-1.15]* |
| I46 | 44,890 | 80,041,190 | 0.67 | 0.54 | 0.54 | 0.13 | 0.13 | 0.09 | 1.24 [1.20-1.28]* | 1.15 [1.11-1.19]* |
| I47 | 49,570 | 79,507,640 | 0.74 | 0.60 | 0.61 | 0.14 | 0.13 | 0.07 | 1.22 [1.18-1.26]* | 1.11 [1.08-1.15]* |
| I48 | 387,130 | 76,982,860 | 5.92 | 4.84 | 5.09 | 1.09 | 0.83 | 0.53 | 1.16 [1.15-1.18]* | 1.10 [1.09-1.11]* |
| I49 | 59,100 | 79,625,140 | 0.87 | 0.71 | 0.73 | 0.16 | 0.14 | 0.07 | 1.19 [1.15-1.22]* | 1.09 [1.06-1.12]* |
| I50 | 293,480 | 78,465,420 | 4.65 | 3.54 | 3.69 | 1.10 | 0.95 | 0.62 | 1.26 [1.24-1.27]* | 1.15 [1.14-1.17]* |
| I51 | 162,320 | 79,188,930 | 2.50 | 1.95 | 2.00 | 0.54 | 0.50 | 0.30 | 1.25 [1.23-1.27]* | 1.14 [1.12-1.16]* |
| I60 | 9,480 | 80,048,470 | 0.13 | 0.12 | 0.11 | 0.02 | 0.02 | 0.02 | 1.16 [1.08-1.25]* | 1.13 [1.05-1.21]* |
| I61 | 23,720 | 80,021,680 | 0.34 | 0.29 | 0.29 | 0.05 | 0.05 | 0.04 | 1.17 [1.12-1.23]* | 1.14 [1.09-1.20]* |
| I62 | 13,630 | 80,089,880 | 0.20 | 0.16 | 0.17 | 0.03 | 0.03 | 0.02 | 1.18 [1.11-1.25]* | 1.13 [1.07-1.21]* |
| I63 | 112,580 | 79,364,690 | 1.56 | 1.39 | 1.43 | 0.17 | 0.13 | 0.07 | 1.09 [1.07-1.12]* | 1.05 [1.03-1.07]* |
| I64 | 29,280 | 79,830,210 | 0.41 | 0.36 | 0.36 | 0.06 | 0.05 | 0.03 | 1.14 [1.09-1.19]* | 1.09 [1.04-1.13]* |
| I65 | 27,100 | 79,907,090 | 0.39 | 0.33 | 0.34 | 0.07 | 0.06 | 0.03 | 1.17 [1.12-1.22]* | 1.09 [1.04-1.14]* |
| I66 | 1,670 | 80,158,680 | 0.02 | 0.02 | 0.02 | 0.00 | 0.00 | 0.00 | 1.14 [0.96-1.36] | 1.08 [0.91-1.29] |
| I67 | 169,440 | 79,369,040 | 2.53 | 2.05 | 2.14 | 0.48 | 0.39 | 0.25 | 1.18 [1.16-1.20]* | 1.11 [1.09-1.13]* |
| I68 | 970 | 80,169,470 | 0.02 | 0.01 | 0.01 | 0.00 | 0.00 | 0.00 | 1.24 [1.00-1.54] | 1.19 [0.95-1.49] |
| I69 | 49,620 | 79,732,260 | 0.73 | 0.60 | 0.61 | 0.13 | 0.12 | 0.07 | 1.19 [1.15-1.23]* | 1.11 [1.07-1.14]* |

| | | | | | | | | | | |
|---|---|---|---|---|---|---|---|---|---|---|
| I70 | 42,110 | 79,860,360 | 0.65 | 0.50 | 0.52 | 0.15 | 0.13 | 0.08 | 1.25 [1.21-1.30]* | 1.14 [1.10-1.18]* |
| I71 | 51,810 | 79,806,720 | 0.75 | 0.63 | 0.64 | 0.12 | 0.11 | 0.07 | 1.17 [1.13-1.21]* | 1.11 [1.07-1.14]* |
| I72 | 12,400 | 80,060,340 | 0.18 | 0.15 | 0.15 | 0.03 | 0.03 | 0.01 | 1.18 [1.11-1.25]* | 1.09 [1.02-1.16]* |
| I73 | 92,740 | 79,255,430 | 1.54 | 1.09 | 1.10 | 0.45 | 0.44 | 0.30 | 1.40 [1.37-1.43]* | 1.24 [1.21-1.27]* |
| I74 | 24,030 | 79,940,130 | 0.35 | 0.29 | 0.30 | 0.05 | 0.05 | 0.02 | 1.17 [1.12-1.22]* | 1.07 [1.02-1.12]* |
| I77 | 31,420 | 79,828,680 | 0.50 | 0.37 | 0.37 | 0.12 | 0.12 | 0.08 | 1.32 [1.27-1.37]* | 1.20 [1.15-1.24]* |
| I78 | 7,740 | 80,069,570 | 0.12 | 0.09 | 0.09 | 0.03 | 0.03 | 0.02 | 1.35 [1.25-1.45]* | 1.24 [1.14-1.34]* |
| I79 | 3,150 | 80,142,840 | 0.05 | 0.04 | 0.04 | 0.01 | 0.01 | 0.01 | 1.35 [1.20-1.52]* | 1.13 [1.00-1.28] |
| I80 | 63,680 | 79,306,270 | 0.97 | 0.77 | 0.78 | 0.20 | 0.19 | 0.13 | 1.24 [1.21-1.28]* | 1.16 [1.12-1.19]* |
| I81 | 3,590 | 80,156,140 | 0.05 | 0.04 | 0.04 | 0.01 | 0.01 | 0.00 | 1.17 [1.04-1.31]* | 1.08 [0.96-1.22] |
| I82 | 8,000 | 80,109,230 | 0.12 | 0.10 | 0.10 | 0.02 | 0.02 | 0.01 | 1.21 [1.12-1.31]* | 1.12 [1.04-1.21]* |
| I83 | 73,640 | 78,408,760 | 1.38 | 0.84 | 0.85 | 0.54 | 0.53 | 0.49 | 1.63 [1.59-1.67]* | 1.55 [1.52-1.59]* |
| I84 | 115,340 | 77,259,280 | 1.89 | 1.41 | 1.38 | 0.48 | 0.51 | 0.37 | 1.37 [1.34-1.39]* | 1.24 [1.22-1.27]* |
| I85 | 9,770 | 80,080,350 | 0.17 | 0.11 | 0.11 | 0.05 | 0.06 | 0.04 | 1.51 [1.41-1.61]* | 1.35 [1.26-1.44]* |
| I86 | 7,630 | 80,052,140 | 0.12 | 0.09 | 0.09 | 0.03 | 0.03 | 0.02 | 1.36 [1.26-1.47]* | 1.22 [1.13-1.32]* |
| I87 | 16,050 | 80,056,380 | 0.29 | 0.18 | 0.18 | 0.11 | 0.11 | 0.09 | 1.61 [1.53-1.69]* | 1.48 [1.40-1.55]* |
| I88 | 2,590 | 80,031,900 | 0.04 | 0.03 | 0.03 | 0.01 | 0.01 | 0.01 | 1.34 [1.18-1.52]* | 1.22 [1.07-1.39]* |
| I89 | 21,540 | 80,031,070 | 0.38 | 0.25 | 0.25 | 0.13 | 0.13 | 0.10 | 1.52 [1.46-1.59]* | 1.37 [1.31-1.44]* |
| I95 | 204,290 | 79,078,050 | 3.11 | 2.47 | 2.57 | 0.63 | 0.53 | 0.34 | 1.21 [1.19-1.23]* | 1.12 [1.11-1.14]* |
| I97 | 2,570 | 80,121,490 | 0.04 | 0.03 | 0.03 | 0.01 | 0.01 | 0.01 | 1.34 [1.18-1.53]* | 1.25 [1.09-1.43]* |
| I98 | 6,690 | 80,125,740 | 0.11 | 0.08 | 0.08 | 0.03 | 0.04 | 0.03 | 1.49 [1.38-1.61]* | 1.31 [1.21-1.42]* |
| I99 | 3,790 | 80,136,630 | 0.06 | 0.05 | 0.05 | 0.01 | 0.01 | 0.01 | 1.22 [1.09-1.37]* | 1.12 [1.00-1.26] |
| 18+ - J00-J99 | | | | | | | | | | |
| J00 | 4,950 | 80,036,670 | 0.08 | 0.06 | 0.06 | 0.02 | 0.02 | 0.01 | 1.35 [1.23-1.48]* | 1.22 [1.11-1.34]* |
| J01 | 2,620 | 80,115,540 | 0.04 | 0.03 | 0.03 | 0.01 | 0.01 | 0.01 | 1.38 [1.21-1.56]* | 1.18 [1.03-1.35] |
| J02 | 15,040 | 79,862,920 | 0.25 | 0.18 | 0.18 | 0.07 | 0.07 | 0.05 | 1.39 [1.31-1.46]* | 1.24 [1.17-1.31]* |
| J03 | 35,700 | 79,006,920 | 0.58 | 0.42 | 0.44 | 0.15 | 0.14 | 0.08 | 1.31 [1.27-1.36]* | 1.17 [1.13-1.21]* |
| J04 | 2,280 | 80,128,880 | 0.04 | 0.03 | 0.03 | 0.01 | 0.01 | 0.01 | 1.47 [1.28-1.68]* | 1.29 [1.12-1.48]* |
| J05 | 1,040 | 80,017,400 | 0.01 | 0.01 | 0.01 | 0.00 | 0.00 | 0.00 | 1.04 [0.84-1.28] | 0.97 [0.78-1.21] |
| J06 | 16,240 | 79,271,580 | 0.27 | 0.19 | 0.19 | 0.08 | 0.07 | 0.04 | 1.38 [1.31-1.45]* | 1.18 [1.12-1.25]* |
| J10 | 16,600 | 80,089,790 | 0.28 | 0.19 | 0.20 | 0.08 | 0.08 | 0.04 | 1.40 [1.33-1.47]* | 1.19 [1.13-1.26]* |
| J11 | 3,580 | 80,116,630 | 0.06 | 0.04 | 0.04 | 0.02 | 0.01 | 0.01 | 1.34 [1.20-1.50]* | 1.17 [1.04-1.32]* |
| J12 | 29,160 | 80,131,670 | 0.47 | 0.34 | 0.36 | 0.12 | 0.10 | 0.06 | 1.29 [1.23-1.34]* | 1.15 [1.10-1.20]* |
| J13 | 6,450 | 80,108,650 | 0.10 | 0.08 | 0.08 | 0.03 | 0.03 | 0.02 | 1.37 [1.26-1.48]* | 1.20 [1.10-1.31]* |
| J14 | 2,740 | 80,150,580 | 0.04 | 0.03 | 0.03 | 0.01 | 0.01 | 0.01 | 1.38 [1.21-1.56]* | 1.14 [0.99-1.30] |
| J15 | 13,530 | 80,056,610 | 0.23 | 0.16 | 0.16 | 0.07 | 0.07 | 0.04 | 1.42 [1.34-1.50]* | 1.24 [1.17-1.32]* |
| J17 | 2,800 | 80,151,940 | 0.05 | 0.03 | 0.03 | 0.02 | 0.02 | 0.01 | 1.49 [1.32-1.68]* | 1.34 [1.18-1.51]* |
| J18 | 409,970 | 77,814,630 | 6.52 | 5.00 | 5.14 | 1.52 | 1.38 | 0.91 | 1.27 [1.26-1.28]* | 1.16 [1.15-1.18]* |
| J20 | 4,980 | 80,109,720 | 0.08 | 0.06 | 0.06 | 0.02 | 0.02 | 0.01 | 1.28 [1.16-1.41]* | 1.12 [1.02-1.24] |
| J21 | 1,320 | 79,993,820 | 0.02 | 0.02 | 0.02 | 0.01 | 0.01 | 0.00 | 1.46 [1.22-1.75]* | 1.21 [1.00-1.46] |
| J22 | 233,090 | 77,686,310 | 3.86 | 2.81 | 2.86 | 1.05 | 1.00 | 0.67 | 1.35 [1.33-1.37]* | 1.21 [1.19-1.23]* |
| J30 | 37,490 | 79,709,390 | 0.75 | 0.41 | 0.42 | 0.34 | 0.32 | 0.18 | 1.76 [1.71-1.82]* | 1.32 [1.27-1.36]* |
| J31 | 9,050 | 79,977,450 | 0.17 | 0.10 | 0.10 | 0.07 | 0.07 | 0.04 | 1.70 [1.59-1.81]* | 1.33 [1.24-1.43]* |
| J32 | 24,430 | 79,663,000 | 0.41 | 0.28 | 0.28 | 0.13 | 0.13 | 0.07 | 1.46 [1.40-1.52]* | 1.20 [1.15-1.26]* |
| J33 | 17,240 | 79,649,340 | 0.30 | 0.20 | 0.19 | 0.10 | 0.11 | 0.06 | 1.54 [1.47-1.62]* | 1.25 [1.19-1.32]* |
| J34 | 49,820 | 78,477,910 | 0.87 | 0.58 | 0.58 | 0.29 | 0.29 | 0.19 | 1.51 [1.46-1.55]* | 1.29 [1.25-1.33]* |
| J35 | 17,950 | 78,443,410 | 0.29 | 0.22 | 0.22 | 0.07 | 0.07 | 0.04 | 1.30 [1.24-1.36]* | 1.17 [1.11-1.23]* |
| J36 | 9,820 | 79,932,360 | 0.14 | 0.12 | 0.12 | 0.02 | 0.02 | 0.01 | 1.16 [1.08-1.24]* | 1.11 [1.03-1.19]* |
| J37 | 550 | 80,154,210 | 0.01 | 0.01 | 0.01 | 0.00 | 0.00 | 0.00 | 1.40 [1.07-1.84] | 1.15 [0.86-1.53] |
| J38 | 18,780 | 79,836,750 | 0.29 | 0.22 | 0.22 | 0.07 | 0.07 | 0.04 | 1.31 [1.25-1.38]* | 1.15 [1.10-1.21]* |
| J39 | 9,340 | 80,035,450 | 0.15 | 0.11 | 0.11 | 0.04 | 0.04 | 0.02 | 1.34 [1.25-1.44]* | 1.18 [1.10-1.26]* |
| J40 | 11,790 | 80,016,140 | 0.20 | 0.14 | 0.14 | 0.06 | 0.06 | 0.04 | 1.44 [1.36-1.53]* | 1.23 [1.16-1.31]* |
| J41 | 580 | 80,165,720 | 0.01 | 0.01 | 0.01 | 0.00 | 0.00 | 0.00 | 1.38 [1.05-1.81] | 1.18 [0.88-1.57] |
| J42 | 4,760 | 80,108,380 | 0.08 | 0.06 | 0.06 | 0.02 | 0.02 | 0.01 | 1.42 [1.29-1.56]* | 1.19 [1.08-1.32]* |
| J43 | 65,760 | 79,737,350 | 1.06 | 0.77 | 0.78 | 0.28 | 0.28 | 0.16 | 1.35 [1.32-1.39]* | 1.18 [1.15-1.21]* |
| J44 | 262,180 | 77,840,340 | 4.57 | 3.11 | 3.14 | 1.46 | 1.43 | 0.77 | 1.45 [1.44-1.47]* | 1.20 [1.19-1.22]* |
| J45 | 392,250 | 70,400,710 | 9.41 | 4.74 | 4.76 | 4.67 | 4.65 | 2.40 | 1.98 [1.96-2.00]* | 1.34 [1.33-1.36]* |
| J46 | 4,510 | 79,914,140 | 0.11 | 0.04 | 0.04 | 0.07 | 0.07 | 0.04 | 2.46 [2.25-2.68]* | 1.55 [1.40-1.72]* |
| J47 | 47,260 | 79,830,350 | 0.83 | 0.54 | 0.55 | 0.29 | 0.28 | 0.17 | 1.51 [1.47-1.56]* | 1.27 [1.23-1.31]* |
| J61 | 6,540 | 80,134,890 | 0.11 | 0.08 | 0.08 | 0.03 | 0.03 | 0.02 | 1.38 [1.27-1.50]* | 1.27 [1.16-1.38]* |
| J67 | 1,920 | 80,158,270 | 0.03 | 0.02 | 0.02 | 0.01 | 0.01 | 0.01 | 1.60 [1.38-1.85]* | 1.38 [1.19-1.61]* |
| J69 | 63,850 | 80,020,380 | 0.94 | 0.77 | 0.80 | 0.18 | 0.14 | 0.10 | 1.18 [1.14-1.21]* | 1.12 [1.09-1.16]* |
| J70 | 1,300 | 80,166,240 | 0.02 | 0.02 | 0.02 | 0.01 | 0.01 | 0.00 | 1.44 [1.21-1.73]* | 1.27 [1.06-1.53] |
| J80 | 4,870 | 80,140,330 | 0.08 | 0.06 | 0.06 | 0.02 | 0.02 | 0.02 | 1.40 [1.27-1.54]* | 1.27 [1.15-1.40]* |
| J81 | 22,160 | 80,027,380 | 0.34 | 0.26 | 0.27 | 0.08 | 0.08 | 0.05 | 1.29 [1.23-1.35]* | 1.18 [1.13-1.24]* |
| J82 | 1,700 | 80,162,140 | 0.04 | 0.02 | 0.02 | 0.02 | 0.02 | 0.01 | 2.31 [2.00-2.68]* | 1.55 [1.31-1.82]* |
| J84 | 40,930 | 79,988,140 | 0.70 | 0.47 | 0.48 | 0.23 | 0.22 | 0.18 | 1.45 [1.41-1.50]* | 1.34 [1.29-1.38]* |
| J85 | 2,090 | 80,155,860 | 0.03 | 0.02 | 0.02 | 0.01 | 0.01 | 0.01 | 1.37 [1.19-1.59]* | 1.23 [1.06-1.43]* |
| J86 | 6,930 | 80,102,480 | 0.10 | 0.08 | 0.08 | 0.02 | 0.02 | 0.01 | 1.24 [1.14-1.35]* | 1.16 [1.07-1.27]* |
| J90 | 207,560 | 79,167,580 | 3.09 | 2.52 | 2.59 | 0.57 | 0.50 | 0.33 | 1.19 [1.17-1.21]* | 1.12 [1.10-1.14]* |
| J91 | 4,960 | 80,153,760 | 0.07 | 0.06 | 0.06 | 0.02 | 0.01 | 0.01 | 1.25 [1.13-1.38]* | 1.17 [1.06-1.30]* |
| J92 | 23,420 | 80,056,920 | 0.36 | 0.28 | 0.28 | 0.08 | 0.08 | 0.06 | 1.28 [1.22-1.34]* | 1.21 [1.15-1.26]* |
| J93 | 19,220 | 79,965,420 | 0.27 | 0.23 | 0.24 | 0.04 | 0.03 | 0.02 | 1.14 [1.08-1.20]* | 1.07 [1.01-1.12] |
| J94 | 7,760 | 80,126,590 | 0.11 | 0.09 | 0.10 | 0.02 | 0.02 | 0.01 | 1.18 [1.09-1.28]* | 1.10 [1.02-1.19] |
| J95 | 6,050 | 80,092,970 | 0.09 | 0.07 | 0.07 | 0.02 | 0.01 | 0.01 | 1.20 [1.10-1.31]* | 1.11 [1.02-1.22] |
| J96 | 132,870 | 79,708,820 | 2.11 | 1.57 | 1.61 | 0.54 | 0.50 | 0.30 | 1.31 [1.29-1.34]* | 1.17 [1.14-1.19]* |
| J98 | 121,570 | 79,447,590 | 1.85 | 1.46 | 1.50 | 0.39 | 0.36 | 0.19 | 1.24 [1.22-1.26]* | 1.12 [1.09-1.14]* |
| J99 | 3,720 | 80,142,050 | 0.07 | 0.04 | 0.04 | 0.03 | 0.03 | 0.02 | 1.66 [1.50-1.83]* | 1.41 [1.27-1.57]* |
| 18+ - K00-K95 | | | | | | | | | | |
| K00 | 6,740 | 79,806,690 | 0.10 | 0.08 | 0.08 | 0.01 | 0.01 | 0.00 | 1.15 [1.06-1.25]* | 1.05 [0.96-1.14] |
| K01 | 43,140 | 78,500,620 | 0.66 | 0.53 | 0.54 | 0.14 | 0.12 | 0.08 | 1.23 [1.19-1.27]* | 1.13 [1.10-1.17]* |
| K02 | 81,570 | 77,772,280 | 1.27 | 1.00 | 1.02 | 0.27 | 0.25 | 0.14 | 1.25 [1.22-1.28]* | 1.12 [1.10-1.15]* |
| K03 | 2,060 | 80,122,560 | 0.03 | 0.02 | 0.02 | 0.01 | 0.01 | 0.01 | 1.46 [1.27-1.68]* | 1.34 [1.16-1.56]* |
| K04 | 27,810 | 79,509,460 | 0.41 | 0.34 | 0.34 | 0.08 | 0.07 | 0.04 | 1.22 [1.17-1.27]* | 1.10 [1.05-1.15]* |
| K05 | 34,970 | 79,511,180 | 0.57 | 0.41 | 0.43 | 0.15 | 0.14 | 0.09 | 1.33 [1.28-1.38]* | 1.20 [1.15-1.24]* |
| K06 | 4,150 | 80,104,060 | 0.07 | 0.05 | 0.05 | 0.02 | 0.02 | 0.01 | 1.37 [1.23-1.51]* | 1.27 [1.15-1.42]* |
| K07 | 17,780 | 79,453,790 | 0.27 | 0.21 | 0.22 | 0.05 | 0.05 | 0.03 | 1.22 [1.16-1.28]* | 1.11 [1.05-1.17]* |
| K08 | 42,180 | 79,275,510 | 0.65 | 0.51 | 0.52 | 0.14 | 0.13 | 0.08 | 1.25 [1.21-1.29]* | 1.14 [1.10-1.17]* |
| K09 | 3,540 | 80,078,270 | 0.05 | 0.04 | 0.04 | 0.01 | 0.01 | 0.01 | 1.26 [1.13-1.41]* | 1.18 [1.05-1.33]* |
| K10 | 5,110 | 80,103,760 | 0.08 | 0.06 | 0.06 | 0.02 | 0.02 | 0.01 | 1.27 [1.15-1.39]* | 1.11 [1.01-1.22] |
| K11 | 10,620 | 79,961,330 | 0.17 | 0.12 | 0.12 | 0.04 | 0.04 | 0.03 | 1.36 [1.28-1.45]* | 1.25 [1.17-1.33]* |
| K12 | 15,950 | 79,997,940 | 0.25 | 0.19 | 0.19 | 0.07 | 0.06 | 0.05 | 1.33 [1.26-1.40]* | 1.22 [1.16-1.29]* |
| K13 | 22,550 | 79,730,660 | 0.37 | 0.26 | 0.27 | 0.10 | 0.10 | 0.08 | 1.38 [1.32-1.44]* | 1.26 [1.21-1.32]* |
| K14 | 11,970 | 79,984,680 | 0.19 | 0.14 | 0.14 | 0.05 | 0.06 | 0.04 | 1.40 [1.32-1.49]* | 1.24 [1.17-1.32]* |
| K20 | 79,860 | 78,715,280 | 1.27 | 0.96 | 0.96 | 0.31 | 0.31 | 0.19 | 1.32 [1.29-1.36]* | 1.18 [1.15-1.20]* |
| K21 | 307,650 | 76,400,580 | 5.22 | 3.77 | 3.82 | 1.45 | 1.39 | 0.79 | 1.36 [1.35-1.38]* | 1.18 [1.17-1.19]* |
| K22 | 118,410 | 78,566,380 | 1.86 | 1.43 | 1.43 | 0.43 | 0.43 | 0.28 | 1.30 [1.27-1.32]* | 1.18 [1.16-1.20]* |
| K23 | 1,410 | 80,162,610 | 0.02 | 0.02 | 0.02 | 0.01 | 0.01 | 0.00 | 1.50 [1.26-1.78]* | 1.23 [1.02-1.47] |
| K25 | 58,620 | 79,335,080 | 0.91 | 0.70 | 0.70 | 0.21 | 0.21 | 0.13 | 1.30 [1.26-1.33]* | 1.17 [1.14-1.20]* |
| K26 | 42,110 | 79,545,760 | 0.62 | 0.51 | 0.51 | 0.11 | 0.11 | 0.07 | 1.21 [1.17-1.25]* | 1.14 [1.10-1.18]* |
| K27 | 7,160 | 80,071,040 | 0.11 | 0.08 | 0.09 | 0.03 | 0.02 | 0.01 | 1.28 [1.19-1.39]* | 1.13 [1.04-1.22]* |
| K28 | 1,020 | 80,160,630 | 0.02 | 0.01 | 0.01 | 0.01 | 0.01 | 0.00 | 1.46 [1.19-1.79]* | 1.19 [0.96-1.47] |
| K29 | 294,590 | 75,378,480 | 4.97 | 3.68 | 3.74 | 1.29 | 1.24 | 0.78 | 1.33 [1.31-1.35]* | 1.19 [1.17-1.20]* |
| K30 | 73,260 | 78,333,000 | 1.19 | 0.88 | 0.88 | 0.31 | 0.31 | 0.20 | 1.36 [1.33-1.39]* | 1.20 [1.17-1.23]* |
| K31 | 100,920 | 79,149,690 | 1.60 | 1.21 | 1.22 | 0.39 | 0.38 | 0.21 | 1.31 [1.28-1.33]* | 1.15 [1.12-1.17]* |
| K35 | 40,380 | 78,970,850 | 0.55 | 0.50 | 0.50 | 0.05 | 0.05 | 0.02 | 1.09 [1.05-1.13]* | 1.03 [1.00-1.07] |
| K36 | 1,000 | 80,147,490 | 0.01 | 0.01 | 0.01 | 0.00 | 0.00 | 0.00 | 1.20 [0.97-1.48] | 1.11 [0.89-1.38] |

| | | | | | | | | | | |
|---|---|---|---|---|---|---|---|---|---|---|
| K37 | 7,580 | 79,951,510 | 0.11 | 0.09 | 0.09 | 0.01 | 0.01 | 0.00 | 1.13 [1.04-1.22]* | 1.01 [0.93-1.10] |
| K38 | 5,800 | 80,057,190 | 0.08 | 0.07 | 0.07 | 0.01 | 0.01 | 0.00 | 1.16 [1.06-1.27]* | 1.06 [0.97-1.16] |
| K40 | 109,720 | 77,749,940 | 1.54 | 1.38 | 1.39 | 0.16 | 0.16 | 0.11 | 1.11 [1.09-1.14]* | 1.08 [1.06-1.10]* |
| K41 | 8,290 | 80,037,240 | 0.12 | 0.10 | 0.10 | 0.02 | 0.02 | 0.01 | 1.18 [1.09-1.27]* | 1.12 [1.04-1.21]* |
| K42 | 48,410 | 79,359,750 | 0.75 | 0.58 | 0.58 | 0.17 | 0.17 | 0.09 | 1.29 [1.26-1.33]* | 1.14 [1.11-1.18]* |
| K43 | 39,560 | 79,538,540 | 0.60 | 0.47 | 0.47 | 0.13 | 0.13 | 0.07 | 1.27 [1.23-1.32]* | 1.14 [1.10-1.18]* |
| K44 | 294,760 | 75,772,100 | 4.89 | 3.67 | 3.71 | 1.22 | 1.18 | 0.73 | 1.32 [1.30-1.33]* | 1.17 [1.16-1.19]* |
| K45 | 1,920 | 80,149,920 | 0.03 | 0.02 | 0.02 | 0.01 | 0.00 | 0.00 | 1.18 [1.01-1.38] | 1.04 [0.88-1.22] |
| K46 | 7,780 | 80,113,930 | 0.12 | 0.09 | 0.09 | 0.03 | 0.03 | 0.02 | 1.34 [1.24-1.45]* | 1.16 [1.07-1.25]* |
| K50 | 20,710 | 79,672,650 | 0.38 | 0.23 | 0.23 | 0.15 | 0.15 | 0.10 | 1.65 [1.58-1.72]* | 1.35 [1.29-1.42]* |
| K51 | 30,970 | 79,504,500 | 0.54 | 0.36 | 0.36 | 0.18 | 0.18 | 0.12 | 1.51 [1.46-1.57]* | 1.29 [1.24-1.34]* |
| K52 | 155,430 | 76,793,940 | 2.60 | 1.90 | 1.88 | 0.71 | 0.73 | 0.51 | 1.39 [1.36-1.41]* | 1.24 [1.22-1.26]* |
| K55 | 25,690 | 79,993,560 | 0.39 | 0.31 | 0.31 | 0.08 | 0.08 | 0.05 | 1.25 [1.20-1.30]* | 1.14 [1.10-1.20]* |
| K56 | 76,910 | 79,455,890 | 1.15 | 0.93 | 0.95 | 0.22 | 0.20 | 0.12 | 1.21 [1.18-1.24]* | 1.11 [1.09-1.14]* |
| K57 | 337,950 | 76,374,500 | 5.37 | 4.22 | 4.30 | 1.15 | 1.07 | 0.65 | 1.25 [1.23-1.26]* | 1.14 [1.12-1.15]* |
| K58 | 94,390 | 78,979,430 | 1.69 | 1.09 | 1.11 | 0.60 | 0.58 | 0.27 | 1.52 [1.49-1.55]* | 1.19 [1.17-1.22]* |
| K59 | 294,580 | 77,471,250 | 4.78 | 3.59 | 3.71 | 1.19 | 1.07 | 0.69 | 1.29 [1.27-1.30]* | 1.17 [1.15-1.18]* |
| K60 | 27,110 | 79,475,140 | 0.48 | 0.31 | 0.31 | 0.16 | 0.17 | 0.13 | 1.54 [1.48-1.60]* | 1.38 [1.32-1.43]* |
| K61 | 16,990 | 79,752,570 | 0.27 | 0.20 | 0.20 | 0.07 | 0.08 | 0.06 | 1.39 [1.33-1.46]* | 1.26 [1.19-1.32]* |
| K62 | 173,220 | 76,810,080 | 2.83 | 2.13 | 2.12 | 0.70 | 0.71 | 0.49 | 1.34 [1.31-1.36]* | 1.21 [1.19-1.23]* |
| K63 | 176,740 | 78,390,690 | 2.74 | 2.15 | 2.16 | 0.60 | 0.58 | 0.34 | 1.27 [1.25-1.29]* | 1.14 [1.12-1.16]* |
| K64 | 125,380 | 79,678,900 | 2.00 | 1.48 | 1.53 | 0.52 | 0.48 | 0.30 | 1.31 [1.29-1.34]* | 1.18 [1.15-1.20]* |
| K65 | 18,740 | 80,011,490 | 0.27 | 0.23 | 0.23 | 0.04 | 0.04 | 0.02 | 1.18 [1.12-1.24]* | 1.10 [1.05-1.16]* |
| K66 | 66,890 | 79,435,870 | 0.99 | 0.81 | 0.82 | 0.18 | 0.16 | 0.08 | 1.20 [1.17-1.23]* | 1.09 [1.06-1.12]* |
| K70 | 22,350 | 79,924,270 | 0.37 | 0.26 | 0.25 | 0.11 | 0.12 | 0.08 | 1.46 [1.40-1.52]* | 1.29 [1.23-1.35]* |
| K71 | 1,400 | 80,156,260 | 0.02 | 0.02 | 0.02 | 0.01 | 0.01 | 0.01 | 1.40 [1.17-1.67]* | 1.28 [1.07-1.54]* |
| K72 | 13,970 | 80,118,260 | 0.22 | 0.16 | 0.16 | 0.06 | 0.06 | 0.04 | 1.35 [1.28-1.43]* | 1.22 [1.15-1.29]* |
| K73 | 1,310 | 80,143,630 | 0.02 | 0.02 | 0.01 | 0.01 | 0.01 | 0.01 | 1.55 [1.30-1.84]* | 1.41 [1.18-1.69]* |
| K74 | 20,660 | 80,017,650 | 0.35 | 0.24 | 0.24 | 0.12 | 0.12 | 0.08 | 1.49 [1.42-1.55]* | 1.31 [1.25-1.37]* |
| K75 | 13,480 | 80,048,610 | 0.22 | 0.16 | 0.16 | 0.07 | 0.07 | 0.05 | 1.42 [1.34-1.50]* | 1.26 [1.19-1.33]* |
| K76 | 99,490 | 79,488,170 | 1.60 | 1.18 | 1.20 | 0.42 | 0.40 | 0.23 | 1.34 [1.31-1.36]* | 1.17 [1.14-1.19]* |
| K80 | 190,040 | 77,339,500 | 2.83 | 2.38 | 2.43 | 0.45 | 0.41 | 0.20 | 1.17 [1.15-1.19]* | 1.08 [1.06-1.09]* |
| K81 | 34,900 | 79,627,020 | 0.51 | 0.42 | 0.43 | 0.09 | 0.08 | 0.04 | 1.19 [1.15-1.24]* | 1.09 [1.05-1.13]* |
| K82 | 26,510 | 79,826,840 | 0.39 | 0.32 | 0.33 | 0.07 | 0.06 | 0.03 | 1.19 [1.14-1.24]* | 1.09 [1.04-1.14]* |
| K83 | 49,650 | 79,717,100 | 0.72 | 0.60 | 0.62 | 0.12 | 0.10 | 0.05 | 1.17 [1.13-1.20]* | 1.08 [1.05-1.12]* |
| K85 | 32,930 | 79,701,870 | 0.49 | 0.40 | 0.40 | 0.09 | 0.09 | 0.05 | 1.21 [1.17-1.26]* | 1.11 [1.07-1.15]* |
| K86 | 27,360 | 79,934,090 | 0.41 | 0.33 | 0.33 | 0.08 | 0.08 | 0.04 | 1.24 [1.19-1.29]* | 1.12 [1.07-1.17]* |
| K90 | 20,980 | 79,831,340 | 0.39 | 0.24 | 0.24 | 0.15 | 0.14 | 0.10 | 1.60 [1.53-1.67]* | 1.36 [1.30-1.42]* |
| K91 | 24,040 | 79,877,100 | 0.38 | 0.28 | 0.29 | 0.09 | 0.09 | 0.06 | 1.32 [1.26-1.37]* | 1.18 [1.13-1.23]* |
| K92 | 216,660 | 78,072,810 | 3.45 | 2.63 | 2.66 | 0.82 | 0.79 | 0.51 | 1.30 [1.28-1.31]* | 1.17 [1.16-1.19]* |
| 18+ - L00-L99 | | | | | | | | | | |
| L01 | 1,070 | 80,125,110 | 0.03 | 0.01 | 0.01 | 0.02 | 0.02 | 0.01 | 2.65 [2.21-3.17]* | 2.19 [1.81-2.65]* |
| L02 | 42,440 | 79,323,530 | 0.70 | 0.50 | 0.50 | 0.19 | 0.19 | 0.13 | 1.38 [1.34-1.43]* | 1.23 [1.19-1.27]* |
| L03 | 160,820 | 78,260,380 | 3.02 | 1.85 | 1.83 | 1.17 | 1.18 | 1.01 | 1.64 [1.62-1.67]* | 1.50 [1.48-1.53]* |
| L04 | 1,280 | 80,128,380 | 0.02 | 0.02 | 0.02 | 0.00 | 0.00 | 0.00 | 1.29 [1.07-1.55]* | 1.21 [1.00-1.46] |
| L05 | 15,430 | 79,764,450 | 0.22 | 0.19 | 0.19 | 0.03 | 0.03 | 0.02 | 1.17 [1.11-1.24]* | 1.08 [1.02-1.14]* |
| L08 | 42,770 | 79,792,010 | 0.75 | 0.49 | 0.50 | 0.26 | 0.25 | 0.21 | 1.51 [1.46-1.56]* | 1.38 [1.34-1.43]* |
| L10 | 550 | 80,166,890 | 0.01 | 0.01 | 0.01 | 0.01 | 0.01 | 0.01 | 2.25 [1.74-2.92]* | 2.06 [1.57-2.69]* |
| L11 | 450 | 80,166,650 | 0.01 | 0.01 | 0.01 | 0.00 | 0.00 | 0.00 | 1.64 [1.21-2.22]* | 1.56 [1.15-2.13]* |
| L12 | 3,760 | 80,149,530 | 0.10 | 0.04 | 0.04 | 0.06 | 0.06 | 0.06 | 2.81 [2.54-3.11]* | 2.65 [2.39-2.93]* |
| L13 | 620 | 80,162,910 | 0.02 | 0.01 | 0.01 | 0.01 | 0.01 | 0.01 | 3.31 [2.62-4.18]* | 2.99 [2.35-3.79]* |
| L20 | 3,160 | 80,035,160 | 0.20 | 0.01 | 0.01 | 0.19 | 0.19 | 0.19 | 31.99 [27.28-37.50]* | 27.25 [23.19-32.01]* |
| L21 | 1,700 | 80,150,390 | 0.05 | 0.02 | 0.02 | 0.03 | 0.03 | 0.03 | 3.13 [2.71-3.62]* | 2.85 [2.45-3.30]* |
| L22 | 800 | 80,163,560 | 0.01 | 0.01 | 0.01 | 0.00 | 0.00 | 0.00 | 1.08 [0.83-1.40] | 1.02 [0.79-1.33] |
| L23 | 1,510 | 80,135,300 | 0.05 | 0.01 | 0.01 | 0.03 | 0.03 | 0.03 | 3.56 [3.07-4.12]* | 3.19 [2.74-3.71]* |
| L24 | 10,580 | 80,154,260 | 0.18 | 0.12 | 0.13 | 0.06 | 0.05 | 0.04 | 1.42 [1.32-1.52]* | 1.31 [1.22-1.41]* |
| L25 | 1,780 | 80,147,250 | 0.05 | 0.02 | 0.02 | 0.04 | 0.04 | 0.03 | 3.18 [2.77-3.66]* | 2.96 [2.57-3.41]* |
| L26 | 210 | 80,170,040 | 0.01 | 0.00 | 0.00 | 0.01 | 0.01 | 0.01 | 6.67 [4.43-10.04]* | 5.73 [3.75-8.75]* |
| L27 | 9,290 | 80,059,650 | 0.17 | 0.10 | 0.11 | 0.07 | 0.06 | 0.05 | 1.60 [1.50-1.71]* | 1.45 [1.36-1.56]* |
| L28 | 3,360 | 80,117,170 | 0.11 | 0.03 | 0.03 | 0.08 | 0.09 | 0.08 | 4.18 [3.79-4.61]* | 3.82 [3.45-4.22]* |
| L29 | 15,850 | 79,897,580 | 0.31 | 0.18 | 0.18 | 0.13 | 0.12 | 0.10 | 1.68 [1.60-1.76]* | 1.50 [1.42-1.58]* |
| L30 | 59,960 | 78,892,840 | 2.94 | 0.30 | 0.29 | 2.64 | 2.65 | 2.61 | 10.25 [9.99-10.52]* | 9.13 [8.89-9.38]* |
| L40 | 41,930 | 79,691,250 | 0.96 | 0.43 | 0.44 | 0.52 | 0.52 | 0.48 | 2.19 [2.13-2.26]* | 2.02 [1.96-2.08]* |
| L42 | 620 | 80,165,640 | 0.01 | 0.01 | 0.01 | 0.00 | 0.00 | 0.00 | 1.63 [1.27-2.11]* | 1.40 [1.07-1.83] |
| L43 | 4,530 | 80,097,550 | 0.10 | 0.05 | 0.05 | 0.05 | 0.05 | 0.05 | 2.15 [1.97-2.35]* | 1.98 [1.81-2.17]* |
| L50 | 6,820 | 80,038,330 | 0.13 | 0.07 | 0.08 | 0.06 | 0.06 | 0.04 | 1.78 [1.65-1.92]* | 1.45 [1.34-1.56]* |
| L51 | 1,340 | 80,134,920 | 0.03 | 0.01 | 0.01 | 0.01 | 0.01 | 0.01 | 1.83 [1.54-2.16]* | 1.60 [1.35-1.91]* |
| L52 | 930 | 80,149,370 | 0.02 | 0.01 | 0.01 | 0.01 | 0.01 | 0.00 | 1.50 [1.22-1.84]* | 1.33 [1.07-1.64] |
| L53 | 13,720 | 80,031,940 | 0.27 | 0.15 | 0.15 | 0.12 | 0.12 | 0.11 | 1.81 [1.71-1.91]* | 1.65 [1.56-1.75]* |
| L56 | 560 | 80,163,460 | 0.01 | 0.01 | 0.01 | 0.01 | 0.01 | 0.01 | 2.85 [2.23-3.63]* | 2.58 [2.01-3.31]* |
| L57 | 34,870 | 79,782,240 | 0.55 | 0.41 | 0.42 | 0.14 | 0.13 | 0.10 | 1.31 [1.26-1.35]* | 1.22 [1.18-1.27]* |
| L59 | 470 | 80,166,960 | 0.01 | 0.01 | 0.01 | 0.00 | 0.00 | 0.00 | 1.44 [1.07-1.93] | 1.41 [1.04-1.91] |
| L60 | 9,780 | 79,755,050 | 0.17 | 0.11 | 0.11 | 0.05 | 0.05 | 0.04 | 1.48 [1.39-1.58]* | 1.33 [1.24-1.42]* |
| L63 | 610 | 80,163,440 | 0.02 | 0.01 | 0.01 | 0.01 | 0.01 | 0.01 | 2.84 [2.24-3.59]* | 2.08 [1.61-2.68]* |
| L65 | 2,040 | 80,146,200 | 0.04 | 0.02 | 0.02 | 0.02 | 0.02 | 0.02 | 2.04 [1.78-2.33]* | 1.73 [1.50-1.99]* |
| L66 | 440 | 80,165,240 | 0.01 | 0.00 | 0.00 | 0.00 | 0.00 | 0.00 | 1.93 [1.44-2.57]* | 1.79 [1.33-2.42]* |
| L68 | 720 | 80,153,740 | 0.01 | 0.01 | 0.01 | 0.00 | 0.00 | 0.00 | 1.28 [1.00-1.64] | 1.12 [0.87-1.45] |
| L70 | 2,600 | 80,136,020 | 0.04 | 0.03 | 0.03 | 0.01 | 0.01 | 0.01 | 1.42 [1.25-1.61]* | 1.22 [1.07-1.39]* |
| L71 | 3,450 | 80,137,240 | 0.06 | 0.04 | 0.04 | 0.02 | 0.02 | 0.02 | 1.59 [1.43-1.78]* | 1.49 [1.33-1.66]* |
| L72 | 49,960 | 78,687,070 | 0.71 | 0.62 | 0.61 | 0.10 | 0.10 | 0.06 | 1.17 [1.14-1.21]* | 1.09 [1.06-1.12]* |
| L73 | 6,480 | 80,036,470 | 0.12 | 0.07 | 0.07 | 0.04 | 0.04 | 0.03 | 1.56 [1.44-1.68]* | 1.35 [1.24-1.46]* |
| L80 | 1,410 | 80,156,560 | 0.03 | 0.02 | 0.02 | 0.01 | 0.01 | 0.01 | 1.85 [1.57-2.18]* | 1.65 [1.40-1.96]* |
| L81 | 7,250 | 80,045,070 | 0.11 | 0.09 | 0.09 | 0.03 | 0.02 | 0.02 | 1.29 [1.19-1.39]* | 1.20 [1.11-1.30]* |
| L82 | 23,170 | 79,791,410 | 0.35 | 0.28 | 0.28 | 0.07 | 0.07 | 0.05 | 1.25 [1.19-1.31]* | 1.17 [1.12-1.22]* |
| L83 | 1,180 | 80,149,670 | 0.02 | 0.01 | 0.01 | 0.01 | 0.01 | 0.01 | 1.82 [1.51-2.18]* | 1.60 [1.32-1.93]* |
| L84 | 2,500 | 80,129,300 | 0.04 | 0.03 | 0.03 | 0.01 | 0.01 | 0.01 | 1.47 [1.29-1.67]* | 1.33 [1.16-1.51]* |
| L85 | 9,540 | 80,051,700 | 0.17 | 0.11 | 0.11 | 0.06 | 0.06 | 0.05 | 1.57 [1.47-1.68]* | 1.46 [1.37-1.57]* |
| L88 | 560 | 80,165,350 | 0.01 | 0.01 | 0.01 | 0.00 | 0.00 | 0.00 | 1.68 [1.29-2.19]* | 1.40 [1.07-1.85] |
| L89 | 121,240 | 79,869,450 | 1.84 | 1.45 | 1.52 | 0.40 | 0.32 | 0.24 | 1.21 [1.19-1.24]* | 1.15 [1.13-1.18]* |
| L90 | 22,670 | 79,644,660 | 0.36 | 0.27 | 0.27 | 0.09 | 0.09 | 0.07 | 1.33 [1.28-1.39]* | 1.24 [1.18-1.29]* |
| L91 | 7,100 | 79,940,370 | 0.11 | 0.08 | 0.08 | 0.03 | 0.03 | 0.02 | 1.35 [1.25-1.46]* | 1.26 [1.16-1.36]* |
| L92 | 5,760 | 80,039,050 | 0.09 | 0.07 | 0.07 | 0.03 | 0.03 | 0.02 | 1.40 [1.28-1.52]* | 1.26 [1.15-1.37]* |
| L93 | 2,660 | 80,135,060 | 0.05 | 0.03 | 0.03 | 0.03 | 0.03 | 0.02 | 1.86 [1.66-2.10]* | 1.61 [1.42-1.82]* |
| L94 | 1,700 | 80,144,470 | 0.03 | 0.02 | 0.02 | 0.01 | 0.01 | 0.01 | 1.55 [1.33-1.81]* | 1.41 [1.20-1.65]* |
| L95 | 2,200 | 80,144,190 | 0.04 | 0.02 | 0.02 | 0.02 | 0.02 | 0.02 | 1.78 [1.56-2.04]* | 1.63 [1.42-1.87]* |
| L97 | 86,550 | 79,669,000 | 1.53 | 0.99 | 1.00 | 0.54 | 0.53 | 0.46 | 1.53 [1.50-1.57]* | 1.42 [1.39-1.46]* |
| L98 | 68,620 | 79,064,650 | 1.13 | 0.81 | 0.81 | 0.32 | 0.32 | 0.26 | 1.40 [1.36-1.44]* | 1.30 [1.27-1.33]* |
| 18+ - M00-M99 | | | | | | | | | | |
| M00 | 7,860 | 80,065,920 | 0.13 | 0.09 | 0.09 | 0.04 | 0.04 | 0.04 | 1.50 [1.39-1.61]* | 1.37 [1.27-1.48]* |
| M02 | 1,200 | 80,149,530 | 0.02 | 0.01 | 0.01 | 0.01 | 0.01 | 0.00 | 1.42 [1.18-1.72]* | 1.29 [1.06-1.57] |
| M05 | 10,240 | 80,043,880 | 0.17 | 0.12 | 0.12 | 0.05 | 0.05 | 0.04 | 1.39 [1.30-1.48]* | 1.30 [1.22-1.39]* |
| M06 | 74,630 | 79,237,340 | 1.24 | 0.88 | 0.89 | 0.36 | 0.35 | 0.26 | 1.40 [1.36-1.43]* | 1.27 [1.23-1.30]* |
| M07 | 8,760 | 80,059,300 | 0.17 | 0.10 | 0.10 | 0.07 | 0.07 | 0.06 | 1.70 [1.59-1.82]* | 1.51 [1.41-1.61]* |

| | | | | | | | | | | |
|---|---|---|---|---|---|---|---|---|---|---|
| M08 | 780 | 80,126,530 | 0.01 | 0.01 | 0.01 | 0.00 | 0.00 | 0.00 | 1.38 [1.09-1.73]* | 1.26 [1.00-1.60] |
| M10 | 84,450 | 79,550,070 | 1.36 | 1.00 | 1.00 | 0.36 | 0.36 | 0.26 | 1.36 [1.33-1.39]* | 1.24 [1.21-1.27]* |
| M11 | 9,430 | 80,089,630 | 0.15 | 0.11 | 0.12 | 0.04 | 0.03 | 0.02 | 1.30 [1.21-1.40]* | 1.19 [1.11-1.28]* |
| M12 | 1,950 | 80,142,740 | 0.03 | 0.02 | 0.02 | 0.01 | 0.01 | 0.00 | 1.33 [1.15-1.54]* | 1.17 [1.01-1.37] |
| M13 | 212,780 | 78,075,530 | 3.49 | 2.56 | 2.63 | 0.94 | 0.87 | 0.56 | 1.33 [1.31-1.35]* | 1.19 [1.17-1.21]* |
| M14 | 2,200 | 80,146,590 | 0.04 | 0.02 | 0.02 | 0.01 | 0.02 | 0.01 | 1.62 [1.42-1.86]* | 1.33 [1.16-1.54]* |
| M15 | 78,160 | 79,437,580 | 1.28 | 0.92 | 0.93 | 0.36 | 0.35 | 0.23 | 1.37 [1.34-1.41]* | 1.22 [1.19-1.25]* |
| M16 | 131,110 | 78,280,840 | 1.86 | 1.63 | 1.66 | 0.22 | 0.20 | 0.12 | 1.12 [1.10-1.14]* | 1.07 [1.05-1.09]* |
| M17 | 211,460 | 76,849,020 | 3.33 | 2.63 | 2.66 | 0.71 | 0.67 | 0.45 | 1.25 [1.23-1.27]* | 1.16 [1.14-1.18]* |
| M18 | 11,580 | 80,019,170 | 0.19 | 0.13 | 0.14 | 0.06 | 0.06 | 0.03 | 1.42 [1.33-1.50]* | 1.22 [1.14-1.29]* |
| M19 | 329,730 | 77,326,230 | 5.56 | 3.99 | 4.08 | 1.57 | 1.47 | 0.98 | 1.36 [1.34-1.38]* | 1.21 [1.20-1.23]* |
| M20 | 47,520 | 79,034,860 | 0.74 | 0.57 | 0.57 | 0.17 | 0.17 | 0.12 | 1.30 [1.26-1.34]* | 1.20 [1.16-1.24]* |
| M21 | 25,190 | 79,820,250 | 0.38 | 0.30 | 0.31 | 0.08 | 0.07 | 0.05 | 1.24 [1.19-1.29]* | 1.14 [1.09-1.19]* |
| M22 | 8,590 | 79,905,460 | 0.14 | 0.10 | 0.10 | 0.03 | 0.03 | 0.02 | 1.35 [1.26-1.44]* | 1.18 [1.10-1.27]* |
| M23 | 93,680 | 77,788,550 | 1.40 | 1.16 | 1.14 | 0.24 | 0.26 | 0.16 | 1.22 [1.20-1.25]* | 1.13 [1.10-1.15]* |
| M24 | 34,170 | 79,538,560 | 0.53 | 0.41 | 0.41 | 0.12 | 0.12 | 0.07 | 1.29 [1.24-1.33]* | 1.16 [1.11-1.20]* |
| M25 | 206,140 | 77,261,860 | 3.34 | 2.52 | 2.57 | 0.81 | 0.77 | 0.48 | 1.30 [1.28-1.32]* | 1.17 [1.15-1.18]* |
| M30 | 910 | 80,152,640 | 0.02 | 0.01 | 0.01 | 0.01 | 0.01 | 0.00 | 1.47 [1.20-1.82]* | 1.23 [0.99-1.53] |
| M31 | 11,820 | 80,051,160 | 0.19 | 0.14 | 0.14 | 0.05 | 0.05 | 0.04 | 1.34 [1.26-1.43]* | 1.24 [1.16-1.32]* |
| M32 | 5,490 | 80,075,430 | 0.11 | 0.06 | 0.06 | 0.05 | 0.05 | 0.04 | 1.75 [1.61-1.90]* | 1.52 [1.39-1.65]* |
| M33 | 1,140 | 80,154,720 | 0.02 | 0.01 | 0.01 | 0.01 | 0.01 | 0.01 | 2.10 [1.76-2.50]* | 1.98 [1.65-2.37]* |
| M34 | 2,190 | 80,141,850 | 0.04 | 0.02 | 0.02 | 0.02 | 0.02 | 0.01 | 1.66 [1.45-1.89]* | 1.49 [1.30-1.71]* |
| M35 | 54,420 | 79,679,330 | 0.93 | 0.63 | 0.64 | 0.30 | 0.28 | 0.22 | 1.44 [1.40-1.49]* | 1.31 [1.27-1.34]* |
| M36 | 450 | 80,164,480 | 0.01 | 0.01 | 0.01 | 0.00 | 0.00 | 0.00 | 1.41 [1.03-1.93] | 1.27 [0.92-1.76] |
| M40 | 13,340 | 80,101,810 | 0.20 | 0.16 | 0.16 | 0.05 | 0.04 | 0.02 | 1.24 [1.17-1.32]* | 1.14 [1.07-1.21]* |
| M41 | 34,460 | 79,855,970 | 0.53 | 0.41 | 0.42 | 0.12 | 0.11 | 0.07 | 1.27 [1.23-1.32]* | 1.16 [1.12-1.21]* |
| M42 | 690 | 80,163,660 | 0.01 | 0.01 | 0.01 | 0.00 | 0.00 | 0.00 | 1.22 [0.94-1.57] | 1.01 [0.77-1.32] |
| M43 | 29,950 | 79,837,360 | 0.47 | 0.36 | 0.36 | 0.11 | 0.11 | 0.06 | 1.29 [1.24-1.34]* | 1.14 [1.09-1.19]* |
| M45 | 7,210 | 80,067,950 | 0.13 | 0.08 | 0.08 | 0.04 | 0.05 | 0.04 | 1.56 [1.45-1.69]* | 1.39 [1.29-1.50]* |
| M46 | 24,580 | 79,925,120 | 0.41 | 0.29 | 0.29 | 0.12 | 0.12 | 0.07 | 1.41 [1.36-1.48]* | 1.22 [1.17-1.28]* |
| M47 | 140,280 | 78,738,700 | 2.30 | 1.67 | 1.69 | 0.63 | 0.61 | 0.37 | 1.36 [1.34-1.38]* | 1.19 [1.17-1.21]* |
| M48 | 82,570 | 79,423,300 | 1.29 | 0.98 | 1.00 | 0.31 | 0.29 | 0.17 | 1.29 [1.26-1.32]* | 1.15 [1.12-1.17]* |
| M49 | 2,980 | 80,158,680 | 0.04 | 0.04 | 0.04 | 0.00 | 0.00 | 0.00 | 1.07 [0.94-1.21] | 1.03 [0.90-1.17] |
| M50 | 21,750 | 79,927,330 | 0.34 | 0.26 | 0.26 | 0.09 | 0.08 | 0.04 | 1.32 [1.26-1.38]* | 1.14 [1.09-1.20]* |
| M51 | 105,430 | 78,739,650 | 1.66 | 1.27 | 1.28 | 0.40 | 0.39 | 0.19 | 1.30 [1.27-1.33]* | 1.13 [1.11-1.15]* |
| M53 | 6,410 | 80,067,990 | 0.10 | 0.08 | 0.08 | 0.02 | 0.02 | 0.01 | 1.27 [1.17-1.38]* | 1.08 [0.99-1.18] |
| M54 | 258,750 | 76,320,770 | 4.25 | 3.20 | 3.28 | 1.05 | 0.98 | 0.50 | 1.30 [1.28-1.32]* | 1.13 [1.12-1.15]* |
| M60 | 4,300 | 80,113,880 | 0.07 | 0.05 | 0.05 | 0.02 | 0.02 | 0.02 | 1.48 [1.34-1.63]* | 1.36 [1.23-1.51]* |
| M62 | 20,940 | 79,964,450 | 0.32 | 0.25 | 0.25 | 0.07 | 0.07 | 0.04 | 1.26 [1.20-1.33]* | 1.16 [1.10-1.22]* |
| M65 | 42,540 | 79,378,090 | 0.68 | 0.50 | 0.50 | 0.18 | 0.18 | 0.12 | 1.35 [1.31-1.39]* | 1.21 [1.17-1.25]* |
| M66 | 4,600 | 80,091,610 | 0.07 | 0.05 | 0.05 | 0.02 | 0.02 | 0.01 | 1.31 [1.18-1.44]* | 1.21 [1.09-1.34]* |
| M67 | 28,010 | 79,452,230 | 0.43 | 0.34 | 0.33 | 0.10 | 0.10 | 0.07 | 1.30 [1.25-1.35]* | 1.19 [1.14-1.24]* |
| M70 | 17,700 | 79,888,590 | 0.30 | 0.21 | 0.20 | 0.09 | 0.09 | 0.06 | 1.44 [1.37-1.52]* | 1.28 [1.21-1.34]* |
| M71 | 11,350 | 80,028,530 | 0.18 | 0.13 | 0.14 | 0.05 | 0.04 | 0.03 | 1.33 [1.25-1.41]* | 1.20 [1.13-1.28]* |
| M72 | 25,760 | 79,685,080 | 0.35 | 0.32 | 0.31 | 0.03 | 0.04 | 0.02 | 1.12 [1.07-1.17]* | 1.07 [1.02-1.11]* |
| M75 | 72,230 | 78,880,850 | 1.15 | 0.87 | 0.87 | 0.28 | 0.28 | 0.16 | 1.32 [1.29-1.35]* | 1.17 [1.14-1.20]* |
| M76 | 5,820 | 80,094,420 | 0.10 | 0.07 | 0.07 | 0.03 | 0.03 | 0.02 | 1.41 [1.29-1.53]* | 1.23 [1.12-1.34]* |
| M77 | 14,060 | 79,903,840 | 0.24 | 0.16 | 0.16 | 0.07 | 0.08 | 0.05 | 1.48 [1.40-1.56]* | 1.29 [1.22-1.36]* |
| M79 | 209,720 | 77,215,060 | 3.61 | 2.52 | 2.57 | 1.09 | 1.04 | 0.66 | 1.41 [1.39-1.43]* | 1.22 [1.21-1.24]* |
| M80 | 26,210 | 79,976,240 | 0.41 | 0.31 | 0.32 | 0.10 | 0.09 | 0.06 | 1.29 [1.23-1.34]* | 1.18 [1.13-1.23]* |
| M81 | 159,490 | 78,992,300 | 2.57 | 1.90 | 1.98 | 0.67 | 0.59 | 0.38 | 1.30 [1.28-1.32]* | 1.17 [1.15-1.19]* |
| M83 | 1,040 | 80,158,720 | 0.02 | 0.01 | 0.01 | 0.00 | 0.00 | 0.00 | 1.23 [1.00-1.52] | 1.13 [0.91-1.39] |
| M84 | 21,980 | 79,809,540 | 0.33 | 0.26 | 0.26 | 0.07 | 0.07 | 0.04 | 1.25 [1.19-1.31]* | 1.14 [1.09-1.20]* |
| M85 | 34,610 | 79,961,330 | 0.56 | 0.41 | 0.42 | 0.15 | 0.13 | 0.08 | 1.32 [1.27-1.37]* | 1.17 [1.13-1.22]* |
| M86 | 14,680 | 80,015,070 | 0.24 | 0.17 | 0.17 | 0.06 | 0.07 | 0.05 | 1.38 [1.31-1.46]* | 1.25 [1.18-1.32]* |
| M87 | 6,690 | 80,072,680 | 0.10 | 0.08 | 0.08 | 0.02 | 0.02 | 0.01 | 1.27 [1.17-1.38]* | 1.16 [1.07-1.26]* |
| M88 | 4,480 | 80,098,880 | 0.06 | 0.05 | 0.05 | 0.01 | 0.01 | 0.01 | 1.15 [1.03-1.28] | 1.11 [0.99-1.24] |
| M89 | 16,290 | 79,858,610 | 0.26 | 0.19 | 0.20 | 0.06 | 0.06 | 0.04 | 1.30 [1.24-1.37]* | 1.17 [1.11-1.24]* |
| M90 | 7,220 | 80,148,000 | 0.09 | 0.09 | 0.09 | 0.00 | 0.00 | 0.00 | 1.03 [0.95-1.13] | 1.02 [0.94-1.11] |
| M91 | 700 | 80,143,080 | 0.01 | 0.01 | 0.01 | 0.00 | 0.00 | 0.00 | 1.27 [0.99-1.64] | 1.21 [0.93-1.56] |
| M92 | 1,740 | 80,130,940 | 0.03 | 0.02 | 0.02 | 0.01 | 0.01 | 0.01 | 1.43 [1.22-1.67]* | 1.27 [1.08-1.49]* |
| M93 | 3,450 | 80,070,960 | 0.05 | 0.04 | 0.04 | 0.01 | 0.01 | 0.01 | 1.30 [1.16-1.46]* | 1.15 [1.02-1.29] |
| M94 | 12,150 | 79,985,750 | 0.20 | 0.14 | 0.14 | 0.06 | 0.06 | 0.04 | 1.41 [1.33-1.50]* | 1.21 [1.14-1.28]* |
| M95 | 3,020 | 80,069,300 | 0.05 | 0.04 | 0.03 | 0.01 | 0.01 | 0.01 | 1.34 [1.19-1.51]* | 1.20 [1.06-1.35]* |
| M96 | 8,700 | 80,087,970 | 0.14 | 0.10 | 0.11 | 0.04 | 0.03 | 0.02 | 1.31 [1.21-1.41]* | 1.18 [1.10-1.28]* |
| M99 | 2,480 | 80,151,010 | 0.04 | 0.03 | 0.03 | 0.01 | 0.01 | 0.00 | 1.32 [1.15-1.51]* | 1.13 [0.98-1.30] |
| 18+ - N00-N99 | | | | | | | | | | |
| N02 | 6,690 | 80,036,670 | 0.11 | 0.08 | 0.08 | 0.03 | 0.03 | 0.02 | 1.36 [1.25-1.47]* | 1.28 [1.17-1.38]* |
| N03 | 26,900 | 79,962,920 | 0.43 | 0.32 | 0.32 | 0.11 | 0.11 | 0.08 | 1.34 [1.29-1.40]* | 1.22 [1.17-1.27]* |
| N04 | 3,680 | 80,110,780 | 0.06 | 0.04 | 0.04 | 0.02 | 0.02 | 0.02 | 1.50 [1.35-1.67]* | 1.37 [1.23-1.52]* |
| N05 | 3,720 | 80,105,790 | 0.06 | 0.04 | 0.04 | 0.02 | 0.02 | 0.02 | 1.49 [1.34-1.66]* | 1.36 [1.22-1.51]* |
| N08 | 12,160 | 80,066,520 | 0.21 | 0.14 | 0.14 | 0.06 | 0.06 | 0.04 | 1.45 [1.36-1.53]* | 1.26 [1.18-1.33]* |
| N10 | 5,120 | 80,082,160 | 0.07 | 0.06 | 0.06 | 0.01 | 0.01 | 0.00 | 1.18 [1.07-1.29]* | 1.04 [0.94-1.15] |
| N11 | 2,520 | 80,122,870 | 0.04 | 0.03 | 0.03 | 0.01 | 0.01 | 0.00 | 1.24 [1.08-1.41]* | 1.14 [1.00-1.31] |
| N12 | 29,140 | 79,734,820 | 0.43 | 0.35 | 0.36 | 0.08 | 0.07 | 0.03 | 1.21 [1.16-1.25]* | 1.07 [1.03-1.12]* |
| N13 | 64,780 | 79,598,450 | 0.87 | 0.80 | 0.81 | 0.07 | 0.06 | 0.03 | 1.07 [1.04-1.10]* | 1.03 [1.01-1.06] |
| N14 | 930 | 80,166,550 | 0.01 | 0.01 | 0.01 | 0.00 | 0.00 | 0.00 | 1.31 [1.05-1.63] | 1.20 [0.96-1.51] |
| N15 | 1,670 | 80,153,520 | 0.02 | 0.02 | 0.02 | 0.00 | 0.00 | 0.00 | 1.21 [1.03-1.43] | 1.08 [0.91-1.29] |
| N16 | 820 | 80,162,670 | 0.01 | 0.01 | 0.01 | 0.01 | 0.01 | 0.01 | 1.65 [1.32-2.06]* | 1.51 [1.20-1.89]* |
| N17 | 356,820 | 78,959,680 | 5.47 | 4.32 | 4.48 | 1.15 | 0.99 | 0.65 | 1.22 [1.21-1.23]* | 1.13 [1.12-1.15]* |
| N18 | 299,100 | 78,798,540 | 4.66 | 3.61 | 3.70 | 1.06 | 0.96 | 0.62 | 1.26 [1.24-1.28]* | 1.15 [1.14-1.17]* |
| N19 | 43,440 | 79,796,710 | 0.68 | 0.52 | 0.51 | 0.16 | 0.16 | 0.12 | 1.31 [1.27-1.36]* | 1.21 [1.17-1.25]* |
| N20 | 54,130 | 79,281,100 | 0.77 | 0.66 | 0.67 | 0.10 | 0.10 | 0.05 | 1.15 [1.12-1.19]* | 1.07 [1.04-1.10]* |
| N21 | 13,440 | 80,001,560 | 0.17 | 0.17 | 0.17 | 0.00 | 0.00 | 0.00 | 1.01 [0.95-1.08] | 0.98 [0.92-1.05] |
| N23 | 12,520 | 79,789,240 | 0.18 | 0.15 | 0.15 | 0.03 | 0.03 | 0.01 | 1.19 [1.12-1.26]* | 1.06 [1.00-1.13] |
| N25 | 2,500 | 80,144,100 | 0.04 | 0.03 | 0.03 | 0.01 | 0.01 | 0.01 | 1.39 [1.22-1.59]* | 1.29 [1.12-1.48]* |
| N26 | 4,990 | 80,141,950 | 0.07 | 0.06 | 0.06 | 0.01 | 0.01 | 0.01 | 1.18 [1.07-1.30]* | 1.07 [0.97-1.19] |
| N27 | 2,460 | 80,144,840 | 0.04 | 0.03 | 0.03 | 0.01 | 0.01 | 0.01 | 1.34 [1.17-1.53]* | 1.25 [1.09-1.43]* |
| N28 | 71,180 | 79,625,890 | 1.05 | 0.86 | 0.88 | 0.19 | 0.17 | 0.09 | 1.19 [1.16-1.22]* | 1.10 [1.07-1.13]* |
| N29 | 520 | 80,165,150 | 0.01 | 0.01 | 0.01 | 0.00 | 0.00 | 0.00 | 1.48 [1.11-1.97]* | 1.35 [1.01-1.82] |
| N30 | 33,380 | 79,610,190 | 0.51 | 0.40 | 0.40 | 0.11 | 0.11 | 0.07 | 1.27 [1.22-1.31]* | 1.17 [1.12-1.21]* |
| N31 | 9,300 | 79,955,110 | 0.15 | 0.11 | 0.11 | 0.04 | 0.04 | 0.03 | 1.39 [1.30-1.48]* | 1.20 [1.12-1.28]* |
| N32 | 96,620 | 78,669,860 | 1.47 | 1.18 | 1.20 | 0.29 | 0.27 | 0.17 | 1.22 [1.20-1.25]* | 1.13 [1.10-1.16]* |
| N34 | 1,460 | 80,134,840 | 0.02 | 0.02 | 0.02 | 0.01 | 0.01 | 0.00 | 1.34 [1.13-1.59]* | 1.25 [1.05-1.49] |
| N35 | 32,380 | 79,524,340 | 0.51 | 0.39 | 0.39 | 0.12 | 0.12 | 0.09 | 1.31 [1.26-1.36]* | 1.21 [1.16-1.26]* |
| N36 | 8,810 | 80,010,340 | 0.13 | 0.10 | 0.10 | 0.03 | 0.03 | 0.02 | 1.29 [1.20-1.38]* | 1.21 [1.12-1.30]* |
| N39 | 423,010 | 75,628,220 | 6.71 | 5.35 | 5.59 | 1.37 | 1.13 | 0.71 | 1.20 [1.19-1.21]* | 1.12 [1.11-1.13]* |
| N40 | 143,600 | 78,410,740 | 2.16 | 1.76 | 1.86 | 0.40 | 0.30 | 0.20 | 1.16 [1.14-1.18]* | 1.10 [1.08-1.12]* |
| N41 | 10,600 | 79,986,600 | 0.14 | 0.13 | 0.13 | 0.01 | 0.02 | 0.01 | 1.14 [1.07-1.22]* | 1.08 [1.00-1.15] |
| N42 | 13,660 | 79,962,430 | 0.19 | 0.17 | 0.17 | 0.02 | 0.02 | 0.01 | 1.11 [1.05-1.18]* | 1.06 [1.00-1.13] |
| N43 | 13,510 | 79,848,640 | 0.19 | 0.16 | 0.16 | 0.03 | 0.03 | 0.03 | 1.21 [1.14-1.28]* | 1.16 [1.09-1.23]* |
| N44 | 1,390 | 80,082,730 | 0.02 | 0.02 | 0.02 | 0.00 | 0.00 | 0.00 | 1.10 [0.92-1.32] | 1.02 [0.85-1.24] |
| N45 | 12,340 | 79,930,080 | 0.18 | 0.15 | 0.15 | 0.04 | 0.04 | 0.02 | 1.26 [1.18-1.33]* | 1.15 [1.08-1.22]* |
| N47 | 29,840 | 79,260,330 | 0.49 | 0.35 | 0.34 | 0.14 | 0.15 | 0.12 | 1.43 [1.38-1.48]* | 1.32 [1.27-1.38]* |
| N48 | 20,130 | 79,733,710 | 0.33 | 0.24 | 0.23 | 0.09 | 0.10 | 0.08 | 1.44 [1.37-1.50]* | 1.32 [1.26-1.39]* |
| N49 | 3,110 | 80,123,630 | 0.05 | 0.04 | 0.04 | 0.01 | 0.01 | 0.01 | 1.38 [1.23-1.56]* | 1.27 [1.13-1.44]* |
| N50 | 24,000 | 79,606,140 | 0.35 | 0.29 | 0.28 | 0.06 | 0.07 | 0.05 | 1.25 [1.20-1.31]* | 1.16 [1.11-1.21]* |

| | | | | | | | | | | |
|---|---|---|---|---|---|---|---|---|---|---|
| N60 | 10,740 | 79,865,610 | 0.15 | 0.13 | 0.13 | 0.02 | 0.02 | 0.01 | 1.14 [1.07-1.22]* | 1.09 [1.02-1.16] |
| N61 | 6,870 | 80,012,930 | 0.11 | 0.08 | 0.08 | 0.03 | 0.03 | 0.02 | 1.38 [1.27-1.49]* | 1.24 [1.14-1.35]* |
| N62 | 5,790 | 79,967,830 | 0.09 | 0.07 | 0.07 | 0.02 | 0.02 | 0.01 | 1.28 [1.17-1.39]* | 1.15 [1.05-1.26]* |
| N63 | 11,310 | 79,859,760 | 0.16 | 0.14 | 0.14 | 0.02 | 0.02 | 0.01 | 1.17 [1.09-1.24]* | 1.09 [1.02-1.16] |
| N64 | 10,770 | 79,941,660 | 0.16 | 0.13 | 0.13 | 0.03 | 0.03 | 0.02 | 1.25 [1.17-1.33]* | 1.15 [1.07-1.23]* |
| N70 | 7,730 | 79,983,400 | 0.11 | 0.09 | 0.10 | 0.01 | 0.01 | 0.00 | 1.08 [1.00-1.17] | 1.01 [0.93-1.09] |
| N71 | 3,730 | 80,084,160 | 0.06 | 0.04 | 0.05 | 0.01 | 0.01 | 0.01 | 1.23 [1.10-1.37]* | 1.12 [1.01-1.26] |
| N72 | 7,410 | 79,893,680 | 0.10 | 0.09 | 0.09 | 0.01 | 0.01 | 0.01 | 1.12 [1.04-1.21]* | 1.06 [0.98-1.15] |
| N73 | 41,930 | 79,139,910 | 0.61 | 0.51 | 0.53 | 0.10 | 0.08 | 0.03 | 1.16 [1.12-1.20]* | 1.06 [1.02-1.09]* |
| N75 | 7,320 | 79,963,230 | 0.10 | 0.09 | 0.09 | 0.01 | 0.00 | 0.00 | 1.04 [0.96-1.13] | 0.99 [0.91-1.07] |
| N76 | 9,810 | 79,914,850 | 0.17 | 0.11 | 0.12 | 0.05 | 0.05 | 0.04 | 1.44 [1.35-1.53]* | 1.29 [1.21-1.38]* |
| N77 | 13,250 | 79,919,600 | 0.21 | 0.16 | 0.16 | 0.05 | 0.05 | 0.02 | 1.30 [1.23-1.38]* | 1.13 [1.06-1.20]* |
| N80 | 41,990 | 79,084,600 | 0.61 | 0.51 | 0.53 | 0.09 | 0.08 | 0.02 | 1.15 [1.11-1.18]* | 1.04 [1.00-1.07] |
| N81 | 68,190 | 78,616,600 | 1.02 | 0.83 | 0.85 | 0.18 | 0.17 | 0.11 | 1.20 [1.17-1.23]* | 1.13 [1.10-1.16]* |
| N82 | 3,220 | 80,127,410 | 0.05 | 0.04 | 0.04 | 0.01 | 0.01 | 0.01 | 1.32 [1.17-1.48]* | 1.21 [1.07-1.36]* |
| N83 | 66,730 | 78,657,570 | 0.97 | 0.82 | 0.85 | 0.15 | 0.13 | 0.04 | 1.15 [1.12-1.18]* | 1.04 [1.01-1.07]* |
| N84 | 56,920 | 78,770,070 | 0.80 | 0.71 | 0.71 | 0.10 | 0.09 | 0.07 | 1.13 [1.09-1.16]* | 1.09 [1.06-1.12]* |
| N85 | 44,850 | 79,242,320 | 0.66 | 0.55 | 0.56 | 0.12 | 0.11 | 0.07 | 1.19 [1.16-1.23]* | 1.11 [1.08-1.15]* |
| N86 | 13,370 | 79,777,460 | 0.20 | 0.16 | 0.17 | 0.04 | 0.03 | 0.02 | 1.20 [1.13-1.27]* | 1.12 [1.05-1.19]* |
| N87 | 16,420 | 79,444,200 | 0.23 | 0.20 | 0.20 | 0.03 | 0.03 | 0.01 | 1.13 [1.07-1.19]* | 1.06 [1.00-1.12] |
| N88 | 13,410 | 79,863,860 | 0.20 | 0.16 | 0.16 | 0.04 | 0.03 | 0.03 | 1.21 [1.14-1.28]* | 1.15 [1.08-1.22]* |
| N89 | 21,720 | 79,582,710 | 0.34 | 0.26 | 0.26 | 0.08 | 0.08 | 0.05 | 1.30 [1.25-1.36]* | 1.18 [1.13-1.24]* |
| N90 | 20,520 | 79,704,310 | 0.34 | 0.24 | 0.25 | 0.10 | 0.09 | 0.07 | 1.37 [1.31-1.43]* | 1.26 [1.20-1.32]* |
| N91 | 2,220 | 80,103,500 | 0.03 | 0.03 | 0.03 | 0.01 | 0.01 | 0.00 | 1.30 [1.13-1.49]* | 1.14 [0.99-1.32] |
| N92 | 90,160 | 77,478,560 | 1.39 | 1.11 | 1.15 | 0.27 | 0.24 | 0.13 | 1.21 [1.18-1.24]* | 1.10 [1.08-1.13]* |
| N93 | 41,760 | 78,933,430 | 0.63 | 0.51 | 0.52 | 0.12 | 0.11 | 0.05 | 1.21 [1.17-1.25]* | 1.10 [1.06-1.13]* |
| N94 | 33,220 | 79,027,970 | 0.52 | 0.40 | 0.41 | 0.12 | 0.12 | 0.06 | 1.28 [1.24-1.33]* | 1.13 [1.09-1.17]* |
| N95 | 48,690 | 78,929,200 | 0.75 | 0.59 | 0.60 | 0.16 | 0.15 | 0.11 | 1.25 [1.21-1.29]* | 1.18 [1.15-1.22]* |
| N96 | 710 | 80,157,340 | 0.01 | 0.01 | 0.01 | 0.00 | 0.00 | 0.00 | 1.11 [0.85-1.44] | 1.07 [0.81-1.41] |
| N97 | 7,810 | 79,616,230 | 0.09 | 0.10 | 0.10 | −0.01 | −0.01 | −0.01 | 0.95 [0.87-1.03] | 0.93 [0.86-1.01] |
| N98 | 970 | 80,143,350 | 0.01 | 0.01 | 0.01 | 0.00 | 0.00 | 0.00 | 0.81 [0.64-1.04] | 0.76 [0.59-0.98] |
| N99 | 11,530 | 79,948,240 | 0.17 | 0.14 | 0.14 | 0.04 | 0.04 | 0.03 | 1.25 [1.18-1.33]* | 1.17 [1.10-1.25]* |
| 18+ - O00-O9A | | | | | | | | | | |
| O00 | 13,100 | 79,801,330 | 0.18 | 0.16 | 0.17 | 0.02 | 0.02 | 0.01 | 1.09 [1.03-1.16]* | 1.03 [0.97-1.10] |
| O01 | 790 | 80,149,540 | 0.01 | 0.01 | 0.01 | 0.00 | 0.00 | 0.00 | 1.18 [0.93-1.51] | 1.17 [0.91-1.50] |
| O02 | 30,130 | 79,092,520 | 0.42 | 0.37 | 0.38 | 0.05 | 0.04 | 0.03 | 1.11 [1.06-1.15]* | 1.07 [1.03-1.11]* |
| O03 | 45,160 | 78,658,480 | 0.63 | 0.56 | 0.59 | 0.07 | 0.04 | 0.02 | 1.07 [1.04-1.11]* | 1.03 [1.00-1.07] |
| O04 | 53,100 | 78,170,760 | 0.74 | 0.67 | 0.71 | 0.08 | 0.03 | 0.00 | 1.04 [1.01-1.07]* | 0.99 [0.96-1.03] |
| O08 | 4,680 | 80,056,880 | 0.06 | 0.06 | 0.06 | 0.01 | 0.01 | 0.00 | 1.09 [0.98-1.20] | 1.01 [0.91-1.12] |
| O10 | 4,860 | 80,062,620 | 0.07 | 0.06 | 0.06 | 0.01 | 0.01 | 0.00 | 1.16 [1.05-1.28]* | 1.07 [0.97-1.19] |
| O11 | 680 | 80,155,860 | 0.01 | 0.01 | 0.01 | 0.00 | 0.00 | 0.00 | 1.26 [0.98-1.63] | 1.16 [0.88-1.52] |
| O12 | 9,940 | 79,813,350 | 0.15 | 0.12 | 0.13 | 0.03 | 0.02 | 0.01 | 1.15 [1.08-1.23]* | 1.06 [0.99-1.14] |
| O13 | 27,630 | 79,231,360 | 0.39 | 0.34 | 0.35 | 0.05 | 0.04 | 0.03 | 1.11 [1.07-1.16]* | 1.08 [1.04-1.13]* |
| O14 | 20,080 | 79,521,210 | 0.29 | 0.24 | 0.26 | 0.04 | 0.03 | 0.02 | 1.12 [1.07-1.18]* | 1.09 [1.03-1.14]* |
| O15 | 780 | 80,146,950 | 0.01 | 0.01 | 0.01 | 0.00 | 0.00 | 0.00 | 1.12 [0.88-1.44] | 1.08 [0.83-1.39] |
| O16 | 22,430 | 79,198,340 | 0.32 | 0.28 | 0.29 | 0.04 | 0.04 | 0.03 | 1.12 [1.07-1.18]* | 1.08 [1.03-1.14]* |
| O20 | 39,250 | 78,815,340 | 0.56 | 0.48 | 0.51 | 0.08 | 0.05 | 0.02 | 1.10 [1.06-1.14]* | 1.03 [1.00-1.07] |
| O21 | 27,820 | 79,355,160 | 0.41 | 0.34 | 0.36 | 0.07 | 0.06 | 0.02 | 1.16 [1.11-1.21]* | 1.05 [1.00-1.09] |
| O22 | 4,630 | 80,063,430 | 0.06 | 0.06 | 0.06 | 0.01 | 0.01 | 0.00 | 1.10 [1.00-1.22] | 1.07 [0.96-1.19] |
| O23 | 30,190 | 79,217,370 | 0.46 | 0.36 | 0.38 | 0.10 | 0.08 | 0.04 | 1.20 [1.15-1.24]* | 1.09 [1.05-1.14]* |
| O24 | 37,780 | 79,520,960 | 0.54 | 0.46 | 0.49 | 0.07 | 0.05 | 0.03 | 1.09 [1.06-1.13]* | 1.07 [1.03-1.11]* |
| O26 | 112,960 | 76,196,660 | 1.81 | 1.41 | 1.52 | 0.40 | 0.29 | 0.16 | 1.19 [1.17-1.22]* | 1.10 [1.07-1.12]* |
| O28 | 11,600 | 79,969,550 | 0.16 | 0.14 | 0.15 | 0.02 | 0.01 | 0.00 | 1.09 [1.02-1.16]* | 1.01 [0.94-1.08] |
| O30 | 14,380 | 79,793,260 | 0.19 | 0.18 | 0.18 | 0.01 | 0.00 | 0.00 | 1.01 [0.96-1.08] | 0.98 [0.92-1.04] |
| O31 | 780 | 80,151,690 | 0.01 | 0.01 | 0.01 | 0.00 | 0.00 | 0.00 | 1.10 [0.86-1.41] | 1.01 [0.78-1.31] |
| O32 | 56,540 | 78,630,390 | 0.79 | 0.70 | 0.74 | 0.08 | 0.04 | 0.03 | 1.06 [1.03-1.09]* | 1.04 [1.01-1.07] |
| O33 | 1,350 | 80,103,240 | 0.02 | 0.02 | 0.02 | 0.00 | 0.00 | 0.00 | 1.19 [0.99-1.44] | 1.16 [0.96-1.40] |
| O34 | 93,700 | 77,911,470 | 1.36 | 1.17 | 1.23 | 0.19 | 0.13 | 0.10 | 1.10 [1.08-1.13]* | 1.08 [1.06-1.11]* |
| O35 | 13,960 | 79,854,480 | 0.19 | 0.17 | 0.18 | 0.02 | 0.01 | 0.01 | 1.06 [1.00-1.13] | 1.03 [0.97-1.10] |
| O36 | 151,870 | 76,116,270 | 2.42 | 1.90 | 2.09 | 0.51 | 0.32 | 0.19 | 1.15 [1.13-1.17]* | 1.09 [1.07-1.11]* |
| O40 | 12,940 | 79,935,190 | 0.18 | 0.16 | 0.17 | 0.03 | 0.02 | 0.01 | 1.10 [1.04-1.17]* | 1.04 [0.98-1.11] |
| O41 | 17,210 | 79,621,930 | 0.24 | 0.21 | 0.22 | 0.03 | 0.03 | 0.02 | 1.12 [1.06-1.18]* | 1.08 [1.02-1.14]* |
| O42 | 92,130 | 77,616,700 | 1.32 | 1.16 | 1.24 | 0.16 | 0.07 | 0.06 | 1.06 [1.04-1.08]* | 1.04 [1.02-1.07]* |
| O43 | 12,820 | 79,767,780 | 0.17 | 0.16 | 0.16 | 0.02 | 0.01 | 0.00 | 1.06 [0.99-1.12] | 1.03 [0.97-1.10] |
| O44 | 8,680 | 79,938,290 | 0.12 | 0.11 | 0.11 | 0.01 | 0.00 | 0.00 | 1.04 [0.96-1.12] | 0.99 [0.92-1.07] |
| O45 | 3,850 | 80,063,590 | 0.05 | 0.05 | 0.05 | 0.00 | 0.00 | 0.00 | 1.05 [0.94-1.17] | 1.01 [0.90-1.14] |
| O46 | 42,510 | 78,789,400 | 0.63 | 0.52 | 0.54 | 0.11 | 0.08 | 0.05 | 1.16 [1.12-1.19]* | 1.09 [1.06-1.13]* |
| O47 | 29,930 | 78,234,860 | 0.44 | 0.37 | 0.38 | 0.07 | 0.07 | 0.04 | 1.17 [1.13-1.22]* | 1.10 [1.06-1.14]* |
| O48 | 56,850 | 78,046,270 | 0.77 | 0.72 | 0.75 | 0.06 | 0.02 | 0.02 | 1.03 [1.00-1.06] | 1.03 [1.00-1.06] |
| O60 | 47,310 | 78,791,130 | 0.68 | 0.58 | 0.61 | 0.10 | 0.07 | 0.04 | 1.11 [1.07-1.14]* | 1.05 [1.02-1.09]* |
| O61 | 9,080 | 79,882,130 | 0.13 | 0.11 | 0.11 | 0.02 | 0.01 | 0.01 | 1.11 [1.03-1.19]* | 1.06 [0.98-1.14] |
| O62 | 36,040 | 79,234,370 | 0.51 | 0.44 | 0.47 | 0.06 | 0.04 | 0.03 | 1.08 [1.04-1.12]* | 1.06 [1.02-1.10]* |
| O63 | 82,080 | 77,220,080 | 1.14 | 1.05 | 1.11 | 0.10 | 0.04 | 0.03 | 1.03 [1.01-1.06] | 1.02 [1.00-1.05] |
| O64 | 16,050 | 79,496,560 | 0.21 | 0.20 | 0.20 | 0.01 | 0.01 | 0.01 | 1.03 [0.97-1.09] | 1.03 [0.97-1.09] |
| O65 | 1,150 | 80,100,630 | 0.01 | 0.01 | 0.01 | 0.00 | 0.00 | 0.00 | 1.01 [0.82-1.24] | 1.04 [0.84-1.29] |
| O66 | 15,500 | 79,643,040 | 0.21 | 0.19 | 0.20 | 0.01 | 0.01 | 0.00 | 1.03 [0.97-1.09] | 1.01 [0.96-1.07] |
| O67 | 4,050 | 80,023,680 | 0.05 | 0.05 | 0.05 | 0.00 | 0.00 | 0.00 | 1.07 [0.96-1.19] | 1.06 [0.95-1.19] |
| O68 | 170,860 | 74,525,240 | 2.63 | 2.22 | 2.44 | 0.41 | 0.18 | 0.14 | 1.08 [1.06-1.09]* | 1.06 [1.04-1.07]* |
| O69 | 15,930 | 79,359,800 | 0.22 | 0.20 | 0.20 | 0.03 | 0.02 | 0.02 | 1.10 [1.04-1.16]* | 1.10 [1.04-1.16]* |
| O70 | 227,580 | 72,189,890 | 3.65 | 3.04 | 3.44 | 0.60 | 0.21 | 0.19 | 1.06 [1.04-1.08]* | 1.06 [1.04-1.07]* |
| O71 | 13,960 | 79,736,300 | 0.19 | 0.17 | 0.18 | 0.01 | 0.01 | 0.01 | 1.05 [0.99-1.11] | 1.05 [0.98-1.11] |
| O72 | 115,960 | 77,496,740 | 1.69 | 1.45 | 1.57 | 0.24 | 0.12 | 0.11 | 1.08 [1.06-1.10]* | 1.07 [1.05-1.09]* |
| O73 | 8,140 | 79,866,150 | 0.11 | 0.10 | 0.10 | 0.01 | 0.01 | 0.01 | 1.11 [1.03-1.20]* | 1.08 [1.00-1.17] |
| O74 | 2,220 | 80,112,630 | 0.03 | 0.03 | 0.03 | 0.01 | 0.00 | 0.00 | 1.15 [1.00-1.34] | 1.10 [0.95-1.28] |
| O75 | 57,770 | 78,605,140 | 0.81 | 0.72 | 0.76 | 0.09 | 0.05 | 0.04 | 1.06 [1.03-1.09]* | 1.06 [1.03-1.09]* |
| O80 | 61,780 | 77,113,120 | 0.89 | 0.78 | 0.82 | 0.11 | 0.07 | 0.07 | 1.09 [1.06-1.12]* | 1.09 [1.06-1.12]* |
| O81 | 950 | 80,059,650 | 0.01 | 0.01 | 0.01 | 0.00 | 0.00 | 0.00 | 1.05 [0.84-1.32] | 1.04 [0.83-1.31] |
| O82 | 9,110 | 79,677,790 | 0.12 | 0.11 | 0.11 | 0.01 | 0.01 | 0.01 | 1.08 [1.00-1.16] | 1.05 [0.98-1.13] |
| O85 | 4,840 | 80,099,980 | 0.07 | 0.06 | 0.06 | 0.01 | 0.01 | 0.00 | 1.08 [0.98-1.20] | 1.02 [0.92-1.13] |
| O86 | 17,630 | 79,721,450 | 0.26 | 0.21 | 0.22 | 0.05 | 0.04 | 0.03 | 1.18 [1.12-1.24]* | 1.13 [1.08-1.20]* |
| O87 | 2,560 | 80,074,810 | 0.04 | 0.03 | 0.03 | 0.01 | 0.01 | 0.01 | 1.24 [1.09-1.42]* | 1.21 [1.06-1.39]* |
| O88 | 920 | 80,153,650 | 0.01 | 0.01 | 0.01 | 0.00 | 0.00 | 0.00 | 1.20 [0.96-1.50] | 1.10 [0.87-1.39] |
| O89 | 760 | 80,148,980 | 0.01 | 0.01 | 0.01 | 0.00 | 0.00 | 0.00 | 1.34 [1.06-1.71] | 1.30 [1.02-1.67] |
| O90 | 20,850 | 79,748,790 | 0.31 | 0.25 | 0.26 | 0.06 | 0.05 | 0.04 | 1.20 [1.14-1.25]* | 1.13 [1.08-1.19]* |
| O91 | 2,110 | 80,129,070 | 0.03 | 0.03 | 0.03 | 0.00 | 0.00 | 0.00 | 1.06 [0.91-1.23] | 1.02 [0.87-1.20] |
| O92 | 3,200 | 80,079,340 | 0.04 | 0.04 | 0.04 | 0.01 | 0.00 | 0.00 | 1.12 [0.99-1.26] | 1.07 [0.95-1.22] |
| O94 | 680 | 80,167,110 | 0.01 | 0.01 | 0.01 | 0.00 | 0.00 | 0.00 | 1.18 [0.92-1.53] | 1.04 [0.79-1.36] |
| O98 | 7,610 | 79,984,320 | 0.12 | 0.09 | 0.10 | 0.03 | 0.02 | 0.01 | 1.21 [1.12-1.31]* | 1.14 [1.05-1.23]* |
| O99 | 125,400 | 76,748,410 | 2.01 | 1.55 | 1.69 | 0.46 | 0.33 | 0.19 | 1.19 [1.17-1.22]* | 1.11 [1.08-1.13]* |
| 18+ - Q00-Q99 | | | | | | | | | | |
| Q03 | 440 | 80,154,340 | 0.01 | 0.00 | 0.01 | 0.00 | 0.00 | 0.00 | 1.58 [1.17-2.12]* | 1.47 [1.08-2.00] |
| Q04 | 850 | 80,150,240 | 0.01 | 0.01 | 0.01 | 0.00 | 0.00 | 0.00 | 1.21 [0.96-1.53] | 1.07 [0.84-1.36] |
| Q05 | 1,800 | 80,123,680 | 0.03 | 0.02 | 0.02 | 0.01 | 0.01 | 0.00 | 1.33 [1.14-1.55]* | 1.20 [1.02-1.40] |
| Q07 | 1,560 | 80,147,510 | 0.03 | 0.02 | 0.02 | 0.01 | 0.01 | 0.00 | 1.34 [1.14-1.58]* | 1.12 [0.94-1.34] |
| Q10 | 460 | 80,149,410 | 0.01 | 0.01 | 0.01 | 0.00 | 0.00 | 0.00 | 1.46 [1.08-1.97] | 1.40 [1.02-1.90] |
| Q12 | 880 | 80,150,000 | 0.01 | 0.01 | 0.01 | 0.00 | 0.00 | 0.00 | 1.26 [1.00-1.58] | 1.19 [0.94-1.51] |

| | | | | | | | | | | |
|---|---|---|---|---|---|---|---|---|---|---|
| Q18 | 1,570 | 80,111,760 | 0.02 | 0.02 | 0.02 | 0.00 | 0.00 | 0.00 | 1.10 [0.93-1.31] | 1.01 [0.84-1.20] |
| Q21 | 7,160 | 80,008,080 | 0.11 | 0.09 | 0.09 | 0.02 | 0.02 | 0.01 | 1.24 [1.14-1.34]* | 1.14 [1.06-1.24]* |
| Q23 | 3,520 | 80,117,360 | 0.05 | 0.04 | 0.04 | 0.01 | 0.01 | 0.00 | 1.17 [1.04-1.31]* | 1.10 [0.98-1.24] |
| Q24 | 2,400 | 80,119,680 | 0.04 | 0.03 | 0.03 | 0.01 | 0.01 | 0.01 | 1.36 [1.19-1.55]* | 1.23 [1.07-1.41]* |
| Q25 | 1,730 | 80,106,240 | 0.03 | 0.02 | 0.02 | 0.00 | 0.00 | 0.00 | 1.19 [1.01-1.40] | 1.11 [0.94-1.31] |
| Q27 | 1,950 | 80,136,840 | 0.03 | 0.02 | 0.02 | 0.00 | 0.00 | 0.00 | 1.20 [1.03-1.40] | 1.13 [0.96-1.32] |
| Q28 | 1,400 | 80,144,200 | 0.02 | 0.02 | 0.02 | 0.01 | 0.01 | 0.01 | 1.46 [1.23-1.74]* | 1.36 [1.14-1.63]* |
| Q38 | 3,260 | 80,062,820 | 0.05 | 0.04 | 0.04 | 0.01 | 0.01 | 0.00 | 1.16 [1.02-1.31] | 1.08 [0.95-1.22] |
| Q39 | 1,580 | 80,152,680 | 0.02 | 0.02 | 0.02 | 0.01 | 0.01 | 0.00 | 1.30 [1.09-1.53]* | 1.16 [0.98-1.38] |
| Q40 | 1,000 | 80,151,610 | 0.01 | 0.01 | 0.01 | 0.00 | 0.00 | 0.00 | 1.20 [0.97-1.48] | 1.03 [0.82-1.28] |
| Q43 | 3,370 | 80,112,740 | 0.05 | 0.04 | 0.04 | 0.01 | 0.01 | 0.00 | 1.23 [1.10-1.38]* | 1.10 [0.98-1.24] |
| Q44 | 1,200 | 80,156,550 | 0.02 | 0.01 | 0.01 | 0.00 | 0.00 | 0.00 | 1.21 [1.00-1.47] | 1.10 [0.91-1.34] |
| Q45 | 500 | 80,167,050 | 0.01 | 0.01 | 0.01 | 0.00 | 0.00 | 0.00 | 1.36 [1.01-1.83] | 1.13 [0.82-1.54] |
| Q50 | 2,780 | 80,102,130 | 0.04 | 0.03 | 0.03 | 0.01 | 0.01 | 0.01 | 1.30 [1.14-1.47]* | 1.19 [1.04-1.35] |
| Q51 | 2,900 | 80,112,880 | 0.04 | 0.04 | 0.04 | 0.00 | 0.00 | 0.00 | 1.10 [0.97-1.25] | 1.02 [0.89-1.16] |
| Q52 | 880 | 80,142,290 | 0.01 | 0.01 | 0.01 | 0.00 | 0.00 | 0.00 | 1.22 [0.97-1.52] | 1.14 [0.90-1.44] |
| Q53 | 640 | 80,025,360 | 0.01 | 0.01 | 0.01 | 0.00 | 0.00 | 0.00 | 1.12 [0.85-1.46] | 1.05 [0.79-1.38] |
| Q54 | 1,410 | 80,114,160 | 0.02 | 0.02 | 0.02 | 0.00 | 0.00 | 0.00 | 1.24 [1.03-1.49] | 1.17 [0.97-1.41] |
| Q55 | 770 | 80,091,590 | 0.01 | 0.01 | 0.01 | 0.00 | 0.00 | 0.00 | 1.22 [0.96-1.55] | 1.10 [0.86-1.42] |
| Q60 | 2,350 | 80,139,320 | 0.04 | 0.03 | 0.03 | 0.01 | 0.01 | 0.00 | 1.20 [1.05-1.38]* | 1.08 [0.94-1.25] |
| Q61 | 6,530 | 80,077,270 | 0.09 | 0.08 | 0.08 | 0.01 | 0.01 | 0.00 | 1.14 [1.05-1.24]* | 1.04 [0.95-1.14] |
| Q62 | 740 | 80,134,140 | 0.01 | 0.01 | 0.01 | 0.00 | 0.00 | 0.00 | 1.01 [0.77-1.31] | 0.98 [0.75-1.29] |
| Q63 | 4,170 | 80,101,230 | 0.06 | 0.05 | 0.05 | 0.01 | 0.01 | 0.00 | 1.17 [1.05-1.30]* | 1.05 [0.94-1.17] |
| Q64 | 530 | 80,151,070 | 0.01 | 0.01 | 0.01 | 0.00 | 0.00 | 0.00 | 1.31 [0.99-1.74] | 1.21 [0.91-1.62] |
| Q65 | 2,480 | 80,103,570 | 0.04 | 0.03 | 0.03 | 0.01 | 0.01 | 0.00 | 1.17 [1.02-1.34] | 1.07 [0.93-1.23] |
| Q66 | 2,290 | 80,040,100 | 0.04 | 0.03 | 0.03 | 0.01 | 0.01 | 0.01 | 1.38 [1.20-1.57]* | 1.25 [1.09-1.44]* |
| Q67 | 1,390 | 80,133,010 | 0.02 | 0.02 | 0.02 | 0.01 | 0.01 | 0.01 | 1.41 [1.19-1.68]* | 1.30 [1.09-1.56]* |
| Q74 | 1,430 | 80,129,100 | 0.02 | 0.02 | 0.02 | 0.01 | 0.00 | 0.00 | 1.28 [1.08-1.53]* | 1.16 [0.97-1.40] |
| Q76 | 2,040 | 80,128,960 | 0.03 | 0.02 | 0.03 | 0.01 | 0.01 | 0.00 | 1.23 [1.06-1.42]* | 1.05 [0.90-1.22] |
| Q78 | 1,040 | 80,150,320 | 0.02 | 0.01 | 0.01 | 0.00 | 0.00 | 0.00 | 1.31 [1.07-1.61] | 1.17 [0.95-1.45] |
| Q79 | 2,710 | 80,131,760 | 0.04 | 0.03 | 0.03 | 0.01 | 0.01 | 0.00 | 1.34 [1.18-1.52]* | 1.01 [0.88-1.16] |
| Q82 | 1,350 | 80,094,200 | 0.03 | 0.02 | 0.01 | 0.01 | 0.01 | 0.01 | 1.75 [1.47-2.07]* | 1.56 [1.31-1.86]* |
| Q83 | 1,400 | 80,111,110 | 0.02 | 0.02 | 0.02 | 0.00 | 0.00 | 0.00 | 1.15 [0.96-1.38] | 1.05 [0.87-1.26] |
| Q85 | 1,680 | 80,132,350 | 0.02 | 0.02 | 0.02 | 0.00 | 0.00 | 0.00 | 1.05 [0.89-1.24] | 0.95 [0.80-1.14] |
| Q87 | 1,700 | 80,116,430 | 0.03 | 0.02 | 0.02 | 0.01 | 0.01 | 0.01 | 1.44 [1.24-1.69]* | 1.30 [1.10-1.52]* |
| Q89 | 1,500 | 80,127,150 | 0.02 | 0.02 | 0.02 | 0.00 | 0.00 | 0.00 | 1.20 [1.01-1.43] | 1.09 [0.91-1.30] |
| Q90 | 1,640 | 80,087,070 | 0.05 | 0.01 | 0.01 | 0.04 | 0.04 | 0.04 | 3.76 [3.28-4.32]* | 3.86 [3.35-4.45]* |
| Q99 | 680 | 80,155,540 | 0.01 | 0.01 | 0.01 | 0.00 | 0.00 | 0.00 | 1.28 [1.00-1.65] | 1.14 [0.88-1.47] |
| 18+ - R00-R99 | | | | | | | | | | |
| R00 | 192,580 | 78,229,360 | 2.95 | 2.36 | 2.42 | 0.59 | 0.53 | 0.29 | 1.22 [1.20-1.24]* | 1.11 [1.09-1.13]* |
| R01 | 25,540 | 79,871,390 | 0.38 | 0.31 | 0.31 | 0.08 | 0.07 | 0.04 | 1.22 [1.17-1.27]* | 1.12 [1.07-1.17]* |
| R02 | 29,260 | 79,998,790 | 0.44 | 0.35 | 0.36 | 0.09 | 0.08 | 0.05 | 1.23 [1.18-1.28]* | 1.13 [1.08-1.18]* |
| R03 | 69,240 | 79,463,910 | 1.03 | 0.84 | 0.86 | 0.19 | 0.16 | 0.11 | 1.19 [1.16-1.22]* | 1.12 [1.09-1.15]* |
| R04 | 70,990 | 79,136,520 | 1.14 | 0.84 | 0.85 | 0.29 | 0.29 | 0.20 | 1.35 [1.31-1.38]* | 1.22 [1.18-1.25]* |
| R05 | 62,990 | 79,430,140 | 1.03 | 0.74 | 0.76 | 0.28 | 0.27 | 0.16 | 1.35 [1.32-1.39]* | 1.19 [1.15-1.22]* |
| R06 | 180,450 | 77,612,430 | 2.97 | 2.19 | 2.22 | 0.78 | 0.75 | 0.45 | 1.34 [1.32-1.36]* | 1.18 [1.16-1.20]* |
| R07 | 330,860 | 73,910,210 | 5.59 | 4.23 | 4.29 | 1.36 | 1.30 | 0.76 | 1.30 [1.29-1.32]* | 1.16 [1.14-1.17]* |
| R09 | 28,530 | 79,896,050 | 0.45 | 0.34 | 0.34 | 0.11 | 0.11 | 0.07 | 1.32 [1.27-1.37]* | 1.17 [1.12-1.22]* |
| R10 | 412,880 | 69,634,660 | 7.38 | 5.60 | 5.76 | 1.78 | 1.62 | 0.91 | 1.28 [1.27-1.29]* | 1.14 [1.13-1.15]* |
| R11 | 227,170 | 76,777,900 | 3.60 | 2.82 | 2.87 | 0.78 | 0.73 | 0.39 | 1.25 [1.23-1.27]* | 1.12 [1.10-1.14]* |
| R12 | 17,770 | 79,855,150 | 0.29 | 0.21 | 0.21 | 0.08 | 0.07 | 0.04 | 1.35 [1.28-1.42]* | 1.18 [1.12-1.24]* |
| R13 | 130,210 | 79,048,840 | 2.09 | 1.55 | 1.59 | 0.54 | 0.50 | 0.32 | 1.32 [1.29-1.34]* | 1.18 [1.16-1.21]* |
| R14 | 29,400 | 79,834,070 | 0.47 | 0.35 | 0.35 | 0.12 | 0.11 | 0.06 | 1.32 [1.27-1.37]* | 1.15 [1.11-1.20]* |
| R15 | 55,150 | 79,891,400 | 0.83 | 0.66 | 0.69 | 0.17 | 0.14 | 0.09 | 1.20 [1.17-1.24]* | 1.13 [1.09-1.16]* |
| R16 | 19,700 | 80,000,540 | 0.33 | 0.23 | 0.23 | 0.10 | 0.10 | 0.07 | 1.44 [1.38-1.51]* | 1.28 [1.22-1.35]* |
| R17 | 17,050 | 79,988,040 | 0.25 | 0.20 | 0.20 | 0.05 | 0.05 | 0.04 | 1.24 [1.18-1.31]* | 1.18 [1.12-1.24]* |
| R18 | 44,410 | 79,927,710 | 0.65 | 0.53 | 0.54 | 0.12 | 0.12 | 0.08 | 1.22 [1.18-1.26]* | 1.15 [1.11-1.19]* |
| R19 | 166,250 | 78,069,200 | 2.74 | 2.00 | 2.02 | 0.74 | 0.71 | 0.47 | 1.35 [1.33-1.37]* | 1.21 [1.19-1.23]* |
| R20 | 45,150 | 79,515,400 | 0.70 | 0.54 | 0.55 | 0.16 | 0.15 | 0.07 | 1.27 [1.23-1.31]* | 1.11 [1.07-1.14]* |
| R21 | 43,940 | 79,405,940 | 0.84 | 0.49 | 0.50 | 0.35 | 0.35 | 0.30 | 1.70 [1.65-1.75]* | 1.55 [1.50-1.60]* |
| R22 | 30,100 | 79,631,430 | 0.49 | 0.35 | 0.36 | 0.13 | 0.13 | 0.09 | 1.37 [1.31-1.42]* | 1.22 [1.17-1.27]* |
| R23 | 16,820 | 79,956,570 | 0.27 | 0.20 | 0.20 | 0.07 | 0.07 | 0.05 | 1.33 [1.26-1.40]* | 1.21 [1.15-1.28]* |
| R25 | 23,960 | 79,919,330 | 0.37 | 0.28 | 0.29 | 0.09 | 0.08 | 0.05 | 1.28 [1.23-1.34]* | 1.14 [1.09-1.19]* |
| R26 | 199,480 | 79,388,970 | 3.06 | 2.40 | 2.54 | 0.66 | 0.52 | 0.33 | 1.20 [1.19-1.22]* | 1.12 [1.10-1.14]* |
| R27 | 6,420 | 80,094,190 | 0.09 | 0.08 | 0.08 | 0.02 | 0.01 | 0.01 | 1.18 [1.08-1.29]* | 1.07 [0.98-1.16] |
| R29 | 269,000 | 79,132,700 | 4.09 | 3.25 | 3.43 | 0.84 | 0.66 | 0.39 | 1.19 [1.18-1.21]* | 1.10 [1.09-1.12]* |
| R30 | 19,180 | 79,829,070 | 0.31 | 0.23 | 0.23 | 0.08 | 0.08 | 0.04 | 1.34 [1.28-1.41]* | 1.17 [1.11-1.23]* |
| R31 | 143,650 | 77,624,320 | 2.16 | 1.78 | 1.81 | 0.37 | 0.35 | 0.23 | 1.19 [1.17-1.21]* | 1.12 [1.10-1.14]* |
| R32 | 104,820 | 79,501,710 | 1.57 | 1.27 | 1.32 | 0.30 | 0.24 | 0.15 | 1.18 [1.16-1.21]* | 1.10 [1.08-1.13]* |
| R33 | 203,160 | 78,648,870 | 3.03 | 2.49 | 2.61 | 0.54 | 0.42 | 0.27 | 1.16 [1.14-1.18]* | 1.10 [1.08-1.11]* |
| R34 | 5,050 | 80,146,660 | 0.08 | 0.06 | 0.06 | 0.02 | 0.02 | 0.01 | 1.30 [1.18-1.43]* | 1.17 [1.06-1.30]* |
| R35 | 32,500 | 79,438,270 | 0.52 | 0.39 | 0.39 | 0.13 | 0.13 | 0.09 | 1.33 [1.29-1.38]* | 1.20 [1.15-1.25]* |
| R36 | 680 | 80,162,720 | 0.01 | 0.01 | 0.01 | 0.00 | 0.00 | 0.00 | 1.35 [1.04-1.75] | 1.23 [0.94-1.61] |
| R39 | 37,920 | 79,427,170 | 0.61 | 0.45 | 0.45 | 0.16 | 0.16 | 0.10 | 1.35 [1.31-1.40]* | 1.21 [1.16-1.25]* |
| R40 | 29,980 | 79,948,600 | 0.46 | 0.36 | 0.37 | 0.10 | 0.08 | 0.05 | 1.23 [1.18-1.28]* | 1.12 [1.07-1.16]* |
| R41 | 194,300 | 79,182,880 | 2.90 | 2.36 | 2.46 | 0.54 | 0.43 | 0.29 | 1.18 [1.16-1.20]* | 1.11 [1.09-1.13]* |
| R42 | 106,100 | 78,959,630 | 1.63 | 1.28 | 1.31 | 0.35 | 0.32 | 0.17 | 1.25 [1.22-1.27]* | 1.12 [1.09-1.14]* |
| R43 | 1,460 | 80,159,530 | 0.02 | 0.02 | 0.02 | 0.01 | 0.00 | 0.00 | 1.28 [1.07-1.52]* | 1.12 [0.93-1.34] |
| R44 | 26,350 | 79,989,560 | 0.40 | 0.31 | 0.32 | 0.08 | 0.07 | 0.04 | 1.22 [1.17-1.28]* | 1.11 [1.07-1.17]* |
| R45 | 55,460 | 79,792,570 | 0.82 | 0.67 | 0.69 | 0.15 | 0.13 | 0.05 | 1.19 [1.15-1.22]* | 1.06 [1.03-1.09]* |
| R46 | 16,950 | 80,079,460 | 0.24 | 0.21 | 0.21 | 0.03 | 0.03 | 0.02 | 1.13 [1.07-1.19]* | 1.07 [1.01-1.13] |
| R47 | 53,720 | 79,774,510 | 0.79 | 0.65 | 0.67 | 0.14 | 0.11 | 0.06 | 1.17 [1.13-1.20]* | 1.08 [1.05-1.12]* |
| R48 | 1,560 | 80,156,470 | 0.02 | 0.02 | 0.02 | 0.01 | 0.01 | 0.00 | 1.32 [1.11-1.56]* | 1.10 [0.92-1.32] |
| R49 | 10,120 | 79,975,150 | 0.16 | 0.12 | 0.12 | 0.04 | 0.04 | 0.02 | 1.35 [1.27-1.44]* | 1.17 [1.10-1.26]* |
| R50 | 75,200 | 79,048,190 | 1.17 | 0.90 | 0.92 | 0.26 | 0.25 | 0.17 | 1.27 [1.24-1.30]* | 1.17 [1.14-1.20]* |
| R51 | 137,650 | 77,590,440 | 2.22 | 1.68 | 1.72 | 0.55 | 0.50 | 0.26 | 1.29 [1.27-1.32]* | 1.13 [1.11-1.15]* |
| R52 | 24,030 | 79,907,530 | 0.40 | 0.28 | 0.28 | 0.12 | 0.12 | 0.07 | 1.42 [1.36-1.48]* | 1.20 [1.15-1.25]* |
| R53 | 96,380 | 79,444,960 | 1.48 | 1.16 | 1.20 | 0.32 | 0.28 | 0.16 | 1.23 [1.20-1.26]* | 1.12 [1.10-1.15]* |
| R54 | 127,790 | 79,721,980 | 1.89 | 1.54 | 1.62 | 0.35 | 0.28 | 0.19 | 1.17 [1.15-1.19]* | 1.11 [1.09-1.14]* |
| R55 | 191,080 | 77,903,180 | 2.90 | 2.36 | 2.42 | 0.55 | 0.49 | 0.31 | 1.20 [1.18-1.22]* | 1.12 [1.10-1.14]* |
| R56 | 53,580 | 79,207,090 | 0.82 | 0.65 | 0.65 | 0.17 | 0.17 | 0.11 | 1.26 [1.22-1.29]* | 1.15 [1.12-1.19]* |
| R57 | 16,660 | 80,130,760 | 0.24 | 0.20 | 0.20 | 0.04 | 0.04 | 0.03 | 1.20 [1.13-1.26]* | 1.12 [1.06-1.18]* |
| R58 | 2,550 | 80,152,890 | 0.04 | 0.03 | 0.03 | 0.01 | 0.01 | 0.01 | 1.34 [1.17-1.52]* | 1.22 [1.07-1.40]* |
| R59 | 48,530 | 79,608,580 | 0.78 | 0.57 | 0.57 | 0.21 | 0.21 | 0.17 | 1.36 [1.32-1.40]* | 1.27 [1.23-1.31]* |
| R60 | 91,280 | 79,680,660 | 1.46 | 1.08 | 1.10 | 0.38 | 0.36 | 0.26 | 1.33 [1.30-1.36]* | 1.22 [1.19-1.24]* |
| R61 | 9,730 | 80,020,340 | 0.16 | 0.11 | 0.11 | 0.04 | 0.04 | 0.03 | 1.38 [1.29-1.48]* | 1.21 [1.13-1.30]* |
| R62 | 2,060 | 80,025,770 | 0.03 | 0.03 | 0.03 | 0.00 | 0.00 | 0.00 | 1.09 [0.93-1.27] | 1.00 [0.85-1.17] |
| R63 | 181,590 | 78,602,310 | 2.82 | 2.20 | 2.26 | 0.62 | 0.56 | 0.35 | 1.25 [1.23-1.27]* | 1.14 [1.12-1.16]* |
| R64 | 5,210 | 80,163,000 | 0.07 | 0.06 | 0.07 | 0.01 | 0.01 | 0.01 | 1.12 [1.01-1.24] | 1.09 [0.99-1.21] |
| R65 | 11,770 | 80,144,340 | 0.18 | 0.14 | 0.15 | 0.04 | 0.03 | 0.02 | 1.22 [1.14-1.30]* | 1.12 [1.05-1.20]* |
| R68 | 17,480 | 80,038,470 | 0.26 | 0.21 | 0.21 | 0.05 | 0.05 | 0.04 | 1.24 [1.18-1.31]* | 1.18 [1.12-1.25]* |
| R69 | 157,270 | 75,394,290 | 2.49 | 2.00 | 2.00 | 0.49 | 0.49 | 0.33 | 1.24 [1.22-1.27]* | 1.15 [1.13-1.17]* |
| R70 | 1,300 | 80,144,060 | 0.02 | 0.01 | 0.02 | 0.01 | 0.01 | 0.01 | 1.46 [1.22-1.75]* | 1.35 [1.13-1.62]* |
| R71 | 1,120 | 80,162,110 | 0.02 | 0.01 | 0.01 | 0.00 | 0.00 | 0.00 | 1.11 [0.90-1.36] | 1.05 [0.85-1.30] |
| R72 | 6,120 | 80,117,630 | 0.09 | 0.07 | 0.08 | 0.02 | 0.01 | 0.01 | 1.20 [1.10-1.31]* | 1.10 [1.01-1.21] |
| R73 | 22,750 | 79,954,770 | 0.36 | 0.27 | 0.27 | 0.09 | 0.09 | 0.05 | 1.32 [1.26-1.38]* | 1.17 [1.12-1.23]* |

| | | | | | | | | | | |
|---|---|---|---|---|---|---|---|---|---|---|
| R74 | 10,150 | 80,091,930 | 0.15 | 0.12 | 0.13 | 0.03 | 0.02 | 0.01 | 1.17 [1.09-1.26]* | 1.06 [0.98-1.14] |
| R76 | 2,640 | 80,133,350 | 0.04 | 0.03 | 0.03 | 0.01 | 0.01 | 0.01 | 1.37 [1.21-1.56]* | 1.26 [1.11-1.44]* |
| R77 | 6,380 | 80,138,030 | 0.10 | 0.08 | 0.08 | 0.02 | 0.02 | 0.01 | 1.29 [1.18-1.40]* | 1.16 [1.07-1.27]* |
| R78 | 850 | 80,158,980 | 0.01 | 0.01 | 0.01 | 0.00 | 0.00 | 0.00 | 1.16 [0.91-1.46] | 1.04 [0.81-1.32] |
| R79 | 126,160 | 79,188,200 | 1.93 | 1.52 | 1.56 | 0.41 | 0.37 | 0.23 | 1.24 [1.21-1.26]* | 1.14 [1.11-1.16]* |
| R80 | 4,430 | 80,085,640 | 0.07 | 0.05 | 0.05 | 0.02 | 0.02 | 0.01 | 1.38 [1.25-1.53]* | 1.26 [1.14-1.40]* |
| R81 | 1,360 | 80,082,290 | 0.02 | 0.02 | 0.02 | 0.00 | 0.00 | 0.00 | 1.15 [0.96-1.38] | 1.09 [0.90-1.31] |
| R82 | 9,840 | 80,060,100 | 0.15 | 0.12 | 0.12 | 0.03 | 0.03 | 0.01 | 1.21 [1.13-1.30]* | 1.10 [1.03-1.18]* |
| R84 | 2,850 | 80,155,190 | 0.05 | 0.03 | 0.03 | 0.01 | 0.01 | 0.01 | 1.36 [1.20-1.54]* | 1.15 [1.01-1.31] |
| R85 | 1,790 | 80,125,620 | 0.03 | 0.02 | 0.02 | 0.01 | 0.01 | 0.01 | 1.39 [1.19-1.62]* | 1.25 [1.06-1.46]* |
| R87 | 9,800 | 79,625,700 | 0.14 | 0.12 | 0.12 | 0.02 | 0.02 | 0.01 | 1.17 [1.09-1.25]* | 1.11 [1.03-1.19]* |
| R89 | 1,580 | 80,155,780 | 0.03 | 0.02 | 0.02 | 0.01 | 0.01 | 0.01 | 1.44 [1.22-1.70]* | 1.31 [1.11-1.55]* |
| R90 | 5,560 | 80,123,330 | 0.08 | 0.07 | 0.07 | 0.01 | 0.01 | 0.01 | 1.19 [1.08-1.30]* | 1.10 [1.00-1.21] |
| R91 | 62,470 | 79,740,830 | 0.95 | 0.75 | 0.76 | 0.21 | 0.19 | 0.12 | 1.25 [1.22-1.29]* | 1.15 [1.12-1.18]* |
| R92 | 570 | 80,165,850 | 0.01 | 0.01 | 0.01 | 0.00 | 0.00 | 0.00 | 1.09 [0.81-1.46] | 0.97 [0.72-1.32] |
| R93 | 63,010 | 79,633,310 | 0.93 | 0.76 | 0.77 | 0.17 | 0.16 | 0.10 | 1.21 [1.18-1.25]* | 1.12 [1.09-1.15]* |
| R94 | 111,220 | 79,247,940 | 1.72 | 1.34 | 1.36 | 0.38 | 0.36 | 0.25 | 1.26 [1.24-1.29]* | 1.17 [1.14-1.19]* |
| 18+ - S00-T88 | | | | | | | | | | |
| S00 | 101,420 | 79,211,750 | 1.52 | 1.23 | 1.28 | 0.29 | 0.24 | 0.15 | 1.19 [1.16-1.21]* | 1.11 [1.08-1.13]* |
| S01 | 102,230 | 78,962,470 | 1.52 | 1.25 | 1.29 | 0.28 | 0.24 | 0.16 | 1.18 [1.16-1.21]* | 1.12 [1.09-1.14]* |
| S02 | 39,290 | 79,240,280 | 0.56 | 0.48 | 0.49 | 0.08 | 0.08 | 0.05 | 1.16 [1.12-1.20]* | 1.09 [1.05-1.13]* |
| S03 | 970 | 80,152,920 | 0.01 | 0.01 | 0.01 | 0.00 | 0.00 | 0.00 | 1.14 [0.91-1.41] | 1.05 [0.84-1.31] |
| S05 | 9,650 | 80,025,460 | 0.14 | 0.12 | 0.12 | 0.02 | 0.02 | 0.01 | 1.15 [1.07-1.23]* | 1.08 [1.01-1.16] |
| S06 | 32,240 | 79,818,420 | 0.46 | 0.39 | 0.40 | 0.06 | 0.05 | 0.03 | 1.13 [1.08-1.17]* | 1.07 [1.03-1.11]* |
| S09 | 48,480 | 79,139,860 | 0.73 | 0.59 | 0.60 | 0.14 | 0.13 | 0.07 | 1.21 [1.18-1.25]* | 1.11 [1.07-1.15]* |
| S10 | 2,900 | 80,122,220 | 0.04 | 0.04 | 0.04 | 0.00 | 0.00 | 0.00 | 1.13 [0.99-1.29] | 1.02 [0.89-1.17] |
| S11 | 1,720 | 80,143,230 | 0.03 | 0.02 | 0.02 | 0.00 | 0.00 | 0.00 | 1.19 [1.01-1.40] | 1.09 [0.92-1.29] |
| S12 | 8,500 | 80,096,430 | 0.12 | 0.10 | 0.11 | 0.02 | 0.02 | 0.01 | 1.18 [1.09-1.27]* | 1.11 [1.02-1.20] |
| S13 | 1,930 | 80,121,470 | 0.03 | 0.02 | 0.02 | 0.00 | 0.00 | 0.00 | 1.20 [1.03-1.40] | 1.09 [0.92-1.27] |
| S14 | 1,600 | 80,149,460 | 0.02 | 0.02 | 0.02 | 0.00 | 0.00 | 0.00 | 1.07 [0.90-1.28] | 1.04 [0.87-1.24] |
| S19 | 2,380 | 80,113,760 | 0.04 | 0.03 | 0.03 | 0.01 | 0.01 | 0.00 | 1.30 [1.13-1.49]* | 1.14 [0.99-1.32] |
| S20 | 11,700 | 80,051,260 | 0.18 | 0.14 | 0.15 | 0.04 | 0.03 | 0.02 | 1.22 [1.15-1.31]* | 1.11 [1.03-1.18]* |
| S21 | 2,280 | 80,131,810 | 0.03 | 0.03 | 0.03 | 0.01 | 0.00 | 0.00 | 1.15 [1.00-1.33] | 1.05 [0.90-1.22] |
| S22 | 41,950 | 79,762,840 | 0.62 | 0.50 | 0.52 | 0.12 | 0.11 | 0.06 | 1.21 [1.17-1.25]* | 1.11 [1.07-1.15]* |
| S27 | 12,860 | 80,013,670 | 0.18 | 0.16 | 0.16 | 0.02 | 0.02 | 0.01 | 1.10 [1.04-1.18]* | 1.04 [0.98-1.11] |
| S29 | 3,190 | 80,133,270 | 0.05 | 0.04 | 0.04 | 0.02 | 0.02 | 0.01 | 1.44 [1.28-1.62]* | 1.28 [1.13-1.44]* |
| S30 | 20,230 | 79,934,440 | 0.30 | 0.24 | 0.25 | 0.06 | 0.05 | 0.03 | 1.22 [1.16-1.28]* | 1.10 [1.04-1.15]* |
| S31 | 5,020 | 80,057,000 | 0.07 | 0.06 | 0.06 | 0.01 | 0.01 | 0.00 | 1.13 [1.02-1.25] | 1.02 [0.93-1.13] |
| S32 | 45,970 | 79,780,790 | 0.66 | 0.56 | 0.58 | 0.10 | 0.08 | 0.05 | 1.14 [1.10-1.17]* | 1.07 [1.04-1.11]* |
| S33 | 590 | 80,162,150 | 0.01 | 0.01 | 0.01 | 0.00 | 0.00 | 0.00 | 1.00 [0.75-1.35] | 0.92 [0.68-1.25] |
| S35 | 550 | 80,166,830 | 0.01 | 0.01 | 0.01 | 0.00 | 0.00 | 0.00 | 1.17 [0.87-1.56] | 1.17 [0.87-1.59] |
| S36 | 4,760 | 80,095,200 | 0.07 | 0.06 | 0.06 | 0.01 | 0.01 | 0.00 | 1.12 [1.01-1.24] | 1.06 [0.96-1.18] |
| S37 | 3,740 | 80,124,410 | 0.05 | 0.05 | 0.05 | 0.00 | 0.00 | 0.00 | 1.05 [0.93-1.18] | 1.01 [0.89-1.14] |
| S39 | 8,050 | 80,021,160 | 0.12 | 0.10 | 0.10 | 0.02 | 0.02 | 0.01 | 1.18 [1.09-1.27]* | 1.06 [0.98-1.15] |
| S40 | 17,010 | 80,045,050 | 0.25 | 0.20 | 0.21 | 0.05 | 0.04 | 0.02 | 1.17 [1.10-1.23]* | 1.07 [1.01-1.13] |
| S41 | 6,640 | 80,102,290 | 0.10 | 0.08 | 0.08 | 0.02 | 0.02 | 0.01 | 1.26 [1.16-1.38]* | 1.14 [1.04-1.25]* |
| S42 | 50,890 | 79,453,520 | 0.72 | 0.62 | 0.64 | 0.10 | 0.09 | 0.05 | 1.14 [1.10-1.17]* | 1.08 [1.04-1.11]* |
| S43 | 9,460 | 80,042,740 | 0.13 | 0.12 | 0.12 | 0.02 | 0.01 | 0.01 | 1.11 [1.03-1.19]* | 1.05 [0.98-1.13] |
| S46 | 3,860 | 80,131,520 | 0.05 | 0.05 | 0.05 | 0.01 | 0.01 | 0.00 | 1.12 [1.00-1.25] | 1.04 [0.92-1.16] |
| S49 | 5,240 | 80,120,950 | 0.08 | 0.06 | 0.06 | 0.02 | 0.02 | 0.01 | 1.28 [1.16-1.40]* | 1.15 [1.04-1.27]* |
| S50 | 17,920 | 80,036,180 | 0.27 | 0.21 | 0.22 | 0.05 | 0.04 | 0.02 | 1.19 [1.13-1.26]* | 1.10 [1.04-1.16]* |
| S51 | 26,820 | 79,918,860 | 0.40 | 0.32 | 0.33 | 0.08 | 0.07 | 0.03 | 1.21 [1.16-1.26]* | 1.09 [1.04-1.14]* |
| S52 | 64,300 | 78,494,210 | 0.89 | 0.80 | 0.81 | 0.09 | 0.08 | 0.05 | 1.10 [1.07-1.13]* | 1.06 [1.03-1.09]* |
| S53 | 1,660 | 80,123,130 | 0.02 | 0.02 | 0.02 | 0.00 | 0.00 | 0.00 | 1.02 [0.86-1.21] | 0.95 [0.79-1.13] |
| S54 | 1,060 | 80,142,700 | 0.02 | 0.01 | 0.01 | 0.00 | 0.00 | 0.00 | 1.23 [1.00-1.51] | 1.09 [0.88-1.36] |
| S55 | 520 | 80,161,200 | 0.01 | 0.01 | 0.01 | 0.00 | 0.00 | 0.00 | 1.47 [1.11-1.95]* | 1.37 [1.03-1.84] |
| S56 | 3,890 | 80,085,310 | 0.05 | 0.05 | 0.05 | 0.00 | 0.00 | 0.00 | 1.09 [0.97-1.22] | 1.04 [0.92-1.16] |
| S59 | 1,990 | 80,149,950 | 0.03 | 0.02 | 0.02 | 0.01 | 0.01 | 0.01 | 1.31 [1.12-1.53]* | 1.19 [1.02-1.40] |
| S60 | 13,840 | 80,030,760 | 0.20 | 0.17 | 0.17 | 0.04 | 0.03 | 0.02 | 1.17 [1.10-1.25]* | 1.09 [1.02-1.15]* |
| S61 | 52,580 | 79,131,000 | 0.72 | 0.65 | 0.66 | 0.07 | 0.06 | 0.02 | 1.09 [1.06-1.13]* | 1.03 [1.00-1.06] |
| S62 | 37,610 | 79,371,480 | 0.51 | 0.47 | 0.47 | 0.04 | 0.04 | 0.01 | 1.08 [1.04-1.12]* | 1.02 [0.98-1.06] |
| S63 | 6,610 | 80,044,760 | 0.10 | 0.08 | 0.08 | 0.02 | 0.02 | 0.01 | 1.20 [1.10-1.31]* | 1.11 [1.02-1.21] |
| S64 | 8,200 | 79,957,580 | 0.10 | 0.10 | 0.10 | 0.00 | 0.00 | 0.00 | 1.01 [0.94-1.09] | 0.96 [0.89-1.04] |
| S65 | 2,260 | 80,119,240 | 0.03 | 0.03 | 0.03 | 0.00 | 0.00 | 0.00 | 1.03 [0.88-1.19] | 0.99 [0.85-1.15] |
| S66 | 15,580 | 79,764,590 | 0.20 | 0.19 | 0.19 | 0.01 | 0.01 | 0.00 | 1.04 [0.99-1.10] | 1.00 [0.94-1.05] |
| S67 | 1,250 | 80,122,960 | 0.01 | 0.02 | 0.02 | 0.00 | 0.00 | 0.00 | 0.78 [0.63-0.97] | 0.79 [0.63-0.98] |
| S68 | 3,810 | 80,066,070 | 0.05 | 0.05 | 0.05 | 0.00 | 0.00 | 0.00 | 0.97 [0.86-1.09] | 0.97 [0.86-1.09] |
| S69 | 5,200 | 80,093,320 | 0.08 | 0.06 | 0.06 | 0.02 | 0.02 | 0.01 | 1.26 [1.14-1.39]* | 1.13 [1.02-1.25] |
| S70 | 16,830 | 80,047,640 | 0.25 | 0.20 | 0.21 | 0.04 | 0.03 | 0.02 | 1.16 [1.10-1.23]* | 1.07 [1.01-1.13] |
| S71 | 3,120 | 80,109,680 | 0.04 | 0.04 | 0.04 | 0.01 | 0.01 | 0.00 | 1.14 [1.01-1.29] | 1.02 [0.89-1.16] |
| S72 | 123,580 | 79,193,870 | 1.78 | 1.51 | 1.59 | 0.26 | 0.18 | 0.14 | 1.11 [1.09-1.14]* | 1.08 [1.06-1.11]* |
| S73 | 1,510 | 80,148,320 | 0.02 | 0.02 | 0.02 | 0.00 | 0.00 | 0.00 | 1.09 [0.91-1.31] | 1.02 [0.85-1.23] |
| S76 | 3,580 | 80,124,650 | 0.05 | 0.04 | 0.04 | 0.01 | 0.01 | 0.00 | 1.16 [1.03-1.30] | 1.08 [0.96-1.22] |
| S79 | 9,020 | 80,101,460 | 0.14 | 0.11 | 0.11 | 0.03 | 0.03 | 0.02 | 1.23 [1.14-1.32]* | 1.14 [1.06-1.23]* |
| S80 | 32,330 | 79,906,560 | 0.48 | 0.39 | 0.41 | 0.09 | 0.07 | 0.03 | 1.18 [1.13-1.22]* | 1.07 [1.03-1.12]* |
| S81 | 24,430 | 79,915,160 | 0.37 | 0.29 | 0.30 | 0.08 | 0.07 | 0.04 | 1.24 [1.18-1.29]* | 1.11 [1.06-1.16]* |
| S82 | 65,090 | 78,708,760 | 0.90 | 0.81 | 0.81 | 0.08 | 0.08 | 0.03 | 1.10 [1.07-1.13]* | 1.04 [1.01-1.06] |
| S83 | 8,820 | 79,959,810 | 0.12 | 0.11 | 0.11 | 0.02 | 0.02 | 0.01 | 1.17 [1.08-1.25]* | 1.07 [1.00-1.16] |
| S86 | 4,840 | 80,047,500 | 0.07 | 0.06 | 0.06 | 0.01 | 0.01 | 0.01 | 1.18 [1.07-1.30]* | 1.10 [1.00-1.22] |
| S89 | 6,780 | 80,089,780 | 0.10 | 0.08 | 0.08 | 0.02 | 0.02 | 0.01 | 1.24 [1.14-1.34]* | 1.12 [1.03-1.22]* |
| S90 | 7,900 | 80,090,010 | 0.11 | 0.10 | 0.10 | 0.01 | 0.01 | 0.00 | 1.13 [1.04-1.22]* | 1.02 [0.94-1.10] |
| S91 | 6,780 | 80,044,570 | 0.10 | 0.08 | 0.08 | 0.02 | 0.02 | 0.01 | 1.23 [1.13-1.34]* | 1.10 [1.01-1.20] |
| S92 | 14,330 | 79,906,560 | 0.21 | 0.17 | 0.18 | 0.03 | 0.03 | 0.01 | 1.15 [1.09-1.22]* | 1.05 [0.99-1.12] |
| S93 | 4,780 | 80,096,520 | 0.07 | 0.06 | 0.06 | 0.01 | 0.01 | 0.00 | 1.13 [1.02-1.25] | 1.01 [0.91-1.12] |
| S96 | 960 | 80,151,660 | 0.01 | 0.01 | 0.01 | 0.00 | 0.00 | 0.00 | 1.11 [0.89-1.38] | 0.95 [0.75-1.20] |
| S99 | 3,340 | 80,128,550 | 0.05 | 0.04 | 0.04 | 0.01 | 0.01 | 0.00 | 1.19 [1.05-1.34]* | 1.02 [0.90-1.16] |
| T00 | 7,150 | 80,116,560 | 0.10 | 0.09 | 0.09 | 0.02 | 0.01 | 0.00 | 1.15 [1.06-1.26]* | 1.05 [0.96-1.15] |
| T01 | 3,530 | 80,140,620 | 0.05 | 0.04 | 0.04 | 0.01 | 0.01 | 0.00 | 1.14 [1.01-1.29] | 0.99 [0.87-1.12] |
| T02 | 2,280 | 80,139,930 | 0.03 | 0.03 | 0.03 | 0.01 | 0.00 | 0.00 | 1.18 [1.02-1.36] | 1.13 [0.98-1.31] |
| T09 | 1,230 | 80,153,370 | 0.02 | 0.01 | 0.01 | 0.00 | 0.00 | 0.00 | 1.20 [0.99-1.47] | 1.11 [0.90-1.36] |
| T11 | 4,870 | 80,128,330 | 0.07 | 0.06 | 0.06 | 0.02 | 0.01 | 0.01 | 1.22 [1.10-1.35]* | 1.09 [0.98-1.21] |
| T13 | 3,110 | 80,137,090 | 0.05 | 0.04 | 0.04 | 0.01 | 0.01 | 0.00 | 1.24 [1.09-1.40]* | 1.12 [0.98-1.27] |
| T14 | 3,010 | 80,080,450 | 0.04 | 0.04 | 0.04 | 0.01 | 0.01 | 0.00 | 1.19 [1.05-1.35]* | 1.07 [0.94-1.21] |
| T16 | 780 | 80,109,200 | 0.01 | 0.01 | 0.01 | 0.00 | 0.00 | 0.00 | 1.14 [0.89-1.47] | 1.01 [0.78-1.31] |
| T17 | 6,670 | 80,059,520 | 0.11 | 0.08 | 0.08 | 0.03 | 0.03 | 0.02 | 1.36 [1.25-1.48]* | 1.22 [1.12-1.33]* |
| T18 | 6,520 | 80,042,250 | 0.10 | 0.08 | 0.08 | 0.02 | 0.02 | 0.00 | 1.31 [1.21-1.42]* | 1.18 [1.08-1.28]* |
| T19 | 780 | 80,155,890 | 0.01 | 0.01 | 0.01 | 0.00 | 0.00 | 0.00 | 1.29 [1.01-1.64] | 1.18 [0.92-1.51] |
| T20 | 2,110 | 80,119,760 | 0.03 | 0.03 | 0.03 | 0.00 | 0.00 | 0.00 | 1.11 [0.96-1.29] | 1.06 [0.91-1.23] |
| T21 | 3,060 | 80,112,730 | 0.04 | 0.04 | 0.04 | 0.01 | 0.01 | 0.00 | 1.14 [1.00-1.29] | 1.07 [0.94-1.22] |
| T22 | 2,680 | 80,115,030 | 0.04 | 0.03 | 0.03 | 0.00 | 0.00 | 0.00 | 1.09 [0.95-1.24] | 1.01 [0.88-1.16] |
| T23 | 2,620 | 80,108,220 | 0.03 | 0.03 | 0.03 | 0.00 | 0.00 | 0.00 | 1.03 [0.90-1.18] | 0.98 [0.85-1.13] |
| T24 | 2,890 | 80,118,950 | 0.04 | 0.04 | 0.04 | 0.00 | 0.00 | 0.00 | 0.98 [0.85-1.12] | 0.94 [0.82-1.08] |
| T25 | 1,170 | 80,149,350 | 0.01 | 0.01 | 0.01 | 0.00 | 0.00 | 0.00 | 0.98 [0.79-1.21] | 0.91 [0.73-1.13] |
| T29 | 560 | 80,160,900 | 0.01 | 0.01 | 0.01 | 0.00 | 0.00 | 0.00 | 1.26 [0.94-1.68] | 1.20 [0.89-1.62] |
| T31 | 6,570 | 80,029,540 | 0.09 | 0.08 | 0.08 | 0.01 | 0.00 | 0.00 | 1.05 [0.97-1.15] | 1.00 [0.91-1.09] |
| T36 | 2,360 | 80,094,790 | 0.04 | 0.03 | 0.03 | 0.01 | 0.01 | 0.00 | 1.30 [1.13-1.49]* | 1.09 [0.94-1.25] |
| T37 | 990 | 80,146,250 | 0.02 | 0.01 | 0.01 | 0.00 | 0.00 | 0.00 | 1.40 [1.14-1.73]* | 1.20 [0.97-1.50] |
| T38 | 4,210 | 80,093,140 | 0.07 | 0.05 | 0.05 | 0.02 | 0.02 | 0.01 | 1.34 [1.21-1.48]* | 1.09 [0.98-1.21] |

| | | | | | | | | | | |
|---|---|---|---|---|---|---|---|---|---|---|
| T39 | 43,010 | 78,802,400 | 0.61 | 0.53 | 0.54 | 0.08 | 0.07 | −0.01 | 1.13 [1.09-1.16]* | 0.99 [0.95-1.02] |
| T40 | 24,340 | 79,720,540 | 0.36 | 0.29 | 0.30 | 0.07 | 0.06 | 0.00 | 1.21 [1.16-1.26]* | 1.00 [0.95-1.04] |
| T41 | 620 | 80,163,860 | 0.01 | 0.01 | 0.01 | 0.00 | 0.00 | 0.00 | 1.17 [0.90-1.54] | 1.06 [0.80-1.40] |
| T42 | 18,000 | 79,684,780 | 0.27 | 0.22 | 0.22 | 0.05 | 0.05 | 0.00 | 1.23 [1.17-1.29]* | 1.00 [0.95-1.06] |
| T43 | 27,020 | 79,440,670 | 0.40 | 0.33 | 0.33 | 0.08 | 0.07 | 0.01 | 1.22 [1.17-1.27]* | 1.02 [0.97-1.06] |
| T44 | 5,520 | 80,053,800 | 0.08 | 0.07 | 0.07 | 0.01 | 0.01 | 0.00 | 1.22 [1.11-1.33]* | 1.04 [0.94-1.14] |
| T45 | 6,510 | 80,030,970 | 0.10 | 0.08 | 0.08 | 0.02 | 0.02 | 0.01 | 1.31 [1.21-1.42]* | 1.10 [1.01-1.20] |
| T46 | 5,400 | 80,098,330 | 0.08 | 0.06 | 0.06 | 0.02 | 0.02 | 0.00 | 1.23 [1.12-1.36]* | 1.06 [0.96-1.17] |
| T47 | 2,740 | 80,119,560 | 0.04 | 0.03 | 0.03 | 0.01 | 0.01 | 0.00 | 1.26 [1.11-1.43]* | 1.00 [0.88-1.15] |
| T48 | 850 | 80,144,340 | 0.01 | 0.01 | 0.01 | 0.00 | 0.00 | 0.00 | 1.42 [1.14-1.77]* | 1.11 [0.87-1.41] |
| T49 | 480 | 80,149,180 | 0.01 | 0.01 | 0.01 | 0.00 | 0.00 | 0.00 | 1.30 [0.96-1.77] | 1.08 [0.78-1.48] |
| T50 | 6,650 | 79,952,190 | 0.10 | 0.08 | 0.08 | 0.02 | 0.02 | 0.00 | 1.23 [1.14-1.34]* | 1.02 [0.94-1.12] |
| T51 | 24,580 | 79,542,270 | 0.35 | 0.30 | 0.30 | 0.05 | 0.05 | −0.01 | 1.16 [1.11-1.21]* | 0.98 [0.94-1.02] |
| T54 | 830 | 80,152,230 | 0.01 | 0.01 | 0.01 | 0.00 | 0.00 | 0.00 | 1.01 [0.79-1.29] | 0.94 [0.73-1.20] |
| T56 | 620 | 80,159,030 | 0.01 | 0.01 | 0.01 | 0.00 | 0.00 | 0.00 | 1.17 [0.90-1.54] | 1.05 [0.80-1.39] |
| T59 | 1,740 | 80,132,410 | 0.02 | 0.02 | 0.02 | 0.00 | 0.00 | 0.00 | 1.08 [0.91-1.27] | 0.97 [0.82-1.16] |
| T63 | 840 | 80,150,410 | 0.01 | 0.01 | 0.01 | 0.00 | 0.00 | 0.00 | 0.99 [0.77-1.26] | 0.88 [0.68-1.13] |
| T65 | 720 | 80,152,180 | 0.01 | 0.01 | 0.01 | 0.00 | 0.00 | 0.00 | 1.14 [0.88-1.47] | 0.96 [0.73-1.25] |
| T68 | 9,240 | 80,137,470 | 0.13 | 0.11 | 0.12 | 0.02 | 0.02 | 0.01 | 1.13 [1.05-1.22]* | 1.12 [1.04-1.21]* |
| T74 | 680 | 80,158,250 | 0.01 | 0.01 | 0.01 | 0.00 | 0.00 | 0.00 | 1.17 [0.90-1.52] | 0.95 [0.72-1.25] |
| T75 | 620 | 80,151,990 | 0.01 | 0.01 | 0.01 | 0.00 | 0.00 | 0.00 | 1.24 [0.95-1.63] | 1.13 [0.85-1.49] |
| T78 | 14,180 | 79,777,970 | 0.30 | 0.15 | 0.15 | 0.15 | 0.15 | 0.11 | 1.99 [1.89-2.09]* | 1.58 [1.49-1.66]* |
| T79 | 11,930 | 80,022,060 | 0.16 | 0.15 | 0.15 | 0.02 | 0.02 | 0.01 | 1.11 [1.04-1.18]* | 1.04 [0.97-1.11] |
| T80 | 4,980 | 80,107,410 | 0.08 | 0.06 | 0.06 | 0.02 | 0.02 | 0.02 | 1.39 [1.27-1.53]* | 1.28 [1.17-1.41]* |
| T81 | 163,730 | 77,368,020 | 2.62 | 2.01 | 2.02 | 0.62 | 0.61 | 0.41 | 1.30 [1.28-1.32]* | 1.19 [1.17-1.21]* |
| T82 | 39,240 | 79,719,890 | 0.63 | 0.46 | 0.47 | 0.16 | 0.16 | 0.11 | 1.34 [1.30-1.39]* | 1.21 [1.17-1.25]* |
| T83 | 41,320 | 79,875,030 | 0.59 | 0.50 | 0.51 | 0.09 | 0.08 | 0.05 | 1.15 [1.12-1.20]* | 1.10 [1.06-1.14]* |
| T84 | 54,600 | 79,256,690 | 0.86 | 0.65 | 0.65 | 0.21 | 0.21 | 0.15 | 1.32 [1.28-1.36]* | 1.21 [1.18-1.25]* |
| T85 | 33,240 | 79,734,140 | 0.52 | 0.39 | 0.40 | 0.13 | 0.13 | 0.09 | 1.32 [1.28-1.37]* | 1.21 [1.17-1.26]* |
| T86 | 4,710 | 80,098,320 | 0.08 | 0.06 | 0.06 | 0.02 | 0.02 | 0.02 | 1.36 [1.24-1.49]* | 1.26 [1.15-1.39]* |
| T87 | 3,050 | 80,131,490 | 0.05 | 0.04 | 0.04 | 0.01 | 0.01 | 0.01 | 1.30 [1.16-1.47]* | 1.14 [1.01-1.29] |
| T88 | 10,200 | 79,964,560 | 0.18 | 0.12 | 0.12 | 0.06 | 0.06 | 0.04 | 1.49 [1.40-1.59]* | 1.29 [1.20-1.37]* |
| T90 | 8,480 | 79,964,840 | 0.13 | 0.10 | 0.10 | 0.03 | 0.03 | 0.02 | 1.31 [1.22-1.41]* | 1.19 [1.11-1.29]* |
| T91 | 4,080 | 80,090,070 | 0.06 | 0.05 | 0.05 | 0.01 | 0.01 | 0.00 | 1.24 [1.11-1.37]* | 1.08 [0.97-1.21] |
| T92 | 6,870 | 79,989,050 | 0.10 | 0.08 | 0.08 | 0.02 | 0.02 | 0.01 | 1.20 [1.11-1.30]* | 1.11 [1.02-1.21] |
| T93 | 9,080 | 79,946,010 | 0.13 | 0.11 | 0.11 | 0.03 | 0.03 | 0.02 | 1.25 [1.16-1.34]* | 1.14 [1.06-1.22]* |
| T94 | 850 | 80,156,110 | 0.01 | 0.01 | 0.01 | 0.00 | 0.00 | 0.00 | 1.24 [0.98-1.56] | 1.12 [0.88-1.42] |
| T95 | 570 | 80,151,750 | 0.01 | 0.01 | 0.01 | 0.00 | 0.00 | 0.00 | 1.31 [0.99-1.72] | 1.26 [0.95-1.68] |
| T98 | 1,420 | 80,146,700 | 0.02 | 0.02 | 0.02 | 0.01 | 0.01 | 0.00 | 1.34 [1.13-1.60]* | 1.21 [1.01-1.44] |
| 18+ - U00-U89 | | | | | | | | | | |
| U07 | 51,760 | 80,122,450 | 0.82 | 0.61 | 0.65 | 0.21 | 0.16 | 0.09 | 1.25 [1.21-1.29]* | 1.13 [1.09-1.17]* |
| U80 | 17,660 | 80,066,780 | 0.31 | 0.20 | 0.20 | 0.11 | 0.10 | 0.08 | 1.50 [1.43-1.57]* | 1.36 [1.29-1.43]* |
| U81 | 1,050 | 80,167,920 | 0.02 | 0.01 | 0.01 | 0.00 | 0.00 | 0.00 | 1.18 [0.95-1.46] | 1.08 [0.87-1.34] |
| U82 | 34,980 | 80,078,740 | 0.59 | 0.40 | 0.42 | 0.19 | 0.17 | 0.12 | 1.39 [1.34-1.44]* | 1.25 [1.20-1.30]* |
| U83 | 20,870 | 80,120,260 | 0.34 | 0.24 | 0.26 | 0.10 | 0.09 | 0.06 | 1.34 [1.28-1.40]* | 1.20 [1.14-1.26]* |
| U88 | 4,320 | 80,150,500 | 0.07 | 0.05 | 0.05 | 0.02 | 0.01 | 0.01 | 1.28 [1.15-1.42]* | 1.16 [1.04-1.29]* |
| U89 | 4,860 | 80,145,240 | 0.08 | 0.06 | 0.06 | 0.02 | 0.02 | 0.01 | 1.34 [1.21-1.47]* | 1.20 [1.08-1.32]* |
| 18+ - V00-Y99 | | | | | | | | | | |
| V03 | 6,030 | 79,967,890 | 0.08 | 0.07 | 0.07 | 0.01 | 0.01 | 0.00 | 1.10 [1.00-1.20] | 1.05 [0.96-1.15] |
| V04 | 500 | 80,157,460 | 0.01 | 0.01 | 0.01 | 0.00 | 0.00 | 0.00 | 1.18 [0.87-1.61] | 1.14 [0.83-1.55] |
| V09 | 1,680 | 80,145,620 | 0.02 | 0.02 | 0.02 | 0.00 | 0.01 | 0.00 | 1.29 [1.09-1.53]* | 1.13 [0.95-1.35] |
| V13 | 2,210 | 80,113,960 | 0.03 | 0.03 | 0.03 | 0.00 | 0.00 | 0.00 | 0.90 [0.77-1.05] | 0.90 [0.77-1.06] |
| V17 | 700 | 80,156,250 | 0.01 | 0.01 | 0.01 | 0.00 | 0.00 | 0.00 | 0.84 [0.63-1.11] | 0.79 [0.59-1.05] |
| V18 | 8,350 | 79,864,680 | 0.10 | 0.10 | 0.10 | 0.00 | 0.00 | 0.00 | 1.02 [0.94-1.10] | 1.00 [0.93-1.09] |
| V19 | 1,320 | 80,128,720 | 0.02 | 0.02 | 0.02 | 0.00 | 0.00 | 0.00 | 1.01 [0.84-1.23] | 0.99 [0.81-1.21] |
| V23 | 3,570 | 80,084,500 | 0.04 | 0.05 | 0.04 | 0.00 | 0.00 | 0.00 | 0.97 [0.86-1.09] | 0.94 [0.83-1.06] |
| V27 | 790 | 80,151,570 | 0.01 | 0.01 | 0.01 | 0.00 | 0.00 | 0.00 | 0.84 [0.65-1.09] | 0.78 [0.59-1.03] |
| V28 | 4,120 | 80,059,270 | 0.05 | 0.05 | 0.05 | 0.00 | 0.00 | 0.00 | 0.98 [0.88-1.09] | 0.93 [0.82-1.04] |
| V29 | 1,050 | 80,136,530 | 0.01 | 0.01 | 0.01 | 0.00 | 0.00 | 0.00 | 1.06 [0.86-1.32] | 1.00 [0.80-1.25] |
| V43 | 8,300 | 79,959,530 | 0.12 | 0.10 | 0.10 | 0.01 | 0.01 | 0.01 | 1.14 [1.06-1.23]* | 1.05 [0.98-1.14] |
| V44 | 790 | 80,152,690 | 0.01 | 0.01 | 0.01 | 0.00 | 0.00 | 0.00 | 1.10 [0.87-1.41] | 1.04 [0.81-1.33] |
| V47 | 3,790 | 80,087,750 | 0.06 | 0.05 | 0.05 | 0.01 | 0.01 | 0.01 | 1.24 [1.11-1.38]* | 1.16 [1.04-1.30] |
| V48 | 4,790 | 80,093,080 | 0.07 | 0.06 | 0.06 | 0.01 | 0.01 | 0.00 | 1.15 [1.05-1.28]* | 1.07 [0.97-1.19] |
| V49 | 3,360 | 80,070,950 | 0.05 | 0.04 | 0.04 | 0.01 | 0.00 | 0.00 | 1.12 [0.99-1.26] | 1.03 [0.92-1.17] |
| V78 | 1,880 | 80,152,350 | 0.03 | 0.02 | 0.02 | 0.00 | 0.00 | 0.00 | 1.19 [1.01-1.40] | 1.12 [0.95-1.32] |
| V79 | 590 | 80,162,900 | 0.01 | 0.01 | 0.01 | 0.00 | 0.00 | 0.00 | 1.02 [0.76-1.36] | 0.89 [0.66-1.21] |
| V80 | 4,020 | 80,057,370 | 0.05 | 0.05 | 0.05 | −0.01 | −0.01 | −0.01 | 0.88 [0.79-0.99] | 0.87 [0.77-0.98] |
| V82 | 470 | 80,166,550 | 0.01 | 0.01 | 0.01 | 0.00 | 0.00 | 0.00 | 1.52 [1.12-2.07]* | 1.47 [1.08-2.00] |
| V86 | 720 | 80,155,990 | 0.01 | 0.01 | 0.01 | 0.00 | 0.00 | 0.00 | 1.04 [0.80-1.35] | 0.96 [0.73-1.26] |
| V89 | 1,460 | 80,099,500 | 0.02 | 0.02 | 0.02 | 0.00 | 0.00 | 0.00 | 1.28 [1.08-1.53]* | 1.19 [0.99-1.42] |
| W00 | 7,320 | 80,044,360 | 0.10 | 0.09 | 0.09 | 0.01 | 0.01 | 0.01 | 1.10 [1.02-1.19] | 1.08 [0.99-1.17] |
| W01 | 142,600 | 78,473,160 | 2.11 | 1.75 | 1.82 | 0.35 | 0.28 | 0.18 | 1.16 [1.13-1.18]* | 1.09 [1.07-1.11]* |
| W02 | 2,860 | 80,021,660 | 0.04 | 0.04 | 0.04 | 0.00 | 0.00 | 0.00 | 1.01 [0.89-1.15] | 0.98 [0.85-1.12] |
| W03 | 5,590 | 79,962,300 | 0.07 | 0.07 | 0.07 | 0.00 | 0.00 | 0.00 | 1.06 [0.96-1.16] | 0.99 [0.90-1.09] |
| W05 | 3,200 | 80,147,700 | 0.05 | 0.04 | 0.04 | 0.01 | 0.01 | 0.01 | 1.26 [1.11-1.42]* | 1.15 [1.01-1.30] |
| W06 | 33,580 | 79,988,210 | 0.51 | 0.40 | 0.42 | 0.10 | 0.08 | 0.05 | 1.19 [1.15-1.24]* | 1.11 [1.07-1.16]* |
| W07 | 18,170 | 80,024,190 | 0.27 | 0.22 | 0.23 | 0.05 | 0.04 | 0.03 | 1.19 [1.13-1.25]* | 1.12 [1.06-1.18]* |
| W08 | 4,390 | 80,115,420 | 0.07 | 0.05 | 0.05 | 0.01 | 0.01 | 0.01 | 1.24 [1.11-1.37]* | 1.14 [1.02-1.27] |
| W09 | 760 | 79,984,690 | 0.01 | 0.01 | 0.01 | 0.00 | 0.00 | 0.00 | 1.23 [0.97-1.57] | 1.17 [0.91-1.50] |
| W10 | 58,160 | 79,379,430 | 0.85 | 0.71 | 0.71 | 0.14 | 0.14 | 0.09 | 1.19 [1.16-1.23]* | 1.11 [1.08-1.15]* |
| W11 | 9,410 | 79,979,400 | 0.12 | 0.12 | 0.11 | 0.00 | 0.01 | 0.00 | 1.06 [0.99-1.14] | 1.02 [0.94-1.10] |
| W12 | 540 | 80,158,480 | 0.01 | 0.01 | 0.01 | 0.00 | 0.00 | 0.00 | 1.15 [0.86-1.54] | 1.14 [0.85-1.54] |
| W13 | 4,150 | 80,018,020 | 0.05 | 0.05 | 0.05 | 0.00 | 0.00 | 0.00 | 1.05 [0.94-1.17] | 0.98 [0.88-1.10] |
| W17 | 7,440 | 79,948,390 | 0.10 | 0.09 | 0.09 | 0.01 | 0.01 | 0.01 | 1.16 [1.07-1.25]* | 1.09 [1.00-1.18] |
| W18 | 96,200 | 79,431,120 | 1.46 | 1.16 | 1.20 | 0.30 | 0.25 | 0.16 | 1.21 [1.18-1.24]* | 1.13 [1.10-1.15]* |
| W19 | 225,320 | 78,191,600 | 3.43 | 2.76 | 2.89 | 0.66 | 0.54 | 0.37 | 1.19 [1.17-1.21]* | 1.12 [1.10-1.14]* |
| W20 | 4,700 | 80,059,100 | 0.06 | 0.06 | 0.06 | 0.00 | 0.00 | 0.00 | 1.08 [0.97-1.19] | 1.01 [0.91-1.12] |
| W21 | 2,580 | 80,062,700 | 0.03 | 0.03 | 0.03 | 0.00 | 0.00 | 0.00 | 1.11 [0.97-1.27] | 1.08 [0.94-1.24] |
| W22 | 22,440 | 79,761,900 | 0.33 | 0.27 | 0.28 | 0.05 | 0.05 | 0.02 | 1.17 [1.12-1.23]* | 1.08 [1.03-1.13]* |
| W23 | 9,910 | 79,916,890 | 0.13 | 0.12 | 0.12 | 0.01 | 0.01 | 0.00 | 1.08 [1.01-1.16] | 1.03 [0.96-1.11] |
| W25 | 8,020 | 79,891,260 | 0.11 | 0.10 | 0.10 | 0.01 | 0.01 | 0.00 | 1.08 [1.00-1.17] | 1.02 [0.94-1.10] |
| W26 | 8,340 | 80,015,690 | 0.11 | 0.10 | 0.10 | 0.01 | 0.01 | 0.00 | 1.12 [1.04-1.20]* | 1.03 [0.96-1.12] |
| W27 | 3,900 | 80,071,850 | 0.05 | 0.05 | 0.05 | 0.00 | 0.00 | 0.00 | 1.03 [0.92-1.16] | 0.98 [0.87-1.10] |
| W29 | 4,750 | 80,086,990 | 0.06 | 0.06 | 0.06 | 0.00 | 0.00 | 0.00 | 0.98 [0.88-1.08] | 0.95 [0.85-1.05] |
| W31 | 4,160 | 80,070,110 | 0.05 | 0.05 | 0.05 | 0.00 | 0.00 | 0.00 | 1.03 [0.92-1.14] | 1.02 [0.91-1.14] |
| W44 | 9,060 | 79,939,830 | 0.14 | 0.11 | 0.11 | 0.03 | 0.03 | 0.02 | 1.30 [1.21-1.40]* | 1.18 [1.09-1.27]* |
| W45 | 4,870 | 80,026,510 | 0.07 | 0.06 | 0.06 | 0.01 | 0.01 | 0.00 | 1.13 [1.03-1.25] | 1.05 [0.95-1.16] |
| W46 | 910 | 80,167,010 | 0.01 | 0.01 | 0.01 | 0.00 | 0.00 | 0.00 | 1.17 [0.93-1.46] | 0.95 [0.75-1.21] |
| W49 | 1,360 | 80,128,420 | 0.02 | 0.02 | 0.02 | 0.00 | 0.00 | 0.00 | 1.29 [1.08-1.54]* | 1.22 [1.01-1.47] |
| W50 | 6,270 | 79,975,140 | 0.09 | 0.08 | 0.08 | 0.01 | 0.01 | 0.01 | 1.17 [1.07-1.27]* | 1.08 [0.99-1.18] |
| W51 | 3,920 | 80,017,140 | 0.05 | 0.05 | 0.05 | 0.01 | 0.01 | 0.00 | 1.11 [1.00-1.24] | 1.05 [0.94-1.17] |
| W54 | 7,810 | 80,028,180 | 0.10 | 0.10 | 0.10 | 0.01 | 0.01 | 0.00 | 1.04 [0.97-1.13] | 0.97 [0.90-1.05] |
| W55 | 4,530 | 80,099,020 | 0.06 | 0.06 | 0.06 | 0.00 | 0.00 | −0.01 | 1.00 [0.90-1.11] | 0.91 [0.81-1.01] |
| W57 | 5,080 | 80,086,930 | 0.08 | 0.06 | 0.06 | 0.03 | 0.03 | 0.02 | 1.45 [1.32-1.59]* | 1.31 [1.19-1.44]* |
| W60 | 500 | 80,164,570 | 0.01 | 0.01 | 0.01 | 0.00 | 0.00 | 0.00 | 1.47 [1.10-1.96]* | 1.37 [1.02-1.84] |
| W64 | 1,130 | 80,160,880 | 0.02 | 0.01 | 0.01 | 0.00 | 0.00 | 0.00 | 1.10 [0.89-1.36] | 1.02 [0.82-1.26] |
| W78 | 3,640 | 80,163,690 | 0.05 | 0.04 | 0.05 | 0.01 | 0.01 | 0.00 | 1.11 [0.98-1.25] | 1.06 [0.94-1.20] |

| | | | | | | | | | | |
|---|---|---|---|---|---|---|---|---|---|---|
| W79 | 5,140 | 80,141,540 | 0.08 | 0.06 | 0.06 | 0.02 | 0.02 | 0.01 | 1.26 [1.14-1.39]* | 1.17 [1.06-1.29]* |
| W80 | 2,090 | 80,165,370 | 0.03 | 0.03 | 0.03 | 0.01 | 0.01 | 0.00 | 1.23 [1.05-1.44]* | 1.12 [0.95-1.31] |
| W84 | 2,140 | 80,167,710 | 0.03 | 0.03 | 0.03 | 0.00 | 0.00 | 0.00 | 1.06 [0.91-1.24] | 0.99 [0.85-1.17] |
| X09 | 700 | 80,156,530 | 0.01 | 0.01 | 0.01 | 0.00 | 0.00 | 0.00 | 1.02 [0.78-1.34] | 0.98 [0.75-1.29] |
| X10 | 1,330 | 80,140,210 | 0.02 | 0.02 | 0.02 | 0.00 | 0.00 | 0.00 | 1.19 [0.99-1.43] | 1.09 [0.90-1.32] |
| X12 | 1,180 | 80,147,230 | 0.02 | 0.01 | 0.02 | 0.00 | 0.00 | 0.00 | 1.01 [0.82-1.24] | 0.93 [0.75-1.15] |
| X16 | 900 | 80,163,270 | 0.01 | 0.01 | 0.01 | 0.00 | 0.00 | 0.00 | 1.14 [0.89-1.45] | 1.05 [0.82-1.35] |
| X23 | 1,070 | 80,151,580 | 0.02 | 0.01 | 0.01 | 0.00 | 0.00 | 0.00 | 1.19 [0.97-1.47] | 1.11 [0.89-1.37] |
| X31 | 1,300 | 80,165,890 | 0.02 | 0.02 | 0.02 | 0.00 | 0.00 | 0.00 | 1.00 [0.81-1.24] | 0.99 [0.80-1.23] |
| X40 | 9,270 | 79,921,370 | 0.13 | 0.11 | 0.12 | 0.02 | 0.02 | 0.00 | 1.16 [1.08-1.24]* | 1.01 [0.94-1.08] |
| X41 | 6,870 | 79,990,790 | 0.10 | 0.08 | 0.08 | 0.02 | 0.02 | 0.01 | 1.26 [1.17-1.37]* | 1.06 [0.97-1.15] |
| X42 | 7,390 | 80,076,470 | 0.11 | 0.09 | 0.09 | 0.03 | 0.02 | 0.00 | 1.28 [1.18-1.38]* | 1.04 [0.96-1.13] |
| X43 | 1,290 | 80,149,660 | 0.02 | 0.02 | 0.02 | 0.00 | 0.00 | 0.00 | 1.20 [0.98-1.46] | 1.08 [0.88-1.32] |
| X44 | 7,950 | 80,014,310 | 0.12 | 0.09 | 0.10 | 0.03 | 0.03 | 0.01 | 1.29 [1.20-1.39]* | 1.10 [1.02-1.19] |
| X45 | 3,020 | 80,086,820 | 0.04 | 0.04 | 0.04 | 0.01 | 0.01 | 0.00 | 1.16 [1.03-1.31] | 0.99 [0.87-1.13] |
| X47 | 1,020 | 80,151,400 | 0.01 | 0.01 | 0.01 | 0.00 | 0.00 | 0.00 | 1.10 [0.89-1.37] | 1.00 [0.80-1.25] |
| X49 | 2,250 | 80,103,320 | 0.04 | 0.03 | 0.03 | 0.01 | 0.01 | 0.01 | 1.38 [1.20-1.58]* | 1.20 [1.04-1.38] |
| X50 | 15,580 | 79,894,920 | 0.22 | 0.19 | 0.19 | 0.03 | 0.03 | 0.01 | 1.15 [1.09-1.21]* | 1.05 [1.00-1.12] |
| X51 | 1,190 | 80,153,010 | 0.02 | 0.01 | 0.01 | 0.00 | 0.00 | 0.00 | 1.14 [0.94-1.39] | 1.10 [0.90-1.34] |
| X58 | 4,120 | 80,114,110 | 0.07 | 0.05 | 0.05 | 0.02 | 0.02 | 0.02 | 1.45 [1.31-1.61]* | 1.28 [1.15-1.43]* |
| X59 | 58,500 | 79,077,690 | 0.89 | 0.71 | 0.72 | 0.18 | 0.17 | 0.10 | 1.24 [1.20-1.27]* | 1.13 [1.10-1.17]* |
| X60 | 35,590 | 79,033,630 | 0.50 | 0.44 | 0.45 | 0.06 | 0.06 | −0.01 | 1.13 [1.09-1.17]* | 0.98 [0.95-1.02] |
| X61 | 30,370 | 79,346,610 | 0.45 | 0.37 | 0.37 | 0.08 | 0.08 | 0.00 | 1.21 [1.16-1.25]* | 1.00 [0.96-1.05] |
| X62 | 16,980 | 79,824,230 | 0.25 | 0.21 | 0.21 | 0.04 | 0.04 | −0.01 | 1.19 [1.13-1.25]* | 0.97 [0.92-1.02] |
| X63 | 4,120 | 80,081,820 | 0.06 | 0.05 | 0.05 | 0.01 | 0.01 | 0.00 | 1.21 [1.09-1.34]* | 1.01 [0.90-1.12] |
| X64 | 13,370 | 79,780,540 | 0.20 | 0.16 | 0.16 | 0.04 | 0.04 | 0.00 | 1.23 [1.16-1.30]* | 1.02 [0.96-1.09] |
| X65 | 22,350 | 79,634,790 | 0.32 | 0.27 | 0.27 | 0.05 | 0.04 | −0.01 | 1.16 [1.11-1.22]* | 0.98 [0.93-1.02] |
| X69 | 1,210 | 80,140,730 | 0.01 | 0.02 | 0.02 | 0.00 | 0.00 | 0.00 | 0.94 [0.77-1.16] | 0.81 [0.65-1.00] |
| X70 | 980 | 80,156,280 | 0.01 | 0.01 | 0.01 | 0.00 | 0.00 | 0.00 | 0.99 [0.79-1.24] | 0.87 [0.69-1.10] |
| X78 | 7,580 | 79,991,860 | 0.10 | 0.09 | 0.09 | 0.01 | 0.01 | −0.01 | 1.09 [1.00-1.17] | 0.90 [0.82-0.97]* |
| X83 | 700 | 80,162,020 | 0.01 | 0.01 | 0.01 | 0.00 | 0.00 | 0.00 | 1.13 [0.88-1.47] | 0.97 [0.74-1.27] |
| X84 | 1,070 | 80,149,800 | 0.01 | 0.01 | 0.01 | 0.00 | 0.00 | 0.00 | 1.04 [0.84-1.28] | 0.81 [0.65-1.02] |
| X99 | 4,390 | 80,057,570 | 0.06 | 0.05 | 0.05 | 0.00 | 0.00 | 0.00 | 1.09 [0.98-1.21] | 1.02 [0.92-1.14] |
| Y00 | 2,320 | 80,099,550 | 0.03 | 0.03 | 0.03 | 0.00 | 0.00 | 0.00 | 1.12 [0.97-1.29] | 0.99 [0.85-1.15] |
| Y04 | 18,850 | 79,553,480 | 0.26 | 0.23 | 0.23 | 0.03 | 0.03 | 0.01 | 1.12 [1.07-1.18]* | 1.02 [0.97-1.08] |
| Y07 | 770 | 80,153,460 | 0.01 | 0.01 | 0.01 | 0.00 | 0.00 | 0.00 | 1.01 [0.78-1.30] | 0.82 [0.63-1.08] |
| Y08 | 940 | 80,148,860 | 0.01 | 0.01 | 0.01 | 0.00 | 0.00 | 0.00 | 1.00 [0.79-1.26] | 0.93 [0.73-1.19] |
| Y09 | 2,820 | 80,069,120 | 0.04 | 0.03 | 0.03 | 0.00 | 0.00 | 0.00 | 1.13 [1.00-1.29] | 1.02 [0.89-1.16] |
| Y40 | 17,840 | 79,974,450 | 0.32 | 0.20 | 0.21 | 0.11 | 0.11 | 0.08 | 1.52 [1.45-1.60]* | 1.34 [1.27-1.41]* |
| Y41 | 3,750 | 80,128,630 | 0.06 | 0.04 | 0.04 | 0.02 | 0.02 | 0.01 | 1.45 [1.30-1.62]* | 1.27 [1.14-1.42]* |
| Y42 | 14,950 | 80,020,910 | 0.25 | 0.17 | 0.18 | 0.08 | 0.08 | 0.04 | 1.43 [1.36-1.51]* | 1.20 [1.14-1.27]* |
| Y43 | 31,040 | 79,949,230 | 0.46 | 0.37 | 0.38 | 0.09 | 0.09 | 0.07 | 1.23 [1.18-1.28]* | 1.18 [1.13-1.22]* |
| Y44 | 10,690 | 80,085,470 | 0.16 | 0.13 | 0.13 | 0.04 | 0.03 | 0.02 | 1.26 [1.18-1.34]* | 1.16 [1.08-1.24]* |
| Y45 | 24,700 | 79,930,580 | 0.40 | 0.29 | 0.29 | 0.11 | 0.10 | 0.07 | 1.35 [1.30-1.41]* | 1.20 [1.15-1.25]* |
| Y46 | 3,410 | 80,132,080 | 0.06 | 0.04 | 0.04 | 0.02 | 0.02 | 0.01 | 1.40 [1.25-1.57]* | 1.25 [1.11-1.40]* |
| Y47 | 1,670 | 80,155,480 | 0.03 | 0.02 | 0.02 | 0.01 | 0.01 | 0.00 | 1.37 [1.16-1.62]* | 1.22 [1.03-1.45] |
| Y48 | 1,920 | 80,147,210 | 0.03 | 0.02 | 0.02 | 0.01 | 0.01 | 0.00 | 1.32 [1.13-1.54]* | 1.16 [0.99-1.35] |
| Y49 | 5,990 | 80,111,520 | 0.10 | 0.07 | 0.07 | 0.03 | 0.02 | 0.02 | 1.35 [1.24-1.47]* | 1.23 [1.12-1.34]* |
| Y51 | 9,330 | 80,088,160 | 0.14 | 0.11 | 0.11 | 0.03 | 0.03 | 0.02 | 1.27 [1.18-1.36]* | 1.18 [1.10-1.27]* |
| Y52 | 22,390 | 80,003,590 | 0.35 | 0.26 | 0.26 | 0.09 | 0.09 | 0.06 | 1.34 [1.28-1.40]* | 1.21 [1.16-1.27]* |
| Y53 | 4,190 | 80,144,200 | 0.07 | 0.05 | 0.05 | 0.02 | 0.02 | 0.01 | 1.39 [1.25-1.54]* | 1.19 [1.07-1.32]* |
| Y54 | 17,560 | 80,071,460 | 0.29 | 0.20 | 0.21 | 0.08 | 0.08 | 0.05 | 1.37 [1.30-1.44]* | 1.23 [1.17-1.30]* |
| Y55 | 2,280 | 80,149,480 | 0.04 | 0.03 | 0.03 | 0.01 | 0.01 | 0.01 | 1.43 [1.25-1.63]* | 1.20 [1.04-1.39] |
| Y56 | 1,780 | 80,138,740 | 0.04 | 0.02 | 0.02 | 0.02 | 0.02 | 0.01 | 1.87 [1.62-2.17]* | 1.63 [1.40-1.89]* |
| Y57 | 9,910 | 80,089,770 | 0.16 | 0.12 | 0.12 | 0.05 | 0.04 | 0.03 | 1.38 [1.29-1.47]* | 1.21 [1.13-1.30]* |
| Y59 | 900 | 80,161,790 | 0.02 | 0.01 | 0.01 | 0.01 | 0.01 | 0.00 | 1.55 [1.25-1.92]* | 1.38 [1.11-1.72]* |
| Y60 | 12,400 | 80,062,560 | 0.19 | 0.15 | 0.15 | 0.04 | 0.04 | 0.03 | 1.27 [1.20-1.35]* | 1.17 [1.10-1.25]* |
| Y65 | 1,620 | 80,154,170 | 0.02 | 0.02 | 0.02 | 0.00 | 0.00 | 0.00 | 1.24 [1.05-1.47] | 1.13 [0.95-1.35] |
| Y71 | 730 | 80,163,590 | 0.01 | 0.01 | 0.01 | 0.00 | 0.00 | 0.00 | 1.59 [1.26-2.02]* | 1.42 [1.11-1.82]* |
| Y73 | 740 | 80,164,620 | 0.01 | 0.01 | 0.01 | 0.00 | 0.00 | 0.00 | 0.94 [0.72-1.23] | 0.89 [0.67-1.17] |
| Y76 | 460 | 80,162,470 | 0.01 | 0.01 | 0.01 | 0.00 | 0.00 | 0.00 | 1.37 [1.01-1.85] | 1.26 [0.92-1.71] |
| Y77 | 820 | 80,164,790 | 0.01 | 0.01 | 0.01 | 0.00 | 0.00 | 0.00 | 0.98 [0.77-1.26] | 0.91 [0.70-1.17] |
| Y79 | 1,750 | 80,143,830 | 0.03 | 0.02 | 0.02 | 0.00 | 0.01 | 0.00 | 1.24 [1.06-1.46]* | 1.16 [0.98-1.37] |
| Y82 | 600 | 80,163,540 | 0.01 | 0.01 | 0.01 | 0.00 | 0.00 | 0.00 | 1.27 [0.97-1.68] | 1.16 [0.88-1.54] |
| Y83 | 217,020 | 76,786,630 | 3.48 | 2.68 | 2.70 | 0.80 | 0.78 | 0.52 | 1.29 [1.27-1.31]* | 1.17 [1.16-1.19]* |
| Y84 | 49,500 | 79,773,520 | 0.75 | 0.59 | 0.60 | 0.16 | 0.15 | 0.10 | 1.25 [1.22-1.29]* | 1.16 [1.12-1.19]* |
| Y85 | 5,330 | 80,057,740 | 0.08 | 0.06 | 0.06 | 0.02 | 0.02 | 0.01 | 1.30 [1.19-1.42]* | 1.15 [1.05-1.27]* |
| Y86 | 9,340 | 79,943,580 | 0.13 | 0.11 | 0.11 | 0.02 | 0.02 | 0.01 | 1.20 [1.12-1.29]* | 1.10 [1.02-1.18] |
| Y87 | 1,590 | 80,133,190 | 0.02 | 0.02 | 0.02 | 0.01 | 0.01 | 0.00 | 1.34 [1.14-1.57]* | 1.18 [0.99-1.40] |
| Y88 | 1,800 | 80,143,330 | 0.03 | 0.02 | 0.02 | 0.00 | 0.00 | 0.00 | 1.18 [1.00-1.38] | 1.01 [0.85-1.20] |
| Y89 | 590 | 80,159,310 | 0.01 | 0.01 | 0.01 | 0.00 | 0.00 | 0.00 | 1.31 [1.00-1.72] | 1.19 [0.90-1.57] |
| Y90 | 1,020 | 80,158,460 | 0.01 | 0.01 | 0.01 | 0.00 | 0.00 | 0.00 | 1.17 [0.94-1.44] | 1.07 [0.86-1.33] |
| Y91 | 2,240 | 80,123,340 | 0.03 | 0.03 | 0.03 | 0.00 | 0.00 | 0.00 | 1.17 [1.01-1.35] | 1.07 [0.92-1.24] |
| Y95 | 117,240 | 79,889,980 | 1.81 | 1.39 | 1.46 | 0.42 | 0.35 | 0.22 | 1.24 [1.21-1.27]* | 1.14 [1.11-1.16]* |
| 18+ - Z00-Z99 | | | | | | | | | | |
| Z00 | 4,760 | 80,053,020 | 0.07 | 0.06 | 0.06 | 0.01 | 0.01 | 0.01 | 1.22 [1.11-1.34]* | 1.13 [1.03-1.25] |
| Z01 | 13,740 | 79,268,950 | 0.23 | 0.16 | 0.16 | 0.06 | 0.07 | 0.04 | 1.41 [1.34-1.49]* | 1.22 [1.15-1.29]* |
| Z02 | 19,070 | 79,974,960 | 0.27 | 0.23 | 0.23 | 0.04 | 0.04 | 0.02 | 1.17 [1.11-1.23]* | 1.07 [1.02-1.13]* |
| Z03 | 131,600 | 77,667,030 | 2.13 | 1.60 | 1.61 | 0.54 | 0.52 | 0.33 | 1.32 [1.30-1.35]* | 1.19 [1.16-1.21]* |
| Z04 | 20,310 | 79,330,850 | 0.31 | 0.24 | 0.24 | 0.07 | 0.07 | 0.04 | 1.27 [1.21-1.33]* | 1.16 [1.10-1.21]* |
| Z08 | 43,850 | 79,379,910 | 0.59 | 0.54 | 0.54 | 0.04 | 0.04 | 0.03 | 1.08 [1.05-1.12]* | 1.05 [1.02-1.09]* |
| Z09 | 80,390 | 78,605,890 | 1.25 | 0.97 | 0.97 | 0.28 | 0.28 | 0.17 | 1.29 [1.26-1.32]* | 1.16 [1.13-1.19]* |
| Z11 | 250,740 | 79,677,600 | 3.81 | 3.00 | 3.14 | 0.81 | 0.67 | 0.33 | 1.21 [1.20-1.23]* | 1.10 [1.08-1.11]* |
| Z12 | 62,920 | 79,548,940 | 0.88 | 0.77 | 0.76 | 0.11 | 0.12 | 0.10 | 1.16 [1.13-1.19]* | 1.13 [1.09-1.16]* |
| Z13 | 159,270 | 78,022,810 | 2.50 | 1.94 | 1.98 | 0.55 | 0.52 | 0.34 | 1.26 [1.24-1.28]* | 1.16 [1.14-1.18]* |
| Z20 | 8,290 | 80,137,560 | 0.13 | 0.10 | 0.10 | 0.03 | 0.02 | 0.02 | 1.24 [1.15-1.34]* | 1.13 [1.05-1.23]* |
| Z22 | 78,700 | 79,358,270 | 1.34 | 0.92 | 0.95 | 0.43 | 0.39 | 0.31 | 1.41 [1.38-1.45]* | 1.29 [1.26-1.33]* |
| Z24 | 1,540 | 80,113,220 | 0.02 | 0.02 | 0.02 | 0.00 | 0.00 | 0.00 | 1.13 [0.95-1.35] | 1.12 [0.93-1.33] |
| Z27 | 3,010 | 80,085,420 | 0.04 | 0.04 | 0.04 | 0.00 | 0.00 | 0.00 | 1.06 [0.93-1.20] | 1.01 [0.89-1.15] |
| Z29 | 42,460 | 79,728,430 | 0.65 | 0.51 | 0.52 | 0.15 | 0.13 | 0.09 | 1.25 [1.21-1.29]* | 1.15 [1.11-1.19]* |
| Z30 | 52,160 | 77,607,840 | 0.76 | 0.65 | 0.65 | 0.11 | 0.11 | 0.07 | 1.17 [1.14-1.21]* | 1.10 [1.06-1.13]* |
| Z31 | 950 | 80,060,180 | 0.01 | 0.01 | 0.01 | 0.00 | 0.00 | 0.00 | 1.06 [0.85-1.33] | 0.98 [0.77-1.23] |
| Z32 | 3,720 | 79,996,490 | 0.06 | 0.04 | 0.05 | 0.01 | 0.01 | 0.01 | 1.23 [1.10-1.37]* | 1.13 [1.01-1.26] |
| Z33 | 34,560 | 79,037,830 | 0.55 | 0.41 | 0.43 | 0.13 | 0.11 | 0.06 | 1.26 [1.22-1.31]* | 1.12 [1.07-1.16]* |
| Z34 | 94,890 | 77,369,210 | 1.43 | 1.18 | 1.28 | 0.25 | 0.15 | 0.09 | 1.12 [1.09-1.14]* | 1.07 [1.04-1.09]* |
| Z35 | 84,850 | 78,583,120 | 1.22 | 1.05 | 1.13 | 0.17 | 0.09 | 0.05 | 1.08 [1.05-1.11]* | 1.05 [1.02-1.07]* |
| Z36 | 17,740 | 78,969,760 | 0.25 | 0.22 | 0.23 | 0.03 | 0.03 | 0.02 | 1.12 [1.07-1.18]* | 1.09 [1.03-1.14]* |
| Z37 | 324,930 | 65,865,400 | 6.34 | 4.62 | 5.67 | 1.72 | 0.67 | 0.49 | 1.12 [1.10-1.13]* | 1.08 [1.07-1.10]* |
| Z38 | 960 | 77,989,370 | 0.01 | 0.01 | 0.01 | 0.00 | 0.00 | 0.00 | 1.18 [0.95-1.47] | 1.18 [0.94-1.48] |
| Z39 | 28,840 | 79,114,330 | 0.41 | 0.35 | 0.38 | 0.06 | 0.04 | 0.03 | 1.09 [1.05-1.14]* | 1.07 [1.03-1.12]* |
| Z40 | 5,140 | 80,095,860 | 0.07 | 0.06 | 0.06 | 0.01 | 0.01 | 0.00 | 1.11 [1.00-1.22] | 1.05 [0.95-1.16] |
| Z41 | 4,360 | 80,004,280 | 0.07 | 0.05 | 0.05 | 0.01 | 0.01 | 0.01 | 1.24 [1.12-1.37]* | 1.13 [1.02-1.26] |
| Z42 | 14,240 | 79,876,310 | 0.20 | 0.17 | 0.17 | 0.03 | 0.03 | 0.02 | 1.17 [1.11-1.24]* | 1.10 [1.04-1.17]* |
| Z43 | 19,240 | 79,830,190 | 0.28 | 0.23 | 0.23 | 0.05 | 0.05 | 0.04 | 1.24 [1.18-1.30]* | 1.17 [1.11-1.23]* |
| Z45 | 42,100 | 79,622,760 | 0.62 | 0.51 | 0.51 | 0.11 | 0.12 | 0.07 | 1.23 [1.19-1.27]* | 1.13 [1.10-1.17]* |
| Z46 | 54,790 | 79,317,560 | 0.78 | 0.67 | 0.67 | 0.10 | 0.10 | 0.07 | 1.15 [1.12-1.19]* | 1.10 [1.06-1.13]* |
| Z47 | 26,200 | 79,136,700 | 0.38 | 0.32 | 0.32 | 0.06 | 0.06 | 0.03 | 1.19 [1.14-1.24]* | 1.10 [1.06-1.15]* |

| | | | | | | | | | | |
|---|---|---|---|---|---|---|---|---|---|---|
| Z48 | 12,110 | 79,815,110 | 0.18 | 0.14 | 0.14 | 0.04 | 0.04 | 0.03 | 1.30 [1.22-1.38]* | 1.21 [1.14-1.29]* |
| Z49 | 8,500 | 80,071,260 | 0.14 | 0.10 | 0.10 | 0.04 | 0.05 | 0.04 | 1.46 [1.37-1.57]* | 1.35 [1.25-1.45]* |
| Z50 | 322,690 | 78,181,860 | 4.91 | 3.96 | 4.13 | 0.95 | 0.77 | 0.48 | 1.19 [1.17-1.20]* | 1.11 [1.09-1.12]* |
| Z51 | 378,850 | 77,113,120 | 5.80 | 4.72 | 4.81 | 1.08 | 1.00 | 0.66 | 1.21 [1.19-1.22]* | 1.13 [1.12-1.14]* |
| Z52 | 2,000 | 80,130,110 | 0.02 | 0.02 | 0.02 | 0.00 | 0.00 | 0.00 | 1.02 [0.87-1.19] | 1.03 [0.88-1.21] |
| Z53 | 304,520 | 74,616,060 | 5.11 | 3.86 | 3.87 | 1.26 | 1.25 | 0.84 | 1.32 [1.31-1.34]* | 1.20 [1.18-1.21]* |
| Z54 | 2,900 | 80,101,430 | 0.05 | 0.03 | 0.03 | 0.01 | 0.01 | 0.01 | 1.35 [1.20-1.53]* | 1.29 [1.14-1.46]* |
| Z56 | 4,120 | 80,117,970 | 0.06 | 0.05 | 0.05 | 0.01 | 0.01 | 0.00 | 1.16 [1.05-1.29]* | 1.00 [0.89-1.11] |
| Z57 | 12,070 | 80,101,130 | 0.18 | 0.14 | 0.14 | 0.04 | 0.04 | 0.03 | 1.26 [1.18-1.34]* | 1.16 [1.09-1.24]* |
| Z58 | 6,910 | 80,132,670 | 0.11 | 0.08 | 0.09 | 0.02 | 0.02 | 0.01 | 1.23 [1.13-1.33]* | 1.09 [1.00-1.19] |
| Z59 | 8,860 | 80,068,980 | 0.12 | 0.11 | 0.11 | 0.01 | 0.01 | 0.00 | 1.09 [1.01-1.17] | 0.98 [0.91-1.06] |
| Z60 | 104,450 | 79,022,890 | 1.53 | 1.28 | 1.30 | 0.26 | 0.24 | 0.15 | 1.18 [1.16-1.21]* | 1.11 [1.08-1.13]* |
| Z61 | 2,500 | 80,118,310 | 0.03 | 0.03 | 0.03 | 0.00 | 0.00 | −0.01 | 1.03 [0.89-1.18] | 0.86 [0.74-0.99] |
| Z63 | 27,110 | 79,826,810 | 0.40 | 0.33 | 0.34 | 0.07 | 0.06 | 0.01 | 1.17 [1.12-1.22]* | 1.02 [0.98-1.07] |
| Z64 | 810 | 80,131,510 | 0.01 | 0.01 | 0.01 | 0.00 | 0.00 | 0.00 | 1.01 [0.79-1.29] | 0.90 [0.69-1.17] |
| Z65 | 2,280 | 80,137,010 | 0.03 | 0.03 | 0.03 | 0.00 | 0.00 | 0.00 | 1.14 [0.99-1.32] | 0.99 [0.86-1.15] |
| Z71 | 26,060 | 79,816,380 | 0.39 | 0.31 | 0.32 | 0.08 | 0.07 | 0.04 | 1.22 [1.16-1.27]* | 1.11 [1.06-1.16]* |
| Z72 | 224,150 | 74,263,690 | 3.46 | 2.92 | 2.87 | 0.54 | 0.60 | 0.29 | 1.21 [1.19-1.22]* | 1.09 [1.07-1.11]* |
| Z73 | 54,340 | 79,913,130 | 0.81 | 0.65 | 0.68 | 0.15 | 0.12 | 0.07 | 1.18 [1.14-1.22]* | 1.09 [1.06-1.12]* |
| Z74 | 41,950 | 79,925,930 | 0.61 | 0.51 | 0.52 | 0.11 | 0.09 | 0.07 | 1.17 [1.13-1.21]* | 1.12 [1.08-1.16]* |
| Z75 | 82,230 | 79,782,870 | 1.18 | 1.00 | 1.06 | 0.18 | 0.12 | 0.08 | 1.12 [1.09-1.15]* | 1.07 [1.04-1.10]* |
| Z76 | 5,930 | 79,953,090 | 0.09 | 0.07 | 0.07 | 0.01 | 0.01 | 0.01 | 1.17 [1.07-1.28]* | 1.15 [1.05-1.26]* |
| Z80 | 89,420 | 78,940,610 | 1.29 | 1.10 | 1.10 | 0.19 | 0.19 | 0.11 | 1.17 [1.15-1.20]* | 1.09 [1.07-1.12]* |
| Z81 | 8,810 | 80,068,070 | 0.13 | 0.11 | 0.11 | 0.02 | 0.02 | 0.00 | 1.16 [1.08-1.24]* | 1.00 [0.92-1.07] |
| Z82 | 166,430 | 77,553,150 | 2.54 | 2.06 | 2.07 | 0.48 | 0.47 | 0.26 | 1.23 [1.21-1.25]* | 1.11 [1.09-1.13]* |
| Z83 | 61,650 | 79,377,470 | 0.94 | 0.74 | 0.77 | 0.20 | 0.17 | 0.11 | 1.22 [1.19-1.26]* | 1.13 [1.10-1.16]* |
| Z84 | 5,110 | 80,078,420 | 0.08 | 0.06 | 0.06 | 0.02 | 0.02 | 0.01 | 1.25 [1.14-1.37]* | 1.18 [1.07-1.30]* |
| Z85 | 318,270 | 76,715,550 | 4.81 | 4.01 | 4.12 | 0.80 | 0.69 | 0.45 | 1.17 [1.15-1.18]* | 1.10 [1.09-1.12]* |
| Z86 | 992,770 | 70,039,250 | 17.52 | 13.44 | 13.85 | 4.07 | 3.67 | 2.15 | 1.26 [1.26-1.27]* | 1.14 [1.13-1.15]* |
| Z87 | 475,440 | 74,393,440 | 8.00 | 6.04 | 6.18 | 1.96 | 1.82 | 1.08 | 1.30 [1.28-1.31]* | 1.16 [1.14-1.17]* |
| Z88 | 493,180 | 73,415,660 | 9.21 | 6.18 | 6.30 | 3.04 | 2.91 | 1.97 | 1.46 [1.45-1.48]* | 1.27 [1.26-1.29]* |
| Z89 | 14,800 | 80,043,520 | 0.23 | 0.18 | 0.18 | 0.05 | 0.05 | 0.03 | 1.29 [1.22-1.36]* | 1.17 [1.10-1.24]* |
| Z90 | 273,740 | 77,718,280 | 4.22 | 3.37 | 3.45 | 0.85 | 0.77 | 0.42 | 1.22 [1.21-1.24]* | 1.11 [1.09-1.12]* |
| Z91 | 184,150 | 77,895,770 | 3.39 | 2.14 | 2.20 | 1.25 | 1.19 | 0.76 | 1.54 [1.52-1.56]* | 1.29 [1.27-1.31]* |
| Z92 | 646,880 | 74,685,820 | 10.53 | 8.26 | 8.50 | 2.27 | 2.02 | 1.21 | 1.24 [1.23-1.25]* | 1.13 [1.12-1.14]* |
| Z93 | 60,630 | 79,615,330 | 0.90 | 0.73 | 0.74 | 0.17 | 0.16 | 0.12 | 1.22 [1.19-1.26]* | 1.15 [1.12-1.18]* |
| Z94 | 11,670 | 79,976,850 | 0.19 | 0.14 | 0.13 | 0.06 | 0.06 | 0.05 | 1.42 [1.33-1.50]* | 1.32 [1.24-1.40]* |
| Z95 | 210,610 | 77,719,030 | 3.26 | 2.59 | 2.64 | 0.67 | 0.62 | 0.37 | 1.23 [1.22-1.25]* | 1.13 [1.11-1.15]* |
| Z96 | 409,870 | 76,418,260 | 6.46 | 5.13 | 5.31 | 1.33 | 1.15 | 0.79 | 1.22 [1.20-1.23]* | 1.14 [1.13-1.15]* |
| Z97 | 39,340 | 79,832,180 | 0.61 | 0.47 | 0.48 | 0.15 | 0.14 | 0.09 | 1.29 [1.24-1.33]* | 1.16 [1.12-1.21]* |
| Z98 | 53,090 | 79,569,170 | 0.84 | 0.63 | 0.64 | 0.21 | 0.20 | 0.11 | 1.32 [1.28-1.36]* | 1.16 [1.12-1.19]* |
| Z99 | 65,050 | 79,711,970 | 1.10 | 0.76 | 0.77 | 0.34 | 0.32 | 0.21 | 1.42 [1.38-1.45]* | 1.23 [1.20-1.27]* |
| 40+ - A00-B99 | | | | | | | | | | |
| A02 | 940 | 51,040,700 | 0.02 | 0.02 | 0.02 | 0.00 | 0.00 | 0.00 | 1.27 [1.02-1.58] | 1.12 [0.90-1.41] |
| A04 | 45,010 | 50,557,300 | 1.10 | 0.85 | 0.86 | 0.25 | 0.24 | 0.16 | 1.28 [1.24-1.32]* | 1.17 [1.14-1.21]* |
| A08 | 20,340 | 50,890,030 | 0.51 | 0.38 | 0.39 | 0.13 | 0.12 | 0.08 | 1.31 [1.25-1.38]* | 1.17 [1.12-1.23]* |
| A09 | 161,490 | 50,169,410 | 4.07 | 3.04 | 3.14 | 1.04 | 0.93 | 0.54 | 1.30 [1.27-1.32]* | 1.15 [1.13-1.17]* |
| A15 | 880 | 51,045,060 | 0.02 | 0.02 | 0.02 | 0.00 | 0.00 | 0.00 | 1.29 [1.03-1.61] | 1.17 [0.93-1.48] |
| A16 | 1,770 | 51,026,060 | 0.05 | 0.03 | 0.03 | 0.02 | 0.01 | 0.01 | 1.43 [1.23-1.67]* | 1.30 [1.11-1.52]* |
| A18 | 850 | 51,036,800 | 0.02 | 0.02 | 0.02 | 0.01 | 0.01 | 0.00 | 1.35 [1.08-1.68]* | 1.28 [1.02-1.61] |
| A31 | 780 | 51,056,100 | 0.02 | 0.01 | 0.01 | 0.01 | 0.01 | 0.01 | 1.69 [1.36-2.12]* | 1.43 [1.13-1.80]* |
| A40 | 8,830 | 51,006,130 | 0.23 | 0.16 | 0.16 | 0.07 | 0.06 | 0.05 | 1.39 [1.29-1.49]* | 1.29 [1.20-1.39]* |
| A41 | 179,460 | 50,379,250 | 4.38 | 3.39 | 3.50 | 0.99 | 0.88 | 0.63 | 1.25 [1.23-1.27]* | 1.17 [1.15-1.19]* |
| A46 | 600 | 51,045,250 | 0.02 | 0.01 | 0.01 | 0.01 | 0.01 | 0.01 | 2.09 [1.64-2.66]* | 1.97 [1.54-2.53]* |
| A48 | 790 | 51,051,600 | 0.02 | 0.02 | 0.02 | 0.00 | 0.00 | 0.00 | 1.21 [0.95-1.55] | 1.16 [0.91-1.49] |
| A49 | 24,020 | 50,862,880 | 0.61 | 0.44 | 0.45 | 0.16 | 0.15 | 0.11 | 1.34 [1.28-1.40]* | 1.23 [1.18-1.29]* |
| A86 | 990 | 51,051,740 | 0.02 | 0.02 | 0.02 | 0.00 | 0.00 | 0.00 | 1.18 [0.95-1.47] | 1.08 [0.86-1.35] |
| A87 | 900 | 51,018,090 | 0.02 | 0.02 | 0.02 | 0.01 | 0.01 | 0.00 | 1.38 [1.12-1.71]* | 1.19 [0.96-1.49] |
| B00 | 4,330 | 51,004,460 | 0.13 | 0.08 | 0.08 | 0.06 | 0.06 | 0.05 | 1.73 [1.58-1.91]* | 1.54 [1.40-1.70]* |
| B01 | 590 | 51,042,270 | 0.02 | 0.01 | 0.01 | 0.00 | 0.00 | 0.00 | 1.43 [1.10-1.87]* | 1.29 [0.99-1.70] |
| B02 | 12,700 | 50,956,510 | 0.33 | 0.23 | 0.23 | 0.10 | 0.09 | 0.07 | 1.40 [1.32-1.49]* | 1.29 [1.21-1.37]* |
| B07 | 4,200 | 50,950,770 | 0.11 | 0.08 | 0.08 | 0.03 | 0.03 | 0.02 | 1.39 [1.26-1.54]* | 1.28 [1.15-1.42]* |
| B15 | 480 | 51,052,240 | 0.01 | 0.01 | 0.01 | 0.00 | 0.00 | 0.00 | 1.35 [1.00-1.82] | 1.26 [0.93-1.72] |
| B16 | 920 | 51,033,550 | 0.02 | 0.02 | 0.02 | 0.01 | 0.01 | 0.01 | 1.54 [1.25-1.90]* | 1.45 [1.17-1.81]* |
| B17 | 1,480 | 51,024,580 | 0.04 | 0.03 | 0.03 | 0.01 | 0.01 | 0.01 | 1.52 [1.29-1.80]* | 1.35 [1.13-1.60]* |
| B18 | 6,420 | 50,927,850 | 0.16 | 0.12 | 0.12 | 0.04 | 0.04 | 0.02 | 1.30 [1.20-1.41]* | 1.18 [1.08-1.29]* |
| B25 | 1,140 | 51,049,320 | 0.03 | 0.02 | 0.02 | 0.01 | 0.01 | 0.01 | 1.45 [1.19-1.75]* | 1.32 [1.09-1.61]* |
| B34 | 17,310 | 50,733,850 | 0.43 | 0.32 | 0.33 | 0.11 | 0.11 | 0.06 | 1.33 [1.27-1.40]* | 1.16 [1.10-1.22]* |
| B35 | 5,050 | 51,011,710 | 0.15 | 0.09 | 0.09 | 0.07 | 0.06 | 0.05 | 1.71 [1.57-1.87]* | 1.52 [1.39-1.67]* |
| B36 | 1,970 | 51,049,810 | 0.06 | 0.03 | 0.03 | 0.03 | 0.03 | 0.02 | 1.77 [1.53-2.05]* | 1.58 [1.36-1.84]* |
| B37 | 65,140 | 50,617,320 | 1.69 | 1.20 | 1.23 | 0.49 | 0.46 | 0.29 | 1.38 [1.34-1.41]* | 1.21 [1.18-1.24]* |
| B44 | 2,580 | 51,040,820 | 0.08 | 0.04 | 0.04 | 0.04 | 0.04 | 0.03 | 1.90 [1.68-2.14]* | 1.50 [1.32-1.70]* |
| B49 | 1,560 | 51,051,710 | 0.04 | 0.03 | 0.03 | 0.01 | 0.01 | 0.01 | 1.51 [1.29-1.78]* | 1.39 [1.18-1.64]* |
| B59 | 860 | 51,057,220 | 0.02 | 0.02 | 0.02 | 0.01 | 0.01 | 0.01 | 1.39 [1.12-1.73]* | 1.30 [1.04-1.62] |
| B80 | 730 | 51,050,390 | 0.02 | 0.01 | 0.01 | 0.00 | 0.00 | 0.00 | 1.21 [0.95-1.54] | 1.14 [0.88-1.47] |
| B86 | 1,410 | 51,045,110 | 0.06 | 0.02 | 0.02 | 0.03 | 0.03 | 0.03 | 2.65 [2.26-3.12]* | 2.49 [2.11-2.94]* |
| B90 | 700 | 51,054,280 | 0.02 | 0.01 | 0.01 | 0.01 | 0.01 | 0.00 | 1.39 [1.08-1.79]* | 1.25 [0.97-1.62] |
| B91 | 1,180 | 51,043,880 | 0.03 | 0.02 | 0.02 | 0.01 | 0.01 | 0.01 | 1.33 [1.09-1.62]* | 1.29 [1.06-1.59] |
| B94 | 1,290 | 51,048,630 | 0.03 | 0.02 | 0.02 | 0.01 | 0.01 | 0.00 | 1.33 [1.11-1.60]* | 1.17 [0.97-1.41] |
| B95 | 88,090 | 50,078,410 | 2.43 | 1.61 | 1.63 | 0.82 | 0.81 | 0.67 | 1.50 [1.46-1.53]* | 1.38 [1.35-1.41]* |
| B96 | 200,020 | 49,133,560 | 4.99 | 3.87 | 4.02 | 1.12 | 0.98 | 0.64 | 1.24 [1.22-1.26]* | 1.15 [1.13-1.17]* |
| B97 | 34,790 | 50,878,560 | 0.88 | 0.64 | 0.68 | 0.24 | 0.20 | 0.11 | 1.30 [1.25-1.35]* | 1.14 [1.10-1.19]* |
| B98 | 20,340 | 50,916,460 | 0.48 | 0.38 | 0.39 | 0.09 | 0.08 | 0.05 | 1.21 [1.15-1.27]* | 1.12 [1.07-1.18]* |
| B99 | 5,750 | 51,040,400 | 0.14 | 0.11 | 0.11 | 0.03 | 0.02 | 0.02 | 1.21 [1.10-1.33]* | 1.12 [1.02-1.24] |
| 40+ - C00-D49 | | | | | | | | | | |
| C00 | 740 | 51,051,840 | 0.02 | 0.01 | 0.01 | 0.00 | 0.00 | 0.00 | 1.10 [0.85-1.42] | 1.05 [0.81-1.37] |
| C01 | 1,620 | 51,050,510 | 0.03 | 0.03 | 0.03 | 0.00 | 0.00 | 0.00 | 1.07 [0.90-1.27] | 1.04 [0.87-1.24] |
| C02 | 2,450 | 51,036,110 | 0.06 | 0.05 | 0.04 | 0.01 | 0.01 | 0.01 | 1.32 [1.16-1.51]* | 1.27 [1.11-1.46]* |
| C03 | 540 | 51,058,060 | 0.01 | 0.01 | 0.01 | 0.01 | 0.00 | 0.00 | 1.44 [1.09-1.91] | 1.42 [1.07-1.88] |
| C04 | 760 | 51,053,070 | 0.02 | 0.01 | 0.01 | 0.00 | 0.00 | 0.00 | 1.12 [0.87-1.44] | 1.06 [0.82-1.37] |
| C05 | 780 | 51,053,970 | 0.02 | 0.01 | 0.01 | 0.00 | 0.00 | 0.00 | 1.32 [1.04-1.68] | 1.27 [0.99-1.62] |
| C06 | 1,110 | 51,052,070 | 0.03 | 0.02 | 0.02 | 0.01 | 0.00 | 0.00 | 1.21 [0.99-1.48] | 1.13 [0.92-1.39] |
| C07 | 1,070 | 51,050,250 | 0.02 | 0.02 | 0.02 | 0.00 | 0.00 | 0.00 | 1.01 [0.81-1.25] | 0.97 [0.78-1.21] |
| C09 | 2,050 | 51,041,720 | 0.04 | 0.04 | 0.04 | 0.00 | 0.01 | 0.00 | 1.15 [0.99-1.33] | 1.09 [0.93-1.27] |
| C10 | 1,040 | 51,056,350 | 0.02 | 0.02 | 0.02 | 0.00 | 0.00 | 0.00 | 1.16 [0.94-1.44] | 1.09 [0.87-1.35] |
| C11 | 510 | 51,057,520 | 0.01 | 0.01 | 0.01 | 0.00 | 0.00 | 0.00 | 1.36 [1.01-1.83] | 1.31 [0.97-1.77] |
| C12 | 560 | 51,058,430 | 0.01 | 0.01 | 0.01 | 0.00 | 0.00 | 0.00 | 1.35 [1.02-1.78] | 1.31 [0.99-1.74] |
| C13 | 640 | 51,059,530 | 0.01 | 0.01 | 0.01 | 0.00 | 0.00 | 0.00 | 1.22 [0.93-1.60] | 1.20 [0.91-1.58] |
| C14 | 1,170 | 51,055,760 | 0.03 | 0.02 | 0.02 | 0.00 | 0.00 | 0.00 | 1.06 [0.87-1.30] | 1.00 [0.81-1.23] |
| C15 | 16,270 | 51,004,350 | 0.36 | 0.31 | 0.31 | 0.05 | 0.05 | 0.04 | 1.17 [1.10-1.23]* | 1.13 [1.07-1.19]* |
| C16 | 13,070 | 51,007,200 | 0.28 | 0.25 | 0.25 | 0.02 | 0.02 | 0.02 | 1.09 [1.02-1.16]* | 1.06 [0.99-1.13] |
| C17 | 2,640 | 51,047,120 | 0.06 | 0.05 | 0.05 | 0.01 | 0.01 | 0.01 | 1.13 [0.99-1.29] | 1.10 [0.96-1.26] |
| C18 | 45,170 | 50,672,280 | 0.91 | 0.89 | 0.90 | 0.02 | 0.01 | 0.00 | 1.01 [0.98-1.05] | 1.00 [0.96-1.03] |
| C19 | 9,190 | 50,989,590 | 0.18 | 0.18 | 0.18 | 0.00 | −0.01 | −0.01 | 0.96 [0.89-1.04] | 0.96 [0.89-1.04] |
| C20 | 19,410 | 50,861,270 | 0.37 | 0.38 | 0.38 | −0.01 | −0.01 | −0.01 | 0.97 [0.92-1.02] | 0.97 [0.92-1.03] |
| C21 | 2,400 | 51,037,490 | 0.05 | 0.05 | 0.05 | 0.00 | 0.00 | 0.00 | 1.09 [0.94-1.25] | 1.05 [0.91-1.21] |

| | | | | | | | | | | |
|---|---|---|---|---|---|---|---|---|---|---|
| C22 | 8,940 | 51,041,580 | 0.21 | 0.17 | 0.17 | 0.04 | 0.04 | 0.03 | 1.23 [1.14-1.32]* | 1.17 [1.08-1.26]* |
| C23 | 1,310 | 51,059,660 | 0.03 | 0.03 | 0.03 | 0.00 | 0.00 | 0.00 | 1.06 [0.87-1.29] | 1.05 [0.86-1.28] |
| C24 | 2,470 | 51,053,600 | 0.05 | 0.05 | 0.05 | 0.00 | 0.00 | 0.00 | 1.07 [0.92-1.23] | 1.02 [0.88-1.18] |
| C25 | 15,060 | 51,035,790 | 0.33 | 0.29 | 0.29 | 0.05 | 0.04 | 0.03 | 1.15 [1.09-1.22]* | 1.11 [1.05-1.18]* |
| C26 | 3,300 | 51,047,880 | 0.07 | 0.06 | 0.07 | 0.01 | 0.01 | 0.01 | 1.12 [0.99-1.26] | 1.08 [0.95-1.22] |
| C30 | 530 | 51,056,660 | 0.01 | 0.01 | 0.01 | 0.00 | 0.00 | 0.00 | 1.16 [0.86-1.56] | 1.12 [0.83-1.52] |
| C32 | 3,470 | 51,023,110 | 0.08 | 0.07 | 0.07 | 0.01 | 0.01 | 0.01 | 1.16 [1.03-1.30] | 1.12 [0.99-1.26] |
| C34 | 65,170 | 50,883,280 | 1.45 | 1.24 | 1.26 | 0.21 | 0.19 | 0.12 | 1.15 [1.12-1.18]* | 1.09 [1.06-1.12]* |
| C38 | 830 | 51,059,370 | 0.02 | 0.02 | 0.02 | 0.00 | 0.00 | 0.00 | 1.00 [0.79-1.27] | 0.99 [0.77-1.26] |
| C41 | 1,520 | 51,050,330 | 0.04 | 0.03 | 0.03 | 0.01 | 0.01 | 0.01 | 1.21 [1.02-1.44] | 1.17 [0.98-1.39] |
| C43 | 16,870 | 50,858,180 | 0.32 | 0.33 | 0.34 | −0.01 | −0.02 | −0.02 | 0.96 [0.90-1.01] | 0.94 [0.89-0.99] |
| C44 | 126,920 | 49,518,040 | 2.82 | 2.51 | 2.56 | 0.32 | 0.26 | 0.17 | 1.10 [1.08-1.13]* | 1.06 [1.04-1.08]* |
| C45 | 4,090 | 51,054,030 | 0.08 | 0.08 | 0.08 | 0.00 | 0.00 | 0.00 | 1.01 [0.90-1.13] | 1.02 [0.91-1.14] |
| C48 | 2,220 | 51,052,900 | 0.05 | 0.04 | 0.04 | 0.00 | 0.00 | 0.00 | 1.06 [0.91-1.23] | 1.04 [0.90-1.21] |
| C49 | 3,550 | 51,028,900 | 0.08 | 0.07 | 0.07 | 0.01 | 0.01 | 0.01 | 1.13 [1.00-1.27] | 1.09 [0.96-1.23] |
| C50 | 74,410 | 49,849,330 | 1.56 | 1.48 | 1.51 | 0.08 | 0.05 | 0.03 | 1.03 [1.01-1.06] | 1.02 [0.99-1.05] |
| C51 | 2,010 | 51,039,090 | 0.05 | 0.04 | 0.04 | 0.01 | 0.01 | 0.01 | 1.27 [1.09-1.47]* | 1.22 [1.05-1.42]* |
| C52 | 670 | 51,057,980 | 0.01 | 0.01 | 0.01 | 0.00 | 0.00 | 0.00 | 0.92 [0.70-1.22] | 0.93 [0.70-1.23] |
| C53 | 3,050 | 51,006,030 | 0.06 | 0.06 | 0.06 | 0.00 | 0.00 | 0.00 | 0.96 [0.84-1.09] | 0.98 [0.86-1.12] |
| C54 | 12,680 | 50,899,930 | 0.26 | 0.25 | 0.25 | 0.01 | 0.01 | 0.01 | 1.03 [0.97-1.09] | 1.05 [0.98-1.11] |
| C55 | 1,740 | 51,048,320 | 0.04 | 0.03 | 0.03 | 0.00 | 0.00 | 0.00 | 1.01 [0.85-1.19] | 1.01 [0.85-1.20] |
| C56 | 10,950 | 50,968,380 | 0.22 | 0.21 | 0.22 | 0.01 | 0.00 | 0.01 | 1.02 [0.96-1.09] | 1.02 [0.96-1.10] |
| C57 | 1,260 | 51,055,660 | 0.03 | 0.02 | 0.02 | 0.01 | 0.00 | 0.01 | 1.19 [0.99-1.44] | 1.21 [1.00-1.47] |
| C60 | 940 | 51,050,040 | 0.02 | 0.02 | 0.02 | 0.00 | 0.00 | 0.00 | 1.21 [0.97-1.51] | 1.24 [0.99-1.56] |
| C61 | 73,070 | 50,383,270 | 1.54 | 1.43 | 1.47 | 0.11 | 0.08 | 0.04 | 1.05 [1.03-1.08]* | 1.02 [1.00-1.05] |
| C62 | 860 | 51,027,880 | 0.02 | 0.02 | 0.02 | 0.00 | 0.00 | 0.00 | 0.99 [0.78-1.26] | 0.95 [0.74-1.22] |
| C64 | 14,700 | 50,946,120 | 0.32 | 0.28 | 0.28 | 0.03 | 0.03 | 0.02 | 1.11 [1.05-1.18]* | 1.06 [1.00-1.12] |
| C65 | 1,340 | 51,051,280 | 0.03 | 0.03 | 0.03 | 0.01 | 0.01 | 0.01 | 1.22 [1.01-1.48] | 1.21 [1.00-1.47] |
| C66 | 1,910 | 51,049,020 | 0.04 | 0.04 | 0.04 | 0.00 | 0.00 | 0.00 | 1.08 [0.92-1.28] | 1.04 [0.88-1.23] |
| C67 | 33,560 | 50,662,650 | 0.71 | 0.65 | 0.66 | 0.06 | 0.06 | 0.04 | 1.08 [1.04-1.13]* | 1.06 [1.02-1.10]* |
| C68 | 730 | 51,056,610 | 0.01 | 0.01 | 0.01 | 0.00 | 0.00 | 0.00 | 1.11 [0.85-1.44] | 1.10 [0.84-1.43] |
| C69 | 1,220 | 51,043,890 | 0.02 | 0.02 | 0.02 | 0.00 | 0.00 | 0.00 | 1.03 [0.84-1.26] | 1.00 [0.81-1.23] |
| C71 | 6,850 | 51,029,800 | 0.15 | 0.13 | 0.13 | 0.02 | 0.02 | 0.01 | 1.13 [1.04-1.23]* | 1.10 [1.01-1.20] |
| C73 | 3,350 | 51,007,370 | 0.07 | 0.07 | 0.06 | 0.00 | 0.00 | 0.00 | 1.06 [0.94-1.20] | 1.03 [0.92-1.17] |
| C75 | 620 | 51,056,320 | 0.01 | 0.01 | 0.01 | 0.00 | 0.00 | 0.00 | 1.07 [0.81-1.41] | 1.00 [0.75-1.33] |
| C76 | 2,190 | 51,047,490 | 0.05 | 0.04 | 0.04 | 0.00 | 0.00 | 0.00 | 1.07 [0.92-1.25] | 1.06 [0.91-1.24] |
| C77 | 73,140 | 50,532,770 | 1.52 | 1.43 | 1.45 | 0.09 | 0.08 | 0.06 | 1.05 [1.03-1.08]* | 1.04 [1.01-1.07]* |
| C78 | 109,940 | 50,850,120 | 2.29 | 2.13 | 2.16 | 0.16 | 0.13 | 0.10 | 1.06 [1.04-1.08]* | 1.04 [1.02-1.07]* |
| C79 | 92,510 | 50,875,120 | 1.95 | 1.79 | 1.81 | 0.16 | 0.14 | 0.11 | 1.08 [1.05-1.10]* | 1.06 [1.03-1.08]* |
| C80 | 32,730 | 50,980,460 | 0.70 | 0.63 | 0.63 | 0.07 | 0.06 | 0.05 | 1.10 [1.06-1.14]* | 1.07 [1.03-1.12]* |
| C81 | 2,300 | 51,021,060 | 0.07 | 0.04 | 0.04 | 0.02 | 0.03 | 0.02 | 1.62 [1.42-1.85]* | 1.52 [1.33-1.74]* |
| C82 | 3,860 | 51,014,590 | 0.08 | 0.07 | 0.08 | 0.01 | 0.01 | 0.00 | 1.10 [0.98-1.23] | 1.05 [0.94-1.17] |
| C83 | 10,750 | 50,979,900 | 0.25 | 0.20 | 0.20 | 0.05 | 0.05 | 0.04 | 1.23 [1.15-1.31]* | 1.19 [1.11-1.27]* |
| C84 | 1,410 | 51,049,430 | 0.05 | 0.02 | 0.02 | 0.03 | 0.03 | 0.03 | 2.56 [2.19-2.99]* | 2.48 [2.11-2.90]* |
| C85 | 13,510 | 50,911,760 | 0.33 | 0.25 | 0.25 | 0.07 | 0.08 | 0.06 | 1.30 [1.23-1.38]* | 1.23 [1.16-1.31]* |
| C88 | 1,850 | 51,049,540 | 0.04 | 0.03 | 0.03 | 0.01 | 0.01 | 0.01 | 1.28 [1.10-1.51]* | 1.24 [1.06-1.46]* |
| C90 | 10,000 | 50,997,600 | 0.22 | 0.19 | 0.19 | 0.03 | 0.03 | 0.02 | 1.14 [1.06-1.22]* | 1.10 [1.03-1.18]* |
| C91 | 9,230 | 50,981,940 | 0.20 | 0.18 | 0.18 | 0.02 | 0.02 | 0.01 | 1.09 [1.02-1.18] | 1.05 [0.98-1.13] |
| C92 | 7,310 | 51,023,350 | 0.18 | 0.14 | 0.14 | 0.04 | 0.04 | 0.04 | 1.28 [1.19-1.39]* | 1.25 [1.15-1.35]* |
| C93 | 1,020 | 51,059,740 | 0.02 | 0.02 | 0.02 | 0.00 | 0.00 | 0.00 | 1.10 [0.88-1.37] | 1.07 [0.85-1.34] |
| C94 | 480 | 51,060,480 | 0.01 | 0.01 | 0.01 | 0.00 | 0.00 | 0.00 | 1.23 [0.90-1.68] | 1.17 [0.85-1.61] |
| C95 | 1,490 | 51,054,140 | 0.04 | 0.03 | 0.03 | 0.01 | 0.01 | 0.01 | 1.32 [1.11-1.57]* | 1.24 [1.03-1.48] |
| C97 | 8,500 | 51,036,540 | 0.18 | 0.16 | 0.17 | 0.01 | 0.01 | 0.00 | 1.05 [0.97-1.14] | 1.02 [0.94-1.10] |
| D00 | 940 | 51,053,050 | 0.02 | 0.02 | 0.02 | 0.00 | 0.00 | 0.00 | 1.07 [0.85-1.34] | 0.98 [0.77-1.23] |
| D01 | 900 | 51,051,330 | 0.02 | 0.02 | 0.02 | 0.00 | 0.00 | 0.00 | 1.13 [0.89-1.42] | 1.04 [0.82-1.31] |
| D02 | 960 | 51,052,740 | 0.02 | 0.02 | 0.02 | 0.00 | 0.00 | 0.00 | 1.22 [0.98-1.52] | 1.14 [0.92-1.43] |
| D03 | 6,530 | 50,979,550 | 0.14 | 0.13 | 0.13 | 0.01 | 0.01 | 0.01 | 1.08 [0.99-1.18] | 1.04 [0.95-1.14] |
| D04 | 14,210 | 50,921,750 | 0.34 | 0.27 | 0.27 | 0.08 | 0.07 | 0.05 | 1.26 [1.19-1.34]* | 1.17 [1.10-1.24]* |
| D05 | 12,310 | 50,833,500 | 0.24 | 0.24 | 0.24 | 0.00 | 0.00 | 0.00 | 1.00 [0.94-1.06] | 0.98 [0.92-1.05] |
| D06 | 2,480 | 50,823,020 | 0.05 | 0.05 | 0.05 | 0.00 | 0.00 | 0.00 | 1.03 [0.90-1.18] | 0.99 [0.86-1.14] |
| D07 | 3,620 | 50,998,440 | 0.08 | 0.07 | 0.07 | 0.01 | 0.01 | 0.01 | 1.22 [1.09-1.37]* | 1.16 [1.03-1.30] |
| D09 | 3,540 | 51,025,720 | 0.08 | 0.07 | 0.07 | 0.01 | 0.01 | 0.01 | 1.13 [1.01-1.27] | 1.10 [0.97-1.23] |
| D10 | 2,590 | 50,994,710 | 0.06 | 0.05 | 0.05 | 0.02 | 0.02 | 0.01 | 1.35 [1.19-1.54]* | 1.21 [1.06-1.38]* |
| D11 | 3,770 | 50,985,820 | 0.08 | 0.07 | 0.07 | 0.00 | 0.00 | 0.00 | 1.04 [0.93-1.16] | 0.99 [0.88-1.11] |
| D12 | 120,430 | 49,508,060 | 2.85 | 2.34 | 2.36 | 0.51 | 0.49 | 0.27 | 1.21 [1.18-1.23]* | 1.11 [1.08-1.13]* |
| D13 | 7,840 | 50,856,160 | 0.18 | 0.15 | 0.15 | 0.03 | 0.03 | 0.01 | 1.21 [1.12-1.31]* | 1.09 [1.01-1.18] |
| D14 | 1,990 | 51,017,110 | 0.05 | 0.04 | 0.04 | 0.01 | 0.01 | 0.01 | 1.32 [1.14-1.53]* | 1.22 [1.05-1.42]* |
| D15 | 920 | 51,052,090 | 0.02 | 0.02 | 0.02 | 0.00 | 0.00 | 0.00 | 1.16 [0.93-1.45] | 1.12 [0.89-1.40] |
| D16 | 1,880 | 51,017,370 | 0.04 | 0.04 | 0.04 | 0.01 | 0.01 | 0.00 | 1.20 [1.03-1.40] | 1.09 [0.93-1.28] |
| D17 | 31,530 | 50,120,670 | 0.69 | 0.62 | 0.61 | 0.08 | 0.08 | 0.04 | 1.13 [1.09-1.18]* | 1.06 [1.02-1.10]* |
| D18 | 12,590 | 50,857,730 | 0.29 | 0.24 | 0.24 | 0.05 | 0.05 | 0.02 | 1.19 [1.12-1.26]* | 1.09 [1.02-1.16]* |
| D21 | 3,320 | 50,985,550 | 0.08 | 0.06 | 0.06 | 0.02 | 0.02 | 0.01 | 1.25 [1.12-1.40]* | 1.17 [1.04-1.31] |
| D22 | 19,170 | 50,412,570 | 0.41 | 0.37 | 0.37 | 0.04 | 0.04 | 0.02 | 1.11 [1.06-1.17]* | 1.05 [0.99-1.10] |
| D23 | 16,910 | 50,487,720 | 0.40 | 0.32 | 0.32 | 0.07 | 0.08 | 0.06 | 1.26 [1.20-1.32]* | 1.17 [1.11-1.24]* |
| D24 | 7,130 | 50,817,110 | 0.15 | 0.14 | 0.14 | 0.01 | 0.00 | 0.00 | 1.02 [0.94-1.11] | 0.98 [0.90-1.07] |
| D25 | 53,810 | 49,537,950 | 1.18 | 1.07 | 1.09 | 0.11 | 0.09 | 0.06 | 1.09 [1.06-1.12]* | 1.05 [1.02-1.08]* |
| D26 | 1,780 | 51,006,230 | 0.04 | 0.03 | 0.03 | 0.00 | 0.00 | 0.00 | 1.08 [0.92-1.27] | 1.04 [0.88-1.23] |
| D27 | 10,290 | 50,823,440 | 0.21 | 0.20 | 0.20 | 0.01 | 0.01 | 0.00 | 1.04 [0.97-1.11] | 0.99 [0.93-1.07] |
| D28 | 1,090 | 51,027,240 | 0.02 | 0.02 | 0.02 | 0.00 | 0.00 | 0.00 | 1.14 [0.93-1.40] | 1.11 [0.90-1.37] |
| D29 | 1,040 | 51,043,870 | 0.02 | 0.02 | 0.02 | 0.00 | 0.00 | 0.00 | 1.14 [0.92-1.42] | 1.10 [0.89-1.38] |
| D30 | 2,220 | 51,032,530 | 0.05 | 0.04 | 0.04 | 0.00 | 0.00 | 0.00 | 1.08 [0.94-1.26] | 1.03 [0.89-1.20] |
| D31 | 1,850 | 51,037,510 | 0.04 | 0.03 | 0.03 | 0.01 | 0.01 | 0.01 | 1.26 [1.07-1.47]* | 1.20 [1.02-1.41] |
| D32 | 8,190 | 50,985,200 | 0.18 | 0.16 | 0.16 | 0.02 | 0.02 | 0.01 | 1.11 [1.03-1.21]* | 1.05 [0.97-1.14] |
| D33 | 2,780 | 51,021,540 | 0.06 | 0.05 | 0.05 | 0.01 | 0.01 | 0.00 | 1.11 [0.97-1.27] | 1.02 [0.89-1.17] |
| D34 | 1,810 | 51,023,250 | 0.04 | 0.04 | 0.04 | 0.00 | 0.00 | 0.00 | 1.04 [0.89-1.23] | 1.00 [0.85-1.19] |
| D35 | 11,130 | 50,923,410 | 0.25 | 0.21 | 0.22 | 0.03 | 0.03 | 0.01 | 1.13 [1.05-1.20]* | 1.04 [0.98-1.12] |
| D36 | 4,300 | 50,955,120 | 0.10 | 0.08 | 0.08 | 0.02 | 0.02 | 0.01 | 1.24 [1.12-1.37]* | 1.13 [1.02-1.26] |
| D37 | 16,360 | 50,898,110 | 0.35 | 0.32 | 0.32 | 0.04 | 0.03 | 0.02 | 1.11 [1.05-1.17]* | 1.06 [1.00-1.12] |
| D38 | 3,860 | 51,034,270 | 0.09 | 0.07 | 0.07 | 0.02 | 0.01 | 0.01 | 1.20 [1.08-1.34]* | 1.14 [1.02-1.27] |
| D39 | 1,880 | 51,026,350 | 0.04 | 0.04 | 0.04 | 0.00 | 0.00 | 0.00 | 0.97 [0.83-1.14] | 0.94 [0.79-1.11] |
| D40 | 620 | 51,049,490 | 0.01 | 0.01 | 0.01 | 0.00 | 0.00 | 0.00 | 1.21 [0.92-1.59] | 1.17 [0.88-1.54] |
| D41 | 11,660 | 50,929,190 | 0.25 | 0.22 | 0.23 | 0.02 | 0.02 | 0.02 | 1.10 [1.03-1.18]* | 1.06 [1.00-1.14] |
| D43 | 3,220 | 51,039,400 | 0.07 | 0.06 | 0.06 | 0.01 | 0.00 | 0.00 | 1.06 [0.94-1.20] | 1.01 [0.89-1.14] |
| D44 | 1,940 | 51,031,000 | 0.04 | 0.04 | 0.03 | 0.01 | 0.01 | 0.01 | 1.26 [1.08-1.47]* | 1.16 [0.99-1.35] |
| D45 | 4,180 | 50,994,040 | 0.09 | 0.08 | 0.08 | 0.01 | 0.01 | 0.00 | 1.06 [0.96-1.19] | 1.00 [0.89-1.11] |
| D46 | 10,030 | 51,012,800 | 0.25 | 0.19 | 0.19 | 0.07 | 0.06 | 0.05 | 1.33 [1.24-1.42]* | 1.26 [1.18-1.35]* |
| D47 | 15,930 | 50,969,740 | 0.39 | 0.30 | 0.31 | 0.09 | 0.08 | 0.05 | 1.25 [1.19-1.32]* | 1.16 [1.10-1.23]* |
| D48 | 6,730 | 50,937,780 | 0.15 | 0.13 | 0.13 | 0.02 | 0.01 | 0.01 | 1.11 [1.02-1.20] | 1.05 [0.96-1.15] |
| 40+ - D50-D89 | | | | | | | | | | |
| D50 | 202,630 | 49,229,630 | 5.26 | 3.87 | 3.98 | 1.39 | 1.28 | 0.88 | 1.32 [1.30-1.34]* | 1.20 [1.18-1.22]* |
| D51 | 18,880 | 50,867,700 | 0.48 | 0.35 | 0.35 | 0.13 | 0.13 | 0.09 | 1.36 [1.30-1.43]* | 1.24 [1.18-1.31]* |
| D52 | 14,730 | 50,991,170 | 0.36 | 0.27 | 0.28 | 0.09 | 0.08 | 0.06 | 1.30 [1.23-1.38]* | 1.20 [1.14-1.28]* |
| D53 | 3,800 | 51,037,310 | 0.10 | 0.07 | 0.07 | 0.04 | 0.04 | 0.03 | 1.53 [1.37-1.71]* | 1.37 [1.23-1.54]* |
| D56 | 4,380 | 50,977,920 | 0.12 | 0.08 | 0.08 | 0.04 | 0.04 | 0.04 | 1.54 [1.40-1.70]* | 1.42 [1.28-1.57]* |
| D57 | 3,780 | 50,962,930 | 0.10 | 0.07 | 0.07 | 0.03 | 0.02 | 0.02 | 1.30 [1.17-1.44]* | 1.23 [1.10-1.37]* |
| D58 | 1,550 | 51,043,150 | 0.04 | 0.03 | 0.03 | 0.01 | 0.01 | 0.00 | 1.18 [0.99-1.40] | 1.12 [0.94-1.34] |
| D59 | 2,340 | 51,038,000 | 0.06 | 0.04 | 0.04 | 0.02 | 0.02 | 0.02 | 1.43 [1.25-1.64]* | 1.38 [1.20-1.58]* |

| | | | | | | | | | | |
|---|---|---|---|---|---|---|---|---|---|---|
| D61 | 12,880 | 51,005,100 | 0.32 | 0.24 | 0.24 | 0.08 | 0.07 | 0.06 | 1.30 [1.22-1.38]* | 1.22 [1.15-1.30]* |
| D62 | 3,370 | 50,981,740 | 0.08 | 0.06 | 0.06 | 0.01 | 0.01 | 0.01 | 1.18 [1.05-1.33]* | 1.12 [0.99-1.26] |
| D63 | 21,270 | 50,943,080 | 0.51 | 0.40 | 0.40 | 0.11 | 0.11 | 0.08 | 1.27 [1.21-1.33]* | 1.18 [1.13-1.24]* |
| D64 | 277,310 | 48,504,870 | 7.21 | 5.40 | 5.54 | 1.82 | 1.67 | 1.24 | 1.30 [1.29-1.32]* | 1.21 [1.19-1.22]* |
| D65 | 840 | 51,058,690 | 0.02 | 0.02 | 0.02 | 0.00 | 0.00 | 0.00 | 1.21 [0.95-1.53] | 1.17 [0.92-1.49] |
| D66 | 610 | 51,049,570 | 0.01 | 0.01 | 0.01 | 0.00 | 0.00 | 0.00 | 1.18 [0.90-1.57] | 1.09 [0.82-1.45] |
| D68 | 11,730 | 50,884,800 | 0.29 | 0.22 | 0.22 | 0.07 | 0.07 | 0.04 | 1.34 [1.26-1.42]* | 1.18 [1.11-1.26]* |
| D69 | 37,580 | 50,794,980 | 0.92 | 0.70 | 0.71 | 0.21 | 0.20 | 0.15 | 1.28 [1.24-1.33]* | 1.20 [1.16-1.25]* |
| D70 | 37,520 | 50,799,270 | 0.88 | 0.71 | 0.71 | 0.17 | 0.17 | 0.14 | 1.23 [1.19-1.28]* | 1.19 [1.15-1.23]* |
| D72 | 7,890 | 51,006,700 | 0.21 | 0.14 | 0.15 | 0.06 | 0.06 | 0.04 | 1.40 [1.30-1.51]* | 1.25 [1.16-1.35]* |
| D73 | 8,890 | 50,984,730 | 0.21 | 0.17 | 0.17 | 0.05 | 0.04 | 0.03 | 1.26 [1.17-1.35]* | 1.16 [1.08-1.25]* |
| D75 | 11,670 | 50,945,750 | 0.27 | 0.22 | 0.23 | 0.05 | 0.05 | 0.03 | 1.21 [1.14-1.29]* | 1.12 [1.05-1.19]* |
| D76 | 490 | 51,051,290 | 0.01 | 0.01 | 0.01 | 0.00 | 0.00 | 0.00 | 1.38 [1.03-1.85] | 1.27 [0.94-1.72] |
| D80 | 2,020 | 51,039,960 | 0.06 | 0.04 | 0.04 | 0.02 | 0.02 | 0.01 | 1.46 [1.27-1.69]* | 1.21 [1.04-1.41] |
| D84 | 850 | 51,052,290 | 0.03 | 0.01 | 0.01 | 0.01 | 0.01 | 0.01 | 1.75 [1.42-2.16]* | 1.38 [1.10-1.72]* |
| D86 | 5,630 | 50,971,560 | 0.15 | 0.10 | 0.10 | 0.05 | 0.05 | 0.04 | 1.51 [1.39-1.64]* | 1.35 [1.23-1.47]* |
| D89 | 4,650 | 51,019,410 | 0.12 | 0.08 | 0.09 | 0.04 | 0.04 | 0.03 | 1.41 [1.28-1.55]* | 1.28 [1.16-1.41]* |
| 40+ - E00-E89 | | | | | | | | | | |
| E02 | 1,720 | 51,054,180 | 0.04 | 0.03 | 0.03 | 0.01 | 0.00 | 0.00 | 1.11 [0.93-1.32] | 1.07 [0.89-1.28] |
| E03 | 198,660 | 48,514,780 | 5.22 | 3.85 | 3.92 | 1.37 | 1.31 | 1.01 | 1.33 [1.31-1.35]* | 1.24 [1.22-1.26]* |
| E04 | 20,340 | 50,767,650 | 0.45 | 0.39 | 0.40 | 0.06 | 0.05 | 0.02 | 1.12 [1.07-1.17]* | 1.04 [0.99-1.09] |
| E05 | 31,650 | 50,660,640 | 0.78 | 0.59 | 0.61 | 0.18 | 0.17 | 0.11 | 1.27 [1.22-1.32]* | 1.17 [1.12-1.21]* |
| E06 | 3,000 | 51,014,260 | 0.07 | 0.06 | 0.06 | 0.01 | 0.01 | 0.01 | 1.24 [1.10-1.40]* | 1.16 [1.02-1.31] |
| E07 | 5,570 | 50,994,750 | 0.13 | 0.10 | 0.11 | 0.03 | 0.03 | 0.02 | 1.27 [1.16-1.39]* | 1.15 [1.05-1.26]* |
| E10 | 33,960 | 50,276,570 | 0.84 | 0.64 | 0.64 | 0.20 | 0.20 | 0.10 | 1.32 [1.27-1.36]* | 1.13 [1.09-1.17]* |
| E11 | 333,460 | 46,606,620 | 8.69 | 6.82 | 6.97 | 1.87 | 1.72 | 0.87 | 1.25 [1.23-1.26]* | 1.11 [1.10-1.12]* |
| E13 | 3,810 | 51,032,600 | 0.10 | 0.07 | 0.07 | 0.03 | 0.03 | 0.02 | 1.43 [1.29-1.59]* | 1.21 [1.09-1.35]* |
| E14 | 45,140 | 50,502,840 | 1.17 | 0.84 | 0.85 | 0.33 | 0.31 | 0.17 | 1.37 [1.33-1.41]* | 1.17 [1.13-1.21]* |
| E16 | 48,180 | 50,807,870 | 1.17 | 0.90 | 0.92 | 0.27 | 0.25 | 0.11 | 1.27 [1.23-1.31]* | 1.10 [1.07-1.14]* |
| E20 | 1,260 | 51,050,580 | 0.03 | 0.02 | 0.02 | 0.01 | 0.01 | 0.01 | 1.53 [1.27-1.84]* | 1.33 [1.10-1.62]* |
| E21 | 16,620 | 50,931,760 | 0.38 | 0.32 | 0.33 | 0.06 | 0.05 | 0.03 | 1.16 [1.10-1.22]* | 1.07 [1.02-1.13] |
| E22 | 10,340 | 50,984,450 | 0.29 | 0.18 | 0.19 | 0.10 | 0.10 | 0.08 | 1.51 [1.41-1.61]* | 1.41 [1.32-1.50]* |
| E23 | 3,970 | 51,001,610 | 0.11 | 0.07 | 0.07 | 0.04 | 0.03 | 0.03 | 1.49 [1.34-1.65]* | 1.33 [1.20-1.48]* |
| E24 | 1,030 | 51,044,870 | 0.03 | 0.02 | 0.02 | 0.01 | 0.01 | 0.01 | 1.61 [1.33-1.96]* | 1.24 [1.01-1.52] |
| E26 | 640 | 51,051,290 | 0.01 | 0.01 | 0.01 | 0.00 | 0.00 | 0.00 | 1.14 [0.87-1.50] | 0.97 [0.73-1.29] |
| E27 | 10,280 | 50,977,920 | 0.27 | 0.19 | 0.19 | 0.09 | 0.08 | 0.05 | 1.45 [1.36-1.54]* | 1.25 [1.17-1.34]* |
| E28 | 4,540 | 50,886,220 | 0.11 | 0.08 | 0.09 | 0.03 | 0.02 | 0.00 | 1.26 [1.15-1.39]* | 1.05 [0.95-1.16] |
| E29 | 1,450 | 51,043,050 | 0.04 | 0.03 | 0.02 | 0.02 | 0.02 | 0.01 | 1.76 [1.50-2.08]* | 1.40 [1.18-1.66]* |
| E34 | 1,080 | 51,044,630 | 0.03 | 0.02 | 0.02 | 0.01 | 0.01 | 0.00 | 1.39 [1.14-1.70]* | 1.23 [1.00-1.50] |
| E41 | 610 | 51,060,640 | 0.01 | 0.01 | 0.01 | 0.00 | 0.00 | 0.00 | 1.10 [0.82-1.48] | 1.09 [0.81-1.48] |
| E43 | 650 | 51,059,840 | 0.01 | 0.01 | 0.01 | 0.00 | 0.00 | 0.00 | 1.24 [0.94-1.63] | 1.20 [0.91-1.60] |
| E46 | 8,690 | 51,026,740 | 0.20 | 0.16 | 0.17 | 0.03 | 0.03 | 0.02 | 1.16 [1.07-1.25]* | 1.09 [1.01-1.18] |
| E51 | 860 | 51,053,310 | 0.02 | 0.02 | 0.02 | 0.01 | 0.01 | 0.00 | 1.48 [1.19-1.84]* | 1.28 [1.02-1.61] |
| E53 | 48,410 | 50,810,220 | 1.20 | 0.90 | 0.93 | 0.30 | 0.26 | 0.17 | 1.28 [1.24-1.33]* | 1.17 [1.13-1.21]* |
| E55 | 61,600 | 50,809,510 | 1.59 | 1.13 | 1.17 | 0.46 | 0.42 | 0.27 | 1.35 [1.32-1.39]* | 1.20 [1.17-1.24]* |
| E56 | 620 | 51,058,320 | 0.01 | 0.01 | 0.01 | 0.00 | 0.00 | 0.00 | 1.21 [0.92-1.60] | 1.10 [0.83-1.46] |
| E61 | 8,900 | 51,010,460 | 0.23 | 0.16 | 0.17 | 0.07 | 0.07 | 0.04 | 1.39 [1.29-1.49]* | 1.22 [1.13-1.31]* |
| E63 | 570 | 51,059,030 | 0.01 | 0.01 | 0.01 | 0.00 | 0.00 | 0.00 | 1.46 [1.11-1.94]* | 1.32 [0.99-1.77] |
| E66 | 271,010 | 48,354,460 | 7.09 | 5.29 | 5.35 | 1.80 | 1.74 | 0.91 | 1.32 [1.31-1.34]* | 1.15 [1.13-1.16]* |
| E72 | 420 | 51,056,390 | 0.01 | 0.01 | 0.01 | 0.00 | 0.00 | 0.00 | 1.52 [1.12-2.08]* | 1.39 [1.01-1.90] |
| E73 | 1,540 | 51,048,610 | 0.04 | 0.03 | 0.03 | 0.01 | 0.01 | 0.00 | 1.35 [1.14-1.59]* | 1.10 [0.92-1.30] |
| E74 | 920 | 51,050,320 | 0.02 | 0.02 | 0.02 | 0.01 | 0.01 | 0.00 | 1.37 [1.10-1.70]* | 1.25 [1.01-1.56] |
| E78 | 441,840 | 45,348,790 | 11.83 | 9.29 | 9.62 | 2.54 | 2.21 | 1.17 | 1.23 [1.22-1.24]* | 1.11 [1.10-1.12]* |
| E79 | 500 | 51,057,690 | 0.01 | 0.01 | 0.01 | 0.00 | 0.00 | 0.00 | 1.43 [1.07-1.92] | 1.25 [0.92-1.69] |
| E80 | 4,020 | 51,017,990 | 0.09 | 0.08 | 0.08 | 0.02 | 0.02 | 0.01 | 1.21 [1.09-1.35]* | 1.13 [1.01-1.26] |
| E83 | 76,890 | 50,728,850 | 1.81 | 1.45 | 1.51 | 0.36 | 0.30 | 0.17 | 1.20 [1.17-1.23]* | 1.10 [1.08-1.13]* |
| E85 | 2,890 | 51,047,110 | 0.07 | 0.05 | 0.06 | 0.01 | 0.01 | 0.01 | 1.18 [1.04-1.35] | 1.10 [0.96-1.25] |
| E86 | 194,120 | 50,308,450 | 4.63 | 3.69 | 3.90 | 0.93 | 0.73 | 0.48 | 1.19 [1.17-1.21]* | 1.12 [1.10-1.14]* |
| E87 | 311,620 | 49,733,080 | 7.89 | 5.92 | 6.15 | 1.98 | 1.74 | 1.21 | 1.28 [1.27-1.30]* | 1.18 [1.17-1.20]* |
| E88 | 13,510 | 51,003,820 | 0.34 | 0.25 | 0.26 | 0.09 | 0.08 | 0.06 | 1.31 [1.23-1.39]* | 1.20 [1.13-1.27]* |
| E89 | 18,800 | 50,855,880 | 0.45 | 0.35 | 0.36 | 0.10 | 0.09 | 0.05 | 1.25 [1.19-1.31]* | 1.13 [1.08-1.19]* |
| 40+ - F01-F99 | | | | | | | | | | |
| F00 | 51,220 | 50,906,960 | 1.13 | 0.98 | 1.06 | 0.16 | 0.07 | 0.05 | 1.07 [1.03-1.10]* | 1.05 [1.01-1.08]* |
| F01 | 58,800 | 50,876,570 | 1.31 | 1.12 | 1.21 | 0.19 | 0.11 | 0.05 | 1.09 [1.06-1.12]* | 1.04 [1.00-1.07] |
| F02 | 10,170 | 51,030,340 | 0.21 | 0.20 | 0.21 | 0.01 | 0.00 | 0.00 | 1.02 [0.95-1.10] | 1.00 [0.93-1.08] |
| F03 | 141,060 | 50,594,100 | 3.16 | 2.71 | 2.95 | 0.45 | 0.20 | 0.13 | 1.07 [1.05-1.09]* | 1.04 [1.02-1.06]* |
| F05 | 128,470 | 50,751,990 | 3.04 | 2.42 | 2.60 | 0.62 | 0.44 | 0.28 | 1.17 [1.15-1.19]* | 1.10 [1.08-1.13]* |
| F06 | 19,870 | 50,969,820 | 0.46 | 0.37 | 0.39 | 0.08 | 0.07 | 0.04 | 1.17 [1.11-1.23]* | 1.11 [1.05-1.16]* |
| F07 | 910 | 51,047,000 | 0.02 | 0.02 | 0.02 | 0.01 | 0.01 | 0.00 | 1.35 [1.09-1.68]* | 1.23 [0.98-1.53] |
| F10 | 76,830 | 49,711,970 | 1.86 | 1.48 | 1.48 | 0.38 | 0.39 | 0.25 | 1.26 [1.23-1.29]* | 1.15 [1.12-1.18]* |
| F11 | 3,890 | 50,914,360 | 0.09 | 0.07 | 0.07 | 0.02 | 0.02 | 0.00 | 1.30 [1.17-1.45]* | 1.05 [0.94-1.17] |
| F12 | 4,870 | 50,981,300 | 0.12 | 0.09 | 0.09 | 0.02 | 0.03 | 0.01 | 1.28 [1.17-1.41]* | 1.06 [0.96-1.17] |
| F13 | 1,040 | 51,041,610 | 0.03 | 0.02 | 0.02 | 0.01 | 0.01 | 0.00 | 1.42 [1.16-1.74]* | 1.14 [0.92-1.41] |
| F14 | 1,740 | 51,018,850 | 0.05 | 0.03 | 0.03 | 0.01 | 0.01 | 0.01 | 1.42 [1.22-1.65]* | 1.13 [0.97-1.33] |
| F15 | 740 | 51,040,460 | 0.02 | 0.01 | 0.01 | 0.00 | 0.00 | 0.00 | 1.16 [0.91-1.48] | 0.98 [0.76-1.27] |
| F17 | 323,530 | 47,834,770 | 7.63 | 6.58 | 6.63 | 1.06 | 1.01 | 0.30 | 1.15 [1.14-1.17]* | 1.04 [1.03-1.05]* |
| F19 | 2,580 | 50,991,480 | 0.06 | 0.05 | 0.05 | 0.01 | 0.01 | 0.00 | 1.22 [1.07-1.39]* | 0.96 [0.84-1.10] |
| F20 | 11,300 | 50,741,280 | 0.28 | 0.21 | 0.21 | 0.07 | 0.07 | 0.06 | 1.32 [1.24-1.40]* | 1.25 [1.18-1.34]* |
| F22 | 5,830 | 50,989,350 | 0.14 | 0.11 | 0.11 | 0.04 | 0.04 | 0.03 | 1.32 [1.21-1.45]* | 1.22 [1.11-1.33]* |
| F23 | 2,300 | 51,003,230 | 0.06 | 0.04 | 0.04 | 0.01 | 0.01 | 0.01 | 1.28 [1.12-1.48]* | 1.20 [1.04-1.38] |
| F25 | 2,230 | 50,991,550 | 0.06 | 0.04 | 0.04 | 0.02 | 0.02 | 0.01 | 1.38 [1.20-1.58]* | 1.27 [1.10-1.46]* |
| F29 | 4,710 | 50,999,260 | 0.12 | 0.09 | 0.09 | 0.04 | 0.04 | 0.02 | 1.40 [1.28-1.54]* | 1.24 [1.13-1.37]* |
| F30 | 1,490 | 51,014,580 | 0.04 | 0.03 | 0.03 | 0.01 | 0.01 | 0.01 | 1.42 [1.21-1.68]* | 1.31 [1.11-1.55]* |
| F31 | 12,140 | 50,810,570 | 0.30 | 0.23 | 0.23 | 0.08 | 0.08 | 0.03 | 1.33 [1.25-1.41]* | 1.12 [1.06-1.20]* |
| F32 | 249,590 | 48,243,350 | 6.81 | 4.82 | 4.90 | 1.99 | 1.91 | 0.90 | 1.39 [1.37-1.41]* | 1.15 [1.14-1.17]* |
| F33 | 8,570 | 50,840,480 | 0.21 | 0.16 | 0.16 | 0.05 | 0.05 | 0.01 | 1.28 [1.19-1.37]* | 1.05 [0.98-1.14] |
| F34 | 1,240 | 51,039,970 | 0.03 | 0.02 | 0.02 | 0.01 | 0.01 | 0.01 | 1.52 [1.27-1.83]* | 1.24 [1.02-1.50] |
| F38 | 590 | 51,055,860 | 0.01 | 0.01 | 0.01 | 0.00 | 0.00 | 0.00 | 1.21 [0.92-1.61] | 1.04 [0.78-1.39] |
| F39 | 1,030 | 51,048,340 | 0.03 | 0.02 | 0.02 | 0.01 | 0.01 | 0.00 | 1.33 [1.08-1.64]* | 1.15 [0.93-1.43] |
| F40 | 19,620 | 50,875,490 | 0.49 | 0.36 | 0.38 | 0.12 | 0.11 | 0.06 | 1.30 [1.24-1.36]* | 1.14 [1.08-1.19]* |
| F41 | 199,420 | 49,443,920 | 5.25 | 3.77 | 3.86 | 1.47 | 1.38 | 0.62 | 1.36 [1.34-1.38]* | 1.13 [1.12-1.15]* |
| F42 | 3,130 | 51,016,800 | 0.10 | 0.05 | 0.05 | 0.05 | 0.05 | 0.03 | 1.84 [1.65-2.06]* | 1.53 [1.37-1.72]* |
| F43 | 8,660 | 50,886,440 | 0.22 | 0.16 | 0.16 | 0.06 | 0.06 | 0.01 | 1.35 [1.26-1.45]* | 1.07 [0.99-1.15] |
| F44 | 2,150 | 51,028,350 | 0.06 | 0.04 | 0.04 | 0.02 | 0.02 | 0.01 | 1.45 [1.27-1.67]* | 1.15 [0.99-1.33] |
| F45 | 3,460 | 50,992,520 | 0.09 | 0.06 | 0.07 | 0.03 | 0.02 | 0.01 | 1.37 [1.23-1.53]* | 1.16 [1.04-1.30] |
| F48 | 830 | 51,048,760 | 0.02 | 0.01 | 0.02 | 0.01 | 0.01 | 0.00 | 1.51 [1.21-1.87]* | 1.20 [0.95-1.51] |
| F50 | 1,890 | 51,023,680 | 0.05 | 0.03 | 0.03 | 0.01 | 0.01 | 0.01 | 1.42 [1.22-1.64]* | 1.19 [1.02-1.39] |
| F51 | 800 | 51,053,200 | 0.02 | 0.01 | 0.01 | 0.01 | 0.01 | 0.00 | 1.50 [1.20-1.88]* | 1.27 [1.00-1.60] |
| F52 | 2,440 | 51,017,710 | 0.06 | 0.04 | 0.04 | 0.02 | 0.02 | 0.01 | 1.48 [1.30-1.68]* | 1.26 [1.10-1.44]* |
| F60 | 5,580 | 50,915,090 | 0.16 | 0.10 | 0.10 | 0.06 | 0.06 | 0.03 | 1.59 [1.46-1.72]* | 1.23 [1.13-1.35]* |
| F69 | 520 | 51,050,000 | 0.02 | 0.01 | 0.01 | 0.01 | 0.01 | 0.01 | 1.81 [1.39-2.36]* | 1.49 [1.13-1.97]* |
| F70 | 1,020 | 51,034,890 | 0.04 | 0.02 | 0.02 | 0.02 | 0.02 | 0.02 | 2.17 [1.80-2.61]* | 1.81 [1.49-2.20]* |
| F71 | 360 | 51,050,760 | 0.02 | 0.01 | 0.01 | 0.01 | 0.01 | 0.01 | 2.90 [2.15-3.91]* | 2.62 [1.92-3.56]* |
| F72 | 600 | 51,044,950 | 0.02 | 0.01 | 0.01 | 0.02 | 0.02 | 0.01 | 2.79 [2.21-3.52]* | 2.58 [2.02-3.29]* |
| F79 | 4,320 | 50,988,970 | 0.15 | 0.07 | 0.07 | 0.08 | 0.08 | 0.07 | 2.13 [1.94-2.33]* | 1.93 [1.76-2.12]* |
| F80 | 2,620 | 51,040,090 | 0.07 | 0.05 | 0.05 | 0.02 | 0.02 | 0.02 | 1.41 [1.24-1.61]* | 1.31 [1.14-1.49]* |

| | | | | | | | | | | |
|---|---|---|---|---|---|---|---|---|---|---|
| F81 | 11,100 | 50,831,980 | 0.38 | 0.18 | 0.18 | 0.20 | 0.21 | 0.19 | 2.15 [2.03-2.27]* | 1.99 [1.88-2.11]* |
| F84 | 2,250 | 51,026,060 | 0.08 | 0.04 | 0.04 | 0.04 | 0.04 | 0.03 | 2.15 [1.90-2.44]* | 1.73 [1.51-1.97]* |
| F90 | 500 | 51,057,130 | 0.02 | 0.01 | 0.01 | 0.01 | 0.01 | 0.00 | 1.75 [1.34-2.29]* | 1.26 [0.94-1.68] |
| F91 | 560 | 51,054,260 | 0.01 | 0.01 | 0.01 | 0.00 | 0.00 | 0.00 | 1.09 [0.81-1.37] | 1.01 [0.74-1.37] |
| F99 | 3,250 | 50,975,700 | 0.08 | 0.06 | 0.06 | 0.02 | 0.02 | 0.01 | 1.30 [1.16-1.46]* | 1.12 [0.99-1.26] |
| 40+ - G00-G99 | | | | | | | | | | |
| G00 | 940 | 51,046,440 | 0.02 | 0.02 | 0.02 | 0.00 | 0.00 | 0.00 | 1.25 [1.01-1.55] | 1.20 [0.96-1.49] |
| G03 | 1,040 | 51,047,570 | 0.02 | 0.02 | 0.02 | 0.00 | 0.00 | 0.00 | 1.13 [0.92-1.40] | 1.00 [0.80-1.24] |
| G04 | 3,180 | 51,031,760 | 0.08 | 0.06 | 0.06 | 0.02 | 0.01 | 0.01 | 1.23 [1.09-1.39]* | 1.12 [0.99-1.27] |
| G05 | 820 | 51,054,380 | 0.02 | 0.01 | 0.01 | 0.01 | 0.01 | 0.01 | 1.60 [1.28-2.00]* | 1.48 [1.18-1.86]* |
| G06 | 1,580 | 51,043,730 | 0.04 | 0.03 | 0.03 | 0.01 | 0.01 | 0.00 | 1.18 [1.00-1.41] | 1.11 [0.93-1.33] |
| G08 | 600 | 51,055,130 | 0.01 | 0.01 | 0.01 | 0.00 | 0.00 | 0.00 | 1.14 [0.86-1.51] | 1.06 [0.79-1.42] |
| G09 | 800 | 51,050,030 | 0.02 | 0.01 | 0.02 | 0.00 | 0.00 | 0.00 | 1.24 [0.98-1.56] | 1.11 [0.87-1.41] |
| G10 | 540 | 51,054,360 | 0.01 | 0.01 | 0.01 | 0.00 | 0.00 | 0.00 | 1.26 [0.94-1.67] | 1.29 [0.97-1.73] |
| G11 | 1,590 | 51,041,700 | 0.04 | 0.03 | 0.03 | 0.01 | 0.01 | 0.00 | 1.20 [1.02-1.43] | 1.10 [0.93-1.31] |
| G12 | 4,150 | 51,043,510 | 0.09 | 0.08 | 0.08 | 0.01 | 0.01 | 0.00 | 1.10 [0.99-1.22] | 1.06 [0.95-1.18] |
| G20 | 39,000 | 50,807,970 | 0.83 | 0.75 | 0.78 | 0.07 | 0.05 | 0.02 | 1.06 [1.02-1.10]* | 1.03 [0.99-1.07] |
| G21 | 3,270 | 51,045,340 | 0.08 | 0.06 | 0.06 | 0.02 | 0.02 | 0.02 | 1.34 [1.19-1.51]* | 1.28 [1.13-1.44]* |
| G23 | 2,060 | 51,054,230 | 0.05 | 0.04 | 0.04 | 0.01 | 0.00 | 0.00 | 1.08 [0.93-1.27] | 1.03 [0.88-1.21] |
| G24 | 3,260 | 51,018,790 | 0.08 | 0.06 | 0.06 | 0.02 | 0.01 | 0.01 | 1.21 [1.08-1.37]* | 1.11 [0.99-1.26] |
| G25 | 19,670 | 50,937,650 | 0.49 | 0.36 | 0.38 | 0.12 | 0.11 | 0.05 | 1.28 [1.22-1.35]* | 1.11 [1.06-1.17]* |
| G30 | 68,520 | 50,819,790 | 1.52 | 1.31 | 1.41 | 0.21 | 0.11 | 0.08 | 1.08 [1.05-1.11]* | 1.06 [1.03-1.09]* |
| G31 | 48,660 | 50,840,080 | 1.12 | 0.92 | 0.96 | 0.20 | 0.16 | 0.10 | 1.17 [1.13-1.20]* | 1.10 [1.07-1.14]* |
| G32 | 690 | 51,054,850 | 0.02 | 0.01 | 0.01 | 0.01 | 0.01 | 0.00 | 1.47 [1.15-1.89]* | 1.30 [1.01-1.68] |
| G35 | 8,150 | 50,835,220 | 0.19 | 0.15 | 0.15 | 0.03 | 0.03 | 0.02 | 1.23 [1.14-1.32]* | 1.09 [1.01-1.18] |
| G37 | 1,770 | 51,015,600 | 0.04 | 0.03 | 0.03 | 0.01 | 0.01 | 0.01 | 1.36 [1.17-1.59]* | 1.24 [1.06-1.46]* |
| G40 | 48,710 | 50,113,760 | 1.26 | 0.91 | 0.91 | 0.35 | 0.35 | 0.21 | 1.38 [1.34-1.42]* | 1.20 [1.16-1.23]* |
| G41 | 3,140 | 51,025,780 | 0.08 | 0.06 | 0.06 | 0.02 | 0.02 | 0.01 | 1.29 [1.15-1.46]* | 1.15 [1.02-1.30] |
| G43 | 38,220 | 50,543,910 | 0.97 | 0.71 | 0.72 | 0.25 | 0.24 | 0.09 | 1.33 [1.29-1.38]* | 1.10 [1.06-1.14]* |
| G44 | 5,910 | 50,957,330 | 0.15 | 0.11 | 0.11 | 0.04 | 0.04 | 0.02 | 1.37 [1.26-1.49]* | 1.15 [1.06-1.26]* |
| G45 | 46,400 | 50,511,080 | 1.07 | 0.89 | 0.91 | 0.19 | 0.16 | 0.09 | 1.18 [1.14-1.22]* | 1.10 [1.06-1.13]* |
| G46 | 3,030 | 51,042,520 | 0.07 | 0.06 | 0.06 | 0.01 | 0.01 | 0.01 | 1.19 [1.05-1.36]* | 1.12 [0.98-1.28] |
| G47 | 52,080 | 50,370,280 | 1.48 | 0.94 | 0.94 | 0.55 | 0.55 | 0.32 | 1.58 [1.54-1.63]* | 1.27 [1.24-1.31]* |
| G50 | 6,930 | 50,974,860 | 0.18 | 0.13 | 0.13 | 0.06 | 0.06 | 0.04 | 1.44 [1.33-1.56]* | 1.24 [1.15-1.35]* |
| G51 | 13,270 | 50,891,810 | 0.31 | 0.25 | 0.26 | 0.06 | 0.05 | 0.03 | 1.21 [1.14-1.29]* | 1.10 [1.04-1.17]* |
| G52 | 1,040 | 51,048,930 | 0.03 | 0.02 | 0.02 | 0.01 | 0.01 | 0.00 | 1.28 [1.04-1.57] | 1.15 [0.93-1.43] |
| G53 | 2,510 | 51,041,530 | 0.06 | 0.05 | 0.05 | 0.02 | 0.02 | 0.01 | 1.35 [1.18-1.55]* | 1.25 [1.09-1.44]* |
| G54 | 4,130 | 51,004,160 | 0.10 | 0.08 | 0.08 | 0.03 | 0.03 | 0.01 | 1.35 [1.22-1.50]* | 1.16 [1.04-1.29]* |
| G55 | 45,780 | 50,421,160 | 1.11 | 0.86 | 0.87 | 0.25 | 0.24 | 0.11 | 1.28 [1.24-1.32]* | 1.11 [1.08-1.15]* |
| G56 | 72,630 | 49,335,500 | 1.85 | 1.39 | 1.39 | 0.46 | 0.46 | 0.27 | 1.33 [1.30-1.36]* | 1.17 [1.14-1.20]* |
| G57 | 8,260 | 50,900,430 | 0.21 | 0.15 | 0.15 | 0.05 | 0.05 | 0.03 | 1.35 [1.26-1.45]* | 1.19 [1.11-1.28]* |
| G58 | 4,040 | 50,998,280 | 0.10 | 0.07 | 0.07 | 0.03 | 0.03 | 0.02 | 1.41 [1.27-1.56]* | 1.22 [1.10-1.36]* |
| G59 | 3,570 | 51,015,790 | 0.10 | 0.06 | 0.06 | 0.03 | 0.03 | 0.01 | 1.49 [1.34-1.66]* | 1.18 [1.05-1.32]* |
| G60 | 1,900 | 51,037,270 | 0.05 | 0.04 | 0.04 | 0.01 | 0.01 | 0.00 | 1.26 [1.09-1.47]* | 1.11 [0.95-1.30] |
| G61 | 2,350 | 51,021,330 | 0.06 | 0.04 | 0.04 | 0.01 | 0.01 | 0.01 | 1.24 [1.08-1.43]* | 1.17 [1.02-1.35] |
| G62 | 28,670 | 50,845,350 | 0.72 | 0.53 | 0.54 | 0.20 | 0.19 | 0.10 | 1.35 [1.30-1.40]* | 1.15 [1.11-1.20]* |
| G63 | 15,330 | 50,958,270 | 0.41 | 0.28 | 0.28 | 0.14 | 0.13 | 0.06 | 1.46 [1.38-1.54]* | 1.17 [1.11-1.24]* |
| G70 | 2,390 | 51,030,860 | 0.05 | 0.05 | 0.05 | 0.01 | 0.01 | 0.00 | 1.17 [1.02-1.35] | 1.09 [0.94-1.26] |
| G71 | 1,360 | 51,036,470 | 0.03 | 0.03 | 0.03 | 0.00 | 0.00 | 0.00 | 1.14 [0.95-1.36] | 1.08 [0.89-1.30] |
| G72 | 2,900 | 51,032,450 | 0.08 | 0.05 | 0.05 | 0.02 | 0.02 | 0.02 | 1.41 [1.25-1.59]* | 1.29 [1.14-1.46]* |
| G80 | 2,150 | 50,996,680 | 0.06 | 0.04 | 0.04 | 0.02 | 0.02 | 0.02 | 1.66 [1.46-1.90]* | 1.59 [1.38-1.82]* |
| G81 | 52,240 | 50,596,550 | 1.21 | 0.99 | 1.03 | 0.22 | 0.18 | 0.11 | 1.17 [1.14-1.21]* | 1.10 [1.07-1.13]* |
| G82 | 4,620 | 50,981,500 | 0.11 | 0.09 | 0.09 | 0.02 | 0.02 | 0.01 | 1.21 [1.10-1.34]* | 1.13 [1.03-1.26] |
| G83 | 7,060 | 50,990,170 | 0.17 | 0.13 | 0.14 | 0.03 | 0.03 | 0.02 | 1.23 [1.13-1.33]* | 1.12 [1.03-1.21]* |
| G90 | 3,600 | 51,024,660 | 0.08 | 0.07 | 0.07 | 0.02 | 0.01 | 0.01 | 1.22 [1.08-1.36]* | 1.08 [0.96-1.22] |
| G91 | 8,690 | 50,982,410 | 0.20 | 0.16 | 0.17 | 0.03 | 0.03 | 0.02 | 1.17 [1.09-1.26]* | 1.13 [1.04-1.21]* |
| G93 | 32,090 | 50,813,540 | 0.76 | 0.60 | 0.61 | 0.16 | 0.15 | 0.08 | 1.25 [1.21-1.30]* | 1.11 [1.07-1.16]* |
| G94 | 1,170 | 51,054,740 | 0.03 | 0.02 | 0.02 | 0.01 | 0.00 | 0.00 | 1.19 [0.98-1.45] | 1.16 [0.95-1.42] |
| G95 | 10,450 | 50,964,070 | 0.24 | 0.20 | 0.20 | 0.05 | 0.04 | 0.03 | 1.22 [1.14-1.31]* | 1.14 [1.06-1.22]* |
| G96 | 2,670 | 51,022,950 | 0.07 | 0.05 | 0.05 | 0.02 | 0.02 | 0.01 | 1.37 [1.21-1.55]* | 1.21 [1.06-1.37]* |
| G97 | 1,820 | 51,023,060 | 0.04 | 0.03 | 0.03 | 0.01 | 0.01 | 0.00 | 1.24 [1.06-1.45]* | 1.09 [0.93-1.28] |
| G98 | 950 | 51,055,440 | 0.03 | 0.02 | 0.02 | 0.01 | 0.01 | 0.00 | 1.53 [1.24-1.88]* | 1.22 [0.98-1.52] |
| G99 | 16,820 | 50,889,580 | 0.40 | 0.32 | 0.32 | 0.08 | 0.08 | 0.04 | 1.26 [1.19-1.32]* | 1.12 [1.06-1.18]* |
| 40+ - H00-H59 | | | | | | | | | | |
| H00 | 7,560 | 50,806,010 | 0.18 | 0.14 | 0.14 | 0.03 | 0.03 | 0.03 | 1.25 [1.15-1.34]* | 1.18 [1.10-1.28]* |
| H01 | 10,160 | 50,945,090 | 0.26 | 0.19 | 0.19 | 0.08 | 0.08 | 0.06 | 1.40 [1.31-1.50]* | 1.31 [1.22-1.40]* |
| H02 | 53,810 | 50,138,060 | 1.35 | 1.01 | 1.03 | 0.34 | 0.32 | 0.24 | 1.31 [1.28-1.35]* | 1.22 [1.19-1.26]* |
| H03 | 510 | 51,053,730 | 0.01 | 0.01 | 0.01 | 0.00 | 0.00 | 0.00 | 1.30 [0.96-1.76] | 1.18 [0.87-1.61] |
| H04 | 25,350 | 50,664,750 | 0.68 | 0.46 | 0.47 | 0.21 | 0.21 | 0.16 | 1.44 [1.39-1.51]* | 1.32 [1.26-1.37]* |
| H05 | 3,780 | 51,003,050 | 0.10 | 0.07 | 0.07 | 0.04 | 0.03 | 0.03 | 1.47 [1.33-1.63]* | 1.35 [1.21-1.50]* |
| H06 | 990 | 51,043,580 | 0.03 | 0.02 | 0.02 | 0.01 | 0.01 | 0.01 | 1.47 [1.20-1.80]* | 1.36 [1.11-1.68]* |
| H10 | 10,670 | 50,988,090 | 0.27 | 0.20 | 0.20 | 0.08 | 0.07 | 0.05 | 1.35 [1.26-1.45]* | 1.24 [1.16-1.33]* |
| H11 | 10,450 | 50,902,940 | 0.25 | 0.20 | 0.20 | 0.06 | 0.05 | 0.04 | 1.27 [1.19-1.36]* | 1.21 [1.13-1.29]* |
| H13 | 550 | 51,054,510 | 0.02 | 0.01 | 0.01 | 0.01 | 0.01 | 0.01 | 1.78 [1.35-2.34]* | 1.63 [1.23-2.16]* |
| H15 | 890 | 51,047,690 | 0.02 | 0.02 | 0.02 | 0.01 | 0.01 | 0.00 | 1.30 [1.04-1.62] | 1.18 [0.94-1.48] |
| H16 | 4,370 | 50,997,020 | 0.12 | 0.08 | 0.08 | 0.05 | 0.05 | 0.04 | 1.58 [1.44-1.75]* | 1.48 [1.34-1.64]* |
| H17 | 2,720 | 51,028,400 | 0.07 | 0.05 | 0.05 | 0.02 | 0.02 | 0.02 | 1.37 [1.21-1.56]* | 1.28 [1.12-1.46]* |
| H18 | 18,490 | 50,891,460 | 0.44 | 0.35 | 0.36 | 0.10 | 0.09 | 0.07 | 1.24 [1.18-1.31]* | 1.19 [1.13-1.25]* |
| H19 | 1,140 | 51,046,730 | 0.03 | 0.02 | 0.02 | 0.01 | 0.01 | 0.01 | 1.77 [1.47-2.14]* | 1.54 [1.27-1.88]* |
| H20 | 4,700 | 50,988,650 | 0.12 | 0.09 | 0.09 | 0.03 | 0.03 | 0.03 | 1.38 [1.25-1.51]* | 1.30 [1.18-1.43]* |
| H21 | 8,860 | 50,977,310 | 0.20 | 0.17 | 0.17 | 0.03 | 0.03 | 0.02 | 1.15 [1.07-1.24]* | 1.09 [1.01-1.18] |
| H25 | 219,740 | 48,641,500 | 5.34 | 4.34 | 4.51 | 1.00 | 0.83 | 0.60 | 1.18 [1.17-1.20]* | 1.13 [1.11-1.14]* |
| H26 | 308,810 | 47,219,390 | 7.92 | 6.24 | 6.55 | 1.67 | 1.36 | 0.98 | 1.21 [1.19-1.22]* | 1.14 [1.13-1.16]* |
| H27 | 3,880 | 51,003,690 | 0.09 | 0.07 | 0.08 | 0.02 | 0.01 | 0.01 | 1.19 [1.07-1.33]* | 1.14 [1.01-1.27] |
| H28 | 1,610 | 51,044,140 | 0.04 | 0.03 | 0.03 | 0.01 | 0.01 | 0.00 | 1.28 [1.08-1.51]* | 1.13 [0.95-1.34] |
| H31 | 5,020 | 51,015,020 | 0.11 | 0.10 | 0.10 | 0.02 | 0.01 | 0.01 | 1.10 [1.00-1.22] | 1.06 [0.96-1.17] |
| H33 | 18,780 | 50,703,150 | 0.38 | 0.37 | 0.37 | 0.01 | 0.02 | 0.01 | 1.04 [0.99-1.10] | 1.02 [0.97-1.08] |
| H34 | 12,450 | 50,937,910 | 0.28 | 0.24 | 0.25 | 0.05 | 0.04 | 0.02 | 1.15 [1.08-1.22]* | 1.09 [1.02-1.16]* |
| H35 | 113,160 | 50,085,220 | 2.65 | 2.18 | 2.28 | 0.47 | 0.37 | 0.26 | 1.16 [1.14-1.19]* | 1.11 [1.09-1.13]* |
| H36 | 44,100 | 50,619,510 | 1.06 | 0.83 | 0.85 | 0.23 | 0.21 | 0.08 | 1.24 [1.20-1.28]* | 1.08 [1.05-1.12]* |
| H40 | 104,530 | 50,002,190 | 2.45 | 2.01 | 2.08 | 0.43 | 0.37 | 0.28 | 1.18 [1.15-1.20]* | 1.13 [1.11-1.16]* |
| H43 | 20,450 | 50,824,110 | 0.46 | 0.39 | 0.40 | 0.06 | 0.06 | 0.03 | 1.14 [1.09-1.20]* | 1.08 [1.03-1.13]* |
| H44 | 6,190 | 50,985,090 | 0.15 | 0.12 | 0.12 | 0.03 | 0.03 | 0.03 | 1.29 [1.19-1.41]* | 1.22 [1.12-1.33]* |
| H46 | 1,450 | 51,039,480 | 0.04 | 0.03 | 0.03 | 0.01 | 0.01 | 0.01 | 1.33 [1.12-1.58]* | 1.20 [1.01-1.44] |
| H47 | 5,330 | 50,998,810 | 0.12 | 0.10 | 0.10 | 0.02 | 0.02 | 0.02 | 1.24 [1.13-1.36]* | 1.18 [1.08-1.30]* |
| H49 | 4,380 | 51,011,310 | 0.09 | 0.08 | 0.08 | 0.01 | 0.01 | 0.00 | 1.13 [1.02-1.26] | 1.05 [0.94-1.17] |
| H50 | 5,860 | 50,919,680 | 0.14 | 0.11 | 0.11 | 0.03 | 0.03 | 0.02 | 1.29 [1.18-1.40]* | 1.20 [1.10-1.31]* |
| H51 | 610 | 51,054,620 | 0.01 | 0.01 | 0.01 | 0.00 | 0.00 | 0.00 | 1.11 [0.84-1.48] | 1.01 [0.76-1.35] |
| H52 | 32,660 | 50,799,520 | 0.73 | 0.63 | 0.65 | 0.10 | 0.08 | 0.05 | 1.12 [1.08-1.17]* | 1.08 [1.04-1.12]* |
| H53 | 49,140 | 50,447,830 | 1.18 | 0.93 | 0.95 | 0.25 | 0.24 | 0.13 | 1.25 [1.21-1.29]* | 1.13 [1.09-1.16]* |
| H54 | 48,680 | 50,759,720 | 1.15 | 0.92 | 0.95 | 0.23 | 0.20 | 0.14 | 1.21 [1.17-1.25]* | 1.14 [1.10-1.17]* |
| H55 | 3,690 | 51,025,380 | 0.09 | 0.07 | 0.07 | 0.02 | 0.01 | 0.00 | 1.15 [1.03-1.29] | 1.04 [0.93-1.17] |
| H57 | 9,250 | 50,965,450 | 0.22 | 0.17 | 0.18 | 0.05 | 0.05 | 0.03 | 1.28 [1.19-1.37]* | 1.16 [1.08-1.25]* |
| H58 | 2,130 | 51,043,570 | 0.05 | 0.04 | 0.04 | 0.01 | 0.01 | 0.00 | 1.17 [1.01-1.36] | 1.06 [0.91-1.23] |
| H59 | 4,040 | 51,002,540 | 0.09 | 0.08 | 0.08 | 0.01 | 0.01 | 0.00 | 1.10 [0.98-1.22] | 1.06 [0.95-1.18] |
| 40+ - H60-H95 | | | | | | | | | | |

| | | | | | | | | | | |
|---|---|---|---|---|---|---|---|---|---|---|
| H60 | 6,300 | 50,931,690 | 0.20 | 0.11 | 0.11 | 0.09 | 0.09 | 0.08 | 1.85 [1.71-2.00]* | 1.67 [1.54-1.81]* |
| H61 | 10,750 | 50,879,750 | 0.29 | 0.20 | 0.20 | 0.09 | 0.09 | 0.07 | 1.45 [1.36-1.55]* | 1.34 [1.26-1.43]* |
| H65 | 5,350 | 50,898,270 | 0.13 | 0.10 | 0.10 | 0.03 | 0.03 | 0.03 | 1.35 [1.24-1.48]* | 1.24 [1.13-1.35]* |
| H66 | 5,900 | 50,915,990 | 0.16 | 0.11 | 0.11 | 0.06 | 0.06 | 0.04 | 1.52 [1.40-1.65]* | 1.37 [1.26-1.50]* |
| H69 | 1,080 | 51,038,920 | 0.03 | 0.02 | 0.02 | 0.01 | 0.01 | 0.01 | 1.47 [1.21-1.79]* | 1.27 [1.04-1.55] |
| H70 | 1,280 | 51,042,520 | 0.03 | 0.02 | 0.03 | 0.01 | 0.01 | 0.00 | 1.24 [1.03-1.50] | 1.13 [0.93-1.38] |
| H71 | 2,670 | 50,978,220 | 0.06 | 0.05 | 0.05 | 0.01 | 0.01 | 0.00 | 1.12 [0.98-1.28] | 1.07 [0.93-1.22] |
| H72 | 5,080 | 50,894,630 | 0.12 | 0.09 | 0.10 | 0.03 | 0.03 | 0.02 | 1.31 [1.19-1.43]* | 1.20 [1.10-1.32]* |
| H73 | 1,990 | 51,000,810 | 0.05 | 0.04 | 0.04 | 0.01 | 0.01 | 0.01 | 1.34 [1.16-1.55]* | 1.23 [1.06-1.42]* |
| H74 | 2,830 | 50,984,940 | 0.07 | 0.05 | 0.05 | 0.02 | 0.02 | 0.01 | 1.33 [1.18-1.51]* | 1.24 [1.09-1.41]* |
| H80 | 1,080 | 51,028,370 | 0.02 | 0.02 | 0.02 | 0.00 | 0.00 | 0.00 | 1.14 [0.93-1.40] | 1.12 [0.90-1.38] |
| H81 | 20,920 | 50,866,910 | 0.51 | 0.39 | 0.40 | 0.12 | 0.10 | 0.06 | 1.26 [1.20-1.32]* | 1.14 [1.09-1.20]* |
| H83 | 7,820 | 50,938,780 | 0.19 | 0.15 | 0.15 | 0.04 | 0.04 | 0.02 | 1.26 [1.17-1.36]* | 1.13 [1.04-1.22]* |
| H90 | 17,240 | 50,907,620 | 0.44 | 0.32 | 0.32 | 0.13 | 0.12 | 0.09 | 1.37 [1.30-1.44]* | 1.24 [1.18-1.31]* |
| H91 | 124,020 | 50,364,520 | 3.01 | 2.35 | 2.44 | 0.66 | 0.57 | 0.41 | 1.23 [1.21-1.26]* | 1.16 [1.13-1.18]* |
| H92 | 4,600 | 50,967,670 | 0.12 | 0.08 | 0.09 | 0.04 | 0.04 | 0.03 | 1.43 [1.30-1.57]* | 1.26 [1.14-1.39]* |
| H93 | 10,700 | 50,948,080 | 0.27 | 0.20 | 0.20 | 0.07 | 0.07 | 0.05 | 1.36 [1.27-1.44]* | 1.22 [1.14-1.30]* |
| H95 | 560 | 51,042,870 | 0.01 | 0.01 | 0.01 | 0.00 | 0.00 | 0.00 | 1.36 [1.04-1.79] | 1.30 [0.99-1.72] |
| 40+ - I00-I99 | | | | | | | | | | |
| I05 | 6,840 | 50,979,600 | 0.17 | 0.13 | 0.13 | 0.04 | 0.04 | 0.03 | 1.32 [1.21-1.43]* | 1.21 [1.11-1.32]* |
| I06 | 680 | 51,049,560 | 0.02 | 0.01 | 0.01 | 0.00 | 0.00 | 0.00 | 1.36 [1.06-1.75] | 1.22 [0.95-1.58] |
| I07 | 17,330 | 50,988,030 | 0.43 | 0.32 | 0.33 | 0.11 | 0.10 | 0.07 | 1.31 [1.25-1.38]* | 1.20 [1.14-1.27]* |
| I08 | 91,440 | 50,611,190 | 2.19 | 1.73 | 1.80 | 0.46 | 0.39 | 0.25 | 1.22 [1.19-1.25]* | 1.13 [1.10-1.16]* |
| I09 | 1,000 | 51,049,990 | 0.03 | 0.02 | 0.02 | 0.01 | 0.01 | 0.01 | 1.52 [1.24-1.86]* | 1.39 [1.13-1.71]* |
| I10 | 896,210 | 37,682,130 | 29.42 | 22.54 | 23.76 | 6.88 | 5.66 | 3.13 | 1.24 [1.23-1.25]* | 1.12 [1.11-1.13]* |
| I11 | 5,190 | 51,002,590 | 0.12 | 0.10 | 0.10 | 0.02 | 0.02 | 0.01 | 1.22 [1.11-1.35]* | 1.10 [0.99-1.21] |
| I12 | 51,700 | 50,598,080 | 1.26 | 0.97 | 0.96 | 0.29 | 0.30 | 0.20 | 1.31 [1.27-1.35]* | 1.19 [1.16-1.23]* |
| I13 | 2,610 | 51,054,060 | 0.06 | 0.05 | 0.05 | 0.02 | 0.01 | 0.01 | 1.28 [1.11-1.47]* | 1.14 [0.99-1.31] |
| I15 | 3,150 | 51,025,550 | 0.08 | 0.06 | 0.06 | 0.02 | 0.02 | 0.01 | 1.26 [1.12-1.42]* | 1.11 [0.98-1.26] |
| I20 | 201,260 | 47,185,870 | 5.27 | 4.05 | 4.12 | 1.22 | 1.14 | 0.64 | 1.28 [1.26-1.30]* | 1.14 [1.12-1.15]* |
| I21 | 137,370 | 49,410,410 | 3.18 | 2.70 | 2.75 | 0.48 | 0.42 | 0.24 | 1.15 [1.13-1.18]* | 1.08 [1.06-1.10]* |
| I22 | 12,570 | 50,887,210 | 0.30 | 0.24 | 0.23 | 0.06 | 0.07 | 0.04 | 1.29 [1.22-1.37]* | 1.16 [1.09-1.24]* |
| I23 | 750 | 51,051,210 | 0.02 | 0.01 | 0.01 | 0.00 | 0.00 | 0.00 | 1.30 [1.02-1.66] | 1.29 [1.00-1.65] |
| I24 | 38,920 | 50,775,070 | 0.94 | 0.73 | 0.76 | 0.21 | 0.18 | 0.09 | 1.24 [1.20-1.28]* | 1.11 [1.07-1.15]* |
| I25 | 345,430 | 46,002,200 | 9.11 | 7.16 | 7.45 | 1.95 | 1.66 | 0.90 | 1.22 [1.21-1.24]* | 1.11 [1.10-1.12]* |
| I26 | 67,350 | 50,474,730 | 1.48 | 1.30 | 1.33 | 0.18 | 0.15 | 0.06 | 1.12 [1.09-1.15]* | 1.04 [1.01-1.07]* |
| I27 | 42,020 | 50,869,870 | 1.05 | 0.78 | 0.80 | 0.27 | 0.25 | 0.16 | 1.31 [1.27-1.36]* | 1.18 [1.14-1.22]* |
| I28 | 950 | 51,053,440 | 0.02 | 0.02 | 0.02 | 0.00 | 0.00 | 0.00 | 1.16 [0.92-1.45] | 1.01 [0.80-1.28] |
| I30 | 1,600 | 51,029,510 | 0.04 | 0.03 | 0.03 | 0.01 | 0.01 | 0.01 | 1.26 [1.07-1.49]* | 1.19 [1.00-1.41] |
| I31 | 23,390 | 50,876,190 | 0.55 | 0.44 | 0.45 | 0.11 | 0.10 | 0.06 | 1.22 [1.17-1.28]* | 1.13 [1.08-1.18]* |
| I33 | 3,960 | 51,027,570 | 0.10 | 0.07 | 0.08 | 0.03 | 0.03 | 0.02 | 1.34 [1.20-1.49]* | 1.25 [1.12-1.39]* |
| I34 | 58,560 | 50,585,480 | 1.36 | 1.11 | 1.14 | 0.25 | 0.22 | 0.13 | 1.19 [1.16-1.23]* | 1.10 [1.07-1.14]* |
| I35 | 86,930 | 50,439,200 | 2.10 | 1.64 | 1.70 | 0.45 | 0.40 | 0.26 | 1.23 [1.20-1.26]* | 1.14 [1.11-1.17]* |
| I36 | 6,100 | 51,033,870 | 0.15 | 0.11 | 0.12 | 0.03 | 0.03 | 0.02 | 1.24 [1.13-1.35]* | 1.13 [1.04-1.24]* |
| I37 | 6,060 | 51,034,000 | 0.15 | 0.11 | 0.11 | 0.03 | 0.03 | 0.02 | 1.29 [1.18-1.41]* | 1.17 [1.07-1.28]* |
| I38 | 8,930 | 50,980,570 | 0.22 | 0.16 | 0.17 | 0.06 | 0.06 | 0.04 | 1.33 [1.24-1.43]* | 1.22 [1.14-1.32]* |
| I40 | 440 | 51,056,380 | 0.01 | 0.01 | 0.01 | 0.00 | 0.00 | 0.00 | 1.50 [1.10-2.04] | 1.39 [1.01-1.91] |
| I42 | 20,590 | 50,844,760 | 0.48 | 0.39 | 0.39 | 0.09 | 0.09 | 0.05 | 1.22 [1.16-1.28]* | 1.10 [1.05-1.16]* |
| I43 | 900 | 51,058,390 | 0.02 | 0.02 | 0.02 | 0.00 | 0.00 | 0.00 | 0.98 [0.77-1.25] | 0.92 [0.72-1.18] |
| I44 | 137,620 | 50,162,310 | 3.29 | 2.63 | 2.73 | 0.66 | 0.56 | 0.36 | 1.20 [1.18-1.23]* | 1.12 [1.10-1.14]* |
| I45 | 82,590 | 50,528,220 | 1.96 | 1.56 | 1.63 | 0.40 | 0.33 | 0.20 | 1.20 [1.17-1.23]* | 1.11 [1.08-1.14]* |
| I46 | 43,180 | 50,937,680 | 1.01 | 0.81 | 0.83 | 0.20 | 0.19 | 0.12 | 1.23 [1.19-1.27]* | 1.14 [1.10-1.18]* |
| I47 | 44,890 | 50,474,580 | 1.05 | 0.85 | 0.87 | 0.20 | 0.19 | 0.10 | 1.22 [1.18-1.26]* | 1.11 [1.07-1.15]* |
| I48 | 383,660 | 47,896,520 | 9.46 | 7.70 | 8.13 | 1.76 | 1.33 | 0.81 | 1.16 [1.15-1.18]* | 1.09 [1.08-1.11]* |
| I49 | 54,720 | 50,555,180 | 1.27 | 1.04 | 1.08 | 0.23 | 0.20 | 0.10 | 1.18 [1.15-1.22]* | 1.09 [1.06-1.12]* |
| I50 | 291,040 | 49,373,470 | 7.36 | 5.58 | 5.82 | 1.78 | 1.53 | 0.98 | 1.26 [1.25-1.28]* | 1.15 [1.14-1.17]* |
| I51 | 158,350 | 50,112,670 | 3.86 | 3.01 | 3.09 | 0.85 | 0.77 | 0.46 | 1.25 [1.23-1.27]* | 1.13 [1.11-1.16]* |
| I60 | 8,810 | 50,943,160 | 0.19 | 0.17 | 0.17 | 0.03 | 0.02 | 0.02 | 1.14 [1.06-1.22]* | 1.11 [1.03-1.19]* |
| I61 | 23,220 | 50,917,940 | 0.53 | 0.44 | 0.45 | 0.09 | 0.08 | 0.06 | 1.17 [1.12-1.22]* | 1.13 [1.08-1.19]* |
| I62 | 13,430 | 50,983,440 | 0.31 | 0.25 | 0.26 | 0.05 | 0.04 | 0.03 | 1.17 [1.10-1.25]* | 1.12 [1.05-1.19]* |
| I63 | 111,470 | 50,267,290 | 2.44 | 2.17 | 2.25 | 0.28 | 0.20 | 0.10 | 1.09 [1.07-1.11]* | 1.04 [1.02-1.07]* |
| I64 | 29,140 | 50,722,790 | 0.65 | 0.56 | 0.57 | 0.09 | 0.08 | 0.05 | 1.15 [1.10-1.19]* | 1.09 [1.05-1.14]* |
| I65 | 27,030 | 50,798,120 | 0.62 | 0.51 | 0.53 | 0.11 | 0.09 | 0.05 | 1.17 [1.12-1.22]* | 1.08 [1.04-1.13]* |
| I66 | 1,630 | 51,048,900 | 0.03 | 0.03 | 0.03 | 0.00 | 0.00 | 0.00 | 1.08 [0.90-1.28] | 1.00 [0.84-1.20] |
| I67 | 168,530 | 50,265,350 | 3.99 | 3.22 | 3.37 | 0.77 | 0.62 | 0.39 | 1.18 [1.16-1.20]* | 1.11 [1.09-1.13]* |
| I68 | 980 | 51,058,710 | 0.02 | 0.02 | 0.02 | 0.00 | 0.00 | 0.00 | 1.22 [0.98-1.52] | 1.16 [0.92-1.45] |
| I69 | 48,840 | 50,632,850 | 1.13 | 0.93 | 0.96 | 0.21 | 0.18 | 0.10 | 1.18 [1.15-1.22]* | 1.10 [1.06-1.13]* |
| I70 | 41,950 | 50,751,360 | 1.02 | 0.78 | 0.81 | 0.24 | 0.21 | 0.13 | 1.27 [1.22-1.31]* | 1.15 [1.11-1.19]* |
| I71 | 51,380 | 50,700,530 | 1.17 | 0.98 | 1.00 | 0.19 | 0.18 | 0.12 | 1.18 [1.14-1.21]* | 1.11 [1.08-1.15]* |
| I72 | 11,930 | 50,953,610 | 0.27 | 0.23 | 0.23 | 0.05 | 0.04 | 0.02 | 1.18 [1.10-1.25]* | 1.09 [1.02-1.16]* |
| I73 | 89,050 | 50,175,750 | 2.33 | 1.66 | 1.68 | 0.68 | 0.65 | 0.44 | 1.39 [1.36-1.42]* | 1.23 [1.20-1.26]* |
| I74 | 23,300 | 50,833,930 | 0.53 | 0.44 | 0.45 | 0.09 | 0.09 | 0.04 | 1.19 [1.14-1.25]* | 1.09 [1.04-1.14]* |
| I77 | 30,470 | 50,728,300 | 0.76 | 0.57 | 0.57 | 0.20 | 0.19 | 0.13 | 1.33 [1.28-1.39]* | 1.20 [1.16-1.25]* |
| I78 | 7,260 | 50,972,070 | 0.18 | 0.13 | 0.14 | 0.05 | 0.04 | 0.03 | 1.31 [1.21-1.42]* | 1.21 [1.12-1.31]* |
| I79 | 3,090 | 51,032,090 | 0.08 | 0.06 | 0.06 | 0.02 | 0.02 | 0.01 | 1.35 [1.20-1.52]* | 1.13 [1.00-1.28] |
| I80 | 58,600 | 50,251,150 | 1.41 | 1.11 | 1.13 | 0.30 | 0.28 | 0.20 | 1.25 [1.21-1.28]* | 1.16 [1.13-1.20]* |
| I81 | 3,330 | 51,046,990 | 0.07 | 0.06 | 0.07 | 0.01 | 0.01 | 0.00 | 1.14 [1.01-1.28] | 1.06 [0.94-1.19] |
| I82 | 6,980 | 51,009,600 | 0.16 | 0.13 | 0.13 | 0.03 | 0.03 | 0.02 | 1.20 [1.11-1.30]* | 1.11 [1.02-1.20] |
| I83 | 64,120 | 49,388,940 | 1.96 | 1.16 | 1.17 | 0.80 | 0.79 | 0.73 | 1.68 [1.64-1.72]* | 1.59 [1.55-1.63]* |
| I84 | 98,060 | 48,352,680 | 2.55 | 1.92 | 1.90 | 0.63 | 0.65 | 0.47 | 1.34 [1.32-1.37]* | 1.22 [1.20-1.25]* |
| I85 | 9,220 | 50,974,330 | 0.25 | 0.17 | 0.17 | 0.08 | 0.08 | 0.06 | 1.47 [1.38-1.57]* | 1.31 [1.22-1.40]* |
| I86 | 5,300 | 50,981,350 | 0.13 | 0.10 | 0.10 | 0.03 | 0.03 | 0.02 | 1.36 [1.24-1.49]* | 1.21 [1.10-1.33]* |
| I87 | 14,890 | 50,953,980 | 0.44 | 0.26 | 0.26 | 0.17 | 0.17 | 0.15 | 1.65 [1.56-1.74]* | 1.51 [1.43-1.60]* |
| I88 | 1,330 | 51,039,960 | 0.04 | 0.02 | 0.02 | 0.01 | 0.01 | 0.01 | 1.53 [1.29-1.82]* | 1.45 [1.22-1.74]* |
| I89 | 20,640 | 50,929,470 | 0.57 | 0.37 | 0.38 | 0.20 | 0.19 | 0.15 | 1.51 [1.44-1.58]* | 1.36 [1.30-1.43]* |
| I95 | 195,670 | 50,039,650 | 4.71 | 3.74 | 3.91 | 0.97 | 0.80 | 0.50 | 1.20 [1.19-1.22]* | 1.12 [1.10-1.14]* |
| I97 | 2,500 | 51,012,680 | 0.06 | 0.05 | 0.05 | 0.01 | 0.01 | 0.01 | 1.28 [1.12-1.46]* | 1.19 [1.04-1.36] |
| I98 | 6,240 | 51,015,930 | 0.16 | 0.11 | 0.11 | 0.05 | 0.05 | 0.03 | 1.45 [1.34-1.58]* | 1.27 [1.17-1.38]* |
| I99 | 3,630 | 51,028,970 | 0.09 | 0.07 | 0.07 | 0.02 | 0.02 | 0.01 | 1.24 [1.11-1.39]* | 1.14 [1.02-1.28] |
| 40+ - J00-J99 | | | | | | | | | | |
| J00 | 3,250 | 50,991,260 | 0.08 | 0.06 | 0.06 | 0.02 | 0.02 | 0.02 | 1.38 [1.23-1.55]* | 1.25 [1.11-1.41]* |
| J01 | 1,650 | 51,024,000 | 0.04 | 0.03 | 0.03 | 0.01 | 0.01 | 0.01 | 1.38 [1.18-1.62]* | 1.16 [0.99-1.37] |
| J02 | 8,890 | 50,911,160 | 0.23 | 0.16 | 0.16 | 0.07 | 0.07 | 0.05 | 1.44 [1.34-1.54]* | 1.26 [1.18-1.36]* |
| J03 | 5,280 | 50,858,200 | 0.13 | 0.10 | 0.10 | 0.03 | 0.02 | 0.01 | 1.25 [1.14-1.36]* | 1.08 [0.98-1.19] |
| J04 | 1,720 | 51,033,090 | 0.04 | 0.03 | 0.03 | 0.01 | 0.01 | 0.01 | 1.41 [1.21-1.65]* | 1.19 [1.01-1.40] |
| J05 | 800 | 51,047,720 | 0.02 | 0.02 | 0.02 | 0.00 | 0.00 | 0.00 | 1.12 [0.88-1.43] | 1.03 [0.80-1.32] |
| J06 | 11,120 | 50,862,820 | 0.28 | 0.21 | 0.20 | 0.07 | 0.07 | 0.04 | 1.36 [1.28-1.45]* | 1.18 [1.11-1.26]* |
| J10 | 13,950 | 51,003,630 | 0.36 | 0.26 | 0.26 | 0.10 | 0.09 | 0.05 | 1.36 [1.28-1.44]* | 1.17 [1.10-1.24]* |
| J11 | 2,600 | 51,024,320 | 0.07 | 0.05 | 0.05 | 0.02 | 0.02 | 0.01 | 1.35 [1.19-1.54]* | 1.20 [1.05-1.37]* |
| J12 | 27,470 | 51,030,540 | 0.70 | 0.50 | 0.53 | 0.19 | 0.16 | 0.10 | 1.31 [1.25-1.36]* | 1.16 [1.11-1.21]* |
| J13 | 5,960 | 51,008,600 | 0.15 | 0.11 | 0.11 | 0.04 | 0.04 | 0.02 | 1.36 [1.24-1.48]* | 1.19 [1.09-1.30]* |
| J14 | 2,560 | 51,043,050 | 0.06 | 0.05 | 0.05 | 0.02 | 0.02 | 0.01 | 1.35 [1.19-1.54]* | 1.11 [0.96-1.27] |
| J15 | 12,540 | 50,973,360 | 0.33 | 0.23 | 0.23 | 0.10 | 0.10 | 0.06 | 1.43 [1.35-1.52]* | 1.23 [1.16-1.31]* |
| J17 | 2,590 | 51,044,320 | 0.07 | 0.05 | 0.05 | 0.02 | 0.02 | 0.02 | 1.46 [1.29-1.65]* | 1.31 [1.15-1.50]* |
| J18 | 393,090 | 49,071,460 | 9.93 | 7.60 | 7.87 | 2.33 | 2.05 | 1.34 | 1.26 [1.25-1.28]* | 1.16 [1.14-1.17]* |
| J20 | 4,360 | 51,017,010 | 0.11 | 0.08 | 0.08 | 0.03 | 0.02 | 0.01 | 1.26 [1.14-1.40]* | 1.12 [1.00-1.24] |
| J21 | 1,190 | 51,051,270 | 0.03 | 0.02 | 0.02 | 0.01 | 0.01 | 0.01 | 1.47 [1.22-1.78]* | 1.25 [1.03-1.53] |

| | | | | | | | | | | |
|---|---|---|---|---|---|---|---|---|---|---|
| J22 | 212,150 | 49,077,990 | 5.50 | 4.07 | 4.17 | 1.43 | 1.32 | 0.89 | 1.32 [1.30-1.34]* | 1.19 [1.17-1.21]* |
| J30 | 22,750 | 50,810,500 | 0.68 | 0.40 | 0.41 | 0.28 | 0.27 | 0.15 | 1.67 [1.60-1.74]* | 1.29 [1.23-1.35]* |
| J31 | 6,340 | 50,935,290 | 0.18 | 0.11 | 0.11 | 0.07 | 0.07 | 0.03 | 1.59 [1.47-1.72]* | 1.24 [1.14-1.34]* |
| J32 | 17,800 | 50,651,120 | 0.46 | 0.33 | 0.33 | 0.14 | 0.14 | 0.07 | 1.42 [1.35-1.49]* | 1.18 [1.13-1.25]* |
| J33 | 13,520 | 50,590,400 | 0.37 | 0.25 | 0.24 | 0.12 | 0.13 | 0.07 | 1.52 [1.44-1.61]* | 1.25 [1.18-1.32]* |
| J34 | 27,720 | 49,934,600 | 0.75 | 0.51 | 0.50 | 0.23 | 0.24 | 0.16 | 1.48 [1.43-1.54]* | 1.28 [1.23-1.33]* |
| J35 | 5,320 | 50,754,380 | 0.13 | 0.10 | 0.10 | 0.02 | 0.02 | 0.01 | 1.23 [1.13-1.35]* | 1.10 [1.01-1.21] |
| J36 | 2,750 | 50,951,020 | 0.06 | 0.05 | 0.05 | 0.01 | 0.01 | 0.00 | 1.14 [1.00-1.30] | 1.09 [0.95-1.25] |
| J37 | 500 | 51,044,810 | 0.01 | 0.01 | 0.01 | 0.00 | 0.00 | 0.00 | 1.31 [0.98-1.75] | 1.10 [0.81-1.49] |
| J38 | 16,580 | 50,765,550 | 0.40 | 0.31 | 0.31 | 0.09 | 0.09 | 0.05 | 1.30 [1.24-1.37]* | 1.16 [1.10-1.22]* |
| J39 | 7,980 | 50,949,860 | 0.20 | 0.15 | 0.15 | 0.05 | 0.05 | 0.03 | 1.32 [1.23-1.43]* | 1.18 [1.09-1.27]* |
| J40 | 10,940 | 50,916,280 | 0.29 | 0.20 | 0.20 | 0.09 | 0.09 | 0.06 | 1.45 [1.36-1.54]* | 1.25 [1.17-1.33]* |
| J41 | 540 | 51,055,440 | 0.01 | 0.01 | 0.01 | 0.00 | 0.00 | 0.00 | 1.36 [1.03-1.81] | 1.18 [0.88-1.58] |
| J42 | 4,500 | 50,999,150 | 0.12 | 0.08 | 0.08 | 0.04 | 0.04 | 0.02 | 1.42 [1.29-1.56]* | 1.18 [1.07-1.31]* |
| J43 | 64,610 | 50,639,430 | 1.65 | 1.20 | 1.22 | 0.45 | 0.43 | 0.25 | 1.36 [1.32-1.39]* | 1.18 [1.15-1.22]* |
| J44 | 260,140 | 48,748,310 | 7.26 | 4.93 | 5.01 | 2.33 | 2.24 | 1.24 | 1.45 [1.43-1.47]* | 1.21 [1.19-1.22]* |
| J45 | 267,020 | 44,751,780 | 9.53 | 5.20 | 5.17 | 4.33 | 4.36 | 2.26 | 1.84 [1.82-1.87]* | 1.31 [1.29-1.33]* |
| J46 | 2,640 | 50,939,850 | 0.09 | 0.04 | 0.04 | 0.05 | 0.05 | 0.02 | 2.08 [1.85-2.33]* | 1.35 [1.18-1.54]* |
| J47 | 46,310 | 50,734,820 | 1.28 | 0.84 | 0.85 | 0.44 | 0.43 | 0.27 | 1.50 [1.45-1.55]* | 1.26 [1.22-1.30]* |
| J61 | 6,480 | 51,024,720 | 0.17 | 0.12 | 0.12 | 0.05 | 0.05 | 0.04 | 1.40 [1.28-1.52]* | 1.27 [1.17-1.38]* |
| J67 | 1,840 | 51,048,120 | 0.05 | 0.03 | 0.03 | 0.02 | 0.02 | 0.01 | 1.59 [1.37-1.84]* | 1.40 [1.20-1.63]* |
| J69 | 61,920 | 50,925,540 | 1.45 | 1.17 | 1.23 | 0.28 | 0.22 | 0.16 | 1.18 [1.14-1.21]* | 1.12 [1.09-1.15]* |
| J70 | 1,250 | 51,055,650 | 0.03 | 0.02 | 0.02 | 0.01 | 0.01 | 0.01 | 1.51 [1.25-1.81]* | 1.36 [1.13-1.64]* |
| J80 | 4,280 | 51,035,810 | 0.11 | 0.08 | 0.08 | 0.03 | 0.03 | 0.03 | 1.45 [1.31-1.60]* | 1.31 [1.18-1.45]* |
| J81 | 21,550 | 50,925,270 | 0.53 | 0.40 | 0.40 | 0.12 | 0.12 | 0.09 | 1.31 [1.25-1.37]* | 1.21 [1.15-1.27]* |
| J82 | 1,430 | 51,052,560 | 0.05 | 0.02 | 0.02 | 0.03 | 0.03 | 0.02 | 2.11 [1.80-2.47]* | 1.49 [1.25-1.77]* |
| J84 | 40,380 | 50,881,670 | 1.09 | 0.73 | 0.75 | 0.36 | 0.33 | 0.27 | 1.44 [1.40-1.49]* | 1.33 [1.28-1.37]* |
| J85 | 1,870 | 51,048,100 | 0.05 | 0.03 | 0.03 | 0.01 | 0.01 | 0.01 | 1.37 [1.18-1.59]* | 1.22 [1.05-1.43] |
| J86 | 6,360 | 51,005,380 | 0.15 | 0.12 | 0.12 | 0.03 | 0.03 | 0.02 | 1.25 [1.15-1.36]* | 1.16 [1.07-1.27]* |
| J90 | 200,180 | 50,136,900 | 4.74 | 3.83 | 3.96 | 0.91 | 0.78 | 0.52 | 1.20 [1.18-1.22]* | 1.12 [1.10-1.14]* |
| J91 | 4,760 | 51,045,380 | 0.11 | 0.09 | 0.09 | 0.02 | 0.02 | 0.01 | 1.21 [1.09-1.34]* | 1.13 [1.02-1.25] |
| J92 | 23,440 | 50,946,430 | 0.56 | 0.44 | 0.45 | 0.12 | 0.11 | 0.09 | 1.26 [1.20-1.31]* | 1.18 [1.13-1.24]* |
| J93 | 16,040 | 50,914,360 | 0.36 | 0.31 | 0.31 | 0.05 | 0.05 | 0.03 | 1.17 [1.10-1.23]* | 1.10 [1.04-1.16]* |
| J94 | 7,380 | 51,021,310 | 0.17 | 0.14 | 0.14 | 0.03 | 0.03 | 0.02 | 1.20 [1.11-1.30]* | 1.12 [1.03-1.22]* |
| J95 | 5,680 | 50,992,850 | 0.13 | 0.11 | 0.11 | 0.02 | 0.03 | 0.02 | 1.24 [1.13-1.35]* | 1.16 [1.06-1.27]* |
| J96 | 128,820 | 50,629,500 | 3.22 | 2.40 | 2.48 | 0.83 | 0.75 | 0.43 | 1.30 [1.28-1.33]* | 1.16 [1.13-1.18]* |
| J98 | 114,890 | 50,436,960 | 2.76 | 2.17 | 2.25 | 0.59 | 0.52 | 0.28 | 1.23 [1.21-1.26]* | 1.11 [1.09-1.13]* |
| J99 | 3,400 | 51,035,020 | 0.10 | 0.06 | 0.06 | 0.04 | 0.04 | 0.03 | 1.71 [1.54-1.91]* | 1.47 [1.32-1.64]* |
| 40+ - K00-K95 | | | | | | | | | | |
| K00 | 1,340 | 51,009,730 | 0.03 | 0.03 | 0.03 | 0.01 | 0.01 | 0.00 | 1.22 [1.02-1.47] | 1.12 [0.93-1.35] |
| K01 | 7,610 | 50,446,540 | 0.17 | 0.15 | 0.15 | 0.02 | 0.02 | 0.02 | 1.15 [1.06-1.24]* | 1.10 [1.02-1.19] |
| K02 | 38,990 | 50,050,970 | 0.96 | 0.74 | 0.75 | 0.22 | 0.21 | 0.12 | 1.28 [1.24-1.32]* | 1.15 [1.11-1.19]* |
| K03 | 1,160 | 51,035,740 | 0.03 | 0.02 | 0.02 | 0.01 | 0.01 | 0.01 | 1.42 [1.18-1.72]* | 1.27 [1.05-1.54] |
| K04 | 16,390 | 50,623,350 | 0.39 | 0.31 | 0.31 | 0.08 | 0.08 | 0.04 | 1.25 [1.19-1.32]* | 1.13 [1.07-1.19]* |
| K05 | 9,760 | 50,756,730 | 0.24 | 0.18 | 0.18 | 0.06 | 0.06 | 0.04 | 1.33 [1.24-1.42]* | 1.21 [1.13-1.29]* |
| K06 | 3,220 | 51,011,860 | 0.08 | 0.06 | 0.06 | 0.02 | 0.02 | 0.02 | 1.37 [1.22-1.54]* | 1.25 [1.11-1.40]* |
| K07 | 4,320 | 50,910,920 | 0.11 | 0.08 | 0.08 | 0.03 | 0.02 | 0.01 | 1.29 [1.17-1.42]* | 1.14 [1.03-1.27] |
| K08 | 26,860 | 50,500,010 | 0.65 | 0.51 | 0.51 | 0.15 | 0.15 | 0.09 | 1.29 [1.24-1.34]* | 1.15 [1.11-1.20]* |
| K09 | 2,490 | 50,993,720 | 0.05 | 0.05 | 0.05 | 0.01 | 0.01 | 0.00 | 1.13 [0.99-1.30] | 1.08 [0.94-1.24] |
| K10 | 4,020 | 51,008,940 | 0.10 | 0.07 | 0.08 | 0.02 | 0.02 | 0.01 | 1.27 [1.15-1.41]* | 1.12 [1.01-1.25] |
| K11 | 8,450 | 50,900,390 | 0.21 | 0.16 | 0.16 | 0.06 | 0.06 | 0.04 | 1.36 [1.27-1.46]* | 1.23 [1.15-1.33]* |
| K12 | 12,360 | 50,947,860 | 0.31 | 0.23 | 0.23 | 0.09 | 0.08 | 0.06 | 1.35 [1.28-1.44]* | 1.25 [1.18-1.33]* |
| K13 | 18,320 | 50,718,530 | 0.47 | 0.34 | 0.34 | 0.13 | 0.13 | 0.10 | 1.38 [1.32-1.45]* | 1.26 [1.20-1.32]* |
| K14 | 10,390 | 50,902,160 | 0.26 | 0.19 | 0.19 | 0.07 | 0.07 | 0.05 | 1.37 [1.29-1.46]* | 1.23 [1.15-1.31]* |
| K20 | 72,930 | 49,690,260 | 1.83 | 1.39 | 1.40 | 0.44 | 0.43 | 0.26 | 1.30 [1.27-1.34]* | 1.16 [1.13-1.19]* |
| K21 | 270,840 | 47,612,590 | 7.35 | 5.33 | 5.42 | 2.02 | 1.93 | 1.09 | 1.36 [1.34-1.37]* | 1.17 [1.16-1.19]* |
| K22 | 110,630 | 49,543,140 | 2.75 | 2.12 | 2.13 | 0.63 | 0.62 | 0.41 | 1.29 [1.26-1.32]* | 1.17 [1.15-1.20]* |
| K23 | 1,340 | 51,052,370 | 0.04 | 0.02 | 0.02 | 0.01 | 0.01 | 0.01 | 1.49 [1.24-1.78]* | 1.27 [1.05-1.52] |
| K25 | 54,880 | 50,258,530 | 1.35 | 1.04 | 1.05 | 0.32 | 0.30 | 0.19 | 1.29 [1.25-1.32]* | 1.17 [1.13-1.20]* |
| K26 | 39,990 | 50,464,380 | 0.92 | 0.76 | 0.77 | 0.16 | 0.15 | 0.10 | 1.20 [1.16-1.24]* | 1.13 [1.09-1.17]* |
| K27 | 6,650 | 50,967,060 | 0.16 | 0.12 | 0.12 | 0.04 | 0.04 | 0.02 | 1.29 [1.19-1.40]* | 1.15 [1.05-1.25]* |
| K28 | 870 | 51,051,410 | 0.02 | 0.02 | 0.02 | 0.01 | 0.01 | 0.00 | 1.38 [1.11-1.72]* | 1.07 [0.84-1.35] |
| K29 | 261,190 | 46,666,980 | 7.11 | 5.27 | 5.35 | 1.84 | 1.76 | 1.12 | 1.33 [1.31-1.35]* | 1.19 [1.17-1.20]* |
| K30 | 62,230 | 49,366,650 | 1.61 | 1.19 | 1.18 | 0.42 | 0.42 | 0.27 | 1.36 [1.32-1.39]* | 1.21 [1.17-1.24]* |
| K31 | 95,600 | 50,087,860 | 2.39 | 1.81 | 1.84 | 0.58 | 0.55 | 0.30 | 1.30 [1.27-1.33]* | 1.14 [1.12-1.17]* |
| K35 | 18,130 | 50,545,970 | 0.37 | 0.36 | 0.36 | 0.02 | 0.02 | 0.00 | 1.05 [1.00-1.11] | 1.01 [0.95-1.06] |
| K37 | 2,660 | 50,979,500 | 0.06 | 0.05 | 0.05 | 0.01 | 0.01 | 0.00 | 1.15 [1.01-1.32] | 1.07 [0.93-1.23] |
| K38 | 3,040 | 51,005,450 | 0.06 | 0.06 | 0.06 | 0.01 | 0.01 | 0.00 | 1.10 [0.97-1.25] | 1.05 [0.92-1.19] |
| K40 | 99,720 | 48,919,820 | 2.24 | 1.99 | 2.02 | 0.25 | 0.22 | 0.15 | 1.11 [1.08-1.13]* | 1.07 [1.05-1.10]* |
| K41 | 7,810 | 50,932,750 | 0.17 | 0.15 | 0.15 | 0.02 | 0.02 | 0.02 | 1.16 [1.08-1.26]* | 1.10 [1.02-1.19] |
| K42 | 41,090 | 50,347,830 | 1.00 | 0.78 | 0.78 | 0.23 | 0.23 | 0.12 | 1.29 [1.25-1.34]* | 1.14 [1.10-1.17]* |
| K43 | 35,500 | 50,475,010 | 0.86 | 0.67 | 0.67 | 0.19 | 0.19 | 0.10 | 1.28 [1.24-1.33]* | 1.14 [1.10-1.18]* |
| K44 | 272,460 | 46,830,490 | 7.27 | 5.50 | 5.59 | 1.77 | 1.69 | 1.02 | 1.30 [1.29-1.32]* | 1.16 [1.15-1.18]* |
| K45 | 1,750 | 51,040,260 | 0.04 | 0.03 | 0.03 | 0.01 | 0.01 | 0.00 | 1.20 [1.02-1.41] | 1.07 [0.90-1.26] |
| K46 | 7,280 | 51,007,690 | 0.18 | 0.13 | 0.13 | 0.05 | 0.05 | 0.03 | 1.35 [1.25-1.46]* | 1.17 [1.08-1.27]* |
| K50 | 13,400 | 50,711,460 | 0.37 | 0.24 | 0.24 | 0.13 | 0.13 | 0.08 | 1.56 [1.47-1.64]* | 1.29 [1.22-1.36]* |
| K51 | 22,750 | 50,516,360 | 0.60 | 0.42 | 0.42 | 0.19 | 0.19 | 0.12 | 1.45 [1.39-1.51]* | 1.24 [1.19-1.30]* |
| K52 | 129,920 | 48,538,230 | 3.41 | 2.52 | 2.50 | 0.89 | 0.91 | 0.63 | 1.36 [1.34-1.39]* | 1.23 [1.21-1.25]* |
| K55 | 24,920 | 50,889,930 | 0.60 | 0.47 | 0.48 | 0.13 | 0.12 | 0.07 | 1.25 [1.20-1.31]* | 1.14 [1.09-1.19]* |
| K56 | 71,360 | 50,423,700 | 1.66 | 1.36 | 1.39 | 0.30 | 0.28 | 0.16 | 1.20 [1.17-1.23]* | 1.11 [1.08-1.14]* |
| K57 | 332,810 | 47,286,730 | 8.57 | 6.71 | 6.86 | 1.86 | 1.71 | 1.03 | 1.25 [1.23-1.26]* | 1.14 [1.12-1.15]* |
| K58 | 65,400 | 50,128,800 | 1.84 | 1.19 | 1.21 | 0.65 | 0.64 | 0.30 | 1.53 [1.49-1.56]* | 1.20 [1.17-1.23]* |
| K59 | 263,370 | 48,994,650 | 6.73 | 5.08 | 5.29 | 1.65 | 1.45 | 0.93 | 1.27 [1.26-1.29]* | 1.16 [1.14-1.18]* |
| K60 | 17,290 | 50,499,480 | 0.48 | 0.31 | 0.31 | 0.16 | 0.16 | 0.13 | 1.53 [1.46-1.60]* | 1.37 [1.30-1.43]* |
| K61 | 9,900 | 50,748,290 | 0.25 | 0.18 | 0.18 | 0.07 | 0.07 | 0.05 | 1.39 [1.31-1.49]* | 1.25 [1.17-1.34]* |
| K62 | 149,260 | 48,028,240 | 3.85 | 2.95 | 2.94 | 0.90 | 0.91 | 0.61 | 1.31 [1.29-1.33]* | 1.19 [1.17-1.21]* |
| K63 | 164,640 | 49,385,930 | 4.01 | 3.19 | 3.21 | 0.82 | 0.80 | 0.45 | 1.25 [1.23-1.27]* | 1.13 [1.11-1.15]* |
| K64 | 109,720 | 50,625,920 | 2.72 | 2.05 | 2.11 | 0.67 | 0.61 | 0.38 | 1.29 [1.27-1.32]* | 1.16 [1.14-1.19]* |
| K65 | 16,840 | 50,926,190 | 0.38 | 0.32 | 0.32 | 0.06 | 0.05 | 0.03 | 1.17 [1.11-1.23]* | 1.10 [1.04-1.16]* |
| K66 | 51,430 | 50,444,680 | 1.17 | 0.99 | 1.00 | 0.19 | 0.18 | 0.08 | 1.18 [1.14-1.21]* | 1.07 [1.04-1.11]* |
| K70 | 20,430 | 50,823,510 | 0.54 | 0.37 | 0.37 | 0.16 | 0.17 | 0.12 | 1.46 [1.40-1.53]* | 1.28 [1.23-1.34]* |
| K71 | 1,100 | 51,049,810 | 0.03 | 0.02 | 0.02 | 0.01 | 0.01 | 0.01 | 1.52 [1.25-1.85]* | 1.39 [1.14-1.70]* |
| K72 | 12,850 | 51,016,820 | 0.33 | 0.24 | 0.24 | 0.09 | 0.09 | 0.07 | 1.40 [1.32-1.48]* | 1.25 [1.18-1.33]* |
| K73 | 1,150 | 51,036,220 | 0.03 | 0.02 | 0.02 | 0.01 | 0.01 | 0.01 | 1.51 [1.25-1.82]* | 1.36 [1.12-1.64]* |
| K74 | 19,570 | 50,915,160 | 0.53 | 0.35 | 0.36 | 0.18 | 0.17 | 0.12 | 1.49 [1.42-1.56]* | 1.30 [1.24-1.37]* |
| K75 | 12,200 | 50,952,900 | 0.32 | 0.22 | 0.23 | 0.10 | 0.09 | 0.06 | 1.41 [1.33-1.49]* | 1.25 [1.18-1.33]* |
| K76 | 90,370 | 50,433,750 | 2.28 | 1.69 | 1.71 | 0.60 | 0.57 | 0.32 | 1.33 [1.30-1.36]* | 1.16 [1.14-1.19]* |
| K80 | 158,440 | 48,531,720 | 3.73 | 3.16 | 3.23 | 0.56 | 0.50 | 0.23 | 1.15 [1.13-1.17]* | 1.07 [1.05-1.08]* |
| K81 | 29,360 | 50,567,190 | 0.68 | 0.56 | 0.57 | 0.12 | 0.11 | 0.05 | 1.19 [1.14-1.24]* | 1.09 [1.04-1.13]* |
| K82 | 21,120 | 50,758,780 | 0.48 | 0.40 | 0.41 | 0.08 | 0.07 | 0.04 | 1.18 [1.13-1.24]* | 1.08 [1.03-1.14]* |
| K83 | 44,220 | 50,685,040 | 1.01 | 0.84 | 0.87 | 0.17 | 0.14 | 0.07 | 1.16 [1.12-1.20]* | 1.07 [1.04-1.11]* |
| K85 | 27,620 | 50,650,630 | 0.63 | 0.53 | 0.53 | 0.11 | 0.10 | 0.05 | 1.19 [1.14-1.24]* | 1.08 [1.04-1.13]* |
| K86 | 25,230 | 50,848,970 | 0.60 | 0.47 | 0.49 | 0.12 | 0.11 | 0.06 | 1.23 [1.18-1.29]* | 1.11 [1.06-1.16]* |
| K90 | 14,920 | 50,814,430 | 0.43 | 0.26 | 0.27 | 0.16 | 0.16 | 0.12 | 1.61 [1.53-1.70]* | 1.37 [1.30-1.44]* |
| K91 | 21,360 | 50,800,860 | 0.51 | 0.40 | 0.40 | 0.11 | 0.11 | 0.07 | 1.28 [1.22-1.34]* | 1.16 [1.11-1.22]* |
| K92 | 186,280 | 49,300,040 | 4.66 | 3.59 | 3.65 | 1.07 | 1.01 | 0.65 | 1.28 [1.26-1.30]* | 1.16 [1.14-1.18]* |
| 40+ - L00-L99 | | | | | | | | | | |

| | | | | | | | | | | |
|---|---|---|---|---|---|---|---|---|---|---|
| L01 | 780 | 51,051,490 | 0.03 | 0.01 | 0.01 | 0.01 | 0.02 | 0.01 | 2.27 [1.83-2.82]* | 1.96 [1.57-2.46]* |
| L02 | 26,710 | 50,491,250 | 0.68 | 0.50 | 0.50 | 0.19 | 0.19 | 0.13 | 1.37 [1.32-1.43]* | 1.23 [1.18-1.28]* |
| L03 | 145,920 | 49,405,490 | 4.36 | 2.65 | 2.66 | 1.70 | 1.69 | 1.44 | 1.64 [1.61-1.66]* | 1.49 [1.47-1.52]* |
| L04 | 850 | 51,043,620 | 0.02 | 0.02 | 0.02 | 0.01 | 0.01 | 0.01 | 1.37 [1.09-1.71]* | 1.32 [1.05-1.66] |
| L05 | 2,570 | 50,911,590 | 0.06 | 0.05 | 0.05 | 0.01 | 0.01 | 0.01 | 1.30 [1.15-1.48]* | 1.17 [1.03-1.34] |
| L08 | 38,510 | 50,758,350 | 1.06 | 0.69 | 0.71 | 0.37 | 0.36 | 0.29 | 1.51 [1.46-1.56]* | 1.37 [1.32-1.42]* |
| L10 | 510 | 51,056,770 | 0.02 | 0.01 | 0.01 | 0.01 | 0.01 | 0.01 | 2.27 [1.74-2.97]* | 2.01 [1.53-2.66]* |
| L11 | 440 | 51,056,040 | 0.01 | 0.01 | 0.01 | 0.01 | 0.01 | 0.01 | 1.81 [1.33-2.46]* | 1.77 [1.29-2.42]* |
| L12 | 3,750 | 51,039,440 | 0.15 | 0.06 | 0.06 | 0.10 | 0.10 | 0.10 | 2.80 [2.54-3.10]* | 2.63 [2.37-2.91]* |
| L13 | 540 | 51,053,390 | 0.02 | 0.01 | 0.01 | 0.02 | 0.02 | 0.02 | 3.00 [2.34-3.86]* | 2.70 [2.08-3.49]* |
| L20 | 2,030 | 51,021,680 | 0.20 | 0.01 | 0.01 | 0.19 | 0.19 | 0.19 | 35.83 [29.13-44.08]* | 31.58 [25.62-38.94]* |
| L21 | 1,520 | 51,045,840 | 0.07 | 0.02 | 0.02 | 0.05 | 0.05 | 0.05 | 3.22 [2.77-3.75]* | 2.91 [2.49-3.40]* |
| L22 | 800 | 51,060,420 | 0.02 | 0.02 | 0.02 | 0.00 | 0.00 | 0.00 | 1.00 [0.77-1.31] | 0.96 [0.73-1.25] |
| L23 | 1,140 | 51,037,130 | 0.05 | 0.02 | 0.01 | 0.04 | 0.04 | 0.04 | 3.72 [3.14-4.40]* | 3.39 [2.86-4.03]* |
| L24 | 10,430 | 51,045,380 | 0.28 | 0.19 | 0.20 | 0.09 | 0.08 | 0.06 | 1.41 [1.32-1.51]* | 1.29 [1.20-1.39]* |
| L25 | 1,380 | 51,043,700 | 0.06 | 0.02 | 0.02 | 0.04 | 0.04 | 0.04 | 3.33 [2.85-3.90]* | 3.10 [2.64-3.64]* |
| L27 | 7,700 | 50,977,330 | 0.22 | 0.14 | 0.14 | 0.09 | 0.08 | 0.07 | 1.60 [1.49-1.72]* | 1.46 [1.36-1.58]* |
| L28 | 3,020 | 51,013,060 | 0.15 | 0.04 | 0.04 | 0.12 | 0.12 | 0.11 | 3.99 [3.59-4.42]* | 3.61 [3.25-4.01]* |
| L29 | 9,110 | 50,891,530 | 0.31 | 0.15 | 0.15 | 0.16 | 0.15 | 0.14 | 1.99 [1.87-2.12]* | 1.78 [1.67-1.90]* |
| L30 | 42,300 | 50,387,880 | 3.15 | 0.35 | 0.33 | 2.80 | 2.81 | 2.78 | 9.44 [9.16-9.73]* | 8.61 [8.35-8.89]* |
| L40 | 34,620 | 50,650,630 | 1.25 | 0.56 | 0.57 | 0.69 | 0.68 | 0.63 | 2.21 [2.14-2.28]* | 2.03 [1.97-2.10]* |
| L42 | 410 | 51,057,560 | 0.01 | 0.01 | 0.01 | 0.01 | 0.01 | 0.01 | 1.79 [1.32-2.43]* | 1.61 [1.18-2.22]* |
| L43 | 4,180 | 50,990,470 | 0.15 | 0.07 | 0.07 | 0.08 | 0.08 | 0.07 | 2.15 [1.96-2.36]* | 1.99 [1.81-2.18]* |
| L50 | 4,600 | 50,989,330 | 0.15 | 0.08 | 0.08 | 0.07 | 0.07 | 0.05 | 1.86 [1.70-2.03]* | 1.50 [1.37-1.65]* |
| L51 | 980 | 51,043,170 | 0.03 | 0.02 | 0.02 | 0.02 | 0.02 | 0.01 | 2.03 [1.67-2.46]* | 1.77 [1.45-2.16]* |
| L52 | 510 | 51,048,170 | 0.01 | 0.01 | 0.01 | 0.01 | 0.00 | 0.00 | 1.49 [1.12-1.96]* | 1.28 [0.96-1.71] |
| L53 | 11,930 | 50,947,250 | 0.37 | 0.20 | 0.21 | 0.17 | 0.17 | 0.15 | 1.82 [1.72-1.93]* | 1.66 [1.57-1.77]* |
| L56 | 470 | 51,054,690 | 0.02 | 0.01 | 0.01 | 0.01 | 0.01 | 0.01 | 2.99 [2.29-3.90]* | 2.67 [2.03-3.51]* |
| L57 | 34,150 | 50,677,710 | 0.86 | 0.63 | 0.65 | 0.22 | 0.21 | 0.16 | 1.32 [1.28-1.37]* | 1.24 [1.19-1.28]* |
| L59 | 440 | 51,056,810 | 0.01 | 0.01 | 0.01 | 0.00 | 0.00 | 0.00 | 1.40 [1.03-1.91] | 1.40 [1.01-1.92] |
| L60 | 6,410 | 50,847,190 | 0.17 | 0.12 | 0.12 | 0.06 | 0.06 | 0.04 | 1.51 [1.39-1.63]* | 1.32 [1.22-1.44]* |
| L63 | 400 | 51,056,070 | 0.02 | 0.01 | 0.01 | 0.01 | 0.01 | 0.01 | 2.62 [1.97-3.48]* | 2.06 [1.53-2.79]* |
| L65 | 1,440 | 51,044,600 | 0.05 | 0.02 | 0.02 | 0.03 | 0.03 | 0.02 | 2.09 [1.78-2.45]* | 1.77 [1.50-2.08]* |
| L66 | 360 | 51,056,530 | 0.01 | 0.01 | 0.01 | 0.01 | 0.01 | 0.00 | 1.82 [1.32-2.51]* | 1.64 [1.18-2.28]* |
| L70 | 710 | 51,048,370 | 0.02 | 0.01 | 0.01 | 0.01 | 0.01 | 0.00 | 1.44 [1.13-1.83]* | 1.24 [0.97-1.60] |
| L71 | 3,220 | 51,027,650 | 0.09 | 0.06 | 0.06 | 0.03 | 0.03 | 0.03 | 1.52 [1.36-1.70]* | 1.40 [1.25-1.57]* |
| L72 | 35,780 | 49,839,570 | 0.81 | 0.70 | 0.69 | 0.11 | 0.12 | 0.07 | 1.17 [1.13-1.21]* | 1.09 [1.06-1.14]* |
| L73 | 3,380 | 50,965,490 | 0.09 | 0.06 | 0.06 | 0.03 | 0.03 | 0.03 | 1.58 [1.42-1.75]* | 1.38 [1.24-1.55]* |
| L80 | 1,170 | 51,048,810 | 0.04 | 0.02 | 0.02 | 0.02 | 0.02 | 0.02 | 1.90 [1.59-2.27]* | 1.67 [1.39-2.01]* |
| L81 | 6,210 | 50,953,950 | 0.15 | 0.12 | 0.12 | 0.03 | 0.03 | 0.02 | 1.27 [1.17-1.38]* | 1.19 [1.09-1.30]* |
| L82 | 22,160 | 50,689,060 | 0.53 | 0.42 | 0.42 | 0.11 | 0.11 | 0.08 | 1.26 [1.20-1.31]* | 1.17 [1.12-1.23]* |
| L83 | 1,040 | 51,044,820 | 0.03 | 0.02 | 0.02 | 0.01 | 0.01 | 0.01 | 1.71 [1.41-2.08]* | 1.56 [1.27-1.90]* |
| L84 | 2,260 | 51,022,610 | 0.06 | 0.04 | 0.04 | 0.02 | 0.02 | 0.02 | 1.50 [1.31-1.72]* | 1.35 [1.18-1.55]* |
| L85 | 9,140 | 50,948,720 | 0.25 | 0.16 | 0.16 | 0.09 | 0.09 | 0.08 | 1.55 [1.45-1.66]* | 1.45 [1.35-1.55]* |
| L88 | 450 | 51,056,190 | 0.01 | 0.01 | 0.01 | 0.01 | 0.01 | 0.00 | 1.68 [1.25-2.26]* | 1.45 [1.07-1.97] |
| L89 | 119,510 | 50,772,560 | 2.88 | 2.24 | 2.37 | 0.64 | 0.51 | 0.38 | 1.22 [1.19-1.24]* | 1.15 [1.13-1.17]* |
| L90 | 17,690 | 50,642,140 | 0.44 | 0.33 | 0.33 | 0.11 | 0.11 | 0.08 | 1.32 [1.25-1.38]* | 1.21 [1.15-1.28]* |
| L91 | 4,370 | 50,903,250 | 0.11 | 0.08 | 0.08 | 0.03 | 0.03 | 0.02 | 1.36 [1.23-1.50]* | 1.25 [1.13-1.38]* |
| L92 | 4,230 | 50,964,650 | 0.11 | 0.08 | 0.08 | 0.03 | 0.03 | 0.02 | 1.37 [1.24-1.52]* | 1.24 [1.12-1.38]* |
| L93 | 2,200 | 51,028,700 | 0.07 | 0.04 | 0.04 | 0.03 | 0.03 | 0.03 | 1.84 [1.62-2.10]* | 1.58 [1.38-1.81]* |
| L94 | 1,340 | 51,039,790 | 0.04 | 0.02 | 0.02 | 0.02 | 0.02 | 0.02 | 1.81 [1.53-2.14]* | 1.60 [1.34-1.90]* |
| L95 | 1,910 | 51,038,980 | 0.06 | 0.03 | 0.03 | 0.03 | 0.02 | 0.02 | 1.70 [1.47-1.96]* | 1.53 [1.32-1.77]* |
| L97 | 85,290 | 50,567,910 | 2.39 | 1.54 | 1.56 | 0.85 | 0.83 | 0.70 | 1.53 [1.50-1.57]* | 1.42 [1.38-1.45]* |
| L98 | 61,110 | 50,117,810 | 1.59 | 1.14 | 1.14 | 0.45 | 0.45 | 0.36 | 1.39 [1.35-1.43]* | 1.29 [1.25-1.33]* |
| 40+ - M00-M99 | | | | | | | | | | |
| M00 | 7,070 | 50,975,720 | 0.19 | 0.13 | 0.13 | 0.07 | 0.07 | 0.06 | 1.54 [1.42-1.66]* | 1.41 [1.30-1.52]* |
| M02 | 840 | 51,047,870 | 0.02 | 0.02 | 0.02 | 0.01 | 0.01 | 0.00 | 1.34 [1.07-1.67] | 1.23 [0.97-1.54] |
| M05 | 9,650 | 50,935,100 | 0.25 | 0.18 | 0.18 | 0.07 | 0.07 | 0.06 | 1.37 [1.28-1.47]* | 1.29 [1.21-1.38]* |
| M06 | 70,210 | 50,160,110 | 1.84 | 1.31 | 1.33 | 0.54 | 0.52 | 0.38 | 1.39 [1.36-1.42]* | 1.26 [1.23-1.29]* |
| M07 | 7,400 | 50,958,680 | 0.22 | 0.13 | 0.13 | 0.09 | 0.09 | 0.07 | 1.70 [1.59-1.83]* | 1.50 [1.40-1.62]* |
| M10 | 83,120 | 50,446,500 | 2.12 | 1.55 | 1.56 | 0.58 | 0.56 | 0.39 | 1.36 [1.33-1.39]* | 1.23 [1.20-1.26]* |
| M11 | 9,460 | 50,978,620 | 0.23 | 0.18 | 0.18 | 0.06 | 0.05 | 0.03 | 1.29 [1.20-1.38]* | 1.17 [1.09-1.26]* |
| M12 | 1,530 | 51,037,560 | 0.04 | 0.03 | 0.03 | 0.01 | 0.01 | 0.01 | 1.36 [1.16-1.61]* | 1.16 [0.98-1.38] |
| M13 | 207,040 | 49,017,270 | 5.44 | 3.96 | 4.08 | 1.47 | 1.36 | 0.87 | 1.33 [1.31-1.35]* | 1.19 [1.17-1.21]* |
| M14 | 2,040 | 51,038,340 | 0.06 | 0.04 | 0.04 | 0.02 | 0.02 | 0.01 | 1.62 [1.41-1.87]* | 1.31 [1.14-1.52]* |
| M15 | 77,580 | 50,329,230 | 2.02 | 1.44 | 1.48 | 0.57 | 0.54 | 0.35 | 1.36 [1.33-1.40]* | 1.21 [1.18-1.24]* |
| M16 | 128,650 | 49,182,620 | 2.92 | 2.55 | 2.62 | 0.36 | 0.30 | 0.16 | 1.11 [1.09-1.14]* | 1.06 [1.04-1.08]* |
| M17 | 204,510 | 47,786,480 | 5.19 | 4.08 | 4.14 | 1.10 | 1.04 | 0.69 | 1.25 [1.23-1.27]* | 1.15 [1.13-1.17]* |
| M18 | 11,340 | 50,909,240 | 0.30 | 0.21 | 0.21 | 0.09 | 0.09 | 0.06 | 1.44 [1.35-1.53]* | 1.24 [1.17-1.32]* |
| M19 | 324,110 | 48,247,880 | 8.79 | 6.28 | 6.49 | 2.51 | 2.30 | 1.49 | 1.35 [1.34-1.37]* | 1.20 [1.19-1.22]* |
| M20 | 41,130 | 50,058,940 | 1.02 | 0.78 | 0.78 | 0.24 | 0.24 | 0.17 | 1.31 [1.26-1.35]* | 1.21 [1.17-1.25]* |
| M21 | 22,370 | 50,786,790 | 0.53 | 0.42 | 0.43 | 0.11 | 0.10 | 0.06 | 1.23 [1.18-1.29]* | 1.13 [1.08-1.19]* |
| M22 | 4,760 | 50,877,800 | 0.12 | 0.09 | 0.09 | 0.03 | 0.03 | 0.02 | 1.33 [1.21-1.46]* | 1.18 [1.07-1.29]* |
| M23 | 68,540 | 49,075,880 | 1.63 | 1.35 | 1.32 | 0.29 | 0.31 | 0.19 | 1.23 [1.20-1.27]* | 1.13 [1.10-1.16]* |
| M24 | 22,110 | 50,648,670 | 0.54 | 0.41 | 0.42 | 0.13 | 0.12 | 0.07 | 1.29 [1.23-1.35]* | 1.16 [1.11-1.21]* |
| M25 | 177,440 | 48,655,010 | 4.55 | 3.45 | 3.54 | 1.10 | 1.01 | 0.61 | 1.29 [1.26-1.31]* | 1.15 [1.14-1.17]* |
| M30 | 810 | 51,050,740 | 0.02 | 0.01 | 0.02 | 0.01 | 0.01 | 0.00 | 1.42 [1.13-1.78]* | 1.18 [0.93-1.50] |
| M31 | 11,460 | 50,945,160 | 0.29 | 0.21 | 0.22 | 0.08 | 0.07 | 0.05 | 1.33 [1.25-1.42]* | 1.22 [1.15-1.30]* |
| M32 | 4,110 | 50,980,650 | 0.12 | 0.07 | 0.07 | 0.05 | 0.05 | 0.04 | 1.69 [1.54-1.86]* | 1.48 [1.34-1.63]* |
| M33 | 1,020 | 51,047,000 | 0.03 | 0.02 | 0.02 | 0.02 | 0.02 | 0.02 | 2.02 [1.67-2.43]* | 1.92 [1.58-2.32]* |
| M34 | 2,030 | 51,033,040 | 0.06 | 0.04 | 0.04 | 0.02 | 0.02 | 0.02 | 1.65 [1.44-1.90]* | 1.49 [1.29-1.72]* |
| M35 | 49,890 | 50,609,480 | 1.33 | 0.91 | 0.93 | 0.42 | 0.40 | 0.31 | 1.43 [1.39-1.47]* | 1.30 [1.26-1.34]* |
| M40 | 12,660 | 50,999,830 | 0.31 | 0.24 | 0.25 | 0.07 | 0.06 | 0.04 | 1.24 [1.17-1.32]* | 1.15 [1.08-1.22]* |
| M41 | 29,740 | 50,846,100 | 0.73 | 0.55 | 0.58 | 0.18 | 0.15 | 0.10 | 1.27 [1.22-1.32]* | 1.16 [1.11-1.21]* |
| M43 | 27,460 | 50,767,230 | 0.69 | 0.51 | 0.53 | 0.18 | 0.16 | 0.09 | 1.30 [1.25-1.35]* | 1.15 [1.10-1.19]* |
| M45 | 6,330 | 50,964,930 | 0.17 | 0.11 | 0.12 | 0.06 | 0.06 | 0.04 | 1.48 [1.37-1.61]* | 1.31 [1.20-1.42]* |
| M46 | 23,210 | 50,824,720 | 0.61 | 0.42 | 0.43 | 0.18 | 0.18 | 0.11 | 1.41 [1.35-1.47]* | 1.21 [1.16-1.27]* |
| M47 | 135,350 | 49,647,300 | 3.53 | 2.55 | 2.62 | 0.98 | 0.91 | 0.53 | 1.35 [1.32-1.37]* | 1.18 [1.16-1.20]* |
| M48 | 79,900 | 50,326,640 | 1.99 | 1.50 | 1.55 | 0.49 | 0.45 | 0.24 | 1.29 [1.26-1.32]* | 1.14 [1.11-1.16]* |
| M49 | 2,780 | 51,049,910 | 0.06 | 0.05 | 0.05 | 0.01 | 0.01 | 0.01 | 1.16 [1.01-1.32] | 1.12 [0.98-1.28] |
| M50 | 20,050 | 50,820,660 | 0.50 | 0.37 | 0.37 | 0.12 | 0.12 | 0.07 | 1.33 [1.27-1.39]* | 1.15 [1.10-1.21]* |
| M51 | 88,370 | 49,743,600 | 2.23 | 1.68 | 1.69 | 0.55 | 0.53 | 0.28 | 1.32 [1.29-1.34]* | 1.14 [1.12-1.17]* |
| M53 | 5,000 | 50,973,140 | 0.12 | 0.09 | 0.09 | 0.03 | 0.03 | 0.01 | 1.30 [1.18-1.42]* | 1.09 [0.99-1.21] |
| M54 | 207,120 | 47,902,330 | 5.44 | 4.08 | 4.18 | 1.35 | 1.26 | 0.66 | 1.30 [1.28-1.32]* | 1.14 [1.12-1.16]* |
| M60 | 3,580 | 51,016,780 | 0.10 | 0.06 | 0.07 | 0.03 | 0.03 | 0.03 | 1.48 [1.33-1.65]* | 1.38 [1.24-1.54]* |
| M62 | 18,120 | 50,894,960 | 0.43 | 0.34 | 0.35 | 0.09 | 0.09 | 0.06 | 1.25 [1.19-1.32]* | 1.16 [1.10-1.23]* |
| M65 | 36,350 | 50,352,890 | 0.92 | 0.68 | 0.68 | 0.24 | 0.24 | 0.15 | 1.35 [1.30-1.39]* | 1.20 [1.16-1.24]* |
| M66 | 4,190 | 50,986,080 | 0.10 | 0.08 | 0.08 | 0.02 | 0.02 | 0.02 | 1.30 [1.17-1.44]* | 1.18 [1.06-1.31]* |
| M67 | 19,770 | 50,512,190 | 0.49 | 0.37 | 0.37 | 0.12 | 0.12 | 0.08 | 1.33 [1.27-1.39]* | 1.21 [1.15-1.26]* |
| M70 | 15,780 | 50,798,130 | 0.42 | 0.29 | 0.29 | 0.13 | 0.13 | 0.09 | 1.43 [1.36-1.51]* | 1.26 [1.20-1.33]* |
| M71 | 10,540 | 50,926,780 | 0.26 | 0.20 | 0.20 | 0.07 | 0.06 | 0.04 | 1.32 [1.24-1.41]* | 1.19 [1.11-1.27]* |
| M72 | 24,430 | 50,585,450 | 0.53 | 0.47 | 0.47 | 0.05 | 0.06 | 0.03 | 1.12 [1.07-1.17]* | 1.07 [1.02-1.12]* |
| M75 | 65,480 | 49,832,850 | 1.65 | 1.24 | 1.25 | 0.41 | 0.40 | 0.24 | 1.32 [1.29-1.36]* | 1.17 [1.14-1.20]* |
| M76 | 4,590 | 50,997,300 | 0.12 | 0.08 | 0.09 | 0.04 | 0.03 | 0.02 | 1.41 [1.28-1.55]* | 1.21 [1.10-1.33]* |
| M77 | 12,120 | 50,806,020 | 0.32 | 0.22 | 0.22 | 0.10 | 0.10 | 0.07 | 1.47 [1.39-1.55]* | 1.27 [1.19-1.35]* |
| M79 | 175,120 | 48,561,580 | 4.81 | 3.35 | 3.40 | 1.47 | 1.41 | 0.91 | 1.41 [1.39-1.44]* | 1.23 [1.21-1.25]* |

| | | | | | | | | | | |
|---|---|---|---|---|---|---|---|---|---|---|
| M80 | 25,810 | 50,870,160 | 0.64 | 0.48 | 0.50 | 0.16 | 0.15 | 0.10 | 1.30 [1.24-1.35]* | 1.19 [1.14-1.24]* |
| M81 | 158,100 | 49,894,880 | 4.03 | 2.98 | 3.10 | 1.05 | 0.93 | 0.60 | 1.30 [1.28-1.32]* | 1.18 [1.16-1.20]* |
| M83 | 920 | 51,049,120 | 0.02 | 0.02 | 0.02 | 0.01 | 0.01 | 0.00 | 1.28 [1.02-1.59] | 1.16 [0.92-1.45] |
| M84 | 17,320 | 50,797,050 | 0.42 | 0.32 | 0.33 | 0.09 | 0.09 | 0.06 | 1.26 [1.20-1.33]* | 1.16 [1.10-1.22]* |
| M85 | 32,980 | 50,873,120 | 0.83 | 0.61 | 0.64 | 0.22 | 0.19 | 0.11 | 1.29 [1.25-1.34]* | 1.16 [1.11-1.20]* |
| M86 | 13,640 | 50,931,610 | 0.35 | 0.25 | 0.25 | 0.10 | 0.09 | 0.06 | 1.37 [1.30-1.45]* | 1.23 [1.16-1.30]* |
| M87 | 5,920 | 50,975,430 | 0.14 | 0.11 | 0.11 | 0.03 | 0.03 | 0.02 | 1.28 [1.17-1.39]* | 1.16 [1.06-1.27]* |
| M88 | 4,220 | 50,989,970 | 0.09 | 0.08 | 0.08 | 0.01 | 0.01 | 0.01 | 1.18 [1.05-1.31]* | 1.13 [1.01-1.27] |
| M89 | 13,010 | 50,826,590 | 0.32 | 0.24 | 0.24 | 0.08 | 0.08 | 0.05 | 1.32 [1.24-1.40]* | 1.18 [1.11-1.25]* |
| M90 | 6,970 | 51,041,940 | 0.14 | 0.14 | 0.14 | 0.01 | 0.01 | 0.00 | 1.04 [0.96-1.13] | 1.02 [0.94-1.12] |
| M92 | 920 | 51,043,500 | 0.02 | 0.02 | 0.02 | 0.01 | 0.01 | 0.01 | 1.45 [1.17-1.78]* | 1.27 [1.02-1.58] |
| M93 | 2,060 | 51,012,090 | 0.05 | 0.04 | 0.04 | 0.01 | 0.01 | 0.01 | 1.30 [1.13-1.50]* | 1.15 [0.99-1.33] |
| M94 | 8,210 | 50,921,420 | 0.22 | 0.15 | 0.15 | 0.08 | 0.07 | 0.05 | 1.49 [1.39-1.60]* | 1.27 [1.18-1.36]* |
| M95 | 1,400 | 51,000,180 | 0.03 | 0.03 | 0.03 | 0.01 | 0.01 | 0.00 | 1.31 [1.10-1.56]* | 1.14 [0.95-1.37] |
| M96 | 8,220 | 50,982,080 | 0.21 | 0.15 | 0.15 | 0.06 | 0.05 | 0.04 | 1.35 [1.25-1.46]* | 1.23 [1.14-1.33]* |
| M99 | 2,340 | 51,040,950 | 0.06 | 0.04 | 0.04 | 0.01 | 0.01 | 0.01 | 1.32 [1.15-1.52]* | 1.11 [0.97-1.29] |
| 40+ - N00-N99 | | | | | | | | | | |
| N02 | 5,750 | 50,941,160 | 0.15 | 0.11 | 0.11 | 0.04 | 0.04 | 0.03 | 1.36 [1.25-1.48]* | 1.27 [1.17-1.39]* |
| N03 | 26,580 | 50,855,870 | 0.66 | 0.49 | 0.49 | 0.17 | 0.17 | 0.12 | 1.35 [1.29-1.41]* | 1.22 [1.17-1.28]* |
| N04 | 3,150 | 51,019,170 | 0.08 | 0.06 | 0.06 | 0.03 | 0.03 | 0.02 | 1.47 [1.31-1.65]* | 1.33 [1.19-1.50]* |
| N05 | 3,130 | 51,012,810 | 0.08 | 0.06 | 0.06 | 0.03 | 0.03 | 0.02 | 1.49 [1.33-1.67]* | 1.36 [1.21-1.53]* |
| N08 | 11,340 | 50,966,170 | 0.30 | 0.21 | 0.21 | 0.10 | 0.09 | 0.06 | 1.43 [1.35-1.52]* | 1.22 [1.15-1.31]* |
| N10 | 2,850 | 51,011,270 | 0.07 | 0.05 | 0.05 | 0.01 | 0.01 | 0.01 | 1.21 [1.07-1.37]* | 1.09 [0.96-1.24] |
| N11 | 1,860 | 51,026,130 | 0.04 | 0.03 | 0.03 | 0.01 | 0.01 | 0.01 | 1.29 [1.11-1.51]* | 1.18 [1.01-1.38] |
| N12 | 15,700 | 50,820,600 | 0.38 | 0.29 | 0.30 | 0.09 | 0.08 | 0.04 | 1.28 [1.21-1.35]* | 1.13 [1.07-1.19]* |
| N13 | 57,260 | 50,595,600 | 1.21 | 1.12 | 1.13 | 0.09 | 0.08 | 0.05 | 1.07 [1.04-1.10]* | 1.04 [1.01-1.07] |
| N14 | 860 | 51,056,380 | 0.02 | 0.02 | 0.02 | 0.01 | 0.01 | 0.00 | 1.41 [1.12-1.77]* | 1.26 [1.00-1.59] |
| N15 | 1,330 | 51,047,440 | 0.03 | 0.03 | 0.03 | 0.01 | 0.01 | 0.00 | 1.21 [1.00-1.46] | 1.10 [0.91-1.34] |
| N16 | 630 | 51,054,080 | 0.02 | 0.01 | 0.01 | 0.01 | 0.01 | 0.00 | 1.51 [1.17-1.95]* | 1.34 [1.03-1.75] |
| N17 | 346,860 | 49,910,840 | 8.45 | 6.63 | 6.90 | 1.83 | 1.55 | 1.02 | 1.23 [1.21-1.24]* | 1.14 [1.12-1.15]* |
| N18 | 296,040 | 49,719,170 | 7.36 | 5.66 | 5.87 | 1.70 | 1.49 | 0.93 | 1.25 [1.24-1.27]* | 1.15 [1.13-1.16]* |
| N19 | 42,040 | 50,712,120 | 1.03 | 0.79 | 0.80 | 0.25 | 0.23 | 0.17 | 1.29 [1.25-1.34]* | 1.19 [1.15-1.23]* |
| N20 | 43,790 | 50,267,470 | 0.98 | 0.85 | 0.85 | 0.13 | 0.13 | 0.06 | 1.15 [1.11-1.19]* | 1.07 [1.03-1.10]* |
| N21 | 12,350 | 50,900,450 | 0.25 | 0.24 | 0.24 | 0.01 | 0.01 | 0.00 | 1.03 [0.96-1.10] | 1.00 [0.94-1.07] |
| N23 | 8,060 | 50,742,700 | 0.18 | 0.16 | 0.15 | 0.02 | 0.02 | 0.00 | 1.14 [1.05-1.23]* | 1.01 [0.94-1.09] |
| N25 | 2,230 | 51,037,330 | 0.06 | 0.04 | 0.04 | 0.02 | 0.02 | 0.01 | 1.39 [1.21-1.60]* | 1.28 [1.11-1.48]* |
| N26 | 4,600 | 51,035,350 | 0.11 | 0.09 | 0.09 | 0.02 | 0.02 | 0.01 | 1.18 [1.06-1.31]* | 1.07 [0.96-1.18] |
| N27 | 2,150 | 51,040,720 | 0.06 | 0.04 | 0.04 | 0.02 | 0.01 | 0.01 | 1.37 [1.19-1.58]* | 1.29 [1.11-1.49]* |
| N28 | 67,240 | 50,564,440 | 1.56 | 1.28 | 1.31 | 0.28 | 0.25 | 0.14 | 1.19 [1.16-1.22]* | 1.10 [1.07-1.13]* |
| N30 | 29,730 | 50,551,720 | 0.70 | 0.56 | 0.57 | 0.13 | 0.13 | 0.08 | 1.23 [1.18-1.28]* | 1.14 [1.09-1.18]* |
| N31 | 8,000 | 50,868,690 | 0.21 | 0.15 | 0.15 | 0.06 | 0.06 | 0.04 | 1.40 [1.30-1.51]* | 1.22 [1.13-1.31]* |
| N32 | 90,090 | 49,632,990 | 2.16 | 1.74 | 1.77 | 0.42 | 0.39 | 0.26 | 1.22 [1.19-1.25]* | 1.14 [1.11-1.16]* |
| N34 | 1,160 | 51,032,240 | 0.03 | 0.02 | 0.02 | 0.01 | 0.01 | 0.00 | 1.25 [1.03-1.52] | 1.19 [0.97-1.45] |
| N35 | 28,420 | 50,487,340 | 0.70 | 0.53 | 0.54 | 0.16 | 0.16 | 0.12 | 1.30 [1.25-1.35]* | 1.21 [1.16-1.26]* |
| N36 | 7,810 | 50,920,200 | 0.18 | 0.15 | 0.15 | 0.04 | 0.03 | 0.02 | 1.23 [1.14-1.32]* | 1.15 [1.06-1.24]* |
| N39 | 387,940 | 47,241,490 | 9.84 | 7.86 | 8.20 | 1.99 | 1.64 | 1.06 | 1.20 [1.19-1.21]* | 1.12 [1.11-1.13]* |
| N40 | 143,300 | 49,304,800 | 3.44 | 2.79 | 2.96 | 0.65 | 0.48 | 0.31 | 1.16 [1.14-1.18]* | 1.10 [1.08-1.12]* |
| N41 | 10,300 | 50,881,250 | 0.22 | 0.20 | 0.20 | 0.02 | 0.02 | 0.01 | 1.11 [1.03-1.18]* | 1.05 [0.98-1.12] |
| N42 | 13,500 | 50,853,590 | 0.29 | 0.26 | 0.26 | 0.03 | 0.03 | 0.02 | 1.13 [1.06-1.20]* | 1.08 [1.01-1.14] |
| N43 | 11,260 | 50,842,980 | 0.25 | 0.21 | 0.21 | 0.04 | 0.04 | 0.03 | 1.19 [1.12-1.27]* | 1.14 [1.07-1.22]* |
| N45 | 8,340 | 50,899,600 | 0.20 | 0.16 | 0.16 | 0.04 | 0.04 | 0.03 | 1.25 [1.16-1.35]* | 1.15 [1.07-1.24]* |
| N47 | 19,550 | 50,699,390 | 0.49 | 0.36 | 0.36 | 0.13 | 0.14 | 0.11 | 1.38 [1.32-1.45]* | 1.28 [1.22-1.35]* |
| N48 | 16,280 | 50,760,480 | 0.42 | 0.30 | 0.30 | 0.12 | 0.12 | 0.09 | 1.40 [1.33-1.48]* | 1.29 [1.23-1.36]* |
| N49 | 2,520 | 51,023,580 | 0.06 | 0.05 | 0.05 | 0.02 | 0.02 | 0.01 | 1.41 [1.23-1.60]* | 1.28 [1.12-1.47]* |
| N50 | 17,650 | 50,664,170 | 0.42 | 0.33 | 0.33 | 0.08 | 0.09 | 0.06 | 1.26 [1.20-1.33]* | 1.17 [1.11-1.23]* |
| N60 | 8,450 | 50,778,840 | 0.18 | 0.16 | 0.17 | 0.02 | 0.02 | 0.01 | 1.11 [1.03-1.19]* | 1.05 [0.97-1.13] |
| N61 | 3,970 | 50,941,190 | 0.10 | 0.07 | 0.07 | 0.03 | 0.03 | 0.02 | 1.39 [1.25-1.54]* | 1.25 [1.12-1.39]* |
| N62 | 3,450 | 50,922,020 | 0.08 | 0.06 | 0.06 | 0.02 | 0.02 | 0.01 | 1.27 [1.13-1.42]* | 1.11 [0.99-1.25] |
| N63 | 9,150 | 50,784,980 | 0.21 | 0.17 | 0.18 | 0.03 | 0.03 | 0.01 | 1.16 [1.08-1.25]* | 1.08 [1.00-1.16] |
| N64 | 8,290 | 50,864,260 | 0.19 | 0.16 | 0.16 | 0.04 | 0.03 | 0.02 | 1.21 [1.12-1.30]* | 1.11 [1.03-1.20]* |
| N70 | 4,170 | 50,911,340 | 0.09 | 0.08 | 0.08 | 0.01 | 0.00 | 0.00 | 1.06 [0.95-1.18] | 1.00 [0.89-1.11] |
| N71 | 2,420 | 50,991,040 | 0.06 | 0.05 | 0.05 | 0.01 | 0.01 | 0.01 | 1.25 [1.09-1.42]* | 1.13 [0.99-1.30] |
| N72 | 4,020 | 50,832,040 | 0.09 | 0.08 | 0.08 | 0.01 | 0.01 | 0.01 | 1.12 [1.01-1.24] | 1.07 [0.96-1.19] |
| N73 | 20,330 | 50,301,110 | 0.45 | 0.39 | 0.40 | 0.06 | 0.05 | 0.02 | 1.13 [1.08-1.18]* | 1.04 [0.99-1.09] |
| N75 | 2,780 | 50,917,450 | 0.06 | 0.05 | 0.05 | 0.00 | 0.00 | 0.00 | 1.06 [0.93-1.20] | 1.02 [0.89-1.16] |
| N76 | 5,800 | 50,888,620 | 0.16 | 0.10 | 0.11 | 0.06 | 0.05 | 0.04 | 1.51 [1.39-1.64]* | 1.37 [1.26-1.49]* |
| N77 | 4,580 | 50,961,170 | 0.12 | 0.08 | 0.09 | 0.04 | 0.03 | 0.02 | 1.40 [1.27-1.54]* | 1.19 [1.07-1.31]* |
| N80 | 19,480 | 50,218,740 | 0.42 | 0.38 | 0.39 | 0.04 | 0.03 | 0.00 | 1.09 [1.03-1.14]* | 1.00 [0.95-1.05] |
| N81 | 62,640 | 49,529,080 | 1.48 | 1.22 | 1.24 | 0.26 | 0.24 | 0.15 | 1.20 [1.17-1.23]* | 1.12 [1.09-1.15]* |
| N82 | 2,740 | 51,021,850 | 0.07 | 0.05 | 0.05 | 0.02 | 0.02 | 0.01 | 1.30 [1.15-1.48]* | 1.19 [1.05-1.36]* |
| N83 | 35,460 | 49,962,450 | 0.79 | 0.69 | 0.71 | 0.10 | 0.08 | 0.03 | 1.12 [1.08-1.16]* | 1.04 [1.00-1.07] |
| N84 | 49,890 | 49,695,760 | 1.11 | 0.98 | 0.99 | 0.13 | 0.12 | 0.09 | 1.12 [1.09-1.16]* | 1.09 [1.06-1.13]* |
| N85 | 35,810 | 50,197,380 | 0.82 | 0.69 | 0.70 | 0.14 | 0.12 | 0.08 | 1.18 [1.14-1.22]* | 1.11 [1.07-1.15]* |
| N86 | 3,500 | 50,806,740 | 0.08 | 0.07 | 0.07 | 0.01 | 0.01 | 0.01 | 1.23 [1.10-1.37]* | 1.17 [1.04-1.31]* |
| N87 | 5,150 | 50,558,960 | 0.11 | 0.10 | 0.10 | 0.01 | 0.01 | 0.00 | 1.06 [0.97-1.17] | 1.01 [0.91-1.11] |
| N88 | 10,980 | 50,776,480 | 0.25 | 0.21 | 0.21 | 0.04 | 0.04 | 0.03 | 1.19 [1.12-1.27]* | 1.12 [1.05-1.20]* |
| N89 | 9,340 | 50,706,230 | 0.23 | 0.17 | 0.18 | 0.06 | 0.05 | 0.04 | 1.30 [1.22-1.39]* | 1.21 [1.13-1.29]* |
| N90 | 13,870 | 50,691,310 | 0.37 | 0.25 | 0.26 | 0.12 | 0.11 | 0.09 | 1.43 [1.36-1.51]* | 1.32 [1.25-1.39]* |
| N91 | 640 | 51,025,050 | 0.02 | 0.01 | 0.01 | 0.01 | 0.01 | 0.01 | 1.52 [1.19-1.93]* | 1.41 [1.10-1.81]* |
| N92 | 58,270 | 48,611,360 | 1.39 | 1.16 | 1.18 | 0.24 | 0.21 | 0.11 | 1.18 [1.15-1.21]* | 1.09 [1.06-1.12]* |
| N93 | 20,700 | 50,194,750 | 0.48 | 0.40 | 0.40 | 0.08 | 0.07 | 0.04 | 1.18 [1.13-1.24]* | 1.09 [1.04-1.14]* |
| N94 | 13,520 | 50,229,810 | 0.33 | 0.26 | 0.26 | 0.07 | 0.07 | 0.04 | 1.27 [1.20-1.34]* | 1.14 [1.08-1.21]* |
| N95 | 48,440 | 49,820,420 | 1.18 | 0.93 | 0.95 | 0.25 | 0.23 | 0.17 | 1.24 [1.21-1.28]* | 1.17 [1.14-1.21]* |
| N97 | 740 | 50,640,650 | 0.01 | 0.01 | 0.01 | 0.00 | 0.00 | 0.00 | 0.97 [0.75-1.26] | 0.98 [0.76-1.27] |
| N99 | 9,740 | 50,859,510 | 0.23 | 0.18 | 0.18 | 0.04 | 0.04 | 0.03 | 1.22 [1.14-1.31]* | 1.14 [1.06-1.22]* |
| 40+ - O00-O9A | | | | | | | | | | |
| O00 | 800 | 50,856,230 | 0.02 | 0.02 | 0.02 | 0.00 | 0.00 | 0.00 | 1.03 [0.81-1.31] | 1.02 [0.80-1.30] |
| O02 | 3,470 | 50,399,380 | 0.07 | 0.07 | 0.07 | 0.00 | 0.00 | 0.00 | 1.02 [0.91-1.15] | 1.00 [0.89-1.13] |
| O03 | 5,060 | 50,210,800 | 0.11 | 0.10 | 0.10 | 0.01 | 0.00 | 0.00 | 1.04 [0.95-1.14] | 1.03 [0.94-1.14] |
| O04 | 3,100 | 50,308,600 | 0.06 | 0.06 | 0.06 | 0.00 | −0.01 | −0.01 | 0.92 [0.81-1.04] | 0.90 [0.79-1.03] |
| O10 | 790 | 50,987,040 | 0.02 | 0.01 | 0.01 | 0.00 | 0.00 | 0.00 | 1.27 [1.01-1.59] | 1.06 [0.83-1.34] |
| O13 | 1,690 | 50,552,980 | 0.03 | 0.03 | 0.03 | 0.00 | 0.00 | 0.00 | 0.96 [0.81-1.14] | 0.96 [0.81-1.14] |
| O14 | 1,180 | 50,732,120 | 0.02 | 0.02 | 0.02 | 0.00 | 0.00 | 0.00 | 0.98 [0.80-1.20] | 0.96 [0.78-1.18] |
| O16 | 1,360 | 50,526,200 | 0.03 | 0.03 | 0.03 | 0.00 | 0.00 | 0.00 | 1.03 [0.86-1.24] | 1.02 [0.84-1.24] |
| O20 | 2,010 | 50,437,370 | 0.04 | 0.04 | 0.04 | 0.00 | 0.00 | 0.00 | 0.97 [0.83-1.13] | 0.98 [0.84-1.15] |
| O23 | 840 | 50,711,160 | 0.02 | 0.02 | 0.02 | 0.00 | 0.00 | 0.00 | 0.93 [0.73-1.19] | 0.89 [0.69-1.14] |
| O24 | 3,710 | 50,710,150 | 0.08 | 0.07 | 0.07 | 0.00 | 0.00 | 0.00 | 1.04 [0.93-1.16] | 1.03 [0.92-1.16] |
| O26 | 4,120 | 49,332,280 | 0.09 | 0.08 | 0.09 | 0.01 | 0.00 | −0.01 | 0.95 [0.85-1.05] | 0.92 [0.83-1.03] |
| O28 | 720 | 50,966,200 | 0.01 | 0.01 | 0.01 | 0.00 | 0.00 | 0.00 | 0.97 [0.75-1.25] | 0.94 [0.72-1.23] |
| O30 | 1,100 | 50,841,560 | 0.02 | 0.02 | 0.02 | 0.00 | 0.00 | 0.00 | 0.86 [0.69-1.07] | 0.88 [0.70-1.10] |
| O32 | 3,370 | 50,219,210 | 0.07 | 0.07 | 0.07 | 0.00 | 0.00 | 0.00 | 0.96 [0.85-1.08] | 0.98 [0.87-1.11] |
| O34 | 7,340 | 49,685,010 | 0.15 | 0.15 | 0.15 | 0.01 | 0.00 | 0.00 | 1.00 [0.93-1.08] | 1.03 [0.95-1.12] |
| O35 | 1,420 | 50,902,510 | 0.03 | 0.03 | 0.03 | 0.00 | 0.00 | 0.00 | 0.97 [0.80-1.16] | 0.95 [0.78-1.15] |
| O36 | 5,790 | 49,331,580 | 0.13 | 0.11 | 0.13 | 0.01 | 0.00 | 0.00 | 1.02 [0.93-1.11] | 1.02 [0.93-1.11] |
| O40 | 940 | 50,942,470 | 0.02 | 0.02 | 0.02 | 0.00 | 0.00 | 0.00 | 0.95 [0.76-1.19] | 0.96 [0.76-1.22] |
| O41 | 770 | 50,802,530 | 0.01 | 0.02 | 0.01 | 0.00 | 0.00 | 0.00 | 0.99 [0.77-1.27] | 1.05 [0.81-1.36] |
| O42 | 3,440 | 49,887,940 | 0.06 | 0.07 | 0.07 | −0.01 | −0.01 | −0.01 | 0.84 [0.75-0.95]* | 0.89 [0.78-1.00] |

| | | | | | | | | | | |
|---|---|---|---|---|---|---|---|---|---|---|
| O43 | 620 | 50,856,460 | 0.01 | 0.01 | 0.01 | 0.00 | 0.00 | 0.00 | 0.94 [0.71-1.25] | 0.92 [0.69-1.23] |
| O44 | 790 | 50,915,210 | 0.01 | 0.02 | 0.02 | 0.00 | 0.00 | 0.00 | 0.98 [0.76-1.25] | 0.97 [0.75-1.26] |
| O46 | 1,800 | 50,423,420 | 0.04 | 0.04 | 0.04 | 0.00 | 0.00 | 0.00 | 1.04 [0.89-1.22] | 1.04 [0.88-1.22] |
| O47 | 850 | 50,093,870 | 0.02 | 0.02 | 0.02 | 0.00 | 0.00 | 0.00 | 0.95 [0.75-1.20] | 0.91 [0.71-1.16] |
| O48 | 1,800 | 50,013,340 | 0.03 | 0.04 | 0.04 | −0.01 | −0.01 | 0.00 | 0.84 [0.71-1.00] | 0.89 [0.75-1.06] |
| O60 | 2,540 | 50,383,810 | 0.05 | 0.05 | 0.05 | 0.00 | 0.00 | 0.00 | 1.06 [0.93-1.21] | 1.03 [0.90-1.18] |
| O61 | 700 | 50,899,040 | 0.01 | 0.01 | 0.01 | 0.00 | 0.00 | 0.00 | 0.98 [0.75-1.27] | 0.92 [0.70-1.21] |
| O62 | 1,570 | 50,623,230 | 0.03 | 0.03 | 0.03 | 0.00 | 0.00 | 0.00 | 0.96 [0.81-1.15] | 0.99 [0.83-1.19] |
| O63 | 2,810 | 49,613,830 | 0.06 | 0.06 | 0.06 | 0.00 | −0.01 | 0.00 | 0.91 [0.80-1.03] | 0.96 [0.84-1.10] |
| O66 | 690 | 50,787,050 | 0.01 | 0.01 | 0.01 | 0.00 | 0.00 | 0.00 | 0.95 [0.73-1.24] | 0.97 [0.74-1.28] |
| O68 | 6,150 | 48,362,750 | 0.14 | 0.13 | 0.15 | 0.01 | −0.01 | −0.01 | 0.93 [0.85-1.01] | 0.95 [0.87-1.04] |
| O69 | 760 | 50,588,540 | 0.02 | 0.01 | 0.01 | 0.00 | 0.00 | 0.00 | 1.18 [0.93-1.49] | 1.22 [0.95-1.56] |
| O70 | 5,740 | 47,338,080 | 0.14 | 0.12 | 0.15 | 0.02 | −0.01 | 0.00 | 0.93 [0.85-1.01] | 1.00 [0.92-1.10] |
| O72 | 5,900 | 49,797,520 | 0.12 | 0.12 | 0.13 | 0.00 | 0.00 | 0.00 | 0.97 [0.89-1.06] | 1.00 [0.92-1.10] |
| O75 | 2,460 | 50,317,650 | 0.05 | 0.05 | 0.05 | 0.00 | 0.00 | 0.00 | 0.91 [0.79-1.05] | 0.97 [0.83-1.12] |
| O80 | 1,620 | 49,503,870 | 0.04 | 0.03 | 0.03 | 0.00 | 0.00 | 0.00 | 1.05 [0.89-1.24] | 1.11 [0.94-1.32] |
| O82 | 780 | 50,708,280 | 0.01 | 0.02 | 0.02 | 0.00 | 0.00 | 0.00 | 0.86 [0.67-1.11] | 0.91 [0.69-1.18] |
| O86 | 980 | 50,851,790 | 0.02 | 0.02 | 0.02 | 0.00 | 0.00 | 0.00 | 1.20 [0.97-1.47] | 1.16 [0.93-1.43] |
| O90 | 1,250 | 50,865,190 | 0.03 | 0.02 | 0.02 | 0.00 | 0.00 | 0.00 | 1.10 [0.91-1.33] | 1.08 [0.89-1.31] |
| O99 | 6,040 | 49,408,590 | 0.14 | 0.12 | 0.13 | 0.02 | 0.01 | 0.00 | 1.06 [0.97-1.15] | 1.02 [0.93-1.11] |
| 40+ - Q00-Q99 | | | | | | | | | | |
| Q05 | 1,210 | 51,029,190 | 0.03 | 0.02 | 0.02 | 0.01 | 0.01 | 0.00 | 1.32 [1.10-1.60]* | 1.16 [0.96-1.41] |
| Q07 | 650 | 51,050,160 | 0.02 | 0.01 | 0.01 | 0.00 | 0.00 | 0.00 | 1.31 [1.02-1.69] | 1.10 [0.84-1.43] |
| Q12 | 610 | 51,048,900 | 0.01 | 0.01 | 0.01 | 0.00 | 0.00 | 0.00 | 1.23 [0.93-1.62] | 1.18 [0.88-1.56] |
| Q18 | 730 | 51,034,680 | 0.02 | 0.01 | 0.01 | 0.00 | 0.00 | 0.00 | 1.18 [0.92-1.50] | 1.10 [0.86-1.42] |
| Q21 | 5,110 | 50,977,840 | 0.12 | 0.10 | 0.10 | 0.03 | 0.03 | 0.02 | 1.28 [1.16-1.40]* | 1.16 [1.06-1.28]* |
| Q23 | 2,820 | 51,028,170 | 0.06 | 0.05 | 0.05 | 0.01 | 0.01 | 0.01 | 1.14 [1.00-1.30] | 1.10 [0.96-1.25] |
| Q24 | 1,510 | 51,038,180 | 0.04 | 0.03 | 0.03 | 0.01 | 0.01 | 0.01 | 1.40 [1.19-1.66]* | 1.24 [1.04-1.47] |
| Q25 | 1,070 | 51,044,910 | 0.02 | 0.02 | 0.02 | 0.00 | 0.00 | 0.00 | 1.19 [0.97-1.46] | 1.08 [0.87-1.33] |
| Q27 | 1,340 | 51,041,150 | 0.03 | 0.03 | 0.03 | 0.00 | 0.00 | 0.00 | 1.19 [0.99-1.43] | 1.11 [0.91-1.34] |
| Q28 | 1,020 | 51,040,250 | 0.03 | 0.02 | 0.02 | 0.01 | 0.01 | 0.01 | 1.36 [1.11-1.66]* | 1.25 [1.02-1.54] |
| Q38 | 2,850 | 51,029,560 | 0.07 | 0.05 | 0.06 | 0.01 | 0.01 | 0.01 | 1.20 [1.05-1.37]* | 1.10 [0.97-1.26] |
| Q39 | 1,500 | 51,045,930 | 0.04 | 0.03 | 0.03 | 0.01 | 0.01 | 0.01 | 1.29 [1.08-1.53]* | 1.16 [0.97-1.39] |
| Q40 | 900 | 51,052,630 | 0.02 | 0.02 | 0.02 | 0.00 | 0.00 | 0.00 | 1.17 [0.94-1.47] | 1.03 [0.82-1.30] |
| Q43 | 2,500 | 51,026,550 | 0.06 | 0.05 | 0.05 | 0.01 | 0.01 | 0.01 | 1.22 [1.07-1.39]* | 1.10 [0.96-1.26] |
| Q44 | 1,010 | 51,050,000 | 0.02 | 0.02 | 0.02 | 0.00 | 0.00 | 0.00 | 1.11 [0.90-1.38] | 1.01 [0.81-1.26] |
| Q50 | 1,050 | 51,015,110 | 0.02 | 0.02 | 0.02 | 0.00 | 0.00 | 0.00 | 1.12 [0.91-1.37] | 1.04 [0.84-1.28] |
| Q51 | 1,300 | 51,021,230 | 0.03 | 0.02 | 0.02 | 0.00 | 0.00 | 0.00 | 1.20 [1.00-1.44] | 1.08 [0.89-1.31] |
| Q52 | 450 | 51,050,330 | 0.01 | 0.01 | 0.01 | 0.00 | 0.00 | 0.00 | 1.45 [1.08-1.96] | 1.32 [0.97-1.80] |
| Q54 | 1,270 | 51,043,560 | 0.03 | 0.02 | 0.02 | 0.01 | 0.00 | 0.00 | 1.17 [0.96-1.41] | 1.12 [0.92-1.36] |
| Q60 | 1,830 | 51,039,870 | 0.04 | 0.03 | 0.04 | 0.01 | 0.01 | 0.00 | 1.23 [1.05-1.44] | 1.13 [0.96-1.32] |
| Q61 | 5,560 | 50,985,020 | 0.13 | 0.11 | 0.11 | 0.02 | 0.02 | 0.01 | 1.19 [1.08-1.30]* | 1.10 [1.01-1.21] |
| Q63 | 2,890 | 51,020,860 | 0.06 | 0.06 | 0.06 | 0.01 | 0.01 | 0.00 | 1.09 [0.96-1.24] | 0.97 [0.85-1.10] |
| Q65 | 1,180 | 51,034,930 | 0.03 | 0.02 | 0.02 | 0.00 | 0.00 | 0.00 | 1.18 [0.97-1.43] | 1.08 [0.88-1.32] |
| Q66 | 1,410 | 51,032,650 | 0.04 | 0.02 | 0.03 | 0.02 | 0.02 | 0.01 | 1.60 [1.36-1.89]* | 1.44 [1.21-1.71]* |
| Q67 | 730 | 51,050,250 | 0.02 | 0.01 | 0.01 | 0.01 | 0.01 | 0.01 | 1.56 [1.23-1.99]* | 1.47 [1.15-1.87]* |
| Q74 | 710 | 51,049,320 | 0.02 | 0.01 | 0.01 | 0.00 | 0.00 | 0.00 | 1.35 [1.06-1.72] | 1.18 [0.91-1.52] |
| Q76 | 1,340 | 51,037,170 | 0.03 | 0.02 | 0.02 | 0.01 | 0.01 | 0.00 | 1.35 [1.13-1.61]* | 1.11 [0.92-1.34] |
| Q78 | 780 | 51,051,370 | 0.02 | 0.01 | 0.02 | 0.01 | 0.00 | 0.00 | 1.30 [1.03-1.63] | 1.12 [0.88-1.42] |
| Q79 | 1,190 | 51,045,600 | 0.03 | 0.02 | 0.02 | 0.00 | 0.00 | 0.00 | 1.19 [0.98-1.45] | 0.91 [0.74-1.12] |
| Q82 | 990 | 51,040,910 | 0.03 | 0.02 | 0.02 | 0.01 | 0.01 | 0.01 | 1.81 [1.49-2.21]* | 1.58 [1.29-1.94]* |
| Q85 | 1,140 | 51,038,160 | 0.02 | 0.02 | 0.02 | 0.00 | 0.00 | 0.00 | 1.08 [0.88-1.32] | 1.00 [0.81-1.24] |
| Q87 | 870 | 51,039,250 | 0.02 | 0.02 | 0.02 | 0.01 | 0.01 | 0.01 | 1.59 [1.29-1.97]* | 1.42 [1.14-1.77]* |
| Q89 | 1,000 | 51,041,070 | 0.02 | 0.02 | 0.02 | 0.00 | 0.00 | 0.00 | 1.15 [0.93-1.43] | 1.08 [0.87-1.34] |
| Q90 | 1,120 | 51,012,000 | 0.06 | 0.01 | 0.01 | 0.04 | 0.04 | 0.04 | 4.16 [3.54-4.89]* | 4.05 [3.44-4.78]* |
| 40+ - R00-R99 | | | | | | | | | | |
| R00 | 154,590 | 49,519,820 | 3.73 | 2.99 | 3.07 | 0.74 | 0.66 | 0.37 | 1.22 [1.20-1.24]* | 1.11 [1.09-1.13]* |
| R01 | 21,740 | 50,859,420 | 0.52 | 0.41 | 0.42 | 0.11 | 0.10 | 0.06 | 1.23 [1.18-1.29]* | 1.14 [1.08-1.19]* |
| R02 | 27,160 | 50,904,940 | 0.64 | 0.51 | 0.53 | 0.14 | 0.12 | 0.07 | 1.22 [1.17-1.28]* | 1.12 [1.07-1.17]* |
| R03 | 54,600 | 50,501,440 | 1.28 | 1.04 | 1.06 | 0.24 | 0.22 | 0.15 | 1.20 [1.17-1.24]* | 1.13 [1.10-1.16]* |
| R04 | 63,950 | 50,218,000 | 1.63 | 1.20 | 1.21 | 0.43 | 0.41 | 0.28 | 1.34 [1.31-1.38]* | 1.21 [1.18-1.24]* |
| R05 | 54,200 | 50,521,300 | 1.39 | 1.00 | 1.03 | 0.39 | 0.37 | 0.22 | 1.36 [1.32-1.39]* | 1.19 [1.15-1.22]* |
| R06 | 153,380 | 49,259,560 | 3.96 | 2.93 | 2.98 | 1.03 | 0.99 | 0.59 | 1.33 [1.31-1.35]* | 1.18 [1.16-1.20]* |
| R07 | 267,960 | 45,545,490 | 7.35 | 5.56 | 5.61 | 1.78 | 1.73 | 1.00 | 1.31 [1.29-1.33]* | 1.16 [1.14-1.17]* |
| R09 | 24,530 | 50,834,620 | 0.60 | 0.46 | 0.46 | 0.15 | 0.14 | 0.09 | 1.31 [1.25-1.36]* | 1.17 [1.11-1.22]* |
| R10 | 243,520 | 44,601,240 | 6.80 | 5.16 | 5.22 | 1.64 | 1.58 | 0.93 | 1.30 [1.29-1.32]* | 1.16 [1.14-1.18]* |
| R11 | 171,590 | 48,900,060 | 4.27 | 3.34 | 3.41 | 0.93 | 0.87 | 0.50 | 1.25 [1.23-1.27]* | 1.13 [1.11-1.15]* |
| R12 | 14,020 | 50,777,760 | 0.35 | 0.26 | 0.26 | 0.09 | 0.09 | 0.05 | 1.34 [1.27-1.42]* | 1.17 [1.11-1.24]* |
| R13 | 121,980 | 50,012,610 | 3.09 | 2.30 | 2.35 | 0.80 | 0.74 | 0.48 | 1.31 [1.29-1.34]* | 1.18 [1.16-1.21]* |
| R14 | 23,820 | 50,780,200 | 0.58 | 0.44 | 0.45 | 0.14 | 0.14 | 0.07 | 1.30 [1.25-1.36]* | 1.15 [1.10-1.20]* |
| R15 | 52,810 | 50,808,340 | 1.25 | 1.00 | 1.05 | 0.25 | 0.20 | 0.14 | 1.19 [1.15-1.23]* | 1.12 [1.09-1.16]* |
| R16 | 16,440 | 50,930,150 | 0.43 | 0.30 | 0.30 | 0.13 | 0.13 | 0.09 | 1.41 [1.34-1.49]* | 1.27 [1.20-1.34]* |
| R17 | 15,360 | 50,905,080 | 0.36 | 0.29 | 0.29 | 0.07 | 0.07 | 0.05 | 1.23 [1.17-1.30]* | 1.17 [1.11-1.24]* |
| R18 | 41,020 | 50,850,550 | 0.95 | 0.78 | 0.78 | 0.18 | 0.17 | 0.13 | 1.22 [1.18-1.26]* | 1.15 [1.11-1.19]* |
| R19 | 141,980 | 49,191,500 | 3.66 | 2.72 | 2.74 | 0.94 | 0.91 | 0.59 | 1.33 [1.31-1.36]* | 1.19 [1.17-1.21]* |
| R20 | 32,460 | 50,545,700 | 0.79 | 0.61 | 0.62 | 0.18 | 0.17 | 0.08 | 1.27 [1.23-1.32]* | 1.11 [1.07-1.15]* |
| R21 | 34,340 | 50,635,460 | 1.07 | 0.60 | 0.60 | 0.47 | 0.47 | 0.41 | 1.78 [1.72-1.84]* | 1.63 [1.58-1.69]* |
| R22 | 23,860 | 50,647,180 | 0.61 | 0.44 | 0.45 | 0.17 | 0.16 | 0.11 | 1.36 [1.31-1.42]* | 1.23 [1.18-1.29]* |
| R23 | 14,860 | 50,940,630 | 0.37 | 0.28 | 0.28 | 0.09 | 0.09 | 0.06 | 1.33 [1.26-1.41]* | 1.21 [1.14-1.28]* |
| R25 | 20,420 | 50,877,980 | 0.50 | 0.38 | 0.39 | 0.12 | 0.11 | 0.06 | 1.28 [1.22-1.35]* | 1.14 [1.09-1.20]* |
| R26 | 195,630 | 50,335,360 | 4.74 | 3.70 | 3.95 | 1.04 | 0.80 | 0.50 | 1.20 [1.18-1.22]* | 1.12 [1.10-1.14]* |
| R27 | 5,600 | 51,005,730 | 0.13 | 0.11 | 0.11 | 0.03 | 0.02 | 0.01 | 1.20 [1.09-1.31]* | 1.09 [0.99-1.20] |
| R29 | 261,830 | 50,125,150 | 6.32 | 4.99 | 5.31 | 1.34 | 1.01 | 0.60 | 1.19 [1.17-1.21]* | 1.11 [1.09-1.12]* |
| R30 | 13,690 | 50,821,060 | 0.35 | 0.25 | 0.26 | 0.10 | 0.09 | 0.05 | 1.36 [1.29-1.44]* | 1.18 [1.11-1.25]* |
| R31 | 130,760 | 48,722,830 | 3.12 | 2.59 | 2.63 | 0.53 | 0.48 | 0.31 | 1.18 [1.16-1.21]* | 1.11 [1.09-1.13]* |
| R32 | 99,600 | 50,469,900 | 2.35 | 1.89 | 2.00 | 0.45 | 0.35 | 0.22 | 1.17 [1.15-1.20]* | 1.10 [1.08-1.13]* |
| R33 | 191,170 | 49,675,710 | 4.52 | 3.70 | 3.90 | 0.81 | 0.62 | 0.40 | 1.16 [1.14-1.18]* | 1.10 [1.08-1.12]* |
| R34 | 4,660 | 51,041,130 | 0.11 | 0.09 | 0.09 | 0.03 | 0.02 | 0.01 | 1.26 [1.14-1.39]* | 1.15 [1.03-1.27]* |
| R35 | 27,640 | 50,422,950 | 0.69 | 0.52 | 0.52 | 0.17 | 0.17 | 0.11 | 1.32 [1.27-1.38]* | 1.19 [1.14-1.24]* |
| R36 | 570 | 51,054,680 | 0.01 | 0.01 | 0.01 | 0.00 | 0.00 | 0.00 | 1.39 [1.05-1.85] | 1.24 [0.92-1.66] |
| R39 | 32,840 | 50,387,690 | 0.82 | 0.62 | 0.62 | 0.20 | 0.20 | 0.13 | 1.33 [1.28-1.38]* | 1.19 [1.15-1.23]* |
| R40 | 26,600 | 50,894,060 | 0.65 | 0.50 | 0.52 | 0.15 | 0.12 | 0.07 | 1.24 [1.18-1.29]* | 1.12 [1.08-1.17]* |
| R41 | 188,980 | 50,162,900 | 4.47 | 3.62 | 3.81 | 0.85 | 0.66 | 0.43 | 1.17 [1.15-1.19]* | 1.11 [1.09-1.13]* |
| R42 | 89,870 | 50,066,350 | 2.19 | 1.71 | 1.76 | 0.48 | 0.43 | 0.23 | 1.24 [1.22-1.27]* | 1.12 [1.09-1.15]* |
| R43 | 1,100 | 51,051,570 | 0.03 | 0.02 | 0.02 | 0.01 | 0.01 | 0.00 | 1.36 [1.11-1.66]* | 1.14 [0.92-1.41] |
| R44 | 23,180 | 50,928,440 | 0.55 | 0.44 | 0.45 | 0.11 | 0.10 | 0.06 | 1.22 [1.17-1.28]* | 1.12 [1.07-1.17]* |
| R45 | 41,290 | 50,843,770 | 0.99 | 0.77 | 0.80 | 0.21 | 0.19 | 0.10 | 1.24 [1.20-1.28]* | 1.11 [1.07-1.15]* |
| R46 | 15,420 | 50,991,930 | 0.34 | 0.29 | 0.30 | 0.05 | 0.04 | 0.02 | 1.12 [1.06-1.19]* | 1.07 [1.01-1.13] |
| R47 | 50,890 | 50,700,370 | 1.17 | 0.97 | 1.01 | 0.20 | 0.16 | 0.08 | 1.16 [1.13-1.20]* | 1.08 [1.04-1.11]* |
| R48 | 780 | 51,055,800 | 0.02 | 0.01 | 0.01 | 0.01 | 0.00 | 0.00 | 1.31 [1.03-1.66] | 1.13 [0.88-1.45] |
| R49 | 9,190 | 50,884,460 | 0.23 | 0.17 | 0.17 | 0.06 | 0.06 | 0.03 | 1.32 [1.24-1.42]* | 1.14 [1.07-1.23]* |
| R50 | 58,530 | 50,402,530 | 1.43 | 1.10 | 1.12 | 0.32 | 0.30 | 0.21 | 1.27 [1.24-1.31]* | 1.17 [1.14-1.21]* |
| R51 | 85,750 | 49,412,160 | 2.20 | 1.64 | 1.66 | 0.56 | 0.54 | 0.29 | 1.32 [1.29-1.35]* | 1.15 [1.13-1.18]* |
| R52 | 20,000 | 50,843,250 | 0.52 | 0.37 | 0.37 | 0.16 | 0.15 | 0.09 | 1.42 [1.35-1.48]* | 1.21 [1.15-1.27]* |
| R53 | 86,400 | 50,484,940 | 2.08 | 1.63 | 1.70 | 0.45 | 0.38 | 0.22 | 1.22 [1.19-1.25]* | 1.12 [1.09-1.14]* |
| R54 | 127,830 | 50,611,500 | 3.00 | 2.43 | 2.57 | 0.57 | 0.43 | 0.29 | 1.17 [1.14-1.19]* | 1.11 [1.08-1.13]* |

| | | | | | | | | | |
|---|---|---|---|---|---|---|---|---|---|
| R55 | 168,480 | 49,214,460 | 4.07 | 3.28 | 3.38 | 0.79 | 0.69 | 0.45 | 1.20 [1.18-1.22]* | 1.12 [1.10-1.14]* |
| R56 | 42,880 | 50,599,490 | 1.03 | 0.81 | 0.82 | 0.23 | 0.21 | 0.15 | 1.26 [1.22-1.30]* | 1.16 [1.13-1.20]* |
| R57 | 15,970 | 51,025,010 | 0.37 | 0.30 | 0.31 | 0.07 | 0.05 | 0.03 | 1.18 [1.11-1.24]* | 1.10 [1.04-1.16]* |
| R58 | 2,310 | 51,046,040 | 0.06 | 0.04 | 0.04 | 0.02 | 0.02 | 0.01 | 1.34 [1.17-1.54]* | 1.22 [1.06-1.41]* |
| R59 | 41,420 | 50,657,440 | 1.03 | 0.77 | 0.78 | 0.26 | 0.25 | 0.20 | 1.32 [1.28-1.37]* | 1.24 [1.20-1.29]* |
| R60 | 88,320 | 50,597,450 | 2.23 | 1.64 | 1.67 | 0.59 | 0.57 | 0.41 | 1.34 [1.31-1.37]* | 1.22 [1.19-1.25]* |
| R61 | 7,100 | 50,951,720 | 0.18 | 0.13 | 0.13 | 0.05 | 0.05 | 0.03 | 1.34 [1.24-1.45]* | 1.17 [1.08-1.27]* |
| R62 | 1,020 | 51,054,970 | 0.03 | 0.02 | 0.02 | 0.01 | 0.01 | 0.00 | 1.26 [1.02-1.56] | 1.18 [0.95-1.46] |
| R63 | 163,750 | 49,789,660 | 4.01 | 3.13 | 3.22 | 0.88 | 0.79 | 0.50 | 1.24 [1.22-1.27]* | 1.14 [1.12-1.16]* |
| R64 | 5,150 | 51,053,120 | 0.11 | 0.10 | 0.10 | 0.01 | 0.01 | 0.01 | 1.10 [1.00-1.22] | 1.08 [0.97-1.20] |
| R65 | 11,030 | 51,037,250 | 0.26 | 0.21 | 0.21 | 0.06 | 0.05 | 0.03 | 1.23 [1.15-1.32]* | 1.13 [1.05-1.21]* |
| R68 | 16,230 | 50,972,260 | 0.39 | 0.30 | 0.31 | 0.08 | 0.07 | 0.06 | 1.24 [1.17-1.31]* | 1.17 [1.11-1.24]* |
| R69 | 128,150 | 47,668,880 | 3.22 | 2.57 | 2.59 | 0.65 | 0.63 | 0.44 | 1.24 [1.22-1.27]* | 1.16 [1.14-1.18]* |
| R70 | 1,220 | 51,035,870 | 0.03 | 0.02 | 0.02 | 0.01 | 0.01 | 0.01 | 1.48 [1.23-1.77]* | 1.37 [1.13-1.65]* |
| R71 | 1,010 | 51,052,870 | 0.02 | 0.02 | 0.02 | 0.00 | 0.00 | 0.00 | 1.13 [0.91-1.40] | 1.06 [0.85-1.33] |
| R72 | 5,180 | 51,020,190 | 0.12 | 0.10 | 0.10 | 0.02 | 0.02 | 0.01 | 1.20 [1.09-1.32]* | 1.13 [1.03-1.25] |
| R73 | 20,780 | 50,879,400 | 0.52 | 0.39 | 0.39 | 0.13 | 0.12 | 0.07 | 1.31 [1.25-1.38]* | 1.16 [1.11-1.22]* |
| R74 | 8,640 | 50,996,020 | 0.20 | 0.16 | 0.17 | 0.04 | 0.03 | 0.02 | 1.21 [1.12-1.30]* | 1.10 [1.02-1.19] |
| R76 | 2,070 | 51,028,640 | 0.05 | 0.04 | 0.04 | 0.02 | 0.01 | 0.01 | 1.38 [1.20-1.60]* | 1.26 [1.09-1.46]* |
| R77 | 5,610 | 51,034,410 | 0.14 | 0.10 | 0.11 | 0.03 | 0.03 | 0.02 | 1.27 [1.16-1.40]* | 1.17 [1.06-1.28]* |
| R78 | 740 | 51,051,060 | 0.02 | 0.01 | 0.02 | 0.00 | 0.00 | 0.00 | 1.12 [0.88-1.45] | 1.02 [0.79-1.32] |
| R79 | 119,070 | 50,142,330 | 2.89 | 2.26 | 2.33 | 0.62 | 0.56 | 0.35 | 1.24 [1.22-1.26]* | 1.14 [1.11-1.16]* |
| R80 | 3,400 | 50,996,480 | 0.09 | 0.06 | 0.06 | 0.03 | 0.03 | 0.02 | 1.45 [1.29-1.62]* | 1.31 [1.17-1.47]* |
| R82 | 7,610 | 50,984,340 | 0.17 | 0.14 | 0.15 | 0.03 | 0.02 | 0.01 | 1.16 [1.07-1.25]* | 1.06 [0.98-1.15] |
| R84 | 2,580 | 51,047,160 | 0.07 | 0.05 | 0.05 | 0.02 | 0.02 | 0.01 | 1.45 [1.28-1.65]* | 1.24 [1.09-1.42]* |
| R85 | 1,410 | 51,019,260 | 0.03 | 0.03 | 0.03 | 0.01 | 0.01 | 0.00 | 1.32 [1.11-1.58]* | 1.16 [0.97-1.39] |
| R87 | 3,940 | 50,652,250 | 0.08 | 0.08 | 0.08 | 0.01 | 0.01 | 0.00 | 1.08 [0.97-1.21] | 1.03 [0.92-1.15] |
| R89 | 1,360 | 51,047,170 | 0.04 | 0.02 | 0.02 | 0.01 | 0.01 | 0.01 | 1.47 [1.23-1.75]* | 1.35 [1.13-1.62]* |
| R90 | 4,770 | 51,022,770 | 0.11 | 0.09 | 0.09 | 0.01 | 0.01 | 0.01 | 1.15 [1.04-1.27]* | 1.08 [0.98-1.20] |
| R91 | 60,460 | 50,650,720 | 1.46 | 1.14 | 1.17 | 0.32 | 0.29 | 0.19 | 1.25 [1.22-1.29]* | 1.15 [1.12-1.18]* |
| R92 | 560 | 51,055,190 | 0.01 | 0.01 | 0.01 | 0.00 | 0.00 | 0.00 | 1.07 [0.80-1.44] | 1.01 [0.75-1.37] |
| R93 | 57,480 | 50,576,680 | 1.34 | 1.09 | 1.12 | 0.25 | 0.23 | 0.14 | 1.20 [1.17-1.24]* | 1.11 [1.08-1.15]* |
| R94 | 97,580 | 50,269,430 | 2.39 | 1.84 | 1.90 | 0.55 | 0.49 | 0.34 | 1.26 [1.23-1.29]* | 1.16 [1.14-1.19]* |
| 40+ - S00-T88 | | | | | | | | | | |
| S00 | 89,310 | 50,460,570 | 2.13 | 1.69 | 1.77 | 0.44 | 0.36 | 0.23 | 1.20 [1.17-1.23]* | 1.12 [1.09-1.15]* |
| S01 | 87,380 | 50,298,170 | 2.09 | 1.66 | 1.74 | 0.43 | 0.36 | 0.25 | 1.20 [1.18-1.23]* | 1.14 [1.11-1.17]* |
| S02 | 20,990 | 50,641,960 | 0.49 | 0.40 | 0.40 | 0.09 | 0.09 | 0.06 | 1.21 [1.16-1.27]* | 1.14 [1.09-1.20]* |
| S03 | 520 | 51,054,710 | 0.01 | 0.01 | 0.01 | 0.00 | 0.00 | 0.00 | 1.16 [0.86-1.57] | 1.07 [0.79-1.45] |
| S05 | 7,860 | 50,966,860 | 0.18 | 0.15 | 0.15 | 0.03 | 0.03 | 0.02 | 1.21 [1.11-1.31]* | 1.14 [1.05-1.23]* |
| S06 | 27,530 | 50,839,170 | 0.62 | 0.52 | 0.55 | 0.09 | 0.07 | 0.05 | 1.14 [1.09-1.19]* | 1.08 [1.03-1.13]* |
| S09 | 39,960 | 50,552,650 | 0.97 | 0.75 | 0.77 | 0.22 | 0.20 | 0.13 | 1.25 [1.21-1.30]* | 1.15 [1.11-1.19]* |
| S10 | 1,920 | 51,034,480 | 0.04 | 0.04 | 0.04 | 0.01 | 0.00 | 0.00 | 1.09 [0.93-1.29] | 0.97 [0.82-1.15] |
| S11 | 910 | 51,046,210 | 0.02 | 0.02 | 0.02 | 0.01 | 0.01 | 0.01 | 1.46 [1.17-1.82]* | 1.34 [1.06-1.68] |
| S12 | 7,500 | 51,002,670 | 0.17 | 0.14 | 0.15 | 0.03 | 0.03 | 0.02 | 1.18 [1.08-1.28]* | 1.10 [1.01-1.20] |
| S13 | 1,250 | 51,030,320 | 0.03 | 0.02 | 0.02 | 0.01 | 0.01 | 0.00 | 1.21 [1.00-1.47] | 1.12 [0.92-1.36] |
| S14 | 1,250 | 51,045,880 | 0.03 | 0.02 | 0.02 | 0.00 | 0.00 | 0.00 | 1.08 [0.89-1.32] | 1.06 [0.87-1.30] |
| S19 | 1,570 | 51,029,330 | 0.04 | 0.03 | 0.03 | 0.01 | 0.01 | 0.00 | 1.28 [1.08-1.52]* | 1.11 [0.93-1.32] |
| S20 | 10,170 | 50,970,910 | 0.25 | 0.19 | 0.20 | 0.06 | 0.05 | 0.03 | 1.25 [1.17-1.34]* | 1.13 [1.05-1.21]* |
| S21 | 1,150 | 51,043,830 | 0.03 | 0.02 | 0.02 | 0.01 | 0.01 | 0.01 | 1.45 [1.19-1.77]* | 1.32 [1.08-1.63]* |
| S22 | 37,920 | 50,710,200 | 0.90 | 0.71 | 0.73 | 0.19 | 0.17 | 0.10 | 1.24 [1.19-1.28]* | 1.13 [1.09-1.17]* |
| S27 | 9,750 | 50,949,500 | 0.21 | 0.19 | 0.19 | 0.03 | 0.03 | 0.02 | 1.15 [1.07-1.23]* | 1.08 [1.00-1.16] |
| S29 | 2,720 | 51,031,230 | 0.07 | 0.05 | 0.05 | 0.02 | 0.02 | 0.02 | 1.48 [1.30-1.68]* | 1.30 [1.14-1.48]* |
| S30 | 16,650 | 50,919,870 | 0.40 | 0.31 | 0.31 | 0.09 | 0.09 | 0.06 | 1.29 [1.22-1.36]* | 1.17 [1.11-1.24]* |
| S31 | 2,520 | 51,015,690 | 0.06 | 0.05 | 0.05 | 0.02 | 0.02 | 0.01 | 1.36 [1.19-1.56]* | 1.21 [1.05-1.39]* |
| S32 | 42,330 | 50,740,110 | 0.97 | 0.81 | 0.84 | 0.16 | 0.13 | 0.09 | 1.16 [1.12-1.20]* | 1.10 [1.06-1.14]* |
| S36 | 2,890 | 51,020,890 | 0.07 | 0.05 | 0.06 | 0.01 | 0.01 | 0.01 | 1.17 [1.03-1.33] | 1.13 [0.99-1.28] |
| S37 | 2,740 | 51,033,330 | 0.06 | 0.05 | 0.06 | 0.00 | 0.00 | 0.00 | 1.02 [0.89-1.17] | 0.98 [0.85-1.13] |
| S39 | 5,970 | 50,975,010 | 0.14 | 0.11 | 0.11 | 0.03 | 0.03 | 0.01 | 1.22 [1.12-1.34]* | 1.10 [1.00-1.21] |
| S40 | 15,260 | 50,973,710 | 0.36 | 0.29 | 0.30 | 0.08 | 0.06 | 0.04 | 1.21 [1.14-1.28]* | 1.11 [1.04-1.18]* |
| S41 | 5,020 | 51,026,960 | 0.13 | 0.09 | 0.10 | 0.04 | 0.03 | 0.02 | 1.34 [1.21-1.48]* | 1.21 [1.09-1.34]* |
| S42 | 45,110 | 50,582,710 | 1.02 | 0.86 | 0.89 | 0.15 | 0.13 | 0.08 | 1.14 [1.10-1.18]* | 1.08 [1.05-1.12]* |
| S43 | 7,940 | 50,958,290 | 0.17 | 0.15 | 0.16 | 0.02 | 0.02 | 0.01 | 1.09 [1.01-1.19] | 1.05 [0.97-1.14] |
| S46 | 3,160 | 51,027,860 | 0.07 | 0.06 | 0.06 | 0.01 | 0.00 | 0.00 | 1.07 [0.95-1.22] | 0.97 [0.85-1.10] |
| S49 | 4,770 | 51,022,330 | 0.12 | 0.09 | 0.09 | 0.03 | 0.02 | 0.02 | 1.27 [1.14-1.40]* | 1.15 [1.04-1.28]* |
| S50 | 15,840 | 50,974,110 | 0.38 | 0.30 | 0.31 | 0.08 | 0.07 | 0.04 | 1.23 [1.16-1.30]* | 1.13 [1.07-1.20]* |
| S51 | 21,730 | 50,917,850 | 0.53 | 0.40 | 0.43 | 0.13 | 0.11 | 0.06 | 1.25 [1.19-1.31]* | 1.13 [1.07-1.19]* |
| S52 | 54,650 | 50,175,110 | 1.20 | 1.07 | 1.08 | 0.13 | 0.12 | 0.07 | 1.11 [1.07-1.14]* | 1.07 [1.03-1.10]* |
| S53 | 1,160 | 51,037,270 | 0.02 | 0.02 | 0.02 | 0.00 | 0.00 | 0.00 | 1.07 [0.87-1.31] | 0.98 [0.80-1.21] |
| S54 | 460 | 51,046,080 | 0.01 | 0.01 | 0.01 | 0.00 | 0.00 | 0.00 | 1.35 [1.00-1.83] | 1.28 [0.94-1.75] |
| S56 | 1,580 | 51,016,180 | 0.03 | 0.03 | 0.03 | 0.00 | 0.00 | 0.00 | 1.09 [0.91-1.29] | 1.05 [0.87-1.25] |
| S59 | 1,720 | 51,048,150 | 0.04 | 0.03 | 0.03 | 0.01 | 0.01 | 0.01 | 1.39 [1.18-1.64]* | 1.28 [1.08-1.51]* |
| S60 | 11,290 | 50,972,580 | 0.26 | 0.21 | 0.22 | 0.05 | 0.04 | 0.03 | 1.20 [1.12-1.28]* | 1.11 [1.04-1.19]* |
| S61 | 31,840 | 50,464,500 | 0.71 | 0.61 | 0.63 | 0.10 | 0.08 | 0.04 | 1.14 [1.09-1.18]* | 1.06 [1.02-1.11]* |
| S62 | 21,090 | 50,650,250 | 0.46 | 0.41 | 0.42 | 0.05 | 0.04 | 0.01 | 1.09 [1.04-1.15]* | 1.03 [0.98-1.08] |
| S63 | 4,110 | 50,979,690 | 0.09 | 0.08 | 0.08 | 0.02 | 0.02 | 0.01 | 1.21 [1.09-1.34]* | 1.14 [1.02-1.27] |
| S64 | 3,710 | 50,939,860 | 0.07 | 0.07 | 0.07 | 0.00 | 0.00 | 0.00 | 1.01 [0.91-1.14] | 0.98 [0.87-1.10] |
| S65 | 1,120 | 51,030,690 | 0.02 | 0.02 | 0.02 | 0.00 | 0.00 | 0.00 | 0.99 [0.80-1.22] | 0.96 [0.78-1.19] |
| S66 | 6,880 | 50,830,300 | 0.14 | 0.14 | 0.13 | 0.00 | 0.00 | 0.00 | 1.00 [0.92-1.09] | 0.97 [0.89-1.05] |
| S68 | 2,540 | 50,992,280 | 0.05 | 0.05 | 0.05 | 0.00 | 0.00 | 0.00 | 0.97 [0.85-1.12] | 0.99 [0.86-1.14] |
| S69 | 3,950 | 51,013,120 | 0.09 | 0.07 | 0.07 | 0.02 | 0.02 | 0.01 | 1.29 [1.15-1.44]* | 1.15 [1.02-1.28] |
| S70 | 15,470 | 50,965,780 | 0.36 | 0.29 | 0.31 | 0.07 | 0.06 | 0.04 | 1.19 [1.13-1.26]* | 1.11 [1.05-1.18]* |
| S71 | 1,480 | 51,036,390 | 0.04 | 0.03 | 0.03 | 0.01 | 0.01 | 0.01 | 1.30 [1.09-1.55]* | 1.20 [1.00-1.44] |
| S72 | 121,160 | 50,172,280 | 2.77 | 2.34 | 2.48 | 0.43 | 0.29 | 0.22 | 1.12 [1.10-1.14]* | 1.08 [1.06-1.11]* |
| S73 | 1,260 | 51,044,420 | 0.03 | 0.02 | 0.02 | 0.00 | 0.00 | 0.00 | 1.17 [0.96-1.42] | 1.07 [0.87-1.31] |
| S76 | 3,010 | 51,022,170 | 0.06 | 0.06 | 0.06 | 0.01 | 0.01 | 0.00 | 1.14 [1.01-1.30] | 1.07 [0.94-1.22] |
| S79 | 8,580 | 51,000,920 | 0.21 | 0.16 | 0.17 | 0.05 | 0.04 | 0.03 | 1.25 [1.15-1.34]* | 1.16 [1.07-1.25]* |
| S80 | 29,510 | 50,864,090 | 0.69 | 0.56 | 0.58 | 0.13 | 0.11 | 0.05 | 1.19 [1.14-1.24]* | 1.07 [1.03-1.12]* |
| S81 | 21,220 | 50,896,270 | 0.52 | 0.40 | 0.40 | 0.12 | 0.12 | 0.07 | 1.29 [1.23-1.36]* | 1.16 [1.10-1.22]* |
| S82 | 49,160 | 50,050,160 | 1.08 | 0.96 | 0.98 | 0.12 | 0.10 | 0.04 | 1.10 [1.07-1.14]* | 1.04 [1.01-1.07] |
| S83 | 4,560 | 50,935,820 | 0.10 | 0.09 | 0.09 | 0.02 | 0.02 | 0.01 | 1.18 [1.06-1.30]* | 1.07 [0.97-1.19] |
| S86 | 3,380 | 50,956,260 | 0.08 | 0.06 | 0.06 | 0.01 | 0.01 | 0.01 | 1.20 [1.07-1.34]* | 1.11 [0.98-1.24] |
| S89 | 6,070 | 51,001,030 | 0.15 | 0.11 | 0.12 | 0.04 | 0.03 | 0.02 | 1.27 [1.16-1.38]* | 1.15 [1.05-1.26]* |
| S90 | 6,700 | 51,007,120 | 0.15 | 0.13 | 0.13 | 0.02 | 0.02 | 0.00 | 1.11 [1.02-1.21] | 1.01 [0.92-1.10] |
| S91 | 4,790 | 50,998,550 | 0.12 | 0.09 | 0.09 | 0.03 | 0.03 | 0.02 | 1.30 [1.17-1.43]* | 1.16 [1.05-1.29]* |
| S92 | 10,420 | 50,873,910 | 0.24 | 0.20 | 0.20 | 0.05 | 0.04 | 0.02 | 1.20 [1.12-1.28]* | 1.09 [1.02-1.17] |
| S93 | 3,350 | 51,007,730 | 0.08 | 0.06 | 0.06 | 0.01 | 0.01 | 0.01 | 1.21 [1.07-1.36]* | 1.08 [0.96-1.22] |
| S96 | 530 | 51,049,950 | 0.01 | 0.01 | 0.01 | 0.00 | 0.00 | 0.00 | 1.19 [0.89-1.59] | 1.00 [0.73-1.36] |
| S99 | 2,730 | 51,031,700 | 0.06 | 0.05 | 0.05 | 0.01 | 0.01 | 0.00 | 1.16 [1.01-1.32] | 1.02 [0.89-1.17] |
| T00 | 6,030 | 51,030,220 | 0.14 | 0.11 | 0.12 | 0.03 | 0.02 | 0.01 | 1.19 [1.08-1.31]* | 1.09 [0.99-1.20] |
| T01 | 2,540 | 51,045,610 | 0.06 | 0.05 | 0.05 | 0.01 | 0.01 | 0.00 | 1.18 [1.02-1.36] | 1.05 [0.90-1.21] |
| T02 | 1,800 | 51,041,080 | 0.04 | 0.03 | 0.03 | 0.01 | 0.01 | 0.01 | 1.31 [1.12-1.54]* | 1.26 [1.07-1.48]* |
| T09 | 950 | 51,049,120 | 0.02 | 0.02 | 0.02 | 0.00 | 0.00 | 0.00 | 1.22 [0.97-1.52] | 1.10 [0.87-1.38] |
| T11 | 4,190 | 51,035,990 | 0.10 | 0.08 | 0.08 | 0.02 | 0.02 | 0.01 | 1.28 [1.14-1.43]* | 1.15 [1.03-1.29] |
| T13 | 2,790 | 51,036,580 | 0.07 | 0.05 | 0.05 | 0.01 | 0.01 | 0.01 | 1.26 [1.10-1.44]* | 1.13 [0.98-1.30] |
| T14 | 2,410 | 50,999,610 | 0.06 | 0.05 | 0.05 | 0.01 | 0.01 | 0.00 | 1.23 [1.07-1.41]* | 1.10 [0.95-1.26] |
| T16 | 460 | 51,054,930 | 0.01 | 0.01 | 0.01 | 0.00 | 0.00 | 0.00 | 1.34 [0.98-1.83] | 1.16 [0.83-1.60] |
| T17 | 5,800 | 50,998,940 | 0.14 | 0.11 | 0.11 | 0.04 | 0.03 | 0.02 | 1.30 [1.19-1.43]* | 1.18 [1.08-1.30]* |

| | | | | | | | | | | |
|---|---|---|---|---|---|---|---|---|---|---|
| T18 | 5,010 | 50,980,710 | 0.12 | 0.09 | 0.09 | 0.02 | 0.02 | 0.01 | 1.26 [1.15-1.39]* | 1.13 [1.03-1.25] |
| T20 | 1,270 | 51,037,870 | 0.03 | 0.02 | 0.02 | 0.01 | 0.01 | 0.00 | 1.28 [1.06-1.54] | 1.20 [0.99-1.46] |
| T21 | 2,270 | 51,033,170 | 0.05 | 0.04 | 0.04 | 0.01 | 0.01 | 0.01 | 1.20 [1.04-1.39] | 1.11 [0.96-1.29] |
| T22 | 1,700 | 51,032,910 | 0.04 | 0.03 | 0.03 | 0.00 | 0.00 | 0.00 | 1.13 [0.96-1.34] | 1.06 [0.89-1.26] |
| T23 | 1,530 | 51,029,930 | 0.03 | 0.03 | 0.03 | 0.00 | 0.00 | 0.00 | 1.09 [0.92-1.31] | 1.02 [0.85-1.22] |
| T24 | 2,090 | 51,034,370 | 0.04 | 0.04 | 0.04 | 0.00 | 0.00 | 0.00 | 0.97 [0.82-1.13] | 0.93 [0.79-1.10] |
| T25 | 800 | 51,050,430 | 0.02 | 0.02 | 0.02 | 0.00 | 0.00 | 0.00 | 0.92 [0.71-1.19] | 0.81 [0.61-1.06] |
| T31 | 4,120 | 50,994,360 | 0.09 | 0.08 | 0.08 | 0.01 | 0.01 | 0.00 | 1.10 [0.99-1.23] | 1.03 [0.93-1.16] |
| T36 | 920 | 51,033,520 | 0.02 | 0.02 | 0.02 | 0.01 | 0.01 | 0.01 | 1.50 [1.22-1.85]* | 1.25 [1.01-1.56] |
| T37 | 460 | 51,050,610 | 0.01 | 0.01 | 0.01 | 0.00 | 0.00 | 0.00 | 1.55 [1.14-2.09]* | 1.28 [0.93-1.76] |
| T38 | 2,880 | 51,014,870 | 0.07 | 0.05 | 0.05 | 0.02 | 0.02 | 0.01 | 1.37 [1.21-1.54]* | 1.11 [0.98-1.26] |
| T39 | 16,440 | 50,481,170 | 0.40 | 0.31 | 0.31 | 0.08 | 0.08 | 0.03 | 1.26 [1.20-1.33]* | 1.08 [1.02-1.14]* |
| T40 | 11,900 | 50,827,710 | 0.29 | 0.22 | 0.22 | 0.07 | 0.07 | 0.02 | 1.32 [1.24-1.40]* | 1.06 [0.99-1.13] |
| T42 | 9,520 | 50,726,630 | 0.23 | 0.18 | 0.18 | 0.05 | 0.05 | 0.01 | 1.30 [1.21-1.39]* | 1.06 [0.99-1.14] |
| T43 | 10,760 | 50,652,530 | 0.26 | 0.20 | 0.20 | 0.06 | 0.06 | 0.02 | 1.32 [1.24-1.40]* | 1.08 [1.01-1.15] |
| T44 | 2,910 | 50,994,120 | 0.07 | 0.05 | 0.05 | 0.01 | 0.01 | 0.00 | 1.23 [1.09-1.40]* | 1.02 [0.90-1.17] |
| T45 | 3,670 | 50,996,490 | 0.09 | 0.07 | 0.07 | 0.02 | 0.02 | 0.01 | 1.35 [1.21-1.51]* | 1.14 [1.02-1.28] |
| T46 | 4,560 | 51,005,860 | 0.11 | 0.09 | 0.09 | 0.02 | 0.02 | 0.01 | 1.26 [1.13-1.39]* | 1.06 [0.96-1.18] |
| T47 | 1,620 | 51,033,280 | 0.04 | 0.03 | 0.03 | 0.01 | 0.01 | 0.00 | 1.26 [1.07-1.49]* | 0.99 [0.83-1.18] |
| T48 | 400 | 51,051,790 | 0.01 | 0.01 | 0.01 | 0.01 | 0.01 | 0.00 | 1.77 [1.30-2.40]* | 1.44 [1.04-1.99] |
| T50 | 3,900 | 50,930,850 | 0.09 | 0.07 | 0.07 | 0.02 | 0.02 | 0.00 | 1.26 [1.13-1.40]* | 1.03 [0.92-1.15] |
| T51 | 9,680 | 50,698,730 | 0.23 | 0.18 | 0.18 | 0.04 | 0.05 | 0.01 | 1.26 [1.18-1.35]* | 1.03 [0.96-1.11] |
| T56 | 500 | 51,051,660 | 0.01 | 0.01 | 0.01 | 0.00 | 0.00 | 0.00 | 1.18 [0.88-1.59] | 1.07 [0.79-1.45] |
| T59 | 1,360 | 51,034,740 | 0.03 | 0.03 | 0.03 | 0.00 | 0.00 | 0.00 | 1.13 [0.93-1.37] | 1.03 [0.84-1.25] |
| T63 | 660 | 51,045,030 | 0.01 | 0.01 | 0.01 | 0.00 | 0.00 | 0.00 | 0.95 [0.72-1.25] | 0.88 [0.66-1.16] |
| T65 | 460 | 51,053,480 | 0.01 | 0.01 | 0.01 | 0.00 | 0.00 | 0.00 | 1.29 [0.95-1.75] | 1.10 [0.80-1.52] |
| T68 | 8,900 | 51,032,870 | 0.20 | 0.17 | 0.18 | 0.03 | 0.02 | 0.02 | 1.11 [1.03-1.20]* | 1.11 [1.02-1.20] |
| T78 | 9,310 | 50,863,760 | 0.28 | 0.16 | 0.16 | 0.12 | 0.12 | 0.09 | 1.72 [1.61-1.83]* | 1.45 [1.36-1.55]* |
| T79 | 9,850 | 50,957,870 | 0.22 | 0.19 | 0.19 | 0.03 | 0.02 | 0.01 | 1.12 [1.04-1.21]* | 1.07 [0.99-1.15] |
| T80 | 4,300 | 51,011,140 | 0.11 | 0.08 | 0.08 | 0.03 | 0.03 | 0.02 | 1.38 [1.25-1.52]* | 1.26 [1.14-1.40]* |
| T81 | 137,430 | 48,761,730 | 3.46 | 2.68 | 2.69 | 0.79 | 0.77 | 0.53 | 1.29 [1.26-1.31]* | 1.18 [1.16-1.20]* |
| T82 | 36,330 | 50,654,740 | 0.92 | 0.67 | 0.68 | 0.24 | 0.23 | 0.16 | 1.34 [1.30-1.39]* | 1.21 [1.17-1.25]* |
| T83 | 37,600 | 50,796,320 | 0.84 | 0.72 | 0.74 | 0.12 | 0.10 | 0.06 | 1.14 [1.09-1.18]* | 1.08 [1.04-1.12]* |
| T84 | 47,800 | 50,248,290 | 1.20 | 0.90 | 0.90 | 0.30 | 0.30 | 0.22 | 1.34 [1.30-1.38]* | 1.23 [1.19-1.27]* |
| T85 | 28,850 | 50,712,970 | 0.71 | 0.54 | 0.54 | 0.17 | 0.17 | 0.12 | 1.31 [1.26-1.36]* | 1.20 [1.16-1.25]* |
| T86 | 3,760 | 51,002,780 | 0.09 | 0.07 | 0.07 | 0.02 | 0.03 | 0.02 | 1.38 [1.24-1.53]* | 1.25 [1.12-1.40]* |
| T87 | 2,860 | 51,024,850 | 0.07 | 0.05 | 0.05 | 0.02 | 0.02 | 0.01 | 1.28 [1.13-1.45]* | 1.08 [0.95-1.23] |
| T88 | 8,270 | 50,896,500 | 0.22 | 0.15 | 0.15 | 0.07 | 0.07 | 0.05 | 1.46 [1.36-1.56]* | 1.28 [1.19-1.38]* |
| T90 | 4,750 | 50,934,430 | 0.12 | 0.09 | 0.09 | 0.03 | 0.03 | 0.02 | 1.38 [1.26-1.52]* | 1.22 [1.11-1.35]* |
| T91 | 3,100 | 50,994,840 | 0.07 | 0.06 | 0.06 | 0.02 | 0.02 | 0.01 | 1.28 [1.13-1.44]* | 1.13 [0.99-1.27] |
| T92 | 4,720 | 50,927,970 | 0.11 | 0.09 | 0.09 | 0.02 | 0.02 | 0.01 | 1.24 [1.12-1.36]* | 1.15 [1.04-1.27]* |
| T93 | 6,020 | 50,898,210 | 0.14 | 0.11 | 0.11 | 0.03 | 0.03 | 0.02 | 1.28 [1.18-1.39]* | 1.16 [1.06-1.26]* |
| T94 | 670 | 51,049,040 | 0.02 | 0.01 | 0.01 | 0.00 | 0.00 | 0.00 | 1.11 [0.85-1.44] | 0.99 [0.76-1.29] |
| T98 | 1,170 | 51,040,320 | 0.03 | 0.02 | 0.02 | 0.01 | 0.01 | 0.01 | 1.35 [1.12-1.63]* | 1.23 [1.01-1.50] |
| 40+ - U00-U89 | | | | | | | | | | |
| U07 | 46,240 | 51,019,040 | 1.15 | 0.85 | 0.90 | 0.29 | 0.25 | 0.15 | 1.28 [1.24-1.32]* | 1.15 [1.11-1.19]* |
| U80 | 16,160 | 50,973,850 | 0.44 | 0.29 | 0.30 | 0.15 | 0.15 | 0.11 | 1.49 [1.41-1.57]* | 1.34 [1.27-1.41]* |
| U81 | 980 | 51,057,570 | 0.02 | 0.02 | 0.02 | 0.00 | 0.00 | 0.00 | 1.11 [0.89-1.39] | 1.04 [0.83-1.31] |
| U82 | 31,620 | 50,982,600 | 0.83 | 0.57 | 0.61 | 0.26 | 0.23 | 0.16 | 1.38 [1.32-1.43]* | 1.24 [1.19-1.29]* |
| U83 | 19,020 | 51,017,570 | 0.50 | 0.35 | 0.37 | 0.15 | 0.13 | 0.09 | 1.35 [1.29-1.42]* | 1.21 [1.15-1.28]* |
| U88 | 4,010 | 51,042,810 | 0.10 | 0.07 | 0.08 | 0.03 | 0.02 | 0.02 | 1.30 [1.17-1.45]* | 1.19 [1.06-1.33]* |
| U89 | 4,450 | 51,039,670 | 0.11 | 0.08 | 0.09 | 0.03 | 0.03 | 0.02 | 1.35 [1.22-1.49]* | 1.21 [1.09-1.34]* |
| 40+ - V00-Y99 | | | | | | | | | | |
| V03 | 4,260 | 50,972,610 | 0.10 | 0.08 | 0.08 | 0.02 | 0.01 | 0.01 | 1.18 [1.06-1.31]* | 1.14 [1.02-1.26] |
| V09 | 1,440 | 51,046,090 | 0.03 | 0.03 | 0.03 | 0.01 | 0.01 | 0.00 | 1.30 [1.08-1.56]* | 1.13 [0.94-1.37] |
| V13 | 1,300 | 51,033,280 | 0.02 | 0.03 | 0.03 | 0.00 | 0.00 | 0.00 | 0.90 [0.74-1.11] | 0.90 [0.73-1.11] |
| V18 | 5,000 | 50,963,010 | 0.10 | 0.10 | 0.10 | 0.00 | 0.00 | 0.00 | 1.05 [0.95-1.16] | 1.04 [0.94-1.15] |
| V19 | 770 | 51,046,250 | 0.02 | 0.01 | 0.01 | 0.00 | 0.00 | 0.00 | 1.10 [0.86-1.41] | 1.10 [0.85-1.42] |
| V23 | 1,310 | 51,014,060 | 0.02 | 0.03 | 0.02 | 0.00 | 0.00 | 0.00 | 0.95 [0.78-1.16] | 0.91 [0.74-1.12] |
| V28 | 1,700 | 51,006,060 | 0.03 | 0.03 | 0.03 | 0.00 | 0.00 | 0.00 | 1.03 [0.87-1.23] | 0.96 [0.81-1.15] |
| V43 | 4,800 | 50,927,570 | 0.11 | 0.09 | 0.09 | 0.02 | 0.01 | 0.01 | 1.16 [1.05-1.28]* | 1.06 [0.96-1.18] |
| V47 | 1,730 | 51,021,580 | 0.04 | 0.03 | 0.03 | 0.01 | 0.01 | 0.01 | 1.26 [1.07-1.48]* | 1.15 [0.98-1.36] |
| V48 | 3,280 | 51,016,860 | 0.08 | 0.06 | 0.06 | 0.01 | 0.01 | 0.01 | 1.21 [1.07-1.36]* | 1.12 [0.99-1.26] |
| V49 | 1,840 | 51,004,350 | 0.04 | 0.03 | 0.03 | 0.01 | 0.01 | 0.00 | 1.21 [1.03-1.41] | 1.12 [0.95-1.32] |
| V78 | 1,770 | 51,043,650 | 0.04 | 0.03 | 0.03 | 0.01 | 0.01 | 0.00 | 1.17 [0.99-1.38] | 1.10 [0.92-1.30] |
| V79 | 550 | 51,054,350 | 0.01 | 0.01 | 0.01 | 0.00 | 0.00 | 0.00 | 1.01 [0.74-1.38] | 0.93 [0.68-1.28] |
| V80 | 2,090 | 51,006,750 | 0.03 | 0.04 | 0.04 | −0.01 | −0.01 | −0.01 | 0.76 [0.64-0.90]* | 0.76 [0.64-0.91]* |
| V82 | 430 | 51,056,820 | 0.01 | 0.01 | 0.01 | 0.00 | 0.00 | 0.00 | 1.37 [0.99-1.90] | 1.33 [0.95-1.85] |
| V89 | 650 | 51,020,080 | 0.02 | 0.01 | 0.01 | 0.00 | 0.00 | 0.00 | 1.39 [1.08-1.79] | 1.23 [0.94-1.59] |
| W00 | 6,330 | 50,952,220 | 0.13 | 0.12 | 0.12 | 0.01 | 0.01 | 0.01 | 1.08 [0.99-1.18] | 1.06 [0.97-1.16] |
| W01 | 134,320 | 49,695,770 | 3.16 | 2.60 | 2.72 | 0.55 | 0.44 | 0.28 | 1.16 [1.14-1.18]* | 1.10 [1.08-1.12]* |
| W02 | 1,300 | 51,024,040 | 0.03 | 0.03 | 0.02 | 0.00 | 0.00 | 0.00 | 1.02 [0.84-1.24] | 1.00 [0.82-1.22] |
| W03 | 1,990 | 51,009,050 | 0.04 | 0.04 | 0.04 | 0.00 | 0.00 | 0.00 | 1.03 [0.88-1.21] | 0.97 [0.82-1.14] |
| W05 | 3,040 | 51,039,900 | 0.07 | 0.06 | 0.06 | 0.02 | 0.02 | 0.01 | 1.26 [1.12-1.43]* | 1.14 [1.01-1.30] |
| W06 | 33,080 | 50,919,760 | 0.79 | 0.62 | 0.66 | 0.17 | 0.13 | 0.08 | 1.19 [1.14-1.24]* | 1.11 [1.07-1.16]* |
| W07 | 17,700 | 50,953,100 | 0.42 | 0.33 | 0.35 | 0.09 | 0.07 | 0.05 | 1.21 [1.15-1.28]* | 1.13 [1.07-1.20]* |
| W08 | 4,120 | 51,030,180 | 0.10 | 0.08 | 0.08 | 0.02 | 0.02 | 0.01 | 1.22 [1.10-1.37]* | 1.13 [1.01-1.26] |
| W10 | 51,500 | 50,425,230 | 1.19 | 0.98 | 1.00 | 0.21 | 0.19 | 0.12 | 1.19 [1.16-1.23]* | 1.11 [1.08-1.15]* |
| W11 | 8,440 | 50,882,950 | 0.17 | 0.16 | 0.17 | 0.01 | 0.00 | 0.00 | 1.02 [0.95-1.10] | 0.98 [0.90-1.05] |
| W13 | 2,200 | 51,000,760 | 0.05 | 0.04 | 0.04 | 0.01 | 0.01 | 0.00 | 1.14 [0.98-1.31] | 1.07 [0.92-1.24] |
| W17 | 5,460 | 50,956,560 | 0.12 | 0.10 | 0.10 | 0.02 | 0.02 | 0.01 | 1.19 [1.09-1.31]* | 1.12 [1.02-1.23] |
| W18 | 91,330 | 50,489,480 | 2.19 | 1.73 | 1.80 | 0.46 | 0.39 | 0.26 | 1.22 [1.19-1.25]* | 1.13 [1.11-1.16]* |
| W19 | 213,280 | 49,475,690 | 5.17 | 4.13 | 4.36 | 1.04 | 0.81 | 0.55 | 1.19 [1.17-1.20]* | 1.12 [1.10-1.14]* |
| W20 | 2,950 | 50,997,110 | 0.06 | 0.06 | 0.06 | 0.01 | 0.01 | 0.00 | 1.13 [1.00-1.28] | 1.04 [0.92-1.19] |
| W21 | 760 | 51,034,420 | 0.01 | 0.01 | 0.02 | 0.00 | 0.00 | 0.00 | 0.94 [0.73-1.22] | 0.88 [0.68-1.15] |
| W22 | 15,230 | 50,865,230 | 0.36 | 0.29 | 0.30 | 0.07 | 0.06 | 0.03 | 1.20 [1.13-1.27]* | 1.09 [1.03-1.16]* |
| W23 | 6,210 | 50,934,810 | 0.13 | 0.12 | 0.12 | 0.01 | 0.02 | 0.01 | 1.14 [1.04-1.25]* | 1.10 [1.01-1.20] |
| W25 | 2,690 | 50,939,990 | 0.06 | 0.05 | 0.05 | 0.01 | 0.01 | 0.00 | 1.16 [1.02-1.32] | 1.06 [0.93-1.22] |
| W26 | 3,720 | 50,971,280 | 0.08 | 0.07 | 0.07 | 0.01 | 0.01 | 0.00 | 1.10 [0.98-1.23] | 1.02 [0.91-1.14] |
| W27 | 2,140 | 50,996,190 | 0.04 | 0.04 | 0.04 | 0.00 | 0.00 | 0.00 | 1.08 [0.93-1.25] | 1.01 [0.87-1.18] |
| W29 | 3,030 | 50,997,270 | 0.06 | 0.06 | 0.06 | 0.00 | 0.00 | 0.00 | 0.99 [0.87-1.13] | 0.96 [0.84-1.10] |
| W31 | 2,670 | 50,982,250 | 0.05 | 0.05 | 0.05 | 0.00 | 0.00 | 0.00 | 0.95 [0.82-1.09] | 0.94 [0.82-1.08] |
| W44 | 6,640 | 50,952,150 | 0.16 | 0.12 | 0.12 | 0.04 | 0.04 | 0.02 | 1.30 [1.19-1.41]* | 1.18 [1.08-1.28]* |
| W45 | 2,840 | 50,978,850 | 0.06 | 0.06 | 0.05 | 0.00 | 0.00 | 0.00 | 1.08 [0.95-1.23] | 0.99 [0.87-1.13] |
| W46 | 510 | 51,058,690 | 0.01 | 0.01 | 0.01 | 0.00 | 0.00 | 0.00 | 1.32 [0.99-1.75] | 1.05 [0.78-1.41] |
| W49 | 830 | 51,036,320 | 0.02 | 0.02 | 0.02 | 0.00 | 0.00 | 0.00 | 1.32 [1.05-1.66] | 1.25 [0.99-1.58] |
| W50 | 1,510 | 51,008,220 | 0.03 | 0.03 | 0.03 | 0.01 | 0.01 | 0.00 | 1.23 [1.03-1.45] | 1.11 [0.93-1.33] |
| W51 | 1,100 | 51,024,990 | 0.02 | 0.02 | 0.02 | 0.00 | 0.00 | 0.00 | 1.10 [0.89-1.35] | 1.06 [0.85-1.31] |
| W54 | 5,280 | 50,977,320 | 0.12 | 0.10 | 0.10 | 0.01 | 0.01 | 0.00 | 1.13 [1.03-1.24]* | 1.04 [0.95-1.15] |
| W55 | 3,450 | 51,008,430 | 0.07 | 0.07 | 0.07 | 0.00 | 0.00 | −0.01 | 0.97 [0.86-1.09] | 0.88 [0.78-1.00] |
| W57 | 3,400 | 51,000,210 | 0.09 | 0.06 | 0.06 | 0.03 | 0.03 | 0.02 | 1.42 [1.27-1.58]* | 1.28 [1.14-1.43]* |
| W60 | 410 | 51,055,620 | 0.01 | 0.01 | 0.01 | 0.00 | 0.00 | 0.00 | 1.46 [1.07-1.99] | 1.38 [1.01-1.89] |
| W64 | 900 | 51,054,220 | 0.02 | 0.02 | 0.02 | 0.00 | 0.00 | 0.00 | 1.13 [0.90-1.43] | 1.06 [0.84-1.35] |
| W78 | 3,490 | 51,054,280 | 0.08 | 0.07 | 0.07 | 0.01 | 0.01 | 0.00 | 1.10 [0.97-1.25] | 1.05 [0.92-1.19] |
| W79 | 4,770 | 51,038,320 | 0.11 | 0.09 | 0.09 | 0.02 | 0.02 | 0.01 | 1.19 [1.08-1.32]* | 1.13 [1.02-1.25] |
| W80 | 1,960 | 51,057,350 | 0.05 | 0.04 | 0.04 | 0.01 | 0.01 | 0.01 | 1.27 [1.08-1.50]* | 1.15 [0.98-1.36] |
| W84 | 2,070 | 51,057,600 | 0.05 | 0.04 | 0.04 | 0.01 | 0.00 | 0.00 | 1.08 [0.92-1.27] | 1.03 [0.87-1.21] |

| | | | | | | | | | | |
|---|---|---|---|---|---|---|---|---|---|---|
| X09 | 520 | 51,052,100 | 0.01 | 0.01 | 0.01 | 0.00 | 0.00 | 0.00 | 1.11 [0.82-1.51] | 1.05 [0.77-1.43] |
| X10 | 940 | 51,048,790 | 0.02 | 0.02 | 0.02 | 0.00 | 0.00 | 0.00 | 1.19 [0.95-1.50] | 1.10 [0.88-1.39] |
| X12 | 810 | 51,051,350 | 0.02 | 0.02 | 0.02 | 0.00 | 0.00 | 0.00 | 1.09 [0.86-1.40] | 0.99 [0.77-1.28] |
| X16 | 800 | 51,055,590 | 0.02 | 0.02 | 0.02 | 0.00 | 0.00 | 0.00 | 1.04 [0.80-1.34] | 0.98 [0.75-1.27] |
| X23 | 860 | 51,045,630 | 0.02 | 0.02 | 0.02 | 0.00 | 0.00 | 0.00 | 1.13 [0.89-1.42] | 1.04 [0.82-1.31] |
| X31 | 1,230 | 51,056,720 | 0.02 | 0.02 | 0.02 | 0.00 | 0.00 | 0.00 | 1.00 [0.80-1.23] | 0.99 [0.80-1.24] |
| X40 | 4,560 | 50,955,290 | 0.11 | 0.09 | 0.09 | 0.02 | 0.02 | 0.01 | 1.26 [1.14-1.39]* | 1.07 [0.96-1.18] |
| X41 | 3,890 | 50,955,540 | 0.09 | 0.07 | 0.07 | 0.02 | 0.02 | 0.01 | 1.29 [1.16-1.43]* | 1.08 [0.96-1.20] |
| X42 | 4,580 | 51,009,200 | 0.12 | 0.08 | 0.08 | 0.03 | 0.03 | 0.01 | 1.38 [1.25-1.52]* | 1.11 [1.00-1.23] |
| X43 | 1,060 | 51,049,480 | 0.02 | 0.02 | 0.02 | 0.00 | 0.00 | 0.00 | 1.16 [0.93-1.45] | 1.03 [0.82-1.29] |
| X44 | 6,190 | 50,979,200 | 0.15 | 0.11 | 0.12 | 0.04 | 0.04 | 0.01 | 1.30 [1.19-1.42]* | 1.11 [1.01-1.21] |
| X45 | 1,170 | 51,015,360 | 0.03 | 0.02 | 0.02 | 0.00 | 0.01 | 0.00 | 1.23 [1.02-1.50] | 1.00 [0.81-1.22] |
| X47 | 760 | 51,048,690 | 0.02 | 0.01 | 0.01 | 0.00 | 0.00 | 0.00 | 1.17 [0.91-1.51] | 1.12 [0.86-1.45] |
| X49 | 1,420 | 51,028,550 | 0.03 | 0.03 | 0.03 | 0.01 | 0.01 | 0.01 | 1.30 [1.09-1.55]* | 1.17 [0.98-1.40] |
| X50 | 10,070 | 50,877,430 | 0.22 | 0.19 | 0.19 | 0.03 | 0.03 | 0.01 | 1.15 [1.08-1.24]* | 1.06 [0.99-1.14] |
| X51 | 970 | 51,045,260 | 0.02 | 0.02 | 0.02 | 0.00 | 0.00 | 0.00 | 1.16 [0.93-1.45] | 1.10 [0.88-1.38] |
| X58 | 3,130 | 51,028,680 | 0.08 | 0.06 | 0.06 | 0.02 | 0.02 | 0.02 | 1.39 [1.23-1.56]* | 1.26 [1.11-1.42]* |
| X59 | 46,780 | 50,405,730 | 1.14 | 0.88 | 0.90 | 0.26 | 0.24 | 0.16 | 1.26 [1.22-1.30]* | 1.16 [1.12-1.20]* |
| X60 | 12,520 | 50,581,340 | 0.30 | 0.24 | 0.24 | 0.07 | 0.07 | 0.03 | 1.29 [1.21-1.36]* | 1.09 [1.03-1.16]* |
| X61 | 12,390 | 50,572,330 | 0.30 | 0.23 | 0.23 | 0.07 | 0.07 | 0.02 | 1.32 [1.25-1.40]* | 1.08 [1.02-1.15] |
| X62 | 6,950 | 50,886,530 | 0.17 | 0.13 | 0.13 | 0.04 | 0.04 | 0.00 | 1.29 [1.20-1.40]* | 1.02 [0.94-1.11] |
| X63 | 1,750 | 51,010,940 | 0.04 | 0.03 | 0.03 | 0.01 | 0.01 | 0.00 | 1.26 [1.08-1.48]* | 1.01 [0.85-1.19] |
| X64 | 5,800 | 50,866,930 | 0.15 | 0.11 | 0.11 | 0.04 | 0.04 | 0.01 | 1.37 [1.26-1.48]* | 1.10 [1.01-1.20] |
| X65 | 8,940 | 50,748,800 | 0.21 | 0.17 | 0.17 | 0.04 | 0.05 | 0.01 | 1.28 [1.20-1.37]* | 1.04 [0.97-1.12] |
| X78 | 2,530 | 50,992,350 | 0.06 | 0.05 | 0.05 | 0.01 | 0.01 | 0.00 | 1.30 [1.14-1.48]* | 1.05 [0.91-1.20] |
| X99 | 850 | 51,013,000 | 0.02 | 0.02 | 0.02 | 0.00 | 0.00 | 0.00 | 1.17 [0.93-1.47] | 1.03 [0.81-1.31] |
| Y00 | 800 | 51,023,090 | 0.02 | 0.02 | 0.01 | 0.00 | 0.00 | 0.00 | 1.25 [0.99-1.57] | 1.06 [0.83-1.35] |
| Y04 | 4,900 | 50,799,460 | 0.11 | 0.09 | 0.09 | 0.02 | 0.02 | 0.01 | 1.23 [1.12-1.35]* | 1.06 [0.96-1.17] |
| Y09 | 900 | 51,008,440 | 0.02 | 0.02 | 0.02 | 0.00 | 0.00 | 0.00 | 1.27 [1.02-1.58] | 1.09 [0.87-1.37] |
| Y40 | 15,490 | 50,903,870 | 0.43 | 0.28 | 0.28 | 0.16 | 0.15 | 0.11 | 1.53 [1.45-1.61]* | 1.36 [1.29-1.43]* |
| Y41 | 3,230 | 51,025,320 | 0.09 | 0.06 | 0.06 | 0.03 | 0.03 | 0.02 | 1.45 [1.29-1.62]* | 1.26 [1.12-1.43]* |
| Y42 | 13,520 | 50,929,340 | 0.35 | 0.25 | 0.25 | 0.11 | 0.10 | 0.05 | 1.40 [1.32-1.48]* | 1.18 [1.11-1.25]* |
| Y43 | 28,560 | 50,875,230 | 0.66 | 0.54 | 0.54 | 0.12 | 0.11 | 0.09 | 1.21 [1.16-1.26]* | 1.16 [1.11-1.21]* |
| Y44 | 10,240 | 50,980,380 | 0.25 | 0.19 | 0.20 | 0.05 | 0.05 | 0.03 | 1.24 [1.16-1.33]* | 1.14 [1.06-1.22]* |
| Y45 | 22,060 | 50,850,030 | 0.56 | 0.41 | 0.42 | 0.15 | 0.14 | 0.09 | 1.34 [1.28-1.40]* | 1.20 [1.14-1.25]* |
| Y46 | 3,100 | 51,027,090 | 0.08 | 0.06 | 0.06 | 0.02 | 0.02 | 0.01 | 1.35 [1.20-1.52]* | 1.20 [1.07-1.36]* |
| Y47 | 1,450 | 51,047,640 | 0.04 | 0.03 | 0.03 | 0.01 | 0.01 | 0.01 | 1.40 [1.17-1.67]* | 1.26 [1.05-1.51] |
| Y48 | 1,520 | 51,041,880 | 0.04 | 0.03 | 0.03 | 0.01 | 0.01 | 0.00 | 1.28 [1.08-1.52]* | 1.13 [0.94-1.35] |
| Y49 | 5,320 | 51,010,810 | 0.13 | 0.10 | 0.10 | 0.04 | 0.03 | 0.03 | 1.35 [1.23-1.48]* | 1.24 [1.13-1.36]* |
| Y51 | 9,110 | 50,980,270 | 0.21 | 0.17 | 0.17 | 0.04 | 0.04 | 0.03 | 1.24 [1.16-1.34]* | 1.16 [1.08-1.25]* |
| Y52 | 22,210 | 50,894,680 | 0.55 | 0.41 | 0.42 | 0.14 | 0.13 | 0.09 | 1.30 [1.24-1.36]* | 1.19 [1.13-1.24]* |
| Y53 | 3,970 | 51,036,550 | 0.10 | 0.07 | 0.08 | 0.03 | 0.03 | 0.02 | 1.37 [1.23-1.53]* | 1.20 [1.07-1.34]* |
| Y54 | 17,330 | 50,961,890 | 0.45 | 0.32 | 0.33 | 0.13 | 0.12 | 0.08 | 1.36 [1.29-1.43]* | 1.23 [1.17-1.30]* |
| Y55 | 960 | 51,049,740 | 0.03 | 0.02 | 0.02 | 0.01 | 0.01 | 0.01 | 1.55 [1.27-1.90]* | 1.24 [1.00-1.53] |
| Y56 | 1,470 | 51,036,640 | 0.05 | 0.02 | 0.03 | 0.02 | 0.02 | 0.02 | 1.88 [1.60-2.21]* | 1.61 [1.37-1.91]* |
| Y57 | 9,140 | 50,991,230 | 0.23 | 0.17 | 0.17 | 0.07 | 0.07 | 0.04 | 1.39 [1.29-1.49]* | 1.22 [1.13-1.31]* |
| Y59 | 720 | 51,055,770 | 0.02 | 0.01 | 0.01 | 0.01 | 0.01 | 0.00 | 1.44 [1.13-1.83]* | 1.29 [1.01-1.65] |
| Y60 | 11,130 | 50,963,400 | 0.27 | 0.21 | 0.21 | 0.06 | 0.05 | 0.04 | 1.25 [1.17-1.33]* | 1.16 [1.09-1.24]* |
| Y65 | 1,420 | 51,046,800 | 0.03 | 0.03 | 0.03 | 0.01 | 0.01 | 0.00 | 1.21 [1.01-1.45] | 1.10 [0.92-1.33] |
| Y71 | 690 | 51,053,850 | 0.02 | 0.01 | 0.01 | 0.01 | 0.01 | 0.01 | 1.56 [1.22-2.00]* | 1.43 [1.11-1.84]* |
| Y73 | 650 | 51,054,920 | 0.01 | 0.01 | 0.01 | 0.00 | 0.00 | 0.00 | 0.99 [0.75-1.30] | 0.90 [0.68-1.20] |
| Y77 | 850 | 51,054,290 | 0.02 | 0.02 | 0.02 | 0.00 | 0.00 | 0.00 | 0.99 [0.78-1.28] | 0.95 [0.74-1.22] |
| Y79 | 1,620 | 51,035,910 | 0.04 | 0.03 | 0.03 | 0.01 | 0.01 | 0.00 | 1.19 [1.00-1.40] | 1.11 [0.94-1.32] |
| Y82 | 510 | 51,054,200 | 0.01 | 0.01 | 0.01 | 0.00 | 0.00 | 0.00 | 1.31 [0.98-1.76] | 1.19 [0.88-1.60] |
| Y83 | 184,430 | 48,241,050 | 4.68 | 3.64 | 3.67 | 1.04 | 1.00 | 0.66 | 1.27 [1.25-1.29]* | 1.17 [1.15-1.18]* |
| Y84 | 46,100 | 50,704,500 | 1.10 | 0.87 | 0.89 | 0.23 | 0.21 | 0.14 | 1.24 [1.20-1.28]* | 1.14 [1.11-1.18]* |
| Y85 | 3,540 | 50,977,960 | 0.09 | 0.07 | 0.06 | 0.02 | 0.02 | 0.02 | 1.38 [1.24-1.54]* | 1.25 [1.12-1.40]* |
| Y86 | 6,050 | 50,903,380 | 0.14 | 0.11 | 0.11 | 0.03 | 0.03 | 0.02 | 1.24 [1.14-1.35]* | 1.13 [1.03-1.23]* |
| Y87 | 610 | 51,039,800 | 0.02 | 0.01 | 0.01 | 0.00 | 0.00 | 0.00 | 1.39 [1.07-1.80] | 1.23 [0.94-1.61] |
| Y88 | 1,490 | 51,037,240 | 0.03 | 0.03 | 0.03 | 0.01 | 0.01 | 0.00 | 1.24 [1.04-1.47] | 1.06 [0.88-1.27] |
| Y90 | 640 | 51,053,610 | 0.01 | 0.01 | 0.01 | 0.00 | 0.00 | 0.00 | 1.19 [0.91-1.55] | 1.05 [0.80-1.38] |
| Y91 | 1,270 | 51,030,540 | 0.03 | 0.02 | 0.02 | 0.01 | 0.01 | 0.00 | 1.27 [1.05-1.53] | 1.10 [0.91-1.34] |
| Y95 | 115,490 | 50,790,640 | 2.82 | 2.16 | 2.26 | 0.66 | 0.55 | 0.33 | 1.24 [1.22-1.27]* | 1.13 [1.11-1.16]* |
| 40+ - Z00-Z99 | | | | | | | | | | |
| Z00 | 3,400 | 50,974,830 | 0.08 | 0.06 | 0.06 | 0.01 | 0.02 | 0.01 | 1.25 [1.11-1.40]* | 1.15 [1.02-1.29] |
| Z01 | 9,340 | 50,494,740 | 0.24 | 0.17 | 0.17 | 0.07 | 0.07 | 0.05 | 1.43 [1.34-1.53]* | 1.26 [1.18-1.35]* |
| Z02 | 14,350 | 50,920,290 | 0.32 | 0.27 | 0.27 | 0.05 | 0.05 | 0.02 | 1.16 [1.10-1.24]* | 1.08 [1.01-1.14] |
| Z03 | 108,930 | 49,056,990 | 2.81 | 2.09 | 2.12 | 0.72 | 0.70 | 0.45 | 1.33 [1.30-1.36]* | 1.19 [1.17-1.22]* |
| Z04 | 13,000 | 50,589,870 | 0.32 | 0.24 | 0.24 | 0.08 | 0.08 | 0.06 | 1.32 [1.24-1.40]* | 1.22 [1.15-1.29]* |
| Z08 | 42,720 | 50,289,960 | 0.91 | 0.84 | 0.84 | 0.07 | 0.07 | 0.04 | 1.08 [1.05-1.12]* | 1.05 [1.02-1.09]* |
| Z09 | 70,680 | 49,668,620 | 1.74 | 1.35 | 1.35 | 0.39 | 0.39 | 0.25 | 1.29 [1.26-1.32]* | 1.17 [1.14-1.20]* |
| Z11 | 184,560 | 50,698,810 | 4.42 | 3.47 | 3.61 | 0.95 | 0.82 | 0.42 | 1.23 [1.21-1.25]* | 1.10 [1.09-1.12]* |
| Z12 | 60,680 | 50,457,040 | 1.35 | 1.17 | 1.17 | 0.18 | 0.17 | 0.14 | 1.15 [1.12-1.18]* | 1.11 [1.08-1.15]* |
| Z13 | 114,180 | 49,557,230 | 2.82 | 2.19 | 2.23 | 0.63 | 0.60 | 0.38 | 1.27 [1.24-1.29]* | 1.15 [1.13-1.18]* |
| Z20 | 6,930 | 51,041,130 | 0.17 | 0.13 | 0.14 | 0.04 | 0.03 | 0.02 | 1.25 [1.15-1.36]* | 1.13 [1.04-1.24]* |
| Z22 | 45,470 | 50,602,950 | 1.24 | 0.83 | 0.84 | 0.42 | 0.40 | 0.31 | 1.48 [1.43-1.53]* | 1.34 [1.30-1.38]* |
| Z29 | 27,530 | 50,822,190 | 0.67 | 0.51 | 0.52 | 0.16 | 0.15 | 0.10 | 1.29 [1.23-1.34]* | 1.17 [1.12-1.22]* |
| Z30 | 16,350 | 48,995,000 | 0.39 | 0.32 | 0.32 | 0.07 | 0.06 | 0.04 | 1.20 [1.14-1.26]* | 1.11 [1.05-1.17]* |
| Z33 | 1,370 | 50,579,860 | 0.03 | 0.03 | 0.03 | 0.01 | 0.01 | 0.00 | 1.19 [1.00-1.42] | 1.08 [0.90-1.29] |
| Z34 | 3,070 | 49,929,150 | 0.06 | 0.06 | 0.06 | 0.00 | 0.00 | 0.00 | 0.97 [0.86-1.09] | 0.96 [0.84-1.09] |
| Z35 | 9,440 | 50,136,550 | 0.19 | 0.19 | 0.20 | 0.00 | −0.01 | −0.01 | 0.96 [0.89-1.03] | 0.97 [0.90-1.05] |
| Z36 | 940 | 50,378,890 | 0.02 | 0.02 | 0.02 | 0.00 | 0.00 | 0.00 | 1.10 [0.89-1.36] | 1.13 [0.91-1.42] |
| Z37 | 7,950 | 44,231,080 | 0.27 | 0.16 | 0.26 | 0.11 | 0.02 | 0.03 | 1.06 [0.99-1.13] | 1.12 [1.04-1.20]* |
| Z39 | 1,310 | 50,519,870 | 0.03 | 0.03 | 0.03 | 0.00 | 0.00 | 0.00 | 1.03 [0.86-1.24] | 1.06 [0.88-1.29] |
| Z40 | 4,140 | 50,990,660 | 0.09 | 0.08 | 0.08 | 0.01 | 0.01 | 0.01 | 1.11 [1.00-1.24] | 1.06 [0.96-1.19] |
| Z41 | 2,520 | 50,959,890 | 0.06 | 0.05 | 0.05 | 0.02 | 0.02 | 0.01 | 1.34 [1.18-1.52]* | 1.23 [1.08-1.40]* |
| Z42 | 11,660 | 50,812,640 | 0.26 | 0.22 | 0.22 | 0.03 | 0.03 | 0.02 | 1.15 [1.09-1.23]* | 1.10 [1.03-1.17]* |
| Z43 | 17,470 | 50,767,660 | 0.39 | 0.33 | 0.33 | 0.06 | 0.06 | 0.04 | 1.18 [1.12-1.24]* | 1.12 [1.07-1.19]* |
| Z45 | 38,570 | 50,608,490 | 0.89 | 0.73 | 0.74 | 0.16 | 0.15 | 0.09 | 1.21 [1.16-1.25]* | 1.11 [1.07-1.15]* |
| Z46 | 50,490 | 50,291,920 | 1.12 | 0.98 | 0.98 | 0.14 | 0.14 | 0.09 | 1.14 [1.10-1.17]* | 1.08 [1.05-1.12]* |
| Z47 | 16,710 | 50,452,450 | 0.38 | 0.32 | 0.32 | 0.06 | 0.07 | 0.04 | 1.21 [1.15-1.28]* | 1.13 [1.07-1.19]* |
| Z48 | 10,030 | 50,797,130 | 0.23 | 0.19 | 0.19 | 0.04 | 0.04 | 0.03 | 1.22 [1.15-1.31]* | 1.15 [1.08-1.23]* |
| Z49 | 7,880 | 50,970,560 | 0.21 | 0.14 | 0.14 | 0.07 | 0.06 | 0.05 | 1.44 [1.34-1.55]* | 1.30 [1.21-1.40]* |
| Z50 | 305,560 | 49,223,110 | 7.39 | 5.95 | 6.22 | 1.44 | 1.17 | 0.73 | 1.19 [1.17-1.20]* | 1.11 [1.09-1.12]* |
| Z51 | 347,970 | 48,474,320 | 8.46 | 6.90 | 7.05 | 1.56 | 1.41 | 0.93 | 1.20 [1.19-1.21]* | 1.12 [1.11-1.14]* |
| Z52 | 1,370 | 51,028,040 | 0.03 | 0.03 | 0.03 | 0.00 | 0.00 | 0.00 | 0.97 [0.80-1.18] | 0.99 [0.82-1.20] |
| Z53 | 255,900 | 46,463,760 | 6.87 | 5.21 | 5.24 | 1.66 | 1.63 | 1.09 | 1.31 [1.29-1.33]* | 1.19 [1.17-1.20]* |
| Z54 | 2,850 | 50,995,890 | 0.07 | 0.05 | 0.05 | 0.02 | 0.02 | 0.02 | 1.40 [1.24-1.59]* | 1.33 [1.17-1.51]* |
| Z56 | 2,280 | 51,023,250 | 0.05 | 0.04 | 0.04 | 0.01 | 0.01 | 0.00 | 1.14 [0.99-1.32] | 0.99 [0.85-1.14] |
| Z57 | 11,760 | 50,992,300 | 0.28 | 0.22 | 0.22 | 0.06 | 0.06 | 0.04 | 1.27 [1.19-1.35]* | 1.17 [1.10-1.25]* |
| Z58 | 5,770 | 51,033,050 | 0.14 | 0.11 | 0.11 | 0.03 | 0.03 | 0.01 | 1.25 [1.14-1.37]* | 1.11 [1.01-1.22] |
| Z59 | 5,820 | 50,991,540 | 0.13 | 0.11 | 0.11 | 0.01 | 0.01 | 0.00 | 1.11 [1.01-1.21] | 1.00 [0.91-1.10] |
| Z60 | 101,090 | 49,981,860 | 2.35 | 1.95 | 1.99 | 0.40 | 0.36 | 0.23 | 1.18 [1.16-1.21]* | 1.11 [1.08-1.13]* |
| Z61 | 830 | 51,048,640 | 0.02 | 0.02 | 0.02 | 0.01 | 0.00 | 0.00 | 1.31 [1.05-1.64] | 1.05 [0.83-1.32] |
| Z63 | 17,920 | 50,867,620 | 0.42 | 0.34 | 0.34 | 0.09 | 0.08 | 0.03 | 1.23 [1.17-1.30]* | 1.08 [1.02-1.14]* |
| Z65 | 970 | 51,044,870 | 0.02 | 0.02 | 0.02 | 0.00 | 0.00 | 0.00 | 1.18 [0.95-1.46] | 1.00 [0.80-1.25] |
| Z71 | 22,080 | 50,826,660 | 0.51 | 0.42 | 0.43 | 0.09 | 0.08 | 0.04 | 1.18 [1.12-1.23]* | 1.09 [1.04-1.15]* |

| | | | | | | | | | | |
|---|---|---|---|---|---|---|---|---|---|---|
| Z72 | 151,770 | 46,608,240 | 3.79 | 3.14 | 3.10 | 0.65 | 0.69 | 0.40 | 1.22 [1.20-1.24]* | 1.12 [1.10-1.14]* |
| Z73 | 51,700 | 50,830,510 | 1.21 | 0.98 | 1.02 | 0.23 | 0.19 | 0.11 | 1.19 [1.15-1.22]* | 1.10 [1.07-1.14]* |
| Z74 | 41,560 | 50,823,900 | 0.95 | 0.79 | 0.82 | 0.17 | 0.14 | 0.10 | 1.17 [1.13-1.21]* | 1.11 [1.08-1.15]* |
| Z75 | 80,540 | 50,705,850 | 1.83 | 1.54 | 1.63 | 0.30 | 0.20 | 0.13 | 1.12 [1.09-1.15]* | 1.07 [1.05-1.10]* |
| Z76 | 1,600 | 50,982,820 | 0.04 | 0.03 | 0.03 | 0.01 | 0.01 | 0.01 | 1.43 [1.21-1.69]* | 1.36 [1.15-1.61]* |
| Z80 | 76,350 | 49,921,660 | 1.72 | 1.49 | 1.50 | 0.23 | 0.22 | 0.12 | 1.15 [1.12-1.18]* | 1.08 [1.05-1.10]* |
| Z81 | 2,460 | 51,024,600 | 0.06 | 0.05 | 0.04 | 0.01 | 0.01 | 0.01 | 1.33 [1.16-1.51]* | 1.14 [1.00-1.31] |
| Z82 | 129,130 | 48,875,960 | 3.13 | 2.54 | 2.53 | 0.59 | 0.59 | 0.32 | 1.23 [1.21-1.26]* | 1.11 [1.09-1.14]* |
| Z83 | 30,150 | 50,623,530 | 0.70 | 0.57 | 0.58 | 0.13 | 0.12 | 0.07 | 1.20 [1.16-1.25]* | 1.11 [1.07-1.15]* |
| Z84 | 1,860 | 51,021,690 | 0.05 | 0.03 | 0.03 | 0.01 | 0.01 | 0.01 | 1.41 [1.22-1.64]* | 1.30 [1.12-1.52]* |
| Z85 | 308,340 | 47,696,830 | 7.49 | 6.24 | 6.41 | 1.25 | 1.08 | 0.70 | 1.17 [1.15-1.18]* | 1.10 [1.09-1.12]* |
| Z86 | 821,250 | 42,469,790 | 23.96 | 18.33 | 18.79 | 5.63 | 5.17 | 3.25 | 1.28 [1.27-1.29]* | 1.16 [1.15-1.17]* |
| Z87 | 371,500 | 46,470,050 | 9.99 | 7.56 | 7.74 | 2.43 | 2.25 | 1.36 | 1.29 [1.28-1.31]* | 1.16 [1.14-1.17]* |
| Z88 | 392,750 | 45,656,180 | 11.78 | 7.91 | 8.09 | 3.87 | 3.70 | 2.56 | 1.46 [1.44-1.47]* | 1.28 [1.26-1.29]* |
| Z89 | 14,220 | 50,938,610 | 0.35 | 0.26 | 0.27 | 0.08 | 0.08 | 0.05 | 1.29 [1.22-1.36]* | 1.15 [1.09-1.22]* |
| Z90 | 246,980 | 48,797,600 | 6.03 | 4.85 | 4.98 | 1.18 | 1.05 | 0.57 | 1.21 [1.20-1.23]* | 1.10 [1.09-1.12]* |
| Z91 | 120,030 | 49,676,090 | 3.47 | 2.19 | 2.23 | 1.28 | 1.24 | 0.86 | 1.56 [1.53-1.59]* | 1.33 [1.30-1.36]* |
| Z92 | 617,510 | 45,851,230 | 16.36 | 12.85 | 13.28 | 3.51 | 3.07 | 1.78 | 1.23 [1.22-1.24]* | 1.12 [1.11-1.13]* |
| Z93 | 57,190 | 50,568,990 | 1.31 | 1.09 | 1.11 | 0.22 | 0.21 | 0.15 | 1.19 [1.16-1.22]* | 1.13 [1.09-1.16]* |
| Z94 | 9,560 | 50,901,700 | 0.24 | 0.18 | 0.17 | 0.07 | 0.07 | 0.06 | 1.40 [1.31-1.50]* | 1.30 [1.21-1.39]* |
| Z95 | 206,250 | 48,662,020 | 5.12 | 4.05 | 4.15 | 1.07 | 0.96 | 0.55 | 1.23 [1.21-1.25]* | 1.12 [1.10-1.14]* |
| Z96 | 400,100 | 47,405,490 | 10.20 | 8.06 | 8.38 | 2.14 | 1.82 | 1.25 | 1.22 [1.20-1.23]* | 1.14 [1.13-1.15]* |
| Z97 | 31,530 | 50,786,970 | 0.77 | 0.59 | 0.61 | 0.18 | 0.16 | 0.11 | 1.26 [1.22-1.31]* | 1.16 [1.12-1.21]* |
| Z98 | 46,690 | 50,543,240 | 1.14 | 0.88 | 0.89 | 0.26 | 0.25 | 0.14 | 1.28 [1.25-1.32]* | 1.14 [1.10-1.17]* |
| Z99 | 61,140 | 50,651,900 | 1.62 | 1.12 | 1.15 | 0.51 | 0.48 | 0.30 | 1.41 [1.38-1.45]* | 1.23 [1.19-1.26]* |
| <18 - A00-B99 | | | | | | | | | | |
| A02 | 640 | 46,396,080 | 0.02 | 0.01 | 0.01 | 0.00 | 0.00 | 0.00 | 1.25 [0.97-1.61] | 1.26 [0.97-1.63] |
| A04 | 3,620 | 46,359,160 | 0.08 | 0.08 | 0.08 | 0.01 | 0.01 | 0.00 | 1.08 [0.97-1.21] | 1.03 [0.91-1.15] |
| A07 | 380 | 46,408,660 | 0.01 | 0.01 | 0.01 | 0.00 | 0.00 | 0.00 | 1.52 [1.10-2.10] | 1.44 [1.03-2.00] |
| A08 | 40,120 | 44,886,310 | 0.98 | 0.87 | 0.86 | 0.11 | 0.12 | 0.10 | 1.14 [1.10-1.17]* | 1.11 [1.08-1.15]* |
| A09 | 43,710 | 45,776,210 | 1.09 | 0.92 | 0.92 | 0.17 | 0.17 | 0.13 | 1.19 [1.15-1.22]* | 1.14 [1.10-1.17]* |
| A38 | 1,980 | 46,393,700 | 0.05 | 0.04 | 0.04 | 0.00 | 0.00 | 0.00 | 1.05 [0.91-1.22] | 1.03 [0.89-1.20] |
| A39 | 1,910 | 46,334,260 | 0.04 | 0.04 | 0.04 | 0.00 | 0.00 | 0.00 | 1.05 [0.91-1.22] | 1.04 [0.89-1.21] |
| A40 | 1,090 | 46,388,560 | 0.02 | 0.02 | 0.02 | 0.00 | 0.00 | 0.00 | 1.11 [0.90-1.35] | 1.05 [0.86-1.30] |
| A41 | 10,020 | 46,237,800 | 0.23 | 0.21 | 0.21 | 0.02 | 0.02 | 0.02 | 1.10 [1.03-1.18]* | 1.08 [1.01-1.16] |
| A49 | 3,100 | 46,336,740 | 0.07 | 0.06 | 0.06 | 0.01 | 0.01 | 0.01 | 1.19 [1.06-1.34]* | 1.17 [1.04-1.32] |
| A87 | 1,420 | 46,372,210 | 0.03 | 0.03 | 0.03 | 0.00 | 0.01 | 0.01 | 1.27 [1.06-1.51]* | 1.24 [1.03-1.49] |
| B00 | 5,190 | 46,273,200 | 0.24 | 0.08 | 0.08 | 0.16 | 0.16 | 0.16 | 3.13 [2.90-3.38]* | 2.97 [2.74-3.22]* |
| B01 | 8,250 | 46,179,670 | 0.25 | 0.16 | 0.15 | 0.09 | 0.10 | 0.09 | 1.63 [1.53-1.74]* | 1.58 [1.48-1.69]* |
| B02 | 1,410 | 46,399,680 | 0.04 | 0.03 | 0.03 | 0.01 | 0.01 | 0.01 | 1.41 [1.20-1.67]* | 1.35 [1.13-1.60]* |
| B07 | 1,380 | 46,395,490 | 0.04 | 0.03 | 0.03 | 0.01 | 0.01 | 0.01 | 1.45 [1.22-1.71]* | 1.32 [1.11-1.58]* |
| B08 | 2,920 | 46,352,060 | 0.08 | 0.06 | 0.05 | 0.03 | 0.03 | 0.03 | 1.53 [1.37-1.71]* | 1.48 [1.32-1.66]* |
| B09 | 1,340 | 46,371,370 | 0.03 | 0.03 | 0.03 | 0.01 | 0.00 | 0.00 | 1.16 [0.97-1.38] | 1.14 [0.95-1.36] |
| B27 | 5,600 | 46,358,420 | 0.13 | 0.12 | 0.12 | 0.02 | 0.02 | 0.01 | 1.16 [1.06-1.26]* | 1.10 [1.01-1.21] |
| B34 | 114,870 | 43,223,860 | 3.72 | 2.37 | 2.36 | 1.35 | 1.36 | 1.28 | 1.57 [1.55-1.60]* | 1.53 [1.50-1.55]* |
| B35 | 860 | 46,403,080 | 0.03 | 0.02 | 0.02 | 0.01 | 0.01 | 0.01 | 1.47 [1.19-1.82]* | 1.38 [1.10-1.72]* |
| B37 | 9,010 | 46,192,150 | 0.20 | 0.19 | 0.18 | 0.01 | 0.02 | 0.01 | 1.11 [1.04-1.20]* | 1.06 [0.99-1.14] |
| B80 | 1,640 | 46,396,700 | 0.04 | 0.03 | 0.03 | 0.01 | 0.01 | 0.00 | 1.18 [1.00-1.38] | 1.12 [0.95-1.32] |
| B85 | 570 | 46,406,160 | 0.01 | 0.01 | 0.01 | 0.00 | 0.00 | 0.00 | 0.95 [0.71-1.26] | 0.95 [0.71-1.26] |
| B94 | 570 | 46,409,480 | 0.01 | 0.01 | 0.01 | 0.00 | 0.00 | 0.00 | 1.23 [0.93-1.61] | 1.25 [0.94-1.65] |
| B95 | 19,300 | 45,943,210 | 0.56 | 0.38 | 0.37 | 0.17 | 0.19 | 0.17 | 1.50 [1.43-1.57]* | 1.45 [1.39-1.52]* |
| B96 | 22,970 | 45,843,080 | 0.52 | 0.50 | 0.49 | 0.02 | 0.03 | 0.02 | 1.07 [1.02-1.12]* | 1.04 [1.00-1.09] |
| B97 | 24,830 | 45,750,600 | 0.67 | 0.51 | 0.50 | 0.16 | 0.17 | 0.15 | 1.34 [1.29-1.39]* | 1.28 [1.23-1.34]* |
| B98 | 1,120 | 46,411,550 | 0.03 | 0.02 | 0.02 | 0.00 | 0.00 | 0.00 | 1.01 [0.82-1.25] | 1.00 [0.80-1.24] |
| B99 | 570 | 46,409,480 | 0.01 | 0.01 | 0.01 | 0.00 | 0.00 | 0.00 | 1.25 [0.95-1.63] | 1.18 [0.89-1.56] |
| <18 - C00-D49 | | | | | | | | | | |
| C71 | 1,170 | 46,403,980 | 0.02 | 0.03 | 0.02 | 0.00 | 0.00 | 0.00 | 0.99 [0.81-1.21] | 1.02 [0.83-1.24] |
| C77 | 640 | 46,414,630 | 0.01 | 0.01 | 0.01 | 0.00 | 0.00 | 0.00 | 1.14 [0.86-1.51] | 1.14 [0.86-1.53] |
| C78 | 710 | 46,414,060 | 0.01 | 0.02 | 0.01 | 0.00 | 0.00 | 0.00 | 1.01 [0.78-1.32] | 1.03 [0.78-1.35] |
| C79 | 810 | 46,413,510 | 0.02 | 0.02 | 0.02 | 0.00 | 0.00 | 0.00 | 0.98 [0.77-1.25] | 0.99 [0.77-1.27] |
| C81 | 720 | 46,410,670 | 0.02 | 0.01 | 0.01 | 0.00 | 0.01 | 0.00 | 1.41 [1.11-1.79]* | 1.33 [1.04-1.71] |
| C91 | 1,520 | 46,392,600 | 0.03 | 0.03 | 0.03 | 0.00 | 0.00 | 0.00 | 0.97 [0.81-1.15] | 0.97 [0.81-1.15] |
| C92 | 470 | 46,412,240 | 0.01 | 0.01 | 0.01 | 0.00 | 0.00 | 0.00 | 1.09 [0.81-1.49] | 1.10 [0.80-1.51] |
| D06 | 1,880 | 46,404,400 | 0.03 | 0.04 | 0.03 | −0.01 | 0.00 | 0.00 | 0.92 [0.77-1.10] | 0.92 [0.77-1.11] |
| D10 | 800 | 46,407,280 | 0.02 | 0.02 | 0.02 | 0.00 | 0.00 | 0.00 | 1.14 [0.90-1.45] | 1.13 [0.88-1.44] |
| D12 | 690 | 46,412,000 | 0.02 | 0.01 | 0.01 | 0.00 | 0.00 | 0.00 | 1.26 [0.96-1.64] | 1.22 [0.93-1.61] |
| D16 | 2,610 | 46,386,870 | 0.06 | 0.06 | 0.06 | 0.01 | 0.00 | 0.00 | 1.07 [0.94-1.22] | 1.06 [0.92-1.21] |
| D17 | 2,390 | 46,387,140 | 0.05 | 0.05 | 0.05 | 0.00 | 0.01 | 0.00 | 1.13 [0.98-1.30] | 1.07 [0.93-1.23] |
| D18 | 3,470 | 46,275,080 | 0.08 | 0.07 | 0.07 | 0.01 | 0.01 | 0.01 | 1.14 [1.02-1.28] | 1.13 [1.01-1.27] |
| D22 | 11,280 | 45,851,480 | 0.26 | 0.24 | 0.23 | 0.02 | 0.03 | 0.02 | 1.15 [1.08-1.22]* | 1.11 [1.04-1.18]* |
| D23 | 6,100 | 46,295,700 | 0.14 | 0.13 | 0.12 | 0.01 | 0.02 | 0.01 | 1.14 [1.05-1.24]* | 1.11 [1.02-1.21] |
| D24 | 3,050 | 46,390,290 | 0.07 | 0.07 | 0.06 | 0.00 | 0.01 | 0.01 | 1.13 [1.00-1.28] | 1.12 [0.98-1.27] |
| D25 | 920 | 46,413,240 | 0.02 | 0.02 | 0.02 | 0.00 | 0.00 | 0.00 | 1.14 [0.89-1.44] | 1.12 [0.87-1.44] |
| D27 | 1,690 | 46,405,030 | 0.04 | 0.04 | 0.03 | 0.00 | 0.00 | 0.00 | 1.07 [0.90-1.28] | 1.04 [0.87-1.25] |
| D31 | 530 | 46,409,600 | 0.01 | 0.01 | 0.01 | 0.00 | 0.00 | 0.00 | 1.33 [1.01-1.74] | 1.29 [0.98-1.70] |
| D36 | 1,010 | 46,403,900 | 0.02 | 0.02 | 0.02 | 0.00 | 0.00 | 0.00 | 1.04 [0.84-1.29] | 0.99 [0.79-1.24] |
| D43 | 680 | 46,410,070 | 0.02 | 0.01 | 0.01 | 0.00 | 0.00 | 0.00 | 1.22 [0.95-1.57] | 1.23 [0.95-1.59] |
| D47 | 390 | 46,414,010 | 0.01 | 0.01 | 0.01 | 0.00 | 0.00 | 0.00 | 1.36 [0.98-1.88] | 1.30 [0.92-1.82] |
| D48 | 1,120 | 46,401,200 | 0.02 | 0.02 | 0.02 | 0.00 | 0.00 | 0.00 | 0.93 [0.75-1.15] | 0.92 [0.74-1.15] |
| <18 - D50-D89 | | | | | | | | | | |
| D50 | 15,810 | 46,230,920 | 0.41 | 0.32 | 0.32 | 0.08 | 0.09 | 0.08 | 1.28 [1.21-1.35]* | 1.24 [1.18-1.31]* |
| D51 | 570 | 46,414,090 | 0.01 | 0.01 | 0.01 | 0.00 | 0.00 | 0.00 | 1.43 [1.07-1.91] | 1.33 [0.99-1.80] |
| D56 | 1,930 | 46,393,150 | 0.05 | 0.04 | 0.04 | 0.01 | 0.01 | 0.01 | 1.34 [1.16-1.56]* | 1.31 [1.12-1.53]* |
| D57 | 3,800 | 46,347,970 | 0.11 | 0.08 | 0.08 | 0.03 | 0.03 | 0.03 | 1.36 [1.23-1.51]* | 1.36 [1.22-1.51]* |
| D58 | 590 | 46,403,380 | 0.01 | 0.01 | 0.01 | 0.00 | 0.00 | 0.00 | 1.21 [0.92-1.59] | 1.20 [0.90-1.58] |
| D59 | 530 | 46,406,910 | 0.01 | 0.01 | 0.01 | 0.00 | 0.00 | 0.00 | 1.27 [0.96-1.67] | 1.25 [0.94-1.67] |
| D61 | 1,150 | 46,405,460 | 0.03 | 0.02 | 0.02 | 0.00 | 0.00 | 0.00 | 1.07 [0.87-1.30] | 1.04 [0.84-1.27] |
| D62 | 1,070 | 46,409,860 | 0.02 | 0.02 | 0.02 | 0.00 | 0.00 | 0.00 | 0.99 [0.79-1.23] | 0.98 [0.78-1.23] |
| D63 | 450 | 46,412,650 | 0.01 | 0.01 | 0.01 | 0.00 | 0.00 | 0.00 | 1.18 [0.86-1.62] | 1.16 [0.83-1.60] |
| D64 | 14,860 | 46,229,200 | 0.35 | 0.31 | 0.30 | 0.04 | 0.05 | 0.04 | 1.17 [1.11-1.23]* | 1.14 [1.08-1.21]* |
| D68 | 2,870 | 46,379,830 | 0.07 | 0.06 | 0.06 | 0.00 | 0.01 | 0.00 | 1.09 [0.96-1.23] | 1.03 [0.90-1.17] |
| D69 | 12,100 | 46,220,060 | 0.28 | 0.26 | 0.26 | 0.02 | 0.02 | 0.02 | 1.06 [1.00-1.13] | 1.06 [1.00-1.13] |
| D70 | 5,720 | 46,329,550 | 0.13 | 0.12 | 0.12 | 0.01 | 0.02 | 0.01 | 1.14 [1.04-1.24]* | 1.11 [1.02-1.22] |
| D72 | 1,330 | 46,404,250 | 0.04 | 0.03 | 0.03 | 0.01 | 0.01 | 0.01 | 1.41 [1.18-1.68]* | 1.31 [1.10-1.58]* |
| D73 | 810 | 46,410,060 | 0.02 | 0.02 | 0.02 | 0.00 | 0.00 | 0.00 | 0.92 [0.72-1.18] | 0.95 [0.74-1.22] |
| D75 | 480 | 46,410,930 | 0.01 | 0.01 | 0.01 | 0.00 | 0.00 | 0.00 | 1.18 [0.86-1.62] | 1.12 [0.81-1.55] |
| D80 | 780 | 46,403,570 | 0.02 | 0.02 | 0.02 | 0.01 | 0.01 | 0.01 | 1.47 [1.18-1.84]* | 1.33 [1.06-1.68] |
| D84 | 580 | 46,409,000 | 0.02 | 0.01 | 0.01 | 0.01 | 0.01 | 0.01 | 1.76 [1.37-2.27]* | 1.59 [1.22-2.06]* |
| D89 | 410 | 46,414,590 | 0.01 | 0.01 | 0.01 | 0.00 | 0.00 | 0.00 | 1.53 [1.13-2.08]* | 1.37 [1.00-1.89] |
| <18 - E00-E89 | | | | | | | | | | |
| E03 | 7,130 | 46,337,800 | 0.16 | 0.15 | 0.14 | 0.01 | 0.03 | 0.03 | 1.20 [1.11-1.31]* | 1.18 [1.09-1.29]* |
| E04 | 780 | 46,411,490 | 0.02 | 0.02 | 0.01 | 0.00 | 0.00 | 0.00 | 1.24 [0.98-1.58] | 1.24 [0.97-1.59] |
| E05 | 1,810 | 46,404,840 | 0.04 | 0.04 | 0.03 | 0.00 | 0.01 | 0.01 | 1.28 [1.09-1.50]* | 1.24 [1.05-1.46] |
| E10 | 10,960 | 46,265,590 | 0.24 | 0.24 | 0.24 | 0.00 | −0.01 | −0.01 | 0.97 [0.91-1.04] | 0.96 [0.90-1.03] |
| E11 | 2,660 | 46,395,940 | 0.06 | 0.06 | 0.05 | 0.01 | 0.01 | 0.01 | 1.25 [1.09-1.43]* | 1.23 [1.08-1.41]* |

| | | | | | | | | | | |
|---|---|---|---|---|---|---|---|---|---|---|
| E14 | 1,180 | 46,404,520 | 0.02 | 0.03 | 0.03 | 0.00 | 0.00 | 0.00 | 0.92 [0.75-1.14] | 0.92 [0.74-1.14] |
| E16 | 7,370 | 46,304,330 | 0.17 | 0.16 | 0.15 | 0.01 | 0.01 | 0.01 | 1.07 [0.99-1.15] | 1.05 [0.97-1.14] |
| E22 | 750 | 46,410,010 | 0.02 | 0.02 | 0.01 | 0.00 | 0.01 | 0.00 | 1.35 [1.06-1.71] | 1.28 [1.00-1.63] |
| E23 | 2,050 | 46,390,370 | 0.05 | 0.04 | 0.04 | 0.01 | 0.01 | 0.01 | 1.24 [1.07-1.43]* | 1.18 [1.02-1.37] |
| E27 | 1,590 | 46,398,330 | 0.05 | 0.03 | 0.03 | 0.01 | 0.01 | 0.01 | 1.46 [1.24-1.71]* | 1.32 [1.12-1.55]* |
| E28 | 10,770 | 46,351,270 | 0.24 | 0.23 | 0.21 | 0.00 | 0.03 | 0.02 | 1.13 [1.06-1.21]* | 1.10 [1.03-1.18]* |
| E30 | 3,130 | 46,379,870 | 0.08 | 0.06 | 0.07 | 0.02 | 0.01 | 0.01 | 1.22 [1.09-1.37]* | 1.17 [1.04-1.32]* |
| E34 | 4,560 | 46,359,100 | 0.12 | 0.09 | 0.10 | 0.02 | 0.02 | 0.01 | 1.19 [1.08-1.31]* | 1.11 [1.01-1.22] |
| E46 | 530 | 46,412,930 | 0.01 | 0.01 | 0.01 | 0.00 | 0.00 | 0.00 | 1.29 [0.97-1.73] | 1.25 [0.93-1.69] |
| E53 | 2,670 | 46,403,450 | 0.07 | 0.06 | 0.05 | 0.01 | 0.01 | 0.01 | 1.27 [1.11-1.45]* | 1.19 [1.04-1.37] |
| E55 | 8,180 | 46,345,750 | 0.23 | 0.16 | 0.17 | 0.07 | 0.06 | 0.05 | 1.34 [1.25-1.45]* | 1.26 [1.17-1.35]* |
| E61 | 1,810 | 46,403,550 | 0.05 | 0.04 | 0.04 | 0.02 | 0.01 | 0.01 | 1.35 [1.16-1.57]* | 1.27 [1.09-1.48]* |
| E66 | 49,920 | 46,117,900 | 1.14 | 1.07 | 0.97 | 0.08 | 0.18 | 0.13 | 1.18 [1.14-1.22]* | 1.13 [1.10-1.17]* |
| E73 | 2,380 | 46,359,090 | 0.07 | 0.05 | 0.05 | 0.02 | 0.02 | 0.01 | 1.36 [1.20-1.55]* | 1.23 [1.08-1.41]* |
| E74 | 470 | 46,409,180 | 0.01 | 0.01 | 0.01 | 0.00 | 0.00 | 0.00 | 1.32 [0.98-1.78] | 1.22 [0.89-1.66] |
| E78 | 1,280 | 46,407,770 | 0.03 | 0.03 | 0.02 | 0.00 | 0.00 | 0.00 | 1.18 [0.98-1.43] | 1.16 [0.95-1.41] |
| E80 | 1,220 | 46,394,600 | 0.03 | 0.03 | 0.02 | 0.00 | 0.00 | 0.00 | 1.12 [0.92-1.36] | 1.05 [0.86-1.29] |
| E83 | 3,900 | 46,360,830 | 0.09 | 0.08 | 0.08 | 0.01 | 0.01 | 0.01 | 1.08 [0.96-1.20] | 1.06 [0.95-1.19] |
| E86 | 21,080 | 45,992,530 | 0.50 | 0.45 | 0.44 | 0.05 | 0.06 | 0.04 | 1.13 [1.08-1.18]* | 1.10 [1.05-1.15]* |
| E87 | 12,040 | 46,201,390 | 0.28 | 0.26 | 0.26 | 0.02 | 0.02 | 0.02 | 1.09 [1.02-1.15]* | 1.06 [1.00-1.13] |
| E88 | 1,770 | 46,393,580 | 0.04 | 0.04 | 0.04 | 0.00 | 0.01 | 0.00 | 1.16 [0.99-1.36] | 1.12 [0.95-1.32] |
| E89 | 620 | 46,413,460 | 0.01 | 0.01 | 0.01 | 0.00 | 0.00 | 0.00 | 1.11 [0.84-1.47] | 1.09 [0.81-1.47] |
| <18 - F01-F99 | | | | | | | | | | |
| F05 | 730 | 46,413,190 | 0.02 | 0.02 | 0.02 | 0.00 | 0.00 | 0.00 | 1.00 [0.77-1.30] | 1.01 [0.78-1.32] |
| F10 | 18,800 | 46,218,030 | 0.37 | 0.42 | 0.37 | −0.05 | −0.01 | −0.01 | 0.98 [0.93-1.04] | 0.97 [0.92-1.02] |
| F11 | 1,080 | 46,407,880 | 0.02 | 0.02 | 0.02 | 0.00 | 0.00 | 0.00 | 1.00 [0.81-1.25] | 1.07 [0.85-1.35] |
| F12 | 8,690 | 46,362,670 | 0.18 | 0.19 | 0.18 | −0.01 | −0.01 | −0.01 | 0.96 [0.89-1.04] | 0.97 [0.89-1.05] |
| F14 | 2,700 | 46,401,920 | 0.05 | 0.06 | 0.05 | −0.01 | 0.00 | 0.00 | 1.02 [0.89-1.16] | 1.03 [0.89-1.18] |
| F15 | 1,050 | 46,409,140 | 0.02 | 0.02 | 0.02 | 0.00 | 0.00 | 0.00 | 0.95 [0.76-1.18] | 0.92 [0.73-1.16] |
| F17 | 107,180 | 45,702,090 | 2.17 | 2.39 | 2.19 | −0.22 | −0.02 | 0.00 | 0.99 [0.97-1.01] | 1.00 [0.98-1.02] |
| F19 | 2,700 | 46,401,130 | 0.05 | 0.06 | 0.06 | −0.01 | 0.00 | 0.00 | 0.93 [0.81-1.06] | 0.96 [0.83-1.10] |
| F20 | 1,640 | 46,405,740 | 0.03 | 0.04 | 0.03 | 0.00 | 0.00 | 0.00 | 1.00 [0.84-1.20] | 1.04 [0.86-1.25] |
| F22 | 540 | 46,414,470 | 0.01 | 0.01 | 0.01 | 0.00 | 0.00 | 0.00 | 1.00 [0.74-1.36] | 0.99 [0.72-1.35] |
| F23 | 1,020 | 46,410,230 | 0.02 | 0.02 | 0.02 | 0.00 | 0.00 | 0.00 | 0.94 [0.75-1.17] | 0.93 [0.74-1.17] |
| F29 | 1,900 | 46,405,370 | 0.04 | 0.04 | 0.04 | 0.00 | 0.00 | 0.00 | 0.91 [0.77-1.07] | 0.92 [0.77-1.09] |
| F31 | 2,680 | 46,400,320 | 0.06 | 0.06 | 0.05 | 0.00 | 0.00 | 0.00 | 1.07 [0.93-1.23] | 1.08 [0.93-1.24] |
| F32 | 49,230 | 46,109,190 | 1.12 | 1.05 | 0.99 | 0.07 | 0.13 | 0.13 | 1.14 [1.10-1.17]* | 1.13 [1.09-1.16]* |
| F33 | 670 | 46,411,700 | 0.01 | 0.01 | 0.01 | 0.00 | 0.00 | 0.00 | 1.10 [0.84-1.44] | 1.18 [0.89-1.56] |
| F40 | 9,680 | 46,347,870 | 0.23 | 0.20 | 0.20 | 0.03 | 0.03 | 0.02 | 1.13 [1.05-1.21]* | 1.08 [1.01-1.16] |
| F41 | 54,020 | 46,147,660 | 1.29 | 1.14 | 1.11 | 0.15 | 0.18 | 0.15 | 1.16 [1.13-1.20]* | 1.14 [1.10-1.17]* |
| F42 | 2,510 | 46,401,930 | 0.06 | 0.05 | 0.05 | 0.01 | 0.01 | 0.01 | 1.25 [1.09-1.43]* | 1.15 [1.00-1.32] |
| F43 | 4,250 | 46,391,970 | 0.09 | 0.09 | 0.09 | 0.00 | 0.00 | 0.00 | 1.00 [0.90-1.12] | 0.99 [0.89-1.11] |
| F44 | 1,130 | 46,410,550 | 0.03 | 0.02 | 0.02 | 0.00 | 0.01 | 0.00 | 1.27 [1.04-1.55] | 1.19 [0.97-1.47] |
| F45 | 860 | 46,409,370 | 0.02 | 0.02 | 0.02 | 0.00 | 0.00 | 0.00 | 1.27 [1.02-1.60] | 1.16 [0.91-1.46] |
| F50 | 4,950 | 46,380,030 | 0.12 | 0.10 | 0.11 | 0.02 | 0.01 | 0.01 | 1.13 [1.02-1.24] | 1.09 [0.99-1.20] |
| F51 | 730 | 46,409,980 | 0.02 | 0.01 | 0.01 | 0.00 | 0.00 | 0.00 | 1.33 [1.05-1.68] | 1.31 [1.03-1.67] |
| F53 | 710 | 46,413,260 | 0.01 | 0.02 | 0.01 | 0.00 | 0.00 | 0.00 | 0.96 [0.73-1.26] | 0.95 [0.71-1.26] |
| F60 | 4,420 | 46,391,610 | 0.09 | 0.10 | 0.09 | 0.00 | 0.00 | 0.00 | 1.02 [0.92-1.14] | 1.02 [0.91-1.14] |
| F70 | 790 | 46,411,970 | 0.02 | 0.02 | 0.02 | 0.00 | 0.00 | 0.00 | 1.19 [0.94-1.51] | 1.14 [0.89-1.46] |
| F72 | 850 | 46,409,990 | 0.02 | 0.02 | 0.02 | 0.00 | 0.00 | 0.00 | 1.03 [0.82-1.29] | 1.01 [0.80-1.29] |
| F79 | 4,870 | 46,371,880 | 0.12 | 0.10 | 0.11 | 0.01 | 0.01 | 0.00 | 1.06 [0.96-1.16] | 1.03 [0.93-1.13] |
| F80 | 9,110 | 46,310,380 | 0.23 | 0.19 | 0.19 | 0.04 | 0.03 | 0.02 | 1.16 [1.09-1.24]* | 1.11 [1.04-1.19]* |
| F81 | 15,820 | 46,230,150 | 0.39 | 0.33 | 0.34 | 0.06 | 0.05 | 0.03 | 1.13 [1.07-1.19]* | 1.08 [1.03-1.14]* |
| F82 | 1,560 | 46,395,720 | 0.04 | 0.03 | 0.03 | 0.00 | 0.00 | 0.00 | 1.12 [0.95-1.33] | 1.07 [0.90-1.26] |
| F83 | 1,490 | 46,402,370 | 0.03 | 0.03 | 0.03 | 0.00 | 0.00 | 0.00 | 0.91 [0.76-1.09] | 0.88 [0.73-1.05] |
| F84 | 24,870 | 46,204,610 | 0.67 | 0.50 | 0.55 | 0.17 | 0.12 | 0.09 | 1.22 [1.17-1.27]* | 1.15 [1.11-1.20]* |
| F89 | 7,040 | 46,361,330 | 0.16 | 0.15 | 0.16 | 0.01 | 0.00 | −0.01 | 0.97 [0.90-1.05] | 0.92 [0.85-1.00] |
| F90 | 13,340 | 46,306,380 | 0.35 | 0.27 | 0.30 | 0.08 | 0.05 | 0.04 | 1.16 [1.09-1.22]* | 1.12 [1.06-1.19]* |
| F91 | 2,570 | 46,376,820 | 0.07 | 0.05 | 0.05 | 0.02 | 0.02 | 0.01 | 1.30 [1.15-1.48]* | 1.24 [1.09-1.41]* |
| F94 | 550 | 46,414,050 | 0.01 | 0.01 | 0.01 | 0.00 | 0.00 | 0.00 | 1.19 [0.90-1.56] | 1.14 [0.86-1.51] |
| F95 | 1,360 | 46,407,270 | 0.04 | 0.03 | 0.03 | 0.01 | 0.01 | 0.01 | 1.30 [1.10-1.55]* | 1.22 [1.02-1.45] |
| F98 | 1,840 | 46,395,030 | 0.05 | 0.04 | 0.04 | 0.01 | 0.01 | 0.01 | 1.30 [1.12-1.51]* | 1.25 [1.08-1.46]* |
| F99 | 1,360 | 46,407,120 | 0.03 | 0.03 | 0.03 | 0.00 | 0.00 | 0.00 | 1.10 [0.91-1.32] | 1.09 [0.90-1.33] |
| <18 - G00-G99 | | | | | | | | | | |
| G00 | 550 | 46,375,780 | 0.01 | 0.01 | 0.01 | 0.00 | 0.00 | 0.00 | 0.92 [0.69-1.24] | 0.90 [0.66-1.21] |
| G01 | 530 | 46,391,620 | 0.01 | 0.01 | 0.01 | 0.00 | 0.00 | 0.00 | 1.09 [0.82-1.45] | 1.11 [0.83-1.49] |
| G03 | 820 | 46,379,080 | 0.02 | 0.02 | 0.02 | 0.00 | 0.01 | 0.00 | 1.32 [1.06-1.65] | 1.27 [1.01-1.60] |
| G04 | 1,200 | 46,399,560 | 0.03 | 0.03 | 0.03 | 0.00 | 0.00 | 0.00 | 1.10 [0.91-1.33] | 1.08 [0.89-1.31] |
| G24 | 1,340 | 46,402,880 | 0.03 | 0.03 | 0.03 | 0.00 | −0.01 | −0.01 | 0.82 [0.67-1.00] | 0.79 [0.64-0.96] |
| G25 | 1,570 | 46,382,060 | 0.04 | 0.03 | 0.03 | 0.00 | 0.00 | 0.00 | 1.07 [0.91-1.27] | 1.04 [0.87-1.24] |
| G31 | 580 | 46,408,680 | 0.01 | 0.01 | 0.01 | 0.00 | 0.00 | 0.00 | 1.04 [0.78-1.37] | 1.01 [0.76-1.35] |
| G35 | 630 | 46,413,860 | 0.01 | 0.01 | 0.01 | 0.00 | 0.00 | 0.00 | 1.14 [0.85-1.53] | 1.15 [0.85-1.55] |
| G37 | 540 | 46,412,650 | 0.01 | 0.01 | 0.01 | 0.00 | 0.00 | 0.00 | 1.04 [0.77-1.40] | 1.02 [0.75-1.39] |
| G40 | 18,330 | 46,093,440 | 0.42 | 0.39 | 0.39 | 0.02 | 0.03 | 0.02 | 1.08 [1.02-1.13]* | 1.06 [1.00-1.11] |
| G41 | 2,260 | 46,377,050 | 0.05 | 0.05 | 0.05 | 0.00 | 0.00 | 0.00 | 0.96 [0.84-1.11] | 0.95 [0.83-1.10] |
| G43 | 16,520 | 46,290,750 | 0.40 | 0.35 | 0.33 | 0.05 | 0.07 | 0.05 | 1.20 [1.14-1.26]* | 1.13 [1.07-1.20]* |
| G44 | 1,990 | 46,403,270 | 0.05 | 0.04 | 0.04 | 0.01 | 0.01 | 0.01 | 1.22 [1.05-1.42] | 1.19 [1.02-1.39] |
| G47 | 33,010 | 46,023,030 | 0.91 | 0.66 | 0.69 | 0.24 | 0.22 | 0.18 | 1.31 [1.27-1.36]* | 1.25 [1.21-1.30]* |
| G51 | 2,660 | 46,381,550 | 0.06 | 0.06 | 0.06 | 0.00 | 0.00 | 0.00 | 1.09 [0.95-1.24] | 1.09 [0.95-1.24] |
| G55 | 2,210 | 46,405,110 | 0.05 | 0.05 | 0.04 | 0.00 | 0.01 | 0.00 | 1.12 [0.96-1.30] | 1.06 [0.90-1.23] |
| G56 | 1,900 | 46,401,820 | 0.04 | 0.04 | 0.04 | 0.00 | 0.01 | 0.01 | 1.23 [1.06-1.44]* | 1.16 [0.98-1.36] |
| G62 | 940 | 46,410,940 | 0.02 | 0.02 | 0.02 | 0.00 | 0.00 | 0.00 | 0.97 [0.78-1.22] | 0.94 [0.74-1.19] |
| G71 | 950 | 46,403,570 | 0.02 | 0.02 | 0.02 | 0.00 | 0.00 | 0.00 | 1.00 [0.81-1.25] | 1.01 [0.81-1.26] |
| G80 | 5,760 | 46,293,480 | 0.10 | 0.13 | 0.13 | −0.03 | −0.03 | −0.03 | 0.79 [0.72-0.87]* | 0.77 [0.70-0.85]* |
| G81 | 2,570 | 46,375,380 | 0.05 | 0.06 | 0.06 | 0.00 | 0.00 | 0.00 | 0.98 [0.86-1.13] | 0.98 [0.85-1.12] |
| G82 | 1,170 | 46,399,780 | 0.02 | 0.03 | 0.03 | −0.01 | −0.01 | −0.01 | 0.75 [0.61-0.93]* | 0.73 [0.59-0.91]* |
| G83 | 820 | 46,406,260 | 0.02 | 0.02 | 0.02 | 0.00 | 0.00 | 0.00 | 0.97 [0.76-1.24] | 0.95 [0.74-1.22] |
| G90 | 660 | 46,411,610 | 0.01 | 0.01 | 0.01 | 0.00 | 0.00 | 0.00 | 1.01 [0.77-1.33] | 0.94 [0.71-1.25] |
| G91 | 1,820 | 46,368,200 | 0.04 | 0.04 | 0.04 | 0.00 | 0.00 | 0.00 | 0.97 [0.82-1.14] | 0.97 [0.82-1.14] |
| G93 | 8,880 | 46,312,950 | 0.21 | 0.19 | 0.18 | 0.02 | 0.02 | 0.02 | 1.13 [1.05-1.22]* | 1.08 [1.01-1.17] |
| G95 | 960 | 46,408,850 | 0.02 | 0.02 | 0.02 | 0.00 | 0.00 | 0.00 | 1.08 [0.87-1.35] | 1.02 [0.82-1.28] |
| G96 | 690 | 46,409,180 | 0.01 | 0.02 | 0.01 | 0.00 | 0.00 | 0.00 | 1.03 [0.79-1.33] | 0.97 [0.74-1.26] |
| G97 | 1,040 | 46,408,540 | 0.02 | 0.02 | 0.02 | 0.00 | 0.00 | 0.00 | 1.12 [0.91-1.39] | 1.09 [0.88-1.35] |
| <18 - H00-H59 | | | | | | | | | | |
| H00 | 7,670 | 46,292,230 | 0.19 | 0.16 | 0.16 | 0.03 | 0.03 | 0.03 | 1.20 [1.11-1.29]* | 1.17 [1.09-1.26]* |
| H01 | 1,020 | 46,399,880 | 0.03 | 0.02 | 0.02 | 0.01 | 0.01 | 0.01 | 1.49 [1.23-1.81]* | 1.39 [1.14-1.70]* |
| H02 | 4,250 | 46,344,630 | 0.10 | 0.09 | 0.09 | 0.02 | 0.02 | 0.02 | 1.22 [1.10-1.35]* | 1.18 [1.06-1.31]* |
| H03 | 360 | 46,411,020 | 0.01 | 0.01 | 0.01 | 0.01 | 0.01 | 0.00 | 1.74 [1.27-2.38]* | 1.60 [1.16-2.21]* |
| H04 | 4,340 | 46,302,430 | 0.10 | 0.09 | 0.09 | 0.01 | 0.02 | 0.01 | 1.20 [1.09-1.32]* | 1.15 [1.04-1.26]* |
| H05 | 6,760 | 46,273,120 | 0.18 | 0.14 | 0.14 | 0.05 | 0.05 | 0.04 | 1.33 [1.23-1.44]* | 1.29 [1.19-1.39]* |
| H10 | 6,180 | 46,168,510 | 0.18 | 0.12 | 0.13 | 0.06 | 0.05 | 0.04 | 1.42 [1.32-1.54]* | 1.32 [1.22-1.43]* |
| H11 | 1,260 | 46,396,840 | 0.03 | 0.03 | 0.03 | 0.00 | 0.00 | 0.00 | 1.10 [0.91-1.32] | 1.07 [0.88-1.30] |
| H16 | 700 | 46,406,540 | 0.02 | 0.01 | 0.01 | 0.01 | 0.01 | 0.01 | 1.92 [1.54-2.41]* | 1.75 [1.39-2.21]* |
| H18 | 2,230 | 46,401,710 | 0.07 | 0.04 | 0.04 | 0.03 | 0.04 | 0.03 | 1.91 [1.67-2.18]* | 1.70 [1.48-1.95]* |
| H20 | 830 | 46,409,100 | 0.02 | 0.02 | 0.02 | 0.01 | 0.01 | 0.01 | 1.40 [1.12-1.74]* | 1.35 [1.08-1.70]* |

| | | | | | | | | | | |
|---|---|---|---|---|---|---|---|---|---|---|
| H26 | 1,510 | 46,395,670 | 0.04 | 0.03 | 0.03 | 0.02 | 0.02 | 0.02 | 1.60 [1.36-1.87]* | 1.56 [1.32-1.83]* |
| H33 | 1,090 | 46,404,830 | 0.02 | 0.02 | 0.02 | 0.00 | 0.00 | 0.00 | 1.10 [0.89-1.35] | 1.08 [0.87-1.34] |
| H35 | 1,320 | 46,353,910 | 0.03 | 0.03 | 0.03 | 0.00 | 0.00 | 0.00 | 1.16 [0.96-1.39] | 1.14 [0.94-1.38] |
| H36 | 570 | 46,415,230 | 0.01 | 0.01 | 0.01 | 0.00 | 0.00 | 0.00 | 1.09 [0.81-1.48] | 1.14 [0.83-1.57] |
| H40 | 650 | 46,408,090 | 0.02 | 0.01 | 0.01 | 0.00 | 0.00 | 0.00 | 1.29 [1.00-1.67] | 1.26 [0.97-1.64] |
| H44 | 580 | 46,391,750 | 0.01 | 0.01 | 0.01 | 0.00 | 0.00 | 0.00 | 1.25 [0.95-1.64] | 1.14 [0.87-1.51] |
| H46 | 410 | 46,414,300 | 0.01 | 0.01 | 0.01 | 0.00 | 0.00 | 0.00 | 1.43 [1.04-1.97] | 1.45 [1.04-2.02] |
| H47 | 3,090 | 46,390,770 | 0.07 | 0.06 | 0.07 | 0.01 | 0.01 | 0.00 | 1.10 [0.97-1.24] | 1.06 [0.93-1.20] |
| H49 | 1,250 | 46,402,370 | 0.03 | 0.03 | 0.03 | 0.00 | 0.00 | 0.00 | 0.97 [0.80-1.18] | 0.92 [0.75-1.12] |
| H50 | 20,760 | 46,019,910 | 0.46 | 0.45 | 0.46 | 0.01 | 0.00 | −0.01 | 1.01 [0.96-1.05] | 0.99 [0.94-1.04] |
| H52 | 4,230 | 46,368,660 | 0.10 | 0.09 | 0.09 | 0.01 | 0.01 | 0.01 | 1.13 [1.02-1.25] | 1.10 [0.99-1.21] |
| H53 | 15,770 | 46,262,430 | 0.38 | 0.33 | 0.32 | 0.05 | 0.06 | 0.04 | 1.18 [1.12-1.24]* | 1.13 [1.07-1.19]* |
| H54 | 3,590 | 46,370,660 | 0.08 | 0.08 | 0.08 | 0.00 | 0.00 | 0.00 | 1.00 [0.89-1.12] | 0.96 [0.86-1.08] |
| H55 | 1,800 | 46,387,640 | 0.04 | 0.04 | 0.04 | 0.00 | 0.00 | 0.00 | 0.95 [0.80-1.11] | 0.92 [0.78-1.09] |
| H57 | 2,400 | 46,376,690 | 0.06 | 0.05 | 0.05 | 0.02 | 0.01 | 0.01 | 1.28 [1.13-1.46]* | 1.22 [1.07-1.40]* |
| <18 - H60-H95 | | | | | | | | | | |
| H60 | 4,500 | 46,338,940 | 0.12 | 0.09 | 0.09 | 0.03 | 0.03 | 0.03 | 1.32 [1.20-1.46]* | 1.28 [1.16-1.41]* |
| H61 | 12,220 | 46,206,730 | 0.32 | 0.25 | 0.25 | 0.07 | 0.06 | 0.06 | 1.26 [1.19-1.33]* | 1.22 [1.15-1.29]* |
| H65 | 62,420 | 44,949,020 | 1.59 | 1.33 | 1.33 | 0.25 | 0.26 | 0.22 | 1.20 [1.17-1.23]* | 1.16 [1.13-1.19]* |
| H66 | 25,910 | 45,698,900 | 0.64 | 0.55 | 0.53 | 0.10 | 0.11 | 0.09 | 1.21 [1.16-1.25]* | 1.17 [1.13-1.22]* |
| H69 | 1,040 | 46,400,890 | 0.03 | 0.02 | 0.02 | 0.01 | 0.01 | 0.01 | 1.43 [1.18-1.73]* | 1.44 [1.18-1.76]* |
| H70 | 1,620 | 46,387,430 | 0.04 | 0.03 | 0.03 | 0.00 | 0.00 | 0.00 | 1.04 [0.88-1.23] | 1.01 [0.85-1.20] |
| H71 | 3,430 | 46,369,850 | 0.08 | 0.07 | 0.08 | 0.01 | 0.00 | 0.00 | 1.07 [0.95-1.19] | 1.05 [0.94-1.18] |
| H72 | 7,870 | 46,291,160 | 0.19 | 0.16 | 0.16 | 0.03 | 0.03 | 0.02 | 1.17 [1.09-1.26]* | 1.14 [1.06-1.23]* |
| H73 | 7,240 | 46,297,110 | 0.18 | 0.15 | 0.15 | 0.03 | 0.03 | 0.03 | 1.19 [1.10-1.28]* | 1.16 [1.07-1.25]* |
| H74 | 3,780 | 46,355,750 | 0.09 | 0.08 | 0.08 | 0.01 | 0.01 | 0.01 | 1.10 [0.99-1.23] | 1.06 [0.95-1.19] |
| H81 | 410 | 46,415,370 | 0.01 | 0.01 | 0.01 | 0.00 | 0.00 | 0.00 | 1.31 [0.94-1.81] | 1.23 [0.87-1.73] |
| H83 | 900 | 46,405,990 | 0.02 | 0.02 | 0.02 | 0.00 | 0.00 | 0.00 | 1.18 [0.95-1.47] | 1.12 [0.89-1.40] |
| H90 | 10,630 | 46,263,300 | 0.26 | 0.22 | 0.23 | 0.04 | 0.04 | 0.03 | 1.16 [1.09-1.23]* | 1.12 [1.05-1.19]* |
| H91 | 13,600 | 46,174,140 | 0.33 | 0.29 | 0.29 | 0.04 | 0.04 | 0.03 | 1.15 [1.08-1.21]* | 1.11 [1.05-1.17]* |
| H92 | 5,810 | 46,313,000 | 0.15 | 0.12 | 0.12 | 0.03 | 0.03 | 0.02 | 1.23 [1.13-1.33]* | 1.18 [1.08-1.29]* |
| H93 | 1,520 | 46,400,020 | 0.04 | 0.03 | 0.03 | 0.01 | 0.01 | 0.01 | 1.20 [1.01-1.43] | 1.16 [0.98-1.39] |
| H95 | 700 | 46,408,100 | 0.02 | 0.01 | 0.01 | 0.00 | 0.00 | 0.00 | 1.20 [0.93-1.54] | 1.17 [0.90-1.51] |
| <18 - I00-I99 | | | | | | | | | | |
| I07 | 620 | 46,411,290 | 0.01 | 0.01 | 0.01 | 0.00 | 0.00 | 0.00 | 0.89 [0.67-1.18] | 0.89 [0.67-1.20] |
| I08 | 800 | 46,411,680 | 0.02 | 0.02 | 0.02 | 0.00 | 0.00 | 0.00 | 1.08 [0.84-1.37] | 1.06 [0.83-1.37] |
| I10 | 7,710 | 46,347,770 | 0.18 | 0.16 | 0.15 | 0.02 | 0.03 | 0.03 | 1.21 [1.12-1.31]* | 1.17 [1.08-1.27]* |
| I15 | 710 | 46,406,890 | 0.02 | 0.01 | 0.02 | 0.00 | 0.00 | 0.00 | 1.07 [0.84-1.38] | 1.08 [0.84-1.39] |
| I25 | 720 | 46,412,220 | 0.01 | 0.02 | 0.01 | 0.00 | 0.00 | 0.00 | 1.06 [0.81-1.37] | 1.08 [0.83-1.41] |
| I26 | 1,370 | 46,409,580 | 0.03 | 0.03 | 0.03 | 0.00 | 0.00 | 0.00 | 1.07 [0.88-1.29] | 1.03 [0.84-1.26] |
| I27 | 580 | 46,392,720 | 0.01 | 0.01 | 0.01 | 0.00 | 0.00 | 0.00 | 0.85 [0.63-1.15] | 0.77 [0.57-1.05] |
| I31 | 2,180 | 46,391,700 | 0.04 | 0.05 | 0.04 | 0.00 | 0.00 | 0.00 | 1.02 [0.88-1.19] | 0.99 [0.85-1.16] |
| I34 | 1,150 | 46,405,630 | 0.03 | 0.02 | 0.03 | 0.00 | 0.00 | 0.00 | 0.99 [0.81-1.21] | 0.93 [0.76-1.15] |
| I35 | 1,130 | 46,404,640 | 0.02 | 0.02 | 0.03 | 0.00 | 0.00 | 0.00 | 0.95 [0.78-1.17] | 0.96 [0.78-1.18] |
| I37 | 820 | 46,407,830 | 0.02 | 0.02 | 0.02 | 0.00 | 0.00 | 0.00 | 0.95 [0.75-1.20] | 0.94 [0.74-1.20] |
| I38 | 680 | 46,390,600 | 0.02 | 0.01 | 0.01 | 0.00 | 0.00 | 0.00 | 1.09 [0.84-1.40] | 1.06 [0.82-1.38] |
| I42 | 1,070 | 46,392,960 | 0.02 | 0.02 | 0.02 | 0.00 | 0.00 | 0.00 | 1.17 [0.96-1.43] | 1.18 [0.95-1.45] |
| I44 | 1,050 | 46,406,130 | 0.03 | 0.02 | 0.02 | 0.00 | 0.01 | 0.01 | 1.29 [1.04-1.58] | 1.26 [1.02-1.56] |
| I45 | 3,300 | 46,386,840 | 0.07 | 0.07 | 0.07 | 0.00 | 0.00 | 0.00 | 1.03 [0.91-1.16] | 0.99 [0.87-1.12] |
| I46 | 1,270 | 46,407,590 | 0.03 | 0.03 | 0.03 | 0.00 | 0.00 | 0.00 | 1.08 [0.89-1.30] | 1.04 [0.85-1.26] |
| I47 | 3,280 | 46,365,780 | 0.08 | 0.07 | 0.07 | 0.01 | 0.01 | 0.01 | 1.16 [1.03-1.31] | 1.11 [0.98-1.25] |
| I48 | 1,020 | 46,407,780 | 0.02 | 0.02 | 0.02 | 0.00 | 0.00 | 0.00 | 1.06 [0.85-1.32] | 1.01 [0.80-1.27] |
| I49 | 3,060 | 46,388,440 | 0.07 | 0.06 | 0.06 | 0.01 | 0.01 | 0.01 | 1.16 [1.03-1.31] | 1.10 [0.96-1.24] |
| I50 | 1,220 | 46,392,750 | 0.03 | 0.03 | 0.02 | 0.00 | 0.00 | 0.00 | 1.09 [0.90-1.33] | 1.10 [0.90-1.35] |
| I51 | 3,230 | 46,377,740 | 0.07 | 0.07 | 0.07 | 0.00 | 0.01 | 0.00 | 1.09 [0.97-1.23] | 1.07 [0.95-1.22] |
| I61 | 560 | 46,409,390 | 0.01 | 0.01 | 0.01 | 0.00 | 0.00 | 0.00 | 0.88 [0.65-1.19] | 0.88 [0.64-1.20] |
| I63 | 720 | 46,405,470 | 0.02 | 0.02 | 0.01 | 0.00 | 0.00 | 0.00 | 1.10 [0.86-1.42] | 1.17 [0.90-1.51] |
| I67 | 640 | 46,412,070 | 0.01 | 0.01 | 0.01 | 0.00 | 0.00 | 0.00 | 1.08 [0.82-1.41] | 1.08 [0.82-1.43] |
| I69 | 770 | 46,405,670 | 0.02 | 0.02 | 0.02 | 0.00 | 0.00 | 0.00 | 0.94 [0.73-1.21] | 0.91 [0.70-1.17] |
| I73 | 1,830 | 46,402,510 | 0.05 | 0.04 | 0.03 | 0.01 | 0.01 | 0.01 | 1.44 [1.24-1.68]* | 1.35 [1.15-1.58]* |
| I77 | 610 | 46,411,330 | 0.01 | 0.01 | 0.01 | 0.00 | 0.00 | 0.00 | 1.19 [0.91-1.56] | 1.13 [0.85-1.49] |
| I78 | 580 | 46,407,870 | 0.01 | 0.01 | 0.01 | 0.00 | 0.00 | 0.00 | 1.03 [0.77-1.36] | 1.01 [0.76-1.35] |
| I80 | 1,680 | 46,402,560 | 0.04 | 0.04 | 0.03 | 0.00 | 0.01 | 0.00 | 1.15 [0.98-1.36] | 1.11 [0.93-1.32] |
| I82 | 710 | 46,410,470 | 0.01 | 0.02 | 0.01 | 0.00 | 0.00 | 0.00 | 1.00 [0.77-1.29] | 0.98 [0.75-1.29] |
| I83 | 1,420 | 46,407,240 | 0.03 | 0.03 | 0.02 | −0.01 | 0.00 | 0.00 | 1.05 [0.86-1.27] | 1.01 [0.82-1.24] |
| I84 | 3,640 | 46,369,670 | 0.08 | 0.08 | 0.07 | 0.00 | 0.01 | 0.01 | 1.21 [1.08-1.36]* | 1.17 [1.04-1.31]* |
| I86 | 1,950 | 46,398,510 | 0.04 | 0.04 | 0.04 | 0.00 | 0.00 | 0.00 | 1.03 [0.88-1.21] | 0.99 [0.84-1.17] |
| I87 | 570 | 46,412,600 | 0.01 | 0.01 | 0.01 | 0.00 | 0.00 | 0.00 | 1.06 [0.79-1.42] | 1.03 [0.76-1.39] |
| I88 | 12,370 | 46,238,610 | 0.33 | 0.25 | 0.26 | 0.08 | 0.07 | 0.06 | 1.26 [1.19-1.33]* | 1.22 [1.15-1.29]* |
| I89 | 760 | 46,405,850 | 0.02 | 0.02 | 0.02 | 0.00 | 0.00 | 0.00 | 1.16 [0.91-1.47] | 1.08 [0.84-1.39] |
| I95 | 6,070 | 46,346,760 | 0.14 | 0.13 | 0.13 | 0.01 | 0.01 | 0.01 | 1.06 [0.97-1.16] | 1.04 [0.95-1.14] |
| <18 - J00-J99 | | | | | | | | | | |
| J00 | 9,280 | 46,179,660 | 0.26 | 0.19 | 0.18 | 0.07 | 0.07 | 0.07 | 1.41 [1.32-1.50]* | 1.35 [1.27-1.44]* |
| J01 | 1,120 | 46,404,840 | 0.03 | 0.02 | 0.02 | 0.01 | 0.01 | 0.01 | 1.44 [1.19-1.74]* | 1.36 [1.12-1.66]* |
| J02 | 13,140 | 46,194,760 | 0.33 | 0.27 | 0.28 | 0.06 | 0.05 | 0.04 | 1.19 [1.13-1.26]* | 1.15 [1.09-1.22]* |
| J03 | 113,070 | 44,582,020 | 2.92 | 2.43 | 2.44 | 0.48 | 0.48 | 0.40 | 1.20 [1.17-1.22]* | 1.16 [1.14-1.18]* |
| J04 | 950 | 46,397,920 | 0.02 | 0.02 | 0.02 | 0.01 | 0.01 | 0.01 | 1.32 [1.07-1.63]* | 1.29 [1.05-1.60] |
| J05 | 21,810 | 45,724,110 | 0.47 | 0.48 | 0.47 | 0.00 | 0.00 | −0.01 | 1.01 [0.96-1.05] | 0.98 [0.94-1.03] |
| J06 | 72,590 | 43,287,340 | 2.00 | 1.59 | 1.55 | 0.41 | 0.45 | 0.39 | 1.29 [1.26-1.32]* | 1.25 [1.22-1.27]* |
| J10 | 4,640 | 46,367,210 | 0.12 | 0.10 | 0.10 | 0.02 | 0.02 | 0.01 | 1.23 [1.12-1.35]* | 1.15 [1.04-1.26]* |
| J11 | 1,070 | 46,399,400 | 0.03 | 0.02 | 0.02 | 0.01 | 0.00 | 0.00 | 1.18 [0.97-1.44] | 1.13 [0.92-1.38] |
| J12 | 2,540 | 46,376,190 | 0.06 | 0.05 | 0.05 | 0.01 | 0.01 | 0.01 | 1.16 [1.02-1.32] | 1.10 [0.96-1.25] |
| J13 | 610 | 46,401,710 | 0.01 | 0.01 | 0.01 | 0.00 | 0.00 | 0.00 | 1.18 [0.90-1.53] | 1.09 [0.83-1.43] |
| J15 | 1,920 | 46,372,000 | 0.05 | 0.04 | 0.04 | 0.01 | 0.01 | 0.01 | 1.22 [1.05-1.41]* | 1.16 [1.00-1.35] |
| J18 | 33,390 | 45,603,240 | 0.87 | 0.69 | 0.67 | 0.18 | 0.20 | 0.18 | 1.30 [1.26-1.35]* | 1.25 [1.21-1.30]* |
| J20 | 1,230 | 46,361,620 | 0.03 | 0.03 | 0.02 | 0.01 | 0.01 | 0.01 | 1.30 [1.09-1.56]* | 1.25 [1.04-1.50] |
| J21 | 3,820 | 44,624,160 | 0.09 | 0.09 | 0.08 | 0.00 | 0.00 | 0.00 | 1.04 [0.94-1.16] | 1.00 [0.90-1.11] |
| J22 | 59,190 | 45,172,410 | 1.70 | 1.20 | 1.19 | 0.50 | 0.51 | 0.44 | 1.43 [1.39-1.46]* | 1.35 [1.31-1.38]* |
| J30 | 20,680 | 46,235,050 | 0.93 | 0.32 | 0.33 | 0.61 | 0.60 | 0.54 | 2.83 [2.72-2.95]* | 2.38 [2.28-2.48]* |
| J31 | 3,600 | 46,361,390 | 0.13 | 0.06 | 0.06 | 0.06 | 0.06 | 0.05 | 1.96 [1.78-2.17]* | 1.68 [1.52-1.87]* |
| J32 | 4,940 | 46,361,870 | 0.13 | 0.10 | 0.10 | 0.03 | 0.03 | 0.02 | 1.31 [1.20-1.44]* | 1.21 [1.10-1.33]* |
| J33 | 1,800 | 46,398,900 | 0.05 | 0.04 | 0.03 | 0.01 | 0.01 | 0.01 | 1.41 [1.21-1.64]* | 1.21 [1.03-1.42] |
| J34 | 23,060 | 46,069,540 | 0.65 | 0.46 | 0.44 | 0.19 | 0.21 | 0.18 | 1.48 [1.42-1.54]* | 1.37 [1.31-1.43]* |
| J35 | 90,040 | 44,505,370 | 2.43 | 1.91 | 1.90 | 0.51 | 0.53 | 0.45 | 1.28 [1.25-1.30]* | 1.23 [1.20-1.26]* |
| J36 | 5,480 | 46,364,010 | 0.12 | 0.12 | 0.11 | 0.00 | 0.01 | 0.01 | 1.11 [1.01-1.22] | 1.10 [1.00-1.21] |
| J38 | 2,730 | 46,339,560 | 0.07 | 0.06 | 0.05 | 0.01 | 0.01 | 0.01 | 1.23 [1.08-1.39]* | 1.12 [0.99-1.28] |
| J39 | 2,310 | 46,373,680 | 0.06 | 0.05 | 0.05 | 0.01 | 0.01 | 0.01 | 1.27 [1.11-1.45]* | 1.17 [1.02-1.34] |
| J40 | 440 | 46,410,240 | 0.01 | 0.01 | 0.01 | 0.00 | 0.00 | 0.00 | 1.32 [0.96-1.82] | 1.24 [0.89-1.73] |
| J43 | 660 | 46,411,630 | 0.02 | 0.01 | 0.01 | 0.00 | 0.00 | 0.00 | 1.21 [0.93-1.57] | 1.17 [0.89-1.53] |
| J44 | 990 | 46,401,090 | 0.03 | 0.02 | 0.02 | 0.01 | 0.01 | 0.01 | 1.81 [1.49-2.20]* | 1.54 [1.26-1.89]* |
| J45 | 161,550 | 43,482,590 | 6.89 | 2.87 | 2.76 | 4.02 | 4.13 | 3.54 | 2.50 [2.46-2.53]* | 2.06 [2.02-2.09]* |
| J46 | 5,670 | 46,251,990 | 0.27 | 0.08 | 0.08 | 0.19 | 0.19 | 0.17 | 3.34 [3.10-3.60]* | 2.76 [2.55-2.99]* |
| J47 | 1,040 | 46,404,390 | 0.03 | 0.02 | 0.02 | 0.01 | 0.01 | 0.00 | 1.29 [1.06-1.58] | 1.15 [0.93-1.42] |
| J69 | 1,690 | 46,398,510 | 0.03 | 0.04 | 0.04 | 0.00 | 0.00 | 0.00 | 0.93 [0.78-1.10] | 0.90 [0.76-1.07] |
| J81 | 520 | 46,410,290 | 0.01 | 0.01 | 0.01 | 0.00 | 0.00 | 0.00 | 0.97 [0.72-1.31] | 0.96 [0.70-1.30] |
| J86 | 1,270 | 46,393,690 | 0.03 | 0.03 | 0.03 | 0.00 | 0.00 | 0.00 | 1.07 [0.89-1.30] | 1.10 [0.91-1.33] |

| | | | | | | | | | | |
|---|---|---|---|---|---|---|---|---|---|---|
| J90 | 5,940 | 46,342,440 | 0.14 | 0.13 | 0.12 | 0.01 | 0.01 | 0.01 | 1.11 [1.01-1.21] | 1.11 [1.02-1.22] |
| J93 | 2,910 | 46,382,590 | 0.06 | 0.06 | 0.06 | 0.00 | 0.00 | 0.00 | 1.05 [0.93-1.20] | 1.06 [0.93-1.21] |
| J95 | 650 | 46,399,290 | 0.01 | 0.01 | 0.01 | 0.00 | 0.00 | 0.00 | 0.98 [0.75-1.27] | 0.91 [0.70-1.20] |
| J96 | 3,060 | 46,383,370 | 0.07 | 0.06 | 0.06 | 0.01 | 0.01 | 0.01 | 1.21 [1.07-1.36]* | 1.12 [0.99-1.26] |
| J98 | 10,240 | 46,193,100 | 0.26 | 0.21 | 0.21 | 0.05 | 0.05 | 0.04 | 1.25 [1.17-1.33]* | 1.17 [1.10-1.25]* |
| <18 - K00-K95 | | | | | | | | | | |
| K00 | 23,020 | 46,121,370 | 0.58 | 0.48 | 0.51 | 0.10 | 0.07 | 0.06 | 1.14 [1.09-1.19]* | 1.11 [1.06-1.16]* |
| K01 | 36,540 | 46,015,180 | 0.88 | 0.77 | 0.75 | 0.11 | 0.13 | 0.11 | 1.18 [1.14-1.22]* | 1.14 [1.10-1.18]* |
| K02 | 124,750 | 44,514,260 | 2.94 | 2.77 | 2.87 | 0.17 | 0.07 | 0.03 | 1.02 [1.00-1.04] | 1.01 [0.99-1.03] |
| K03 | 3,080 | 46,391,910 | 0.07 | 0.06 | 0.07 | 0.01 | 0.00 | 0.00 | 1.00 [0.89-1.13] | 0.98 [0.86-1.11] |
| K04 | 18,040 | 46,189,010 | 0.40 | 0.39 | 0.40 | 0.01 | 0.00 | 0.00 | 1.00 [0.95-1.05] | 0.99 [0.94-1.05] |
| K05 | 12,650 | 46,314,290 | 0.29 | 0.27 | 0.24 | 0.02 | 0.05 | 0.04 | 1.21 [1.14-1.29]* | 1.16 [1.09-1.24]* |
| K06 | 1,160 | 46,402,690 | 0.03 | 0.02 | 0.02 | 0.00 | 0.00 | 0.00 | 1.19 [0.98-1.44] | 1.12 [0.92-1.37] |
| K07 | 28,850 | 46,042,070 | 0.71 | 0.60 | 0.62 | 0.10 | 0.09 | 0.07 | 1.15 [1.10-1.19]* | 1.11 [1.07-1.16]* |
| K08 | 21,350 | 46,141,220 | 0.48 | 0.46 | 0.46 | 0.02 | 0.02 | 0.01 | 1.04 [0.99-1.09] | 1.02 [0.98-1.07] |
| K09 | 1,230 | 46,390,240 | 0.03 | 0.03 | 0.03 | 0.01 | 0.01 | 0.01 | 1.23 [1.02-1.48] | 1.19 [0.98-1.44] |
| K10 | 920 | 46,408,610 | 0.02 | 0.02 | 0.02 | 0.00 | 0.00 | 0.00 | 1.26 [1.02-1.57] | 1.21 [0.97-1.52] |
| K11 | 3,480 | 46,369,940 | 0.09 | 0.07 | 0.07 | 0.01 | 0.01 | 0.01 | 1.16 [1.04-1.30]* | 1.12 [1.00-1.25] |
| K12 | 6,550 | 46,319,930 | 0.16 | 0.14 | 0.14 | 0.03 | 0.03 | 0.02 | 1.19 [1.10-1.29]* | 1.17 [1.08-1.27]* |
| K13 | 6,360 | 46,324,230 | 0.17 | 0.13 | 0.13 | 0.04 | 0.04 | 0.03 | 1.30 [1.20-1.41]* | 1.26 [1.16-1.37]* |
| K14 | 1,920 | 46,387,870 | 0.05 | 0.04 | 0.04 | 0.01 | 0.01 | 0.01 | 1.32 [1.14-1.53]* | 1.26 [1.09-1.47]* |
| K20 | 4,150 | 46,378,550 | 0.12 | 0.08 | 0.08 | 0.03 | 0.04 | 0.03 | 1.49 [1.35-1.64]* | 1.34 [1.21-1.49]* |
| K21 | 20,450 | 45,682,780 | 0.50 | 0.43 | 0.40 | 0.07 | 0.09 | 0.05 | 1.23 [1.18-1.29]* | 1.11 [1.06-1.17]* |
| K22 | 4,700 | 46,359,560 | 0.11 | 0.10 | 0.09 | 0.01 | 0.02 | 0.01 | 1.21 [1.10-1.33]* | 1.12 [1.01-1.24] |
| K25 | 1,540 | 46,405,300 | 0.04 | 0.03 | 0.03 | 0.01 | 0.01 | 0.01 | 1.30 [1.10-1.54]* | 1.27 [1.07-1.52]* |
| K26 | 1,110 | 46,408,850 | 0.03 | 0.02 | 0.02 | 0.00 | 0.00 | 0.00 | 1.21 [0.99-1.48] | 1.18 [0.95-1.46] |
| K29 | 22,420 | 46,150,230 | 0.55 | 0.47 | 0.45 | 0.08 | 0.10 | 0.08 | 1.23 [1.17-1.28]* | 1.16 [1.11-1.22]* |
| K30 | 3,350 | 46,382,730 | 0.07 | 0.07 | 0.06 | 0.00 | 0.01 | 0.01 | 1.19 [1.06-1.34]* | 1.12 [0.99-1.26] |
| K31 | 2,820 | 46,389,310 | 0.06 | 0.06 | 0.06 | 0.00 | 0.01 | 0.00 | 1.11 [0.97-1.26] | 1.04 [0.91-1.19] |
| K35 | 35,200 | 45,972,980 | 0.79 | 0.76 | 0.75 | 0.03 | 0.03 | 0.02 | 1.04 [1.00-1.08] | 1.02 [0.98-1.06] |
| K36 | 580 | 46,411,650 | 0.01 | 0.01 | 0.01 | 0.00 | 0.00 | 0.00 | 1.35 [1.03-1.76] | 1.36 [1.03-1.79] |
| K37 | 7,090 | 46,334,220 | 0.16 | 0.15 | 0.15 | 0.01 | 0.01 | 0.01 | 1.08 [0.99-1.17] | 1.04 [0.95-1.13] |
| K38 | 3,670 | 46,381,200 | 0.08 | 0.08 | 0.08 | 0.01 | 0.01 | 0.00 | 1.08 [0.97-1.21] | 1.05 [0.94-1.18] |
| K40 | 14,550 | 45,882,350 | 0.31 | 0.32 | 0.31 | 0.00 | 0.01 | −0.01 | 1.02 [0.96-1.08] | 0.98 [0.93-1.04] |
| K42 | 6,960 | 46,207,300 | 0.17 | 0.15 | 0.14 | 0.02 | 0.03 | 0.01 | 1.19 [1.10-1.28]* | 1.09 [1.00-1.18] |
| K43 | 2,800 | 46,368,610 | 0.06 | 0.06 | 0.05 | 0.00 | 0.01 | 0.00 | 1.13 [1.00-1.28] | 1.05 [0.92-1.20] |
| K44 | 7,250 | 46,367,100 | 0.17 | 0.15 | 0.13 | 0.02 | 0.04 | 0.03 | 1.27 [1.17-1.38]* | 1.17 [1.08-1.28]* |
| K50 | 5,460 | 46,361,370 | 0.17 | 0.10 | 0.10 | 0.07 | 0.08 | 0.07 | 1.80 [1.65-1.96]* | 1.67 [1.53-1.82]* |
| K51 | 4,730 | 46,374,900 | 0.13 | 0.10 | 0.09 | 0.03 | 0.04 | 0.03 | 1.44 [1.31-1.58]* | 1.36 [1.23-1.50]* |
| K52 | 31,400 | 45,010,920 | 0.77 | 0.68 | 0.63 | 0.09 | 0.14 | 0.12 | 1.22 [1.18-1.27]* | 1.19 [1.14-1.23]* |
| K56 | 5,940 | 46,320,500 | 0.14 | 0.13 | 0.13 | 0.01 | 0.01 | 0.01 | 1.08 [0.99-1.18] | 1.04 [0.95-1.14] |
| K57 | 1,080 | 46,411,950 | 0.02 | 0.02 | 0.02 | 0.00 | 0.00 | 0.00 | 1.15 [0.93-1.43] | 1.06 [0.84-1.32] |
| K58 | 12,600 | 46,336,490 | 0.30 | 0.26 | 0.24 | 0.04 | 0.07 | 0.05 | 1.29 [1.21-1.37]* | 1.21 [1.14-1.29]* |
| K59 | 50,040 | 45,515,250 | 1.26 | 1.06 | 1.05 | 0.20 | 0.20 | 0.16 | 1.19 [1.16-1.23]* | 1.15 [1.11-1.18]* |
| K60 | 4,900 | 46,337,650 | 0.13 | 0.10 | 0.09 | 0.03 | 0.04 | 0.03 | 1.40 [1.28-1.53]* | 1.34 [1.22-1.47]* |
| K61 | 4,140 | 46,356,790 | 0.10 | 0.09 | 0.08 | 0.02 | 0.03 | 0.03 | 1.32 [1.20-1.47]* | 1.32 [1.19-1.47]* |
| K62 | 11,350 | 46,253,380 | 0.29 | 0.23 | 0.21 | 0.05 | 0.07 | 0.06 | 1.34 [1.26-1.42]* | 1.26 [1.18-1.35]* |
| K63 | 5,740 | 46,365,800 | 0.15 | 0.12 | 0.11 | 0.03 | 0.04 | 0.03 | 1.40 [1.29-1.53]* | 1.30 [1.19-1.42]* |
| K64 | 6,340 | 46,392,880 | 0.16 | 0.13 | 0.12 | 0.03 | 0.04 | 0.03 | 1.34 [1.23-1.46]* | 1.27 [1.16-1.39]* |
| K65 | 1,420 | 46,403,100 | 0.03 | 0.03 | 0.03 | 0.00 | 0.00 | 0.00 | 1.05 [0.88-1.26] | 1.06 [0.88-1.28] |
| K66 | 6,890 | 46,368,100 | 0.15 | 0.15 | 0.14 | 0.00 | 0.02 | 0.01 | 1.11 [1.02-1.21] | 1.07 [0.99-1.17] |
| K72 | 650 | 46,410,730 | 0.01 | 0.01 | 0.01 | 0.00 | 0.00 | 0.00 | 0.89 [0.67-1.18] | 0.88 [0.66-1.17] |
| K75 | 890 | 46,408,700 | 0.02 | 0.02 | 0.02 | 0.01 | 0.01 | 0.01 | 1.33 [1.07-1.66] | 1.29 [1.03-1.63] |
| K76 | 4,220 | 46,386,910 | 0.10 | 0.09 | 0.09 | 0.01 | 0.01 | 0.01 | 1.12 [1.00-1.24] | 1.07 [0.96-1.20] |
| K80 | 9,980 | 46,342,480 | 0.20 | 0.22 | 0.19 | −0.02 | 0.01 | 0.00 | 1.05 [0.97-1.12] | 1.02 [0.95-1.10] |
| K81 | 1,700 | 46,406,240 | 0.03 | 0.04 | 0.03 | 0.00 | 0.00 | 0.00 | 1.06 [0.89-1.26] | 1.03 [0.86-1.24] |
| K82 | 1,790 | 46,406,070 | 0.03 | 0.04 | 0.04 | 0.00 | 0.00 | 0.00 | 0.99 [0.84-1.17] | 0.96 [0.81-1.15] |
| K83 | 1,910 | 46,396,030 | 0.04 | 0.04 | 0.04 | 0.00 | 0.00 | 0.00 | 1.05 [0.89-1.23] | 1.01 [0.85-1.19] |
| K85 | 2,460 | 46,399,400 | 0.05 | 0.05 | 0.05 | 0.00 | 0.00 | 0.00 | 1.03 [0.89-1.18] | 1.02 [0.88-1.18] |
| K86 | 1,330 | 46,405,120 | 0.02 | 0.03 | 0.03 | −0.01 | 0.00 | 0.00 | 0.86 [0.70-1.05] | 0.84 [0.69-1.03] |
| K90 | 10,230 | 46,202,780 | 0.36 | 0.18 | 0.18 | 0.18 | 0.19 | 0.16 | 2.04 [1.93-2.17]* | 1.76 [1.65-1.87]* |
| K91 | 2,600 | 46,377,470 | 0.06 | 0.06 | 0.06 | 0.00 | 0.00 | 0.00 | 1.06 [0.93-1.21] | 0.99 [0.86-1.13] |
| K92 | 19,970 | 46,176,390 | 0.47 | 0.42 | 0.40 | 0.05 | 0.08 | 0.05 | 1.19 [1.14-1.25]* | 1.13 [1.08-1.19]* |
| <18 - L00-L99 | | | | | | | | | | |
| L00 | 690 | 46,378,430 | 0.02 | 0.01 | 0.01 | 0.01 | 0.01 | 0.01 | 1.55 [1.23-1.94]* | 1.54 [1.22-1.94]* |
| L01 | 2,750 | 46,316,580 | 0.10 | 0.05 | 0.05 | 0.06 | 0.06 | 0.05 | 2.22 [2.00-2.48]* | 2.09 [1.87-2.33]* |
| L02 | 14,140 | 46,174,860 | 0.36 | 0.29 | 0.28 | 0.07 | 0.08 | 0.07 | 1.29 [1.22-1.36]* | 1.26 [1.19-1.33]* |
| L03 | 19,260 | 46,107,130 | 0.53 | 0.39 | 0.38 | 0.14 | 0.14 | 0.13 | 1.38 [1.32-1.44]* | 1.34 [1.28-1.40]* |
| L04 | 3,950 | 46,337,620 | 0.10 | 0.08 | 0.08 | 0.01 | 0.01 | 0.01 | 1.14 [1.03-1.26] | 1.12 [1.01-1.25] |
| L05 | 10,020 | 46,196,660 | 0.22 | 0.22 | 0.20 | 0.00 | 0.02 | 0.01 | 1.09 [1.02-1.17] | 1.06 [0.98-1.14] |
| L08 | 6,040 | 46,276,120 | 0.19 | 0.11 | 0.11 | 0.07 | 0.08 | 0.08 | 1.72 [1.59-1.86]* | 1.68 [1.55-1.82]* |
| L20 | 4,620 | 46,203,740 | 0.39 | 0.02 | 0.02 | 0.37 | 0.37 | 0.37 | 18.85 [16.88-21.04]* | 16.21 [14.49-18.13]* |
| L21 | 250 | 46,378,120 | 0.01 | 0.00 | 0.00 | 0.01 | 0.01 | 0.01 | 3.22 [2.24-4.64]* | 3.12 [2.12-4.57]* |
| L22 | 1,270 | 46,310,890 | 0.03 | 0.03 | 0.03 | 0.00 | 0.00 | 0.00 | 1.01 [0.84-1.21] | 0.95 [0.78-1.15] |
| L23 | 880 | 46,399,270 | 0.05 | 0.01 | 0.01 | 0.03 | 0.04 | 0.03 | 4.13 [3.41-5.01]* | 3.36 [2.75-4.12]* |
| L25 | 480 | 46,408,790 | 0.02 | 0.01 | 0.01 | 0.01 | 0.01 | 0.01 | 1.95 [1.49-2.56]* | 1.80 [1.36-2.37]* |
| L27 | 2,290 | 46,374,940 | 0.08 | 0.04 | 0.04 | 0.04 | 0.04 | 0.04 | 2.18 [1.93-2.47]* | 1.91 [1.68-2.18]* |
| L28 | 330 | 46,412,720 | 0.02 | 0.00 | 0.00 | 0.02 | 0.02 | 0.02 | 6.73 [4.77-9.50]* | 6.04 [4.24-8.60]* |
| L29 | 3,750 | 46,384,700 | 0.10 | 0.08 | 0.07 | 0.03 | 0.03 | 0.03 | 1.46 [1.31-1.62]* | 1.36 [1.21-1.51]* |
| L30 | 57,680 | 45,032,470 | 4.41 | 0.45 | 0.44 | 3.96 | 3.96 | 3.92 | 9.92 [9.66-10.19]* | 9.12 [8.88-9.37]* |
| L40 | 3,650 | 46,386,640 | 0.12 | 0.07 | 0.06 | 0.05 | 0.06 | 0.06 | 2.04 [1.84-2.26]* | 2.01 [1.80-2.23]* |
| L50 | 7,050 | 46,279,420 | 0.25 | 0.12 | 0.13 | 0.13 | 0.13 | 0.11 | 2.03 [1.89-2.17]* | 1.75 [1.62-1.88]* |
| L51 | 2,000 | 46,370,980 | 0.04 | 0.04 | 0.04 | 0.00 | 0.00 | 0.00 | 0.97 [0.83-1.13] | 0.95 [0.82-1.11] |
| L52 | 510 | 46,411,610 | 0.01 | 0.01 | 0.01 | 0.00 | 0.00 | 0.00 | 1.47 [1.11-1.96]* | 1.36 [1.01-1.82] |
| L53 | 2,830 | 46,358,490 | 0.08 | 0.06 | 0.06 | 0.02 | 0.02 | 0.02 | 1.45 [1.29-1.63]* | 1.37 [1.21-1.54]* |
| L60 | 6,790 | 46,320,020 | 0.18 | 0.14 | 0.13 | 0.04 | 0.04 | 0.03 | 1.32 [1.22-1.43]* | 1.24 [1.14-1.34]* |
| L65 | 730 | 46,410,610 | 0.02 | 0.01 | 0.01 | 0.01 | 0.01 | 0.01 | 1.78 [1.41-2.24]* | 1.64 [1.29-2.08]* |
| L70 | 1,880 | 46,402,950 | 0.05 | 0.04 | 0.04 | 0.01 | 0.01 | 0.01 | 1.37 [1.17-1.59]* | 1.27 [1.08-1.49]* |
| L72 | 8,300 | 46,302,020 | 0.18 | 0.18 | 0.16 | 0.00 | 0.02 | 0.01 | 1.10 [1.02-1.19] | 1.07 [0.99-1.16] |
| L73 | 1,620 | 46,403,510 | 0.04 | 0.03 | 0.03 | 0.01 | 0.01 | 0.01 | 1.32 [1.12-1.55]* | 1.24 [1.05-1.47] |
| L80 | 280 | 46,415,240 | 0.01 | 0.00 | 0.00 | 0.01 | 0.01 | 0.01 | 2.46 [1.72-3.53]* | 2.34 [1.60-3.42]* |
| L81 | 1,330 | 46,389,820 | 0.04 | 0.03 | 0.03 | 0.01 | 0.01 | 0.01 | 1.50 [1.26-1.78]* | 1.45 [1.21-1.72]* |
| L83 | 470 | 46,413,550 | 0.02 | 0.01 | 0.01 | 0.01 | 0.01 | 0.01 | 1.58 [1.18-2.12]* | 1.51 [1.11-2.04]* |
| L85 | 490 | 46,411,350 | 0.02 | 0.01 | 0.01 | 0.01 | 0.01 | 0.01 | 2.18 [1.67-2.84]* | 2.18 [1.66-2.87]* |
| L89 | 1,310 | 46,405,190 | 0.03 | 0.03 | 0.03 | 0.00 | 0.00 | 0.00 | 0.99 [0.82-1.20] | 0.97 [0.80-1.18] |
| L90 | 4,070 | 46,358,720 | 0.09 | 0.09 | 0.09 | 0.00 | 0.00 | 0.00 | 1.04 [0.94-1.16] | 1.02 [0.92-1.14] |
| L91 | 3,380 | 46,356,840 | 0.08 | 0.07 | 0.07 | 0.01 | 0.02 | 0.02 | 1.26 [1.12-1.40]* | 1.25 [1.11-1.40]* |
| L92 | 2,300 | 46,369,450 | 0.05 | 0.05 | 0.05 | 0.01 | 0.01 | 0.00 | 1.11 [0.97-1.28] | 1.04 [0.90-1.20] |
| L94 | 440 | 46,413,380 | 0.01 | 0.01 | 0.01 | 0.01 | 0.00 | 0.00 | 1.53 [1.14-2.06]* | 1.54 [1.13-2.09]* |
| L97 | 450 | 46,413,760 | 0.01 | 0.01 | 0.01 | 0.00 | 0.00 | 0.00 | 1.28 [0.94-1.75] | 1.34 [0.97-1.85] |
| L98 | 7,640 | 46,283,450 | 0.19 | 0.16 | 0.15 | 0.04 | 0.04 | 0.04 | 1.28 [1.19-1.38]* | 1.25 [1.16-1.35]* |
| <18 - M00-M99 | | | | | | | | | | |
| M00 | 1,530 | 46,383,650 | 0.04 | 0.03 | 0.03 | 0.01 | 0.01 | 0.01 | 1.40 [1.20-1.64]* | 1.37 [1.16-1.61]* |
| M02 | 1,340 | 46,397,250 | 0.03 | 0.03 | 0.03 | 0.00 | 0.00 | 0.00 | 1.13 [0.94-1.34] | 1.11 [0.93-1.33] |
| M06 | 1,190 | 46,409,890 | 0.03 | 0.02 | 0.02 | 0.00 | 0.01 | 0.00 | 1.29 [1.06-1.57] | 1.21 [0.99-1.49] |

| | | | | | | | | | | |
|---|---|---|---|---|---|---|---|---|---|---|
| M07 | 520 | 46,414,600 | 0.01 | 0.01 | 0.01 | 0.00 | 0.00 | 0.00 | 1.52 [1.14-2.03]* | 1.46 [1.08-1.97] |
| M08 | 2,670 | 46,373,880 | 0.07 | 0.05 | 0.05 | 0.02 | 0.02 | 0.02 | 1.34 [1.19-1.51]* | 1.30 [1.15-1.47]* |
| M13 | 2,500 | 46,388,440 | 0.06 | 0.05 | 0.05 | 0.01 | 0.02 | 0.01 | 1.36 [1.19-1.55]* | 1.27 [1.11-1.45]* |
| M16 | 570 | 46,414,570 | 0.01 | 0.01 | 0.01 | 0.00 | 0.00 | 0.00 | 1.12 [0.84-1.49] | 1.08 [0.80-1.46] |
| M17 | 1,760 | 46,404,790 | 0.04 | 0.04 | 0.03 | 0.00 | 0.00 | 0.00 | 1.14 [0.96-1.35] | 1.09 [0.91-1.30] |
| M19 | 1,450 | 46,409,170 | 0.03 | 0.03 | 0.03 | 0.00 | 0.01 | 0.00 | 1.26 [1.05-1.51] | 1.14 [0.94-1.39] |
| M20 | 4,620 | 46,360,320 | 0.11 | 0.10 | 0.09 | 0.01 | 0.02 | 0.01 | 1.19 [1.08-1.32]* | 1.15 [1.04-1.27]* |
| M21 | 6,080 | 46,345,830 | 0.14 | 0.13 | 0.14 | 0.01 | 0.00 | 0.00 | 1.00 [0.92-1.09] | 0.97 [0.88-1.06] |
| M22 | 2,600 | 46,391,030 | 0.06 | 0.05 | 0.05 | 0.01 | 0.01 | 0.01 | 1.26 [1.10-1.44]* | 1.18 [1.03-1.35] |
| M23 | 14,610 | 46,282,650 | 0.32 | 0.31 | 0.28 | 0.00 | 0.04 | 0.03 | 1.14 [1.08-1.21]* | 1.09 [1.03-1.16]* |
| M24 | 10,700 | 46,244,310 | 0.24 | 0.23 | 0.21 | 0.01 | 0.03 | 0.02 | 1.15 [1.08-1.23]* | 1.10 [1.03-1.18]* |
| M25 | 29,840 | 46,051,740 | 0.74 | 0.62 | 0.62 | 0.12 | 0.13 | 0.10 | 1.21 [1.16-1.25]* | 1.15 [1.10-1.19]* |
| M30 | 1,200 | 46,390,000 | 0.03 | 0.02 | 0.02 | 0.01 | 0.01 | 0.01 | 1.28 [1.07-1.53]* | 1.29 [1.08-1.55]* |
| M32 | 510 | 46,413,280 | 0.02 | 0.01 | 0.01 | 0.01 | 0.01 | 0.01 | 1.73 [1.30-2.30]* | 1.73 [1.29-2.32]* |
| M35 | 5,300 | 46,382,020 | 0.15 | 0.10 | 0.10 | 0.04 | 0.04 | 0.04 | 1.43 [1.31-1.56]* | 1.32 [1.21-1.45]* |
| M40 | 810 | 46,410,480 | 0.02 | 0.02 | 0.02 | 0.00 | 0.00 | 0.00 | 1.04 [0.82-1.32] | 0.98 [0.77-1.26] |
| M41 | 7,660 | 46,342,280 | 0.18 | 0.16 | 0.17 | 0.01 | 0.01 | 0.01 | 1.07 [0.99-1.15] | 1.03 [0.96-1.12] |
| M43 | 2,600 | 46,382,240 | 0.06 | 0.06 | 0.06 | 0.00 | 0.00 | 0.00 | 1.02 [0.90-1.17] | 0.98 [0.85-1.12] |
| M46 | 600 | 46,411,340 | 0.01 | 0.01 | 0.01 | 0.00 | 0.00 | 0.00 | 1.10 [0.83-1.45] | 1.08 [0.81-1.44] |
| M47 | 1,000 | 46,412,300 | 0.02 | 0.02 | 0.02 | 0.00 | 0.00 | 0.00 | 1.07 [0.85-1.35] | 1.00 [0.79-1.27] |
| M48 | 900 | 46,412,470 | 0.02 | 0.02 | 0.02 | 0.00 | 0.00 | 0.00 | 1.12 [0.88-1.41] | 1.05 [0.82-1.33] |
| M51 | 4,230 | 46,391,040 | 0.09 | 0.09 | 0.08 | 0.00 | 0.02 | 0.01 | 1.24 [1.11-1.38]* | 1.17 [1.04-1.30]* |
| M53 | 670 | 46,410,130 | 0.01 | 0.01 | 0.01 | 0.00 | 0.00 | 0.00 | 1.03 [0.78-1.36] | 0.99 [0.75-1.31] |
| M54 | 26,110 | 46,193,160 | 0.59 | 0.56 | 0.52 | 0.04 | 0.08 | 0.05 | 1.15 [1.10-1.19]* | 1.09 [1.04-1.14]* |
| M60 | 1,490 | 46,401,730 | 0.04 | 0.03 | 0.03 | 0.01 | 0.01 | 0.01 | 1.35 [1.15-1.59]* | 1.31 [1.11-1.55]* |
| M62 | 3,390 | 46,380,020 | 0.07 | 0.07 | 0.08 | 0.00 | 0.00 | 0.00 | 0.99 [0.88-1.12] | 0.94 [0.83-1.06] |
| M65 | 6,860 | 46,323,100 | 0.17 | 0.14 | 0.14 | 0.03 | 0.04 | 0.03 | 1.26 [1.17-1.37]* | 1.23 [1.13-1.33]* |
| M67 | 8,910 | 46,302,020 | 0.21 | 0.19 | 0.18 | 0.02 | 0.02 | 0.02 | 1.13 [1.05-1.21]* | 1.08 [1.01-1.16] |
| M70 | 890 | 46,408,790 | 0.02 | 0.02 | 0.02 | 0.00 | 0.00 | 0.00 | 1.16 [0.93-1.46] | 1.13 [0.89-1.43] |
| M71 | 400 | 46,413,360 | 0.01 | 0.01 | 0.01 | 0.00 | 0.00 | 0.00 | 1.51 [1.09-2.08] | 1.44 [1.04-2.02] |
| M72 | 500 | 46,412,970 | 0.01 | 0.01 | 0.01 | 0.00 | 0.00 | 0.00 | 1.23 [0.91-1.67] | 1.18 [0.85-1.62] |
| M75 | 2,060 | 46,397,090 | 0.04 | 0.04 | 0.04 | 0.00 | 0.01 | 0.00 | 1.18 [1.01-1.37] | 1.12 [0.96-1.31] |
| M76 | 610 | 46,412,830 | 0.02 | 0.01 | 0.01 | 0.00 | 0.00 | 0.00 | 1.24 [0.95-1.63] | 1.13 [0.85-1.50] |
| M77 | 680 | 46,412,700 | 0.01 | 0.01 | 0.01 | 0.00 | 0.00 | 0.00 | 0.95 [0.73-1.26] | 0.88 [0.66-1.17] |
| M79 | 24,580 | 46,138,930 | 0.60 | 0.52 | 0.50 | 0.08 | 0.10 | 0.08 | 1.20 [1.15-1.25]* | 1.15 [1.10-1.20]* |
| M81 | 780 | 46,411,960 | 0.02 | 0.01 | 0.01 | 0.00 | 0.01 | 0.00 | 1.38 [1.09-1.75]* | 1.30 [1.02-1.66] |
| M84 | 4,090 | 46,364,800 | 0.09 | 0.09 | 0.08 | 0.00 | 0.01 | 0.01 | 1.11 [1.00-1.23] | 1.08 [0.96-1.20] |
| M85 | 1,820 | 46,396,390 | 0.04 | 0.04 | 0.04 | 0.01 | 0.01 | 0.01 | 1.18 [1.01-1.39] | 1.13 [0.96-1.33] |
| M86 | 2,210 | 46,377,340 | 0.06 | 0.04 | 0.04 | 0.02 | 0.02 | 0.02 | 1.36 [1.19-1.56]* | 1.33 [1.16-1.53]* |
| M87 | 620 | 46,410,470 | 0.01 | 0.01 | 0.01 | 0.00 | 0.00 | 0.00 | 1.22 [0.93-1.59] | 1.16 [0.88-1.53] |
| M89 | 3,700 | 46,367,470 | 0.09 | 0.08 | 0.08 | 0.01 | 0.01 | 0.01 | 1.12 [1.01-1.25] | 1.09 [0.97-1.22] |
| M91 | 1,170 | 46,394,180 | 0.03 | 0.02 | 0.02 | 0.00 | 0.00 | 0.00 | 1.16 [0.96-1.39] | 1.12 [0.92-1.35] |
| M92 | 1,360 | 46,403,520 | 0.04 | 0.03 | 0.03 | 0.01 | 0.01 | 0.01 | 1.31 [1.10-1.56]* | 1.19 [0.99-1.43] |
| M93 | 2,310 | 46,386,780 | 0.06 | 0.05 | 0.05 | 0.01 | 0.01 | 0.01 | 1.25 [1.09-1.43]* | 1.18 [1.03-1.36] |
| M94 | 2,870 | 46,395,940 | 0.07 | 0.06 | 0.06 | 0.01 | 0.01 | 0.01 | 1.24 [1.09-1.40]* | 1.18 [1.04-1.35]* |
| M95 | 1,220 | 46,402,980 | 0.03 | 0.03 | 0.02 | 0.00 | 0.00 | 0.00 | 1.21 [1.00-1.46] | 1.20 [0.98-1.46] |
| <18 - N00-N99 | | | | | | | | | | |
| N02 | 740 | 46,409,020 | 0.01 | 0.02 | 0.02 | 0.00 | 0.00 | 0.00 | 0.86 [0.66-1.12] | 0.84 [0.64-1.10] |
| N04 | 1,260 | 46,393,120 | 0.03 | 0.03 | 0.03 | 0.00 | 0.00 | 0.00 | 0.99 [0.82-1.20] | 0.98 [0.81-1.19] |
| N05 | 1,200 | 46,399,460 | 0.03 | 0.02 | 0.02 | 0.01 | 0.01 | 0.01 | 1.36 [1.13-1.63]* | 1.32 [1.10-1.60]* |
| N10 | 1,850 | 46,399,340 | 0.04 | 0.04 | 0.04 | 0.00 | 0.00 | 0.00 | 1.07 [0.92-1.26] | 1.05 [0.89-1.24] |
| N12 | 10,270 | 46,324,510 | 0.22 | 0.22 | 0.21 | 0.00 | 0.01 | 0.00 | 1.05 [0.98-1.12] | 1.02 [0.95-1.10] |
| N13 | 6,560 | 46,284,850 | 0.14 | 0.14 | 0.13 | 0.00 | 0.01 | 0.00 | 1.05 [0.97-1.15] | 1.02 [0.94-1.12] |
| N17 | 6,260 | 46,371,570 | 0.14 | 0.13 | 0.13 | 0.00 | 0.01 | 0.01 | 1.07 [0.98-1.17] | 1.07 [0.97-1.17] |
| N18 | 1,730 | 46,396,910 | 0.04 | 0.04 | 0.03 | 0.00 | 0.00 | 0.00 | 1.14 [0.97-1.34] | 1.12 [0.95-1.33] |
| N19 | 1,440 | 46,390,540 | 0.03 | 0.03 | 0.03 | 0.00 | 0.00 | 0.00 | 1.08 [0.91-1.29] | 1.05 [0.87-1.26] |
| N20 | 4,090 | 46,384,780 | 0.07 | 0.09 | 0.08 | −0.02 | −0.01 | −0.01 | 0.94 [0.84-1.05] | 0.92 [0.81-1.03] |
| N23 | 1,700 | 46,401,970 | 0.03 | 0.04 | 0.03 | 0.00 | 0.00 | 0.00 | 1.03 [0.87-1.22] | 0.98 [0.83-1.18] |
| N27 | 580 | 46,408,770 | 0.01 | 0.01 | 0.01 | 0.00 | 0.00 | 0.00 | 0.96 [0.73-1.27] | 0.94 [0.71-1.25] |
| N28 | 3,820 | 46,342,330 | 0.08 | 0.08 | 0.08 | 0.00 | 0.00 | 0.00 | 0.99 [0.89-1.11] | 0.98 [0.88-1.10] |
| N30 | 2,060 | 46,397,820 | 0.05 | 0.04 | 0.04 | 0.01 | 0.01 | 0.01 | 1.40 [1.21-1.62]* | 1.33 [1.15-1.55]* |
| N31 | 1,520 | 46,396,590 | 0.03 | 0.03 | 0.03 | 0.00 | 0.00 | 0.00 | 0.98 [0.82-1.16] | 0.95 [0.79-1.13] |
| N32 | 4,700 | 46,371,770 | 0.11 | 0.10 | 0.10 | 0.02 | 0.02 | 0.01 | 1.16 [1.05-1.27]* | 1.10 [0.99-1.21] |
| N35 | 3,260 | 46,375,500 | 0.08 | 0.07 | 0.06 | 0.02 | 0.02 | 0.02 | 1.33 [1.19-1.49]* | 1.29 [1.15-1.45]* |
| N36 | 1,260 | 46,398,140 | 0.03 | 0.03 | 0.03 | 0.00 | 0.00 | 0.00 | 1.03 [0.85-1.24] | 0.99 [0.82-1.21] |
| N39 | 39,040 | 45,467,300 | 0.91 | 0.84 | 0.81 | 0.07 | 0.10 | 0.08 | 1.12 [1.09-1.16]* | 1.09 [1.05-1.13]* |
| N43 | 9,220 | 46,226,250 | 0.21 | 0.20 | 0.20 | 0.01 | 0.01 | 0.01 | 1.05 [0.98-1.12] | 1.02 [0.96-1.10] |
| N44 | 6,960 | 46,332,880 | 0.18 | 0.14 | 0.16 | 0.03 | 0.02 | 0.02 | 1.14 [1.05-1.23]* | 1.11 [1.03-1.21]* |
| N45 | 5,160 | 46,357,660 | 0.12 | 0.11 | 0.11 | 0.01 | 0.01 | 0.01 | 1.12 [1.02-1.23] | 1.09 [0.99-1.20] |
| N47 | 35,010 | 45,707,570 | 1.01 | 0.70 | 0.69 | 0.31 | 0.31 | 0.29 | 1.45 [1.41-1.50]* | 1.41 [1.36-1.46]* |
| N48 | 12,360 | 46,206,640 | 0.34 | 0.25 | 0.25 | 0.09 | 0.09 | 0.08 | 1.35 [1.28-1.43]* | 1.32 [1.24-1.40]* |
| N49 | 730 | 46,405,580 | 0.02 | 0.02 | 0.02 | 0.00 | 0.00 | 0.00 | 1.13 [0.88-1.44] | 1.11 [0.87-1.43] |
| N50 | 12,460 | 46,248,750 | 0.30 | 0.26 | 0.27 | 0.04 | 0.03 | 0.02 | 1.12 [1.05-1.19]* | 1.08 [1.02-1.15] |
| N60 | 510 | 46,413,220 | 0.01 | 0.01 | 0.01 | 0.00 | 0.00 | 0.00 | 1.37 [1.01-1.85] | 1.34 [0.98-1.84] |
| N61 | 1,100 | 46,402,960 | 0.03 | 0.02 | 0.02 | 0.00 | 0.01 | 0.00 | 1.26 [1.03-1.54] | 1.22 [0.99-1.51] |
| N62 | 1,400 | 46,398,490 | 0.03 | 0.03 | 0.03 | 0.01 | 0.01 | 0.01 | 1.30 [1.09-1.55]* | 1.26 [1.05-1.51] |
| N63 | 740 | 46,409,290 | 0.02 | 0.02 | 0.01 | 0.00 | 0.00 | 0.00 | 1.19 [0.93-1.53] | 1.16 [0.90-1.50] |
| N64 | 900 | 46,409,530 | 0.02 | 0.02 | 0.02 | 0.00 | 0.01 | 0.01 | 1.37 [1.09-1.71]* | 1.35 [1.07-1.70] |
| N70 | 900 | 46,411,510 | 0.02 | 0.02 | 0.02 | 0.00 | 0.00 | 0.00 | 1.08 [0.85-1.37] | 1.06 [0.83-1.35] |
| N72 | 690 | 46,413,020 | 0.01 | 0.02 | 0.01 | 0.00 | 0.00 | 0.00 | 0.95 [0.72-1.26] | 0.96 [0.72-1.28] |
| N73 | 6,510 | 46,366,710 | 0.14 | 0.14 | 0.12 | 0.00 | 0.01 | 0.01 | 1.11 [1.01-1.21] | 1.08 [0.98-1.18] |
| N75 | 1,660 | 46,404,420 | 0.03 | 0.04 | 0.03 | −0.01 | 0.00 | 0.00 | 0.93 [0.77-1.11] | 0.93 [0.77-1.12] |
| N76 | 3,120 | 46,378,000 | 0.08 | 0.06 | 0.06 | 0.01 | 0.01 | 0.01 | 1.22 [1.09-1.38]* | 1.17 [1.04-1.32]* |
| N77 | 4,470 | 46,375,890 | 0.09 | 0.10 | 0.08 | 0.00 | 0.01 | 0.01 | 1.10 [0.99-1.22] | 1.06 [0.95-1.18] |
| N80 | 6,590 | 46,372,750 | 0.15 | 0.14 | 0.13 | 0.00 | 0.02 | 0.02 | 1.16 [1.06-1.27]* | 1.12 [1.02-1.22] |
| N81 | 600 | 46,414,220 | 0.01 | 0.01 | 0.01 | 0.00 | 0.00 | 0.00 | 1.27 [0.95-1.69] | 1.26 [0.94-1.70] |
| N83 | 14,260 | 46,300,080 | 0.32 | 0.31 | 0.28 | 0.01 | 0.03 | 0.02 | 1.11 [1.05-1.17]* | 1.06 [1.00-1.13] |
| N84 | 930 | 46,411,810 | 0.02 | 0.02 | 0.02 | 0.00 | 0.00 | 0.00 | 1.17 [0.93-1.48] | 1.09 [0.86-1.39] |
| N85 | 2,370 | 46,403,460 | 0.05 | 0.05 | 0.04 | 0.00 | 0.01 | 0.01 | 1.17 [1.01-1.35] | 1.13 [0.97-1.32] |
| N86 | 3,150 | 46,394,140 | 0.06 | 0.07 | 0.06 | −0.01 | 0.00 | 0.00 | 1.07 [0.94-1.22] | 1.04 [0.91-1.19] |
| N87 | 2,350 | 46,399,970 | 0.05 | 0.05 | 0.04 | −0.01 | 0.01 | 0.01 | 1.13 [0.98-1.31] | 1.15 [0.98-1.34] |
| N88 | 470 | 46,414,920 | 0.01 | 0.01 | 0.01 | 0.00 | 0.00 | 0.00 | 1.08 [0.78-1.49] | 1.10 [0.78-1.55] |
| N89 | 5,590 | 46,356,520 | 0.12 | 0.12 | 0.11 | 0.00 | 0.01 | 0.01 | 1.10 [1.00-1.21] | 1.07 [0.97-1.17] |
| N90 | 2,440 | 46,389,420 | 0.06 | 0.05 | 0.05 | 0.00 | 0.01 | 0.01 | 1.23 [1.08-1.41]* | 1.22 [1.06-1.41]* |
| N91 | 1,030 | 46,409,930 | 0.02 | 0.02 | 0.02 | 0.00 | 0.00 | 0.00 | 1.14 [0.92-1.41] | 1.11 [0.89-1.38] |
| N92 | 9,390 | 46,352,420 | 0.23 | 0.20 | 0.19 | 0.03 | 0.04 | 0.03 | 1.21 [1.13-1.30]* | 1.16 [1.08-1.25]* |
| N93 | 8,160 | 46,339,660 | 0.18 | 0.18 | 0.15 | 0.00 | 0.02 | 0.02 | 1.14 [1.06-1.23]* | 1.11 [1.02-1.20] |
| N94 | 7,730 | 46,350,760 | 0.18 | 0.16 | 0.15 | 0.02 | 0.04 | 0.03 | 1.24 [1.15-1.34]* | 1.18 [1.09-1.28]* |
| N99 | 950 | 46,403,080 | 0.02 | 0.02 | 0.02 | 0.00 | 0.00 | 0.00 | 1.01 [0.80-1.26] | 0.99 [0.79-1.25] |
| <18 - O00-O9A | | | | | | | | | | |
| O00 | 3,220 | 46,393,840 | 0.06 | 0.07 | 0.06 | −0.01 | 0.00 | 0.00 | 1.02 [0.90-1.16] | 0.99 [0.87-1.13] |
| O02 | 6,150 | 46,364,790 | 0.12 | 0.14 | 0.11 | −0.02 | 0.01 | 0.00 | 1.05 [0.95-1.15] | 1.04 [0.94-1.14] |
| O03 | 11,330 | 46,320,250 | 0.22 | 0.25 | 0.22 | −0.04 | 0.00 | 0.00 | 1.00 [0.94-1.07] | 0.99 [0.92-1.06] |
| O04 | 28,250 | 46,116,890 | 0.55 | 0.63 | 0.56 | −0.08 | −0.01 | −0.01 | 0.98 [0.94-1.03] | 0.98 [0.94-1.03] |
| O08 | 1,690 | 46,404,950 | 0.03 | 0.04 | 0.03 | 0.00 | 0.00 | 0.00 | 0.99 [0.83-1.18] | 0.99 [0.83-1.18] |
| O10 | 710 | 46,413,940 | 0.01 | 0.02 | 0.01 | 0.00 | 0.00 | 0.00 | 0.95 [0.72-1.25] | 0.95 [0.71-1.27] |

| | | | | | | | | | | |
|---|---|---|---|---|---|---|---|---|---|---|
| O12 | 3,090 | 46,391,400 | 0.06 | 0.07 | 0.06 | 0.00 | 0.01 | 0.00 | 1.10 [0.97-1.25] | 1.08 [0.95-1.23] |
| O13 | 7,440 | 46,356,100 | 0.14 | 0.17 | 0.14 | −0.02 | 0.01 | 0.00 | 1.04 [0.96-1.13] | 1.02 [0.93-1.11] |
| O14 | 6,260 | 46,367,010 | 0.13 | 0.14 | 0.12 | −0.01 | 0.01 | 0.01 | 1.12 [1.02-1.22] | 1.12 [1.02-1.22] |
| O16 | 5,430 | 46,362,320 | 0.11 | 0.12 | 0.10 | −0.01 | 0.01 | 0.01 | 1.10 [1.00-1.21] | 1.08 [0.98-1.19] |
| O20 | 12,990 | 46,303,170 | 0.25 | 0.29 | 0.25 | −0.04 | 0.00 | 0.00 | 1.00 [0.94-1.07] | 1.00 [0.93-1.06] |
| O21 | 12,120 | 46,321,330 | 0.24 | 0.27 | 0.24 | −0.02 | 0.01 | 0.00 | 1.03 [0.97-1.10] | 1.01 [0.94-1.08] |
| O22 | 880 | 46,412,180 | 0.02 | 0.02 | 0.02 | 0.00 | 0.00 | 0.00 | 0.95 [0.74-1.22] | 0.90 [0.69-1.17] |
| O23 | 12,510 | 46,303,890 | 0.25 | 0.28 | 0.24 | −0.02 | 0.01 | 0.01 | 1.03 [0.97-1.10] | 1.02 [0.96-1.09] |
| O24 | 8,020 | 46,373,460 | 0.15 | 0.18 | 0.15 | −0.03 | 0.00 | 0.00 | 1.00 [0.92-1.09] | 1.02 [0.94-1.11] |
| O26 | 41,270 | 46,042,770 | 0.84 | 0.91 | 0.80 | −0.07 | 0.04 | 0.02 | 1.05 [1.01-1.08] | 1.02 [0.99-1.06] |
| O28 | 4,070 | 46,396,240 | 0.08 | 0.09 | 0.08 | −0.01 | 0.00 | 0.00 | 0.98 [0.87-1.10] | 0.97 [0.86-1.09] |
| O30 | 2,880 | 46,397,210 | 0.06 | 0.06 | 0.05 | −0.01 | 0.00 | 0.00 | 1.08 [0.94-1.23] | 1.08 [0.94-1.24] |
| O32 | 14,040 | 46,318,610 | 0.26 | 0.31 | 0.27 | −0.05 | −0.01 | −0.02 | 0.96 [0.90-1.02] | 0.95 [0.89-1.01] |
| O34 | 16,530 | 46,311,030 | 0.31 | 0.37 | 0.31 | −0.07 | −0.01 | −0.01 | 0.98 [0.92-1.04] | 0.97 [0.91-1.03] |
| O35 | 3,620 | 46,390,580 | 0.07 | 0.08 | 0.07 | −0.01 | 0.00 | 0.00 | 1.00 [0.89-1.13] | 0.99 [0.88-1.13] |
| O36 | 57,740 | 45,989,100 | 1.18 | 1.28 | 1.15 | −0.09 | 0.04 | 0.02 | 1.03 [1.00-1.06] | 1.02 [0.99-1.05] |
| O40 | 3,440 | 46,396,580 | 0.07 | 0.08 | 0.07 | −0.01 | 0.00 | 0.00 | 1.04 [0.92-1.18] | 1.06 [0.93-1.20] |
| O41 | 5,210 | 46,375,470 | 0.10 | 0.12 | 0.10 | −0.02 | 0.00 | 0.00 | 0.99 [0.89-1.09] | 1.00 [0.90-1.10] |
| O42 | 27,500 | 46,200,250 | 0.53 | 0.61 | 0.53 | −0.09 | 0.00 | 0.00 | 1.00 [0.96-1.05] | 1.00 [0.95-1.04] |
| O43 | 3,490 | 46,390,040 | 0.06 | 0.08 | 0.07 | −0.02 | 0.00 | 0.00 | 0.94 [0.83-1.06] | 0.96 [0.84-1.09] |
| O44 | 1,530 | 46,407,940 | 0.03 | 0.03 | 0.03 | −0.01 | 0.00 | 0.00 | 0.90 [0.74-1.09] | 0.87 [0.71-1.06] |
| O45 | 1,010 | 46,410,090 | 0.02 | 0.02 | 0.02 | 0.00 | 0.00 | 0.00 | 1.01 [0.80-1.26] | 1.03 [0.82-1.31] |
| O46 | 13,090 | 46,299,330 | 0.26 | 0.29 | 0.25 | −0.03 | 0.01 | 0.00 | 1.04 [0.97-1.10] | 1.02 [0.96-1.09] |
| O47 | 7,840 | 46,321,600 | 0.15 | 0.17 | 0.14 | −0.03 | 0.01 | 0.01 | 1.05 [0.97-1.14] | 1.04 [0.95-1.13] |
| O48 | 15,840 | 46,271,490 | 0.29 | 0.36 | 0.30 | −0.06 | −0.01 | −0.01 | 0.97 [0.92-1.03] | 0.97 [0.91-1.03] |
| O60 | 13,890 | 46,305,570 | 0.27 | 0.31 | 0.27 | −0.04 | 0.00 | 0.00 | 1.02 [0.96-1.08] | 1.02 [0.96-1.09] |
| O61 | 2,280 | 46,401,390 | 0.05 | 0.05 | 0.04 | 0.00 | 0.00 | 0.00 | 1.10 [0.94-1.27] | 1.05 [0.90-1.23] |
| O62 | 10,920 | 46,340,220 | 0.21 | 0.24 | 0.21 | −0.04 | −0.01 | −0.01 | 0.97 [0.91-1.04] | 0.97 [0.90-1.04] |
| O63 | 23,750 | 46,215,960 | 0.45 | 0.53 | 0.45 | −0.08 | 0.00 | −0.01 | 1.00 [0.95-1.05] | 0.98 [0.93-1.03] |
| O64 | 3,630 | 46,379,830 | 0.07 | 0.08 | 0.07 | −0.01 | 0.01 | 0.00 | 1.08 [0.96-1.22] | 1.07 [0.94-1.20] |
| O66 | 4,050 | 46,385,170 | 0.07 | 0.09 | 0.08 | −0.02 | −0.01 | −0.01 | 0.90 [0.80-1.01] | 0.90 [0.80-1.01] |
| O67 | 900 | 46,410,690 | 0.02 | 0.02 | 0.02 | 0.00 | 0.00 | 0.00 | 1.07 [0.85-1.36] | 1.10 [0.86-1.41] |
| O68 | 55,640 | 45,962,070 | 1.10 | 1.24 | 1.10 | −0.14 | 0.00 | 0.00 | 1.00 [0.97-1.03] | 1.00 [0.97-1.03] |
| O69 | 3,910 | 46,384,670 | 0.07 | 0.09 | 0.07 | −0.02 | 0.00 | 0.00 | 0.99 [0.88-1.11] | 0.99 [0.88-1.12] |
| O70 | 80,450 | 45,732,820 | 1.57 | 1.81 | 1.61 | −0.25 | −0.05 | −0.04 | 0.97 [0.95-1.00] | 0.98 [0.95-1.00] |
| O71 | 4,190 | 46,380,220 | 0.08 | 0.09 | 0.08 | −0.01 | 0.00 | 0.00 | 1.00 [0.90-1.12] | 1.02 [0.91-1.14] |
| O72 | 33,630 | 46,182,990 | 0.65 | 0.75 | 0.66 | −0.11 | −0.01 | −0.02 | 0.98 [0.94-1.02] | 0.97 [0.93-1.02] |
| O73 | 1,760 | 46,403,150 | 0.03 | 0.04 | 0.03 | −0.01 | 0.00 | 0.00 | 1.10 [0.93-1.31] | 1.11 [0.93-1.32] |
| O74 | 690 | 46,413,310 | 0.01 | 0.02 | 0.01 | 0.00 | 0.00 | 0.00 | 0.88 [0.66-1.16] | 0.87 [0.65-1.16] |
| O75 | 16,610 | 46,292,450 | 0.31 | 0.37 | 0.33 | −0.06 | −0.02 | −0.02 | 0.95 [0.90-1.01] | 0.95 [0.89-1.01] |
| O80 | 17,100 | 46,236,870 | 0.30 | 0.39 | 0.32 | −0.09 | −0.02 | −0.01 | 0.94 [0.89-1.00] | 0.96 [0.91-1.02] |
| O82 | 1,060 | 46,408,780 | 0.02 | 0.02 | 0.02 | 0.00 | 0.00 | 0.00 | 1.07 [0.86-1.33] | 1.07 [0.85-1.34] |
| O85 | 1,680 | 46,408,060 | 0.03 | 0.04 | 0.04 | 0.00 | 0.00 | 0.00 | 0.93 [0.78-1.11] | 0.90 [0.74-1.08] |
| O86 | 5,280 | 46,378,130 | 0.11 | 0.12 | 0.10 | −0.01 | 0.00 | 0.00 | 1.03 [0.93-1.13] | 1.02 [0.92-1.13] |
| O90 | 5,960 | 46,378,100 | 0.12 | 0.13 | 0.12 | −0.01 | 0.01 | 0.00 | 1.06 [0.97-1.17] | 1.04 [0.95-1.15] |
| O91 | 510 | 46,414,720 | 0.01 | 0.01 | 0.01 | 0.00 | 0.00 | 0.00 | 1.07 [0.78-1.46] | 1.10 [0.79-1.53] |
| O92 | 770 | 46,412,400 | 0.01 | 0.02 | 0.02 | 0.00 | 0.00 | 0.00 | 0.96 [0.74-1.24] | 0.91 [0.69-1.20] |
| O98 | 2,600 | 46,400,190 | 0.05 | 0.06 | 0.05 | 0.00 | 0.00 | 0.00 | 1.06 [0.92-1.22] | 1.04 [0.90-1.20] |
| O99 | 41,790 | 46,125,480 | 0.86 | 0.92 | 0.82 | −0.06 | 0.04 | 0.02 | 1.05 [1.01-1.08] | 1.03 [0.99-1.07] |
| <18 - P00-P96 | | | | | | | | | | |
| P83 | 1,640 | 45,386,040 | 0.04 | 0.04 | 0.04 | 0.00 | 0.00 | 0.00 | 1.03 [0.88-1.21] | 1.01 [0.86-1.19] |
| <18 - Q00-Q99 | | | | | | | | | | |
| Q02 | 1,930 | 46,369,840 | 0.04 | 0.04 | 0.04 | 0.00 | 0.00 | 0.00 | 1.08 [0.93-1.25] | 1.05 [0.90-1.22] |
| Q04 | 1,520 | 46,356,970 | 0.03 | 0.03 | 0.03 | 0.00 | 0.00 | 0.00 | 1.01 [0.85-1.20] | 0.97 [0.81-1.16] |
| Q05 | 520 | 46,401,240 | 0.01 | 0.01 | 0.01 | 0.00 | 0.00 | 0.00 | 1.08 [0.81-1.45] | 1.03 [0.76-1.39] |
| Q07 | 1,390 | 46,401,650 | 0.03 | 0.03 | 0.03 | 0.00 | 0.00 | 0.00 | 1.02 [0.85-1.23] | 0.96 [0.80-1.16] |
| Q10 | 2,240 | 46,360,730 | 0.05 | 0.05 | 0.05 | 0.01 | 0.01 | 0.01 | 1.14 [1.00-1.31] | 1.12 [0.98-1.29] |
| Q12 | 620 | 46,396,860 | 0.02 | 0.01 | 0.01 | 0.00 | 0.00 | 0.00 | 1.25 [0.97-1.63] | 1.21 [0.93-1.57] |
| Q17 | 6,100 | 46,209,660 | 0.14 | 0.13 | 0.13 | 0.01 | 0.01 | 0.01 | 1.07 [0.98-1.16] | 1.06 [0.97-1.15] |
| Q18 | 2,220 | 46,360,690 | 0.05 | 0.05 | 0.05 | 0.01 | 0.01 | 0.01 | 1.19 [1.04-1.36] | 1.17 [1.02-1.34] |
| Q21 | 4,270 | 46,098,230 | 0.09 | 0.09 | 0.09 | 0.00 | 0.00 | 0.00 | 1.01 [0.91-1.12] | 0.98 [0.88-1.09] |
| Q22 | 980 | 46,362,480 | 0.02 | 0.02 | 0.02 | 0.00 | 0.00 | 0.00 | 0.87 [0.69-1.09] | 0.87 [0.70-1.10] |
| Q23 | 1,580 | 46,371,540 | 0.04 | 0.03 | 0.03 | 0.00 | 0.00 | 0.00 | 1.07 [0.91-1.27] | 1.04 [0.87-1.23] |
| Q24 | 1,660 | 46,338,740 | 0.04 | 0.04 | 0.04 | 0.00 | 0.00 | 0.00 | 0.94 [0.79-1.11] | 0.92 [0.77-1.09] |
| Q25 | 2,510 | 46,120,720 | 0.05 | 0.06 | 0.05 | 0.00 | 0.00 | 0.00 | 0.96 [0.84-1.10] | 0.93 [0.81-1.07] |
| Q27 | 1,460 | 46,384,050 | 0.03 | 0.03 | 0.03 | 0.00 | 0.00 | 0.00 | 1.01 [0.84-1.20] | 1.00 [0.83-1.19] |
| Q28 | 460 | 46,411,140 | 0.01 | 0.01 | 0.01 | 0.00 | 0.00 | 0.00 | 0.98 [0.71-1.35] | 0.98 [0.71-1.36] |
| Q31 | 1,040 | 46,336,300 | 0.02 | 0.02 | 0.02 | 0.00 | 0.00 | 0.00 | 0.98 [0.80-1.21] | 0.87 [0.70-1.08] |
| Q35 | 1,290 | 46,354,740 | 0.03 | 0.03 | 0.03 | 0.00 | 0.00 | 0.00 | 1.10 [0.92-1.32] | 1.04 [0.87-1.26] |
| Q38 | 7,430 | 45,808,640 | 0.18 | 0.16 | 0.16 | 0.02 | 0.02 | 0.02 | 1.11 [1.03-1.20]* | 1.09 [1.01-1.18] |
| Q43 | 1,430 | 46,346,450 | 0.03 | 0.03 | 0.03 | 0.00 | 0.00 | 0.00 | 0.98 [0.81-1.17] | 0.95 [0.79-1.14] |
| Q50 | 840 | 46,408,370 | 0.02 | 0.02 | 0.02 | 0.00 | 0.01 | 0.00 | 1.41 [1.12-1.77]* | 1.28 [1.00-1.63] |
| Q51 | 600 | 46,413,090 | 0.01 | 0.01 | 0.01 | 0.00 | 0.00 | 0.00 | 1.09 [0.83-1.44] | 1.08 [0.81-1.43] |
| Q52 | 960 | 46,388,380 | 0.02 | 0.02 | 0.02 | 0.00 | 0.00 | 0.00 | 1.13 [0.91-1.40] | 1.07 [0.86-1.33] |
| Q53 | 13,320 | 45,876,510 | 0.30 | 0.29 | 0.30 | 0.01 | 0.00 | −0.01 | 0.99 [0.93-1.05] | 0.97 [0.91-1.03] |
| Q54 | 2,650 | 46,192,910 | 0.06 | 0.06 | 0.06 | 0.00 | 0.00 | 0.00 | 1.07 [0.95-1.22] | 1.07 [0.94-1.22] |
| Q55 | 8,260 | 46,244,830 | 0.20 | 0.17 | 0.18 | 0.03 | 0.02 | 0.02 | 1.11 [1.03-1.19]* | 1.08 [1.01-1.16] |
| Q60 | 800 | 46,392,770 | 0.02 | 0.02 | 0.02 | 0.00 | 0.00 | 0.00 | 0.92 [0.72-1.18] | 0.92 [0.72-1.17] |
| Q61 | 1,080 | 46,373,710 | 0.02 | 0.02 | 0.02 | 0.00 | 0.00 | 0.00 | 1.05 [0.86-1.29] | 1.00 [0.82-1.23] |
| Q62 | 1,130 | 46,229,470 | 0.02 | 0.02 | 0.02 | 0.00 | 0.00 | 0.00 | 0.99 [0.81-1.21] | 0.97 [0.79-1.19] |
| Q63 | 2,050 | 46,337,060 | 0.05 | 0.04 | 0.04 | 0.01 | 0.01 | 0.01 | 1.16 [1.00-1.34] | 1.12 [0.96-1.30] |
| Q64 | 810 | 46,373,830 | 0.02 | 0.02 | 0.02 | 0.00 | 0.00 | 0.00 | 1.06 [0.84-1.33] | 1.02 [0.80-1.29] |
| Q65 | 2,750 | 46,268,650 | 0.05 | 0.06 | 0.06 | −0.01 | −0.01 | −0.01 | 0.89 [0.78-1.02] | 0.86 [0.75-0.99] |
| Q66 | 3,680 | 45,607,920 | 0.09 | 0.08 | 0.08 | 0.01 | 0.01 | 0.01 | 1.12 [1.01-1.25] | 1.08 [0.97-1.21] |
| Q67 | 2,600 | 46,332,630 | 0.06 | 0.05 | 0.06 | 0.01 | 0.00 | 0.00 | 1.08 [0.95-1.23] | 1.01 [0.89-1.16] |
| Q68 | 900 | 46,381,530 | 0.02 | 0.02 | 0.02 | 0.00 | 0.00 | 0.00 | 1.11 [0.90-1.39] | 1.06 [0.85-1.33] |
| Q69 | 600 | 46,312,180 | 0.01 | 0.01 | 0.01 | 0.00 | 0.00 | 0.00 | 0.91 [0.69-1.20] | 0.89 [0.67-1.18] |
| Q74 | 2,690 | 46,351,650 | 0.07 | 0.06 | 0.06 | 0.01 | 0.01 | 0.01 | 1.17 [1.03-1.33] | 1.15 [1.01-1.30] |
| Q75 | 1,850 | 46,333,260 | 0.04 | 0.04 | 0.04 | 0.00 | 0.00 | 0.00 | 1.00 [0.86-1.17] | 0.97 [0.83-1.13] |
| Q76 | 1,150 | 46,386,030 | 0.02 | 0.02 | 0.03 | 0.00 | 0.00 | 0.00 | 0.91 [0.75-1.12] | 0.87 [0.71-1.07] |
| Q78 | 910 | 46,399,590 | 0.02 | 0.02 | 0.02 | 0.00 | 0.00 | 0.00 | 1.06 [0.85-1.32] | 1.03 [0.82-1.29] |
| Q79 | 1,780 | 46,360,580 | 0.05 | 0.04 | 0.04 | 0.01 | 0.01 | 0.00 | 1.24 [1.06-1.45]* | 1.12 [0.95-1.32] |
| Q82 | 1,940 | 45,727,890 | 0.05 | 0.04 | 0.04 | 0.01 | 0.01 | 0.01 | 1.19 [1.03-1.38] | 1.16 [1.00-1.34] |
| Q83 | 800 | 46,401,820 | 0.02 | 0.02 | 0.02 | 0.00 | 0.00 | 0.00 | 0.97 [0.76-1.25] | 0.93 [0.73-1.20] |
| Q85 | 1,180 | 46,395,920 | 0.02 | 0.03 | 0.03 | 0.00 | 0.00 | 0.00 | 0.96 [0.79-1.17] | 0.94 [0.76-1.15] |
| Q87 | 2,310 | 46,357,240 | 0.05 | 0.05 | 0.05 | 0.00 | 0.00 | 0.00 | 1.05 [0.91-1.21] | 1.02 [0.88-1.17] |
| Q89 | 2,050 | 46,368,770 | 0.05 | 0.04 | 0.04 | 0.00 | 0.00 | 0.00 | 1.02 [0.88-1.18] | 1.01 [0.87-1.17] |
| Q90 | 810 | 46,348,380 | 0.02 | 0.02 | 0.02 | 0.00 | 0.00 | 0.00 | 1.03 [0.81-1.31] | 1.00 [0.79-1.29] |
| Q92 | 510 | 46,410,650 | 0.01 | 0.01 | 0.01 | 0.00 | 0.00 | 0.00 | 0.98 [0.73-1.31] | 0.95 [0.71-1.29] |
| Q93 | 1,560 | 46,394,160 | 0.04 | 0.03 | 0.03 | 0.01 | 0.01 | 0.00 | 1.19 [1.01-1.41] | 1.12 [0.95-1.32] |
| Q99 | 1,650 | 46,392,600 | 0.04 | 0.03 | 0.04 | 0.01 | 0.00 | 0.00 | 1.12 [0.95-1.31] | 1.07 [0.91-1.26] |
| <18 - R00-R99 | | | | | | | | | | |
| R00 | 30,990 | 46,118,240 | 0.73 | 0.66 | 0.64 | 0.08 | 0.09 | 0.07 | 1.14 [1.10-1.18]* | 1.10 [1.06-1.15]* |
| R01 | 9,310 | 46,076,890 | 0.24 | 0.19 | 0.19 | 0.04 | 0.04 | 0.03 | 1.21 [1.13-1.29]* | 1.16 [1.09-1.24]* |
| R02 | 1,690 | 46,401,560 | 0.04 | 0.04 | 0.04 | 0.00 | 0.00 | 0.00 | 1.08 [0.92-1.28] | 1.07 [0.91-1.27] |

| | | | | | | | | | | |
|---|---|---|---|---|---|---|---|---|---|---|
| R03 | 7,750 | 46,354,640 | 0.17 | 0.17 | 0.15 | 0.01 | 0.02 | 0.02 | 1.13 [1.04-1.22]* | 1.10 [1.02-1.20] |
| R04 | 12,720 | 46,211,430 | 0.32 | 0.26 | 0.26 | 0.06 | 0.06 | 0.05 | 1.24 [1.17-1.31]* | 1.19 [1.12-1.26]* |
| R05 | 25,410 | 45,768,050 | 0.70 | 0.51 | 0.51 | 0.19 | 0.19 | 0.16 | 1.37 [1.32-1.43]* | 1.30 [1.25-1.35]* |
| R06 | 111,440 | 43,508,850 | 3.86 | 2.21 | 2.20 | 1.65 | 1.66 | 1.58 | 1.76 [1.72-1.79]* | 1.70 [1.67-1.73]* |
| R07 | 33,900 | 46,143,470 | 0.78 | 0.72 | 0.68 | 0.06 | 0.10 | 0.07 | 1.14 [1.10-1.18]* | 1.10 [1.06-1.15]* |
| R09 | 3,660 | 46,360,440 | 0.09 | 0.08 | 0.07 | 0.01 | 0.02 | 0.01 | 1.21 [1.08-1.34]* | 1.15 [1.03-1.29] |
| R10 | 155,120 | 44,478,720 | 3.89 | 3.38 | 3.28 | 0.52 | 0.61 | 0.48 | 1.19 [1.17-1.21]* | 1.14 [1.12-1.16]* |
| R11 | 90,830 | 44,502,810 | 2.29 | 1.97 | 1.96 | 0.32 | 0.33 | 0.26 | 1.17 [1.14-1.19]* | 1.13 [1.10-1.15]* |
| R12 | 1,560 | 46,406,290 | 0.03 | 0.03 | 0.03 | 0.00 | 0.00 | 0.00 | 1.08 [0.90-1.28] | 1.00 [0.83-1.20] |
| R13 | 5,820 | 46,360,480 | 0.14 | 0.12 | 0.12 | 0.02 | 0.02 | 0.02 | 1.21 [1.11-1.32]* | 1.12 [1.02-1.23] |
| R14 | 3,820 | 46,321,680 | 0.09 | 0.08 | 0.08 | 0.01 | 0.01 | 0.01 | 1.16 [1.04-1.30]* | 1.11 [0.99-1.24] |
| R15 | 2,370 | 46,392,990 | 0.06 | 0.05 | 0.05 | 0.01 | 0.01 | 0.00 | 1.14 [0.99-1.31] | 1.09 [0.95-1.25] |
| R16 | 3,680 | 46,364,740 | 0.08 | 0.08 | 0.08 | 0.01 | 0.01 | 0.01 | 1.11 [0.99-1.24] | 1.08 [0.96-1.21] |
| R17 | 1,550 | 46,354,710 | 0.04 | 0.03 | 0.03 | 0.00 | 0.00 | 0.00 | 1.12 [0.94-1.33] | 1.13 [0.95-1.34] |
| R18 | 2,190 | 46,395,170 | 0.04 | 0.05 | 0.05 | 0.00 | 0.00 | 0.00 | 0.94 [0.81-1.10] | 0.93 [0.80-1.09] |
| R19 | 15,580 | 46,235,200 | 0.37 | 0.33 | 0.31 | 0.04 | 0.06 | 0.05 | 1.19 [1.13-1.26]* | 1.14 [1.08-1.20]* |
| R20 | 7,360 | 46,360,970 | 0.17 | 0.16 | 0.15 | 0.01 | 0.02 | 0.01 | 1.15 [1.06-1.24]* | 1.09 [1.00-1.18] |
| R21 | 36,580 | 45,262,870 | 0.91 | 0.78 | 0.78 | 0.13 | 0.14 | 0.11 | 1.17 [1.14-1.21]* | 1.13 [1.10-1.17]* |
| R22 | 9,420 | 46,226,520 | 0.26 | 0.19 | 0.18 | 0.08 | 0.08 | 0.07 | 1.42 [1.33-1.52]* | 1.36 [1.27-1.45]* |
| R23 | 14,500 | 45,984,010 | 0.36 | 0.30 | 0.31 | 0.05 | 0.05 | 0.04 | 1.16 [1.10-1.22]* | 1.12 [1.06-1.18]* |
| R25 | 5,600 | 46,109,690 | 0.13 | 0.12 | 0.12 | 0.01 | 0.01 | 0.01 | 1.10 [1.01-1.20] | 1.06 [0.97-1.16] |
| R26 | 10,160 | 46,302,530 | 0.24 | 0.21 | 0.22 | 0.03 | 0.02 | 0.02 | 1.10 [1.03-1.17]* | 1.07 [1.00-1.15] |
| R27 | 2,550 | 46,384,800 | 0.06 | 0.05 | 0.05 | 0.01 | 0.01 | 0.01 | 1.17 [1.03-1.33] | 1.12 [0.98-1.28] |
| R29 | 6,690 | 45,882,410 | 0.15 | 0.14 | 0.14 | 0.01 | 0.01 | 0.00 | 1.07 [0.99-1.17] | 1.02 [0.94-1.11] |
| R30 | 5,820 | 46,354,070 | 0.14 | 0.12 | 0.12 | 0.02 | 0.02 | 0.02 | 1.18 [1.08-1.29]* | 1.14 [1.05-1.25]* |
| R31 | 9,870 | 46,294,670 | 0.23 | 0.21 | 0.20 | 0.02 | 0.03 | 0.02 | 1.14 [1.06-1.22]* | 1.09 [1.02-1.17] |
| R32 | 6,830 | 46,350,530 | 0.17 | 0.14 | 0.15 | 0.03 | 0.02 | 0.02 | 1.15 [1.07-1.25]* | 1.10 [1.02-1.20] |
| R33 | 7,920 | 46,333,920 | 0.18 | 0.17 | 0.16 | 0.02 | 0.03 | 0.02 | 1.16 [1.08-1.25]* | 1.10 [1.02-1.19] |
| R34 | 680 | 46,407,710 | 0.01 | 0.01 | 0.02 | 0.00 | 0.00 | 0.00 | 0.95 [0.73-1.23] | 0.92 [0.71-1.21] |
| R35 | 4,290 | 46,366,990 | 0.11 | 0.09 | 0.09 | 0.02 | 0.02 | 0.02 | 1.22 [1.10-1.35]* | 1.19 [1.07-1.32]* |
| R39 | 4,040 | 46,357,980 | 0.10 | 0.08 | 0.08 | 0.02 | 0.02 | 0.02 | 1.31 [1.19-1.45]* | 1.25 [1.13-1.39]* |
| R40 | 4,860 | 46,346,960 | 0.11 | 0.10 | 0.11 | 0.01 | 0.01 | 0.00 | 1.05 [0.95-1.16] | 1.02 [0.92-1.13] |
| R41 | 7,630 | 46,334,260 | 0.18 | 0.16 | 0.16 | 0.02 | 0.01 | 0.01 | 1.09 [1.01-1.18] | 1.04 [0.96-1.13] |
| R42 | 12,210 | 46,318,570 | 0.29 | 0.26 | 0.26 | 0.03 | 0.03 | 0.02 | 1.11 [1.05-1.18]* | 1.07 [1.01-1.14] |
| R44 | 4,580 | 46,383,030 | 0.11 | 0.10 | 0.10 | 0.02 | 0.01 | 0.01 | 1.14 [1.03-1.26]* | 1.10 [1.00-1.22] |
| R45 | 20,300 | 46,278,510 | 0.49 | 0.42 | 0.46 | 0.07 | 0.03 | 0.02 | 1.06 [1.02-1.12]* | 1.04 [0.99-1.10] |
| R46 | 3,120 | 46,386,690 | 0.07 | 0.07 | 0.07 | 0.01 | 0.00 | 0.00 | 1.06 [0.94-1.19] | 1.00 [0.89-1.14] |
| R47 | 3,190 | 46,387,090 | 0.07 | 0.07 | 0.07 | 0.00 | 0.01 | 0.00 | 1.09 [0.97-1.22] | 1.04 [0.92-1.18] |
| R48 | 1,200 | 46,409,960 | 0.03 | 0.02 | 0.03 | 0.01 | 0.00 | 0.00 | 1.16 [0.96-1.40] | 1.09 [0.90-1.32] |
| R49 | 1,440 | 46,394,180 | 0.04 | 0.03 | 0.03 | 0.01 | 0.01 | 0.01 | 1.25 [1.05-1.48] | 1.20 [1.01-1.42] |
| R50 | 57,030 | 45,017,920 | 1.42 | 1.22 | 1.24 | 0.20 | 0.19 | 0.15 | 1.15 [1.12-1.18]* | 1.12 [1.09-1.15]* |
| R51 | 47,230 | 45,915,700 | 1.17 | 0.99 | 0.97 | 0.18 | 0.20 | 0.15 | 1.20 [1.17-1.24]* | 1.15 [1.12-1.19]* |
| R52 | 2,700 | 46,394,350 | 0.06 | 0.06 | 0.06 | 0.00 | 0.01 | 0.00 | 1.12 [0.98-1.28] | 1.06 [0.93-1.22] |
| R53 | 13,740 | 46,207,110 | 0.35 | 0.28 | 0.28 | 0.06 | 0.06 | 0.05 | 1.22 [1.16-1.29]* | 1.18 [1.11-1.25]* |
| R55 | 23,120 | 46,162,510 | 0.54 | 0.49 | 0.49 | 0.04 | 0.05 | 0.03 | 1.10 [1.05-1.14]* | 1.05 [1.01-1.10] |
| R56 | 47,630 | 44,996,960 | 1.05 | 1.06 | 1.03 | −0.01 | 0.02 | 0.01 | 1.02 [0.99-1.05] | 1.01 [0.98-1.04] |
| R57 | 820 | 46,406,860 | 0.02 | 0.02 | 0.02 | 0.00 | 0.00 | 0.00 | 1.01 [0.80-1.29] | 1.03 [0.80-1.32] |
| R59 | 13,730 | 46,204,070 | 0.36 | 0.28 | 0.28 | 0.08 | 0.08 | 0.07 | 1.29 [1.22-1.36]* | 1.25 [1.18-1.32]* |
| R60 | 2,030 | 46,391,110 | 0.05 | 0.04 | 0.04 | 0.01 | 0.01 | 0.01 | 1.21 [1.05-1.40] | 1.16 [1.00-1.35] |
| R61 | 1,920 | 46,396,990 | 0.05 | 0.04 | 0.04 | 0.01 | 0.01 | 0.01 | 1.24 [1.07-1.45]* | 1.18 [1.01-1.37] |
| R62 | 15,210 | 45,834,030 | 0.34 | 0.33 | 0.33 | 0.02 | 0.02 | 0.01 | 1.06 [1.00-1.12] | 1.02 [0.96-1.07] |
| R63 | 26,750 | 45,601,210 | 0.66 | 0.57 | 0.57 | 0.09 | 0.08 | 0.06 | 1.14 [1.10-1.19]* | 1.11 [1.06-1.15]* |
| R65 | 530 | 46,415,020 | 0.01 | 0.01 | 0.01 | 0.00 | 0.00 | 0.00 | 1.21 [0.90-1.63] | 1.20 [0.88-1.63] |
| R68 | 2,700 | 46,173,350 | 0.06 | 0.06 | 0.06 | 0.00 | 0.00 | 0.00 | 1.03 [0.91-1.17] | 1.01 [0.89-1.15] |
| R69 | 20,610 | 42,786,150 | 0.52 | 0.47 | 0.43 | 0.05 | 0.09 | 0.07 | 1.20 [1.15-1.26]* | 1.17 [1.11-1.22]* |
| R72 | 870 | 46,407,240 | 0.02 | 0.02 | 0.02 | 0.00 | 0.00 | 0.00 | 1.22 [0.97-1.52] | 1.16 [0.92-1.46] |
| R73 | 1,950 | 46,379,520 | 0.05 | 0.04 | 0.04 | 0.01 | 0.01 | 0.01 | 1.20 [1.04-1.40] | 1.13 [0.97-1.32] |
| R74 | 1,580 | 46,399,680 | 0.04 | 0.03 | 0.03 | 0.01 | 0.01 | 0.00 | 1.24 [1.05-1.46] | 1.14 [0.96-1.36] |
| R76 | 740 | 46,410,740 | 0.02 | 0.01 | 0.01 | 0.00 | 0.00 | 0.00 | 1.29 [1.02-1.64] | 1.24 [0.97-1.59] |
| R77 | 690 | 46,408,180 | 0.02 | 0.01 | 0.02 | 0.00 | 0.00 | 0.00 | 1.12 [0.87-1.45] | 1.12 [0.86-1.46] |
| R79 | 6,350 | 46,329,520 | 0.15 | 0.13 | 0.13 | 0.02 | 0.02 | 0.02 | 1.18 [1.09-1.28]* | 1.14 [1.05-1.25]* |
| R80 | 1,460 | 46,398,560 | 0.04 | 0.03 | 0.03 | 0.01 | 0.01 | 0.01 | 1.18 [0.99-1.40] | 1.17 [0.98-1.40] |
| R81 | 470 | 46,409,460 | 0.01 | 0.01 | 0.01 | 0.00 | 0.00 | 0.00 | 1.24 [0.92-1.67] | 1.24 [0.91-1.69] |
| R82 | 2,430 | 46,382,320 | 0.06 | 0.05 | 0.05 | 0.01 | 0.01 | 0.01 | 1.18 [1.03-1.35] | 1.15 [1.00-1.32] |
| R84 | 970 | 46,408,040 | 0.02 | 0.02 | 0.02 | 0.00 | 0.00 | 0.00 | 1.05 [0.84-1.30] | 0.99 [0.80-1.24] |
| R87 | 1,030 | 46,409,240 | 0.02 | 0.02 | 0.02 | 0.00 | 0.00 | 0.00 | 1.09 [0.87-1.37] | 1.08 [0.86-1.37] |
| R89 | 350 | 46,413,860 | 0.01 | 0.01 | 0.01 | 0.00 | 0.00 | 0.00 | 1.53 [1.10-2.14] | 1.40 [0.99-1.97] |
| R90 | 990 | 46,402,320 | 0.02 | 0.02 | 0.02 | 0.00 | 0.00 | 0.00 | 1.17 [0.95-1.45] | 1.14 [0.92-1.41] |
| R91 | 1,520 | 46,387,970 | 0.04 | 0.03 | 0.03 | 0.01 | 0.01 | 0.01 | 1.29 [1.09-1.53]* | 1.26 [1.06-1.50]* |
| R93 | 4,080 | 46,335,540 | 0.09 | 0.09 | 0.09 | 0.01 | 0.01 | 0.00 | 1.07 [0.96-1.19] | 1.04 [0.93-1.16] |
| R94 | 9,160 | 46,320,910 | 0.22 | 0.19 | 0.19 | 0.02 | 0.03 | 0.02 | 1.16 [1.08-1.24]* | 1.12 [1.04-1.21]* |
| <18 - S00-T88 | | | | | | | | | | |
| S00 | 31,040 | 45,843,480 | 0.66 | 0.68 | 0.68 | −0.02 | −0.02 | −0.03 | 0.97 [0.93-1.01] | 0.95 [0.92-0.99] |
| S01 | 40,950 | 45,768,950 | 0.86 | 0.90 | 0.89 | −0.04 | −0.02 | −0.03 | 0.97 [0.94-1.01] | 0.96 [0.93-1.00] |
| S02 | 23,260 | 46,081,850 | 0.49 | 0.51 | 0.48 | −0.02 | 0.01 | 0.00 | 1.02 [0.98-1.07] | 1.01 [0.96-1.06] |
| S03 | 1,500 | 46,399,700 | 0.03 | 0.03 | 0.03 | 0.00 | 0.00 | 0.00 | 1.04 [0.88-1.23] | 1.02 [0.85-1.21] |
| S05 | 2,620 | 46,376,700 | 0.05 | 0.06 | 0.06 | 0.00 | 0.00 | 0.00 | 0.95 [0.83-1.09] | 0.93 [0.81-1.07] |
| S06 | 6,820 | 46,304,130 | 0.15 | 0.15 | 0.14 | 0.00 | 0.01 | 0.01 | 1.05 [0.97-1.14] | 1.04 [0.95-1.13] |
| S09 | 28,170 | 45,654,460 | 0.63 | 0.61 | 0.59 | 0.01 | 0.03 | 0.02 | 1.05 [1.01-1.10]* | 1.04 [0.99-1.08] |
| S10 | 1,500 | 46,400,010 | 0.03 | 0.03 | 0.03 | 0.00 | 0.00 | 0.00 | 0.96 [0.81-1.15] | 0.96 [0.80-1.15] |
| S11 | 810 | 46,409,930 | 0.02 | 0.02 | 0.02 | 0.00 | 0.00 | 0.00 | 1.05 [0.82-1.33] | 1.01 [0.79-1.30] |
| S12 | 740 | 46,411,490 | 0.01 | 0.02 | 0.01 | 0.00 | 0.00 | 0.00 | 1.00 [0.77-1.29] | 1.01 [0.77-1.32] |
| S13 | 730 | 46,408,810 | 0.02 | 0.02 | 0.02 | 0.00 | 0.00 | 0.00 | 1.01 [0.79-1.30] | 0.98 [0.76-1.27] |
| S19 | 990 | 46,404,100 | 0.02 | 0.02 | 0.02 | 0.00 | 0.00 | 0.00 | 1.10 [0.89-1.36] | 1.06 [0.86-1.32] |
| S20 | 2,120 | 46,391,390 | 0.04 | 0.05 | 0.05 | −0.01 | 0.00 | −0.01 | 0.90 [0.77-1.05] | 0.89 [0.76-1.04] |
| S21 | 1,300 | 46,405,640 | 0.02 | 0.03 | 0.03 | 0.00 | 0.00 | −0.01 | 0.85 [0.70-1.04] | 0.83 [0.67-1.02] |
| S22 | 2,630 | 46,390,750 | 0.05 | 0.06 | 0.05 | 0.00 | 0.00 | 0.00 | 1.05 [0.91-1.20] | 1.05 [0.91-1.21] |
| S27 | 2,720 | 46,396,010 | 0.06 | 0.06 | 0.06 | 0.00 | 0.00 | 0.00 | 1.02 [0.89-1.16] | 1.02 [0.89-1.18] |
| S30 | 6,800 | 46,322,450 | 0.14 | 0.15 | 0.15 | 0.00 | 0.00 | −0.01 | 0.97 [0.89-1.05] | 0.95 [0.87-1.03] |
| S31 | 4,460 | 46,357,270 | 0.10 | 0.10 | 0.09 | 0.00 | 0.00 | 0.00 | 1.03 [0.93-1.15] | 1.03 [0.93-1.14] |
| S32 | 3,030 | 46,388,820 | 0.06 | 0.07 | 0.06 | −0.01 | 0.00 | 0.00 | 0.95 [0.84-1.08] | 0.95 [0.83-1.08] |
| S36 | 2,200 | 46,396,890 | 0.05 | 0.05 | 0.05 | 0.00 | 0.00 | 0.00 | 0.99 [0.85-1.15] | 1.00 [0.86-1.16] |
| S37 | 1,090 | 46,406,280 | 0.02 | 0.02 | 0.02 | 0.00 | 0.00 | 0.00 | 0.99 [0.80-1.22] | 1.00 [0.80-1.24] |
| S39 | 2,950 | 46,374,070 | 0.07 | 0.06 | 0.06 | 0.01 | 0.01 | 0.01 | 1.18 [1.04-1.33]* | 1.13 [0.99-1.28] |
| S40 | 3,090 | 46,380,130 | 0.06 | 0.07 | 0.07 | 0.00 | −0.01 | −0.01 | 0.92 [0.81-1.04] | 0.90 [0.79-1.02] |
| S41 | 2,180 | 46,394,730 | 0.05 | 0.05 | 0.05 | 0.00 | 0.00 | 0.00 | 0.98 [0.85-1.14] | 0.98 [0.84-1.14] |
| S42 | 20,750 | 46,090,680 | 0.47 | 0.44 | 0.45 | 0.03 | 0.02 | 0.02 | 1.05 [1.00-1.10] | 1.04 [0.99-1.09] |
| S43 | 1,080 | 46,408,230 | 0.02 | 0.02 | 0.02 | 0.00 | 0.00 | 0.00 | 1.02 [0.82-1.26] | 1.01 [0.81-1.26] |
| S49 | 610 | 46,410,090 | 0.01 | 0.01 | 0.01 | 0.00 | 0.00 | 0.00 | 1.17 [0.89-1.53] | 1.14 [0.86-1.51] |
| S50 | 4,550 | 46,370,800 | 0.10 | 0.10 | 0.10 | 0.00 | −0.01 | −0.01 | 0.93 [0.84-1.03] | 0.90 [0.81-1.00] |
| S51 | 6,390 | 46,357,540 | 0.13 | 0.14 | 0.14 | 0.00 | 0.00 | 0.00 | 0.98 [0.90-1.07] | 0.97 [0.89-1.06] |
| S52 | 54,280 | 45,503,840 | 1.22 | 1.19 | 1.21 | 0.04 | 0.01 | 0.00 | 1.01 [0.98-1.04] | 1.00 [0.97-1.03] |
| S53 | 1,750 | 46,389,450 | 0.04 | 0.04 | 0.04 | 0.00 | 0.00 | 0.00 | 0.96 [0.82-1.13] | 0.96 [0.81-1.13] |
| S54 | 610 | 46,411,580 | 0.01 | 0.01 | 0.01 | 0.00 | 0.00 | 0.00 | 1.00 [0.75-1.33] | 1.00 [0.75-1.34] |
| S56 | 2,010 | 46,399,110 | 0.04 | 0.04 | 0.04 | 0.00 | 0.00 | 0.00 | 0.97 [0.83-1.14] | 0.96 [0.82-1.14] |
| S59 | 580 | 46,408,360 | 0.01 | 0.01 | 0.01 | 0.00 | 0.00 | 0.00 | 1.02 [0.77-1.35] | 0.95 [0.71-1.27] |
| S60 | 4,580 | 46,366,140 | 0.10 | 0.10 | 0.10 | 0.00 | 0.00 | 0.00 | 1.00 [0.91-1.11] | 0.99 [0.89-1.10] |

| | | | | | | | | | | |
|---|---|---|---|---|---|---|---|---|---|---|
| S61 | 30,870 | 46,031,600 | 0.64 | 0.68 | 0.66 | −0.04 | −0.03 | −0.03 | 0.96 [0.92-1.00] | 0.96 [0.92-1.00] |
| S62 | 23,030 | 46,159,790 | 0.49 | 0.50 | 0.49 | −0.01 | 0.00 | 0.00 | 1.01 [0.97-1.06] | 1.00 [0.95-1.04] |
| S63 | 2,310 | 46,392,870 | 0.05 | 0.05 | 0.05 | 0.00 | 0.01 | 0.00 | 1.14 [0.99-1.31] | 1.10 [0.95-1.27] |
| S64 | 4,030 | 46,373,980 | 0.08 | 0.09 | 0.08 | −0.01 | 0.00 | 0.00 | 0.94 [0.84-1.05] | 0.94 [0.84-1.06] |
| S65 | 1,060 | 46,406,780 | 0.02 | 0.02 | 0.02 | 0.00 | 0.00 | 0.00 | 1.03 [0.83-1.27] | 1.04 [0.83-1.30] |
| S66 | 7,300 | 46,339,330 | 0.15 | 0.16 | 0.15 | −0.01 | 0.00 | 0.00 | 0.99 [0.91-1.07] | 0.98 [0.90-1.07] |
| S67 | 1,790 | 46,383,930 | 0.04 | 0.04 | 0.04 | 0.00 | 0.00 | 0.00 | 0.92 [0.79-1.09] | 0.92 [0.78-1.09] |
| S68 | 3,160 | 46,362,860 | 0.07 | 0.07 | 0.07 | 0.00 | 0.00 | 0.00 | 0.99 [0.88-1.12] | 0.98 [0.87-1.11] |
| S69 | 2,140 | 46,389,480 | 0.05 | 0.05 | 0.05 | 0.00 | 0.00 | 0.00 | 1.02 [0.88-1.18] | 1.01 [0.87-1.17] |
| S70 | 2,750 | 46,384,640 | 0.06 | 0.06 | 0.06 | 0.00 | 0.00 | 0.00 | 0.94 [0.82-1.07] | 0.94 [0.82-1.07] |
| S71 | 2,670 | 46,390,560 | 0.06 | 0.06 | 0.06 | 0.00 | 0.00 | 0.00 | 0.99 [0.86-1.13] | 0.95 [0.82-1.09] |
| S72 | 5,860 | 46,300,920 | 0.12 | 0.13 | 0.12 | −0.01 | 0.00 | 0.00 | 0.98 [0.90-1.07] | 0.97 [0.89-1.06] |
| S76 | 710 | 46,412,370 | 0.02 | 0.01 | 0.02 | 0.00 | 0.00 | 0.00 | 1.09 [0.84-1.40] | 1.05 [0.80-1.36] |
| S79 | 760 | 46,408,100 | 0.02 | 0.02 | 0.01 | 0.00 | 0.00 | 0.00 | 1.17 [0.92-1.49] | 1.21 [0.94-1.55] |
| S80 | 5,830 | 46,344,620 | 0.12 | 0.13 | 0.13 | −0.01 | −0.01 | −0.01 | 0.93 [0.85-1.02] | 0.91 [0.83-0.99] |
| S81 | 5,340 | 46,352,400 | 0.12 | 0.11 | 0.12 | 0.01 | 0.01 | 0.00 | 1.06 [0.97-1.17] | 1.04 [0.95-1.14] |
| S82 | 23,290 | 46,089,130 | 0.51 | 0.51 | 0.49 | 0.00 | 0.02 | 0.01 | 1.04 [0.99-1.08] | 1.02 [0.97-1.07] |
| S83 | 4,140 | 46,379,270 | 0.09 | 0.09 | 0.08 | 0.00 | 0.01 | 0.01 | 1.11 [1.00-1.24] | 1.07 [0.96-1.19] |
| S86 | 870 | 46,409,480 | 0.02 | 0.02 | 0.02 | 0.00 | 0.00 | 0.00 | 1.04 [0.82-1.31] | 1.01 [0.79-1.28] |
| S89 | 1,300 | 46,401,060 | 0.03 | 0.03 | 0.03 | 0.00 | 0.00 | 0.00 | 1.05 [0.87-1.26] | 1.03 [0.85-1.25] |
| S90 | 2,310 | 46,385,620 | 0.05 | 0.05 | 0.05 | 0.00 | 0.00 | 0.00 | 1.02 [0.88-1.17] | 1.01 [0.88-1.17] |
| S91 | 4,910 | 46,345,950 | 0.11 | 0.11 | 0.11 | 0.00 | 0.00 | 0.00 | 0.99 [0.90-1.09] | 0.99 [0.89-1.09] |
| S92 | 4,000 | 46,373,850 | 0.08 | 0.09 | 0.08 | −0.01 | 0.00 | 0.00 | 0.99 [0.88-1.10] | 0.97 [0.86-1.08] |
| S93 | 1,250 | 46,405,770 | 0.03 | 0.03 | 0.03 | 0.00 | 0.01 | 0.00 | 1.20 [1.00-1.45] | 1.16 [0.96-1.41] |
| S96 | 460 | 46,412,210 | 0.01 | 0.01 | 0.01 | 0.00 | 0.00 | 0.00 | 1.22 [0.90-1.65] | 1.21 [0.88-1.65] |
| S99 | 970 | 46,405,130 | 0.02 | 0.02 | 0.02 | 0.00 | 0.00 | 0.00 | 1.17 [0.94-1.45] | 1.14 [0.91-1.42] |
| T00 | 2,150 | 46,391,690 | 0.05 | 0.05 | 0.05 | 0.00 | 0.00 | 0.00 | 0.98 [0.84-1.13] | 0.98 [0.84-1.14] |
| T01 | 1,180 | 46,409,690 | 0.03 | 0.02 | 0.03 | 0.00 | 0.00 | 0.00 | 1.07 [0.87-1.30] | 1.05 [0.86-1.29] |
| T02 | 780 | 46,409,320 | 0.02 | 0.02 | 0.02 | 0.00 | 0.00 | 0.00 | 0.97 [0.76-1.24] | 0.94 [0.73-1.20] |
| T11 | 1,230 | 46,403,130 | 0.03 | 0.03 | 0.03 | 0.00 | 0.00 | 0.00 | 0.95 [0.78-1.16] | 0.93 [0.76-1.15] |
| T13 | 660 | 46,407,050 | 0.01 | 0.01 | 0.01 | 0.00 | 0.00 | 0.00 | 0.83 [0.63-1.10] | 0.83 [0.62-1.10] |
| T14 | 920 | 46,390,750 | 0.02 | 0.02 | 0.02 | 0.00 | 0.00 | 0.00 | 0.89 [0.71-1.13] | 0.86 [0.68-1.09] |
| T15 | 510 | 46,408,110 | 0.01 | 0.01 | 0.01 | 0.00 | 0.00 | 0.00 | 1.00 [0.74-1.35] | 0.98 [0.72-1.32] |
| T16 | 8,310 | 46,276,250 | 0.19 | 0.18 | 0.18 | 0.01 | 0.01 | 0.01 | 1.05 [0.97-1.12] | 1.03 [0.96-1.11] |
| T17 | 7,190 | 46,262,300 | 0.15 | 0.16 | 0.16 | 0.00 | 0.00 | −0.01 | 0.99 [0.91-1.07] | 0.97 [0.89-1.05] |
| T18 | 4,880 | 46,333,000 | 0.11 | 0.10 | 0.11 | 0.00 | 0.00 | 0.00 | 1.01 [0.91-1.11] | 0.98 [0.89-1.08] |
| T20 | 2,960 | 46,348,910 | 0.06 | 0.07 | 0.06 | −0.01 | −0.01 | −0.01 | 0.90 [0.80-1.02] | 0.90 [0.79-1.02] |
| T21 | 3,960 | 46,322,580 | 0.08 | 0.09 | 0.09 | −0.01 | −0.01 | −0.01 | 0.91 [0.82-1.01] | 0.91 [0.81-1.01] |
| T22 | 3,380 | 46,342,080 | 0.07 | 0.07 | 0.07 | −0.01 | 0.00 | 0.00 | 0.94 [0.84-1.06] | 0.94 [0.84-1.06] |
| T23 | 3,600 | 46,342,650 | 0.08 | 0.08 | 0.08 | 0.00 | 0.00 | 0.00 | 0.97 [0.87-1.09] | 0.95 [0.85-1.07] |
| T24 | 2,310 | 46,367,560 | 0.05 | 0.05 | 0.05 | 0.00 | −0.01 | 0.00 | 0.90 [0.78-1.04] | 0.91 [0.79-1.05] |
| T25 | 1,480 | 46,385,470 | 0.03 | 0.03 | 0.03 | 0.00 | 0.00 | 0.00 | 0.92 [0.77-1.10] | 0.90 [0.75-1.08] |
| T29 | 590 | 46,404,510 | 0.01 | 0.01 | 0.01 | 0.00 | 0.00 | 0.00 | 0.81 [0.61-1.08] | 0.80 [0.60-1.07] |
| T31 | 9,970 | 46,212,710 | 0.20 | 0.22 | 0.22 | −0.02 | −0.02 | −0.02 | 0.92 [0.86-0.99] | 0.92 [0.86-0.98] |
| T36 | 1,850 | 46,392,570 | 0.04 | 0.04 | 0.04 | 0.00 | 0.01 | 0.01 | 1.18 [1.01-1.38] | 1.17 [1.00-1.37] |
| T37 | 580 | 46,409,190 | 0.01 | 0.01 | 0.01 | 0.00 | 0.00 | 0.00 | 1.14 [0.86-1.50] | 1.17 [0.88-1.55] |
| T38 | 1,840 | 46,394,820 | 0.04 | 0.04 | 0.04 | 0.00 | 0.00 | 0.00 | 1.05 [0.89-1.22] | 1.02 [0.87-1.20] |
| T39 | 35,760 | 46,003,750 | 0.78 | 0.78 | 0.77 | 0.01 | 0.02 | 0.01 | 1.02 [0.98-1.06] | 1.01 [0.97-1.05] |
| T40 | 9,850 | 46,326,630 | 0.20 | 0.22 | 0.21 | −0.01 | −0.01 | 0.00 | 0.97 [0.90-1.04] | 0.98 [0.91-1.05] |
| T42 | 5,430 | 46,358,520 | 0.11 | 0.12 | 0.11 | −0.01 | 0.00 | 0.00 | 0.96 [0.87-1.05] | 0.98 [0.88-1.08] |
| T43 | 13,380 | 46,284,310 | 0.28 | 0.29 | 0.28 | −0.02 | −0.01 | −0.01 | 0.97 [0.91-1.03] | 0.98 [0.92-1.04] |
| T44 | 2,770 | 46,383,170 | 0.06 | 0.06 | 0.06 | 0.00 | 0.00 | 0.00 | 1.01 [0.89-1.15] | 1.00 [0.88-1.15] |
| T45 | 4,250 | 46,363,010 | 0.10 | 0.09 | 0.09 | 0.02 | 0.02 | 0.01 | 1.18 [1.07-1.31]* | 1.14 [1.03-1.27] |
| T46 | 1,240 | 46,397,710 | 0.03 | 0.03 | 0.03 | 0.00 | 0.00 | 0.00 | 1.00 [0.83-1.21] | 0.98 [0.81-1.19] |
| T47 | 1,380 | 46,402,500 | 0.03 | 0.03 | 0.03 | 0.00 | 0.00 | 0.00 | 0.95 [0.79-1.15] | 0.94 [0.77-1.14] |
| T48 | 1,280 | 46,389,260 | 0.03 | 0.03 | 0.03 | 0.00 | 0.00 | 0.00 | 1.05 [0.87-1.26] | 1.01 [0.83-1.22] |
| T49 | 1,210 | 46,389,840 | 0.02 | 0.03 | 0.03 | 0.00 | 0.00 | 0.00 | 0.95 [0.79-1.16] | 0.92 [0.76-1.12] |
| T50 | 2,970 | 46,372,590 | 0.06 | 0.07 | 0.06 | −0.01 | 0.00 | 0.00 | 0.96 [0.85-1.09] | 0.95 [0.83-1.08] |
| T51 | 9,260 | 46,327,950 | 0.18 | 0.20 | 0.19 | −0.02 | 0.00 | 0.00 | 0.98 [0.91-1.05] | 1.00 [0.93-1.08] |
| T52 | 1,260 | 46,384,930 | 0.02 | 0.03 | 0.03 | −0.01 | 0.00 | 0.00 | 0.83 [0.68-1.01] | 0.83 [0.68-1.01] |
| T54 | 1,060 | 46,403,010 | 0.02 | 0.02 | 0.03 | 0.00 | 0.00 | 0.00 | 0.86 [0.70-1.07] | 0.85 [0.68-1.06] |
| T59 | 500 | 46,405,480 | 0.01 | 0.01 | 0.01 | 0.00 | 0.00 | 0.00 | 1.04 [0.77-1.41] | 1.05 [0.77-1.43] |
| T62 | 500 | 46,405,960 | 0.01 | 0.01 | 0.01 | 0.00 | 0.00 | 0.00 | 1.09 [0.82-1.46] | 1.06 [0.79-1.43] |
| T65 | 1,050 | 46,394,280 | 0.02 | 0.02 | 0.02 | 0.00 | 0.00 | 0.00 | 0.99 [0.80-1.21] | 0.97 [0.78-1.19] |
| T74 | 1,040 | 46,394,520 | 0.02 | 0.02 | 0.02 | 0.00 | 0.00 | 0.00 | 0.85 [0.68-1.06] | 0.82 [0.66-1.03] |
| T75 | 610 | 46,404,280 | 0.01 | 0.01 | 0.01 | 0.00 | 0.00 | 0.00 | 1.01 [0.76-1.32] | 0.97 [0.73-1.28] |
| T78 | 12,590 | 46,119,040 | 0.71 | 0.15 | 0.15 | 0.56 | 0.57 | 0.53 | 4.81 [4.57-5.06]* | 3.86 [3.66-4.07]* |
| T79 | 2,030 | 46,388,240 | 0.04 | 0.04 | 0.04 | 0.00 | 0.00 | 0.00 | 1.05 [0.90-1.22] | 1.03 [0.89-1.21] |
| T80 | 1,160 | 46,390,000 | 0.03 | 0.02 | 0.03 | 0.00 | 0.00 | 0.00 | 1.09 [0.90-1.33] | 1.08 [0.89-1.32] |
| T81 | 26,500 | 46,025,840 | 0.66 | 0.55 | 0.54 | 0.11 | 0.12 | 0.10 | 1.22 [1.17-1.27]* | 1.17 [1.12-1.22]* |
| T82 | 4,550 | 46,338,940 | 0.10 | 0.10 | 0.10 | 0.00 | 0.00 | 0.00 | 1.01 [0.91-1.11] | 0.99 [0.89-1.09] |
| T83 | 1,840 | 46,400,870 | 0.04 | 0.04 | 0.04 | 0.00 | 0.00 | 0.00 | 1.02 [0.87-1.20] | 0.96 [0.81-1.14] |
| T84 | 5,300 | 46,367,010 | 0.12 | 0.11 | 0.11 | 0.01 | 0.01 | 0.01 | 1.14 [1.04-1.25]* | 1.11 [1.01-1.22] |
| T85 | 6,280 | 46,316,080 | 0.15 | 0.13 | 0.13 | 0.02 | 0.02 | 0.02 | 1.17 [1.08-1.27]* | 1.12 [1.03-1.22] |
| T86 | 940 | 46,406,040 | 0.02 | 0.02 | 0.02 | 0.00 | 0.00 | 0.00 | 1.02 [0.82-1.28] | 1.01 [0.81-1.27] |
| T88 | 2,250 | 46,354,530 | 0.06 | 0.04 | 0.04 | 0.02 | 0.02 | 0.01 | 1.40 [1.23-1.60]* | 1.29 [1.13-1.48]* |
| T90 | 3,150 | 46,378,010 | 0.07 | 0.07 | 0.06 | 0.01 | 0.01 | 0.01 | 1.15 [1.02-1.29] | 1.13 [1.00-1.27] |
| T91 | 540 | 46,412,570 | 0.01 | 0.01 | 0.01 | 0.00 | 0.00 | 0.00 | 1.07 [0.79-1.43] | 1.08 [0.79-1.46] |
| T92 | 1,840 | 46,393,390 | 0.04 | 0.04 | 0.04 | 0.00 | 0.00 | 0.00 | 1.07 [0.91-1.26] | 1.04 [0.88-1.23] |
| T93 | 2,520 | 46,389,440 | 0.05 | 0.05 | 0.05 | 0.00 | 0.00 | 0.00 | 1.10 [0.96-1.26] | 1.05 [0.91-1.21] |
| T95 | 580 | 46,406,960 | 0.01 | 0.01 | 0.01 | 0.00 | 0.00 | 0.00 | 0.80 [0.60-1.07] | 0.82 [0.61-1.11] |
| <18 - U00-U89 | | | | | | | | | | |
| U07 | 3,600 | 46,412,830 | 0.08 | 0.08 | 0.07 | 0.01 | 0.01 | 0.01 | 1.14 [1.02-1.29] | 1.09 [0.96-1.23] |
| U80 | 1,500 | 46,390,800 | 0.04 | 0.03 | 0.03 | 0.01 | 0.01 | 0.01 | 1.41 [1.20-1.65]* | 1.33 [1.12-1.57]* |
| U82 | 3,720 | 46,397,420 | 0.10 | 0.07 | 0.08 | 0.02 | 0.02 | 0.02 | 1.30 [1.17-1.45]* | 1.26 [1.13-1.41]* |
| U83 | 2,190 | 46,406,430 | 0.06 | 0.04 | 0.05 | 0.01 | 0.01 | 0.01 | 1.22 [1.06-1.41]* | 1.17 [1.01-1.35] |
| U89 | 480 | 46,410,560 | 0.01 | 0.01 | 0.01 | 0.00 | 0.00 | 0.00 | 1.33 [1.00-1.77] | 1.28 [0.96-1.72] |
| <18 - V00-Y99 | | | | | | | | | | |
| V03 | 6,080 | 46,309,040 | 0.12 | 0.13 | 0.13 | −0.01 | −0.01 | −0.01 | 0.93 [0.85-1.02] | 0.93 [0.85-1.02] |
| V09 | 910 | 46,404,240 | 0.02 | 0.02 | 0.02 | 0.00 | 0.00 | 0.00 | 1.12 [0.90-1.39] | 1.08 [0.87-1.35] |
| V13 | 1,430 | 46,396,740 | 0.03 | 0.03 | 0.03 | 0.00 | 0.00 | 0.00 | 0.94 [0.78-1.13] | 0.97 [0.80-1.17] |
| V17 | 740 | 46,408,690 | 0.02 | 0.02 | 0.02 | 0.00 | 0.00 | 0.00 | 0.97 [0.75-1.25] | 0.96 [0.75-1.24] |
| V18 | 12,720 | 46,208,550 | 0.28 | 0.27 | 0.27 | 0.01 | 0.01 | 0.00 | 1.03 [0.97-1.09] | 1.00 [0.94-1.07] |
| V19 | 1,880 | 46,390,410 | 0.04 | 0.04 | 0.04 | 0.00 | 0.00 | 0.00 | 1.02 [0.87-1.19] | 1.01 [0.87-1.19] |
| V23 | 1,890 | 46,402,640 | 0.04 | 0.04 | 0.04 | −0.01 | 0.00 | 0.00 | 0.93 [0.79-1.10] | 0.95 [0.80-1.13] |
| V27 | 570 | 46,412,740 | 0.01 | 0.01 | 0.01 | 0.00 | 0.00 | 0.00 | 0.91 [0.67-1.23] | 0.89 [0.65-1.22] |
| V28 | 2,710 | 46,389,470 | 0.05 | 0.06 | 0.05 | −0.01 | 0.00 | 0.00 | 1.01 [0.88-1.15] | 0.98 [0.85-1.12] |
| V29 | 660 | 46,410,560 | 0.01 | 0.01 | 0.01 | 0.00 | 0.00 | 0.00 | 0.91 [0.69-1.20] | 0.91 [0.69-1.21] |
| V43 | 2,640 | 46,386,510 | 0.05 | 0.06 | 0.05 | 0.00 | 0.00 | 0.00 | 1.04 [0.91-1.19] | 1.02 [0.88-1.17] |
| V47 | 1,670 | 46,401,250 | 0.04 | 0.04 | 0.03 | 0.00 | 0.00 | 0.00 | 1.04 [0.88-1.23] | 1.02 [0.86-1.22] |
| V48 | 1,300 | 46,402,250 | 0.03 | 0.03 | 0.02 | 0.00 | 0.00 | 0.00 | 1.13 [0.94-1.36] | 1.12 [0.92-1.36] |
| V49 | 1,300 | 46,399,510 | 0.03 | 0.03 | 0.03 | 0.00 | 0.00 | 0.00 | 1.06 [0.88-1.28] | 1.06 [0.87-1.29] |
| V80 | 2,920 | 46,378,460 | 0.06 | 0.06 | 0.06 | 0.00 | 0.00 | 0.00 | 1.03 [0.90-1.17] | 1.01 [0.88-1.15] |
| V86 | 920 | 46,409,230 | 0.02 | 0.02 | 0.02 | 0.00 | 0.00 | 0.00 | 1.14 [0.91-1.42] | 1.10 [0.87-1.38] |
| V89 | 610 | 46,404,490 | 0.01 | 0.01 | 0.01 | 0.00 | 0.00 | 0.00 | 0.90 [0.67-1.20] | 0.85 [0.63-1.15] |
| W00 | 820 | 46,407,680 | 0.02 | 0.02 | 0.02 | 0.00 | 0.00 | 0.00 | 1.19 [0.94-1.50] | 1.19 [0.93-1.51] |

| | | | | | | | | | | |
|---|---|---|---|---|---|---|---|---|---|---|
| W01 | 23,640 | 46,019,140 | 0.53 | 0.51 | 0.51 | 0.02 | 0.02 | 0.01 | 1.03 [0.99-1.08] | 1.02 [0.97-1.06] |
| W02 | 6,950 | 46,315,910 | 0.16 | 0.15 | 0.15 | 0.01 | 0.00 | 0.00 | 1.00 [0.92-1.09] | 1.00 [0.92-1.08] |
| W03 | 9,120 | 46,299,380 | 0.20 | 0.20 | 0.20 | 0.01 | 0.00 | 0.00 | 1.02 [0.95-1.10] | 1.00 [0.93-1.07] |
| W04 | 1,570 | 46,307,360 | 0.03 | 0.03 | 0.03 | 0.00 | 0.00 | 0.00 | 0.97 [0.82-1.15] | 0.97 [0.81-1.15] |
| W06 | 5,250 | 46,257,500 | 0.11 | 0.11 | 0.11 | 0.00 | 0.00 | 0.00 | 1.03 [0.94-1.13] | 1.02 [0.93-1.12] |
| W07 | 5,040 | 46,287,490 | 0.11 | 0.11 | 0.11 | 0.00 | 0.00 | 0.00 | 0.97 [0.88-1.06] | 0.96 [0.87-1.06] |
| W08 | 3,700 | 46,322,300 | 0.07 | 0.08 | 0.08 | −0.01 | −0.01 | −0.01 | 0.89 [0.80-1.00] | 0.89 [0.80-1.00] |
| W09 | 22,650 | 46,068,570 | 0.53 | 0.48 | 0.50 | 0.05 | 0.03 | 0.02 | 1.05 [1.01-1.10] | 1.04 [0.99-1.09] |
| W10 | 10,850 | 46,193,640 | 0.23 | 0.24 | 0.23 | −0.01 | 0.00 | 0.00 | 1.02 [0.95-1.08] | 1.00 [0.94-1.07] |
| W11 | 620 | 46,409,890 | 0.01 | 0.01 | 0.01 | 0.00 | 0.00 | 0.00 | 1.05 [0.80-1.38] | 1.06 [0.80-1.40] |
| W13 | 5,180 | 46,322,050 | 0.11 | 0.11 | 0.11 | −0.01 | −0.01 | −0.01 | 0.95 [0.87-1.05] | 0.94 [0.85-1.04] |
| W14 | 2,640 | 46,368,580 | 0.06 | 0.06 | 0.06 | 0.00 | 0.00 | 0.00 | 0.95 [0.83-1.08] | 0.94 [0.82-1.08] |
| W17 | 9,320 | 46,235,120 | 0.20 | 0.20 | 0.20 | 0.00 | −0.01 | −0.01 | 0.97 [0.91-1.04] | 0.95 [0.88-1.02] |
| W18 | 14,030 | 46,200,880 | 0.32 | 0.30 | 0.30 | 0.02 | 0.02 | 0.01 | 1.06 [1.00-1.12] | 1.05 [0.99-1.11] |
| W19 | 22,970 | 46,019,900 | 0.51 | 0.50 | 0.49 | 0.01 | 0.02 | 0.02 | 1.04 [1.00-1.09] | 1.03 [0.98-1.08] |
| W20 | 3,410 | 46,359,720 | 0.07 | 0.07 | 0.07 | 0.00 | 0.00 | 0.00 | 0.99 [0.88-1.11] | 0.96 [0.85-1.08] |
| W21 | 5,110 | 46,348,870 | 0.12 | 0.11 | 0.11 | 0.01 | 0.01 | 0.01 | 1.12 [1.02-1.23] | 1.10 [1.00-1.21] |
| W22 | 17,590 | 46,139,140 | 0.39 | 0.38 | 0.37 | 0.01 | 0.02 | 0.01 | 1.05 [1.00-1.10] | 1.03 [0.98-1.09] |
| W23 | 15,360 | 46,171,690 | 0.33 | 0.33 | 0.33 | 0.00 | 0.00 | 0.00 | 1.00 [0.95-1.05] | 0.99 [0.94-1.05] |
| W25 | 6,660 | 46,323,140 | 0.13 | 0.15 | 0.14 | −0.01 | −0.01 | −0.01 | 0.95 [0.87-1.03] | 0.94 [0.86-1.02] |
| W26 | 4,540 | 46,384,190 | 0.10 | 0.10 | 0.10 | 0.00 | 0.00 | 0.00 | 1.04 [0.93-1.15] | 1.03 [0.93-1.14] |
| W27 | 1,590 | 46,395,240 | 0.03 | 0.04 | 0.03 | −0.01 | 0.00 | 0.00 | 0.89 [0.74-1.06] | 0.89 [0.74-1.06] |
| W29 | 1,090 | 46,407,250 | 0.02 | 0.02 | 0.02 | 0.00 | 0.00 | 0.00 | 0.98 [0.79-1.21] | 0.98 [0.79-1.23] |
| W31 | 1,170 | 46,406,300 | 0.03 | 0.02 | 0.02 | 0.00 | 0.00 | 0.00 | 1.17 [0.96-1.42] | 1.16 [0.95-1.43] |
| W34 | 530 | 46,411,290 | 0.01 | 0.01 | 0.01 | 0.00 | 0.00 | 0.00 | 1.04 [0.77-1.41] | 1.04 [0.76-1.43] |
| W44 | 17,070 | 46,123,250 | 0.38 | 0.37 | 0.37 | 0.01 | 0.01 | 0.00 | 1.02 [0.97-1.08] | 1.01 [0.96-1.06] |
| W45 | 4,220 | 46,350,020 | 0.10 | 0.09 | 0.09 | 0.01 | 0.01 | 0.00 | 1.06 [0.96-1.18] | 1.05 [0.94-1.16] |
| W49 | 980 | 46,399,910 | 0.02 | 0.02 | 0.02 | 0.00 | 0.00 | 0.00 | 1.01 [0.81-1.26] | 1.01 [0.81-1.26] |
| W50 | 8,540 | 46,310,840 | 0.19 | 0.18 | 0.18 | 0.01 | 0.01 | 0.01 | 1.08 [1.00-1.16] | 1.06 [0.98-1.14] |
| W51 | 6,930 | 46,320,010 | 0.16 | 0.15 | 0.15 | 0.01 | 0.01 | 0.00 | 1.04 [0.96-1.13] | 1.01 [0.93-1.10] |
| W54 | 5,490 | 46,339,890 | 0.11 | 0.12 | 0.12 | −0.01 | −0.01 | −0.02 | 0.88 [0.80-0.97]* | 0.86 [0.78-0.95]* |
| W55 | 1,240 | 46,403,170 | 0.03 | 0.03 | 0.03 | 0.00 | 0.00 | 0.00 | 0.99 [0.81-1.20] | 0.98 [0.80-1.20] |
| W57 | 2,140 | 46,394,660 | 0.05 | 0.04 | 0.04 | 0.01 | 0.01 | 0.01 | 1.26 [1.09-1.45]* | 1.24 [1.08-1.44]* |
| W79 | 670 | 46,403,460 | 0.02 | 0.01 | 0.01 | 0.01 | 0.01 | 0.01 | 1.96 [1.56-2.47]* | 1.68 [1.32-2.14]* |
| X10 | 3,280 | 46,339,990 | 0.07 | 0.07 | 0.07 | 0.00 | 0.00 | 0.00 | 0.98 [0.87-1.10] | 0.98 [0.87-1.10] |
| X11 | 870 | 46,391,750 | 0.02 | 0.02 | 0.02 | 0.00 | 0.00 | 0.00 | 1.01 [0.81-1.26] | 0.99 [0.79-1.24] |
| X12 | 1,660 | 46,368,680 | 0.03 | 0.04 | 0.04 | 0.00 | 0.00 | −0.01 | 0.87 [0.74-1.03] | 0.87 [0.73-1.03] |
| X15 | 1,560 | 46,386,830 | 0.03 | 0.03 | 0.03 | 0.00 | 0.00 | 0.00 | 0.99 [0.83-1.17] | 0.99 [0.83-1.17] |
| X40 | 8,890 | 46,270,870 | 0.19 | 0.19 | 0.19 | −0.01 | 0.00 | 0.00 | 1.00 [0.94-1.08] | 1.00 [0.93-1.08] |
| X41 | 3,680 | 46,353,390 | 0.08 | 0.08 | 0.08 | 0.00 | 0.00 | 0.00 | 0.98 [0.88-1.10] | 0.99 [0.88-1.11] |
| X42 | 2,200 | 46,393,440 | 0.04 | 0.05 | 0.05 | 0.00 | 0.00 | 0.00 | 0.98 [0.85-1.14] | 0.98 [0.84-1.14] |
| X43 | 780 | 46,399,740 | 0.02 | 0.02 | 0.02 | 0.00 | 0.00 | 0.00 | 0.95 [0.75-1.22] | 0.92 [0.71-1.18] |
| X44 | 5,840 | 46,282,630 | 0.13 | 0.13 | 0.12 | 0.00 | 0.00 | 0.00 | 1.03 [0.94-1.12] | 1.01 [0.92-1.10] |
| X45 | 1,440 | 46,398,890 | 0.03 | 0.03 | 0.03 | 0.00 | 0.00 | 0.00 | 0.92 [0.76-1.11] | 0.94 [0.78-1.14] |
| X46 | 910 | 46,391,450 | 0.02 | 0.02 | 0.02 | 0.00 | 0.00 | 0.00 | 0.83 [0.66-1.04] | 0.82 [0.65-1.04] |
| X49 | 3,310 | 46,340,160 | 0.09 | 0.07 | 0.07 | 0.02 | 0.02 | 0.02 | 1.37 [1.23-1.52]* | 1.28 [1.14-1.43]* |
| X50 | 5,630 | 46,362,320 | 0.12 | 0.12 | 0.12 | 0.00 | 0.01 | 0.00 | 1.05 [0.96-1.15] | 1.01 [0.92-1.11] |
| X58 | 1,720 | 46,387,930 | 0.06 | 0.03 | 0.03 | 0.03 | 0.03 | 0.03 | 2.06 [1.78-2.38]* | 1.79 [1.54-2.08]* |
| X59 | 24,560 | 45,922,830 | 0.55 | 0.53 | 0.51 | 0.02 | 0.04 | 0.03 | 1.08 [1.03-1.12]* | 1.05 [1.00-1.09] |
| X60 | 27,590 | 46,146,220 | 0.61 | 0.60 | 0.59 | 0.01 | 0.01 | 0.01 | 1.02 [0.98-1.07] | 1.01 [0.97-1.06] |
| X61 | 13,160 | 46,306,860 | 0.27 | 0.29 | 0.28 | −0.01 | −0.01 | 0.00 | 0.97 [0.92-1.04] | 0.98 [0.92-1.05] |
| X62 | 7,750 | 46,351,590 | 0.16 | 0.17 | 0.16 | −0.01 | 0.00 | 0.00 | 0.98 [0.90-1.06] | 0.98 [0.91-1.07] |
| X63 | 1,970 | 46,402,370 | 0.04 | 0.04 | 0.04 | 0.00 | 0.00 | 0.00 | 1.05 [0.90-1.23] | 1.06 [0.90-1.24] |
| X64 | 8,140 | 46,336,520 | 0.18 | 0.17 | 0.17 | 0.01 | 0.01 | 0.01 | 1.05 [0.98-1.14] | 1.03 [0.95-1.11] |
| X65 | 7,880 | 46,347,290 | 0.16 | 0.17 | 0.16 | −0.02 | 0.00 | 0.00 | 0.99 [0.92-1.08] | 1.02 [0.94-1.11] |
| X69 | 1,000 | 46,409,450 | 0.02 | 0.02 | 0.02 | 0.00 | 0.00 | 0.00 | 1.03 [0.83-1.27] | 1.01 [0.81-1.26] |
| X70 | 700 | 46,413,150 | 0.01 | 0.02 | 0.01 | 0.00 | 0.00 | 0.00 | 0.98 [0.75-1.27] | 1.04 [0.80-1.37] |
| X78 | 7,320 | 46,361,400 | 0.17 | 0.16 | 0.16 | 0.01 | 0.00 | 0.00 | 1.03 [0.95-1.11] | 1.01 [0.93-1.09] |
| X83 | 570 | 46,414,240 | 0.01 | 0.01 | 0.01 | 0.00 | 0.00 | 0.00 | 0.87 [0.65-1.17] | 0.84 [0.62-1.13] |
| X84 | 860 | 46,411,570 | 0.02 | 0.02 | 0.02 | 0.00 | 0.00 | 0.00 | 0.95 [0.75-1.22] | 0.95 [0.74-1.22] |
| X99 | 3,030 | 46,394,160 | 0.06 | 0.07 | 0.06 | 0.00 | 0.00 | 0.00 | 1.02 [0.90-1.16] | 1.02 [0.90-1.17] |
| Y00 | 1,070 | 46,407,070 | 0.02 | 0.02 | 0.02 | 0.00 | 0.00 | 0.00 | 0.91 [0.73-1.13] | 0.89 [0.71-1.11] |
| Y04 | 11,900 | 46,284,710 | 0.23 | 0.26 | 0.23 | −0.03 | 0.00 | 0.00 | 1.00 [0.94-1.07] | 0.99 [0.93-1.06] |
| Y07 | 1,020 | 46,393,990 | 0.02 | 0.02 | 0.02 | 0.00 | 0.00 | 0.00 | 0.92 [0.74-1.15] | 0.92 [0.74-1.15] |
| Y09 | 1,280 | 46,402,880 | 0.02 | 0.03 | 0.02 | 0.00 | 0.00 | 0.00 | 1.08 [0.89-1.32] | 1.06 [0.86-1.30] |
| Y40 | 2,930 | 46,371,570 | 0.08 | 0.06 | 0.06 | 0.02 | 0.02 | 0.01 | 1.30 [1.15-1.47]* | 1.25 [1.10-1.41]* |
| Y41 | 430 | 46,412,940 | 0.01 | 0.01 | 0.01 | 0.00 | 0.00 | 0.00 | 1.17 [0.85-1.62] | 1.09 [0.78-1.54] |
| Y42 | 1,290 | 46,405,360 | 0.03 | 0.03 | 0.02 | 0.01 | 0.01 | 0.01 | 1.36 [1.14-1.63]* | 1.30 [1.08-1.57]* |
| Y43 | 3,240 | 46,380,350 | 0.07 | 0.07 | 0.07 | 0.01 | 0.01 | 0.01 | 1.11 [0.99-1.24] | 1.08 [0.96-1.22] |
| Y44 | 400 | 46,414,730 | 0.01 | 0.01 | 0.01 | 0.00 | 0.00 | 0.00 | 1.40 [1.03-1.92] | 1.38 [1.00-1.90] |
| Y45 | 1,620 | 46,402,410 | 0.04 | 0.03 | 0.03 | 0.01 | 0.01 | 0.01 | 1.29 [1.10-1.52]* | 1.22 [1.03-1.45] |
| Y46 | 440 | 46,412,890 | 0.01 | 0.01 | 0.01 | 0.00 | 0.00 | 0.00 | 1.20 [0.88-1.65] | 1.16 [0.84-1.60] |
| Y55 | 860 | 46,411,760 | 0.02 | 0.02 | 0.02 | 0.00 | 0.01 | 0.00 | 1.31 [1.05-1.64] | 1.22 [0.96-1.55] |
| Y57 | 650 | 46,407,380 | 0.02 | 0.01 | 0.01 | 0.01 | 0.01 | 0.01 | 1.84 [1.44-2.33]* | 1.65 [1.28-2.13]* |
| Y59 | 490 | 46,399,430 | 0.02 | 0.01 | 0.01 | 0.01 | 0.01 | 0.01 | 1.62 [1.23-2.13]* | 1.50 [1.13-1.99]* |
| Y60 | 850 | 46,410,280 | 0.02 | 0.02 | 0.02 | 0.00 | 0.00 | 0.00 | 1.10 [0.87-1.39] | 1.07 [0.85-1.36] |
| Y83 | 32,540 | 45,969,670 | 0.81 | 0.68 | 0.66 | 0.13 | 0.15 | 0.12 | 1.22 [1.18-1.27]* | 1.17 [1.13-1.22]* |
| Y84 | 3,140 | 46,367,110 | 0.08 | 0.07 | 0.06 | 0.01 | 0.01 | 0.01 | 1.21 [1.08-1.36]* | 1.16 [1.03-1.31] |
| Y85 | 980 | 46,407,380 | 0.02 | 0.02 | 0.02 | 0.00 | 0.00 | 0.00 | 1.16 [0.93-1.44] | 1.16 [0.93-1.46] |
| Y86 | 2,910 | 46,381,050 | 0.06 | 0.06 | 0.06 | 0.00 | 0.00 | 0.00 | 0.99 [0.87-1.13] | 0.97 [0.85-1.11] |
| Y87 | 580 | 46,412,060 | 0.01 | 0.01 | 0.01 | 0.00 | 0.00 | 0.00 | 1.05 [0.78-1.40] | 0.98 [0.72-1.33] |
| Y95 | 930 | 46,411,760 | 0.02 | 0.02 | 0.02 | 0.01 | 0.01 | 0.01 | 1.39 [1.12-1.73]* | 1.43 [1.14-1.79]* |
| <18 - Z00-Z99 | | | | | | | | | | |
| Z00 | 920 | 46,355,600 | 0.02 | 0.02 | 0.02 | 0.00 | 0.00 | 0.00 | 1.18 [0.95-1.46] | 1.17 [0.94-1.46] |
| Z01 | 19,200 | 45,677,820 | 0.90 | 0.29 | 0.29 | 0.61 | 0.61 | 0.54 | 3.12 [3.00-3.25]* | 2.48 [2.38-2.59]* |
| Z02 | 1,810 | 46,400,720 | 0.03 | 0.04 | 0.03 | −0.01 | 0.00 | 0.00 | 1.06 [0.89-1.25] | 1.04 [0.87-1.24] |
| Z03 | 33,510 | 44,686,530 | 0.94 | 0.70 | 0.68 | 0.24 | 0.26 | 0.22 | 1.38 [1.34-1.43]* | 1.30 [1.25-1.34]* |
| Z04 | 11,460 | 45,929,180 | 0.26 | 0.25 | 0.23 | 0.01 | 0.03 | 0.03 | 1.14 [1.07-1.21]* | 1.11 [1.04-1.18]* |
| Z08 | 1,400 | 46,400,340 | 0.03 | 0.03 | 0.03 | 0.00 | 0.00 | 0.00 | 0.98 [0.82-1.18] | 1.01 [0.84-1.22] |
| Z09 | 10,380 | 46,205,250 | 0.25 | 0.22 | 0.21 | 0.03 | 0.04 | 0.03 | 1.18 [1.11-1.26]* | 1.14 [1.06-1.21]* |
| Z11 | 57,710 | 46,286,310 | 1.39 | 1.21 | 1.19 | 0.19 | 0.21 | 0.17 | 1.17 [1.14-1.21]* | 1.14 [1.11-1.17]* |
| Z12 | 550 | 46,409,370 | 0.01 | 0.01 | 0.01 | 0.00 | 0.00 | 0.00 | 1.30 [0.98-1.74] | 1.23 [0.91-1.65] |
| Z13 | 33,760 | 45,650,510 | 0.89 | 0.70 | 0.65 | 0.20 | 0.24 | 0.20 | 1.37 [1.32-1.42]* | 1.29 [1.24-1.33]* |
| Z20 | 1,950 | 46,370,790 | 0.05 | 0.04 | 0.04 | 0.01 | 0.01 | 0.01 | 1.22 [1.06-1.42]* | 1.19 [1.03-1.39] |
| Z22 | 13,220 | 46,267,650 | 0.29 | 0.28 | 0.26 | 0.00 | 0.03 | 0.03 | 1.13 [1.06-1.20]* | 1.10 [1.03-1.17]* |
| Z24 | 810 | 46,349,990 | 0.02 | 0.02 | 0.02 | 0.00 | 0.00 | 0.00 | 1.02 [0.80-1.31] | 0.98 [0.76-1.27] |
| Z25 | 640 | 46,398,460 | 0.03 | 0.01 | 0.01 | 0.02 | 0.02 | 0.02 | 3.36 [2.69-4.21]* | 2.52 [1.98-3.21]* |
| Z27 | 3,260 | 46,278,730 | 0.14 | 0.05 | 0.04 | 0.09 | 0.10 | 0.09 | 3.25 [2.94-3.59]* | 2.71 [2.44-3.01]* |
| Z29 | 8,640 | 45,598,000 | 0.20 | 0.19 | 0.17 | 0.02 | 0.03 | 0.03 | 1.17 [1.09-1.26]* | 1.14 [1.06-1.23]* |
| Z30 | 5,960 | 46,348,660 | 0.11 | 0.13 | 0.11 | −0.02 | 0.00 | 0.00 | 1.03 [0.94-1.13] | 1.03 [0.94-1.13] |
| Z32 | 1,150 | 46,404,670 | 0.02 | 0.03 | 0.02 | 0.00 | 0.00 | 0.00 | 1.06 [0.86-1.31] | 1.07 [0.86-1.34] |
| Z33 | 12,380 | 46,309,960 | 0.26 | 0.27 | 0.23 | −0.01 | 0.03 | 0.02 | 1.12 [1.05-1.20]* | 1.10 [1.03-1.18]* |
| Z34 | 32,090 | 46,123,090 | 0.63 | 0.71 | 0.63 | −0.09 | −0.01 | −0.01 | 0.99 [0.95-1.03] | 0.99 [0.95-1.03] |
| Z35 | 20,940 | 46,284,360 | 0.40 | 0.47 | 0.41 | −0.07 | −0.01 | −0.02 | 0.97 [0.92-1.02] | 0.96 [0.91-1.01] |
| Z36 | 3,280 | 46,372,610 | 0.06 | 0.07 | 0.06 | −0.01 | 0.00 | 0.00 | 0.99 [0.87-1.12] | 0.97 [0.85-1.10] |
| Z37 | 138,770 | 45,098,260 | 2.81 | 3.15 | 2.87 | −0.34 | −0.06 | −0.06 | 0.98 [0.96-1.00] | 0.98 [0.96-1.00] |

| | | | | | | | | | | |
|---|---|---|---|---|---|---|---|---|---|---|
| Z39 | 7,720 | 46,344,620 | 0.15 | 0.17 | 0.14 | −0.02 | 0.01 | 0.01 | 1.05 [0.97-1.14] | 1.06 [0.97-1.15] |
| Z40 | 840 | 46,412,250 | 0.02 | 0.02 | 0.02 | 0.00 | 0.00 | 0.00 | 1.07 [0.85-1.35] | 1.05 [0.83-1.33] |
| Z41 | 2,760 | 46,314,530 | 0.08 | 0.06 | 0.05 | 0.02 | 0.02 | 0.02 | 1.39 [1.24-1.57]* | 1.39 [1.23-1.57]* |
| Z42 | 1,680 | 46,391,800 | 0.04 | 0.04 | 0.03 | 0.00 | 0.01 | 0.00 | 1.15 [0.98-1.36] | 1.15 [0.97-1.36] |
| Z43 | 3,640 | 46,342,440 | 0.07 | 0.08 | 0.08 | −0.01 | −0.01 | −0.01 | 0.93 [0.83-1.04] | 0.87 [0.77-0.98] |
| Z45 | 5,990 | 46,312,530 | 0.14 | 0.13 | 0.13 | 0.01 | 0.01 | 0.01 | 1.08 [0.99-1.18] | 1.04 [0.96-1.14] |
| Z46 | 4,270 | 46,345,420 | 0.09 | 0.09 | 0.09 | 0.00 | 0.00 | 0.00 | 1.05 [0.95-1.17] | 1.02 [0.92-1.14] |
| Z47 | 24,700 | 46,002,290 | 0.55 | 0.53 | 0.53 | 0.02 | 0.02 | 0.01 | 1.03 [0.99-1.08] | 1.02 [0.97-1.06] |
| Z48 | 5,650 | 46,265,370 | 0.13 | 0.12 | 0.12 | 0.01 | 0.01 | 0.01 | 1.10 [1.01-1.20] | 1.06 [0.97-1.16] |
| Z50 | 16,820 | 46,298,690 | 0.40 | 0.35 | 0.36 | 0.04 | 0.04 | 0.03 | 1.12 [1.06-1.18]* | 1.07 [1.02-1.13] |
| Z51 | 24,270 | 46,015,510 | 0.60 | 0.51 | 0.49 | 0.09 | 0.11 | 0.08 | 1.22 [1.17-1.27]* | 1.17 [1.12-1.22]* |
| Z53 | 54,150 | 45,537,250 | 1.38 | 1.14 | 1.11 | 0.25 | 0.27 | 0.22 | 1.25 [1.21-1.28]* | 1.19 [1.16-1.23]* |
| Z55 | 1,380 | 46,405,520 | 0.04 | 0.03 | 0.03 | 0.01 | 0.01 | 0.01 | 1.23 [1.03-1.46] | 1.19 [0.99-1.42] |
| Z56 | 820 | 46,413,400 | 0.02 | 0.02 | 0.02 | 0.00 | 0.00 | 0.00 | 1.00 [0.78-1.29] | 0.96 [0.73-1.25] |
| Z58 | 2,680 | 46,376,250 | 0.08 | 0.05 | 0.05 | 0.02 | 0.02 | 0.02 | 1.44 [1.27-1.62]* | 1.36 [1.20-1.54]* |
| Z59 | 1,780 | 46,399,370 | 0.04 | 0.04 | 0.04 | 0.00 | 0.00 | 0.00 | 1.02 [0.87-1.21] | 1.08 [0.90-1.28] |
| Z60 | 2,590 | 46,363,180 | 0.05 | 0.06 | 0.06 | 0.00 | 0.00 | 0.00 | 0.92 [0.80-1.06] | 0.92 [0.80-1.06] |
| Z61 | 3,240 | 46,379,370 | 0.07 | 0.07 | 0.07 | 0.00 | 0.00 | 0.00 | 0.98 [0.87-1.11] | 0.97 [0.86-1.10] |
| Z62 | 890 | 46,398,870 | 0.02 | 0.02 | 0.02 | 0.00 | 0.00 | 0.00 | 0.96 [0.77-1.21] | 0.94 [0.75-1.19] |
| Z63 | 12,700 | 46,233,570 | 0.27 | 0.28 | 0.29 | −0.01 | −0.02 | −0.02 | 0.94 [0.88-1.00] | 0.92 [0.86-0.98] |
| Z65 | 1,190 | 46,404,610 | 0.02 | 0.03 | 0.03 | 0.00 | 0.00 | 0.00 | 0.96 [0.78-1.17] | 0.99 [0.80-1.23] |
| Z71 | 10,530 | 46,194,010 | 0.47 | 0.16 | 0.16 | 0.31 | 0.31 | 0.27 | 2.95 [2.79-3.12]* | 2.36 [2.22-2.50]* |
| Z72 | 31,140 | 46,088,140 | 0.61 | 0.69 | 0.59 | −0.09 | 0.01 | 0.01 | 1.02 [0.98-1.06] | 1.02 [0.98-1.07] |
| Z73 | 1,840 | 46,404,950 | 0.04 | 0.04 | 0.04 | 0.00 | 0.00 | 0.00 | 1.08 [0.91-1.27] | 1.05 [0.88-1.24] |
| Z75 | 1,700 | 46,387,370 | 0.04 | 0.04 | 0.04 | 0.00 | 0.00 | 0.00 | 1.13 [0.96-1.32] | 1.11 [0.94-1.31] |
| Z76 | 1,810 | 45,502,160 | 0.04 | 0.04 | 0.03 | 0.00 | 0.01 | 0.00 | 1.19 [1.01-1.39] | 1.13 [0.95-1.33] |
| Z80 | 5,360 | 46,371,520 | 0.12 | 0.11 | 0.10 | 0.01 | 0.02 | 0.02 | 1.22 [1.11-1.34]* | 1.17 [1.06-1.29]* |
| Z81 | 9,030 | 46,231,030 | 0.22 | 0.19 | 0.19 | 0.03 | 0.03 | 0.02 | 1.13 [1.05-1.21]* | 1.09 [1.02-1.18] |
| Z82 | 41,460 | 45,416,680 | 1.24 | 0.83 | 0.79 | 0.41 | 0.44 | 0.41 | 1.56 [1.51-1.61]* | 1.50 [1.45-1.54]* |
| Z83 | 25,420 | 45,277,060 | 0.65 | 0.54 | 0.53 | 0.12 | 0.12 | 0.10 | 1.24 [1.19-1.29]* | 1.19 [1.14-1.24]* |
| Z84 | 7,180 | 46,195,140 | 0.25 | 0.13 | 0.13 | 0.12 | 0.12 | 0.12 | 1.96 [1.83-2.10]* | 1.88 [1.75-2.02]* |
| Z85 | 5,130 | 46,365,630 | 0.11 | 0.11 | 0.11 | 0.00 | 0.01 | 0.01 | 1.05 [0.96-1.16] | 1.06 [0.96-1.17] |
| Z86 | 88,100 | 45,725,420 | 1.97 | 1.92 | 1.78 | 0.05 | 0.18 | 0.16 | 1.10 [1.08-1.13]* | 1.09 [1.06-1.11]* |
| Z87 | 99,180 | 45,070,060 | 2.57 | 2.10 | 2.05 | 0.47 | 0.52 | 0.41 | 1.25 [1.23-1.28]* | 1.19 [1.17-1.22]* |
| Z88 | 69,650 | 45,689,740 | 1.95 | 1.41 | 1.34 | 0.55 | 0.61 | 0.49 | 1.46 [1.42-1.49]* | 1.34 [1.30-1.37]* |
| Z90 | 13,130 | 46,324,620 | 0.31 | 0.28 | 0.26 | 0.04 | 0.05 | 0.04 | 1.21 [1.14-1.28]* | 1.16 [1.09-1.23]* |
| Z91 | 74,680 | 45,741,780 | 2.78 | 1.32 | 1.30 | 1.46 | 1.48 | 1.28 | 2.13 [2.08-2.18]* | 1.86 [1.81-1.90]* |
| Z92 | 19,570 | 46,208,820 | 0.48 | 0.41 | 0.40 | 0.07 | 0.08 | 0.06 | 1.19 [1.13-1.25]* | 1.14 [1.08-1.19]* |
| Z93 | 5,700 | 46,310,300 | 0.12 | 0.12 | 0.12 | −0.01 | −0.01 | −0.01 | 0.96 [0.87-1.05] | 0.90 [0.82-0.99] |
| Z94 | 2,000 | 46,390,380 | 0.05 | 0.04 | 0.04 | 0.00 | 0.00 | 0.00 | 1.09 [0.94-1.26] | 1.06 [0.91-1.24] |
| Z95 | 4,930 | 46,356,700 | 0.10 | 0.11 | 0.11 | 0.00 | −0.01 | −0.01 | 0.95 [0.86-1.05] | 0.92 [0.83-1.02] |
| Z96 | 8,770 | 46,337,410 | 0.20 | 0.19 | 0.18 | 0.02 | 0.02 | 0.02 | 1.12 [1.04-1.20]* | 1.08 [1.01-1.17] |
| Z97 | 4,970 | 46,380,650 | 0.11 | 0.11 | 0.10 | 0.01 | 0.01 | 0.01 | 1.12 [1.01-1.23] | 1.07 [0.97-1.18] |
| Z98 | 4,700 | 46,335,100 | 0.10 | 0.10 | 0.10 | 0.00 | 0.01 | 0.00 | 1.08 [0.98-1.19] | 1.03 [0.93-1.14] |
| Z99 | 4,940 | 46,357,320 | 0.10 | 0.11 | 0.11 | 0.00 | −0.01 | −0.01 | 0.94 [0.86-1.04] | 0.91 [0.82-1.01] |
| hospitalised - A00-B99 | | | | | | | | | | |
| A02 | 1,530 | 81,557,590 | 0.02 | 0.02 | 0.02 | 0.00 | 0.00 | 0.00 | 1.11 [0.93-1.31] | 1.05 [0.89-1.25] |
| A04 | 46,880 | 80,951,040 | 0.62 | 0.57 | 0.52 | 0.05 | 0.10 | 0.08 | 1.19 [1.16-1.23]* | 1.15 [1.12-1.19]* |
| A05 | 490 | 81,603,180 | 0.01 | 0.01 | 0.01 | 0.00 | 0.00 | 0.00 | 1.19 [0.87-1.62] | 1.20 [0.87-1.64] |
| A07 | 860 | 81,590,390 | 0.01 | 0.01 | 0.01 | 0.00 | 0.00 | 0.00 | 1.17 [0.93-1.47] | 1.14 [0.90-1.44] |
| A08 | 57,140 | 79,844,050 | 0.83 | 0.69 | 0.72 | 0.14 | 0.11 | 0.09 | 1.15 [1.12-1.18]* | 1.12 [1.09-1.16]* |
| A09 | 212,790 | 79,837,570 | 2.98 | 2.59 | 2.45 | 0.39 | 0.52 | 0.41 | 1.21 [1.20-1.23]* | 1.16 [1.14-1.18]* |
| A15 | 1,090 | 81,580,310 | 0.01 | 0.01 | 0.01 | 0.00 | 0.00 | 0.00 | 1.15 [0.94-1.42] | 1.12 [0.90-1.38] |
| A16 | 2,140 | 81,548,890 | 0.03 | 0.03 | 0.02 | 0.01 | 0.01 | 0.01 | 1.30 [1.13-1.50]* | 1.24 [1.07-1.44]* |
| A18 | 1,070 | 81,568,590 | 0.01 | 0.01 | 0.01 | 0.00 | 0.00 | 0.00 | 1.22 [1.00-1.50] | 1.18 [0.96-1.45] |
| A31 | 1,090 | 81,599,450 | 0.02 | 0.01 | 0.01 | 0.00 | 0.00 | 0.00 | 1.35 [1.11-1.65]* | 1.21 [0.99-1.48] |
| A38 | 1,840 | 81,585,520 | 0.03 | 0.02 | 0.03 | 0.01 | 0.00 | 0.00 | 1.03 [0.89-1.20] | 1.02 [0.87-1.19] |
| A39 | 1,850 | 81,503,120 | 0.03 | 0.02 | 0.02 | 0.00 | 0.00 | 0.00 | 1.05 [0.90-1.22] | 1.03 [0.88-1.21] |
| A40 | 9,810 | 81,514,230 | 0.13 | 0.12 | 0.11 | 0.02 | 0.03 | 0.03 | 1.26 [1.18-1.35]* | 1.24 [1.16-1.33]* |
| A41 | 184,770 | 80,642,010 | 2.47 | 2.25 | 2.07 | 0.23 | 0.40 | 0.38 | 1.19 [1.18-1.21]* | 1.18 [1.16-1.20]* |
| A46 | 580 | 81,589,290 | 0.01 | 0.01 | 0.01 | 0.00 | 0.01 | 0.01 | 1.97 [1.53-2.53]* | 1.93 [1.50-2.49]* |
| A48 | 870 | 81,594,320 | 0.01 | 0.01 | 0.01 | 0.00 | 0.00 | 0.00 | 1.23 [0.98-1.54] | 1.27 [1.01-1.60] |
| A49 | 26,570 | 81,284,930 | 0.36 | 0.32 | 0.29 | 0.05 | 0.08 | 0.07 | 1.27 [1.22-1.32]* | 1.24 [1.19-1.29]* |
| A63 | 620 | 81,506,810 | 0.01 | 0.01 | 0.01 | 0.00 | 0.00 | 0.00 | 1.45 [1.12-1.86]* | 1.38 [1.07-1.80] |
| A69 | 700 | 81,599,440 | 0.01 | 0.01 | 0.01 | 0.00 | 0.00 | 0.00 | 1.14 [0.89-1.47] | 1.07 [0.82-1.38] |
| A86 | 1,260 | 81,589,510 | 0.02 | 0.02 | 0.01 | 0.00 | 0.00 | 0.00 | 1.03 [0.84-1.25] | 0.99 [0.81-1.21] |
| A87 | 3,260 | 81,483,260 | 0.04 | 0.04 | 0.04 | 0.00 | 0.01 | 0.00 | 1.16 [1.03-1.30] | 1.09 [0.96-1.22] |
| B00 | 9,070 | 81,373,920 | 0.20 | 0.09 | 0.09 | 0.11 | 0.11 | 0.11 | 2.31 [2.17-2.46]* | 2.18 [2.05-2.33]* |
| B01 | 8,220 | 81,337,150 | 0.15 | 0.09 | 0.10 | 0.06 | 0.05 | 0.05 | 1.53 [1.44-1.64]* | 1.49 [1.40-1.60]* |
| B02 | 13,560 | 81,467,790 | 0.20 | 0.16 | 0.15 | 0.04 | 0.05 | 0.04 | 1.32 [1.24-1.40]* | 1.26 [1.19-1.33]* |
| B07 | 5,490 | 81,435,720 | 0.08 | 0.06 | 0.06 | 0.02 | 0.02 | 0.02 | 1.32 [1.21-1.45]* | 1.25 [1.14-1.37]* |
| B08 | 3,000 | 81,528,610 | 0.05 | 0.03 | 0.03 | 0.02 | 0.02 | 0.02 | 1.54 [1.38-1.72]* | 1.49 [1.33-1.67]* |
| B09 | 1,310 | 81,561,170 | 0.02 | 0.02 | 0.02 | 0.00 | 0.00 | 0.00 | 1.17 [0.98-1.40] | 1.16 [0.97-1.39] |
| B15 | 660 | 81,592,280 | 0.01 | 0.01 | 0.01 | 0.00 | 0.00 | 0.00 | 1.06 [0.82-1.39] | 1.03 [0.79-1.35] |
| B16 | 1,290 | 81,566,860 | 0.02 | 0.02 | 0.01 | 0.00 | 0.00 | 0.00 | 1.17 [0.97-1.41] | 1.17 [0.97-1.42] |
| B17 | 1,990 | 81,558,400 | 0.03 | 0.02 | 0.02 | 0.00 | 0.01 | 0.00 | 1.24 [1.07-1.44]* | 1.20 [1.03-1.40] |
| B18 | 8,230 | 81,428,980 | 0.10 | 0.10 | 0.09 | −0.01 | 0.01 | 0.01 | 1.07 [0.99-1.16] | 1.09 [1.01-1.18] |
| B19 | 470 | 81,599,640 | 0.01 | 0.01 | 0.01 | 0.00 | 0.00 | 0.00 | 1.18 [0.87-1.60] | 1.14 [0.84-1.57] |
| B25 | 1,690 | 81,585,200 | 0.02 | 0.02 | 0.02 | 0.00 | 0.00 | 0.00 | 1.23 [1.05-1.44] | 1.18 [1.01-1.40] |
| B26 | 500 | 81,595,840 | 0.01 | 0.01 | 0.01 | 0.00 | 0.00 | 0.00 | 1.13 [0.84-1.52] | 1.08 [0.79-1.46] |
| B27 | 4,620 | 81,485,940 | 0.07 | 0.05 | 0.06 | 0.01 | 0.01 | 0.00 | 1.08 [0.98-1.19] | 1.04 [0.94-1.15] |
| B30 | 620 | 81,596,460 | 0.01 | 0.01 | 0.01 | 0.00 | 0.00 | 0.00 | 1.16 [0.90-1.51] | 1.09 [0.83-1.43] |
| B34 | 124,310 | 77,872,300 | 2.32 | 1.42 | 1.56 | 0.90 | 0.76 | 0.70 | 1.49 [1.46-1.51]* | 1.44 [1.41-1.46]* |
| B35 | 5,770 | 81,534,900 | 0.10 | 0.06 | 0.06 | 0.03 | 0.04 | 0.03 | 1.65 [1.51-1.79]* | 1.55 [1.42-1.69]* |
| B36 | 2,230 | 81,590,440 | 0.04 | 0.02 | 0.02 | 0.01 | 0.01 | 0.01 | 1.68 [1.46-1.92]* | 1.60 [1.39-1.84]* |
| B37 | 77,600 | 80,744,150 | 1.08 | 0.93 | 0.86 | 0.15 | 0.22 | 0.17 | 1.26 [1.23-1.29]* | 1.19 [1.16-1.22]* |
| B44 | 3,100 | 81,579,850 | 0.05 | 0.04 | 0.03 | 0.01 | 0.02 | 0.01 | 1.51 [1.35-1.70]* | 1.27 [1.13-1.43]* |
| B49 | 1,810 | 81,594,820 | 0.03 | 0.02 | 0.02 | 0.01 | 0.01 | 0.01 | 1.51 [1.30-1.76]* | 1.45 [1.25-1.69]* |
| B50 | 690 | 81,574,190 | 0.01 | 0.01 | 0.01 | 0.00 | 0.00 | 0.00 | 0.83 [0.63-1.10] | 0.86 [0.65-1.14] |
| B59 | 920 | 81,603,630 | 0.01 | 0.01 | 0.01 | 0.00 | 0.00 | 0.00 | 1.36 [1.09-1.68]* | 1.30 [1.04-1.62] |
| B80 | 2,300 | 81,565,690 | 0.03 | 0.03 | 0.03 | 0.01 | 0.00 | 0.00 | 1.16 [1.01-1.33] | 1.11 [0.96-1.27] |
| B85 | 930 | 81,590,850 | 0.01 | 0.01 | 0.01 | 0.00 | 0.00 | 0.00 | 1.04 [0.83-1.30] | 1.03 [0.82-1.29] |
| B86 | 1,590 | 81,570,060 | 0.03 | 0.02 | 0.01 | 0.02 | 0.02 | 0.02 | 2.19 [1.87-2.55]* | 2.20 [1.88-2.58]* |
| B90 | 720 | 81,601,060 | 0.01 | 0.01 | 0.01 | 0.00 | 0.00 | 0.00 | 1.41 [1.11-1.81]* | 1.31 [1.02-1.68] |
| B91 | 1,070 | 81,590,030 | 0.01 | 0.01 | 0.01 | 0.00 | 0.00 | 0.00 | 1.14 [0.92-1.41] | 1.16 [0.94-1.44] |
| B94 | 1,940 | 81,584,660 | 0.03 | 0.02 | 0.02 | 0.00 | 0.01 | 0.00 | 1.24 [1.07-1.44]* | 1.18 [1.01-1.37] |
| B95 | 107,700 | 79,824,640 | 1.66 | 1.27 | 1.17 | 0.39 | 0.48 | 0.46 | 1.41 [1.38-1.44]* | 1.38 [1.35-1.41]* |
| B96 | 216,040 | 78,712,320 | 2.94 | 2.70 | 2.52 | 0.24 | 0.42 | 0.36 | 1.17 [1.15-1.18]* | 1.14 [1.12-1.16]* |
| B97 | 60,280 | 80,666,480 | 0.89 | 0.71 | 0.71 | 0.18 | 0.18 | 0.14 | 1.25 [1.22-1.29]* | 1.19 [1.16-1.23]* |
| B98 | 21,450 | 81,435,090 | 0.27 | 0.26 | 0.24 | 0.01 | 0.03 | 0.02 | 1.13 [1.08-1.18]* | 1.10 [1.04-1.15]* |
| B99 | 6,270 | 81,575,560 | 0.08 | 0.08 | 0.07 | 0.01 | 0.01 | 0.01 | 1.20 [1.10-1.32]* | 1.19 [1.09-1.31]* |
| hospitalised - C00-D49 | | | | | | | | | | |
| C00 | 700 | 81,598,920 | 0.01 | 0.01 | 0.01 | 0.00 | 0.00 | 0.00 | 1.04 [0.80-1.36] | 1.03 [0.79-1.35] |
| C01 | 1,420 | 81,596,880 | 0.02 | 0.02 | 0.01 | 0.00 | 0.00 | 0.00 | 1.12 [0.93-1.35] | 1.17 [0.97-1.41] |
| C02 | 2,190 | 81,580,900 | 0.03 | 0.03 | 0.02 | 0.00 | 0.01 | 0.01 | 1.30 [1.13-1.50]* | 1.31 [1.13-1.51]* |
| C03 | 510 | 81,605,650 | 0.01 | 0.01 | 0.01 | 0.00 | 0.00 | 0.00 | 1.32 [0.99-1.76] | 1.32 [0.98-1.77] |
| C04 | 690 | 81,600,140 | 0.01 | 0.01 | 0.01 | 0.00 | 0.00 | 0.00 | 1.10 [0.84-1.43] | 1.13 [0.86-1.48] |

| | | | | | | | | | | |
|---|---|---|---|---|---|---|---|---|---|---|
| C05 | 690 | 81,601,330 | 0.01 | 0.01 | 0.01 | 0.00 | 0.00 | 0.00 | 1.14 [0.89-1.47] | 1.20 [0.93-1.56] |
| C06 | 1,030 | 81,599,480 | 0.01 | 0.01 | 0.01 | 0.00 | 0.00 | 0.00 | 1.18 [0.96-1.46] | 1.19 [0.96-1.47] |
| C07 | 1,000 | 81,596,020 | 0.01 | 0.01 | 0.01 | 0.00 | 0.00 | 0.00 | 1.04 [0.83-1.31] | 1.03 [0.82-1.30] |
| C09 | 1,740 | 81,586,910 | 0.02 | 0.02 | 0.02 | 0.00 | 0.00 | 0.00 | 1.07 [0.90-1.26] | 1.08 [0.91-1.28] |
| C10 | 950 | 81,603,520 | 0.01 | 0.01 | 0.01 | 0.00 | 0.00 | 0.00 | 1.08 [0.86-1.36] | 1.09 [0.87-1.38] |
| C12 | 500 | 81,606,400 | 0.01 | 0.01 | 0.01 | 0.00 | 0.00 | 0.00 | 1.26 [0.93-1.70] | 1.33 [0.98-1.81] |
| C13 | 570 | 81,607,400 | 0.01 | 0.01 | 0.01 | 0.00 | 0.00 | 0.00 | 1.19 [0.89-1.58] | 1.30 [0.97-1.75] |
| C14 | 1,120 | 81,603,030 | 0.01 | 0.01 | 0.01 | 0.00 | 0.00 | 0.00 | 0.96 [0.77-1.18] | 0.97 [0.78-1.20] |
| C15 | 13,720 | 81,546,480 | 0.17 | 0.17 | 0.15 | 0.00 | 0.02 | 0.02 | 1.13 [1.07-1.20]* | 1.14 [1.07-1.21]* |
| C16 | 11,120 | 81,548,420 | 0.13 | 0.14 | 0.12 | −0.01 | 0.01 | 0.01 | 1.06 [0.99-1.14] | 1.06 [0.99-1.14] |
| C17 | 2,410 | 81,593,690 | 0.03 | 0.03 | 0.03 | 0.00 | 0.00 | 0.00 | 1.08 [0.94-1.24] | 1.07 [0.93-1.23] |
| C18 | 37,720 | 81,181,490 | 0.42 | 0.47 | 0.43 | −0.05 | 0.00 | 0.00 | 1.00 [0.96-1.04] | 1.00 [0.96-1.03] |
| C19 | 7,770 | 81,530,200 | 0.08 | 0.10 | 0.09 | −0.02 | −0.01 | 0.00 | 0.94 [0.87-1.02] | 0.95 [0.88-1.04] |
| C20 | 15,290 | 81,389,270 | 0.16 | 0.19 | 0.17 | −0.03 | −0.01 | −0.01 | 0.95 [0.90-1.01] | 0.97 [0.91-1.03] |
| C21 | 2,100 | 81,583,730 | 0.02 | 0.03 | 0.02 | 0.00 | 0.00 | 0.00 | 1.04 [0.89-1.21] | 1.04 [0.89-1.21] |
| C22 | 8,170 | 81,586,170 | 0.11 | 0.10 | 0.09 | 0.01 | 0.02 | 0.02 | 1.18 [1.09-1.27]* | 1.17 [1.08-1.26]* |
| C23 | 1,100 | 81,607,650 | 0.01 | 0.01 | 0.01 | 0.00 | 0.00 | 0.00 | 1.03 [0.83-1.28] | 1.04 [0.84-1.29] |
| C24 | 2,140 | 81,600,390 | 0.02 | 0.03 | 0.02 | 0.00 | 0.00 | 0.00 | 1.04 [0.89-1.22] | 1.03 [0.88-1.20] |
| C25 | 12,960 | 81,580,950 | 0.16 | 0.16 | 0.14 | 0.00 | 0.02 | 0.02 | 1.13 [1.07-1.20]* | 1.13 [1.07-1.21]* |
| C26 | 3,170 | 81,594,500 | 0.04 | 0.04 | 0.04 | 0.00 | 0.00 | 0.00 | 1.06 [0.93-1.20] | 1.05 [0.92-1.19] |
| C32 | 3,070 | 81,566,270 | 0.04 | 0.04 | 0.03 | 0.00 | 0.00 | 0.00 | 1.08 [0.95-1.22] | 1.11 [0.98-1.26] |
| C34 | 56,780 | 81,413,950 | 0.70 | 0.70 | 0.63 | 0.00 | 0.07 | 0.07 | 1.11 [1.08-1.14]* | 1.12 [1.09-1.15]* |
| C38 | 780 | 81,606,300 | 0.01 | 0.01 | 0.01 | 0.00 | 0.00 | 0.00 | 1.02 [0.80-1.32] | 1.04 [0.80-1.34] |
| C40 | 550 | 81,600,130 | 0.01 | 0.01 | 0.01 | 0.00 | 0.00 | 0.00 | 1.02 [0.75-1.37] | 1.04 [0.77-1.41] |
| C41 | 1,600 | 81,592,870 | 0.02 | 0.02 | 0.02 | 0.00 | 0.00 | 0.00 | 1.18 [1.00-1.40] | 1.19 [1.00-1.41] |
| C43 | 15,430 | 81,375,510 | 0.17 | 0.20 | 0.17 | −0.03 | −0.01 | −0.01 | 0.97 [0.91-1.02] | 0.96 [0.90-1.02] |
| C44 | 110,000 | 79,937,130 | 1.34 | 1.39 | 1.27 | −0.05 | 0.07 | 0.04 | 1.06 [1.03-1.08]* | 1.03 [1.01-1.06]* |
| C45 | 3,400 | 81,601,410 | 0.04 | 0.04 | 0.04 | 0.00 | 0.00 | 0.00 | 1.02 [0.91-1.16] | 1.04 [0.92-1.18] |
| C48 | 1,890 | 81,598,880 | 0.02 | 0.02 | 0.02 | 0.00 | 0.00 | 0.00 | 1.04 [0.89-1.22] | 1.03 [0.88-1.21] |
| C49 | 3,580 | 81,565,410 | 0.04 | 0.04 | 0.04 | 0.00 | 0.00 | 0.00 | 1.10 [0.98-1.24] | 1.09 [0.97-1.23] |
| C50 | 61,740 | 80,280,890 | 0.72 | 0.78 | 0.70 | −0.06 | 0.02 | 0.02 | 1.02 [0.99-1.05] | 1.03 [1.00-1.06] |
| C51 | 1,730 | 81,584,860 | 0.02 | 0.02 | 0.02 | 0.00 | 0.00 | 0.00 | 1.21 [1.03-1.43] | 1.20 [1.02-1.42] |
| C52 | 590 | 81,605,690 | 0.01 | 0.01 | 0.01 | 0.00 | 0.00 | 0.00 | 0.93 [0.69-1.24] | 0.93 [0.69-1.25] |
| C53 | 3,410 | 81,539,120 | 0.04 | 0.04 | 0.04 | 0.00 | 0.00 | 0.00 | 1.02 [0.90-1.15] | 1.05 [0.93-1.19] |
| C54 | 9,850 | 81,431,870 | 0.11 | 0.12 | 0.11 | −0.01 | 0.00 | 0.00 | 1.03 [0.96-1.10] | 1.04 [0.97-1.12] |
| C55 | 1,470 | 81,594,570 | 0.02 | 0.02 | 0.02 | 0.00 | 0.00 | 0.00 | 0.96 [0.80-1.15] | 0.95 [0.79-1.15] |
| C56 | 9,020 | 81,499,100 | 0.10 | 0.11 | 0.10 | −0.01 | 0.00 | 0.00 | 1.00 [0.93-1.08] | 1.01 [0.94-1.09] |
| C57 | 1,070 | 81,603,150 | 0.01 | 0.01 | 0.01 | 0.00 | 0.00 | 0.00 | 1.15 [0.94-1.42] | 1.17 [0.95-1.44] |
| C60 | 810 | 81,597,070 | 0.01 | 0.01 | 0.01 | 0.00 | 0.00 | 0.00 | 1.17 [0.92-1.49] | 1.21 [0.94-1.55] |
| C61 | 61,890 | 80,881,010 | 0.72 | 0.78 | 0.71 | −0.06 | 0.00 | 0.00 | 1.01 [0.98-1.04] | 1.00 [0.97-1.03] |
| C62 | 1,320 | 81,554,190 | 0.02 | 0.02 | 0.01 | 0.00 | 0.00 | 0.00 | 1.02 [0.84-1.24] | 1.03 [0.84-1.25] |
| C64 | 13,300 | 81,477,050 | 0.16 | 0.17 | 0.15 | −0.01 | 0.01 | 0.01 | 1.06 [0.99-1.12] | 1.05 [0.99-1.12] |
| C65 | 1,240 | 81,598,470 | 0.01 | 0.02 | 0.01 | 0.00 | 0.00 | 0.00 | 1.11 [0.91-1.36] | 1.15 [0.94-1.41] |
| C66 | 1,750 | 81,595,990 | 0.02 | 0.02 | 0.02 | 0.00 | 0.00 | 0.00 | 0.99 [0.83-1.17] | 1.00 [0.84-1.18] |
| C67 | 28,770 | 81,173,620 | 0.34 | 0.36 | 0.33 | −0.02 | 0.01 | 0.01 | 1.03 [0.99-1.07] | 1.03 [0.99-1.08] |
| C68 | 700 | 81,604,130 | 0.01 | 0.01 | 0.01 | 0.00 | 0.00 | 0.00 | 0.94 [0.71-1.23] | 0.95 [0.72-1.26] |
| C69 | 1,210 | 81,583,290 | 0.01 | 0.02 | 0.01 | 0.00 | 0.00 | 0.00 | 0.96 [0.79-1.18] | 0.97 [0.79-1.19] |
| C71 | 6,810 | 81,552,500 | 0.08 | 0.08 | 0.08 | 0.00 | 0.01 | 0.01 | 1.09 [1.00-1.19] | 1.09 [1.00-1.18] |
| C73 | 3,670 | 81,538,910 | 0.04 | 0.05 | 0.04 | 0.00 | 0.00 | 0.00 | 1.02 [0.91-1.15] | 1.01 [0.90-1.14] |
| C74 | 620 | 81,601,690 | 0.01 | 0.01 | 0.01 | 0.00 | 0.00 | 0.00 | 1.09 [0.83-1.42] | 1.08 [0.82-1.42] |
| C75 | 660 | 81,602,410 | 0.01 | 0.01 | 0.01 | 0.00 | 0.00 | 0.00 | 0.99 [0.75-1.30] | 1.00 [0.76-1.32] |
| C76 | 2,110 | 81,590,460 | 0.03 | 0.03 | 0.02 | 0.00 | 0.00 | 0.00 | 1.09 [0.93-1.26] | 1.09 [0.94-1.28] |
| C77 | 64,290 | 81,010,120 | 0.74 | 0.81 | 0.72 | −0.06 | 0.03 | 0.04 | 1.04 [1.01-1.07]* | 1.05 [1.02-1.08]* |
| C78 | 97,790 | 81,367,710 | 1.14 | 1.22 | 1.09 | −0.08 | 0.05 | 0.07 | 1.05 [1.02-1.07]* | 1.07 [1.04-1.09]* |
| C79 | 84,090 | 81,397,000 | 0.98 | 1.05 | 0.93 | −0.06 | 0.05 | 0.07 | 1.05 [1.03-1.08]* | 1.08 [1.05-1.11]* |
| C80 | 27,990 | 81,516,900 | 0.33 | 0.35 | 0.31 | −0.01 | 0.03 | 0.03 | 1.09 [1.04-1.13]* | 1.09 [1.05-1.14]* |
| C81 | 2,800 | 81,539,400 | 0.04 | 0.03 | 0.03 | 0.01 | 0.01 | 0.01 | 1.46 [1.30-1.65]* | 1.43 [1.26-1.61]* |
| C82 | 3,520 | 81,555,970 | 0.04 | 0.04 | 0.04 | 0.00 | 0.00 | 0.00 | 1.04 [0.93-1.17] | 1.02 [0.90-1.15] |
| C83 | 10,050 | 81,512,390 | 0.13 | 0.12 | 0.11 | 0.01 | 0.02 | 0.02 | 1.19 [1.11-1.27]* | 1.17 [1.09-1.25]* |
| C84 | 1,330 | 81,592,440 | 0.03 | 0.01 | 0.01 | 0.01 | 0.01 | 0.01 | 2.15 [1.83-2.53]* | 2.17 [1.84-2.57]* |
| C85 | 12,520 | 81,430,380 | 0.16 | 0.15 | 0.13 | 0.01 | 0.03 | 0.03 | 1.23 [1.16-1.30]* | 1.21 [1.13-1.28]* |
| C88 | 1,760 | 81,596,430 | 0.02 | 0.02 | 0.02 | 0.00 | 0.00 | 0.00 | 1.14 [0.96-1.34] | 1.12 [0.95-1.33] |
| C90 | 8,640 | 81,537,510 | 0.10 | 0.11 | 0.10 | 0.00 | 0.01 | 0.01 | 1.10 [1.02-1.19] | 1.08 [1.00-1.17] |
| C91 | 9,190 | 81,491,170 | 0.11 | 0.11 | 0.11 | 0.00 | 0.00 | 0.00 | 1.04 [0.97-1.12] | 1.03 [0.95-1.11] |
| C92 | 6,960 | 81,553,870 | 0.09 | 0.08 | 0.07 | 0.01 | 0.02 | 0.02 | 1.24 [1.15-1.35]* | 1.24 [1.14-1.35]* |
| C93 | 980 | 81,607,480 | 0.01 | 0.01 | 0.01 | 0.00 | 0.00 | 0.00 | 1.11 [0.89-1.40] | 1.15 [0.91-1.44] |
| C95 | 1,690 | 81,596,680 | 0.02 | 0.02 | 0.02 | 0.00 | 0.00 | 0.00 | 1.17 [0.99-1.38] | 1.16 [0.98-1.37] |
| C97 | 8,170 | 81,583,760 | 0.09 | 0.10 | 0.09 | −0.01 | 0.00 | 0.00 | 1.02 [0.94-1.10] | 1.01 [0.93-1.09] |
| D00 | 900 | 81,600,100 | 0.01 | 0.01 | 0.01 | 0.00 | 0.00 | 0.00 | 1.02 [0.81-1.29] | 1.01 [0.80-1.28] |
| D01 | 900 | 81,598,370 | 0.01 | 0.01 | 0.01 | 0.00 | 0.00 | 0.00 | 1.00 [0.79-1.27] | 0.96 [0.76-1.22] |
| D02 | 890 | 81,599,930 | 0.01 | 0.01 | 0.01 | 0.00 | 0.00 | 0.00 | 1.14 [0.91-1.43] | 1.13 [0.90-1.43] |
| D03 | 5,970 | 81,516,900 | 0.07 | 0.07 | 0.07 | −0.01 | 0.00 | 0.00 | 1.02 [0.93-1.12] | 1.00 [0.91-1.09] |
| D04 | 13,490 | 81,458,650 | 0.18 | 0.16 | 0.15 | 0.01 | 0.03 | 0.02 | 1.20 [1.13-1.28]* | 1.16 [1.09-1.23]* |
| D05 | 10,490 | 81,358,630 | 0.11 | 0.13 | 0.12 | −0.02 | 0.00 | 0.00 | 0.96 [0.90-1.03] | 0.96 [0.89-1.03] |
| D06 | 9,040 | 81,199,100 | 0.10 | 0.11 | 0.10 | −0.01 | 0.00 | 0.00 | 0.99 [0.92-1.07] | 1.00 [0.93-1.08] |
| D07 | 3,510 | 81,538,570 | 0.04 | 0.04 | 0.04 | 0.00 | 0.01 | 0.00 | 1.13 [1.01-1.27] | 1.10 [0.98-1.24] |
| D09 | 3,380 | 81,570,810 | 0.04 | 0.04 | 0.04 | 0.00 | 0.00 | 0.00 | 1.05 [0.93-1.19] | 1.05 [0.93-1.19] |
| D10 | 3,320 | 81,512,670 | 0.05 | 0.04 | 0.04 | 0.01 | 0.01 | 0.01 | 1.25 [1.12-1.40]* | 1.20 [1.07-1.35]* |
| D11 | 3,690 | 81,518,610 | 0.04 | 0.05 | 0.04 | 0.00 | 0.00 | 0.00 | 1.02 [0.91-1.15] | 1.02 [0.91-1.15] |
| D12 | 111,980 | 79,893,120 | 1.40 | 1.40 | 1.24 | 0.00 | 0.17 | 0.12 | 1.13 [1.11-1.16]* | 1.09 [1.07-1.12]* |
| D13 | 7,610 | 81,376,500 | 0.09 | 0.09 | 0.08 | 0.00 | 0.01 | 0.00 | 1.10 [1.02-1.19] | 1.04 [0.96-1.13] |
| D14 | 2,380 | 81,545,640 | 0.03 | 0.03 | 0.03 | 0.00 | 0.00 | 0.00 | 1.16 [1.01-1.33] | 1.12 [0.97-1.29] |
| D15 | 940 | 81,596,760 | 0.01 | 0.01 | 0.01 | 0.00 | 0.00 | 0.00 | 1.07 [0.85-1.34] | 1.06 [0.84-1.33] |
| D16 | 3,980 | 81,505,160 | 0.05 | 0.05 | 0.05 | 0.01 | 0.00 | 0.00 | 1.08 [0.98-1.21] | 1.05 [0.94-1.17] |
| D17 | 32,970 | 80,452,790 | 0.39 | 0.41 | 0.36 | −0.02 | 0.03 | 0.01 | 1.07 [1.03-1.11]* | 1.03 [0.99-1.07] |
| D18 | 16,030 | 81,207,280 | 0.20 | 0.20 | 0.18 | 0.01 | 0.02 | 0.01 | 1.11 [1.05-1.17]* | 1.07 [1.01-1.13] |
| D21 | 3,680 | 81,509,190 | 0.05 | 0.04 | 0.04 | 0.00 | 0.01 | 0.01 | 1.21 [1.08-1.35]* | 1.16 [1.04-1.30]* |
| D22 | 31,790 | 80,076,170 | 0.40 | 0.40 | 0.37 | 0.00 | 0.03 | 0.02 | 1.07 [1.03-1.12]* | 1.04 [1.00-1.08] |
| D23 | 21,570 | 80,752,420 | 0.28 | 0.26 | 0.25 | 0.01 | 0.03 | 0.02 | 1.13 [1.08-1.19]* | 1.09 [1.04-1.15]* |
| D24 | 10,600 | 81,228,790 | 0.12 | 0.13 | 0.12 | −0.01 | 0.00 | 0.00 | 0.99 [0.93-1.06] | 0.97 [0.90-1.04] |
| D25 | 51,800 | 79,878,700 | 0.62 | 0.65 | 0.59 | −0.03 | 0.03 | 0.02 | 1.06 [1.03-1.09]* | 1.03 [1.00-1.06] |
| D26 | 1,800 | 81,543,660 | 0.02 | 0.02 | 0.02 | 0.00 | 0.00 | 0.00 | 1.09 [0.93-1.29] | 1.07 [0.90-1.26] |
| D27 | 12,610 | 81,291,980 | 0.15 | 0.16 | 0.14 | −0.01 | 0.00 | 0.00 | 1.03 [0.96-1.09] | 1.01 [0.95-1.07] |
| D28 | 1,400 | 81,565,610 | 0.02 | 0.02 | 0.01 | 0.00 | 0.00 | 0.00 | 1.20 [1.00-1.44] | 1.17 [0.97-1.40] |
| D29 | 1,100 | 81,585,520 | 0.01 | 0.01 | 0.01 | 0.00 | 0.00 | 0.00 | 1.17 [0.95-1.44] | 1.14 [0.92-1.40] |
| D30 | 2,160 | 81,577,330 | 0.02 | 0.03 | 0.02 | 0.00 | 0.00 | 0.00 | 1.03 [0.88-1.20] | 1.01 [0.87-1.17] |
| D31 | 2,160 | 81,570,730 | 0.03 | 0.03 | 0.02 | 0.01 | 0.01 | 0.01 | 1.28 [1.11-1.48]* | 1.26 [1.09-1.46]* |
| D32 | 7,880 | 81,523,740 | 0.09 | 0.10 | 0.09 | 0.00 | 0.01 | 0.01 | 1.09 [1.01-1.18] | 1.07 [0.99-1.17] |
| D33 | 3,070 | 81,555,970 | 0.04 | 0.04 | 0.04 | 0.00 | 0.00 | 0.00 | 1.03 [0.91-1.17] | 0.99 [0.87-1.12] |
| D34 | 1,990 | 81,562,410 | 0.02 | 0.02 | 0.02 | 0.00 | 0.00 | 0.00 | 1.06 [0.91-1.24] | 1.01 [0.86-1.19] |
| D35 | 11,720 | 81,442,730 | 0.13 | 0.15 | 0.13 | −0.01 | 0.00 | 0.00 | 1.04 [0.97-1.11] | 1.01 [0.95-1.08] |
| D36 | 5,360 | 81,460,900 | 0.07 | 0.07 | 0.06 | 0.00 | 0.01 | 0.01 | 1.12 [1.02-1.23] | 1.08 [0.98-1.19] |
| D37 | 14,600 | 81,421,620 | 0.17 | 0.18 | 0.16 | −0.01 | 0.02 | 0.01 | 1.09 [1.03-1.16]* | 1.09 [1.03-1.15]* |
| D38 | 3,480 | 81,577,490 | 0.04 | 0.04 | 0.04 | 0.00 | 0.01 | 0.01 | 1.15 [1.03-1.29] | 1.17 [1.04-1.31]* |
| D39 | 1,960 | 81,564,350 | 0.02 | 0.02 | 0.02 | 0.00 | 0.00 | 0.00 | 1.09 [0.93-1.28] | 1.12 [0.95-1.31] |
| D40 | 670 | 81,592,810 | 0.01 | 0.01 | 0.01 | 0.00 | 0.00 | 0.00 | 1.13 [0.86-1.47] | 1.11 [0.85-1.46] |
| D41 | 10,510 | 81,461,220 | 0.12 | 0.13 | 0.12 | −0.01 | 0.00 | 0.00 | 1.02 [0.95-1.09] | 1.02 [0.95-1.09] |
| D43 | 3,550 | 81,572,440 | 0.04 | 0.04 | 0.04 | 0.00 | 0.00 | 0.00 | 1.09 [0.97-1.22] | 1.08 [0.96-1.22] |

| | | | | | | | | | | |
|---|---|---|---|---|---|---|---|---|---|---|
| D44 | 2,210 | 81,568,090 | 0.03 | 0.03 | 0.03 | 0.00 | 0.00 | 0.00 | 1.08 [0.94-1.26] | 1.06 [0.91-1.23] |
| D45 | 3,850 | 81,531,490 | 0.04 | 0.05 | 0.04 | 0.00 | 0.00 | 0.00 | 1.06 [0.95-1.19] | 1.04 [0.93-1.17] |
| D46 | 9,430 | 81,553,650 | 0.13 | 0.11 | 0.10 | 0.02 | 0.03 | 0.03 | 1.26 [1.17-1.35]* | 1.24 [1.16-1.34]* |
| D47 | 15,950 | 81,504,200 | 0.21 | 0.19 | 0.18 | 0.02 | 0.03 | 0.03 | 1.19 [1.13-1.26]* | 1.16 [1.09-1.22]* |
| D48 | 7,410 | 81,439,490 | 0.09 | 0.09 | 0.08 | 0.00 | 0.00 | 0.00 | 1.03 [0.95-1.11] | 1.01 [0.93-1.09] |
| hospitalised - D50-D89 | | | | | | | | | | |
| D50 | 215,590 | 79,258,590 | 3.05 | 2.64 | 2.43 | 0.41 | 0.62 | 0.52 | 1.25 [1.23-1.27]* | 1.20 [1.19-1.22]* |
| D51 | 19,160 | 81,384,470 | 0.26 | 0.23 | 0.20 | 0.03 | 0.06 | 0.05 | 1.29 [1.23-1.35]* | 1.23 [1.17-1.29]* |
| D52 | 14,790 | 81,528,460 | 0.20 | 0.18 | 0.16 | 0.02 | 0.04 | 0.04 | 1.25 [1.18-1.32]* | 1.24 [1.17-1.31]* |
| D53 | 4,040 | 81,578,680 | 0.06 | 0.05 | 0.04 | 0.01 | 0.01 | 0.01 | 1.35 [1.21-1.50]* | 1.30 [1.17-1.45]* |
| D55 | 620 | 81,597,900 | 0.01 | 0.01 | 0.01 | 0.00 | 0.00 | 0.00 | 1.40 [1.08-1.82] | 1.41 [1.08-1.83] |
| D56 | 7,820 | 81,445,640 | 0.11 | 0.09 | 0.09 | 0.02 | 0.02 | 0.02 | 1.27 [1.18-1.37]* | 1.25 [1.16-1.35]* |
| D57 | 8,570 | 81,368,410 | 0.12 | 0.10 | 0.10 | 0.02 | 0.02 | 0.02 | 1.22 [1.13-1.31]* | 1.21 [1.12-1.30]* |
| D58 | 2,230 | 81,567,830 | 0.03 | 0.03 | 0.02 | 0.00 | 0.00 | 0.00 | 1.17 [1.01-1.35] | 1.17 [1.01-1.35] |
| D59 | 2,730 | 81,568,660 | 0.04 | 0.03 | 0.03 | 0.01 | 0.01 | 0.01 | 1.36 [1.20-1.54]* | 1.36 [1.20-1.55]* |
| D61 | 13,800 | 81,526,800 | 0.19 | 0.17 | 0.15 | 0.02 | 0.04 | 0.04 | 1.25 [1.18-1.32]* | 1.24 [1.17-1.31]* |
| D62 | 5,860 | 81,472,720 | 0.07 | 0.07 | 0.07 | 0.00 | 0.01 | 0.01 | 1.07 [0.98-1.18] | 1.08 [0.99-1.19] |
| D63 | 20,860 | 81,470,620 | 0.28 | 0.25 | 0.23 | 0.02 | 0.05 | 0.05 | 1.20 [1.15-1.26]* | 1.20 [1.14-1.26]* |
| D64 | 282,260 | 78,408,510 | 3.97 | 3.51 | 3.24 | 0.47 | 0.74 | 0.66 | 1.23 [1.21-1.24]* | 1.20 [1.18-1.21]* |
| D65 | 940 | 81,604,210 | 0.01 | 0.01 | 0.01 | 0.00 | 0.00 | 0.00 | 1.16 [0.93-1.45] | 1.17 [0.93-1.46] |
| D66 | 960 | 81,581,630 | 0.01 | 0.01 | 0.01 | 0.00 | 0.00 | 0.00 | 1.20 [0.97-1.50] | 1.17 [0.94-1.47] |
| D68 | 16,700 | 81,332,350 | 0.22 | 0.20 | 0.18 | 0.02 | 0.03 | 0.02 | 1.19 [1.13-1.25]* | 1.13 [1.07-1.19]* |
| D69 | 48,690 | 81,047,650 | 0.65 | 0.59 | 0.56 | 0.07 | 0.10 | 0.09 | 1.18 [1.14-1.21]* | 1.17 [1.13-1.20]* |
| D70 | 40,740 | 81,194,780 | 0.53 | 0.49 | 0.45 | 0.04 | 0.08 | 0.08 | 1.18 [1.14-1.22]* | 1.17 [1.13-1.21]* |
| D72 | 9,300 | 81,527,550 | 0.14 | 0.11 | 0.10 | 0.03 | 0.04 | 0.03 | 1.35 [1.26-1.45]* | 1.26 [1.17-1.35]* |
| D73 | 9,950 | 81,507,860 | 0.13 | 0.12 | 0.11 | 0.01 | 0.02 | 0.02 | 1.16 [1.08-1.24]* | 1.14 [1.07-1.23]* |
| D75 | 11,780 | 81,468,040 | 0.15 | 0.14 | 0.13 | 0.01 | 0.02 | 0.02 | 1.16 [1.09-1.24]* | 1.15 [1.07-1.22]* |
| D76 | 790 | 81,588,920 | 0.01 | 0.01 | 0.01 | 0.00 | 0.00 | 0.00 | 1.28 [1.01-1.61] | 1.21 [0.96-1.54] |
| D80 | 2,940 | 81,567,680 | 0.05 | 0.03 | 0.03 | 0.01 | 0.01 | 0.01 | 1.44 [1.28-1.62]* | 1.30 [1.15-1.47]* |
| D82 | 720 | 81,595,290 | 0.01 | 0.01 | 0.01 | 0.00 | 0.00 | 0.00 | 1.27 [1.00-1.62] | 1.19 [0.92-1.53] |
| D83 | 390 | 81,603,620 | 0.01 | 0.00 | 0.00 | 0.00 | 0.00 | 0.00 | 1.74 [1.28-2.37]* | 1.55 [1.12-2.13]* |
| D84 | 1,580 | 81,586,570 | 0.03 | 0.02 | 0.02 | 0.01 | 0.01 | 0.01 | 1.55 [1.32-1.81]* | 1.33 [1.13-1.56]* |
| D86 | 5,880 | 81,501,790 | 0.09 | 0.07 | 0.06 | 0.02 | 0.03 | 0.02 | 1.43 [1.32-1.56]* | 1.34 [1.23-1.46]* |
| D89 | 4,920 | 81,558,150 | 0.07 | 0.06 | 0.05 | 0.01 | 0.02 | 0.01 | 1.32 [1.20-1.45]* | 1.26 [1.14-1.38]* |
| hospitalised - E00-E89 | | | | | | | | | | |
| E02 | 1,770 | 81,600,700 | 0.02 | 0.02 | 0.02 | 0.00 | 0.00 | 0.00 | 1.11 [0.93-1.31] | 1.11 [0.93-1.31] |
| E03 | 188,000 | 78,608,480 | 2.65 | 2.33 | 2.10 | 0.32 | 0.55 | 0.47 | 1.26 [1.24-1.28]* | 1.22 [1.20-1.24]* |
| E04 | 20,750 | 81,261,940 | 0.25 | 0.26 | 0.23 | 0.00 | 0.02 | 0.02 | 1.09 [1.04-1.15]* | 1.06 [1.01-1.12] |
| E05 | 34,380 | 81,111,240 | 0.45 | 0.42 | 0.38 | 0.04 | 0.08 | 0.06 | 1.21 [1.17-1.25]* | 1.16 [1.12-1.21]* |
| E06 | 3,700 | 81,547,400 | 0.05 | 0.04 | 0.04 | 0.00 | 0.01 | 0.01 | 1.20 [1.07-1.34]* | 1.14 [1.02-1.28] |
| E07 | 6,000 | 81,529,610 | 0.08 | 0.07 | 0.07 | 0.01 | 0.01 | 0.01 | 1.18 [1.08-1.29]* | 1.11 [1.01-1.21] |
| E10 | 41,150 | 80,467,140 | 0.54 | 0.50 | 0.47 | 0.04 | 0.07 | 0.05 | 1.14 [1.11-1.18]* | 1.11 [1.07-1.15]* |
| E11 | 275,400 | 76,698,820 | 3.84 | 3.53 | 3.21 | 0.31 | 0.64 | 0.53 | 1.20 [1.18-1.21]* | 1.16 [1.15-1.18]* |
| E13 | 4,460 | 81,572,120 | 0.06 | 0.05 | 0.05 | 0.01 | 0.01 | 0.01 | 1.29 [1.16-1.42]* | 1.19 [1.08-1.32]* |
| E14 | 45,900 | 80,960,140 | 0.63 | 0.55 | 0.49 | 0.07 | 0.13 | 0.11 | 1.27 [1.23-1.31]* | 1.21 [1.17-1.25]* |
| E16 | 56,760 | 81,191,820 | 0.75 | 0.69 | 0.64 | 0.06 | 0.10 | 0.09 | 1.16 [1.13-1.20]* | 1.14 [1.10-1.17]* |
| E20 | 1,520 | 81,593,770 | 0.02 | 0.02 | 0.02 | 0.00 | 0.01 | 0.00 | 1.32 [1.11-1.56]* | 1.23 [1.03-1.46] |
| E21 | 16,730 | 81,461,500 | 0.20 | 0.21 | 0.19 | 0.00 | 0.02 | 0.01 | 1.08 [1.03-1.15]* | 1.06 [1.00-1.12] |
| E22 | 10,920 | 81,510,590 | 0.16 | 0.13 | 0.12 | 0.04 | 0.05 | 0.04 | 1.39 [1.30-1.48]* | 1.34 [1.26-1.43]* |
| E23 | 6,370 | 81,498,450 | 0.09 | 0.07 | 0.07 | 0.02 | 0.02 | 0.02 | 1.25 [1.15-1.36]* | 1.20 [1.10-1.30]* |
| E24 | 1,360 | 81,585,690 | 0.02 | 0.02 | 0.01 | 0.00 | 0.01 | 0.00 | 1.40 [1.17-1.67]* | 1.21 [1.01-1.45] |
| E26 | 720 | 81,596,600 | 0.01 | 0.01 | 0.01 | 0.00 | 0.00 | 0.00 | 1.16 [0.90-1.50] | 1.14 [0.88-1.48] |
| E27 | 12,390 | 81,486,410 | 0.18 | 0.14 | 0.13 | 0.04 | 0.05 | 0.04 | 1.37 [1.29-1.45]* | 1.27 [1.19-1.35]* |
| E28 | 27,200 | 81,137,860 | 0.36 | 0.33 | 0.31 | 0.04 | 0.05 | 0.04 | 1.18 [1.13-1.23]* | 1.11 [1.06-1.16]* |
| E29 | 1,790 | 81,584,470 | 0.03 | 0.02 | 0.02 | 0.01 | 0.01 | 0.01 | 1.45 [1.25-1.69]* | 1.32 [1.13-1.55]* |
| E30 | 2,720 | 81,563,210 | 0.05 | 0.03 | 0.04 | 0.01 | 0.01 | 0.01 | 1.15 [1.02-1.30] | 1.13 [0.99-1.28] |
| E34 | 5,390 | 81,514,690 | 0.08 | 0.06 | 0.07 | 0.02 | 0.01 | 0.01 | 1.16 [1.06-1.27]* | 1.11 [1.02-1.22] |
| E41 | 580 | 81,608,220 | 0.01 | 0.01 | 0.01 | 0.00 | 0.00 | 0.00 | 1.15 [0.86-1.55] | 1.20 [0.89-1.61] |
| E43 | 750 | 81,606,700 | 0.01 | 0.01 | 0.01 | 0.00 | 0.00 | 0.00 | 1.11 [0.86-1.43] | 1.13 [0.87-1.47] |
| E46 | 9,190 | 81,561,900 | 0.12 | 0.11 | 0.10 | 0.00 | 0.02 | 0.02 | 1.16 [1.08-1.25]* | 1.17 [1.08-1.26]* |
| E51 | 900 | 81,600,720 | 0.01 | 0.01 | 0.01 | 0.00 | 0.00 | 0.00 | 1.36 [1.09-1.70]* | 1.38 [1.10-1.73]* |
| E53 | 52,200 | 81,301,980 | 0.71 | 0.63 | 0.57 | 0.09 | 0.14 | 0.12 | 1.24 [1.20-1.28]* | 1.19 [1.16-1.23]* |
| E55 | 72,780 | 81,242,350 | 1.05 | 0.86 | 0.81 | 0.20 | 0.24 | 0.19 | 1.30 [1.26-1.33]* | 1.23 [1.19-1.26]* |
| E56 | 760 | 81,604,260 | 0.01 | 0.01 | 0.01 | 0.00 | 0.00 | 0.00 | 1.21 [0.94-1.54] | 1.18 [0.92-1.52] |
| E58 | 530 | 81,605,810 | 0.01 | 0.01 | 0.01 | 0.00 | 0.00 | 0.00 | 1.55 [1.17-2.06]* | 1.50 [1.13-1.99]* |
| E61 | 11,430 | 81,531,550 | 0.16 | 0.13 | 0.13 | 0.03 | 0.03 | 0.03 | 1.27 [1.19-1.35]* | 1.19 [1.12-1.27]* |
| E63 | 700 | 81,604,840 | 0.01 | 0.01 | 0.01 | 0.00 | 0.00 | 0.00 | 1.23 [0.95-1.59] | 1.18 [0.91-1.53] |
| E66 | 358,810 | 77,812,920 | 5.01 | 4.51 | 4.09 | 0.50 | 0.92 | 0.67 | 1.22 [1.21-1.24]* | 1.15 [1.14-1.17]* |
| E70 | 420 | 81,595,160 | 0.01 | 0.00 | 0.01 | 0.00 | 0.00 | 0.00 | 1.25 [0.91-1.72] | 1.22 [0.88-1.69] |
| E72 | 840 | 81,594,490 | 0.01 | 0.01 | 0.01 | 0.00 | 0.00 | 0.00 | 1.13 [0.90-1.43] | 1.12 [0.89-1.43] |
| E73 | 4,060 | 81,531,880 | 0.06 | 0.05 | 0.05 | 0.02 | 0.02 | 0.01 | 1.32 [1.19-1.45]* | 1.16 [1.05-1.29]* |
| E74 | 1,420 | 81,585,670 | 0.02 | 0.02 | 0.02 | 0.00 | 0.00 | 0.00 | 1.29 [1.08-1.53]* | 1.23 [1.03-1.47] |
| E75 | 600 | 81,599,130 | 0.01 | 0.01 | 0.01 | 0.00 | 0.00 | 0.00 | 1.11 [0.84-1.47] | 1.12 [0.85-1.48] |
| E78 | 386,100 | 75,439,450 | 5.43 | 5.04 | 4.71 | 0.39 | 0.72 | 0.59 | 1.15 [1.14-1.17]* | 1.12 [1.11-1.14]* |
| E79 | 590 | 81,604,050 | 0.01 | 0.01 | 0.01 | 0.00 | 0.00 | 0.00 | 1.19 [0.90-1.58] | 1.13 [0.85-1.50] |
| E80 | 5,630 | 81,524,740 | 0.07 | 0.07 | 0.06 | 0.00 | 0.01 | 0.00 | 1.08 [0.99-1.19] | 1.03 [0.94-1.13] |
| E83 | 79,560 | 81,168,710 | 1.02 | 0.97 | 0.89 | 0.05 | 0.13 | 0.12 | 1.15 [1.12-1.18]* | 1.13 [1.10-1.16]* |
| E84 | 630 | 81,573,470 | 0.01 | 0.01 | 0.01 | 0.00 | 0.00 | 0.00 | 1.14 [0.87-1.50] | 1.03 [0.78-1.36] |
| E85 | 2,880 | 81,591,010 | 0.04 | 0.03 | 0.03 | 0.00 | 0.01 | 0.00 | 1.18 [1.04-1.34] | 1.14 [1.00-1.30] |
| E86 | 208,290 | 80,284,600 | 2.77 | 2.55 | 2.43 | 0.21 | 0.34 | 0.32 | 1.14 [1.12-1.16]* | 1.13 [1.11-1.15]* |
| E87 | 310,120 | 79,913,950 | 4.28 | 3.78 | 3.51 | 0.50 | 0.77 | 0.70 | 1.22 [1.21-1.24]* | 1.20 [1.18-1.21]* |
| E88 | 15,120 | 81,513,790 | 0.21 | 0.18 | 0.17 | 0.03 | 0.04 | 0.03 | 1.23 [1.16-1.30]* | 1.20 [1.13-1.27]* |
| E89 | 20,670 | 81,369,430 | 0.26 | 0.25 | 0.23 | 0.01 | 0.03 | 0.03 | 1.15 [1.10-1.21]* | 1.11 [1.06-1.16]* |
| hospitalised - F01-F99 | | | | | | | | | | |
| F00 | 47,170 | 81,452,030 | 0.58 | 0.58 | 0.56 | 0.01 | 0.03 | 0.03 | 1.05 [1.02-1.09]* | 1.05 [1.01-1.08]* |
| F01 | 55,520 | 81,418,320 | 0.69 | 0.68 | 0.65 | 0.01 | 0.04 | 0.04 | 1.06 [1.03-1.10]* | 1.06 [1.03-1.09]* |
| F02 | 9,690 | 81,576,050 | 0.11 | 0.12 | 0.11 | −0.01 | −0.01 | 0.00 | 0.95 [0.88-1.03] | 0.97 [0.90-1.05] |
| F03 | 129,530 | 81,128,370 | 1.62 | 1.59 | 1.55 | 0.02 | 0.06 | 0.07 | 1.04 [1.02-1.06]* | 1.04 [1.02-1.07]* |
| F05 | 123,790 | 81,281,940 | 1.62 | 1.50 | 1.42 | 0.13 | 0.20 | 0.19 | 1.14 [1.12-1.16]* | 1.13 [1.11-1.16]* |
| F06 | 19,490 | 81,507,640 | 0.25 | 0.24 | 0.22 | 0.01 | 0.02 | 0.02 | 1.10 [1.04-1.16]* | 1.09 [1.04-1.15]* |
| F07 | 1,320 | 81,585,890 | 0.02 | 0.02 | 0.02 | 0.00 | 0.00 | 0.00 | 1.09 [0.90-1.32] | 1.08 [0.89-1.31] |
| F10 | 91,770 | 79,520,110 | 1.18 | 1.15 | 1.03 | 0.03 | 0.14 | 0.15 | 1.14 [1.11-1.16]* | 1.15 [1.12-1.18]* |
| F11 | 7,480 | 81,345,860 | 0.09 | 0.09 | 0.08 | −0.01 | 0.01 | 0.00 | 1.07 [0.99-1.16] | 1.06 [0.97-1.15] |
| F12 | 16,290 | 81,375,920 | 0.19 | 0.20 | 0.20 | −0.01 | −0.01 | −0.01 | 0.95 [0.90-1.01] | 0.96 [0.91-1.02] |
| F13 | 1,830 | 81,577,800 | 0.02 | 0.02 | 0.02 | 0.00 | 0.00 | 0.00 | 1.10 [0.93-1.29] | 1.06 [0.89-1.24] |
| F14 | 6,700 | 81,501,640 | 0.08 | 0.08 | 0.08 | −0.01 | 0.00 | 0.00 | 0.98 [0.90-1.07] | 0.99 [0.91-1.08] |
| F15 | 2,280 | 81,555,960 | 0.03 | 0.03 | 0.03 | 0.00 | 0.00 | 0.00 | 0.93 [0.80-1.08] | 0.94 [0.81-1.10] |
| F16 | 680 | 81,589,220 | 0.01 | 0.01 | 0.01 | 0.00 | 0.00 | 0.00 | 0.87 [0.66-1.14] | 0.88 [0.66-1.17] |
| F17 | 459,920 | 76,318,170 | 5.87 | 6.07 | 5.53 | −0.19 | 0.34 | 0.49 | 1.06 [1.05-1.07]* | 1.09 [1.08-1.10]* |
| F19 | 7,480 | 81,462,220 | 0.08 | 0.09 | 0.09 | −0.01 | 0.00 | 0.00 | 0.96 [0.89-1.05] | 0.98 [0.90-1.07] |
| F20 | 13,370 | 81,160,100 | 0.17 | 0.16 | 0.15 | 0.00 | 0.02 | 0.02 | 1.15 [1.08-1.22]* | 1.17 [1.10-1.24]* |
| F22 | 6,480 | 81,515,060 | 0.09 | 0.08 | 0.07 | 0.01 | 0.01 | 0.01 | 1.19 [1.10-1.30]* | 1.19 [1.09-1.29]* |
| F23 | 3,690 | 81,508,600 | 0.04 | 0.05 | 0.04 | 0.00 | 0.00 | 0.00 | 1.07 [0.96-1.20] | 1.08 [0.96-1.21] |
| F25 | 3,420 | 81,513,540 | 0.04 | 0.04 | 0.04 | 0.00 | 0.00 | 0.01 | 1.11 [0.99-1.25] | 1.15 [1.02-1.30] |
| F29 | 7,550 | 81,501,580 | 0.10 | 0.09 | 0.09 | 0.01 | 0.01 | 0.01 | 1.11 [1.03-1.20]* | 1.11 [1.02-1.20] |

| | | | | | | | | | | |
|---|---|---|---|---|---|---|---|---|---|---|
| F30 | 1,870 | 81,545,300 | 0.02 | 0.02 | 0.02 | 0.00 | 0.00 | 0.00 | 1.19 [1.02-1.39] | 1.18 [1.01-1.38] |
| F31 | 18,490 | 81,255,810 | 0.23 | 0.23 | 0.20 | 0.01 | 0.03 | 0.03 | 1.16 [1.10-1.22]* | 1.12 [1.06-1.18]* |
| F32 | 328,920 | 77,616,790 | 4.74 | 4.11 | 3.76 | 0.63 | 0.99 | 0.78 | 1.26 [1.25-1.28]* | 1.20 [1.18-1.21]* |
| F33 | 10,430 | 81,330,010 | 0.13 | 0.13 | 0.11 | 0.00 | 0.02 | 0.02 | 1.19 [1.11-1.27]* | 1.15 [1.07-1.23]* |
| F34 | 1,800 | 81,578,730 | 0.02 | 0.02 | 0.02 | 0.00 | 0.00 | 0.00 | 1.23 [1.05-1.44]* | 1.13 [0.96-1.32] |
| F38 | 820 | 81,600,480 | 0.01 | 0.01 | 0.01 | 0.00 | 0.00 | 0.00 | 1.16 [0.92-1.47] | 1.10 [0.86-1.41] |
| F39 | 1,500 | 81,590,650 | 0.02 | 0.02 | 0.02 | 0.00 | 0.00 | 0.00 | 1.24 [1.04-1.47] | 1.17 [0.98-1.40] |
| F40 | 30,890 | 81,275,260 | 0.43 | 0.37 | 0.36 | 0.06 | 0.07 | 0.05 | 1.19 [1.14-1.24]* | 1.13 [1.09-1.18]* |
| F41 | 281,700 | 79,236,760 | 4.01 | 3.44 | 3.22 | 0.56 | 0.79 | 0.58 | 1.25 [1.23-1.26]* | 1.17 [1.16-1.19]* |
| F42 | 6,960 | 81,521,290 | 0.11 | 0.08 | 0.08 | 0.03 | 0.04 | 0.03 | 1.47 [1.36-1.58]* | 1.34 [1.24-1.46]* |
| F43 | 16,460 | 81,327,920 | 0.21 | 0.20 | 0.19 | 0.02 | 0.03 | 0.02 | 1.16 [1.10-1.22]* | 1.09 [1.04-1.15]* |
| F44 | 4,100 | 81,555,090 | 0.06 | 0.05 | 0.05 | 0.01 | 0.01 | 0.01 | 1.23 [1.11-1.36]* | 1.11 [1.00-1.24] |
| F45 | 4,700 | 81,515,050 | 0.07 | 0.06 | 0.05 | 0.01 | 0.02 | 0.01 | 1.29 [1.17-1.42]* | 1.17 [1.06-1.29]* |
| F48 | 1,350 | 81,589,030 | 0.02 | 0.02 | 0.01 | 0.00 | 0.00 | 0.00 | 1.24 [1.04-1.48] | 1.12 [0.93-1.35] |
| F50 | 7,180 | 81,486,360 | 0.11 | 0.08 | 0.09 | 0.02 | 0.02 | 0.01 | 1.19 [1.10-1.28]* | 1.13 [1.04-1.22]* |
| F51 | 1,650 | 81,588,390 | 0.03 | 0.02 | 0.02 | 0.01 | 0.01 | 0.01 | 1.37 [1.17-1.60]* | 1.27 [1.08-1.50]* |
| F52 | 2,630 | 81,558,310 | 0.04 | 0.03 | 0.03 | 0.00 | 0.01 | 0.01 | 1.32 [1.16-1.50]* | 1.24 [1.09-1.42]* |
| F53 | 2,750 | 81,558,040 | 0.04 | 0.03 | 0.03 | 0.00 | 0.01 | 0.00 | 1.20 [1.06-1.36]* | 1.15 [1.01-1.31] |
| F60 | 14,290 | 81,347,590 | 0.19 | 0.17 | 0.16 | 0.02 | 0.03 | 0.02 | 1.21 [1.14-1.27]* | 1.13 [1.07-1.20]* |
| F69 | 1,130 | 81,585,390 | 0.02 | 0.01 | 0.01 | 0.00 | 0.00 | 0.00 | 1.39 [1.15-1.68]* | 1.29 [1.06-1.57] |
| F70 | 2,060 | 81,563,140 | 0.03 | 0.02 | 0.02 | 0.01 | 0.01 | 0.01 | 1.50 [1.30-1.72]* | 1.39 [1.20-1.61]* |
| F71 | 820 | 81,589,730 | 0.01 | 0.01 | 0.01 | 0.01 | 0.01 | 0.01 | 1.73 [1.39-2.15]* | 1.67 [1.34-2.08]* |
| F72 | 1,640 | 81,576,620 | 0.03 | 0.02 | 0.02 | 0.01 | 0.01 | 0.01 | 1.42 [1.21-1.66]* | 1.34 [1.14-1.57]* |
| F79 | 9,790 | 81,460,770 | 0.16 | 0.11 | 0.12 | 0.05 | 0.04 | 0.04 | 1.38 [1.30-1.48]* | 1.31 [1.23-1.40]* |
| F80 | 11,110 | 81,474,720 | 0.16 | 0.13 | 0.14 | 0.03 | 0.02 | 0.02 | 1.14 [1.08-1.22]* | 1.12 [1.05-1.19]* |
| F81 | 26,620 | 81,066,040 | 0.43 | 0.30 | 0.31 | 0.12 | 0.11 | 0.10 | 1.36 [1.31-1.42]* | 1.29 [1.24-1.35]* |
| F82 | 1,570 | 81,584,850 | 0.02 | 0.02 | 0.02 | 0.00 | 0.00 | 0.00 | 1.06 [0.90-1.25] | 1.02 [0.86-1.20] |
| F83 | 1,490 | 81,593,500 | 0.02 | 0.02 | 0.02 | 0.00 | 0.00 | 0.00 | 0.88 [0.74-1.06] | 0.84 [0.70-1.01] |
| F84 | 25,430 | 81,309,560 | 0.43 | 0.28 | 0.35 | 0.14 | 0.08 | 0.05 | 1.23 [1.18-1.28]* | 1.14 [1.09-1.19]* |
| F89 | 7,210 | 81,549,550 | 0.10 | 0.09 | 0.10 | 0.01 | −0.01 | −0.01 | 0.94 [0.87-1.01] | 0.88 [0.82-0.96]* |
| F90 | 13,160 | 81,467,110 | 0.21 | 0.15 | 0.19 | 0.06 | 0.03 | 0.02 | 1.14 [1.08-1.21]* | 1.11 [1.04-1.17]* |
| F91 | 2,870 | 81,547,410 | 0.04 | 0.03 | 0.04 | 0.01 | 0.01 | 0.00 | 1.16 [1.03-1.31] | 1.12 [0.99-1.26] |
| F94 | 520 | 81,604,790 | 0.01 | 0.01 | 0.01 | 0.00 | 0.00 | 0.00 | 1.07 [0.81-1.43] | 1.04 [0.77-1.39] |
| F95 | 1,520 | 81,591,710 | 0.03 | 0.02 | 0.02 | 0.01 | 0.01 | 0.00 | 1.27 [1.08-1.50]* | 1.20 [1.01-1.42] |
| F98 | 1,990 | 81,577,510 | 0.03 | 0.02 | 0.03 | 0.01 | 0.01 | 0.00 | 1.22 [1.05-1.41]* | 1.15 [0.99-1.34] |
| F99 | 5,580 | 81,473,030 | 0.07 | 0.07 | 0.06 | 0.00 | 0.01 | 0.01 | 1.12 [1.02-1.23] | 1.09 [0.99-1.19] |
| hospitalised - G00-G99 | | | | | | | | | | |
| G00 | 1,450 | 81,545,990 | 0.02 | 0.02 | 0.02 | 0.00 | 0.00 | 0.00 | 1.02 [0.85-1.22] | 0.98 [0.81-1.18] |
| G01 | 700 | 81,569,010 | 0.01 | 0.01 | 0.01 | 0.00 | 0.00 | 0.00 | 1.10 [0.85-1.41] | 1.08 [0.84-1.39] |
| G02 | 690 | 81,584,660 | 0.01 | 0.01 | 0.01 | 0.00 | 0.00 | 0.00 | 1.05 [0.81-1.37] | 1.00 [0.76-1.30] |
| G03 | 1,960 | 81,549,040 | 0.02 | 0.02 | 0.02 | 0.00 | 0.00 | 0.00 | 1.05 [0.90-1.23] | 1.02 [0.87-1.19] |
| G04 | 4,270 | 81,552,860 | 0.06 | 0.05 | 0.05 | 0.01 | 0.01 | 0.01 | 1.15 [1.04-1.28]* | 1.12 [1.01-1.24] |
| G05 | 1,030 | 81,593,120 | 0.02 | 0.01 | 0.01 | 0.00 | 0.00 | 0.00 | 1.44 [1.18-1.77]* | 1.38 [1.12-1.70]* |
| G06 | 1,840 | 81,581,090 | 0.02 | 0.02 | 0.02 | 0.00 | 0.00 | 0.00 | 1.20 [1.03-1.41] | 1.20 [1.02-1.41] |
| G08 | 950 | 81,595,360 | 0.01 | 0.01 | 0.01 | 0.00 | 0.00 | 0.00 | 1.29 [1.04-1.60] | 1.29 [1.03-1.61] |
| G09 | 1,280 | 81,585,170 | 0.02 | 0.02 | 0.01 | 0.00 | 0.00 | 0.00 | 1.17 [0.98-1.41] | 1.14 [0.95-1.38] |
| G10 | 470 | 81,600,980 | 0.01 | 0.01 | 0.01 | 0.00 | 0.00 | 0.00 | 1.25 [0.92-1.69] | 1.27 [0.93-1.73] |
| G11 | 1,950 | 81,579,880 | 0.02 | 0.02 | 0.02 | 0.00 | 0.00 | 0.00 | 1.15 [0.99-1.34] | 1.14 [0.97-1.33] |
| G12 | 3,840 | 81,583,480 | 0.05 | 0.05 | 0.04 | 0.00 | 0.00 | 0.00 | 1.06 [0.95-1.19] | 1.05 [0.94-1.18] |
| G20 | 33,700 | 81,335,820 | 0.40 | 0.42 | 0.39 | −0.02 | 0.01 | 0.01 | 1.03 [0.99-1.07] | 1.03 [0.99-1.07] |
| G21 | 3,370 | 81,590,570 | 0.05 | 0.04 | 0.04 | 0.01 | 0.01 | 0.01 | 1.22 [1.08-1.38]* | 1.24 [1.10-1.40]* |
| G23 | 2,030 | 81,600,540 | 0.02 | 0.03 | 0.02 | 0.00 | 0.00 | 0.00 | 1.02 [0.87-1.19] | 1.01 [0.86-1.18] |
| G24 | 4,670 | 81,540,800 | 0.06 | 0.06 | 0.06 | 0.00 | 0.00 | 0.00 | 1.04 [0.94-1.15] | 1.02 [0.92-1.13] |
| G25 | 21,460 | 81,434,560 | 0.29 | 0.26 | 0.24 | 0.03 | 0.05 | 0.03 | 1.20 [1.15-1.26]* | 1.13 [1.07-1.18]* |
| G30 | 62,200 | 81,361,150 | 0.77 | 0.76 | 0.73 | 0.01 | 0.04 | 0.04 | 1.06 [1.03-1.09]* | 1.05 [1.02-1.09]* |
| G31 | 46,520 | 81,362,890 | 0.59 | 0.57 | 0.52 | 0.03 | 0.07 | 0.07 | 1.13 [1.10-1.17]* | 1.13 [1.09-1.17]* |
| G32 | 760 | 81,602,140 | 0.01 | 0.01 | 0.01 | 0.00 | 0.00 | 0.00 | 1.29 [1.01-1.64] | 1.29 [1.01-1.65] |
| G35 | 8,460 | 81,328,400 | 0.10 | 0.10 | 0.09 | 0.00 | 0.01 | 0.01 | 1.14 [1.06-1.22]* | 1.12 [1.04-1.21]* |
| G37 | 2,540 | 81,543,150 | 0.03 | 0.03 | 0.03 | 0.00 | 0.01 | 0.01 | 1.22 [1.07-1.40]* | 1.19 [1.04-1.36] |
| G40 | 65,210 | 79,984,660 | 0.89 | 0.80 | 0.75 | 0.09 | 0.14 | 0.11 | 1.19 [1.16-1.22]* | 1.14 [1.11-1.17]* |
| G41 | 5,580 | 81,518,390 | 0.07 | 0.07 | 0.07 | 0.01 | 0.01 | 0.01 | 1.09 [1.00-1.20] | 1.07 [0.98-1.18] |
| G43 | 65,590 | 80,732,890 | 0.91 | 0.79 | 0.75 | 0.12 | 0.16 | 0.09 | 1.21 [1.18-1.25]* | 1.12 [1.09-1.15]* |
| G44 | 9,500 | 81,449,610 | 0.13 | 0.11 | 0.11 | 0.02 | 0.02 | 0.02 | 1.23 [1.15-1.32]* | 1.15 [1.07-1.23]* |
| G45 | 43,040 | 81,007,210 | 0.54 | 0.53 | 0.48 | 0.01 | 0.06 | 0.05 | 1.12 [1.08-1.15]* | 1.09 [1.06-1.13]* |
| G46 | 2,910 | 81,589,260 | 0.04 | 0.04 | 0.03 | 0.00 | 0.00 | 0.00 | 1.15 [1.01-1.30] | 1.13 [0.99-1.29] |
| G47 | 85,780 | 80,393,660 | 1.34 | 1.00 | 0.97 | 0.34 | 0.37 | 0.28 | 1.38 [1.35-1.41]* | 1.27 [1.24-1.30]* |
| G50 | 7,350 | 81,509,750 | 0.10 | 0.09 | 0.08 | 0.02 | 0.02 | 0.02 | 1.32 [1.22-1.42]* | 1.22 [1.13-1.32]* |
| G51 | 15,430 | 81,363,460 | 0.20 | 0.19 | 0.17 | 0.01 | 0.03 | 0.02 | 1.15 [1.09-1.21]* | 1.11 [1.05-1.17]* |
| G52 | 1,180 | 81,593,630 | 0.02 | 0.01 | 0.01 | 0.00 | 0.00 | 0.00 | 1.26 [1.03-1.53] | 1.20 [0.98-1.46] |
| G53 | 2,550 | 81,586,630 | 0.04 | 0.03 | 0.03 | 0.00 | 0.01 | 0.01 | 1.26 [1.10-1.44]* | 1.21 [1.05-1.39]* |
| G54 | 4,830 | 81,538,200 | 0.06 | 0.06 | 0.05 | 0.00 | 0.01 | 0.01 | 1.24 [1.13-1.37]* | 1.18 [1.07-1.30]* |
| G55 | 52,950 | 80,844,380 | 0.67 | 0.65 | 0.57 | 0.02 | 0.10 | 0.07 | 1.18 [1.14-1.21]* | 1.11 [1.08-1.15]* |
| G56 | 74,220 | 79,648,020 | 1.00 | 0.91 | 0.81 | 0.09 | 0.19 | 0.14 | 1.24 [1.21-1.27]* | 1.16 [1.13-1.19]* |
| G57 | 9,170 | 81,419,860 | 0.12 | 0.11 | 0.10 | 0.01 | 0.02 | 0.02 | 1.25 [1.17-1.34]* | 1.17 [1.09-1.26]* |
| G58 | 4,820 | 81,529,180 | 0.06 | 0.06 | 0.05 | 0.01 | 0.01 | 0.01 | 1.24 [1.12-1.36]* | 1.16 [1.05-1.28]* |
| G59 | 3,930 | 81,559,050 | 0.05 | 0.05 | 0.04 | 0.01 | 0.01 | 0.01 | 1.27 [1.14-1.41]* | 1.20 [1.07-1.33]* |
| G60 | 2,250 | 81,575,250 | 0.03 | 0.03 | 0.03 | 0.00 | 0.00 | 0.00 | 1.17 [1.01-1.35] | 1.07 [0.93-1.24] |
| G61 | 2,540 | 81,556,420 | 0.03 | 0.03 | 0.03 | 0.00 | 0.00 | 0.00 | 1.14 [0.99-1.30] | 1.10 [0.96-1.26] |
| G62 | 30,530 | 81,359,790 | 0.41 | 0.37 | 0.33 | 0.04 | 0.08 | 0.06 | 1.23 [1.19-1.28]* | 1.17 [1.12-1.22]* |
| G63 | 16,580 | 81,494,480 | 0.23 | 0.20 | 0.17 | 0.03 | 0.06 | 0.04 | 1.32 [1.26-1.39]* | 1.24 [1.18-1.31]* |
| G70 | 2,550 | 81,567,030 | 0.03 | 0.03 | 0.03 | 0.00 | 0.00 | 0.00 | 1.14 [0.99-1.30] | 1.10 [0.95-1.26] |
| G71 | 2,280 | 81,559,740 | 0.03 | 0.03 | 0.03 | 0.00 | 0.00 | 0.00 | 1.04 [0.90-1.20] | 1.03 [0.89-1.19] |
| G72 | 3,320 | 81,570,070 | 0.05 | 0.04 | 0.04 | 0.01 | 0.01 | 0.01 | 1.28 [1.14-1.43]* | 1.23 [1.09-1.38]* |
| G80 | 7,370 | 81,371,710 | 0.09 | 0.09 | 0.10 | 0.00 | −0.01 | −0.01 | 0.92 [0.85-1.00] | 0.90 [0.83-0.97]* |
| G81 | 52,100 | 81,045,370 | 0.66 | 0.64 | 0.59 | 0.02 | 0.07 | 0.07 | 1.12 [1.08-1.15]* | 1.11 [1.08-1.15]* |
| G82 | 5,750 | 81,488,740 | 0.07 | 0.07 | 0.07 | 0.00 | 0.00 | 0.00 | 1.04 [0.95-1.14] | 1.03 [0.94-1.13] |
| G83 | 8,530 | 81,509,330 | 0.10 | 0.11 | 0.09 | 0.00 | 0.01 | 0.01 | 1.09 [1.01-1.18] | 1.06 [0.98-1.15] |
| G90 | 4,510 | 81,559,250 | 0.06 | 0.05 | 0.05 | 0.00 | 0.01 | 0.00 | 1.13 [1.02-1.25] | 1.06 [0.95-1.18] |
| G91 | 9,760 | 81,457,550 | 0.12 | 0.12 | 0.11 | 0.01 | 0.02 | 0.02 | 1.14 [1.06-1.22]* | 1.14 [1.06-1.22]* |
| G93 | 42,270 | 81,167,670 | 0.56 | 0.51 | 0.48 | 0.05 | 0.08 | 0.06 | 1.17 [1.14-1.21]* | 1.12 [1.08-1.16]* |
| G94 | 1,450 | 81,595,890 | 0.02 | 0.02 | 0.02 | 0.00 | 0.00 | 0.00 | 1.11 [0.93-1.32] | 1.13 [0.94-1.36] |
| G95 | 11,170 | 81,485,510 | 0.14 | 0.14 | 0.12 | 0.00 | 0.01 | 0.01 | 1.11 [1.04-1.19]* | 1.09 [1.02-1.17] |
| G96 | 3,680 | 81,552,160 | 0.05 | 0.04 | 0.04 | 0.00 | 0.01 | 0.01 | 1.20 [1.08-1.34]* | 1.15 [1.03-1.29] |
| G97 | 3,480 | 81,544,530 | 0.05 | 0.04 | 0.04 | 0.00 | 0.00 | 0.00 | 1.11 [0.99-1.24] | 1.05 [0.94-1.19] |
| G98 | 1,320 | 81,600,510 | 0.02 | 0.02 | 0.01 | 0.00 | 0.01 | 0.00 | 1.41 [1.18-1.69]* | 1.29 [1.07-1.55]* |
| G99 | 17,400 | 81,413,380 | 0.22 | 0.21 | 0.19 | 0.01 | 0.04 | 0.03 | 1.19 [1.13-1.26]* | 1.15 [1.09-1.21]* |
| hospitalised - H00-H59 | | | | | | | | | | |
| H00 | 13,330 | 81,108,730 | 0.18 | 0.16 | 0.16 | 0.02 | 0.03 | 0.02 | 1.17 [1.11-1.24]* | 1.14 [1.08-1.21]* |
| H01 | 10,440 | 81,456,960 | 0.15 | 0.12 | 0.11 | 0.03 | 0.04 | 0.03 | 1.32 [1.24-1.41]* | 1.26 [1.18-1.34]* |
| H02 | 53,490 | 80,471,540 | 0.72 | 0.65 | 0.59 | 0.07 | 0.13 | 0.11 | 1.22 [1.19-1.26]* | 1.17 [1.14-1.21]* |
| H03 | 790 | 81,591,260 | 0.01 | 0.01 | 0.01 | 0.00 | 0.00 | 0.00 | 1.34 [1.06-1.68] | 1.23 [0.97-1.57] |
| H04 | 27,450 | 81,046,430 | 0.39 | 0.32 | 0.30 | 0.07 | 0.09 | 0.08 | 1.31 [1.26-1.37]* | 1.24 [1.19-1.29]* |
| H05 | 9,520 | 81,380,630 | 0.15 | 0.11 | 0.11 | 0.04 | 0.04 | 0.03 | 1.32 [1.24-1.41]* | 1.29 [1.21-1.38]* |
| H06 | 1,100 | 81,588,950 | 0.02 | 0.01 | 0.01 | 0.00 | 0.00 | 0.00 | 1.37 [1.13-1.67]* | 1.34 [1.10-1.63]* |
| H10 | 16,230 | 81,269,940 | 0.25 | 0.19 | 0.18 | 0.06 | 0.06 | 0.05 | 1.35 [1.28-1.42]* | 1.28 [1.22-1.35]* |
| H11 | 11,120 | 81,396,020 | 0.14 | 0.13 | 0.12 | 0.01 | 0.02 | 0.02 | 1.17 [1.10-1.25]* | 1.15 [1.08-1.23]* |

| | | | | | | | | | | |
|---|---|---|---|---|---|---|---|---|---|---|
| H13 | 890 | 81,589,830 | 0.01 | 0.01 | 0.01 | 0.00 | 0.00 | 0.00 | 1.36 [1.10-1.69]* | 1.29 [1.03-1.61] |
| H15 | 1,070 | 81,591,570 | 0.01 | 0.01 | 0.01 | 0.00 | 0.00 | 0.00 | 1.32 [1.08-1.62]* | 1.24 [1.01-1.53] |
| H16 | 4,970 | 81,516,860 | 0.08 | 0.06 | 0.05 | 0.02 | 0.03 | 0.03 | 1.59 [1.45-1.74]* | 1.53 [1.39-1.68]* |
| H17 | 2,940 | 81,567,980 | 0.04 | 0.03 | 0.03 | 0.01 | 0.01 | 0.01 | 1.39 [1.23-1.57]* | 1.33 [1.18-1.50]* |
| H18 | 19,510 | 81,388,660 | 0.28 | 0.23 | 0.21 | 0.05 | 0.06 | 0.05 | 1.29 [1.23-1.36]* | 1.24 [1.18-1.30]* |
| H19 | 1,250 | 81,589,910 | 0.02 | 0.01 | 0.01 | 0.01 | 0.01 | 0.01 | 1.67 [1.40-1.99]* | 1.51 [1.26-1.81]* |
| H20 | 5,370 | 81,513,290 | 0.08 | 0.06 | 0.06 | 0.02 | 0.02 | 0.02 | 1.38 [1.26-1.50]* | 1.34 [1.23-1.47]* |
| H21 | 8,490 | 81,512,120 | 0.11 | 0.10 | 0.09 | 0.01 | 0.01 | 0.01 | 1.15 [1.06-1.24]* | 1.12 [1.04-1.21]* |
| H22 | 430 | 81,604,970 | 0.01 | 0.01 | 0.00 | 0.00 | 0.00 | 0.00 | 1.36 [0.99-1.87] | 1.33 [0.96-1.84] |
| H25 | 188,180 | 79,053,140 | 2.51 | 2.35 | 2.23 | 0.16 | 0.28 | 0.22 | 1.12 [1.11-1.14]* | 1.10 [1.08-1.12]* |
| H26 | 262,240 | 77,501,040 | 3.67 | 3.31 | 3.18 | 0.36 | 0.49 | 0.41 | 1.16 [1.14-1.17]* | 1.13 [1.11-1.14]* |
| H27 | 3,970 | 81,535,230 | 0.05 | 0.05 | 0.04 | 0.00 | 0.01 | 0.01 | 1.15 [1.03-1.28] | 1.13 [1.01-1.27] |
| H28 | 1,520 | 81,590,210 | 0.02 | 0.02 | 0.02 | 0.00 | 0.00 | 0.00 | 1.27 [1.07-1.51]* | 1.24 [1.04-1.48] |
| H30 | 480 | 81,600,150 | 0.01 | 0.01 | 0.01 | 0.00 | 0.00 | 0.00 | 1.34 [1.00-1.81] | 1.30 [0.96-1.76] |
| H31 | 4,820 | 81,557,210 | 0.06 | 0.06 | 0.06 | 0.00 | 0.00 | 0.00 | 1.05 [0.95-1.16] | 1.04 [0.94-1.15] |
| H33 | 17,000 | 81,178,500 | 0.19 | 0.21 | 0.19 | −0.02 | 0.01 | 0.01 | 1.03 [0.98-1.09] | 1.03 [0.97-1.09] |
| H34 | 11,420 | 81,476,640 | 0.14 | 0.14 | 0.13 | 0.00 | 0.01 | 0.01 | 1.11 [1.04-1.18]* | 1.09 [1.02-1.17] |
| H35 | 102,790 | 80,493,180 | 1.33 | 1.26 | 1.18 | 0.06 | 0.15 | 0.12 | 1.12 [1.10-1.15]* | 1.10 [1.08-1.13]* |
| H36 | 43,800 | 81,114,960 | 0.56 | 0.53 | 0.48 | 0.03 | 0.08 | 0.07 | 1.17 [1.13-1.21]* | 1.15 [1.11-1.19]* |
| H40 | 91,610 | 80,457,410 | 1.18 | 1.13 | 1.04 | 0.05 | 0.14 | 0.12 | 1.13 [1.11-1.16]* | 1.11 [1.08-1.14]* |
| H43 | 19,620 | 81,340,400 | 0.24 | 0.24 | 0.22 | −0.01 | 0.02 | 0.01 | 1.08 [1.03-1.13]* | 1.06 [1.01-1.11] |
| H44 | 6,430 | 81,493,780 | 0.09 | 0.08 | 0.07 | 0.01 | 0.02 | 0.01 | 1.24 [1.14-1.35]* | 1.19 [1.09-1.30]* |
| H46 | 1,920 | 81,575,540 | 0.03 | 0.02 | 0.02 | 0.00 | 0.00 | 0.00 | 1.22 [1.05-1.42] | 1.18 [1.01-1.38] |
| H47 | 8,370 | 81,497,530 | 0.11 | 0.10 | 0.10 | 0.01 | 0.01 | 0.01 | 1.15 [1.07-1.24]* | 1.12 [1.04-1.21]* |
| H49 | 5,380 | 81,529,730 | 0.06 | 0.07 | 0.06 | 0.00 | 0.00 | 0.00 | 1.02 [0.93-1.13] | 1.00 [0.91-1.10] |
| H50 | 23,520 | 80,971,190 | 0.32 | 0.28 | 0.31 | 0.04 | 0.01 | 0.00 | 1.03 [0.99-1.08] | 1.01 [0.97-1.06] |
| H51 | 1,050 | 81,592,920 | 0.01 | 0.01 | 0.01 | 0.00 | 0.00 | 0.00 | 1.05 [0.85-1.30] | 1.00 [0.81-1.24] |
| H52 | 33,260 | 81,265,310 | 0.42 | 0.41 | 0.38 | 0.01 | 0.04 | 0.03 | 1.09 [1.05-1.13]* | 1.07 [1.02-1.11]* |
| H53 | 68,880 | 80,602,830 | 0.92 | 0.84 | 0.79 | 0.08 | 0.13 | 0.09 | 1.16 [1.13-1.19]* | 1.11 [1.08-1.14]* |
| H54 | 50,950 | 81,220,010 | 0.66 | 0.62 | 0.58 | 0.04 | 0.08 | 0.07 | 1.14 [1.11-1.18]* | 1.12 [1.09-1.16]* |
| H55 | 5,510 | 81,533,360 | 0.07 | 0.07 | 0.07 | 0.00 | 0.00 | 0.00 | 1.06 [0.97-1.17] | 1.04 [0.94-1.14] |
| H57 | 11,680 | 81,449,570 | 0.16 | 0.14 | 0.13 | 0.02 | 0.03 | 0.02 | 1.20 [1.12-1.27]* | 1.14 [1.07-1.21]* |
| H58 | 2,210 | 81,586,400 | 0.03 | 0.03 | 0.02 | 0.00 | 0.00 | 0.00 | 1.16 [1.00-1.35] | 1.13 [0.98-1.32] |
| H59 | 3,880 | 81,542,340 | 0.05 | 0.05 | 0.04 | 0.00 | 0.00 | 0.00 | 1.12 [1.00-1.25] | 1.11 [0.99-1.25] |
| hospitalised - H60-H95 | | | | | | | | | | |
| H60 | 11,080 | 81,348,560 | 0.18 | 0.13 | 0.12 | 0.05 | 0.06 | 0.05 | 1.48 [1.39-1.57]* | 1.42 [1.34-1.51]* |
| H61 | 21,580 | 81,159,580 | 0.33 | 0.25 | 0.26 | 0.08 | 0.07 | 0.06 | 1.27 [1.22-1.33]* | 1.24 [1.18-1.30]* |
| H65 | 55,860 | 79,788,940 | 0.87 | 0.66 | 0.75 | 0.22 | 0.12 | 0.10 | 1.17 [1.14-1.20]* | 1.14 [1.11-1.17]* |
| H66 | 29,670 | 80,658,090 | 0.45 | 0.35 | 0.37 | 0.10 | 0.08 | 0.07 | 1.21 [1.16-1.25]* | 1.18 [1.13-1.22]* |
| H69 | 2,250 | 81,558,720 | 0.03 | 0.03 | 0.03 | 0.01 | 0.01 | 0.01 | 1.32 [1.16-1.52]* | 1.25 [1.08-1.43]* |
| H70 | 2,840 | 81,551,950 | 0.04 | 0.03 | 0.03 | 0.00 | 0.00 | 0.00 | 1.10 [0.97-1.24] | 1.07 [0.94-1.22] |
| H71 | 6,110 | 81,436,460 | 0.08 | 0.07 | 0.08 | 0.01 | 0.00 | 0.00 | 1.04 [0.95-1.13] | 1.05 [0.96-1.15] |
| H72 | 12,800 | 81,225,210 | 0.18 | 0.15 | 0.16 | 0.03 | 0.02 | 0.02 | 1.13 [1.07-1.20]* | 1.11 [1.05-1.18]* |
| H73 | 9,050 | 81,384,620 | 0.13 | 0.11 | 0.12 | 0.03 | 0.02 | 0.01 | 1.14 [1.06-1.22]* | 1.12 [1.05-1.20]* |
| H74 | 6,640 | 81,428,060 | 0.09 | 0.08 | 0.08 | 0.01 | 0.01 | 0.01 | 1.12 [1.03-1.21]* | 1.09 [1.00-1.18] |
| H80 | 1,230 | 81,569,180 | 0.01 | 0.02 | 0.01 | 0.00 | 0.00 | 0.00 | 1.11 [0.92-1.35] | 1.09 [0.89-1.33] |
| H81 | 20,710 | 81,392,500 | 0.27 | 0.25 | 0.23 | 0.02 | 0.04 | 0.03 | 1.17 [1.11-1.23]* | 1.11 [1.06-1.16]* |
| H83 | 8,560 | 81,452,320 | 0.11 | 0.10 | 0.09 | 0.01 | 0.02 | 0.01 | 1.17 [1.09-1.26]* | 1.11 [1.03-1.19]* |
| H90 | 27,840 | 81,256,260 | 0.40 | 0.33 | 0.33 | 0.07 | 0.07 | 0.05 | 1.20 [1.16-1.25]* | 1.15 [1.11-1.20]* |
| H91 | 130,360 | 80,566,200 | 1.77 | 1.58 | 1.51 | 0.19 | 0.26 | 0.22 | 1.18 [1.15-1.20]* | 1.14 [1.12-1.16]* |
| H92 | 10,520 | 81,366,910 | 0.16 | 0.12 | 0.12 | 0.03 | 0.03 | 0.03 | 1.26 [1.18-1.34]* | 1.20 [1.13-1.28]* |
| H93 | 12,160 | 81,457,620 | 0.17 | 0.14 | 0.13 | 0.02 | 0.04 | 0.03 | 1.28 [1.21-1.36]* | 1.21 [1.14-1.29]* |
| H95 | 1,450 | 81,571,870 | 0.02 | 0.02 | 0.02 | 0.00 | 0.00 | 0.00 | 1.12 [0.94-1.33] | 1.13 [0.94-1.35] |
| hospitalised - I00-I99 | | | | | | | | | | |
| I05 | 6,750 | 81,517,090 | 0.09 | 0.08 | 0.07 | 0.01 | 0.01 | 0.01 | 1.19 [1.10-1.30]* | 1.16 [1.07-1.26]* |
| I06 | 670 | 81,596,240 | 0.01 | 0.01 | 0.01 | 0.00 | 0.00 | 0.00 | 1.18 [0.91-1.52] | 1.12 [0.86-1.45] |
| I07 | 17,640 | 81,520,100 | 0.24 | 0.21 | 0.19 | 0.03 | 0.05 | 0.04 | 1.23 [1.17-1.30]* | 1.20 [1.14-1.27]* |
| I08 | 88,830 | 81,118,690 | 1.16 | 1.08 | 1.01 | 0.08 | 0.15 | 0.13 | 1.15 [1.12-1.18]* | 1.12 [1.10-1.15]* |
| I09 | 1,030 | 81,596,700 | 0.01 | 0.01 | 0.01 | 0.00 | 0.00 | 0.00 | 1.37 [1.11-1.68]* | 1.33 [1.07-1.64]* |
| I10 | 652,690 | 67,379,090 | 10.99 | 9.35 | 9.37 | 1.65 | 1.62 | 1.38 | 1.17 [1.16-1.18]* | 1.14 [1.13-1.15]* |
| I11 | 4,980 | 81,543,760 | 0.06 | 0.06 | 0.06 | 0.00 | 0.00 | 0.00 | 1.06 [0.96-1.17] | 1.04 [0.94-1.15] |
| I12 | 50,020 | 81,093,880 | 0.64 | 0.61 | 0.54 | 0.03 | 0.10 | 0.10 | 1.19 [1.16-1.23]* | 1.18 [1.14-1.22]* |
| I13 | 2,620 | 81,601,500 | 0.03 | 0.03 | 0.03 | 0.00 | 0.01 | 0.00 | 1.17 [1.02-1.35] | 1.14 [0.99-1.31] |
| I15 | 3,920 | 81,555,910 | 0.05 | 0.05 | 0.04 | 0.00 | 0.00 | 0.00 | 1.11 [1.00-1.24] | 1.09 [0.98-1.22] |
| I20 | 176,160 | 77,429,990 | 2.43 | 2.24 | 2.08 | 0.20 | 0.35 | 0.26 | 1.17 [1.15-1.19]* | 1.12 [1.10-1.14]* |
| I21 | 117,600 | 79,809,470 | 1.50 | 1.47 | 1.35 | 0.03 | 0.15 | 0.13 | 1.11 [1.09-1.13]* | 1.10 [1.07-1.12]* |
| I22 | 11,880 | 81,420,530 | 0.15 | 0.14 | 0.13 | 0.01 | 0.02 | 0.02 | 1.18 [1.11-1.25]* | 1.15 [1.08-1.23]* |
| I23 | 650 | 81,598,080 | 0.01 | 0.01 | 0.01 | 0.00 | 0.00 | 0.00 | 1.31 [1.02-1.70] | 1.29 [1.00-1.68] |
| I24 | 37,560 | 81,297,100 | 0.49 | 0.46 | 0.42 | 0.03 | 0.07 | 0.06 | 1.17 [1.13-1.21]* | 1.14 [1.10-1.18]* |
| I25 | 296,940 | 76,169,720 | 4.19 | 3.82 | 3.67 | 0.36 | 0.52 | 0.41 | 1.14 [1.13-1.16]* | 1.11 [1.10-1.12]* |
| I26 | 63,850 | 80,929,310 | 0.77 | 0.79 | 0.72 | −0.02 | 0.05 | 0.03 | 1.07 [1.04-1.10]* | 1.04 [1.01-1.07]* |
| I27 | 41,440 | 81,375,400 | 0.56 | 0.50 | 0.45 | 0.07 | 0.11 | 0.09 | 1.25 [1.21-1.29]* | 1.20 [1.16-1.24]* |
| I28 | 1,290 | 81,596,300 | 0.02 | 0.02 | 0.02 | 0.00 | 0.00 | 0.00 | 0.99 [0.82-1.21] | 0.94 [0.77-1.15] |
| I30 | 1,900 | 81,562,200 | 0.02 | 0.02 | 0.02 | 0.00 | 0.00 | 0.00 | 1.18 [1.01-1.37] | 1.15 [0.98-1.34] |
| I31 | 25,010 | 81,357,660 | 0.31 | 0.31 | 0.28 | 0.01 | 0.04 | 0.03 | 1.13 [1.08-1.18]* | 1.10 [1.05-1.15]* |
| I33 | 4,130 | 81,566,040 | 0.06 | 0.05 | 0.04 | 0.01 | 0.01 | 0.01 | 1.25 [1.13-1.39]* | 1.22 [1.10-1.36]* |
| I34 | 56,820 | 81,068,350 | 0.72 | 0.70 | 0.64 | 0.02 | 0.08 | 0.06 | 1.13 [1.09-1.16]* | 1.10 [1.07-1.13]* |
| I35 | 82,070 | 80,919,520 | 1.08 | 1.00 | 0.93 | 0.08 | 0.16 | 0.13 | 1.17 [1.14-1.20]* | 1.13 [1.10-1.16]* |
| I36 | 6,360 | 81,576,210 | 0.08 | 0.08 | 0.07 | 0.01 | 0.01 | 0.01 | 1.17 [1.07-1.28]* | 1.13 [1.04-1.24]* |
| I37 | 6,970 | 81,564,250 | 0.09 | 0.08 | 0.08 | 0.01 | 0.01 | 0.01 | 1.13 [1.04-1.23]* | 1.10 [1.01-1.20] |
| I38 | 9,400 | 81,489,180 | 0.12 | 0.11 | 0.10 | 0.01 | 0.02 | 0.02 | 1.21 [1.13-1.30]* | 1.17 [1.09-1.26]* |
| I40 | 620 | 81,600,160 | 0.01 | 0.01 | 0.01 | 0.00 | 0.00 | 0.00 | 1.35 [1.04-1.76] | 1.33 [1.02-1.75] |
| I42 | 21,160 | 81,333,670 | 0.27 | 0.26 | 0.23 | 0.01 | 0.03 | 0.03 | 1.15 [1.10-1.20]* | 1.11 [1.06-1.16]* |
| I43 | 920 | 81,605,810 | 0.01 | 0.01 | 0.01 | 0.00 | 0.00 | 0.00 | 0.97 [0.76-1.23] | 0.93 [0.73-1.18] |
| I44 | 129,810 | 80,634,060 | 1.70 | 1.59 | 1.47 | 0.11 | 0.22 | 0.19 | 1.15 [1.13-1.17]* | 1.13 [1.10-1.15]* |
| I45 | 82,070 | 80,977,230 | 1.06 | 1.00 | 0.92 | 0.06 | 0.14 | 0.11 | 1.15 [1.12-1.18]* | 1.12 [1.09-1.15]* |
| I46 | 41,380 | 81,459,640 | 0.53 | 0.50 | 0.45 | 0.03 | 0.08 | 0.08 | 1.18 [1.14-1.22]* | 1.17 [1.13-1.21]* |
| I47 | 46,510 | 80,859,610 | 0.59 | 0.57 | 0.52 | 0.02 | 0.07 | 0.05 | 1.14 [1.10-1.18]* | 1.10 [1.06-1.13]* |
| I48 | 331,760 | 78,203,070 | 4.50 | 4.18 | 4.02 | 0.32 | 0.48 | 0.40 | 1.12 [1.11-1.13]* | 1.10 [1.09-1.11]* |
| I49 | 56,460 | 81,011,750 | 0.72 | 0.69 | 0.64 | 0.02 | 0.08 | 0.05 | 1.12 [1.09-1.15]* | 1.08 [1.05-1.11]* |
| I50 | 269,500 | 79,775,020 | 3.72 | 3.29 | 3.09 | 0.43 | 0.63 | 0.54 | 1.20 [1.19-1.22]* | 1.17 [1.15-1.19]* |
| I51 | 156,150 | 80,532,850 | 2.07 | 1.91 | 1.75 | 0.16 | 0.31 | 0.26 | 1.18 [1.16-1.20]* | 1.14 [1.12-1.16]* |
| I60 | 8,000 | 81,472,570 | 0.10 | 0.10 | 0.09 | 0.00 | 0.01 | 0.01 | 1.10 [1.02-1.18] | 1.10 [1.02-1.20] |
| I61 | 20,910 | 81,439,670 | 0.26 | 0.26 | 0.23 | 0.01 | 0.03 | 0.03 | 1.14 [1.09-1.20]* | 1.14 [1.09-1.20]* |
| I62 | 12,680 | 81,515,050 | 0.16 | 0.15 | 0.14 | 0.00 | 0.01 | 0.01 | 1.10 [1.03-1.17]* | 1.10 [1.04-1.18]* |
| I63 | 99,330 | 80,740,600 | 1.21 | 1.23 | 1.14 | −0.02 | 0.08 | 0.08 | 1.07 [1.04-1.09]* | 1.07 [1.04-1.09]* |
| I64 | 24,590 | 81,239,710 | 0.31 | 0.30 | 0.27 | 0.00 | 0.03 | 0.03 | 1.12 [1.07-1.17]* | 1.12 [1.07-1.17]* |
| I65 | 25,030 | 81,324,150 | 0.31 | 0.31 | 0.28 | 0.00 | 0.03 | 0.03 | 1.11 [1.06-1.16]* | 1.10 [1.05-1.15]* |
| I66 | 1,610 | 81,594,550 | 0.02 | 0.02 | 0.02 | 0.00 | 0.00 | 0.00 | 1.12 [0.94-1.34] | 1.09 [0.91-1.30] |
| I67 | 160,410 | 80,761,490 | 2.08 | 1.96 | 1.83 | 0.12 | 0.25 | 0.23 | 1.14 [1.12-1.16]* | 1.13 [1.11-1.15]* |
| I68 | 950 | 81,606,610 | 0.01 | 0.01 | 0.01 | 0.00 | 0.00 | 0.00 | 1.25 [1.00-1.56] | 1.22 [0.97-1.53] |
| I69 | 48,360 | 81,129,260 | 0.60 | 0.60 | 0.54 | 0.00 | 0.05 | 0.05 | 1.10 [1.06-1.13]* | 1.09 [1.06-1.13]* |
| I70 | 40,220 | 81,273,150 | 0.53 | 0.49 | 0.45 | 0.04 | 0.08 | 0.08 | 1.18 [1.14-1.22]* | 1.17 [1.13-1.21]* |
| I71 | 47,430 | 81,217,710 | 0.59 | 0.58 | 0.53 | 0.01 | 0.06 | 0.06 | 1.11 [1.07-1.15]* | 1.11 [1.08-1.15]* |
| I72 | 11,840 | 81,486,870 | 0.14 | 0.15 | 0.13 | 0.00 | 0.01 | 0.01 | 1.11 [1.04-1.18]* | 1.10 [1.03-1.17]* |
| I73 | 87,780 | 80,614,700 | 1.24 | 1.05 | 0.96 | 0.18 | 0.28 | 0.25 | 1.29 [1.26-1.32]* | 1.25 [1.22-1.28]* |
| I74 | 22,460 | 81,355,910 | 0.28 | 0.28 | 0.25 | 0.00 | 0.03 | 0.03 | 1.11 [1.06-1.17]* | 1.11 [1.06-1.17]* |

| | | | | | | | | | | |
|---|---|---|---|---|---|---|---|---|---|---|
| I77 | 30,020 | 81,232,370 | 0.40 | 0.36 | 0.33 | 0.03 | 0.07 | 0.07 | 1.22 [1.17-1.27]* | 1.21 [1.16-1.26]* |
| I78 | 7,780 | 81,493,780 | 0.10 | 0.09 | 0.08 | 0.01 | 0.02 | 0.02 | 1.24 [1.15-1.34]* | 1.21 [1.12-1.31]* |
| I79 | 3,170 | 81,578,080 | 0.04 | 0.04 | 0.03 | 0.00 | 0.01 | 0.01 | 1.19 [1.06-1.35]* | 1.15 [1.02-1.30] |
| I80 | 56,610 | 80,660,280 | 0.74 | 0.69 | 0.63 | 0.05 | 0.12 | 0.10 | 1.19 [1.15-1.22]* | 1.15 [1.12-1.19]* |
| I81 | 3,460 | 81,591,460 | 0.04 | 0.04 | 0.04 | 0.00 | 0.00 | 0.00 | 1.04 [0.92-1.17] | 1.03 [0.91-1.16] |
| I82 | 7,740 | 81,536,300 | 0.10 | 0.09 | 0.09 | 0.00 | 0.01 | 0.01 | 1.13 [1.04-1.22]* | 1.10 [1.02-1.19] |
| I83 | 63,260 | 79,639,390 | 1.00 | 0.74 | 0.66 | 0.26 | 0.34 | 0.33 | 1.52 [1.48-1.56]* | 1.48 [1.45-1.52]* |
| I84 | 100,420 | 78,371,030 | 1.37 | 1.26 | 1.09 | 0.11 | 0.28 | 0.22 | 1.25 [1.23-1.28]* | 1.19 [1.17-1.22]* |
| I85 | 9,510 | 81,505,490 | 0.13 | 0.11 | 0.10 | 0.02 | 0.04 | 0.03 | 1.37 [1.28-1.47]* | 1.34 [1.25-1.43]* |
| I86 | 7,570 | 81,471,110 | 0.10 | 0.09 | 0.08 | 0.01 | 0.02 | 0.02 | 1.24 [1.15-1.34]* | 1.21 [1.12-1.31]* |
| I87 | 15,640 | 81,481,180 | 0.24 | 0.18 | 0.16 | 0.06 | 0.08 | 0.08 | 1.49 [1.42-1.57]* | 1.45 [1.38-1.53]* |
| I88 | 11,850 | 81,361,660 | 0.19 | 0.13 | 0.16 | 0.06 | 0.04 | 0.03 | 1.23 [1.16-1.31]* | 1.20 [1.13-1.28]* |
| I89 | 21,140 | 81,449,470 | 0.31 | 0.25 | 0.22 | 0.07 | 0.09 | 0.08 | 1.42 [1.35-1.48]* | 1.37 [1.31-1.43]* |
| I95 | 197,290 | 80,402,540 | 2.58 | 2.42 | 2.25 | 0.15 | 0.33 | 0.29 | 1.15 [1.13-1.16]* | 1.13 [1.11-1.15]* |
| I97 | 2,450 | 81,550,980 | 0.03 | 0.03 | 0.03 | 0.00 | 0.00 | 0.00 | 1.17 [1.02-1.34] | 1.17 [1.02-1.34] |
| I98 | 6,550 | 81,556,540 | 0.09 | 0.08 | 0.07 | 0.01 | 0.02 | 0.02 | 1.36 [1.26-1.48]* | 1.31 [1.21-1.43]* |
| I99 | 3,590 | 81,568,850 | 0.04 | 0.04 | 0.04 | 0.00 | 0.01 | 0.00 | 1.13 [1.00-1.27] | 1.11 [0.99-1.25] |
| hospitalised - J00-J99 | | | | | | | | | | |
| J00 | 12,330 | 81,270,110 | 0.20 | 0.14 | 0.15 | 0.06 | 0.05 | 0.05 | 1.36 [1.28-1.44]* | 1.31 [1.23-1.39]* |
| J01 | 3,070 | 81,543,010 | 0.04 | 0.04 | 0.03 | 0.01 | 0.01 | 0.01 | 1.28 [1.14-1.44]* | 1.19 [1.05-1.34]* |
| J02 | 22,430 | 81,142,530 | 0.33 | 0.26 | 0.27 | 0.07 | 0.06 | 0.05 | 1.23 [1.18-1.28]* | 1.18 [1.13-1.23]* |
| J03 | 108,380 | 79,148,740 | 1.70 | 1.29 | 1.46 | 0.42 | 0.25 | 0.20 | 1.17 [1.15-1.19]* | 1.14 [1.11-1.16]* |
| J04 | 2,750 | 81,550,670 | 0.04 | 0.03 | 0.03 | 0.01 | 0.01 | 0.01 | 1.33 [1.17-1.50]* | 1.23 [1.08-1.40]* |
| J05 | 20,010 | 80,887,380 | 0.27 | 0.24 | 0.27 | 0.02 | 0.00 | −0.01 | 0.98 [0.94-1.03] | 0.97 [0.92-1.02] |
| J06 | 76,210 | 78,184,710 | 1.23 | 0.91 | 1.00 | 0.32 | 0.24 | 0.20 | 1.24 [1.21-1.27]* | 1.19 [1.17-1.22]* |
| J10 | 19,480 | 81,483,750 | 0.28 | 0.23 | 0.22 | 0.05 | 0.06 | 0.04 | 1.28 [1.22-1.34]* | 1.18 [1.13-1.24]* |
| J11 | 3,970 | 81,536,410 | 0.06 | 0.05 | 0.04 | 0.01 | 0.01 | 0.01 | 1.26 [1.13-1.40]* | 1.18 [1.06-1.32]* |
| J12 | 29,960 | 81,533,820 | 0.41 | 0.36 | 0.34 | 0.06 | 0.08 | 0.06 | 1.23 [1.18-1.28]* | 1.17 [1.12-1.22]* |
| J13 | 6,360 | 81,528,740 | 0.09 | 0.08 | 0.07 | 0.01 | 0.02 | 0.01 | 1.27 [1.17-1.38]* | 1.19 [1.09-1.29]* |
| J14 | 2,750 | 81,581,780 | 0.04 | 0.03 | 0.03 | 0.00 | 0.01 | 0.00 | 1.23 [1.08-1.40]* | 1.11 [0.97-1.26] |
| J15 | 14,180 | 81,453,670 | 0.20 | 0.17 | 0.15 | 0.04 | 0.05 | 0.04 | 1.32 [1.24-1.39]* | 1.23 [1.16-1.30]* |
| J17 | 2,820 | 81,583,460 | 0.04 | 0.03 | 0.03 | 0.01 | 0.01 | 0.01 | 1.35 [1.20-1.53]* | 1.28 [1.13-1.45]* |
| J18 | 402,130 | 78,500,770 | 5.65 | 4.99 | 4.67 | 0.66 | 0.98 | 0.83 | 1.21 [1.20-1.22]* | 1.17 [1.16-1.18]* |
| J20 | 5,620 | 81,499,840 | 0.08 | 0.07 | 0.06 | 0.01 | 0.01 | 0.01 | 1.22 [1.12-1.34]* | 1.15 [1.05-1.26]* |
| J21 | 4,670 | 79,843,560 | 0.07 | 0.06 | 0.06 | 0.01 | 0.01 | 0.00 | 1.09 [0.99-1.20] | 1.03 [0.93-1.13] |
| J22 | 263,300 | 77,974,730 | 3.95 | 3.23 | 3.09 | 0.72 | 0.87 | 0.71 | 1.28 [1.26-1.30]* | 1.22 [1.20-1.23]* |
| J30 | 45,100 | 81,034,840 | 0.95 | 0.46 | 0.47 | 0.49 | 0.48 | 0.38 | 2.03 [1.98-2.09]* | 1.66 [1.61-1.71]* |
| J31 | 10,340 | 81,371,670 | 0.18 | 0.11 | 0.11 | 0.06 | 0.07 | 0.05 | 1.62 [1.52-1.72]* | 1.34 [1.26-1.43]* |
| J32 | 24,280 | 81,027,590 | 0.35 | 0.29 | 0.26 | 0.06 | 0.09 | 0.05 | 1.32 [1.27-1.38]* | 1.17 [1.12-1.22]* |
| J33 | 14,260 | 81,018,130 | 0.21 | 0.17 | 0.15 | 0.04 | 0.06 | 0.04 | 1.42 [1.35-1.50]* | 1.20 [1.14-1.27]* |
| J34 | 51,120 | 79,591,450 | 0.80 | 0.60 | 0.57 | 0.19 | 0.22 | 0.17 | 1.39 [1.35-1.43]* | 1.26 [1.23-1.30]* |
| J35 | 79,650 | 78,709,280 | 1.33 | 0.93 | 1.07 | 0.39 | 0.26 | 0.22 | 1.24 [1.21-1.27]* | 1.20 [1.17-1.23]* |
| J36 | 8,570 | 81,334,380 | 0.11 | 0.10 | 0.10 | 0.01 | 0.01 | 0.01 | 1.07 [1.00-1.15] | 1.06 [0.99-1.15] |
| J37 | 580 | 81,589,640 | 0.01 | 0.01 | 0.01 | 0.00 | 0.00 | 0.00 | 1.19 [0.90-1.58] | 1.09 [0.82-1.45] |
| J38 | 18,970 | 81,182,600 | 0.25 | 0.23 | 0.21 | 0.02 | 0.04 | 0.03 | 1.21 [1.15-1.27]* | 1.15 [1.09-1.21]* |
| J39 | 10,390 | 81,428,470 | 0.14 | 0.12 | 0.12 | 0.02 | 0.02 | 0.02 | 1.20 [1.12-1.28]* | 1.12 [1.05-1.20]* |
| J40 | 11,310 | 81,436,530 | 0.16 | 0.13 | 0.12 | 0.02 | 0.04 | 0.03 | 1.30 [1.23-1.39]* | 1.19 [1.12-1.27]* |
| J41 | 540 | 81,602,340 | 0.01 | 0.01 | 0.01 | 0.00 | 0.00 | 0.00 | 1.18 [0.88-1.57] | 1.13 [0.84-1.52] |
| J42 | 4,580 | 81,539,790 | 0.06 | 0.05 | 0.05 | 0.01 | 0.02 | 0.01 | 1.31 [1.19-1.44]* | 1.17 [1.05-1.29]* |
| J43 | 62,250 | 81,136,800 | 0.85 | 0.75 | 0.67 | 0.11 | 0.19 | 0.15 | 1.28 [1.25-1.32]* | 1.21 [1.18-1.25]* |
| J44 | 237,140 | 79,075,120 | 3.51 | 2.87 | 2.59 | 0.64 | 0.93 | 0.64 | 1.36 [1.34-1.38]* | 1.22 [1.21-1.24]* |
| J45 | 385,090 | 69,544,600 | 8.73 | 4.73 | 4.51 | 3.99 | 4.21 | 2.96 | 1.93 [1.91-1.95]* | 1.51 [1.50-1.53]* |
| J46 | 8,330 | 81,244,880 | 0.21 | 0.08 | 0.08 | 0.13 | 0.13 | 0.10 | 2.59 [2.43-2.76]* | 2.01 [1.88-2.15]* |
| J47 | 46,340 | 81,234,330 | 0.69 | 0.54 | 0.49 | 0.15 | 0.19 | 0.12 | 1.39 [1.35-1.44]* | 1.22 [1.18-1.26]* |
| J61 | 6,350 | 81,569,760 | 0.09 | 0.08 | 0.07 | 0.01 | 0.02 | 0.02 | 1.32 [1.21-1.43]* | 1.25 [1.15-1.37]* |
| J67 | 1,780 | 81,594,100 | 0.03 | 0.02 | 0.02 | 0.01 | 0.01 | 0.01 | 1.44 [1.24-1.67]* | 1.30 [1.11-1.52]* |
| J69 | 61,760 | 81,434,640 | 0.79 | 0.75 | 0.70 | 0.04 | 0.09 | 0.09 | 1.12 [1.09-1.16]* | 1.13 [1.10-1.16]* |
| J70 | 1,280 | 81,602,840 | 0.02 | 0.01 | 0.01 | 0.00 | 0.00 | 0.00 | 1.35 [1.13-1.62]* | 1.28 [1.06-1.54]* |
| J80 | 4,710 | 81,571,130 | 0.06 | 0.06 | 0.05 | 0.01 | 0.01 | 0.01 | 1.27 [1.15-1.40]* | 1.25 [1.13-1.38]* |
| J81 | 20,970 | 81,448,390 | 0.27 | 0.25 | 0.23 | 0.02 | 0.04 | 0.04 | 1.19 [1.14-1.25]* | 1.17 [1.11-1.23]* |
| J82 | 1,730 | 81,598,430 | 0.04 | 0.02 | 0.02 | 0.02 | 0.02 | 0.01 | 2.24 [1.93-2.59]* | 1.69 [1.44-1.98]* |
| J84 | 38,840 | 81,407,170 | 0.57 | 0.46 | 0.41 | 0.11 | 0.15 | 0.14 | 1.37 [1.32-1.42]* | 1.32 [1.27-1.36]* |
| J85 | 2,070 | 81,589,020 | 0.03 | 0.02 | 0.02 | 0.00 | 0.01 | 0.00 | 1.23 [1.06-1.42]* | 1.18 [1.01-1.37] |
| J86 | 7,140 | 81,514,590 | 0.09 | 0.09 | 0.08 | 0.00 | 0.01 | 0.01 | 1.13 [1.05-1.23]* | 1.13 [1.04-1.23]* |
| J90 | 193,490 | 80,481,220 | 2.51 | 2.38 | 2.21 | 0.13 | 0.31 | 0.28 | 1.14 [1.12-1.16]* | 1.12 [1.11-1.14]* |
| J91 | 4,560 | 81,588,560 | 0.06 | 0.05 | 0.05 | 0.01 | 0.01 | 0.01 | 1.20 [1.09-1.34]* | 1.19 [1.08-1.32]* |
| J92 | 22,160 | 81,485,720 | 0.29 | 0.27 | 0.24 | 0.03 | 0.05 | 0.05 | 1.21 [1.15-1.27]* | 1.19 [1.13-1.25]* |
| J93 | 17,710 | 81,364,400 | 0.22 | 0.22 | 0.20 | 0.00 | 0.02 | 0.01 | 1.08 [1.02-1.14]* | 1.07 [1.01-1.12] |
| J94 | 7,400 | 81,556,750 | 0.09 | 0.09 | 0.08 | 0.00 | 0.01 | 0.01 | 1.09 [1.01-1.18] | 1.08 [0.99-1.17] |
| J95 | 5,870 | 81,509,350 | 0.07 | 0.07 | 0.06 | 0.00 | 0.01 | 0.01 | 1.11 [1.01-1.21] | 1.10 [1.00-1.20] |
| J96 | 127,850 | 81,090,280 | 1.74 | 1.54 | 1.40 | 0.20 | 0.34 | 0.27 | 1.24 [1.22-1.26]* | 1.18 [1.16-1.21]* |
| J98 | 122,090 | 80,669,870 | 1.62 | 1.49 | 1.37 | 0.13 | 0.25 | 0.19 | 1.18 [1.16-1.21]* | 1.14 [1.11-1.16]* |
| J99 | 3,880 | 81,575,380 | 0.06 | 0.04 | 0.04 | 0.01 | 0.02 | 0.01 | 1.44 [1.30-1.60]* | 1.28 [1.15-1.42]* |
| hospitalised - K00-K95 | | | | | | | | | | |
| K00 | 20,070 | 81,116,610 | 0.31 | 0.23 | 0.28 | 0.08 | 0.03 | 0.02 | 1.09 [1.05-1.15]* | 1.07 [1.02-1.13]* |
| K01 | 44,940 | 79,720,850 | 0.65 | 0.54 | 0.57 | 0.10 | 0.08 | 0.06 | 1.13 [1.10-1.17]* | 1.10 [1.06-1.13]* |
| K02 | 155,560 | 77,937,500 | 2.23 | 1.94 | 2.12 | 0.29 | 0.10 | 0.06 | 1.05 [1.03-1.07]* | 1.03 [1.01-1.05]* |
| K03 | 4,190 | 81,543,490 | 0.06 | 0.05 | 0.06 | 0.01 | 0.01 | 0.00 | 1.11 [1.00-1.23] | 1.07 [0.97-1.19] |
| K04 | 35,360 | 80,753,310 | 0.46 | 0.43 | 0.43 | 0.03 | 0.03 | 0.02 | 1.07 [1.03-1.11]* | 1.04 [1.00-1.08] |
| K05 | 29,470 | 80,848,610 | 0.40 | 0.36 | 0.34 | 0.04 | 0.06 | 0.05 | 1.19 [1.15-1.24]* | 1.15 [1.10-1.19]* |
| K06 | 4,340 | 81,528,490 | 0.06 | 0.05 | 0.05 | 0.01 | 0.01 | 0.01 | 1.24 [1.12-1.37]* | 1.19 [1.07-1.32]* |
| K07 | 28,280 | 80,772,930 | 0.42 | 0.33 | 0.38 | 0.09 | 0.05 | 0.04 | 1.12 [1.08-1.17]* | 1.09 [1.05-1.14]* |
| K08 | 49,670 | 80,505,860 | 0.66 | 0.61 | 0.59 | 0.06 | 0.07 | 0.05 | 1.12 [1.08-1.15]* | 1.08 [1.05-1.12]* |
| K09 | 3,380 | 81,489,100 | 0.05 | 0.04 | 0.04 | 0.00 | 0.01 | 0.01 | 1.17 [1.05-1.32]* | 1.15 [1.02-1.29] |
| K10 | 5,280 | 81,530,860 | 0.07 | 0.06 | 0.06 | 0.00 | 0.01 | 0.01 | 1.15 [1.04-1.26]* | 1.10 [1.00-1.21] |
| K11 | 11,490 | 81,351,840 | 0.16 | 0.14 | 0.13 | 0.02 | 0.03 | 0.03 | 1.24 [1.16-1.32]* | 1.19 [1.12-1.27]* |
| K12 | 18,940 | 81,355,520 | 0.27 | 0.22 | 0.22 | 0.04 | 0.05 | 0.04 | 1.23 [1.17-1.29]* | 1.20 [1.14-1.26]* |
| K13 | 23,250 | 81,082,660 | 0.34 | 0.27 | 0.26 | 0.06 | 0.08 | 0.07 | 1.30 [1.25-1.36]* | 1.26 [1.20-1.31]* |
| K14 | 12,090 | 81,387,660 | 0.17 | 0.14 | 0.13 | 0.02 | 0.04 | 0.03 | 1.29 [1.21-1.37]* | 1.21 [1.14-1.29]* |
| K20 | 74,210 | 80,000,990 | 1.00 | 0.91 | 0.82 | 0.09 | 0.19 | 0.13 | 1.23 [1.20-1.26]* | 1.15 [1.12-1.18]* |
| K21 | 293,970 | 76,908,000 | 4.20 | 3.73 | 3.37 | 0.48 | 0.84 | 0.54 | 1.25 [1.23-1.26]* | 1.15 [1.13-1.16]* |
| K22 | 108,790 | 79,821,530 | 1.45 | 1.34 | 1.20 | 0.11 | 0.25 | 0.19 | 1.21 [1.18-1.23]* | 1.15 [1.13-1.17]* |
| K23 | 1,370 | 81,599,500 | 0.02 | 0.02 | 0.01 | 0.00 | 0.01 | 0.01 | 1.47 [1.23-1.75]* | 1.33 [1.11-1.60]* |
| K25 | 53,830 | 80,693,520 | 0.71 | 0.66 | 0.59 | 0.05 | 0.12 | 0.10 | 1.21 [1.17-1.24]* | 1.16 [1.13-1.20]* |
| K26 | 37,060 | 80,917,980 | 0.47 | 0.46 | 0.41 | 0.01 | 0.06 | 0.06 | 1.15 [1.11-1.19]* | 1.14 [1.10-1.18]* |
| K27 | 7,060 | 81,498,750 | 0.09 | 0.09 | 0.08 | 0.00 | 0.01 | 0.01 | 1.18 [1.09-1.28]* | 1.13 [1.04-1.23]* |
| K28 | 1,050 | 81,597,060 | 0.02 | 0.01 | 0.01 | 0.00 | 0.00 | 0.00 | 1.43 [1.17-1.75]* | 1.29 [1.05-1.59] |
| K29 | 274,710 | 76,266,830 | 3.92 | 3.52 | 3.21 | 0.40 | 0.71 | 0.54 | 1.22 [1.21-1.24]* | 1.16 [1.15-1.18]* |
| K30 | 65,390 | 79,576,250 | 0.89 | 0.81 | 0.71 | 0.08 | 0.18 | 0.13 | 1.25 [1.22-1.28]* | 1.16 [1.13-1.20]* |
| K31 | 97,970 | 80,488,750 | 1.30 | 1.20 | 1.08 | 0.10 | 0.22 | 0.15 | 1.21 [1.18-1.23]* | 1.13 [1.10-1.15]* |
| K35 | 47,670 | 80,165,410 | 0.62 | 0.59 | 0.61 | 0.03 | 0.01 | 0.00 | 1.01 [0.98-1.04] | 0.99 [0.96-1.03] |
| K36 | 1,100 | 81,581,340 | 0.01 | 0.01 | 0.01 | 0.00 | 0.00 | 0.00 | 1.15 [0.94-1.40] | 1.12 [0.91-1.37] |
| K37 | 9,380 | 81,349,570 | 0.13 | 0.11 | 0.12 | 0.02 | 0.01 | 0.00 | 1.06 [0.99-1.13] | 1.02 [0.95-1.09] |
| K38 | 6,680 | 81,476,850 | 0.09 | 0.08 | 0.08 | 0.01 | 0.00 | 0.00 | 1.06 [0.98-1.15] | 1.04 [0.95-1.13] |
| K40 | 93,390 | 78,560,130 | 1.19 | 1.19 | 1.12 | 0.00 | 0.07 | 0.04 | 1.06 [1.04-1.09]* | 1.04 [1.02-1.06]* |
| K41 | 7,330 | 81,459,960 | 0.09 | 0.09 | 0.08 | 0.00 | 0.01 | 0.01 | 1.12 [1.03-1.21]* | 1.07 [0.98-1.16] |
| K42 | 48,630 | 80,553,380 | 0.65 | 0.59 | 0.54 | 0.05 | 0.11 | 0.07 | 1.20 [1.17-1.24]* | 1.13 [1.09-1.16]* |

| | | | | | | | | | | |
|---|---|---|---|---|---|---|---|---|---|---|
| K43 | 40,740 | 80,887,220 | 0.52 | 0.50 | 0.44 | 0.02 | 0.08 | 0.06 | 1.18 [1.14-1.22]* | 1.12 [1.09-1.16]* |
| K44 | 264,170 | 76,819,440 | 3.71 | 3.37 | 3.05 | 0.34 | 0.66 | 0.45 | 1.22 [1.20-1.23]* | 1.14 [1.12-1.15]* |
| K45 | 1,890 | 81,584,640 | 0.02 | 0.02 | 0.02 | 0.00 | 0.00 | 0.00 | 1.12 [0.96-1.32] | 1.04 [0.88-1.23] |
| K46 | 7,950 | 81,542,930 | 0.10 | 0.10 | 0.09 | 0.01 | 0.02 | 0.01 | 1.20 [1.11-1.30]* | 1.12 [1.04-1.21]* |
| K50 | 20,250 | 81,049,270 | 0.33 | 0.23 | 0.21 | 0.09 | 0.12 | 0.09 | 1.55 [1.48-1.62]* | 1.38 [1.32-1.45]* |
| K51 | 28,680 | 80,863,330 | 0.42 | 0.34 | 0.31 | 0.08 | 0.11 | 0.08 | 1.36 [1.31-1.42]* | 1.25 [1.20-1.30]* |
| K52 | 157,320 | 76,930,080 | 2.26 | 1.99 | 1.81 | 0.26 | 0.45 | 0.37 | 1.25 [1.23-1.27]* | 1.20 [1.18-1.22]* |
| K55 | 24,820 | 81,412,770 | 0.32 | 0.30 | 0.28 | 0.02 | 0.04 | 0.04 | 1.15 [1.10-1.21]* | 1.13 [1.08-1.19]* |
| K56 | 75,850 | 80,772,830 | 0.97 | 0.93 | 0.85 | 0.03 | 0.12 | 0.09 | 1.14 [1.11-1.17]* | 1.10 [1.07-1.13]* |
| K57 | 304,890 | 77,511,870 | 4.12 | 3.89 | 3.53 | 0.23 | 0.59 | 0.42 | 1.17 [1.15-1.18]* | 1.11 [1.10-1.13]* |
| K58 | 92,310 | 80,313,960 | 1.38 | 1.09 | 0.99 | 0.28 | 0.38 | 0.25 | 1.39 [1.36-1.42]* | 1.22 [1.19-1.25]* |
| K59 | 309,970 | 78,138,110 | 4.39 | 3.86 | 3.67 | 0.53 | 0.73 | 0.58 | 1.20 [1.18-1.21]* | 1.15 [1.14-1.17]* |
| K60 | 25,530 | 80,789,230 | 0.38 | 0.30 | 0.27 | 0.08 | 0.11 | 0.10 | 1.42 [1.37-1.48]* | 1.35 [1.30-1.41]* |
| K61 | 15,240 | 81,109,590 | 0.21 | 0.18 | 0.17 | 0.03 | 0.05 | 0.04 | 1.29 [1.22-1.36]* | 1.26 [1.19-1.33]* |
| K62 | 157,110 | 77,845,850 | 2.16 | 1.98 | 1.76 | 0.17 | 0.40 | 0.32 | 1.22 [1.20-1.25]* | 1.17 [1.15-1.19]* |
| K63 | 166,330 | 79,651,920 | 2.18 | 2.07 | 1.83 | 0.11 | 0.34 | 0.26 | 1.19 [1.17-1.21]* | 1.14 [1.12-1.16]* |
| K64 | 121,010 | 81,082,530 | 1.61 | 1.46 | 1.31 | 0.15 | 0.30 | 0.22 | 1.22 [1.20-1.25]* | 1.16 [1.14-1.18]* |
| K65 | 17,640 | 81,428,450 | 0.22 | 0.22 | 0.19 | 0.00 | 0.02 | 0.02 | 1.12 [1.07-1.18]* | 1.11 [1.06-1.17]* |
| K66 | 64,960 | 80,792,520 | 0.81 | 0.80 | 0.73 | 0.01 | 0.08 | 0.06 | 1.11 [1.08-1.14]* | 1.08 [1.05-1.11]* |
| K70 | 20,580 | 81,331,930 | 0.28 | 0.25 | 0.21 | 0.04 | 0.07 | 0.07 | 1.35 [1.29-1.41]* | 1.34 [1.28-1.41]* |
| K71 | 1,420 | 81,591,530 | 0.02 | 0.02 | 0.02 | 0.00 | 0.00 | 0.00 | 1.18 [0.98-1.41] | 1.16 [0.97-1.39] |
| K72 | 13,290 | 81,546,100 | 0.18 | 0.16 | 0.14 | 0.02 | 0.04 | 0.04 | 1.30 [1.23-1.38]* | 1.27 [1.20-1.35]* |
| K73 | 1,310 | 81,577,450 | 0.02 | 0.02 | 0.01 | 0.00 | 0.01 | 0.00 | 1.40 [1.17-1.67]* | 1.32 [1.10-1.59]* |
| K74 | 20,290 | 81,438,270 | 0.29 | 0.24 | 0.21 | 0.05 | 0.08 | 0.07 | 1.37 [1.31-1.43]* | 1.32 [1.26-1.38]* |
| K75 | 13,090 | 81,470,520 | 0.19 | 0.15 | 0.14 | 0.03 | 0.05 | 0.04 | 1.33 [1.26-1.41]* | 1.28 [1.20-1.35]* |
| K76 | 96,960 | 80,852,320 | 1.31 | 1.17 | 1.05 | 0.13 | 0.26 | 0.20 | 1.25 [1.22-1.28]* | 1.19 [1.16-1.21]* |
| K80 | 172,840 | 78,512,740 | 2.19 | 2.20 | 2.00 | −0.01 | 0.19 | 0.13 | 1.10 [1.08-1.12]* | 1.06 [1.04-1.08]* |
| K81 | 32,140 | 81,008,170 | 0.40 | 0.40 | 0.35 | 0.01 | 0.05 | 0.04 | 1.13 [1.09-1.18]* | 1.10 [1.05-1.14]* |
| K82 | 24,800 | 81,230,060 | 0.30 | 0.31 | 0.27 | 0.00 | 0.03 | 0.02 | 1.11 [1.06-1.16]* | 1.07 [1.02-1.12]* |
| K83 | 46,670 | 81,107,480 | 0.58 | 0.57 | 0.52 | 0.01 | 0.06 | 0.05 | 1.12 [1.09-1.16]* | 1.10 [1.06-1.14]* |
| K85 | 30,510 | 81,088,150 | 0.38 | 0.38 | 0.33 | 0.00 | 0.04 | 0.04 | 1.13 [1.09-1.18]* | 1.10 [1.06-1.15]* |
| K86 | 26,820 | 81,342,850 | 0.34 | 0.33 | 0.30 | 0.01 | 0.04 | 0.03 | 1.14 [1.10-1.19]* | 1.11 [1.07-1.16]* |
| K90 | 25,860 | 81,075,820 | 0.46 | 0.28 | 0.28 | 0.18 | 0.18 | 0.15 | 1.67 [1.61-1.73]* | 1.49 [1.43-1.55]* |
| K91 | 24,720 | 81,262,820 | 0.32 | 0.30 | 0.27 | 0.02 | 0.05 | 0.04 | 1.19 [1.14-1.24]* | 1.13 [1.09-1.18]* |
| K92 | 207,230 | 79,207,000 | 2.80 | 2.57 | 2.34 | 0.23 | 0.46 | 0.38 | 1.20 [1.18-1.22]* | 1.16 [1.14-1.17]* |
| hospitalised - L00-L99 | | | | | | | | | | |
| L00 | 540 | 81,568,910 | 0.01 | 0.01 | 0.01 | 0.00 | 0.00 | 0.00 | 1.36 [1.04-1.77] | 1.33 [1.01-1.74] |
| L01 | 3,140 | 81,487,780 | 0.07 | 0.03 | 0.03 | 0.04 | 0.04 | 0.04 | 2.16 [1.95-2.39]* | 2.01 [1.81-2.23]* |
| L02 | 43,330 | 80,534,190 | 0.61 | 0.52 | 0.49 | 0.09 | 0.13 | 0.12 | 1.26 [1.22-1.30]* | 1.23 [1.19-1.27]* |
| L03 | 158,270 | 79,318,120 | 2.54 | 1.86 | 1.70 | 0.68 | 0.84 | 0.78 | 1.49 [1.47-1.52]* | 1.44 [1.42-1.46]* |
| L04 | 4,470 | 81,501,380 | 0.07 | 0.05 | 0.06 | 0.01 | 0.01 | 0.01 | 1.16 [1.06-1.28]* | 1.15 [1.05-1.27]* |
| L05 | 12,140 | 81,015,590 | 0.16 | 0.15 | 0.15 | 0.02 | 0.01 | 0.01 | 1.09 [1.02-1.16]* | 1.09 [1.02-1.16] |
| L08 | 44,280 | 81,095,370 | 0.68 | 0.51 | 0.47 | 0.17 | 0.21 | 0.20 | 1.45 [1.41-1.50]* | 1.42 [1.38-1.47]* |
| L10 | 500 | 81,603,670 | 0.01 | 0.01 | 0.00 | 0.00 | 0.00 | 0.00 | 2.00 [1.52-2.64]* | 1.86 [1.40-2.46]* |
| L11 | 410 | 81,603,630 | 0.01 | 0.00 | 0.00 | 0.00 | 0.00 | 0.00 | 1.64 [1.20-2.25]* | 1.58 [1.15-2.17]* |
| L12 | 3,500 | 81,583,340 | 0.08 | 0.03 | 0.03 | 0.04 | 0.05 | 0.05 | 2.64 [2.37-2.93]* | 2.56 [2.30-2.85]* |
| L13 | 610 | 81,598,590 | 0.01 | 0.01 | 0.01 | 0.01 | 0.01 | 0.01 | 2.83 [2.23-3.59]* | 2.63 [2.06-3.35]* |
| L20 | 6,010 | 81,313,990 | 0.32 | 0.01 | 0.01 | 0.31 | 0.31 | 0.30 | 22.58 [20.37-25.03]* | 19.70 [17.74-21.86]* |
| L21 | 1,740 | 81,550,310 | 0.04 | 0.02 | 0.01 | 0.03 | 0.03 | 0.03 | 2.86 [2.48-3.29]* | 2.72 [2.36-3.15]* |
| L22 | 1,990 | 81,502,350 | 0.03 | 0.02 | 0.02 | 0.00 | 0.00 | 0.00 | 1.01 [0.86-1.18] | 0.99 [0.84-1.16] |
| L23 | 2,010 | 81,557,180 | 0.06 | 0.02 | 0.02 | 0.04 | 0.04 | 0.04 | 3.34 [2.94-3.78]* | 3.01 [2.64-3.42]* |
| L24 | 10,460 | 81,588,620 | 0.16 | 0.12 | 0.11 | 0.03 | 0.04 | 0.04 | 1.36 [1.27-1.45]* | 1.34 [1.25-1.44]* |
| L25 | 1,880 | 81,576,020 | 0.04 | 0.02 | 0.02 | 0.03 | 0.03 | 0.03 | 2.69 [2.35-3.08]* | 2.61 [2.27-2.99]* |
| L26 | 190 | 81,607,090 | 0.01 | 0.00 | 0.00 | 0.01 | 0.01 | 0.01 | 6.43 [4.17-9.91]* | 6.03 [3.88-9.38]* |
| L27 | 9,970 | 81,459,700 | 0.17 | 0.11 | 0.10 | 0.06 | 0.07 | 0.06 | 1.62 [1.52-1.73]* | 1.53 [1.43-1.63]* |
| L28 | 3,240 | 81,544,760 | 0.09 | 0.03 | 0.02 | 0.07 | 0.07 | 0.07 | 3.75 [3.39-4.14]* | 3.51 [3.17-3.89]* |
| L29 | 16,050 | 81,299,320 | 0.27 | 0.18 | 0.17 | 0.09 | 0.10 | 0.09 | 1.59 [1.51-1.67]* | 1.50 [1.42-1.58]* |
| L30 | 95,050 | 79,174,070 | 4.24 | 0.47 | 0.46 | 3.78 | 3.78 | 3.74 | 9.24 [9.06-9.43]* | 8.55 [8.37-8.72]* |
| L40 | 37,810 | 81,058,980 | 0.74 | 0.40 | 0.35 | 0.34 | 0.39 | 0.38 | 2.11 [2.04-2.17]* | 2.04 [1.98-2.11]* |
| L42 | 660 | 81,600,970 | 0.01 | 0.01 | 0.01 | 0.00 | 0.00 | 0.00 | 1.42 [1.10-1.83]* | 1.32 [1.02-1.72] |
| L43 | 4,160 | 81,526,160 | 0.08 | 0.04 | 0.04 | 0.03 | 0.04 | 0.04 | 1.99 [1.81-2.18]* | 1.89 [1.72-2.08]* |
| L50 | 11,590 | 81,366,990 | 0.23 | 0.12 | 0.12 | 0.11 | 0.11 | 0.09 | 1.89 [1.79-2.00]* | 1.60 [1.51-1.70]* |
| L51 | 2,740 | 81,533,400 | 0.04 | 0.03 | 0.03 | 0.01 | 0.01 | 0.01 | 1.24 [1.10-1.40]* | 1.18 [1.05-1.34]* |
| L52 | 1,060 | 81,581,960 | 0.02 | 0.01 | 0.01 | 0.00 | 0.00 | 0.00 | 1.36 [1.12-1.66]* | 1.24 [1.01-1.52] |
| L53 | 14,760 | 81,409,400 | 0.25 | 0.17 | 0.15 | 0.08 | 0.09 | 0.09 | 1.61 [1.53-1.69]* | 1.56 [1.47-1.64]* |
| L55 | 460 | 81,600,140 | 0.01 | 0.01 | 0.01 | 0.00 | 0.00 | 0.00 | 1.12 [0.81-1.53] | 1.10 [0.79-1.52] |
| L56 | 560 | 81,598,890 | 0.01 | 0.01 | 0.01 | 0.01 | 0.01 | 0.01 | 2.36 [1.85-3.00]* | 2.22 [1.73-2.86]* |
| L57 | 31,910 | 81,190,610 | 0.43 | 0.38 | 0.35 | 0.05 | 0.09 | 0.07 | 1.25 [1.20-1.30]* | 1.20 [1.15-1.24]* |
| L59 | 470 | 81,603,390 | 0.01 | 0.01 | 0.01 | 0.00 | 0.00 | 0.00 | 1.21 [0.89-1.65] | 1.25 [0.91-1.72] |
| L60 | 11,930 | 81,135,920 | 0.18 | 0.14 | 0.14 | 0.04 | 0.05 | 0.04 | 1.33 [1.25-1.41]* | 1.24 [1.17-1.32]* |
| L63 | 630 | 81,599,290 | 0.02 | 0.01 | 0.01 | 0.01 | 0.01 | 0.01 | 2.70 [2.15-3.40]* | 2.19 [1.71-2.79]* |
| L65 | 2,280 | 81,578,110 | 0.04 | 0.02 | 0.02 | 0.02 | 0.02 | 0.02 | 1.89 [1.66-2.15]* | 1.68 [1.47-1.92]* |
| L66 | 460 | 81,601,690 | 0.01 | 0.00 | 0.00 | 0.00 | 0.00 | 0.00 | 1.72 [1.29-2.29]* | 1.62 [1.21-2.17]* |
| L68 | 760 | 81,588,530 | 0.01 | 0.01 | 0.01 | 0.00 | 0.00 | 0.00 | 1.10 [0.86-1.41] | 1.05 [0.82-1.35] |
| L70 | 2,830 | 81,569,000 | 0.04 | 0.03 | 0.03 | 0.01 | 0.01 | 0.01 | 1.29 [1.14-1.46]* | 1.21 [1.07-1.37]* |
| L71 | 3,200 | 81,570,380 | 0.05 | 0.04 | 0.03 | 0.01 | 0.02 | 0.01 | 1.50 [1.34-1.68]* | 1.43 [1.27-1.60]* |
| L72 | 43,240 | 79,909,640 | 0.53 | 0.54 | 0.49 | −0.01 | 0.04 | 0.03 | 1.09 [1.06-1.13]* | 1.07 [1.03-1.10]* |
| L73 | 6,240 | 81,455,450 | 0.09 | 0.07 | 0.07 | 0.02 | 0.03 | 0.02 | 1.37 [1.26-1.49]* | 1.31 [1.20-1.42]* |
| L80 | 1,440 | 81,591,570 | 0.03 | 0.02 | 0.01 | 0.01 | 0.01 | 0.01 | 1.83 [1.56-2.15]* | 1.73 [1.46-2.04]* |
| L81 | 7,390 | 81,450,080 | 0.10 | 0.09 | 0.08 | 0.01 | 0.02 | 0.02 | 1.24 [1.15-1.34]* | 1.19 [1.10-1.29]* |
| L82 | 20,650 | 81,194,070 | 0.26 | 0.25 | 0.22 | 0.01 | 0.04 | 0.03 | 1.18 [1.13-1.24]* | 1.13 [1.07-1.18]* |
| L83 | 1,400 | 81,583,830 | 0.02 | 0.02 | 0.02 | 0.01 | 0.01 | 0.01 | 1.55 [1.31-1.83]* | 1.46 [1.23-1.74]* |
| L84 | 2,480 | 81,563,320 | 0.04 | 0.03 | 0.03 | 0.01 | 0.01 | 0.01 | 1.32 [1.16-1.51]* | 1.26 [1.10-1.44]* |
| L85 | 9,000 | 81,475,470 | 0.14 | 0.10 | 0.09 | 0.04 | 0.05 | 0.05 | 1.56 [1.46-1.67]* | 1.51 [1.41-1.62]* |
| L88 | 590 | 81,602,150 | 0.01 | 0.01 | 0.01 | 0.00 | 0.00 | 0.00 | 1.33 [1.02-1.74] | 1.21 [0.92-1.60] |
| L89 | 115,450 | 81,284,880 | 1.54 | 1.39 | 1.30 | 0.14 | 0.24 | 0.24 | 1.18 [1.16-1.21]* | 1.18 [1.16-1.21]* |
| L90 | 23,090 | 81,008,110 | 0.31 | 0.28 | 0.25 | 0.03 | 0.05 | 0.04 | 1.20 [1.15-1.26]* | 1.16 [1.11-1.21]* |
| L91 | 7,930 | 81,322,600 | 0.11 | 0.09 | 0.09 | 0.02 | 0.02 | 0.02 | 1.22 [1.13-1.32]* | 1.18 [1.09-1.27]* |
| L92 | 6,690 | 81,429,620 | 0.09 | 0.08 | 0.08 | 0.01 | 0.02 | 0.01 | 1.21 [1.11-1.31]* | 1.15 [1.06-1.25]* |
| L93 | 2,610 | 81,568,110 | 0.04 | 0.03 | 0.03 | 0.01 | 0.02 | 0.02 | 1.68 [1.49-1.90]* | 1.55 [1.36-1.75]* |
| L94 | 1,870 | 81,577,330 | 0.03 | 0.02 | 0.02 | 0.01 | 0.01 | 0.01 | 1.46 [1.26-1.69]* | 1.37 [1.18-1.59]* |
| L95 | 2,240 | 81,575,110 | 0.04 | 0.03 | 0.02 | 0.01 | 0.01 | 0.01 | 1.51 [1.32-1.73]* | 1.42 [1.23-1.63]* |
| L97 | 81,580 | 81,066,480 | 1.24 | 0.95 | 0.85 | 0.29 | 0.39 | 0.37 | 1.45 [1.42-1.49]* | 1.43 [1.40-1.46]* |
| L98 | 66,300 | 80,323,000 | 0.94 | 0.80 | 0.72 | 0.15 | 0.22 | 0.20 | 1.30 [1.27-1.34]* | 1.26 [1.23-1.29]* |
| hospitalised - M00-M99 | | | | | | | | | | |
| M00 | 8,550 | 81,467,400 | 0.12 | 0.10 | 0.09 | 0.02 | 0.03 | 0.03 | 1.35 [1.26-1.45]* | 1.31 [1.22-1.41]* |
| M02 | 2,050 | 81,569,700 | 0.03 | 0.02 | 0.02 | 0.00 | 0.00 | 0.00 | 1.17 [1.01-1.35] | 1.14 [0.98-1.32] |
| M05 | 9,850 | 81,467,650 | 0.13 | 0.12 | 0.11 | 0.01 | 0.03 | 0.02 | 1.24 [1.16-1.32]* | 1.21 [1.13-1.29]* |
| M06 | 68,390 | 80,587,150 | 0.96 | 0.82 | 0.73 | 0.14 | 0.23 | 0.18 | 1.31 [1.28-1.34]* | 1.24 [1.21-1.27]* |
| M07 | 8,350 | 81,482,310 | 0.13 | 0.09 | 0.08 | 0.04 | 0.05 | 0.04 | 1.60 [1.49-1.71]* | 1.50 [1.40-1.61]* |
| M08 | 2,740 | 81,540,120 | 0.04 | 0.03 | 0.04 | 0.01 | 0.01 | 0.01 | 1.25 [1.10-1.41]* | 1.23 [1.08-1.39]* |
| M10 | 79,130 | 80,936,360 | 1.08 | 0.95 | 0.85 | 0.13 | 0.23 | 0.20 | 1.27 [1.24-1.30]* | 1.22 [1.19-1.25]* |
| M11 | 9,170 | 81,519,410 | 0.12 | 0.11 | 0.10 | 0.01 | 0.03 | 0.02 | 1.26 [1.17-1.35]* | 1.20 [1.12-1.29]* |
| M12 | 1,980 | 81,575,720 | 0.03 | 0.02 | 0.02 | 0.00 | 0.01 | 0.00 | 1.25 [1.08-1.45]* | 1.21 [1.04-1.40] |
| M13 | 201,300 | 79,359,340 | 2.82 | 2.47 | 2.24 | 0.35 | 0.57 | 0.44 | 1.26 [1.24-1.27]* | 1.18 [1.17-1.20]* |

| | | | | | | | | | | |
|---|---|---|---|---|---|---|---|---|---|---|
| M14 | 2,380 | 81,580,580 | 0.03 | 0.03 | 0.02 | 0.00 | 0.01 | 0.01 | 1.32 [1.15-1.51]* | 1.25 [1.09-1.43]* |
| M15 | 76,670 | 80,825,760 | 1.05 | 0.92 | 0.83 | 0.13 | 0.22 | 0.17 | 1.27 [1.24-1.30]* | 1.19 [1.16-1.22]* |
| M16 | 114,100 | 79,576,170 | 1.40 | 1.44 | 1.31 | −0.04 | 0.09 | 0.06 | 1.07 [1.05-1.09]* | 1.05 [1.03-1.07]* |
| M17 | 188,250 | 78,007,630 | 2.52 | 2.39 | 2.15 | 0.14 | 0.37 | 0.27 | 1.17 [1.15-1.19]* | 1.12 [1.10-1.14]* |
| M18 | 11,450 | 81,444,030 | 0.16 | 0.14 | 0.12 | 0.02 | 0.03 | 0.02 | 1.28 [1.20-1.36]* | 1.17 [1.10-1.25]* |
| M19 | 315,810 | 78,559,980 | 4.49 | 3.90 | 3.57 | 0.59 | 0.93 | 0.71 | 1.26 [1.24-1.27]* | 1.19 [1.17-1.20]* |
| M20 | 43,580 | 80,353,420 | 0.58 | 0.53 | 0.48 | 0.04 | 0.09 | 0.07 | 1.19 [1.16-1.23]* | 1.14 [1.10-1.17]* |
| M21 | 28,510 | 81,198,690 | 0.37 | 0.35 | 0.33 | 0.02 | 0.04 | 0.03 | 1.11 [1.07-1.16]* | 1.07 [1.03-1.12]* |
| M22 | 8,240 | 81,310,780 | 0.11 | 0.10 | 0.09 | 0.01 | 0.02 | 0.02 | 1.26 [1.17-1.36]* | 1.17 [1.09-1.26]* |
| M23 | 78,520 | 78,921,090 | 0.99 | 1.00 | 0.88 | 0.00 | 0.11 | 0.06 | 1.12 [1.10-1.15]* | 1.07 [1.04-1.09]* |
| M24 | 35,360 | 80,836,880 | 0.46 | 0.43 | 0.40 | 0.03 | 0.06 | 0.04 | 1.15 [1.11-1.19]* | 1.10 [1.06-1.14]* |
| M25 | 209,400 | 78,278,930 | 2.91 | 2.62 | 2.43 | 0.29 | 0.48 | 0.37 | 1.20 [1.18-1.22]* | 1.14 [1.13-1.16]* |
| M30 | 1,800 | 81,567,190 | 0.03 | 0.02 | 0.02 | 0.01 | 0.01 | 0.01 | 1.32 [1.13-1.53]* | 1.23 [1.06-1.43]* |
| M31 | 11,030 | 81,477,980 | 0.15 | 0.13 | 0.12 | 0.02 | 0.03 | 0.02 | 1.25 [1.17-1.33]* | 1.18 [1.10-1.26]* |
| M32 | 5,260 | 81,502,600 | 0.09 | 0.06 | 0.05 | 0.03 | 0.03 | 0.03 | 1.62 [1.49-1.76]* | 1.49 [1.36-1.63]* |
| M33 | 1,060 | 81,588,360 | 0.02 | 0.01 | 0.01 | 0.01 | 0.01 | 0.01 | 1.90 [1.58-2.29]* | 1.81 [1.50-2.19]* |
| M34 | 2,070 | 81,574,960 | 0.03 | 0.02 | 0.02 | 0.01 | 0.01 | 0.01 | 1.55 [1.35-1.78]* | 1.46 [1.26-1.68]* |
| M35 | 52,950 | 81,063,260 | 0.78 | 0.62 | 0.58 | 0.16 | 0.20 | 0.16 | 1.35 [1.31-1.39]* | 1.27 [1.23-1.30]* |
| M36 | 530 | 81,600,460 | 0.01 | 0.01 | 0.01 | 0.00 | 0.00 | 0.00 | 1.29 [0.97-1.73] | 1.24 [0.92-1.67] |
| M40 | 13,350 | 81,531,760 | 0.18 | 0.16 | 0.15 | 0.02 | 0.03 | 0.02 | 1.18 [1.11-1.25]* | 1.13 [1.06-1.20]* |
| M41 | 37,300 | 81,254,980 | 0.50 | 0.45 | 0.44 | 0.05 | 0.06 | 0.04 | 1.14 [1.10-1.18]* | 1.09 [1.05-1.13]* |
| M42 | 730 | 81,600,300 | 0.01 | 0.01 | 0.01 | 0.00 | 0.00 | 0.00 | 1.14 [0.89-1.46] | 1.02 [0.79-1.32] |
| M43 | 30,620 | 81,228,660 | 0.40 | 0.37 | 0.34 | 0.03 | 0.06 | 0.04 | 1.18 [1.13-1.22]* | 1.10 [1.06-1.15]* |
| M45 | 6,740 | 81,493,240 | 0.10 | 0.08 | 0.07 | 0.02 | 0.03 | 0.02 | 1.42 [1.31-1.54]* | 1.33 [1.23-1.44]* |
| M46 | 24,550 | 81,341,170 | 0.34 | 0.29 | 0.26 | 0.05 | 0.08 | 0.06 | 1.29 [1.24-1.35]* | 1.20 [1.15-1.26]* |
| M47 | 136,980 | 80,068,320 | 1.88 | 1.67 | 1.50 | 0.21 | 0.38 | 0.29 | 1.26 [1.23-1.28]* | 1.18 [1.16-1.20]* |
| M48 | 80,880 | 80,800,370 | 1.06 | 0.99 | 0.89 | 0.07 | 0.17 | 0.13 | 1.20 [1.17-1.23]* | 1.14 [1.11-1.16]* |
| M49 | 2,660 | 81,594,130 | 0.03 | 0.03 | 0.03 | 0.00 | 0.00 | 0.00 | 1.08 [0.94-1.24] | 1.09 [0.95-1.25] |
| M50 | 21,390 | 81,338,810 | 0.28 | 0.26 | 0.23 | 0.02 | 0.05 | 0.04 | 1.21 [1.16-1.27]* | 1.15 [1.09-1.20]* |
| M51 | 100,560 | 80,026,400 | 1.32 | 1.24 | 1.10 | 0.08 | 0.22 | 0.15 | 1.20 [1.18-1.23]* | 1.13 [1.11-1.16]* |
| M53 | 6,530 | 81,492,480 | 0.08 | 0.08 | 0.07 | 0.00 | 0.01 | 0.00 | 1.15 [1.06-1.25]* | 1.06 [0.97-1.16] |
| M54 | 250,260 | 77,375,330 | 3.46 | 3.18 | 2.91 | 0.28 | 0.55 | 0.38 | 1.19 [1.17-1.20]* | 1.12 [1.11-1.14]* |
| M60 | 5,040 | 81,536,490 | 0.08 | 0.06 | 0.05 | 0.02 | 0.02 | 0.02 | 1.38 [1.26-1.52]* | 1.35 [1.23-1.49]* |
| M62 | 21,830 | 81,363,300 | 0.28 | 0.26 | 0.25 | 0.02 | 0.03 | 0.03 | 1.14 [1.09-1.19]* | 1.11 [1.06-1.17]* |
| M65 | 43,470 | 80,681,090 | 0.59 | 0.53 | 0.47 | 0.07 | 0.12 | 0.09 | 1.26 [1.22-1.30]* | 1.19 [1.15-1.22]* |
| M66 | 4,260 | 81,520,580 | 0.05 | 0.05 | 0.05 | 0.00 | 0.01 | 0.01 | 1.16 [1.05-1.29]* | 1.11 [1.00-1.24] |
| M67 | 29,840 | 80,756,360 | 0.39 | 0.36 | 0.34 | 0.03 | 0.06 | 0.04 | 1.17 [1.12-1.21]* | 1.11 [1.07-1.16]* |
| M70 | 16,970 | 81,292,580 | 0.23 | 0.20 | 0.18 | 0.03 | 0.05 | 0.04 | 1.30 [1.23-1.37]* | 1.21 [1.15-1.27]* |
| M71 | 10,840 | 81,450,590 | 0.15 | 0.13 | 0.11 | 0.01 | 0.03 | 0.02 | 1.26 [1.18-1.35]* | 1.19 [1.11-1.27]* |
| M72 | 22,100 | 81,074,170 | 0.26 | 0.28 | 0.24 | −0.02 | 0.01 | 0.01 | 1.06 [1.01-1.11] | 1.04 [0.99-1.09] |
| M75 | 67,990 | 80,184,840 | 0.89 | 0.84 | 0.74 | 0.05 | 0.15 | 0.10 | 1.20 [1.17-1.23]* | 1.13 [1.10-1.16]* |
| M76 | 5,770 | 81,524,080 | 0.08 | 0.07 | 0.06 | 0.01 | 0.02 | 0.01 | 1.32 [1.21-1.44]* | 1.22 [1.11-1.33]* |
| M77 | 13,560 | 81,312,190 | 0.19 | 0.16 | 0.14 | 0.03 | 0.05 | 0.04 | 1.34 [1.26-1.41]* | 1.24 [1.17-1.31]* |
| M79 | 209,940 | 78,285,910 | 3.05 | 2.59 | 2.38 | 0.46 | 0.68 | 0.52 | 1.28 [1.27-1.30]* | 1.20 [1.19-1.22]* |
| M80 | 24,740 | 81,395,890 | 0.33 | 0.30 | 0.27 | 0.04 | 0.06 | 0.05 | 1.22 [1.17-1.28]* | 1.17 [1.12-1.22]* |
| M81 | 151,170 | 80,350,480 | 2.08 | 1.83 | 1.70 | 0.25 | 0.39 | 0.29 | 1.23 [1.21-1.25]* | 1.16 [1.14-1.18]* |
| M83 | 960 | 81,595,120 | 0.01 | 0.01 | 0.01 | 0.00 | 0.00 | 0.00 | 1.29 [1.04-1.60] | 1.22 [0.98-1.52] |
| M84 | 21,330 | 81,190,800 | 0.27 | 0.26 | 0.24 | 0.01 | 0.04 | 0.03 | 1.15 [1.10-1.20]* | 1.13 [1.07-1.18]* |
| M85 | 35,070 | 81,372,430 | 0.48 | 0.42 | 0.39 | 0.06 | 0.09 | 0.07 | 1.24 [1.19-1.28]* | 1.16 [1.12-1.20]* |
| M86 | 15,880 | 81,411,250 | 0.22 | 0.19 | 0.17 | 0.03 | 0.05 | 0.04 | 1.28 [1.21-1.35]* | 1.26 [1.19-1.33]* |
| M87 | 6,730 | 81,497,510 | 0.09 | 0.08 | 0.07 | 0.00 | 0.01 | 0.01 | 1.16 [1.07-1.26]* | 1.13 [1.04-1.23]* |
| M88 | 3,790 | 81,528,080 | 0.05 | 0.05 | 0.04 | 0.00 | 0.00 | 0.00 | 1.07 [0.96-1.21] | 1.05 [0.93-1.18] |
| M89 | 17,420 | 81,246,350 | 0.23 | 0.21 | 0.20 | 0.02 | 0.03 | 0.03 | 1.18 [1.12-1.24]* | 1.13 [1.07-1.19]* |
| M90 | 6,560 | 81,581,130 | 0.07 | 0.08 | 0.07 | −0.01 | 0.00 | 0.00 | 1.02 [0.93-1.11] | 1.04 [0.95-1.14] |
| M91 | 1,450 | 81,565,990 | 0.02 | 0.02 | 0.02 | 0.00 | 0.00 | 0.00 | 1.13 [0.95-1.34] | 1.11 [0.93-1.32] |
| M92 | 2,200 | 81,562,280 | 0.03 | 0.03 | 0.03 | 0.01 | 0.01 | 0.01 | 1.31 [1.14-1.51]* | 1.18 [1.02-1.36] |
| M93 | 4,210 | 81,493,300 | 0.06 | 0.05 | 0.05 | 0.01 | 0.01 | 0.01 | 1.18 [1.07-1.31]* | 1.12 [1.00-1.24] |
| M94 | 12,620 | 81,403,730 | 0.18 | 0.15 | 0.14 | 0.03 | 0.04 | 0.03 | 1.31 [1.23-1.39]* | 1.21 [1.14-1.29]* |
| M95 | 2,960 | 81,491,560 | 0.04 | 0.04 | 0.03 | 0.01 | 0.01 | 0.01 | 1.21 [1.07-1.37]* | 1.16 [1.03-1.32] |
| M96 | 9,030 | 81,516,410 | 0.12 | 0.11 | 0.10 | 0.01 | 0.02 | 0.02 | 1.20 [1.11-1.29]* | 1.17 [1.08-1.26]* |
| M99 | 2,560 | 81,586,100 | 0.03 | 0.03 | 0.03 | 0.00 | 0.00 | 0.00 | 1.16 [1.01-1.33] | 1.09 [0.95-1.25] |
| hospitalised - N00-N99 | | | | | | | | | | |
| N00 | 530 | 81,599,820 | 0.01 | 0.01 | 0.01 | 0.00 | 0.00 | 0.00 | 1.15 [0.87-1.53] | 1.14 [0.85-1.54] |
| N02 | 6,600 | 81,456,850 | 0.09 | 0.08 | 0.07 | 0.01 | 0.01 | 0.01 | 1.19 [1.10-1.30]* | 1.17 [1.07-1.27]* |
| N03 | 26,170 | 81,385,950 | 0.35 | 0.31 | 0.28 | 0.04 | 0.07 | 0.06 | 1.26 [1.21-1.32]* | 1.22 [1.17-1.28]* |
| N04 | 4,280 | 81,526,240 | 0.06 | 0.05 | 0.05 | 0.01 | 0.01 | 0.01 | 1.23 [1.12-1.37]* | 1.20 [1.08-1.33]* |
| N05 | 4,380 | 81,527,020 | 0.06 | 0.05 | 0.05 | 0.01 | 0.02 | 0.02 | 1.37 [1.24-1.51]* | 1.32 [1.19-1.46]* |
| N08 | 12,730 | 81,491,740 | 0.17 | 0.15 | 0.14 | 0.02 | 0.04 | 0.04 | 1.28 [1.21-1.36]* | 1.26 [1.19-1.34]* |
| N10 | 5,320 | 81,507,240 | 0.07 | 0.06 | 0.06 | 0.00 | 0.00 | 0.00 | 1.06 [0.97-1.17] | 1.03 [0.93-1.13] |
| N11 | 2,580 | 81,553,650 | 0.03 | 0.03 | 0.03 | 0.00 | 0.00 | 0.00 | 1.08 [0.94-1.24] | 1.09 [0.95-1.25] |
| N12 | 29,720 | 81,114,050 | 0.38 | 0.36 | 0.35 | 0.02 | 0.03 | 0.02 | 1.10 [1.05-1.14]* | 1.06 [1.02-1.11]* |
| N13 | 61,440 | 80,899,320 | 0.72 | 0.77 | 0.71 | −0.05 | 0.01 | 0.01 | 1.01 [0.99-1.04] | 1.02 [0.99-1.05] |
| N14 | 930 | 81,603,530 | 0.01 | 0.01 | 0.01 | 0.00 | 0.00 | 0.00 | 1.25 [1.00-1.55] | 1.22 [0.98-1.53] |
| N15 | 1,710 | 81,587,830 | 0.02 | 0.02 | 0.02 | 0.00 | 0.00 | 0.00 | 1.08 [0.91-1.28] | 1.06 [0.90-1.26] |
| N16 | 870 | 81,598,350 | 0.01 | 0.01 | 0.01 | 0.00 | 0.00 | 0.00 | 1.29 [1.03-1.62] | 1.30 [1.03-1.64] |
| N17 | 338,580 | 80,306,460 | 4.52 | 4.14 | 3.85 | 0.37 | 0.67 | 0.63 | 1.17 [1.16-1.19]* | 1.16 [1.15-1.18]* |
| N18 | 289,170 | 80,151,460 | 3.88 | 3.54 | 3.26 | 0.34 | 0.61 | 0.54 | 1.19 [1.17-1.20]* | 1.16 [1.15-1.18]* |
| N19 | 40,720 | 81,185,420 | 0.54 | 0.49 | 0.44 | 0.05 | 0.10 | 0.09 | 1.23 [1.19-1.27]* | 1.20 [1.16-1.24]* |
| N20 | 48,820 | 80,605,500 | 0.58 | 0.61 | 0.54 | −0.03 | 0.04 | 0.03 | 1.07 [1.04-1.10]* | 1.05 [1.02-1.08]* |
| N21 | 12,260 | 81,422,080 | 0.13 | 0.16 | 0.14 | −0.02 | −0.01 | −0.01 | 0.95 [0.89-1.02] | 0.96 [0.90-1.02] |
| N23 | 11,140 | 81,174,260 | 0.13 | 0.14 | 0.12 | −0.01 | 0.01 | 0.00 | 1.05 [0.98-1.12] | 1.01 [0.94-1.08] |
| N25 | 2,650 | 81,575,090 | 0.03 | 0.03 | 0.03 | 0.00 | 0.00 | 0.00 | 1.15 [1.00-1.31] | 1.14 [1.00-1.31] |
| N26 | 5,190 | 81,575,220 | 0.06 | 0.06 | 0.06 | 0.00 | 0.01 | 0.01 | 1.10 [1.00-1.21] | 1.09 [0.99-1.21] |
| N27 | 2,770 | 81,573,760 | 0.04 | 0.03 | 0.03 | 0.00 | 0.00 | 0.00 | 1.12 [0.98-1.28] | 1.12 [0.98-1.27] |
| N28 | 69,620 | 80,965,660 | 0.87 | 0.86 | 0.79 | 0.02 | 0.09 | 0.07 | 1.11 [1.08-1.14]* | 1.09 [1.06-1.12]* |
| N29 | 660 | 81,598,300 | 0.01 | 0.01 | 0.01 | 0.00 | 0.00 | 0.00 | 1.18 [0.91-1.55] | 1.16 [0.88-1.52] |
| N30 | 32,110 | 80,992,360 | 0.41 | 0.39 | 0.35 | 0.02 | 0.06 | 0.05 | 1.16 [1.12-1.21]* | 1.13 [1.09-1.17]* |
| N31 | 10,300 | 81,363,560 | 0.14 | 0.12 | 0.11 | 0.01 | 0.02 | 0.02 | 1.19 [1.12-1.27]* | 1.13 [1.06-1.21]* |
| N32 | 91,150 | 79,970,370 | 1.18 | 1.13 | 1.03 | 0.05 | 0.15 | 0.12 | 1.14 [1.12-1.17]* | 1.11 [1.09-1.14]* |
| N34 | 1,580 | 81,565,930 | 0.02 | 0.02 | 0.02 | 0.00 | 0.00 | 0.00 | 1.12 [0.94-1.33] | 1.09 [0.91-1.29] |
| N35 | 31,140 | 80,880,560 | 0.41 | 0.38 | 0.34 | 0.03 | 0.07 | 0.06 | 1.20 [1.16-1.25]* | 1.17 [1.12-1.22]* |
| N36 | 9,020 | 81,421,600 | 0.11 | 0.11 | 0.10 | 0.00 | 0.01 | 0.01 | 1.14 [1.06-1.23]* | 1.11 [1.03-1.19]* |
| N39 | 403,030 | 76,126,900 | 5.63 | 5.21 | 4.96 | 0.42 | 0.67 | 0.58 | 1.14 [1.12-1.15]* | 1.11 [1.10-1.13]* |
| N40 | 124,410 | 79,729,070 | 1.64 | 1.54 | 1.49 | 0.10 | 0.15 | 0.11 | 1.10 [1.08-1.12]* | 1.07 [1.05-1.09]* |
| N41 | 9,450 | 81,405,900 | 0.11 | 0.12 | 0.10 | −0.01 | 0.00 | 0.00 | 1.04 [0.97-1.12] | 1.01 [0.93-1.08] |
| N42 | 12,520 | 81,381,370 | 0.14 | 0.16 | 0.14 | −0.01 | 0.00 | 0.00 | 1.03 [0.97-1.10] | 1.01 [0.95-1.08] |
| N43 | 17,630 | 81,128,920 | 0.23 | 0.21 | 0.21 | 0.02 | 0.02 | 0.02 | 1.10 [1.05-1.16]* | 1.09 [1.03-1.15]* |
| N44 | 5,530 | 81,482,120 | 0.09 | 0.06 | 0.08 | 0.03 | 0.01 | 0.01 | 1.15 [1.06-1.26]* | 1.14 [1.04-1.25]* |
| N45 | 13,290 | 81,316,900 | 0.17 | 0.16 | 0.15 | 0.01 | 0.02 | 0.01 | 1.11 [1.04-1.18]* | 1.09 [1.02-1.15]* |
| N47 | 44,480 | 80,256,610 | 0.74 | 0.51 | 0.54 | 0.23 | 0.20 | 0.19 | 1.37 [1.33-1.42]* | 1.34 [1.30-1.38]* |
| N48 | 26,020 | 80,994,840 | 0.39 | 0.30 | 0.30 | 0.08 | 0.09 | 0.08 | 1.28 [1.23-1.34]* | 1.26 [1.21-1.31]* |
| N49 | 3,220 | 81,547,360 | 0.04 | 0.04 | 0.04 | 0.00 | 0.01 | 0.01 | 1.18 [1.05-1.33]* | 1.19 [1.05-1.34]* |
| N50 | 27,610 | 80,905,870 | 0.38 | 0.33 | 0.33 | 0.04 | 0.05 | 0.04 | 1.15 [1.10-1.19]* | 1.12 [1.08-1.17]* |
| N60 | 9,260 | 81,273,960 | 0.11 | 0.11 | 0.10 | 0.00 | 0.01 | 0.01 | 1.10 [1.02-1.18]* | 1.09 [1.01-1.17] |
| N61 | 6,460 | 81,427,950 | 0.09 | 0.08 | 0.07 | 0.01 | 0.02 | 0.02 | 1.27 [1.17-1.38]* | 1.26 [1.15-1.37]* |
| N62 | 5,140 | 81,377,660 | 0.07 | 0.06 | 0.06 | 0.00 | 0.01 | 0.01 | 1.16 [1.05-1.27]* | 1.12 [1.01-1.23] |
| N63 | 9,880 | 81,264,630 | 0.12 | 0.12 | 0.11 | 0.00 | 0.01 | 0.01 | 1.10 [1.02-1.18]* | 1.08 [1.00-1.16] |

| | | | | | | | | | | |
|---|---|---|---|---|---|---|---|---|---|---|
| N64 | 9,960 | 81,355,960 | 0.13 | 0.12 | 0.11 | 0.01 | 0.02 | 0.02 | 1.18 [1.11-1.27]* | 1.15 [1.08-1.24]* |
| N70 | 6,970 | 81,401,480 | 0.08 | 0.09 | 0.08 | −0.01 | 0.00 | 0.00 | 1.00 [0.92-1.09] | 0.98 [0.90-1.07] |
| N71 | 3,460 | 81,512,870 | 0.04 | 0.04 | 0.04 | 0.00 | 0.01 | 0.00 | 1.14 [1.02-1.28] | 1.08 [0.96-1.22] |
| N72 | 6,140 | 81,302,250 | 0.07 | 0.08 | 0.07 | −0.01 | 0.00 | 0.00 | 1.04 [0.96-1.14] | 1.02 [0.93-1.12] |
| N73 | 38,640 | 80,480,810 | 0.47 | 0.48 | 0.44 | −0.01 | 0.03 | 0.02 | 1.07 [1.03-1.11]* | 1.03 [1.00-1.07] |
| N75 | 6,090 | 81,377,660 | 0.07 | 0.08 | 0.07 | −0.01 | 0.00 | 0.00 | 1.01 [0.92-1.10] | 1.00 [0.91-1.10] |
| N76 | 10,140 | 81,313,920 | 0.15 | 0.12 | 0.11 | 0.03 | 0.04 | 0.03 | 1.32 [1.24-1.41]* | 1.27 [1.19-1.36]* |
| N77 | 12,940 | 81,328,630 | 0.18 | 0.16 | 0.15 | 0.02 | 0.03 | 0.02 | 1.19 [1.12-1.26]* | 1.13 [1.06-1.20]* |
| N80 | 36,000 | 80,412,160 | 0.45 | 0.45 | 0.41 | 0.00 | 0.03 | 0.01 | 1.08 [1.04-1.12]* | 1.03 [0.99-1.07] |
| N81 | 59,120 | 79,925,540 | 0.76 | 0.73 | 0.66 | 0.02 | 0.10 | 0.07 | 1.15 [1.12-1.18]* | 1.10 [1.07-1.13]* |
| N82 | 3,080 | 81,559,930 | 0.04 | 0.04 | 0.03 | 0.00 | 0.01 | 0.01 | 1.23 [1.09-1.38]* | 1.19 [1.05-1.34]* |
| N83 | 60,910 | 79,951,970 | 0.77 | 0.76 | 0.71 | 0.01 | 0.06 | 0.03 | 1.08 [1.05-1.11]* | 1.04 [1.01-1.07]* |
| N84 | 45,630 | 80,062,610 | 0.56 | 0.57 | 0.51 | −0.02 | 0.04 | 0.03 | 1.08 [1.05-1.12]* | 1.07 [1.03-1.10]* |
| N85 | 39,540 | 80,593,480 | 0.50 | 0.49 | 0.44 | 0.01 | 0.05 | 0.04 | 1.12 [1.09-1.16]* | 1.09 [1.05-1.13]* |
| N86 | 11,210 | 81,174,440 | 0.14 | 0.14 | 0.13 | 0.01 | 0.02 | 0.01 | 1.13 [1.06-1.20]* | 1.08 [1.01-1.15] |
| N87 | 12,780 | 80,802,690 | 0.15 | 0.16 | 0.14 | −0.01 | 0.01 | 0.01 | 1.05 [0.99-1.12] | 1.05 [0.98-1.11] |
| N88 | 11,860 | 81,269,870 | 0.15 | 0.15 | 0.13 | 0.00 | 0.02 | 0.01 | 1.12 [1.05-1.19]* | 1.10 [1.03-1.17]* |
| N89 | 20,720 | 80,955,580 | 0.27 | 0.25 | 0.23 | 0.02 | 0.04 | 0.03 | 1.17 [1.12-1.23]* | 1.12 [1.07-1.18]* |
| N90 | 19,020 | 81,089,870 | 0.26 | 0.23 | 0.21 | 0.03 | 0.06 | 0.05 | 1.27 [1.21-1.34]* | 1.22 [1.16-1.28]* |
| N91 | 2,230 | 81,534,480 | 0.03 | 0.03 | 0.03 | 0.00 | 0.00 | 0.00 | 1.15 [1.00-1.32] | 1.10 [0.95-1.27] |
| N92 | 79,150 | 78,677,320 | 1.05 | 1.00 | 0.92 | 0.05 | 0.13 | 0.09 | 1.15 [1.12-1.17]* | 1.10 [1.07-1.13]* |
| N93 | 38,310 | 80,244,560 | 0.49 | 0.48 | 0.44 | 0.01 | 0.05 | 0.03 | 1.11 [1.07-1.14]* | 1.07 [1.04-1.11]* |
| N94 | 30,520 | 80,349,940 | 0.40 | 0.37 | 0.34 | 0.03 | 0.06 | 0.04 | 1.17 [1.13-1.22]* | 1.11 [1.07-1.16]* |
| N95 | 40,150 | 80,260,210 | 0.53 | 0.49 | 0.44 | 0.04 | 0.09 | 0.07 | 1.19 [1.15-1.23]* | 1.16 [1.12-1.20]* |
| N96 | 670 | 81,593,170 | 0.01 | 0.01 | 0.01 | 0.00 | 0.00 | 0.00 | 0.99 [0.75-1.30] | 1.00 [0.76-1.33] |
| N97 | 4,700 | 80,991,400 | 0.05 | 0.06 | 0.05 | −0.01 | 0.00 | 0.00 | 0.97 [0.88-1.08] | 0.96 [0.86-1.07] |
| N98 | 730 | 81,578,260 | 0.01 | 0.01 | 0.01 | 0.00 | 0.00 | 0.00 | 0.80 [0.60-1.06] | 0.77 [0.57-1.02] |
| N99 | 11,620 | 81,356,340 | 0.14 | 0.14 | 0.13 | 0.00 | 0.02 | 0.01 | 1.13 [1.06-1.20]* | 1.11 [1.04-1.18]* |
| hospitalised - O00-O9A | | | | | | | | | | |
| O00 | 11,450 | 81,201,520 | 0.13 | 0.14 | 0.13 | −0.01 | 0.00 | 0.00 | 0.99 [0.92-1.05] | 0.98 [0.92-1.05] |
| O01 | 650 | 81,584,750 | 0.01 | 0.01 | 0.01 | 0.00 | 0.00 | 0.00 | 1.14 [0.87-1.48] | 1.21 [0.92-1.59] |
| O02 | 25,360 | 80,417,910 | 0.30 | 0.32 | 0.29 | −0.02 | 0.01 | 0.01 | 1.04 [1.00-1.09] | 1.03 [0.99-1.08] |
| O03 | 38,390 | 79,946,380 | 0.46 | 0.49 | 0.45 | −0.03 | 0.00 | 0.00 | 1.01 [0.97-1.05] | 1.00 [0.97-1.04] |
| O04 | 43,540 | 79,428,140 | 0.54 | 0.55 | 0.57 | −0.01 | −0.03 | −0.03 | 0.94 [0.91-0.98]* | 0.95 [0.92-0.98]* |
| O08 | 4,280 | 81,483,310 | 0.05 | 0.05 | 0.05 | 0.00 | 0.00 | 0.00 | 0.97 [0.87-1.09] | 0.97 [0.87-1.09] |
| O10 | 4,440 | 81,487,960 | 0.05 | 0.05 | 0.05 | 0.00 | 0.00 | 0.00 | 1.09 [0.98-1.21] | 1.06 [0.96-1.19] |
| O11 | 620 | 81,591,780 | 0.01 | 0.01 | 0.01 | 0.00 | 0.00 | 0.00 | 1.12 [0.85-1.48] | 1.09 [0.82-1.44] |
| O12 | 8,260 | 81,213,360 | 0.10 | 0.10 | 0.09 | 0.00 | 0.01 | 0.00 | 1.09 [1.01-1.17] | 1.05 [0.97-1.14] |
| O13 | 20,670 | 80,566,070 | 0.25 | 0.26 | 0.24 | 0.00 | 0.01 | 0.01 | 1.05 [1.00-1.10] | 1.03 [0.98-1.08] |
| O14 | 14,620 | 80,888,980 | 0.18 | 0.18 | 0.17 | 0.00 | 0.01 | 0.01 | 1.06 [1.00-1.13] | 1.03 [0.97-1.09] |
| O15 | 580 | 81,580,460 | 0.01 | 0.01 | 0.01 | 0.00 | 0.00 | 0.00 | 1.05 [0.79-1.40] | 1.00 [0.74-1.35] |
| O16 | 16,400 | 80,530,040 | 0.20 | 0.20 | 0.19 | 0.00 | 0.02 | 0.01 | 1.08 [1.02-1.14]* | 1.06 [1.01-1.12] |
| O20 | 33,410 | 80,135,800 | 0.41 | 0.42 | 0.41 | −0.01 | 0.00 | 0.00 | 1.00 [0.96-1.04] | 0.99 [0.95-1.03] |
| O21 | 24,680 | 80,711,630 | 0.30 | 0.31 | 0.30 | 0.00 | 0.01 | 0.00 | 1.02 [0.97-1.06] | 0.99 [0.94-1.03] |
| O22 | 4,390 | 81,489,790 | 0.05 | 0.05 | 0.05 | 0.00 | 0.00 | 0.00 | 1.00 [0.90-1.12] | 1.00 [0.90-1.12] |
| O23 | 26,820 | 80,563,170 | 0.34 | 0.33 | 0.32 | 0.01 | 0.03 | 0.02 | 1.08 [1.04-1.13]* | 1.06 [1.02-1.11]* |
| O24 | 33,740 | 80,897,110 | 0.40 | 0.42 | 0.39 | −0.02 | 0.01 | 0.00 | 1.02 [0.98-1.06] | 1.01 [0.97-1.05] |
| O26 | 91,380 | 77,302,320 | 1.27 | 1.16 | 1.18 | 0.11 | 0.09 | 0.05 | 1.07 [1.05-1.10]* | 1.04 [1.02-1.07]* |
| O28 | 10,310 | 81,388,470 | 0.12 | 0.13 | 0.12 | −0.01 | 0.00 | 0.00 | 0.99 [0.92-1.06] | 0.96 [0.90-1.03] |
| O30 | 12,370 | 81,191,910 | 0.14 | 0.16 | 0.14 | −0.02 | −0.01 | −0.01 | 0.95 [0.89-1.02] | 0.95 [0.88-1.01] |
| O31 | 720 | 81,587,380 | 0.01 | 0.01 | 0.01 | 0.00 | 0.00 | 0.00 | 1.08 [0.84-1.39] | 1.07 [0.82-1.39] |
| O32 | 45,580 | 79,917,320 | 0.54 | 0.58 | 0.54 | −0.04 | −0.01 | −0.01 | 0.99 [0.96-1.02] | 0.98 [0.95-1.02] |
| O33 | 1,140 | 81,534,310 | 0.01 | 0.01 | 0.01 | 0.00 | 0.00 | 0.00 | 1.02 [0.83-1.26] | 1.01 [0.82-1.24] |
| O34 | 90,710 | 79,146,910 | 1.09 | 1.16 | 1.07 | −0.07 | 0.02 | 0.02 | 1.02 [1.00-1.04] | 1.02 [0.99-1.04] |
| O35 | 12,260 | 81,259,790 | 0.14 | 0.15 | 0.14 | −0.01 | 0.00 | 0.00 | 0.99 [0.93-1.05] | 0.98 [0.92-1.05] |
| O36 | 119,100 | 77,225,730 | 1.65 | 1.52 | 1.58 | 0.13 | 0.07 | 0.05 | 1.04 [1.02-1.07]* | 1.03 [1.01-1.05]* |
| O40 | 11,950 | 81,352,870 | 0.14 | 0.15 | 0.14 | −0.01 | 0.01 | 0.00 | 1.04 [0.98-1.11] | 1.02 [0.96-1.09] |
| O41 | 13,900 | 81,005,190 | 0.17 | 0.17 | 0.16 | 0.00 | 0.01 | 0.01 | 1.04 [0.98-1.11] | 1.04 [0.98-1.10] |
| O42 | 68,050 | 78,812,600 | 0.86 | 0.87 | 0.85 | −0.01 | 0.01 | 0.01 | 1.01 [0.98-1.04] | 1.02 [0.99-1.05] |
| O43 | 10,540 | 81,165,940 | 0.12 | 0.13 | 0.12 | −0.01 | 0.00 | 0.00 | 1.01 [0.94-1.08] | 1.00 [0.94-1.08] |
| O44 | 7,850 | 81,352,440 | 0.09 | 0.10 | 0.09 | −0.01 | 0.00 | 0.00 | 1.01 [0.94-1.10] | 1.00 [0.92-1.08] |
| O45 | 3,300 | 81,489,740 | 0.04 | 0.04 | 0.04 | 0.00 | 0.00 | 0.00 | 1.00 [0.88-1.13] | 0.99 [0.88-1.13] |
| O46 | 35,020 | 80,086,420 | 0.44 | 0.44 | 0.41 | 0.00 | 0.03 | 0.02 | 1.06 [1.02-1.10]* | 1.05 [1.01-1.09] |
| O47 | 23,500 | 79,470,910 | 0.30 | 0.30 | 0.28 | 0.00 | 0.02 | 0.01 | 1.07 [1.02-1.12]* | 1.05 [1.00-1.10] |
| O48 | 40,410 | 79,270,940 | 0.48 | 0.52 | 0.49 | −0.04 | −0.02 | −0.01 | 0.97 [0.93-1.00] | 0.97 [0.94-1.01] |
| O60 | 39,610 | 80,086,560 | 0.48 | 0.50 | 0.47 | −0.02 | 0.01 | 0.01 | 1.03 [0.99-1.06] | 1.02 [0.98-1.06] |
| O61 | 6,700 | 81,288,040 | 0.08 | 0.08 | 0.08 | 0.00 | 0.00 | 0.00 | 1.05 [0.97-1.15] | 1.02 [0.93-1.11] |
| O62 | 27,980 | 80,578,560 | 0.34 | 0.35 | 0.33 | −0.01 | 0.01 | 0.01 | 1.02 [0.98-1.07] | 1.03 [0.98-1.07] |
| O63 | 50,500 | 78,365,170 | 0.64 | 0.65 | 0.63 | −0.01 | 0.01 | 0.00 | 1.01 [0.98-1.04] | 1.00 [0.97-1.03] |
| O64 | 11,000 | 80,863,800 | 0.13 | 0.14 | 0.13 | −0.01 | 0.00 | 0.00 | 1.01 [0.94-1.07] | 1.00 [0.94-1.07] |
| O65 | 820 | 81,531,580 | 0.01 | 0.01 | 0.01 | 0.00 | 0.00 | 0.00 | 0.99 [0.78-1.26] | 1.02 [0.79-1.31] |
| O66 | 11,740 | 81,025,240 | 0.13 | 0.15 | 0.14 | −0.01 | 0.00 | 0.00 | 0.99 [0.93-1.06] | 0.99 [0.93-1.06] |
| O67 | 3,230 | 81,445,820 | 0.04 | 0.04 | 0.04 | 0.00 | 0.00 | 0.00 | 1.01 [0.89-1.14] | 0.99 [0.88-1.13] |
| O68 | 113,140 | 75,488,680 | 1.58 | 1.48 | 1.57 | 0.10 | 0.01 | 0.00 | 1.01 [0.99-1.03] | 1.00 [0.98-1.02] |
| O69 | 12,920 | 80,716,360 | 0.15 | 0.16 | 0.15 | −0.01 | 0.01 | 0.01 | 1.03 [0.97-1.10] | 1.04 [0.98-1.11] |
| O70 | 149,700 | 73,066,490 | 2.23 | 2.00 | 2.28 | 0.23 | −0.05 | −0.02 | 0.98 [0.96-0.99] | 0.99 [0.97-1.01] |
| O71 | 11,070 | 81,128,280 | 0.13 | 0.14 | 0.13 | −0.01 | 0.00 | 0.00 | 1.00 [0.93-1.06] | 1.00 [0.94-1.07] |
| O72 | 88,670 | 78,684,440 | 1.13 | 1.13 | 1.10 | 0.00 | 0.02 | 0.02 | 1.02 [1.00-1.04] | 1.02 [0.99-1.04] |
| O73 | 6,700 | 81,269,990 | 0.08 | 0.08 | 0.07 | −0.01 | 0.00 | 0.00 | 1.04 [0.95-1.13] | 1.03 [0.95-1.13] |
| O74 | 1,850 | 81,544,010 | 0.02 | 0.02 | 0.02 | 0.00 | 0.00 | 0.00 | 1.12 [0.96-1.31] | 1.10 [0.94-1.30] |
| O75 | 45,770 | 79,886,760 | 0.55 | 0.58 | 0.55 | −0.03 | 0.00 | 0.01 | 1.01 [0.97-1.04] | 1.01 [0.98-1.05] |
| O80 | 52,830 | 78,240,240 | 0.64 | 0.68 | 0.66 | −0.04 | −0.02 | 0.00 | 0.97 [0.94-1.00] | 1.00 [0.97-1.03] |
| O82 | 7,740 | 81,056,600 | 0.09 | 0.10 | 0.09 | −0.01 | 0.00 | 0.00 | 1.02 [0.94-1.10] | 1.01 [0.93-1.09] |
| O85 | 4,170 | 81,530,810 | 0.05 | 0.05 | 0.05 | 0.00 | 0.00 | 0.00 | 1.03 [0.93-1.15] | 1.01 [0.90-1.12] |
| O86 | 14,470 | 81,111,350 | 0.18 | 0.18 | 0.16 | 0.01 | 0.02 | 0.01 | 1.11 [1.05-1.18]* | 1.08 [1.02-1.15]* |
| O87 | 2,160 | 81,502,560 | 0.03 | 0.03 | 0.02 | 0.00 | 0.00 | 0.00 | 1.17 [1.01-1.35] | 1.16 [0.99-1.35] |
| O88 | 900 | 81,589,330 | 0.01 | 0.01 | 0.01 | 0.00 | 0.00 | 0.00 | 1.14 [0.91-1.44] | 1.09 [0.86-1.38] |
| O89 | 640 | 81,583,300 | 0.01 | 0.01 | 0.01 | 0.00 | 0.00 | 0.00 | 1.17 [0.90-1.53] | 1.14 [0.87-1.49] |
| O90 | 17,470 | 81,144,550 | 0.22 | 0.21 | 0.20 | 0.01 | 0.02 | 0.02 | 1.11 [1.05-1.17]* | 1.08 [1.02-1.14]* |
| O91 | 1,740 | 81,561,350 | 0.02 | 0.02 | 0.02 | 0.00 | 0.00 | 0.00 | 0.98 [0.82-1.16] | 0.95 [0.80-1.14] |
| O92 | 2,610 | 81,508,320 | 0.03 | 0.03 | 0.03 | 0.00 | 0.00 | 0.00 | 1.04 [0.91-1.19] | 1.03 [0.89-1.18] |
| O94 | 660 | 81,603,890 | 0.01 | 0.01 | 0.01 | 0.00 | 0.00 | 0.00 | 1.15 [0.88-1.50] | 1.11 [0.84-1.46] |
| O98 | 6,820 | 81,400,060 | 0.09 | 0.08 | 0.08 | 0.00 | 0.01 | 0.01 | 1.10 [1.01-1.19] | 1.06 [0.98-1.16] |
| O99 | 104,040 | 77,881,300 | 1.43 | 1.31 | 1.31 | 0.11 | 0.11 | 0.08 | 1.08 [1.06-1.11]* | 1.06 [1.04-1.08]* |
| hospitalised - P00-P96 | | | | | | | | | | |
| P83 | 1,370 | 80,594,280 | 0.02 | 0.02 | 0.02 | 0.00 | 0.00 | 0.00 | 1.00 [0.84-1.20] | 1.01 [0.84-1.21] |
| hospitalised - Q00-Q99 | | | | | | | | | | |
| Q02 | 1,950 | 81,557,430 | 0.03 | 0.02 | 0.03 | 0.00 | 0.00 | 0.00 | 1.02 [0.88-1.18] | 1.00 [0.86-1.16] |
| Q03 | 760 | 81,567,870 | 0.01 | 0.01 | 0.01 | 0.00 | 0.00 | 0.00 | 1.22 [0.96-1.55] | 1.21 [0.95-1.55] |
| Q04 | 2,090 | 81,536,420 | 0.03 | 0.03 | 0.03 | 0.00 | 0.00 | 0.00 | 1.02 [0.88-1.18] | 0.98 [0.84-1.14] |
| Q05 | 1,870 | 81,544,140 | 0.02 | 0.02 | 0.02 | 0.00 | 0.00 | 0.00 | 1.18 [1.01-1.37] | 1.10 [0.94-1.28] |
| Q06 | 600 | 81,597,540 | 0.01 | 0.01 | 0.01 | 0.00 | 0.00 | 0.00 | 0.85 [0.63-1.13] | 0.81 [0.60-1.08] |
| Q07 | 2,370 | 81,573,650 | 0.03 | 0.03 | 0.03 | 0.00 | 0.00 | 0.00 | 1.11 [0.97-1.27] | 1.04 [0.90-1.19] |
| Q10 | 2,280 | 81,541,940 | 0.03 | 0.03 | 0.03 | 0.01 | 0.00 | 0.00 | 1.14 [1.00-1.31] | 1.12 [0.98-1.29] |
| Q12 | 1,180 | 81,570,370 | 0.02 | 0.01 | 0.01 | 0.00 | 0.00 | 0.00 | 1.16 [0.95-1.40] | 1.13 [0.93-1.38] |
| Q13 | 480 | 81,593,240 | 0.01 | 0.01 | 0.01 | 0.00 | 0.00 | 0.00 | 1.22 [0.90-1.64] | 1.14 [0.84-1.55] |

| | | | | | | | | | | |
|---|---|---|---|---|---|---|---|---|---|---|
| Q14 | 620 | 81,597,600 | 0.01 | 0.01 | 0.01 | 0.00 | 0.00 | 0.00 | 1.08 [0.82-1.42] | 1.08 [0.82-1.43] |
| Q17 | 4,170 | 81,342,050 | 0.06 | 0.05 | 0.06 | 0.01 | 0.00 | 0.00 | 1.06 [0.96-1.17] | 1.06 [0.95-1.17] |
| Q18 | 2,770 | 81,506,030 | 0.04 | 0.03 | 0.03 | 0.01 | 0.00 | 0.00 | 1.14 [1.01-1.29] | 1.13 [1.00-1.29] |
| Q20 | 570 | 81,565,000 | 0.01 | 0.01 | 0.01 | 0.00 | 0.00 | 0.00 | 1.03 [0.77-1.37] | 1.00 [0.75-1.34] |
| Q21 | 9,280 | 81,167,760 | 0.12 | 0.11 | 0.11 | 0.01 | 0.01 | 0.01 | 1.12 [1.04-1.20]* | 1.07 [1.00-1.15] |
| Q22 | 1,220 | 81,544,750 | 0.01 | 0.02 | 0.02 | 0.00 | 0.00 | 0.00 | 0.94 [0.77-1.15] | 0.95 [0.78-1.17] |
| Q23 | 4,210 | 81,516,460 | 0.05 | 0.05 | 0.05 | 0.00 | 0.00 | 0.00 | 1.04 [0.94-1.16] | 1.02 [0.92-1.14] |
| Q24 | 3,430 | 81,491,750 | 0.04 | 0.04 | 0.04 | 0.00 | 0.00 | 0.00 | 1.08 [0.96-1.22] | 1.03 [0.92-1.16] |
| Q25 | 3,480 | 81,285,750 | 0.04 | 0.04 | 0.04 | 0.00 | 0.00 | 0.00 | 1.02 [0.91-1.14] | 0.98 [0.88-1.11] |
| Q26 | 620 | 81,591,250 | 0.01 | 0.01 | 0.01 | 0.00 | 0.00 | 0.00 | 0.97 [0.74-1.27] | 0.93 [0.71-1.23] |
| Q27 | 2,950 | 81,545,950 | 0.04 | 0.04 | 0.04 | 0.00 | 0.00 | 0.00 | 1.03 [0.91-1.17] | 1.02 [0.90-1.16] |
| Q28 | 1,440 | 81,574,030 | 0.02 | 0.02 | 0.02 | 0.00 | 0.00 | 0.00 | 1.18 [0.99-1.40] | 1.17 [0.98-1.39] |
| Q31 | 1,320 | 81,524,350 | 0.02 | 0.02 | 0.02 | 0.00 | 0.00 | 0.00 | 0.92 [0.77-1.11] | 0.84 [0.69-1.02] |
| Q32 | 540 | 81,592,100 | 0.01 | 0.01 | 0.01 | 0.00 | 0.00 | 0.00 | 0.92 [0.68-1.23] | 0.80 [0.59-1.09] |
| Q33 | 520 | 81,575,570 | 0.01 | 0.01 | 0.01 | 0.00 | 0.00 | 0.00 | 1.01 [0.75-1.36] | 0.90 [0.66-1.23] |
| Q35 | 1,350 | 81,537,590 | 0.02 | 0.02 | 0.02 | 0.00 | 0.00 | 0.00 | 1.04 [0.87-1.24] | 1.01 [0.84-1.22] |
| Q38 | 8,660 | 80,950,640 | 0.12 | 0.10 | 0.11 | 0.02 | 0.01 | 0.01 | 1.10 [1.02-1.18] | 1.07 [1.00-1.15] |
| Q39 | 1,650 | 81,575,410 | 0.02 | 0.02 | 0.02 | 0.00 | 0.00 | 0.00 | 1.11 [0.93-1.31] | 1.03 [0.87-1.23] |
| Q40 | 1,010 | 81,513,210 | 0.01 | 0.01 | 0.01 | 0.00 | 0.00 | 0.00 | 1.20 [0.97-1.48] | 1.12 [0.90-1.39] |
| Q43 | 3,920 | 81,489,790 | 0.05 | 0.05 | 0.04 | 0.00 | 0.01 | 0.00 | 1.13 [1.01-1.26] | 1.07 [0.96-1.19] |
| Q44 | 1,320 | 81,582,930 | 0.02 | 0.02 | 0.01 | 0.00 | 0.00 | 0.00 | 1.08 [0.89-1.31] | 1.05 [0.86-1.27] |
| Q45 | 540 | 81,600,990 | 0.01 | 0.01 | 0.01 | 0.00 | 0.00 | 0.00 | 1.37 [1.03-1.81] | 1.29 [0.97-1.72] |
| Q50 | 2,630 | 81,529,660 | 0.03 | 0.03 | 0.03 | 0.00 | 0.00 | 0.00 | 1.13 [0.99-1.29] | 1.11 [0.97-1.27] |
| Q51 | 2,580 | 81,544,310 | 0.03 | 0.03 | 0.03 | 0.00 | 0.00 | 0.00 | 1.06 [0.93-1.22] | 1.03 [0.90-1.18] |
| Q52 | 1,260 | 81,558,060 | 0.02 | 0.01 | 0.02 | 0.00 | 0.00 | 0.00 | 1.23 [1.03-1.48] | 1.20 [0.99-1.44] |
| Q53 | 11,030 | 81,025,560 | 0.15 | 0.13 | 0.16 | 0.02 | −0.01 | −0.01 | 0.96 [0.90-1.02] | 0.96 [0.90-1.02] |
| Q54 | 3,240 | 81,360,470 | 0.04 | 0.04 | 0.04 | 0.00 | 0.00 | 0.00 | 1.04 [0.93-1.17] | 1.06 [0.94-1.19] |
| Q55 | 7,100 | 81,403,860 | 0.11 | 0.08 | 0.10 | 0.03 | 0.01 | 0.01 | 1.13 [1.04-1.22]* | 1.10 [1.02-1.19] |
| Q60 | 2,720 | 81,554,670 | 0.03 | 0.03 | 0.03 | 0.00 | 0.00 | 0.00 | 1.07 [0.94-1.23] | 1.06 [0.92-1.21] |
| Q61 | 6,690 | 81,470,580 | 0.08 | 0.08 | 0.08 | 0.00 | 0.01 | 0.00 | 1.07 [0.99-1.17] | 1.05 [0.96-1.15] |
| Q62 | 1,520 | 81,409,110 | 0.02 | 0.02 | 0.02 | 0.00 | 0.00 | 0.00 | 0.96 [0.81-1.15] | 0.96 [0.80-1.15] |
| Q63 | 5,300 | 81,470,370 | 0.07 | 0.06 | 0.06 | 0.00 | 0.00 | 0.00 | 1.05 [0.96-1.16] | 1.00 [0.91-1.10] |
| Q64 | 1,140 | 81,551,510 | 0.02 | 0.01 | 0.02 | 0.00 | 0.00 | 0.00 | 1.03 [0.85-1.26] | 1.01 [0.82-1.24] |
| Q65 | 4,290 | 81,417,150 | 0.05 | 0.05 | 0.05 | 0.00 | 0.00 | 0.00 | 0.93 [0.84-1.04] | 0.91 [0.82-1.02] |
| Q66 | 4,730 | 80,756,330 | 0.07 | 0.06 | 0.06 | 0.01 | 0.01 | 0.01 | 1.12 [1.02-1.23] | 1.09 [0.99-1.20] |
| Q67 | 3,130 | 81,501,880 | 0.04 | 0.04 | 0.04 | 0.01 | 0.00 | 0.00 | 1.10 [0.98-1.24] | 1.07 [0.95-1.21] |
| Q68 | 1,080 | 81,562,170 | 0.01 | 0.01 | 0.01 | 0.00 | 0.00 | 0.00 | 1.03 [0.84-1.27] | 1.00 [0.81-1.24] |
| Q72 | 480 | 81,592,300 | 0.01 | 0.01 | 0.01 | 0.00 | 0.00 | 0.00 | 1.06 [0.78-1.43] | 1.04 [0.76-1.42] |
| Q74 | 3,260 | 81,517,730 | 0.05 | 0.04 | 0.04 | 0.01 | 0.00 | 0.00 | 1.11 [0.98-1.24] | 1.08 [0.96-1.21] |
| Q75 | 1,830 | 81,521,550 | 0.02 | 0.02 | 0.03 | 0.00 | 0.00 | 0.00 | 0.98 [0.84-1.14] | 0.95 [0.81-1.11] |
| Q76 | 2,710 | 81,540,610 | 0.03 | 0.03 | 0.03 | 0.00 | 0.00 | 0.00 | 1.07 [0.94-1.21] | 1.02 [0.89-1.16] |
| Q78 | 1,760 | 81,573,860 | 0.02 | 0.02 | 0.02 | 0.00 | 0.00 | 0.00 | 1.12 [0.95-1.31] | 1.07 [0.91-1.26] |
| Q79 | 3,540 | 81,523,670 | 0.05 | 0.04 | 0.04 | 0.01 | 0.01 | 0.00 | 1.19 [1.07-1.33]* | 1.04 [0.93-1.17] |
| Q80 | 250 | 81,602,560 | 0.01 | 0.00 | 0.00 | 0.00 | 0.00 | 0.00 | 2.74 [1.90-3.95]* | 2.50 [1.71-3.64]* |
| Q82 | 2,680 | 80,900,600 | 0.04 | 0.03 | 0.03 | 0.01 | 0.01 | 0.01 | 1.32 [1.17-1.49]* | 1.26 [1.11-1.43]* |
| Q83 | 1,260 | 81,537,110 | 0.02 | 0.02 | 0.01 | 0.00 | 0.00 | 0.00 | 1.04 [0.86-1.26] | 0.99 [0.82-1.21] |
| Q85 | 2,330 | 81,551,960 | 0.03 | 0.03 | 0.03 | 0.00 | 0.00 | 0.00 | 0.94 [0.81-1.08] | 0.92 [0.79-1.06] |
| Q87 | 3,320 | 81,507,090 | 0.05 | 0.04 | 0.04 | 0.01 | 0.01 | 0.00 | 1.14 [1.01-1.28] | 1.10 [0.98-1.24] |
| Q89 | 2,890 | 81,526,250 | 0.04 | 0.03 | 0.04 | 0.00 | 0.00 | 0.00 | 1.06 [0.93-1.20] | 1.04 [0.91-1.18] |
| Q90 | 1,160 | 81,459,150 | 0.02 | 0.01 | 0.01 | 0.01 | 0.01 | 0.01 | 2.00 [1.68-2.37]* | 1.90 [1.59-2.27]* |
| Q92 | 620 | 81,601,510 | 0.01 | 0.01 | 0.01 | 0.00 | 0.00 | 0.00 | 1.18 [0.91-1.54] | 1.15 [0.88-1.51] |
| Q93 | 1,690 | 81,582,690 | 0.03 | 0.02 | 0.02 | 0.01 | 0.01 | 0.00 | 1.25 [1.07-1.46]* | 1.17 [1.00-1.38] |
| Q98 | 460 | 81,596,960 | 0.01 | 0.01 | 0.00 | 0.00 | 0.00 | 0.00 | 1.63 [1.22-2.17]* | 1.50 [1.12-2.02]* |
| Q99 | 2,040 | 81,574,330 | 0.03 | 0.02 | 0.03 | 0.01 | 0.00 | 0.00 | 1.10 [0.95-1.27] | 1.05 [0.90-1.21] |
| hospitalised - R00-R99 | | | | | | | | | | |
| R00 | 188,760 | 79,371,840 | 2.50 | 2.35 | 2.20 | 0.15 | 0.30 | 0.22 | 1.14 [1.12-1.16]* | 1.10 [1.08-1.12]* |
| R01 | 30,680 | 81,007,020 | 0.42 | 0.37 | 0.36 | 0.05 | 0.06 | 0.05 | 1.17 [1.13-1.22]* | 1.13 [1.09-1.18]* |
| R02 | 28,210 | 81,413,330 | 0.36 | 0.34 | 0.31 | 0.02 | 0.05 | 0.05 | 1.16 [1.11-1.21]* | 1.16 [1.11-1.21]* |
| R03 | 64,760 | 80,822,970 | 0.83 | 0.79 | 0.74 | 0.03 | 0.09 | 0.07 | 1.12 [1.09-1.15]* | 1.10 [1.07-1.13]* |
| R04 | 73,250 | 80,377,220 | 1.01 | 0.89 | 0.82 | 0.13 | 0.19 | 0.16 | 1.23 [1.20-1.26]* | 1.19 [1.16-1.22]* |
| R05 | 79,910 | 80,300,020 | 1.18 | 0.95 | 0.92 | 0.23 | 0.27 | 0.21 | 1.29 [1.26-1.32]* | 1.21 [1.18-1.24]* |
| R06 | 260,570 | 76,492,040 | 4.42 | 3.16 | 3.10 | 1.26 | 1.32 | 1.18 | 1.43 [1.41-1.44]* | 1.36 [1.35-1.38]* |
| R07 | 301,820 | 74,726,870 | 4.31 | 3.97 | 3.64 | 0.34 | 0.67 | 0.48 | 1.19 [1.17-1.20]* | 1.13 [1.11-1.14]* |
| R09 | 28,890 | 81,267,600 | 0.39 | 0.35 | 0.32 | 0.04 | 0.07 | 0.05 | 1.22 [1.17-1.27]* | 1.16 [1.12-1.21]* |
| R10 | 408,530 | 69,657,160 | 6.53 | 5.69 | 5.54 | 0.83 | 0.99 | 0.71 | 1.18 [1.17-1.19]* | 1.12 [1.11-1.13]* |
| R11 | 263,030 | 76,697,280 | 3.74 | 3.35 | 3.25 | 0.39 | 0.49 | 0.36 | 1.15 [1.13-1.17]* | 1.11 [1.09-1.12]* |
| R12 | 16,740 | 81,258,260 | 0.22 | 0.20 | 0.18 | 0.02 | 0.04 | 0.03 | 1.23 [1.17-1.30]* | 1.14 [1.08-1.20]* |
| R13 | 126,880 | 80,364,500 | 1.73 | 1.54 | 1.41 | 0.19 | 0.32 | 0.25 | 1.23 [1.21-1.25]* | 1.17 [1.15-1.19]* |
| R14 | 29,600 | 81,168,960 | 0.40 | 0.36 | 0.33 | 0.04 | 0.07 | 0.05 | 1.22 [1.17-1.26]* | 1.14 [1.10-1.19]* |
| R15 | 54,570 | 81,298,980 | 0.71 | 0.66 | 0.62 | 0.04 | 0.09 | 0.08 | 1.14 [1.10-1.17]* | 1.13 [1.09-1.16]* |
| R16 | 20,570 | 81,386,920 | 0.29 | 0.24 | 0.23 | 0.04 | 0.06 | 0.05 | 1.27 [1.21-1.33]* | 1.23 [1.17-1.29]* |
| R17 | 15,740 | 81,357,570 | 0.20 | 0.19 | 0.17 | 0.01 | 0.03 | 0.03 | 1.19 [1.12-1.25]* | 1.18 [1.12-1.25]* |
| R18 | 40,870 | 81,331,060 | 0.52 | 0.50 | 0.44 | 0.02 | 0.08 | 0.08 | 1.17 [1.13-1.21]* | 1.17 [1.13-1.21]* |
| R19 | 158,450 | 79,215,830 | 2.19 | 1.95 | 1.76 | 0.24 | 0.44 | 0.33 | 1.25 [1.23-1.27]* | 1.18 [1.16-1.20]* |
| R20 | 45,150 | 80,884,180 | 0.59 | 0.55 | 0.50 | 0.03 | 0.08 | 0.05 | 1.16 [1.13-1.20]* | 1.10 [1.06-1.13]* |
| R21 | 68,450 | 79,860,330 | 1.10 | 0.80 | 0.81 | 0.30 | 0.30 | 0.27 | 1.37 [1.34-1.40]* | 1.32 [1.29-1.35]* |
| R22 | 33,100 | 80,883,670 | 0.48 | 0.39 | 0.37 | 0.09 | 0.11 | 0.10 | 1.31 [1.27-1.36]* | 1.25 [1.20-1.29]* |
| R23 | 27,980 | 81,023,600 | 0.40 | 0.33 | 0.34 | 0.07 | 0.06 | 0.05 | 1.18 [1.14-1.23]* | 1.14 [1.10-1.19]* |
| R25 | 26,650 | 81,075,410 | 0.35 | 0.32 | 0.30 | 0.03 | 0.05 | 0.04 | 1.16 [1.12-1.21]* | 1.11 [1.07-1.16]* |
| R26 | 199,250 | 80,702,410 | 2.66 | 2.42 | 2.31 | 0.24 | 0.34 | 0.31 | 1.15 [1.13-1.17]* | 1.13 [1.11-1.15]* |
| R27 | 8,000 | 81,506,630 | 0.11 | 0.10 | 0.09 | 0.01 | 0.01 | 0.01 | 1.12 [1.04-1.21]* | 1.09 [1.01-1.18] |
| R29 | 261,880 | 80,059,520 | 3.49 | 3.22 | 3.05 | 0.27 | 0.44 | 0.38 | 1.14 [1.13-1.16]* | 1.12 [1.11-1.14]* |
| R30 | 20,910 | 81,210,550 | 0.29 | 0.25 | 0.24 | 0.04 | 0.05 | 0.04 | 1.21 [1.16-1.27]* | 1.14 [1.09-1.20]* |
| R31 | 129,590 | 78,761,260 | 1.68 | 1.64 | 1.51 | 0.04 | 0.17 | 0.14 | 1.11 [1.09-1.13]* | 1.09 [1.07-1.11]* |
| R32 | 104,490 | 80,866,240 | 1.36 | 1.27 | 1.21 | 0.09 | 0.15 | 0.14 | 1.13 [1.10-1.15]* | 1.11 [1.09-1.14]* |
| R33 | 188,060 | 79,946,970 | 2.46 | 2.32 | 2.20 | 0.14 | 0.27 | 0.23 | 1.12 [1.10-1.14]* | 1.10 [1.09-1.12]* |
| R34 | 5,190 | 81,574,200 | 0.07 | 0.06 | 0.06 | 0.01 | 0.01 | 0.01 | 1.23 [1.11-1.35]* | 1.20 [1.09-1.32]* |
| R35 | 31,880 | 80,787,160 | 0.44 | 0.38 | 0.35 | 0.05 | 0.08 | 0.06 | 1.23 [1.19-1.28]* | 1.17 [1.12-1.21]* |
| R36 | 700 | 81,597,920 | 0.01 | 0.01 | 0.01 | 0.00 | 0.00 | 0.00 | 1.19 [0.92-1.54] | 1.15 [0.88-1.48] |
| R39 | 37,540 | 80,759,680 | 0.50 | 0.46 | 0.41 | 0.04 | 0.09 | 0.07 | 1.22 [1.18-1.26]* | 1.16 [1.12-1.20]* |
| R40 | 31,520 | 81,326,580 | 0.42 | 0.38 | 0.36 | 0.04 | 0.06 | 0.05 | 1.15 [1.11-1.20]* | 1.13 [1.08-1.17]* |
| R41 | 183,140 | 80,526,070 | 2.41 | 2.24 | 2.12 | 0.17 | 0.29 | 0.27 | 1.14 [1.12-1.16]* | 1.13 [1.11-1.15]* |
| R42 | 104,580 | 80,284,090 | 1.38 | 1.28 | 1.20 | 0.10 | 0.19 | 0.13 | 1.16 [1.13-1.18]* | 1.10 [1.08-1.13]* |
| R43 | 1,410 | 81,595,460 | 0.02 | 0.02 | 0.02 | 0.00 | 0.00 | 0.00 | 1.24 [1.04-1.49] | 1.17 [0.98-1.41] |
| R44 | 27,780 | 81,405,630 | 0.36 | 0.34 | 0.32 | 0.03 | 0.05 | 0.04 | 1.15 [1.10-1.19]* | 1.12 [1.08-1.17]* |
| R45 | 63,000 | 81,172,120 | 0.85 | 0.76 | 0.76 | 0.09 | 0.09 | 0.08 | 1.12 [1.09-1.15]* | 1.10 [1.07-1.13]* |
| R46 | 18,010 | 81,493,330 | 0.23 | 0.22 | 0.21 | 0.01 | 0.02 | 0.02 | 1.10 [1.04-1.16]* | 1.11 [1.05-1.17]* |
| R47 | 52,310 | 81,169,530 | 0.66 | 0.64 | 0.59 | 0.02 | 0.07 | 0.06 | 1.11 [1.08-1.15]* | 1.10 [1.06-1.13]* |
| R48 | 2,070 | 81,591,160 | 0.03 | 0.02 | 0.03 | 0.01 | 0.01 | 0.00 | 1.23 [1.06-1.42]* | 1.16 [1.00-1.35] |
| R49 | 10,460 | 81,381,110 | 0.14 | 0.13 | 0.12 | 0.02 | 0.03 | 0.02 | 1.23 [1.15-1.31]* | 1.15 [1.08-1.23]* |
| R50 | 112,410 | 79,302,880 | 1.60 | 1.37 | 1.38 | 0.23 | 0.22 | 0.18 | 1.16 [1.14-1.18]* | 1.13 [1.11-1.15]* |
| R51 | 148,830 | 78,643,690 | 2.10 | 1.84 | 1.77 | 0.25 | 0.33 | 0.23 | 1.19 [1.17-1.21]* | 1.12 [1.10-1.14]* |
| R52 | 25,280 | 81,315,440 | 0.35 | 0.30 | 0.27 | 0.05 | 0.08 | 0.06 | 1.28 [1.23-1.34]* | 1.20 [1.15-1.25]* |
| R53 | 100,410 | 80,687,600 | 1.35 | 1.22 | 1.16 | 0.13 | 0.19 | 0.15 | 1.17 [1.14-1.19]* | 1.13 [1.10-1.16]* |
| R54 | 118,780 | 81,141,370 | 1.55 | 1.44 | 1.38 | 0.11 | 0.17 | 0.17 | 1.13 [1.10-1.15]* | 1.12 [1.10-1.14]* |
| R55 | 181,620 | 79,085,190 | 2.42 | 2.27 | 2.13 | 0.15 | 0.29 | 0.25 | 1.14 [1.12-1.16]* | 1.11 [1.09-1.13]* |
| R56 | 85,100 | 79,525,880 | 1.13 | 1.05 | 1.06 | 0.08 | 0.08 | 0.07 | 1.07 [1.05-1.10]* | 1.06 [1.04-1.09]* |

| | | | | | | | | | | |
|---|---|---|---|---|---|---|---|---|---|---|
| R57 | 16,150 | 81,556,560 | 0.20 | 0.20 | 0.18 | 0.01 | 0.03 | 0.03 | 1.15 [1.09-1.21]* | 1.15 [1.08-1.21]* |
| R58 | 2,600 | 81,584,790 | 0.04 | 0.03 | 0.03 | 0.01 | 0.01 | 0.01 | 1.30 [1.14-1.48]* | 1.29 [1.13-1.48]* |
| R59 | 52,360 | 80,851,360 | 0.75 | 0.62 | 0.59 | 0.13 | 0.16 | 0.15 | 1.27 [1.24-1.31]* | 1.26 [1.22-1.29]* |
| R60 | 87,450 | 81,069,580 | 1.21 | 1.05 | 0.95 | 0.16 | 0.25 | 0.22 | 1.27 [1.24-1.30]* | 1.22 [1.19-1.25]* |
| R61 | 9,720 | 81,434,020 | 0.13 | 0.12 | 0.11 | 0.02 | 0.03 | 0.02 | 1.23 [1.15-1.32]* | 1.16 [1.09-1.25]* |
| R62 | 15,270 | 80,998,400 | 0.21 | 0.18 | 0.21 | 0.02 | 0.00 | −0.01 | 0.99 [0.94-1.05] | 0.97 [0.92-1.02] |
| R63 | 183,790 | 79,277,050 | 2.50 | 2.27 | 2.12 | 0.23 | 0.38 | 0.32 | 1.18 [1.16-1.20]* | 1.14 [1.13-1.16]* |
| R64 | 4,830 | 81,600,160 | 0.06 | 0.06 | 0.06 | 0.00 | 0.01 | 0.01 | 1.09 [0.99-1.21] | 1.12 [1.01-1.25] |
| R65 | 11,520 | 81,579,380 | 0.15 | 0.14 | 0.13 | 0.01 | 0.02 | 0.02 | 1.18 [1.11-1.26]* | 1.15 [1.08-1.23]* |
| R68 | 18,200 | 81,260,350 | 0.24 | 0.22 | 0.21 | 0.02 | 0.03 | 0.03 | 1.17 [1.11-1.23]* | 1.16 [1.10-1.23]* |
| R69 | 139,970 | 73,746,920 | 2.01 | 1.87 | 1.73 | 0.14 | 0.28 | 0.24 | 1.16 [1.14-1.18]* | 1.13 [1.11-1.15]* |
| R70 | 1,240 | 81,577,230 | 0.02 | 0.01 | 0.01 | 0.00 | 0.00 | 0.00 | 1.35 [1.12-1.62]* | 1.28 [1.06-1.55]* |
| R71 | 1,080 | 81,596,870 | 0.01 | 0.01 | 0.01 | 0.00 | 0.00 | 0.00 | 1.11 [0.90-1.36] | 1.09 [0.88-1.35] |
| R72 | 5,980 | 81,544,140 | 0.08 | 0.07 | 0.07 | 0.00 | 0.01 | 0.01 | 1.15 [1.05-1.26]* | 1.13 [1.03-1.23] |
| R73 | 22,880 | 81,348,630 | 0.31 | 0.27 | 0.25 | 0.04 | 0.06 | 0.05 | 1.25 [1.20-1.31]* | 1.19 [1.14-1.25]* |
| R74 | 10,260 | 81,511,670 | 0.13 | 0.12 | 0.12 | 0.01 | 0.02 | 0.01 | 1.15 [1.08-1.23]* | 1.12 [1.04-1.20]* |
| R76 | 2,720 | 81,563,050 | 0.04 | 0.03 | 0.03 | 0.01 | 0.01 | 0.01 | 1.29 [1.14-1.46]* | 1.23 [1.08-1.40]* |
| R77 | 6,370 | 81,565,830 | 0.09 | 0.08 | 0.07 | 0.01 | 0.02 | 0.01 | 1.23 [1.13-1.34]* | 1.18 [1.08-1.29]* |
| R78 | 820 | 81,593,170 | 0.01 | 0.01 | 0.01 | 0.00 | 0.00 | 0.00 | 1.04 [0.82-1.33] | 1.05 [0.82-1.35] |
| R79 | 122,040 | 80,488,140 | 1.60 | 1.50 | 1.37 | 0.11 | 0.23 | 0.19 | 1.17 [1.15-1.19]* | 1.14 [1.11-1.16]* |
| R80 | 4,940 | 81,504,550 | 0.07 | 0.06 | 0.06 | 0.01 | 0.01 | 0.01 | 1.23 [1.12-1.36]* | 1.20 [1.09-1.32]* |
| R81 | 1,250 | 81,508,030 | 0.02 | 0.02 | 0.01 | 0.00 | 0.00 | 0.00 | 1.20 [0.99-1.44] | 1.17 [0.97-1.42] |
| R82 | 10,600 | 81,465,750 | 0.14 | 0.13 | 0.12 | 0.01 | 0.01 | 0.01 | 1.12 [1.05-1.20]* | 1.10 [1.02-1.17]* |
| R84 | 3,530 | 81,583,100 | 0.05 | 0.04 | 0.04 | 0.01 | 0.01 | 0.00 | 1.19 [1.06-1.33]* | 1.08 [0.96-1.21] |
| R85 | 1,780 | 81,557,380 | 0.03 | 0.02 | 0.02 | 0.00 | 0.01 | 0.00 | 1.32 [1.13-1.54]* | 1.22 [1.04-1.43] |
| R87 | 7,550 | 81,001,980 | 0.09 | 0.09 | 0.08 | 0.00 | 0.01 | 0.00 | 1.07 [0.99-1.16] | 1.06 [0.97-1.15] |
| R89 | 1,720 | 81,588,300 | 0.02 | 0.02 | 0.02 | 0.00 | 0.01 | 0.01 | 1.31 [1.11-1.53]* | 1.26 [1.07-1.49]* |
| R90 | 5,490 | 81,545,570 | 0.07 | 0.07 | 0.06 | 0.00 | 0.01 | 0.01 | 1.17 [1.07-1.28]* | 1.14 [1.04-1.26]* |
| R91 | 57,990 | 81,118,200 | 0.77 | 0.70 | 0.64 | 0.06 | 0.13 | 0.11 | 1.20 [1.17-1.24]* | 1.17 [1.14-1.21]* |
| R93 | 60,270 | 80,965,800 | 0.77 | 0.74 | 0.67 | 0.03 | 0.10 | 0.08 | 1.14 [1.11-1.18]* | 1.12 [1.08-1.15]* |
| R94 | 106,370 | 80,560,420 | 1.42 | 1.30 | 1.19 | 0.12 | 0.22 | 0.20 | 1.19 [1.16-1.21]* | 1.16 [1.14-1.19]* |
| hospitalised - S00-T88 | | | | | | | | | | |
| S00 | 114,120 | 80,202,090 | 1.50 | 1.40 | 1.38 | 0.09 | 0.11 | 0.10 | 1.08 [1.06-1.11]* | 1.07 [1.05-1.09]* |
| S01 | 118,230 | 79,884,540 | 1.57 | 1.46 | 1.45 | 0.11 | 0.12 | 0.11 | 1.08 [1.06-1.11]* | 1.07 [1.05-1.09]* |
| S02 | 40,280 | 80,453,570 | 0.51 | 0.50 | 0.49 | 0.01 | 0.02 | 0.01 | 1.04 [1.00-1.07] | 1.02 [0.99-1.06] |
| S03 | 1,880 | 81,578,420 | 0.03 | 0.02 | 0.02 | 0.00 | 0.00 | 0.00 | 1.07 [0.92-1.25] | 1.06 [0.90-1.24] |
| S05 | 9,820 | 81,429,500 | 0.12 | 0.12 | 0.11 | 0.00 | 0.01 | 0.01 | 1.10 [1.02-1.18] | 1.09 [1.02-1.17] |
| S06 | 31,970 | 81,170,680 | 0.40 | 0.39 | 0.38 | 0.01 | 0.02 | 0.02 | 1.06 [1.01-1.10]* | 1.05 [1.01-1.09] |
| S09 | 62,400 | 80,015,610 | 0.84 | 0.77 | 0.76 | 0.07 | 0.08 | 0.07 | 1.11 [1.08-1.14]* | 1.08 [1.05-1.12]* |
| S10 | 3,500 | 81,547,010 | 0.04 | 0.04 | 0.04 | 0.00 | 0.00 | 0.00 | 0.97 [0.86-1.10] | 0.96 [0.85-1.09] |
| S11 | 1,770 | 81,575,210 | 0.02 | 0.02 | 0.02 | 0.00 | 0.00 | 0.00 | 1.16 [0.98-1.36] | 1.16 [0.98-1.38] |
| S12 | 7,770 | 81,525,610 | 0.10 | 0.09 | 0.09 | 0.01 | 0.01 | 0.01 | 1.15 [1.06-1.25]* | 1.15 [1.06-1.24]* |
| S13 | 1,940 | 81,551,570 | 0.03 | 0.02 | 0.02 | 0.00 | 0.00 | 0.00 | 1.14 [0.98-1.33] | 1.10 [0.94-1.29] |
| S14 | 1,440 | 81,583,080 | 0.02 | 0.02 | 0.02 | 0.00 | 0.00 | 0.00 | 1.01 [0.84-1.22] | 1.02 [0.84-1.24] |
| S19 | 2,620 | 81,541,410 | 0.03 | 0.03 | 0.03 | 0.00 | 0.00 | 0.00 | 1.16 [1.02-1.32] | 1.08 [0.95-1.24] |
| S20 | 12,180 | 81,464,340 | 0.16 | 0.15 | 0.14 | 0.01 | 0.02 | 0.01 | 1.12 [1.05-1.19]* | 1.08 [1.02-1.16] |
| S21 | 2,320 | 81,560,180 | 0.03 | 0.03 | 0.03 | 0.00 | 0.00 | 0.00 | 1.05 [0.91-1.22] | 1.06 [0.91-1.23] |
| S22 | 38,960 | 81,151,780 | 0.50 | 0.48 | 0.43 | 0.02 | 0.06 | 0.05 | 1.15 [1.11-1.19]* | 1.12 [1.08-1.16]* |
| S27 | 11,590 | 81,429,970 | 0.14 | 0.14 | 0.13 | −0.01 | 0.01 | 0.01 | 1.04 [0.97-1.11] | 1.04 [0.97-1.11] |
| S29 | 3,150 | 81,566,220 | 0.04 | 0.04 | 0.03 | 0.01 | 0.01 | 0.01 | 1.32 [1.17-1.48]* | 1.24 [1.10-1.41]* |
| S30 | 23,110 | 81,302,490 | 0.30 | 0.28 | 0.27 | 0.02 | 0.03 | 0.02 | 1.10 [1.05-1.16]* | 1.07 [1.02-1.12]* |
| S31 | 6,600 | 81,456,140 | 0.09 | 0.08 | 0.08 | 0.01 | 0.00 | 0.00 | 1.05 [0.97-1.15] | 1.05 [0.96-1.14] |
| S32 | 42,660 | 81,179,250 | 0.53 | 0.52 | 0.49 | 0.01 | 0.04 | 0.03 | 1.08 [1.04-1.12]* | 1.06 [1.02-1.09]* |
| S33 | 530 | 81,598,500 | 0.01 | 0.01 | 0.01 | 0.00 | 0.00 | 0.00 | 1.00 [0.74-1.36] | 0.92 [0.68-1.26] |
| S36 | 4,790 | 81,520,250 | 0.06 | 0.06 | 0.06 | 0.00 | 0.00 | 0.00 | 1.05 [0.95-1.16] | 1.05 [0.95-1.16] |
| S37 | 3,730 | 81,553,990 | 0.04 | 0.05 | 0.04 | −0.01 | 0.00 | 0.00 | 0.96 [0.85-1.08] | 0.95 [0.84-1.08] |
| S39 | 8,850 | 81,430,610 | 0.12 | 0.11 | 0.10 | 0.01 | 0.01 | 0.01 | 1.12 [1.04-1.20]* | 1.07 [0.99-1.15] |
| S40 | 17,780 | 81,454,700 | 0.23 | 0.22 | 0.21 | 0.01 | 0.02 | 0.01 | 1.10 [1.04-1.16]* | 1.07 [1.01-1.13] |
| S41 | 6,940 | 81,526,630 | 0.09 | 0.08 | 0.08 | 0.01 | 0.01 | 0.01 | 1.14 [1.05-1.24]* | 1.10 [1.01-1.20] |
| S42 | 58,220 | 80,647,580 | 0.76 | 0.71 | 0.70 | 0.04 | 0.05 | 0.05 | 1.08 [1.05-1.11]* | 1.07 [1.04-1.10]* |
| S43 | 8,340 | 81,465,720 | 0.10 | 0.10 | 0.10 | 0.00 | 0.01 | 0.00 | 1.06 [0.98-1.14] | 1.05 [0.97-1.13] |
| S46 | 3,610 | 81,563,490 | 0.04 | 0.04 | 0.04 | 0.00 | 0.00 | 0.00 | 1.06 [0.95-1.20] | 1.02 [0.90-1.15] |
| S49 | 5,290 | 81,551,120 | 0.07 | 0.06 | 0.06 | 0.01 | 0.01 | 0.01 | 1.18 [1.07-1.30]* | 1.14 [1.03-1.26]* |
| S50 | 19,430 | 81,446,070 | 0.25 | 0.23 | 0.23 | 0.02 | 0.02 | 0.01 | 1.09 [1.03-1.14]* | 1.06 [1.01-1.12] |
| S51 | 27,290 | 81,320,790 | 0.36 | 0.33 | 0.32 | 0.03 | 0.04 | 0.03 | 1.13 [1.09-1.18]* | 1.10 [1.05-1.14]* |
| S52 | 87,620 | 79,385,340 | 1.17 | 1.09 | 1.14 | 0.08 | 0.03 | 0.01 | 1.02 [1.00-1.05] | 1.01 [0.99-1.04] |
| S53 | 2,490 | 81,542,770 | 0.03 | 0.03 | 0.03 | 0.00 | 0.00 | 0.00 | 1.03 [0.90-1.18] | 1.01 [0.88-1.16] |
| S54 | 1,060 | 81,574,880 | 0.01 | 0.01 | 0.01 | 0.00 | 0.00 | 0.00 | 1.20 [0.98-1.47] | 1.15 [0.93-1.43] |
| S55 | 450 | 81,595,920 | 0.01 | 0.01 | 0.00 | 0.00 | 0.00 | 0.00 | 1.35 [0.99-1.84] | 1.43 [1.04-1.97] |
| S56 | 3,610 | 81,506,350 | 0.04 | 0.05 | 0.04 | 0.00 | 0.00 | 0.00 | 0.92 [0.82-1.04] | 0.93 [0.82-1.05] |
| S59 | 2,260 | 81,580,010 | 0.03 | 0.03 | 0.03 | 0.00 | 0.00 | 0.00 | 1.15 [0.99-1.33] | 1.11 [0.95-1.29] |
| S60 | 15,390 | 81,432,450 | 0.20 | 0.19 | 0.19 | 0.01 | 0.01 | 0.01 | 1.07 [1.01-1.13] | 1.05 [0.99-1.11] |
| S61 | 58,400 | 80,262,880 | 0.72 | 0.73 | 0.73 | −0.01 | 0.00 | −0.01 | 0.99 [0.97-1.02] | 0.98 [0.95-1.01] |
| S62 | 39,560 | 80,638,870 | 0.49 | 0.49 | 0.49 | 0.00 | −0.01 | −0.01 | 0.99 [0.95-1.02] | 0.98 [0.95-1.02] |
| S63 | 6,140 | 81,459,670 | 0.08 | 0.07 | 0.07 | 0.00 | 0.01 | 0.01 | 1.13 [1.03-1.23]* | 1.09 [1.00-1.19] |
| S64 | 7,530 | 81,354,260 | 0.08 | 0.09 | 0.09 | −0.01 | −0.01 | −0.01 | 0.90 [0.83-0.98] | 0.90 [0.83-0.98] |
| S65 | 2,080 | 81,546,380 | 0.02 | 0.03 | 0.02 | 0.00 | 0.00 | 0.00 | 0.96 [0.83-1.13] | 0.98 [0.84-1.15] |
| S66 | 13,460 | 81,129,770 | 0.15 | 0.17 | 0.16 | −0.02 | −0.01 | −0.01 | 0.95 [0.89-1.00] | 0.95 [0.89-1.01] |
| S67 | 2,110 | 81,533,390 | 0.02 | 0.03 | 0.03 | 0.00 | 0.00 | 0.00 | 0.87 [0.75-1.02] | 0.88 [0.75-1.03] |
| S68 | 4,860 | 81,455,230 | 0.06 | 0.06 | 0.06 | 0.00 | 0.00 | 0.00 | 0.92 [0.83-1.02] | 0.92 [0.84-1.02] |
| S69 | 5,950 | 81,509,390 | 0.08 | 0.07 | 0.07 | 0.01 | 0.01 | 0.01 | 1.17 [1.07-1.28]* | 1.13 [1.03-1.24]* |
| S70 | 17,690 | 81,459,540 | 0.23 | 0.21 | 0.21 | 0.01 | 0.02 | 0.01 | 1.08 [1.02-1.14]* | 1.06 [1.00-1.12] |
| S71 | 3,880 | 81,533,700 | 0.05 | 0.05 | 0.05 | 0.00 | 0.00 | 0.00 | 1.03 [0.92-1.15] | 1.01 [0.90-1.13] |
| S72 | 110,390 | 80,501,790 | 1.42 | 1.36 | 1.31 | 0.06 | 0.11 | 0.11 | 1.08 [1.06-1.11]* | 1.09 [1.06-1.11]* |
| S73 | 1,700 | 81,580,490 | 0.02 | 0.02 | 0.02 | 0.00 | 0.00 | 0.00 | 1.05 [0.88-1.24] | 1.02 [0.86-1.22] |
| S76 | 3,310 | 81,555,670 | 0.04 | 0.04 | 0.04 | 0.00 | 0.00 | 0.00 | 1.09 [0.96-1.23] | 1.05 [0.92-1.18] |
| S79 | 9,110 | 81,530,370 | 0.12 | 0.11 | 0.10 | 0.01 | 0.02 | 0.02 | 1.17 [1.09-1.27]* | 1.15 [1.07-1.24]* |
| S80 | 34,070 | 81,288,440 | 0.44 | 0.42 | 0.40 | 0.02 | 0.03 | 0.02 | 1.09 [1.04-1.13]* | 1.04 [1.00-1.08] |
| S81 | 25,060 | 81,310,840 | 0.33 | 0.30 | 0.29 | 0.03 | 0.04 | 0.03 | 1.15 [1.10-1.20]* | 1.08 [1.03-1.13]* |
| S82 | 63,730 | 79,877,290 | 0.79 | 0.80 | 0.76 | 0.00 | 0.03 | 0.02 | 1.04 [1.02-1.07]* | 1.03 [1.00-1.06] |
| S83 | 8,470 | 81,365,050 | 0.11 | 0.10 | 0.10 | 0.00 | 0.01 | 0.00 | 1.06 [0.98-1.14] | 1.02 [0.94-1.10] |
| S86 | 4,080 | 81,463,130 | 0.05 | 0.05 | 0.05 | 0.00 | 0.00 | 0.00 | 1.10 [0.99-1.22] | 1.04 [0.93-1.16] |
| S89 | 7,170 | 81,512,030 | 0.09 | 0.09 | 0.08 | 0.01 | 0.01 | 0.01 | 1.12 [1.04-1.22]* | 1.08 [0.99-1.17] |
| S90 | 8,850 | 81,502,400 | 0.11 | 0.11 | 0.10 | 0.00 | 0.00 | 0.00 | 1.04 [0.96-1.12] | 1.01 [0.93-1.09] |
| S91 | 9,090 | 81,433,920 | 0.12 | 0.11 | 0.11 | 0.01 | 0.01 | 0.01 | 1.08 [1.01-1.17] | 1.06 [0.98-1.14] |
| S92 | 13,660 | 81,303,660 | 0.17 | 0.17 | 0.16 | 0.01 | 0.02 | 0.01 | 1.10 [1.04-1.17]* | 1.08 [1.01-1.14] |
| S93 | 4,540 | 81,523,720 | 0.06 | 0.06 | 0.05 | 0.00 | 0.01 | 0.00 | 1.11 [1.00-1.23] | 1.05 [0.95-1.17] |
| S96 | 1,010 | 81,585,440 | 0.01 | 0.01 | 0.01 | 0.00 | 0.00 | 0.00 | 1.16 [0.94-1.43] | 1.06 [0.85-1.32] |
| S99 | 3,650 | 81,557,000 | 0.05 | 0.04 | 0.04 | 0.00 | 0.00 | 0.00 | 1.08 [0.97-1.21] | 1.03 [0.92-1.16] |
| T00 | 7,860 | 81,536,810 | 0.10 | 0.10 | 0.09 | 0.01 | 0.01 | 0.00 | 1.08 [0.99-1.17] | 1.04 [0.96-1.13] |
| T01 | 3,780 | 81,574,850 | 0.05 | 0.05 | 0.05 | 0.00 | 0.00 | 0.00 | 1.09 [0.97-1.22] | 1.05 [0.93-1.18] |
| T02 | 2,340 | 81,571,610 | 0.03 | 0.03 | 0.03 | 0.00 | 0.00 | 0.00 | 1.05 [0.91-1.21] | 1.05 [0.90-1.21] |
| T09 | 1,270 | 81,586,350 | 0.02 | 0.02 | 0.01 | 0.00 | 0.00 | 0.00 | 1.11 [0.92-1.35] | 1.09 [0.89-1.33] |
| T11 | 5,270 | 81,557,050 | 0.07 | 0.06 | 0.06 | 0.01 | 0.01 | 0.01 | 1.15 [1.05-1.27]* | 1.11 [1.00-1.23] |
| T13 | 3,300 | 81,565,470 | 0.04 | 0.04 | 0.04 | 0.00 | 0.00 | 0.00 | 1.11 [0.98-1.26] | 1.06 [0.94-1.21] |
| T14 | 3,080 | 81,493,750 | 0.04 | 0.04 | 0.04 | 0.00 | 0.00 | 0.00 | 1.02 [0.90-1.16] | 0.98 [0.87-1.12] |
| T15 | 710 | 81,586,950 | 0.01 | 0.01 | 0.01 | 0.00 | 0.00 | 0.00 | 0.95 [0.74-1.23] | 0.95 [0.74-1.23] |

| | | | | | | | | | | |
|---|---|---|---|---|---|---|---|---|---|---|
| T16 | 7,430 | 81,446,920 | 0.10 | 0.09 | 0.10 | 0.02 | 0.00 | 0.00 | 1.02 [0.94-1.10] | 1.00 [0.93-1.08] |
| T17 | 11,950 | 81,369,370 | 0.16 | 0.14 | 0.15 | 0.01 | 0.01 | 0.00 | 1.07 [1.00-1.14] | 1.03 [0.97-1.10] |
| T18 | 9,140 | 81,411,020 | 0.12 | 0.11 | 0.11 | 0.01 | 0.01 | 0.00 | 1.07 [0.99-1.14] | 1.02 [0.95-1.10] |
| T19 | 870 | 81,590,050 | 0.01 | 0.01 | 0.01 | 0.00 | 0.00 | 0.00 | 1.17 [0.93-1.47] | 1.15 [0.91-1.45] |
| T20 | 3,920 | 81,500,220 | 0.05 | 0.05 | 0.05 | 0.00 | 0.00 | 0.00 | 0.99 [0.88-1.10] | 0.99 [0.89-1.11] |
| T21 | 5,800 | 81,472,200 | 0.07 | 0.07 | 0.07 | 0.00 | 0.00 | 0.00 | 0.99 [0.91-1.09] | 1.00 [0.91-1.09] |
| T22 | 4,860 | 81,487,670 | 0.06 | 0.06 | 0.06 | 0.00 | 0.00 | 0.00 | 0.94 [0.85-1.04] | 0.95 [0.85-1.05] |
| T23 | 4,790 | 81,482,320 | 0.06 | 0.06 | 0.06 | 0.00 | 0.00 | 0.00 | 0.97 [0.88-1.07] | 0.97 [0.88-1.08] |
| T24 | 4,100 | 81,517,750 | 0.05 | 0.05 | 0.05 | 0.00 | −0.01 | −0.01 | 0.89 [0.79-0.99] | 0.90 [0.80-1.00] |
| T25 | 2,120 | 81,560,260 | 0.02 | 0.03 | 0.03 | 0.00 | 0.00 | 0.00 | 0.89 [0.76-1.03] | 0.87 [0.75-1.02] |
| T29 | 920 | 81,586,850 | 0.01 | 0.01 | 0.01 | 0.00 | 0.00 | 0.00 | 0.94 [0.75-1.19] | 0.94 [0.74-1.19] |
| T31 | 13,100 | 81,297,810 | 0.16 | 0.16 | 0.17 | 0.00 | −0.01 | −0.01 | 0.94 [0.89-1.00] | 0.94 [0.89-1.00] |
| T36 | 2,880 | 81,521,450 | 0.04 | 0.03 | 0.03 | 0.01 | 0.00 | 0.00 | 1.14 [1.00-1.29] | 1.11 [0.98-1.26] |
| T37 | 1,210 | 81,578,700 | 0.02 | 0.01 | 0.01 | 0.00 | 0.00 | 0.00 | 1.21 [1.00-1.47] | 1.20 [0.98-1.46] |
| T38 | 5,210 | 81,516,650 | 0.07 | 0.06 | 0.06 | 0.01 | 0.01 | 0.01 | 1.14 [1.04-1.25]* | 1.09 [0.99-1.20] |
| T39 | 51,500 | 80,036,120 | 0.67 | 0.64 | 0.66 | 0.04 | 0.01 | 0.01 | 1.02 [0.99-1.05] | 1.01 [0.98-1.04] |
| T40 | 25,930 | 81,110,930 | 0.33 | 0.32 | 0.30 | 0.01 | 0.03 | 0.02 | 1.08 [1.04-1.13]* | 1.06 [1.01-1.10] |
| T41 | 620 | 81,599,690 | 0.01 | 0.01 | 0.01 | 0.00 | 0.00 | 0.00 | 1.14 [0.87-1.49] | 1.11 [0.84-1.47] |
| T42 | 18,920 | 81,057,790 | 0.23 | 0.23 | 0.22 | −0.01 | 0.01 | 0.01 | 1.05 [1.00-1.10] | 1.03 [0.98-1.08] |
| T43 | 28,310 | 80,786,820 | 0.36 | 0.35 | 0.34 | 0.01 | 0.02 | 0.01 | 1.06 [1.01-1.10]* | 1.04 [1.00-1.09] |
| T44 | 6,370 | 81,468,970 | 0.08 | 0.08 | 0.07 | 0.00 | 0.01 | 0.00 | 1.07 [0.98-1.16] | 1.06 [0.97-1.16] |
| T45 | 8,230 | 81,438,110 | 0.11 | 0.10 | 0.10 | 0.02 | 0.01 | 0.01 | 1.15 [1.07-1.24]* | 1.11 [1.03-1.20]* |
| T46 | 5,970 | 81,519,360 | 0.08 | 0.07 | 0.07 | 0.00 | 0.01 | 0.01 | 1.12 [1.02-1.23] | 1.09 [0.99-1.19] |
| T47 | 3,370 | 81,548,510 | 0.04 | 0.04 | 0.04 | 0.00 | 0.00 | 0.00 | 1.08 [0.96-1.22] | 1.04 [0.92-1.17] |
| T48 | 1,680 | 81,562,120 | 0.02 | 0.02 | 0.02 | 0.00 | 0.00 | 0.00 | 1.11 [0.95-1.30] | 1.01 [0.85-1.19] |
| T49 | 1,410 | 81,564,820 | 0.02 | 0.02 | 0.02 | 0.00 | 0.00 | 0.00 | 0.99 [0.82-1.18] | 0.97 [0.81-1.16] |
| T50 | 7,580 | 81,350,760 | 0.09 | 0.09 | 0.09 | 0.00 | 0.00 | 0.00 | 1.05 [0.97-1.14] | 1.02 [0.94-1.11] |
| T51 | 23,490 | 80,912,660 | 0.29 | 0.29 | 0.27 | −0.01 | 0.02 | 0.02 | 1.06 [1.01-1.11] | 1.06 [1.01-1.11] |
| T52 | 1,350 | 81,568,250 | 0.02 | 0.02 | 0.02 | 0.00 | 0.00 | 0.00 | 0.87 [0.72-1.05] | 0.86 [0.71-1.04] |
| T54 | 1,360 | 81,579,330 | 0.02 | 0.02 | 0.02 | 0.00 | 0.00 | 0.00 | 0.95 [0.79-1.15] | 0.94 [0.78-1.14] |
| T56 | 700 | 81,594,260 | 0.01 | 0.01 | 0.01 | 0.00 | 0.00 | 0.00 | 1.10 [0.85-1.43] | 1.12 [0.86-1.45] |
| T59 | 1,860 | 81,559,170 | 0.02 | 0.02 | 0.02 | 0.00 | 0.00 | 0.00 | 1.04 [0.89-1.23] | 1.03 [0.87-1.21] |
| T62 | 630 | 81,587,760 | 0.01 | 0.01 | 0.01 | 0.00 | 0.00 | 0.00 | 1.21 [0.93-1.56] | 1.15 [0.88-1.50] |
| T63 | 820 | 81,583,150 | 0.01 | 0.01 | 0.01 | 0.00 | 0.00 | 0.00 | 1.00 [0.78-1.28] | 0.95 [0.74-1.22] |
| T65 | 1,480 | 81,571,230 | 0.02 | 0.02 | 0.02 | 0.00 | 0.00 | 0.00 | 1.01 [0.85-1.20] | 0.99 [0.83-1.18] |
| T68 | 8,650 | 81,569,910 | 0.11 | 0.11 | 0.10 | 0.00 | 0.01 | 0.01 | 1.07 [0.99-1.16] | 1.11 [1.03-1.21]* |
| T71 | 760 | 81,597,110 | 0.01 | 0.01 | 0.01 | 0.00 | 0.00 | 0.00 | 0.88 [0.68-1.14] | 0.89 [0.68-1.16] |
| T74 | 1,400 | 81,578,440 | 0.02 | 0.02 | 0.02 | 0.00 | 0.00 | 0.00 | 0.91 [0.75-1.09] | 0.89 [0.73-1.07] |
| T75 | 900 | 81,579,060 | 0.01 | 0.01 | 0.01 | 0.00 | 0.00 | 0.00 | 1.00 [0.80-1.26] | 0.96 [0.76-1.22] |
| T78 | 20,970 | 81,000,580 | 0.55 | 0.19 | 0.19 | 0.36 | 0.36 | 0.32 | 2.90 [2.78-3.01]* | 2.42 [2.32-2.52]* |
| T79 | 11,120 | 81,432,590 | 0.13 | 0.14 | 0.13 | 0.00 | 0.01 | 0.01 | 1.04 [0.98-1.12] | 1.04 [0.97-1.12] |
| T80 | 5,420 | 81,518,400 | 0.08 | 0.06 | 0.06 | 0.01 | 0.02 | 0.01 | 1.26 [1.15-1.38]* | 1.24 [1.13-1.36]* |
| T81 | 161,790 | 78,353,140 | 2.23 | 2.02 | 1.85 | 0.20 | 0.38 | 0.31 | 1.20 [1.18-1.22]* | 1.16 [1.14-1.18]* |
| T82 | 42,020 | 81,058,860 | 0.55 | 0.51 | 0.46 | 0.04 | 0.09 | 0.08 | 1.19 [1.16-1.23]* | 1.17 [1.13-1.21]* |
| T83 | 40,610 | 81,279,850 | 0.49 | 0.50 | 0.46 | −0.01 | 0.04 | 0.03 | 1.08 [1.04-1.12]* | 1.07 [1.03-1.11]* |
| T84 | 55,170 | 80,604,440 | 0.73 | 0.67 | 0.60 | 0.05 | 0.12 | 0.10 | 1.20 [1.17-1.24]* | 1.17 [1.13-1.20]* |
| T85 | 36,850 | 81,075,090 | 0.49 | 0.45 | 0.41 | 0.04 | 0.08 | 0.07 | 1.19 [1.15-1.23]* | 1.15 [1.11-1.20]* |
| T86 | 5,300 | 81,521,380 | 0.07 | 0.06 | 0.06 | 0.00 | 0.01 | 0.01 | 1.17 [1.07-1.28]* | 1.17 [1.06-1.29]* |
| T87 | 3,180 | 81,563,140 | 0.04 | 0.04 | 0.03 | 0.00 | 0.01 | 0.01 | 1.21 [1.07-1.36]* | 1.17 [1.04-1.32] |
| T88 | 10,810 | 81,338,860 | 0.16 | 0.13 | 0.12 | 0.03 | 0.04 | 0.03 | 1.36 [1.28-1.45]* | 1.27 [1.19-1.35]* |
| T90 | 8,670 | 81,367,960 | 0.11 | 0.10 | 0.10 | 0.01 | 0.02 | 0.01 | 1.16 [1.08-1.25]* | 1.14 [1.06-1.23]* |
| T91 | 4,180 | 81,515,540 | 0.05 | 0.05 | 0.04 | 0.00 | 0.01 | 0.00 | 1.14 [1.02-1.26] | 1.08 [0.97-1.20] |
| T92 | 6,780 | 81,397,600 | 0.08 | 0.08 | 0.08 | 0.00 | 0.01 | 0.01 | 1.09 [1.00-1.18] | 1.07 [0.98-1.16] |
| T93 | 8,770 | 81,354,830 | 0.11 | 0.11 | 0.10 | 0.00 | 0.01 | 0.01 | 1.14 [1.06-1.22]* | 1.09 [1.01-1.18] |
| T94 | 830 | 81,591,120 | 0.01 | 0.01 | 0.01 | 0.00 | 0.00 | 0.00 | 1.11 [0.87-1.41] | 1.06 [0.83-1.35] |
| T95 | 970 | 81,580,890 | 0.01 | 0.01 | 0.01 | 0.00 | 0.00 | 0.00 | 1.06 [0.85-1.31] | 1.08 [0.86-1.34] |
| T98 | 1,560 | 81,579,630 | 0.02 | 0.02 | 0.02 | 0.00 | 0.00 | 0.00 | 1.08 [0.91-1.29] | 1.04 [0.87-1.24] |
| hospitalised - U00-U89 | | | | | | | | | | |
| U07 | 51,060 | 81,559,080 | 0.69 | 0.61 | 0.57 | 0.08 | 0.12 | 0.09 | 1.21 [1.17-1.25]* | 1.15 [1.12-1.19]* |
| U80 | 18,360 | 81,477,260 | 0.27 | 0.21 | 0.19 | 0.06 | 0.08 | 0.07 | 1.40 [1.33-1.47]* | 1.35 [1.29-1.42]* |
| U81 | 1,070 | 81,604,370 | 0.01 | 0.01 | 0.01 | 0.00 | 0.00 | 0.00 | 1.16 [0.94-1.44] | 1.14 [0.92-1.42] |
| U82 | 36,620 | 81,498,750 | 0.53 | 0.43 | 0.40 | 0.10 | 0.13 | 0.12 | 1.32 [1.27-1.37]* | 1.28 [1.23-1.33]* |
| U83 | 21,740 | 81,548,800 | 0.31 | 0.26 | 0.24 | 0.06 | 0.07 | 0.06 | 1.29 [1.23-1.35]* | 1.25 [1.20-1.31]* |
| U84 | 460 | 81,609,200 | 0.01 | 0.01 | 0.01 | 0.00 | 0.00 | 0.00 | 1.20 [0.87-1.66] | 1.19 [0.86-1.66] |
| U88 | 4,470 | 81,582,070 | 0.06 | 0.05 | 0.05 | 0.01 | 0.01 | 0.01 | 1.16 [1.05-1.29]* | 1.12 [1.01-1.24] |
| U89 | 5,130 | 81,575,890 | 0.07 | 0.06 | 0.06 | 0.01 | 0.02 | 0.01 | 1.29 [1.17-1.42]* | 1.25 [1.13-1.38]* |
| hospitalised - V00-Y99 | | | | | | | | | | |
| V01 | 560 | 81,597,380 | 0.01 | 0.01 | 0.01 | 0.00 | 0.00 | 0.00 | 1.02 [0.76-1.36] | 1.00 [0.74-1.33] |
| V03 | 8,530 | 81,340,650 | 0.11 | 0.10 | 0.11 | 0.00 | 0.00 | 0.00 | 0.98 [0.91-1.06] | 0.97 [0.90-1.05] |
| V04 | 540 | 81,592,510 | 0.01 | 0.01 | 0.01 | 0.00 | 0.00 | 0.00 | 1.34 [1.00-1.78] | 1.33 [0.99-1.78] |
| V09 | 2,220 | 81,572,890 | 0.03 | 0.03 | 0.03 | 0.00 | 0.00 | 0.00 | 1.19 [1.03-1.37] | 1.13 [0.98-1.31] |
| V11 | 490 | 81,600,550 | 0.01 | 0.01 | 0.01 | 0.00 | 0.00 | 0.00 | 0.97 [0.71-1.31] | 0.96 [0.71-1.32] |
| V13 | 2,270 | 81,539,150 | 0.02 | 0.03 | 0.03 | 0.00 | −0.01 | −0.01 | 0.81 [0.70-0.95]* | 0.82 [0.70-0.96] |
| V17 | 1,010 | 81,588,190 | 0.01 | 0.01 | 0.01 | 0.00 | 0.00 | 0.00 | 0.87 [0.70-1.09] | 0.88 [0.70-1.11] |
| V18 | 13,980 | 81,195,160 | 0.18 | 0.17 | 0.18 | 0.01 | 0.00 | 0.00 | 0.99 [0.93-1.05] | 0.98 [0.92-1.04] |
| V19 | 2,200 | 81,550,890 | 0.03 | 0.03 | 0.03 | 0.00 | 0.00 | 0.00 | 0.95 [0.82-1.10] | 0.94 [0.81-1.09] |
| V23 | 2,820 | 81,508,900 | 0.03 | 0.04 | 0.03 | −0.01 | 0.00 | 0.00 | 0.89 [0.77-1.01] | 0.91 [0.79-1.05] |
| V27 | 720 | 81,585,660 | 0.01 | 0.01 | 0.01 | 0.00 | 0.00 | 0.00 | 0.80 [0.61-1.06] | 0.78 [0.59-1.04] |
| V28 | 4,050 | 81,478,400 | 0.04 | 0.05 | 0.05 | −0.01 | 0.00 | 0.00 | 0.92 [0.82-1.03] | 0.91 [0.81-1.03] |
| V29 | 1,010 | 81,567,890 | 0.01 | 0.01 | 0.01 | 0.00 | 0.00 | 0.00 | 1.08 [0.87-1.33] | 1.06 [0.85-1.32] |
| V43 | 7,300 | 81,366,180 | 0.09 | 0.09 | 0.08 | 0.00 | 0.01 | 0.00 | 1.07 [0.99-1.16] | 1.04 [0.95-1.12] |
| V44 | 660 | 81,585,990 | 0.01 | 0.01 | 0.01 | 0.00 | 0.00 | 0.00 | 1.15 [0.88-1.49] | 1.13 [0.86-1.48] |
| V47 | 3,130 | 81,513,390 | 0.04 | 0.04 | 0.04 | 0.00 | 0.01 | 0.01 | 1.18 [1.05-1.33]* | 1.16 [1.02-1.31] |
| V48 | 4,310 | 81,517,960 | 0.05 | 0.05 | 0.05 | 0.00 | 0.00 | 0.00 | 1.08 [0.97-1.20] | 1.05 [0.94-1.17] |
| V49 | 3,100 | 81,492,160 | 0.04 | 0.04 | 0.04 | 0.00 | 0.00 | 0.00 | 1.06 [0.94-1.21] | 1.06 [0.93-1.20] |
| V78 | 1,750 | 81,587,700 | 0.02 | 0.02 | 0.02 | 0.00 | 0.00 | 0.00 | 1.16 [0.98-1.37] | 1.13 [0.96-1.34] |
| V79 | 570 | 81,598,950 | 0.01 | 0.01 | 0.01 | 0.00 | 0.00 | 0.00 | 1.07 [0.80-1.44] | 1.00 [0.74-1.36] |
| V80 | 4,290 | 81,476,150 | 0.05 | 0.05 | 0.05 | −0.01 | −0.01 | −0.01 | 0.88 [0.79-0.98] | 0.87 [0.78-0.98] |
| V82 | 440 | 81,603,210 | 0.01 | 0.01 | 0.00 | 0.00 | 0.00 | 0.00 | 1.39 [1.02-1.90] | 1.41 [1.02-1.93] |
| V86 | 1,110 | 81,589,010 | 0.01 | 0.01 | 0.01 | 0.00 | 0.00 | 0.00 | 1.05 [0.85-1.29] | 1.03 [0.84-1.27] |
| V89 | 1,290 | 81,523,200 | 0.02 | 0.02 | 0.01 | 0.00 | 0.00 | 0.00 | 1.15 [0.96-1.39] | 1.10 [0.91-1.34] |
| W00 | 6,310 | 81,465,120 | 0.07 | 0.08 | 0.07 | −0.01 | 0.00 | 0.00 | 1.04 [0.96-1.14] | 1.03 [0.95-1.13] |
| W01 | 140,330 | 79,568,580 | 1.85 | 1.74 | 1.68 | 0.10 | 0.17 | 0.14 | 1.10 [1.08-1.12]* | 1.08 [1.06-1.10]* |
| W02 | 6,510 | 81,410,890 | 0.09 | 0.08 | 0.09 | 0.01 | 0.00 | 0.00 | 0.99 [0.91-1.07] | 0.97 [0.89-1.06] |
| W03 | 8,960 | 81,344,760 | 0.12 | 0.11 | 0.12 | 0.01 | 0.00 | −0.01 | 0.98 [0.91-1.05] | 0.95 [0.88-1.02] |
| W04 | 1,660 | 81,492,410 | 0.02 | 0.02 | 0.02 | 0.00 | 0.00 | 0.00 | 1.01 [0.86-1.20] | 1.03 [0.88-1.22] |
| W05 | 3,340 | 81,581,160 | 0.04 | 0.04 | 0.04 | 0.00 | 0.01 | 0.01 | 1.15 [1.02-1.30] | 1.14 [1.01-1.29] |
| W06 | 36,400 | 81,292,760 | 0.48 | 0.44 | 0.43 | 0.04 | 0.05 | 0.04 | 1.12 [1.08-1.16]* | 1.10 [1.06-1.14]* |
| W07 | 21,230 | 81,352,910 | 0.27 | 0.26 | 0.25 | 0.02 | 0.02 | 0.02 | 1.08 [1.03-1.13]* | 1.06 [1.01-1.12] |
| W08 | 7,150 | 81,474,430 | 0.09 | 0.09 | 0.09 | 0.01 | 0.00 | 0.00 | 1.04 [0.96-1.13] | 1.03 [0.95-1.12] |
| W09 | 18,400 | 81,204,130 | 0.27 | 0.22 | 0.27 | 0.06 | 0.01 | 0.00 | 1.02 [0.97-1.07] | 1.01 [0.96-1.06] |
| W10 | 58,540 | 80,599,250 | 0.75 | 0.72 | 0.67 | 0.02 | 0.07 | 0.06 | 1.11 [1.08-1.14]* | 1.09 [1.06-1.12]* |
| W11 | 7,720 | 81,390,510 | 0.08 | 0.10 | 0.09 | −0.01 | 0.00 | 0.00 | 0.99 [0.91-1.07] | 0.97 [0.89-1.05] |
| W13 | 6,440 | 81,394,090 | 0.08 | 0.08 | 0.08 | 0.00 | 0.00 | 0.00 | 0.97 [0.89-1.06] | 0.97 [0.89-1.06] |
| W14 | 2,090 | 81,536,070 | 0.03 | 0.02 | 0.03 | 0.01 | 0.00 | 0.00 | 1.00 [0.87-1.16] | 1.01 [0.87-1.17] |
| W17 | 12,640 | 81,258,120 | 0.16 | 0.15 | 0.16 | 0.01 | 0.00 | 0.00 | 1.00 [0.94-1.06] | 0.99 [0.93-1.05] |
| W18 | 98,890 | 80,698,910 | 1.31 | 1.20 | 1.15 | 0.11 | 0.16 | 0.14 | 1.14 [1.11-1.16]* | 1.12 [1.09-1.14]* |

| | | | | | | | | | | |
|---|---|---|---|---|---|---|---|---|---|---|
| W19 | 215,560 | 79,286,030 | 2.92 | 2.67 | 2.57 | 0.24 | 0.34 | 0.31 | 1.13 [1.12-1.15]* | 1.12 [1.10-1.14]* |
| W20 | 5,830 | 81,449,950 | 0.07 | 0.07 | 0.07 | 0.00 | 0.00 | 0.00 | 0.99 [0.90-1.08] | 0.97 [0.89-1.07] |
| W21 | 4,730 | 81,466,520 | 0.07 | 0.06 | 0.07 | 0.01 | 0.00 | 0.00 | 1.04 [0.95-1.15] | 1.02 [0.92-1.13] |
| W22 | 30,700 | 81,000,470 | 0.41 | 0.37 | 0.38 | 0.03 | 0.03 | 0.02 | 1.07 [1.03-1.12]* | 1.05 [1.01-1.09] |
| W23 | 19,030 | 81,166,550 | 0.24 | 0.23 | 0.25 | 0.01 | −0.01 | −0.01 | 0.97 [0.92-1.02] | 0.96 [0.92-1.01] |
| W25 | 8,820 | 81,258,060 | 0.11 | 0.11 | 0.11 | 0.00 | 0.00 | 0.00 | 0.98 [0.91-1.05] | 0.97 [0.90-1.05] |
| W26 | 8,380 | 81,424,060 | 0.10 | 0.10 | 0.10 | 0.00 | 0.00 | 0.00 | 0.99 [0.92-1.07] | 0.98 [0.91-1.06] |
| W27 | 3,770 | 81,487,560 | 0.04 | 0.05 | 0.04 | 0.00 | 0.00 | 0.00 | 0.97 [0.87-1.09] | 0.96 [0.85-1.08] |
| W29 | 3,910 | 81,509,180 | 0.04 | 0.05 | 0.04 | −0.01 | 0.00 | 0.00 | 0.90 [0.80-1.01] | 0.91 [0.81-1.02] |
| W31 | 3,430 | 81,487,010 | 0.04 | 0.04 | 0.04 | −0.01 | 0.00 | 0.00 | 0.93 [0.82-1.05] | 0.92 [0.81-1.04] |
| W44 | 21,220 | 81,150,610 | 0.29 | 0.26 | 0.27 | 0.03 | 0.01 | 0.00 | 1.05 [1.00-1.10] | 1.02 [0.97-1.06] |
| W45 | 6,550 | 81,410,540 | 0.08 | 0.08 | 0.08 | 0.00 | 0.00 | 0.00 | 1.01 [0.93-1.10] | 0.99 [0.90-1.07] |
| W46 | 1,020 | 81,603,260 | 0.01 | 0.01 | 0.01 | 0.00 | 0.00 | 0.00 | 0.86 [0.69-1.09] | 0.85 [0.67-1.08] |
| W49 | 1,660 | 81,551,350 | 0.02 | 0.02 | 0.02 | 0.00 | 0.00 | 0.00 | 1.20 [1.02-1.42] | 1.18 [1.00-1.39] |
| W50 | 8,910 | 81,359,480 | 0.12 | 0.11 | 0.11 | 0.02 | 0.01 | 0.01 | 1.06 [0.99-1.14] | 1.05 [0.98-1.13] |
| W51 | 6,820 | 81,403,420 | 0.09 | 0.08 | 0.09 | 0.01 | 0.00 | 0.00 | 1.05 [0.97-1.13] | 1.02 [0.94-1.11] |
| W54 | 9,950 | 81,409,710 | 0.12 | 0.12 | 0.13 | −0.01 | −0.01 | −0.01 | 0.93 [0.86-0.99] | 0.91 [0.85-0.98]* |
| W55 | 4,440 | 81,522,680 | 0.05 | 0.06 | 0.05 | −0.01 | 0.00 | −0.01 | 0.92 [0.83-1.03] | 0.88 [0.79-0.98] |
| W57 | 5,650 | 81,503,830 | 0.08 | 0.07 | 0.06 | 0.01 | 0.02 | 0.02 | 1.29 [1.19-1.41]* | 1.24 [1.13-1.36]* |
| W60 | 510 | 81,600,230 | 0.01 | 0.01 | 0.01 | 0.00 | 0.00 | 0.00 | 1.29 [0.97-1.71] | 1.24 [0.93-1.66] |
| W64 | 1,160 | 81,595,720 | 0.01 | 0.01 | 0.01 | 0.00 | 0.00 | 0.00 | 1.11 [0.91-1.36] | 1.11 [0.91-1.37] |
| W78 | 3,580 | 81,596,640 | 0.04 | 0.04 | 0.04 | 0.00 | 0.00 | 0.00 | 1.05 [0.93-1.19] | 1.06 [0.94-1.20] |
| W79 | 5,380 | 81,565,780 | 0.07 | 0.06 | 0.06 | 0.01 | 0.01 | 0.01 | 1.22 [1.11-1.34]* | 1.18 [1.07-1.30]* |
| W80 | 2,250 | 81,596,270 | 0.03 | 0.03 | 0.03 | 0.00 | 0.00 | 0.00 | 1.10 [0.95-1.29] | 1.06 [0.91-1.24] |
| W84 | 2,100 | 81,604,260 | 0.03 | 0.03 | 0.02 | 0.00 | 0.00 | 0.00 | 1.02 [0.87-1.20] | 1.01 [0.86-1.19] |
| X08 | 600 | 81,596,800 | 0.01 | 0.01 | 0.01 | 0.00 | 0.00 | 0.00 | 0.98 [0.74-1.31] | 0.96 [0.72-1.29] |
| X09 | 830 | 81,588,930 | 0.01 | 0.01 | 0.01 | 0.00 | 0.00 | 0.00 | 0.96 [0.75-1.24] | 0.94 [0.73-1.21] |
| X10 | 3,920 | 81,512,100 | 0.05 | 0.05 | 0.05 | 0.00 | 0.00 | 0.00 | 0.99 [0.89-1.11] | 0.99 [0.89-1.11] |
| X11 | 1,130 | 81,575,610 | 0.01 | 0.01 | 0.01 | 0.00 | 0.00 | 0.00 | 0.95 [0.77-1.17] | 0.93 [0.75-1.15] |
| X12 | 2,280 | 81,545,120 | 0.03 | 0.03 | 0.03 | 0.00 | 0.00 | 0.00 | 0.92 [0.80-1.06] | 0.91 [0.78-1.05] |
| X15 | 1,720 | 81,573,520 | 0.02 | 0.02 | 0.02 | 0.00 | 0.00 | 0.00 | 1.00 [0.85-1.17] | 1.00 [0.85-1.18] |
| X16 | 1,180 | 81,588,880 | 0.01 | 0.01 | 0.01 | 0.00 | 0.00 | 0.00 | 0.95 [0.77-1.17] | 0.95 [0.77-1.18] |
| X19 | 740 | 81,594,220 | 0.01 | 0.01 | 0.01 | 0.00 | 0.00 | 0.00 | 1.02 [0.80-1.31] | 0.99 [0.77-1.28] |
| X23 | 1,070 | 81,584,320 | 0.01 | 0.01 | 0.01 | 0.00 | 0.00 | 0.00 | 1.24 [1.01-1.52] | 1.18 [0.96-1.46] |
| X31 | 1,180 | 81,602,500 | 0.01 | 0.01 | 0.01 | 0.00 | 0.00 | 0.00 | 0.97 [0.78-1.21] | 1.01 [0.80-1.26] |
| X40 | 13,870 | 81,251,810 | 0.17 | 0.17 | 0.17 | 0.00 | 0.00 | 0.00 | 1.02 [0.96-1.08] | 1.00 [0.94-1.06] |
| X41 | 8,480 | 81,375,840 | 0.11 | 0.10 | 0.10 | 0.00 | 0.01 | 0.00 | 1.06 [0.99-1.14] | 1.05 [0.97-1.13] |
| X42 | 7,960 | 81,495,760 | 0.10 | 0.10 | 0.09 | 0.01 | 0.01 | 0.01 | 1.13 [1.05-1.22]* | 1.10 [1.02-1.19] |
| X43 | 1,800 | 81,573,040 | 0.02 | 0.02 | 0.02 | 0.00 | 0.00 | 0.00 | 1.02 [0.86-1.21] | 1.02 [0.86-1.20] |
| X44 | 12,040 | 81,346,450 | 0.16 | 0.15 | 0.14 | 0.01 | 0.01 | 0.01 | 1.09 [1.03-1.16]* | 1.06 [1.00-1.13] |
| X45 | 3,180 | 81,509,430 | 0.04 | 0.04 | 0.04 | 0.00 | 0.00 | 0.00 | 1.01 [0.89-1.14] | 1.00 [0.88-1.14] |
| X46 | 920 | 81,579,010 | 0.01 | 0.01 | 0.01 | 0.00 | 0.00 | 0.00 | 0.91 [0.72-1.14] | 0.90 [0.71-1.12] |
| X47 | 1,120 | 81,581,920 | 0.01 | 0.01 | 0.01 | 0.00 | 0.00 | 0.00 | 1.08 [0.88-1.33] | 1.06 [0.86-1.31] |
| X49 | 4,490 | 81,478,370 | 0.07 | 0.05 | 0.05 | 0.02 | 0.02 | 0.01 | 1.31 [1.19-1.44]* | 1.24 [1.12-1.36]* |
| X50 | 15,540 | 81,285,960 | 0.19 | 0.19 | 0.18 | 0.00 | 0.01 | 0.00 | 1.05 [0.99-1.11] | 1.01 [0.95-1.07] |
| X51 | 1,030 | 81,587,710 | 0.01 | 0.01 | 0.01 | 0.00 | 0.00 | 0.00 | 1.06 [0.86-1.32] | 1.04 [0.84-1.30] |
| X58 | 4,900 | 81,528,870 | 0.08 | 0.06 | 0.05 | 0.03 | 0.03 | 0.02 | 1.50 [1.36-1.64]* | 1.38 [1.26-1.52]* |
| X59 | 65,630 | 80,132,300 | 0.89 | 0.80 | 0.78 | 0.09 | 0.11 | 0.09 | 1.14 [1.11-1.17]* | 1.11 [1.08-1.14]* |
| X60 | 40,040 | 80,374,800 | 0.52 | 0.49 | 0.51 | 0.03 | 0.01 | 0.01 | 1.03 [0.99-1.06] | 1.01 [0.98-1.05] |
| X61 | 30,720 | 80,704,550 | 0.38 | 0.38 | 0.37 | 0.00 | 0.02 | 0.01 | 1.05 [1.01-1.09] | 1.04 [1.00-1.08] |
| X62 | 18,130 | 81,234,150 | 0.23 | 0.22 | 0.21 | 0.00 | 0.01 | 0.01 | 1.07 [1.01-1.12] | 1.04 [0.98-1.09] |
| X63 | 4,510 | 81,512,000 | 0.06 | 0.05 | 0.05 | 0.00 | 0.00 | 0.00 | 1.06 [0.95-1.17] | 1.05 [0.95-1.17] |
| X64 | 15,320 | 81,181,840 | 0.20 | 0.19 | 0.19 | 0.02 | 0.02 | 0.01 | 1.09 [1.03-1.15]* | 1.07 [1.01-1.13] |
| X65 | 21,240 | 81,021,790 | 0.26 | 0.26 | 0.24 | 0.00 | 0.02 | 0.02 | 1.08 [1.03-1.13]* | 1.07 [1.02-1.13]* |
| X69 | 1,580 | 81,574,240 | 0.02 | 0.02 | 0.02 | 0.00 | 0.00 | 0.00 | 0.95 [0.79-1.13] | 0.94 [0.79-1.13] |
| X70 | 1,210 | 81,591,880 | 0.01 | 0.02 | 0.02 | 0.00 | 0.00 | 0.00 | 0.88 [0.71-1.08] | 0.89 [0.72-1.09] |
| X78 | 10,230 | 81,414,310 | 0.13 | 0.12 | 0.13 | 0.01 | 0.00 | 0.00 | 1.01 [0.95-1.09] | 0.99 [0.92-1.06] |
| X83 | 980 | 81,597,870 | 0.01 | 0.01 | 0.01 | 0.00 | 0.00 | 0.00 | 0.85 [0.67-1.07] | 0.82 [0.65-1.04] |
| X84 | 1,430 | 81,585,310 | 0.02 | 0.02 | 0.02 | 0.00 | 0.00 | 0.00 | 0.97 [0.80-1.17] | 0.91 [0.75-1.10] |
| X99 | 3,780 | 81,475,740 | 0.04 | 0.05 | 0.05 | 0.00 | 0.00 | 0.00 | 0.96 [0.85-1.07] | 0.97 [0.87-1.10] |
| Y00 | 2,170 | 81,526,480 | 0.02 | 0.03 | 0.03 | 0.00 | 0.00 | 0.00 | 0.95 [0.81-1.11] | 0.97 [0.83-1.14] |
| Y04 | 17,110 | 80,902,910 | 0.20 | 0.21 | 0.21 | −0.01 | −0.01 | −0.01 | 0.97 [0.92-1.03] | 0.97 [0.92-1.03] |
| Y07 | 1,500 | 81,573,720 | 0.02 | 0.02 | 0.02 | 0.00 | 0.00 | 0.00 | 0.88 [0.73-1.05] | 0.87 [0.72-1.04] |
| Y08 | 850 | 81,581,940 | 0.01 | 0.01 | 0.01 | 0.00 | 0.00 | 0.00 | 0.86 [0.67-1.11] | 0.89 [0.68-1.16] |
| Y09 | 2,510 | 81,489,410 | 0.03 | 0.03 | 0.03 | 0.00 | 0.00 | 0.00 | 1.02 [0.88-1.17] | 1.02 [0.88-1.17] |
| Y28 | 640 | 81,587,680 | 0.01 | 0.01 | 0.01 | 0.00 | 0.00 | 0.00 | 0.93 [0.71-1.24] | 0.87 [0.65-1.16] |
| Y34 | 560 | 81,584,930 | 0.01 | 0.01 | 0.01 | 0.00 | 0.00 | 0.00 | 1.02 [0.77-1.37] | 0.99 [0.74-1.34] |
| Y40 | 18,560 | 81,367,390 | 0.28 | 0.22 | 0.20 | 0.06 | 0.08 | 0.07 | 1.40 [1.33-1.47]* | 1.32 [1.26-1.39]* |
| Y41 | 3,870 | 81,559,070 | 0.06 | 0.05 | 0.04 | 0.01 | 0.01 | 0.01 | 1.29 [1.16-1.44]* | 1.21 [1.08-1.35]* |
| Y42 | 15,240 | 81,442,400 | 0.22 | 0.18 | 0.16 | 0.04 | 0.05 | 0.04 | 1.33 [1.26-1.40]* | 1.22 [1.15-1.28]* |
| Y43 | 30,430 | 81,341,340 | 0.39 | 0.37 | 0.34 | 0.02 | 0.05 | 0.05 | 1.15 [1.11-1.19]* | 1.14 [1.10-1.19]* |
| Y44 | 10,610 | 81,515,460 | 0.14 | 0.13 | 0.12 | 0.01 | 0.02 | 0.02 | 1.19 [1.11-1.27]* | 1.15 [1.07-1.23]* |
| Y45 | 24,130 | 81,343,150 | 0.33 | 0.29 | 0.27 | 0.04 | 0.07 | 0.05 | 1.25 [1.19-1.30]* | 1.19 [1.14-1.25]* |
| Y46 | 3,680 | 81,563,690 | 0.05 | 0.04 | 0.04 | 0.01 | 0.01 | 0.01 | 1.23 [1.11-1.38]* | 1.19 [1.06-1.33]* |
| Y47 | 1,840 | 81,588,900 | 0.02 | 0.02 | 0.02 | 0.00 | 0.00 | 0.00 | 1.17 [0.99-1.37] | 1.13 [0.96-1.33] |
| Y48 | 2,100 | 81,579,960 | 0.03 | 0.03 | 0.02 | 0.00 | 0.00 | 0.00 | 1.20 [1.04-1.39] | 1.14 [0.98-1.32] |
| Y49 | 5,880 | 81,544,440 | 0.08 | 0.07 | 0.06 | 0.01 | 0.02 | 0.02 | 1.28 [1.18-1.40]* | 1.27 [1.16-1.39]* |
| Y51 | 8,950 | 81,520,030 | 0.12 | 0.11 | 0.10 | 0.01 | 0.02 | 0.02 | 1.19 [1.10-1.28]* | 1.18 [1.10-1.27]* |
| Y52 | 21,640 | 81,430,220 | 0.29 | 0.26 | 0.23 | 0.03 | 0.06 | 0.05 | 1.24 [1.18-1.30]* | 1.21 [1.15-1.27]* |
| Y53 | 4,150 | 81,577,550 | 0.06 | 0.05 | 0.05 | 0.01 | 0.01 | 0.01 | 1.31 [1.18-1.46]* | 1.22 [1.10-1.36]* |
| Y54 | 16,690 | 81,501,780 | 0.23 | 0.20 | 0.18 | 0.04 | 0.05 | 0.04 | 1.29 [1.22-1.36]* | 1.24 [1.17-1.30]* |
| Y55 | 2,280 | 81,583,440 | 0.03 | 0.03 | 0.03 | 0.01 | 0.01 | 0.00 | 1.32 [1.15-1.51]* | 1.16 [1.01-1.34] |
| Y56 | 1,880 | 81,570,290 | 0.03 | 0.02 | 0.02 | 0.01 | 0.01 | 0.01 | 1.65 [1.43-1.90]* | 1.51 [1.31-1.75]* |
| Y57 | 10,020 | 81,514,730 | 0.14 | 0.12 | 0.11 | 0.02 | 0.03 | 0.03 | 1.33 [1.24-1.42]* | 1.25 [1.17-1.34]* |
| Y59 | 1,260 | 81,583,470 | 0.02 | 0.01 | 0.01 | 0.01 | 0.01 | 0.00 | 1.41 [1.18-1.69]* | 1.30 [1.08-1.57]* |
| Y60 | 12,010 | 81,487,110 | 0.16 | 0.15 | 0.13 | 0.01 | 0.02 | 0.02 | 1.17 [1.10-1.24]* | 1.13 [1.06-1.21]* |
| Y65 | 1,660 | 81,587,730 | 0.02 | 0.02 | 0.02 | 0.00 | 0.00 | 0.00 | 1.10 [0.93-1.30] | 1.08 [0.91-1.28] |
| Y71 | 800 | 81,598,670 | 0.01 | 0.01 | 0.01 | 0.00 | 0.00 | 0.00 | 1.38 [1.09-1.74]* | 1.32 [1.04-1.67] |
| Y73 | 740 | 81,600,600 | 0.01 | 0.01 | 0.01 | 0.00 | 0.00 | 0.00 | 0.92 [0.70-1.21] | 0.89 [0.67-1.18] |
| Y77 | 760 | 81,601,740 | 0.01 | 0.01 | 0.01 | 0.00 | 0.00 | 0.00 | 1.05 [0.81-1.37] | 1.03 [0.79-1.35] |
| Y79 | 1,690 | 81,577,640 | 0.02 | 0.02 | 0.02 | 0.00 | 0.00 | 0.00 | 1.15 [0.97-1.36] | 1.10 [0.93-1.31] |
| Y82 | 650 | 81,598,200 | 0.01 | 0.01 | 0.01 | 0.00 | 0.00 | 0.00 | 1.20 [0.92-1.56] | 1.19 [0.91-1.55] |
| Y83 | 218,840 | 77,712,220 | 3.01 | 2.77 | 2.54 | 0.24 | 0.47 | 0.38 | 1.18 [1.17-1.20]* | 1.15 [1.13-1.16]* |
| Y84 | 49,800 | 81,143,380 | 0.64 | 0.61 | 0.55 | 0.04 | 0.09 | 0.08 | 1.16 [1.13-1.20]* | 1.14 [1.10-1.18]* |
| Y85 | 5,210 | 81,479,470 | 0.07 | 0.06 | 0.06 | 0.00 | 0.01 | 0.01 | 1.18 [1.07-1.29]* | 1.13 [1.02-1.24] |
| Y86 | 9,340 | 81,348,370 | 0.12 | 0.11 | 0.11 | 0.00 | 0.01 | 0.01 | 1.09 [1.02-1.17] | 1.06 [0.98-1.14] |
| Y87 | 1,570 | 81,564,890 | 0.02 | 0.02 | 0.02 | 0.00 | 0.00 | 0.00 | 1.16 [0.98-1.37] | 1.14 [0.96-1.36] |
| Y88 | 1,990 | 81,577,140 | 0.02 | 0.02 | 0.02 | 0.00 | 0.00 | 0.00 | 1.02 [0.87-1.20] | 0.97 [0.83-1.14] |
| Y89 | 590 | 81,594,950 | 0.01 | 0.01 | 0.01 | 0.00 | 0.00 | 0.00 | 1.06 [0.80-1.40] | 1.06 [0.80-1.42] |
| Y90 | 1,080 | 81,594,690 | 0.01 | 0.01 | 0.01 | 0.00 | 0.00 | 0.00 | 0.98 [0.80-1.21] | 0.99 [0.80-1.23] |
| Y91 | 1,970 | 81,555,200 | 0.02 | 0.02 | 0.02 | 0.00 | 0.00 | 0.00 | 1.10 [0.94-1.29] | 1.11 [0.95-1.31] |
| Y95 | 113,580 | 81,310,160 | 1.52 | 1.37 | 1.27 | 0.15 | 0.25 | 0.22 | 1.20 [1.17-1.22]* | 1.17 [1.14-1.19]* |
| hospitalised - Z00-Z99 | | | | | | | | | | |
| Z00 | 4,520 | 81,426,880 | 0.06 | 0.06 | 0.05 | 0.00 | 0.01 | 0.00 | 1.10 [1.00-1.22] | 1.07 [0.97-1.19] |
| Z01 | 26,600 | 80,106,450 | 0.63 | 0.26 | 0.27 | 0.37 | 0.35 | 0.30 | 2.28 [2.20-2.36]* | 1.90 [1.83-1.98]* |
| Z02 | 17,230 | 81,402,100 | 0.21 | 0.21 | 0.19 | 0.00 | 0.02 | 0.01 | 1.10 [1.04-1.16]* | 1.07 [1.01-1.13] |

| | | | | | | | | | | |
|---|---|---|---|---|---|---|---|---|---|---|
| Z03 | 140,810 | 77,422,570 | 2.08 | 1.75 | 1.65 | 0.32 | 0.42 | 0.34 | 1.26 [1.23-1.28]* | 1.20 [1.17-1.22]* |
| Z04 | 24,330 | 80,370,990 | 0.32 | 0.30 | 0.29 | 0.03 | 0.04 | 0.03 | 1.12 [1.08-1.17]* | 1.09 [1.04-1.14]* |
| Z08 | 41,140 | 80,740,920 | 0.48 | 0.52 | 0.46 | −0.03 | 0.02 | 0.02 | 1.05 [1.01-1.09]* | 1.05 [1.01-1.09]* |
| Z09 | 83,650 | 79,753,210 | 1.10 | 1.04 | 0.94 | 0.07 | 0.17 | 0.12 | 1.18 [1.15-1.20]* | 1.12 [1.10-1.15]* |
| Z11 | 260,640 | 81,037,100 | 3.49 | 3.15 | 3.04 | 0.34 | 0.46 | 0.37 | 1.15 [1.13-1.17]* | 1.12 [1.10-1.13]* |
| Z12 | 53,890 | 80,923,370 | 0.64 | 0.67 | 0.58 | −0.04 | 0.06 | 0.04 | 1.10 [1.06-1.13]* | 1.07 [1.04-1.11]* |
| Z13 | 152,810 | 78,842,970 | 2.13 | 1.89 | 1.79 | 0.24 | 0.34 | 0.27 | 1.19 [1.17-1.21]* | 1.14 [1.12-1.17]* |
| Z20 | 9,230 | 81,532,300 | 0.12 | 0.11 | 0.11 | 0.01 | 0.02 | 0.01 | 1.16 [1.08-1.25]* | 1.12 [1.04-1.21]* |
| Z22 | 77,930 | 80,655,920 | 1.12 | 0.93 | 0.86 | 0.19 | 0.26 | 0.23 | 1.30 [1.27-1.33]* | 1.26 [1.23-1.29]* |
| Z24 | 1,350 | 81,489,410 | 0.02 | 0.02 | 0.02 | 0.00 | 0.00 | 0.00 | 1.07 [0.89-1.29] | 1.09 [0.90-1.32] |
| Z25 | 800 | 81,588,150 | 0.02 | 0.01 | 0.01 | 0.01 | 0.01 | 0.01 | 2.63 [2.15-3.22]* | 2.17 [1.75-2.69]* |
| Z27 | 3,890 | 81,426,870 | 0.08 | 0.04 | 0.04 | 0.05 | 0.05 | 0.04 | 2.26 [2.06-2.48]* | 2.04 [1.85-2.25]* |
| Z29 | 42,620 | 80,382,840 | 0.58 | 0.52 | 0.48 | 0.06 | 0.10 | 0.08 | 1.20 [1.16-1.24]* | 1.16 [1.12-1.20]* |
| Z30 | 42,980 | 78,753,240 | 0.52 | 0.55 | 0.49 | −0.03 | 0.03 | 0.02 | 1.06 [1.02-1.09]* | 1.05 [1.01-1.08] |
| Z31 | 590 | 81,484,240 | 0.01 | 0.01 | 0.01 | 0.00 | 0.00 | 0.00 | 1.20 [0.91-1.57] | 1.15 [0.86-1.53] |
| Z32 | 3,340 | 81,416,190 | 0.04 | 0.04 | 0.04 | 0.00 | 0.00 | 0.00 | 1.09 [0.97-1.23] | 1.07 [0.95-1.21] |
| Z33 | 31,360 | 80,365,070 | 0.41 | 0.38 | 0.37 | 0.03 | 0.05 | 0.03 | 1.12 [1.08-1.17]* | 1.08 [1.04-1.12]* |
| Z34 | 73,730 | 78,565,720 | 0.96 | 0.93 | 0.94 | 0.03 | 0.03 | 0.02 | 1.03 [1.00-1.05] | 1.02 [0.99-1.04] |
| Z35 | 79,400 | 79,883,840 | 0.96 | 1.00 | 0.96 | −0.05 | 0.00 | 0.00 | 1.00 [0.98-1.02] | 1.00 [0.98-1.03] |
| Z36 | 12,550 | 80,304,960 | 0.16 | 0.16 | 0.15 | 0.00 | 0.01 | 0.01 | 1.05 [0.99-1.11] | 1.05 [0.98-1.11] |
| Z37 | 153,410 | 66,916,600 | 2.98 | 2.11 | 2.99 | 0.87 | −0.01 | 0.01 | 1.00 [0.98-1.01] | 1.00 [0.99-1.02] |
| Z38 | 800 | 57,877,130 | 0.01 | 0.01 | 0.01 | 0.00 | 0.00 | 0.00 | 1.02 [0.80-1.31] | 1.01 [0.79-1.29] |
| Z39 | 21,870 | 80,456,560 | 0.27 | 0.27 | 0.26 | 0.00 | 0.01 | 0.01 | 1.03 [0.98-1.08] | 1.03 [0.98-1.08] |
| Z40 | 5,470 | 81,522,190 | 0.06 | 0.07 | 0.06 | 0.00 | 0.00 | 0.00 | 1.02 [0.93-1.12] | 1.01 [0.92-1.12] |
| Z41 | 5,670 | 81,358,940 | 0.08 | 0.07 | 0.07 | 0.01 | 0.01 | 0.01 | 1.22 [1.12-1.33]* | 1.21 [1.11-1.32]* |
| Z42 | 14,460 | 81,276,510 | 0.18 | 0.18 | 0.16 | 0.00 | 0.02 | 0.02 | 1.11 [1.05-1.17]* | 1.11 [1.05-1.18]* |
| Z43 | 20,920 | 81,181,220 | 0.26 | 0.26 | 0.24 | 0.00 | 0.02 | 0.02 | 1.09 [1.04-1.14]* | 1.08 [1.03-1.13]* |
| Z45 | 45,600 | 80,955,240 | 0.56 | 0.56 | 0.51 | 0.00 | 0.05 | 0.04 | 1.10 [1.06-1.13]* | 1.07 [1.04-1.11]* |
| Z46 | 51,820 | 80,642,250 | 0.63 | 0.65 | 0.59 | −0.02 | 0.04 | 0.04 | 1.07 [1.04-1.10]* | 1.06 [1.03-1.09]* |
| Z47 | 38,390 | 80,279,350 | 0.52 | 0.47 | 0.48 | 0.05 | 0.04 | 0.03 | 1.08 [1.05-1.12]* | 1.06 [1.03-1.10]* |
| Z48 | 14,840 | 81,121,260 | 0.20 | 0.18 | 0.17 | 0.02 | 0.03 | 0.02 | 1.15 [1.08-1.21]* | 1.12 [1.06-1.19]* |
| Z49 | 8,790 | 81,492,870 | 0.12 | 0.11 | 0.09 | 0.01 | 0.02 | 0.02 | 1.25 [1.17-1.34]* | 1.26 [1.17-1.36]* |
| Z50 | 303,650 | 79,443,320 | 4.05 | 3.77 | 3.56 | 0.28 | 0.49 | 0.42 | 1.14 [1.12-1.15]* | 1.12 [1.10-1.13]* |
| Z51 | 356,290 | 78,073,660 | 4.78 | 4.51 | 4.14 | 0.27 | 0.64 | 0.56 | 1.16 [1.14-1.17]* | 1.13 [1.12-1.15]* |
| Z52 | 1,590 | 81,560,440 | 0.02 | 0.02 | 0.02 | 0.00 | 0.00 | 0.00 | 0.94 [0.79-1.13] | 0.96 [0.80-1.15] |
| Z53 | 305,670 | 75,087,910 | 4.45 | 3.97 | 3.66 | 0.48 | 0.79 | 0.64 | 1.21 [1.20-1.23]* | 1.17 [1.15-1.18]* |
| Z54 | 2,320 | 81,529,700 | 0.03 | 0.03 | 0.03 | 0.00 | 0.01 | 0.01 | 1.27 [1.11-1.47]* | 1.27 [1.10-1.46]* |
| Z55 | 1,320 | 81,588,450 | 0.02 | 0.01 | 0.02 | 0.01 | 0.00 | 0.00 | 1.18 [0.99-1.41] | 1.15 [0.96-1.38] |
| Z56 | 3,760 | 81,549,650 | 0.04 | 0.05 | 0.04 | 0.00 | 0.00 | 0.00 | 1.07 [0.96-1.20] | 1.04 [0.92-1.16] |
| Z57 | 11,140 | 81,532,410 | 0.14 | 0.13 | 0.12 | 0.01 | 0.03 | 0.02 | 1.21 [1.14-1.29]* | 1.16 [1.09-1.24]* |
| Z58 | 8,480 | 81,532,670 | 0.12 | 0.10 | 0.10 | 0.02 | 0.02 | 0.02 | 1.24 [1.15-1.33]* | 1.16 [1.08-1.25]* |
| Z59 | 8,840 | 81,488,960 | 0.10 | 0.11 | 0.10 | −0.01 | 0.00 | 0.00 | 1.00 [0.93-1.08] | 1.03 [0.96-1.12] |
| Z60 | 93,120 | 80,346,040 | 1.19 | 1.15 | 1.07 | 0.04 | 0.13 | 0.11 | 1.12 [1.09-1.15]* | 1.10 [1.08-1.13]* |
| Z61 | 4,270 | 81,538,500 | 0.06 | 0.05 | 0.06 | 0.00 | 0.00 | 0.00 | 0.95 [0.86-1.06] | 0.94 [0.85-1.05] |
| Z62 | 930 | 81,585,740 | 0.01 | 0.01 | 0.01 | 0.00 | 0.00 | 0.00 | 0.87 [0.69-1.08] | 0.84 [0.67-1.06] |
| Z63 | 32,370 | 81,131,070 | 0.41 | 0.40 | 0.39 | 0.02 | 0.02 | 0.01 | 1.05 [1.01-1.09] | 1.03 [0.99-1.07] |
| Z64 | 670 | 81,563,980 | 0.01 | 0.01 | 0.01 | 0.00 | 0.00 | 0.00 | 0.99 [0.75-1.30] | 0.98 [0.74-1.30] |
| Z65 | 2,730 | 81,564,730 | 0.03 | 0.03 | 0.03 | 0.00 | 0.00 | 0.00 | 1.00 [0.87-1.14] | 1.01 [0.88-1.16] |
| Z71 | 32,190 | 81,070,080 | 0.55 | 0.36 | 0.35 | 0.19 | 0.20 | 0.16 | 1.57 [1.51-1.62]* | 1.43 [1.38-1.48]* |
| Z72 | 180,940 | 75,101,660 | 2.38 | 2.42 | 2.15 | −0.03 | 0.23 | 0.25 | 1.11 [1.09-1.12]* | 1.11 [1.10-1.13]* |
| Z73 | 51,680 | 81,334,090 | 0.67 | 0.63 | 0.59 | 0.04 | 0.08 | 0.06 | 1.13 [1.09-1.16]* | 1.11 [1.07-1.14]* |
| Z74 | 39,090 | 81,339,670 | 0.49 | 0.48 | 0.44 | 0.01 | 0.05 | 0.05 | 1.12 [1.08-1.16]* | 1.12 [1.08-1.16]* |
| Z75 | 77,150 | 81,182,540 | 0.98 | 0.94 | 0.89 | 0.04 | 0.09 | 0.09 | 1.10 [1.07-1.13]* | 1.10 [1.07-1.13]* |
| Z76 | 5,500 | 80,573,310 | 0.07 | 0.07 | 0.06 | 0.00 | 0.01 | 0.01 | 1.15 [1.04-1.26]* | 1.14 [1.04-1.25]* |
| Z80 | 80,340 | 80,228,440 | 0.98 | 1.01 | 0.90 | −0.03 | 0.08 | 0.05 | 1.09 [1.06-1.12]* | 1.06 [1.03-1.08]* |
| Z81 | 13,500 | 81,348,130 | 0.19 | 0.16 | 0.17 | 0.02 | 0.01 | 0.01 | 1.07 [1.01-1.13] | 1.05 [0.99-1.11] |
| Z82 | 173,450 | 77,952,170 | 2.45 | 2.17 | 2.02 | 0.28 | 0.43 | 0.36 | 1.21 [1.19-1.23]* | 1.17 [1.15-1.19]* |
| Z83 | 66,650 | 79,761,760 | 0.93 | 0.81 | 0.80 | 0.12 | 0.14 | 0.11 | 1.17 [1.14-1.20]* | 1.13 [1.10-1.16]* |
| Z84 | 10,020 | 81,317,680 | 0.19 | 0.11 | 0.12 | 0.08 | 0.07 | 0.07 | 1.61 [1.52-1.71]* | 1.57 [1.47-1.67]* |
| Z85 | 298,230 | 77,867,340 | 3.87 | 3.82 | 3.53 | 0.05 | 0.34 | 0.32 | 1.10 [1.08-1.11]* | 1.09 [1.08-1.10]* |
| Z86 | 903,550 | 70,578,800 | 13.69 | 12.57 | 11.69 | 1.12 | 1.99 | 1.68 | 1.17 [1.16-1.18]* | 1.14 [1.13-1.15]* |
| Z87 | 511,880 | 74,515,470 | 7.47 | 6.72 | 6.29 | 0.76 | 1.18 | 0.89 | 1.19 [1.18-1.20]* | 1.13 [1.12-1.15]* |
| Z88 | 464,510 | 74,024,570 | 7.54 | 5.96 | 5.50 | 1.58 | 2.04 | 1.59 | 1.37 [1.36-1.38]* | 1.27 [1.25-1.28]* |
| Z89 | 14,930 | 81,465,650 | 0.19 | 0.18 | 0.16 | 0.01 | 0.03 | 0.03 | 1.18 [1.11-1.24]* | 1.17 [1.11-1.24]* |
| Z90 | 272,590 | 78,940,640 | 3.53 | 3.43 | 3.12 | 0.10 | 0.41 | 0.31 | 1.13 [1.12-1.15]* | 1.10 [1.08-1.11]* |
| Z91 | 212,370 | 78,905,790 | 3.80 | 2.42 | 2.35 | 1.38 | 1.45 | 1.20 | 1.61 [1.59-1.64]* | 1.46 [1.44-1.48]* |
| Z92 | 608,550 | 75,665,020 | 8.47 | 7.93 | 7.39 | 0.54 | 1.08 | 0.88 | 1.15 [1.14-1.16]* | 1.12 [1.11-1.13]* |
| Z93 | 61,670 | 80,925,830 | 0.77 | 0.76 | 0.69 | 0.01 | 0.08 | 0.08 | 1.12 [1.09-1.15]* | 1.11 [1.08-1.14]* |
| Z94 | 12,980 | 81,377,700 | 0.17 | 0.16 | 0.14 | 0.01 | 0.03 | 0.03 | 1.21 [1.14-1.28]* | 1.20 [1.13-1.27]* |
| Z95 | 206,260 | 78,942,430 | 2.69 | 2.59 | 2.40 | 0.09 | 0.29 | 0.25 | 1.12 [1.10-1.14]* | 1.10 [1.08-1.12]* |
| Z96 | 390,130 | 77,603,750 | 5.27 | 4.97 | 4.63 | 0.31 | 0.64 | 0.54 | 1.14 [1.13-1.15]* | 1.11 [1.10-1.13]* |
| Z97 | 39,920 | 81,226,280 | 0.53 | 0.48 | 0.45 | 0.05 | 0.08 | 0.06 | 1.18 [1.14-1.22]* | 1.14 [1.10-1.18]* |
| Z98 | 55,790 | 80,908,410 | 0.73 | 0.68 | 0.61 | 0.05 | 0.12 | 0.08 | 1.19 [1.16-1.23]* | 1.13 [1.10-1.16]* |
| Z99 | 69,870 | 81,074,510 | 0.96 | 0.84 | 0.77 | 0.12 | 0.19 | 0.15 | 1.25 [1.22-1.28]* | 1.19 [1.16-1.22]* |

[1] Hazard ratios (99% confidence intervals) estimated from Cox models comparing people with eczema to those without eczema. * indicates that the result is significant under Bonferroni-correction.

[2] Rate difference calculated using the estimated rate in the unexposed (rate in the exposed * (1/hazard ratio))
