## Supplementary Table 3 for "Mapping risks of hospital-recorded health conditions in people with eczema"

### Supplementary Table 3: Hazard ratios and events from all cohorts

| | Hazard ratio (99% confidence interval) | | | | | | | | | | | | | | |
|---|---|---|---|---|---|---|---|---|---|---|---|---|---|---|---|
| | crude | | | | | adjusted | | | | | Events (in exposed) | | | | |
| Outcome | any age | 18+ | 40+ | <18 | hosp. | any age | 18+ | 40+ | <18 | hosp. | any age | 18+ | 40+ | <18 | hosp. |
| Certain infectious and parasitic diseases (A00-B99) | | | | | | | | | | | | | | | |
| A02 | 1.23 [1.05-1.44] | 1.17 [0.97-1.42] | 1.27 [1.02-1.58] | 1.25 [0.97-1.61] | 1.11 [0.93-1.31] | 1.15 [0.98-1.36] | 1.04 [0.86-1.27] | 1.12 [0.90-1.41] | 1.26 [0.97-1.63] | 1.05 [0.89-1.25] | 395 | 259 | 203 | 160 | 324 |
| A04 | 1.27 [1.23-1.31] | 1.28 [1.24-1.32] | 1.28 [1.24-1.32] | 1.08 [0.97-1.21] | 1.19 [1.16-1.23] | 1.21 [1.17-1.24] | 1.18 [1.14-1.22] | 1.17 [1.14-1.21] | 1.03 [0.91-1.15] | 1.15 [1.12-1.19] | 11,231 | 10,718 | 9,774 | 814 | 9,899 |
| A05 | 1.26 [0.93-1.69] | 1.31 [0.96-1.80] | | | 1.19 [0.87-1.62] | 1.20 [0.89-1.63] | 1.23 [0.89-1.70] | | | 1.20 [0.87-1.64] | 118 | 102 | NA | NA | 104 |
| A07 | 1.21 [0.98-1.50] | 1.19 [0.91-1.56] | | 1.52 [1.10-2.10] | 1.17 [0.93-1.47] | 1.14 [0.92-1.42] | 1.04 [0.79-1.38] | | 1.44 [1.03-2.00] | 1.14 [0.90-1.44] | 216 | 132 | NA | 102 | 183 |
| A08 | 1.20 [1.17-1.23] | 1.31 [1.26-1.37] | 1.31 [1.25-1.38] | 1.14 [1.10-1.17] | 1.15 [1.12-1.18] | 1.15 [1.12-1.18] | 1.17 [1.12-1.22] | 1.17 [1.12-1.23] | 1.11 [1.08-1.15] | 1.12 [1.09-1.16] | 14,673 | 5,645 | 4,555 | 9,469 | 13,130 |
| A09 | 1.29 [1.28-1.31] | 1.32 [1.30-1.34] | 1.30 [1.27-1.32] | 1.19 [1.15-1.22] | 1.21 [1.20-1.23] | 1.20 [1.18-1.22] | 1.17 [1.15-1.19] | 1.15 [1.13-1.17] | 1.14 [1.10-1.17] | 1.16 [1.14-1.18] | 51,508 | 44,465 | 35,930 | 10,685 | 47,052 |
| A15 | 1.09 [0.90-1.32] | 1.13 [0.93-1.37] | 1.29 [1.03-1.61] | | 1.15 [0.94-1.42] | 1.06 [0.87-1.29] | 1.08 [0.88-1.32] | 1.17 [0.93-1.48] | | 1.12 [0.90-1.38] | 261 | 240 | 186 | NA | 217 |
| A16 | 1.33 [1.17-1.51] | 1.41 [1.23-1.61] | 1.43 [1.23-1.67] | | 1.30 [1.13-1.50] | 1.28 [1.12-1.46] | 1.30 [1.14-1.50] | 1.30 [1.11-1.52] | | 1.24 [1.07-1.44] | 610 | 554 | 429 | NA | 497 |
| A18 | 1.19 [0.99-1.42] | 1.35 [1.11-1.63] | 1.35 [1.08-1.68] | | 1.22 [1.00-1.50] | 1.14 [0.95-1.38] | 1.31 [1.08-1.60] | 1.28 [1.02-1.61] | | 1.18 [0.96-1.45] | 286 | 263 | 193 | NA | 228 |
| A31 | 1.41 [1.16-1.71] | 1.63 [1.32-2.01] | 1.69 [1.36-2.12] | | 1.35 [1.11-1.65] | 1.26 [1.03-1.54] | 1.39 [1.12-1.73] | 1.43 [1.13-1.80] | | 1.21 [0.99-1.48] | 266 | 235 | 208 | NA | 251 |
| A38 | 1.08 [0.93-1.25] | | | 1.05 [0.91-1.22] | 1.03 [0.89-1.20] | 1.06 [0.91-1.22] | | | 1.03 [0.89-1.20] | 1.02 [0.87-1.19] | 469 | NA | NA | 456 | 430 |
| A39 | 1.06 [0.92-1.21] | 1.10 [0.82-1.46] | | 1.05 [0.91-1.22] | 1.05 [0.90-1.22] | 1.03 [0.90-1.18] | 1.06 [0.79-1.42] | | 1.04 [0.89-1.21] | 1.03 [0.88-1.21] | 511 | 116 | NA | 421 | 415 |
| A40 | 1.33 [1.25-1.43] | 1.35 [1.26-1.45] | 1.39 [1.29-1.49] | 1.11 [0.90-1.35] | 1.26 [1.18-1.35] | 1.28 [1.20-1.37] | 1.27 [1.18-1.36] | 1.29 [1.20-1.39] | 1.05 [0.86-1.30] | 1.24 [1.16-1.33] | 2,387 | 2,187 | 2,041 | 247 | 2,169 |
| A41 | 1.25 [1.23-1.27] | 1.26 [1.24-1.28] | 1.25 [1.23-1.27] | 1.10 [1.03-1.18] | 1.19 [1.18-1.21] | 1.21 [1.19-1.23] | 1.17 [1.15-1.19] | 1.17 [1.15-1.19] | 1.08 [1.01-1.16] | 1.18 [1.16-1.20] | 42,650 | 41,175 | 38,875 | 2,275 | 39,444 |
| A46 | 2.16 [1.72-2.72] | 2.10 [1.67-2.65] | 2.09 [1.64-2.66] | | 1.97 [1.53-2.53] | 2.11 [1.67-2.66] | 1.95 [1.54-2.48] | 1.97 [1.54-2.53] | | 1.93 [1.50-2.49] | 211 | 204 | 182 | NA | 171 |
| A48 | 1.21 [0.98-1.49] | 1.26 [1.00-1.59] | 1.21 [0.95-1.55] | | 1.23 [0.98-1.54] | 1.22 [0.99-1.51] | 1.20 [0.95-1.52] | 1.16 [0.91-1.49] | | 1.27 [1.01-1.60] | 217 | 179 | 164 | NA | 190 |
| A49 | 1.34 [1.29-1.40] | 1.36 [1.31-1.42] | 1.34 [1.28-1.40] | 1.19 [1.06-1.34] | 1.27 [1.22-1.32] | 1.28 [1.23-1.34] | 1.25 [1.20-1.31] | 1.23 [1.18-1.29] | 1.17 [1.04-1.32] | 1.24 [1.19-1.29] | 6,459 | 5,849 | 5,422 | 740 | 5,854 |
| A63 | 1.49 [1.21-1.83] | 1.47 [1.19-1.81] | | | 1.45 [1.12-1.86] | 1.35 [1.09-1.67] | 1.31 [1.06-1.63] | | | 1.38 [1.07-1.80] | 216 | 205 | NA | NA | 141 |
| A69 | 1.25 [0.99-1.59] | 1.23 [0.94-1.61] | | | 1.14 [0.89-1.47] | 1.15 [0.90-1.47] | 1.06 [0.80-1.41] | | | 1.07 [0.82-1.38] | 169 | 129 | NA | NA | 146 |
| A86 | 1.01 [0.84-1.21] | 1.09 [0.89-1.33] | 1.18 [0.95-1.47] | | 1.03 [0.84-1.25] | 0.98 [0.81-1.17] | 1.01 [0.82-1.23] | 1.08 [0.86-1.35] | | 0.99 [0.81-1.21] | 291 | 238 | 199 | NA | 243 |
| A87 | 1.31 [1.17-1.45] | 1.40 [1.25-1.58] | 1.38 [1.12-1.71] | 1.27 [1.06-1.51] | 1.16 [1.03-1.30] | 1.20 [1.08-1.34] | 1.22 [1.08-1.38] | 1.19 [0.96-1.49] | 1.24 [1.03-1.49] | 1.09 [0.96-1.22] | 873 | 732 | 200 | 342 | 707 |
| B00 | 2.52 [2.38-2.67] | 2.06 [1.91-2.24] | 1.73 [1.58-1.91] | 3.13 [2.90-3.38] | 2.31 [2.17-2.46] | 2.34 [2.21-2.49] | 1.80 [1.66-1.96] | 1.54 [1.40-1.70] | 2.97 [2.74-3.22] | 2.18 [2.05-2.33] | 3,846 | 1,786 | 1,174 | 2,331 | 3,252 |
| B01 | 1.61 [1.52-1.72] | 1.39 [1.14-1.68] | 1.43 [1.10-1.87] | 1.63 [1.53-1.74] | 1.53 [1.44-1.64] | 1.56 [1.46-1.66] | 1.27 [1.04-1.55] | 1.29 [0.99-1.70] | 1.58 [1.48-1.69] | 1.49 [1.40-1.60] | 2,710 | 260 | 139 | 2,482 | 2,395 |
| B02 | 1.42 [1.34-1.50] | 1.41 [1.33-1.49] | 1.40 [1.32-1.49] | 1.41 [1.20-1.67] | 1.32 [1.24-1.40] | 1.34 [1.26-1.42] | 1.29 [1.21-1.37] | 1.29 [1.21-1.37] | 1.35 [1.13-1.60] | 1.26 [1.19-1.33] | 3,475 | 3,158 | 2,946 | 394 | 3,139 |
| B07 | 1.41 [1.30-1.53] | 1.40 [1.28-1.53] | 1.39 [1.26-1.54] | 1.45 [1.22-1.71] | 1.32 [1.21-1.45] | 1.30 [1.20-1.42] | 1.25 [1.14-1.37] | 1.28 [1.15-1.42] | 1.32 [1.11-1.58] | 1.25 [1.14-1.37] | 1,541 | 1,240 | 948 | 379 | 1,278 |
| B08 | 1.60 [1.44-1.77] | 1.95 [1.48-2.56] | | 1.53 [1.37-1.71] | 1.54 [1.38-1.72] | 1.52 [1.37-1.69] | 1.70 [1.27-2.27] | | 1.48 [1.32-1.66] | 1.49 [1.33-1.67] | 951 | 143 | NA | 826 | 862 |
| B09 | 1.19 [1.01-1.41] | | | 1.16 [0.97-1.38] | 1.17 [0.98-1.40] | 1.18 [0.99-1.40] | | | 1.14 [0.95-1.36] | 1.16 [0.97-1.39] | 351 | NA | NA | 326 | 315 |
| B15 | 1.10 [0.86-1.39] | 1.24 [0.95-1.60] | 1.35 [1.00-1.82] | | 1.06 [0.82-1.39] | 1.06 [0.83-1.36] | 1.17 [0.89-1.53] | 1.26 [0.93-1.72] | | 1.03 [0.79-1.35] | 171 | 142 | 108 | NA | 139 |
| B16 | 1.34 [1.12-1.59] | 1.39 [1.17-1.66] | 1.54 [1.25-1.90] | | 1.17 [0.97-1.41] | 1.31 [1.09-1.56] | 1.37 [1.14-1.64] | 1.45 [1.17-1.81] | | 1.17 [0.97-1.42] | 314 | 309 | 217 | NA | 256 |
| B17 | 1.39 [1.21-1.60] | 1.41 [1.22-1.62] | 1.52 [1.29-1.80] | | 1.24 [1.07-1.44] | 1.33 [1.15-1.53] | 1.26 [1.08-1.46] | 1.35 [1.13-1.60] | | 1.20 [1.03-1.40] | 499 | 463 | 353 | NA | 421 |
| B18 | 1.19 [1.11-1.28] | 1.23 [1.14-1.32] | 1.30 [1.20-1.41] | | 1.07 [0.99-1.16] | 1.16 [1.08-1.25] | 1.13 [1.05-1.22] | 1.18 [1.08-1.29] | | 1.09 [1.01-1.18] | 1,820 | 1,807 | 1,396 | NA | 1,544 |
| B19 | 1.16 [0.87-1.54] | | | | 1.18 [0.87-1.60] | 1.07 [0.80-1.43] | | | | 1.14 [0.84-1.57] | 123 | NA | NA | NA | 102 |
| B25 | 1.41 [1.21-1.64] | 1.40 [1.19-1.65] | 1.45 [1.19-1.75] | | 1.23 [1.05-1.44] | 1.35 [1.15-1.58] | 1.27 [1.07-1.50] | 1.32 [1.09-1.61] | | 1.18 [1.01-1.40] | 413 | 357 | 264 | NA | 367 |
| B26 | 1.20 [0.92-1.56] | | | | 1.13 [0.84-1.52] | 1.21 [0.92-1.58] | | | | 1.08 [0.79-1.46] | 144 | NA | NA | NA | 109 |
| B27 | 1.15 [1.06-1.24] | 1.11 [1.00-1.24] | | 1.16 [1.06-1.26] | 1.08 [0.98-1.19] | 1.10 [1.01-1.19] | 1.02 [0.91-1.14] | | 1.10 [1.01-1.21] | 1.04 [0.94-1.15] | 1,551 | 787 | NA | 1,319 | 1,099 |
| B30 | 1.26 [0.98-1.62] | | | | 1.16 [0.90-1.51] | 1.16 [0.89-1.50] | | | | 1.09 [0.83-1.43] | 156 | NA | NA | NA | 144 |
| B33 | 1.24 [0.93-1.66] | | | | | 1.22 [0.91-1.64] | | | | | 109 | NA | NA | NA | NA |
| B34 | 1.54 [1.52-1.57] | 1.33 [1.28-1.38] | 1.33 [1.27-1.40] | 1.57 [1.55-1.60] | 1.49 [1.46-1.51] | 1.48 [1.45-1.50] | 1.15 [1.11-1.20] | 1.16 [1.10-1.22] | 1.53 [1.50-1.55] | 1.44 [1.41-1.46] | 40,130 | 6,707 | 3,879 | 34,560 | 35,691 |
| B35 | 1.72 [1.58-1.86] | 1.77 [1.62-1.93] | 1.71 [1.57-1.87] | 1.47 [1.19-1.82] | 1.65 [1.51-1.79] | 1.58 [1.45-1.71] | 1.56 [1.43-1.71] | 1.52 [1.39-1.67] | 1.38 [1.10-1.72] | 1.55 [1.42-1.69] | 1,735 | 1,527 | 1,376 | 248 | 1,546 |
| B36 | 1.87 [1.63-2.14] | 1.86 [1.62-2.14] | 1.77 [1.53-2.05] | | 1.68 [1.46-1.92] | 1.72 [1.50-1.97] | 1.66 [1.44-1.92] | 1.58 [1.36-1.84] | | 1.60 [1.39-1.84] | 651 | 592 | 538 | NA | 596 |
| B37 | 1.36 [1.33-1.39] | 1.37 [1.34-1.40] | 1.38 [1.34-1.41] | 1.11 [1.04-1.20] | 1.26 [1.23-1.29] | 1.25 [1.22-1.28] | 1.20 [1.17-1.23] | 1.21 [1.18-1.24] | 1.06 [0.99-1.14] | 1.19 [1.16-1.22] | 18,911 | 17,959 | 15,080 | 1,965 | 17,282 |
| B44 | 1.69 [1.51-1.88] | 1.84 [1.64-2.06] | 1.90 [1.68-2.14] | | 1.51 [1.35-1.70] | 1.40 [1.24-1.57] | 1.47 [1.30-1.66] | 1.50 [1.32-1.70] | | 1.27 [1.13-1.43] | 857 | 811 | 745 | NA | 784 |
| B49 | 1.63 [1.41-1.88] | 1.53 [1.30-1.78] | 1.51 [1.29-1.78] | | 1.51 [1.30-1.76] | 1.54 [1.33-1.79] | 1.40 [1.19-1.65] | 1.39 [1.18-1.64] | | 1.45 [1.25-1.69] | 500 | 418 | 376 | NA | 461 |
| B50 | 0.95 [0.76-1.19] | 0.98 [0.76-1.26] | | | 0.83 [0.63-1.10] | 0.97 [0.77-1.22] | 1.05 [0.81-1.37] | | | 0.86 [0.65-1.14] | 194 | 143 | NA | NA | 116 |
| B59 | 1.53 [1.24-1.88] | 1.55 [1.26-1.91] | 1.39 [1.12-1.73] | | 1.36 [1.09-1.68] | 1.44 [1.16-1.78] | 1.44 [1.16-1.78] | 1.30 [1.04-1.62] | | 1.30 [1.04-1.62] | 236 | 228 | 205 | NA | 210 |
| B80 | 1.21 [1.06-1.37] | 1.24 [1.03-1.50] | 1.21 [0.95-1.54] | 1.18 [1.00-1.38] | 1.16 [1.01-1.33] | 1.14 [1.00-1.30] | 1.13 [0.93-1.37] | 1.14 [0.88-1.47] | 1.12 [0.95-1.32] | 1.11 [0.96-1.27] | 614 | 261 | 148 | 405 | 532 |
| B85 | 1.12 [0.91-1.37] | 1.19 [0.89-1.59] | | 0.95 [0.71-1.26] | 1.04 [0.83-1.30] | 1.11 [0.90-1.36] | 1.09 [0.81-1.47] | | 0.95 [0.71-1.26] | 1.03 [0.82-1.29] | 228 | 117 | NA | 115 | 189 |
| B86 | 2.53 [2.19-2.93] | 2.59 [2.22-3.04] | 2.65 [2.26-3.12] | | 2.19 [1.87-2.55] | 2.49 [2.15-2.89] | 2.43 [2.07-2.86] | 2.49 [2.11-2.94] | | 2.20 [1.88-2.58] | 607 | 519 | 496 | NA | 516 |
| B90 | 1.60 [1.25-2.03] | 1.43 [1.12-1.82] | 1.39 [1.08-1.79] | | 1.41 [1.11-1.81] | 1.46 [1.14-1.87] | 1.29 [1.00-1.65] | 1.25 [0.97-1.62] | | 1.31 [1.02-1.68] | 184 | 182 | 167 | NA | 171 |
| B91 | 1.30 [1.07-1.58] | 1.29 [1.06-1.57] | 1.33 [1.09-1.62] | | 1.14 [0.92-1.41] | 1.30 [1.06-1.58] | 1.27 [1.04-1.56] | 1.29 [1.06-1.59] | | 1.16 [0.94-1.44] | 257 | 256 | 250 | NA | 210 |
| B94 | 1.32 [1.14-1.52] | 1.31 [1.11-1.55] | 1.33 [1.11-1.60] | 1.23 [0.93-1.61] | 1.24 [1.07-1.44] | 1.19 [1.02-1.39] | 1.14 [0.96-1.36] | 1.17 [0.97-1.41] | 1.25 [0.94-1.65] | 1.18 [1.01-1.37] | 468 | 348 | 296 | 137 | 438 |
| B95 | 1.51 [1.48-1.54] | 1.50 [1.47-1.54] | 1.50 [1.46-1.53] | 1.50 [1.43-1.57] | 1.41 [1.38-1.44] | 1.44 [1.42-1.47] | 1.39 [1.36-1.42] | 1.38 [1.35-1.41] | 1.45 [1.39-1.52] | 1.38 [1.35-1.41] | 29,775 | 25,667 | 21,484 | 5,462 | 26,256 |
| B96 | 1.23 [1.21-1.24] | 1.24 [1.22-1.26] | 1.24 [1.22-1.26] | 1.07 [1.02-1.12] | 1.17 [1.15-1.18] | 1.17 [1.16-1.19] | 1.14 [1.13-1.16] | 1.15 [1.13-1.17] | 1.04 [1.00-1.09] | 1.14 [1.12-1.16] | 51,288 | 47,700 | 43,442 | 5,099 | 46,091 |
| B97 | 1.32 [1.29-1.35] | 1.29 [1.25-1.34] | 1.30 [1.25-1.35] | 1.34 [1.29-1.39] | 1.25 [1.22-1.29] | 1.23 [1.20-1.27] | 1.15 [1.11-1.19] | 1.14 [1.10-1.19] | 1.28 [1.23-1.34] | 1.19 [1.16-1.23] | 15,607 | 9,623 | 7,862 | 6,512 | 14,153 |
| B98 | 1.20 [1.14-1.25] | 1.22 [1.16-1.27] | 1.21 [1.15-1.27] | 1.01 [0.82-1.25] | 1.13 [1.08-1.18] | 1.14 [1.09-1.19] | 1.13 [1.08-1.18] | 1.12 [1.07-1.18] | 1.00 [0.80-1.24] | 1.10 [1.04-1.15] | 4,905 | 4,763 | 4,267 | 248 | 4,320 |
| B99 | 1.24 [1.14-1.36] | 1.26 [1.16-1.38] | 1.21 [1.10-1.33] | 1.25 [0.95-1.63] | 1.20 [1.10-1.32] | 1.20 [1.10-1.31] | 1.17 [1.07-1.28] | 1.12 [1.02-1.24] | 1.18 [0.89-1.56] | 1.19 [1.09-1.31] | 1,443 | 1,335 | 1,229 | 144 | 1,356 |
| Neoplasms (C00-D49) | | | | | | | | | | | | | | | |
| C00 | 1.04 [0.81-1.35] | 1.08 [0.84-1.40] | 1.10 [0.85-1.42] | | 1.04 [0.80-1.36] | 1.03 [0.80-1.33] | 1.05 [0.81-1.36] | 1.05 [0.81-1.37] | | 1.03 [0.79-1.35] | 146 | 146 | 144 | NA | 128 |
| C01 | 1.08 [0.91-1.28] | 1.09 [0.92-1.30] | 1.07 [0.90-1.27] | | 1.12 [0.93-1.35] | 1.08 [0.90-1.28] | 1.05 [0.88-1.26] | 1.04 [0.87-1.24] | | 1.17 [0.97-1.41] | 306 | 306 | 303 | NA | 266 |
| C02 | 1.28 [1.13-1.46] | 1.31 [1.15-1.50] | 1.32 [1.16-1.51] | | 1.30 [1.13-1.50] | 1.27 [1.11-1.45] | 1.24 [1.08-1.42] | 1.27 [1.11-1.46] | | 1.31 [1.13-1.51] | 548 | 548 | 530 | NA | 473 |
| C03 | 1.47 [1.12-1.92] | 1.38 [1.06-1.81] | 1.44 [1.09-1.91] | | 1.32 [0.99-1.76] | 1.46 [1.11-1.91] | 1.38 [1.05-1.82] | 1.42 [1.07-1.88] | | 1.32 [0.98-1.77] | 139 | 139 | 132 | NA | 118 |
| C04 | 1.13 [0.88-1.45] | 1.15 [0.90-1.48] | 1.12 [0.87-1.44] | | 1.10 [0.84-1.43] | 1.14 [0.89-1.47] | 1.08 [0.84-1.39] | 1.06 [0.82-1.37] | | 1.13 [0.86-1.48] | 151 | 150 | 145 | NA | 128 |
| C05 | 1.28 [1.01-1.62] | 1.32 [1.05-1.67] | 1.32 [1.04-1.68] | | 1.14 [0.89-1.47] | 1.29 [1.02-1.64] | 1.29 [1.01-1.63] | 1.27 [0.99-1.62] | | 1.20 [0.93-1.56] | 176 | 177 | 168 | NA | 147 |
| C06 | 1.25 [1.03-1.52] | 1.24 [1.02-1.52] | 1.21 [0.99-1.48] | | 1.18 [0.96-1.46] | 1.24 [1.01-1.51] | 1.18 [0.96-1.44] | 1.13 [0.92-1.39] | | 1.19 [0.96-1.47] | 244 | 246 | 233 | NA | 214 |
| C07 | 1.03 [0.84-1.27] | 1.05 [0.85-1.29] | 1.01 [0.81-1.25] | | 1.04 [0.83-1.31] | 1.02 [0.83-1.25] | 1.00 [0.80-1.24] | 0.97 [0.78-1.21] | | 1.03 [0.82-1.30] | 219 | 212 | 192 | NA | 184 |
| C09 | 1.07 [0.92-1.25] | 1.12 [0.97-1.31] | 1.15 [0.99-1.33] | | 1.07 [0.90-1.26] | 1.04 [0.89-1.22] | 1.06 [0.91-1.24] | 1.09 [0.93-1.27] | | 1.08 [0.91-1.28] | 391 | 391 | 391 | NA | 316 |
| C10 | 1.11 [0.90-1.37] | 1.18 [0.95-1.45] | 1.16 [0.94-1.44] | | 1.08 [0.86-1.36] | 1.09 [0.88-1.35] | 1.10 [0.88-1.36] | 1.09 [0.87-1.35] | | 1.09 [0.87-1.38] | 204 | 204 | 203 | NA | 176 |
| C11 | 1.19 [0.90-1.58] | 1.20 [0.90-1.60] | 1.36 [1.01-1.83] | | | 1.19 [0.89-1.58] | 1.17 [0.87-1.56] | 1.31 [0.97-1.77] | | | 115 | 111 | 108 | NA | NA |
| C12 | 1.24 [0.94-1.63] | 1.29 [0.98-1.70] | 1.35 [1.02-1.78] | | 1.26 [0.93-1.70] | 1.25 [0.94-1.65] | 1.24 [0.93-1.65] | 1.31 [0.99-1.74] | | 1.33 [0.98-1.81] | 122 | 122 | 122 | NA | 103 |
| C13 | 1.16 [0.89-1.52] | 1.25 [0.95-1.63] | 1.22 [0.93-1.60] | | 1.19 [0.89-1.58] | 1.20 [0.91-1.57] | 1.24 [0.94-1.63] | 1.20 [0.91-1.58] | | 1.30 [0.97-1.75] | 132 | 132 | 132 | NA | 116 |
| C14 | 1.10 [0.90-1.34] | 1.12 [0.91-1.36] | 1.06 [0.87-1.30] | | 0.96 [0.77-1.18] | 1.08 [0.88-1.33] | 1.06 [0.87-1.31] | 1.00 [0.81-1.23] | | 0.97 [0.78-1.20] | 228 | 228 | 227 | NA | 196 |
| C15 | 1.18 [1.12-1.25] | 1.15 [1.09-1.22] | 1.17 [1.10-1.23] | | 1.13 [1.07-1.20] | 1.17 [1.11-1.24] | 1.12 [1.06-1.18] | 1.13 [1.07-1.19] | | 1.14 [1.07-1.21] | 3,259 | 3,259 | 3,242 | NA | 2,722 |
| C16 | 1.08 [1.01-1.15] | 1.07 [1.00-1.13] | 1.09 [1.02-1.16] | | 1.06 [0.99-1.14] | 1.07 [1.01-1.14] | 1.04 [0.98-1.11] | 1.06 [0.99-1.13] | | 1.06 [0.99-1.14] | 2,508 | 2,508 | 2,476 | NA | 2,102 |
| C17 | 1.11 [0.98-1.27] | 1.15 [1.01-1.32] | 1.13 [0.99-1.29] | | 1.08 [0.94-1.24] | 1.10 [0.96-1.26] | 1.09 [0.95-1.25] | 1.10 [0.96-1.26] | | 1.07 [0.93-1.23] | 528 | 527 | 520 | NA | 468 |
| C18 | 1.01 [0.98-1.05] | 1.01 [0.98-1.05] | 1.01 [0.98-1.05] | | 1.00 [0.96-1.04] | 1.00 [0.97-1.04] | 1.00 [0.97-1.03] | 1.00 [0.96-1.03] | | 1.00 [0.96-1.03] | 8,255 | 8,246 | 8,116 | NA | 6,819 |
| C19 | 0.95 [0.88-1.02] | 0.96 [0.89-1.04] | 0.96 [0.89-1.04] | | 0.94 [0.87-1.02] | 0.95 [0.88-1.02] | 0.96 [0.89-1.04] | 0.96 [0.89-1.04] | | 0.95 [0.88-1.04] | 1,621 | 1,619 | 1,583 | NA | 1,334 |
| C20 | 0.97 [0.92-1.02] | 0.97 [0.92-1.02] | 0.97 [0.92-1.02] | | 0.95 [0.90-1.01] | 0.98 [0.93-1.03] | 0.98 [0.93-1.03] | 0.97 [0.92-1.03] | | 0.97 [0.91-1.03] | 3,365 | 3,365 | 3,318 | NA | 2,627 |
| C21 | 1.08 [0.94-1.24] | 1.08 [0.94-1.25] | 1.09 [0.94-1.25] | | 1.04 [0.89-1.21] | 1.07 [0.93-1.23] | 1.05 [0.91-1.21] | 1.05 [0.91-1.21] | | 1.04 [0.89-1.21] | 458 | 458 | 451 | NA | 390 |
| C22 | 1.25 [1.16-1.34] | 1.25 [1.16-1.34] | 1.23 [1.14-1.32] | | 1.18 [1.09-1.27] | 1.21 [1.13-1.31] | 1.18 [1.10-1.27] | 1.17 [1.08-1.26] | | 1.17 [1.08-1.26] | 1,925 | 1,919 | 1,894 | NA | 1,708 |
| C23 | 1.07 [0.88-1.31] | 1.06 [0.87-1.29] | 1.06 [0.87-1.29] | | 1.03 [0.83-1.28] | 1.07 [0.88-1.30] | 1.04 [0.85-1.26] | 1.05 [0.86-1.28] | | 1.04 [0.84-1.29] | 242 | 242 | 242 | NA | 204 |
| C24 | 1.09 [0.94-1.25] | 1.07 [0.93-1.24] | 1.07 [0.92-1.23] | | 1.04 [0.89-1.22] | 1.08 [0.94-1.25] | 1.04 [0.90-1.20] | 1.02 [0.88-1.18] | | 1.03 [0.88-1.20] | 464 | 465 | 458 | NA | 389 |
| C25 | 1.16 [1.10-1.23] | 1.14 [1.08-1.21] | 1.15 [1.09-1.22] | | 1.13 [1.07-1.20] | 1.15 [1.09-1.22] | 1.10 [1.04-1.17] | 1.11 [1.05-1.18] | | 1.13 [1.07-1.21] | 3,016 | 3,008 | 2,998 | NA | 2,573 |
| C26 | 1.13 [1.00-1.28] | 1.10 [0.97-1.24] | 1.12 [0.99-1.26] | | 1.06 [0.93-1.20] | 1.10 [0.98-1.25] | 1.05 [0.93-1.19] | 1.08 [0.95-1.22] | | 1.05 [0.92-1.19] | 678 | 677 | 659 | NA | 626 |
| C30 | 1.07 [0.80-1.43] | 1.12 [0.84-1.51] | 1.16 [0.86-1.56] | | | 1.06 [0.79-1.42] | 1.09 [0.81-1.46] | 1.12 [0.83-1.52] | | | 109 | 107 | 107 | NA | NA |
| C32 | 1.14 [1.02-1.28] | 1.17 [1.04-1.32] | 1.16 [1.03-1.30] | | 1.08 [0.95-1.22] | 1.16 [1.03-1.30] | 1.14 [1.01-1.28] | 1.12 [0.99-1.26] | | 1.11 [0.98-1.26] | 690 | 689 | 680 | NA | 586 |
| C34 | 1.15 [1.12-1.18] | 1.15 [1.12-1.18] | 1.15 [1.12-1.18] | | 1.11 [1.08-1.14] | 1.14 [1.11-1.17] | 1.09 [1.06-1.12] | 1.09 [1.06-1.12] | | 1.12 [1.09-1.15] | 13,074 | 13,057 | 13,024 | NA | 11,181 |
| C38 | 1.07 [0.85-1.35] | 0.97 [0.76-1.22] | 1.00 [0.79-1.27] | | 1.02 [0.80-1.32] | 1.08 [0.85-1.37] | 0.95 [0.74-1.20] | 0.99 [0.77-1.26] | | 1.04 [0.80-1.34] | 171 | 165 | 157 | NA | 144 |
| C40 | 1.05 [0.80-1.37] | | | | 1.02 [0.75-1.37] | 1.02 [0.78-1.34] | | | | 1.04 [0.77-1.41] | 134 | NA | NA | NA | 104 |
| C41 | 1.25 [1.07-1.46] | 1.23 [1.04-1.45] | 1.21 [1.02-1.44] | | 1.18 [1.00-1.40] | 1.24 [1.06-1.46] | 1.19 [1.00-1.40] | 1.17 [0.98-1.39] | | 1.19 [1.00-1.41] | 397 | 353 | 316 | NA | 332 |
| C43 | 0.95 [0.90-1.01] | 0.97 [0.92-1.02] | 0.96 [0.90-1.01] | | 0.97 [0.91-1.02] | 0.94 [0.89-1.00] | 0.95 [0.90-1.01] | 0.94 [0.89-0.99] | | 0.96 [0.90-1.02] | 3,170 | 3,159 | 2,886 | NA | 2,682 |
| C44 | 1.10 [1.08-1.12] | 1.11 [1.08-1.13] | 1.10 [1.08-1.13] | | 1.06 [1.03-1.08] | 1.07 [1.05-1.09] | 1.07 [1.04-1.09] | 1.06 [1.04-1.08] | | 1.03 [1.01-1.06] | 25,094 | 25,077 | 24,709 | NA | 21,264 |
| C45 | 0.98 [0.88-1.10] | 1.00 [0.89-1.12] | 1.01 [0.90-1.13] | | 1.02 [0.91-1.16] | 0.99 [0.89-1.11] | 1.01 [0.90-1.13] | 1.02 [0.91-1.14] | | 1.04 [0.92-1.18] | 725 | 723 | 725 | NA | 614 |
| C48 | 1.05 [0.90-1.21] | 1.07 [0.93-1.24] | 1.06 [0.91-1.23] | | 1.04 [0.89-1.22] | 1.03 [0.89-1.20] | 1.07 [0.92-1.24] | 1.04 [0.90-1.21] | | 1.03 [0.88-1.21] | 420 | 413 | 409 | NA | 351 |
| C49 | 1.08 [0.97-1.20] | 1.13 [1.01-1.26] | 1.13 [1.00-1.27] | | 1.10 [0.98-1.24] | 1.06 [0.95-1.18] | 1.10 [0.99-1.24] | 1.09 [0.96-1.23] | | 1.09 [0.97-1.23] | 820 | 758 | 682 | NA | 697 |
| C50 | 1.04 [1.02-1.07] | 1.04 [1.01-1.06] | 1.03 [1.01-1.06] | | 1.02 [0.99-1.05] | 1.04 [1.01-1.07] | 1.03 [1.00-1.05] | 1.02 [0.99-1.05] | | 1.03 [1.00-1.06] | 14,359 | 14,360 | 13,755 | NA | 11,473 |
| C51 | 1.20 [1.04-1.39] | 1.24 [1.07-1.44] | 1.27 [1.09-1.47] | | 1.21 [1.03-1.43] | 1.18 [1.02-1.38] | 1.22 [1.05-1.42] | 1.22 [1.05-1.42] | | 1.20 [1.02-1.42] | 434 | 433 | 424 | NA | 361 |
| C52 | 0.98 [0.74-1.29] | 0.90 [0.68-1.19] | 0.92 [0.70-1.22] | | 0.93 [0.69-1.24] | 1.00 [0.76-1.33] | 0.90 [0.68-1.19] | 0.93 [0.70-1.23] | | 0.93 [0.69-1.25] | 117 | 116 | 114 | NA | 103 |
| C53 | 0.96 [0.86-1.07] | 0.96 [0.86-1.07] | 0.96 [0.84-1.09] | | 1.02 [0.90-1.15] | 0.97 [0.87-1.09] | 0.97 [0.87-1.08] | 0.98 [0.86-1.12] | | 1.05 [0.93-1.19] | 755 | 752 | 523 | NA | 615 |
| C54 | 1.04 [0.98-1.11] | 1.04 [0.98-1.11] | 1.03 [0.97-1.09] | | 1.03 [0.96-1.10] | 1.06 [0.99-1.12] | 1.06 [1.00-1.13] | 1.05 [0.98-1.11] | | 1.04 [0.97-1.12] | 2,358 | 2,355 | 2,327 | NA | 1,822 |
| C55 | 1.06 [0.90-1.25] | 1.02 [0.86-1.20] | 1.01 [0.85-1.19] | | 0.96 [0.80-1.15] | 1.05 [0.88-1.24] | 1.01 [0.85-1.19] | 1.01 [0.85-1.20] | | 0.95 [0.79-1.15] | 325 | 323 | 315 | NA | 263 |
| C56 | 1.03 [0.97-1.10] | 1.01 [0.95-1.08] | 1.02 [0.96-1.09] | | 1.00 [0.93-1.08] | 1.04 [0.97-1.11] | 1.02 [0.95-1.09] | 1.02 [0.96-1.10] | | 1.01 [0.94-1.09] | 2,094 | 2,078 | 1,979 | NA | 1,630 |
| C57 | 1.18 [0.97-1.42] | 1.19 [0.99-1.44] | 1.19 [0.99-1.44] | | 1.15 [0.94-1.42] | 1.20 [0.99-1.46] | 1.22 [1.00-1.47] | 1.21 [1.00-1.47] | | 1.17 [0.95-1.44] | 262 | 262 | 260 | NA | 215 |
| C60 | 1.19 [0.95-1.48] | 1.30 [1.04-1.62] | 1.21 [0.97-1.51] | | 1.17 [0.92-1.49] | 1.20 [0.96-1.50] | 1.29 [1.03-1.62] | 1.24 [0.99-1.56] | | 1.21 [0.94-1.55] | 195 | 195 | 190 | NA | 162 |
| C61 | 1.06 [1.03-1.09] | 1.06 [1.03-1.09] | 1.05 [1.03-1.08] | | 1.01 [0.98-1.04] | 1.04 [1.01-1.06] | 1.03 [1.00-1.06] | 1.02 [1.00-1.05] | | 1.00 [0.97-1.03] | 13,721 | 13,725 | 13,723 | NA | 11,486 |
| C62 | 1.04 [0.89-1.22] | 1.02 [0.87-1.20] | 0.99 [0.78-1.26] | | 1.02 [0.84-1.24] | 1.02 [0.87-1.20] | 0.99 [0.84-1.17] | 0.95 [0.74-1.22] | | 1.03 [0.84-1.25] | 362 | 345 | 146 | NA | 242 |
| C64 | 1.11 [1.05-1.18] | 1.10 [1.04-1.17] | 1.11 [1.05-1.18] | | 1.06 [0.99-1.12] | 1.09 [1.03-1.15] | 1.05 [0.99-1.11] | 1.06 [1.00-1.12] | | 1.05 [0.99-1.12] | 2,914 | 2,867 | 2,824 | NA | 2,501 |
| C65 | 1.22 [1.01-1.47] | 1.22 [1.01-1.47] | 1.22 [1.01-1.48] | | 1.11 [0.91-1.36] | 1.24 [1.02-1.50] | 1.18 [0.98-1.43] | 1.21 [1.00-1.47] | | 1.15 [0.94-1.41] | 282 | 280 | 277 | NA | 241 |
| C66 | 1.10 [0.93-1.29] | 1.09 [0.92-1.28] | 1.08 [0.92-1.28] | | 0.99 [0.83-1.17] | 1.09 [0.93-1.29] | 1.05 [0.89-1.24] | 1.04 [0.88-1.23] | | 1.00 [0.84-1.18] | 357 | 357 | 357 | NA | 316 |
| C67 | 1.08 [1.04-1.12] | 1.09 [1.05-1.13] | 1.08 [1.04-1.13] | | 1.03 [0.99-1.07] | 1.07 [1.03-1.12] | 1.06 [1.02-1.10] | 1.06 [1.02-1.10] | | 1.03 [0.99-1.08] | 6,425 | 6,415 | 6,366 | NA | 5,376 |
| C68 | 1.03 [0.79-1.35] | 1.05 [0.81-1.37] | 1.11 [0.85-1.44] | | 0.94 [0.71-1.23] | 1.02 [0.78-1.33] | 1.04 [0.80-1.36] | 1.10 [0.84-1.43] | | 0.95 [0.72-1.26] | 134 | 135 | 132 | NA | 118 |
| C69 | 0.98 [0.81-1.18] | 0.97 [0.80-1.17] | 1.03 [0.84-1.26] | | 0.96 [0.79-1.18] | 0.96 [0.79-1.16] | 0.95 [0.78-1.16] | 1.00 [0.81-1.23] | | 0.97 [0.79-1.19] | 261 | 238 | 222 | NA | 221 |
| C71 | 1.12 [1.04-1.21] | 1.15 [1.06-1.24] | 1.13 [1.04-1.23] | 0.99 [0.81-1.21] | 1.09 [1.00-1.19] | 1.11 [1.03-1.19] | 1.12 [1.03-1.21] | 1.10 [1.01-1.20] | 1.02 [0.83-1.24] | 1.09 [1.00-1.18] | 1,672 | 1,469 | 1,338 | 243 | 1,326 |
| C72 | 1.20 [0.90-1.59] | | | | | 1.11 [0.82-1.49] | | | | | 120 | NA | NA | NA | NA |
| C73 | 1.06 [0.96-1.18] | 1.06 [0.95-1.18] | 1.06 [0.94-1.20] | | 1.02 [0.91-1.15] | 1.04 [0.93-1.16] | 1.02 [0.92-1.14] | 1.03 [0.92-1.17] | | 1.01 [0.90-1.14] | 819 | 798 | 618 | NA | 675 |
| C74 | 1.20 [0.93-1.55] | 1.13 [0.83-1.52] | | | 1.09 [0.83-1.42] | 1.19 [0.92-1.54] | 1.06 [0.77-1.44] | | | 1.08 [0.82-1.42] | 145 | 107 | NA | NA | 129 |
| C75 | 1.07 [0.83-1.39] | 1.00 [0.76-1.32] | 1.07 [0.81-1.41] | | 0.99 [0.75-1.30] | 1.04 [0.80-1.36] | 0.96 [0.73-1.27] | 1.00 [0.75-1.33] | | 1.00 [0.76-1.32] | 137 | 121 | 112 | NA | 121 |
| C76 | 1.10 [0.96-1.27] | 1.11 [0.95-1.28] | 1.07 [0.92-1.25] | | 1.09 [0.93-1.26] | 1.11 [0.96-1.28] | 1.09 [0.94-1.27] | 1.06 [0.91-1.24] | | 1.09 [0.94-1.28] | 472 | 431 | 415 | NA | 412 |
| C77 | 1.06 [1.03-1.09] | 1.05 [1.03-1.08] | 1.05 [1.03-1.08] | 1.14 [0.86-1.51] | 1.04 [1.01-1.07] | 1.06 [1.04-1.09] | 1.04 [1.01-1.07] | 1.04 [1.01-1.07] | 1.14 [0.86-1.53] | 1.05 [1.02-1.08] | 14,180 | 14,120 | 13,563 | 140 | 11,924 |
| C78 | 1.06 [1.04-1.09] | 1.05 [1.03-1.08] | 1.06 [1.04-1.08] | 1.01 [0.78-1.32] | 1.05 [1.02-1.07] | 1.07 [1.04-1.09] | 1.04 [1.01-1.06] | 1.04 [1.02-1.07] | 1.03 [0.78-1.35] | 1.07 [1.04-1.09] | 20,975 | 20,875 | 20,532 | 147 | 18,290 |
| C79 | 1.08 [1.05-1.10] | 1.07 [1.05-1.10] | 1.08 [1.05-1.10] | 0.98 [0.77-1.25] | 1.05 [1.03-1.08] | 1.08 [1.06-1.11] | 1.05 [1.03-1.08] | 1.06 [1.03-1.08] | 0.99 [0.77-1.27] | 1.08 [1.05-1.11] | 17,880 | 17,755 | 17,443 | 168 | 15,788 |
| C80 | 1.10 [1.06-1.15] | 1.09 [1.05-1.13] | 1.10 [1.06-1.14] | | 1.09 [1.04-1.13] | 1.10 [1.06-1.14] | 1.06 [1.02-1.10] | 1.07 [1.03-1.12] | | 1.09 [1.05-1.14] | 6,357 | 6,332 | 6,242 | NA | 5,385 |
| C81 | 1.71 [1.54-1.90] | 1.72 [1.54-1.91] | 1.62 [1.42-1.85] | 1.41 [1.11-1.79] | 1.46 [1.30-1.65] | 1.64 [1.48-1.83] | 1.65 [1.48-1.85] | 1.52 [1.33-1.74] | 1.33 [1.04-1.71] | 1.43 [1.26-1.61] | 969 | 870 | 588 | 185 | 701 |
| C82 | 1.16 [1.04-1.29] | 1.10 [0.99-1.23] | 1.10 [0.98-1.23] | | 1.04 [0.93-1.17] | 1.10 [0.99-1.23] | 1.04 [0.93-1.16] | 1.05 [0.94-1.17] | | 1.02 [0.90-1.15] | 790 | 782 | 744 | NA | 658 |
| C83 | 1.24 [1.17-1.32] | 1.26 [1.18-1.34] | 1.23 [1.15-1.31] | | 1.19 [1.11-1.27] | 1.21 [1.13-1.29] | 1.22 [1.14-1.30] | 1.19 [1.11-1.27] | | 1.17 [1.09-1.25] | 2,429 | 2,371 | 2,250 | NA | 2,081 |
| C84 | 2.44 [2.11-2.83] | 2.71 [2.33-3.15] | 2.56 [2.19-2.99] | | 2.15 [1.83-2.53] | 2.41 [2.08-2.80] | 2.64 [2.26-3.08] | 2.48 [2.11-2.90] | | 2.17 [1.84-2.57] | 545 | 530 | 489 | NA | 437 |
| C85 | 1.33 [1.25-1.40] | 1.31 [1.24-1.39] | 1.30 [1.23-1.38] | | 1.23 [1.16-1.30] | 1.28 [1.21-1.35] | 1.26 [1.19-1.33] | 1.23 [1.16-1.31] | | 1.21 [1.13-1.28] | 3,161 | 3,108 | 2,917 | NA | 2,644 |
| C88 | 1.28 [1.10-1.50] | 1.23 [1.06-1.44] | 1.28 [1.10-1.51] | | 1.14 [0.96-1.34] | 1.26 [1.08-1.48] | 1.21 [1.03-1.42] | 1.24 [1.06-1.46] | | 1.12 [0.95-1.33] | 410 | 407 | 395 | NA | 365 |
| C90 | 1.13 [1.06-1.22] | 1.15 [1.07-1.23] | 1.14 [1.06-1.22] | | 1.10 [1.02-1.19] | 1.11 [1.03-1.19] | 1.13 [1.05-1.21] | 1.10 [1.03-1.18] | | 1.08 [1.00-1.17] | 1,995 | 1,988 | 1,969 | NA | 1,686 |
| C91 | 1.07 [1.00-1.15] | 1.12 [1.04-1.21] | 1.09 [1.02-1.18] | 0.97 [0.81-1.15] | 1.04 [0.97-1.12] | 1.05 [0.99-1.13] | 1.08 [1.01-1.17] | 1.05 [0.98-1.13] | 0.97 [0.81-1.15] | 1.03 [0.95-1.11] | 2,123 | 1,828 | 1,763 | 321 | 1,756 |
| C92 | 1.28 [1.18-1.38] | 1.29 [1.20-1.39] | 1.28 [1.19-1.39] | 1.09 [0.81-1.49] | 1.24 [1.15-1.35] | 1.25 [1.16-1.35] | 1.26 [1.16-1.36] | 1.25 [1.15-1.35] | 1.10 [0.80-1.51] | 1.24 [1.14-1.35] | 1,752 | 1,680 | 1,574 | 107 | 1,488 |
| C93 | 1.17 [0.94-1.46] | 1.18 [0.95-1.47] | 1.10 [0.88-1.37] | | 1.11 [0.89-1.40] | 1.19 [0.95-1.48] | 1.14 [0.91-1.42] | 1.07 [0.85-1.34] | | 1.15 [0.91-1.44] | 216 | 213 | 211 | NA | 199 |
| C94 | 1.25 [0.92-1.70] | 1.31 [0.96-1.80] | 1.23 [0.90-1.68] | | | 1.24 [0.91-1.70] | 1.27 [0.92-1.74] | 1.17 [0.85-1.61] | | | 105 | 104 | 103 | NA | NA |
| C95 | 1.22 [1.04-1.43] | 1.34 [1.13-1.60] | 1.32 [1.11-1.57] | | 1.17 [0.99-1.38] | 1.19 [1.02-1.40] | 1.27 [1.06-1.52] | 1.24 [1.03-1.48] | | 1.16 [0.98-1.37] | 395 | 351 | 336 | NA | 357 |
| C97 | 1.06 [0.98-1.15] | 1.08 [1.00-1.17] | 1.05 [0.97-1.14] | | 1.02 [0.94-1.10] | 1.04 [0.96-1.12] | 1.04 [0.96-1.13] | 1.02 [0.94-1.10] | | 1.01 [0.93-1.09] | 1,611 | 1,610 | 1,596 | NA | 1,510 |
| D00 | 1.09 [0.87-1.36] | 1.05 [0.84-1.32] | 1.07 [0.85-1.34] | | 1.02 [0.81-1.29] | 1.05 [0.84-1.32] | 0.99 [0.79-1.24] | 0.98 [0.77-1.23] | | 1.01 [0.80-1.28] | 181 | 181 | 179 | NA | 168 |
| D01 | 1.10 [0.88-1.38] | 1.11 [0.89-1.40] | 1.13 [0.89-1.42] | | 1.00 [0.79-1.27] | 1.04 [0.82-1.30] | 1.02 [0.81-1.29] | 1.04 [0.82-1.31] | | 0.96 [0.76-1.22] | 180 | 180 | 170 | NA | 162 |
| D02 | 1.27 [1.02-1.57] | 1.20 [0.97-1.49] | 1.22 [0.98-1.52] | | 1.14 [0.91-1.43] | 1.23 [0.99-1.54] | 1.13 [0.91-1.41] | 1.14 [0.92-1.43] | | 1.13 [0.90-1.43] | 209 | 208 | 203 | NA | 184 |
| D03 | 1.06 [0.97-1.15] | 1.08 [0.99-1.18] | 1.08 [0.99-1.18] | | 1.02 [0.93-1.12] | 1.02 [0.94-1.11] | 1.04 [0.95-1.13] | 1.04 [0.95-1.14] | | 1.00 [0.91-1.09] | 1,295 | 1,292 | 1,226 | NA | 1,095 |
| D04 | 1.25 [1.18-1.33] | 1.26 [1.19-1.34] | 1.26 [1.19-1.34] | | 1.20 [1.13-1.28] | 1.19 [1.12-1.27] | 1.18 [1.11-1.25] | 1.17 [1.10-1.24] | | 1.16 [1.09-1.23] | 3,098 | 3,093 | 3,069 | NA | 2,827 |
| D05 | 1.01 [0.95-1.07] | 1.01 [0.95-1.07] | 1.00 [0.94-1.06] | | 0.96 [0.90-1.03] | 0.99 [0.93-1.06] | 1.00 [0.93-1.06] | 0.98 [0.92-1.05] | | 0.96 [0.89-1.03] | 2,263 | 2,265 | 2,184 | NA | 1,812 |
| D06 | 1.05 [0.98-1.12] | 1.06 [0.99-1.13] | 1.03 [0.90-1.18] | 0.92 [0.77-1.10] | 0.99 [0.92-1.07] | 1.03 [0.96-1.11] | 1.00 [0.93-1.07] | 0.99 [0.86-1.14] | 0.92 [0.77-1.11] | 1.00 [0.93-1.08] | 2,060 | 2,068 | 455 | 308 | 1,642 |
| D07 | 1.20 [1.08-1.33] | 1.28 [1.15-1.42] | 1.22 [1.09-1.37] | | 1.13 [1.01-1.27] | 1.14 [1.02-1.28] | 1.20 [1.07-1.34] | 1.16 [1.03-1.30] | | 1.10 [0.98-1.24] | 799 | 797 | 738 | NA | 700 |
| D09 | 1.12 [1.00-1.26] | 1.13 [1.00-1.27] | 1.13 [1.01-1.27] | | 1.05 [0.93-1.19] | 1.11 [0.98-1.25] | 1.08 [0.96-1.21] | 1.10 [0.97-1.23] | | 1.05 [0.93-1.19] | 694 | 697 | 693 | NA | 635 |

#### Supplementary Table 3: Hazard ratios and events from all cohorts

| | Hazard ratio (99% confidence interval) | | | | | | | | | | Events (in exposed) | | | | |
|---|---|---|---|---|---|---|---|---|---|---|---|---|---|---|---|
| | crude | | | | | adjusted | | | | | | | | | |
| Outcome | any age | 18+ | 40+ | <18 | hosp. | any age | 18+ | 40+ | <18 | hosp. | any age | 18+ | 40+ | <18 | hosp. |
| D10 | 1.27 [1.15-1.41] | 1.33 [1.20-1.49] | 1.35 [1.19-1.54] | 1.14 [0.90-1.45] | 1.25 [1.12-1.40] | 1.21 [1.09-1.35] | 1.19 [1.06-1.33] | 1.21 [1.06-1.38] | 1.13 [0.88-1.44] | 1.20 [1.07-1.35] | 906 | 794 | 577 | 181 | 728 |
| D11 | 1.04 [0.94-1.16] | 1.04 [0.94-1.16] | 1.04 [0.93-1.16] | | 1.02 [0.91-1.15] | 1.04 [0.93-1.16] | 1.00 [0.90-1.12] | 0.99 [0.88-1.11] | | 1.02 [0.91-1.15] | 812 | 795 | 689 | NA | 664 |
| D12 | 1.21 [1.19-1.24] | 1.21 [1.19-1.24] | 1.21 [1.18-1.23] | 1.26 [0.96-1.64] | 1.13 [1.11-1.16] | 1.14 [1.12-1.16] | 1.11 [1.09-1.13] | 1.11 [1.08-1.13] | 1.22 [0.93-1.61] | 1.09 [1.07-1.12] | 25,696 | 25,649 | 24,951 | 150 | 22,280 |
| D13 | 1.22 [1.13-1.31] | 1.21 [1.12-1.30] | 1.21 [1.12-1.31] | | 1.10 [1.02-1.19] | 1.12 [1.04-1.21] | 1.09 [1.01-1.17] | 1.09 [1.01-1.18] | | 1.04 [0.96-1.13] | 1,755 | 1,739 | 1,613 | NA | 1,479 |
| D14 | 1.31 [1.15-1.48] | 1.30 [1.14-1.48] | 1.32 [1.14-1.53] | | 1.16 [1.01-1.33] | 1.23 [1.08-1.40] | 1.16 [1.02-1.33] | 1.22 [1.05-1.42] | | 1.12 [0.97-1.29] | 609 | 560 | 433 | NA | 489 |
| D15 | 1.15 [0.93-1.42] | 1.15 [0.92-1.43] | 1.16 [0.93-1.45] | | 1.07 [0.85-1.34] | 1.14 [0.92-1.41] | 1.11 [0.89-1.38] | 1.12 [0.89-1.40] | | 1.06 [0.84-1.33] | 214 | 200 | 186 | NA | 176 |
| D16 | 1.18 [1.07-1.29] | 1.27 [1.13-1.43] | 1.20 [1.03-1.40] | 1.07 [0.94-1.22] | 1.08 [0.98-1.21] | 1.14 [1.03-1.25] | 1.18 [1.04-1.33] | 1.09 [0.93-1.28] | 1.06 [0.92-1.21] | 1.05 [0.94-1.17] | 1,123 | 647 | 390 | 604 | 878 |
| D17 | 1.16 [1.12-1.20] | 1.16 [1.12-1.20] | 1.13 [1.09-1.18] | 1.13 [0.98-1.30] | 1.07 [1.03-1.11] | 1.10 [1.07-1.14] | 1.08 [1.05-1.12] | 1.06 [1.02-1.10] | 1.07 [0.93-1.23] | 1.03 [0.99-1.07] | 7,797 | 7,547 | 6,147 | 516 | 6,248 |
| D18 | 1.17 [1.11-1.23] | 1.16 [1.10-1.23] | 1.19 [1.12-1.26] | 1.14 [1.02-1.28] | 1.11 [1.05-1.17] | 1.11 [1.05-1.17] | 1.07 [1.01-1.13] | 1.09 [1.02-1.16] | 1.13 [1.01-1.27] | 1.07 [1.01-1.13] | 3,729 | 3,083 | 2,567 | 790 | 3,237 |
| D21 | 1.28 [1.16-1.42] | 1.34 [1.21-1.49] | 1.25 [1.12-1.40] | | 1.21 [1.08-1.35] | 1.23 [1.11-1.36] | 1.24 [1.11-1.38] | 1.17 [1.04-1.31] | | 1.16 [1.04-1.30] | 949 | 880 | 712 | NA | 765 |
| D22 | 1.14 [1.10-1.18] | 1.15 [1.11-1.20] | 1.11 [1.06-1.17] | 1.15 [1.08-1.22] | 1.07 [1.03-1.12] | 1.09 [1.05-1.13] | 1.09 [1.05-1.13] | 1.05 [0.99-1.10] | 1.11 [1.04-1.18] | 1.04 [1.00-1.08] | 8,229 | 6,589 | 3,685 | 2,539 | 6,285 |
| D23 | 1.23 [1.18-1.28] | 1.26 [1.20-1.32] | 1.26 [1.20-1.32] | 1.14 [1.05-1.24] | 1.13 [1.08-1.19] | 1.17 [1.13-1.22] | 1.17 [1.12-1.23] | 1.17 [1.11-1.24] | 1.11 [1.02-1.21] | 1.09 [1.04-1.15] | 5,653 | 4,444 | 3,532 | 1,395 | 4,449 |
| D24 | 1.05 [0.99-1.11] | 1.04 [0.98-1.11] | 1.02 [0.94-1.11] | 1.13 [1.00-1.28] | 0.99 [0.93-1.06] | 1.01 [0.95-1.07] | 0.99 [0.93-1.06] | 0.98 [0.90-1.07] | 1.12 [0.98-1.27] | 0.97 [0.90-1.04] | 2,632 | 2,432 | 1,299 | 675 | 1,975 |
| D25 | 1.07 [1.04-1.10] | 1.08 [1.05-1.11] | 1.09 [1.06-1.12] | 1.14 [0.89-1.44] | 1.06 [1.03-1.09] | 1.04 [1.01-1.07] | 1.05 [1.02-1.08] | 1.05 [1.02-1.08] | 1.12 [0.87-1.44] | 1.03 [1.00-1.06] | 12,552 | 12,549 | 10,372 | 190 | 9,894 |
| D26 | 1.12 [0.96-1.29] | 1.10 [0.95-1.27] | 1.08 [0.92-1.27] | | 1.09 [0.93-1.29] | 1.09 [0.94-1.27] | 1.08 [0.93-1.25] | 1.04 [0.88-1.23] | | 1.07 [0.90-1.26] | 424 | 420 | 330 | NA | 342 |
| D27 | 1.04 [0.98-1.10] | 1.06 [1.00-1.12] | 1.04 [0.97-1.11] | 1.07 [0.90-1.28] | 1.03 [0.96-1.09] | 1.02 [0.96-1.08] | 1.01 [0.96-1.07] | 0.99 [0.93-1.07] | 1.04 [0.87-1.25] | 1.01 [0.95-1.07] | 2,900 | 2,818 | 1,877 | 349 | 2,332 |
| D28 | 1.25 [1.06-1.47] | 1.26 [1.06-1.49] | 1.14 [0.93-1.40] | | 1.20 [1.00-1.44] | 1.19 [1.00-1.41] | 1.19 [1.00-1.41] | 1.11 [0.90-1.37] | | 1.17 [0.97-1.40] | 347 | 332 | 218 | NA | 287 |
| D29 | 1.23 [1.02-1.49] | 1.21 [0.99-1.49] | 1.14 [0.92-1.42] | | 1.17 [0.95-1.44] | 1.19 [0.98-1.45] | 1.16 [0.95-1.43] | 1.10 [0.89-1.38] | | 1.14 [0.92-1.40] | 263 | 232 | 208 | NA | 225 |
| D30 | 1.11 [0.96-1.28] | 1.08 [0.94-1.25] | 1.08 [0.94-1.26] | | 1.03 [0.88-1.20] | 1.08 [0.93-1.25] | 1.02 [0.88-1.18] | 1.03 [0.89-1.20] | | 1.01 [0.87-1.17] | 450 | 443 | 422 | NA | 400 |
| D31 | 1.28 [1.12-1.46] | 1.30 [1.12-1.50] | 1.26 [1.07-1.47] | 1.33 [1.01-1.74] | 1.28 [1.11-1.48] | 1.25 [1.09-1.43] | 1.23 [1.06-1.43] | 1.20 [1.02-1.41] | 1.29 [0.98-1.70] | 1.26 [1.09-1.46] | 571 | 459 | 394 | 143 | 495 |
| D32 | 1.13 [1.05-1.22] | 1.09 [1.01-1.18] | 1.11 [1.03-1.21] | | 1.09 [1.01-1.18] | 1.09 [1.01-1.18] | 1.03 [0.95-1.11] | 1.05 [0.97-1.14] | | 1.07 [0.99-1.17] | 1,701 | 1,692 | 1,611 | NA | 1,525 |
| D33 | 1.13 [1.00-1.27] | 1.08 [0.96-1.22] | 1.11 [0.97-1.27] | | 1.03 [0.91-1.17] | 1.05 [0.93-1.18] | 0.98 [0.87-1.11] | 1.02 [0.89-1.17] | | 0.99 [0.87-1.12] | 685 | 612 | 539 | NA | 594 |
| D34 | 1.07 [0.93-1.24] | 1.04 [0.90-1.21] | 1.04 [0.89-1.23] | | 1.06 [0.91-1.24] | 1.03 [0.88-1.19] | 0.98 [0.84-1.14] | 1.00 [0.85-1.19] | | 1.01 [0.86-1.19] | 432 | 419 | 333 | NA | 381 |
| D35 | 1.13 [1.06-1.20] | 1.13 [1.06-1.20] | 1.13 [1.05-1.20] | | 1.04 [0.97-1.11] | 1.08 [1.02-1.15] | 1.05 [0.99-1.12] | 1.04 [0.98-1.12] | | 1.01 [0.95-1.08] | 2,516 | 2,486 | 2,196 | NA | 2,170 |
| D36 | 1.22 [1.12-1.33] | 1.23 [1.12-1.34] | 1.24 [1.12-1.37] | 1.04 [0.84-1.29] | 1.12 [1.02-1.23] | 1.14 [1.04-1.24] | 1.13 [1.03-1.24] | 1.13 [1.02-1.26] | 0.99 [0.79-1.24] | 1.08 [0.98-1.19] | 1,284 | 1,144 | 903 | 213 | 1,064 |
| D37 | 1.10 [1.04-1.16] | 1.10 [1.05-1.16] | 1.11 [1.05-1.17] | | 1.09 [1.03-1.16] | 1.07 [1.01-1.13] | 1.06 [1.00-1.12] | 1.06 [1.00-1.12] | | 1.09 [1.03-1.15] | 3,261 | 3,236 | 3,142 | NA | 2,802 |
| D38 | 1.22 [1.10-1.36] | 1.21 [1.09-1.35] | 1.20 [1.08-1.34] | | 1.15 [1.03-1.29] | 1.20 [1.08-1.34] | 1.15 [1.03-1.28] | 1.14 [1.02-1.27] | | 1.17 [1.04-1.31] | 833 | 823 | 792 | NA | 707 |
| D39 | 1.06 [0.92-1.22] | 1.02 [0.88-1.18] | 0.97 [0.83-1.14] | | 1.09 [0.93-1.28] | 1.06 [0.92-1.22] | 1.02 [0.88-1.18] | 0.94 [0.79-1.11] | | 1.12 [0.95-1.31] | 459 | 438 | 329 | NA | 373 |
| D40 | 1.12 [0.88-1.43] | 1.13 [0.88-1.45] | 1.21 [0.92-1.59] | | 1.13 [0.86-1.47] | 1.11 [0.87-1.43] | 1.11 [0.86-1.43] | 1.17 [0.88-1.54] | | 1.11 [0.85-1.46] | 158 | 151 | 122 | NA | 130 |
| D41 | 1.09 [1.02-1.16] | 1.09 [1.02-1.17] | 1.10 [1.03-1.18] | | 1.02 [0.95-1.09] | 1.07 [1.00-1.15] | 1.06 [0.99-1.13] | 1.06 [1.00-1.14] | | 1.02 [0.95-1.09] | 2,275 | 2,266 | 2,225 | NA | 1,938 |
| D43 | 1.13 [1.02-1.26] | 1.11 [0.99-1.25] | 1.06 [0.94-1.20] | 1.22 [0.95-1.57] | 1.09 [0.97-1.22] | 1.11 [0.99-1.23] | 1.07 [0.95-1.21] | 1.01 [0.89-1.14] | 1.23 [0.95-1.59] | 1.08 [0.96-1.22] | 833 | 701 | 611 | 164 | 702 |
| D44 | 1.17 [1.02-1.34] | 1.16 [1.01-1.34] | 1.26 [1.08-1.47] | | 1.08 [0.94-1.26] | 1.10 [0.96-1.27] | 1.07 [0.92-1.24] | 1.16 [0.99-1.35] | | 1.06 [0.91-1.23] | 512 | 465 | 395 | NA | 439 |
| D45 | 1.10 [0.99-1.23] | 1.13 [1.01-1.25] | 1.06 [0.96-1.19] | | 1.06 [0.95-1.19] | 1.09 [0.98-1.21] | 1.06 [0.95-1.18] | 1.00 [0.89-1.11] | | 1.04 [0.93-1.17] | 839 | 837 | 776 | NA | 704 |
| D46 | 1.32 [1.24-1.42] | 1.34 [1.26-1.44] | 1.33 [1.24-1.42] | | 1.26 [1.17-1.35] | 1.29 [1.20-1.38] | 1.28 [1.19-1.37] | 1.26 [1.18-1.35] | | 1.24 [1.16-1.34] | 2,285 | 2,273 | 2,247 | NA | 2,056 |
| D47 | 1.26 [1.20-1.33] | 1.27 [1.20-1.34] | 1.25 [1.19-1.32] | 1.36 [0.98-1.88] | 1.19 [1.13-1.26] | 1.20 [1.13-1.26] | 1.18 [1.11-1.24] | 1.16 [1.10-1.23] | 1.30 [0.92-1.82] | 1.16 [1.09-1.22] | 3,677 | 3,627 | 3,488 | 101 | 3,408 |
| D48 | 1.09 [1.01-1.17] | 1.12 [1.03-1.21] | 1.11 [1.02-1.20] | 0.93 [0.75-1.15] | 1.03 [0.95-1.11] | 1.05 [0.97-1.13] | 1.06 [0.98-1.15] | 1.05 [0.96-1.15] | 0.92 [0.74-1.15] | 1.01 [0.93-1.09] | 1,721 | 1,557 | 1,319 | 229 | 1,400 |
| Diseases of the blood and blood-forming organs and certain disorders involving the immune mechanism (D50-D89) | | | | | | | | | | | | | | | |
| D50 | 1.32 [1.31-1.34] | 1.32 [1.30-1.34] | 1.32 [1.30-1.34] | 1.28 [1.21-1.35] | 1.25 [1.23-1.27] | 1.25 [1.23-1.27] | 1.20 [1.18-1.22] | 1.20 [1.18-1.22] | 1.24 [1.18-1.31] | 1.20 [1.19-1.22] | 53,162 | 50,841 | 45,714 | 4,007 | 47,985 |
| D51 | 1.38 [1.32-1.44] | 1.35 [1.29-1.42] | 1.36 [1.30-1.43] | 1.43 [1.07-1.91] | 1.29 [1.23-1.35] | 1.29 [1.23-1.35] | 1.23 [1.17-1.29] | 1.24 [1.18-1.31] | 1.33 [0.99-1.80] | 1.23 [1.17-1.29] | 4,681 | 4,667 | 4,277 | 132 | 4,210 |
| D52 | 1.28 [1.21-1.35] | 1.31 [1.24-1.38] | 1.30 [1.23-1.38] | | 1.25 [1.18-1.32] | 1.23 [1.17-1.31] | 1.22 [1.15-1.29] | 1.20 [1.14-1.28] | | 1.24 [1.17-1.31] | 3,437 | 3,417 | 3,254 | NA | 3,181 |
| D53 | 1.42 [1.28-1.58] | 1.43 [1.28-1.59] | 1.53 [1.37-1.71] | | 1.35 [1.21-1.50] | 1.34 [1.20-1.49] | 1.30 [1.16-1.45] | 1.37 [1.23-1.54] | | 1.30 [1.17-1.45] | 1,003 | 976 | 937 | NA | 928 |
| D55 | 1.50 [1.17-1.91] | 1.61 [1.18-2.18] | | | 1.40 [1.08-1.82] | 1.47 [1.14-1.89] | 1.50 [1.09-2.06] | | | 1.41 [1.08-1.83] | 181 | 108 | NA | NA | 157 |
| D56 | 1.40 [1.31-1.50] | 1.42 [1.32-1.53] | 1.54 [1.40-1.70] | 1.34 [1.16-1.56] | 1.27 [1.18-1.37] | 1.37 [1.27-1.47] | 1.37 [1.27-1.48] | 1.42 [1.28-1.57] | 1.31 [1.12-1.53] | 1.25 [1.16-1.35] | 2,201 | 1,898 | 1,101 | 518 | 1,811 |
| D57 | 1.31 [1.23-1.40] | 1.37 [1.27-1.47] | 1.30 [1.17-1.44] | 1.36 [1.23-1.51] | 1.22 [1.13-1.31] | 1.28 [1.20-1.37] | 1.32 [1.22-1.43] | 1.23 [1.10-1.37] | 1.36 [1.22-1.51] | 1.21 [1.12-1.30] | 2,519 | 1,759 | 859 | 1,054 | 1,979 |
| D58 | 1.21 [1.05-1.38] | 1.24 [1.07-1.44] | 1.18 [0.99-1.40] | 1.21 [0.92-1.59] | 1.17 [1.01-1.35] | 1.19 [1.04-1.37] | 1.19 [1.02-1.39] | 1.12 [0.94-1.34] | 1.20 [0.90-1.58] | 1.17 [1.01-1.35] | 534 | 433 | 317 | 137 | 466 |
| D59 | 1.44 [1.28-1.62] | 1.47 [1.30-1.67] | 1.43 [1.25-1.64] | 1.27 [0.96-1.67] | 1.36 [1.20-1.54] | 1.42 [1.26-1.59] | 1.43 [1.25-1.63] | 1.38 [1.20-1.58] | 1.25 [0.94-1.67] | 1.36 [1.20-1.55] | 745 | 639 | 566 | 133 | 654 |
| D61 | 1.31 [1.24-1.39] | 1.33 [1.25-1.41] | 1.30 [1.22-1.38] | 1.07 [0.87-1.30] | 1.25 [1.18-1.32] | 1.27 [1.20-1.35] | 1.26 [1.19-1.34] | 1.22 [1.15-1.30] | 1.04 [0.84-1.27] | 1.24 [1.17-1.31] | 3,263 | 3,099 | 2,850 | 249 | 2,999 |
| D62 | 1.13 [1.04-1.23] | 1.13 [1.04-1.23] | 1.18 [1.05-1.33] | 0.99 [0.79-1.23] | 1.07 [0.98-1.18] | 1.12 [1.03-1.22] | 1.10 [1.01-1.20] | 1.12 [0.99-1.26] | 0.98 [0.78-1.23] | 1.08 [0.99-1.19] | 1,430 | 1,401 | 686 | 211 | 1,170 |
| D63 | 1.28 [1.23-1.34] | 1.29 [1.23-1.35] | 1.27 [1.21-1.33] | 1.18 [0.86-1.62] | 1.20 [1.15-1.26] | 1.24 [1.19-1.30] | 1.21 [1.15-1.27] | 1.18 [1.13-1.24] | 1.16 [0.83-1.60] | 1.20 [1.14-1.26] | 4,840 | 4,774 | 4,591 | 103 | 4,427 |
| D64 | 1.30 [1.28-1.31] | 1.30 [1.29-1.32] | 1.30 [1.29-1.32] | 1.17 [1.11-1.23] | 1.23 [1.21-1.24] | 1.24 [1.23-1.26] | 1.21 [1.19-1.22] | 1.21 [1.19-1.22] | 1.14 [1.08-1.21] | 1.20 [1.18-1.21] | 69,443 | 67,794 | 61,959 | 3,476 | 62,148 |
| D65 | 1.19 [0.96-1.46] | 1.25 [1.00-1.57] | 1.21 [0.95-1.53] | | 1.16 [0.93-1.45] | 1.17 [0.94-1.44] | 1.21 [0.96-1.51] | 1.17 [0.92-1.49] | | 1.17 [0.93-1.46] | 221 | 199 | 174 | NA | 190 |
| D66 | 1.15 [0.94-1.40] | 1.13 [0.90-1.43] | 1.18 [0.90-1.57] | | 1.20 [0.97-1.50] | 1.11 [0.90-1.36] | 1.03 [0.81-1.31] | 1.09 [0.82-1.45] | | 1.17 [0.94-1.47] | 242 | 177 | 121 | NA | 204 |
| D68 | 1.29 [1.22-1.35] | 1.32 [1.25-1.39] | 1.34 [1.26-1.42] | 1.09 [0.96-1.23] | 1.19 [1.13-1.25] | 1.19 [1.13-1.25] | 1.18 [1.11-1.24] | 1.18 [1.11-1.26] | 1.03 [0.90-1.17] | 1.13 [1.07-1.19] | 3,974 | 3,599 | 2,602 | 650 | 3,500 |
| D69 | 1.23 [1.20-1.27] | 1.27 [1.23-1.31] | 1.28 [1.24-1.33] | 1.06 [1.00-1.13] | 1.18 [1.14-1.21] | 1.20 [1.16-1.23] | 1.20 [1.16-1.24] | 1.20 [1.16-1.25] | 1.06 [1.00-1.13] | 1.17 [1.13-1.20] | 11,780 | 9,406 | 8,184 | 2,747 | 10,458 |
| D70 | 1.23 [1.19-1.27] | 1.24 [1.20-1.28] | 1.23 [1.19-1.28] | 1.14 [1.04-1.24] | 1.18 [1.14-1.22] | 1.20 [1.16-1.24] | 1.19 [1.15-1.23] | 1.19 [1.15-1.23] | 1.11 [1.02-1.22] | 1.17 [1.13-1.21] | 9,777 | 8,724 | 7,840 | 1,334 | 8,508 |
| D72 | 1.43 [1.34-1.53] | 1.44 [1.35-1.55] | 1.40 [1.30-1.51] | 1.41 [1.18-1.68] | 1.35 [1.26-1.45] | 1.31 [1.22-1.40] | 1.27 [1.18-1.37] | 1.25 [1.16-1.35] | 1.31 [1.10-1.58] | 1.26 [1.17-1.35] | 2,421 | 2,176 | 1,866 | 358 | 2,199 |
| D73 | 1.25 [1.17-1.34] | 1.24 [1.16-1.32] | 1.26 [1.17-1.35] | 0.92 [0.72-1.18] | 1.16 [1.08-1.24] | 1.20 [1.12-1.28] | 1.15 [1.07-1.23] | 1.16 [1.08-1.25] | 0.95 [0.74-1.22] | 1.14 [1.07-1.23] | 2,222 | 2,116 | 1,898 | 169 | 2,030 |
| D75 | 1.21 [1.14-1.29] | 1.22 [1.15-1.30] | 1.21 [1.14-1.29] | 1.18 [0.86-1.62] | 1.16 [1.09-1.24] | 1.17 [1.10-1.24] | 1.12 [1.05-1.19] | 1.12 [1.05-1.19] | 1.12 [0.81-1.55] | 1.15 [1.07-1.22] | 2,692 | 2,637 | 2,448 | 107 | 2,406 |
| D76 | 1.27 [1.02-1.57] | 1.37 [1.06-1.78] | 1.38 [1.03-1.85] | | 1.28 [1.01-1.61] | 1.20 [0.97-1.50] | 1.26 [0.96-1.65] | 1.27 [0.94-1.72] | | 1.21 [0.96-1.54] | 213 | 141 | 108 | NA | 178 |
| D80 | 1.54 [1.37-1.73] | 1.56 [1.36-1.77] | 1.46 [1.27-1.69] | 1.47 [1.18-1.84] | 1.44 [1.28-1.62] | 1.36 [1.20-1.53] | 1.32 [1.15-1.52] | 1.21 [1.04-1.41] | 1.33 [1.06-1.68] | 1.30 [1.15-1.47] | 784 | 597 | 494 | 223 | 735 |
| D82 | 1.31 [1.04-1.65] | 1.51 [1.12-2.02] | | | 1.27 [1.00-1.62] | 1.19 [0.94-1.51] | 1.33 [0.98-1.80] | | | 1.19 [0.92-1.53] | 188 | 116 | NA | NA | 169 |
| D83 | 2.01 [1.46-2.75] | 2.05 [1.48-2.85] | | | 1.74 [1.28-2.37] | 1.76 [1.27-2.43] | 1.75 [1.24-2.46] | | | 1.55 [1.12-2.13] | 119 | 104 | NA | NA | 114 |
| D84 | 1.71 [1.47-1.99] | 1.70 [1.41-2.05] | 1.75 [1.42-2.16] | 1.76 [1.37-2.27] | 1.55 [1.32-1.81] | 1.43 [1.22-1.69] | 1.35 [1.10-1.65] | 1.38 [1.10-1.72] | 1.59 [1.22-2.06] | 1.33 [1.13-1.56] | 464 | 296 | 234 | 186 | 433 |
| D86 | 1.58 [1.46-1.71] | 1.56 [1.44-1.69] | 1.51 [1.39-1.64] | | 1.43 [1.32-1.56] | 1.47 [1.35-1.59] | 1.41 [1.30-1.54] | 1.35 [1.23-1.47] | | 1.34 [1.23-1.46] | 1,617 | 1,586 | 1,381 | NA | 1,402 |
| D89 | 1.43 [1.31-1.57] | 1.42 [1.29-1.56] | 1.41 [1.28-1.55] | 1.53 [1.13-2.08] | 1.32 [1.20-1.45] | 1.33 [1.21-1.46] | 1.29 [1.17-1.42] | 1.28 [1.16-1.41] | 1.37 [1.00-1.89] | 1.26 [1.14-1.38] | 1,274 | 1,180 | 1,098 | 120 | 1,133 |
| Endocrine, nutritional and metabolic diseases (E00-E89) | | | | | | | | | | | | | | | |
| E02 | 1.14 [0.97-1.35] | 1.20 [1.02-1.41] | 1.11 [0.93-1.32] | | 1.11 [0.93-1.31] | 1.12 [0.95-1.33] | 1.16 [0.98-1.37] | 1.07 [0.89-1.28] | | 1.11 [0.93-1.31] | 395 | 392 | 347 | NA | 364 |
| E03 | 1.34 [1.32-1.36] | 1.34 [1.32-1.36] | 1.33 [1.31-1.35] | 1.20 [1.11-1.31] | 1.26 [1.24-1.28] | 1.28 [1.26-1.30] | 1.24 [1.23-1.26] | 1.24 [1.22-1.26] | 1.18 [1.09-1.29] | 1.22 [1.20-1.24] | 50,515 | 49,910 | 44,793 | 1,612 | 41,507 |
| E04 | 1.15 [1.10-1.20] | 1.13 [1.08-1.18] | 1.12 [1.07-1.17] | 1.24 [0.98-1.58] | 1.09 [1.04-1.15] | 1.09 [1.04-1.14] | 1.05 [1.01-1.10] | 1.04 [0.99-1.09] | 1.24 [0.97-1.59] | 1.06 [1.01-1.12] | 4,652 | 4,564 | 4,023 | 181 | 4,054 |
| E05 | 1.29 [1.25-1.34] | 1.29 [1.24-1.33] | 1.27 [1.22-1.32] | 1.28 [1.09-1.50] | 1.21 [1.17-1.25] | 1.23 [1.18-1.27] | 1.18 [1.14-1.22] | 1.17 [1.12-1.21] | 1.24 [1.05-1.46] | 1.16 [1.12-1.21] | 8,314 | 8,168 | 6,931 | 419 | 7,289 |
| E06 | 1.22 [1.10-1.35] | 1.22 [1.10-1.36] | 1.24 [1.10-1.40] | | 1.20 [1.07-1.34] | 1.17 [1.05-1.30] | 1.13 [1.01-1.26] | 1.16 [1.02-1.31] | | 1.14 [1.02-1.28] | 883 | 841 | 636 | NA | 787 |
| E07 | 1.26 [1.16-1.37] | 1.29 [1.19-1.41] | 1.27 [1.16-1.39] | | 1.18 [1.08-1.29] | 1.17 [1.07-1.27] | 1.17 [1.08-1.28] | 1.15 [1.05-1.26] | | 1.11 [1.01-1.21] | 1,392 | 1,364 | 1,204 | NA | 1,266 |
| E10 | 1.23 [1.19-1.26] | 1.30 [1.25-1.34] | 1.32 [1.27-1.36] | 0.97 [0.91-1.04] | 1.14 [1.11-1.18] | 1.15 [1.12-1.19] | 1.12 [1.09-1.16] | 1.13 [1.09-1.17] | 0.96 [0.90-1.03] | 1.11 [1.07-1.15] | 10,587 | 8,646 | 7,458 | 2,324 | 8,645 |
| E11 | 1.26 [1.24-1.27] | 1.26 [1.24-1.27] | 1.25 [1.23-1.26] | 1.25 [1.09-1.43] | 1.20 [1.18-1.21] | 1.19 [1.18-1.20] | 1.12 [1.11-1.14] | 1.11 [1.10-1.12] | 1.23 [1.08-1.41] | 1.16 [1.15-1.18] | 74,974 | 74,783 | 72,124 | 622 | 59,224 |
| E13 | 1.41 [1.28-1.55] | 1.39 [1.26-1.54] | 1.43 [1.29-1.59] | | 1.29 [1.16-1.42] | 1.25 [1.13-1.39] | 1.18 [1.06-1.31] | 1.21 [1.09-1.35] | | 1.19 [1.08-1.32] | 1,051 | 1,010 | 890 | NA | 972 |
| E14 | 1.37 [1.33-1.41] | 1.38 [1.34-1.42] | 1.37 [1.33-1.41] | 0.92 [0.75-1.14] | 1.27 [1.23-1.31] | 1.25 [1.22-1.29] | 1.19 [1.15-1.23] | 1.17 [1.13-1.21] | 0.92 [0.74-1.14] | 1.21 [1.17-1.25] | 11,113 | 11,014 | 10,369 | 228 | 10,005 |
| E16 | 1.25 [1.22-1.29] | 1.27 [1.24-1.31] | 1.27 [1.23-1.31] | 1.07 [0.99-1.15] | 1.16 [1.13-1.20] | 1.18 [1.14-1.21] | 1.11 [1.08-1.14] | 1.10 [1.07-1.14] | 1.05 [0.97-1.14] | 1.14 [1.10-1.17] | 12,636 | 11,294 | 10,504 | 1,633 | 11,955 |
| E20 | 1.44 [1.21-1.70] | 1.49 [1.25-1.77] | 1.53 [1.27-1.84] | | 1.32 [1.11-1.56] | 1.26 [1.06-1.50] | 1.30 [1.09-1.56] | 1.33 [1.10-1.62] | | 1.23 [1.03-1.46] | 372 | 343 | 302 | NA | 349 |
| E21 | 1.17 [1.11-1.23] | 1.17 [1.11-1.23] | 1.16 [1.10-1.22] | | 1.08 [1.03-1.15] | 1.12 [1.06-1.18] | 1.08 [1.02-1.14] | 1.07 [1.02-1.13] | | 1.06 [1.00-1.12] | 3,577 | 3,548 | 3,397 | NA | 3,267 |
| E22 | 1.48 [1.39-1.57] | 1.47 [1.38-1.56] | 1.51 [1.41-1.61] | 1.35 [1.06-1.71] | 1.39 [1.30-1.48] | 1.41 [1.32-1.50] | 1.37 [1.28-1.46] | 1.41 [1.32-1.50] | 1.28 [1.00-1.63] | 1.34 [1.26-1.43] | 2,906 | 2,778 | 2,574 | 199 | 2,627 |
| E23 | 1.39 [1.28-1.50] | 1.43 [1.30-1.56] | 1.49 [1.34-1.65] | 1.24 [1.07-1.43] | 1.25 [1.15-1.36] | 1.29 [1.19-1.40] | 1.27 [1.15-1.39] | 1.33 [1.20-1.48] | 1.18 [1.02-1.37] | 1.20 [1.10-1.30] | 1,650 | 1,222 | 958 | 519 | 1,470 |
| E24 | 1.60 [1.35-1.90] | 1.46 [1.22-1.74] | 1.61 [1.33-1.96] | | 1.40 [1.17-1.67] | 1.32 [1.10-1.58] | 1.12 [0.93-1.36] | 1.24 [1.01-1.52] | | 1.21 [1.01-1.45] | 357 | 319 | 263 | NA | 327 |
| E26 | 1.21 [0.95-1.54] | 1.16 [0.91-1.49] | 1.14 [0.87-1.50] | | 1.16 [0.90-1.50] | 1.13 [0.88-1.44] | 1.04 [0.80-1.34] | 0.97 [0.73-1.29] | | 1.14 [0.88-1.48] | 163 | 150 | 124 | NA | 142 |
| E27 | 1.49 [1.40-1.57] | 1.48 [1.39-1.57] | 1.45 [1.36-1.54] | 1.46 [1.24-1.71] | 1.37 [1.29-1.45] | 1.34 [1.26-1.42] | 1.28 [1.20-1.36] | 1.25 [1.17-1.34] | 1.32 [1.12-1.55] | 1.27 [1.19-1.35] | 3,166 | 2,827 | 2,446 | 456 | 2,935 |
| E28 | 1.21 [1.17-1.26] | 1.24 [1.20-1.29] | 1.26 [1.15-1.39] | 1.13 [1.06-1.21] | 1.18 [1.13-1.23] | 1.11 [1.07-1.15] | 1.08 [1.04-1.12] | 1.05 [0.95-1.16] | 1.10 [1.03-1.18] | 1.11 [1.06-1.16] | 7,155 | 6,884 | 991 | 2,338 | 5,846 |
| E29 | 1.71 [1.47-1.98] | 1.76 [1.51-2.05] | 1.76 [1.50-2.08] | | 1.45 [1.25-1.69] | 1.50 [1.28-1.75] | 1.42 [1.21-1.67] | 1.40 [1.18-1.66] | | 1.32 [1.13-1.55] | 466 | 445 | 385 | NA | 430 |
| E30 | 1.24 [1.11-1.39] | | | 1.22 [1.09-1.37] | 1.15 [1.02-1.30] | 1.19 [1.06-1.34] | | | 1.17 [1.04-1.32] | 1.13 [0.99-1.28] | 843 | NA | NA | 827 | 725 |
| E34 | 1.23 [1.13-1.34] | 1.31 [1.12-1.53] | 1.39 [1.14-1.70] | 1.19 [1.08-1.31] | 1.16 [1.06-1.27] | 1.14 [1.05-1.24] | 1.13 [0.96-1.34] | 1.23 [1.00-1.50] | 1.11 [1.01-1.22] | 1.11 [1.02-1.22] | 1,485 | 395 | 242 | 1,140 | 1,328 |
| E41 | 1.07 [0.80-1.42] | 1.05 [0.78-1.41] | 1.10 [0.82-1.48] | | 1.15 [0.86-1.55] | 1.10 [0.82-1.47] | 1.06 [0.78-1.42] | 1.09 [0.81-1.48] | | 1.20 [0.89-1.61] | 123 | 121 | 113 | NA | 118 |
| E43 | 1.20 [0.94-1.54] | 1.09 [0.84-1.40] | 1.24 [0.94-1.63] | | 1.11 [0.86-1.43] | 1.18 [0.92-1.53] | 1.00 [0.77-1.30] | 1.20 [0.91-1.60] | | 1.13 [0.87-1.47] | 158 | 148 | 130 | NA | 149 |
| E46 | 1.22 [1.13-1.31] | 1.18 [1.10-1.27] | 1.16 [1.07-1.25] | 1.29 [0.97-1.73] | 1.16 [1.08-1.25] | 1.19 [1.11-1.28] | 1.10 [1.02-1.18] | 1.09 [1.01-1.18] | 1.25 [0.93-1.69] | 1.17 [1.08-1.26] | 2,023 | 1,956 | 1,779 | 135 | 1,874 |
| E51 | 1.47 [1.19-1.82] | 1.41 [1.14-1.74] | 1.48 [1.19-1.84] | | 1.36 [1.09-1.70] | 1.39 [1.12-1.73] | 1.23 [0.99-1.54] | 1.28 [1.02-1.61] | | 1.38 [1.10-1.73] | 216 | 213 | 200 | NA | 197 |
| E53 | 1.30 [1.27-1.34] | 1.31 [1.27-1.35] | 1.28 [1.24-1.33] | 1.27 [1.11-1.45] | 1.24 [1.20-1.28] | 1.23 [1.20-1.27] | 1.18 [1.15-1.22] | 1.17 [1.13-1.21] | 1.19 [1.04-1.37] | 1.19 [1.16-1.23] | 12,236 | 12,126 | 10,701 | 648 | 11,410 |
| E55 | 1.37 [1.34-1.40] | 1.38 [1.34-1.42] | 1.35 [1.32-1.39] | 1.34 [1.25-1.45] | 1.30 [1.26-1.33] | 1.26 [1.23-1.29] | 1.22 [1.19-1.25] | 1.20 [1.17-1.24] | 1.26 [1.17-1.35] | 1.23 [1.19-1.26] | 18,051 | 16,575 | 14,208 | 2,279 | 16,868 |
| E56 | 1.27 [1.00-1.62] | 1.29 [1.00-1.67] | 1.21 [0.92-1.60] | | 1.21 [0.94-1.54] | 1.19 [0.93-1.53] | 1.14 [0.87-1.48] | 1.10 [0.83-1.46] | | 1.18 [0.92-1.52] | 176 | 155 | 132 | NA | 165 |
| E58 | 1.55 [1.18-2.04] | 1.41 [1.06-1.88] | | | 1.55 [1.17-2.06] | 1.48 [1.11-1.95] | 1.25 [0.93-1.68] | | | 1.50 [1.13-1.99] | 141 | 124 | NA | NA | 134 |
| E61 | 1.38 [1.30-1.47] | 1.39 [1.30-1.48] | 1.39 [1.29-1.49] | 1.35 [1.16-1.57] | 1.27 [1.19-1.35] | 1.26 [1.18-1.34] | 1.21 [1.14-1.30] | 1.22 [1.13-1.31] | 1.27 [1.09-1.48] | 1.19 [1.12-1.27] | 2,835 | 2,508 | 2,090 | 509 | 2,635 |
| E63 | 1.30 [1.01-1.67] | 1.17 [0.89-1.52] | 1.46 [1.11-1.94] | | 1.23 [0.95-1.59] | 1.24 [0.96-1.60] | 1.08 [0.82-1.41] | 1.32 [0.99-1.77] | | 1.18 [0.91-1.53] | 165 | 143 | 134 | NA | 157 |
| E65 | 1.45 [1.08-1.95] | 1.54 [1.14-2.08] | | | | 1.33 [0.98-1.80] | 1.39 [1.02-1.90] | | | | 113 | 107 | NA | NA | NA |
| E66 | 1.30 [1.29-1.32] | 1.32 [1.30-1.33] | 1.32 [1.31-1.34] | 1.18 [1.14-1.22] | 1.22 [1.21-1.24] | 1.20 [1.19-1.21] | 1.15 [1.14-1.17] | 1.15 [1.13-1.16] | 1.13 [1.10-1.17] | 1.15 [1.14-1.17] | 88,230 | 86,031 | 60,487 | 11,250 | 77,698 |
| E70 | 1.35 [1.02-1.79] | | | | 1.25 [0.91-1.72] | 1.30 [0.98-1.74] | | | | 1.22 [0.88-1.69] | 128 | NA | NA | NA | 101 |
| E72 | 1.17 [0.93-1.46] | 1.39 [1.07-1.81] | 1.52 [1.12-2.08] | | 1.13 [0.90-1.43] | 1.09 [0.87-1.37] | 1.24 [0.94-1.62] | 1.39 [1.01-1.90] | | 1.12 [0.89-1.43] | 189 | 139 | 104 | NA | 175 |
| E73 | 1.41 [1.28-1.55] | 1.38 [1.21-1.57] | 1.35 [1.14-1.59] | 1.36 [1.20-1.55] | 1.32 [1.19-1.45] | 1.22 [1.11-1.35] | 1.11 [0.96-1.27] | 1.10 [0.92-1.30] | 1.23 [1.08-1.41] | 1.16 [1.05-1.29] | 1,148 | 578 | 356 | 660 | 1,014 |
| E74 | 1.35 [1.14-1.60] | 1.34 [1.10-1.63] | 1.37 [1.10-1.70] | 1.32 [0.98-1.78] | 1.29 [1.08-1.53] | 1.24 [1.05-1.48] | 1.22 [1.00-1.49] | 1.25 [1.01-1.56] | 1.22 [0.89-1.66] | 1.23 [1.03-1.47] | 365 | 257 | 210 | 126 | 324 |
| E75 | 1.20 [0.93-1.55] | | | | 1.11 [0.84-1.47] | 1.17 [0.90-1.52] | | | | 1.12 [0.85-1.48] | 144 | NA | NA | NA | 125 |
| E78 | 1.23 [1.22-1.24] | 1.23 [1.22-1.24] | 1.23 [1.22-1.24] | 1.18 [0.98-1.43] | 1.15 [1.14-1.17] | 1.17 [1.16-1.18] | 1.12 [1.10-1.13] | 1.11 [1.10-1.12] | 1.16 [0.95-1.41] | 1.12 [1.11-1.14] | 97,183 | 97,116 | 95,613 | 285 | 82,546 |
| E79 | 1.32 [1.01-1.72] | 1.35 [1.02-1.77] | 1.43 [1.07-1.92] | | 1.19 [0.90-1.58] | 1.25 [0.95-1.65] | 1.21 [0.91-1.60] | 1.25 [0.92-1.69] | | 1.13 [0.85-1.50] | 141 | 132 | 115 | NA | 123 |
| E80 | 1.17 [1.07-1.28] | 1.21 [1.11-1.33] | 1.21 [1.09-1.35] | 1.12 [0.92-1.36] | 1.08 [0.99-1.19] | 1.09 [1.00-1.20] | 1.11 [1.01-1.22] | 1.13 [1.01-1.26] | 1.05 [0.86-1.29] | 1.03 [0.94-1.13] | 1,292 | 1,155 | 824 | 270 | 1,115 |
| E83 | 1.21 [1.18-1.24] | 1.22 [1.19-1.25] | 1.20 [1.17-1.23] | 1.08 [0.96-1.20] | 1.15 [1.12-1.18] | 1.16 [1.13-1.19] | 1.12 [1.09-1.15] | 1.10 [1.08-1.13] | 1.06 [0.95-1.19] | 1.13 [1.10-1.16] | 17,831 | 17,352 | 16,193 | 875 | 16,376 |
| E84 | 1.23 [0.96-1.58] | | | | 1.14 [0.87-1.50] | 1.11 [0.86-1.43] | | | | 1.03 [0.78-1.36] | 160 | NA | NA | NA | 134 |
| E85 | 1.25 [1.10-1.41] | 1.22 [1.07-1.38] | 1.18 [1.04-1.35] | | 1.18 [1.04-1.34] | 1.20 [1.06-1.36] | 1.14 [1.00-1.29] | 1.10 [0.96-1.25] | | 1.14 [1.00-1.30] | 655 | 623 | 590 | NA | 596 |
| E86 | 1.20 [1.18-1.21] | 1.19 [1.17-1.21] | 1.19 [1.17-1.21] | 1.13 [1.08-1.18] | 1.14 [1.12-1.16] | 1.16 [1.14-1.18] | 1.12 [1.10-1.14] | 1.12 [1.10-1.14] | 1.10 [1.05-1.15] | 1.13 [1.11-1.15] | 47,668 | 43,859 | 41,005 | 4,893 | 43,955 |
| E87 | 1.28 [1.27-1.30] | 1.29 [1.27-1.30] | 1.28 [1.27-1.30] | 1.09 [1.02-1.15] | 1.22 [1.21-1.24] | 1.23 [1.22-1.25] | 1.18 [1.17-1.20] | 1.18 [1.17-1.20] | 1.06 [1.00-1.13] | 1.20 [1.18-1.21] | 73,651 | 71,910 | 69,111 | 2,751 | 67,831 |
| E88 | 1.29 [1.23-1.36] | 1.32 [1.24-1.39] | 1.31 [1.23-1.39] | 1.16 [0.99-1.36] | 1.23 [1.16-1.30] | 1.23 [1.17-1.30] | 1.21 [1.14-1.28] | 1.20 [1.13-1.27] | 1.12 [0.95-1.32] | 1.20 [1.13-1.27] | 3,596 | 3,299 | 3,016 | 408 | 3,317 |
| E89 | 1.25 [1.19-1.31] | 1.26 [1.20-1.32] | 1.25 [1.19-1.31] | 1.11 [0.84-1.47] | 1.15 [1.10-1.21] | 1.17 [1.12-1.23] | 1.14 [1.09-1.20] | 1.13 [1.08-1.19] | 1.09 [0.81-1.47] | 1.11 [1.06-1.16] | 4,522 | 4,479 | 4,023 | 133 | 4,194 |
| Mental, Behavioral and Neurodevelopmental disorders (F01-F99) | | | | | | | | | | | | | | | |
| F00 | 1.06 [1.03-1.10] | 1.06 [1.03-1.10] | 1.07 [1.03-1.10] | | 1.05 [1.02-1.09] | 1.06 [1.03-1.10] | 1.04 [1.01-1.08] | 1.05 [1.01-1.08] | | 1.05 [1.01-1.08] | 10,162 | 10,154 | 10,158 | NA | 9,384 |
| F01 | 1.10 [1.07-1.13] | 1.10 [1.06-1.13] | 1.09 [1.06-1.12] | | 1.06 [1.03-1.10] | 1.08 [1.05-1.12] | 1.05 [1.02-1.08] | 1.04 [1.00-1.07] | | 1.06 [1.03-1.09] | 11,780 | 11,779 | 11,773 | NA | 11,049 |
| F02 | 1.01 [0.94-1.09] | 1.01 [0.94-1.09] | 1.02 [0.95-1.10] | | 0.95 [0.88-1.03] | 1.02 [0.95-1.10] | 0.99 [0.92-1.07] | 1.00 [0.93-1.08] | | 0.97 [0.90-1.05] | 1,911 | 1,907 | 1,898 | NA | 1,762 |
| F03 | 1.07 [1.05-1.09] | 1.06 [1.04-1.09] | 1.07 [1.05-1.09] | | 1.04 [1.02-1.06] | 1.07 [1.05-1.09] | 1.04 [1.02-1.06] | 1.04 [1.02-1.06] | | 1.04 [1.02-1.07] | 28,174 | 28,162 | 28,133 | NA | 25,894 |
| F05 | 1.17 [1.15-1.20] | 1.17 [1.15-1.19] | 1.17 [1.15-1.19] | 1.00 [0.77-1.30] | 1.14 [1.12-1.16] | 1.14 [1.12-1.17] | 1.10 [1.08-1.13] | 1.10 [1.08-1.13] | 1.01 [0.78-1.32] | 1.13 [1.11-1.16] | 27,443 | 27,341 | 27,145 | 161 | 26,053 |
| F06 | 1.16 [1.10-1.22] | 1.16 [1.10-1.22] | 1.17 [1.11-1.23] | | 1.10 [1.04-1.16] | 1.13 [1.07-1.19] | 1.10 [1.04-1.16] | 1.11 [1.05-1.16] | | 1.09 [1.04-1.15] | 4,231 | 4,198 | 4,118 | NA | 3,968 |
| F07 | 1.28 [1.07-1.53] | 1.29 [1.07-1.55] | 1.35 [1.09-1.68] | | 1.09 [0.90-1.32] | 1.18 [0.99-1.42] | 1.16 [0.96-1.41] | 1.23 [0.98-1.53] | | 1.08 [0.89-1.31] | 319 | 278 | 207 | NA | 271 |
| F10 | 1.20 [1.18-1.23] | 1.23 [1.20-1.25] | 1.26 [1.23-1.29] | 0.98 [0.93-1.04] | 1.14 [1.11-1.16] | 1.17 [1.15-1.19] | 1.11 [1.09-1.14] | 1.15 [1.12-1.18] | 0.97 [0.92-1.02] | 1.15 [1.12-1.18] | 23,382 | 22,021 | 16,404 | 3,621 | 18,640 |
| F11 | 1.17 [1.08-1.26] | 1.18 [1.10-1.28] | 1.30 [1.17-1.45] | 1.00 [0.81-1.25] | 1.07 [0.99-1.16] | 1.08 [1.00-1.17] | 0.95 [0.88-1.03] | 1.05 [0.94-1.17] | 1.07 [0.85-1.35] | 1.06 [0.97-1.15] | 1,667 | 1,649 | 834 | 200 | 1,382 |
| F12 | 1.08 [1.03-1.14] | 1.12 [1.06-1.18] | 1.28 [1.17-1.41] | 0.96 [0.89-1.04] | 0.95 [0.90-1.01] | 1.01 [0.96-1.07] | 0.95 [0.90-1.00] | 1.06 [0.96-1.17] | 0.97 [0.89-1.05] | 0.96 [0.91-1.02] | 3,794 | 3,351 | 1,033 | 1,749 | 3,051 |
| F13 | 1.26 [1.08-1.47] | 1.24 [1.06-1.45] | 1.42 [1.16-1.74] | | 1.10 [0.93-1.29] | 1.14 [0.97-1.33] | 0.98 [0.83-1.15] | 1.14 [0.92-1.41] | | 1.06 [0.89-1.24] | 416 | 387 | 236 | NA | 367 |
| F14 | 1.12 [1.03-1.21] | 1.16 [1.07-1.26] | 1.42 [1.22-1.65] | 1.02 [0.89-1.16] | 0.98 [0.90-1.07] | 1.04 [0.95-1.13] | 0.97 [0.89-1.05] | 1.13 [0.97-1.33] | 1.03 [0.89-1.18] | 0.99 [0.91-1.08] | 1,455 | 1,412 | 404 | 530 | 1,208 |
| F15 | 1.03 [0.90-1.19] | 1.04 [0.90-1.20] | 1.16 [0.91-1.48] | 0.95 [0.76-1.18] | 0.93 [0.80-1.08] | 0.97 [0.85-1.12] | 0.93 [0.80-1.08] | 0.98 [0.76-1.27] | 0.92 [0.73-1.16] | 0.94 [0.81-1.10] | 503 | 438 | 148 | 200 | 403 |
| F16 | 0.95 [0.73-1.22] | 0.87 [0.66-1.15] | | | 0.87 [0.66-1.14] | 0.91 [0.70-1.19] | 0.78 [0.59-1.03] | | | 0.88 [0.66-1.17] | 153 | 114 | NA | NA | 120 |
| F17 | 1.13 [1.12-1.14] | 1.15 [1.14-1.16] | 1.15 [1.14-1.17] | 0.99 [0.97-1.01] | 1.06 [1.05-1.07] | 1.11 [1.10-1.12] | 1.02 [1.01-1.03] | 1.04 [1.03-1.05] | 1.00 [0.98-1.02] | 1.09 [1.08-1.10] | 108,659 | 105,861 | 64,812 | 21,247 | 90,044 |
| F19 | 1.07 [0.99-1.16] | 1.08 [1.00-1.17] | 1.22 [1.07-1.39] | 0.93 [0.81-1.06] | 0.96 [0.89-1.05] | 1.00 [0.92-1.09] | 0.89 [0.82-0.97] | 0.96 [0.84-1.10] | 0.96 [0.83-1.10] | 0.98 [0.90-1.07] | 1,561 | 1,442 | 535 | 525 | 1,355 |
| F20 | 1.28 [1.21-1.34] | 1.27 [1.20-1.34] | 1.32 [1.24-1.40] | 1.00 [0.84-1.20] | 1.15 [1.08-1.22] | 1.26 [1.19-1.33] | 1.17 [1.11-1.24] | 1.25 [1.18-1.34] | 1.04 [0.86-1.25] | 1.17 [1.10-1.24] | 3,454 | 3,406 | 2,516 | 313 | 2,702 |
| F22 | 1.29 [1.19-1.40] | 1.28 [1.18-1.39] | 1.32 [1.21-1.45] | 1.00 [0.74-1.36] | 1.19 [1.10-1.30] | 1.23 [1.14-1.34] | 1.18 [1.08-1.28] | 1.22 [1.11-1.33] | 0.99 [0.72-1.35] | 1.19 [1.09-1.29] | 1,560 | 1,515 | 1,291 | 113 | 1,372 |
| F23 | 1.16 [1.05-1.29] | 1.12 [1.01-1.24] | 1.28 [1.12-1.48] | 0.94 [0.75-1.17] | 1.07 [0.96-1.20] | 1.13 [1.02-1.26] | 1.05 [0.94-1.17] | 1.20 [1.04-1.38] | 0.93 [0.74-1.17] | 1.08 [0.96-1.21] | 916 | 838 | 494 | 194 | 720 |
| F25 | 1.36 [1.21-1.52] | 1.34 [1.20-1.50] | 1.38 [1.20-1.58] | | 1.11 [0.99-1.25] | 1.32 [1.17-1.49] | 1.19 [1.06-1.34] | 1.27 [1.10-1.46] | | 1.15 [1.02-1.30] | 747 | 741 | 510 | NA | 670 |
| F29 | 1.22 [1.13-1.32] | 1.26 [1.17-1.36] | 1.40 [1.28-1.54] | 0.91 [0.77-1.07] | 1.11 [1.03-1.20] | 1.16 [1.07-1.25] | 1.11 [1.03-1.20] | 1.24 [1.13-1.37] | 0.92 [0.77-1.09] | 1.11 [1.02-1.20] | 1,809 | 1,696 | 1,103 | 375 | 1,564 |
| F30 | 1.34 [1.16-1.54] | 1.33 [1.15-1.54] | 1.42 [1.21-1.68] | | 1.19 [1.02-1.39] | 1.28 [1.11-1.48] | 1.21 [1.05-1.41] | 1.31 [1.11-1.55] | | 1.18 [1.01-1.38] | 482 | 465 | 350 | NA | 391 |
| F31 | 1.31 [1.24-1.37] | 1.32 [1.26-1.39] | 1.33 [1.25-1.41] | 1.07 [0.93-1.23] | 1.16 [1.10-1.22] | 1.19 [1.13-1.25] | 1.08 [1.03-1.14] | 1.12 [1.06-1.20] | 1.08 [0.93-1.24] | 1.12 [1.06-1.18] | 4,288 | 4,215 | 2,717 | 559 | 3,768 |
| F32 | 1.37 [1.35-1.38] | 1.38 [1.36-1.40] | 1.39 [1.37-1.41] | 1.14 [1.10-1.17] | 1.26 [1.25-1.28] | 1.23 [1.22-1.25] | 1.13 [1.12-1.14] | 1.15 [1.14-1.17] | 1.13 [1.09-1.16] | 1.20 [1.18-1.21] | 83,130 | 80,435 | 58,205 | 11,060 | 73,576 |
| F33 | 1.31 [1.23-1.39] | 1.30 [1.22-1.39] | 1.28 [1.19-1.37] | 1.10 [0.84-1.44] | 1.19 [1.11-1.27] | 1.18 [1.10-1.26] | 1.07 [1.00-1.14] | 1.05 [0.98-1.14] | 1.18 [0.89-1.56] | 1.15 [1.07-1.23] | 2,416 | 2,378 | 1,853 | 139 | 2,125 |
| F34 | 1.44 [1.24-1.68] | 1.47 [1.26-1.72] | 1.52 [1.27-1.83] | | 1.23 [1.05-1.44] | 1.25 [1.06-1.46] | 1.18 [1.00-1.39] | 1.24 [1.02-1.50] | | 1.13 [0.96-1.32] | 448 | 416 | 299 | NA | 392 |
| F38 | 1.30 [1.04-1.64] | 1.27 [1.00-1.62] | 1.21 [0.92-1.61] | | 1.16 [0.92-1.47] | 1.19 [0.94-1.51] | 1.09 [0.85-1.40] | 1.04 [0.78-1.39] | | 1.10 [0.86-1.41] | 199 | 176 | 126 | NA | 179 |
| F39 | 1.33 [1.12-1.57] | 1.35 [1.13-1.61] | 1.33 [1.08-1.64] | | 1.24 [1.04-1.47] | 1.20 [1.00-1.42] | 1.10 [0.92-1.33] | 1.15 [0.93-1.43] | | 1.17 [0.98-1.40] | 368 | 318 | 237 | NA | 337 |
| F40 | 1.26 [1.21-1.30] | 1.30 [1.25-1.35] | 1.30 [1.24-1.36] | 1.13 [1.05-1.21] | 1.19 [1.14-1.24] | 1.16 [1.12-1.21] | 1.13 [1.09-1.18] | 1.14 [1.08-1.19] | 1.08 [1.01-1.16] | 1.13 [1.09-1.18] | 7,877 | 6,569 | 4,366 | 2,284 | 6,831 |
| F41 | 1.34 [1.32-1.36] | 1.35 [1.33-1.37] | 1.36 [1.34-1.38] | 1.16 [1.13-1.20] | 1.25 [1.23-1.26] | 1.20 [1.19-1.22] | 1.11 [1.10-1.13] | 1.13 [1.12-1.15] | 1.14 [1.10-1.17] | 1.17 [1.16-1.19] | 70,774 | 66,825 | 45,718 | 12,693 | 62,988 |

#### Supplementary Table 3: Hazard ratios and events from all cohorts

| Outcome | Hazard ratio (99% confidence interval): crude: any age | crude: 18+ | crude: 40+ | crude: <18 | crude: hosp. | adjusted: any age | adjusted: 18+ | adjusted: 40+ | adjusted: <18 | adjusted: hosp. | Events (in exposed): any age | 18+ | 40+ | <18 | hosp. |
|---|---|---|---|---|---|---|---|---|---|---|---|---|---|---|---|
| F42 | 1.64 [1.52-1.76] | 1.67 [1.54-1.80] | 1.84 [1.65-2.06] | 1.25 [1.09-1.43] | 1.47 [1.36-1.58] | 1.43 [1.32-1.54] | 1.33 [1.22-1.44] | 1.53 [1.37-1.72] | 1.15 [1.00-1.32] | 1.34 [1.24-1.46] | 2,087 | 1,795 | 888 | 635 | 1,802 |
| F43 | 1.26 [1.20-1.33] | 1.28 [1.21-1.35] | 1.35 [1.26-1.45] | 1.00 [0.90-1.12] | 1.16 [1.10-1.22] | 1.11 [1.05-1.17] | 1.01 [0.95-1.06] | 1.07 [0.99-1.15] | 0.99 [0.89-1.11] | 1.09 [1.04-1.15] | 3,877 | 3,597 | 1,949 | 901 | 3,448 |
| F44 | 1.40 [1.26-1.55] | 1.43 [1.28-1.59] | 1.45 [1.27-1.67] | 1.27 [1.04-1.55] | 1.23 [1.11-1.36] | 1.21 [1.09-1.34] | 1.11 [1.00-1.25] | 1.15 [0.99-1.33] | 1.19 [0.97-1.47] | 1.11 [1.00-1.24] | 1,011 | 905 | 522 | 279 | 924 |
| F45 | 1.45 [1.32-1.59] | 1.40 [1.27-1.54] | 1.37 [1.23-1.53] | 1.27 [1.02-1.60] | 1.29 [1.17-1.42] | 1.28 [1.16-1.41] | 1.15 [1.04-1.27] | 1.16 [1.04-1.30] | 1.16 [0.91-1.46] | 1.17 [1.06-1.29] | 1,216 | 1,079 | 802 | 215 | 1,083 |
| F48 | 1.55 [1.30-1.84] | 1.56 [1.30-1.87] | 1.51 [1.21-1.87] | | 1.24 [1.04-1.48] | 1.35 [1.12-1.62] | 1.27 [1.05-1.54] | 1.20 [0.95-1.51] | | 1.12 [0.93-1.35] | 346 | 306 | 206 | NA | 298 |
| F50 | 1.28 [1.19-1.37] | 1.29 [1.18-1.41] | 1.42 [1.22-1.64] | 1.13 [1.02-1.24] | 1.19 [1.10-1.28] | 1.17 [1.08-1.26] | 1.08 [0.99-1.18] | 1.19 [1.02-1.39] | 1.09 [0.99-1.20] | 1.13 [1.04-1.22] | 2,058 | 1,258 | 443 | 1,189 | 1,711 |
| F51 | 1.43 [1.23-1.67] | 1.57 [1.30-1.89] | 1.50 [1.20-1.88] | 1.33 [1.05-1.68] | 1.37 [1.17-1.60] | 1.30 [1.11-1.52] | 1.32 [1.09-1.60] | 1.27 [1.00-1.60] | 1.31 [1.03-1.67] | 1.27 [1.08-1.50] | 458 | 302 | 201 | 193 | 411 |
| F52 | 1.49 [1.32-1.69] | 1.48 [1.30-1.67] | 1.48 [1.30-1.68] | | 1.32 [1.16-1.50] | 1.35 [1.19-1.53] | 1.25 [1.10-1.42] | 1.26 [1.10-1.44] | | 1.24 [1.09-1.42] | 643 | 635 | 577 | NA | 569 |
| F53 | 1.35 [1.19-1.53] | 1.34 [1.18-1.52] | | 0.96 [0.73-1.26] | 1.20 [1.06-1.36] | 1.24 [1.08-1.41] | 1.14 [0.99-1.30] | | 0.95 [0.71-1.26] | 1.15 [1.01-1.31] | 619 | 613 | NA | 135 | 586 |
| F60 | 1.34 [1.27-1.42] | 1.39 [1.31-1.47] | 1.59 [1.46-1.72] | 1.02 [0.92-1.14] | 1.21 [1.14-1.27] | 1.15 [1.09-1.22] | 1.05 [0.99-1.11] | 1.23 [1.13-1.35] | 1.02 [0.91-1.14] | 1.13 [1.07-1.20] | 3,390 | 3,274 | 1,412 | 915 | 3,113 |
| F61 | 1.13 [0.85-1.51] | 1.17 [0.88-1.56] | | | | 0.99 [0.73-1.35] | 0.97 [0.71-1.32] | | | | 112 | 111 | NA | NA | NA |
| F69 | 1.47 [1.22-1.77] | 1.62 [1.31-1.99] | 1.81 [1.39-2.36] | | 1.39 [1.15-1.68] | 1.27 [1.05-1.55] | 1.27 [1.01-1.58] | 1.49 [1.13-1.97] | | 1.29 [1.06-1.57] | 302 | 234 | 147 | NA | 275 |
| F70 | 1.72 [1.51-1.97] | 1.87 [1.62-2.16] | 2.17 [1.80-2.61] | 1.19 [0.94-1.51] | 1.50 [1.30-1.72] | 1.54 [1.34-1.76] | 1.65 [1.42-1.92] | 1.81 [1.49-2.20] | 1.14 [0.89-1.46] | 1.39 [1.20-1.61] | 625 | 518 | 317 | 183 | 533 |
| F71 | 2.16 [1.76-2.65] | 2.40 [1.92-3.00] | 2.90 [2.15-3.91] | | 1.73 [1.39-2.15] | 1.93 [1.56-2.39] | 2.23 [1.77-2.82] | 2.62 [1.92-3.56] | | 1.67 [1.34-2.08] | 280 | 231 | 137 | NA | 237 |
| F72 | 1.67 [1.43-1.94] | 2.15 [1.80-2.57] | 2.79 [2.21-3.52] | 1.03 [0.82-1.29] | 1.42 [1.21-1.66] | 1.51 [1.29-1.76] | 2.09 [1.74-2.52] | 2.58 [2.02-3.29] | 1.01 [0.80-1.29] | 1.34 [1.14-1.57] | 485 | 355 | 219 | 197 | 439 |
| F79 | 1.56 [1.47-1.66] | 1.89 [1.76-2.04] | 2.13 [1.94-2.33] | 1.06 [0.96-1.16] | 1.38 [1.30-1.48] | 1.44 [1.35-1.54] | 1.74 [1.61-1.88] | 1.93 [1.76-2.12] | 1.03 [0.93-1.13] | 1.31 [1.23-1.40] | 2,891 | 2,009 | 1,368 | 1,151 | 2,559 |
| F80 | 1.22 [1.15-1.29] | 1.38 [1.22-1.56] | 1.41 [1.24-1.61] | 1.16 [1.09-1.24] | 1.14 [1.08-1.22] | 1.16 [1.10-1.24] | 1.29 [1.13-1.46] | 1.31 [1.14-1.49] | 1.11 [1.04-1.19] | 1.12 [1.05-1.19] | 2,868 | 666 | 603 | 2,234 | 2,631 |
| F81 | 1.56 [1.50-1.61] | 1.92 [1.83-2.00] | 2.15 [2.03-2.27] | 1.13 [1.07-1.19] | 1.36 [1.31-1.42] | 1.44 [1.39-1.50] | 1.79 [1.71-1.88] | 1.99 [1.88-2.11] | 1.08 [1.03-1.14] | 1.29 [1.24-1.35] | 8,353 | 5,304 | 3,443 | 3,825 | 6,839 |
| F82 | 1.19 [1.02-1.39] | | | 1.12 [0.95-1.33] | 1.06 [0.90-1.25] | 1.11 [0.95-1.31] | | | 1.07 [0.90-1.26] | 1.02 [0.86-1.20] | 403 | NA | NA | 372 | 363 |
| F83 | 0.95 [0.80-1.13] | | | 0.91 [0.76-1.09] | 0.88 [0.74-1.06] | 0.89 [0.74-1.06] | | | 0.88 [0.73-1.05] | 0.84 [0.70-1.01] | 326 | NA | NA | 311 | 311 |
| F84 | 1.32 [1.28-1.38] | 1.54 [1.44-1.65] | 2.15 [1.90-2.44] | 1.22 [1.17-1.27] | 1.23 [1.18-1.28] | 1.18 [1.14-1.23] | 1.34 [1.24-1.44] | 1.73 [1.51-1.97] | 1.15 [1.11-1.20] | 1.14 [1.09-1.19] | 7,896 | 2,095 | 695 | 6,653 | 6,860 |
| F89 | 1.02 [0.95-1.10] | 1.53 [1.20-1.96] | | 0.97 [0.90-1.05] | 0.94 [0.87-1.01] | 0.94 [0.87-1.02] | 1.33 [1.02-1.73] | | 0.92 [0.85-1.00] | 0.88 [0.82-0.96] | 1,669 | 166 | NA | 1,560 | 1,570 |
| F90 | 1.22 [1.15-1.28] | 1.23 [1.12-1.35] | 1.75 [1.34-2.29] | 1.16 [1.09-1.22] | 1.14 [1.08-1.21] | 1.12 [1.06-1.19] | 1.09 [0.99-1.21] | 1.26 [0.94-1.68] | 1.12 [1.06-1.19] | 1.11 [1.04-1.17] | 3,883 | 1,020 | 139 | 3,444 | 3,393 |
| F91 | 1.27 [1.13-1.42] | 1.19 [0.93-1.52] | 1.09 [0.81-1.47] | 1.30 [1.15-1.48] | 1.16 [1.03-1.31] | 1.19 [1.06-1.34] | 1.08 [0.84-1.39] | 1.01 [0.74-1.37] | 1.24 [1.09-1.41] | 1.12 [0.99-1.26] | 820 | 160 | 106 | 694 | 709 |
| F94 | 1.22 [0.94-1.58] | | | 1.19 [0.90-1.56] | 1.07 [0.81-1.43] | 1.15 [0.88-1.51] | | | 1.14 [0.86-1.51] | 1.04 [0.77-1.39] | 161 | NA | NA | 148 | 136 |
| F95 | 1.38 [1.18-1.61] | 1.52 [1.16-1.98] | | 1.30 [1.10-1.55] | 1.27 [1.08-1.50] | 1.23 [1.05-1.45] | 1.23 [0.93-1.64] | | 1.22 [1.02-1.45] | 1.20 [1.01-1.42] | 474 | 137 | NA | 380 | 414 |
| F98 | 1.32 [1.15-1.52] | 1.26 [0.95-1.66] | | 1.30 [1.12-1.51] | 1.22 [1.05-1.41] | 1.22 [1.06-1.41] | 1.08 [0.81-1.44] | | 1.25 [1.08-1.46] | 1.15 [0.99-1.34] | 567 | 128 | NA | 495 | 513 |
| F99 | 1.24 [1.14-1.36] | 1.27 [1.16-1.39] | 1.30 [1.16-1.46] | 1.10 [0.91-1.32] | 1.12 [1.02-1.23] | 1.16 [1.06-1.27] | 1.08 [0.99-1.19] | 1.12 [0.99-1.26] | 1.09 [0.90-1.33] | 1.09 [0.99-1.19] | 1,303 | 1,179 | 715 | 299 | 1,144 |
| Diseases of the nervous system (G00-G99) | | | | | | | | | | | | | | | |
| G00 | 1.12 [0.95-1.31] | 1.20 [1.00-1.44] | 1.25 [1.01-1.55] | 0.92 [0.69-1.24] | 1.02 [0.85-1.22] | 1.07 [0.90-1.26] | 1.12 [0.92-1.35] | 1.20 [0.96-1.49] | 0.90 [0.66-1.21] | 0.98 [0.81-1.18] | 351 | 275 | 200 | 108 | 283 |
| G01 | 1.11 [0.89-1.39] | | | 1.09 [0.82-1.45] | 1.10 [0.85-1.41] | 1.06 [0.84-1.33] | | | 1.11 [0.83-1.49] | 1.08 [0.84-1.39] | 193 | NA | NA | 116 | 157 |
| G02 | 1.08 [0.85-1.39] | 1.19 [0.90-1.56] | | | 1.05 [0.81-1.37] | 0.98 [0.76-1.27] | 1.04 [0.78-1.39] | | | 1.00 [0.76-1.30] | 165 | 130 | NA | NA | 143 |
| G03 | 1.24 [1.07-1.43] | 1.17 [0.98-1.39] | 1.13 [0.92-1.40] | 1.32 [1.06-1.65] | 1.05 [0.90-1.23] | 1.15 [1.00-1.34] | 1.03 [0.86-1.24] | 1.00 [0.80-1.24] | 1.27 [1.01-1.60] | 1.02 [0.87-1.19] | 464 | 307 | 200 | 205 | 396 |
| G04 | 1.25 [1.13-1.38] | 1.28 [1.15-1.43] | 1.23 [1.09-1.39] | 1.10 [0.91-1.33] | 1.15 [1.04-1.28] | 1.19 [1.08-1.32] | 1.17 [1.04-1.30] | 1.12 [0.99-1.27] | 1.08 [0.89-1.31] | 1.12 [1.01-1.24] | 1,050 | 822 | 674 | 279 | 908 |
| G05 | 1.42 [1.17-1.71] | 1.55 [1.26-1.91] | 1.60 [1.28-2.00] | | 1.44 [1.18-1.77] | 1.37 [1.14-1.66] | 1.46 [1.18-1.81] | 1.48 [1.18-1.86] | | 1.38 [1.12-1.70] | 290 | 232 | 204 | NA | 251 |
| G06 | 1.25 [1.08-1.45] | 1.20 [1.02-1.41] | 1.18 [1.00-1.41] | | 1.20 [1.03-1.41] | 1.20 [1.03-1.40] | 1.14 [0.97-1.34] | 1.11 [0.93-1.33] | | 1.20 [1.02-1.41] | 446 | 365 | 319 | NA | 385 |
| G08 | 1.16 [0.95-1.42] | 1.32 [1.06-1.65] | 1.14 [0.86-1.51] | | 1.29 [1.04-1.60] | 1.12 [0.92-1.38] | 1.26 [1.00-1.58] | 1.06 [0.79-1.42] | | 1.29 [1.03-1.61] | 239 | 199 | 116 | NA | 206 |
| G09 | 1.36 [1.14-1.63] | 1.38 [1.12-1.70] | 1.24 [0.98-1.56] | | 1.17 [0.98-1.41] | 1.28 [1.06-1.54] | 1.27 [1.03-1.57] | 1.11 [0.87-1.41] | | 1.14 [0.95-1.38] | 311 | 232 | 176 | NA | 280 |
| G10 | 1.40 [1.07-1.84] | 1.33 [1.02-1.74] | 1.26 [0.94-1.67] | | 1.25 [0.92-1.69] | 1.42 [1.08-1.87] | 1.39 [1.05-1.83] | 1.29 [0.97-1.73] | | 1.27 [0.93-1.73] | 135 | 132 | 113 | NA | 101 |
| G11 | 1.17 [1.01-1.36] | 1.25 [1.07-1.46] | 1.20 [1.02-1.43] | | 1.15 [0.99-1.34] | 1.13 [0.97-1.31] | 1.16 [0.99-1.36] | 1.10 [0.93-1.31] | | 1.14 [0.97-1.33] | 454 | 380 | 328 | NA | 400 |
| G12 | 1.08 [0.97-1.20] | 1.12 [1.00-1.24] | 1.10 [0.99-1.22] | | 1.06 [0.95-1.19] | 1.04 [0.93-1.15] | 1.07 [0.96-1.20] | 1.06 [0.95-1.18] | | 1.05 [0.94-1.18] | 857 | 824 | 796 | NA | 732 |
| G20 | 1.06 [1.02-1.09] | 1.05 [1.01-1.09] | 1.06 [1.02-1.10] | | 1.03 [0.99-1.07] | 1.05 [1.01-1.09] | 1.02 [0.99-1.06] | 1.03 [0.99-1.07] | | 1.03 [0.99-1.07] | 7,432 | 7,435 | 7,409 | NA | 6,407 |
| G21 | 1.34 [1.19-1.50] | 1.27 [1.13-1.43] | 1.34 [1.19-1.51] | | 1.22 [1.08-1.38] | 1.31 [1.17-1.48] | 1.20 [1.06-1.35] | 1.28 [1.13-1.44] | | 1.24 [1.10-1.40] | 774 | 773 | 752 | NA | 736 |
| G23 | 1.17 [1.00-1.36] | 1.12 [0.96-1.30] | 1.08 [0.93-1.27] | | 1.02 [0.87-1.19] | 1.14 [0.97-1.33] | 1.07 [0.92-1.25] | 1.03 [0.88-1.21] | | 1.01 [0.86-1.18] | 428 | 418 | 405 | NA | 393 |
| G24 | 1.14 [1.03-1.25] | 1.20 [1.08-1.34] | 1.21 [1.08-1.37] | 0.82 [0.67-1.00] | 1.04 [0.94-1.15] | 1.09 [0.98-1.20] | 1.10 [0.98-1.23] | 1.11 [0.99-1.26] | 0.79 [0.64-0.96] | 1.02 [0.92-1.13] | 1,028 | 821 | 698 | 250 | 933 |
| G25 | 1.28 [1.23-1.34] | 1.30 [1.24-1.36] | 1.28 [1.22-1.35] | 1.07 [0.91-1.27] | 1.20 [1.15-1.26] | 1.16 [1.11-1.22] | 1.12 [1.07-1.17] | 1.11 [1.06-1.17] | 1.04 [0.87-1.24] | 1.13 [1.07-1.18] | 4,923 | 4,667 | 4,349 | 353 | 4,613 |
| G30 | 1.07 [1.04-1.10] | 1.07 [1.04-1.10] | 1.08 [1.05-1.11] | | 1.06 [1.03-1.09] | 1.06 [1.03-1.09] | 1.05 [1.02-1.08] | 1.06 [1.03-1.09] | | 1.05 [1.02-1.09] | 13,635 | 13,626 | 13,622 | NA | 12,389 |
| G31 | 1.17 [1.13-1.21] | 1.18 [1.14-1.22] | 1.17 [1.13-1.20] | 1.04 [0.78-1.37] | 1.13 [1.10-1.17] | 1.14 [1.10-1.18] | 1.12 [1.08-1.16] | 1.10 [1.07-1.14] | 1.01 [0.76-1.35] | 1.13 [1.09-1.17] | 10,278 | 10,176 | 10,013 | 126 | 9,507 |
| G32 | 1.39 [1.10-1.76] | 1.29 [1.02-1.63] | 1.47 [1.15-1.89] | | 1.29 [1.01-1.64] | 1.30 [1.02-1.66] | 1.12 [0.88-1.43] | 1.30 [1.01-1.68] | | 1.29 [1.01-1.65] | 174 | 171 | 160 | NA | 161 |
| G35 | 1.23 [1.15-1.31] | 1.22 [1.15-1.30] | 1.23 [1.14-1.32] | 1.14 [0.85-1.53] | 1.14 [1.06-1.22] | 1.16 [1.08-1.24] | 1.08 [1.01-1.15] | 1.09 [1.01-1.18] | 1.15 [0.85-1.55] | 1.12 [1.04-1.21] | 2,212 | 2,199 | 1,684 | 125 | 1,677 |
| G37 | 1.28 [1.14-1.45] | 1.30 [1.14-1.47] | 1.36 [1.17-1.59] | 1.04 [0.77-1.40] | 1.22 [1.07-1.40] | 1.22 [1.07-1.38] | 1.16 [1.02-1.33] | 1.24 [1.06-1.46] | 1.02 [0.75-1.39] | 1.19 [1.04-1.36] | 646 | 580 | 389 | 113 | 544 |
| G40 | 1.31 [1.28-1.34] | 1.36 [1.32-1.40] | 1.38 [1.34-1.42] | 1.08 [1.02-1.13] | 1.19 [1.16-1.22] | 1.21 [1.18-1.24] | 1.19 [1.15-1.22] | 1.20 [1.16-1.23] | 1.06 [1.00-1.11] | 1.14 [1.11-1.17] | 17,275 | 14,171 | 11,140 | 4,096 | 14,157 |
| G41 | 1.20 [1.10-1.31] | 1.33 [1.20-1.48] | 1.29 [1.15-1.46] | 0.96 [0.84-1.11] | 1.09 [1.00-1.20] | 1.13 [1.04-1.24] | 1.20 [1.07-1.34] | 1.15 [1.02-1.30] | 0.95 [0.83-1.10] | 1.07 [0.98-1.18] | 1,276 | 866 | 693 | 479 | 1,177 |
| G43 | 1.32 [1.28-1.35] | 1.32 [1.29-1.36] | 1.33 [1.29-1.38] | 1.20 [1.14-1.26] | 1.21 [1.18-1.25] | 1.16 [1.13-1.19] | 1.08 [1.05-1.11] | 1.10 [1.06-1.14] | 1.13 [1.07-1.20] | 1.12 [1.09-1.15] | 16,417 | 14,623 | 8,600 | 3,948 | 14,478 |
| G44 | 1.33 [1.24-1.42] | 1.31 [1.23-1.40] | 1.37 [1.26-1.49] | 1.22 [1.05-1.42] | 1.23 [1.15-1.32] | 1.20 [1.12-1.28] | 1.12 [1.04-1.20] | 1.15 [1.06-1.26] | 1.19 [1.02-1.39] | 1.15 [1.07-1.23] | 2,317 | 2,109 | 1,353 | 470 | 2,084 |
| G45 | 1.18 [1.15-1.22] | 1.18 [1.15-1.22] | 1.18 [1.14-1.22] | | 1.12 [1.08-1.15] | 1.14 [1.10-1.17] | 1.10 [1.07-1.14] | 1.10 [1.06-1.13] | | 1.09 [1.06-1.13] | 9,727 | 9,696 | 9,541 | NA | 8,585 |
| G46 | 1.22 [1.07-1.38] | 1.18 [1.04-1.34] | 1.19 [1.05-1.36] | | 1.15 [1.01-1.30] | 1.19 [1.04-1.35] | 1.12 [0.98-1.27] | 1.12 [0.98-1.28] | | 1.13 [0.99-1.29] | 647 | 644 | 635 | NA | 596 |
| G47 | 1.48 [1.45-1.51] | 1.58 [1.54-1.62] | 1.58 [1.54-1.63] | 1.31 [1.27-1.36] | 1.38 [1.35-1.41] | 1.32 [1.30-1.35] | 1.28 [1.24-1.31] | 1.27 [1.24-1.31] | 1.25 [1.21-1.30] | 1.27 [1.24-1.30] | 23,676 | 15,345 | 13,168 | 8,935 | 21,280 |
| G50 | 1.42 [1.32-1.53] | 1.43 [1.33-1.55] | 1.44 [1.33-1.56] | | 1.32 [1.22-1.42] | 1.29 [1.20-1.39] | 1.22 [1.13-1.32] | 1.24 [1.15-1.35] | | 1.22 [1.13-1.32] | 1,837 | 1,821 | 1,646 | NA | 1,651 |
| G51 | 1.21 [1.15-1.28] | 1.24 [1.17-1.31] | 1.21 [1.14-1.29] | 1.09 [0.95-1.24] | 1.15 [1.09-1.21] | 1.15 [1.09-1.21] | 1.13 [1.06-1.19] | 1.10 [1.04-1.17] | 1.09 [0.95-1.24] | 1.11 [1.05-1.17] | 3,683 | 3,203 | 2,781 | 595 | 3,191 |
| G52 | 1.23 [1.02-1.49] | 1.32 [1.08-1.60] | 1.28 [1.04-1.57] | | 1.26 [1.03-1.53] | 1.17 [0.97-1.42] | 1.18 [0.97-1.45] | 1.15 [0.93-1.43] | | 1.20 [0.98-1.46] | 276 | 259 | 230 | NA | 254 |
| G53 | 1.33 [1.17-1.52] | 1.32 [1.15-1.51] | 1.35 [1.18-1.55] | | 1.26 [1.10-1.44] | 1.26 [1.10-1.44] | 1.21 [1.06-1.39] | 1.25 [1.09-1.44] | | 1.21 [1.05-1.39] | 615 | 595 | 570 | NA | 564 |
| G54 | 1.33 [1.21-1.46] | 1.29 [1.17-1.42] | 1.35 [1.22-1.50] | | 1.24 [1.13-1.37] | 1.21 [1.10-1.33] | 1.10 [1.00-1.22] | 1.16 [1.04-1.29] | | 1.18 [1.07-1.30] | 1,109 | 1,087 | 912 | NA | 1,017 |
| G55 | 1.27 [1.24-1.31] | 1.27 [1.23-1.31] | 1.28 [1.24-1.32] | 1.12 [0.96-1.30] | 1.18 [1.14-1.21] | 1.17 [1.14-1.21] | 1.11 [1.07-1.14] | 1.11 [1.08-1.15] | 1.06 [0.90-1.23] | 1.11 [1.08-1.15] | 11,907 | 11,866 | 9,868 | 458 | 10,751 |
| G56 | 1.35 [1.32-1.38] | 1.34 [1.31-1.37] | 1.33 [1.30-1.36] | 1.23 [1.06-1.44] | 1.24 [1.21-1.27] | 1.24 [1.21-1.27] | 1.18 [1.15-1.21] | 1.17 [1.14-1.20] | 1.16 [0.98-1.36] | 1.16 [1.13-1.19] | 18,507 | 18,382 | 16,157 | 428 | 15,834 |
| G57 | 1.39 [1.30-1.48] | 1.40 [1.31-1.50] | 1.35 [1.26-1.45] | | 1.25 [1.17-1.34] | 1.28 [1.19-1.37] | 1.25 [1.17-1.34] | 1.19 [1.11-1.28] | | 1.17 [1.09-1.26] | 2,204 | 2,164 | 1,841 | NA | 1,944 |
| G58 | 1.32 [1.20-1.45] | 1.40 [1.27-1.54] | 1.41 [1.27-1.56] | | 1.24 [1.12-1.36] | 1.20 [1.09-1.32] | 1.21 [1.09-1.33] | 1.22 [1.10-1.36] | | 1.16 [1.05-1.28] | 1,125 | 1,087 | 932 | NA | 1,032 |
| G59 | 1.49 [1.34-1.65] | 1.47 [1.33-1.64] | 1.49 [1.34-1.66] | | 1.27 [1.14-1.41] | 1.31 [1.18-1.47] | 1.18 [1.05-1.32] | 1.18 [1.05-1.32] | | 1.20 [1.07-1.33] | 888 | 886 | 856 | NA | 843 |
| G60 | 1.28 [1.11-1.46] | 1.26 [1.10-1.46] | 1.26 [1.09-1.47] | | 1.17 [1.01-1.35] | 1.17 [1.02-1.34] | 1.12 [0.97-1.30] | 1.11 [0.95-1.30] | | 1.07 [0.93-1.24] | 529 | 483 | 419 | NA | 473 |
| G61 | 1.25 [1.11-1.41] | 1.26 [1.11-1.43] | 1.24 [1.08-1.43] | | 1.14 [0.99-1.30] | 1.21 [1.07-1.37] | 1.19 [1.05-1.36] | 1.17 [1.02-1.35] | | 1.10 [0.96-1.26] | 627 | 574 | 494 | NA | 517 |
| G62 | 1.35 [1.30-1.40] | 1.36 [1.30-1.41] | 1.35 [1.30-1.40] | 0.97 [0.78-1.22] | 1.23 [1.19-1.28] | 1.23 [1.18-1.28] | 1.16 [1.12-1.21] | 1.15 [1.11-1.20] | 0.94 [0.74-1.19] | 1.17 [1.12-1.22] | 6,954 | 6,849 | 6,481 | 198 | 6,516 |
| G63 | 1.45 [1.38-1.53] | 1.44 [1.37-1.52] | 1.46 [1.38-1.54] | | 1.32 [1.26-1.39] | 1.30 [1.23-1.37] | 1.16 [1.10-1.23] | 1.17 [1.11-1.24] | | 1.24 [1.18-1.31] | 3,841 | 3,821 | 3,700 | NA | 3,718 |
| G70 | 1.12 [0.99-1.28] | 1.17 [1.02-1.33] | 1.17 [1.02-1.35] | | 1.14 [0.99-1.30] | 1.07 [0.94-1.22] | 1.09 [0.95-1.25] | 1.09 [0.94-1.26] | | 1.10 [0.95-1.26] | 582 | 535 | 478 | NA | 511 |
| G71 | 1.18 [1.04-1.35] | 1.22 [1.04-1.43] | 1.14 [0.95-1.36] | 1.00 [0.81-1.25] | 1.04 [0.90-1.20] | 1.15 [1.00-1.31] | 1.16 [0.98-1.36] | 1.08 [0.89-1.30] | 1.01 [0.81-1.26] | 1.03 [0.89-1.19] | 555 | 387 | 271 | 201 | 459 |
| G72 | 1.41 [1.26-1.58] | 1.42 [1.27-1.60] | 1.41 [1.25-1.59] | | 1.28 [1.14-1.43] | 1.32 [1.18-1.48] | 1.28 [1.13-1.44] | 1.29 [1.14-1.46] | | 1.23 [1.09-1.38] | 815 | 748 | 682 | NA | 743 |
| G80 | 1.01 [0.94-1.09] | 1.39 [1.25-1.55] | 1.66 [1.46-1.90] | 0.79 [0.72-0.87] | 0.92 [0.85-1.00] | 0.96 [0.89-1.04] | 1.33 [1.19-1.49] | 1.59 [1.38-1.82] | 0.77 [0.70-0.85] | 0.90 [0.83-0.97] | 1,750 | 801 | 552 | 1,019 | 1,431 |
| G81 | 1.17 [1.14-1.20] | 1.19 [1.15-1.23] | 1.17 [1.14-1.21] | 0.98 [0.86-1.13] | 1.12 [1.08-1.15] | 1.14 [1.11-1.18] | 1.12 [1.08-1.15] | 1.10 [1.07-1.13] | 0.98 [0.85-1.12] | 1.11 [1.08-1.15] | 11,632 | 11,192 | 10,789 | 541 | 10,542 |
| G82 | 1.15 [1.05-1.25] | 1.22 [1.11-1.34] | 1.21 [1.10-1.34] | 0.75 [0.61-0.93] | 1.04 [0.95-1.14] | 1.10 [1.01-1.20] | 1.14 [1.04-1.25] | 1.13 [1.03-1.26] | 0.73 [0.59-0.91] | 1.03 [0.94-1.13] | 1,261 | 1,119 | 951 | 198 | 1,098 |
| G83 | 1.21 [1.12-1.30] | 1.20 [1.12-1.30] | 1.23 [1.13-1.33] | 0.97 [0.76-1.24] | 1.09 [1.01-1.18] | 1.15 [1.06-1.23] | 1.09 [1.01-1.17] | 1.12 [1.03-1.21] | 0.95 [0.74-1.22] | 1.06 [0.98-1.15] | 1,842 | 1,735 | 1,491 | 165 | 1,652 |
| G90 | 1.26 [1.15-1.40] | 1.23 [1.11-1.36] | 1.22 [1.08-1.36] | 1.01 [0.77-1.33] | 1.13 [1.02-1.25] | 1.17 [1.05-1.29] | 1.07 [0.97-1.19] | 1.08 [0.96-1.22] | 0.94 [0.71-1.25] | 1.06 [0.95-1.18] | 1,018 | 945 | 747 | 143 | 926 |
| G91 | 1.17 [1.10-1.25] | 1.21 [1.13-1.30] | 1.17 [1.09-1.26] | 0.97 [0.82-1.14] | 1.14 [1.06-1.22] | 1.15 [1.08-1.23] | 1.16 [1.08-1.25] | 1.13 [1.04-1.21] | 0.97 [0.82-1.14] | 1.14 [1.06-1.22] | 2,341 | 2,035 | 1,777 | 376 | 2,007 |
| G93 | 1.26 [1.22-1.30] | 1.27 [1.23-1.31] | 1.25 [1.21-1.30] | 1.13 [1.05-1.22] | 1.17 [1.14-1.21] | 1.17 [1.13-1.21] | 1.12 [1.08-1.16] | 1.11 [1.07-1.16] | 1.08 [1.01-1.17] | 1.12 [1.08-1.16] | 10,133 | 8,873 | 6,837 | 2,055 | 9,031 |
| G94 | 1.18 [1.00-1.39] | 1.25 [1.04-1.50] | 1.19 [0.98-1.45] | | 1.11 [0.93-1.32] | 1.17 [0.99-1.38] | 1.22 [1.01-1.47] | 1.16 [0.95-1.42] | | 1.13 [0.94-1.36] | 361 | 272 | 245 | NA | 295 |
| G95 | 1.19 [1.12-1.27] | 1.22 [1.14-1.29] | 1.22 [1.14-1.31] | 1.08 [0.87-1.35] | 1.11 [1.04-1.19] | 1.14 [1.07-1.21] | 1.13 [1.06-1.21] | 1.14 [1.06-1.22] | 1.02 [0.82-1.28] | 1.09 [1.02-1.17] | 2,508 | 2,358 | 2,173 | 207 | 2,215 |
| G96 | 1.30 [1.17-1.44] | 1.35 [1.20-1.50] | 1.37 [1.21-1.55] | 1.03 [0.79-1.33] | 1.20 [1.08-1.34] | 1.20 [1.07-1.34] | 1.18 [1.05-1.32] | 1.21 [1.06-1.37] | 0.97 [0.74-1.26] | 1.15 [1.03-1.29] | 875 | 780 | 621 | 145 | 781 |
| G97 | 1.22 [1.09-1.36] | 1.26 [1.12-1.42] | 1.24 [1.06-1.45] | 1.12 [0.91-1.39] | 1.11 [0.99-1.24] | 1.11 [0.99-1.24] | 1.09 [0.97-1.23] | 1.09 [0.93-1.28] | 1.09 [0.88-1.35] | 1.05 [0.94-1.19] | 818 | 695 | 378 | 237 | 726 |
| G98 | 1.49 [1.25-1.79] | 1.46 [1.20-1.76] | 1.53 [1.24-1.88] | | 1.41 [1.18-1.69] | 1.33 [1.10-1.60] | 1.18 [0.97-1.45] | 1.22 [0.98-1.52] | | 1.29 [1.07-1.55] | 323 | 279 | 243 | NA | 314 |
| G99 | 1.26 [1.20-1.33] | 1.28 [1.21-1.34] | 1.26 [1.19-1.32] | | 1.19 [1.13-1.26] | 1.18 [1.12-1.24] | 1.13 [1.08-1.19] | 1.12 [1.06-1.18] | | 1.15 [1.09-1.21] | 3,910 | 3,886 | 3,580 | NA | 3,567 |
| Diseases of the eye and adnexa (H00-H59) | | | | | | | | | | | | | | | |
| H00 | 1.25 [1.19-1.31] | 1.27 [1.19-1.35] | 1.25 [1.15-1.34] | 1.20 [1.11-1.29] | 1.17 [1.11-1.24] | 1.20 [1.14-1.27] | 1.22 [1.14-1.29] | 1.18 [1.10-1.28] | 1.17 [1.09-1.26] | 1.14 [1.08-1.21] | 3,999 | 2,448 | 1,574 | 1,871 | 2,955 |
| H01 | 1.42 [1.33-1.51] | 1.39 [1.30-1.48] | 1.40 [1.31-1.50] | 1.49 [1.23-1.81] | 1.32 [1.24-1.41] | 1.33 [1.25-1.42] | 1.29 [1.21-1.38] | 1.31 [1.22-1.40] | 1.39 [1.14-1.70] | 1.26 [1.18-1.34] | 2,757 | 2,521 | 2,368 | 291 | 2,435 |
| H02 | 1.31 [1.27-1.34] | 1.32 [1.28-1.35] | 1.31 [1.28-1.35] | 1.22 [1.10-1.35] | 1.22 [1.19-1.26] | 1.24 [1.21-1.28] | 1.22 [1.19-1.26] | 1.22 [1.19-1.26] | 1.18 [1.06-1.31] | 1.17 [1.14-1.21] | 13,523 | 12,689 | 11,932 | 1,036 | 11,492 |
| H03 | 1.61 [1.30-1.98] | 1.41 [1.07-1.84] | 1.30 [0.96-1.76] | 1.74 [1.27-2.38] | 1.34 [1.06-1.68] | 1.50 [1.20-1.86] | 1.25 [0.95-1.65] | 1.18 [0.87-1.61] | 1.60 [1.16-2.21] | 1.23 [0.97-1.57] | 247 | 137 | 107 | 122 | 202 |
| H04 | 1.40 [1.35-1.45] | 1.44 [1.38-1.50] | 1.44 [1.39-1.51] | 1.20 [1.09-1.32] | 1.31 [1.26-1.37] | 1.30 [1.25-1.35] | 1.32 [1.27-1.37] | 1.32 [1.26-1.37] | 1.15 [1.04-1.26] | 1.24 [1.19-1.29] | 7,312 | 6,384 | 6,029 | 1,027 | 6,318 |
| H05 | 1.40 [1.32-1.49] | 1.43 [1.30-1.57] | 1.47 [1.33-1.63] | 1.33 [1.23-1.44] | 1.32 [1.24-1.41] | 1.36 [1.28-1.44] | 1.31 [1.19-1.45] | 1.35 [1.21-1.50] | 1.29 [1.19-1.39] | 1.29 [1.21-1.38] | 2,893 | 1,170 | 926 | 1,807 | 2,409 |
| H06 | 1.49 [1.23-1.79] | 1.45 [1.21-1.75] | 1.47 [1.20-1.80] | | 1.37 [1.13-1.67] | 1.43 [1.18-1.73] | 1.28 [1.05-1.55] | 1.36 [1.11-1.68] | | 1.34 [1.10-1.63] | 292 | 288 | 242 | NA | 250 |
| H10 | 1.43 [1.36-1.50] | 1.43 [1.34-1.52] | 1.35 [1.26-1.45] | 1.42 [1.32-1.54] | 1.35 [1.28-1.42] | 1.34 [1.27-1.41] | 1.29 [1.21-1.38] | 1.24 [1.16-1.33] | 1.32 [1.22-1.43] | 1.28 [1.22-1.35] | 4,406 | 2,770 | 2,435 | 1,760 | 3,999 |
| H11 | 1.24 [1.16-1.31] | 1.24 [1.16-1.32] | 1.27 [1.19-1.36] | 1.10 [0.91-1.32] | 1.17 [1.10-1.25] | 1.19 [1.12-1.26] | 1.18 [1.11-1.26] | 1.21 [1.13-1.29] | 1.07 [0.88-1.30] | 1.15 [1.08-1.23] | 2,738 | 2,541 | 2,263 | 289 | 2,305 |
| H13 | 1.47 [1.20-1.80] | 1.59 [1.24-2.03] | 1.78 [1.35-2.34] | | 1.36 [1.10-1.69] | 1.37 [1.11-1.69] | 1.50 [1.16-1.93] | 1.63 [1.23-2.16] | | 1.29 [1.03-1.61] | 252 | 172 | 143 | NA | 216 |
| H15 | 1.41 [1.17-1.71] | 1.45 [1.19-1.77] | 1.30 [1.04-1.62] | | 1.32 [1.08-1.62] | 1.31 [1.08-1.60] | 1.28 [1.05-1.58] | 1.18 [0.94-1.48] | | 1.24 [1.01-1.53] | 277 | 260 | 208 | NA | 241 |
| H16 | 1.71 [1.57-1.86] | 1.64 [1.50-1.80] | 1.58 [1.44-1.75] | 1.92 [1.54-2.41] | 1.59 [1.45-1.74] | 1.60 [1.47-1.75] | 1.52 [1.39-1.67] | 1.48 [1.34-1.64] | 1.75 [1.39-2.21] | 1.53 [1.39-1.68] | 1,515 | 1,344 | 1,111 | 243 | 1,297 |
| H17 | 1.41 [1.26-1.59] | 1.40 [1.24-1.57] | 1.37 [1.21-1.56] | | 1.39 [1.23-1.57] | 1.34 [1.19-1.51] | 1.30 [1.15-1.48] | 1.28 [1.12-1.46] | | 1.33 [1.18-1.50] | 774 | 722 | 635 | NA | 694 |
| H18 | 1.38 [1.32-1.44] | 1.36 [1.30-1.43] | 1.24 [1.18-1.31] | 1.91 [1.67-2.18] | 1.29 [1.23-1.36] | 1.31 [1.25-1.37] | 1.27 [1.22-1.34] | 1.19 [1.13-1.25] | 1.70 [1.48-1.95] | 1.24 [1.18-1.30] | 5,208 | 4,964 | 3,982 | 735 | 4,460 |
| H19 | 1.81 [1.53-2.14] | 1.85 [1.55-2.19] | 1.77 [1.47-2.14] | | 1.67 [1.40-1.99] | 1.60 [1.34-1.90] | 1.59 [1.33-1.90] | 1.54 [1.27-1.88] | | 1.51 [1.26-1.81] | 401 | 366 | 307 | NA | 339 |
| H20 | 1.43 [1.32-1.56] | 1.38 [1.26-1.50] | 1.38 [1.25-1.51] | 1.40 [1.12-1.74] | 1.38 [1.26-1.50] | 1.38 [1.27-1.50] | 1.29 [1.18-1.41] | 1.30 [1.18-1.43] | 1.35 [1.08-1.70] | 1.34 [1.23-1.47] | 1,446 | 1,262 | 1,077 | 233 | 1,273 |
| H21 | 1.18 [1.10-1.27] | 1.18 [1.09-1.26] | 1.15 [1.07-1.24] | | 1.15 [1.06-1.24] | 1.14 [1.06-1.23] | 1.12 [1.04-1.21] | 1.09 [1.01-1.18] | | 1.12 [1.04-1.21] | 1,915 | 1,866 | 1,792 | NA | 1,746 |
| H22 | 1.46 [1.07-1.99] | 1.45 [1.06-1.98] | | | 1.36 [0.99-1.87] | 1.40 [1.02-1.91] | 1.28 [0.92-1.77] | | | 1.33 [0.96-1.84] | 112 | 103 | NA | NA | 102 |
| H25 | 1.17 [1.15-1.19] | 1.17 [1.16-1.19] | 1.18 [1.17-1.20] | | 1.12 [1.11-1.14] | 1.13 [1.12-1.15] | 1.12 [1.10-1.13] | 1.13 [1.11-1.14] | | 1.10 [1.08-1.12] | 46,074 | 46,065 | 45,897 | NA | 39,385 |
| H26 | 1.21 [1.19-1.23] | 1.21 [1.19-1.22] | 1.21 [1.19-1.22] | 1.60 [1.36-1.87] | 1.16 [1.14-1.17] | 1.17 [1.15-1.18] | 1.14 [1.13-1.16] | 1.14 [1.13-1.16] | 1.56 [1.32-1.83] | 1.13 [1.11-1.14] | 67,194 | 66,929 | 66,284 | 444 | 56,865 |
| H27 | 1.19 [1.08-1.32] | 1.22 [1.09-1.35] | 1.19 [1.07-1.33] | | 1.15 [1.03-1.28] | 1.16 [1.05-1.29] | 1.16 [1.04-1.29] | 1.14 [1.01-1.27] | | 1.13 [1.01-1.27] | 935 | 877 | 810 | NA | 824 |
| H28 | 1.32 [1.12-1.55] | 1.35 [1.14-1.59] | 1.28 [1.08-1.51] | | 1.27 [1.07-1.51] | 1.24 [1.05-1.47] | 1.22 [1.03-1.44] | 1.13 [0.95-1.34] | | 1.24 [1.04-1.48] | 365 | 361 | 350 | NA | 329 |
| H30 | 1.33 [1.00-1.76] | 1.29 [0.96-1.74] | | | 1.34 [1.00-1.81] | 1.26 [0.94-1.68] | 1.17 [0.87-1.59] | | | 1.30 [0.96-1.76] | 125 | 109 | NA | NA | 109 |
| H31 | 1.11 [1.01-1.22] | 1.13 [1.02-1.24] | 1.10 [1.00-1.22] | | 1.05 [0.95-1.16] | 1.09 [0.99-1.20] | 1.09 [0.98-1.20] | 1.06 [0.96-1.17] | | 1.04 [0.94-1.15] | 1,062 | 1,040 | 999 | NA | 953 |
| H33 | 1.05 [1.00-1.10] | 1.08 [1.02-1.13] | 1.04 [0.99-1.10] | 1.10 [0.89-1.35] | 1.03 [0.98-1.09] | 1.03 [0.98-1.08] | 1.05 [1.00-1.11] | 1.02 [0.97-1.08] | 1.08 [0.87-1.34] | 1.03 [0.97-1.09] | 3,916 | 3,790 | 3,415 | 243 | 3,116 |
| H34 | 1.17 [1.10-1.24] | 1.17 [1.10-1.24] | 1.15 [1.08-1.22] | | 1.11 [1.04-1.18] | 1.14 [1.07-1.21] | 1.11 [1.04-1.18] | 1.09 [1.02-1.16] | | 1.09 [1.02-1.17] | 2,582 | 2,577 | 2,534 | NA | 2,300 |
| H35 | 1.17 [1.14-1.19] | 1.17 [1.15-1.20] | 1.16 [1.14-1.19] | 1.16 [0.96-1.39] | 1.12 [1.10-1.15] | 1.13 [1.11-1.16] | 1.12 [1.10-1.15] | 1.11 [1.09-1.13] | 1.14 [0.94-1.38] | 1.10 [1.08-1.13] | 23,935 | 23,741 | 23,343 | 308 | 21,152 |
| H36 | 1.25 [1.21-1.29] | 1.25 [1.21-1.29] | 1.24 [1.20-1.28] | 1.09 [0.81-1.48] | 1.17 [1.13-1.21] | 1.18 [1.14-1.22] | 1.09 [1.06-1.13] | 1.08 [1.05-1.12] | 1.14 [0.83-1.57] | 1.15 [1.11-1.19] | 9,878 | 9,864 | 9,452 | 111 | 9,023 |
| H40 | 1.18 [1.15-1.20] | 1.18 [1.15-1.20] | 1.18 [1.15-1.20] | 1.29 [1.00-1.67] | 1.13 [1.11-1.16] | 1.14 [1.12-1.17] | 1.13 [1.11-1.16] | 1.13 [1.11-1.16] | 1.26 [0.97-1.64] | 1.11 [1.08-1.14] | 21,887 | 21,771 | 21,555 | 166 | 18,784 |
| H43 | 1.13 [1.07-1.18] | 1.13 [1.07-1.18] | 1.14 [1.09-1.20] | | 1.08 [1.03-1.13] | 1.08 [1.03-1.14] | 1.07 [1.01-1.12] | 1.08 [1.03-1.13] | | 1.06 [1.01-1.11] | 4,302 | 4,261 | 4,076 | NA | 3,808 |
| H44 | 1.30 [1.20-1.40] | 1.32 [1.22-1.43] | 1.29 [1.19-1.41] | 1.25 [0.95-1.64] | 1.24 [1.14-1.35] | 1.24 [1.14-1.34] | 1.25 [1.15-1.36] | 1.22 [1.12-1.33] | 1.14 [0.87-1.51] | 1.19 [1.09-1.30] | 1,563 | 1,446 | 1,338 | 145 | 1,383 |
| H46 | 1.37 [1.19-1.58] | 1.41 [1.22-1.63] | 1.33 [1.12-1.58] | 1.43 [1.04-1.97] | 1.22 [1.05-1.42] | 1.33 [1.15-1.54] | 1.27 [1.09-1.48] | 1.20 [1.01-1.44] | 1.45 [1.04-2.02] | 1.18 [1.01-1.38] | 513 | 464 | 327 | 108 | 429 |
| H47 | 1.18 [1.10-1.26] | 1.23 [1.13-1.33] | 1.24 [1.13-1.36] | 1.10 [0.97-1.24] | 1.15 [1.07-1.24] | 1.13 [1.06-1.22] | 1.13 [1.04-1.23] | 1.18 [1.08-1.30] | 1.06 [0.93-1.20] | 1.12 [1.04-1.21] | 2,095 | 1,584 | 1,118 | 730 | 1,845 |
| H49 | 1.06 [0.97-1.16] | 1.10 [1.00-1.21] | 1.13 [1.02-1.26] | 0.97 [0.80-1.18] | 1.02 [0.93-1.13] | 1.01 [0.92-1.11] | 1.02 [0.92-1.13] | 1.05 [0.94-1.17] | 0.92 [0.75-1.12] | 1.00 [0.91-1.10] | 1,185 | 965 | 849 | 258 | 1,024 |
| H50 | 1.07 [1.03-1.12] | 1.27 [1.18-1.37] | 1.29 [1.18-1.40] | 1.01 [0.96-1.05] | 1.03 [0.99-1.08] | 1.04 [1.00-1.08] | 1.19 [1.11-1.28] | 1.20 [1.10-1.31] | 0.99 [0.94-1.04] | 1.01 [0.97-1.06] | 6,189 | 1,873 | 1,274 | 4,525 | 5,152 |
| H51 | 1.20 [0.98-1.46] | 1.25 [0.98-1.59] | 1.11 [0.84-1.48] | | 1.05 [0.85-1.30] | 1.12 [0.91-1.38] | 1.12 [0.88-1.44] | 1.01 [0.76-1.35] | | 1.00 [0.81-1.24] | 240 | 170 | 124 | NA | 223 |
| H52 | 1.13 [1.09-1.17] | 1.13 [1.09-1.17] | 1.12 [1.08-1.17] | 1.13 [1.02-1.25] | 1.09 [1.05-1.13] | 1.10 [1.06-1.14] | 1.09 [1.05-1.13] | 1.08 [1.04-1.12] | 1.10 [0.99-1.21] | 1.07 [1.02-1.11] | 7,679 | 6,786 | 6,490 | 1,014 | 6,731 |
| H53 | 1.24 [1.21-1.27] | 1.25 [1.22-1.28] | 1.25 [1.21-1.29] | 1.18 [1.12-1.24] | 1.16 [1.13-1.19] | 1.16 [1.13-1.18] | 1.11 [1.08-1.14] | 1.13 [1.09-1.16] | 1.13 [1.07-1.19] | 1.11 [1.08-1.14] | 16,638 | 14,327 | 10,510 | 3,753 | 14,631 |
| H54 | 1.20 [1.16-1.24] | 1.22 [1.18-1.25] | 1.21 [1.17-1.25] | 1.00 [0.89-1.12] | 1.14 [1.11-1.18] | 1.16 [1.12-1.20] | 1.14 [1.11-1.18] | 1.14 [1.10-1.17] | 0.96 [0.86-1.08] | 1.12 [1.09-1.16] | 11,421 | 10,807 | 10,273 | 773 | 10,552 |
| H55 | 1.12 [1.03-1.23] | 1.19 [1.07-1.32] | 1.15 [1.03-1.29] | 0.95 [0.80-1.11] | 1.06 [0.97-1.17] | 1.07 [0.98-1.17] | 1.09 [0.98-1.21] | 1.04 [0.93-1.17] | 0.92 [0.78-1.09] | 1.04 [0.94-1.14] | 1,222 | 903 | 763 | 368 | 1,120 |
| H57 | 1.27 [1.20-1.35] | 1.27 [1.19-1.36] | 1.28 [1.19-1.37] | 1.28 [1.13-1.46] | 1.20 [1.12-1.27] | 1.19 [1.12-1.27] | 1.15 [1.07-1.23] | 1.16 [1.08-1.25] | 1.22 [1.07-1.40] | 1.14 [1.07-1.21] | 2,834 | 2,314 | 2,013 | 631 | 2,526 |
| H58 | 1.23 [1.07-1.42] | 1.26 [1.09-1.46] | 1.17 [1.01-1.36] | | 1.16 [1.00-1.35] | 1.18 [1.02-1.36] | 1.15 [1.00-1.34] | 1.06 [0.91-1.23] | | 1.13 [0.98-1.32] | 515 | 483 | 451 | NA | 455 |
| H59 | 1.17 [1.05-1.30] | 1.16 [1.05-1.30] | 1.10 [0.98-1.22] | | 1.12 [1.00-1.25] | 1.14 [1.03-1.27] | 1.13 [1.01-1.26] | 1.06 [0.95-1.18] | | 1.11 [0.99-1.25] | 842 | 811 | 779 | NA | 758 |
| Diseases of the ear and mastoid process (H60-H95) | | | | | | | | | | | | | | | |
| H60 | 1.60 [1.51-1.69] | 1.71 [1.60-1.83] | 1.85 [1.71-2.00] | 1.32 [1.20-1.46] | 1.48 [1.39-1.57] | 1.50 [1.42-1.59] | 1.54 [1.43-1.64] | 1.67 [1.54-1.81] | 1.28 [1.16-1.41] | 1.42 [1.34-1.51] | 3,336 | 2,433 | 1,757 | 1,163 | 2,866 |
| H61 | 1.35 [1.29-1.40] | 1.45 [1.37-1.54] | 1.45 [1.36-1.55] | 1.26 [1.19-1.33] | 1.27 [1.22-1.33] | 1.30 [1.24-1.35] | 1.35 [1.27-1.43] | 1.34 [1.26-1.43] | 1.22 [1.15-1.29] | 1.24 [1.18-1.30] | 5,991 | 3,047 | 2,555 | 3,129 | 5,243 |
| H65 | 1.21 [1.18-1.24] | 1.35 [1.25-1.46] | 1.35 [1.24-1.48] | 1.20 [1.17-1.23] | 1.17 [1.14-1.20] | 1.17 [1.14-1.20] | 1.22 [1.12-1.32] | 1.24 [1.13-1.35] | 1.16 [1.13-1.19] | 1.14 [1.11-1.17] | 16,730 | 1,582 | 1,177 | 15,291 | 13,784 |
| H66 | 1.26 [1.22-1.31] | 1.53 [1.43-1.64] | 1.52 [1.40-1.65] | 1.21 [1.16-1.25] | 1.21 [1.16-1.25] | 1.21 [1.17-1.26] | 1.39 [1.29-1.49] | 1.37 [1.26-1.50] | 1.17 [1.13-1.22] | 1.18 [1.13-1.22] | 8,169 | 2,125 | 1,460 | 6,303 | 7,174 |
| H69 | 1.46 [1.28-1.66] | 1.49 [1.27-1.74] | 1.47 [1.21-1.79] | 1.43 [1.18-1.73] | 1.32 [1.16-1.52] | 1.36 [1.19-1.56] | 1.28 [1.09-1.51] | 1.27 [1.04-1.55] | 1.44 [1.18-1.76] | 1.25 [1.08-1.43] | 634 | 400 | 254 | 297 | 544 |
| H70 | 1.18 [1.05-1.33] | 1.38 [1.17-1.63] | 1.24 [1.03-1.50] | 1.04 [0.88-1.23] | 1.10 [0.97-1.24] | 1.13 [1.00-1.28] | 1.23 [1.04-1.46] | 1.13 [0.93-1.38] | 1.01 [0.85-1.20] | 1.07 [0.94-1.22] | 696 | 376 | 280 | 350 | 607 |
| H71 | 1.10 [1.02-1.19] | 1.12 [1.01-1.24] | 1.12 [0.98-1.28] | 1.07 [0.95-1.19] | 1.04 [0.95-1.13] | 1.08 [0.99-1.17] | 1.08 [0.98-1.21] | 1.07 [0.93-1.22] | 1.05 [0.94-1.18] | 1.05 [0.96-1.15] | 1,527 | 865 | 512 | 792 | 1,274 |
| H72 | 1.24 [1.18-1.31] | 1.25 [1.16-1.35] | 1.31 [1.19-1.43] | 1.17 [1.09-1.26] | 1.13 [1.07-1.20] | 1.19 [1.13-1.26] | 1.18 [1.09-1.27] | 1.20 [1.10-1.32] | 1.14 [1.06-1.23] | 1.11 [1.05-1.18] | 3,386 | 1,686 | 1,115 | 1,886 | 2,890 |
| H73 | 1.22 [1.14-1.30] | 1.32 [1.18-1.48] | 1.34 [1.16-1.55] | 1.19 [1.10-1.28] | 1.14 [1.06-1.22] | 1.17 [1.10-1.25] | 1.23 [1.09-1.38] | 1.23 [1.06-1.42] | 1.16 [1.07-1.25] | 1.12 [1.05-1.20] | 2,421 | 755 | 449 | 1,806 | 2,153 |
| H74 | 1.19 [1.10-1.29] | 1.24 [1.12-1.37] | 1.33 [1.18-1.51] | 1.10 [0.99-1.23] | 1.12 [1.03-1.21] | 1.15 [1.06-1.24] | 1.16 [1.04-1.29] | 1.24 [1.09-1.41] | 1.06 [0.95-1.19] | 1.09 [1.00-1.18] | 1,687 | 906 | 622 | 880 | 1,471 |
| H80 | 1.09 [0.91-1.30] | 1.12 [0.93-1.34] | 1.14 [0.93-1.40] | | 1.11 [0.92-1.35] | 1.07 [0.90-1.29] | 1.09 [0.91-1.32] | 1.12 [0.90-1.38] | | 1.09 [0.89-1.33] | 291 | 286 | 209 | NA | 238 |
| H81 | 1.26 [1.20-1.32] | 1.26 [1.20-1.32] | 1.26 [1.20-1.32] | 1.31 [0.94-1.81] | 1.17 [1.11-1.23] | 1.17 [1.12-1.23] | 1.14 [1.09-1.20] | 1.14 [1.09-1.20] | 1.23 [0.87-1.73] | 1.11 [1.06-1.16] | 4,814 | 4,767 | 4,531 | 103 | 4,323 |
| H83 | 1.26 [1.18-1.35] | 1.28 [1.19-1.38] | 1.26 [1.17-1.36] | 1.18 [0.95-1.47] | 1.17 [1.09-1.26] | 1.17 [1.09-1.25] | 1.16 [1.08-1.25] | 1.13 [1.04-1.22] | 1.12 [0.89-1.40] | 1.11 [1.03-1.19] | 2,065 | 1,914 | 1,663 | 213 | 1,781 |

#### Supplementary Table 3: Hazard ratios and events from all cohorts

| | Hazard ratio (99% confidence interval) | | | | | | | | | | | | | | |
|---|---|---|---|---|---|---|---|---|---|---|---|---|---|---|---|
| | crude | | | | | adjusted | | | | | Events (in exposed) | | | | |
| Outcome | any age | 18+ | 40+ | <18 | hosp. | any age | 18+ | 40+ | <18 | hosp. | any age | 18+ | 40+ | <18 | hosp. |
| H90 | 1.29 [1.24-1.34] | 1.36 [1.30-1.43] | 1.37 [1.30-1.44] | 1.16 [1.09-1.23] | 1.20 [1.16-1.25] | 1.22 [1.17-1.27] | 1.23 [1.17-1.29] | 1.24 [1.18-1.31] | 1.12 [1.05-1.19] | 1.15 [1.11-1.20] | 6,956 | 4,601 | 3,963 | 2,583 | 6,392 |
| H91 | 1.23 [1.20-1.25] | 1.24 [1.21-1.26] | 1.23 [1.21-1.26] | 1.15 [1.08-1.21] | 1.18 [1.15-1.20] | 1.17 [1.15-1.20] | 1.16 [1.14-1.19] | 1.16 [1.13-1.18] | 1.11 [1.05-1.17] | 1.14 [1.12-1.16] | 30,611 | 27,774 | 26,657 | 3,227 | 28,186 |
| H92 | 1.34 [1.26-1.42] | 1.49 [1.37-1.61] | 1.43 [1.30-1.57] | 1.23 [1.13-1.33] | 1.26 [1.18-1.34] | 1.25 [1.18-1.33] | 1.31 [1.20-1.42] | 1.26 [1.14-1.39] | 1.18 [1.08-1.29] | 1.20 [1.13-1.28] | 2,840 | 1,565 | 1,090 | 1,443 | 2,500 |
| H93 | 1.35 [1.27-1.43] | 1.37 [1.29-1.46] | 1.36 [1.27-1.44] | 1.20 [1.01-1.43] | 1.28 [1.21-1.36] | 1.26 [1.19-1.34] | 1.23 [1.16-1.31] | 1.22 [1.14-1.30] | 1.16 [0.98-1.39] | 1.21 [1.14-1.29] | 3,029 | 2,778 | 2,419 | 378 | 2,708 |
| H95 | 1.27 [1.07-1.50] | 1.36 [1.10-1.67] | 1.36 [1.04-1.79] | 1.20 [0.93-1.54] | 1.12 [0.94-1.33] | 1.24 [1.05-1.48] | 1.27 [1.03-1.58] | 1.30 [0.99-1.72] | 1.17 [0.90-1.51] | 1.13 [0.94-1.35] | 349 | 219 | 121 | 168 | 319 |
| Diseases of the circulatory system (I00-I99) | | | | | | | | | | | | | | | |
| I05 | 1.30 [1.20-1.40] | 1.27 [1.17-1.37] | 1.32 [1.21-1.43] | | 1.19 [1.10-1.30] | 1.22 [1.12-1.32] | 1.17 [1.07-1.27] | 1.21 [1.11-1.32] | | 1.16 [1.07-1.26] | 1,575 | 1,563 | 1,520 | NA | 1,409 |
| I06 | 1.34 [1.05-1.73] | 1.25 [0.97-1.60] | 1.36 [1.06-1.75] | | 1.18 [0.91-1.52] | 1.26 [0.98-1.63] | 1.14 [0.89-1.48] | 1.22 [0.95-1.58] | | 1.12 [0.86-1.45] | 155 | 153 | 153 | NA | 141 |
| I07 | 1.31 [1.24-1.38] | 1.30 [1.23-1.37] | 1.31 [1.25-1.38] | 0.89 [0.67-1.18] | 1.23 [1.17-1.30] | 1.25 [1.19-1.32] | 1.20 [1.14-1.26] | 1.20 [1.14-1.27] | 0.89 [0.67-1.20] | 1.20 [1.14-1.27] | 4,088 | 4,013 | 3,880 | 118 | 3,843 |
| I08 | 1.21 [1.19-1.24] | 1.22 [1.19-1.25] | 1.22 [1.19-1.25] | 1.08 [0.84-1.37] | 1.15 [1.12-1.18] | 1.17 [1.14-1.20] | 1.13 [1.11-1.16] | 1.13 [1.10-1.16] | 1.06 [0.83-1.37] | 1.12 [1.10-1.15] | 19,877 | 19,808 | 19,471 | 177 | 18,578 |
| I09 | 1.42 [1.17-1.74] | 1.43 [1.17-1.74] | 1.52 [1.24-1.86] | | 1.37 [1.11-1.68] | 1.36 [1.11-1.67] | 1.35 [1.10-1.65] | 1.39 [1.13-1.71] | | 1.33 [1.07-1.64] | 249 | 247 | 239 | NA | 230 |
| I10 | 1.24 [1.23-1.25] | 1.24 [1.23-1.25] | 1.24 [1.23-1.25] | 1.21 [1.12-1.31] | 1.17 [1.16-1.18] | 1.19 [1.18-1.20] | 1.13 [1.12-1.14] | 1.12 [1.11-1.13] | 1.17 [1.08-1.27] | 1.14 [1.13-1.15] | 207,382 | 206,548 | 200,764 | 1,786 | 153,264 |
| I11 | 1.21 [1.10-1.33] | 1.21 [1.10-1.33] | 1.22 [1.11-1.35] | | 1.06 [0.96-1.17] | 1.15 [1.04-1.27] | 1.08 [0.98-1.19] | 1.10 [0.99-1.21] | | 1.04 [0.94-1.15] | 1,103 | 1,092 | 1,084 | NA | 972 |
| I12 | 1.29 [1.26-1.33] | 1.30 [1.27-1.34] | 1.31 [1.27-1.35] | | 1.19 [1.16-1.23] | 1.25 [1.21-1.29] | 1.19 [1.15-1.23] | 1.19 [1.16-1.23] | | 1.18 [1.14-1.22] | 11,437 | 11,390 | 11,255 | NA | 10,333 |
| I13 | 1.30 [1.13-1.49] | 1.29 [1.12-1.48] | 1.28 [1.11-1.47] | | 1.17 [1.02-1.35] | 1.22 [1.06-1.40] | 1.15 [1.00-1.33] | 1.14 [0.99-1.31] | | 1.14 [0.99-1.31] | 579 | 577 | 573 | NA | 561 |
| I15 | 1.28 [1.15-1.42] | 1.31 [1.17-1.47] | 1.26 [1.12-1.42] | 1.07 [0.84-1.38] | 1.11 [1.00-1.24] | 1.22 [1.09-1.36] | 1.17 [1.04-1.32] | 1.11 [0.98-1.26] | 1.08 [0.84-1.39] | 1.09 [0.98-1.22] | 875 | 727 | 675 | 163 | 797 |
| I20 | 1.28 [1.26-1.29] | 1.27 [1.26-1.29] | 1.28 [1.26-1.30] | | 1.17 [1.15-1.19] | 1.20 [1.18-1.21] | 1.14 [1.12-1.15] | 1.14 [1.12-1.15] | | 1.12 [1.10-1.14] | 44,789 | 44,808 | 44,361 | NA | 37,819 |
| I21 | 1.16 [1.13-1.18] | 1.15 [1.13-1.18] | 1.15 [1.13-1.18] | | 1.11 [1.09-1.13] | 1.13 [1.11-1.15] | 1.08 [1.06-1.11] | 1.08 [1.06-1.10] | | 1.10 [1.07-1.12] | 28,070 | 28,078 | 27,805 | NA | 23,835 |
| I22 | 1.28 [1.20-1.36] | 1.28 [1.21-1.36] | 1.29 [1.22-1.37] | | 1.18 [1.11-1.25] | 1.22 [1.15-1.30] | 1.16 [1.09-1.23] | 1.16 [1.09-1.24] | | 1.15 [1.08-1.23] | 2,692 | 2,699 | 2,683 | NA | 2,441 |
| I23 | 1.24 [0.98-1.57] | 1.23 [0.97-1.56] | 1.30 [1.02-1.66] | | 1.31 [1.02-1.70] | 1.22 [0.95-1.55] | 1.15 [0.90-1.47] | 1.29 [1.00-1.65] | | 1.29 [1.00-1.68] | 167 | 166 | 162 | NA | 144 |
| I24 | 1.24 [1.20-1.28] | 1.25 [1.21-1.29] | 1.24 [1.20-1.28] | | 1.17 [1.13-1.21] | 1.18 [1.13-1.22] | 1.13 [1.09-1.17] | 1.11 [1.07-1.15] | | 1.14 [1.10-1.18] | 8,471 | 8,469 | 8,361 | NA | 7,812 |
| I25 | 1.22 [1.21-1.23] | 1.22 [1.21-1.24] | 1.22 [1.21-1.24] | 1.06 [0.81-1.37] | 1.14 [1.13-1.16] | 1.16 [1.15-1.18] | 1.11 [1.10-1.13] | 1.11 [1.10-1.12] | 1.08 [0.83-1.41] | 1.11 [1.10-1.12] | 75,749 | 75,716 | 75,062 | 144 | 64,357 |
| I26 | 1.13 [1.10-1.16] | 1.12 [1.09-1.15] | 1.12 [1.09-1.15] | 1.07 [0.88-1.29] | 1.07 [1.04-1.10] | 1.08 [1.05-1.11] | 1.05 [1.02-1.08] | 1.04 [1.01-1.07] | 1.03 [0.84-1.26] | 1.04 [1.01-1.07] | 14,114 | 14,073 | 13,190 | 277 | 12,322 |
| I27 | 1.32 [1.28-1.37] | 1.32 [1.28-1.37] | 1.31 [1.27-1.36] | 0.85 [0.63-1.15] | 1.25 [1.21-1.29] | 1.25 [1.21-1.29] | 1.19 [1.15-1.23] | 1.18 [1.14-1.22] | 0.77 [0.57-1.05] | 1.20 [1.16-1.24] | 9,635 | 9,575 | 9,398 | 106 | 9,065 |
| I28 | 1.16 [0.96-1.41] | 1.19 [0.97-1.46] | 1.16 [0.92-1.45] | | 0.99 [0.82-1.21] | 1.07 [0.88-1.30] | 1.06 [0.86-1.31] | 1.01 [0.80-1.28] | | 0.94 [0.77-1.15] | 269 | 232 | 190 | NA | 246 |
| I30 | 1.24 [1.07-1.42] | 1.20 [1.04-1.39] | 1.26 [1.07-1.49] | | 1.18 [1.01-1.37] | 1.19 [1.03-1.37] | 1.13 [0.98-1.31] | 1.19 [1.00-1.41] | | 1.15 [0.98-1.34] | 475 | 445 | 333 | NA | 388 |
| I31 | 1.22 [1.17-1.28] | 1.23 [1.18-1.28] | 1.22 [1.17-1.28] | 1.02 [0.88-1.19] | 1.13 [1.08-1.18] | 1.17 [1.12-1.22] | 1.13 [1.08-1.18] | 1.13 [1.08-1.18] | 0.99 [0.85-1.16] | 1.10 [1.05-1.15] | 5,730 | 5,512 | 4,908 | 442 | 5,048 |
| I33 | 1.35 [1.23-1.50] | 1.36 [1.23-1.51] | 1.34 [1.20-1.49] | | 1.25 [1.13-1.39] | 1.30 [1.17-1.44] | 1.26 [1.13-1.40] | 1.25 [1.12-1.39] | | 1.22 [1.10-1.36] | 1,011 | 982 | 902 | NA | 891 |
| I34 | 1.18 [1.14-1.21] | 1.19 [1.16-1.22] | 1.19 [1.16-1.23] | 0.99 [0.81-1.21] | 1.13 [1.09-1.16] | 1.13 [1.10-1.16] | 1.11 [1.08-1.14] | 1.10 [1.07-1.14] | 0.93 [0.76-1.15] | 1.10 [1.07-1.13] | 12,668 | 12,517 | 12,151 | 250 | 11,507 |
| I35 | 1.23 [1.20-1.26] | 1.23 [1.20-1.26] | 1.23 [1.20-1.26] | 0.95 [0.78-1.17] | 1.17 [1.14-1.20] | 1.17 [1.15-1.20] | 1.14 [1.11-1.17] | 1.14 [1.11-1.17] | 0.96 [0.78-1.18] | 1.13 [1.10-1.16] | 19,023 | 18,862 | 18,630 | 238 | 17,300 |
| I36 | 1.23 [1.12-1.34] | 1.21 [1.11-1.32] | 1.24 [1.13-1.35] | | 1.17 [1.07-1.28] | 1.16 [1.06-1.26] | 1.12 [1.02-1.22] | 1.13 [1.04-1.24] | | 1.13 [1.04-1.24] | 1,417 | 1,373 | 1,313 | NA | 1,334 |
| I37 | 1.22 [1.12-1.32] | 1.26 [1.15-1.37] | 1.29 [1.18-1.41] | 0.95 [0.75-1.20] | 1.13 [1.04-1.23] | 1.15 [1.06-1.25] | 1.17 [1.07-1.27] | 1.17 [1.07-1.28] | 0.94 [0.74-1.20] | 1.10 [1.01-1.20] | 1,558 | 1,448 | 1,318 | 167 | 1,463 |
| I38 | 1.29 [1.21-1.38] | 1.30 [1.21-1.40] | 1.33 [1.24-1.43] | 1.09 [0.84-1.40] | 1.21 [1.13-1.30] | 1.23 [1.15-1.32] | 1.20 [1.11-1.29] | 1.22 [1.14-1.32] | 1.06 [0.82-1.38] | 1.17 [1.09-1.26] | 2,228 | 2,082 | 2,008 | 158 | 2,005 |
| I40 | 1.29 [1.01-1.64] | 1.23 [0.96-1.58] | 1.50 [1.10-2.04] | | 1.35 [1.04-1.76] | 1.23 [0.96-1.59] | 1.19 [0.92-1.54] | 1.39 [1.01-1.91] | | 1.33 [1.02-1.75] | 173 | 156 | 108 | NA | 152 |
| I42 | 1.21 [1.16-1.27] | 1.23 [1.18-1.29] | 1.22 [1.16-1.28] | 1.17 [0.96-1.43] | 1.15 [1.10-1.20] | 1.15 [1.10-1.20] | 1.12 [1.07-1.17] | 1.10 [1.05-1.16] | 1.18 [0.95-1.45] | 1.11 [1.06-1.16] | 4,772 | 4,639 | 4,297 | 242 | 4,273 |
| I43 | 0.98 [0.78-1.25] | 1.07 [0.85-1.36] | 0.98 [0.77-1.25] | | 0.97 [0.76-1.23] | 0.94 [0.74-1.20] | 0.99 [0.78-1.27] | 0.92 [0.72-1.18] | | 0.93 [0.73-1.18] | 178 | 172 | 160 | NA | 167 |
| I44 | 1.20 [1.18-1.22] | 1.20 [1.18-1.23] | 1.20 [1.18-1.23] | 1.29 [1.04-1.58] | 1.15 [1.13-1.17] | 1.16 [1.13-1.18] | 1.13 [1.10-1.15] | 1.12 [1.10-1.14] | 1.26 [1.02-1.56] | 1.13 [1.10-1.15] | 29,534 | 29,415 | 29,080 | 256 | 27,092 |
| I45 | 1.20 [1.18-1.23] | 1.21 [1.18-1.24] | 1.20 [1.17-1.23] | 1.03 [0.91-1.16] | 1.15 [1.12-1.18] | 1.15 [1.12-1.18] | 1.12 [1.09-1.15] | 1.11 [1.08-1.14] | 0.99 [0.87-1.12] | 1.12 [1.09-1.15] | 18,619 | 18,306 | 17,448 | 697 | 17,020 |
| I46 | 1.23 [1.19-1.27] | 1.24 [1.20-1.28] | 1.23 [1.19-1.27] | 1.08 [0.89-1.30] | 1.18 [1.14-1.22] | 1.19 [1.15-1.23] | 1.15 [1.11-1.19] | 1.14 [1.10-1.18] | 1.04 [0.85-1.26] | 1.17 [1.13-1.21] | 9,596 | 9,458 | 9,070 | 275 | 8,594 |
| I47 | 1.22 [1.19-1.26] | 1.22 [1.18-1.26] | 1.22 [1.18-1.26] | 1.16 [1.03-1.31] | 1.14 [1.10-1.18] | 1.15 [1.12-1.19] | 1.11 [1.08-1.15] | 1.11 [1.07-1.15] | 1.11 [0.98-1.25] | 1.10 [1.06-1.13] | 10,826 | 10,396 | 9,373 | 764 | 9,433 |
| I48 | 1.16 [1.15-1.18] | 1.16 [1.15-1.18] | 1.16 [1.15-1.18] | 1.06 [0.85-1.32] | 1.12 [1.11-1.13] | 1.13 [1.11-1.14] | 1.10 [1.09-1.11] | 1.09 [1.08-1.11] | 1.01 [0.80-1.27] | 1.10 [1.09-1.11] | 81,212 | 81,194 | 80,479 | 207 | 70,255 |
| I49 | 1.18 [1.15-1.22] | 1.19 [1.15-1.22] | 1.18 [1.15-1.22] | 1.16 [1.03-1.31] | 1.12 [1.09-1.15] | 1.12 [1.09-1.15] | 1.09 [1.06-1.12] | 1.09 [1.06-1.12] | 1.10 [0.96-1.24] | 1.08 [1.05-1.11] | 12,655 | 12,285 | 11,350 | 733 | 11,482 |
| I50 | 1.26 [1.25-1.28] | 1.26 [1.24-1.27] | 1.26 [1.25-1.28] | 1.09 [0.90-1.33] | 1.20 [1.19-1.22] | 1.20 [1.19-1.22] | 1.15 [1.14-1.17] | 1.15 [1.14-1.17] | 1.10 [0.90-1.35] | 1.17 [1.15-1.19] | 64,824 | 64,744 | 64,197 | 270 | 58,996 |
| I51 | 1.25 [1.23-1.27] | 1.25 [1.23-1.27] | 1.25 [1.23-1.27] | 1.09 [0.97-1.23] | 1.18 [1.16-1.20] | 1.18 [1.16-1.21] | 1.14 [1.12-1.16] | 1.13 [1.11-1.16] | 1.07 [0.95-1.22] | 1.14 [1.12-1.16] | 35,441 | 35,043 | 34,064 | 721 | 32,954 |
| I60 | 1.16 [1.08-1.24] | 1.16 [1.08-1.25] | 1.14 [1.06-1.22] | | 1.10 [1.02-1.18] | 1.15 [1.07-1.24] | 1.13 [1.05-1.21] | 1.11 [1.03-1.19] | | 1.10 [1.02-1.20] | 1,910 | 1,880 | 1,738 | NA | 1,556 |
| I61 | 1.17 [1.12-1.23] | 1.17 [1.12-1.23] | 1.17 [1.12-1.22] | 0.88 [0.65-1.19] | 1.14 [1.09-1.20] | 1.16 [1.11-1.22] | 1.14 [1.09-1.20] | 1.13 [1.08-1.19] | 0.88 [0.64-1.20] | 1.14 [1.09-1.20] | 4,912 | 4,837 | 4,715 | 104 | 4,230 |
| I62 | 1.16 [1.09-1.23] | 1.18 [1.11-1.25] | 1.17 [1.10-1.25] | | 1.10 [1.03-1.17] | 1.14 [1.07-1.21] | 1.13 [1.07-1.21] | 1.12 [1.05-1.19] | | 1.10 [1.04-1.18] | 2,840 | 2,794 | 2,737 | NA | 2,549 |
| I63 | 1.09 [1.07-1.11] | 1.09 [1.07-1.12] | 1.09 [1.07-1.11] | 1.10 [0.86-1.42] | 1.07 [1.04-1.09] | 1.08 [1.06-1.10] | 1.05 [1.03-1.07] | 1.04 [1.02-1.07] | 1.17 [0.90-1.51] | 1.07 [1.04-1.09] | 22,057 | 21,969 | 21,680 | 159 | 19,420 |
| I64 | 1.15 [1.10-1.19] | 1.14 [1.09-1.19] | 1.15 [1.10-1.19] | | 1.12 [1.07-1.17] | 1.13 [1.08-1.18] | 1.09 [1.04-1.13] | 1.09 [1.05-1.14] | | 1.12 [1.07-1.17] | 5,844 | 5,845 | 5,801 | NA | 4,912 |
| I65 | 1.17 [1.13-1.22] | 1.17 [1.12-1.22] | 1.17 [1.12-1.22] | | 1.11 [1.06-1.16] | 1.14 [1.10-1.19] | 1.09 [1.04-1.14] | 1.08 [1.04-1.13] | | 1.10 [1.05-1.15] | 5,593 | 5,582 | 5,554 | NA | 4,973 |
| I66 | 1.15 [0.97-1.36] | 1.14 [0.96-1.36] | 1.08 [0.90-1.28] | | 1.12 [0.94-1.34] | 1.11 [0.94-1.32] | 1.08 [0.91-1.29] | 1.00 [0.84-1.20] | | 1.09 [0.91-1.30] | 358 | 333 | 309 | NA | 327 |
| I67 | 1.18 [1.16-1.20] | 1.18 [1.16-1.20] | 1.18 [1.16-1.20] | 1.08 [0.82-1.41] | 1.14 [1.12-1.16] | 1.15 [1.13-1.17] | 1.11 [1.09-1.13] | 1.11 [1.09-1.13] | 1.08 [0.82-1.43] | 1.13 [1.11-1.15] | 35,644 | 35,555 | 35,303 | 135 | 33,238 |
| I68 | 1.22 [0.99-1.52] | 1.24 [1.00-1.54] | 1.22 [0.98-1.52] | | 1.25 [1.00-1.56] | 1.20 [0.96-1.49] | 1.19 [0.95-1.49] | 1.16 [0.92-1.45] | | 1.22 [0.97-1.53] | 216 | 214 | 208 | NA | 202 |
| I69 | 1.18 [1.15-1.22] | 1.19 [1.15-1.23] | 1.18 [1.15-1.22] | 0.94 [0.73-1.21] | 1.10 [1.06-1.13] | 1.15 [1.12-1.19] | 1.11 [1.07-1.14] | 1.10 [1.06-1.13] | 0.91 [0.70-1.17] | 1.09 [1.06-1.13] | 10,401 | 10,290 | 10,123 | 153 | 9,596 |
| I70 | 1.26 [1.21-1.30] | 1.25 [1.21-1.30] | 1.27 [1.22-1.31] | | 1.18 [1.14-1.22] | 1.21 [1.17-1.26] | 1.14 [1.10-1.18] | 1.15 [1.11-1.19] | | 1.17 [1.13-1.21] | 9,179 | 9,177 | 9,126 | NA | 8,437 |
| I71 | 1.17 [1.13-1.20] | 1.17 [1.13-1.21] | 1.18 [1.14-1.21] | | 1.11 [1.07-1.15] | 1.15 [1.12-1.19] | 1.11 [1.07-1.14] | 1.11 [1.08-1.15] | | 1.11 [1.08-1.15] | 10,600 | 10,544 | 10,459 | NA | 9,458 |
| I72 | 1.17 [1.10-1.25] | 1.18 [1.11-1.25] | 1.18 [1.10-1.25] | | 1.11 [1.04-1.18] | 1.14 [1.07-1.21] | 1.09 [1.02-1.16] | 1.09 [1.02-1.16] | | 1.10 [1.03-1.17] | 2,555 | 2,547 | 2,443 | NA | 2,312 |
| I73 | 1.40 [1.37-1.43] | 1.40 [1.37-1.43] | 1.39 [1.36-1.42] | 1.44 [1.24-1.68] | 1.29 [1.26-1.32] | 1.32 [1.29-1.35] | 1.24 [1.21-1.27] | 1.23 [1.20-1.26] | 1.35 [1.15-1.58] | 1.25 [1.22-1.28] | 21,801 | 21,629 | 20,652 | 481 | 19,729 |
| I74 | 1.18 [1.13-1.23] | 1.17 [1.12-1.22] | 1.19 [1.14-1.25] | | 1.11 [1.06-1.17] | 1.14 [1.09-1.19] | 1.07 [1.02-1.12] | 1.09 [1.04-1.14] | | 1.11 [1.06-1.17] | 4,919 | 4,896 | 4,775 | NA | 4,434 |
| I77 | 1.32 [1.27-1.37] | 1.32 [1.27-1.37] | 1.33 [1.28-1.39] | 1.19 [0.91-1.56] | 1.22 [1.17-1.27] | 1.26 [1.21-1.31] | 1.20 [1.15-1.24] | 1.20 [1.16-1.25] | 1.13 [0.85-1.49] | 1.21 [1.16-1.26] | 7,078 | 7,005 | 6,801 | 142 | 6,373 |
| I78 | 1.30 [1.21-1.40] | 1.35 [1.25-1.45] | 1.31 [1.21-1.42] | 1.03 [0.77-1.36] | 1.24 [1.15-1.34] | 1.24 [1.15-1.34] | 1.24 [1.14-1.34] | 1.21 [1.12-1.31] | 1.01 [0.76-1.35] | 1.21 [1.12-1.31] | 1,829 | 1,727 | 1,615 | 121 | 1,655 |
| I79 | 1.34 [1.19-1.51] | 1.35 [1.20-1.52] | 1.35 [1.20-1.52] | | 1.19 [1.06-1.35] | 1.24 [1.10-1.40] | 1.13 [1.00-1.28] | 1.13 [1.00-1.28] | | 1.15 [1.02-1.30] | 702 | 702 | 687 | NA | 651 |
| I80 | 1.25 [1.22-1.29] | 1.24 [1.21-1.28] | 1.25 [1.21-1.28] | 1.15 [0.98-1.36] | 1.19 [1.15-1.22] | 1.20 [1.17-1.23] | 1.16 [1.12-1.19] | 1.16 [1.13-1.20] | 1.11 [0.93-1.32] | 1.15 [1.12-1.19] | 13,733 | 13,610 | 12,516 | 371 | 11,862 |
| I81 | 1.14 [1.01-1.28] | 1.17 [1.04-1.31] | 1.14 [1.01-1.28] | | 1.04 [0.92-1.17] | 1.08 [0.96-1.22] | 1.08 [0.96-1.22] | 1.06 [0.94-1.19] | | 1.03 [0.91-1.16] | 730 | 720 | 670 | NA | 649 |
| I82 | 1.21 [1.12-1.30] | 1.21 [1.12-1.31] | 1.20 [1.11-1.30] | 1.00 [0.77-1.29] | 1.13 [1.04-1.22] | 1.15 [1.07-1.25] | 1.12 [1.04-1.21] | 1.11 [1.02-1.20] | 0.98 [0.75-1.29] | 1.10 [1.02-1.19] | 1,750 | 1,683 | 1,445 | 141 | 1,556 |
| I83 | 1.63 [1.59-1.67] | 1.63 [1.59-1.67] | 1.68 [1.64-1.72] | 1.05 [0.86-1.27] | 1.52 [1.48-1.56] | 1.57 [1.54-1.61] | 1.55 [1.52-1.59] | 1.59 [1.55-1.63] | 1.01 [0.82-1.24] | 1.48 [1.45-1.52] | 19,256 | 19,220 | 17,136 | 254 | 15,892 |
| I84 | 1.37 [1.34-1.39] | 1.37 [1.34-1.39] | 1.34 [1.32-1.37] | 1.21 [1.08-1.36] | 1.25 [1.23-1.28] | 1.28 [1.25-1.30] | 1.24 [1.22-1.27] | 1.22 [1.20-1.25] | 1.17 [1.04-1.31] | 1.19 [1.17-1.22] | 26,186 | 25,999 | 21,897 | 783 | 21,420 |
| I85 | 1.46 [1.37-1.55] | 1.51 [1.41-1.61] | 1.47 [1.38-1.57] | | 1.37 [1.28-1.47] | 1.37 [1.29-1.46] | 1.35 [1.26-1.44] | 1.31 [1.22-1.40] | | 1.34 [1.25-1.43] | 2,382 | 2,351 | 2,208 | NA | 2,141 |
| I86 | 1.31 [1.22-1.41] | 1.36 [1.26-1.47] | 1.36 [1.24-1.49] | 1.03 [0.88-1.21] | 1.24 [1.15-1.34] | 1.22 [1.14-1.32] | 1.22 [1.13-1.32] | 1.21 [1.10-1.33] | 0.99 [0.84-1.17] | 1.21 [1.12-1.31] | 1,918 | 1,691 | 1,175 | 412 | 1,611 |
| I87 | 1.60 [1.52-1.69] | 1.61 [1.53-1.69] | 1.65 [1.56-1.74] | 1.06 [0.79-1.42] | 1.49 [1.42-1.57] | 1.52 [1.44-1.60] | 1.48 [1.40-1.55] | 1.51 [1.43-1.60] | 1.03 [0.76-1.39] | 1.45 [1.38-1.53] | 4,228 | 4,167 | 3,906 | 125 | 3,889 |
| I88 | 1.28 [1.22-1.36] | 1.34 [1.18-1.52] | 1.53 [1.29-1.82] | 1.26 [1.19-1.33] | 1.23 [1.16-1.31] | 1.25 [1.18-1.32] | 1.22 [1.07-1.39] | 1.45 [1.22-1.74] | 1.22 [1.15-1.29] | 1.20 [1.13-1.28] | 3,739 | 602 | 327 | 3,277 | 3,123 |
| I89 | 1.53 [1.46-1.59] | 1.52 [1.46-1.59] | 1.51 [1.44-1.58] | 1.16 [0.91-1.47] | 1.42 [1.35-1.48] | 1.43 [1.37-1.50] | 1.37 [1.31-1.44] | 1.36 [1.30-1.43] | 1.08 [0.84-1.39] | 1.37 [1.31-1.43] | 5,461 | 5,350 | 5,091 | 177 | 5,058 |
| I95 | 1.20 [1.18-1.22] | 1.21 [1.19-1.23] | 1.20 [1.19-1.22] | 1.06 [0.97-1.16] | 1.15 [1.13-1.16] | 1.16 [1.14-1.17] | 1.12 [1.11-1.14] | 1.12 [1.10-1.14] | 1.04 [0.95-1.14] | 1.13 [1.11-1.15] | 44,151 | 43,521 | 41,490 | 1,343 | 41,026 |
| I97 | 1.33 [1.17-1.51] | 1.34 [1.18-1.53] | 1.28 [1.12-1.46] | | 1.17 [1.02-1.34] | 1.26 [1.10-1.43] | 1.25 [1.09-1.43] | 1.19 [1.04-1.36] | | 1.17 [1.02-1.34] | 573 | 563 | 546 | NA | 510 |
| I98 | 1.45 [1.34-1.56] | 1.49 [1.38-1.61] | 1.45 [1.34-1.58] | | 1.36 [1.26-1.48] | 1.34 [1.24-1.45] | 1.31 [1.21-1.42] | 1.27 [1.17-1.38] | | 1.31 [1.21-1.43] | 1,591 | 1,586 | 1,468 | NA | 1,455 |
| I99 | 1.25 [1.11-1.39] | 1.22 [1.09-1.37] | 1.24 [1.11-1.39] | | 1.13 [1.00-1.27] | 1.20 [1.07-1.34] | 1.12 [1.00-1.26] | 1.14 [1.02-1.28] | | 1.11 [0.99-1.25] | 808 | 790 | 767 | NA | 718 |
| Diseases of the respiratory system (J00-J99) | | | | | | | | | | | | | | | |
| J00 | 1.41 [1.33-1.48] | 1.35 [1.23-1.48] | 1.38 [1.23-1.55] | 1.41 [1.32-1.50] | 1.36 [1.28-1.44] | 1.33 [1.26-1.40] | 1.22 [1.11-1.34] | 1.25 [1.11-1.41] | 1.35 [1.27-1.44] | 1.31 [1.23-1.39] | 3,524 | 1,113 | 728 | 2,539 | 3,159 |
| J01 | 1.46 [1.31-1.63] | 1.38 [1.21-1.56] | 1.38 [1.18-1.62] | 1.44 [1.19-1.74] | 1.28 [1.14-1.44] | 1.31 [1.17-1.47] | 1.18 [1.03-1.35] | 1.16 [0.99-1.37] | 1.36 [1.12-1.66] | 1.19 [1.05-1.34] | 851 | 617 | 388 | 306 | 714 |
| J02 | 1.32 [1.27-1.38] | 1.39 [1.31-1.46] | 1.44 [1.34-1.54] | 1.19 [1.13-1.26] | 1.23 [1.18-1.28] | 1.24 [1.19-1.29] | 1.24 [1.17-1.31] | 1.26 [1.18-1.36] | 1.15 [1.09-1.22] | 1.18 [1.13-1.23] | 6,206 | 3,525 | 2,082 | 3,263 | 5,339 |
| J03 | 1.22 [1.20-1.24] | 1.31 [1.27-1.36] | 1.25 [1.14-1.36] | 1.20 [1.17-1.22] | 1.17 [1.15-1.19] | 1.17 [1.15-1.19] | 1.17 [1.13-1.21] | 1.08 [0.98-1.19] | 1.16 [1.14-1.18] | 1.14 [1.11-1.16] | 32,053 | 8,122 | 1,121 | 27,861 | 26,714 |
| J04 | 1.34 [1.19-1.51] | 1.47 [1.28-1.68] | 1.41 [1.21-1.65] | 1.32 [1.07-1.63] | 1.33 [1.17-1.50] | 1.23 [1.09-1.39] | 1.29 [1.12-1.48] | 1.19 [1.01-1.40] | 1.29 [1.05-1.60] | 1.23 [1.08-1.40] | 724 | 538 | 396 | 242 | 633 |
| J05 | 1.02 [0.97-1.06] | 1.04 [0.84-1.28] | 1.12 [0.88-1.43] | 1.01 [0.96-1.05] | 0.98 [0.94-1.03] | 0.99 [0.95-1.04] | 0.97 [0.78-1.21] | 1.03 [0.80-1.32] | 0.98 [0.94-1.03] | 0.97 [0.92-1.02] | 4,821 | 201 | 157 | 4,640 | 4,236 |
| J06 | 1.30 [1.27-1.33] | 1.38 [1.31-1.45] | 1.36 [1.28-1.45] | 1.29 [1.26-1.32] | 1.24 [1.21-1.27] | 1.24 [1.21-1.27] | 1.18 [1.12-1.25] | 1.18 [1.11-1.26] | 1.25 [1.22-1.27] | 1.19 [1.17-1.22] | 22,016 | 3,769 | 2,499 | 18,724 | 19,142 |
| J10 | 1.35 [1.28-1.41] | 1.40 [1.33-1.47] | 1.36 [1.28-1.44] | 1.23 [1.12-1.35] | 1.28 [1.22-1.34] | 1.22 [1.16-1.28] | 1.19 [1.13-1.26] | 1.17 [1.10-1.24] | 1.15 [1.04-1.26] | 1.18 [1.13-1.24] | 4,807 | 3,923 | 3,220 | 1,161 | 4,498 |
| J11 | 1.35 [1.22-1.49] | 1.34 [1.20-1.50] | 1.35 [1.19-1.54] | 1.18 [0.97-1.44] | 1.26 [1.13-1.40] | 1.24 [1.12-1.38] | 1.17 [1.04-1.32] | 1.20 [1.05-1.37] | 1.13 [0.92-1.38] | 1.18 [1.06-1.32] | 1,009 | 809 | 587 | 269 | 902 |
| J12 | 1.28 [1.23-1.33] | 1.29 [1.23-1.34] | 1.31 [1.25-1.36] | 1.16 [1.02-1.32] | 1.23 [1.18-1.28] | 1.19 [1.14-1.24] | 1.15 [1.10-1.20] | 1.16 [1.11-1.21] | 1.10 [0.96-1.25] | 1.17 [1.12-1.22] | 7,080 | 6,617 | 6,240 | 592 | 6,651 |
| J13 | 1.32 [1.22-1.43] | 1.37 [1.26-1.48] | 1.36 [1.24-1.48] | 1.18 [0.90-1.53] | 1.27 [1.17-1.38] | 1.21 [1.12-1.32] | 1.20 [1.10-1.31] | 1.19 [1.09-1.30] | 1.09 [0.83-1.43] | 1.19 [1.09-1.29] | 1,584 | 1,485 | 1,350 | 141 | 1,388 |
| J14 | 1.33 [1.17-1.50] | 1.38 [1.21-1.56] | 1.35 [1.19-1.54] | | 1.23 [1.08-1.40] | 1.17 [1.03-1.33] | 1.14 [0.99-1.30] | 1.11 [0.96-1.27] | | 1.11 [0.97-1.26] | 656 | 626 | 583 | NA | 596 |
| J15 | 1.38 [1.31-1.45] | 1.42 [1.34-1.50] | 1.43 [1.35-1.52] | 1.22 [1.05-1.41] | 1.32 [1.24-1.39] | 1.26 [1.19-1.33] | 1.24 [1.17-1.32] | 1.23 [1.16-1.31] | 1.16 [1.00-1.35] | 1.23 [1.16-1.30] | 3,592 | 3,195 | 2,976 | 465 | 3,254 |
| J17 | 1.49 [1.32-1.67] | 1.49 [1.32-1.68] | 1.46 [1.29-1.65] | | 1.35 [1.20-1.53] | 1.38 [1.23-1.56] | 1.34 [1.18-1.51] | 1.31 [1.15-1.50] | | 1.28 [1.13-1.45] | 737 | 687 | 628 | NA | 661 |
| J18 | 1.27 [1.26-1.28] | 1.27 [1.26-1.28] | 1.26 [1.25-1.28] | 1.30 [1.26-1.35] | 1.21 [1.20-1.22] | 1.21 [1.20-1.22] | 1.16 [1.15-1.18] | 1.16 [1.14-1.17] | 1.25 [1.21-1.30] | 1.17 [1.16-1.18] | 97,392 | 90,121 | 86,034 | 8,531 | 88,216 |
| J20 | 1.29 [1.18-1.40] | 1.28 [1.16-1.41] | 1.26 [1.14-1.40] | 1.30 [1.09-1.56] | 1.22 [1.12-1.34] | 1.18 [1.08-1.29] | 1.12 [1.02-1.24] | 1.12 [1.00-1.24] | 1.25 [1.04-1.50] | 1.15 [1.05-1.26] | 1,357 | 1,100 | 960 | 309 | 1,246 |
| J21 | 1.14 [1.05-1.25] | 1.46 [1.22-1.75] | 1.47 [1.22-1.78] | 1.04 [0.94-1.16] | 1.09 [0.99-1.20] | 1.07 [0.97-1.17] | 1.21 [1.00-1.46] | 1.25 [1.03-1.53] | 1.00 [0.90-1.11] | 1.03 [0.93-1.13] | 1,139 | 318 | 287 | 830 | 1,033 |
| J22 | 1.36 [1.35-1.38] | 1.35 [1.33-1.37] | 1.32 [1.30-1.34] | 1.43 [1.39-1.46] | 1.28 [1.26-1.30] | 1.27 [1.26-1.29] | 1.21 [1.19-1.23] | 1.19 [1.17-1.21] | 1.35 [1.31-1.38] | 1.22 [1.20-1.23] | 68,041 | 53,382 | 47,758 | 16,442 | 61,394 |
| J30 | 2.14 [2.08-2.20] | 1.76 [1.71-1.82] | 1.67 [1.60-1.74] | 2.83 [2.72-2.95] | 2.03 [1.98-2.09] | 1.73 [1.68-1.78] | 1.32 [1.27-1.36] | 1.29 [1.23-1.35] | 2.38 [2.28-2.48] | 1.66 [1.61-1.71] | 17,750 | 10,536 | 6,066 | 9,124 | 15,193 |
| J31 | 1.75 [1.66-1.86] | 1.70 [1.59-1.81] | 1.59 [1.47-1.72] | 1.96 [1.78-2.17] | 1.62 [1.52-1.72] | 1.44 [1.35-1.53] | 1.33 [1.24-1.43] | 1.24 [1.14-1.34] | 1.68 [1.52-1.87] | 1.34 [1.26-1.43] | 3,383 | 2,421 | 1,625 | 1,259 | 2,875 |
| J32 | 1.42 [1.36-1.47] | 1.46 [1.40-1.52] | 1.42 [1.35-1.49] | 1.31 [1.20-1.44] | 1.32 [1.27-1.38] | 1.22 [1.18-1.28] | 1.20 [1.15-1.26] | 1.18 [1.13-1.25] | 1.21 [1.10-1.33] | 1.17 [1.12-1.22] | 6,587 | 5,843 | 4,146 | 1,286 | 5,596 |
| J33 | 1.56 [1.49-1.64] | 1.54 [1.47-1.62] | 1.52 [1.44-1.61] | 1.41 [1.21-1.64] | 1.42 [1.35-1.50] | 1.29 [1.23-1.36] | 1.25 [1.19-1.32] | 1.25 [1.18-1.32] | 1.21 [1.03-1.42] | 1.20 [1.14-1.27] | 4,446 | 4,224 | 3,253 | 471 | 3,366 |
| J34 | 1.51 [1.47-1.54] | 1.51 [1.46-1.55] | 1.48 [1.43-1.54] | 1.48 [1.42-1.54] | 1.39 [1.35-1.43] | 1.35 [1.31-1.38] | 1.29 [1.25-1.33] | 1.28 [1.23-1.33] | 1.37 [1.31-1.43] | 1.26 [1.23-1.30] | 16,429 | 12,145 | 6,606 | 6,409 | 12,605 |
| J35 | 1.28 [1.26-1.31] | 1.30 [1.24-1.36] | 1.23 [1.13-1.35] | 1.28 [1.25-1.30] | 1.24 [1.21-1.27] | 1.23 [1.21-1.26] | 1.17 [1.11-1.23] | 1.10 [1.01-1.21] | 1.23 [1.20-1.26] | 1.20 [1.17-1.23] | 25,871 | 4,000 | 1,120 | 23,193 | 20,766 |
| J36 | 1.14 [1.07-1.22] | 1.16 [1.08-1.24] | 1.14 [1.00-1.30] | 1.11 [1.01-1.22] | 1.07 [1.00-1.15] | 1.11 [1.04-1.18] | 1.11 [1.03-1.19] | 1.09 [0.95-1.25] | 1.10 [1.00-1.21] | 1.06 [0.99-1.15] | 2,461 | 1,982 | 525 | 1,193 | 1,763 |
| J37 | 1.39 [1.07-1.81] | 1.40 [1.07-1.84] | 1.31 [0.98-1.75] | | 1.19 [0.90-1.58] | 1.24 [0.95-1.63] | 1.15 [0.86-1.53] | 1.10 [0.81-1.49] | | 1.09 [0.82-1.45] | 143 | 129 | 108 | NA | 118 |
| J38 | 1.30 [1.24-1.36] | 1.31 [1.25-1.38] | 1.30 [1.24-1.37] | 1.23 [1.08-1.39] | 1.21 [1.15-1.27] | 1.21 [1.15-1.27] | 1.15 [1.10-1.21] | 1.16 [1.10-1.22] | 1.12 [0.99-1.28] | 1.15 [1.09-1.21] | 4,666 | 4,151 | 3,615 | 666 | 4,048 |
| J39 | 1.33 [1.25-1.42] | 1.34 [1.25-1.44] | 1.32 [1.23-1.43] | 1.27 [1.11-1.45] | 1.20 [1.12-1.28] | 1.23 [1.15-1.31] | 1.18 [1.10-1.26] | 1.18 [1.09-1.27] | 1.17 [1.02-1.34] | 1.12 [1.05-1.20] | 2,592 | 2,111 | 1,790 | 588 | 2,276 |
| J40 | 1.44 [1.36-1.53] | 1.44 [1.36-1.53] | 1.45 [1.36-1.54] | 1.32 [0.96-1.82] | 1.30 [1.23-1.39] | 1.29 [1.21-1.37] | 1.23 [1.16-1.31] | 1.25 [1.17-1.33] | 1.24 [0.89-1.73] | 1.19 [1.12-1.27] | 2,869 | 2,824 | 2,580 | 108 | 2,545 |
| J41 | 1.44 [1.09-1.89] | 1.38 [1.05-1.81] | 1.36 [1.03-1.81] | | 1.18 [0.88-1.57] | 1.32 [0.99-1.74] | 1.18 [0.88-1.57] | 1.18 [0.88-1.58] | | 1.13 [0.84-1.52] | 131 | 130 | 120 | NA | 112 |
| J42 | 1.45 [1.32-1.60] | 1.42 [1.29-1.56] | 1.42 [1.29-1.56] | | 1.31 [1.19-1.44] | 1.28 [1.16-1.41] | 1.19 [1.08-1.32] | 1.18 [1.07-1.31] | | 1.17 [1.05-1.29] | 1,161 | 1,122 | 1,077 | NA | 1,039 |
| J43 | 1.35 [1.32-1.39] | 1.35 [1.32-1.39] | 1.36 [1.32-1.39] | 1.21 [0.93-1.57] | 1.28 [1.25-1.32] | 1.25 [1.22-1.29] | 1.18 [1.15-1.21] | 1.18 [1.15-1.22] | 1.17 [0.89-1.53] | 1.21 [1.18-1.25] | 15,001 | 14,949 | 14,688 | 151 | 13,692 |
| J44 | 1.46 [1.44-1.48] | 1.45 [1.44-1.47] | 1.45 [1.43-1.47] | 1.81 [1.49-2.20] | 1.36 [1.34-1.38] | 1.29 [1.27-1.31] | 1.20 [1.19-1.22] | 1.21 [1.19-1.22] | 1.54 [1.26-1.89] | 1.22 [1.21-1.24] | 63,230 | 63,040 | 62,368 | 310 | 55,206 |
| J45 | 2.12 [2.11-2.14] | 1.98 [1.96-2.00] | 1.84 [1.82-1.87] | 2.50 [2.46-2.53] | 1.93 [1.91-1.95] | 1.61 [1.59-1.62] | 1.34 [1.33-1.36] | 1.31 [1.29-1.33] | 2.06 [2.02-2.09] | 1.51 [1.50-1.53] | 165,066 | 117,603 | 75,502 | 63,223 | 122,304 |
| J46 | 2.95 [2.79-3.13] | 2.46 [2.25-2.68] | 2.08 [1.85-2.33] | 3.34 [3.10-3.60] | 2.59 [2.43-2.76] | 2.29 [2.15-2.44] | 1.55 [1.40-1.72] | 1.35 [1.18-1.54] | 2.76 [2.55-2.99] | 2.01 [1.88-2.15] | 3,968 | 1,559 | 809 | 2,673 | 3,324 |
| J47 | 1.49 [1.45-1.54] | 1.51 [1.47-1.56] | 1.50 [1.45-1.55] | 1.29 [1.06-1.58] | 1.39 [1.35-1.44] | 1.29 [1.25-1.33] | 1.27 [1.23-1.31] | 1.26 [1.22-1.30] | 1.15 [0.93-1.42] | 1.22 [1.18-1.26] | 11,900 | 11,734 | 11,381 | 269 | 11,013 |
| J61 | 1.39 [1.28-1.52] | 1.38 [1.27-1.50] | 1.40 [1.28-1.52] | | 1.32 [1.21-1.43] | 1.32 [1.21-1.43] | 1.27 [1.16-1.38] | 1.27 [1.17-1.38] | | 1.25 [1.15-1.37] | 1,512 | 1,513 | 1,512 | NA | 1,416 |
| J67 | 1.61 [1.39-1.86] | 1.60 [1.38-1.85] | 1.59 [1.37-1.84] | | 1.44 [1.24-1.67] | 1.44 [1.24-1.67] | 1.38 [1.19-1.61] | 1.40 [1.20-1.63] | | 1.30 [1.11-1.52] | 497 | 492 | 464 | NA | 442 |
| J69 | 1.17 [1.14-1.20] | 1.18 [1.14-1.21] | 1.18 [1.14-1.21] | 0.93 [0.78-1.10] | 1.12 [1.09-1.16] | 1.15 [1.12-1.18] | 1.12 [1.09-1.16] | 1.12 [1.09-1.15] | 0.90 [0.76-1.07] | 1.13 [1.10-1.16] | 13,535 | 13,347 | 12,953 | 323 | 12,687 |
| J70 | 1.52 [1.27-1.82] | 1.44 [1.21-1.73] | 1.51 [1.25-1.81] | | 1.35 [1.13-1.62] | 1.42 [1.18-1.70] | 1.27 [1.06-1.53] | 1.36 [1.13-1.64] | | 1.28 [1.06-1.54] | 321 | 309 | 303 | NA | 300 |
| J80 | 1.37 [1.25-1.51] | 1.40 [1.27-1.54] | 1.45 [1.31-1.60] | | 1.27 [1.15-1.40] | 1.31 [1.19-1.44] | 1.27 [1.15-1.40] | 1.31 [1.18-1.45] | | 1.25 [1.13-1.38] | 1,168 | 1,119 | 1,008 | NA | 1,042 |
| J81 | 1.30 [1.24-1.36] | 1.29 [1.23-1.35] | 1.31 [1.25-1.37] | 0.97 [0.72-1.31] | 1.19 [1.14-1.25] | 1.25 [1.20-1.31] | 1.18 [1.13-1.24] | 1.21 [1.15-1.27] | 0.96 [0.70-1.30] | 1.17 [1.11-1.23] | 4,916 | 4,853 | 4,709 | 107 | 4,416 |
| J82 | 2.40 [2.08-2.78] | 2.31 [2.00-2.68] | 2.11 [1.80-2.47] | | 2.24 [1.93-2.59] | 1.72 [1.47-2.02] | 1.55 [1.31-1.82] | 1.49 [1.25-1.77] | | 1.69 [1.44-1.98] | 600 | 574 | 451 | NA | 570 |
| J84 | 1.44 [1.39-1.49] | 1.45 [1.41-1.50] | 1.44 [1.40-1.49] | | 1.37 [1.32-1.42] | 1.36 [1.31-1.41] | 1.34 [1.29-1.38] | 1.33 [1.28-1.37] | | 1.32 [1.27-1.36] | 9,901 | 9,866 | 9,723 | NA | 9,106 |
| J85 | 1.31 [1.14-1.51] | 1.37 [1.19-1.59] | 1.37 [1.18-1.59] | | 1.23 [1.06-1.42] | 1.21 [1.05-1.39] | 1.23 [1.06-1.43] | 1.22 [1.05-1.43] | | 1.18 [1.01-1.37] | 509 | 469 | 429 | NA | 442 |
| J86 | 1.20 [1.12-1.30] | 1.24 [1.14-1.35] | 1.25 [1.15-1.36] | 1.07 [0.89-1.30] | 1.13 [1.05-1.23] | 1.17 [1.08-1.26] | 1.16 [1.07-1.27] | 1.16 [1.07-1.27] | 1.10 [0.91-1.33] | 1.13 [1.04-1.23] | 1,711 | 1,474 | 1,351 | 269 | 1,449 |
| J90 | 1.19 [1.17-1.21] | 1.19 [1.17-1.21] | 1.20 [1.18-1.22] | 1.11 [1.01-1.21] | 1.14 [1.12-1.16] | 1.16 [1.14-1.18] | 1.12 [1.10-1.14] | 1.12 [1.10-1.14] | 1.11 [1.02-1.22] | 1.12 [1.11-1.14] | 44,263 | 43,369 | 41,910 | 1,336 | 40,046 |
| J91 | 1.24 [1.12-1.37] | 1.25 [1.13-1.38] | 1.21 [1.09-1.34] | | 1.20 [1.09-1.34] | 1.21 [1.09-1.33] | 1.17 [1.06-1.30] | 1.13 [1.02-1.25] | | 1.19 [1.08-1.32] | 1,085 | 1,059 | 1,021 | NA | 984 |
| J92 | 1.26 [1.20-1.31] | 1.28 [1.22-1.34] | 1.26 [1.20-1.31] | | 1.21 [1.15-1.27] | 1.22 [1.16-1.27] | 1.21 [1.15-1.26] | 1.18 [1.13-1.24] | | 1.19 [1.13-1.25] | 5,072 | 5,066 | 5,035 | NA | 4,697 |
| J93 | 1.15 [1.09-1.20] | 1.14 [1.08-1.20] | 1.17 [1.10-1.23] | 1.05 [0.93-1.20] | 1.08 [1.02-1.14] | 1.12 [1.06-1.17] | 1.07 [1.01-1.12] | 1.10 [1.04-1.16] | 1.06 [0.93-1.21] | 1.07 [1.01-1.12] | 4,150 | 3,817 | 3,221 | 627 | 3,472 |
| J94 | 1.18 [1.09-1.28] | 1.18 [1.09-1.28] | 1.20 [1.11-1.30] | | 1.09 [1.01-1.18] | 1.15 [1.07-1.25] | 1.10 [1.02-1.19] | 1.12 [1.03-1.22] | | 1.08 [0.99-1.17] | 1,641 | 1,604 | 1,549 | NA | 1,469 |
| J95 | 1.21 [1.11-1.31] | 1.20 [1.10-1.31] | 1.24 [1.13-1.35] | 0.98 [0.75-1.27] | 1.11 [1.01-1.21] | 1.17 [1.08-1.27] | 1.11 [1.02-1.22] | 1.16 [1.06-1.27] | 0.91 [0.70-1.20] | 1.10 [1.00-1.20] | 1,356 | 1,249 | 1,173 | 134 | 1,156 |
| J96 | 1.32 [1.30-1.34] | 1.31 [1.29-1.34] | 1.30 [1.28-1.33] | 1.21 [1.07-1.36] | 1.24 [1.22-1.26] | 1.23 [1.21-1.25] | 1.17 [1.14-1.19] | 1.16 [1.13-1.18] | 1.12 [0.99-1.26] | 1.18 [1.16-1.21] | 30,130 | 29,782 | 28,727 | 709 | 27,859 |
| J98 | 1.25 [1.23-1.28] | 1.24 [1.22-1.26] | 1.23 [1.21-1.26] | 1.25 [1.17-1.33] | 1.18 [1.16-1.21] | 1.17 [1.15-1.20] | 1.12 [1.09-1.14] | 1.11 [1.09-1.13] | 1.17 [1.10-1.25] | 1.14 [1.11-1.16] | 28,140 | 26,101 | 24,543 | 2,583 | 25,833 |
| J99 | 1.60 [1.45-1.77] | 1.66 [1.50-1.83] | 1.71 [1.54-1.91] | | 1.44 [1.30-1.60] | 1.39 [1.25-1.54] | 1.41 [1.27-1.57] | 1.47 [1.32-1.64] | | 1.28 [1.15-1.42] | 1,026 | 987 | 897 | NA | 950 |
| Diseases of the digestive system (K00-K95) | | | | | | | | | | | | | | | |
| K00 | 1.16 [1.12-1.21] | 1.15 [1.06-1.25] | 1.22 [1.02-1.47] | 1.14 [1.09-1.19] | 1.09 [1.05-1.15] | 1.12 [1.08-1.17] | 1.05 [0.96-1.14] | 1.12 [0.93-1.35] | 1.11 [1.06-1.16] | 1.07 [1.02-1.13] | 6,500 | 1,359 | 276 | 5,703 | 4,997 |
| K01 | 1.21 [1.18-1.25] | 1.23 [1.19-1.27] | 1.15 [1.06-1.24] | 1.18 [1.14-1.22] | 1.13 [1.10-1.17] | 1.16 [1.13-1.19] | 1.13 [1.10-1.17] | 1.10 [1.02-1.19] | 1.14 [1.10-1.18] | 1.10 [1.06-1.13] | 14,936 | 9,240 | 1,484 | 8,671 | 10,238 |
| K02 | 1.11 [1.09-1.13] | 1.25 [1.22-1.28] | 1.28 [1.24-1.32] | 1.02 [1.00-1.04] | 1.05 [1.03-1.07] | 1.07 [1.05-1.09] | 1.12 [1.10-1.15] | 1.15 [1.11-1.19] | 1.01 [0.99-1.03] | 1.03 [1.01-1.05] | 42,689 | 17,603 | 8,465 | 28,068 | 34,584 |
| K03 | 1.17 [1.07-1.29] | 1.46 [1.27-1.68] | 1.42 [1.18-1.72] | 1.00 [0.89-1.13] | 1.11 [1.00-1.23] | 1.12 [1.01-1.23] | 1.34 [1.16-1.56] | 1.27 [1.05-1.54] | 0.98 [0.86-1.11] | 1.07 [0.97-1.19] | 1,129 | 494 | 271 | 736 | 995 |
| K04 | 1.14 [1.10-1.18] | 1.22 [1.17-1.27] | 1.25 [1.19-1.32] | 1.00 [0.95-1.05] | 1.07 [1.03-1.11] | 1.09 [1.05-1.13] | 1.10 [1.05-1.15] | 1.13 [1.07-1.19] | 0.99 [0.94-1.05] | 1.04 [1.00-1.08] | 8,999 | 5,838 | 3,471 | 3,950 | 7,404 |
| K05 | 1.29 [1.25-1.34] | 1.33 [1.28-1.38] | 1.33 [1.24-1.42] | 1.21 [1.14-1.29] | 1.19 [1.15-1.24] | 1.22 [1.17-1.26] | 1.20 [1.15-1.24] | 1.21 [1.13-1.29] | 1.16 [1.09-1.24] | 1.15 [1.10-1.19] | 8,466 | 7,996 | 2,186 | 2,842 | 6,400 |
| K06 | 1.38 [1.26-1.52] | 1.37 [1.23-1.51] | 1.37 [1.22-1.54] | 1.19 [0.98-1.44] | 1.24 [1.12-1.37] | 1.31 [1.19-1.44] | 1.27 [1.15-1.42] | 1.25 [1.11-1.40] | 1.12 [0.92-1.37] | 1.19 [1.07-1.32] | 1,145 | 937 | 728 | 279 | 970 |
| K07 | 1.19 [1.15-1.23] | 1.22 [1.16-1.28] | 1.29 [1.17-1.42] | 1.15 [1.10-1.19] | 1.12 [1.08-1.17] | 1.14 [1.10-1.18] | 1.11 [1.05-1.17] | 1.14 [1.03-1.27] | 1.11 [1.07-1.16] | 1.09 [1.05-1.14] | 9,242 | 3,771 | 948 | 6,972 | 6,750 |
| K08 | 1.19 [1.15-1.22] | 1.25 [1.21-1.29] | 1.29 [1.24-1.34] | 1.04 [0.99-1.09] | 1.12 [1.08-1.15] | 1.13 [1.10-1.16] | 1.14 [1.10-1.17] | 1.15 [1.11-1.20] | 1.02 [0.98-1.07] | 1.08 [1.05-1.12] | 12,768 | 9,116 | 5,826 | 4,744 | 10,589 |
| K09 | 1.26 [1.14-1.40] | 1.26 [1.13-1.41] | 1.13 [0.99-1.30] | 1.23 [1.02-1.48] | 1.17 [1.05-1.32] | 1.24 [1.12-1.38] | 1.18 [1.05-1.33] | 1.08 [0.94-1.24] | 1.19 [0.98-1.44] | 1.15 [1.02-1.29] | 971 | 742 | 489 | 311 | 725 |
| K10 | 1.25 [1.14-1.36] | 1.27 [1.15-1.39] | 1.27 [1.15-1.41] | 1.26 [1.02-1.57] | 1.15 [1.04-1.26] | 1.15 [1.05-1.26] | 1.11 [1.01-1.22] | 1.12 [1.01-1.25] | 1.21 [0.97-1.52] | 1.10 [1.00-1.21] | 1,235 | 1,114 | 877 | 227 | 1,102 |
| K11 | 1.31 [1.24-1.39] | 1.36 [1.28-1.45] | 1.36 [1.27-1.46] | 1.16 [1.04-1.30] | 1.24 [1.16-1.32] | 1.24 [1.17-1.32] | 1.25 [1.17-1.33] | 1.23 [1.15-1.33] | 1.12 [1.00-1.25] | 1.19 [1.12-1.27] | 3,061 | 2,405 | 1,914 | 848 | 2,583 |
| K12 | 1.29 [1.24-1.35] | 1.33 [1.26-1.40] | 1.35 [1.28-1.44] | 1.19 [1.10-1.29] | 1.23 [1.17-1.29] | 1.24 [1.19-1.30] | 1.22 [1.16-1.29] | 1.25 [1.18-1.33] | 1.17 [1.08-1.27] | 1.20 [1.14-1.26] | 4,859 | 3,595 | 2,811 | 1,596 | 4,287 |

#### Supplementary Table 3: Hazard ratios and events from all cohorts

| | Hazard ratio (99% confidence interval) | | | | | | | | | | | | | | |
|---|---|---|---|---|---|---|---|---|---|---|---|---|---|---|---|
| | crude | | | | | adjusted | | | | | Events (in exposed) | | | | |
| Outcome | any age | 18+ | 40+ | <18 | hosp. | any age | 18+ | 40+ | <18 | hosp. | any age | 18+ | 40+ | <18 | hosp. |
| K13 | 1.37 [1.31-1.42] | 1.38 [1.32-1.44] | 1.38 [1.32-1.45] | 1.30 [1.20-1.41] | 1.30 [1.25-1.36] | 1.30 [1.25-1.35] | 1.26 [1.21-1.32] | 1.26 [1.20-1.32] | 1.26 [1.16-1.37] | 1.26 [1.20-1.31] | 6,539 | 5,201 | 4,172 | 1,657 | 5,395 |
| K14 | 1.39 [1.31-1.47] | 1.40 [1.32-1.49] | 1.37 [1.29-1.46] | 1.32 [1.14-1.53] | 1.29 [1.21-1.37] | 1.28 [1.21-1.36] | 1.24 [1.17-1.32] | 1.23 [1.15-1.31] | 1.26 [1.09-1.47] | 1.21 [1.14-1.29] | 3,114 | 2,746 | 2,364 | 495 | 2,705 |
| K20 | 1.33 [1.30-1.36] | 1.32 [1.29-1.36] | 1.30 [1.27-1.34] | 1.49 [1.35-1.64] | 1.23 [1.20-1.26] | 1.22 [1.19-1.25] | 1.18 [1.15-1.20] | 1.16 [1.13-1.19] | 1.34 [1.21-1.49] | 1.15 [1.12-1.18] | 18,438 | 17,762 | 16,047 | 1,158 | 15,910 |
| K21 | 1.36 [1.34-1.37] | 1.36 [1.35-1.38] | 1.36 [1.34-1.37] | 1.23 [1.18-1.29] | 1.25 [1.23-1.26] | 1.21 [1.20-1.23] | 1.18 [1.17-1.19] | 1.17 [1.16-1.19] | 1.11 [1.06-1.17] | 1.15 [1.13-1.16] | 73,030 | 70,858 | 62,008 | 4,889 | 64,783 |
| K22 | 1.29 [1.27-1.32] | 1.30 [1.27-1.32] | 1.29 [1.26-1.32] | 1.21 [1.10-1.33] | 1.21 [1.18-1.23] | 1.21 [1.18-1.23] | 1.18 [1.16-1.20] | 1.17 [1.15-1.20] | 1.12 [1.01-1.24] | 1.15 [1.13-1.17] | 26,465 | 25,926 | 24,113 | 1,101 | 23,013 |
| K23 | 1.54 [1.30-1.84] | 1.50 [1.26-1.78] | 1.49 [1.24-1.78] | | 1.47 [1.23-1.75] | 1.38 [1.16-1.65] | 1.23 [1.02-1.47] | 1.27 [1.05-1.52] | | 1.33 [1.11-1.60] | 347 | 345 | 330 | NA | 325 |
| K25 | 1.29 [1.26-1.33] | 1.30 [1.26-1.33] | 1.29 [1.25-1.32] | 1.30 [1.10-1.54] | 1.21 [1.17-1.24] | 1.22 [1.18-1.25] | 1.17 [1.14-1.20] | 1.17 [1.13-1.20] | 1.27 [1.07-1.52] | 1.16 [1.13-1.20] | 13,006 | 12,851 | 12,008 | 368 | 11,348 |
| K26 | 1.19 [1.15-1.23] | 1.21 [1.17-1.25] | 1.20 [1.16-1.24] | 1.21 [0.99-1.48] | 1.15 [1.11-1.19] | 1.16 [1.12-1.20] | 1.14 [1.10-1.18] | 1.13 [1.09-1.17] | 1.18 [0.95-1.46] | 1.14 [1.10-1.18] | 8,838 | 8,712 | 8,214 | 259 | 7,495 |
| K27 | 1.29 [1.20-1.40] | 1.28 [1.19-1.39] | 1.29 [1.19-1.40] | | 1.18 [1.09-1.28] | 1.20 [1.10-1.30] | 1.13 [1.04-1.22] | 1.15 [1.05-1.25] | | 1.13 [1.04-1.23] | 1,584 | 1,563 | 1,448 | NA | 1,439 |
| K28 | 1.47 [1.20-1.79] | 1.46 [1.19-1.79] | 1.38 [1.11-1.72] | | 1.43 [1.17-1.75] | 1.24 [1.00-1.52] | 1.19 [0.96-1.47] | 1.07 [0.84-1.35] | | 1.29 [1.05-1.59] | 255 | 248 | 201 | NA | 247 |
| K29 | 1.32 [1.31-1.34] | 1.33 [1.31-1.35] | 1.33 [1.31-1.35] | 1.23 [1.17-1.28] | 1.22 [1.21-1.24] | 1.22 [1.21-1.24] | 1.19 [1.17-1.20] | 1.19 [1.17-1.20] | 1.16 [1.11-1.22] | 1.16 [1.15-1.18] | 69,990 | 66,962 | 59,204 | 5,411 | 60,277 |
| K30 | 1.36 [1.33-1.39] | 1.36 [1.33-1.39] | 1.36 [1.32-1.39] | 1.19 [1.06-1.34] | 1.25 [1.22-1.28] | 1.24 [1.21-1.27] | 1.20 [1.17-1.23] | 1.21 [1.17-1.24] | 1.12 [0.99-1.26] | 1.16 [1.13-1.20] | 16,797 | 16,597 | 14,037 | 731 | 14,039 |
| K31 | 1.30 [1.27-1.33] | 1.31 [1.28-1.33] | 1.30 [1.27-1.33] | 1.11 [0.97-1.26] | 1.21 [1.18-1.23] | 1.18 [1.16-1.21] | 1.15 [1.12-1.17] | 1.14 [1.12-1.17] | 1.04 [0.91-1.19] | 1.13 [1.10-1.15] | 22,728 | 22,438 | 21,097 | 633 | 20,728 |
| K35 | 1.08 [1.05-1.11] | 1.09 [1.05-1.13] | 1.05 [1.00-1.11] | 1.04 [1.00-1.08] | 1.01 [0.98-1.04] | 1.04 [1.01-1.07] | 1.03 [1.00-1.07] | 1.01 [0.95-1.06] | 1.02 [0.98-1.06] | 0.99 [0.96-1.03] | 13,415 | 7,735 | 3,340 | 7,734 | 9,842 |
| K36 | 1.17 [0.98-1.40] | 1.20 [0.97-1.48] | | 1.35 [1.03-1.76] | 1.15 [0.94-1.40] | 1.15 [0.96-1.38] | 1.11 [0.89-1.38] | | 1.36 [1.03-1.79] | 1.12 [0.91-1.37] | 297 | 201 | NA | 148 | 237 |
| K37 | 1.14 [1.07-1.21] | 1.13 [1.04-1.22] | 1.15 [1.01-1.32] | 1.08 [0.99-1.17] | 1.06 [0.99-1.13] | 1.07 [1.01-1.14] | 1.01 [0.93-1.10] | 1.07 [0.93-1.23] | 1.04 [0.95-1.13] | 1.02 [0.95-1.09] | 2,653 | 1,504 | 518 | 1,617 | 2,057 |
| K38 | 1.15 [1.07-1.24] | 1.16 [1.06-1.27] | 1.10 [0.97-1.25] | 1.08 [0.97-1.21] | 1.06 [0.98-1.15] | 1.10 [1.01-1.18] | 1.06 [0.97-1.16] | 1.05 [0.92-1.19] | 1.05 [0.94-1.18] | 1.04 [0.95-1.13] | 1,735 | 1,171 | 581 | 837 | 1,394 |
| K40 | 1.10 [1.07-1.12] | 1.11 [1.09-1.14] | 1.11 [1.08-1.13] | 1.02 [0.96-1.08] | 1.06 [1.04-1.09] | 1.06 [1.04-1.08] | 1.08 [1.06-1.10] | 1.07 [1.05-1.10] | 0.98 [0.93-1.04] | 1.04 [1.02-1.06] | 23,888 | 21,407 | 19,508 | 3,081 | 18,644 |
| K41 | 1.19 [1.10-1.28] | 1.18 [1.09-1.27] | 1.16 [1.08-1.26] | | 1.12 [1.03-1.21] | 1.13 [1.05-1.22] | 1.12 [1.04-1.21] | 1.10 [1.02-1.19] | | 1.07 [0.98-1.16] | 1,699 | 1,670 | 1,555 | NA | 1,454 |
| K42 | 1.29 [1.25-1.32] | 1.29 [1.26-1.33] | 1.29 [1.25-1.34] | 1.19 [1.10-1.28] | 1.20 [1.17-1.24] | 1.17 [1.14-1.21] | 1.14 [1.11-1.18] | 1.14 [1.10-1.17] | 1.09 [1.00-1.18] | 1.13 [1.09-1.16] | 11,993 | 10,583 | 8,923 | 1,665 | 10,301 |
| K43 | 1.26 [1.22-1.30] | 1.27 [1.23-1.32] | 1.28 [1.24-1.33] | 1.13 [1.00-1.28] | 1.18 [1.14-1.22] | 1.16 [1.12-1.20] | 1.14 [1.10-1.18] | 1.14 [1.10-1.18] | 1.05 [0.92-1.20] | 1.12 [1.09-1.16] | 8,989 | 8,524 | 7,619 | 615 | 8,303 |
| K44 | 1.31 [1.29-1.32] | 1.32 [1.30-1.33] | 1.30 [1.29-1.32] | 1.27 [1.17-1.38] | 1.22 [1.20-1.23] | 1.20 [1.18-1.21] | 1.17 [1.16-1.19] | 1.16 [1.15-1.18] | 1.17 [1.08-1.28] | 1.14 [1.12-1.15] | 66,300 | 66,090 | 60,673 | 1,682 | 57,225 |
| K45 | 1.22 [1.04-1.42] | 1.18 [1.01-1.38] | 1.20 [1.02-1.41] | | 1.12 [0.96-1.32] | 1.12 [0.95-1.31] | 1.04 [0.88-1.22] | 1.07 [0.90-1.26] | | 1.04 [0.88-1.23] | 407 | 401 | 361 | NA | 378 |
| K46 | 1.31 [1.21-1.41] | 1.34 [1.24-1.45] | 1.35 [1.25-1.46] | | 1.20 [1.11-1.30] | 1.18 [1.09-1.28] | 1.16 [1.07-1.25] | 1.17 [1.08-1.27] | | 1.12 [1.04-1.21] | 1,772 | 1,736 | 1,636 | NA | 1,684 |
| K50 | 1.70 [1.63-1.77] | 1.65 [1.58-1.72] | 1.56 [1.47-1.64] | 1.80 [1.65-1.96] | 1.55 [1.48-1.62] | 1.47 [1.41-1.54] | 1.35 [1.29-1.42] | 1.29 [1.22-1.36] | 1.67 [1.53-1.82] | 1.38 [1.32-1.45] | 6,324 | 5,403 | 3,344 | 1,728 | 5,217 |
| K51 | 1.51 [1.46-1.56] | 1.51 [1.46-1.57] | 1.45 [1.39-1.51] | 1.44 [1.31-1.58] | 1.36 [1.31-1.42] | 1.35 [1.30-1.40] | 1.29 [1.24-1.34] | 1.24 [1.19-1.30] | 1.36 [1.23-1.50] | 1.25 [1.20-1.30] | 8,057 | 7,573 | 5,375 | 1,254 | 6,659 |
| K52 | 1.35 [1.33-1.38] | 1.39 [1.36-1.41] | 1.36 [1.34-1.39] | 1.22 [1.18-1.27] | 1.25 [1.23-1.27] | 1.27 [1.25-1.29] | 1.24 [1.22-1.26] | 1.23 [1.21-1.25] | 1.19 [1.14-1.23] | 1.20 [1.18-1.22] | 41,353 | 35,695 | 29,410 | 7,425 | 34,827 |
| K55 | 1.25 [1.20-1.30] | 1.25 [1.20-1.30] | 1.25 [1.20-1.31] | | 1.15 [1.10-1.21] | 1.19 [1.14-1.24] | 1.14 [1.10-1.20] | 1.14 [1.09-1.19] | | 1.13 [1.08-1.19] | 5,570 | 5,532 | 5,358 | NA | 5,112 |
| K56 | 1.21 [1.18-1.24] | 1.21 [1.18-1.24] | 1.20 [1.17-1.23] | 1.08 [0.99-1.18] | 1.14 [1.11-1.17] | 1.15 [1.12-1.17] | 1.11 [1.09-1.14] | 1.11 [1.08-1.14] | 1.04 [0.95-1.14] | 1.10 [1.07-1.13] | 17,069 | 16,153 | 14,775 | 1,354 | 15,433 |
| K57 | 1.25 [1.24-1.27] | 1.25 [1.23-1.26] | 1.25 [1.23-1.26] | 1.15 [0.93-1.43] | 1.17 [1.15-1.18] | 1.17 [1.16-1.19] | 1.14 [1.12-1.15] | 1.14 [1.12-1.15] | 1.06 [0.84-1.32] | 1.11 [1.10-1.13] | 72,988 | 72,950 | 71,810 | 235 | 63,834 |
| K58 | 1.50 [1.47-1.53] | 1.52 [1.49-1.55] | 1.53 [1.49-1.56] | 1.29 [1.21-1.37] | 1.39 [1.36-1.42] | 1.27 [1.24-1.30] | 1.19 [1.17-1.22] | 1.20 [1.17-1.23] | 1.21 [1.14-1.29] | 1.22 [1.19-1.25] | 24,237 | 23,632 | 16,259 | 3,011 | 21,882 |
| K59 | 1.27 [1.26-1.29] | 1.29 [1.27-1.30] | 1.27 [1.26-1.29] | 1.19 [1.16-1.23] | 1.20 [1.18-1.21] | 1.20 [1.18-1.21] | 1.17 [1.15-1.18] | 1.16 [1.14-1.18] | 1.15 [1.11-1.18] | 1.15 [1.14-1.17] | 75,465 | 65,892 | 58,307 | 12,257 | 68,411 |
| K60 | 1.52 [1.47-1.58] | 1.54 [1.48-1.60] | 1.53 [1.46-1.60] | 1.40 [1.28-1.53] | 1.42 [1.37-1.48] | 1.41 [1.36-1.46] | 1.38 [1.32-1.43] | 1.37 [1.30-1.43] | 1.34 [1.22-1.47] | 1.35 [1.30-1.41] | 7,311 | 6,701 | 4,241 | 1,289 | 6,070 |
| K61 | 1.39 [1.33-1.46] | 1.39 [1.33-1.46] | 1.39 [1.31-1.49] | 1.32 [1.20-1.47] | 1.29 [1.22-1.36] | 1.32 [1.26-1.39] | 1.26 [1.19-1.32] | 1.25 [1.17-1.34] | 1.32 [1.19-1.47] | 1.26 [1.19-1.33] | 4,318 | 3,877 | 2,232 | 1,034 | 3,412 |
| K62 | 1.33 [1.31-1.35] | 1.34 [1.31-1.36] | 1.31 [1.29-1.33] | 1.34 [1.26-1.42] | 1.22 [1.20-1.25] | 1.25 [1.23-1.27] | 1.21 [1.19-1.23] | 1.19 [1.17-1.21] | 1.26 [1.18-1.35] | 1.17 [1.15-1.19] | 39,989 | 38,779 | 32,850 | 2,822 | 33,631 |
| K63 | 1.27 [1.25-1.30] | 1.27 [1.25-1.29] | 1.25 [1.23-1.27] | 1.40 [1.29-1.53] | 1.19 [1.17-1.21] | 1.19 [1.17-1.21] | 1.14 [1.12-1.16] | 1.13 [1.11-1.15] | 1.30 [1.19-1.42] | 1.14 [1.12-1.16] | 38,712 | 38,142 | 34,930 | 1,491 | 34,387 |
| K64 | 1.31 [1.29-1.34] | 1.31 [1.29-1.34] | 1.29 [1.27-1.32] | 1.34 [1.23-1.46] | 1.22 [1.20-1.25] | 1.21 [1.19-1.23] | 1.18 [1.15-1.20] | 1.16 [1.14-1.19] | 1.27 [1.16-1.39] | 1.16 [1.14-1.18] | 28,431 | 28,219 | 24,209 | 1,558 | 25,763 |
| K65 | 1.19 [1.13-1.25] | 1.18 [1.12-1.24] | 1.17 [1.11-1.23] | 1.05 [0.88-1.26] | 1.12 [1.07-1.18] | 1.16 [1.10-1.21] | 1.10 [1.05-1.16] | 1.10 [1.04-1.16] | 1.06 [0.88-1.28] | 1.11 [1.06-1.17] | 4,002 | 3,821 | 3,397 | 319 | 3,510 |
| K66 | 1.19 [1.16-1.22] | 1.20 [1.17-1.23] | 1.18 [1.14-1.21] | 1.11 [1.02-1.21] | 1.11 [1.08-1.14] | 1.12 [1.09-1.15] | 1.09 [1.06-1.12] | 1.07 [1.04-1.11] | 1.07 [0.99-1.17] | 1.08 [1.05-1.11] | 14,367 | 13,909 | 10,436 | 1,499 | 12,929 |
| K70 | 1.44 [1.38-1.51] | 1.46 [1.40-1.52] | 1.46 [1.40-1.53] | | 1.35 [1.29-1.41] | 1.37 [1.31-1.43] | 1.29 [1.23-1.35] | 1.28 [1.23-1.34] | | 1.34 [1.28-1.41] | 5,266 | 5,262 | 4,799 | NA | 4,536 |
| K71 | 1.37 [1.15-1.62] | 1.40 [1.17-1.67] | 1.52 [1.25-1.85] | | 1.18 [0.98-1.41] | 1.29 [1.08-1.54] | 1.28 [1.07-1.54] | 1.39 [1.14-1.70] | | 1.16 [0.97-1.39] | 337 | 324 | 261 | NA | 300 |
| K72 | 1.36 [1.28-1.43] | 1.35 [1.28-1.43] | 1.40 [1.32-1.48] | 0.89 [0.67-1.18] | 1.30 [1.23-1.38] | 1.28 [1.21-1.36] | 1.22 [1.15-1.29] | 1.25 [1.18-1.33] | 0.88 [0.66-1.17] | 1.27 [1.20-1.35] | 3,232 | 3,161 | 2,960 | 125 | 2,923 |
| K73 | 1.48 [1.25-1.75] | 1.55 [1.30-1.84] | 1.51 [1.25-1.82] | | 1.40 [1.17-1.67] | 1.39 [1.16-1.65] | 1.41 [1.18-1.69] | 1.36 [1.12-1.64] | | 1.32 [1.10-1.59] | 334 | 319 | 276 | NA | 299 |
| K74 | 1.46 [1.39-1.52] | 1.49 [1.42-1.55] | 1.49 [1.42-1.56] | | 1.37 [1.31-1.43] | 1.35 [1.29-1.42] | 1.31 [1.25-1.37] | 1.30 [1.24-1.37] | | 1.32 [1.26-1.38] | 5,049 | 5,005 | 4,744 | NA | 4,649 |
| K75 | 1.40 [1.33-1.48] | 1.42 [1.34-1.50] | 1.41 [1.33-1.49] | 1.33 [1.07-1.66] | 1.33 [1.26-1.41] | 1.30 [1.23-1.38] | 1.26 [1.19-1.33] | 1.25 [1.18-1.33] | 1.29 [1.03-1.63] | 1.28 [1.20-1.35] | 3,307 | 3,184 | 2,850 | 231 | 3,003 |
| K76 | 1.33 [1.31-1.36] | 1.34 [1.31-1.36] | 1.33 [1.30-1.36] | 1.12 [1.00-1.24] | 1.25 [1.22-1.28] | 1.23 [1.20-1.25] | 1.17 [1.14-1.19] | 1.16 [1.14-1.19] | 1.07 [0.96-1.20] | 1.19 [1.16-1.21] | 22,874 | 22,511 | 20,279 | 956 | 20,897 |
| K80 | 1.16 [1.14-1.18] | 1.17 [1.15-1.19] | 1.15 [1.13-1.17] | 1.05 [0.97-1.12] | 1.10 [1.08-1.12] | 1.10 [1.09-1.12] | 1.08 [1.06-1.09] | 1.07 [1.05-1.08] | 1.02 [0.95-1.10] | 1.06 [1.04-1.08] | 39,352 | 39,031 | 32,181 | 1,957 | 34,401 |
| K81 | 1.19 [1.15-1.23] | 1.19 [1.15-1.24] | 1.19 [1.14-1.24] | 1.06 [0.89-1.26] | 1.13 [1.09-1.18] | 1.12 [1.08-1.17] | 1.09 [1.05-1.13] | 1.09 [1.04-1.13] | 1.03 [0.86-1.24] | 1.10 [1.05-1.14] | 7,288 | 7,250 | 6,041 | 326 | 6,430 |
| K82 | 1.18 [1.13-1.23] | 1.19 [1.14-1.24] | 1.18 [1.13-1.24] | 0.99 [0.84-1.17] | 1.11 [1.06-1.16] | 1.11 [1.07-1.16] | 1.09 [1.04-1.14] | 1.08 [1.03-1.14] | 0.96 [0.81-1.15] | 1.07 [1.02-1.12] | 5,528 | 5,479 | 4,323 | 344 | 4,861 |
| K83 | 1.16 [1.12-1.19] | 1.17 [1.13-1.20] | 1.16 [1.12-1.20] | 1.05 [0.89-1.23] | 1.12 [1.09-1.16] | 1.11 [1.08-1.15] | 1.08 [1.05-1.12] | 1.07 [1.04-1.11] | 1.01 [0.85-1.19] | 1.10 [1.06-1.14] | 10,270 | 10,156 | 9,035 | 390 | 9,314 |
| K85 | 1.20 [1.16-1.25] | 1.21 [1.17-1.26] | 1.19 [1.14-1.24] | 1.03 [0.89-1.18] | 1.13 [1.09-1.18] | 1.14 [1.10-1.18] | 1.11 [1.07-1.15] | 1.08 [1.04-1.13] | 1.02 [0.88-1.18] | 1.10 [1.06-1.15] | 7,005 | 6,863 | 5,652 | 497 | 6,079 |
| K86 | 1.21 [1.17-1.27] | 1.24 [1.19-1.29] | 1.23 [1.18-1.29] | 0.86 [0.70-1.05] | 1.14 [1.10-1.19] | 1.15 [1.10-1.19] | 1.12 [1.07-1.17] | 1.11 [1.06-1.16] | 0.84 [0.69-1.03] | 1.11 [1.07-1.16] | 5,937 | 5,821 | 5,358 | 244 | 5,452 |
| K90 | 1.78 [1.72-1.85] | 1.60 [1.53-1.67] | 1.61 [1.53-1.70] | 2.04 [1.93-2.17] | 1.67 [1.61-1.73] | 1.56 [1.50-1.62] | 1.36 [1.30-1.42] | 1.37 [1.30-1.44] | 1.76 [1.65-1.87] | 1.49 [1.43-1.55] | 8,443 | 5,443 | 3,833 | 3,599 | 7,356 |
| K91 | 1.29 [1.24-1.35] | 1.32 [1.26-1.37] | 1.28 [1.22-1.34] | 1.06 [0.93-1.21] | 1.19 [1.14-1.24] | 1.20 [1.15-1.25] | 1.18 [1.13-1.23] | 1.16 [1.11-1.22] | 0.99 [0.86-1.13] | 1.13 [1.09-1.18] | 5,721 | 5,347 | 4,601 | 581 | 5,203 |
| K92 | 1.28 [1.27-1.30] | 1.30 [1.28-1.31] | 1.28 [1.26-1.30] | 1.19 [1.14-1.25] | 1.20 [1.18-1.22] | 1.21 [1.19-1.23] | 1.17 [1.16-1.19] | 1.16 [1.14-1.18] | 1.13 [1.08-1.19] | 1.16 [1.14-1.17] | 49,810 | 47,775 | 40,587 | 4,646 | 44,150 |
| Diseases of the skin and subcutaneous tissue (L00-L99) | | | | | | | | | | | | | | | |
| L00 | 1.54 [1.22-1.93] | | | 1.55 [1.23-1.94] | 1.36 [1.04-1.77] | 1.54 [1.22-1.93] | | | 1.54 [1.22-1.94] | 1.33 [1.01-1.74] | 205 | NA | NA | 195 | 145 |
| L01 | 2.28 [2.08-2.51] | 2.65 [2.21-3.17] | 2.27 [1.83-2.82] | 2.22 [2.00-2.48] | 2.16 [1.95-2.39] | 2.10 [1.91-2.32] | 2.19 [1.81-2.65] | 1.96 [1.57-2.46] | 2.09 [1.87-2.33] | 2.01 [1.81-2.23] | 1,345 | 383 | 245 | 1,019 | 1,124 |
| L02 | 1.38 [1.34-1.42] | 1.38 [1.34-1.43] | 1.37 [1.32-1.43] | 1.29 [1.22-1.36] | 1.26 [1.22-1.30] | 1.30 [1.26-1.34] | 1.23 [1.19-1.27] | 1.23 [1.18-1.28] | 1.26 [1.19-1.33] | 1.23 [1.19-1.27] | 12,020 | 9,781 | 6,084 | 3,557 | 9,776 |
| L03 | 1.61 [1.58-1.63] | 1.64 [1.62-1.67] | 1.64 [1.61-1.66] | 1.38 [1.32-1.44] | 1.49 [1.47-1.52] | 1.53 [1.50-1.55] | 1.50 [1.48-1.53] | 1.49 [1.47-1.52] | 1.34 [1.28-1.40] | 1.44 [1.42-1.46] | 45,791 | 41,851 | 37,940 | 5,185 | 40,007 |
| L04 | 1.21 [1.11-1.33] | 1.29 [1.07-1.55] | 1.37 [1.09-1.71] | 1.14 [1.03-1.26] | 1.16 [1.06-1.28] | 1.19 [1.08-1.30] | 1.21 [1.00-1.46] | 1.32 [1.05-1.66] | 1.12 [1.01-1.25] | 1.15 [1.05-1.27] | 1,202 | 280 | 190 | 947 | 1,062 |
| L05 | 1.15 [1.09-1.21] | 1.17 [1.11-1.24] | 1.30 [1.15-1.48] | 1.09 [1.02-1.17] | 1.09 [1.02-1.16] | 1.11 [1.05-1.17] | 1.08 [1.02-1.14] | 1.17 [1.03-1.34] | 1.06 [0.98-1.14] | 1.09 [1.02-1.16] | 3,813 | 3,135 | 557 | 2,154 | 2,607 |
| L08 | 1.54 [1.50-1.59] | 1.51 [1.46-1.56] | 1.51 [1.46-1.56] | 1.72 [1.59-1.86] | 1.45 [1.41-1.50] | 1.47 [1.43-1.52] | 1.38 [1.34-1.43] | 1.37 [1.32-1.42] | 1.68 [1.55-1.82] | 1.42 [1.38-1.47] | 12,090 | 10,568 | 9,505 | 1,865 | 10,924 |
| L10 | 2.28 [1.75-2.95] | 2.25 [1.74-2.92] | 2.27 [1.74-2.97] | | 2.00 [1.52-2.64] | 2.11 [1.62-2.76] | 2.06 [1.57-2.69] | 2.01 [1.53-2.66] | | 1.86 [1.40-2.46] | 180 | 175 | 164 | NA | 156 |
| L11 | 1.67 [1.24-2.25] | 1.64 [1.21-2.22] | 1.81 [1.33-2.46] | | 1.64 [1.20-2.25] | 1.57 [1.16-2.13] | 1.56 [1.15-2.13] | 1.77 [1.29-2.42] | | 1.58 [1.15-2.17] | 118 | 117 | 114 | NA | 107 |
| L12 | 2.86 [2.59-3.16] | 2.81 [2.54-3.11] | 2.80 [2.54-3.10] | | 2.64 [2.37-2.93] | 2.75 [2.48-3.04] | 2.65 [2.39-2.93] | 2.63 [2.37-2.91] | | 2.56 [2.30-2.85] | 1,416 | 1,403 | 1,391 | NA | 1,268 |
| L13 | 3.14 [2.51-3.92] | 3.31 [2.62-4.18] | 3.00 [2.34-3.86] | | 2.83 [2.23-3.59] | 2.84 [2.26-3.58] | 2.99 [2.35-3.79] | 2.70 [2.08-3.49] | | 2.63 [2.06-3.35] | 282 | 253 | 215 | NA | 239 |
| L20 | 24.72 [22.44-27.23] | 31.99 [27.28-37.50] | 35.83 [29.13-44.08] | 18.85 [16.88-21.04] | 22.58 [20.37-25.03] | 21.74 [19.72-23.97] | 27.25 [23.19-32.01] | 31.58 [25.62-38.94] | 16.21 [14.49-18.13] | 19.70 [17.74-21.86] | 6,205 | 2,770 | 1,797 | 3,853 | 5,147 |
| L21 | 3.29 [2.87-3.77] | 3.13 [2.71-3.62] | 3.22 [2.77-3.75] | 3.22 [2.24-4.64] | 2.86 [2.48-3.29] | 3.09 [2.69-3.56] | 2.85 [2.45-3.30] | 2.91 [2.49-3.40] | 3.12 [2.12-4.57] | 2.72 [2.36-3.15] | 776 | 682 | 616 | 114 | 678 |
| L22 | 1.05 [0.90-1.22] | 1.08 [0.83-1.40] | 1.00 [0.77-1.31] | 1.01 [0.84-1.21] | 1.01 [0.86-1.18] | 1.01 [0.87-1.18] | 1.02 [0.79-1.33] | 0.96 [0.73-1.25] | 0.95 [0.78-1.15] | 0.99 [0.84-1.16] | 424 | 164 | 160 | 264 | 405 |
| L23 | 3.75 [3.33-4.22] | 3.56 [3.07-4.12] | 3.72 [3.14-4.40] | 4.13 [3.41-5.01] | 3.34 [2.94-3.78] | 3.33 [2.94-3.76] | 3.19 [2.74-3.71] | 3.39 [2.86-4.03] | 3.36 [2.75-4.12] | 3.01 [2.64-3.42] | 1,066 | 656 | 492 | 460 | 893 |
| L24 | 1.45 [1.35-1.55] | 1.42 [1.32-1.52] | 1.41 [1.32-1.51] | | 1.36 [1.27-1.45] | 1.39 [1.30-1.49] | 1.31 [1.22-1.41] | 1.29 [1.20-1.39] | | 1.34 [1.25-1.44] | 2,601 | 2,559 | 2,515 | NA | 2,507 |
| L25 | 2.99 [2.63-3.39] | 3.18 [2.77-3.66] | 3.33 [2.85-3.90] | 1.95 [1.49-2.56] | 2.69 [2.35-3.08] | 2.84 [2.49-3.23] | 2.96 [2.57-3.41] | 3.10 [2.64-3.64] | 1.80 [1.36-2.37] | 2.61 [2.27-2.99] | 847 | 730 | 573 | 164 | 713 |
| L26 | 6.48 [4.30-9.76] | 6.67 [4.43-10.04] | | | 6.43 [4.17-9.91] | 6.06 [3.99-9.21] | 5.73 [3.75-8.75] | | | 6.03 [3.88-9.38] | 121 | 118 | NA | NA | 102 |
| L27 | 1.75 [1.65-1.86] | 1.60 [1.50-1.71] | 1.60 [1.49-1.72] | 2.18 [1.93-2.47] | 1.62 [1.52-1.73] | 1.61 [1.51-1.71] | 1.45 [1.36-1.56] | 1.46 [1.36-1.58] | 1.91 [1.68-2.18] | 1.53 [1.43-1.63] | 3,074 | 2,429 | 2,008 | 807 | 2,727 |
| L28 | 4.46 [4.06-4.91] | 4.18 [3.79-4.61] | 3.99 [3.59-4.42] | 6.73 [4.77-9.50] | 3.75 [3.39-4.14] | 4.14 [3.75-4.56] | 3.82 [3.45-4.22] | 3.61 [3.25-4.01] | 6.04 [4.24-8.60] | 3.51 [3.17-3.89] | 1,757 | 1,585 | 1,387 | 208 | 1,480 |
| L29 | 1.72 [1.64-1.80] | 1.68 [1.60-1.76] | 1.99 [1.87-2.12] | 1.46 [1.31-1.62] | 1.59 [1.51-1.67] | 1.60 [1.52-1.68] | 1.50 [1.42-1.58] | 1.78 [1.67-1.90] | 1.36 [1.21-1.51] | 1.50 [1.42-1.58] | 4,866 | 4,324 | 2,764 | 1,002 | 4,307 |
| L30 | 10.30 [10.10-10.50] | 10.25 [9.99-10.52] | 9.44 [9.16-9.73] | 9.92 [9.66-10.19] | 9.24 [9.06-9.43] | 9.48 [9.30-9.67] | 9.13 [8.89-9.38] | 8.61 [8.35-8.89] | 9.12 [8.88-9.37] | 8.55 [8.37-8.72] | 77,273 | 40,673 | 27,658 | 41,677 | 65,364 |
| L40 | 2.23 [2.17-2.30] | 2.19 [2.13-2.26] | 2.21 [2.14-2.28] | 2.04 [1.84-2.26] | 2.11 [2.04-2.17] | 2.13 [2.07-2.20] | 2.02 [1.96-2.08] | 2.03 [1.97-2.10] | 2.01 [1.80-2.23] | 2.04 [1.98-2.11] | 14,006 | 13,506 | 11,116 | 1,195 | 11,883 |
| L42 | 1.54 [1.22-1.95] | 1.63 [1.27-2.11] | 1.79 [1.32-2.43] | | 1.42 [1.10-1.83] | 1.44 [1.13-1.83] | 1.40 [1.07-1.83] | 1.61 [1.18-2.22] | | 1.32 [1.02-1.72] | 194 | 166 | 121 | NA | 163 |
| L43 | 2.18 [1.99-2.38] | 2.15 [1.97-2.35] | 2.15 [1.96-2.36] | | 1.99 [1.81-2.18] | 2.03 [1.86-2.23] | 1.98 [1.81-2.17] | 1.99 [1.81-2.18] | | 1.89 [1.72-2.08] | 1,468 | 1,442 | 1,332 | NA | 1,256 |
| L44 | 3.12 [2.21-4.41] | | | | | 2.92 [2.05-4.16] | | | | | 109 | NA | NA | NA | NA |
| L50 | 1.93 [1.83-2.03] | 1.78 [1.65-1.92] | 1.86 [1.70-2.03] | 2.03 [1.89-2.17] | 1.89 [1.79-2.00] | 1.64 [1.55-1.73] | 1.45 [1.34-1.56] | 1.50 [1.37-1.65] | 1.75 [1.62-1.88] | 1.60 [1.51-1.70] | 4,198 | 1,906 | 1,317 | 2,516 | 3,728 |
| L51 | 1.28 [1.14-1.43] | 1.83 [1.54-2.16] | 2.03 [1.67-2.46] | 0.97 [0.83-1.13] | 1.24 [1.10-1.40] | 1.21 [1.07-1.35] | 1.60 [1.35-1.91] | 1.77 [1.45-2.16] | 0.95 [0.82-1.11] | 1.18 [1.05-1.34] | 767 | 378 | 296 | 416 | 656 |
| L52 | 1.49 [1.25-1.79] | 1.50 [1.22-1.84] | 1.49 [1.12-1.96] | 1.47 [1.11-1.96] | 1.36 [1.12-1.66] | 1.38 [1.15-1.66] | 1.33 [1.07-1.64] | 1.28 [0.96-1.71] | 1.36 [1.01-1.82] | 1.24 [1.01-1.52] | 329 | 233 | 127 | 139 | 264 |
| L53 | 1.75 [1.66-1.84] | 1.81 [1.71-1.91] | 1.82 [1.72-1.93] | 1.45 [1.29-1.63] | 1.61 [1.53-1.69] | 1.66 [1.58-1.75] | 1.65 [1.56-1.75] | 1.66 [1.57-1.77] | 1.37 [1.21-1.54] | 1.56 [1.47-1.64] | 4,468 | 3,852 | 3,354 | 791 | 3,949 |
| L55 | 1.15 [0.86-1.53] | | | | 1.12 [0.81-1.53] | 1.10 [0.82-1.48] | | | | 1.10 [0.79-1.52] | 118 | NA | NA | NA | 101 |
| L56 | 2.80 [2.22-3.53] | 2.85 [2.23-3.63] | 2.99 [2.29-3.90] | | 2.36 [1.85-3.00] | 2.65 [2.09-3.36] | 2.58 [2.01-3.31] | 2.67 [2.03-3.51] | | 2.22 [1.73-2.86] | 238 | 212 | 176 | NA | 203 |
| L57 | 1.32 [1.27-1.37] | 1.31 [1.26-1.35] | 1.32 [1.28-1.37] | | 1.25 [1.20-1.30] | 1.25 [1.21-1.30] | 1.22 [1.18-1.27] | 1.24 [1.19-1.28] | | 1.20 [1.15-1.24] | 7,794 | 7,772 | 7,653 | NA | 6,959 |
| L59 | 1.42 [1.07-1.89] | 1.44 [1.07-1.93] | 1.40 [1.03-1.91] | | 1.21 [0.89-1.65] | 1.38 [1.02-1.85] | 1.41 [1.04-1.91] | 1.40 [1.01-1.92] | | 1.25 [0.91-1.72] | 126 | 117 | 104 | NA | 104 |
| L60 | 1.41 [1.34-1.49] | 1.48 [1.39-1.58] | 1.51 [1.39-1.63] | 1.32 [1.22-1.43] | 1.33 [1.25-1.41] | 1.30 [1.23-1.38] | 1.33 [1.24-1.42] | 1.32 [1.22-1.44] | 1.24 [1.14-1.34] | 1.24 [1.17-1.32] | 3,703 | 2,359 | 1,553 | 1,740 | 2,908 |
| L63 | 2.87 [2.32-3.56] | 2.84 [2.24-3.59] | 2.62 [1.97-3.48] | | 2.70 [2.15-3.40] | 2.27 [1.80-2.85] | 2.08 [1.61-2.68] | 2.06 [1.53-2.79] | | 2.19 [1.71-2.79] | 290 | 234 | 151 | NA | 243 |
| L65 | 1.99 [1.76-2.25] | 2.04 [1.78-2.33] | 2.09 [1.78-2.45] | 1.78 [1.41-2.24] | 1.89 [1.66-2.15] | 1.75 [1.54-1.99] | 1.73 [1.50-1.99] | 1.77 [1.50-2.08] | 1.64 [1.29-2.08] | 1.68 [1.47-1.92] | 805 | 634 | 451 | 229 | 693 |
| L66 | 1.73 [1.31-2.27] | 1.93 [1.44-2.57] | 1.82 [1.32-2.51] | | 1.72 [1.29-2.29] | 1.62 [1.22-2.14] | 1.79 [1.33-2.42] | 1.64 [1.18-2.28] | | 1.62 [1.21-2.17] | 151 | 135 | 105 | NA | 132 |
| L68 | 1.25 [1.00-1.56] | 1.28 [1.00-1.64] | | | 1.10 [0.86-1.41] | 1.19 [0.95-1.49] | 1.12 [0.87-1.45] | | | 1.05 [0.82-1.35] | 211 | 157 | NA | NA | 167 |
| L70 | 1.45 [1.30-1.62] | 1.42 [1.25-1.61] | 1.44 [1.13-1.83] | 1.37 [1.17-1.59] | 1.29 [1.14-1.46] | 1.32 [1.18-1.49] | 1.22 [1.07-1.39] | 1.24 [0.97-1.60] | 1.27 [1.08-1.49] | 1.21 [1.07-1.37] | 887 | 622 | 168 | 512 | 718 |
| L71 | 1.55 [1.39-1.72] | 1.59 [1.43-1.78] | 1.52 [1.36-1.70] | | 1.50 [1.34-1.68] | 1.44 [1.29-1.61] | 1.49 [1.33-1.66] | 1.40 [1.25-1.57] | | 1.43 [1.27-1.60] | 890 | 873 | 798 | NA | 784 |
| L72 | 1.16 [1.13-1.19] | 1.17 [1.14-1.21] | 1.17 [1.13-1.21] | 1.10 [1.02-1.19] | 1.09 [1.06-1.13] | 1.11 [1.08-1.14] | 1.09 [1.06-1.12] | 1.09 [1.06-1.14] | 1.07 [0.99-1.16] | 1.07 [1.03-1.10] | 10,923 | 10,002 | 7,136 | 1,763 | 8,434 |
| L73 | 1.57 [1.46-1.70] | 1.56 [1.44-1.68] | 1.58 [1.42-1.75] | 1.32 [1.12-1.55] | 1.37 [1.26-1.49] | 1.44 [1.33-1.56] | 1.35 [1.24-1.46] | 1.38 [1.24-1.55] | 1.24 [1.05-1.47] | 1.31 [1.20-1.42] | 1,777 | 1,630 | 848 | 404 | 1,497 |
| L80 | 1.97 [1.69-2.29] | 1.85 [1.57-2.18] | 1.90 [1.59-2.27] | 2.46 [1.72-3.53] | 1.83 [1.56-2.15] | 1.80 [1.54-2.11] | 1.65 [1.40-1.96] | 1.67 [1.39-2.01] | 2.34 [1.60-3.42] | 1.73 [1.46-2.04] | 492 | 410 | 343 | 106 | 427 |
| L81 | 1.29 [1.20-1.38] | 1.29 [1.19-1.39] | 1.27 [1.17-1.38] | 1.50 [1.26-1.78] | 1.24 [1.15-1.34] | 1.23 [1.14-1.32] | 1.20 [1.11-1.30] | 1.19 [1.09-1.30] | 1.45 [1.21-1.72] | 1.19 [1.10-1.29] | 1,903 | 1,576 | 1,340 | 395 | 1,627 |
| L82 | 1.22 [1.17-1.28] | 1.25 [1.19-1.31] | 1.26 [1.20-1.31] | | 1.18 [1.13-1.24] | 1.16 [1.11-1.21] | 1.17 [1.12-1.22] | 1.17 [1.12-1.23] | | 1.13 [1.07-1.18] | 4,915 | 4,889 | 4,688 | NA | 4,220 |
| L83 | 1.67 [1.43-1.96] | 1.82 [1.51-2.18] | 1.71 [1.41-2.08] | 1.58 [1.18-2.12] | 1.55 [1.31-1.83] | 1.56 [1.33-1.84] | 1.60 [1.32-1.93] | 1.56 [1.27-1.90] | 1.51 [1.11-2.04] | 1.46 [1.23-1.74] | 456 | 322 | 278 | 150 | 387 |
| L84 | 1.49 [1.31-1.69] | 1.47 [1.29-1.67] | 1.50 [1.31-1.72] | | 1.32 [1.16-1.51] | 1.39 [1.22-1.58] | 1.33 [1.16-1.51] | 1.35 [1.18-1.55] | | 1.26 [1.10-1.44] | 634 | 609 | 550 | NA | 566 |
| L85 | 1.62 [1.52-1.73] | 1.57 [1.47-1.68] | 1.55 [1.45-1.66] | 2.18 [1.67-2.84] | 1.56 [1.46-1.67] | 1.54 [1.44-1.65] | 1.46 [1.37-1.57] | 1.45 [1.35-1.55] | 2.18 [1.66-2.87] | 1.51 [1.41-1.62] | 2,552 | 2,405 | 2,271 | 183 | 2,257 |
| L88 | 1.56 [1.20-2.04] | 1.68 [1.29-2.19] | 1.68 [1.25-2.26] | | 1.33 [1.02-1.74] | 1.37 [1.04-1.80] | 1.40 [1.07-1.85] | 1.45 [1.07-1.97] | | 1.21 [0.92-1.60] | 151 | 150 | 119 | NA | 139 |
| L89 | 1.22 [1.20-1.25] | 1.21 [1.19-1.24] | 1.22 [1.19-1.24] | 0.99 [0.82-1.20] | 1.18 [1.16-1.21] | 1.20 [1.17-1.22] | 1.15 [1.13-1.18] | 1.15 [1.13-1.17] | 0.97 [0.80-1.18] | 1.18 [1.16-1.21] | 26,249 | 26,064 | 25,754 | 284 | 24,650 |
| L90 | 1.29 [1.24-1.34] | 1.33 [1.28-1.39] | 1.32 [1.25-1.38] | 1.04 [0.94-1.16] | 1.20 [1.15-1.26] | 1.22 [1.17-1.27] | 1.24 [1.18-1.29] | 1.21 [1.15-1.28] | 1.02 [0.92-1.14] | 1.16 [1.11-1.21] | 5,613 | 5,036 | 3,915 | 880 | 4,909 |
| L91 | 1.35 [1.26-1.44] | 1.35 [1.25-1.46] | 1.36 [1.23-1.50] | 1.26 [1.12-1.40] | 1.22 [1.13-1.32] | 1.27 [1.19-1.36] | 1.26 [1.16-1.36] | 1.25 [1.13-1.38] | 1.25 [1.11-1.40] | 1.18 [1.09-1.27] | 2,210 | 1,609 | 978 | 826 | 1,765 |
| L92 | 1.30 [1.20-1.40] | 1.40 [1.28-1.52] | 1.37 [1.24-1.52] | 1.11 [0.97-1.28] | 1.21 [1.11-1.31] | 1.21 [1.12-1.31] | 1.26 [1.15-1.37] | 1.24 [1.12-1.38] | 1.04 [0.90-1.20] | 1.15 [1.06-1.25] | 1,723 | 1,322 | 957 | 531 | 1,468 |
| L93 | 1.86 [1.66-2.10] | 1.86 [1.66-2.10] | 1.84 [1.62-2.10] | | 1.68 [1.49-1.90] | 1.67 [1.48-1.88] | 1.61 [1.42-1.82] | 1.58 [1.38-1.81] | | 1.55 [1.36-1.75] | 786 | 768 | 620 | NA | 700 |
| L94 | 1.55 [1.35-1.78] | 1.55 [1.33-1.81] | 1.81 [1.53-2.14] | 1.53 [1.14-2.06] | 1.46 [1.26-1.69] | 1.45 [1.26-1.68] | 1.41 [1.20-1.65] | 1.60 [1.34-1.90] | 1.54 [1.13-2.09] | 1.37 [1.18-1.59] | 541 | 426 | 368 | 136 | 472 |
| L95 | 1.61 [1.42-1.83] | 1.78 [1.56-2.04] | 1.70 [1.47-1.96] | | 1.51 [1.32-1.73] | 1.50 [1.31-1.71] | 1.63 [1.42-1.87] | 1.53 [1.32-1.77] | | 1.42 [1.23-1.63] | 663 | 606 | 526 | NA | 572 |
| L97 | 1.54 [1.51-1.58] | 1.53 [1.50-1.57] | 1.53 [1.50-1.57] | 1.28 [0.94-1.75] | 1.45 [1.42-1.49] | 1.49 [1.46-1.53] | 1.42 [1.39-1.46] | 1.42 [1.38-1.45] | 1.34 [0.97-1.85] | 1.43 [1.40-1.46] | 21,630 | 21,576 | 21,276 | 108 | 19,873 |
| L98 | 1.39 [1.36-1.42] | 1.40 [1.36-1.44] | 1.39 [1.35-1.43] | 1.28 [1.19-1.38] | 1.30 [1.27-1.34] | 1.32 [1.29-1.36] | 1.30 [1.27-1.33] | 1.29 [1.25-1.33] | 1.25 [1.16-1.35] | 1.26 [1.23-1.29] | 17,334 | 15,883 | 14,075 | 1,917 | 15,018 |
| Diseases of the musculoskeletal system and connective tissue (M00-M99) | | | | | | | | | | | | | | | |
| M00 | 1.48 [1.39-1.59] | 1.50 [1.39-1.61] | 1.54 [1.42-1.66] | 1.40 [1.20-1.64] | 1.35 [1.26-1.45] | 1.41 [1.32-1.51] | 1.37 [1.27-1.48] | 1.41 [1.30-1.52] | 1.37 [1.16-1.61] | 1.31 [1.22-1.41] | 2,261 | 1,904 | 1,742 | 421 | 1,990 |
| M02 | 1.22 [1.07-1.40] | 1.42 [1.18-1.72] | 1.34 [1.07-1.67] | 1.13 [0.94-1.34] | 1.17 [1.01-1.35] | 1.18 [1.03-1.35] | 1.29 [1.06-1.57] | 1.23 [0.97-1.54] | 1.11 [0.93-1.33] | 1.14 [0.98-1.32] | 556 | 279 | 191 | 313 | 464 |
| M05 | 1.40 [1.32-1.50] | 1.39 [1.30-1.48] | 1.37 [1.28-1.47] | | 1.24 [1.16-1.32] | 1.35 [1.27-1.44] | 1.30 [1.22-1.39] | 1.29 [1.21-1.38] | | 1.21 [1.13-1.29] | 2,360 | 2,359 | 2,210 | NA | 2,108 |
| M06 | 1.40 [1.37-1.44] | 1.40 [1.36-1.43] | 1.39 [1.36-1.42] | 1.29 [1.06-1.57] | 1.31 [1.28-1.34] | 1.31 [1.28-1.34] | 1.27 [1.23-1.30] | 1.26 [1.23-1.29] | 1.21 [0.99-1.49] | 1.24 [1.21-1.27] | 17,481 | 17,434 | 16,286 | 282 | 15,317 |
| M07 | 1.75 [1.64-1.87] | 1.70 [1.59-1.82] | 1.70 [1.59-1.83] | 1.52 [1.14-2.03] | 1.60 [1.49-1.71] | 1.62 [1.52-1.74] | 1.51 [1.41-1.61] | 1.50 [1.40-1.62] | 1.46 [1.08-1.97] | 1.50 [1.40-1.61] | 2,456 | 2,412 | 1,965 | 138 | 2,147 |
| M08 | 1.33 [1.19-1.48] | 1.38 [1.09-1.73] | | 1.34 [1.19-1.51] | 1.25 [1.10-1.41] | 1.28 [1.14-1.43] | 1.26 [1.00-1.60] | | 1.30 [1.15-1.47] | 1.23 [1.08-1.39] | 852 | 183 | NA | 723 | 706 |
| M10 | 1.36 [1.33-1.39] | 1.36 [1.33-1.39] | 1.36 [1.33-1.39] | | 1.27 [1.24-1.30] | 1.29 [1.26-1.32] | 1.24 [1.21-1.27] | 1.23 [1.20-1.26] | | 1.22 [1.19-1.25] | 19,181 | 19,179 | 18,861 | NA | 17,324 |
| M11 | 1.29 [1.20-1.39] | 1.30 [1.21-1.40] | 1.29 [1.20-1.38] | | 1.26 [1.17-1.35] | 1.21 [1.12-1.30] | 1.19 [1.11-1.28] | 1.17 [1.09-1.26] | | 1.20 [1.12-1.29] | 2,128 | 2,129 | 2,100 | NA | 1,994 |
| M12 | 1.40 [1.21-1.61] | 1.33 [1.15-1.54] | 1.36 [1.16-1.61] | | 1.25 [1.08-1.45] | 1.27 [1.10-1.48] | 1.17 [1.01-1.37] | 1.16 [0.98-1.38] | | 1.21 [1.04-1.40] | 475 | 439 | 350 | NA | 430 |
| M13 | 1.34 [1.32-1.36] | 1.33 [1.31-1.35] | 1.33 [1.31-1.35] | 1.36 [1.19-1.55] | 1.26 [1.24-1.27] | 1.24 [1.22-1.26] | 1.19 [1.17-1.21] | 1.19 [1.17-1.21] | 1.27 [1.11-1.45] | 1.18 [1.17-1.20] | 48,665 | 48,345 | 46,962 | 640 | 44,324 |
| M14 | 1.61 [1.41-1.85] | 1.62 [1.42-1.86] | 1.62 [1.41-1.87] | | 1.32 [1.15-1.51] | 1.46 [1.28-1.68] | 1.33 [1.16-1.54] | 1.31 [1.14-1.52] | | 1.25 [1.09-1.43] | 567 | 557 | 526 | NA | 528 |
| M15 | 1.38 [1.34-1.41] | 1.37 [1.34-1.41] | 1.36 [1.33-1.40] | | 1.27 [1.24-1.30] | 1.26 [1.23-1.29] | 1.22 [1.19-1.25] | 1.21 [1.18-1.24] | | 1.19 [1.16-1.22] | 18,016 | 18,006 | 17,845 | NA | 16,796 |
| M16 | 1.13 [1.10-1.15] | 1.12 [1.10-1.14] | 1.11 [1.09-1.14] | 1.12 [0.84-1.49] | 1.07 [1.05-1.09] | 1.09 [1.07-1.11] | 1.07 [1.05-1.09] | 1.06 [1.04-1.08] | 1.08 [0.80-1.46] | 1.05 [1.03-1.07] | 25,902 | 25,881 | 25,397 | 127 | 22,202 |
| M17 | 1.25 [1.23-1.27] | 1.25 [1.23-1.27] | 1.25 [1.23-1.27] | 1.14 [0.96-1.35] | 1.17 [1.15-1.19] | 1.18 [1.16-1.20] | 1.16 [1.14-1.18] | 1.15 [1.13-1.17] | 1.09 [0.91-1.30] | 1.12 [1.10-1.14] | 45,645 | 45,589 | 44,018 | 348 | 39,337 |
| M18 | 1.43 [1.35-1.52] | 1.42 [1.33-1.50] | 1.44 [1.35-1.53] | | 1.28 [1.20-1.36] | 1.27 [1.19-1.35] | 1.22 [1.14-1.29] | 1.24 [1.17-1.32] | | 1.17 [1.10-1.25] | 2,713 | 2,711 | 2,671 | NA | 2,503 |
| M19 | 1.36 [1.34-1.37] | 1.36 [1.34-1.38] | 1.35 [1.34-1.37] | 1.26 [1.05-1.51] | 1.26 [1.24-1.27] | 1.25 [1.24-1.27] | 1.21 [1.20-1.23] | 1.20 [1.19-1.22] | 1.14 [0.94-1.39] | 1.19 [1.17-1.20] | 76,155 | 76,146 | 74,683 | 319 | 70,181 |
| M20 | 1.30 [1.26-1.34] | 1.30 [1.26-1.34] | 1.31 [1.26-1.35] | 1.19 [1.08-1.32] | 1.19 [1.16-1.23] | 1.22 [1.18-1.26] | 1.20 [1.16-1.24] | 1.21 [1.17-1.25] | 1.15 [1.04-1.27] | 1.14 [1.10-1.17] | 11,087 | 10,431 | 8,993 | 1,070 | 9,169 |
| M21 | 1.19 [1.15-1.24] | 1.24 [1.19-1.29] | 1.23 [1.18-1.29] | 1.00 [0.92-1.09] | 1.11 [1.07-1.16] | 1.12 [1.07-1.16] | 1.14 [1.09-1.19] | 1.13 [1.08-1.19] | 0.97 [0.88-1.06] | 1.07 [1.03-1.12] | 6,537 | 5,393 | 4,757 | 1,371 | 5,909 |
| M22 | 1.32 [1.24-1.41] | 1.35 [1.26-1.44] | 1.33 [1.21-1.46] | 1.26 [1.10-1.44] | 1.26 [1.17-1.36] | 1.20 [1.12-1.29] | 1.18 [1.10-1.27] | 1.18 [1.07-1.29] | 1.18 [1.03-1.35] | 1.17 [1.09-1.26] | 2,218 | 1,919 | 1,042 | 618 | 1,797 |
| M23 | 1.22 [1.19-1.25] | 1.22 [1.20-1.25] | 1.23 [1.20-1.27] | 1.14 [1.08-1.21] | 1.12 [1.10-1.15] | 1.14 [1.12-1.17] | 1.13 [1.10-1.15] | 1.13 [1.10-1.16] | 1.09 [1.03-1.16] | 1.07 [1.04-1.09] | 20,542 | 19,394 | 14,216 | 3,153 | 15,635 |
| M24 | 1.26 [1.22-1.30] | 1.29 [1.24-1.33] | 1.29 [1.23-1.35] | 1.15 [1.08-1.23] | 1.15 [1.11-1.19] | 1.17 [1.13-1.21] | 1.16 [1.11-1.20] | 1.16 [1.11-1.21] | 1.10 [1.03-1.18] | 1.10 [1.06-1.14] | 8,753 | 7,438 | 4,819 | 2,395 | 7,397 |
| M25 | 1.29 [1.27-1.31] | 1.30 [1.28-1.32] | 1.29 [1.26-1.31] | 1.21 [1.16-1.25] | 1.20 [1.18-1.22] | 1.20 [1.18-1.22] | 1.17 [1.15-1.18] | 1.15 [1.14-1.17] | 1.15 [1.10-1.19] | 1.14 [1.13-1.16] | 51,040 | 45,901 | 39,263 | 7,314 | 45,475 |
| M30 | 1.39 [1.21-1.59] | 1.47 [1.20-1.82] | 1.42 [1.13-1.78] | 1.28 [1.07-1.53] | 1.32 [1.13-1.53] | 1.28 [1.11-1.48] | 1.23 [0.99-1.53] | 1.18 [0.93-1.50] | 1.29 [1.08-1.55] | 1.23 [1.06-1.43] | 523 | 227 | 192 | 309 | 454 |
| M31 | 1.34 [1.26-1.42] | 1.34 [1.26-1.43] | 1.33 [1.25-1.42] | | 1.25 [1.17-1.33] | 1.26 [1.18-1.34] | 1.24 [1.16-1.32] | 1.22 [1.15-1.30] | | 1.18 [1.10-1.26] | 2,711 | 2,681 | 2,594 | NA | 2,394 |
| M32 | 1.79 [1.65-1.95] | 1.75 [1.61-1.90] | 1.69 [1.54-1.86] | 1.73 [1.30-2.30] | 1.62 [1.49-1.76] | 1.63 [1.49-1.77] | 1.52 [1.39-1.65] | 1.48 [1.34-1.63] | 1.73 [1.29-2.32] | 1.49 [1.36-1.63] | 1,578 | 1,517 | 1,106 | 150 | 1,379 |
| M33 | 2.07 [1.75-2.45] | 2.10 [1.76-2.50] | 2.02 [1.67-2.43] | | 1.90 [1.58-2.29] | 2.00 [1.69-2.38] | 1.98 [1.65-2.37] | 1.92 [1.58-2.32] | | 1.81 [1.50-2.19] | 403 | 354 | 306 | NA | 315 |
| M34 | 1.75 [1.53-2.00] | 1.66 [1.45-1.89] | 1.65 [1.44-1.90] | | 1.55 [1.35-1.78] | 1.62 [1.41-1.85] | 1.49 [1.30-1.71] | 1.49 [1.29-1.72] | | 1.46 [1.26-1.68] | 605 | 585 | 529 | NA | 525 |
| M35 | 1.44 [1.40-1.48] | 1.44 [1.40-1.49] | 1.43 [1.39-1.47] | 1.43 [1.31-1.56] | 1.35 [1.31-1.39] | 1.33 [1.29-1.36] | 1.31 [1.27-1.34] | 1.30 [1.26-1.34] | 1.32 [1.21-1.45] | 1.27 [1.23-1.30] | 14,083 | 13,063 | 11,863 | 1,478 | 12,463 |
| M36 | 1.43 [1.07-1.89] | 1.41 [1.03-1.93] | | | 1.29 [0.97-1.73] | 1.30 [0.97-1.74] | 1.27 [0.92-1.76] | | | 1.24 [0.92-1.67] | 123 | 102 | NA | NA | 118 |

#### Supplementary Table 3: Hazard ratios and events from all cohorts

| | Hazard ratio (99% confidence interval) | | | | | | | | | | Events (in exposed) | | | | |
|---|---|---|---|---|---|---|---|---|---|---|---|---|---|---|---|
| | crude | | | | | adjusted | | | | | | | | | |
| Outcome | any age | 18+ | 40+ | <18 | hosp. | any age | 18+ | 40+ | <18 | hosp. | any age | 18+ | 40+ | <18 | hosp. |
| M40 | 1.25 [1.18-1.33] | 1.24 [1.17-1.32] | 1.24 [1.17-1.32] | 1.04 [0.82-1.32] | 1.18 [1.11-1.25] | 1.18 [1.11-1.25] | 1.14 [1.07-1.21] | 1.15 [1.08-1.22] | 0.98 [0.77-1.26] | 1.13 [1.06-1.20] | 3,017 | 2,897 | 2,744 | 180 | 2,846 |
| M41 | 1.24 [1.20-1.28] | 1.27 [1.23-1.32] | 1.27 [1.22-1.32] | 1.07 [0.99-1.15] | 1.14 [1.10-1.18] | 1.16 [1.12-1.20] | 1.16 [1.12-1.21] | 1.16 [1.11-1.21] | 1.03 [0.96-1.12] | 1.09 [1.05-1.13] | 8,862 | 7,531 | 6,535 | 1,747 | 8,018 |
| M42 | 1.13 [0.89-1.44] | 1.22 [0.94-1.57] | | | 1.14 [0.89-1.46] | 0.98 [0.76-1.26] | 1.01 [0.77-1.32] | | | 1.02 [0.79-1.32] | 167 | 144 | NA | NA | 151 |
| M43 | 1.29 [1.24-1.34] | 1.29 [1.24-1.34] | 1.30 [1.25-1.35] | 1.02 [0.90-1.17] | 1.18 [1.13-1.22] | 1.18 [1.14-1.23] | 1.14 [1.09-1.19] | 1.15 [1.10-1.19] | 0.98 [0.85-1.12] | 1.10 [1.06-1.15] | 7,046 | 6,623 | 6,121 | 575 | 6,468 |
| M45 | 1.50 [1.40-1.62] | 1.56 [1.45-1.69] | 1.48 [1.37-1.61] | | 1.42 [1.31-1.54] | 1.39 [1.29-1.50] | 1.39 [1.29-1.50] | 1.31 [1.20-1.42] | | 1.33 [1.23-1.44] | 1,799 | 1,796 | 1,542 | NA | 1,578 |
| M46 | 1.42 [1.36-1.48] | 1.41 [1.36-1.48] | 1.41 [1.35-1.47] | 1.10 [0.83-1.45] | 1.29 [1.24-1.35] | 1.28 [1.22-1.33] | 1.22 [1.17-1.28] | 1.21 [1.16-1.27] | 1.08 [0.81-1.44] | 1.20 [1.15-1.26] | 5,856 | 5,775 | 5,440 | 135 | 5,426 |
| M47 | 1.35 [1.33-1.38] | 1.36 [1.34-1.38] | 1.35 [1.32-1.37] | 1.07 [0.85-1.35] | 1.26 [1.23-1.28] | 1.23 [1.21-1.26] | 1.19 [1.17-1.21] | 1.18 [1.16-1.20] | 1.00 [0.79-1.27] | 1.18 [1.16-1.20] | 32,121 | 32,101 | 30,887 | 193 | 29,828 |
| M48 | 1.29 [1.26-1.32] | 1.29 [1.26-1.32] | 1.29 [1.26-1.32] | 1.12 [0.88-1.41] | 1.20 [1.17-1.23] | 1.19 [1.16-1.22] | 1.15 [1.12-1.17] | 1.14 [1.11-1.16] | 1.05 [0.82-1.33] | 1.14 [1.11-1.16] | 18,247 | 18,194 | 17,646 | 194 | 16,947 |
| M49 | 1.08 [0.95-1.23] | 1.07 [0.94-1.21] | 1.16 [1.01-1.32] | | 1.08 [0.94-1.24] | 1.06 [0.93-1.21] | 1.03 [0.90-1.17] | 1.12 [0.98-1.28] | | 1.09 [0.95-1.25] | 585 | 575 | 548 | NA | 515 |
| M50 | 1.33 [1.27-1.40] | 1.32 [1.26-1.38] | 1.33 [1.27-1.39] | | 1.21 [1.16-1.27] | 1.22 [1.17-1.28] | 1.14 [1.09-1.20] | 1.15 [1.10-1.21] | | 1.15 [1.09-1.20] | 4,856 | 4,851 | 4,448 | NA | 4,460 |
| M51 | 1.30 [1.28-1.33] | 1.30 [1.27-1.33] | 1.32 [1.29-1.34] | 1.24 [1.11-1.38] | 1.20 [1.18-1.23] | 1.19 [1.16-1.21] | 1.13 [1.11-1.15] | 1.14 [1.12-1.17] | 1.17 [1.04-1.30] | 1.13 [1.11-1.16] | 23,386 | 23,257 | 19,588 | 925 | 21,018 |
| M53 | 1.26 [1.16-1.36] | 1.27 [1.17-1.38] | 1.30 [1.18-1.42] | 1.03 [0.78-1.36] | 1.15 [1.06-1.25] | 1.12 [1.03-1.22] | 1.08 [0.99-1.18] | 1.09 [0.99-1.21] | 0.99 [0.75-1.31] | 1.06 [0.97-1.16] | 1,485 | 1,418 | 1,097 | 137 | 1,341 |
| M54 | 1.30 [1.28-1.32] | 1.30 [1.28-1.32] | 1.30 [1.28-1.32] | 1.15 [1.10-1.19] | 1.19 [1.17-1.20] | 1.19 [1.17-1.21] | 1.13 [1.12-1.15] | 1.14 [1.12-1.16] | 1.09 [1.04-1.14] | 1.12 [1.11-1.14] | 60,233 | 57,895 | 46,321 | 5,857 | 53,676 |
| M60 | 1.45 [1.33-1.58] | 1.48 [1.34-1.63] | 1.48 [1.33-1.65] | 1.35 [1.15-1.59] | 1.38 [1.26-1.52] | 1.38 [1.27-1.51] | 1.36 [1.23-1.51] | 1.38 [1.24-1.54] | 1.31 [1.11-1.55] | 1.35 [1.23-1.49] | 1,388 | 1,020 | 865 | 410 | 1,221 |
| M62 | 1.22 [1.17-1.28] | 1.26 [1.20-1.33] | 1.25 [1.19-1.32] | 0.99 [0.88-1.12] | 1.14 [1.09-1.19] | 1.16 [1.11-1.22] | 1.16 [1.10-1.22] | 1.16 [1.10-1.23] | 0.94 [0.83-1.06] | 1.11 [1.06-1.17] | 5,043 | 4,523 | 3,883 | 738 | 4,560 |
| M65 | 1.36 [1.32-1.40] | 1.35 [1.31-1.39] | 1.35 [1.30-1.39] | 1.26 [1.17-1.37] | 1.26 [1.22-1.30] | 1.25 [1.21-1.29] | 1.21 [1.17-1.25] | 1.20 [1.16-1.24] | 1.23 [1.13-1.33] | 1.19 [1.15-1.22] | 10,874 | 9,568 | 8,130 | 1,723 | 9,468 |
| M66 | 1.29 [1.17-1.42] | 1.31 [1.18-1.44] | 1.30 [1.17-1.44] | | 1.16 [1.05-1.29] | 1.21 [1.10-1.34] | 1.21 [1.09-1.34] | 1.18 [1.06-1.31] | | 1.11 [1.00-1.24] | 1,022 | 1,012 | 919 | NA | 866 |
| M67 | 1.27 [1.22-1.31] | 1.30 [1.25-1.35] | 1.33 [1.27-1.39] | 1.13 [1.05-1.21] | 1.17 [1.12-1.21] | 1.19 [1.14-1.23] | 1.19 [1.14-1.24] | 1.21 [1.15-1.26] | 1.08 [1.01-1.16] | 1.11 [1.07-1.16] | 7,633 | 6,111 | 4,364 | 2,036 | 6,296 |
| M70 | 1.44 [1.37-1.51] | 1.44 [1.37-1.52] | 1.43 [1.36-1.51] | 1.16 [0.93-1.46] | 1.30 [1.23-1.37] | 1.31 [1.25-1.38] | 1.28 [1.21-1.34] | 1.26 [1.20-1.33] | 1.13 [0.89-1.43] | 1.21 [1.15-1.27] | 4,283 | 4,180 | 3,719 | 208 | 3,753 |
| M71 | 1.34 [1.26-1.42] | 1.33 [1.25-1.41] | 1.32 [1.24-1.41] | 1.51 [1.09-2.08] | 1.26 [1.18-1.35] | 1.24 [1.16-1.32] | 1.20 [1.13-1.28] | 1.19 [1.11-1.27] | 1.44 [1.04-2.02] | 1.19 [1.11-1.27] | 2,600 | 2,544 | 2,355 | 107 | 2,333 |
| M72 | 1.12 [1.07-1.17] | 1.12 [1.07-1.17] | 1.12 [1.07-1.17] | 1.23 [0.91-1.67] | 1.06 [1.01-1.11] | 1.08 [1.03-1.13] | 1.07 [1.02-1.11] | 1.07 [1.02-1.12] | 1.18 [0.85-1.62] | 1.04 [0.99-1.09] | 5,021 | 4,955 | 4,699 | 114 | 4,137 |
| M75 | 1.32 [1.28-1.35] | 1.32 [1.29-1.35] | 1.32 [1.29-1.36] | 1.18 [1.01-1.37] | 1.20 [1.17-1.23] | 1.20 [1.17-1.23] | 1.17 [1.14-1.20] | 1.17 [1.14-1.20] | 1.12 [0.96-1.31] | 1.13 [1.10-1.16] | 16,137 | 16,023 | 14,523 | 440 | 14,089 |
| M76 | 1.42 [1.31-1.54] | 1.41 [1.29-1.53] | 1.41 [1.28-1.55] | 1.24 [0.95-1.63] | 1.32 [1.21-1.44] | 1.28 [1.18-1.40] | 1.23 [1.12-1.34] | 1.21 [1.10-1.33] | 1.13 [0.85-1.50] | 1.22 [1.11-1.33] | 1,443 | 1,367 | 1,085 | 151 | 1,302 |
| M77 | 1.47 [1.40-1.56] | 1.48 [1.40-1.56] | 1.47 [1.39-1.55] | 0.95 [0.73-1.26] | 1.34 [1.26-1.41] | 1.33 [1.26-1.40] | 1.29 [1.22-1.36] | 1.27 [1.19-1.35] | 0.88 [0.66-1.17] | 1.24 [1.17-1.31] | 3,405 | 3,360 | 2,884 | 136 | 3,032 |
| M79 | 1.39 [1.37-1.41] | 1.41 [1.39-1.43] | 1.41 [1.39-1.44] | 1.20 [1.15-1.25] | 1.28 [1.27-1.30] | 1.27 [1.25-1.29] | 1.22 [1.21-1.24] | 1.23 [1.21-1.25] | 1.15 [1.10-1.20] | 1.20 [1.19-1.22] | 53,229 | 49,622 | 41,362 | 5,874 | 47,643 |
| M80 | 1.28 [1.23-1.34] | 1.29 [1.23-1.34] | 1.30 [1.24-1.35] | | 1.22 [1.17-1.28] | 1.22 [1.16-1.27] | 1.18 [1.13-1.23] | 1.19 [1.14-1.24] | | 1.17 [1.12-1.22] | 5,833 | 5,806 | 5,748 | NA | 5,339 |
| M81 | 1.31 [1.28-1.33] | 1.30 [1.28-1.32] | 1.30 [1.28-1.32] | 1.38 [1.09-1.75] | 1.23 [1.21-1.25] | 1.22 [1.20-1.24] | 1.17 [1.15-1.19] | 1.18 [1.16-1.20] | 1.30 [1.02-1.66] | 1.16 [1.14-1.18] | 35,991 | 35,913 | 35,416 | 198 | 33,098 |
| M83 | 1.34 [1.09-1.65] | 1.23 [1.00-1.52] | 1.28 [1.02-1.59] | | 1.29 [1.04-1.60] | 1.22 [0.98-1.51] | 1.13 [0.91-1.39] | 1.16 [0.92-1.45] | | 1.22 [0.98-1.52] | 232 | 231 | 208 | NA | 216 |
| M84 | 1.23 [1.17-1.28] | 1.25 [1.19-1.31] | 1.26 [1.20-1.33] | 1.11 [1.00-1.23] | 1.15 [1.10-1.20] | 1.16 [1.11-1.21] | 1.14 [1.09-1.20] | 1.16 [1.10-1.22] | 1.08 [0.96-1.20] | 1.13 [1.07-1.18] | 5,167 | 4,683 | 3,741 | 886 | 4,392 |
| M85 | 1.31 [1.27-1.36] | 1.32 [1.27-1.37] | 1.29 [1.25-1.34] | 1.18 [1.01-1.39] | 1.24 [1.19-1.28] | 1.20 [1.16-1.25] | 1.17 [1.13-1.22] | 1.16 [1.11-1.20] | 1.13 [0.96-1.33] | 1.16 [1.12-1.20] | 8,162 | 7,888 | 7,406 | 435 | 7,679 |
| M86 | 1.37 [1.30-1.44] | 1.38 [1.31-1.46] | 1.37 [1.30-1.45] | 1.36 [1.19-1.56] | 1.28 [1.21-1.35] | 1.31 [1.25-1.38] | 1.25 [1.18-1.32] | 1.23 [1.16-1.30] | 1.33 [1.16-1.53] | 1.26 [1.19-1.33] | 3,878 | 3,351 | 3,129 | 604 | 3,536 |
| M87 | 1.28 [1.18-1.39] | 1.27 [1.17-1.38] | 1.28 [1.17-1.39] | 1.22 [0.93-1.59] | 1.16 [1.07-1.26] | 1.21 [1.11-1.31] | 1.16 [1.07-1.26] | 1.16 [1.06-1.27] | 1.16 [0.88-1.53] | 1.13 [1.04-1.23] | 1,542 | 1,451 | 1,285 | 146 | 1,373 |
| M88 | 1.17 [1.05-1.30] | 1.15 [1.03-1.28] | 1.18 [1.05-1.31] | | 1.07 [0.96-1.21] | 1.13 [1.01-1.26] | 1.11 [0.99-1.24] | 1.13 [1.01-1.27] | | 1.05 [0.93-1.18] | 890 | 887 | 851 | NA | 738 |
| M89 | 1.29 [1.23-1.35] | 1.30 [1.24-1.37] | 1.32 [1.24-1.40] | 1.12 [1.01-1.25] | 1.18 [1.12-1.24] | 1.20 [1.14-1.26] | 1.17 [1.11-1.24] | 1.18 [1.11-1.25] | 1.09 [0.97-1.22] | 1.13 [1.07-1.19] | 4,230 | 3,617 | 2,882 | 849 | 3,693 |
| M90 | 1.06 [0.97-1.15] | 1.03 [0.95-1.13] | 1.04 [0.96-1.13] | | 1.02 [0.93-1.11] | 1.06 [0.97-1.15] | 1.02 [0.94-1.11] | 1.02 [0.94-1.12] | | 1.04 [0.95-1.14] | 1,351 | 1,318 | 1,268 | NA | 1,201 |
| M91 | 1.19 [1.02-1.39] | 1.27 [0.99-1.64] | | 1.16 [0.96-1.39] | 1.13 [0.95-1.34] | 1.16 [0.99-1.36] | 1.21 [0.93-1.56] | | 1.12 [0.92-1.35] | 1.11 [0.93-1.32] | 412 | 151 | NA | 281 | 337 |
| M92 | 1.45 [1.28-1.64] | 1.43 [1.22-1.67] | 1.45 [1.17-1.78] | 1.31 [1.10-1.56] | 1.31 [1.14-1.51] | 1.31 [1.16-1.50] | 1.27 [1.08-1.49] | 1.27 [1.02-1.58] | 1.19 [0.99-1.43] | 1.18 [1.02-1.36] | 680 | 406 | 214 | 360 | 533 |
| M93 | 1.31 [1.20-1.44] | 1.30 [1.16-1.46] | 1.30 [1.13-1.50] | 1.25 [1.09-1.43] | 1.18 [1.07-1.31] | 1.22 [1.11-1.34] | 1.15 [1.02-1.29] | 1.15 [0.99-1.33] | 1.18 [1.03-1.36] | 1.12 [1.00-1.24] | 1,199 | 751 | 441 | 579 | 938 |
| M94 | 1.41 [1.33-1.49] | 1.41 [1.33-1.50] | 1.49 [1.39-1.60] | 1.24 [1.09-1.40] | 1.31 [1.23-1.39] | 1.26 [1.19-1.34] | 1.21 [1.14-1.28] | 1.27 [1.18-1.36] | 1.18 [1.04-1.35] | 1.21 [1.14-1.29] | 3,261 | 2,901 | 2,006 | 715 | 2,911 |
| M95 | 1.36 [1.21-1.51] | 1.34 [1.19-1.51] | 1.31 [1.10-1.56] | 1.21 [1.00-1.46] | 1.21 [1.07-1.37] | 1.27 [1.13-1.42] | 1.20 [1.06-1.35] | 1.14 [0.95-1.37] | 1.20 [0.98-1.46] | 1.16 [1.03-1.32] | 806 | 662 | 311 | 284 | 654 |
| M96 | 1.28 [1.19-1.38] | 1.31 [1.21-1.41] | 1.35 [1.25-1.46] | | 1.20 [1.11-1.29] | 1.20 [1.12-1.30] | 1.18 [1.10-1.28] | 1.23 [1.14-1.33] | | 1.17 [1.08-1.26] | 1,995 | 1,962 | 1,855 | NA | 1,905 |
| M99 | 1.34 [1.17-1.53] | 1.32 [1.15-1.51] | 1.32 [1.15-1.52] | | 1.16 [1.01-1.33] | 1.20 [1.05-1.38] | 1.13 [0.98-1.30] | 1.11 [0.97-1.29] | | 1.09 [0.95-1.25] | 559 | 555 | 521 | NA | 519 |
| Diseases of the genitourinary system (N00-N99) | | | | | | | | | | | | | | | |
| N00 | 1.32 [1.01-1.73] | | | | 1.15 [0.87-1.53] | 1.30 [0.98-1.71] | | | | 1.14 [0.85-1.54] | 140 | NA | NA | NA | 128 |
| N02 | 1.32 [1.22-1.43] | 1.36 [1.25-1.47] | 1.36 [1.25-1.48] | 0.86 [0.66-1.12] | 1.19 [1.10-1.30] | 1.27 [1.17-1.37] | 1.28 [1.17-1.38] | 1.27 [1.17-1.39] | 0.84 [0.64-1.10] | 1.17 [1.07-1.27] | 1,604 | 1,516 | 1,301 | 139 | 1,399 |
| N03 | 1.34 [1.28-1.39] | 1.34 [1.29-1.40] | 1.35 [1.29-1.41] | | 1.26 [1.21-1.32] | 1.27 [1.22-1.32] | 1.22 [1.17-1.27] | 1.22 [1.17-1.28] | | 1.22 [1.17-1.28] | 6,043 | 6,015 | 5,937 | NA | 5,629 |
| N04 | 1.34 [1.22-1.48] | 1.50 [1.35-1.67] | 1.47 [1.31-1.65] | 0.99 [0.82-1.20] | 1.23 [1.12-1.37] | 1.28 [1.16-1.41] | 1.37 [1.23-1.52] | 1.33 [1.19-1.50] | 0.98 [0.81-1.19] | 1.20 [1.08-1.33] | 1,140 | 901 | 762 | 273 | 962 |
| N05 | 1.48 [1.35-1.63] | 1.49 [1.34-1.66] | 1.49 [1.33-1.67] | 1.36 [1.13-1.63] | 1.37 [1.24-1.51] | 1.42 [1.29-1.56] | 1.36 [1.22-1.51] | 1.36 [1.21-1.53] | 1.32 [1.10-1.60] | 1.32 [1.19-1.46] | 1,170 | 893 | 754 | 319 | 1,039 |
| N08 | 1.43 [1.35-1.52] | 1.45 [1.36-1.53] | 1.43 [1.35-1.52] | | 1.28 [1.21-1.36] | 1.35 [1.27-1.43] | 1.26 [1.18-1.33] | 1.22 [1.15-1.31] | | 1.26 [1.19-1.34] | 2,958 | 2,908 | 2,705 | NA | 2,814 |
| N10 | 1.17 [1.07-1.28] | 1.18 [1.07-1.29] | 1.21 [1.07-1.37] | 1.07 [0.92-1.26] | 1.06 [0.97-1.17] | 1.10 [1.01-1.21] | 1.04 [0.94-1.15] | 1.09 [0.96-1.24] | 1.05 [0.89-1.24] | 1.03 [0.93-1.13] | 1,271 | 1,059 | 594 | 411 | 1,091 |
| N11 | 1.25 [1.10-1.42] | 1.24 [1.08-1.41] | 1.29 [1.11-1.51] | | 1.08 [0.94-1.24] | 1.20 [1.05-1.37] | 1.14 [1.00-1.31] | 1.18 [1.01-1.38] | | 1.09 [0.95-1.25] | 572 | 519 | 401 | NA | 508 |
| N12 | 1.18 [1.13-1.22] | 1.21 [1.16-1.25] | 1.28 [1.21-1.35] | 1.05 [0.98-1.12] | 1.10 [1.05-1.14] | 1.10 [1.06-1.15] | 1.07 [1.03-1.12] | 1.13 [1.07-1.19] | 1.02 [0.95-1.10] | 1.06 [1.02-1.11] | 7,028 | 6,129 | 3,401 | 2,174 | 6,119 |
| N13 | 1.06 [1.03-1.09] | 1.07 [1.04-1.10] | 1.07 [1.04-1.10] | 1.05 [0.97-1.15] | 1.01 [0.99-1.04] | 1.05 [1.02-1.08] | 1.03 [1.01-1.06] | 1.04 [1.01-1.07] | 1.02 [0.94-1.12] | 1.02 [0.99-1.05] | 13,110 | 12,270 | 10,762 | 1,400 | 11,521 |
| N14 | 1.34 [1.08-1.66] | 1.31 [1.05-1.63] | 1.41 [1.12-1.77] | | 1.25 [1.00-1.55] | 1.29 [1.03-1.61] | 1.20 [0.96-1.51] | 1.26 [1.00-1.59] | | 1.22 [0.98-1.53] | 220 | 210 | 202 | NA | 212 |
| N15 | 1.20 [1.02-1.41] | 1.21 [1.03-1.43] | 1.21 [1.00-1.46] | | 1.08 [0.91-1.28] | 1.13 [0.96-1.33] | 1.08 [0.91-1.29] | 1.10 [0.91-1.34] | | 1.06 [0.90-1.26] | 372 | 346 | 272 | NA | 338 |
| N16 | 1.61 [1.29-2.00] | 1.65 [1.32-2.06] | 1.51 [1.17-1.95] | | 1.29 [1.03-1.62] | 1.53 [1.22-1.92] | 1.51 [1.20-1.89] | 1.34 [1.03-1.75] | | 1.30 [1.03-1.64] | 221 | 212 | 155 | NA | 195 |
| N17 | 1.23 [1.21-1.24] | 1.22 [1.21-1.23] | 1.23 [1.21-1.24] | 1.07 [0.98-1.17] | 1.17 [1.16-1.19] | 1.19 [1.17-1.20] | 1.13 [1.12-1.15] | 1.14 [1.12-1.15] | 1.07 [0.97-1.17] | 1.16 [1.15-1.18] | 76,985 | 76,432 | 74,248 | 1,369 | 71,764 |
| N18 | 1.26 [1.24-1.28] | 1.26 [1.24-1.28] | 1.25 [1.24-1.27] | 1.14 [0.97-1.34] | 1.19 [1.17-1.20] | 1.21 [1.19-1.22] | 1.15 [1.14-1.17] | 1.15 [1.13-1.16] | 1.12 [0.95-1.33] | 1.16 [1.15-1.18] | 65,241 | 65,046 | 64,301 | 393 | 61,464 |
| N19 | 1.30 [1.26-1.34] | 1.31 [1.27-1.36] | 1.29 [1.25-1.34] | 1.08 [0.91-1.29] | 1.23 [1.19-1.27] | 1.24 [1.20-1.28] | 1.21 [1.17-1.25] | 1.19 [1.15-1.23] | 1.05 [0.87-1.26] | 1.20 [1.16-1.24] | 9,785 | 9,554 | 9,235 | 316 | 8,723 |
| N20 | 1.15 [1.11-1.18] | 1.15 [1.12-1.19] | 1.15 [1.11-1.19] | 0.94 [0.84-1.05] | 1.07 [1.04-1.10] | 1.10 [1.06-1.13] | 1.07 [1.04-1.10] | 1.07 [1.03-1.10] | 0.92 [0.81-1.03] | 1.05 [1.02-1.08] | 10,970 | 10,810 | 8,690 | 738 | 9,284 |
| N21 | 1.02 [0.96-1.08] | 1.01 [0.95-1.08] | 1.03 [0.96-1.10] | | 0.95 [0.89-1.02] | 1.00 [0.94-1.07] | 0.98 [0.92-1.05] | 1.00 [0.94-1.07] | | 0.96 [0.90-1.02] | 2,439 | 2,414 | 2,229 | NA | 2,124 |
| N23 | 1.17 [1.11-1.25] | 1.19 [1.12-1.26] | 1.14 [1.05-1.23] | 1.03 [0.87-1.22] | 1.05 [0.98-1.12] | 1.08 [1.02-1.15] | 1.06 [1.00-1.13] | 1.01 [0.94-1.09] | 0.98 [0.83-1.18] | 1.01 [0.94-1.08] | 2,585 | 2,517 | 1,569 | 334 | 2,068 |
| N25 | 1.31 [1.15-1.48] | 1.39 [1.22-1.59] | 1.39 [1.21-1.60] | | 1.15 [1.00-1.31] | 1.25 [1.10-1.43] | 1.29 [1.12-1.48] | 1.28 [1.11-1.48] | | 1.14 [1.00-1.31] | 592 | 557 | 509 | NA | 546 |
| N26 | 1.20 [1.09-1.32] | 1.18 [1.07-1.30] | 1.18 [1.06-1.31] | | 1.10 [1.00-1.21] | 1.14 [1.03-1.26] | 1.07 [0.97-1.19] | 1.07 [0.96-1.18] | | 1.09 [0.99-1.21] | 1,094 | 1,042 | 952 | NA | 1,040 |
| N27 | 1.30 [1.15-1.47] | 1.34 [1.17-1.53] | 1.37 [1.19-1.58] | 0.96 [0.73-1.27] | 1.12 [0.98-1.28] | 1.27 [1.12-1.44] | 1.25 [1.09-1.43] | 1.29 [1.11-1.49] | 0.94 [0.71-1.25] | 1.12 [0.98-1.27] | 659 | 560 | 496 | 129 | 591 |
| N28 | 1.18 [1.15-1.22] | 1.19 [1.16-1.22] | 1.19 [1.16-1.22] | 0.99 [0.89-1.11] | 1.11 [1.08-1.14] | 1.13 [1.11-1.16] | 1.10 [1.07-1.13] | 1.10 [1.07-1.13] | 0.98 [0.88-1.10] | 1.09 [1.06-1.12] | 15,323 | 14,766 | 13,912 | 808 | 13,992 |
| N29 | 1.32 [1.02-1.71] | 1.48 [1.11-1.97] | | | 1.18 [0.91-1.55] | 1.30 [1.00-1.68] | 1.35 [1.01-1.82] | | | 1.16 [0.88-1.52] | 156 | 121 | NA | NA | 139 |
| N30 | 1.27 [1.22-1.32] | 1.27 [1.22-1.31] | 1.23 [1.18-1.28] | 1.40 [1.21-1.62] | 1.16 [1.12-1.21] | 1.20 [1.16-1.25] | 1.17 [1.12-1.21] | 1.14 [1.09-1.18] | 1.33 [1.15-1.55] | 1.13 [1.09-1.17] | 7,370 | 7,189 | 6,222 | 517 | 6,559 |
| N31 | 1.35 [1.26-1.43] | 1.39 [1.30-1.48] | 1.40 [1.30-1.51] | 0.98 [0.82-1.16] | 1.19 [1.12-1.27] | 1.24 [1.16-1.32] | 1.20 [1.12-1.28] | 1.22 [1.13-1.31] | 0.95 [0.79-1.13] | 1.13 [1.06-1.21] | 2,392 | 2,148 | 1,849 | 319 | 2,190 |
| N32 | 1.23 [1.20-1.26] | 1.22 [1.20-1.25] | 1.22 [1.19-1.25] | 1.16 [1.05-1.27] | 1.14 [1.12-1.17] | 1.17 [1.14-1.19] | 1.13 [1.10-1.16] | 1.14 [1.11-1.16] | 1.10 [0.99-1.21] | 1.11 [1.09-1.14] | 21,199 | 20,497 | 18,959 | 1,124 | 18,793 |
| N34 | 1.35 [1.16-1.58] | 1.34 [1.13-1.59] | 1.25 [1.03-1.52] | | 1.12 [0.94-1.33] | 1.30 [1.11-1.53] | 1.25 [1.05-1.49] | 1.19 [0.97-1.45] | | 1.09 [0.91-1.29] | 404 | 334 | 251 | NA | 327 |
| N35 | 1.31 [1.27-1.36] | 1.31 [1.26-1.36] | 1.30 [1.25-1.35] | 1.33 [1.19-1.49] | 1.20 [1.16-1.25] | 1.25 [1.20-1.30] | 1.21 [1.16-1.26] | 1.21 [1.16-1.26] | 1.29 [1.15-1.45] | 1.17 [1.12-1.22] | 7,679 | 7,156 | 6,216 | 840 | 6,589 |
| N36 | 1.23 [1.15-1.32] | 1.29 [1.20-1.38] | 1.23 [1.14-1.32] | 1.03 [0.85-1.24] | 1.14 [1.06-1.23] | 1.17 [1.10-1.26] | 1.21 [1.12-1.30] | 1.15 [1.06-1.24] | 0.99 [0.82-1.21] | 1.11 [1.03-1.19] | 2,100 | 1,897 | 1,635 | 285 | 1,834 |
| N39 | 1.20 [1.19-1.21] | 1.20 [1.19-1.21] | 1.20 [1.19-1.21] | 1.12 [1.09-1.16] | 1.14 [1.12-1.15] | 1.15 [1.14-1.17] | 1.12 [1.11-1.13] | 1.12 [1.11-1.13] | 1.09 [1.05-1.13] | 1.11 [1.10-1.13] | 96,916 | 90,817 | 82,908 | 8,906 | 86,335 |
| N40 | 1.16 [1.14-1.19] | 1.16 [1.14-1.18] | 1.16 [1.14-1.18] | | 1.10 [1.08-1.12] | 1.12 [1.10-1.14] | 1.10 [1.08-1.12] | 1.10 [1.08-1.12] | | 1.07 [1.05-1.09] | 30,069 | 30,057 | 29,975 | NA | 26,000 |
| N41 | 1.15 [1.08-1.23] | 1.14 [1.07-1.22] | 1.11 [1.03-1.18] | | 1.04 [0.97-1.12] | 1.10 [1.02-1.17] | 1.08 [1.00-1.15] | 1.05 [0.98-1.12] | | 1.01 [0.93-1.08] | 2,047 | 2,045 | 1,963 | NA | 1,726 |
| N42 | 1.11 [1.05-1.18] | 1.11 [1.05-1.18] | 1.13 [1.06-1.20] | | 1.03 [0.97-1.10] | 1.07 [1.00-1.13] | 1.06 [1.00-1.13] | 1.08 [1.01-1.14] | | 1.01 [0.95-1.08] | 2,640 | 2,638 | 2,612 | NA | 2,322 |
| N43 | 1.13 [1.08-1.18] | 1.21 [1.14-1.28] | 1.19 [1.12-1.27] | 1.05 [0.98-1.12] | 1.10 [1.05-1.16] | 1.10 [1.05-1.15] | 1.16 [1.09-1.23] | 1.14 [1.07-1.22] | 1.02 [0.96-1.10] | 1.09 [1.03-1.15] | 4,566 | 2,712 | 2,271 | 2,081 | 3,718 |
| N44 | 1.18 [1.09-1.27] | 1.10 [0.92-1.32] | | 1.14 [1.05-1.23] | 1.15 [1.06-1.26] | 1.16 [1.07-1.25] | 1.02 [0.85-1.24] | | 1.11 [1.03-1.21] | 1.14 [1.04-1.25] | 1,858 | 267 | NA | 1,759 | 1,460 |
| N45 | 1.21 [1.15-1.28] | 1.26 [1.18-1.33] | 1.25 [1.16-1.35] | 1.12 [1.02-1.23] | 1.11 [1.04-1.18] | 1.15 [1.09-1.22] | 1.15 [1.08-1.22] | 1.15 [1.07-1.24] | 1.09 [0.99-1.20] | 1.09 [1.02-1.15] | 3,388 | 2,598 | 1,768 | 1,211 | 2,758 |
| N47 | 1.46 [1.42-1.50] | 1.43 [1.38-1.48] | 1.38 [1.32-1.45] | 1.45 [1.41-1.50] | 1.37 [1.33-1.42] | 1.40 [1.37-1.44] | 1.32 [1.27-1.38] | 1.28 [1.22-1.35] | 1.41 [1.36-1.46] | 1.34 [1.30-1.38] | 15,415 | 6,884 | 4,410 | 9,851 | 11,705 |
| N48 | 1.40 [1.35-1.46] | 1.44 [1.37-1.50] | 1.40 [1.33-1.48] | 1.35 [1.28-1.43] | 1.28 [1.23-1.34] | 1.34 [1.29-1.40] | 1.32 [1.26-1.39] | 1.29 [1.23-1.36] | 1.32 [1.24-1.40] | 1.26 [1.21-1.31] | 7,614 | 4,656 | 3,736 | 3,369 | 6,219 |
| N49 | 1.35 [1.21-1.50] | 1.38 [1.23-1.56] | 1.41 [1.23-1.60] | 1.13 [0.88-1.44] | 1.18 [1.05-1.33] | 1.29 [1.16-1.45] | 1.27 [1.13-1.44] | 1.28 [1.12-1.47] | 1.11 [0.87-1.43] | 1.19 [1.05-1.34] | 822 | 691 | 574 | 171 | 680 |
| N50 | 1.22 [1.17-1.26] | 1.25 [1.20-1.31] | 1.26 [1.20-1.33] | 1.12 [1.05-1.19] | 1.15 [1.10-1.19] | 1.16 [1.12-1.21] | 1.16 [1.11-1.21] | 1.17 [1.11-1.23] | 1.08 [1.02-1.15] | 1.12 [1.08-1.17] | 7,328 | 5,008 | 3,731 | 2,956 | 6,009 |
| N51 | 1.48 [1.08-2.03] | | | | | 1.37 [0.99-1.90] | | | | | 108 | NA | NA | NA | NA |
| N60 | 1.17 [1.10-1.25] | 1.14 [1.07-1.22] | 1.11 [1.03-1.19] | 1.37 [1.01-1.85] | 1.10 [1.02-1.18] | 1.13 [1.06-1.21] | 1.09 [1.02-1.16] | 1.05 [0.97-1.13] | 1.34 [0.98-1.84] | 1.09 [1.01-1.17] | 2,183 | 2,148 | 1,645 | 118 | 1,789 |
| N61 | 1.34 [1.24-1.44] | 1.38 [1.27-1.49] | 1.39 [1.25-1.54] | 1.26 [1.03-1.54] | 1.27 [1.17-1.38] | 1.28 [1.18-1.38] | 1.24 [1.14-1.35] | 1.25 [1.12-1.39] | 1.22 [0.99-1.51] | 1.26 [1.15-1.37] | 1,671 | 1,585 | 906 | 262 | 1,429 |
| N62 | 1.28 [1.18-1.39] | 1.28 [1.17-1.39] | 1.27 [1.13-1.42] | 1.30 [1.09-1.55] | 1.16 [1.05-1.27] | 1.21 [1.11-1.32] | 1.15 [1.05-1.26] | 1.11 [0.99-1.25] | 1.26 [1.05-1.51] | 1.12 [1.01-1.23] | 1,388 | 1,242 | 739 | 339 | 1,060 |
| N63 | 1.16 [1.09-1.23] | 1.17 [1.09-1.24] | 1.16 [1.08-1.25] | 1.19 [0.93-1.53] | 1.10 [1.02-1.18] | 1.11 [1.04-1.18] | 1.09 [1.02-1.16] | 1.08 [1.00-1.16] | 1.16 [0.90-1.50] | 1.08 [1.00-1.16] | 2,320 | 2,264 | 1,839 | 163 | 1,932 |
| N64 | 1.25 [1.18-1.34] | 1.25 [1.17-1.33] | 1.21 [1.12-1.30] | 1.37 [1.09-1.71] | 1.18 [1.11-1.27] | 1.19 [1.12-1.27] | 1.15 [1.07-1.23] | 1.11 [1.03-1.20] | 1.35 [1.07-1.70] | 1.15 [1.08-1.24] | 2,374 | 2,310 | 1,728 | 217 | 2,070 |
| N70 | 1.07 [0.99-1.15] | 1.08 [1.00-1.17] | 1.06 [0.95-1.18] | 1.08 [0.85-1.37] | 1.00 [0.92-1.09] | 1.03 [0.95-1.11] | 1.01 [0.93-1.09] | 1.00 [0.89-1.11] | 1.06 [0.83-1.35] | 0.98 [0.90-1.07] | 1,521 | 1,491 | 779 | 181 | 1,282 |
| N71 | 1.17 [1.05-1.31] | 1.23 [1.10-1.37] | 1.25 [1.09-1.42] | | 1.14 [1.02-1.28] | 1.08 [0.97-1.21] | 1.12 [1.01-1.26] | 1.13 [0.99-1.30] | | 1.08 [0.96-1.22] | 792 | 791 | 507 | NA | 699 |
| N72 | 1.09 [1.01-1.18] | 1.12 [1.04-1.21] | 1.12 [1.01-1.24] | 0.95 [0.72-1.26] | 1.04 [0.96-1.14] | 1.04 [0.96-1.13] | 1.06 [0.98-1.15] | 1.07 [0.96-1.19] | 0.96 [0.72-1.28] | 1.02 [0.93-1.12] | 1,444 | 1,441 | 782 | 126 | 1,136 |
| N73 | 1.14 [1.10-1.17] | 1.16 [1.12-1.20] | 1.13 [1.08-1.18] | 1.11 [1.01-1.21] | 1.07 [1.03-1.11] | 1.07 [1.04-1.11] | 1.06 [1.02-1.09] | 1.04 [0.99-1.09] | 1.08 [0.98-1.18] | 1.03 [1.00-1.07] | 8,766 | 8,580 | 4,006 | 1,360 | 7,511 |
| N75 | 1.06 [0.97-1.15] | 1.04 [0.96-1.13] | 1.06 [0.93-1.20] | 0.93 [0.77-1.11] | 1.01 [0.92-1.10] | 1.04 [0.95-1.12] | 0.99 [0.91-1.07] | 1.02 [0.89-1.16] | 0.93 [0.77-1.12] | 1.00 [0.91-1.10] | 1,430 | 1,376 | 516 | 298 | 1,118 |
| N76 | 1.39 [1.31-1.47] | 1.44 [1.35-1.53] | 1.51 [1.39-1.64] | 1.22 [1.09-1.38] | 1.32 [1.24-1.41] | 1.31 [1.23-1.39] | 1.29 [1.21-1.38] | 1.37 [1.26-1.49] | 1.17 [1.04-1.32] | 1.27 [1.19-1.36] | 2,790 | 2,348 | 1,438 | 759 | 2,369 |
| N77 | 1.28 [1.21-1.36] | 1.30 [1.23-1.38] | 1.40 [1.27-1.54] | 1.10 [0.99-1.22] | 1.19 [1.12-1.26] | 1.19 [1.12-1.26] | 1.13 [1.06-1.20] | 1.19 [1.07-1.31] | 1.06 [0.95-1.18] | 1.13 [1.06-1.20] | 3,197 | 2,984 | 1,073 | 924 | 2,821 |
| N80 | 1.13 [1.09-1.17] | 1.15 [1.11-1.18] | 1.09 [1.03-1.14] | 1.16 [1.06-1.27] | 1.08 [1.04-1.12] | 1.06 [1.02-1.10] | 1.04 [1.00-1.07] | 1.00 [0.95-1.05] | 1.12 [1.02-1.22] | 1.03 [0.99-1.07] | 8,651 | 8,539 | 3,717 | 1,438 | 7,115 |
| N81 | 1.21 [1.18-1.24] | 1.20 [1.17-1.23] | 1.20 [1.17-1.23] | 1.27 [0.95-1.69] | 1.15 [1.12-1.18] | 1.14 [1.12-1.17] | 1.13 [1.10-1.16] | 1.12 [1.09-1.15] | 1.26 [0.94-1.70] | 1.10 [1.07-1.13] | 14,255 | 14,238 | 12,975 | 129 | 12,058 |
| N82 | 1.32 [1.18-1.49] | 1.32 [1.17-1.48] | 1.30 [1.15-1.48] | | 1.23 [1.09-1.38] | 1.24 [1.11-1.40] | 1.21 [1.07-1.36] | 1.19 [1.05-1.36] | | 1.19 [1.05-1.34] | 730 | 718 | 593 | NA | 667 |
| N83 | 1.13 [1.10-1.16] | 1.15 [1.12-1.18] | 1.12 [1.08-1.16] | 1.11 [1.05-1.17] | 1.08 [1.05-1.11] | 1.07 [1.04-1.10] | 1.04 [1.01-1.07] | 1.04 [1.00-1.07] | 1.06 [1.00-1.13] | 1.04 [1.01-1.07] | 14,639 | 13,619 | 6,998 | 3,120 | 12,225 |
| N84 | 1.12 [1.09-1.16] | 1.13 [1.09-1.16] | 1.12 [1.09-1.16] | 1.17 [0.93-1.48] | 1.08 [1.05-1.12] | 1.10 [1.07-1.13] | 1.09 [1.06-1.12] | 1.09 [1.06-1.13] | 1.09 [0.86-1.39] | 1.07 [1.03-1.10] | 11,263 | 11,250 | 9,786 | 199 | 8,858 |
| N85 | 1.17 [1.14-1.21] | 1.19 [1.16-1.23] | 1.18 [1.14-1.22] | 1.17 [1.01-1.35] | 1.12 [1.09-1.16] | 1.13 [1.09-1.16] | 1.11 [1.08-1.15] | 1.11 [1.07-1.15] | 1.13 [0.97-1.32] | 1.09 [1.05-1.13] | 9,361 | 9,313 | 7,312 | 516 | 7,912 |
| N86 | 1.19 [1.12-1.26] | 1.20 [1.13-1.27] | 1.23 [1.10-1.37] | 1.07 [0.94-1.22] | 1.13 [1.06-1.20] | 1.13 [1.07-1.20] | 1.12 [1.05-1.19] | 1.17 [1.04-1.31] | 1.04 [0.91-1.19] | 1.08 [1.01-1.15] | 2,864 | 2,823 | 716 | 627 | 2,300 |
| N87 | 1.10 [1.05-1.17] | 1.13 [1.07-1.19] | 1.06 [0.97-1.17] | 1.13 [0.98-1.31] | 1.05 [0.99-1.12] | 1.06 [1.01-1.12] | 1.06 [1.00-1.12] | 1.01 [0.91-1.11] | 1.15 [0.98-1.34] | 1.05 [0.98-1.11] | 3,251 | 3,250 | 962 | 461 | 2,421 |
| N88 | 1.21 [1.14-1.28] | 1.21 [1.14-1.28] | 1.19 [1.12-1.27] | 1.08 [0.78-1.49] | 1.12 [1.05-1.19] | 1.16 [1.10-1.23] | 1.15 [1.08-1.22] | 1.12 [1.05-1.20] | 1.10 [0.78-1.55] | 1.10 [1.03-1.17] | 2,805 | 2,806 | 2,255 | 101 | 2,351 |
| N89 | 1.29 [1.23-1.34] | 1.30 [1.25-1.36] | 1.30 [1.22-1.39] | 1.10 [1.00-1.21] | 1.17 [1.12-1.23] | 1.21 [1.16-1.27] | 1.18 [1.13-1.24] | 1.21 [1.13-1.29] | 1.07 [0.97-1.17] | 1.12 [1.07-1.18] | 5,156 | 4,822 | 2,065 | 1,179 | 4,386 |
| N90 | 1.38 [1.32-1.45] | 1.37 [1.31-1.43] | 1.43 [1.36-1.51] | 1.23 [1.08-1.41] | 1.27 [1.21-1.34] | 1.31 [1.25-1.37] | 1.26 [1.20-1.32] | 1.32 [1.25-1.39] | 1.22 [1.06-1.41] | 1.22 [1.16-1.28] | 4,972 | 4,765 | 3,303 | 553 | 4,201 |
| N91 | 1.29 [1.13-1.47] | 1.30 [1.13-1.49] | 1.52 [1.19-1.93] | 1.14 [0.92-1.41] | 1.15 [1.00-1.32] | 1.22 [1.06-1.39] | 1.14 [0.99-1.32] | 1.41 [1.10-1.81] | 1.11 [0.89-1.38] | 1.10 [0.95-1.27] | 609 | 491 | 160 | 239 | 485 |
| N92 | 1.21 [1.18-1.23] | 1.21 [1.18-1.24] | 1.18 [1.15-1.21] | 1.21 [1.13-1.30] | 1.15 [1.12-1.17] | 1.13 [1.11-1.16] | 1.10 [1.08-1.13] | 1.09 [1.06-1.12] | 1.16 [1.08-1.25] | 1.10 [1.07-1.13] | 19,969 | 19,184 | 12,084 | 2,231 | 16,515 |
| N93 | 1.20 [1.16-1.24] | 1.21 [1.17-1.25] | 1.18 [1.13-1.24] | 1.14 [1.06-1.23] | 1.11 [1.07-1.14] | 1.14 [1.10-1.17] | 1.10 [1.06-1.13] | 1.09 [1.04-1.14] | 1.11 [1.02-1.20] | 1.07 [1.04-1.11] | 9,184 | 8,788 | 4,249 | 1,741 | 7,743 |
| N94 | 1.26 [1.21-1.30] | 1.28 [1.24-1.33] | 1.27 [1.20-1.34] | 1.24 [1.15-1.34] | 1.17 [1.13-1.22] | 1.16 [1.12-1.20] | 1.13 [1.09-1.17] | 1.14 [1.08-1.21] | 1.18 [1.09-1.28] | 1.11 [1.07-1.16] | 7,792 | 7,307 | 2,895 | 1,794 | 6,450 |
| N95 | 1.25 [1.21-1.29] | 1.25 [1.21-1.29] | 1.24 [1.21-1.28] | | 1.19 [1.15-1.23] | 1.20 [1.16-1.24] | 1.18 [1.15-1.22] | 1.17 [1.14-1.21] | | 1.16 [1.12-1.20] | 10,449 | 10,449 | 10,388 | NA | 8,443 |
| N96 | 1.08 [0.83-1.41] | 1.11 [0.85-1.44] | | | 0.99 [0.75-1.30] | 1.07 [0.81-1.41] | 1.07 [0.81-1.41] | | | 1.00 [0.76-1.33] | 134 | 134 | NA | NA | 120 |
| N97 | 0.93 [0.86-1.01] | 0.95 [0.87-1.03] | 0.97 [0.75-1.26] | | 0.97 [0.88-1.08] | 0.92 [0.85-1.00] | 0.93 [0.86-1.01] | 0.98 [0.76-1.27] | | 0.96 [0.86-1.07] | 1,306 | 1,306 | 125 | NA | 800 |
| N98 | 0.80 [0.63-1.03] | 0.81 [0.64-1.04] | | | 0.80 [0.60-1.06] | 0.76 [0.59-0.98] | 0.76 [0.59-0.98] | | | 0.77 [0.57-1.02] | 143 | 143 | NA | NA | 105 |
| N99 | 1.22 [1.15-1.30] | 1.25 [1.18-1.33] | 1.22 [1.14-1.31] | 1.01 [0.80-1.26] | 1.13 [1.06-1.20] | 1.17 [1.10-1.25] | 1.17 [1.10-1.25] | 1.14 [1.06-1.22] | 0.99 [0.79-1.25] | 1.11 [1.04-1.18] | 2,593 | 2,456 | 2,020 | 199 | 2,309 |
| Pregnancy, childbirth and the puerperium (O00-O9A) | | | | | | | | | | | | | | | |
| O00 | 1.04 [0.98-1.11] | 1.09 [1.03-1.16] | 1.03 [0.81-1.31] | 1.02 [0.90-1.16] | 0.99 [0.92-1.05] | 1.02 [0.95-1.08] | 1.03 [0.97-1.10] | 1.02 [0.80-1.30] | 0.99 [0.87-1.13] | 0.98 [0.92-1.05] | 2,598 | 2,578 | 146 | 620 | 2,122 |
| O01 | 1.17 [0.92-1.49] | 1.18 [0.93-1.51] | | | 1.14 [0.87-1.48] | 1.19 [0.92-1.53] | 1.17 [0.91-1.50] | | | 1.21 [0.92-1.59] | 165 | 161 | NA | NA | 132 |
| O02 | 1.10 [1.05-1.14] | 1.11 [1.06-1.15] | 1.02 [0.91-1.15] | 1.05 [0.95-1.15] | 1.04 [1.00-1.09] | 1.07 [1.02-1.11] | 1.07 [1.03-1.11] | 1.00 [0.89-1.13] | 1.04 [0.94-1.14] | 1.03 [0.99-1.08] | 6,033 | 5,967 | 628 | 1,190 | 4,823 |
| O03 | 1.06 [1.02-1.09] | 1.07 [1.04-1.11] | 1.04 [0.95-1.14] | 1.00 [0.94-1.07] | 1.01 [0.97-1.05] | 1.04 [1.00-1.07] | 1.03 [1.00-1.07] | 1.03 [0.94-1.14] | 0.99 [0.92-1.06] | 1.00 [0.97-1.04] | 9,004 | 8,798 | 949 | 2,142 | 7,259 |
| O04 | 1.02 [1.00-1.05] | 1.04 [1.01-1.07] | 0.92 [0.81-1.04] | 0.98 [0.94-1.03] | 0.94 [0.91-0.98] | 1.01 [0.98-1.04] | 0.99 [0.96-1.03] | 0.90 [0.79-1.03] | 0.98 [0.94-1.03] | 0.95 [0.92-0.98] | 11,751 | 10,339 | 521 | 5,427 | 8,510 |
| O08 | 1.05 [0.95-1.16] | 1.09 [0.98-1.20] | | 0.99 [0.83-1.18] | 0.97 [0.87-1.09] | 1.01 [0.91-1.12] | 1.01 [0.91-1.12] | | 0.99 [0.83-1.18] | 0.97 [0.87-1.09] | 942 | 916 | NA | 336 | 792 |
| O10 | 1.13 [1.02-1.25] | 1.16 [1.05-1.28] | 1.27 [1.01-1.59] | 0.95 [0.72-1.25] | 1.09 [0.98-1.21] | 1.10 [0.99-1.21] | 1.07 [0.97-1.19] | 1.06 [0.83-1.34] | 0.95 [0.71-1.27] | 1.06 [0.96-1.19] | 994 | 995 | 170 | 132 | 851 |
| O11 | 1.17 [0.90-1.51] | 1.26 [0.98-1.63] | | | 1.12 [0.85-1.48] | 1.12 [0.86-1.45] | 1.16 [0.88-1.52] | | | 1.09 [0.82-1.44] | 149 | 148 | NA | NA | 127 |
| O12 | 1.13 [1.06-1.21] | 1.15 [1.08-1.23] | | 1.10 [0.97-1.25] | 1.09 [1.01-1.17] | 1.08 [1.01-1.16] | 1.06 [0.99-1.14] | | 1.08 [0.95-1.23] | 1.05 [0.97-1.14] | 2,093 | 2,054 | NA | 628 | 1,647 |
| O13 | 1.08 [1.04-1.13] | 1.11 [1.07-1.16] | 0.96 [0.81-1.14] | 1.04 [0.96-1.13] | 1.05 [1.00-1.10] | 1.06 [1.01-1.11] | 1.08 [1.04-1.13] | 0.96 [0.81-1.14] | 1.02 [0.93-1.11] | 1.03 [0.98-1.08] | 5,577 | 5,523 | 288 | 1,424 | 4,037 |
| O14 | 1.10 [1.05-1.16] | 1.12 [1.07-1.18] | 0.98 [0.80-1.20] | 1.12 [1.02-1.22] | 1.06 [1.00-1.13] | 1.07 [1.02-1.13] | 1.09 [1.03-1.14] | 0.96 [0.78-1.18] | 1.12 [1.02-1.22] | 1.03 [0.97-1.09] | 4,133 | 4,062 | 205 | 1,285 | 2,898 |
| O15 | 1.10 [0.86-1.41] | 1.12 [0.88-1.44] | | | 1.05 [0.79-1.40] | 1.08 [0.84-1.39] | 1.08 [0.83-1.39] | | | 1.00 [0.74-1.35] | 159 | 153 | NA | NA | 108 |
| O16 | 1.12 [1.07-1.17] | 1.12 [1.07-1.18] | 1.03 [0.86-1.24] | 1.10 [1.00-1.21] | 1.08 [1.02-1.14] | 1.09 [1.04-1.14] | 1.08 [1.03-1.14] | 1.02 [0.84-1.24] | 1.08 [0.98-1.19] | 1.06 [1.01-1.12] | 4,610 | 4,502 | 243 | 1,090 | 3,230 |
| O20 | 1.06 [1.03-1.10] | 1.10 [1.06-1.14] | 0.97 [0.83-1.13] | 1.00 [0.94-1.07] | 1.00 [0.96-1.04] | 1.03 [0.99-1.07] | 1.03 [1.00-1.07] | 0.98 [0.84-1.15] | 1.00 [0.93-1.06] | 0.99 [0.95-1.03] | 8,064 | 7,869 | 356 | 2,475 | 6,494 |
| O21 | 1.11 [1.06-1.15] | 1.16 [1.11-1.21] | | 1.03 [0.97-1.10] | 1.02 [0.97-1.06] | 1.04 [1.00-1.09] | 1.05 [1.00-1.09] | | 1.01 [0.94-1.08] | 0.99 [0.94-1.03] | 5,975 | 5,800 | NA | 2,422 | 4,869 |
| O22 | 1.11 [1.00-1.23] | 1.10 [1.00-1.22] | | 0.95 [0.74-1.22] | 1.00 [0.90-1.12] | 1.09 [0.98-1.21] | 1.07 [0.96-1.19] | | 0.90 [0.69-1.17] | 1.00 [0.90-1.12] | 915 | 913 | NA | 157 | 805 |
| O23 | 1.16 [1.12-1.20] | 1.20 [1.15-1.24] | 0.93 [0.73-1.19] | 1.03 [0.97-1.10] | 1.08 [1.04-1.13] | 1.11 [1.07-1.15] | 1.09 [1.05-1.14] | 0.89 [0.69-1.14] | 1.02 [0.96-1.09] | 1.06 [1.02-1.11] | 6,682 | 6,472 | 135 | 2,480 | 5,486 |
| O24 | 1.07 [1.03-1.11] | 1.09 [1.06-1.13] | 1.04 [0.93-1.16] | 1.00 [0.92-1.09] | 1.02 [0.98-1.06] | 1.05 [1.01-1.09] | 1.07 [1.03-1.11] | 1.03 [0.92-1.16] | 1.02 [0.94-1.11] | 1.01 [0.97-1.05] | 7,581 | 7,552 | 683 | 1,507 | 6,382 |
| O26 | 1.16 [1.13-1.18] | 1.19 [1.17-1.22] | 0.95 [0.85-1.05] | 1.05 [1.01-1.08] | 1.07 [1.05-1.10] | 1.10 [1.08-1.13] | 1.10 [1.07-1.12] | 0.92 [0.83-1.03] | 1.02 [0.99-1.06] | 1.04 [1.02-1.07] | 25,422 | 24,733 | 756 | 8,267 | 19,725 |
| O28 | 1.06 [0.99-1.13] | 1.09 [1.02-1.16] | 0.97 [0.75-1.25] | 0.98 [0.87-1.10] | 0.99 [0.92-1.06] | 1.01 [0.95-1.08] | 1.01 [0.94-1.08] | 0.94 [0.72-1.23] | 0.97 [0.86-1.09] | 0.96 [0.90-1.03] | 2,337 | 2,318 | 122 | 778 | 1,957 |
| O30 | 1.00 [0.94-1.06] | 1.01 [0.96-1.08] | 0.86 [0.69-1.07] | 1.08 [0.94-1.23] | 0.95 [0.89-1.02] | 0.98 [0.92-1.04] | 0.98 [0.92-1.04] | 0.88 [0.70-1.10] | 1.08 [0.94-1.24] | 0.95 [0.88-1.01] | 2,655 | 2,647 | 170 | 573 | 2,178 |
| O31 | 1.05 [0.82-1.34] | 1.10 [0.86-1.41] | | | 1.08 [0.84-1.39] | 1.01 [0.78-1.30] | 1.01 [0.78-1.31] | | | 1.07 [0.82-1.39] | 158 | 157 | NA | NA | 140 |
| O32 | 1.03 [1.00-1.06] | 1.06 [1.03-1.09] | 0.96 [0.85-1.08] | 0.96 [0.90-1.02] | 0.99 [0.96-1.02] | 1.02 [0.99-1.05] | 1.04 [1.01-1.07] | 0.98 [0.87-1.11] | 0.95 [0.89-1.01] | 0.98 [0.95-1.02] | 11,077 | 11,025 | 591 | 2,583 | 8,577 |
| O33 | 1.15 [0.95-1.39] | 1.19 [0.99-1.44] | | | 1.02 [0.83-1.26] | 1.13 [0.93-1.37] | 1.16 [0.96-1.40] | | | 1.01 [0.82-1.24] | 265 | 266 | NA | NA | 209 |
| O34 | 1.09 [1.06-1.11] | 1.10 [1.08-1.13] | 1.00 [0.93-1.08] | 0.98 [0.92-1.04] | 1.02 [1.00-1.04] | 1.07 [1.05-1.10] | 1.08 [1.06-1.11] | 1.03 [0.95-1.12] | 0.97 [0.91-1.03] | 1.02 [0.99-1.04] | 18,898 | 18,872 | 1,365 | 3,020 | 17,262 |
| O35 | 1.02 [0.96-1.08] | 1.06 [1.00-1.13] | 0.97 [0.80-1.16] | 1.00 [0.89-1.13] | 0.99 [0.93-1.05] | 1.00 [0.94-1.06] | 1.03 [0.97-1.10] | 0.95 [0.78-1.15] | 0.99 [0.88-1.13] | 0.98 [0.92-1.05] | 2,724 | 2,691 | 243 | 688 | 2,259 |
| O36 | 1.12 [1.10-1.14] | 1.15 [1.13-1.17] | 1.02 [0.93-1.11] | 1.03 [1.00-1.06] | 1.04 [1.02-1.07] | 1.08 [1.06-1.10] | 1.09 [1.07-1.11] | 1.02 [0.93-1.11] | 1.02 [0.99-1.05] | 1.03 [1.01-1.05] | 33,543 | 32,909 | 1,131 | 11,623 | 25,591 |
| O40 | 1.09 [1.03-1.16] | 1.10 [1.04-1.17] | 0.95 [0.76-1.19] | 1.04 [0.92-1.18] | 1.04 [0.98-1.11] | 1.05 [0.99-1.12] | 1.04 [0.98-1.11] | 0.96 [0.76-1.22] | 1.06 [0.93-1.20] | 1.02 [0.96-1.09] | 2,620 | 2,607 | 158 | 685 | 2,291 |
| O41 | 1.10 [1.04-1.16] | 1.12 [1.06-1.18] | 0.99 [0.77-1.27] | 0.99 [0.89-1.09] | 1.04 [0.98-1.11] | 1.08 [1.02-1.14] | 1.08 [1.02-1.14] | 1.05 [0.81-1.36] | 1.00 [0.90-1.10] | 1.04 [0.98-1.10] | 3,493 | 3,440 | 132 | 985 | 2,693 |

#### Supplementary Table 3: Hazard ratios and events from all cohorts

| | Hazard ratio (99% confidence interval) | | | | | | | | | | Events (in exposed) | | | | |
|---|---|---|---|---|---|---|---|---|---|---|---|---|---|---|---|
| | crude | | | | | adjusted | | | | | | | | | |
| Outcome | any age | 18+ | 40+ | <18 | hosp. | any age | 18+ | 40+ | <18 | hosp. | any age | 18+ | 40+ | <18 | hosp. |
| O42 | 1.03 [1.01-1.06] | 1.06 [1.04-1.08] | 0.84 [0.75-0.95] | 1.00 [0.96-1.05] | 1.01 [0.98-1.04] | 1.03 [1.01-1.05] | 1.04 [1.02-1.07] | 0.89 [0.78-1.00] | 1.00 [0.95-1.04] | 1.02 [0.99-1.05] | 18,440 | 18,245 | 557 | 5,216 | 13,509 |
| O43 | 1.03 [0.97-1.10] | 1.06 [0.99-1.12] | 0.94 [0.71-1.25] | 0.94 [0.83-1.06] | 1.01 [0.94-1.08] | 1.02 [0.96-1.09] | 1.03 [0.97-1.10] | 0.92 [0.69-1.23] | 0.96 [0.84-1.09] | 1.00 [0.94-1.08] | 2,479 | 2,451 | 103 | 627 | 1,950 |
| O44 | 1.03 [0.95-1.11] | 1.04 [0.96-1.12] | 0.98 [0.76-1.25] | 0.90 [0.74-1.09] | 1.01 [0.94-1.10] | 0.99 [0.91-1.07] | 0.99 [0.92-1.07] | 0.97 [0.75-1.26] | 0.87 [0.71-1.06] | 1.00 [0.92-1.08] | 1,648 | 1,640 | 134 | 264 | 1,419 |
| O45 | 1.02 [0.91-1.14] | 1.05 [0.94-1.17] | | 1.01 [0.80-1.26] | 1.00 [0.88-1.13] | 1.00 [0.89-1.12] | 1.01 [0.90-1.14] | | 1.03 [0.82-1.31] | 0.99 [0.88-1.13] | 734 | 733 | NA | 192 | 605 |
| O46 | 1.12 [1.09-1.16] | 1.16 [1.12-1.19] | 1.04 [0.89-1.22] | 1.04 [0.97-1.10] | 1.06 [1.02-1.10] | 1.09 [1.05-1.13] | 1.09 [1.06-1.13] | 1.04 [0.88-1.22] | 1.02 [0.96-1.09] | 1.05 [1.01-1.09] | 8,965 | 8,804 | 333 | 2,539 | 6,973 |
| O47 | 1.16 [1.12-1.21] | 1.17 [1.13-1.22] | 0.95 [0.75-1.20] | 1.05 [0.97-1.14] | 1.07 [1.02-1.12] | 1.12 [1.07-1.16] | 1.10 [1.06-1.14] | 0.91 [0.71-1.16] | 1.04 [0.95-1.13] | 1.05 [1.00-1.10] | 6,347 | 6,178 | 146 | 1,471 | 4,703 |
| O48 | 1.02 [0.99-1.05] | 1.03 [1.00-1.06] | 0.84 [0.71-1.00] | 0.97 [0.92-1.03] | 0.97 [0.93-1.00] | 1.02 [0.99-1.05] | 1.03 [1.00-1.06] | 0.89 [0.75-1.06] | 0.97 [0.91-1.03] | 0.97 [0.94-1.01] | 10,930 | 10,772 | 279 | 2,881 | 7,536 |
| O60 | 1.08 [1.04-1.11] | 1.11 [1.07-1.14] | 1.06 [0.93-1.21] | 1.02 [0.96-1.08] | 1.03 [0.99-1.06] | 1.05 [1.01-1.08] | 1.05 [1.02-1.09] | 1.03 [0.90-1.18] | 1.02 [0.96-1.09] | 1.02 [0.98-1.06] | 9,664 | 9,541 | 479 | 2,675 | 7,666 |
| O61 | 1.08 [1.01-1.17] | 1.11 [1.03-1.19] | 0.98 [0.75-1.27] | 1.10 [0.94-1.27] | 1.05 [0.97-1.15] | 1.05 [0.97-1.13] | 1.06 [0.98-1.14] | 0.92 [0.70-1.21] | 1.05 [0.90-1.23] | 1.02 [0.93-1.11] | 1,800 | 1,784 | 120 | 471 | 1,298 |
| O62 | 1.05 [1.01-1.09] | 1.08 [1.04-1.12] | 0.96 [0.81-1.15] | 0.97 [0.91-1.04] | 1.02 [0.98-1.07] | 1.05 [1.01-1.09] | 1.06 [1.02-1.10] | 0.99 [0.83-1.19] | 0.97 [0.90-1.04] | 1.03 [0.98-1.07] | 7,213 | 7,138 | 271 | 2,058 | 5,453 |
| O63 | 1.02 [0.99-1.04] | 1.03 [1.01-1.06] | 0.91 [0.80-1.03] | 1.00 [0.95-1.05] | 1.01 [0.98-1.04] | 1.01 [0.98-1.03] | 1.02 [1.00-1.05] | 0.96 [0.84-1.10] | 0.98 [0.93-1.03] | 1.00 [0.97-1.03] | 15,999 | 15,795 | 487 | 4,449 | 10,045 |
| O64 | 1.02 [0.97-1.08] | 1.03 [0.97-1.09] | | 1.08 [0.96-1.22] | 1.01 [0.94-1.07] | 1.02 [0.96-1.08] | 1.03 [0.97-1.09] | | 1.07 [0.94-1.20] | 1.00 [0.94-1.07] | 3,014 | 2,962 | NA | 700 | 2,033 |
| O65 | 0.98 [0.80-1.21] | 1.01 [0.82-1.24] | | | 0.99 [0.78-1.26] | 0.99 [0.80-1.22] | 1.04 [0.84-1.29] | | | 1.02 [0.79-1.31] | 207 | 205 | NA | NA | 144 |
| O66 | 1.01 [0.96-1.07] | 1.03 [0.97-1.09] | 0.95 [0.73-1.24] | 0.90 [0.80-1.01] | 0.99 [0.93-1.06] | 1.00 [0.94-1.06] | 1.01 [0.96-1.07] | 0.97 [0.74-1.28] | 0.90 [0.80-1.01] | 0.99 [0.93-1.06] | 2,954 | 2,917 | 115 | 705 | 2,146 |
| O67 | 1.02 [0.91-1.13] | 1.07 [0.96-1.19] | | 1.07 [0.85-1.36] | 1.01 [0.89-1.14] | 1.01 [0.90-1.13] | 1.06 [0.95-1.19] | | 1.10 [0.86-1.41] | 0.99 [0.88-1.13] | 776 | 771 | NA | 178 | 596 |
| O68 | 1.04 [1.03-1.06] | 1.08 [1.06-1.09] | 0.93 [0.85-1.01] | 1.00 [0.97-1.03] | 1.01 [0.99-1.03] | 1.03 [1.02-1.05] | 1.06 [1.04-1.07] | 0.95 [0.87-1.04] | 1.00 [0.97-1.03] | 1.00 [0.98-1.02] | 35,814 | 35,282 | 1,175 | 10,799 | 24,226 |
| O69 | 1.09 [1.03-1.15] | 1.10 [1.04-1.16] | 1.18 [0.93-1.49] | 0.99 [0.88-1.11] | 1.03 [0.97-1.10] | 1.08 [1.02-1.14] | 1.10 [1.04-1.16] | 1.22 [0.95-1.56] | 0.99 [0.88-1.12] | 1.04 [0.98-1.11] | 3,144 | 3,121 | 152 | 714 | 2,453 |
| O70 | 1.03 [1.02-1.05] | 1.06 [1.04-1.08] | 0.93 [0.85-1.01] | 0.97 [0.95-1.00] | 0.98 [0.96-0.99] | 1.04 [1.02-1.05] | 1.06 [1.04-1.07] | 1.00 [0.92-1.10] | 0.98 [0.95-1.00] | 0.99 [0.97-1.01] | 48,711 | 47,702 | 1,170 | 15,359 | 33,463 |
| O71 | 1.01 [0.96-1.08] | 1.05 [0.99-1.11] | | 1.00 [0.90-1.12] | 1.00 [0.93-1.06] | 1.01 [0.95-1.07] | 1.05 [0.98-1.11] | | 1.02 [0.91-1.14] | 1.00 [0.94-1.07] | 2,697 | 2,647 | NA | 802 | 2,035 |
| O72 | 1.05 [1.03-1.08] | 1.08 [1.06-1.10] | 0.97 [0.89-1.06] | 0.98 [0.94-1.02] | 1.02 [1.00-1.04] | 1.05 [1.02-1.07] | 1.07 [1.05-1.09] | 1.00 [0.92-1.10] | 0.97 [0.93-1.02] | 1.02 [0.99-1.04] | 23,598 | 23,373 | 1,079 | 6,369 | 17,714 |
| O73 | 1.10 [1.02-1.19] | 1.11 [1.03-1.20] | | 1.10 [0.93-1.31] | 1.04 [0.95-1.13] | 1.09 [1.01-1.18] | 1.08 [1.00-1.17] | | 1.11 [0.93-1.32] | 1.03 [0.95-1.13] | 1,593 | 1,576 | NA | 333 | 1,234 |
| O74 | 1.16 [1.00-1.34] | 1.15 [1.00-1.34] | | 0.88 [0.66-1.16] | 1.12 [0.96-1.31] | 1.12 [0.96-1.30] | 1.10 [0.95-1.28] | | 0.87 [0.65-1.16] | 1.10 [0.94-1.30] | 464 | 453 | NA | 122 | 378 |
| O75 | 1.04 [1.01-1.07] | 1.06 [1.03-1.09] | 0.91 [0.79-1.05] | 0.95 [0.90-1.01] | 1.01 [0.97-1.04] | 1.04 [1.01-1.07] | 1.06 [1.03-1.09] | 0.97 [0.83-1.12] | 0.95 [0.89-1.01] | 1.01 [0.98-1.05] | 11,471 | 11,349 | 410 | 3,085 | 8,786 |
| O80 | 1.08 [1.05-1.11] | 1.09 [1.06-1.12] | 1.05 [0.89-1.24] | 0.94 [0.89-1.00] | 0.97 [0.94-1.00] | 1.09 [1.06-1.12] | 1.09 [1.06-1.12] | 1.11 [0.94-1.32] | 0.96 [0.91-1.02] | 1.00 [0.97-1.03] | 12,482 | 12,266 | 312 | 2,989 | 10,079 |
| O81 | 1.07 [0.86-1.34] | 1.05 [0.84-1.32] | | | | 1.06 [0.84-1.33] | 1.04 [0.83-1.31] | | | | 177 | 173 | NA | NA | NA |
| O82 | 1.10 [1.03-1.19] | 1.08 [1.00-1.16] | 0.86 [0.67-1.11] | 1.07 [0.86-1.33] | 1.02 [0.94-1.10] | 1.08 [1.00-1.16] | 1.05 [0.98-1.13] | 0.91 [0.69-1.18] | 1.07 [0.85-1.34] | 1.01 [0.93-1.09] | 1,727 | 1,717 | 120 | 211 | 1,404 |
| O85 | 1.07 [0.97-1.19] | 1.08 [0.98-1.20] | | 0.93 [0.78-1.11] | 1.03 [0.93-1.15] | 1.03 [0.93-1.15] | 1.02 [0.92-1.13] | | 0.90 [0.74-1.08] | 1.01 [0.90-1.12] | 988 | 977 | NA | 328 | 816 |
| O86 | 1.15 [1.09-1.21] | 1.18 [1.12-1.24] | 1.20 [0.97-1.47] | 1.03 [0.93-1.13] | 1.11 [1.05-1.18] | 1.12 [1.06-1.18] | 1.13 [1.08-1.20] | 1.16 [0.93-1.43] | 1.02 [0.92-1.13] | 1.08 [1.02-1.15] | 3,748 | 3,716 | 201 | 1,055 | 2,931 |
| O87 | 1.19 [1.04-1.36] | 1.24 [1.09-1.42] | | | 1.17 [1.01-1.35] | 1.17 [1.02-1.34] | 1.21 [1.06-1.39] | | | 1.16 [0.99-1.35] | 542 | 540 | NA | NA | 440 |
| O88 | 1.15 [0.92-1.44] | 1.20 [0.96-1.50] | | | 1.14 [0.91-1.44] | 1.07 [0.85-1.35] | 1.10 [0.87-1.39] | | | 1.09 [0.86-1.38] | 201 | 200 | NA | NA | 184 |
| O89 | 1.32 [1.03-1.68] | 1.34 [1.06-1.71] | | | 1.17 [0.90-1.53] | 1.25 [0.97-1.62] | 1.30 [1.02-1.67] | | | 1.14 [0.87-1.49] | 174 | 174 | NA | NA | 133 |
| O90 | 1.18 [1.12-1.24] | 1.20 [1.14-1.25] | 1.10 [0.91-1.33] | 1.06 [0.97-1.17] | 1.11 [1.05-1.17] | 1.14 [1.08-1.19] | 1.13 [1.08-1.19] | 1.08 [0.89-1.31] | 1.04 [0.95-1.15] | 1.08 [1.02-1.14] | 4,483 | 4,439 | 242 | 1,218 | 3,558 |
| O91 | 1.02 [0.87-1.18] | 1.06 [0.91-1.23] | | 1.07 [0.78-1.46] | 0.98 [0.82-1.16] | 0.99 [0.84-1.16] | 1.02 [0.87-1.20] | | 1.10 [0.79-1.53] | 0.95 [0.80-1.14] | 413 | 407 | NA | 104 | 311 |
| O92 | 1.11 [0.98-1.25] | 1.12 [0.99-1.26] | | 0.96 [0.74-1.24] | 1.04 [0.91-1.19] | 1.07 [0.94-1.21] | 1.07 [0.95-1.22] | | 0.91 [0.69-1.20] | 1.03 [0.89-1.18] | 638 | 631 | NA | 146 | 493 |
| O94 | 1.26 [0.97-1.63] | 1.18 [0.92-1.53] | | | 1.15 [0.88-1.50] | 1.14 [0.86-1.50] | 1.04 [0.79-1.36] | | | 1.11 [0.84-1.46] | 149 | 149 | NA | NA | 138 |
| O98 | 1.15 [1.07-1.25] | 1.21 [1.12-1.31] | | 1.06 [0.92-1.22] | 1.10 [1.01-1.19] | 1.11 [1.02-1.20] | 1.14 [1.05-1.23] | | 1.04 [0.90-1.20] | 1.06 [0.98-1.16] | 1,674 | 1,653 | NA | 524 | 1,404 |
| O99 | 1.16 [1.14-1.19] | 1.19 [1.17-1.22] | 1.06 [0.97-1.15] | 1.05 [1.01-1.08] | 1.08 [1.06-1.11] | 1.11 [1.09-1.13] | 1.11 [1.08-1.13] | 1.02 [0.93-1.11] | 1.03 [0.99-1.07] | 1.06 [1.04-1.08] | 27,987 | 27,633 | 1,210 | 8,440 | 22,273 |
| Certain conditions originating in the perinatal period (P00-P96) | | | | | | | | | | | | | | | |
| P83 | 1.02 [0.87-1.20] | | | 1.03 [0.88-1.21] | 1.00 [0.84-1.20] | 1.02 [0.86-1.20] | | | 1.01 [0.86-1.19] | 1.01 [0.84-1.21] | 369 | NA | NA | 366 | 299 |
| Congenital malformations, deformations and chromosomal abnormalities (Q00-Q99) | | | | | | | | | | | | | | | |
| Q02 | 1.10 [0.96-1.27] | | | 1.08 [0.93-1.25] | 1.02 [0.88-1.18] | 1.05 [0.91-1.21] | | | 1.05 [0.90-1.22] | 1.00 [0.86-1.16] | 474 | NA | NA | 436 | 438 |
| Q03 | 1.20 [0.96-1.50] | 1.58 [1.17-2.12] | | | 1.22 [0.96-1.55] | 1.15 [0.91-1.45] | 1.47 [1.08-2.00] | | | 1.21 [0.95-1.55] | 187 | 112 | NA | NA | 164 |
| Q04 | 1.08 [0.94-1.25] | 1.21 [0.96-1.53] | | 1.01 [0.85-1.20] | 1.02 [0.88-1.18] | 1.01 [0.87-1.17] | 1.07 [0.84-1.36] | | 0.97 [0.81-1.16] | 0.98 [0.84-1.14] | 468 | 178 | NA | 324 | 430 |
| Q05 | 1.29 [1.12-1.49] | 1.33 [1.14-1.55] | 1.32 [1.10-1.60] | 1.08 [0.81-1.45] | 1.18 [1.01-1.37] | 1.18 [1.02-1.37] | 1.20 [1.02-1.40] | 1.16 [0.96-1.41] | 1.03 [0.76-1.39] | 1.10 [0.94-1.28] | 474 | 403 | 268 | 114 | 393 |
| Q06 | 0.91 [0.70-1.20] | | | | 0.85 [0.63-1.13] | 0.88 [0.66-1.16] | | | | 0.81 [0.60-1.08] | 126 | NA | NA | NA | 112 |
| Q07 | 1.20 [1.05-1.36] | 1.34 [1.14-1.58] | 1.31 [1.02-1.69] | 1.02 [0.85-1.23] | 1.11 [0.97-1.27] | 1.09 [0.95-1.24] | 1.12 [0.94-1.34] | 1.10 [0.84-1.43] | 0.96 [0.80-1.16] | 1.04 [0.90-1.19] | 582 | 360 | 142 | 309 | 524 |
| Q10 | 1.19 [1.05-1.35] | 1.46 [1.08-1.97] | | 1.14 [1.00-1.31] | 1.14 [1.00-1.31] | 1.18 [1.04-1.34] | 1.40 [1.02-1.90] | | 1.12 [0.98-1.29] | 1.12 [0.98-1.29] | 610 | 107 | NA | 520 | 530 |
| Q12 | 1.22 [1.02-1.46] | 1.26 [1.00-1.58] | 1.23 [0.93-1.62] | 1.25 [0.97-1.63] | 1.16 [0.95-1.40] | 1.17 [0.98-1.41] | 1.19 [0.94-1.51] | 1.18 [0.88-1.56] | 1.21 [0.93-1.57] | 1.13 [0.93-1.38] | 313 | 185 | 123 | 150 | 261 |
| Q13 | 1.33 [1.00-1.77] | | | | 1.22 [0.90-1.64] | 1.21 [0.90-1.62] | | | | 1.14 [0.84-1.55] | 120 | NA | NA | NA | 106 |
| Q14 | 1.11 [0.86-1.44] | | | | 1.08 [0.82-1.42] | 1.07 [0.82-1.39] | | | | 1.08 [0.82-1.43] | 142 | NA | NA | NA | 126 |
| Q16 | 1.28 [0.93-1.75] | | | | | 1.24 [0.90-1.70] | | | | | 104 | NA | NA | NA | NA |
| Q17 | 1.08 [0.99-1.17] | | | 1.07 [0.98-1.16] | 1.06 [0.96-1.17] | 1.06 [0.97-1.15] | | | 1.06 [0.97-1.15] | 1.06 [0.95-1.17] | 1,396 | NA | NA | 1,359 | 980 |
| Q18 | 1.16 [1.04-1.30] | 1.10 [0.93-1.31] | 1.18 [0.92-1.50] | 1.19 [1.04-1.36] | 1.14 [1.01-1.29] | 1.13 [1.01-1.27] | 1.01 [0.84-1.20] | 1.10 [0.86-1.42] | 1.17 [1.02-1.34] | 1.13 [1.00-1.29] | 788 | 305 | 147 | 538 | 631 |
| Q20 | 1.01 [0.77-1.32] | | | | 1.03 [0.77-1.37] | 0.98 [0.75-1.30] | | | | 1.00 [0.75-1.34] | 128 | NA | NA | NA | 116 |
| Q21 | 1.16 [1.09-1.24] | 1.24 [1.14-1.34] | 1.28 [1.16-1.40] | 1.01 [0.91-1.12] | 1.12 [1.04-1.20] | 1.10 [1.03-1.18] | 1.14 [1.06-1.24] | 1.16 [1.06-1.28] | 0.98 [0.88-1.09] | 1.07 [1.00-1.15] | 2,318 | 1,521 | 1,110 | 923 | 1,960 |
| Q22 | 0.96 [0.79-1.15] | | | 0.87 [0.69-1.09] | 0.94 [0.77-1.15] | 0.96 [0.79-1.17] | | | 0.87 [0.70-1.10] | 0.95 [0.78-1.17] | 259 | NA | NA | 181 | 231 |
| Q23 | 1.15 [1.04-1.26] | 1.17 [1.04-1.31] | 1.14 [1.00-1.30] | 1.07 [0.91-1.27] | 1.04 [0.94-1.16] | 1.11 [1.01-1.23] | 1.10 [0.98-1.24] | 1.10 [0.96-1.25] | 1.04 [0.87-1.23] | 1.02 [0.92-1.14] | 998 | 703 | 552 | 354 | 846 |
| Q24 | 1.17 [1.05-1.31] | 1.36 [1.19-1.55] | 1.40 [1.19-1.66] | 0.94 [0.79-1.11] | 1.08 [0.96-1.22] | 1.10 [0.99-1.23] | 1.23 [1.07-1.41] | 1.24 [1.04-1.47] | 0.92 [0.77-1.09] | 1.03 [0.92-1.16] | 815 | 538 | 344 | 347 | 722 |
| Q25 | 1.06 [0.95-1.18] | 1.19 [1.01-1.40] | 1.19 [0.97-1.46] | 0.96 [0.84-1.10] | 1.02 [0.91-1.14] | 1.03 [0.92-1.14] | 1.11 [0.94-1.31] | 1.08 [0.87-1.33] | 0.93 [0.81-1.07] | 0.98 [0.88-1.11] | 821 | 365 | 217 | 512 | 717 |
| Q26 | 1.11 [0.85-1.44] | | | | 0.97 [0.74-1.27] | 1.04 [0.80-1.37] | | | | 0.93 [0.71-1.23] | 141 | NA | NA | NA | 129 |
| Q27 | 1.08 [0.96-1.22] | 1.20 [1.03-1.40] | 1.19 [0.99-1.43] | 1.01 [0.84-1.20] | 1.03 [0.91-1.17] | 1.04 [0.92-1.18] | 1.13 [0.96-1.32] | 1.11 [0.91-1.34] | 1.00 [0.83-1.19] | 1.02 [0.90-1.16] | 668 | 394 | 273 | 324 | 608 |
| Q28 | 1.32 [1.12-1.54] | 1.46 [1.23-1.74] | 1.36 [1.11-1.66] | 0.98 [0.71-1.35] | 1.18 [0.99-1.40] | 1.27 [1.08-1.49] | 1.36 [1.14-1.63] | 1.25 [1.02-1.54] | 0.98 [0.71-1.36] | 1.17 [0.98-1.39] | 391 | 323 | 235 | 101 | 314 |
| Q31 | 1.03 [0.86-1.24] | | | 0.98 [0.80-1.21] | 0.92 [0.77-1.11] | 0.94 [0.78-1.14] | | | 0.87 [0.70-1.08] | 0.84 [0.69-1.02] | 274 | NA | NA | 210 | 254 |
| Q32 | 0.96 [0.73-1.27] | | | | 0.92 [0.68-1.23] | 0.85 [0.64-1.13] | | | | 0.80 [0.59-1.09] | 118 | NA | NA | NA | 109 |
| Q33 | 1.05 [0.79-1.41] | | | | 1.01 [0.75-1.36] | 0.93 [0.69-1.26] | | | | 0.90 [0.66-1.23] | 112 | NA | NA | NA | 105 |
| Q35 | 1.11 [0.94-1.32] | | | 1.10 [0.92-1.32] | 1.04 [0.87-1.24] | 1.06 [0.89-1.26] | | | 1.04 [0.87-1.26] | 1.01 [0.84-1.22] | 344 | NA | NA | 302 | 304 |
| Q38 | 1.14 [1.07-1.21] | 1.16 [1.02-1.31] | 1.20 [1.05-1.37] | 1.11 [1.03-1.20] | 1.10 [1.02-1.18] | 1.11 [1.04-1.18] | 1.08 [0.95-1.22] | 1.10 [0.97-1.26] | 1.09 [1.01-1.18] | 1.07 [1.00-1.15] | 2,375 | 673 | 605 | 1,736 | 1,978 |
| Q39 | 1.27 [1.08-1.49] | 1.30 [1.09-1.53] | 1.29 [1.08-1.53] | | 1.11 [0.93-1.31] | 1.17 [0.99-1.39] | 1.16 [0.98-1.38] | 1.16 [0.97-1.39] | | 1.03 [0.87-1.23] | 376 | 352 | 326 | NA | 334 |
| Q40 | 1.24 [1.01-1.52] | 1.20 [0.97-1.48] | 1.17 [0.94-1.47] | | 1.20 [0.97-1.48] | 1.11 [0.90-1.38] | 1.03 [0.82-1.28] | 1.03 [0.82-1.30] | | 1.12 [0.90-1.39] | 227 | 207 | 185 | NA | 211 |
| Q43 | 1.17 [1.06-1.29] | 1.23 [1.10-1.38] | 1.22 [1.07-1.39] | 0.98 [0.81-1.17] | 1.13 [1.01-1.26] | 1.10 [0.99-1.22] | 1.10 [0.98-1.24] | 1.10 [0.96-1.26] | 0.95 [0.79-1.14] | 1.07 [0.96-1.19] | 950 | 723 | 519 | 301 | 815 |
| Q44 | 1.15 [0.95-1.38] | 1.21 [1.00-1.47] | 1.11 [0.90-1.38] | | 1.08 [0.89-1.31] | 1.09 [0.90-1.31] | 1.10 [0.91-1.34] | 1.01 [0.81-1.26] | | 1.05 [0.86-1.27] | 289 | 254 | 200 | NA | 253 |
| Q45 | 1.34 [1.02-1.76] | 1.36 [1.01-1.83] | | | 1.37 [1.03-1.81] | 1.24 [0.93-1.64] | 1.13 [0.82-1.54] | | | 1.29 [0.97-1.72] | 135 | 111 | NA | NA | 126 |
| Q50 | 1.25 [1.11-1.41] | 1.30 [1.14-1.47] | 1.12 [0.91-1.37] | 1.41 [1.12-1.77] | 1.13 [0.99-1.29] | 1.19 [1.06-1.35] | 1.19 [1.04-1.35] | 1.04 [0.84-1.28] | 1.28 [1.00-1.63] | 1.11 [0.97-1.27] | 682 | 605 | 206 | 214 | 550 |
| Q51 | 1.06 [0.94-1.21] | 1.10 [0.97-1.25] | 1.20 [1.00-1.44] | 1.09 [0.83-1.44] | 1.06 [0.93-1.22] | 1.01 [0.89-1.15] | 1.02 [0.89-1.16] | 1.08 [0.89-1.31] | 1.08 [0.81-1.43] | 1.03 [0.90-1.18] | 599 | 567 | 260 | 135 | 499 |
| Q52 | 1.21 [1.03-1.42] | 1.22 [0.97-1.52] | 1.45 [1.08-1.96] | 1.13 [0.91-1.40] | 1.23 [1.03-1.48] | 1.12 [0.94-1.32] | 1.14 [0.90-1.44] | 1.32 [0.97-1.80] | 1.07 [0.86-1.33] | 1.20 [0.99-1.44] | 380 | 187 | 108 | 225 | 302 |
| Q53 | 1.00 [0.94-1.06] | 1.12 [0.85-1.46] | | 0.99 [0.93-1.05] | 0.96 [0.90-1.02] | 0.98 [0.92-1.04] | 1.05 [0.79-1.38] | | 0.97 [0.91-1.03] | 0.96 [0.90-1.02] | 3,035 | 126 | NA | 2,935 | 2,479 |
| Q54 | 1.11 [1.00-1.23] | 1.24 [1.03-1.49] | 1.17 [0.96-1.41] | 1.07 [0.95-1.22] | 1.04 [0.93-1.17] | 1.11 [0.99-1.23] | 1.17 [0.97-1.41] | 1.12 [0.92-1.36] | 1.07 [0.94-1.22] | 1.06 [0.94-1.19] | 866 | 292 | 262 | 588 | 694 |
| Q55 | 1.13 [1.06-1.22] | 1.22 [0.96-1.55] | | 1.11 [1.03-1.19] | 1.13 [1.04-1.22] | 1.11 [1.03-1.19] | 1.10 [0.86-1.42] | | 1.08 [1.01-1.16] | 1.10 [1.02-1.19] | 2,125 | 159 | NA | 2,015 | 1,779 |
| Q60 | 1.17 [1.03-1.33] | 1.20 [1.05-1.38] | 1.23 [1.05-1.44] | 0.92 [0.72-1.18] | 1.07 [0.94-1.23] | 1.12 [0.99-1.28] | 1.08 [0.94-1.25] | 1.13 [0.96-1.32] | 0.92 [0.72-1.17] | 1.06 [0.92-1.21] | 628 | 502 | 394 | 166 | 552 |
| Q61 | 1.16 [1.07-1.26] | 1.14 [1.05-1.24] | 1.19 [1.08-1.30] | 1.05 [0.86-1.29] | 1.07 [0.99-1.17] | 1.11 [1.03-1.21] | 1.04 [0.95-1.14] | 1.10 [1.01-1.21] | 1.00 [0.82-1.23] | 1.05 [0.96-1.15] | 1,502 | 1,317 | 1,141 | 239 | 1,307 |
| Q62 | 1.00 [0.85-1.18] | 1.01 [0.77-1.31] | | 0.99 [0.81-1.21] | 0.96 [0.81-1.15] | 0.98 [0.83-1.16] | 0.98 [0.75-1.29] | | 0.97 [0.79-1.19] | 0.96 [0.80-1.15] | 338 | 130 | NA | 228 | 300 |
| Q63 | 1.16 [1.06-1.27] | 1.17 [1.05-1.30] | 1.09 [0.96-1.24] | 1.16 [1.00-1.34] | 1.05 [0.96-1.16] | 1.09 [0.99-1.19] | 1.05 [0.94-1.17] | 0.97 [0.85-1.10] | 1.12 [0.96-1.30] | 1.00 [0.91-1.10] | 1,233 | 857 | 567 | 497 | 1,083 |
| Q64 | 1.15 [0.95-1.38] | 1.31 [0.99-1.74] | | 1.06 [0.84-1.33] | 1.03 [0.85-1.26] | 1.09 [0.90-1.32] | 1.21 [0.91-1.62] | | 1.02 [0.80-1.29] | 1.01 [0.82-1.24] | 290 | 116 | NA | 192 | 253 |
| Q65 | 1.04 [0.94-1.15] | 1.17 [1.02-1.34] | 1.18 [0.97-1.43] | 0.89 [0.78-1.02] | 0.93 [0.84-1.04] | 0.99 [0.89-1.09] | 1.07 [0.93-1.23] | 1.08 [0.88-1.32] | 0.86 [0.75-0.99] | 0.91 [0.82-1.02] | 947 | 502 | 238 | 525 | 815 |
| Q66 | 1.26 [1.15-1.37] | 1.38 [1.20-1.57] | 1.60 [1.36-1.89] | 1.12 [1.01-1.25] | 1.12 [1.02-1.23] | 1.19 [1.09-1.30] | 1.25 [1.09-1.44] | 1.44 [1.21-1.71] | 1.08 [0.97-1.21] | 1.09 [0.99-1.20] | 1,328 | 528 | 363 | 871 | 1,111 |
| Q67 | 1.20 [1.07-1.33] | 1.41 [1.19-1.68] | 1.56 [1.23-1.99] | 1.08 [0.95-1.23] | 1.10 [0.98-1.24] | 1.14 [1.02-1.28] | 1.30 [1.09-1.56] | 1.47 [1.15-1.87] | 1.01 [0.89-1.16] | 1.07 [0.95-1.21] | 831 | 319 | 177 | 598 | 720 |
| Q68 | 1.21 [1.00-1.46] | | | 1.11 [0.90-1.39] | 1.03 [0.84-1.27] | 1.16 [0.96-1.40] | | | 1.06 [0.85-1.33] | 1.00 [0.81-1.24] | 288 | NA | NA | 211 | 241 |
| Q69 | 0.90 [0.69-1.18] | | | 0.91 [0.69-1.20] | | 0.88 [0.67-1.15] | | | 0.89 [0.67-1.18] | | 125 | NA | NA | 118 | NA |
| Q70 | 0.87 [0.65-1.16] | | | | | 0.87 [0.65-1.16] | | | | | 113 | NA | NA | NA | NA |
| Q72 | 1.18 [0.88-1.57] | | | | 1.06 [0.78-1.43] | 1.15 [0.86-1.54] | | | | 1.04 [0.76-1.42] | 128 | NA | NA | NA | 113 |
| Q74 | 1.21 [1.09-1.35] | 1.28 [1.08-1.53] | 1.35 [1.06-1.72] | 1.17 [1.03-1.33] | 1.11 [0.98-1.24] | 1.17 [1.05-1.31] | 1.16 [0.97-1.40] | 1.18 [0.91-1.52] | 1.15 [1.01-1.30] | 1.08 [0.96-1.21] | 889 | 316 | 160 | 646 | 741 |
| Q75 | 1.03 [0.89-1.19] | | | 1.00 [0.86-1.17] | 0.98 [0.84-1.14] | 0.99 [0.85-1.15] | | | 0.97 [0.83-1.13] | 0.95 [0.81-1.11] | 433 | NA | NA | 401 | 396 |
| Q76 | 1.15 [1.02-1.31] | 1.23 [1.06-1.42] | 1.35 [1.13-1.61] | 0.91 [0.75-1.12] | 1.07 [0.94-1.21] | 1.06 [0.94-1.21] | 1.05 [0.90-1.22] | 1.11 [0.92-1.34] | 0.87 [0.71-1.07] | 1.02 [0.89-1.16] | 639 | 444 | 293 | 242 | 556 |
| Q78 | 1.22 [1.05-1.42] | 1.31 [1.07-1.61] | 1.30 [1.03-1.63] | 1.06 [0.85-1.32] | 1.12 [0.95-1.31] | 1.15 [0.99-1.35] | 1.17 [0.95-1.45] | 1.12 [0.88-1.42] | 1.03 [0.82-1.29] | 1.07 [0.91-1.26] | 429 | 237 | 179 | 205 | 385 |
| Q79 | 1.35 [1.22-1.51] | 1.34 [1.18-1.52] | 1.19 [0.98-1.45] | 1.24 [1.06-1.45] | 1.19 [1.07-1.33] | 1.13 [1.01-1.26] | 1.01 [0.88-1.16] | 0.91 [0.74-1.12] | 1.12 [0.95-1.32] | 1.04 [0.93-1.17] | 920 | 629 | 242 | 451 | 807 |
| Q80 | 2.79 [1.97-3.95] | | | | 2.74 [1.90-3.95] | 2.58 [1.81-3.68] | | | | 2.50 [1.71-3.64] | 119 | NA | NA | NA | 105 |
| Q82 | 1.36 [1.21-1.52] | 1.75 [1.47-2.07] | 1.81 [1.49-2.21] | 1.19 [1.03-1.38] | 1.32 [1.17-1.49] | 1.28 [1.14-1.44] | 1.56 [1.31-1.86] | 1.58 [1.29-1.94] | 1.16 [1.00-1.34] | 1.26 [1.11-1.43] | 802 | 360 | 273 | 475 | 672 |
| Q83 | 1.13 [0.96-1.33] | 1.15 [0.96-1.38] | | 0.97 [0.76-1.25] | 1.04 [0.86-1.26] | 1.06 [0.90-1.26] | 1.05 [0.87-1.26] | | 0.93 [0.73-1.20] | 0.99 [0.82-1.21] | 352 | 278 | NA | 154 | 250 |
| Q85 | 1.02 [0.89-1.16] | 1.05 [0.89-1.24] | 1.08 [0.88-1.32] | 0.96 [0.79-1.17] | 0.94 [0.81-1.08] | 0.97 [0.85-1.12] | 0.95 [0.80-1.14] | 1.00 [0.81-1.24] | 0.94 [0.76-1.15] | 0.92 [0.79-1.06] | 515 | 317 | 215 | 240 | 444 |
| Q87 | 1.23 [1.10-1.37] | 1.44 [1.24-1.69] | 1.59 [1.29-1.97] | 1.05 [0.91-1.21] | 1.14 [1.01-1.28] | 1.15 [1.03-1.29] | 1.30 [1.10-1.52] | 1.42 [1.14-1.77] | 1.02 [0.88-1.17] | 1.10 [0.98-1.24] | 847 | 406 | 215 | 512 | 751 |
| Q89 | 1.09 [0.97-1.22] | 1.20 [1.01-1.43] | 1.15 [0.93-1.43] | 1.02 [0.88-1.18] | 1.06 [0.93-1.20] | 1.06 [0.94-1.19] | 1.09 [0.91-1.30] | 1.08 [0.87-1.34] | 1.01 [0.87-1.17] | 1.04 [0.91-1.18] | 716 | 315 | 199 | 451 | 615 |
| Q90 | 2.77 [2.46-3.12] | 3.76 [3.28-4.32] | 4.16 [3.54-4.89] | 1.03 [0.81-1.31] | 2.00 [1.68-2.37] | 2.68 [2.37-3.02] | 3.86 [3.35-4.45] | 4.05 [3.44-4.78] | 1.00 [0.79-1.29] | 1.90 [1.59-2.27] | 855 | 725 | 526 | 161 | 367 |
| Q92 | 1.12 [0.87-1.45] | | | 0.98 [0.73-1.31] | 1.18 [0.91-1.54] | 1.07 [0.83-1.40] | | | 0.95 [0.71-1.29] | 1.15 [0.88-1.51] | 151 | NA | NA | 113 | 143 |
| Q93 | 1.27 [1.09-1.47] | | | 1.19 [1.01-1.41] | 1.25 [1.07-1.46] | 1.17 [1.00-1.36] | | | 1.12 [0.95-1.32] | 1.17 [1.00-1.38] | 466 | NA | NA | 398 | 436 |
| Q96 | 1.62 [1.23-2.13] | | | | | 1.65 [1.24-2.19] | | | | | 140 | NA | NA | NA | NA |
| Q98 | 1.77 [1.35-2.32] | | | | 1.63 [1.22-2.17] | 1.60 [1.21-2.11] | | | | 1.50 [1.12-2.02] | 149 | NA | NA | NA | 129 |
| Q99 | 1.16 [1.00-1.33] | 1.28 [1.00-1.65] | | 1.12 [0.95-1.31] | 1.10 [0.95-1.27] | 1.08 [0.94-1.25] | 1.14 [0.88-1.47] | | 1.07 [0.91-1.26] | 1.05 [0.90-1.21] | 517 | 153 | NA | 395 | 472 |
| Symptoms, signs and abnormal clinical and laboratory findings, not elsewhere classified (R00-R99) | | | | | | | | | | | | | | | |
| R00 | 1.21 [1.19-1.23] | 1.22 [1.20-1.24] | 1.22 [1.20-1.24] | 1.14 [1.10-1.18] | 1.14 [1.12-1.16] | 1.14 [1.13-1.16] | 1.11 [1.09-1.13] | 1.11 [1.09-1.13] | 1.10 [1.06-1.15] | 1.10 [1.08-1.12] | 44,637 | 40,927 | 32,645 | 7,228 | 39,435 |
| R01 | 1.21 [1.17-1.26] | 1.22 [1.17-1.27] | 1.23 [1.18-1.29] | 1.21 [1.13-1.29] | 1.17 [1.13-1.22] | 1.15 [1.11-1.19] | 1.12 [1.07-1.17] | 1.14 [1.08-1.19] | 1.16 [1.09-1.24] | 1.13 [1.09-1.18] | 7,450 | 5,428 | 4,624 | 2,324 | 6,736 |
| R02 | 1.22 [1.17-1.27] | 1.23 [1.18-1.28] | 1.22 [1.17-1.28] | 1.08 [0.92-1.28] | 1.16 [1.11-1.21] | 1.19 [1.14-1.24] | 1.13 [1.08-1.18] | 1.12 [1.07-1.17] | 1.07 [0.91-1.27] | 1.16 [1.11-1.21] | 6,432 | 6,203 | 5,777 | 376 | 5,857 |
| R03 | 1.18 [1.15-1.22] | 1.19 [1.16-1.22] | 1.20 [1.17-1.24] | 1.13 [1.04-1.22] | 1.12 [1.09-1.15] | 1.14 [1.11-1.17] | 1.12 [1.09-1.15] | 1.13 [1.10-1.16] | 1.10 [1.02-1.20] | 1.10 [1.07-1.13] | 15,117 | 14,446 | 11,408 | 1,719 | 13,251 |
| R04 | 1.33 [1.30-1.36] | 1.35 [1.31-1.38] | 1.34 [1.31-1.38] | 1.24 [1.17-1.31] | 1.23 [1.20-1.26] | 1.25 [1.22-1.28] | 1.22 [1.18-1.25] | 1.21 [1.18-1.24] | 1.19 [1.12-1.26] | 1.19 [1.16-1.22] | 18,561 | 15,989 | 14,425 | 3,187 | 16,162 |
| R05 | 1.38 [1.35-1.41] | 1.35 [1.32-1.39] | 1.36 [1.32-1.39] | 1.37 [1.32-1.43] | 1.29 [1.26-1.32] | 1.27 [1.24-1.30] | 1.19 [1.15-1.22] | 1.19 [1.15-1.22] | 1.30 [1.25-1.35] | 1.21 [1.18-1.24] | 20,701 | 14,452 | 12,383 | 6,900 | 18,750 |
| R06 | 1.52 [1.50-1.54] | 1.34 [1.32-1.36] | 1.33 [1.31-1.35] | 1.76 [1.72-1.79] | 1.43 [1.41-1.44] | 1.43 [1.41-1.44] | 1.18 [1.16-1.20] | 1.18 [1.16-1.20] | 1.70 [1.67-1.73] | 1.36 [1.35-1.38] | 74,870 | 41,003 | 34,545 | 35,908 | 67,136 |
| R07 | 1.29 [1.28-1.31] | 1.30 [1.29-1.32] | 1.31 [1.29-1.33] | 1.14 [1.10-1.18] | 1.19 [1.17-1.20] | 1.20 [1.18-1.21] | 1.16 [1.14-1.17] | 1.16 [1.14-1.17] | 1.10 [1.06-1.15] | 1.13 [1.11-1.14] | 77,085 | 74,192 | 60,170 | 7,686 | 65,437 |
| R09 | 1.31 [1.26-1.36] | 1.32 [1.27-1.37] | 1.31 [1.25-1.36] | 1.21 [1.08-1.34] | 1.22 [1.17-1.27] | 1.22 [1.17-1.26] | 1.17 [1.12-1.22] | 1.17 [1.11-1.22] | 1.15 [1.03-1.29] | 1.16 [1.12-1.21] | 6,918 | 6,381 | 5,412 | 887 | 6,244 |
| R10 | 1.27 [1.26-1.28] | 1.28 [1.27-1.29] | 1.30 [1.29-1.32] | 1.19 [1.17-1.21] | 1.18 [1.17-1.19] | 1.18 [1.17-1.19] | 1.14 [1.13-1.15] | 1.16 [1.14-1.18] | 1.14 [1.12-1.16] | 1.12 [1.11-1.13] | 116,393 | 93,858 | 55,106 | 37,214 | 94,513 |
| R11 | 1.23 [1.22-1.25] | 1.25 [1.23-1.27] | 1.25 [1.23-1.27] | 1.17 [1.14-1.19] | 1.15 [1.13-1.17] | 1.16 [1.14-1.17] | 1.12 [1.10-1.14] | 1.13 [1.11-1.15] | 1.13 [1.10-1.15] | 1.11 [1.09-1.12] | 66,001 | 49,305 | 37,038 | 21,941 | 57,542 |
| R12 | 1.35 [1.29-1.42] | 1.35 [1.28-1.42] | 1.34 [1.27-1.42] | 1.08 [0.90-1.28] | 1.23 [1.17-1.30] | 1.22 [1.16-1.28] | 1.18 [1.12-1.24] | 1.17 [1.11-1.24] | 1.00 [0.83-1.20] | 1.14 [1.08-1.20] | 4,130 | 4,046 | 3,156 | 336 | 3,611 |
| R13 | 1.31 [1.29-1.34] | 1.32 [1.29-1.34] | 1.31 [1.29-1.34] | 1.21 [1.11-1.32] | 1.23 [1.21-1.25] | 1.22 [1.19-1.24] | 1.18 [1.16-1.21] | 1.18 [1.16-1.21] | 1.12 [1.02-1.23] | 1.17 [1.15-1.19] | 30,132 | 29,322 | 27,273 | 1,407 | 27,584 |
| R14 | 1.31 [1.27-1.36] | 1.32 [1.27-1.37] | 1.30 [1.25-1.36] | 1.16 [1.04-1.30] | 1.22 [1.17-1.26] | 1.20 [1.16-1.25] | 1.15 [1.11-1.20] | 1.15 [1.10-1.20] | 1.11 [0.99-1.24] | 1.14 [1.10-1.19] | 7,064 | 6,608 | 5,225 | 903 | 6,356 |
| R15 | 1.20 [1.16-1.24] | 1.20 [1.17-1.24] | 1.19 [1.15-1.23] | 1.14 [0.99-1.31] | 1.14 [1.10-1.17] | 1.16 [1.13-1.20] | 1.13 [1.09-1.16] | 1.12 [1.09-1.16] | 1.09 [0.95-1.25] | 1.13 [1.09-1.16] | 12,111 | 11,699 | 11,132 | 560 | 11,344 |
| R16 | 1.38 [1.32-1.45] | 1.44 [1.38-1.51] | 1.41 [1.34-1.49] | 1.11 [0.99-1.24] | 1.27 [1.21-1.33] | 1.30 [1.24-1.36] | 1.28 [1.22-1.35] | 1.27 [1.20-1.34] | 1.08 [0.96-1.21] | 1.23 [1.17-1.29] | 5,156 | 4,624 | 3,839 | 829 | 4,592 |
| R17 | 1.22 [1.16-1.28] | 1.24 [1.18-1.31] | 1.23 [1.17-1.30] | 1.12 [0.94-1.33] | 1.19 [1.12-1.25] | 1.19 [1.13-1.25] | 1.18 [1.12-1.24] | 1.17 [1.11-1.24] | 1.13 [0.95-1.34] | 1.18 [1.12-1.25] | 3,810 | 3,566 | 3,200 | 362 | 3,246 |
| R18 | 1.20 [1.17-1.24] | 1.22 [1.18-1.26] | 1.22 [1.18-1.26] | 0.94 [0.81-1.10] | 1.17 [1.13-1.21] | 1.17 [1.14-1.21] | 1.15 [1.11-1.19] | 1.15 [1.11-1.19] | 0.93 [0.80-1.09] | 1.17 [1.13-1.21] | 9,491 | 9,236 | 8,539 | 441 | 8,376 |
| R19 | 1.34 [1.31-1.36] | 1.35 [1.33-1.37] | 1.33 [1.31-1.36] | 1.19 [1.13-1.26] | 1.25 [1.23-1.27] | 1.23 [1.21-1.25] | 1.21 [1.19-1.23] | 1.19 [1.17-1.21] | 1.14 [1.08-1.20] | 1.18 [1.16-1.20] | 39,491 | 37,905 | 31,773 | 3,664 | 34,526 |
| R20 | 1.27 [1.23-1.30] | 1.27 [1.23-1.31] | 1.27 [1.23-1.32] | 1.15 [1.06-1.24] | 1.16 [1.13-1.20] | 1.16 [1.12-1.20] | 1.11 [1.07-1.14] | 1.11 [1.07-1.15] | 1.09 [1.00-1.18] | 1.10 [1.06-1.13] | 10,595 | 9,813 | 7,028 | 1,678 | 9,380 |
| R21 | 1.46 [1.42-1.49] | 1.70 [1.65-1.75] | 1.78 [1.72-1.84] | 1.17 [1.14-1.21] | 1.37 [1.34-1.40] | 1.39 [1.35-1.42] | 1.55 [1.50-1.60] | 1.63 [1.58-1.69] | 1.13 [1.10-1.17] | 1.32 [1.29-1.35] | 19,887 | 11,833 | 9,506 | 8,865 | 17,409 |
| R22 | 1.38 [1.34-1.43] | 1.37 [1.31-1.42] | 1.36 [1.31-1.42] | 1.42 [1.33-1.52] | 1.31 [1.27-1.36] | 1.29 [1.25-1.34] | 1.22 [1.17-1.27] | 1.23 [1.18-1.29] | 1.36 [1.27-1.45] | 1.25 [1.20-1.29] | 8,990 | 6,898 | 5,433 | 2,598 | 7,743 |
| R23 | 1.25 [1.20-1.29] | 1.33 [1.26-1.40] | 1.33 [1.26-1.41] | 1.16 [1.10-1.22] | 1.18 [1.14-1.23] | 1.19 [1.14-1.24] | 1.21 [1.15-1.28] | 1.21 [1.14-1.28] | 1.12 [1.06-1.18] | 1.14 [1.10-1.19] | 7,127 | 3,782 | 3,302 | 3,510 | 6,405 |
| R25 | 1.24 [1.19-1.29] | 1.28 [1.23-1.34] | 1.28 [1.22-1.35] | 1.10 [1.01-1.20] | 1.16 [1.12-1.21] | 1.15 [1.10-1.20] | 1.14 [1.09-1.19] | 1.14 [1.09-1.20] | 1.06 [0.97-1.16] | 1.11 [1.07-1.16] | 6,270 | 5,246 | 4,471 | 1,304 | 5,683 |
| R26 | 1.21 [1.19-1.23] | 1.20 [1.19-1.22] | 1.20 [1.18-1.22] | 1.10 [1.03-1.17] | 1.15 [1.13-1.17] | 1.16 [1.15-1.18] | 1.12 [1.10-1.14] | 1.12 [1.10-1.14] | 1.07 [1.00-1.15] | 1.13 [1.11-1.15] | 45,075 | 43,012 | 42,038 | 2,374 | 42,416 |
| R27 | 1.18 [1.10-1.27] | 1.18 [1.08-1.29] | 1.20 [1.09-1.31] | 1.17 [1.03-1.33] | 1.12 [1.04-1.21] | 1.12 [1.04-1.21] | 1.07 [0.98-1.16] | 1.09 [0.99-1.20] | 1.12 [0.98-1.28] | 1.09 [1.01-1.18] | 1,904 | 1,344 | 1,171 | 629 | 1,698 |
| R29 | 1.19 [1.18-1.21] | 1.19 [1.18-1.21] | 1.19 [1.17-1.21] | 1.07 [0.99-1.17] | 1.14 [1.13-1.16] | 1.15 [1.13-1.16] | 1.10 [1.09-1.12] | 1.11 [1.09-1.12] | 1.02 [0.94-1.11] | 1.12 [1.11-1.14] | 58,332 | 57,338 | 55,769 | 1,516 | 55,239 |
| R30 | 1.35 [1.29-1.41] | 1.34 [1.28-1.41] | 1.36 [1.29-1.44] | 1.18 [1.08-1.29] | 1.21 [1.16-1.27] | 1.24 [1.19-1.30] | 1.17 [1.11-1.23] | 1.18 [1.11-1.25] | 1.14 [1.05-1.25] | 1.14 [1.09-1.20] | 5,315 | 4,379 | 3,136 | 1,412 | 4,639 |
| R31 | 1.19 [1.17-1.22] | 1.19 [1.17-1.21] | 1.18 [1.16-1.21] | 1.14 [1.06-1.22] | 1.11 [1.09-1.13] | 1.15 [1.13-1.17] | 1.12 [1.10-1.14] | 1.11 [1.09-1.13] | 1.09 [1.02-1.17] | 1.09 [1.07-1.11] | 31,241 | 29,856 | 26,969 | 2,259 | 26,387 |
| R32 | 1.19 [1.17-1.22] | 1.18 [1.16-1.21] | 1.17 [1.15-1.20] | 1.15 [1.07-1.25] | 1.13 [1.10-1.15] | 1.15 [1.13-1.18] | 1.10 [1.08-1.13] | 1.10 [1.08-1.13] | 1.10 [1.02-1.20] | 1.11 [1.09-1.14] | 23,400 | 22,061 | 20,863 | 1,701 | 21,813 |
| R33 | 1.17 [1.15-1.19] | 1.16 [1.14-1.18] | 1.16 [1.14-1.18] | 1.16 [1.08-1.25] | 1.12 [1.10-1.14] | 1.13 [1.11-1.15] | 1.10 [1.08-1.11] | 1.10 [1.08-1.12] | 1.10 [1.02-1.19] | 1.10 [1.09-1.12] | 43,140 | 42,266 | 39,647 | 1,817 | 39,109 |
| R34 | 1.21 [1.11-1.33] | 1.30 [1.18-1.43] | 1.26 [1.14-1.39] | 0.95 [0.73-1.23] | 1.23 [1.11-1.35] | 1.15 [1.05-1.27] | 1.17 [1.06-1.30] | 1.15 [1.03-1.27] | 0.92 [0.71-1.21] | 1.20 [1.09-1.32] | 1,225 | 1,113 | 1,027 | 145 | 1,131 |
| R35 | 1.35 [1.30-1.40] | 1.33 [1.29-1.38] | 1.32 [1.27-1.38] | 1.22 [1.10-1.35] | 1.23 [1.19-1.28] | 1.26 [1.21-1.30] | 1.20 [1.15-1.25] | 1.19 [1.14-1.24] | 1.19 [1.07-1.32] | 1.17 [1.12-1.21] | 7,985 | 7,287 | 6,145 | 1,046 | 6,959 |
| R36 | 1.28 [1.00-1.63] | 1.35 [1.04-1.75] | 1.39 [1.05-1.85] | | 1.19 [0.92-1.54] | 1.25 [0.97-1.60] | 1.23 [0.94-1.61] | 1.24 [0.92-1.66] | | 1.15 [0.88-1.48] | 176 | 156 | 132 | NA | 155 |
| R39 | 1.35 [1.30-1.39] | 1.35 [1.31-1.40] | 1.33 [1.28-1.38] | 1.31 [1.19-1.45] | 1.22 [1.18-1.26] | 1.25 [1.21-1.29] | 1.21 [1.16-1.25] | 1.19 [1.15-1.23] | 1.25 [1.13-1.39] | 1.16 [1.12-1.20] | 9,110 | 8,523 | 7,284 | 1,019 | 7,999 |
| R40 | 1.21 [1.17-1.26] | 1.23 [1.18-1.28] | 1.24 [1.18-1.29] | 1.05 [0.95-1.16] | 1.15 [1.11-1.20] | 1.15 [1.11-1.20] | 1.12 [1.07-1.16] | 1.12 [1.08-1.17] | 1.02 [0.92-1.13] | 1.13 [1.08-1.17] | 7,295 | 6,480 | 5,786 | 1,095 | 6,730 |
| R41 | 1.17 [1.16-1.19] | 1.18 [1.16-1.20] | 1.17 [1.15-1.19] | 1.09 [1.01-1.18] | 1.14 [1.12-1.16] | 1.14 [1.12-1.16] | 1.11 [1.09-1.13] | 1.11 [1.09-1.13] | 1.04 [0.96-1.13] | 1.13 [1.11-1.15] | 41,977 | 40,673 | 39,528 | 1,764 | 38,419 |
| R42 | 1.24 [1.21-1.26] | 1.25 [1.22-1.27] | 1.24 [1.22-1.27] | 1.11 [1.05-1.18] | 1.16 [1.13-1.18] | 1.15 [1.13-1.18] | 1.12 [1.09-1.14] | 1.12 [1.09-1.15] | 1.07 [1.01-1.14] | 1.10 [1.08-1.13] | 24,441 | 22,855 | 19,322 | 2,819 | 22,003 |

#### Supplementary Table 3: Hazard ratios and events from all cohorts

| | Hazard ratio (99% confidence interval) | | | | | | | | | | Events (in exposed) | | | | |
|---|---|---|---|---|---|---|---|---|---|---|---|---|---|---|---|
| | crude | | | | | adjusted | | | | | | | | | |
| Outcome | any age | 18+ | 40+ | <18 | hosp. | any age | 18+ | 40+ | <18 | hosp. | any age | 18+ | 40+ | <18 | hosp. |
| R43 | 1.35 [1.14-1.61] | 1.28 [1.07-1.52] | 1.36 [1.11-1.66] | | 1.24 [1.04-1.49] | 1.23 [1.03-1.47] | 1.12 [0.93-1.34] | 1.14 [0.92-1.41] | | 1.17 [0.98-1.41] | 348 | 318 | 243 | NA | 305 |
| R44 | 1.21 [1.16-1.26] | 1.22 [1.17-1.28] | 1.22 [1.17-1.28] | 1.14 [1.03-1.26] | 1.15 [1.10-1.19] | 1.15 [1.11-1.20] | 1.11 [1.07-1.17] | 1.12 [1.07-1.17] | 1.10 [1.00-1.22] | 1.12 [1.08-1.17] | 6,419 | 5,617 | 4,922 | 1,109 | 5,851 |
| R45 | 1.19 [1.16-1.22] | 1.19 [1.15-1.22] | 1.24 [1.20-1.28] | 1.06 [1.02-1.12] | 1.12 [1.09-1.15] | 1.13 [1.10-1.16] | 1.06 [1.03-1.09] | 1.11 [1.07-1.15] | 1.04 [0.99-1.10] | 1.10 [1.07-1.13] | 15,065 | 11,627 | 8,843 | 4,850 | 13,574 |
| R46 | 1.13 [1.08-1.19] | 1.13 [1.07-1.19] | 1.12 [1.06-1.19] | 1.06 [0.94-1.19] | 1.10 [1.04-1.16] | 1.11 [1.06-1.17] | 1.07 [1.01-1.13] | 1.07 [1.01-1.13] | 1.00 [0.89-1.14] | 1.11 [1.05-1.17] | 3,990 | 3,358 | 3,052 | 731 | 3,662 |
| R47 | 1.17 [1.13-1.20] | 1.17 [1.13-1.20] | 1.16 [1.13-1.20] | 1.09 [0.97-1.22] | 1.11 [1.08-1.15] | 1.13 [1.09-1.16] | 1.08 [1.05-1.12] | 1.08 [1.04-1.11] | 1.04 [0.92-1.18] | 1.10 [1.06-1.13] | 11,628 | 11,115 | 10,439 | 719 | 10,584 |
| R48 | 1.29 [1.12-1.48] | 1.32 [1.11-1.56] | 1.31 [1.03-1.66] | 1.16 [0.96-1.40] | 1.23 [1.06-1.42] | 1.15 [1.00-1.33] | 1.10 [0.92-1.32] | 1.13 [0.88-1.45] | 1.09 [0.90-1.32] | 1.16 [1.00-1.35] | 570 | 352 | 175 | 309 | 512 |
| R49 | 1.35 [1.27-1.44] | 1.35 [1.27-1.44] | 1.32 [1.24-1.42] | 1.25 [1.05-1.48] | 1.23 [1.15-1.31] | 1.23 [1.16-1.32] | 1.17 [1.10-1.26] | 1.14 [1.07-1.23] | 1.20 [1.01-1.42] | 1.15 [1.08-1.23] | 2,591 | 2,269 | 2,050 | 364 | 2,279 |
| R50 | 1.22 [1.20-1.25] | 1.27 [1.24-1.30] | 1.27 [1.24-1.31] | 1.15 [1.12-1.18] | 1.16 [1.14-1.18] | 1.17 [1.15-1.19] | 1.17 [1.14-1.20] | 1.17 [1.14-1.21] | 1.12 [1.09-1.15] | 1.13 [1.11-1.15] | 28,721 | 16,386 | 12,681 | 13,748 | 25,177 |
| R51 | 1.28 [1.26-1.30] | 1.29 [1.27-1.32] | 1.32 [1.29-1.35] | 1.20 [1.17-1.24] | 1.19 [1.17-1.21] | 1.18 [1.16-1.20] | 1.13 [1.11-1.15] | 1.15 [1.13-1.18] | 1.15 [1.12-1.19] | 1.12 [1.10-1.14] | 38,088 | 30,723 | 19,242 | 11,486 | 32,875 |
| R52 | 1.40 [1.34-1.46] | 1.42 [1.36-1.48] | 1.42 [1.35-1.48] | 1.12 [0.98-1.28] | 1.28 [1.23-1.34] | 1.26 [1.21-1.31] | 1.20 [1.15-1.25] | 1.21 [1.15-1.27] | 1.06 [0.93-1.22] | 1.20 [1.15-1.25] | 6,030 | 5,669 | 4,672 | 610 | 5,631 |
| R53 | 1.24 [1.21-1.27] | 1.23 [1.20-1.26] | 1.22 [1.19-1.25] | 1.22 [1.16-1.29] | 1.17 [1.14-1.19] | 1.18 [1.15-1.20] | 1.12 [1.10-1.15] | 1.12 [1.09-1.14] | 1.18 [1.11-1.25] | 1.13 [1.10-1.16] | 23,449 | 20,786 | 18,510 | 3,424 | 21,508 |
| R54 | 1.17 [1.14-1.19] | 1.17 [1.15-1.19] | 1.17 [1.14-1.19] | | 1.13 [1.10-1.15] | 1.14 [1.12-1.17] | 1.11 [1.09-1.14] | 1.11 [1.08-1.13] | | 1.12 [1.10-1.14] | 26,745 | 26,733 | 26,699 | NA | 24,911 |
| R55 | 1.20 [1.18-1.21] | 1.20 [1.18-1.22] | 1.20 [1.18-1.22] | 1.10 [1.05-1.14] | 1.14 [1.12-1.16] | 1.15 [1.13-1.17] | 1.12 [1.10-1.14] | 1.12 [1.10-1.14] | 1.05 [1.01-1.10] | 1.11 [1.09-1.13] | 43,614 | 40,258 | 35,478 | 5,290 | 38,136 |
| R56 | 1.14 [1.11-1.16] | 1.26 [1.22-1.29] | 1.26 [1.22-1.30] | 1.02 [0.99-1.05] | 1.07 [1.05-1.10] | 1.10 [1.08-1.13] | 1.15 [1.12-1.19] | 1.16 [1.13-1.20] | 1.01 [0.98-1.04] | 1.06 [1.04-1.09] | 20,741 | 11,494 | 9,214 | 10,145 | 17,909 |
| R57 | 1.18 [1.12-1.25] | 1.20 [1.13-1.26] | 1.18 [1.11-1.24] | 1.01 [0.80-1.29] | 1.15 [1.09-1.21] | 1.15 [1.09-1.22] | 1.12 [1.06-1.18] | 1.10 [1.04-1.16] | 1.03 [0.80-1.32] | 1.15 [1.08-1.21] | 3,599 | 3,468 | 3,306 | 179 | 3,298 |
| R58 | 1.35 [1.19-1.53] | 1.34 [1.17-1.52] | 1.34 [1.17-1.54] | | 1.30 [1.14-1.48] | 1.30 [1.14-1.48] | 1.22 [1.07-1.40] | 1.22 [1.06-1.41] | | 1.29 [1.13-1.48] | 644 | 582 | 533 | NA | 597 |
| R59 | 1.35 [1.32-1.39] | 1.36 [1.32-1.40] | 1.32 [1.28-1.37] | 1.29 [1.22-1.36] | 1.27 [1.24-1.31] | 1.31 [1.28-1.35] | 1.27 [1.23-1.31] | 1.24 [1.20-1.29] | 1.25 [1.18-1.32] | 1.26 [1.22-1.29] | 13,940 | 11,011 | 9,187 | 3,575 | 11,969 |
| R60 | 1.33 [1.30-1.36] | 1.33 [1.30-1.36] | 1.34 [1.31-1.37] | 1.21 [1.05-1.40] | 1.27 [1.24-1.30] | 1.26 [1.23-1.29] | 1.22 [1.19-1.24] | 1.22 [1.19-1.25] | 1.16 [1.00-1.35] | 1.22 [1.19-1.25] | 20,865 | 20,570 | 19,881 | 481 | 19,325 |
| R61 | 1.36 [1.27-1.45] | 1.38 [1.29-1.48] | 1.34 [1.24-1.45] | 1.24 [1.07-1.45] | 1.23 [1.15-1.32] | 1.24 [1.16-1.32] | 1.21 [1.13-1.30] | 1.17 [1.08-1.27] | 1.18 [1.01-1.37] | 1.16 [1.09-1.25] | 2,502 | 2,240 | 1,590 | 476 | 2,119 |
| R62 | 1.07 [1.01-1.12] | 1.09 [0.93-1.27] | 1.26 [1.02-1.56] | 1.06 [1.00-1.12] | 0.99 [0.94-1.05] | 1.02 [0.97-1.07] | 1.00 [0.85-1.17] | 1.18 [0.95-1.46] | 1.02 [0.96-1.07] | 0.97 [0.92-1.02] | 3,726 | 407 | 228 | 3,386 | 3,335 |
| R63 | 1.24 [1.22-1.26] | 1.25 [1.23-1.27] | 1.24 [1.22-1.27] | 1.14 [1.10-1.19] | 1.18 [1.16-1.20] | 1.18 [1.16-1.20] | 1.14 [1.12-1.16] | 1.14 [1.12-1.16] | 1.11 [1.06-1.15] | 1.14 [1.13-1.16] | 44,131 | 39,308 | 35,255 | 6,420 | 39,412 |
| R64 | 1.16 [1.05-1.28] | 1.12 [1.01-1.24] | 1.10 [1.00-1.22] | | 1.09 [0.99-1.21] | 1.17 [1.06-1.30] | 1.09 [0.99-1.21] | 1.08 [0.97-1.20] | | 1.12 [1.01-1.25] | 1,044 | 1,034 | 1,014 | NA | 970 |
| R65 | 1.23 [1.16-1.31] | 1.22 [1.14-1.30] | 1.23 [1.15-1.32] | 1.21 [0.90-1.63] | 1.18 [1.11-1.26] | 1.18 [1.10-1.26] | 1.12 [1.05-1.20] | 1.13 [1.05-1.21] | 1.20 [0.88-1.63] | 1.15 [1.08-1.23] | 2,566 | 2,506 | 2,352 | 123 | 2,426 |
| R68 | 1.21 [1.15-1.27] | 1.24 [1.18-1.31] | 1.24 [1.17-1.31] | 1.03 [0.91-1.17] | 1.17 [1.11-1.23] | 1.19 [1.13-1.25] | 1.18 [1.12-1.25] | 1.17 [1.11-1.24] | 1.01 [0.89-1.15] | 1.16 [1.10-1.23] | 4,208 | 3,725 | 3,469 | 576 | 3,872 |
| R69 | 1.25 [1.23-1.27] | 1.24 [1.22-1.27] | 1.24 [1.22-1.27] | 1.20 [1.15-1.26] | 1.16 [1.14-1.18] | 1.19 [1.18-1.22] | 1.15 [1.13-1.17] | 1.16 [1.14-1.18] | 1.17 [1.11-1.22] | 1.13 [1.11-1.15] | 37,013 | 33,821 | 27,560 | 4,814 | 29,988 |
| R70 | 1.50 [1.27-1.78] | 1.46 [1.22-1.75] | 1.48 [1.23-1.77] | | 1.35 [1.12-1.62] | 1.43 [1.20-1.70] | 1.35 [1.13-1.62] | 1.37 [1.13-1.65] | | 1.28 [1.06-1.55] | 359 | 318 | 293 | NA | 297 |
| R71 | 1.22 [1.00-1.49] | 1.11 [0.90-1.36] | 1.13 [0.91-1.40] | | 1.11 [0.90-1.36] | 1.18 [0.97-1.45] | 1.05 [0.85-1.30] | 1.06 [0.85-1.33] | | 1.09 [0.88-1.35] | 249 | 223 | 200 | NA | 217 |
| R72 | 1.23 [1.13-1.34] | 1.20 [1.10-1.31] | 1.20 [1.09-1.32] | 1.22 [0.97-1.52] | 1.15 [1.05-1.26] | 1.18 [1.09-1.29] | 1.10 [1.01-1.21] | 1.13 [1.03-1.25] | 1.16 [0.92-1.46] | 1.13 [1.03-1.23] | 1,398 | 1,274 | 1,059 | 214 | 1,232 |
| R73 | 1.31 [1.26-1.37] | 1.32 [1.26-1.38] | 1.31 [1.25-1.38] | 1.20 [1.04-1.40] | 1.25 [1.20-1.31] | 1.21 [1.16-1.27] | 1.17 [1.12-1.23] | 1.16 [1.11-1.22] | 1.13 [0.97-1.32] | 1.19 [1.14-1.25] | 5,465 | 5,101 | 4,622 | 481 | 4,997 |
| R74 | 1.21 [1.14-1.30] | 1.17 [1.09-1.26] | 1.21 [1.12-1.30] | 1.24 [1.05-1.46] | 1.15 [1.08-1.23] | 1.15 [1.07-1.23] | 1.06 [0.98-1.14] | 1.10 [1.02-1.19] | 1.14 [0.96-1.36] | 1.12 [1.04-1.20] | 2,353 | 2,104 | 1,804 | 390 | 2,149 |
| R76 | 1.38 [1.23-1.56] | 1.37 [1.21-1.56] | 1.38 [1.20-1.60] | 1.29 [1.02-1.64] | 1.29 [1.14-1.46] | 1.31 [1.16-1.47] | 1.26 [1.11-1.44] | 1.26 [1.09-1.46] | 1.24 [0.97-1.59] | 1.23 [1.08-1.40] | 749 | 608 | 480 | 189 | 641 |
| R77 | 1.26 [1.16-1.37] | 1.29 [1.18-1.40] | 1.27 [1.16-1.40] | 1.12 [0.87-1.45] | 1.23 [1.13-1.34] | 1.18 [1.09-1.29] | 1.16 [1.07-1.27] | 1.17 [1.06-1.28] | 1.12 [0.86-1.46] | 1.18 [1.08-1.29] | 1,496 | 1,395 | 1,220 | 167 | 1,385 |
| R78 | 1.19 [0.95-1.49] | 1.16 [0.91-1.46] | 1.12 [0.88-1.45] | | 1.04 [0.82-1.33] | 1.15 [0.91-1.45] | 1.04 [0.81-1.32] | 1.02 [0.79-1.32] | | 1.05 [0.82-1.35] | 186 | 174 | 153 | NA | 155 |
| R79 | 1.24 [1.22-1.26] | 1.24 [1.21-1.26] | 1.24 [1.22-1.26] | 1.18 [1.09-1.28] | 1.17 [1.15-1.19] | 1.18 [1.15-1.20] | 1.14 [1.11-1.16] | 1.14 [1.11-1.16] | 1.14 [1.05-1.25] | 1.14 [1.11-1.16] | 28,047 | 27,092 | 25,510 | 1,523 | 25,528 |
| R80 | 1.33 [1.22-1.45] | 1.38 [1.25-1.53] | 1.45 [1.29-1.62] | 1.18 [0.99-1.40] | 1.23 [1.12-1.36] | 1.28 [1.17-1.40] | 1.26 [1.14-1.40] | 1.31 [1.17-1.47] | 1.17 [0.98-1.40] | 1.20 [1.09-1.32] | 1,292 | 1,006 | 788 | 366 | 1,101 |
| R81 | 1.20 [1.02-1.42] | 1.15 [0.96-1.38] | | 1.24 [0.92-1.67] | 1.20 [0.99-1.44] | 1.15 [0.97-1.36] | 1.09 [0.90-1.31] | | 1.24 [0.91-1.69] | 1.17 [0.97-1.42] | 340 | 262 | NA | 113 | 265 |
| R82 | 1.20 [1.13-1.28] | 1.21 [1.13-1.30] | 1.16 [1.07-1.25] | 1.18 [1.03-1.35] | 1.12 [1.05-1.20] | 1.15 [1.07-1.22] | 1.10 [1.03-1.18] | 1.06 [0.98-1.15] | 1.15 [1.00-1.32] | 1.10 [1.02-1.17] | 2,432 | 2,073 | 1,563 | 562 | 2,206 |
| R83 | 1.20 [0.89-1.62] | | | | | 1.14 [0.84-1.56] | | | | | 101 | NA | NA | NA | NA |
| R84 | 1.29 [1.15-1.44] | 1.36 [1.20-1.54] | 1.45 [1.28-1.65] | 1.05 [0.84-1.30] | 1.19 [1.06-1.33] | 1.16 [1.03-1.30] | 1.15 [1.01-1.31] | 1.24 [1.09-1.42] | 0.99 [0.80-1.24] | 1.08 [0.96-1.21] | 858 | 668 | 615 | 216 | 782 |
| R85 | 1.32 [1.14-1.53] | 1.39 [1.19-1.62] | 1.32 [1.11-1.58] | | 1.32 [1.13-1.54] | 1.21 [1.04-1.40] | 1.25 [1.06-1.46] | 1.16 [0.97-1.39] | | 1.22 [1.04-1.43] | 460 | 417 | 314 | NA | 404 |
| R87 | 1.16 [1.08-1.24] | 1.17 [1.09-1.25] | 1.08 [0.97-1.21] | 1.09 [0.87-1.37] | 1.07 [0.99-1.16] | 1.12 [1.05-1.20] | 1.11 [1.03-1.19] | 1.03 [0.92-1.15] | 1.08 [0.86-1.37] | 1.06 [0.97-1.15] | 1,971 | 1,966 | 747 | 186 | 1,436 |
| R89 | 1.48 [1.27-1.73] | 1.44 [1.22-1.70] | 1.47 [1.23-1.75] | 1.53 [1.10-2.14] | 1.31 [1.11-1.53] | 1.41 [1.21-1.65] | 1.31 [1.11-1.55] | 1.35 [1.13-1.62] | 1.40 [0.99-1.97] | 1.26 [1.07-1.49] | 452 | 376 | 326 | 101 | 401 |
| R90 | 1.21 [1.11-1.32] | 1.19 [1.08-1.30] | 1.15 [1.04-1.27] | 1.17 [0.95-1.45] | 1.17 [1.07-1.28] | 1.16 [1.06-1.27] | 1.10 [1.00-1.21] | 1.08 [0.98-1.20] | 1.14 [0.92-1.41] | 1.14 [1.04-1.26] | 1,302 | 1,132 | 949 | 229 | 1,147 |
| R91 | 1.26 [1.22-1.29] | 1.25 [1.22-1.29] | 1.25 [1.22-1.29] | 1.29 [1.09-1.53] | 1.20 [1.17-1.24] | 1.20 [1.17-1.24] | 1.15 [1.12-1.18] | 1.15 [1.12-1.18] | 1.26 [1.06-1.50] | 1.17 [1.14-1.21] | 13,726 | 13,469 | 13,019 | 377 | 12,284 |
| R92 | 1.09 [0.81-1.46] | 1.09 [0.81-1.46] | 1.07 [0.80-1.44] | | | 1.06 [0.78-1.43] | 0.97 [0.72-1.32] | 1.01 [0.75-1.37] | | | 109 | 110 | 108 | NA | NA |
| R93 | 1.21 [1.18-1.25] | 1.21 [1.18-1.25] | 1.20 [1.17-1.24] | 1.07 [0.96-1.19] | 1.14 [1.11-1.18] | 1.16 [1.13-1.19] | 1.12 [1.09-1.15] | 1.11 [1.08-1.15] | 1.04 [0.93-1.16] | 1.12 [1.08-1.15] | 13,695 | 13,179 | 11,961 | 913 | 12,289 |
| R94 | 1.26 [1.24-1.29] | 1.26 [1.24-1.29] | 1.26 [1.23-1.29] | 1.16 [1.08-1.24] | 1.19 [1.16-1.21] | 1.21 [1.18-1.23] | 1.17 [1.14-1.19] | 1.16 [1.14-1.19] | 1.12 [1.04-1.21] | 1.16 [1.14-1.19] | 25,164 | 24,148 | 21,195 | 2,130 | 22,573 |
| Injury, poisoning and certain other consequences of external causes (S00-T88) | | | | | | | | | | | | | | | |
| S00 | 1.13 [1.11-1.15] | 1.19 [1.16-1.21] | 1.20 [1.17-1.23] | 0.97 [0.93-1.01] | 1.08 [1.06-1.11] | 1.10 [1.08-1.12] | 1.11 [1.08-1.13] | 1.12 [1.09-1.15] | 0.95 [0.92-0.99] | 1.07 [1.05-1.09] | 26,675 | 21,323 | 18,953 | 6,459 | 23,816 |
| S01 | 1.13 [1.11-1.15] | 1.18 [1.16-1.21] | 1.20 [1.18-1.23] | 0.97 [0.94-1.01] | 1.08 [1.06-1.11] | 1.10 [1.08-1.12] | 1.12 [1.09-1.14] | 1.14 [1.11-1.17] | 0.96 [0.93-1.00] | 1.07 [1.05-1.09] | 28,522 | 21,362 | 18,583 | 8,473 | 24,850 |
| S02 | 1.11 [1.07-1.14] | 1.16 [1.12-1.20] | 1.21 [1.16-1.27] | 1.02 [0.98-1.07] | 1.04 [1.00-1.07] | 1.08 [1.04-1.11] | 1.09 [1.05-1.13] | 1.14 [1.09-1.20] | 1.01 [0.96-1.06] | 1.02 [0.99-1.06] | 10,815 | 7,891 | 4,370 | 4,829 | 8,181 |
| S03 | 1.08 [0.94-1.25] | 1.14 [0.91-1.41] | 1.16 [0.86-1.57] | 1.04 [0.88-1.23] | 1.07 [0.92-1.25] | 1.04 [0.90-1.21] | 1.05 [0.84-1.31] | 1.07 [0.79-1.45] | 1.02 [0.85-1.21] | 1.06 [0.90-1.24] | 483 | 198 | 110 | 336 | 419 |
| S05 | 1.14 [1.06-1.21] | 1.15 [1.07-1.23] | 1.21 [1.11-1.31] | 0.95 [0.83-1.09] | 1.10 [1.02-1.18] | 1.11 [1.03-1.18] | 1.08 [1.01-1.16] | 1.14 [1.05-1.23] | 0.93 [0.81-1.07] | 1.09 [1.02-1.17] | 2,317 | 1,933 | 1,618 | 523 | 1,974 |
| S06 | 1.11 [1.07-1.15] | 1.13 [1.08-1.17] | 1.14 [1.09-1.19] | 1.05 [0.97-1.14] | 1.06 [1.01-1.10] | 1.09 [1.05-1.13] | 1.07 [1.03-1.11] | 1.08 [1.03-1.13] | 1.04 [0.95-1.13] | 1.05 [1.01-1.09] | 7,461 | 6,446 | 5,546 | 1,470 | 6,400 |
| S09 | 1.16 [1.13-1.19] | 1.21 [1.18-1.25] | 1.25 [1.21-1.30] | 1.05 [1.01-1.10] | 1.11 [1.08-1.14] | 1.11 [1.08-1.14] | 1.11 [1.07-1.15] | 1.15 [1.11-1.19] | 1.04 [0.99-1.08] | 1.08 [1.05-1.12] | 15,718 | 10,281 | 8,656 | 6,130 | 13,300 |
| S10 | 1.05 [0.94-1.17] | 1.13 [0.99-1.29] | 1.09 [0.93-1.29] | 0.96 [0.81-1.15] | 0.97 [0.86-1.10] | 1.00 [0.89-1.12] | 1.02 [0.89-1.17] | 0.97 [0.82-1.15] | 0.96 [0.80-1.15] | 0.96 [0.85-1.09] | 794 | 563 | 376 | 321 | 682 |
| S11 | 1.19 [1.03-1.38] | 1.19 [1.01-1.40] | 1.46 [1.17-1.82] | 1.05 [0.82-1.33] | 1.16 [0.98-1.36] | 1.13 [0.97-1.31] | 1.09 [0.92-1.29] | 1.34 [1.06-1.68] | 1.01 [0.79-1.30] | 1.16 [0.98-1.38] | 456 | 361 | 215 | 174 | 377 |
| S12 | 1.18 [1.09-1.27] | 1.18 [1.09-1.27] | 1.18 [1.08-1.28] | 1.00 [0.77-1.29] | 1.15 [1.06-1.25] | 1.15 [1.06-1.24] | 1.11 [1.02-1.20] | 1.10 [1.01-1.20] | 1.01 [0.77-1.32] | 1.15 [1.06-1.24] | 1,813 | 1,759 | 1,558 | 146 | 1,611 |
| S13 | 1.20 [1.04-1.38] | 1.20 [1.03-1.40] | 1.21 [1.00-1.47] | 1.01 [0.79-1.30] | 1.14 [0.98-1.33] | 1.12 [0.97-1.30] | 1.09 [0.92-1.27] | 1.12 [0.92-1.36] | 0.98 [0.76-1.27] | 1.10 [0.94-1.29] | 493 | 393 | 268 | 156 | 411 |
| S14 | 1.07 [0.90-1.27] | 1.07 [0.90-1.28] | 1.08 [0.89-1.32] | | 1.01 [0.84-1.22] | 1.07 [0.90-1.27] | 1.04 [0.87-1.24] | 1.06 [0.87-1.30] | | 1.02 [0.84-1.24] | 332 | 301 | 238 | NA | 261 |
| S19 | 1.24 [1.10-1.41] | 1.30 [1.13-1.49] | 1.28 [1.08-1.52] | 1.10 [0.89-1.36] | 1.16 [1.02-1.32] | 1.15 [1.02-1.30] | 1.14 [0.99-1.32] | 1.11 [0.93-1.32] | 1.06 [0.86-1.32] | 1.08 [0.95-1.24] | 660 | 511 | 333 | 222 | 557 |
| S20 | 1.18 [1.11-1.25] | 1.22 [1.15-1.31] | 1.25 [1.17-1.34] | 0.90 [0.77-1.05] | 1.12 [1.05-1.19] | 1.12 [1.05-1.19] | 1.11 [1.03-1.18] | 1.13 [1.05-1.21] | 0.89 [0.76-1.04] | 1.08 [1.02-1.16] | 2,809 | 2,536 | 2,233 | 409 | 2,554 |
| S21 | 1.11 [0.97-1.26] | 1.15 [1.00-1.33] | 1.45 [1.19-1.77] | 0.85 [0.70-1.04] | 1.05 [0.91-1.22] | 1.06 [0.93-1.21] | 1.05 [0.90-1.22] | 1.32 [1.08-1.63] | 0.83 [0.67-1.02] | 1.06 [0.91-1.23] | 604 | 466 | 266 | 247 | 477 |
| S22 | 1.21 [1.17-1.25] | 1.21 [1.17-1.25] | 1.24 [1.19-1.28] | 1.05 [0.91-1.20] | 1.15 [1.11-1.19] | 1.15 [1.12-1.19] | 1.11 [1.07-1.15] | 1.13 [1.09-1.17] | 1.05 [0.91-1.21] | 1.12 [1.08-1.16] | 9,018 | 8,827 | 8,049 | 540 | 7,993 |
| S27 | 1.11 [1.05-1.18] | 1.10 [1.04-1.18] | 1.15 [1.07-1.23] | 1.02 [0.89-1.16] | 1.04 [0.97-1.11] | 1.08 [1.02-1.15] | 1.04 [0.98-1.11] | 1.08 [1.00-1.16] | 1.02 [0.89-1.18] | 1.04 [0.97-1.11] | 2,716 | 2,484 | 1,917 | 559 | 2,223 |
| S29 | 1.38 [1.23-1.55] | 1.44 [1.28-1.62] | 1.48 [1.30-1.68] | | 1.32 [1.17-1.48] | 1.27 [1.13-1.43] | 1.28 [1.13-1.44] | 1.30 [1.14-1.48] | | 1.24 [1.10-1.41] | 809 | 759 | 661 | NA | 723 |
| S30 | 1.18 [1.13-1.23] | 1.22 [1.16-1.28] | 1.29 [1.22-1.36] | 0.97 [0.89-1.05] | 1.10 [1.05-1.16] | 1.12 [1.07-1.17] | 1.10 [1.04-1.15] | 1.17 [1.11-1.24] | 0.95 [0.87-1.03] | 1.07 [1.02-1.12] | 5,394 | 4,265 | 3,620 | 1,415 | 4,816 |
| S31 | 1.11 [1.03-1.19] | 1.13 [1.02-1.25] | 1.36 [1.19-1.56] | 1.03 [0.93-1.15] | 1.05 [0.97-1.15] | 1.07 [0.99-1.16] | 1.02 [0.93-1.13] | 1.21 [1.05-1.39] | 1.03 [0.93-1.14] | 1.05 [0.96-1.14] | 1,721 | 989 | 558 | 960 | 1,380 |
| S32 | 1.13 [1.09-1.16] | 1.14 [1.10-1.17] | 1.16 [1.12-1.20] | 0.95 [0.84-1.08] | 1.08 [1.04-1.12] | 1.09 [1.05-1.13] | 1.07 [1.04-1.11] | 1.10 [1.06-1.14] | 0.95 [0.83-1.08] | 1.06 [1.02-1.09] | 9,580 | 9,297 | 8,663 | 604 | 8,496 |
| S33 | 1.08 [0.81-1.45] | 1.00 [0.75-1.35] | | | 1.00 [0.74-1.36] | 1.02 [0.76-1.37] | 0.92 [0.68-1.25] | | | 0.92 [0.68-1.26] | 120 | 108 | NA | NA | 103 |
| S35 | 1.08 [0.82-1.43] | 1.17 [0.87-1.56] | | | | 1.09 [0.82-1.45] | 1.17 [0.87-1.59] | | | | 124 | 113 | NA | NA | NA |
| S36 | 1.11 [1.01-1.22] | 1.12 [1.01-1.24] | 1.17 [1.03-1.33] | 0.99 [0.85-1.15] | 1.05 [0.95-1.16] | 1.09 [1.00-1.20] | 1.06 [0.96-1.18] | 1.13 [0.99-1.28] | 1.00 [0.86-1.16] | 1.05 [0.95-1.16] | 1,192 | 927 | 589 | 453 | 953 |
| S37 | 1.00 [0.90-1.12] | 1.05 [0.93-1.18] | 1.02 [0.89-1.17] | 0.99 [0.80-1.22] | 0.96 [0.85-1.08] | 0.98 [0.87-1.09] | 1.01 [0.89-1.14] | 0.98 [0.85-1.13] | 1.00 [0.80-1.24] | 0.95 [0.84-1.08] | 813 | 690 | 509 | 219 | 671 |
| S39 | 1.17 [1.09-1.25] | 1.18 [1.09-1.27] | 1.22 [1.12-1.34] | 1.18 [1.04-1.33] | 1.12 [1.04-1.20] | 1.09 [1.02-1.17] | 1.06 [0.98-1.15] | 1.10 [1.00-1.21] | 1.13 [0.99-1.28] | 1.07 [0.99-1.15] | 2,149 | 1,654 | 1,258 | 681 | 1,862 |
| S40 | 1.14 [1.08-1.20] | 1.17 [1.10-1.23] | 1.21 [1.14-1.28] | 0.92 [0.81-1.04] | 1.10 [1.04-1.16] | 1.09 [1.03-1.15] | 1.07 [1.01-1.13] | 1.11 [1.04-1.18] | 0.90 [0.79-1.02] | 1.07 [1.01-1.13] | 4,026 | 3,550 | 3,241 | 625 | 3,692 |
| S41 | 1.22 [1.13-1.32] | 1.26 [1.16-1.38] | 1.34 [1.21-1.48] | 0.98 [0.85-1.14] | 1.14 [1.05-1.24] | 1.16 [1.07-1.26] | 1.14 [1.04-1.25] | 1.21 [1.09-1.34] | 0.98 [0.84-1.14] | 1.10 [1.01-1.20] | 1,733 | 1,450 | 1,152 | 451 | 1,504 |
| S42 | 1.11 [1.08-1.14] | 1.14 [1.10-1.17] | 1.14 [1.10-1.18] | 1.05 [1.00-1.10] | 1.08 [1.05-1.11] | 1.08 [1.06-1.11] | 1.08 [1.04-1.11] | 1.08 [1.05-1.12] | 1.04 [0.99-1.09] | 1.07 [1.04-1.10] | 14,330 | 10,210 | 9,071 | 4,662 | 12,069 |
| S43 | 1.10 [1.02-1.18] | 1.11 [1.03-1.19] | 1.09 [1.01-1.19] | 1.02 [0.82-1.26] | 1.06 [0.98-1.14] | 1.07 [0.99-1.15] | 1.05 [0.98-1.13] | 1.05 [0.97-1.14] | 1.01 [0.81-1.26] | 1.05 [0.97-1.13] | 1,936 | 1,858 | 1,556 | 216 | 1,623 |
| S44 | 1.24 [0.93-1.66] | | | | | 1.21 [0.90-1.63] | | | | | 120 | NA | NA | NA | NA |
| S46 | 1.10 [0.99-1.23] | 1.12 [1.00-1.25] | 1.07 [0.95-1.22] | | 1.06 [0.95-1.20] | 1.05 [0.94-1.18] | 1.04 [0.92-1.16] | 0.97 [0.85-1.10] | | 1.02 [0.90-1.15] | 789 | 762 | 618 | NA | 686 |
| S49 | 1.20 [1.09-1.31] | 1.28 [1.16-1.40] | 1.27 [1.14-1.40] | 1.17 [0.89-1.53] | 1.18 [1.07-1.30] | 1.12 [1.02-1.23] | 1.15 [1.04-1.27] | 1.15 [1.04-1.28] | 1.14 [0.86-1.51] | 1.14 [1.03-1.26] | 1,232 | 1,141 | 1,037 | 143 | 1,120 |
| S50 | 1.16 [1.10-1.22] | 1.19 [1.13-1.26] | 1.23 [1.16-1.30] | 0.93 [0.84-1.03] | 1.09 [1.03-1.14] | 1.11 [1.05-1.16] | 1.10 [1.04-1.16] | 1.13 [1.07-1.20] | 0.90 [0.81-1.00] | 1.06 [1.01-1.12] | 4,521 | 3,790 | 3,397 | 947 | 4,090 |
| S51 | 1.15 [1.11-1.20] | 1.21 [1.16-1.26] | 1.25 [1.19-1.31] | 0.98 [0.90-1.07] | 1.13 [1.09-1.18] | 1.09 [1.04-1.14] | 1.09 [1.04-1.14] | 1.13 [1.07-1.19] | 0.97 [0.89-1.06] | 1.10 [1.05-1.14] | 6,586 | 5,723 | 4,777 | 1,332 | 5,836 |
| S52 | 1.06 [1.04-1.08] | 1.10 [1.07-1.13] | 1.11 [1.07-1.14] | 1.01 [0.98-1.04] | 1.02 [1.00-1.05] | 1.04 [1.02-1.06] | 1.06 [1.03-1.09] | 1.07 [1.03-1.10] | 1.00 [0.97-1.03] | 1.01 [0.99-1.04] | 23,610 | 12,452 | 10,625 | 11,920 | 18,431 |
| S53 | 0.98 [0.87-1.11] | 1.02 [0.86-1.21] | 1.07 [0.87-1.31] | 0.96 [0.82-1.13] | 1.03 [0.90-1.18] | 0.95 [0.84-1.08] | 0.95 [0.79-1.13] | 0.98 [0.80-1.21] | 0.96 [0.81-1.13] | 1.01 [0.88-1.16] | 648 | 310 | 218 | 376 | 519 |
| S54 | 1.13 [0.94-1.35] | 1.23 [1.00-1.51] | 1.35 [1.00-1.83] | 1.00 [0.75-1.33] | 1.20 [0.98-1.47] | 1.09 [0.90-1.32] | 1.09 [0.88-1.36] | 1.28 [0.94-1.75] | 1.00 [0.75-1.34] | 1.15 [0.93-1.43] | 290 | 222 | 106 | 126 | 238 |
| S55 | 1.34 [1.02-1.75] | 1.47 [1.11-1.95] | | | 1.35 [0.99-1.84] | 1.33 [1.01-1.76] | 1.37 [1.03-1.84] | | | 1.43 [1.04-1.97] | 139 | 121 | NA | NA | 105 |
| S56 | 1.00 [0.90-1.11] | 1.09 [0.97-1.22] | 1.09 [0.91-1.29] | 0.97 [0.83-1.14] | 0.92 [0.82-1.04] | 0.98 [0.88-1.09] | 1.04 [0.92-1.16] | 1.05 [0.87-1.25] | 0.96 [0.82-1.14] | 0.93 [0.82-1.05] | 883 | 718 | 286 | 395 | 649 |
| S59 | 1.24 [1.08-1.43] | 1.31 [1.12-1.53] | 1.39 [1.18-1.64] | 1.02 [0.77-1.35] | 1.15 [0.99-1.33] | 1.17 [1.01-1.35] | 1.19 [1.02-1.40] | 1.28 [1.08-1.51] | 0.95 [0.71-1.27] | 1.11 [0.95-1.29] | 547 | 443 | 399 | 126 | 485 |
| S60 | 1.14 [1.08-1.20] | 1.17 [1.10-1.25] | 1.20 [1.12-1.28] | 1.00 [0.91-1.11] | 1.07 [1.01-1.13] | 1.10 [1.04-1.16] | 1.09 [1.02-1.15] | 1.11 [1.04-1.19] | 0.99 [0.89-1.10] | 1.05 [0.99-1.11] | 3,606 | 2,885 | 2,342 | 993 | 3,200 |
| S61 | 1.05 [1.02-1.08] | 1.09 [1.06-1.13] | 1.14 [1.09-1.18] | 0.96 [0.92-1.00] | 0.99 [0.97-1.02] | 1.02 [0.99-1.04] | 1.03 [1.00-1.06] | 1.06 [1.02-1.11] | 0.96 [0.92-1.00] | 0.98 [0.95-1.01] | 14,534 | 10,128 | 6,327 | 6,292 | 11,519 |
| S62 | 1.05 [1.02-1.08] | 1.08 [1.04-1.12] | 1.09 [1.04-1.15] | 1.01 [0.97-1.06] | 0.99 [0.95-1.02] | 1.02 [0.98-1.05] | 1.02 [0.98-1.06] | 1.03 [0.98-1.08] | 1.00 [0.95-1.04] | 0.98 [0.95-1.02] | 10,253 | 7,126 | 4,076 | 4,846 | 7,794 |
| S63 | 1.19 [1.11-1.29] | 1.20 [1.10-1.31] | 1.21 [1.09-1.34] | 1.14 [0.99-1.31] | 1.13 [1.03-1.23] | 1.14 [1.06-1.24] | 1.11 [1.02-1.21] | 1.14 [1.02-1.27] | 1.10 [0.95-1.27] | 1.09 [1.00-1.19] | 1,608 | 1,353 | 847 | 515 | 1,263 |
| S64 | 1.00 [0.93-1.07] | 1.01 [0.94-1.09] | 1.01 [0.91-1.14] | 0.94 [0.84-1.05] | 0.90 [0.83-0.98] | 0.98 [0.91-1.06] | 0.96 [0.89-1.04] | 0.98 [0.87-1.10] | 0.94 [0.84-1.06] | 0.90 [0.83-0.98] | 1,854 | 1,453 | 665 | 775 | 1,331 |
| S65 | 1.02 [0.89-1.17] | 1.03 [0.88-1.19] | 0.99 [0.80-1.22] | 1.03 [0.83-1.27] | 0.96 [0.83-1.13] | 1.03 [0.90-1.18] | 0.99 [0.85-1.15] | 0.96 [0.78-1.19] | 1.04 [0.83-1.30] | 0.98 [0.84-1.15] | 507 | 397 | 196 | 219 | 379 |
| S66 | 1.03 [0.98-1.09] | 1.04 [0.99-1.10] | 1.00 [0.92-1.09] | 0.99 [0.91-1.07] | 0.95 [0.89-1.00] | 1.01 [0.95-1.06] | 1.00 [0.94-1.05] | 0.97 [0.89-1.05] | 0.98 [0.90-1.07] | 0.95 [0.89-1.01] | 3,464 | 2,821 | 1,209 | 1,443 | 2,435 |
| S67 | 0.87 [0.76-1.00] | 0.78 [0.63-0.97] | | 0.92 [0.79-1.09] | 0.87 [0.75-1.02] | 0.86 [0.75-0.98] | 0.79 [0.63-0.98] | | 0.92 [0.78-1.09] | 0.88 [0.75-1.03] | 501 | 180 | NA | 358 | 396 |
| S68 | 0.98 [0.90-1.07] | 0.97 [0.86-1.09] | 0.97 [0.85-1.12] | 0.99 [0.88-1.12] | 0.92 [0.83-1.02] | 0.98 [0.90-1.08] | 0.97 [0.86-1.09] | 0.99 [0.86-1.14] | 0.98 [0.87-1.11] | 0.92 [0.84-1.02] | 1,198 | 650 | 432 | 652 | 899 |
| S69 | 1.21 [1.11-1.32] | 1.26 [1.14-1.39] | 1.29 [1.15-1.44] | 1.02 [0.88-1.18] | 1.17 [1.07-1.28] | 1.15 [1.05-1.25] | 1.13 [1.02-1.25] | 1.15 [1.02-1.28] | 1.01 [0.87-1.17] | 1.13 [1.03-1.24] | 1,477 | 1,110 | 848 | 460 | 1,287 |
| S70 | 1.16 [1.10-1.22] | 1.16 [1.10-1.23] | 1.19 [1.13-1.26] | 0.94 [0.82-1.07] | 1.08 [1.02-1.14] | 1.11 [1.05-1.17] | 1.07 [1.01-1.13] | 1.11 [1.05-1.18] | 0.94 [0.82-1.07] | 1.06 [1.00-1.12] | 3,968 | 3,487 | 3,264 | 576 | 3,637 |
| S71 | 1.07 [0.97-1.18] | 1.14 [1.01-1.29] | 1.30 [1.09-1.55] | 0.99 [0.86-1.13] | 1.03 [0.92-1.15] | 1.02 [0.92-1.13] | 1.02 [0.89-1.16] | 1.20 [1.00-1.44] | 0.95 [0.82-1.09] | 1.01 [0.90-1.13] | 1,033 | 612 | 320 | 578 | 818 |
| S72 | 1.10 [1.08-1.12] | 1.11 [1.09-1.14] | 1.12 [1.10-1.14] | 0.98 [0.90-1.07] | 1.08 [1.06-1.11] | 1.09 [1.07-1.11] | 1.08 [1.06-1.11] | 1.08 [1.06-1.11] | 0.97 [0.89-1.06] | 1.09 [1.06-1.11] | 25,942 | 24,948 | 24,537 | 1,190 | 22,614 |
| S73 | 1.09 [0.93-1.28] | 1.09 [0.91-1.31] | 1.17 [0.96-1.42] | | 1.05 [0.88-1.24] | 1.04 [0.88-1.22] | 1.02 [0.85-1.23] | 1.07 [0.87-1.31] | | 1.02 [0.86-1.22] | 369 | 293 | 255 | NA | 339 |
| S76 | 1.14 [1.02-1.27] | 1.16 [1.03-1.30] | 1.14 [1.01-1.30] | 1.09 [0.84-1.40] | 1.09 [0.96-1.23] | 1.07 [0.96-1.20] | 1.08 [0.96-1.22] | 1.07 [0.94-1.22] | 1.05 [0.80-1.36] | 1.05 [0.92-1.18] | 811 | 711 | 581 | 167 | 656 |
| S79 | 1.25 [1.16-1.34] | 1.23 [1.14-1.32] | 1.25 [1.15-1.34] | 1.17 [0.92-1.49] | 1.17 [1.09-1.27] | 1.20 [1.11-1.29] | 1.14 [1.06-1.23] | 1.16 [1.07-1.25] | 1.21 [0.94-1.55] | 1.15 [1.07-1.24] | 2,077 | 1,935 | 1,848 | 174 | 1,915 |
| S80 | 1.14 [1.10-1.18] | 1.18 [1.13-1.22] | 1.19 [1.14-1.24] | 0.93 [0.85-1.02] | 1.09 [1.04-1.13] | 1.08 [1.04-1.12] | 1.07 [1.03-1.12] | 1.07 [1.03-1.12] | 0.91 [0.83-0.99] | 1.04 [1.00-1.08] | 7,694 | 6,760 | 6,187 | 1,180 | 6,986 |
| S81 | 1.20 [1.15-1.26] | 1.24 [1.18-1.29] | 1.29 [1.23-1.36] | 1.06 [0.97-1.17] | 1.15 [1.10-1.20] | 1.12 [1.07-1.17] | 1.11 [1.06-1.16] | 1.16 [1.10-1.22] | 1.04 [0.95-1.14] | 1.08 [1.03-1.13] | 6,108 | 5,213 | 4,640 | 1,211 | 5,362 |
| S82 | 1.09 [1.06-1.11] | 1.10 [1.07-1.13] | 1.10 [1.07-1.14] | 1.04 [0.99-1.08] | 1.04 [1.02-1.07] | 1.05 [1.03-1.08] | 1.04 [1.01-1.06] | 1.04 [1.01-1.07] | 1.02 [0.97-1.07] | 1.03 [1.00-1.06] | 16,239 | 12,559 | 9,534 | 4,983 | 12,630 |
| S83 | 1.15 [1.08-1.23] | 1.17 [1.08-1.25] | 1.18 [1.06-1.30] | 1.11 [1.00-1.24] | 1.06 [0.98-1.14] | 1.08 [1.01-1.16] | 1.07 [1.00-1.16] | 1.07 [0.97-1.19] | 1.07 [0.96-1.19] | 1.02 [0.94-1.10] | 2,258 | 1,762 | 925 | 922 | 1,721 |
| S86 | 1.14 [1.04-1.25] | 1.18 [1.07-1.30] | 1.20 [1.07-1.34] | 1.04 [0.82-1.31] | 1.10 [0.99-1.22] | 1.08 [0.98-1.19] | 1.10 [1.00-1.22] | 1.11 [0.98-1.24] | 1.01 [0.79-1.28] | 1.04 [0.93-1.16] | 1,067 | 972 | 693 | 193 | 806 |
| S89 | 1.23 [1.14-1.33] | 1.24 [1.14-1.34] | 1.27 [1.16-1.38] | 1.05 [0.87-1.26] | 1.12 [1.04-1.22] | 1.15 [1.07-1.25] | 1.12 [1.03-1.22] | 1.15 [1.05-1.26] | 1.03 [0.85-1.25] | 1.08 [0.99-1.17] | 1,684 | 1,472 | 1,332 | 274 | 1,509 |
| S90 | 1.11 [1.04-1.20] | 1.13 [1.04-1.22] | 1.11 [1.02-1.21] | 1.02 [0.88-1.17] | 1.04 [0.96-1.12] | 1.06 [0.98-1.14] | 1.02 [0.94-1.10] | 1.01 [0.92-1.10] | 1.01 [0.88-1.17] | 1.01 [0.93-1.09] | 1,979 | 1,574 | 1,345 | 496 | 1,751 |
| S91 | 1.12 [1.05-1.19] | 1.23 [1.13-1.34] | 1.30 [1.17-1.43] | 0.99 [0.90-1.09] | 1.08 [1.01-1.17] | 1.07 [1.00-1.15] | 1.10 [1.01-1.20] | 1.16 [1.05-1.29] | 0.99 [0.89-1.09] | 1.06 [0.98-1.14] | 2,322 | 1,449 | 1,066 | 1,076 | 1,955 |
| S92 | 1.13 [1.07-1.19] | 1.15 [1.09-1.22] | 1.20 [1.12-1.28] | 0.99 [0.88-1.10] | 1.10 [1.04-1.17] | 1.08 [1.02-1.14] | 1.05 [0.99-1.12] | 1.09 [1.02-1.17] | 0.97 [0.86-1.08] | 1.08 [1.01-1.14] | 3,393 | 2,905 | 2,176 | 808 | 2,775 |
| S93 | 1.19 [1.09-1.31] | 1.13 [1.02-1.25] | 1.21 [1.07-1.36] | 1.20 [1.00-1.45] | 1.11 [1.00-1.23] | 1.12 [1.01-1.23] | 1.01 [0.91-1.12] | 1.08 [0.96-1.22] | 1.16 [0.96-1.41] | 1.05 [0.95-1.17] | 1,122 | 945 | 687 | 299 | 937 |
| S96 | 1.14 [0.95-1.38] | 1.11 [0.89-1.38] | 1.19 [0.89-1.59] | 1.22 [0.90-1.65] | 1.16 [0.94-1.43] | 1.06 [0.87-1.29] | 0.95 [0.75-1.20] | 1.00 [0.73-1.36] | 1.21 [0.88-1.65] | 1.06 [0.85-1.32] | 264 | 185 | 106 | 114 | 218 |
| S99 | 1.15 [1.03-1.28] | 1.19 [1.05-1.34] | 1.16 [1.01-1.32] | 1.17 [0.94-1.45] | 1.08 [0.97-1.21] | 1.06 [0.94-1.18] | 1.02 [0.90-1.16] | 1.02 [0.89-1.17] | 1.14 [0.91-1.42] | 1.03 [0.92-1.16] | 852 | 701 | 558 | 220 | 754 |
| T00 | 1.12 [1.04-1.21] | 1.15 [1.06-1.26] | 1.19 [1.08-1.31] | 0.98 [0.84-1.13] | 1.08 [0.99-1.17] | 1.07 [0.99-1.16] | 1.05 [0.96-1.15] | 1.09 [0.99-1.20] | 0.98 [0.84-1.14] | 1.04 [0.96-1.13] | 1,813 | 1,458 | 1,266 | 446 | 1,632 |
| T01 | 1.12 [1.00-1.25] | 1.14 [1.01-1.29] | 1.18 [1.02-1.36] | 1.07 [0.87-1.30] | 1.09 [0.97-1.22] | 1.05 [0.94-1.18] | 0.99 [0.87-1.12] | 1.05 [0.90-1.21] | 1.05 [0.86-1.29] | 1.05 [0.93-1.18] | 889 | 724 | 543 | 271 | 799 |
| T02 | 1.17 [1.03-1.33] | 1.18 [1.02-1.36] | 1.31 [1.12-1.54] | 0.97 [0.76-1.24] | 1.05 [0.91-1.21] | 1.15 [1.01-1.32] | 1.13 [0.98-1.31] | 1.26 [1.07-1.48] | 0.94 [0.73-1.20] | 1.05 [0.90-1.21] | 593 | 466 | 389 | 164 | 471 |
| T09 | 1.22 [1.02-1.47] | 1.20 [0.99-1.47] | 1.22 [0.97-1.52] | | 1.11 [0.92-1.35] | 1.14 [0.95-1.38] | 1.11 [0.90-1.36] | 1.10 [0.87-1.38] | | 1.09 [0.89-1.33] | 303 | 247 | 203 | NA | 257 |
| T11 | 1.19 [1.08-1.31] | 1.22 [1.10-1.35] | 1.28 [1.14-1.43] | 0.95 [0.78-1.16] | 1.15 [1.05-1.27] | 1.12 [1.01-1.23] | 1.09 [0.98-1.21] | 1.15 [1.03-1.29] | 0.93 [0.76-1.15] | 1.11 [1.00-1.23] | 1,227 | 1,043 | 910 | 253 | 1,123 |
| T13 | 1.17 [1.04-1.31] | 1.24 [1.09-1.40] | 1.26 [1.10-1.44] | 0.83 [0.63-1.10] | 1.11 [0.98-1.26] | 1.10 [0.97-1.24] | 1.12 [0.98-1.27] | 1.13 [0.98-1.30] | 0.83 [0.62-1.10] | 1.06 [0.94-1.21] | 755 | 659 | 595 | 121 | 675 |
| T14 | 1.16 [1.03-1.29] | 1.19 [1.05-1.35] | 1.23 [1.07-1.41] | 0.89 [0.71-1.13] | 1.02 [0.90-1.16] | 1.09 [0.97-1.22] | 1.07 [0.94-1.21] | 1.10 [0.95-1.26] | 0.86 [0.68-1.09] | 0.98 [0.87-1.12] | 756 | 614 | 504 | 169 | 604 |
| T15 | 1.05 [0.83-1.32] | | | 1.00 [0.74-1.35] | 0.95 [0.74-1.23] | 1.03 [0.82-1.30] | | | 0.98 [0.72-1.32] | 0.95 [0.74-1.23] | 180 | NA | NA | 106 | 149 |
| T16 | 1.05 [0.98-1.12] | 1.14 [0.89-1.47] | 1.34 [0.98-1.83] | 1.05 [0.97-1.12] | 1.02 [0.94-1.10] | 1.03 [0.96-1.11] | 1.01 [0.78-1.31] | 1.16 [0.83-1.60] | 1.03 [0.96-1.11] | 1.00 [0.93-1.08] | 2,000 | 153 | 102 | 1,875 | 1,686 |
| T17 | 1.14 [1.07-1.20] | 1.36 [1.25-1.48] | 1.30 [1.19-1.43] | 0.99 [0.91-1.07] | 1.07 [1.00-1.14] | 1.08 [1.02-1.15] | 1.22 [1.12-1.33] | 1.18 [1.08-1.30] | 0.97 [0.89-1.05] | 1.03 [0.97-1.10] | 2,952 | 1,523 | 1,296 | 1,516 | 2,532 |
| T18 | 1.16 [1.09-1.24] | 1.31 [1.21-1.42] | 1.26 [1.15-1.39] | 1.01 [0.91-1.11] | 1.07 [0.99-1.14] | 1.09 [1.02-1.16] | 1.18 [1.08-1.28] | 1.13 [1.03-1.25] | 0.98 [0.89-1.08] | 1.02 [0.95-1.10] | 2,326 | 1,419 | 1,052 | 1,062 | 1,901 |
| T19 | 1.19 [0.96-1.46] | 1.29 [1.01-1.64] | | | 1.17 [0.93-1.47] | 1.11 [0.89-1.37] | 1.18 [0.92-1.51] | | | 1.15 [0.91-1.45] | 221 | 163 | NA | NA | 186 |
| T20 | 0.99 [0.89-1.09] | 1.11 [0.96-1.29] | 1.28 [1.06-1.54] | 0.90 [0.80-1.02] | 0.99 [0.88-1.10] | 0.98 [0.88-1.08] | 1.06 [0.91-1.23] | 1.20 [0.99-1.46] | 0.90 [0.79-1.02] | 0.99 [0.89-1.11] | 918 | 406 | 267 | 575 | 787 |
| T21 | 1.00 [0.92-1.08] | 1.14 [1.00-1.29] | 1.20 [1.04-1.39] | 0.91 [0.82-1.01] | 0.99 [0.91-1.09] | 0.99 [0.91-1.08] | 1.07 [0.94-1.22] | 1.11 [0.96-1.29] | 0.91 [0.81-1.01] | 1.00 [0.91-1.09] | 1,351 | 614 | 476 | 791 | 1,177 |
| T22 | 1.01 [0.92-1.11] | 1.09 [0.95-1.24] | 1.13 [0.96-1.34] | 0.94 [0.84-1.06] | 0.94 [0.85-1.04] | 1.00 [0.91-1.09] | 1.01 [0.88-1.16] | 1.06 [0.89-1.26] | 0.94 [0.84-1.06] | 0.95 [0.85-1.05] | 1,111 | 507 | 334 | 678 | 943 |
| T23 | 1.01 [0.93-1.11] | 1.03 [0.90-1.18] | 1.09 [0.92-1.31] | 0.97 [0.87-1.09] | 0.97 [0.88-1.07] | 1.00 [0.91-1.09] | 0.98 [0.85-1.13] | 1.02 [0.85-1.22] | 0.95 [0.85-1.07] | 0.97 [0.88-1.08] | 1,140 | 479 | 291 | 746 | 952 |
| T24 | 0.92 [0.83-1.02] | 0.98 [0.85-1.12] | 0.97 [0.82-1.13] | 0.90 [0.78-1.04] | 0.89 [0.79-0.99] | 0.92 [0.83-1.02] | 0.94 [0.82-1.08] | 0.93 [0.79-1.10] | 0.91 [0.79-1.05] | 0.90 [0.80-1.00] | 916 | 513 | 370 | 469 | 763 |
| T25 | 0.88 [0.77-1.02] | 0.98 [0.79-1.21] | 0.92 [0.71-1.19] | 0.92 [0.77-1.10] | 0.89 [0.76-1.03] | 0.85 [0.74-0.98] | 0.91 [0.73-1.13] | 0.81 [0.61-1.06] | 0.90 [0.75-1.08] | 0.87 [0.75-1.02] | 461 | 205 | 136 | 289 | 396 |
| T29 | 0.93 [0.75-1.15] | 1.26 [0.94-1.68] | | 0.81 [0.61-1.08] | 0.94 [0.75-1.19] | 0.90 [0.73-1.12] | 1.20 [0.89-1.62] | | 0.80 [0.60-1.07] | 0.94 [0.74-1.19] | 202 | 114 | NA | 109 | 175 |
| T31 | 0.97 [0.92-1.02] | 1.05 [0.97-1.15] | 1.10 [0.99-1.23] | 0.92 [0.86-0.99] | 0.94 [0.89-1.00] | 0.95 [0.90-1.01] | 1.00 [0.91-1.09] | 1.03 [0.93-1.16] | 0.92 [0.86-0.98] | 0.94 [0.89-1.00] | 3,030 | 1,224 | 799 | 2,007 | 2,568 |
| T36 | 1.26 [1.13-1.41] | 1.30 [1.13-1.49] | 1.50 [1.22-1.85] | 1.18 [1.01-1.38] | 1.14 [1.00-1.29] | 1.17 [1.04-1.32] | 1.09 [0.94-1.25] | 1.25 [1.01-1.56] | 1.17 [1.00-1.37] | 1.11 [0.98-1.26] | 783 | 514 | 224 | 430 | 637 |
| T37 | 1.30 [1.08-1.56] | 1.40 [1.14-1.73] | 1.55 [1.14-2.09] | 1.14 [0.86-1.50] | 1.21 [1.00-1.47] | 1.21 [1.00-1.46] | 1.20 [0.97-1.50] | 1.28 [0.93-1.76] | 1.17 [0.88-1.55] | 1.20 [0.98-1.46] | 306 | 227 | 111 | 135 | 264 |
| T38 | 1.26 [1.15-1.37] | 1.34 [1.21-1.48] | 1.37 [1.21-1.54] | 1.05 [0.89-1.22] | 1.14 [1.04-1.25] | 1.14 [1.04-1.25] | 1.09 [0.98-1.21] | 1.11 [0.98-1.26] | 1.02 [0.87-1.20] | 1.09 [0.99-1.20] | 1,234 | 955 | 666 | 419 | 1,125 |
| T39 | 1.10 [1.07-1.13] | 1.13 [1.09-1.16] | 1.26 [1.20-1.33] | 1.02 [0.98-1.06] | 1.02 [0.99-1.05] | 1.05 [1.02-1.08] | 0.99 [0.95-1.02] | 1.08 [1.02-1.14] | 1.01 [0.97-1.05] | 1.01 [0.98-1.04] | 13,727 | 8,586 | 3,525 | 7,704 | 10,684 |
| T40 | 1.18 [1.13-1.22] | 1.21 [1.16-1.26] | 1.32 [1.24-1.40] | 0.97 [0.90-1.04] | 1.08 [1.04-1.13] | 1.08 [1.03-1.12] | 1.00 [0.95-1.04] | 1.06 [0.99-1.13] | 0.98 [0.91-1.05] | 1.06 [1.01-1.10] | 5,986 | 5,084 | 2,615 | 2,003 | 5,251 |
| T41 | 1.18 [0.92-1.50] | 1.17 [0.90-1.54] | | | 1.14 [0.87-1.49] | 1.06 [0.82-1.37] | 1.06 [0.80-1.40] | | | 1.11 [0.84-1.47] | 163 | 126 | NA | NA | 134 |
| T42 | 1.17 [1.12-1.23] | 1.23 [1.17-1.29] | 1.30 [1.21-1.39] | 0.96 [0.87-1.05] | 1.05 [1.00-1.10] | 1.08 [1.02-1.13] | 1.00 [0.95-1.06] | 1.06 [0.99-1.14] | 0.98 [0.88-1.08] | 1.03 [0.98-1.08] | 4,246 | 3,759 | 2,072 | 1,090 | 3,661 |
| T43 | 1.16 [1.12-1.21] | 1.22 [1.17-1.27] | 1.32 [1.24-1.40] | 0.97 [0.91-1.03] | 1.06 [1.01-1.10] | 1.09 [1.05-1.13] | 1.02 [0.97-1.06] | 1.08 [1.01-1.15] | 0.98 [0.92-1.04] | 1.04 [1.00-1.09] | 6,905 | 5,667 | 2,343 | 2,734 | 5,711 |
| T44 | 1.16 [1.07-1.26] | 1.22 [1.11-1.33] | 1.23 [1.09-1.40] | 1.01 [0.89-1.15] | 1.07 [0.98-1.16] | 1.10 [1.01-1.19] | 1.04 [0.94-1.14] | 1.02 [0.90-1.17] | 1.00 [0.88-1.15] | 1.06 [0.97-1.16] | 1,483 | 1,137 | 600 | 578 | 1,285 |
| T45 | 1.26 [1.18-1.35] | 1.31 [1.21-1.42] | 1.35 [1.21-1.51] | 1.18 [1.07-1.31] | 1.15 [1.07-1.24] | 1.15 [1.08-1.24] | 1.10 [1.01-1.20] | 1.14 [1.02-1.28] | 1.14 [1.03-1.27] | 1.11 [1.03-1.20] | 2,123 | 1,430 | 819 | 1,026 | 1,822 |
| T46 | 1.19 [1.10-1.30] | 1.23 [1.12-1.36] | 1.26 [1.13-1.39] | 1.00 [0.83-1.21] | 1.12 [1.02-1.23] | 1.11 [1.02-1.22] | 1.06 [0.96-1.17] | 1.06 [0.96-1.18] | 0.98 [0.81-1.19] | 1.09 [0.99-1.19] | 1,326 | 1,135 | 968 | 264 | 1,210 |

#### Supplementary Table 3: Hazard ratios and events from all cohorts

| | Hazard ratio (99% confidence interval) | | | | | | | | | | Events (in exposed) | | | | |
|---|---|---|---|---|---|---|---|---|---|---|---|---|---|---|---|
| | crude | | | | | adjusted | | | | | | | | | |
| Outcome | any age | 18+ | 40+ | <18 | hosp. | any age | 18+ | 40+ | <18 | hosp. | any age | 18+ | 40+ | <18 | hosp. |
| T47 | 1.15 [1.03-1.29] | 1.26 [1.11-1.43] | 1.26 [1.07-1.49] | 0.95 [0.79-1.15] | 1.08 [0.96-1.22] | 1.04 [0.92-1.17] | 1.00 [0.88-1.15] | 0.99 [0.83-1.18] | 0.94 [0.77-1.14] | 1.04 [0.92-1.17] | 777 | 579 | 350 | 291 | 698 |
| T48 | 1.20 [1.04-1.40] | 1.42 [1.14-1.77] | 1.77 [1.30-2.40] | 1.05 [0.87-1.26] | 1.11 [0.95-1.30] | 1.08 [0.93-1.26] | 1.11 [0.87-1.41] | 1.44 [1.04-1.99] | 1.01 [0.83-1.22] | 1.01 [0.85-1.19] | 450 | 198 | 111 | 286 | 381 |
| T49 | 1.02 [0.87-1.21] | 1.30 [0.96-1.77] | | 0.95 [0.79-1.16] | 0.99 [0.82-1.18] | 0.98 [0.82-1.16] | 1.08 [0.78-1.48] | | 0.92 [0.76-1.12] | 0.97 [0.81-1.16] | 333 | 103 | NA | 244 | 288 |
| T50 | 1.18 [1.09-1.27] | 1.23 [1.14-1.34] | 1.26 [1.13-1.40] | 0.96 [0.85-1.09] | 1.05 [0.97-1.14] | 1.08 [1.00-1.17] | 1.02 [0.94-1.12] | 1.03 [0.92-1.15] | 0.95 [0.83-1.08] | 1.02 [0.94-1.11] | 1,779 | 1,390 | 840 | 589 | 1,496 |
| T51 | 1.14 [1.10-1.19] | 1.16 [1.11-1.21] | 1.26 [1.18-1.35] | 0.98 [0.91-1.05] | 1.06 [1.01-1.11] | 1.07 [1.03-1.12] | 0.98 [0.94-1.02] | 1.03 [0.96-1.11] | 1.00 [0.93-1.08] | 1.06 [1.01-1.11] | 5,485 | 4,942 | 2,032 | 1,804 | 4,568 |
| T52 | 0.87 [0.74-1.04] | | | 0.83 [0.68-1.01] | 0.87 [0.72-1.05] | 0.85 [0.72-1.02] | | | 0.83 [0.68-1.01] | 0.86 [0.71-1.04] | 292 | NA | NA | 228 | 249 |
| T54 | 0.98 [0.82-1.16] | 1.01 [0.79-1.29] | | 0.86 [0.70-1.07] | 0.95 [0.79-1.15] | 0.95 [0.80-1.14] | 0.94 [0.73-1.20] | | 0.85 [0.68-1.06] | 0.94 [0.78-1.14] | 337 | 161 | NA | 215 | 270 |
| T56 | 1.17 [0.91-1.50] | 1.17 [0.90-1.54] | 1.18 [0.88-1.59] | | 1.10 [0.85-1.43] | 1.10 [0.85-1.42] | 1.05 [0.80-1.39] | 1.07 [0.79-1.45] | | 1.12 [0.86-1.45] | 153 | 131 | 102 | NA | 138 |
| T58 | 1.08 [0.81-1.43] | | | | | 1.07 [0.80-1.43] | | | | | 115 | NA | NA | NA | NA |
| T59 | 1.05 [0.90-1.23] | 1.08 [0.91-1.27] | 1.13 [0.93-1.37] | 1.04 [0.77-1.41] | 1.04 [0.89-1.23] | 1.02 [0.87-1.19] | 0.97 [0.82-1.16] | 1.03 [0.84-1.25] | 1.05 [0.77-1.43] | 1.03 [0.87-1.21] | 405 | 330 | 256 | 102 | 360 |
| T62 | 1.24 [0.98-1.56] | | | 1.09 [0.82-1.46] | 1.21 [0.93-1.56] | 1.17 [0.92-1.48] | | | 1.06 [0.79-1.43] | 1.15 [0.88-1.50] | 179 | NA | NA | 112 | 146 |
| T63 | 1.10 [0.89-1.37] | 0.99 [0.77-1.26] | 0.95 [0.72-1.25] | | 1.00 [0.78-1.28] | 1.02 [0.82-1.27] | 0.88 [0.68-1.13] | 0.88 [0.66-1.16] | | 0.95 [0.74-1.22] | 200 | 146 | 111 | NA | 152 |
| T65 | 1.05 [0.89-1.24] | 1.14 [0.88-1.47] | 1.29 [0.95-1.75] | 0.99 [0.80-1.21] | 1.01 [0.85-1.20] | 1.00 [0.84-1.18] | 0.96 [0.73-1.25] | 1.10 [0.80-1.52] | 0.97 [0.78-1.19] | 0.99 [0.83-1.18] | 338 | 142 | 103 | 215 | 300 |
| T68 | 1.10 [1.02-1.19] | 1.13 [1.05-1.22] | 1.11 [1.03-1.20] | | 1.07 [0.99-1.16] | 1.12 [1.04-1.21] | 1.12 [1.04-1.21] | 1.11 [1.02-1.20] | | 1.11 [1.03-1.21] | 1,908 | 1,849 | 1,772 | NA | 1,768 |
| T71 | 0.96 [0.76-1.22] | | | | 0.88 [0.68-1.14] | 0.94 [0.74-1.21] | | | | 0.89 [0.68-1.16] | 167 | NA | NA | NA | 140 |
| T74 | 0.91 [0.76-1.08] | 1.17 [0.90-1.52] | | 0.85 [0.68-1.06] | 0.91 [0.75-1.09] | 0.86 [0.72-1.03] | 0.95 [0.72-1.25] | | 0.82 [0.66-1.03] | 0.89 [0.73-1.07] | 311 | 139 | NA | 195 | 276 |
| T75 | 1.09 [0.89-1.34] | 1.24 [0.95-1.63] | | 1.01 [0.76-1.32] | 1.00 [0.80-1.26] | 1.02 [0.83-1.26] | 1.13 [0.85-1.49] | | 0.97 [0.73-1.28] | 0.96 [0.76-1.22] | 226 | 130 | NA | 130 | 173 |
| T78 | 3.07 [2.96-3.18] | 1.99 [1.89-2.09] | 1.72 [1.61-1.83] | 4.81 [4.57-5.06] | 2.90 [2.78-3.01] | 2.54 [2.45-2.64] | 1.58 [1.49-1.66] | 1.45 [1.36-1.55] | 3.86 [3.66-4.07] | 2.42 [2.32-2.52] | 10,510 | 4,247 | 2,499 | 7,022 | 8,795 |
| T79 | 1.09 [1.03-1.17] | 1.11 [1.04-1.18] | 1.12 [1.04-1.21] | 1.05 [0.90-1.22] | 1.04 [0.98-1.12] | 1.07 [1.00-1.14] | 1.04 [0.97-1.11] | 1.07 [0.99-1.15] | 1.03 [0.89-1.21] | 1.04 [0.97-1.12] | 2,569 | 2,323 | 1,932 | 438 | 2,157 |
| T80 | 1.33 [1.23-1.45] | 1.39 [1.27-1.53] | 1.38 [1.25-1.52] | 1.09 [0.90-1.33] | 1.26 [1.15-1.38] | 1.27 [1.16-1.39] | 1.28 [1.17-1.41] | 1.26 [1.14-1.40] | 1.08 [0.89-1.32] | 1.24 [1.13-1.36] | 1,377 | 1,160 | 991 | 275 | 1,230 |
| T81 | 1.29 [1.27-1.31] | 1.30 [1.28-1.32] | 1.29 [1.26-1.31] | 1.22 [1.17-1.27] | 1.20 [1.18-1.22] | 1.22 [1.20-1.24] | 1.19 [1.17-1.21] | 1.18 [1.16-1.20] | 1.17 [1.12-1.22] | 1.16 [1.14-1.18] | 40,521 | 36,170 | 29,984 | 6,499 | 34,896 |
| T82 | 1.31 [1.27-1.35] | 1.34 [1.30-1.39] | 1.34 [1.30-1.39] | 1.01 [0.91-1.11] | 1.19 [1.16-1.23] | 1.25 [1.21-1.29] | 1.21 [1.17-1.25] | 1.21 [1.17-1.25] | 0.99 [0.89-1.09] | 1.17 [1.13-1.21] | 9,620 | 8,858 | 8,174 | 979 | 8,863 |
| T83 | 1.14 [1.10-1.18] | 1.15 [1.12-1.20] | 1.14 [1.09-1.18] | 1.02 [0.87-1.20] | 1.08 [1.04-1.12] | 1.11 [1.07-1.15] | 1.10 [1.06-1.14] | 1.08 [1.04-1.12] | 0.96 [0.81-1.14] | 1.07 [1.03-1.11] | 8,504 | 8,359 | 7,523 | 379 | 7,942 |
| T84 | 1.31 [1.28-1.35] | 1.32 [1.28-1.36] | 1.34 [1.30-1.38] | 1.14 [1.04-1.25] | 1.20 [1.17-1.24] | 1.24 [1.20-1.27] | 1.21 [1.18-1.25] | 1.23 [1.19-1.27] | 1.11 [1.01-1.22] | 1.17 [1.13-1.20] | 12,672 | 12,081 | 10,652 | 1,194 | 11,593 |
| T85 | 1.30 [1.26-1.35] | 1.32 [1.28-1.37] | 1.31 [1.26-1.36] | 1.17 [1.08-1.27] | 1.19 [1.15-1.23] | 1.23 [1.19-1.28] | 1.21 [1.17-1.26] | 1.20 [1.16-1.25] | 1.12 [1.03-1.22] | 1.15 [1.11-1.20] | 8,581 | 7,413 | 6,359 | 1,493 | 7,828 |
| T86 | 1.33 [1.22-1.46] | 1.36 [1.24-1.49] | 1.38 [1.24-1.53] | 1.02 [0.82-1.28] | 1.17 [1.07-1.28] | 1.29 [1.18-1.41] | 1.26 [1.15-1.39] | 1.25 [1.12-1.40] | 1.01 [0.81-1.27] | 1.17 [1.06-1.29] | 1,200 | 1,070 | 842 | 200 | 1,095 |
| T87 | 1.35 [1.20-1.52] | 1.30 [1.16-1.47] | 1.28 [1.13-1.45] | | 1.21 [1.07-1.36] | 1.25 [1.11-1.41] | 1.14 [1.01-1.29] | 1.08 [0.95-1.23] | | 1.17 [1.04-1.32] | 716 | 681 | 622 | NA | 663 |
| T88 | 1.47 [1.39-1.56] | 1.49 [1.40-1.59] | 1.46 [1.36-1.56] | 1.40 [1.23-1.60] | 1.36 [1.28-1.45] | 1.33 [1.25-1.41] | 1.29 [1.20-1.37] | 1.28 [1.19-1.38] | 1.29 [1.13-1.48] | 1.27 [1.19-1.35] | 2,944 | 2,490 | 1,972 | 623 | 2,573 |
| T90 | 1.29 [1.21-1.38] | 1.31 [1.22-1.41] | 1.38 [1.26-1.52] | 1.15 [1.02-1.29] | 1.16 [1.08-1.25] | 1.23 [1.15-1.31] | 1.19 [1.11-1.29] | 1.22 [1.11-1.35] | 1.13 [1.00-1.27] | 1.14 [1.06-1.23] | 2,230 | 1,854 | 1,061 | 718 | 1,836 |
| T91 | 1.30 [1.17-1.44] | 1.24 [1.11-1.37] | 1.28 [1.13-1.44] | 1.07 [0.79-1.43] | 1.14 [1.02-1.26] | 1.19 [1.07-1.32] | 1.08 [0.97-1.21] | 1.13 [0.99-1.27] | 1.08 [0.79-1.46] | 1.08 [0.97-1.20] | 924 | 867 | 662 | 113 | 820 |
| T92 | 1.17 [1.09-1.26] | 1.20 [1.11-1.30] | 1.24 [1.12-1.36] | 1.07 [0.91-1.26] | 1.09 [1.00-1.18] | 1.12 [1.04-1.21] | 1.11 [1.02-1.21] | 1.15 [1.04-1.27] | 1.04 [0.88-1.23] | 1.07 [0.98-1.16] | 1,618 | 1,399 | 980 | 379 | 1,338 |
| T93 | 1.22 [1.15-1.31] | 1.25 [1.16-1.34] | 1.28 [1.18-1.39] | 1.10 [0.96-1.26] | 1.14 [1.06-1.22] | 1.15 [1.07-1.23] | 1.14 [1.06-1.22] | 1.16 [1.06-1.26] | 1.05 [0.91-1.21] | 1.09 [1.01-1.18] | 2,204 | 1,911 | 1,283 | 541 | 1,790 |
| T94 | 1.26 [1.01-1.57] | 1.24 [0.98-1.56] | 1.11 [0.85-1.44] | | 1.11 [0.87-1.41] | 1.21 [0.96-1.52] | 1.12 [0.88-1.42] | 0.99 [0.76-1.29] | | 1.06 [0.83-1.35] | 196 | 181 | 139 | NA | 168 |
| T95 | 0.97 [0.79-1.19] | 1.31 [0.99-1.72] | | 0.80 [0.60-1.07] | 1.06 [0.85-1.31] | 0.96 [0.78-1.18] | 1.26 [0.95-1.68] | | 0.82 [0.61-1.11] | 1.08 [0.86-1.34] | 216 | 120 | NA | 105 | 200 |
| T98 | 1.19 [1.01-1.41] | 1.34 [1.13-1.60] | 1.35 [1.12-1.63] | | 1.08 [0.91-1.29] | 1.10 [0.92-1.30] | 1.21 [1.01-1.44] | 1.23 [1.01-1.50] | | 1.04 [0.87-1.24] | 344 | 320 | 271 | NA | 309 |
| Codes for special purposes (U00-U89) | | | | | | | | | | | | | | | |
| U07 | 1.27 [1.23-1.31] | 1.25 [1.21-1.29] | 1.28 [1.24-1.32] | 1.14 [1.02-1.29] | 1.21 [1.17-1.25] | 1.18 [1.15-1.22] | 1.13 [1.09-1.17] | 1.15 [1.11-1.19] | 1.09 [0.96-1.23] | 1.15 [1.12-1.19] | 11,865 | 11,558 | 10,299 | 834 | 11,149 |
| U80 | 1.49 [1.42-1.56] | 1.50 [1.43-1.57] | 1.49 [1.41-1.57] | 1.41 [1.20-1.65] | 1.40 [1.33-1.47] | 1.40 [1.33-1.47] | 1.36 [1.29-1.43] | 1.34 [1.27-1.41] | 1.33 [1.12-1.57] | 1.35 [1.29-1.42] | 4,665 | 4,351 | 3,975 | 426 | 4,395 |
| U81 | 1.19 [0.96-1.46] | 1.18 [0.95-1.46] | 1.11 [0.89-1.39] | | 1.16 [0.94-1.44] | 1.14 [0.92-1.41] | 1.08 [0.87-1.34] | 1.04 [0.83-1.31] | | 1.14 [0.92-1.42] | 227 | 221 | 203 | NA | 215 |
| U82 | 1.40 [1.35-1.45] | 1.39 [1.34-1.44] | 1.38 [1.32-1.43] | 1.30 [1.17-1.45] | 1.32 [1.27-1.37] | 1.32 [1.27-1.37] | 1.25 [1.20-1.30] | 1.24 [1.19-1.29] | 1.26 [1.13-1.41] | 1.28 [1.23-1.33] | 8,962 | 8,351 | 7,484 | 984 | 8,556 |
| U83 | 1.35 [1.29-1.41] | 1.34 [1.28-1.40] | 1.35 [1.29-1.42] | 1.22 [1.06-1.41] | 1.29 [1.23-1.35] | 1.28 [1.22-1.34] | 1.20 [1.14-1.26] | 1.21 [1.15-1.28] | 1.17 [1.01-1.35] | 1.25 [1.20-1.31] | 5,253 | 4,881 | 4,456 | 564 | 5,000 |
| U84 | 1.40 [1.02-1.93] | | | | 1.20 [0.87-1.66] | 1.35 [0.97-1.87] | | | | 1.19 [0.86-1.66] | 107 | NA | NA | NA | 101 |
| U88 | 1.29 [1.16-1.42] | 1.28 [1.15-1.42] | 1.30 [1.17-1.45] | | 1.16 [1.05-1.29] | 1.22 [1.10-1.35] | 1.16 [1.04-1.29] | 1.19 [1.06-1.33] | | 1.12 [1.01-1.24] | 1,024 | 969 | 908 | NA | 973 |
| U89 | 1.34 [1.22-1.47] | 1.34 [1.21-1.47] | 1.35 [1.22-1.49] | 1.33 [1.00-1.77] | 1.29 [1.17-1.42] | 1.26 [1.14-1.38] | 1.20 [1.08-1.32] | 1.21 [1.09-1.34] | 1.28 [0.96-1.72] | 1.25 [1.13-1.38] | 1,213 | 1,118 | 1,031 | 127 | 1,151 |
| External causes of morbidity (V00-Y99) | | | | | | | | | | | | | | | |
| V01 | 0.99 [0.76-1.29] | | | | 1.02 [0.76-1.36] | 0.96 [0.73-1.26] | | | | 1.00 [0.74-1.33] | 135 | NA | NA | NA | 112 |
| V02 | 0.99 [0.74-1.32] | | | | | 0.99 [0.74-1.33] | | | | | 110 | NA | NA | NA | NA |
| V03 | 1.02 [0.96-1.09] | 1.10 [1.00-1.20] | 1.18 [1.06-1.31] | 0.93 [0.85-1.02] | 0.98 [0.91-1.06] | 1.00 [0.94-1.07] | 1.05 [0.96-1.15] | 1.14 [1.02-1.26] | 0.93 [0.85-1.02] | 0.97 [0.90-1.05] | 2,231 | 1,164 | 862 | 1,202 | 1,719 |
| V04 | 1.32 [1.03-1.70] | 1.18 [0.87-1.61] | | | 1.34 [1.00-1.78] | 1.27 [0.98-1.64] | 1.14 [0.83-1.55] | | | 1.33 [0.99-1.78] | 160 | 104 | NA | NA | 124 |
| V09 | 1.21 [1.05-1.38] | 1.29 [1.09-1.53] | 1.30 [1.08-1.56] | 1.12 [0.90-1.39] | 1.19 [1.03-1.37] | 1.13 [0.98-1.30] | 1.13 [0.95-1.35] | 1.13 [0.94-1.37] | 1.08 [0.87-1.35] | 1.13 [0.98-1.31] | 559 | 353 | 310 | 223 | 495 |
| V11 | 1.00 [0.76-1.31] | | | | 0.97 [0.71-1.31] | 0.95 [0.71-1.26] | | | | 0.96 [0.71-1.32] | 128 | NA | NA | NA | 106 |
| V13 | 0.91 [0.80-1.03] | 0.90 [0.77-1.05] | 0.90 [0.74-1.11] | 0.94 [0.78-1.13] | 0.81 [0.70-0.95] | 0.91 [0.80-1.03] | 0.90 [0.77-1.06] | 0.90 [0.73-1.11] | 0.97 [0.80-1.17] | 0.82 [0.70-0.96] | 573 | 356 | 207 | 285 | 384 |
| V17 | 0.98 [0.80-1.19] | 0.84 [0.63-1.11] | | 0.97 [0.75-1.25] | 0.87 [0.70-1.09] | 0.97 [0.79-1.18] | 0.79 [0.59-1.05] | | 0.96 [0.75-1.24] | 0.88 [0.70-1.11] | 245 | 108 | NA | 157 | 180 |
| V18 | 1.02 [0.97-1.07] | 1.02 [0.94-1.10] | 1.05 [0.95-1.16] | 1.03 [0.97-1.09] | 0.99 [0.93-1.05] | 1.00 [0.95-1.05] | 1.00 [0.93-1.09] | 1.04 [0.94-1.15] | 1.00 [0.94-1.07] | 0.98 [0.92-1.04] | 3,972 | 1,477 | 895 | 2,787 | 2,917 |
| V19 | 1.07 [0.95-1.22] | 1.01 [0.84-1.23] | 1.10 [0.86-1.41] | 1.02 [0.87-1.19] | 0.95 [0.82-1.10] | 1.05 [0.92-1.19] | 0.99 [0.81-1.21] | 1.10 [0.85-1.42] | 1.01 [0.87-1.19] | 0.94 [0.81-1.09] | 620 | 239 | 143 | 418 | 450 |
| V23 | 0.96 [0.86-1.08] | 0.97 [0.86-1.09] | 0.95 [0.78-1.16] | 0.93 [0.79-1.10] | 0.89 [0.77-1.01] | 0.96 [0.86-1.08] | 0.94 [0.83-1.06] | 0.91 [0.74-1.12] | 0.95 [0.80-1.13] | 0.91 [0.79-1.05] | 719 | 596 | 211 | 355 | 490 |
| V27 | 0.87 [0.69-1.10] | 0.84 [0.65-1.09] | | 0.91 [0.67-1.23] | 0.80 [0.61-1.06] | 0.82 [0.64-1.04] | 0.78 [0.59-1.03] | | 0.89 [0.65-1.22] | 0.78 [0.59-1.04] | 176 | 125 | NA | 110 | 125 |
| V28 | 1.02 [0.92-1.12] | 0.98 [0.88-1.09] | 1.03 [0.87-1.23] | 1.01 [0.88-1.15] | 0.92 [0.82-1.03] | 0.99 [0.90-1.09] | 0.93 [0.82-1.04] | 0.96 [0.81-1.15] | 0.98 [0.85-1.12] | 0.91 [0.81-1.03] | 1,012 | 699 | 298 | 536 | 715 |
| V29 | 1.05 [0.87-1.26] | 1.06 [0.86-1.32] | | 0.91 [0.69-1.20] | 1.08 [0.87-1.33] | 1.01 [0.83-1.22] | 1.00 [0.80-1.25] | | 0.91 [0.69-1.21] | 1.06 [0.85-1.32] | 264 | 191 | NA | 125 | 198 |
| V43 | 1.14 [1.06-1.22] | 1.14 [1.06-1.23] | 1.16 [1.05-1.28] | 1.04 [0.91-1.19] | 1.07 [0.99-1.16] | 1.09 [1.01-1.17] | 1.05 [0.98-1.14] | 1.06 [0.96-1.18] | 1.02 [0.88-1.17] | 1.04 [0.95-1.12] | 1,868 | 1,641 | 960 | 537 | 1,439 |
| V44 | 1.19 [0.95-1.50] | 1.10 [0.87-1.41] | | | 1.15 [0.88-1.49] | 1.11 [0.87-1.40] | 1.04 [0.81-1.33] | | | 1.13 [0.86-1.48] | 178 | 155 | NA | NA | 133 |
| V47 | 1.16 [1.04-1.28] | 1.24 [1.11-1.38] | 1.26 [1.07-1.48] | 1.04 [0.88-1.23] | 1.18 [1.05-1.33] | 1.11 [1.00-1.24] | 1.16 [1.04-1.30] | 1.15 [0.98-1.36] | 1.02 [0.86-1.22] | 1.16 [1.02-1.31] | 892 | 791 | 376 | 351 | 679 |
| V48 | 1.14 [1.03-1.25] | 1.15 [1.05-1.28] | 1.21 [1.07-1.36] | 1.13 [0.94-1.36] | 1.08 [0.97-1.20] | 1.09 [0.99-1.21] | 1.07 [0.97-1.19] | 1.12 [0.99-1.26] | 1.12 [0.92-1.36] | 1.05 [0.94-1.17] | 1,068 | 964 | 679 | 277 | 867 |
| V49 | 1.11 [0.99-1.23] | 1.12 [0.99-1.26] | 1.21 [1.03-1.41] | 1.06 [0.88-1.28] | 1.06 [0.94-1.21] | 1.08 [0.96-1.20] | 1.03 [0.92-1.17] | 1.12 [0.95-1.32] | 1.06 [0.87-1.29] | 1.06 [0.93-1.20] | 797 | 656 | 376 | 267 | 608 |
| V78 | 1.14 [0.98-1.34] | 1.19 [1.01-1.40] | 1.17 [0.99-1.38] | | 1.16 [0.98-1.37] | 1.09 [0.93-1.29] | 1.12 [0.95-1.32] | 1.10 [0.92-1.30] | | 1.13 [0.96-1.34] | 389 | 378 | 357 | NA | 360 |
| V79 | 1.08 [0.83-1.42] | 1.02 [0.76-1.36] | 1.01 [0.74-1.38] | | 1.07 [0.80-1.44] | 1.03 [0.78-1.35] | 0.89 [0.66-1.21] | 0.93 [0.68-1.28] | | 1.00 [0.74-1.36] | 130 | 111 | 101 | NA | 117 |
| V80 | 0.95 [0.86-1.04] | 0.88 [0.79-0.99] | 0.76 [0.64-0.90] | 1.03 [0.90-1.17] | 0.88 [0.79-0.98] | 0.93 [0.85-1.02] | 0.87 [0.77-0.98] | 0.76 [0.64-0.91] | 1.01 [0.88-1.15] | 0.87 [0.78-0.98] | 1,094 | 648 | 288 | 614 | 774 |
| V82 | 1.42 [1.05-1.92] | 1.52 [1.12-2.07] | 1.37 [0.99-1.90] | | 1.39 [1.02-1.90] | 1.42 [1.05-1.93] | 1.47 [1.08-2.00] | 1.33 [0.95-1.85] | | 1.41 [1.02-1.93] | 120 | 114 | 104 | NA | 108 |
| V86 | 1.08 [0.90-1.29] | 1.04 [0.80-1.35] | | 1.14 [0.91-1.42] | 1.05 [0.85-1.29] | 1.02 [0.85-1.23] | 0.96 [0.73-1.26] | | 1.10 [0.87-1.38] | 1.03 [0.84-1.27] | 300 | 128 | NA | 215 | 237 |
| V89 | 1.20 [1.02-1.40] | 1.28 [1.08-1.53] | 1.39 [1.08-1.79] | 0.90 [0.67-1.20] | 1.15 [0.96-1.39] | 1.12 [0.95-1.32] | 1.19 [0.99-1.42] | 1.23 [0.94-1.59] | 0.85 [0.63-1.15] | 1.10 [0.91-1.34] | 370 | 315 | 145 | 111 | 262 |
| W00 | 1.09 [1.01-1.18] | 1.10 [1.02-1.19] | 1.08 [0.99-1.18] | 1.19 [0.94-1.50] | 1.04 [0.96-1.14] | 1.07 [0.99-1.16] | 1.08 [0.99-1.17] | 1.06 [0.97-1.16] | 1.19 [0.93-1.51] | 1.03 [0.95-1.13] | 1,522 | 1,387 | 1,193 | 184 | 1,177 |
| W01 | 1.14 [1.12-1.16] | 1.16 [1.13-1.18] | 1.16 [1.14-1.18] | 1.03 [0.99-1.08] | 1.10 [1.08-1.12] | 1.10 [1.08-1.12] | 1.09 [1.07-1.11] | 1.10 [1.08-1.12] | 1.02 [0.97-1.06] | 1.08 [1.06-1.10] | 34,003 | 29,400 | 27,753 | 5,207 | 29,222 |
| W02 | 1.01 [0.94-1.09] | 1.01 [0.89-1.15] | 1.02 [0.84-1.24] | 1.00 [0.92-1.09] | 0.99 [0.91-1.07] | 0.99 [0.92-1.07] | 0.98 [0.85-1.12] | 1.00 [0.82-1.22] | 1.00 [0.92-1.08] | 0.97 [0.89-1.06] | 1,916 | 513 | 226 | 1,534 | 1,445 |
| W03 | 1.05 [0.99-1.11] | 1.06 [0.96-1.16] | 1.03 [0.88-1.21] | 1.02 [0.95-1.10] | 0.98 [0.91-1.05] | 1.01 [0.95-1.08] | 0.99 [0.90-1.09] | 0.97 [0.82-1.14] | 1.00 [0.93-1.07] | 0.95 [0.88-1.02] | 2,654 | 1,038 | 363 | 1,996 | 1,897 |
| W04 | 1.01 [0.87-1.18] | | | 0.97 [0.82-1.15] | 1.01 [0.86-1.20] | 1.00 [0.86-1.17] | | | 0.97 [0.81-1.15] | 1.03 [0.88-1.22] | 409 | NA | NA | 327 | 358 |
| W05 | 1.24 [1.10-1.39] | 1.26 [1.11-1.42] | 1.26 [1.12-1.43] | | 1.15 [1.02-1.30] | 1.19 [1.05-1.34] | 1.15 [1.01-1.30] | 1.14 [1.01-1.30] | | 1.14 [1.01-1.29] | 722 | 693 | 670 | NA | 685 |
| W06 | 1.17 [1.12-1.21] | 1.19 [1.15-1.24] | 1.19 [1.14-1.24] | 1.03 [0.94-1.13] | 1.12 [1.08-1.16] | 1.13 [1.09-1.17] | 1.11 [1.07-1.16] | 1.11 [1.07-1.16] | 1.02 [0.93-1.12] | 1.10 [1.06-1.14] | 8,242 | 7,160 | 7,055 | 1,123 | 7,666 |
| W07 | 1.14 [1.09-1.19] | 1.19 [1.13-1.25] | 1.21 [1.15-1.28] | 0.97 [0.88-1.06] | 1.08 [1.03-1.13] | 1.11 [1.06-1.16] | 1.12 [1.06-1.18] | 1.13 [1.07-1.20] | 0.96 [0.87-1.06] | 1.06 [1.01-1.12] | 4,869 | 3,857 | 3,763 | 1,039 | 4,389 |
| W08 | 1.09 [1.00-1.17] | 1.24 [1.11-1.37] | 1.22 [1.10-1.37] | 0.89 [0.80-1.00] | 1.04 [0.96-1.13] | 1.06 [0.98-1.15] | 1.14 [1.02-1.27] | 1.13 [1.01-1.26] | 0.89 [0.80-1.00] | 1.03 [0.95-1.12] | 1,652 | 949 | 893 | 723 | 1,500 |
| W09 | 1.05 [1.01-1.10] | 1.23 [0.97-1.57] | | 1.05 [1.01-1.10] | 1.02 [0.97-1.07] | 1.04 [0.99-1.08] | 1.17 [0.91-1.50] | | 1.04 [0.99-1.09] | 1.01 [0.96-1.06] | 5,335 | 162 | NA | 5,222 | 4,350 |
| W10 | 1.15 [1.12-1.18] | 1.19 [1.16-1.23] | 1.19 [1.16-1.23] | 1.02 [0.95-1.08] | 1.11 [1.08-1.14] | 1.11 [1.08-1.14] | 1.11 [1.08-1.15] | 1.11 [1.08-1.15] | 1.00 [0.94-1.07] | 1.09 [1.06-1.12] | 13,793 | 11,995 | 10,622 | 2,267 | 11,894 |
| W11 | 1.06 [0.98-1.13] | 1.06 [0.99-1.14] | 1.02 [0.95-1.10] | 1.05 [0.80-1.38] | 0.99 [0.91-1.07] | 1.02 [0.94-1.09] | 1.02 [0.94-1.10] | 0.98 [0.90-1.05] | 1.06 [0.80-1.40] | 0.97 [0.89-1.05] | 1,786 | 1,714 | 1,537 | 133 | 1,366 |
| W12 | 1.19 [0.90-1.57] | 1.15 [0.86-1.54] | | | | 1.16 [0.88-1.55] | 1.14 [0.85-1.54] | | | | 124 | 105 | NA | NA | NA |
| W13 | 0.99 [0.92-1.07] | 1.05 [0.94-1.17] | 1.14 [0.98-1.31] | 0.95 [0.87-1.05] | 0.97 [0.89-1.06] | 0.97 [0.90-1.05] | 0.98 [0.88-1.10] | 1.07 [0.92-1.24] | 0.94 [0.85-1.04] | 0.97 [0.89-1.06] | 1,646 | 763 | 427 | 1,057 | 1,283 |
| W14 | 0.99 [0.87-1.12] | | | 0.95 [0.83-1.08] | 1.00 [0.87-1.16] | 0.99 [0.87-1.12] | | | 0.94 [0.82-1.08] | 1.01 [0.87-1.17] | 625 | NA | NA | 562 | 482 |
| W17 | 1.05 [0.99-1.11] | 1.16 [1.07-1.25] | 1.19 [1.09-1.31] | 0.97 [0.91-1.04] | 1.00 [0.94-1.06] | 1.02 [0.97-1.08] | 1.09 [1.00-1.18] | 1.12 [1.02-1.23] | 0.95 [0.88-1.02] | 0.99 [0.93-1.05] | 3,262 | 1,473 | 1,105 | 1,964 | 2,609 |
| W18 | 1.19 [1.16-1.22] | 1.21 [1.18-1.24] | 1.22 [1.19-1.25] | 1.06 [1.00-1.12] | 1.14 [1.11-1.16] | 1.15 [1.12-1.17] | 1.13 [1.10-1.15] | 1.13 [1.11-1.16] | 1.05 [0.99-1.11] | 1.12 [1.09-1.14] | 23,221 | 20,500 | 19,473 | 3,144 | 20,939 |
| W19 | 1.17 [1.16-1.19] | 1.19 [1.17-1.21] | 1.19 [1.17-1.20] | 1.04 [1.00-1.09] | 1.13 [1.12-1.15] | 1.14 [1.12-1.16] | 1.12 [1.10-1.14] | 1.12 [1.10-1.14] | 1.03 [0.98-1.08] | 1.12 [1.10-1.14] | 51,848 | 47,686 | 45,247 | 5,023 | 46,001 |
| W20 | 1.03 [0.95-1.11] | 1.08 [0.97-1.19] | 1.13 [1.00-1.28] | 0.99 [0.88-1.11] | 0.99 [0.90-1.08] | 0.99 [0.91-1.07] | 1.01 [0.91-1.12] | 1.04 [0.92-1.19] | 0.96 [0.85-1.08] | 0.97 [0.89-1.07] | 1,439 | 883 | 576 | 699 | 1,132 |
| W21 | 1.11 [1.03-1.21] | 1.11 [0.97-1.27] | 0.94 [0.73-1.22] | 1.12 [1.02-1.23] | 1.04 [0.95-1.15] | 1.08 [0.99-1.18] | 1.08 [0.94-1.24] | 0.88 [0.68-1.15] | 1.10 [1.00-1.21] | 1.02 [0.92-1.13] | 1,513 | 494 | 131 | 1,206 | 1,094 |
| W22 | 1.12 [1.08-1.16] | 1.17 [1.12-1.23] | 1.20 [1.13-1.27] | 1.05 [1.00-1.10] | 1.07 [1.03-1.12] | 1.08 [1.04-1.12] | 1.08 [1.03-1.13] | 1.09 [1.03-1.16] | 1.03 [0.98-1.09] | 1.05 [1.01-1.09] | 7,690 | 4,615 | 3,190 | 3,846 | 6,511 |
| W23 | 1.02 [0.98-1.07] | 1.08 [1.01-1.16] | 1.14 [1.04-1.25] | 1.00 [0.95-1.05] | 0.97 [0.92-1.02] | 1.00 [0.96-1.05] | 1.03 [0.96-1.11] | 1.10 [1.01-1.20] | 0.99 [0.94-1.05] | 0.96 [0.92-1.01] | 4,806 | 1,867 | 1,202 | 3,288 | 3,875 |
| W25 | 1.02 [0.96-1.09] | 1.08 [1.00-1.17] | 1.16 [1.02-1.32] | 0.95 [0.87-1.03] | 0.98 [0.91-1.05] | 1.00 [0.94-1.07] | 1.02 [0.94-1.10] | 1.06 [0.93-1.22] | 0.94 [0.86-1.02] | 0.97 [0.90-1.05] | 2,334 | 1,509 | 529 | 1,307 | 1,703 |
| W26 | 1.06 [0.99-1.13] | 1.12 [1.04-1.20] | 1.10 [0.98-1.23] | 1.04 [0.93-1.15] | 0.99 [0.92-1.07] | 1.02 [0.95-1.09] | 1.03 [0.96-1.12] | 1.02 [0.91-1.14] | 1.03 [0.93-1.14] | 0.98 [0.91-1.06] | 2,119 | 1,614 | 709 | 977 | 1,663 |
| W27 | 0.99 [0.89-1.10] | 1.03 [0.92-1.16] | 1.08 [0.93-1.25] | 0.89 [0.74-1.06] | 0.97 [0.87-1.09] | 0.95 [0.86-1.06] | 0.98 [0.87-1.10] | 1.01 [0.87-1.18] | 0.89 [0.74-1.06] | 0.96 [0.85-1.08] | 881 | 690 | 391 | 300 | 681 |
| W28 | 0.85 [0.63-1.13] | | | | | 0.83 [0.62-1.11] | | | | | 102 | NA | NA | NA | NA |
| W29 | 0.99 [0.89-1.09] | 0.98 [0.88-1.08] | 0.99 [0.87-1.13] | 0.98 [0.79-1.21] | 0.90 [0.80-1.01] | 0.98 [0.88-1.08] | 0.95 [0.85-1.05] | 0.96 [0.84-1.10] | 0.98 [0.79-1.23] | 0.91 [0.81-1.02] | 893 | 814 | 516 | 215 | 651 |
| W31 | 1.02 [0.92-1.13] | 1.03 [0.92-1.14] | 0.95 [0.82-1.09] | 1.17 [0.96-1.42] | 0.93 [0.82-1.05] | 1.02 [0.92-1.13] | 1.02 [0.91-1.14] | 0.94 [0.82-1.08] | 1.16 [0.95-1.43] | 0.92 [0.81-1.04] | 840 | 721 | 444 | 262 | 587 |
| W34 | 1.06 [0.83-1.35] | | | 1.04 [0.77-1.41] | | 1.03 [0.80-1.33] | | | 1.04 [0.76-1.43] | | 153 | NA | NA | 104 | NA |
| W44 | 1.10 [1.06-1.15] | 1.30 [1.21-1.40] | 1.30 [1.19-1.41] | 1.02 [0.97-1.08] | 1.05 [1.00-1.10] | 1.06 [1.02-1.11] | 1.18 [1.09-1.27] | 1.18 [1.08-1.28] | 1.01 [0.96-1.06] | 1.02 [0.97-1.06] | 5,499 | 1,986 | 1,443 | 3,746 | 4,587 |
| W45 | 1.07 [0.99-1.15] | 1.13 [1.03-1.25] | 1.08 [0.95-1.23] | 1.06 [0.96-1.18] | 1.01 [0.93-1.10] | 1.03 [0.96-1.12] | 1.05 [0.95-1.16] | 0.99 [0.87-1.13] | 1.05 [0.94-1.16] | 0.99 [0.90-1.07] | 1,720 | 944 | 526 | 961 | 1,327 |
| W46 | 1.08 [0.86-1.34] | 1.17 [0.93-1.46] | 1.32 [0.99-1.75] | | 0.86 [0.69-1.09] | 0.99 [0.78-1.24] | 0.95 [0.75-1.21] | 1.05 [0.78-1.41] | | 0.85 [0.67-1.08] | 191 | 179 | 116 | NA | 168 |
| W49 | 1.19 [1.03-1.38] | 1.29 [1.08-1.54] | 1.32 [1.05-1.66] | 1.01 [0.81-1.26] | 1.20 [1.02-1.42] | 1.18 [1.02-1.37] | 1.22 [1.01-1.47] | 1.25 [0.99-1.58] | 1.01 [0.81-1.26] | 1.18 [1.00-1.39] | 450 | 287 | 180 | 197 | 361 |
| W50 | 1.11 [1.05-1.18] | 1.17 [1.07-1.27] | 1.23 [1.03-1.45] | 1.08 [1.00-1.16] | 1.06 [0.99-1.14] | 1.08 [1.01-1.15] | 1.08 [0.99-1.18] | 1.11 [0.93-1.33] | 1.06 [0.98-1.14] | 1.05 [0.98-1.13] | 2,632 | 1,264 | 312 | 1,914 | 1,958 |
| W51 | 1.06 [0.99-1.14] | 1.11 [1.00-1.24] | 1.10 [0.89-1.35] | 1.04 [0.96-1.13] | 1.05 [0.97-1.13] | 1.03 [0.96-1.10] | 1.05 [0.94-1.17] | 1.06 [0.85-1.31] | 1.01 [0.93-1.10] | 1.02 [0.94-1.11] | 1,993 | 764 | 216 | 1,541 | 1,526 |
| W54 | 0.98 [0.92-1.05] | 1.04 [0.97-1.13] | 1.13 [1.03-1.24] | 0.88 [0.80-0.97] | 0.93 [0.86-0.99] | 0.95 [0.89-1.01] | 0.97 [0.90-1.05] | 1.04 [0.95-1.15] | 0.86 [0.78-0.95] | 0.91 [0.85-0.98] | 2,316 | 1,462 | 1,040 | 1,062 | 1,865 |
| W55 | 0.98 [0.89-1.08] | 1.00 [0.90-1.11] | 0.97 [0.86-1.09] | 0.99 [0.81-1.20] | 0.92 [0.83-1.03] | 0.92 [0.83-1.02] | 0.91 [0.81-1.01] | 0.88 [0.78-1.00] | 0.98 [0.80-1.20] | 0.88 [0.79-0.98] | 968 | 800 | 601 | 255 | 781 |
| W57 | 1.40 [1.29-1.52] | 1.45 [1.32-1.59] | 1.42 [1.27-1.58] | 1.26 [1.09-1.45] | 1.29 [1.19-1.41] | 1.31 [1.21-1.43] | 1.31 [1.19-1.44] | 1.28 [1.14-1.43] | 1.24 [1.08-1.44] | 1.24 [1.13-1.36] | 1,567 | 1,194 | 788 | 533 | 1,301 |
| W60 | 1.35 [1.04-1.76] | 1.47 [1.10-1.96] | 1.46 [1.07-1.99] | | 1.29 [0.97-1.71] | 1.27 [0.97-1.67] | 1.37 [1.02-1.84] | 1.38 [1.01-1.89] | | 1.24 [0.93-1.66] | 144 | 121 | 102 | NA | 121 |
| W64 | 1.04 [0.86-1.26] | 1.10 [0.89-1.36] | 1.13 [0.90-1.43] | | 1.11 [0.91-1.36] | 1.03 [0.85-1.25] | 1.02 [0.82-1.26] | 1.06 [0.84-1.35] | | 1.11 [0.91-1.37] | 276 | 220 | 181 | NA | 241 |
| W78 | 1.10 [0.97-1.23] | 1.11 [0.98-1.25] | 1.10 [0.97-1.25] | | 1.05 [0.93-1.19] | 1.07 [0.95-1.21] | 1.06 [0.94-1.20] | 1.05 [0.92-1.19] | | 1.06 [0.94-1.20] | 744 | 724 | 692 | NA | 703 |
| W79 | 1.27 [1.16-1.39] | 1.26 [1.14-1.39] | 1.19 [1.08-1.32] | 1.96 [1.56-2.47] | 1.22 [1.11-1.34] | 1.21 [1.10-1.32] | 1.17 [1.06-1.29] | 1.13 [1.02-1.25] | 1.68 [1.32-2.14] | 1.18 [1.07-1.30] | 1,299 | 1,114 | 994 | 227 | 1,180 |
| W80 | 1.18 [1.02-1.37] | 1.23 [1.05-1.44] | 1.27 [1.08-1.50] | | 1.10 [0.95-1.29] | 1.12 [0.96-1.30] | 1.12 [0.95-1.31] | 1.15 [0.98-1.36] | | 1.06 [0.91-1.24] | 484 | 437 | 415 | NA | 446 |
| W84 | 1.06 [0.90-1.24] | 1.06 [0.91-1.24] | 1.08 [0.92-1.27] | | 1.02 [0.87-1.20] | 1.03 [0.88-1.21] | 0.99 [0.85-1.17] | 1.03 [0.87-1.21] | | 1.01 [0.86-1.19] | 431 | 418 | 408 | NA | 406 |
| X04 | 1.02 [0.78-1.33] | | | | | 0.99 [0.75-1.29] | | | | | 132 | NA | NA | NA | NA |
| X08 | 1.04 [0.79-1.37] | | | | 0.98 [0.74-1.31] | 1.04 [0.79-1.37] | | | | 0.96 [0.72-1.29] | 128 | NA | NA | NA | 111 |
| X09 | 1.01 [0.80-1.27] | 1.02 [0.78-1.34] | 1.11 [0.82-1.51] | | 0.96 [0.75-1.24] | 0.98 [0.78-1.25] | 0.98 [0.75-1.29] | 1.05 [0.77-1.43] | | 0.94 [0.73-1.21] | 177 | 131 | 101 | NA | 151 |
| X10 | 1.00 [0.91-1.11] | 1.19 [0.99-1.43] | 1.19 [0.95-1.50] | 0.98 [0.87-1.10] | 0.99 [0.89-1.11] | 0.99 [0.89-1.10] | 1.09 [0.90-1.32] | 1.10 [0.88-1.39] | 0.98 [0.87-1.10] | 0.99 [0.89-1.11] | 921 | 278 | 193 | 691 | 814 |
| X11 | 0.97 [0.81-1.17] | | | 1.01 [0.81-1.26] | 0.95 [0.77-1.17] | 0.94 [0.78-1.14] | | | 0.99 [0.79-1.24] | 0.93 [0.75-1.15] | 273 | NA | NA | 190 | 222 |
| X12 | 0.94 [0.82-1.08] | 1.01 [0.82-1.24] | 1.09 [0.86-1.40] | 0.87 [0.74-1.03] | 0.92 [0.80-1.06] | 0.92 [0.81-1.06] | 0.93 [0.75-1.15] | 0.99 [0.77-1.28] | 0.87 [0.73-1.03] | 0.91 [0.78-1.05] | 529 | 218 | 162 | 330 | 445 |
| X15 | 1.04 [0.89-1.22] | | | 0.99 [0.83-1.17] | 1.00 [0.85-1.17] | 1.03 [0.88-1.21] | | | 0.99 [0.83-1.17] | 1.00 [0.85-1.18] | 394 | NA | NA | 323 | 357 |
| X16 | 0.99 [0.82-1.21] | 1.14 [0.89-1.45] | 1.04 [0.80-1.34] | | 0.95 [0.77-1.17] | 0.96 [0.79-1.17] | 1.05 [0.82-1.35] | 0.98 [0.75-1.27] | | 0.95 [0.77-1.18] | 255 | 173 | 146 | NA | 222 |
| X19 | 1.12 [0.89-1.41] | | | | 1.02 [0.80-1.31] | 1.07 [0.85-1.36] | | | | 0.99 [0.77-1.28] | 178 | NA | NA | NA | 152 |
| X23 | 1.17 [0.97-1.41] | 1.19 [0.97-1.47] | 1.13 [0.89-1.42] | | 1.24 [1.01-1.52] | 1.09 [0.90-1.32] | 1.11 [0.89-1.37] | 1.04 [0.82-1.31] | | 1.18 [0.96-1.46] | 280 | 217 | 169 | NA | 226 |
| X31 | 1.05 [0.85-1.29] | 1.00 [0.81-1.24] | 1.00 [0.80-1.23] | | 0.97 [0.78-1.21] | 1.06 [0.86-1.31] | 0.99 [0.80-1.23] | 0.99 [0.80-1.24] | | 1.01 [0.80-1.26] | 244 | 233 | 221 | NA | 210 |
| X40 | 1.07 [1.02-1.13] | 1.16 [1.08-1.24] | 1.26 [1.14-1.39] | 1.00 [0.94-1.08] | 1.02 [0.96-1.08] | 1.03 [0.97-1.09] | 1.01 [0.94-1.08] | 1.07 [0.96-1.18] | 1.00 [0.93-1.08] | 1.00 [0.94-1.06] | 3,316 | 1,892 | 979 | 1,855 | 2,782 |
| X41 | 1.15 [1.08-1.24] | 1.26 [1.17-1.37] | 1.29 [1.16-1.43] | 0.98 [0.88-1.10] | 1.06 [0.99-1.14] | 1.08 [1.01-1.16] | 1.06 [0.97-1.15] | 1.08 [0.96-1.20] | 0.99 [0.88-1.11] | 1.05 [0.97-1.13] | 1,950 | 1,468 | 844 | 752 | 1,690 |
| X42 | 1.22 [1.13-1.31] | 1.28 [1.18-1.38] | 1.38 [1.25-1.52] | 0.98 [0.85-1.14] | 1.13 [1.05-1.22] | 1.11 [1.03-1.20] | 1.04 [0.96-1.13] | 1.11 [1.00-1.23] | 0.98 [0.84-1.14] | 1.10 [1.02-1.19] | 1,798 | 1,604 | 1,050 | 440 | 1,636 |
| X43 | 1.10 [0.94-1.29] | 1.20 [0.98-1.46] | 1.16 [0.93-1.45] | 0.95 [0.75-1.22] | 1.02 [0.86-1.21] | 1.05 [0.90-1.24] | 1.08 [0.88-1.32] | 1.03 [0.82-1.29] | 0.92 [0.71-1.18] | 1.02 [0.86-1.20] | 395 | 259 | 210 | 155 | 344 |
| X44 | 1.17 [1.10-1.24] | 1.29 [1.20-1.39] | 1.30 [1.19-1.42] | 1.03 [0.94-1.12] | 1.09 [1.03-1.16] | 1.09 [1.03-1.16] | 1.10 [1.02-1.19] | 1.11 [1.01-1.21] | 1.01 [0.92-1.10] | 1.06 [1.00-1.13] | 2,841 | 1,751 | 1,372 | 1,261 | 2,516 |
| X45 | 1.12 [1.00-1.26] | 1.16 [1.03-1.31] | 1.23 [1.02-1.50] | 0.92 [0.76-1.11] | 1.01 [0.89-1.14] | 1.07 [0.95-1.21] | 0.99 [0.87-1.13] | 1.00 [0.81-1.22] | 0.94 [0.78-1.14] | 1.00 [0.88-1.14] | 720 | 595 | 243 | 276 | 588 |
| X46 | 0.91 [0.73-1.12] | | | 0.83 [0.66-1.04] | 0.91 [0.72-1.14] | 0.88 [0.71-1.09] | | | 0.82 [0.65-1.04] | 0.90 [0.71-1.12] | 200 | NA | NA | 158 | 174 |
| X47 | 1.11 [0.92-1.34] | 1.10 [0.89-1.37] | 1.17 [0.91-1.51] | | 1.08 [0.88-1.33] | 1.08 [0.89-1.31] | 1.00 [0.80-1.25] | 1.12 [0.86-1.45] | | 1.06 [0.86-1.31] | 265 | 197 | 148 | NA | 228 |
| X49 | 1.35 [1.24-1.47] | 1.38 [1.20-1.58] | 1.30 [1.09-1.55] | 1.37 [1.23-1.52] | 1.31 [1.19-1.44] | 1.27 [1.16-1.38] | 1.20 [1.04-1.38] | 1.17 [0.98-1.40] | 1.28 [1.14-1.43] | 1.24 [1.12-1.36] | 1,326 | 533 | 310 | 892 | 1,128 |
| X50 | 1.11 [1.06-1.17] | 1.15 [1.09-1.21] | 1.15 [1.08-1.24] | 1.05 [0.96-1.15] | 1.05 [0.99-1.11] | 1.05 [1.00-1.10] | 1.05 [1.00-1.12] | 1.06 [0.99-1.14] | 1.01 [0.92-1.11] | 1.01 [0.95-1.07] | 3,812 | 3,084 | 1,998 | 1,224 | 3,066 |
| X51 | 1.19 [0.98-1.44] | 1.14 [0.94-1.39] | 1.16 [0.93-1.45] | | 1.06 [0.86-1.32] | 1.15 [0.95-1.40] | 1.10 [0.90-1.34] | 1.10 [0.88-1.38] | | 1.04 [0.84-1.30] | 254 | 237 | 186 | NA | 197 |
| X58 | 1.60 [1.47-1.75] | 1.45 [1.31-1.61] | 1.39 [1.23-1.56] | 2.06 [1.78-2.38] | 1.50 [1.36-1.64] | 1.44 [1.32-1.58] | 1.28 [1.15-1.43] | 1.26 [1.11-1.42] | 1.79 [1.54-2.08] | 1.38 [1.26-1.52] | 1,466 | 973 | 729 | 602 | 1,302 |
| X59 | 1.19 [1.16-1.22] | 1.24 [1.20-1.27] | 1.26 [1.22-1.30] | 1.08 [1.03-1.12] | 1.14 [1.11-1.17] | 1.14 [1.11-1.17] | 1.13 [1.10-1.17] | 1.16 [1.12-1.20] | 1.05 [1.00-1.09] | 1.11 [1.08-1.14] | 16,982 | 12,513 | 10,137 | 5,445 | 14,181 |
| X60 | 1.11 [1.08-1.15] | 1.13 [1.09-1.17] | 1.29 [1.21-1.36] | 1.02 [0.98-1.07] | 1.03 [0.99-1.06] | 1.06 [1.02-1.09] | 0.98 [0.95-1.02] | 1.09 [1.03-1.16] | 1.01 [0.97-1.06] | 1.01 [0.98-1.05] | 10,841 | 7,074 | 2,711 | 5,994 | 8,352 |
| X61 | 1.16 [1.12-1.21] | 1.21 [1.16-1.25] | 1.32 [1.25-1.40] | 0.97 [0.92-1.04] | 1.05 [1.01-1.09] | 1.08 [1.04-1.13] | 1.00 [0.96-1.05] | 1.08 [1.02-1.15] | 0.98 [0.92-1.05] | 1.04 [1.00-1.08] | 7,448 | 6,319 | 2,718 | 2,707 | 6,147 |
| X62 | 1.16 [1.10-1.22] | 1.19 [1.13-1.25] | 1.29 [1.20-1.40] | 0.98 [0.90-1.06] | 1.07 [1.01-1.12] | 1.05 [1.00-1.11] | 0.97 [0.92-1.02] | 1.02 [0.94-1.11] | 0.98 [0.91-1.07] | 1.04 [0.98-1.09] | 4,194 | 3,494 | 1,503 | 1,589 | 3,638 |
| X63 | 1.17 [1.06-1.28] | 1.21 [1.09-1.34] | 1.26 [1.08-1.48] | 1.05 [0.90-1.23] | 1.06 [0.95-1.17] | 1.10 [1.00-1.21] | 1.01 [0.90-1.12] | 1.01 [0.85-1.19] | 1.06 [0.90-1.24] | 1.05 [0.95-1.17] | 1,055 | 846 | 366 | 422 | 911 |
| X64 | 1.20 [1.14-1.26] | 1.23 [1.16-1.30] | 1.37 [1.26-1.48] | 1.05 [0.98-1.14] | 1.09 [1.03-1.15] | 1.10 [1.05-1.16] | 1.02 [0.96-1.09] | 1.10 [1.01-1.20] | 1.03 [0.95-1.11] | 1.07 [1.01-1.13] | 3,886 | 2,796 | 1,307 | 1,811 | 3,242 |
| X65 | 1.15 [1.10-1.21] | 1.16 [1.11-1.22] | 1.28 [1.20-1.37] | 0.99 [0.92-1.08] | 1.08 [1.03-1.13] | 1.08 [1.03-1.13] | 0.98 [0.93-1.02] | 1.04 [0.97-1.12] | 1.02 [0.94-1.11] | 1.07 [1.02-1.13] | 4,934 | 4,511 | 1,899 | 1,543 | 4,167 |
| X69 | 1.07 [0.91-1.26] | 0.94 [0.77-1.16] | | 1.03 [0.83-1.27] | 0.95 [0.79-1.13] | 1.02 [0.86-1.20] | 0.81 [0.65-1.00] | | 1.01 [0.81-1.26] | 0.94 [0.79-1.13] | 386 | 211 | NA | 227 | 312 |
| X70 | 0.97 [0.81-1.17] | 0.99 [0.79-1.24] | | 0.98 [0.75-1.27] | 0.88 [0.71-1.08] | 0.96 [0.79-1.17] | 0.87 [0.69-1.10] | | 1.04 [0.80-1.37] | 0.89 [0.72-1.09] | 269 | 174 | NA | 145 | 222 |
| X78 | 1.10 [1.03-1.17] | 1.09 [1.00-1.17] | 1.30 [1.14-1.48] | 1.03 [0.95-1.11] | 1.01 [0.95-1.09] | 1.02 [0.96-1.09] | 0.90 [0.82-0.97] | 1.05 [0.91-1.20] | 1.01 [0.93-1.09] | 0.99 [0.92-1.06] | 2,640 | 1,451 | 548 | 1,650 | 2,162 |

#### Supplementary Table 3: Hazard ratios and events from all cohorts

| | Hazard ratio (99% confidence interval) | | | | | | | | | | | | | | |
|---|---|---|---|---|---|---|---|---|---|---|---|---|---|---|---|
| | crude | | | | | adjusted | | | | | Events (in exposed) | | | | |
| Outcome | any age | 18+ | 40+ | <18 | hosp. | any age | 18+ | 40+ | <18 | hosp. | any age | 18+ | 40+ | <18 | hosp. |
| X79 | 1.23 [0.90-1.67] | | | | | 1.08 [0.78-1.50] | | | | | 109 | NA | NA | NA | NA |
| X83 | 1.03 [0.83-1.28] | 1.13 [0.88-1.47] | | 0.87 [0.65-1.17] | 0.85 [0.67-1.07] | 0.95 [0.76-1.18] | 0.97 [0.74-1.27] | | 0.84 [0.62-1.13] | 0.82 [0.65-1.04] | 219 | 138 | NA | 119 | 187 |
| X84 | 1.04 [0.87-1.24] | 1.04 [0.84-1.28] | | 0.95 [0.75-1.22] | 0.97 [0.80-1.17] | 0.94 [0.78-1.13] | 0.81 [0.65-1.02] | | 0.95 [0.74-1.22] | 0.91 [0.75-1.10] | 316 | 206 | NA | 174 | 277 |
| X99 | 1.06 [0.96-1.16] | 1.09 [0.98-1.21] | 1.17 [0.93-1.47] | 1.02 [0.90-1.16] | 0.96 [0.85-1.07] | 1.04 [0.94-1.14] | 1.02 [0.92-1.14] | 1.03 [0.81-1.31] | 1.02 [0.90-1.17] | 0.97 [0.87-1.10] | 1,053 | 826 | 168 | 620 | 720 |
| Y00 | 1.08 [0.94-1.23] | 1.12 [0.97-1.29] | 1.25 [0.99-1.57] | 0.91 [0.73-1.13] | 0.95 [0.81-1.11] | 1.03 [0.89-1.18] | 0.99 [0.85-1.15] | 1.06 [0.83-1.35] | 0.89 [0.71-1.11] | 0.97 [0.83-1.14] | 520 | 450 | 167 | 201 | 395 |
| Y04 | 1.07 [1.02-1.12] | 1.12 [1.07-1.18] | 1.23 [1.12-1.35] | 1.00 [0.94-1.07] | 0.97 [0.92-1.03] | 1.03 [0.98-1.08] | 1.02 [0.97-1.08] | 1.06 [0.96-1.17] | 0.99 [0.93-1.06] | 0.97 [0.92-1.03] | 4,568 | 3,655 | 1,001 | 2,300 | 3,215 |
| Y07 | 0.92 [0.77-1.09] | 1.01 [0.78-1.30] | | 0.92 [0.74-1.15] | 0.88 [0.73-1.05] | 0.88 [0.74-1.05] | 0.82 [0.63-1.08] | | 0.92 [0.74-1.15] | 0.87 [0.72-1.04] | 317 | 141 | NA | 199 | 279 |
| Y08 | 0.92 [0.74-1.15] | 1.00 [0.79-1.26] | | | 0.86 [0.67-1.11] | 0.90 [0.71-1.12] | 0.93 [0.73-1.19] | | | 0.89 [0.68-1.16] | 188 | 165 | NA | NA | 141 |
| Y09 | 1.09 [0.96-1.23] | 1.13 [1.00-1.29] | 1.27 [1.02-1.58] | 1.08 [0.89-1.32] | 1.02 [0.88-1.17] | 1.05 [0.92-1.19] | 1.02 [0.89-1.16] | 1.09 [0.87-1.37] | 1.06 [0.86-1.30] | 1.02 [0.88-1.17] | 611 | 542 | 185 | 245 | 464 |
| Y28 | 1.04 [0.81-1.33] | | | | 0.93 [0.71-1.24] | 0.96 [0.75-1.24] | | | | 0.87 [0.65-1.16] | 160 | NA | NA | NA | 115 |
| Y34 | 1.12 [0.86-1.44] | | | | 1.02 [0.77-1.37] | 1.03 [0.79-1.34] | | | | 0.99 [0.74-1.34] | 146 | NA | NA | NA | 115 |
| Y40 | 1.51 [1.44-1.58] | 1.52 [1.45-1.60] | 1.53 [1.45-1.61] | 1.30 [1.15-1.47] | 1.40 [1.33-1.47] | 1.39 [1.32-1.45] | 1.34 [1.27-1.41] | 1.36 [1.29-1.43] | 1.25 [1.10-1.41] | 1.32 [1.26-1.39] | 4,982 | 4,461 | 3,875 | 743 | 4,487 |
| Y41 | 1.39 [1.26-1.55] | 1.45 [1.30-1.62] | 1.45 [1.29-1.62] | 1.17 [0.85-1.62] | 1.29 [1.16-1.44] | 1.26 [1.14-1.41] | 1.27 [1.14-1.42] | 1.26 [1.12-1.43] | 1.09 [0.78-1.54] | 1.21 [1.08-1.35] | 981 | 919 | 783 | 106 | 888 |
| Y42 | 1.45 [1.38-1.53] | 1.43 [1.36-1.51] | 1.40 [1.32-1.48] | 1.36 [1.14-1.63] | 1.33 [1.26-1.40] | 1.29 [1.22-1.36] | 1.20 [1.14-1.27] | 1.18 [1.11-1.25] | 1.30 [1.08-1.57] | 1.22 [1.15-1.28] | 3,793 | 3,579 | 3,180 | 335 | 3,504 |
| Y43 | 1.22 [1.17-1.26] | 1.23 [1.18-1.28] | 1.21 [1.16-1.26] | 1.11 [0.99-1.24] | 1.15 [1.11-1.19] | 1.18 [1.14-1.23] | 1.18 [1.13-1.22] | 1.16 [1.11-1.21] | 1.08 [0.96-1.22] | 1.14 [1.10-1.19] | 7,081 | 6,538 | 5,898 | 741 | 6,207 |
| Y44 | 1.27 [1.18-1.35] | 1.26 [1.18-1.34] | 1.24 [1.16-1.33] | 1.40 [1.03-1.92] | 1.19 [1.11-1.27] | 1.20 [1.13-1.29] | 1.16 [1.08-1.24] | 1.14 [1.06-1.22] | 1.38 [1.00-1.90] | 1.15 [1.07-1.23] | 2,396 | 2,327 | 2,205 | 114 | 2,249 |
| Y45 | 1.36 [1.31-1.42] | 1.35 [1.30-1.41] | 1.34 [1.28-1.40] | 1.29 [1.10-1.52] | 1.25 [1.19-1.30] | 1.26 [1.21-1.32] | 1.20 [1.15-1.25] | 1.20 [1.14-1.25] | 1.22 [1.03-1.45] | 1.19 [1.14-1.25] | 5,815 | 5,643 | 5,011 | 392 | 5,308 |
| Y46 | 1.40 [1.25-1.56] | 1.40 [1.25-1.57] | 1.35 [1.20-1.52] | 1.20 [0.88-1.65] | 1.23 [1.11-1.38] | 1.28 [1.14-1.43] | 1.25 [1.11-1.40] | 1.20 [1.07-1.36] | 1.16 [0.84-1.60] | 1.19 [1.06-1.33] | 870 | 793 | 722 | 103 | 812 |
| Y47 | 1.27 [1.09-1.49] | 1.37 [1.16-1.62] | 1.40 [1.17-1.67] | | 1.17 [0.99-1.37] | 1.17 [1.00-1.38] | 1.22 [1.03-1.45] | 1.26 [1.05-1.51] | | 1.13 [0.96-1.33] | 419 | 376 | 331 | NA | 380 |
| Y48 | 1.32 [1.14-1.52] | 1.32 [1.13-1.54] | 1.28 [1.08-1.52] | | 1.20 [1.04-1.39] | 1.20 [1.04-1.39] | 1.16 [0.99-1.35] | 1.13 [0.94-1.35] | | 1.14 [0.98-1.32] | 499 | 429 | 328 | NA | 443 |
| Y49 | 1.35 [1.24-1.47] | 1.35 [1.24-1.47] | 1.35 [1.23-1.48] | | 1.28 [1.18-1.40] | 1.29 [1.18-1.41] | 1.23 [1.12-1.34] | 1.24 [1.13-1.36] | | 1.27 [1.16-1.39] | 1,408 | 1,363 | 1,208 | NA | 1,292 |
| Y51 | 1.22 [1.13-1.31] | 1.27 [1.18-1.36] | 1.24 [1.16-1.34] | | 1.19 [1.10-1.28] | 1.18 [1.10-1.27] | 1.18 [1.10-1.27] | 1.16 [1.08-1.25] | | 1.18 [1.10-1.27] | 1,998 | 1,976 | 1,919 | NA | 1,853 |
| Y52 | 1.32 [1.26-1.39] | 1.34 [1.28-1.40] | 1.30 [1.24-1.36] | | 1.24 [1.18-1.30] | 1.26 [1.20-1.32] | 1.21 [1.16-1.27] | 1.19 [1.13-1.24] | | 1.21 [1.15-1.27] | 5,017 | 5,007 | 4,955 | NA | 4,688 |
| Y53 | 1.38 [1.25-1.53] | 1.39 [1.25-1.54] | 1.37 [1.23-1.53] | | 1.31 [1.18-1.46] | 1.25 [1.13-1.39] | 1.19 [1.07-1.32] | 1.20 [1.07-1.34] | | 1.22 [1.10-1.36] | 1,024 | 992 | 928 | NA | 967 |
| Y54 | 1.35 [1.28-1.42] | 1.37 [1.30-1.44] | 1.36 [1.29-1.43] | | 1.29 [1.22-1.36] | 1.27 [1.21-1.34] | 1.23 [1.17-1.30] | 1.23 [1.17-1.30] | | 1.24 [1.17-1.30] | 4,092 | 4,067 | 4,022 | NA | 3,764 |
| Y55 | 1.46 [1.28-1.66] | 1.43 [1.25-1.63] | 1.55 [1.27-1.90] | 1.31 [1.05-1.64] | 1.32 [1.15-1.51] | 1.28 [1.12-1.46] | 1.20 [1.04-1.39] | 1.24 [1.00-1.53] | 1.22 [0.96-1.55] | 1.16 [1.01-1.34] | 644 | 559 | 245 | 217 | 541 |
| Y56 | 1.80 [1.57-2.06] | 1.87 [1.62-2.17] | 1.88 [1.60-2.21] | | 1.65 [1.43-1.90] | 1.62 [1.41-1.87] | 1.63 [1.40-1.89] | 1.61 [1.37-1.91] | | 1.51 [1.31-1.75] | 594 | 516 | 423 | NA | 520 |
| Y57 | 1.43 [1.34-1.53] | 1.38 [1.29-1.47] | 1.39 [1.29-1.49] | 1.84 [1.44-2.33] | 1.33 [1.24-1.42] | 1.31 [1.22-1.40] | 1.21 [1.13-1.30] | 1.22 [1.13-1.31] | 1.65 [1.28-2.13] | 1.25 [1.17-1.34] | 2,451 | 2,304 | 2,107 | 210 | 2,276 |
| Y59 | 1.52 [1.28-1.81] | 1.55 [1.25-1.92] | 1.44 [1.13-1.83] | 1.62 [1.23-2.13] | 1.41 [1.18-1.69] | 1.40 [1.17-1.68] | 1.38 [1.11-1.72] | 1.29 [1.01-1.65] | 1.50 [1.13-1.99] | 1.30 [1.08-1.57] | 348 | 225 | 173 | 150 | 320 |
| Y60 | 1.24 [1.17-1.32] | 1.27 [1.20-1.35] | 1.25 [1.17-1.33] | 1.10 [0.87-1.39] | 1.17 [1.10-1.24] | 1.18 [1.11-1.25] | 1.17 [1.10-1.25] | 1.16 [1.09-1.24] | 1.07 [0.85-1.36] | 1.13 [1.06-1.21] | 2,782 | 2,690 | 2,394 | 198 | 2,495 |
| Y65 | 1.20 [1.02-1.41] | 1.24 [1.05-1.47] | 1.21 [1.01-1.45] | | 1.10 [0.93-1.30] | 1.14 [0.97-1.34] | 1.13 [0.95-1.35] | 1.10 [0.92-1.33] | | 1.08 [0.91-1.28] | 377 | 338 | 293 | NA | 335 |
| Y71 | 1.59 [1.27-1.99] | 1.59 [1.26-2.02] | 1.56 [1.22-2.00] | | 1.38 [1.09-1.74] | 1.51 [1.20-1.90] | 1.42 [1.11-1.82] | 1.43 [1.11-1.84] | | 1.32 [1.04-1.67] | 200 | 181 | 168 | NA | 181 |
| Y73 | 0.97 [0.75-1.25] | 0.94 [0.72-1.23] | 0.99 [0.75-1.30] | | 0.92 [0.70-1.21] | 0.93 [0.71-1.21] | 0.89 [0.67-1.17] | 0.90 [0.68-1.20] | | 0.89 [0.67-1.18] | 143 | 132 | 120 | NA | 126 |
| Y76 | 1.35 [1.00-1.82] | 1.37 [1.01-1.85] | | | | 1.32 [0.97-1.79] | 1.26 [0.92-1.71] | | | | 109 | 110 | NA | NA | NA |
| Y77 | 1.01 [0.78-1.30] | 0.98 [0.77-1.26] | 0.99 [0.78-1.28] | | 1.05 [0.81-1.37] | 0.98 [0.76-1.27] | 0.91 [0.70-1.17] | 0.95 [0.74-1.22] | | 1.03 [0.79-1.35] | 154 | 149 | 147 | NA | 138 |
| Y79 | 1.20 [1.02-1.41] | 1.24 [1.06-1.46] | 1.19 [1.00-1.40] | | 1.15 [0.97-1.36] | 1.15 [0.97-1.35] | 1.16 [0.98-1.37] | 1.11 [0.94-1.32] | | 1.10 [0.93-1.31] | 383 | 366 | 338 | NA | 338 |
| Y82 | 1.31 [1.01-1.68] | 1.27 [0.97-1.68] | 1.31 [0.98-1.76] | | 1.20 [0.92-1.56] | 1.24 [0.96-1.61] | 1.16 [0.88-1.54] | 1.19 [0.88-1.60] | | 1.19 [0.91-1.55] | 149 | 129 | 113 | NA | 135 |
| Y83 | 1.28 [1.26-1.30] | 1.29 [1.27-1.31] | 1.27 [1.25-1.29] | 1.22 [1.18-1.27] | 1.18 [1.17-1.20] | 1.21 [1.19-1.22] | 1.17 [1.16-1.19] | 1.17 [1.15-1.18] | 1.17 [1.13-1.22] | 1.15 [1.13-1.16] | 53,081 | 47,693 | 40,066 | 7,973 | 46,779 |
| Y84 | 1.24 [1.21-1.28] | 1.25 [1.22-1.29] | 1.24 [1.20-1.28] | 1.21 [1.08-1.36] | 1.16 [1.13-1.20] | 1.19 [1.16-1.23] | 1.16 [1.12-1.19] | 1.14 [1.11-1.18] | 1.16 [1.03-1.31] | 1.14 [1.10-1.18] | 11,136 | 10,644 | 9,831 | 763 | 10,301 |
| Y85 | 1.30 [1.19-1.42] | 1.30 [1.19-1.42] | 1.38 [1.24-1.54] | 1.16 [0.93-1.44] | 1.18 [1.07-1.29] | 1.23 [1.12-1.34] | 1.15 [1.05-1.27] | 1.25 [1.12-1.40] | 1.16 [0.93-1.46] | 1.13 [1.02-1.24] | 1,249 | 1,152 | 803 | 217 | 1,067 |
| Y86 | 1.16 [1.09-1.24] | 1.20 [1.12-1.29] | 1.24 [1.14-1.35] | 0.99 [0.87-1.13] | 1.09 [1.02-1.17] | 1.11 [1.03-1.18] | 1.10 [1.02-1.18] | 1.13 [1.03-1.23] | 0.97 [0.85-1.11] | 1.06 [0.98-1.14] | 2,237 | 1,891 | 1,258 | 595 | 1,858 |
| Y87 | 1.23 [1.05-1.44] | 1.34 [1.14-1.57] | 1.39 [1.07-1.80] | 1.05 [0.78-1.40] | 1.16 [0.98-1.37] | 1.15 [0.97-1.35] | 1.18 [0.99-1.40] | 1.23 [0.94-1.61] | 0.98 [0.72-1.33] | 1.14 [0.96-1.36] | 387 | 353 | 139 | 115 | 331 |
| Y88 | 1.17 [1.00-1.37] | 1.18 [1.00-1.38] | 1.24 [1.04-1.47] | | 1.02 [0.87-1.20] | 1.07 [0.91-1.26] | 1.01 [0.85-1.20] | 1.06 [0.88-1.27] | | 0.97 [0.83-1.14] | 397 | 357 | 312 | NA | 373 |
| Y89 | 1.26 [0.96-1.64] | 1.31 [1.00-1.72] | | | 1.06 [0.80-1.40] | 1.21 [0.92-1.59] | 1.19 [0.90-1.57] | | | 1.06 [0.80-1.42] | 142 | 129 | NA | NA | 118 |
| Y90 | 1.13 [0.93-1.39] | 1.17 [0.94-1.44] | 1.19 [0.91-1.55] | | 0.98 [0.80-1.21] | 1.10 [0.90-1.36] | 1.07 [0.86-1.33] | 1.05 [0.80-1.38] | | 0.99 [0.80-1.23] | 228 | 201 | 127 | NA | 200 |
| Y91 | 1.15 [1.00-1.33] | 1.17 [1.01-1.35] | 1.27 [1.05-1.53] | | 1.10 [0.94-1.29] | 1.12 [0.96-1.29] | 1.07 [0.92-1.24] | 1.10 [0.91-1.34] | | 1.11 [0.95-1.31] | 460 | 441 | 265 | NA | 374 |
| Y95 | 1.25 [1.22-1.27] | 1.24 [1.21-1.27] | 1.24 [1.22-1.27] | 1.39 [1.12-1.73] | 1.20 [1.17-1.22] | 1.19 [1.16-1.21] | 1.14 [1.11-1.16] | 1.13 [1.11-1.16] | 1.43 [1.14-1.79] | 1.17 [1.14-1.19] | 25,680 | 25,635 | 25,150 | 241 | 24,404 |
| Factors influencing health status and contact with health services (Z00-Z99) | | | | | | | | | | | | | | | |
| Z00 | 1.19 [1.08-1.30] | 1.22 [1.11-1.34] | 1.25 [1.11-1.40] | 1.18 [0.95-1.46] | 1.10 [1.00-1.22] | 1.12 [1.02-1.23] | 1.13 [1.03-1.25] | 1.15 [1.02-1.29] | 1.17 [0.94-1.46] | 1.07 [0.97-1.19] | 1,116 | 978 | 701 | 218 | 893 |
| Z01 | 2.34 [2.27-2.42] | 1.41 [1.34-1.49] | 1.43 [1.34-1.53] | 3.12 [3.00-3.25] | 2.28 [2.20-2.36] | 1.96 [1.90-2.03] | 1.22 [1.15-1.29] | 1.26 [1.18-1.35] | 2.48 [2.38-2.59] | 1.90 [1.83-1.98] | 11,656 | 3,191 | 2,154 | 8,751 | 9,914 |
| Z02 | 1.16 [1.10-1.22] | 1.17 [1.11-1.23] | 1.16 [1.10-1.24] | 1.06 [0.89-1.25] | 1.10 [1.04-1.16] | 1.11 [1.05-1.17] | 1.07 [1.02-1.13] | 1.08 [1.01-1.14] | 1.04 [0.87-1.24] | 1.07 [1.01-1.13] | 3,845 | 3,798 | 2,860 | 338 | 3,365 |
| Z03 | 1.34 [1.32-1.36] | 1.32 [1.30-1.35] | 1.33 [1.30-1.36] | 1.38 [1.34-1.43] | 1.26 [1.23-1.28] | 1.25 [1.23-1.27] | 1.19 [1.16-1.21] | 1.19 [1.17-1.22] | 1.30 [1.25-1.34] | 1.20 [1.17-1.22] | 36,893 | 29,542 | 24,467 | 9,073 | 32,200 |
| Z04 | 1.22 [1.17-1.27] | 1.27 [1.21-1.33] | 1.32 [1.24-1.40] | 1.14 [1.07-1.21] | 1.12 [1.08-1.17] | 1.16 [1.11-1.21] | 1.16 [1.10-1.21] | 1.22 [1.15-1.29] | 1.11 [1.04-1.18] | 1.09 [1.04-1.14] | 6,480 | 4,365 | 2,846 | 2,567 | 5,152 |
| Z08 | 1.08 [1.05-1.12] | 1.08 [1.05-1.12] | 1.08 [1.05-1.12] | 0.98 [0.82-1.18] | 1.05 [1.01-1.09] | 1.07 [1.03-1.10] | 1.05 [1.02-1.09] | 1.05 [1.02-1.09] | 1.01 [0.84-1.22] | 1.05 [1.01-1.09] | 8,526 | 8,285 | 8,052 | 288 | 7,717 |
| Z09 | 1.28 [1.25-1.31] | 1.29 [1.26-1.32] | 1.29 [1.26-1.32] | 1.18 [1.11-1.26] | 1.18 [1.15-1.20] | 1.19 [1.17-1.22] | 1.16 [1.13-1.19] | 1.17 [1.14-1.20] | 1.14 [1.06-1.21] | 1.12 [1.10-1.15] | 19,415 | 17,514 | 15,298 | 2,487 | 17,468 |
| Z11 | 1.22 [1.20-1.23] | 1.21 [1.20-1.23] | 1.23 [1.21-1.25] | 1.17 [1.14-1.21] | 1.15 [1.13-1.17] | 1.15 [1.13-1.16] | 1.10 [1.08-1.11] | 1.10 [1.09-1.12] | 1.14 [1.11-1.17] | 1.12 [1.10-1.13] | 60,391 | 53,760 | 39,449 | 13,770 | 55,884 |
| Z12 | 1.16 [1.13-1.19] | 1.16 [1.13-1.19] | 1.15 [1.12-1.18] | 1.30 [0.98-1.74] | 1.10 [1.06-1.13] | 1.12 [1.09-1.15] | 1.13 [1.09-1.16] | 1.11 [1.08-1.15] | 1.23 [0.91-1.65] | 1.07 [1.04-1.11] | 12,500 | 12,458 | 11,973 | 125 | 10,207 |
| Z13 | 1.29 [1.27-1.31] | 1.26 [1.24-1.28] | 1.27 [1.24-1.29] | 1.37 [1.32-1.42] | 1.19 [1.17-1.21] | 1.22 [1.19-1.24] | 1.16 [1.14-1.18] | 1.15 [1.13-1.18] | 1.29 [1.24-1.33] | 1.14 [1.12-1.17] | 39,843 | 34,601 | 24,707 | 8,731 | 33,396 |
| Z20 | 1.25 [1.17-1.34] | 1.24 [1.15-1.34] | 1.25 [1.15-1.36] | 1.22 [1.06-1.42] | 1.16 [1.08-1.25] | 1.19 [1.10-1.28] | 1.13 [1.05-1.23] | 1.13 [1.04-1.24] | 1.19 [1.03-1.39] | 1.12 [1.04-1.21] | 2,183 | 1,817 | 1,522 | 495 | 2,010 |
| Z22 | 1.40 [1.36-1.43] | 1.41 [1.38-1.45] | 1.48 [1.43-1.53] | 1.13 [1.06-1.20] | 1.30 [1.27-1.33] | 1.32 [1.29-1.35] | 1.29 [1.26-1.33] | 1.34 [1.30-1.38] | 1.10 [1.03-1.17] | 1.26 [1.23-1.29] | 19,582 | 18,903 | 11,068 | 2,859 | 17,798 |
| Z24 | 1.07 [0.91-1.26] | 1.13 [0.95-1.35] | | 1.02 [0.80-1.31] | 1.07 [0.89-1.29] | 1.07 [0.90-1.27] | 1.12 [0.93-1.33] | | 0.98 [0.76-1.27] | 1.09 [0.90-1.32] | 353 | 301 | NA | 158 | 279 |
| Z25 | 2.82 [2.31-3.44] | | | 3.36 [2.69-4.21] | 2.63 [2.15-3.22] | 2.27 [1.84-2.80] | | | 2.52 [1.98-3.21] | 2.17 [1.75-2.69] | 375 | NA | NA | 324 | 349 |
| Z27 | 2.36 [2.17-2.57] | 1.06 [0.93-1.20] | | 3.25 [2.94-3.59] | 2.26 [2.06-2.48] | 2.10 [1.92-2.29] | 1.01 [0.89-1.15] | | 2.71 [2.44-3.01] | 2.04 [1.85-2.25] | 1,707 | 585 | NA | 1,371 | 1,355 |
| Z29 | 1.25 [1.21-1.29] | 1.25 [1.21-1.29] | 1.29 [1.23-1.34] | 1.17 [1.09-1.26] | 1.20 [1.16-1.24] | 1.19 [1.15-1.23] | 1.15 [1.11-1.19] | 1.17 [1.12-1.22] | 1.14 [1.06-1.23] | 1.16 [1.12-1.20] | 10,193 | 9,224 | 6,022 | 1,986 | 9,202 |
| Z30 | 1.16 [1.13-1.20] | 1.17 [1.14-1.21] | 1.20 [1.14-1.26] | 1.03 [0.94-1.13] | 1.06 [1.02-1.09] | 1.12 [1.09-1.15] | 1.10 [1.06-1.13] | 1.11 [1.05-1.17] | 1.03 [0.94-1.13] | 1.05 [1.01-1.08] | 10,748 | 10,535 | 3,391 | 1,118 | 8,192 |
| Z31 | 1.02 [0.81-1.28] | 1.06 [0.85-1.33] | | | 1.20 [0.91-1.57] | 0.97 [0.77-1.23] | 0.98 [0.77-1.23] | | | 1.15 [0.86-1.53] | 170 | 170 | NA | NA | 114 |
| Z32 | 1.23 [1.10-1.37] | 1.23 [1.10-1.37] | | 1.06 [0.86-1.31] | 1.09 [0.97-1.23] | 1.17 [1.05-1.31] | 1.13 [1.01-1.26] | | 1.07 [0.86-1.34] | 1.07 [0.95-1.21] | 827 | 786 | NA | 218 | 661 |
| Z33 | 1.23 [1.18-1.27] | 1.26 [1.22-1.31] | 1.19 [1.00-1.42] | 1.12 [1.05-1.20] | 1.12 [1.08-1.17] | 1.14 [1.10-1.19] | 1.12 [1.07-1.16] | 1.08 [0.90-1.29] | 1.10 [1.03-1.18] | 1.08 [1.04-1.12] | 7,874 | 7,664 | 280 | 2,540 | 6,613 |
| Z34 | 1.09 [1.07-1.11] | 1.12 [1.09-1.14] | 0.97 [0.85-1.09] | 0.99 [0.95-1.03] | 1.03 [1.00-1.05] | 1.06 [1.04-1.09] | 1.07 [1.04-1.09] | 0.96 [0.84-1.09] | 0.99 [0.95-1.03] | 1.02 [0.99-1.04] | 20,140 | 19,711 | 555 | 6,167 | 15,116 |
| Z35 | 1.07 [1.04-1.09] | 1.08 [1.05-1.11] | 0.96 [0.89-1.03] | 0.97 [0.92-1.02] | 1.00 [0.98-1.02] | 1.04 [1.02-1.07] | 1.05 [1.02-1.07] | 0.97 [0.90-1.05] | 0.96 [0.91-1.01] | 1.00 [0.98-1.03] | 17,420 | 17,053 | 1,680 | 3,937 | 15,199 |
| Z36 | 1.12 [1.07-1.18] | 1.12 [1.07-1.18] | 1.10 [0.89-1.36] | 0.99 [0.87-1.12] | 1.05 [0.99-1.11] | 1.10 [1.04-1.16] | 1.09 [1.03-1.14] | 1.13 [0.91-1.42] | 0.97 [0.85-1.10] | 1.05 [0.98-1.11] | 3,620 | 3,545 | 185 | 590 | 2,480 |
| Z37 | 1.07 [1.06-1.09] | 1.12 [1.10-1.13] | 1.06 [0.99-1.13] | 0.98 [0.96-1.00] | 1.00 [0.98-1.01] | 1.06 [1.05-1.08] | 1.08 [1.07-1.10] | 1.12 [1.04-1.20] | 0.98 [0.96-1.00] | 1.00 [0.99-1.02] | 78,196 | 76,161 | 2,190 | 27,275 | 41,423 |
| Z38 | 1.09 [0.88-1.35] | 1.18 [0.95-1.47] | | | 1.02 [0.80-1.31] | 1.09 [0.88-1.36] | 1.18 [0.94-1.48] | | | 1.01 [0.79-1.29] | 203 | 194 | NA | NA | 148 |
| Z39 | 1.08 [1.04-1.12] | 1.09 [1.05-1.14] | 1.03 [0.86-1.24] | 1.05 [0.97-1.14] | 1.03 [0.98-1.08] | 1.07 [1.03-1.12] | 1.07 [1.03-1.12] | 1.06 [0.88-1.29] | 1.06 [0.97-1.15] | 1.03 [0.98-1.08] | 5,909 | 5,809 | 242 | 1,473 | 4,324 |
| Z40 | 1.12 [1.02-1.22] | 1.11 [1.00-1.22] | 1.11 [1.00-1.24] | 1.07 [0.85-1.35] | 1.02 [0.93-1.12] | 1.10 [1.00-1.20] | 1.05 [0.95-1.16] | 1.06 [0.96-1.19] | 1.05 [0.83-1.33] | 1.01 [0.92-1.12] | 1,164 | 999 | 793 | 209 | 1,030 |
| Z41 | 1.34 [1.24-1.45] | 1.24 [1.12-1.37] | 1.34 [1.18-1.52] | 1.39 [1.24-1.57] | 1.22 [1.12-1.33] | 1.29 [1.19-1.40] | 1.13 [1.02-1.26] | 1.23 [1.08-1.40] | 1.39 [1.23-1.57] | 1.21 [1.11-1.32] | 1,558 | 932 | 558 | 744 | 1,282 |
| Z42 | 1.18 [1.12-1.25] | 1.17 [1.11-1.24] | 1.15 [1.09-1.23] | 1.15 [0.98-1.36] | 1.11 [1.05-1.17] | 1.16 [1.09-1.22] | 1.10 [1.04-1.17] | 1.10 [1.03-1.17] | 1.15 [0.97-1.36] | 1.11 [1.05-1.18] | 3,092 | 2,864 | 2,308 | 375 | 2,820 |
| Z43 | 1.15 [1.10-1.20] | 1.24 [1.18-1.30] | 1.18 [1.12-1.24] | 0.93 [0.83-1.04] | 1.09 [1.04-1.14] | 1.11 [1.06-1.16] | 1.17 [1.11-1.23] | 1.12 [1.07-1.19] | 0.87 [0.77-0.98] | 1.08 [1.03-1.13] | 4,563 | 3,992 | 3,496 | 722 | 4,114 |
| Z44 | 1.32 [0.98-1.79] | | | | | 1.28 [0.93-1.74] | | | | | 109 | NA | NA | NA | NA |
| Z45 | 1.20 [1.16-1.24] | 1.23 [1.19-1.27] | 1.21 [1.16-1.25] | 1.08 [0.99-1.18] | 1.10 [1.06-1.13] | 1.14 [1.11-1.18] | 1.13 [1.10-1.17] | 1.11 [1.07-1.15] | 1.04 [0.96-1.14] | 1.07 [1.04-1.11] | 9,851 | 8,771 | 7,938 | 1,342 | 8,975 |
| Z46 | 1.14 [1.11-1.17] | 1.15 [1.12-1.19] | 1.14 [1.10-1.17] | 1.05 [0.95-1.17] | 1.07 [1.04-1.10] | 1.11 [1.08-1.14] | 1.10 [1.06-1.13] | 1.08 [1.05-1.12] | 1.02 [0.92-1.14] | 1.06 [1.03-1.09] | 11,527 | 10,919 | 9,939 | 913 | 10,056 |
| Z47 | 1.12 [1.08-1.15] | 1.19 [1.14-1.24] | 1.21 [1.15-1.28] | 1.03 [0.99-1.08] | 1.08 [1.05-1.12] | 1.07 [1.04-1.11] | 1.10 [1.06-1.15] | 1.13 [1.07-1.19] | 1.02 [0.97-1.06] | 1.06 [1.03-1.10] | 9,978 | 5,300 | 3,416 | 5,434 | 8,236 |
| Z48 | 1.20 [1.14-1.26] | 1.30 [1.22-1.38] | 1.22 [1.15-1.31] | 1.10 [1.01-1.20] | 1.15 [1.08-1.21] | 1.15 [1.10-1.22] | 1.21 [1.14-1.29] | 1.15 [1.08-1.23] | 1.06 [0.97-1.16] | 1.12 [1.06-1.19] | 3,747 | 2,608 | 2,063 | 1,292 | 3,160 |
| Z49 | 1.44 [1.34-1.54] | 1.46 [1.37-1.57] | 1.44 [1.34-1.55] | | 1.25 [1.17-1.34] | 1.40 [1.31-1.50] | 1.35 [1.25-1.45] | 1.30 [1.21-1.40] | | 1.26 [1.17-1.36] | 2,075 | 2,028 | 1,871 | NA | 1,896 |
| Z50 | 1.19 [1.17-1.20] | 1.19 [1.17-1.20] | 1.19 [1.17-1.20] | 1.12 [1.06-1.18] | 1.14 [1.12-1.15] | 1.14 [1.13-1.16] | 1.11 [1.09-1.12] | 1.11 [1.09-1.12] | 1.07 [1.02-1.13] | 1.12 [1.10-1.13] | 70,554 | 68,190 | 64,410 | 3,939 | 63,974 |
| Z51 | 1.21 [1.20-1.23] | 1.21 [1.19-1.22] | 1.20 [1.19-1.21] | 1.22 [1.17-1.27] | 1.16 [1.14-1.17] | 1.17 [1.16-1.18] | 1.13 [1.12-1.14] | 1.12 [1.11-1.14] | 1.17 [1.12-1.22] | 1.13 [1.12-1.15] | 83,777 | 79,689 | 72,749 | 5,874 | 74,534 |
| Z52 | 1.00 [0.86-1.17] | 1.02 [0.87-1.19] | 0.97 [0.80-1.18] | | 0.94 [0.79-1.13] | 1.01 [0.86-1.18] | 1.03 [0.88-1.21] | 0.99 [0.82-1.20] | | 0.96 [0.80-1.15] | 394 | 353 | 231 | NA | 273 |
| Z53 | 1.31 [1.30-1.33] | 1.32 [1.31-1.34] | 1.31 [1.29-1.33] | 1.25 [1.21-1.28] | 1.21 [1.20-1.23] | 1.23 [1.22-1.25] | 1.20 [1.18-1.21] | 1.19 [1.17-1.20] | 1.19 [1.16-1.23] | 1.17 [1.15-1.18] | 78,316 | 68,368 | 57,099 | 13,474 | 67,570 |
| Z54 | 1.40 [1.24-1.58] | 1.35 [1.20-1.53] | 1.40 [1.24-1.59] | | 1.27 [1.11-1.47] | 1.36 [1.20-1.54] | 1.29 [1.14-1.46] | 1.33 [1.17-1.51] | | 1.27 [1.10-1.46] | 669 | 661 | 652 | NA | 518 |
| Z55 | 1.33 [1.13-1.56] | | | 1.23 [1.03-1.46] | 1.18 [0.99-1.41] | 1.25 [1.06-1.48] | | | 1.19 [0.99-1.42] | 1.15 [0.96-1.38] | 436 | NA | NA | 379 | 359 |
| Z56 | 1.17 [1.06-1.31] | 1.16 [1.05-1.29] | 1.14 [0.99-1.32] | 1.00 [0.78-1.29] | 1.07 [0.96-1.20] | 1.09 [0.98-1.22] | 1.00 [0.89-1.11] | 0.99 [0.85-1.14] | 0.96 [0.73-1.25] | 1.04 [0.92-1.16] | 836 | 828 | 444 | 163 | 713 |
| Z57 | 1.27 [1.20-1.35] | 1.26 [1.18-1.34] | 1.27 [1.19-1.35] | | 1.21 [1.14-1.29] | 1.21 [1.14-1.29] | 1.16 [1.09-1.24] | 1.17 [1.10-1.25] | | 1.16 [1.09-1.24] | 2,575 | 2,573 | 2,527 | NA | 2,317 |
| Z58 | 1.32 [1.23-1.41] | 1.23 [1.13-1.33] | 1.25 [1.14-1.37] | 1.44 [1.27-1.62] | 1.24 [1.15-1.33] | 1.22 [1.14-1.31] | 1.09 [1.00-1.19] | 1.11 [1.01-1.22] | 1.36 [1.20-1.54] | 1.16 [1.08-1.25] | 2,140 | 1,496 | 1,248 | 755 | 1,967 |
| Z59 | 1.09 [1.01-1.17] | 1.09 [1.01-1.17] | 1.11 [1.01-1.21] | 1.02 [0.87-1.21] | 1.00 [0.93-1.08] | 1.05 [0.97-1.13] | 0.98 [0.91-1.06] | 1.00 [0.91-1.10] | 1.08 [0.90-1.28] | 1.03 [0.96-1.12] | 1,829 | 1,710 | 1,134 | 364 | 1,639 |
| Z60 | 1.18 [1.15-1.21] | 1.18 [1.16-1.21] | 1.18 [1.16-1.21] | 0.92 [0.80-1.06] | 1.12 [1.09-1.15] | 1.14 [1.11-1.16] | 1.11 [1.08-1.13] | 1.11 [1.08-1.13] | 0.92 [0.80-1.06] | 1.10 [1.08-1.13] | 21,873 | 21,488 | 20,779 | 524 | 19,012 |
| Z61 | 1.06 [0.96-1.17] | 1.03 [0.89-1.18] | 1.31 [1.05-1.64] | 0.98 [0.87-1.11] | 0.95 [0.86-1.06] | 1.00 [0.90-1.11] | 0.86 [0.74-0.99] | 1.05 [0.83-1.32] | 0.97 [0.86-1.10] | 0.94 [0.85-1.05] | 1,042 | 473 | 184 | 706 | 885 |
| Z62 | 1.01 [0.82-1.23] | | | 0.96 [0.77-1.21] | 0.87 [0.69-1.08] | 0.93 [0.76-1.15] | | | 0.94 [0.75-1.19] | 0.84 [0.67-1.06] | 238 | NA | NA | 194 | 198 |
| Z63 | 1.12 [1.08-1.16] | 1.17 [1.12-1.22] | 1.23 [1.17-1.30] | 0.94 [0.88-1.00] | 1.05 [1.01-1.09] | 1.05 [1.01-1.09] | 1.02 [0.98-1.07] | 1.08 [1.02-1.14] | 0.92 [0.86-0.98] | 1.03 [0.99-1.07] | 7,491 | 5,594 | 3,787 | 2,657 | 6,615 |
| Z64 | 0.99 [0.78-1.25] | 1.01 [0.79-1.29] | | | 0.99 [0.75-1.30] | 0.94 [0.74-1.20] | 0.90 [0.69-1.17] | | | 0.98 [0.74-1.30] | 168 | 148 | NA | NA | 122 |
| Z65 | 1.09 [0.96-1.24] | 1.14 [0.99-1.32] | 1.18 [0.95-1.46] | 0.96 [0.78-1.17] | 1.00 [0.87-1.14] | 1.04 [0.91-1.18] | 0.99 [0.86-1.15] | 1.00 [0.80-1.25] | 0.99 [0.80-1.23] | 1.01 [0.88-1.16] | 604 | 459 | 202 | 247 | 541 |
| Z71 | 1.65 [1.59-1.70] | 1.22 [1.16-1.27] | 1.18 [1.12-1.23] | 2.95 [2.79-3.12] | 1.57 [1.51-1.62] | 1.49 [1.44-1.55] | 1.11 [1.06-1.16] | 1.09 [1.04-1.15] | 2.36 [2.22-2.50] | 1.43 [1.38-1.48] | 9,814 | 5,511 | 4,554 | 4,667 | 8,735 |
| Z72 | 1.20 [1.18-1.22] | 1.21 [1.19-1.22] | 1.22 [1.20-1.24] | 1.02 [0.98-1.06] | 1.11 [1.09-1.12] | 1.17 [1.15-1.18] | 1.09 [1.07-1.11] | 1.12 [1.10-1.14] | 1.02 [0.98-1.07] | 1.11 [1.10-1.13] | 47,679 | 46,403 | 31,807 | 5,984 | 36,499 |
| Z73 | 1.18 [1.14-1.22] | 1.18 [1.14-1.22] | 1.19 [1.15-1.22] | 1.08 [0.91-1.27] | 1.13 [1.09-1.16] | 1.13 [1.10-1.17] | 1.09 [1.06-1.12] | 1.10 [1.07-1.14] | 1.05 [0.88-1.24] | 1.11 [1.07-1.14] | 11,575 | 11,392 | 10,814 | 405 | 10,721 |
| Z74 | 1.18 [1.14-1.22] | 1.17 [1.13-1.21] | 1.17 [1.13-1.21] | | 1.12 [1.08-1.16] | 1.16 [1.12-1.20] | 1.12 [1.08-1.16] | 1.11 [1.08-1.15] | | 1.12 [1.08-1.16] | 8,723 | 8,665 | 8,539 | NA | 7,908 |
| Z75 | 1.13 [1.10-1.16] | 1.12 [1.09-1.15] | 1.12 [1.09-1.15] | 1.13 [0.96-1.32] | 1.10 [1.07-1.13] | 1.11 [1.08-1.14] | 1.07 [1.04-1.10] | 1.07 [1.05-1.10] | 1.11 [0.94-1.31] | 1.10 [1.07-1.13] | 16,971 | 16,704 | 16,369 | 397 | 15,746 |
| Z76 | 1.20 [1.10-1.30] | 1.17 [1.07-1.28] | 1.43 [1.21-1.69] | 1.19 [1.01-1.39] | 1.15 [1.04-1.26] | 1.18 [1.08-1.28] | 1.15 [1.05-1.26] | 1.36 [1.15-1.61] | 1.13 [0.95-1.33] | 1.14 [1.04-1.25] | 1,414 | 1,206 | 381 | 403 | 1,143 |
| Z80 | 1.17 [1.14-1.20] | 1.17 [1.15-1.20] | 1.15 [1.12-1.18] | 1.22 [1.11-1.34] | 1.09 [1.06-1.12] | 1.11 [1.09-1.14] | 1.09 [1.07-1.12] | 1.08 [1.05-1.10] | 1.17 [1.06-1.29] | 1.06 [1.03-1.08] | 18,501 | 18,055 | 15,189 | 1,231 | 15,525 |
| Z81 | 1.18 [1.11-1.24] | 1.16 [1.08-1.24] | 1.33 [1.16-1.51] | 1.13 [1.05-1.21] | 1.07 [1.01-1.13] | 1.11 [1.05-1.17] | 1.00 [0.92-1.07] | 1.14 [1.00-1.31] | 1.09 [1.02-1.18] | 1.05 [0.99-1.11] | 3,400 | 1,813 | 534 | 2,171 | 2,979 |
| Z82 | 1.30 [1.28-1.32] | 1.23 [1.21-1.25] | 1.23 [1.21-1.26] | 1.56 [1.51-1.61] | 1.21 [1.19-1.23] | 1.23 [1.21-1.24] | 1.11 [1.09-1.13] | 1.11 [1.09-1.14] | 1.50 [1.45-1.54] | 1.17 [1.15-1.19] | 44,479 | 35,084 | 27,088 | 12,010 | 38,153 |
| Z83 | 1.23 [1.21-1.27] | 1.22 [1.19-1.26] | 1.20 [1.16-1.25] | 1.24 [1.19-1.29] | 1.17 [1.14-1.20] | 1.17 [1.14-1.20] | 1.13 [1.10-1.16] | 1.11 [1.07-1.15] | 1.19 [1.14-1.24] | 1.13 [1.10-1.16] | 17,209 | 13,223 | 6,277 | 6,352 | 14,733 |
| Z84 | 1.76 [1.66-1.87] | 1.25 [1.14-1.37] | 1.41 [1.22-1.64] | 1.96 [1.83-2.10] | 1.61 [1.52-1.71] | 1.69 [1.59-1.80] | 1.18 [1.07-1.30] | 1.30 [1.12-1.52] | 1.88 [1.75-2.02] | 1.57 [1.47-1.67] | 3,330 | 1,131 | 433 | 2,435 | 2,975 |
| Z85 | 1.17 [1.15-1.18] | 1.17 [1.15-1.18] | 1.17 [1.15-1.18] | 1.05 [0.96-1.16] | 1.10 [1.08-1.11] | 1.13 [1.12-1.15] | 1.10 [1.09-1.12] | 1.10 [1.09-1.12] | 1.06 [0.96-1.17] | 1.09 [1.08-1.10] | 66,459 | 65,798 | 63,639 | 1,095 | 60,350 |
| Z86 | 1.26 [1.25-1.27] | 1.26 [1.26-1.27] | 1.28 [1.27-1.29] | 1.10 [1.08-1.13] | 1.17 [1.16-1.18] | 1.19 [1.19-1.20] | 1.14 [1.13-1.15] | 1.16 [1.15-1.17] | 1.09 [1.06-1.11] | 1.14 [1.13-1.15] | 226,366 | 219,938 | 182,267 | 19,235 | 197,681 |
| Z87 | 1.29 [1.28-1.30] | 1.30 [1.28-1.31] | 1.29 [1.28-1.31] | 1.25 [1.23-1.28] | 1.19 [1.18-1.20] | 1.20 [1.19-1.21] | 1.16 [1.14-1.17] | 1.16 [1.14-1.17] | 1.19 [1.17-1.22] | 1.13 [1.12-1.15] | 123,301 | 106,361 | 82,698 | 24,819 | 112,318 |
| Z88 | 1.47 [1.45-1.48] | 1.46 [1.45-1.48] | 1.46 [1.44-1.47] | 1.46 [1.42-1.49] | 1.37 [1.36-1.38] | 1.33 [1.31-1.34] | 1.27 [1.26-1.29] | 1.28 [1.26-1.29] | 1.34 [1.30-1.37] | 1.27 [1.25-1.28] | 131,321 | 120,607 | 95,640 | 19,072 | 112,610 |
| Z89 | 1.28 [1.21-1.35] | 1.29 [1.22-1.36] | 1.29 [1.22-1.36] | | 1.18 [1.11-1.24] | 1.23 [1.16-1.30] | 1.17 [1.10-1.24] | 1.15 [1.09-1.22] | | 1.17 [1.11-1.24] | 3,283 | 3,232 | 3,109 | NA | 3,063 |
| Z90 | 1.22 [1.21-1.24] | 1.22 [1.21-1.24] | 1.21 [1.20-1.23] | 1.21 [1.14-1.28] | 1.13 [1.12-1.15] | 1.15 [1.13-1.16] | 1.11 [1.09-1.12] | 1.10 [1.09-1.12] | 1.16 [1.09-1.23] | 1.10 [1.08-1.11] | 59,573 | 58,287 | 52,051 | 3,107 | 55,520 |
| Z91 | 1.76 [1.73-1.78] | 1.54 [1.52-1.56] | 1.56 [1.53-1.59] | 2.13 [2.08-2.18] | 1.61 [1.59-1.64] | 1.54 [1.52-1.56] | 1.29 [1.27-1.31] | 1.33 [1.30-1.36] | 1.86 [1.81-1.90] | 1.46 [1.44-1.48] | 66,982 | 46,826 | 30,367 | 27,043 | 59,336 |
| Z92 | 1.24 [1.23-1.25] | 1.24 [1.23-1.25] | 1.23 [1.22-1.24] | 1.19 [1.13-1.25] | 1.15 [1.14-1.16] | 1.17 [1.16-1.18] | 1.13 [1.12-1.14] | 1.12 [1.11-1.13] | 1.14 [1.08-1.19] | 1.12 [1.11-1.13] | 142,365 | 139,836 | 132,899 | 4,690 | 128,638 |
| Z93 | 1.19 [1.16-1.23] | 1.22 [1.19-1.26] | 1.19 [1.16-1.22] | 0.96 [0.87-1.05] | 1.12 [1.09-1.15] | 1.15 [1.12-1.18] | 1.15 [1.12-1.18] | 1.13 [1.09-1.16] | 0.90 [0.82-0.99] | 1.11 [1.08-1.14] | 13,568 | 12,723 | 11,716 | 1,158 | 12,368 |
| Z94 | 1.36 [1.28-1.44] | 1.42 [1.33-1.50] | 1.40 [1.31-1.50] | 1.09 [0.94-1.26] | 1.21 [1.14-1.28] | 1.31 [1.23-1.38] | 1.32 [1.24-1.40] | 1.30 [1.21-1.39] | 1.06 [0.91-1.24] | 1.20 [1.13-1.27] | 2,996 | 2,708 | 2,181 | 449 | 2,741 |
| Z95 | 1.22 [1.21-1.24] | 1.23 [1.22-1.25] | 1.23 [1.21-1.25] | 0.95 [0.86-1.05] | 1.12 [1.10-1.14] | 1.17 [1.15-1.19] | 1.13 [1.11-1.15] | 1.12 [1.10-1.14] | 0.92 [0.83-1.02] | 1.10 [1.08-1.12] | 45,729 | 45,044 | 44,079 | 1,021 | 42,251 |
| Z96 | 1.22 [1.20-1.23] | 1.22 [1.20-1.23] | 1.22 [1.20-1.23] | 1.12 [1.04-1.20] | 1.14 [1.13-1.15] | 1.17 [1.15-1.18] | 1.14 [1.13-1.15] | 1.14 [1.13-1.15] | 1.08 [1.01-1.17] | 1.11 [1.10-1.13] | 88,982 | 87,678 | 85,483 | 2,009 | 81,648 |
| Z97 | 1.26 [1.22-1.31] | 1.29 [1.24-1.33] | 1.26 [1.22-1.31] | 1.12 [1.01-1.23] | 1.18 [1.14-1.22] | 1.18 [1.14-1.23] | 1.16 [1.12-1.21] | 1.16 [1.12-1.21] | 1.07 [0.97-1.18] | 1.14 [1.10-1.18] | 9,194 | 8,666 | 6,867 | 1,135 | 8,518 |
| Z98 | 1.29 [1.26-1.33] | 1.32 [1.28-1.36] | 1.28 [1.25-1.32] | 1.08 [0.98-1.19] | 1.19 [1.16-1.23] | 1.18 [1.15-1.22] | 1.16 [1.12-1.19] | 1.14 [1.10-1.17] | 1.03 [0.93-1.14] | 1.13 [1.10-1.16] | 12,376 | 11,819 | 10,149 | 1,038 | 11,673 |
| Z99 | 1.39 [1.36-1.43] | 1.42 [1.38-1.45] | 1.41 [1.38-1.45] | 0.94 [0.86-1.04] | 1.25 [1.22-1.28] | 1.28 [1.25-1.31] | 1.23 [1.20-1.27] | 1.23 [1.19-1.26] | 0.91 [0.82-1.01] | 1.19 [1.16-1.22] | 16,202 | 15,470 | 14,479 | 1,037 | 15,347 |
