## Supplementary Table 4 for "Mapping risks of hospital-recorded health conditions in people with eczema"

### Supplementary Table 4: Hazard ratios and events from all cohorts (excluding non-consulters)

| | Hazard ratio (99% confidence interval) | | | | | | | | | | | | | | |
|---|---|---|---|---|---|---|---|---|---|---|---|---|---|---|---|
| | crude | | | | | adjusted | | | | | Events (in exposed) | | | | |
| Outcome | any age | 18+ | 40+ | <18 | hosp. | any age | 18+ | 40+ | <18 | hosp. | any age | 18+ | 40+ | <18 | hosp. |
| Certain infectious and parasitic diseases (A00-B99) | | | | | | | | | | | | | | | |
| A02 | 0.98 [0.77-1.25] | 0.95 [0.74-1.22] | 1.04 [0.79-1.36] | | 0.93 [0.72-1.19] | 0.92 [0.71-1.18] | 0.88 [0.68-1.14] | 0.95 [0.72-1.26] | | 0.88 [0.68-1.14] | 192 | 175 | 151 | NA | 169 |
| A04 | 1.18 [1.13-1.22] | 1.17 [1.13-1.21] | 1.17 [1.12-1.22] | 0.95 [0.75-1.22] | 1.15 [1.11-1.20] | 1.13 [1.09-1.18] | 1.11 [1.07-1.15] | 1.10 [1.06-1.15] | 0.90 [0.70-1.16] | 1.12 [1.08-1.17] | 8,234 | 8,145 | 7,600 | 217 | 7,681 |
| A07 | 1.06 [0.77-1.46] | | | | 1.24 [0.90-1.71] | 0.99 [0.72-1.37] | | | | 1.22 [0.88-1.68] | 119 | NA | NA | NA | 110 |
| A08 | 1.18 [1.13-1.23] | 1.18 [1.12-1.24] | 1.21 [1.15-1.28] | 1.16 [1.08-1.24] | 1.16 [1.11-1.21] | 1.14 [1.09-1.19] | 1.09 [1.03-1.15] | 1.12 [1.06-1.19] | 1.14 [1.07-1.22] | 1.13 [1.08-1.18] | 6,714 | 4,327 | 3,700 | 2,622 | 6,347 |
| A09 | 1.18 [1.16-1.21] | 1.19 [1.17-1.21] | 1.18 [1.16-1.21] | 1.04 [0.98-1.12] | 1.15 [1.13-1.17] | 1.12 [1.10-1.14] | 1.09 [1.07-1.11] | 1.09 [1.07-1.11] | 1.02 [0.95-1.09] | 1.11 [1.08-1.13] | 34,612 | 33,538 | 28,367 | 2,970 | 33,614 |
| A15 | 1.15 [0.88-1.50] | 1.16 [0.89-1.51] | 1.23 [0.93-1.64] | | 1.15 [0.87-1.51] | 1.12 [0.85-1.46] | 1.14 [0.87-1.49] | 1.16 [0.87-1.55] | | 1.10 [0.83-1.46] | 162 | 160 | 138 | NA | 151 |
| A16 | 1.30 [1.10-1.55] | 1.27 [1.07-1.52] | 1.32 [1.09-1.60] | | 1.28 [1.07-1.53] | 1.25 [1.05-1.49] | 1.21 [1.01-1.45] | 1.25 [1.03-1.53] | | 1.23 [1.02-1.48] | 409 | 395 | 323 | NA | 344 |
| A18 | 1.13 [0.88-1.44] | 1.25 [0.97-1.61] | 1.24 [0.94-1.64] | | 1.14 [0.88-1.47] | 1.07 [0.83-1.38] | 1.21 [0.94-1.57] | 1.19 [0.90-1.58] | | 1.11 [0.85-1.44] | 187 | 187 | 146 | NA | 161 |
| A31 | 1.50 [1.16-1.93] | 1.52 [1.16-1.98] | 1.48 [1.11-1.96] | | 1.49 [1.16-1.92] | 1.40 [1.07-1.82] | 1.33 [1.01-1.76] | 1.36 [1.02-1.82] | | 1.35 [1.04-1.75] | 183 | 171 | 151 | NA | 172 |
| A38 | 1.17 [0.85-1.62] | | | 1.07 [0.77-1.49] | 1.20 [0.86-1.66] | 1.12 [0.81-1.56] | | | 1.05 [0.75-1.46] | 1.17 [0.84-1.63] | 130 | NA | NA | 124 | 131 |
| A39 | 0.98 [0.75-1.30] | | | | 0.92 [0.69-1.23] | 0.95 [0.72-1.26] | | | | 0.90 [0.67-1.20] | 151 | NA | NA | NA | 136 |
| A40 | 1.22 [1.12-1.33] | 1.25 [1.15-1.36] | 1.27 [1.17-1.39] | | 1.24 [1.14-1.34] | 1.19 [1.09-1.30] | 1.20 [1.10-1.31] | 1.21 [1.11-1.32] | | 1.22 [1.12-1.33] | 1,706 | 1,687 | 1,607 | NA | 1,673 |
| A41 | 1.16 [1.14-1.18] | 1.17 [1.14-1.19] | 1.17 [1.14-1.19] | 1.05 [0.91-1.20] | 1.15 [1.13-1.17] | 1.13 [1.11-1.16] | 1.11 [1.09-1.14] | 1.11 [1.09-1.13] | 1.04 [0.89-1.20] | 1.14 [1.12-1.16] | 32,658 | 32,404 | 31,120 | 638 | 31,785 |
| A46 | 1.82 [1.33-2.49] | 1.89 [1.38-2.60] | 1.98 [1.42-2.75] | | 1.81 [1.31-2.49] | 1.76 [1.28-2.41] | 1.78 [1.29-2.47] | 1.90 [1.36-2.65] | | 1.78 [1.29-2.45] | 135 | 139 | 124 | NA | 119 |
| A48 | 1.14 [0.85-1.52] | 0.98 [0.73-1.32] | 1.12 [0.83-1.52] | | 1.27 [0.95-1.69] | 1.14 [0.85-1.53] | 0.96 [0.71-1.29] | 1.09 [0.80-1.48] | | 1.30 [0.97-1.74] | 139 | 123 | 123 | NA | 133 |
| A49 | 1.23 [1.17-1.29] | 1.24 [1.18-1.31] | 1.22 [1.16-1.29] | 1.00 [0.76-1.31] | 1.20 [1.14-1.26] | 1.19 [1.13-1.25] | 1.16 [1.10-1.23] | 1.15 [1.09-1.21] | 0.98 [0.75-1.30] | 1.17 [1.11-1.23] | 4,598 | 4,477 | 4,236 | 177 | 4,369 |
| A63 | 1.34 [0.97-1.85] | 1.32 [0.96-1.82] | | | | 1.17 [0.82-1.65] | 1.17 [0.84-1.64] | | | | 108 | 107 | NA | NA | NA |
| A86 | 1.12 [0.87-1.44] | 1.04 [0.81-1.34] | 1.17 [0.89-1.53] | | 1.07 [0.82-1.39] | 1.09 [0.85-1.40] | 0.99 [0.77-1.28] | 1.10 [0.84-1.46] | | 1.03 [0.79-1.34] | 179 | 170 | 149 | NA | 159 |
| A87 | 1.20 [1.01-1.42] | 1.26 [1.07-1.49] | 1.50 [1.12-2.00] | | 1.15 [0.97-1.36] | 1.10 [0.93-1.31] | 1.15 [0.97-1.37] | 1.35 [1.00-1.82] | | 1.10 [0.93-1.31] | 437 | 431 | 144 | NA | 404 |
| B00 | 1.93 [1.76-2.12] | 1.66 [1.49-1.85] | 1.52 [1.35-1.72] | 3.30 [2.76-3.95] | 1.82 [1.65-2.00] | 1.82 [1.65-2.00] | 1.52 [1.36-1.70] | 1.40 [1.24-1.58] | 3.18 [2.65-3.83] | 1.72 [1.57-1.90] | 1,698 | 1,150 | 857 | 651 | 1,547 |
| B01 | 1.56 [1.38-1.78] | 0.97 [0.73-1.30] | | 1.70 [1.47-1.96] | 1.53 [1.34-1.74] | 1.53 [1.34-1.74] | 0.91 [0.67-1.23] | | 1.67 [1.44-1.93] | 1.51 [1.31-1.73] | 831 | 136 | NA | 694 | 762 |
| B02 | 1.32 [1.23-1.42] | 1.29 [1.20-1.38] | 1.28 [1.19-1.37] | | 1.28 [1.19-1.37] | 1.26 [1.18-1.36] | 1.21 [1.12-1.30] | 1.21 [1.12-1.30] | | 1.23 [1.15-1.32] | 2,484 | 2,436 | 2,314 | NA | 2,389 |
| B07 | 1.24 [1.10-1.40] | 1.24 [1.10-1.40] | 1.31 [1.15-1.49] | | 1.25 [1.11-1.41] | 1.19 [1.05-1.34] | 1.14 [1.01-1.29] | 1.23 [1.08-1.40] | | 1.20 [1.07-1.36] | 859 | 826 | 673 | NA | 784 |
| B08 | 1.63 [1.33-2.00] | | | 1.59 [1.25-2.01] | 1.72 [1.39-2.11] | 1.54 [1.25-1.89] | | | 1.51 [1.18-1.92] | 1.61 [1.30-2.00] | 335 | NA | NA | 248 | 316 |
| B09 | 1.12 [0.80-1.58] | | | | | 1.11 [0.78-1.56] | | | | | 103 | NA | NA | NA | NA |
| B16 | 1.11 [0.88-1.41] | 1.13 [0.89-1.45] | 1.29 [0.97-1.71] | | 0.99 [0.77-1.26] | 1.10 [0.86-1.40] | 1.14 [0.89-1.47] | 1.28 [0.96-1.71] | | 0.99 [0.77-1.27] | 197 | 188 | 143 | NA | 170 |
| B17 | 1.17 [0.96-1.44] | 1.28 [1.05-1.57] | 1.39 [1.11-1.73] | | 1.11 [0.91-1.36] | 1.13 [0.92-1.39] | 1.21 [0.98-1.48] | 1.28 [1.02-1.61] | | 1.08 [0.88-1.33] | 286 | 285 | 238 | NA | 265 |
| B18 | 1.07 [0.97-1.18] | 1.09 [0.99-1.21] | 1.14 [1.02-1.27] | | 1.00 [0.90-1.10] | 1.06 [0.96-1.18] | 1.06 [0.95-1.17] | 1.09 [0.98-1.22] | | 1.02 [0.92-1.13] | 1,173 | 1,167 | 965 | NA | 1,052 |
| B25 | 1.26 [1.01-1.58] | 1.18 [0.94-1.47] | 1.32 [1.02-1.71] | | 1.08 [0.86-1.34] | 1.24 [0.98-1.55] | 1.10 [0.87-1.38] | 1.29 [0.99-1.68] | | 1.04 [0.83-1.31] | 240 | 231 | 186 | NA | 219 |
| B27 | 1.10 [0.92-1.31] | 0.98 [0.81-1.18] | | 1.12 [0.90-1.40] | 1.03 [0.84-1.26] | 1.03 [0.86-1.25] | 0.89 [0.73-1.09] | | 1.08 [0.85-1.36] | 0.96 [0.77-1.19] | 422 | 333 | NA | 278 | 302 |
| B34 | 1.50 [1.46-1.55] | 1.19 [1.13-1.25] | 1.24 [1.16-1.32] | 1.66 [1.60-1.73] | 1.46 [1.42-1.51] | 1.44 [1.39-1.48] | 1.06 [1.01-1.12] | 1.12 [1.05-1.19] | 1.61 [1.55-1.68] | 1.40 [1.36-1.45] | 14,598 | 4,503 | 2,897 | 10,637 | 13,533 |
| B35 | 1.56 [1.40-1.74] | 1.63 [1.46-1.82] | 1.62 [1.45-1.81] | | 1.61 [1.45-1.79] | 1.46 [1.31-1.63] | 1.49 [1.33-1.67] | 1.50 [1.34-1.68] | | 1.52 [1.37-1.70] | 1,152 | 1,122 | 1,049 | NA | 1,121 |
| B36 | 1.76 [1.48-2.08] | 1.74 [1.46-2.06] | 1.61 [1.35-1.92] | | 1.63 [1.38-1.93] | 1.64 [1.38-1.96] | 1.58 [1.33-1.89] | 1.49 [1.24-1.78] | | 1.58 [1.33-1.87] | 473 | 460 | 431 | NA | 455 |
| B37 | 1.27 [1.23-1.31] | 1.27 [1.23-1.30] | 1.29 [1.25-1.33] | 0.98 [0.85-1.14] | 1.22 [1.19-1.26] | 1.19 [1.15-1.22] | 1.15 [1.11-1.18] | 1.17 [1.14-1.21] | 0.94 [0.81-1.10] | 1.16 [1.12-1.19] | 13,890 | 13,960 | 12,039 | 586 | 13,510 |
| B44 | 1.55 [1.33-1.79] | 1.53 [1.32-1.77] | 1.58 [1.36-1.85] | | 1.46 [1.26-1.69] | 1.30 [1.12-1.52] | 1.31 [1.12-1.53] | 1.30 [1.11-1.53] | | 1.20 [1.02-1.40] | 580 | 575 | 543 | NA | 572 |
| B49 | 1.41 [1.16-1.73] | 1.42 [1.16-1.74] | 1.30 [1.06-1.60] | | 1.36 [1.12-1.66] | 1.35 [1.10-1.65] | 1.34 [1.09-1.65] | 1.23 [0.99-1.52] | | 1.33 [1.09-1.62] | 310 | 297 | 275 | NA | 295 |
| B59 | 1.39 [1.06-1.81] | 1.46 [1.12-1.90] | 1.20 [0.91-1.56] | | 1.26 [0.97-1.64] | 1.31 [1.00-1.72] | 1.38 [1.05-1.81] | 1.15 [0.87-1.51] | | 1.19 [0.90-1.56] | 164 | 167 | 153 | NA | 162 |
| B80 | 1.24 [0.99-1.57] | 1.18 [0.91-1.53] | | | 1.18 [0.94-1.50] | 1.22 [0.96-1.55] | 1.08 [0.82-1.42] | | | 1.13 [0.89-1.44] | 231 | 170 | NA | NA | 215 |
| B85 | 1.20 [0.87-1.66] | | | | 1.29 [0.92-1.80] | 1.17 [0.84-1.64] | | | | 1.29 [0.91-1.82] | 114 | NA | NA | NA | 105 |
| B86 | 2.66 [2.19-3.23] | 2.59 [2.13-3.15] | 2.52 [2.07-3.07] | | 2.42 [2.00-2.94] | 2.64 [2.16-3.21] | 2.48 [2.03-3.03] | 2.41 [1.97-2.95] | | 2.47 [2.03-3.00] | 416 | 398 | 384 | NA | 396 |
| B90 | 1.56 [1.17-2.08] | 1.35 [1.01-1.80] | 1.42 [1.05-1.92] | | 1.36 [1.02-1.81] | 1.45 [1.08-1.95] | 1.25 [0.93-1.68] | 1.33 [0.98-1.81] | | 1.28 [0.96-1.71] | 147 | 147 | 135 | NA | 140 |
| B91 | 1.21 [0.95-1.55] | 1.11 [0.87-1.41] | 1.17 [0.91-1.50] | | 1.07 [0.84-1.38] | 1.24 [0.97-1.58] | 1.13 [0.88-1.44] | 1.17 [0.91-1.51] | | 1.09 [0.85-1.41] | 195 | 192 | 192 | NA | 168 |
| B94 | 1.17 [0.96-1.44] | 1.21 [0.98-1.51] | 1.15 [0.92-1.44] | | 1.12 [0.91-1.37] | 1.07 [0.86-1.32] | 1.08 [0.87-1.36] | 1.08 [0.85-1.36] | | 1.05 [0.85-1.30] | 278 | 251 | 224 | NA | 275 |
| B95 | 1.37 [1.34-1.41] | 1.37 [1.34-1.41] | 1.37 [1.33-1.41] | 1.33 [1.20-1.48] | 1.34 [1.31-1.38] | 1.33 [1.30-1.36] | 1.29 [1.26-1.33] | 1.30 [1.26-1.33] | 1.30 [1.17-1.45] | 1.32 [1.29-1.35] | 19,482 | 18,791 | 16,505 | 1,326 | 18,546 |
| B96 | 1.15 [1.13-1.17] | 1.15 [1.13-1.18] | 1.16 [1.13-1.18] | 1.00 [0.90-1.10] | 1.14 [1.12-1.16] | 1.12 [1.10-1.14] | 1.09 [1.07-1.11] | 1.09 [1.07-1.11] | 0.99 [0.90-1.10] | 1.12 [1.10-1.14] | 37,843 | 37,301 | 34,633 | 1,355 | 35,990 |
| B97 | 1.20 [1.16-1.25] | 1.17 [1.12-1.22] | 1.17 [1.12-1.22] | 1.23 [1.14-1.34] | 1.17 [1.13-1.22] | 1.13 [1.09-1.17] | 1.07 [1.03-1.12] | 1.07 [1.02-1.12] | 1.19 [1.10-1.29] | 1.12 [1.08-1.16] | 9,079 | 7,386 | 6,260 | 1,997 | 8,738 |
| B98 | 1.10 [1.04-1.16] | 1.10 [1.04-1.16] | 1.09 [1.03-1.16] | | 1.06 [1.00-1.12] | 1.06 [1.00-1.12] | 1.05 [0.99-1.11] | 1.04 [0.98-1.11] | | 1.04 [0.98-1.10] | 3,561 | 3,535 | 3,250 | NA | 3,313 |
| B99 | 1.16 [1.05-1.30] | 1.15 [1.03-1.28] | 1.11 [0.99-1.24] | | 1.10 [0.99-1.22] | 1.14 [1.02-1.27] | 1.09 [0.97-1.21] | 1.06 [0.94-1.18] | | 1.10 [0.98-1.22] | 1,050 | 1,022 | 972 | NA | 1,024 |
| Neoplasms (C00-D49) | | | | | | | | | | | | | | | |
| C00 | 1.03 [0.76-1.41] | 1.26 [0.91-1.73] | 1.07 [0.78-1.47] | | 1.11 [0.81-1.51] | 1.03 [0.75-1.41] | 1.26 [0.91-1.73] | 1.04 [0.76-1.44] | | 1.10 [0.81-1.50] | 110 | 111 | 106 | NA | 106 |
| C01 | 1.03 [0.82-1.29] | 1.07 [0.85-1.35] | 1.04 [0.82-1.31] | | 1.12 [0.89-1.42] | 1.01 [0.80-1.28] | 1.07 [0.84-1.35] | 1.05 [0.83-1.33] | | 1.17 [0.93-1.49] | 207 | 202 | 199 | NA | 192 |
| C02 | 1.27 [1.07-1.51] | 1.29 [1.08-1.52] | 1.33 [1.12-1.58] | | 1.38 [1.16-1.65] | 1.26 [1.06-1.50] | 1.23 [1.04-1.46] | 1.30 [1.09-1.55] | | 1.40 [1.17-1.67] | 396 | 397 | 380 | NA | 375 |
| C04 | 1.07 [0.77-1.49] | 1.12 [0.81-1.55] | | | | 1.07 [0.77-1.50] | 1.04 [0.75-1.46] | | | | 101 | 102 | NA | NA | NA |
| C05 | 1.29 [0.95-1.73] | 1.42 [1.05-1.92] | 1.46 [1.08-1.97] | | 1.14 [0.84-1.55] | 1.30 [0.96-1.76] | 1.38 [1.02-1.87] | 1.42 [1.04-1.92] | | 1.17 [0.86-1.61] | 132 | 134 | 128 | NA | 115 |
| C06 | 1.15 [0.90-1.47] | 1.19 [0.93-1.53] | 1.16 [0.91-1.50] | | 1.24 [0.96-1.59] | 1.14 [0.89-1.46] | 1.15 [0.89-1.48] | 1.09 [0.85-1.42] | | 1.22 [0.94-1.57] | 180 | 178 | 175 | NA | 172 |
| C07 | 0.95 [0.73-1.24] | 0.95 [0.72-1.25] | 0.93 [0.71-1.22] | | 0.96 [0.72-1.26] | 0.94 [0.71-1.22] | 0.92 [0.69-1.21] | 0.91 [0.69-1.19] | | 0.94 [0.71-1.26] | 149 | 150 | 144 | NA | 136 |
| C09 | 1.01 [0.81-1.24] | 1.00 [0.81-1.24] | 1.05 [0.85-1.30] | | 0.96 [0.77-1.20] | 0.99 [0.79-1.23] | 0.97 [0.78-1.20] | 1.02 [0.82-1.27] | | 0.98 [0.78-1.23] | 238 | 234 | 233 | NA | 205 |
| C10 | 1.21 [0.92-1.59] | 1.17 [0.89-1.54] | 1.17 [0.89-1.54] | | 1.12 [0.85-1.48] | 1.19 [0.90-1.58] | 1.12 [0.85-1.48] | 1.12 [0.84-1.48] | | 1.16 [0.87-1.54] | 145 | 143 | 140 | NA | 134 |
| C14 | 1.02 [0.79-1.31] | 1.01 [0.78-1.30] | 0.99 [0.77-1.28] | | 1.02 [0.79-1.32] | 1.02 [0.79-1.31] | 0.98 [0.76-1.27] | 0.96 [0.74-1.24] | | 1.05 [0.81-1.35] | 165 | 165 | 167 | NA | 158 |
| C15 | 1.14 [1.06-1.22] | 1.14 [1.06-1.22] | 1.13 [1.06-1.21] | | 1.10 [1.03-1.18] | 1.14 [1.06-1.22] | 1.11 [1.04-1.19] | 1.11 [1.04-1.19] | | 1.10 [1.03-1.18] | 2,425 | 2,422 | 2,427 | NA | 2,178 |
| C16 | 1.04 [0.96-1.12] | 1.00 [0.92-1.08] | 1.02 [0.95-1.10] | | 1.04 [0.96-1.12] | 1.03 [0.95-1.12] | 0.98 [0.91-1.06] | 1.00 [0.93-1.09] | | 1.03 [0.96-1.12] | 1,850 | 1,863 | 1,848 | NA | 1,705 |
| C17 | 1.07 [0.90-1.26] | 1.07 [0.90-1.26] | 1.04 [0.88-1.23] | | 1.07 [0.91-1.27] | 1.06 [0.90-1.26] | 1.03 [0.87-1.22] | 1.03 [0.87-1.21] | | 1.06 [0.90-1.26] | 401 | 399 | 398 | NA | 375 |
| C18 | 1.00 [0.95-1.04] | 1.00 [0.96-1.04] | 1.01 [0.97-1.05] | | 1.00 [0.95-1.04] | 0.99 [0.95-1.03] | 0.99 [0.95-1.03] | 1.00 [0.96-1.04] | | 0.99 [0.95-1.04] | 6,291 | 6,266 | 6,194 | NA | 5,511 |
| C19 | 0.92 [0.84-1.01] | 0.95 [0.86-1.04] | 0.96 [0.87-1.06] | | 0.96 [0.87-1.06] | 0.93 [0.84-1.02] | 0.96 [0.87-1.05] | 0.97 [0.88-1.07] | | 0.97 [0.88-1.07] | 1,210 | 1,195 | 1,190 | NA | 1,073 |
| C20 | 0.93 [0.87-1.00] | 0.96 [0.90-1.02] | 0.94 [0.88-1.01] | | 0.94 [0.88-1.01] | 0.94 [0.88-1.01] | 0.97 [0.91-1.04] | 0.96 [0.89-1.02] | | 0.96 [0.89-1.03] | 2,422 | 2,423 | 2,391 | NA | 2,051 |
| C21 | 1.03 [0.86-1.23] | 1.05 [0.88-1.27] | 1.01 [0.84-1.21] | | 0.96 [0.79-1.15] | 1.03 [0.86-1.23] | 1.04 [0.86-1.25] | 0.97 [0.81-1.17] | | 0.95 [0.79-1.15] | 328 | 327 | 319 | NA | 291 |
| C22 | 1.22 [1.12-1.33] | 1.19 [1.09-1.30] | 1.19 [1.09-1.30] | | 1.14 [1.05-1.25] | 1.19 [1.09-1.30] | 1.15 [1.06-1.26] | 1.16 [1.06-1.26] | | 1.14 [1.05-1.25] | 1,536 | 1,531 | 1,519 | NA | 1,432 |
| C23 | 1.01 [0.79-1.28] | 1.05 [0.83-1.33] | 0.98 [0.77-1.24] | | 1.02 [0.80-1.31] | 1.01 [0.80-1.29] | 1.04 [0.82-1.32] | 0.98 [0.77-1.25] | | 1.05 [0.82-1.35] | 191 | 188 | 184 | NA | 170 |
| C24 | 1.08 [0.91-1.28] | 1.04 [0.87-1.24] | 1.02 [0.86-1.21] | | 1.05 [0.87-1.25] | 1.09 [0.91-1.30] | 1.03 [0.86-1.22] | 0.99 [0.83-1.18] | | 1.04 [0.87-1.25] | 360 | 360 | 356 | NA | 319 |
| C25 | 1.12 [1.05-1.20] | 1.08 [1.01-1.16] | 1.10 [1.03-1.18] | | 1.11 [1.03-1.19] | 1.11 [1.04-1.20] | 1.06 [0.99-1.14] | 1.08 [1.00-1.15] | | 1.11 [1.03-1.19] | 2,317 | 2,322 | 2,310 | NA | 2,117 |
| C26 | 1.08 [0.93-1.26] | 1.06 [0.91-1.23] | 1.11 [0.95-1.29] | | 1.01 [0.87-1.17] | 1.07 [0.92-1.25] | 1.02 [0.88-1.19] | 1.08 [0.93-1.26] | | 0.99 [0.86-1.15] | 538 | 535 | 526 | NA | 516 |
| C32 | 1.09 [0.93-1.26] | 1.08 [0.93-1.26] | 1.08 [0.93-1.25] | | 1.08 [0.93-1.26] | 1.09 [0.93-1.27] | 1.07 [0.92-1.25] | 1.06 [0.91-1.23] | | 1.10 [0.94-1.29] | 468 | 465 | 467 | NA | 432 |
| C34 | 1.12 [1.09-1.16] | 1.11 [1.07-1.14] | 1.12 [1.08-1.16] | | 1.09 [1.06-1.13] | 1.12 [1.08-1.15] | 1.06 [1.03-1.10] | 1.07 [1.04-1.11] | | 1.10 [1.06-1.14] | 9,955 | 9,936 | 9,945 | NA | 9,046 |
| C38 | 1.03 [0.76-1.40] | 0.93 [0.68-1.27] | 0.97 [0.72-1.31] | | 0.95 [0.70-1.30] | 1.05 [0.77-1.43] | 0.92 [0.67-1.26] | 0.96 [0.71-1.30] | | 0.96 [0.70-1.31] | 119 | 115 | 113 | NA | 102 |
| C41 | 1.27 [1.03-1.57] | 1.20 [0.97-1.48] | 1.18 [0.95-1.46] | | 1.38 [1.12-1.71] | 1.29 [1.04-1.59] | 1.19 [0.96-1.47] | 1.17 [0.94-1.45] | | 1.40 [1.12-1.74] | 264 | 261 | 241 | NA | 242 |
| C43 | 0.92 [0.86-0.99] | 0.91 [0.85-0.97] | 0.89 [0.83-0.96] | | 0.95 [0.88-1.02] | 0.92 [0.85-0.98] | 0.90 [0.84-0.97] | 0.89 [0.82-0.95] | | 0.95 [0.88-1.02] | 2,218 | 2,224 | 2,043 | NA | 1,999 |
| C44 | 1.07 [1.04-1.09] | 1.07 [1.04-1.10] | 1.07 [1.04-1.10] | | 1.04 [1.02-1.07] | 1.05 [1.02-1.07] | 1.04 [1.02-1.07] | 1.04 [1.02-1.07] | | 1.03 [1.00-1.05] | 18,993 | 19,024 | 18,771 | NA | 17,078 |
| C45 | 0.96 [0.83-1.10] | 0.97 [0.84-1.11] | 1.02 [0.89-1.18] | | 1.01 [0.87-1.16] | 0.96 [0.83-1.11] | 0.98 [0.85-1.12] | 1.03 [0.89-1.18] | | 1.02 [0.88-1.18] | 535 | 536 | 542 | NA | 484 |
| C48 | 0.96 [0.80-1.16] | 1.02 [0.85-1.23] | 1.02 [0.84-1.23] | | 1.00 [0.83-1.22] | 0.95 [0.78-1.15] | 1.01 [0.84-1.22] | 1.00 [0.82-1.21] | | 0.99 [0.81-1.21] | 298 | 292 | 287 | NA | 258 |
| C49 | 1.04 [0.90-1.19] | 1.08 [0.94-1.24] | 1.11 [0.96-1.29] | | 1.11 [0.96-1.28] | 1.02 [0.89-1.18] | 1.06 [0.91-1.22] | 1.08 [0.93-1.25] | | 1.09 [0.94-1.26] | 555 | 552 | 506 | NA | 512 |
| C50 | 1.03 [1.00-1.07] | 1.02 [0.99-1.06] | 1.03 [0.99-1.06] | | 1.02 [0.99-1.06] | 1.03 [1.00-1.07] | 1.02 [0.98-1.05] | 1.02 [0.99-1.06] | | 1.03 [1.00-1.07] | 10,383 | 10,406 | 9,875 | NA | 8,735 |
| C51 | 1.12 [0.93-1.35] | 1.17 [0.97-1.41] | 1.19 [0.98-1.44] | | 1.21 [0.99-1.47] | 1.11 [0.92-1.34] | 1.14 [0.95-1.38] | 1.17 [0.96-1.41] | | 1.19 [0.97-1.45] | 318 | 316 | 307 | NA | 282 |
| C53 | 0.99 [0.86-1.14] | 1.03 [0.89-1.18] | 1.04 [0.88-1.23] | | 1.01 [0.87-1.17] | 1.00 [0.87-1.16] | 1.03 [0.89-1.19] | 1.03 [0.87-1.23] | | 1.04 [0.89-1.21] | 531 | 540 | 365 | NA | 453 |
| C54 | 1.05 [0.97-1.13] | 1.04 [0.97-1.13] | 1.03 [0.95-1.11] | | 1.05 [0.96-1.14] | 1.06 [0.98-1.15] | 1.07 [0.99-1.16] | 1.05 [0.97-1.14] | | 1.07 [0.98-1.16] | 1,784 | 1,784 | 1,764 | NA | 1,474 |
| C55 | 1.11 [0.91-1.36] | 1.01 [0.82-1.23] | 1.03 [0.84-1.27] | | 0.95 [0.76-1.17] | 1.09 [0.89-1.34] | 0.99 [0.81-1.22] | 1.02 [0.83-1.26] | | 0.95 [0.76-1.18] | 252 | 251 | 249 | NA | 215 |
| C56 | 1.03 [0.95-1.12] | 1.03 [0.95-1.12] | 1.02 [0.94-1.11] | | 1.00 [0.92-1.10] | 1.04 [0.95-1.13] | 1.03 [0.94-1.12] | 1.02 [0.94-1.11] | | 1.01 [0.92-1.11] | 1,503 | 1,516 | 1,429 | NA | 1,231 |
| C57 | 1.12 [0.89-1.42] | 1.23 [0.97-1.56] | 1.30 [1.02-1.65] | | 1.17 [0.91-1.50] | 1.16 [0.91-1.47] | 1.25 [0.99-1.60] | 1.33 [1.04-1.69] | | 1.18 [0.92-1.52] | 192 | 193 | 190 | NA | 170 |
| C60 | 1.12 [0.84-1.49] | 1.21 [0.91-1.62] | 1.16 [0.87-1.55] | | 1.20 [0.90-1.61] | 1.14 [0.85-1.52] | 1.22 [0.91-1.63] | 1.21 [0.90-1.63] | | 1.27 [0.94-1.71] | 136 | 138 | 132 | NA | 128 |
| C61 | 1.03 [0.99-1.06] | 1.01 [0.98-1.04] | 1.01 [0.98-1.04] | | 0.99 [0.96-1.02] | 1.01 [0.98-1.05] | 1.00 [0.96-1.03] | 1.00 [0.96-1.03] | | 0.99 [0.95-1.02] | 10,385 | 10,370 | 10,387 | NA | 9,295 |
| C62 | 0.95 [0.70-1.28] | 1.02 [0.75-1.40] | | | 0.86 [0.62-1.20] | 0.95 [0.70-1.30] | 1.03 [0.74-1.43] | | | 0.88 [0.63-1.23] | 135 | 131 | NA | NA | 105 |
| C64 | 1.04 [0.97-1.11] | 1.03 [0.96-1.10] | 1.03 [0.96-1.10] | | 1.03 [0.96-1.11] | 1.03 [0.96-1.11] | 1.00 [0.93-1.07] | 1.00 [0.93-1.07] | | 1.04 [0.96-1.11] | 2,215 | 2,207 | 2,184 | NA | 2,005 |
| C65 | 1.26 [1.00-1.59] | 1.24 [0.99-1.56] | 1.11 [0.88-1.39] | | 1.16 [0.92-1.46] | 1.29 [1.02-1.62] | 1.24 [0.99-1.55] | 1.12 [0.89-1.41] | | 1.21 [0.96-1.54] | 226 | 226 | 219 | NA | 192 |
| C66 | 1.10 [0.90-1.34] | 1.03 [0.85-1.25] | 1.00 [0.82-1.21] | | 1.03 [0.84-1.24] | 1.09 [0.89-1.34] | 1.01 [0.83-1.23] | 0.98 [0.80-1.19] | | 1.03 [0.85-1.26] | 278 | 279 | 277 | NA | 266 |
| C67 | 1.03 [0.98-1.08] | 1.02 [0.98-1.07] | 1.02 [0.97-1.07] | | 1.01 [0.96-1.06] | 1.03 [0.98-1.08] | 1.01 [0.96-1.06] | 1.01 [0.96-1.06] | | 1.01 [0.96-1.06] | 4,864 | 4,851 | 4,838 | NA | 4,344 |
| C69 | 1.00 [0.78-1.28] | 0.91 [0.71-1.17] | 1.07 [0.83-1.38] | | 0.93 [0.72-1.21] | 1.00 [0.77-1.28] | 0.90 [0.70-1.15] | 1.04 [0.80-1.34] | | 0.94 [0.73-1.23] | 172 | 166 | 160 | NA | 156 |
| C71 | 1.11 [1.00-1.22] | 1.10 [0.99-1.22] | 1.10 [0.99-1.22] | | 1.10 [0.99-1.23] | 1.11 [1.00-1.23] | 1.08 [0.97-1.20] | 1.08 [0.97-1.20] | | 1.10 [0.98-1.23] | 1,062 | 1,018 | 961 | NA | 908 |
| C73 | 0.97 [0.85-1.12] | 0.98 [0.85-1.12] | 1.04 [0.89-1.21] | | 0.99 [0.86-1.15] | 0.97 [0.84-1.12] | 0.96 [0.83-1.10] | 1.03 [0.88-1.21] | | 0.98 [0.84-1.13] | 562 | 577 | 448 | NA | 509 |
| C76 | 1.08 [0.90-1.30] | 1.07 [0.89-1.30] | 1.06 [0.87-1.28] | | 1.06 [0.88-1.27] | 1.09 [0.91-1.32] | 1.09 [0.90-1.32] | 1.08 [0.89-1.31] | | 1.07 [0.89-1.29] | 320 | 308 | 297 | NA | 306 |
| C77 | 1.04 [1.01-1.07] | 1.04 [1.00-1.07] | 1.05 [1.01-1.08] | | 1.03 [1.00-1.07] | 1.04 [1.01-1.08] | 1.03 [1.00-1.06] | 1.04 [1.01-1.08] | | 1.05 [1.01-1.08] | 10,224 | 10,231 | 9,817 | NA | 9,166 |
| C78 | 1.04 [1.02-1.07] | 1.03 [1.00-1.06] | 1.04 [1.01-1.07] | | 1.04 [1.01-1.07] | 1.05 [1.02-1.08] | 1.02 [0.99-1.04] | 1.03 [1.00-1.06] | | 1.06 [1.03-1.09] | 15,762 | 15,708 | 15,524 | NA | 14,573 |
| C79 | 1.05 [1.02-1.08] | 1.04 [1.01-1.07] | 1.05 [1.02-1.09] | | 1.04 [1.01-1.07] | 1.06 [1.03-1.09] | 1.03 [1.00-1.06] | 1.04 [1.01-1.07] | | 1.06 [1.03-1.09] | 13,276 | 13,254 | 13,086 | NA | 12,441 |
| C80 | 1.07 [1.02-1.12] | 1.05 [1.00-1.11] | 1.07 [1.02-1.12] | | 1.08 [1.02-1.13] | 1.07 [1.01-1.12] | 1.03 [0.98-1.08] | 1.05 [1.00-1.10] | | 1.08 [1.03-1.14] | 4,746 | 4,731 | 4,704 | NA | 4,291 |
| C81 | 1.51 [1.30-1.77] | 1.46 [1.25-1.71] | 1.57 [1.32-1.87] | | 1.28 [1.09-1.52] | 1.47 [1.26-1.72] | 1.43 [1.22-1.67] | 1.49 [1.25-1.78] | | 1.26 [1.07-1.50] | 537 | 523 | 410 | NA | 420 |
| C82 | 1.14 [1.00-1.31] | 1.11 [0.97-1.28] | 1.09 [0.95-1.25] | | 1.07 [0.93-1.23] | 1.11 [0.97-1.28] | 1.08 [0.94-1.24] | 1.06 [0.92-1.21] | | 1.04 [0.90-1.20] | 573 | 575 | 557 | NA | 505 |
| C83 | 1.18 [1.09-1.28] | 1.20 [1.11-1.30] | 1.20 [1.11-1.30] | | 1.15 [1.06-1.25] | 1.16 [1.07-1.25] | 1.18 [1.09-1.28] | 1.18 [1.09-1.28] | | 1.13 [1.04-1.23] | 1,731 | 1,726 | 1,678 | NA | 1,567 |
| C84 | 2.29 [1.87-2.81] | 2.57 [2.09-3.16] | 2.26 [1.83-2.77] | | 2.13 [1.74-2.61] | 2.27 [1.85-2.79] | 2.52 [2.05-3.11] | 2.22 [1.80-2.73] | | 2.14 [1.74-2.63] | 361 | 356 | 337 | NA | 320 |
| C85 | 1.24 [1.16-1.33] | 1.23 [1.14-1.32] | 1.24 [1.16-1.34] | | 1.19 [1.11-1.28] | 1.21 [1.12-1.30] | 1.19 [1.11-1.28] | 1.20 [1.11-1.29] | | 1.17 [1.09-1.26] | 2,245 | 2,223 | 2,151 | NA | 2,026 |
| C88 | 1.25 [1.03-1.52] | 1.23 [1.01-1.50] | 1.26 [1.03-1.54] | | 1.14 [0.93-1.39] | 1.25 [1.02-1.52] | 1.22 [1.00-1.48] | 1.22 [1.00-1.50] | | 1.11 [0.91-1.36] | 296 | 297 | 290 | NA | 278 |
| C90 | 1.11 [1.02-1.21] | 1.12 [1.03-1.22] | 1.10 [1.01-1.20] | | 1.09 [1.00-1.19] | 1.09 [1.00-1.19] | 1.11 [1.02-1.21] | 1.09 [1.00-1.19] | | 1.08 [0.98-1.18] | 1,521 | 1,527 | 1,517 | NA | 1,390 |
| C91 | 1.04 [0.95-1.13] | 1.06 [0.97-1.16] | 1.03 [0.94-1.13] | | 1.01 [0.93-1.11] | 1.02 [0.93-1.12] | 1.04 [0.95-1.13] | 1.01 [0.92-1.11] | | 1.00 [0.92-1.10] | 1,411 | 1,356 | 1,334 | NA | 1,247 |
| C92 | 1.23 [1.12-1.36] | 1.25 [1.13-1.37] | 1.27 [1.15-1.40] | | 1.22 [1.11-1.35] | 1.22 [1.11-1.34] | 1.23 [1.11-1.35] | 1.25 [1.14-1.39] | | 1.21 [1.10-1.34] | 1,236 | 1,235 | 1,191 | NA | 1,137 |
| C93 | 1.09 [0.84-1.42] | 1.05 [0.81-1.36] | 1.06 [0.82-1.37] | | 1.03 [0.80-1.34] | 1.10 [0.85-1.43] | 1.02 [0.78-1.32] | 1.03 [0.80-1.34] | | 1.04 [0.80-1.35] | 173 | 171 | 172 | NA | 162 |
| C95 | 1.26 [1.03-1.55] | 1.20 [0.97-1.48] | 1.22 [0.99-1.52] | | 1.19 [0.98-1.46] | 1.26 [1.02-1.54] | 1.15 [0.93-1.42] | 1.17 [0.94-1.46] | | 1.19 [0.97-1.46] | 280 | 273 | 265 | NA | 273 |
| C97 | 1.03 [0.93-1.13] | 1.06 [0.96-1.17] | 1.02 [0.92-1.12] | | 1.04 [0.95-1.14] | 1.01 [0.92-1.11] | 1.03 [0.93-1.13] | 0.99 [0.90-1.09] | | 1.03 [0.94-1.13] | 1,250 | 1,261 | 1,232 | NA | 1,249 |
| D00 | 0.93 [0.70-1.23] | 0.97 [0.72-1.29] | 0.89 [0.67-1.19] | | 1.04 [0.78-1.39] | 0.91 [0.68-1.22] | 0.93 [0.69-1.25] | 0.84 [0.63-1.13] | | 1.03 [0.77-1.37] | 125 | 123 | 119 | NA | 125 |
| D01 | 1.01 [0.76-1.35] | 1.08 [0.81-1.45] | 1.01 [0.75-1.35] | | 0.98 [0.74-1.30] | 0.98 [0.74-1.31] | 1.04 [0.77-1.39] | 0.94 [0.69-1.26] | | 0.94 [0.71-1.25] | 132 | 129 | 120 | NA | 127 |
| D02 | 1.02 [0.77-1.34] | 1.09 [0.82-1.45] | 1.05 [0.80-1.39] | | 1.03 [0.78-1.37] | 0.98 [0.74-1.30] | 1.05 [0.79-1.40] | 1.01 [0.76-1.34] | | 1.02 [0.77-1.36] | 144 | 146 | 142 | NA | 131 |
| D03 | 1.03 [0.93-1.15] | 1.04 [0.93-1.15] | 1.03 [0.92-1.15] | | 1.03 [0.92-1.15] | 1.02 [0.92-1.14] | 1.01 [0.91-1.13] | 1.01 [0.90-1.12] | | 1.01 [0.90-1.13] | 927 | 937 | 893 | NA | 854 |
| D04 | 1.18 [1.10-1.27] | 1.19 [1.11-1.28] | 1.20 [1.12-1.29] | | 1.21 [1.13-1.30] | 1.14 [1.06-1.22] | 1.13 [1.06-1.22] | 1.13 [1.06-1.22] | | 1.17 [1.09-1.25] | 2,461 | 2,451 | 2,425 | NA | 2,348 |
| D05 | 0.99 [0.91-1.07] | 0.98 [0.90-1.06] | 0.97 [0.89-1.06] | | 0.94 [0.86-1.03] | 0.98 [0.90-1.07] | 0.97 [0.89-1.05] | 0.96 [0.88-1.05] | | 0.94 [0.86-1.03] | 1,529 | 1,546 | 1,469 | NA | 1,310 |
| D06 | 1.02 [0.93-1.12] | 1.02 [0.93-1.11] | 0.98 [0.81-1.18] | 0.79 [0.59-1.04] | 0.96 [0.87-1.05] | 1.01 [0.92-1.11] | 0.98 [0.89-1.07] | 0.96 [0.79-1.16] | 0.77 [0.57-1.05] | 0.97 [0.88-1.06] | 1,411 | 1,492 | 288 | 154 | 1,211 |
| D07 | 1.12 [0.97-1.28] | 1.19 [1.04-1.38] | 1.14 [0.99-1.32] | | 1.10 [0.96-1.27] | 1.07 [0.93-1.23] | 1.13 [0.98-1.31] | 1.10 [0.94-1.27] | | 1.07 [0.93-1.24] | 559 | 559 | 506 | NA | 522 |
| D09 | 1.01 [0.87-1.16] | 1.04 [0.90-1.20] | 1.02 [0.89-1.18] | | 0.98 [0.85-1.13] | 0.99 [0.86-1.15] | 1.01 [0.87-1.17] | 1.01 [0.88-1.17] | | 0.98 [0.85-1.13] | 523 | 534 | 525 | NA | 519 |
| D10 | 1.17 [1.00-1.37] | 1.20 [1.03-1.40] | 1.26 [1.06-1.50] | | 1.18 [1.00-1.38] | 1.12 [0.96-1.31] | 1.09 [0.93-1.28] | 1.14 [0.95-1.36] | | 1.12 [0.95-1.32] | 489 | 489 | 382 | NA | 439 |
| D11 | 1.04 [0.91-1.20] | 1.00 [0.87-1.15] | 1.03 [0.89-1.19] | | 1.09 [0.94-1.26] | 1.04 [0.90-1.20] | 0.97 [0.84-1.12] | 0.99 [0.86-1.15] | | 1.09 [0.94-1.26] | 553 | 554 | 501 | NA | 489 |
| D12 | 1.13 [1.10-1.15] | 1.12 [1.10-1.15] | 1.13 [1.10-1.16] | | 1.10 [1.08-1.13] | 1.07 [1.05-1.10] | 1.06 [1.03-1.09] | 1.06 [1.04-1.09] | | 1.07 [1.04-1.09] | 18,892 | 18,874 | 18,493 | NA | 17,428 |
| D13 | 1.13 [1.03-1.24] | 1.14 [1.04-1.25] | 1.14 [1.04-1.26] | | 1.10 [1.00-1.21] | 1.06 [0.97-1.17] | 1.05 [0.96-1.16] | 1.05 [0.96-1.16] | | 1.06 [0.96-1.16] | 1,308 | 1,300 | 1,229 | NA | 1,191 |
| D14 | 1.21 [1.02-1.45] | 1.09 [0.91-1.30] | 1.21 [1.00-1.47] | | 1.17 [0.97-1.40] | 1.15 [0.96-1.38] | 0.99 [0.82-1.19] | 1.15 [0.94-1.40] | | 1.14 [0.94-1.37] | 375 | 355 | 303 | NA | 333 |
| D15 | 1.05 [0.79-1.40] | 1.04 [0.78-1.37] | 1.02 [0.77-1.35] | | 1.01 [0.76-1.35] | 1.06 [0.79-1.41] | 1.02 [0.77-1.35] | 1.01 [0.76-1.35] | | 1.01 [0.75-1.35] | 140 | 139 | 125 | NA | 125 |
| D16 | 1.12 [0.94-1.32] | 1.17 [0.98-1.39] | 1.05 [0.86-1.29] | | 1.01 [0.85-1.20] | 1.11 [0.93-1.32] | 1.12 [0.93-1.34] | 1.01 [0.82-1.24] | | 0.99 [0.83-1.18] | 427 | 376 | 263 | NA | 371 |
| D17 | 1.07 [1.02-1.12] | 1.06 [1.01-1.11] | 1.07 [1.02-1.13] | 1.01 [0.73-1.38] | 1.03 [0.98-1.09] | 1.03 [0.98-1.08] | 1.01 [0.96-1.06] | 1.03 [0.98-1.08] | 0.94 [0.68-1.30] | 1.00 [0.95-1.05] | 4,816 | 4,804 | 4,102 | 139 | 4,278 |
| D18 | 1.10 [1.03-1.18] | 1.10 [1.02-1.18] | 1.15 [1.06-1.24] | 1.02 [0.80-1.32] | 1.06 [0.99-1.14] | 1.05 [0.98-1.13] | 1.03 [0.96-1.11] | 1.06 [0.98-1.15] | 1.02 [0.79-1.32] | 1.02 [0.95-1.10] | 2,318 | 2,180 | 1,893 | 203 | 2,182 |
| D21 | 1.23 [1.07-1.42] | 1.24 [1.08-1.43] | 1.24 [1.07-1.44] | | 1.19 [1.03-1.38] | 1.20 [1.04-1.38] | 1.17 [1.02-1.35] | 1.18 [1.02-1.38] | | 1.14 [0.98-1.32] | 599 | 589 | 509 | NA | 527 |
| D22 | 1.07 [1.02-1.13] | 1.08 [1.03-1.14] | 1.05 [0.98-1.12] | 1.10 [0.95-1.28] | 1.05 [0.99-1.10] | 1.04 [0.99-1.10] | 1.04 [0.99-1.10] | 1.01 [0.94-1.08] | 1.08 [0.92-1.26] | 1.03 [0.97-1.08] | 4,336 | 4,215 | 2,454 | 578 | 3,746 |
| D23 | 1.12 [1.05-1.18] | 1.16 [1.09-1.24] | 1.16 [1.09-1.24] | 1.05 [0.85-1.29] | 1.12 [1.06-1.20] | 1.07 [1.01-1.14] | 1.11 [1.05-1.19] | 1.11 [1.03-1.19] | 1.02 [0.82-1.26] | 1.09 [1.02-1.16] | 3,067 | 2,866 | 2,409 | 291 | 2,680 |
| D24 | 1.02 [0.94-1.12] | 1.00 [0.92-1.08] | 1.04 [0.93-1.16] | 1.00 [0.76-1.30] | 0.99 [0.91-1.09] | 1.00 [0.91-1.09] | 0.95 [0.87-1.04] | 1.00 [0.90-1.12] | 0.98 [0.74-1.31] | 0.97 [0.89-1.07] | 1,501 | 1,591 | 881 | 175 | 1,283 |
| D25 | 1.02 [0.99-1.06] | 1.05 [1.01-1.09] | 1.04 [1.00-1.08] | | 1.04 [1.00-1.08] | 1.01 [0.97-1.05] | 1.04 [1.00-1.08] | 1.02 [0.98-1.06] | | 1.03 [0.99-1.07] | 8,252 | 8,312 | 6,739 | NA | 6,943 |
| D26 | 1.05 [0.86-1.28] | 1.12 [0.92-1.36] | 0.97 [0.78-1.21] | | 1.01 [0.82-1.24] | 1.03 [0.84-1.26] | 1.10 [0.90-1.34] | 0.95 [0.75-1.18] | | 0.98 [0.80-1.21] | 285 | 290 | 211 | NA | 245 |
| D27 | 1.00 [0.93-1.08] | 1.01 [0.94-1.09] | 1.00 [0.91-1.09] | 0.97 [0.70-1.36] | 0.99 [0.91-1.07] | 0.99 [0.92-1.07] | 0.99 [0.91-1.06] | 0.97 [0.89-1.06] | 0.97 [0.68-1.38] | 0.98 [0.90-1.06] | 1,914 | 1,975 | 1,296 | 116 | 1,619 |
| D28 | 1.20 [0.96-1.50] | 1.18 [0.94-1.47] | 1.16 [0.89-1.52] | | 1.24 [0.98-1.56] | 1.15 [0.91-1.44] | 1.14 [0.91-1.42] | 1.16 [0.88-1.53] | | 1.17 [0.92-1.49] | 237 | 234 | 154 | NA | 202 |
| D29 | 0.97 [0.74-1.28] | 1.04 [0.79-1.37] | 0.99 [0.74-1.31] | | 1.04 [0.79-1.36] | 0.96 [0.73-1.27] | 1.03 [0.78-1.37] | 0.98 [0.74-1.30] | | 1.04 [0.79-1.37] | 150 | 147 | 140 | NA | 144 |
| D30 | 0.99 [0.83-1.19] | 0.98 [0.82-1.17] | 0.93 [0.77-1.11] | | 0.96 [0.80-1.15] | 0.97 [0.81-1.17] | 0.95 [0.79-1.14] | 0.91 [0.75-1.09] | | 0.95 [0.80-1.14] | 330 | 329 | 306 | NA | 316 |
| D31 | 1.13 [0.94-1.35] | 1.13 [0.94-1.37] | 1.11 [0.91-1.36] | | 1.16 [0.96-1.40] | 1.12 [0.93-1.35] | 1.09 [0.90-1.33] | 1.08 [0.88-1.33] | | 1.15 [0.95-1.39] | 346 | 321 | 288 | NA | 321 |

### Supplementary Table 4: Hazard ratios and events from all cohorts (excluding non-consulters)

| | Hazard ratio (99% confidence interval) | | | | | | | | | | Events (in exposed) | | | | |
|---|---|---|---|---|---|---|---|---|---|---|---|---|---|---|---|
| | crude | | | | | adjusted | | | | | | | | | |
| Outcome | any age | 18+ | 40+ | <18 | hosp. | any age | 18+ | 40+ | <18 | hosp. | any age | 18+ | 40+ | <18 | hosp. |
| D32 | 1.08 [0.98-1.18] | 1.04 [0.95-1.14] | 1.09 [0.99-1.20] | | 1.07 [0.97-1.17] | 1.05 [0.96-1.15] | 1.00 [0.91-1.10] | 1.04 [0.95-1.15] | | 1.06 [0.96-1.16] | 1,362 | 1,355 | 1,300 | NA | 1,259 |
| D33 | 0.95 [0.81-1.12] | 1.00 [0.85-1.17] | 1.04 [0.89-1.23] | | 0.96 [0.82-1.13] | 0.90 [0.76-1.05] | 0.93 [0.79-1.09] | 0.98 [0.83-1.16] | | 0.93 [0.79-1.10] | 440 | 434 | 397 | NA | 408 |
| D34 | 1.02 [0.84-1.24] | 0.95 [0.79-1.16] | 0.92 [0.75-1.14] | | 0.98 [0.80-1.19] | 0.98 [0.81-1.20] | 0.91 [0.75-1.11] | 0.89 [0.72-1.11] | | 0.93 [0.76-1.14] | 293 | 295 | 232 | NA | 265 |
| D35 | 1.05 [0.97-1.13] | 1.01 [0.94-1.09] | 1.03 [0.95-1.11] | | 1.00 [0.92-1.08] | 1.02 [0.94-1.10] | 0.97 [0.90-1.05] | 0.99 [0.91-1.07] | | 0.98 [0.91-1.06] | 1,885 | 1,872 | 1,698 | NA | 1,715 |
| D36 | 1.16 [1.02-1.30] | 1.16 [1.02-1.31] | 1.20 [1.05-1.38] | | 1.14 [1.01-1.29] | 1.09 [0.97-1.24] | 1.07 [0.95-1.22] | 1.11 [0.97-1.28] | | 1.08 [0.95-1.23] | 783 | 764 | 631 | NA | 709 |
| D37 | 1.03 [0.97-1.10] | 1.05 [0.98-1.12] | 1.04 [0.97-1.11] | | 1.08 [1.01-1.15] | 1.02 [0.95-1.09] | 1.03 [0.96-1.10] | 1.01 [0.94-1.08] | | 1.08 [1.00-1.15] | 2,430 | 2,436 | 2,352 | NA | 2,221 |
| D38 | 1.17 [1.02-1.34] | 1.11 [0.97-1.27] | 1.16 [1.01-1.32] | | 1.14 [1.00-1.31] | 1.16 [1.01-1.33] | 1.07 [0.94-1.23] | 1.12 [0.97-1.28] | | 1.15 [1.00-1.32] | 603 | 608 | 590 | NA | 549 |
| D39 | 1.06 [0.87-1.28] | 1.02 [0.84-1.24] | 0.99 [0.80-1.22] | | 0.99 [0.81-1.21] | 1.06 [0.88-1.29] | 1.01 [0.83-1.23] | 0.96 [0.78-1.19] | | 1.00 [0.81-1.22] | 291 | 301 | 235 | NA | 253 |
| D41 | 1.06 [0.98-1.14] | 1.04 [0.96-1.13] | 1.02 [0.95-1.11] | | 0.99 [0.92-1.08] | 1.05 [0.97-1.14] | 1.02 [0.94-1.11] | 1.00 [0.92-1.08] | | 1.00 [0.92-1.08] | 1,747 | 1,762 | 1,725 | NA | 1,570 |
| D43 | 1.09 [0.94-1.26] | 1.04 [0.89-1.20] | 1.00 [0.86-1.16] | | 0.98 [0.84-1.14] | 1.08 [0.93-1.25] | 1.02 [0.88-1.19] | 0.97 [0.83-1.13] | | 0.98 [0.84-1.14] | 520 | 499 | 455 | NA | 462 |
| D44 | 1.13 [0.94-1.34] | 1.09 [0.91-1.30] | 1.19 [0.99-1.44] | | 1.08 [0.90-1.29] | 1.06 [0.89-1.27] | 1.02 [0.85-1.23] | 1.13 [0.93-1.37] | | 1.06 [0.89-1.27] | 355 | 347 | 313 | NA | 326 |
| D45 | 0.96 [0.84-1.10] | 0.98 [0.86-1.13] | 0.95 [0.83-1.09] | | 0.97 [0.85-1.12] | 0.96 [0.83-1.10] | 0.94 [0.82-1.08] | 0.91 [0.79-1.05] | | 0.97 [0.84-1.11] | 576 | 578 | 549 | NA | 526 |
| D46 | 1.22 [1.13-1.33] | 1.28 [1.18-1.39] | 1.25 [1.15-1.35] | | 1.21 [1.12-1.32] | 1.20 [1.10-1.30] | 1.24 [1.14-1.34] | 1.21 [1.11-1.31] | | 1.20 [1.10-1.30] | 1,810 | 1,813 | 1,802 | NA | 1,715 |
| D47 | 1.20 [1.12-1.27] | 1.21 [1.14-1.29] | 1.21 [1.13-1.29] | | 1.18 [1.11-1.25] | 1.15 [1.08-1.23] | 1.14 [1.07-1.22] | 1.15 [1.07-1.22] | | 1.14 [1.07-1.22] | 2,913 | 2,936 | 2,826 | NA | 2,857 |
| D48 | 1.01 [0.91-1.12] | 1.03 [0.93-1.14] | 1.03 [0.92-1.15] | | 1.00 [0.90-1.11] | 0.98 [0.89-1.09] | 1.00 [0.90-1.11] | 1.00 [0.89-1.11] | | 0.98 [0.88-1.09] | 1,068 | 1,062 | 927 | NA | 965 |
| C03 | | 1.50 [1.06-2.13] | | | | | 1.50 [1.06-2.13] | | | | NA | 103 | NA | NA | NA |
| Diseases of the blood and blood-forming organs and certain disorders involving the immune mechanism (D50-D89) | | | | | | | | | | | | | | | |
| D50 | 1.22 [1.20-1.24] | 1.21 [1.19-1.24] | 1.22 [1.20-1.24] | 1.15 [1.03-1.29] | 1.19 [1.17-1.21] | 1.17 [1.15-1.19] | 1.13 [1.11-1.16] | 1.13 [1.11-1.15] | 1.13 [1.01-1.27] | 1.15 [1.13-1.17] | 39,867 | 39,616 | 36,296 | 1,149 | 38,121 |
| D51 | 1.25 [1.18-1.33] | 1.22 [1.15-1.29] | 1.25 [1.18-1.32] | | 1.22 [1.16-1.30] | 1.19 [1.12-1.26] | 1.13 [1.07-1.20] | 1.17 [1.10-1.24] | | 1.18 [1.11-1.25] | 3,681 | 3,708 | 3,427 | NA | 3,492 |
| D52 | 1.20 [1.12-1.29] | 1.24 [1.16-1.33] | 1.25 [1.16-1.34] | | 1.23 [1.15-1.31] | 1.17 [1.10-1.26] | 1.18 [1.10-1.26] | 1.19 [1.11-1.27] | | 1.22 [1.14-1.31] | 2,700 | 2,724 | 2,620 | NA | 2,664 |
| D53 | 1.31 [1.15-1.48] | 1.31 [1.16-1.48] | 1.41 [1.24-1.61] | | 1.29 [1.14-1.47] | 1.24 [1.09-1.41] | 1.22 [1.07-1.38] | 1.29 [1.14-1.48] | | 1.26 [1.11-1.43] | 801 | 801 | 780 | NA | 774 |
| D56 | 1.22 [1.11-1.34] | 1.20 [1.09-1.32] | 1.32 [1.18-1.49] | 1.04 [0.77-1.40] | 1.19 [1.08-1.30] | 1.21 [1.10-1.33] | 1.19 [1.08-1.31] | 1.25 [1.11-1.42] | 1.03 [0.75-1.41] | 1.18 [1.07-1.31] | 1,404 | 1,350 | 846 | 151 | 1,257 |
| D57 | 1.15 [1.04-1.27] | 1.15 [1.03-1.27] | 1.06 [0.93-1.22] | 1.41 [1.12-1.78] | 1.12 [1.01-1.24] | 1.15 [1.04-1.27] | 1.16 [1.04-1.29] | 1.05 [0.92-1.21] | 1.50 [1.17-1.92] | 1.13 [1.02-1.25] | 1,281 | 1,115 | 625 | 278 | 1,122 |
| D58 | 1.05 [0.87-1.26] | 1.05 [0.86-1.27] | 1.09 [0.89-1.35] | | 1.15 [0.95-1.39] | 1.06 [0.87-1.28] | 1.03 [0.85-1.26] | 1.08 [0.87-1.34] | | 1.16 [0.96-1.41] | 328 | 307 | 248 | NA | 311 |
| D59 | 1.29 [1.10-1.51] | 1.30 [1.10-1.52] | 1.29 [1.09-1.52] | | 1.30 [1.11-1.52] | 1.27 [1.09-1.49] | 1.29 [1.09-1.52] | 1.27 [1.07-1.50] | | 1.31 [1.11-1.54] | 470 | 455 | 417 | NA | 459 |
| D61 | 1.21 [1.12-1.30] | 1.23 [1.14-1.32] | 1.21 [1.12-1.30] | | 1.20 [1.12-1.29] | 1.18 [1.10-1.27] | 1.18 [1.10-1.27] | 1.16 [1.08-1.25] | | 1.20 [1.11-1.28] | 2,343 | 2,320 | 2,209 | NA | 2,302 |
| D62 | 1.04 [0.93-1.16] | 1.04 [0.94-1.16] | 1.09 [0.94-1.27] | | 1.03 [0.92-1.15] | 1.06 [0.95-1.18] | 1.03 [0.93-1.15] | 1.07 [0.92-1.24] | | 1.04 [0.93-1.17] | 977 | 1,021 | 518 | NA | 870 |
| D63 | 1.19 [1.13-1.26] | 1.22 [1.15-1.29] | 1.16 [1.10-1.23] | | 1.13 [1.07-1.20] | 1.17 [1.11-1.24] | 1.17 [1.10-1.23] | 1.11 [1.05-1.17] | | 1.13 [1.07-1.20] | 3,874 | 3,858 | 3,727 | NA | 3,695 |
| D64 | 1.21 [1.19-1.22] | 1.20 [1.19-1.22] | 1.21 [1.19-1.23] | 1.00 [0.89-1.13] | 1.18 [1.16-1.19] | 1.17 [1.15-1.19] | 1.14 [1.12-1.16] | 1.15 [1.13-1.17] | 0.99 [0.87-1.11] | 1.16 [1.14-1.17] | 53,146 | 53,064 | 49,352 | 1,024 | 50,319 |
| D65 | 1.21 [0.93-1.57] | 1.28 [0.97-1.69] | 1.15 [0.86-1.54] | | 1.36 [1.03-1.78] | 1.21 [0.93-1.58] | 1.27 [0.96-1.68] | 1.13 [0.84-1.52] | | 1.36 [1.04-1.79] | 158 | 152 | 136 | NA | 147 |
| D66 | 1.04 [0.78-1.39] | 1.03 [0.76-1.38] | | | 1.24 [0.92-1.67] | 1.01 [0.75-1.36] | 0.96 [0.71-1.31] | | | 1.22 [0.90-1.67] | 134 | 125 | NA | NA | 124 |
| D68 | 1.21 [1.13-1.29] | 1.21 [1.13-1.29] | 1.23 [1.14-1.33] | 0.92 [0.70-1.20] | 1.16 [1.09-1.24] | 1.13 [1.06-1.21] | 1.11 [1.03-1.19] | 1.12 [1.03-1.21] | 0.88 [0.66-1.17] | 1.12 [1.04-1.19] | 2,654 | 2,617 | 1,958 | 179 | 2,511 |
| D69 | 1.18 [1.14-1.23] | 1.16 [1.11-1.21] | 1.20 [1.15-1.26] | 0.98 [0.85-1.14] | 1.15 [1.10-1.20] | 1.16 [1.11-1.20] | 1.12 [1.08-1.17] | 1.16 [1.11-1.21] | 1.00 [0.86-1.16] | 1.14 [1.10-1.19] | 7,552 | 7,088 | 6,388 | 615 | 7,171 |
| D70 | 1.17 [1.12-1.22] | 1.16 [1.12-1.22] | 1.18 [1.13-1.23] | 1.09 [0.89-1.35] | 1.14 [1.09-1.19] | 1.15 [1.10-1.20] | 1.14 [1.09-1.19] | 1.16 [1.11-1.21] | 1.07 [0.86-1.32] | 1.13 [1.09-1.18] | 6,386 | 6,187 | 5,705 | 313 | 5,917 |
| D72 | 1.28 [1.17-1.39] | 1.30 [1.19-1.42] | 1.26 [1.15-1.38] | 1.53 [1.03-2.27] | 1.28 [1.18-1.40] | 1.18 [1.08-1.29] | 1.18 [1.08-1.30] | 1.17 [1.06-1.28] | 1.38 [0.91-2.09] | 1.21 [1.11-1.32] | 1,675 | 1,643 | 1,446 | 101 | 1,633 |
| D73 | 1.14 [1.04-1.24] | 1.14 [1.04-1.24] | 1.13 [1.03-1.23] | | 1.12 [1.02-1.22] | 1.10 [1.01-1.20] | 1.08 [0.99-1.18] | 1.07 [0.97-1.17] | | 1.10 [1.01-1.20] | 1,545 | 1,531 | 1,426 | NA | 1,495 |
| D75 | 1.13 [1.04-1.22] | 1.12 [1.04-1.21] | 1.12 [1.04-1.22] | | 1.10 [1.02-1.19] | 1.10 [1.02-1.19] | 1.06 [0.98-1.15] | 1.07 [0.99-1.16] | | 1.09 [1.01-1.18] | 1,964 | 1,954 | 1,841 | NA | 1,863 |
| D76 | 1.26 [0.90-1.77] | | | | | 1.18 [0.83-1.68] | | | | | 101 | NA | NA | NA | NA |
| D80 | 1.27 [1.08-1.49] | 1.31 [1.11-1.55] | 1.20 [1.01-1.43] | | 1.29 [1.10-1.51] | 1.15 [0.97-1.36] | 1.17 [0.98-1.39] | 1.04 [0.87-1.25] | | 1.17 [1.00-1.37] | 466 | 425 | 374 | NA | 470 |
| D84 | 1.36 [1.09-1.70] | 1.45 [1.13-1.85] | 1.51 [1.16-1.97] | | 1.46 [1.18-1.82] | 1.17 [0.93-1.48] | 1.25 [0.96-1.62] | 1.21 [0.91-1.62] | | 1.25 [0.99-1.57] | 263 | 217 | 176 | NA | 255 |
| D86 | 1.35 [1.21-1.50] | 1.35 [1.22-1.50] | 1.38 [1.24-1.54] | | 1.31 [1.18-1.45] | 1.27 [1.14-1.41] | 1.26 [1.13-1.41] | 1.28 [1.15-1.43] | | 1.25 [1.12-1.39] | 1,104 | 1,100 | 1,006 | NA | 1,041 |
| D89 | 1.34 [1.20-1.50] | 1.28 [1.14-1.44] | 1.32 [1.17-1.48] | | 1.25 [1.12-1.40] | 1.26 [1.12-1.41] | 1.21 [1.07-1.35] | 1.22 [1.08-1.37] | | 1.20 [1.07-1.35] | 939 | 919 | 870 | NA | 878 |
| Endocrine, nutritional and metabolic diseases (E00-E89) | | | | | | | | | | | | | | | |
| E02 | 1.11 [0.91-1.36] | 1.17 [0.96-1.43] | 1.10 [0.89-1.35] | | 1.14 [0.93-1.39] | 1.10 [0.90-1.35] | 1.14 [0.93-1.41] | 1.08 [0.87-1.33] | | 1.14 [0.93-1.40] | 300 | 306 | 281 | NA | 292 |
| E03 | 1.20 [1.18-1.23] | 1.20 [1.18-1.22] | 1.20 [1.18-1.22] | 0.88 [0.76-1.04] | 1.18 [1.16-1.20] | 1.17 [1.15-1.19] | 1.15 [1.13-1.17] | 1.15 [1.13-1.18] | 0.91 [0.77-1.07] | 1.16 [1.13-1.18] | 38,325 | 38,453 | 34,872 | 506 | 33,143 |
| E04 | 1.06 [1.00-1.12] | 1.04 [0.98-1.10] | 1.02 [0.96-1.09] | | 1.04 [0.98-1.10] | 1.02 [0.96-1.08] | 0.99 [0.93-1.05] | 0.97 [0.91-1.03] | | 1.02 [0.96-1.08] | 3,405 | 3,385 | 3,022 | NA | 3,186 |
| E05 | 1.15 [1.10-1.20] | 1.14 [1.09-1.19] | 1.14 [1.08-1.19] | 0.97 [0.71-1.32] | 1.12 [1.07-1.17] | 1.11 [1.06-1.16] | 1.08 [1.03-1.13] | 1.07 [1.02-1.12] | 0.98 [0.71-1.36] | 1.09 [1.04-1.14] | 6,227 | 6,290 | 5,358 | 137 | 5,817 |
| E06 | 1.14 [0.99-1.30] | 1.13 [0.99-1.30] | 1.16 [0.99-1.35] | | 1.19 [1.04-1.36] | 1.10 [0.96-1.27] | 1.07 [0.93-1.23] | 1.10 [0.94-1.29] | | 1.14 [0.99-1.31] | 636 | 642 | 478 | NA | 616 |
| E07 | 1.17 [1.06-1.30] | 1.19 [1.07-1.32] | 1.15 [1.03-1.29] | | 1.15 [1.03-1.27] | 1.11 [1.00-1.23] | 1.11 [1.00-1.23] | 1.08 [0.97-1.21] | | 1.09 [0.98-1.21] | 1,083 | 1,084 | 963 | NA | 1,031 |
| E10 | 1.06 [1.02-1.10] | 1.07 [1.03-1.11] | 1.10 [1.05-1.14] | 0.77 [0.64-0.92] | 1.07 [1.02-1.11] | 1.01 [0.97-1.05] | 0.97 [0.94-1.02] | 1.00 [0.95-1.04] | 0.77 [0.64-0.92] | 1.05 [1.00-1.09] | 7,209 | 6,882 | 6,311 | 419 | 6,308 |
| E11 | 1.08 [1.06-1.09] | 1.08 [1.06-1.09] | 1.07 [1.06-1.09] | 0.78 [0.60-1.03] | 1.09 [1.07-1.10] | 1.04 [1.03-1.06] | 1.01 [0.99-1.02] | 1.00 [0.99-1.02] | 0.82 [0.61-1.09] | 1.07 [1.05-1.09] | 60,214 | 60,130 | 58,773 | 189 | 49,844 |
| E13 | 1.27 [1.12-1.44] | 1.19 [1.05-1.35] | 1.24 [1.09-1.41] | | 1.23 [1.09-1.39] | 1.14 [1.01-1.30] | 1.04 [0.92-1.19] | 1.08 [0.94-1.23] | | 1.13 [1.00-1.29] | 766 | 765 | 691 | NA | 750 |
| E14 | 1.17 [1.13-1.22] | 1.18 [1.14-1.22] | 1.18 [1.14-1.22] | | 1.16 [1.12-1.21] | 1.11 [1.07-1.15] | 1.07 [1.03-1.11] | 1.06 [1.02-1.10] | | 1.12 [1.08-1.16] | 9,171 | 9,150 | 8,778 | NA | 8,648 |
| E16 | 1.14 [1.10-1.18] | 1.12 [1.08-1.16] | 1.12 [1.08-1.16] | 0.95 [0.80-1.11] | 1.09 [1.05-1.13] | 1.09 [1.05-1.13] | 1.02 [0.99-1.06] | 1.02 [0.99-1.06] | 0.96 [0.81-1.13] | 1.08 [1.04-1.11] | 10,080 | 9,757 | 9,237 | 449 | 9,884 |
| E20 | 1.30 [1.06-1.60] | 1.27 [1.03-1.56] | 1.39 [1.12-1.72] | | 1.31 [1.07-1.60] | 1.18 [0.96-1.47] | 1.16 [0.93-1.43] | 1.26 [1.01-1.58] | | 1.23 [1.00-1.51] | 286 | 284 | 263 | NA | 286 |
| E21 | 1.06 [1.00-1.13] | 1.05 [0.99-1.12] | 1.06 [0.99-1.13] | | 1.03 [0.97-1.10] | 1.03 [0.97-1.10] | 1.00 [0.94-1.06] | 1.01 [0.94-1.08] | | 1.01 [0.95-1.08] | 2,849 | 2,860 | 2,785 | NA | 2,762 |
| E22 | 1.38 [1.28-1.49] | 1.38 [1.28-1.49] | 1.41 [1.30-1.52] | | 1.38 [1.28-1.49] | 1.33 [1.23-1.44] | 1.31 [1.22-1.42] | 1.34 [1.24-1.45] | | 1.34 [1.24-1.44] | 2,225 | 2,199 | 2,061 | NA | 2,120 |
| E23 | 1.19 [1.06-1.33] | 1.28 [1.13-1.44] | 1.32 [1.16-1.50] | 1.01 [0.72-1.42] | 1.24 [1.11-1.39] | 1.12 [1.00-1.26] | 1.17 [1.04-1.32] | 1.24 [1.09-1.41] | 0.96 [0.68-1.36] | 1.20 [1.07-1.35] | 932 | 844 | 714 | 117 | 890 |
| E24 | 1.31 [1.05-1.64] | 1.13 [0.91-1.42] | 1.38 [1.09-1.75] | | 1.23 [0.99-1.53] | 1.13 [0.90-1.43] | 0.90 [0.71-1.14] | 1.13 [0.87-1.45] | | 1.09 [0.87-1.37] | 245 | 236 | 199 | NA | 244 |
| E26 | 1.03 [0.76-1.39] | 0.98 [0.72-1.33] | 0.99 [0.72-1.36] | | 1.11 [0.83-1.49] | 0.99 [0.73-1.35] | 0.93 [0.68-1.27] | 0.89 [0.64-1.24] | | 1.11 [0.82-1.49] | 126 | 121 | 108 | NA | 121 |
| E27 | 1.32 [1.23-1.43] | 1.30 [1.20-1.40] | 1.29 [1.19-1.39] | 1.30 [0.90-1.87] | 1.26 [1.17-1.36] | 1.22 [1.13-1.32] | 1.16 [1.07-1.25] | 1.17 [1.07-1.26] | 1.18 [0.81-1.72] | 1.18 [1.10-1.28] | 2,178 | 2,118 | 1,877 | 124 | 2,151 |
| E28 | 1.10 [1.05-1.16] | 1.13 [1.08-1.19] | 1.17 [1.03-1.32] | 0.94 [0.83-1.06] | 1.11 [1.05-1.17] | 1.03 [0.98-1.09] | 1.02 [0.97-1.07] | 1.01 [0.89-1.15] | 0.95 [0.84-1.08] | 1.05 [1.00-1.11] | 4,613 | 5,088 | 734 | 906 | 4,090 |
| E29 | 1.43 [1.17-1.76] | 1.37 [1.12-1.68] | 1.41 [1.14-1.73] | | 1.27 [1.04-1.55] | 1.25 [1.01-1.55] | 1.15 [0.93-1.43] | 1.18 [0.95-1.47] | | 1.17 [0.95-1.44] | 303 | 299 | 286 | NA | 294 |
| E30 | 1.29 [0.96-1.72] | | | 1.11 [0.82-1.49] | 1.25 [0.92-1.69] | 1.25 [0.92-1.69] | | | 1.07 [0.78-1.46] | 1.21 [0.89-1.65] | 187 | NA | NA | 172 | 172 |
| E34 | 1.06 [0.90-1.23] | 1.13 [0.92-1.38] | 1.21 [0.95-1.55] | 1.07 [0.86-1.34] | 1.12 [0.96-1.32] | 0.99 [0.85-1.17] | 0.98 [0.79-1.21] | 1.14 [0.88-1.46] | 1.03 [0.82-1.29] | 1.08 [0.92-1.27] | 531 | 279 | 186 | 276 | 486 |
| E43 | 1.24 [0.90-1.72] | 0.97 [0.71-1.34] | | | 1.04 [0.75-1.43] | 1.21 [0.86-1.68] | 0.92 [0.67-1.27] | | | 1.07 [0.77-1.48] | 111 | 112 | NA | NA | 112 |
| E46 | 1.19 [1.08-1.30] | 1.13 [1.03-1.24] | 1.15 [1.04-1.26] | | 1.15 [1.05-1.25] | 1.17 [1.07-1.29] | 1.08 [0.99-1.19] | 1.10 [1.01-1.21] | | 1.15 [1.05-1.26] | 1,485 | 1,468 | 1,370 | NA | 1,450 |
| E51 | 1.42 [1.07-1.88] | 1.18 [0.89-1.57] | 1.42 [1.07-1.88] | | 1.34 [1.01-1.78] | 1.38 [1.03-1.84] | 1.09 [0.82-1.46] | 1.30 [0.97-1.74] | | 1.40 [1.05-1.86] | 152 | 144 | 152 | NA | 143 |
| E53 | 1.19 [1.15-1.23] | 1.18 [1.14-1.22] | 1.17 [1.13-1.22] | 1.11 [0.87-1.42] | 1.18 [1.14-1.22] | 1.14 [1.10-1.19] | 1.10 [1.06-1.14] | 1.10 [1.06-1.14] | 1.07 [0.82-1.39] | 1.15 [1.11-1.19] | 9,508 | 9,630 | 8,639 | 233 | 9,356 |
| E55 | 1.23 [1.19-1.27] | 1.21 [1.18-1.25] | 1.22 [1.18-1.26] | 1.19 [1.02-1.39] | 1.20 [1.16-1.24] | 1.16 [1.12-1.20] | 1.12 [1.08-1.16] | 1.12 [1.08-1.16] | 1.13 [0.96-1.33] | 1.15 [1.12-1.19] | 13,124 | 12,882 | 11,410 | 648 | 12,776 |
| E56 | 1.14 [0.83-1.57] | 1.10 [0.79-1.53] | | | 1.05 [0.77-1.44] | 1.08 [0.78-1.50] | 1.01 [0.72-1.42] | | | 1.05 [0.77-1.43] | 117 | 109 | NA | NA | 112 |
| E58 | 1.36 [0.97-1.90] | | | | 1.45 [1.02-2.05] | 1.31 [0.93-1.85] | | | | 1.43 [1.01-2.03] | 106 | NA | NA | NA | 102 |
| E61 | 1.22 [1.12-1.32] | 1.19 [1.10-1.29] | 1.21 [1.11-1.32] | 1.16 [0.86-1.58] | 1.18 [1.09-1.27] | 1.14 [1.05-1.24] | 1.09 [1.00-1.19] | 1.10 [1.01-1.21] | 1.08 [0.79-1.50] | 1.13 [1.04-1.22] | 1,962 | 1,918 | 1,636 | 151 | 1,931 |
| E63 | 1.35 [0.98-1.87] | 1.19 [0.86-1.65] | 1.33 [0.94-1.88] | | 1.35 [0.98-1.86] | 1.34 [0.96-1.87] | 1.12 [0.80-1.57] | 1.26 [0.89-1.80] | | 1.30 [0.94-1.80] | 122 | 116 | 109 | NA | 117 |
| E66 | 1.18 [1.16-1.20] | 1.19 [1.18-1.21] | 1.19 [1.17-1.21] | 1.05 [0.99-1.11] | 1.16 [1.14-1.17] | 1.11 [1.09-1.12] | 1.08 [1.06-1.10] | 1.07 [1.06-1.09] | 1.04 [0.97-1.11] | 1.10 [1.09-1.12] | 61,881 | 63,421 | 46,219 | 3,733 | 58,549 |
| E72 | 1.24 [0.90-1.70] | 1.31 [0.95-1.82] | | | 1.32 [0.96-1.81] | 1.18 [0.86-1.64] | 1.21 [0.87-1.70] | | | 1.35 [0.98-1.86] | 118 | 107 | NA | NA | 115 |
| E73 | 1.25 [1.08-1.45] | 1.21 [1.02-1.43] | 1.18 [0.96-1.45] | 1.27 [0.98-1.64] | 1.18 [1.02-1.37] | 1.06 [0.91-1.24] | 1.03 [0.86-1.23] | 0.97 [0.78-1.21] | 1.17 [0.89-1.52] | 1.05 [0.90-1.23] | 568 | 410 | 270 | 213 | 519 |
| E74 | 1.04 [0.82-1.32] | 1.05 [0.83-1.34] | 1.00 [0.76-1.30] | | 1.13 [0.89-1.43] | 1.00 [0.79-1.28] | 0.99 [0.78-1.27] | 0.95 [0.72-1.25] | | 1.08 [0.85-1.37] | 202 | 184 | 150 | NA | 196 |
| E78 | 1.09 [1.07-1.10] | 1.08 [1.07-1.10] | 1.09 [1.07-1.10] | | 1.07 [1.06-1.09] | 1.06 [1.04-1.07] | 1.02 [1.01-1.04] | 1.02 [1.01-1.04] | | 1.06 [1.05-1.07] | 77,578 | 77,625 | 76,760 | NA | 69,482 |
| E79 | 1.16 [0.84-1.61] | 1.14 [0.82-1.60] | | | | 1.11 [0.79-1.55] | 1.07 [0.76-1.51] | | | | 107 | 104 | NA | NA | NA |
| E80 | 1.08 [0.96-1.21] | 1.10 [0.98-1.24] | 1.09 [0.95-1.25] | | 1.01 [0.89-1.14] | 1.04 [0.92-1.17] | 1.06 [0.94-1.20] | 1.06 [0.93-1.22] | | 0.98 [0.87-1.11] | 804 | 786 | 608 | NA | 747 |
| E83 | 1.12 [1.08-1.15] | 1.12 [1.09-1.15] | 1.12 [1.09-1.15] | 0.94 [0.74-1.20] | 1.10 [1.07-1.13] | 1.09 [1.05-1.12] | 1.06 [1.03-1.09] | 1.06 [1.03-1.09] | 0.92 [0.72-1.18] | 1.09 [1.06-1.12] | 13,585 | 13,541 | 12,929 | 228 | 13,207 |
| E85 | 1.15 [0.99-1.34] | 1.13 [0.97-1.31] | 1.10 [0.94-1.28] | | 1.08 [0.93-1.26] | 1.10 [0.95-1.29] | 1.06 [0.91-1.24] | 1.02 [0.88-1.20] | | 1.04 [0.89-1.22] | 493 | 482 | 470 | NA | 473 |
| E86 | 1.12 [1.10-1.15] | 1.12 [1.10-1.14] | 1.12 [1.10-1.14] | 1.05 [0.95-1.16] | 1.11 [1.09-1.13] | 1.11 [1.09-1.13] | 1.07 [1.05-1.09] | 1.07 [1.05-1.09] | 1.03 [0.93-1.14] | 1.10 [1.08-1.12] | 35,956 | 35,245 | 33,527 | 1,281 | 34,906 |
| E87 | 1.20 [1.18-1.22] | 1.20 [1.18-1.21] | 1.20 [1.18-1.21] | 0.98 [0.86-1.11] | 1.18 [1.16-1.20] | 1.17 [1.15-1.18] | 1.13 [1.11-1.15] | 1.13 [1.11-1.15] | 0.97 [0.84-1.11] | 1.16 [1.14-1.18] | 58,707 | 58,443 | 56,881 | 764 | 56,673 |
| E88 | 1.22 [1.14-1.30] | 1.19 [1.11-1.28] | 1.23 [1.14-1.32] | 1.13 [0.80-1.59] | 1.18 [1.10-1.26] | 1.18 [1.10-1.27] | 1.13 [1.06-1.22] | 1.16 [1.08-1.25] | 1.04 [0.73-1.49] | 1.16 [1.08-1.24] | 2,559 | 2,509 | 2,353 | 112 | 2,518 |
| E89 | 1.12 [1.06-1.19] | 1.12 [1.06-1.19] | 1.13 [1.07-1.20] | | 1.09 [1.03-1.15] | 1.08 [1.02-1.14] | 1.05 [0.99-1.11] | 1.06 [0.99-1.12] | | 1.05 [0.99-1.12] | 3,465 | 3,453 | 3,140 | NA | 3,350 |
| Mental, Behavioral and Neurodevelopmental disorders (F01-F99) | | | | | | | | | | | | | | | |
| F00 | 1.02 [0.98-1.06] | 1.02 [0.98-1.06] | 1.03 [0.99-1.06] | | 1.04 [1.00-1.08] | 1.03 [0.99-1.07] | 1.01 [0.98-1.05] | 1.02 [0.98-1.06] | | 1.04 [1.00-1.08] | 8,435 | 8,435 | 8,431 | NA | 8,103 |
| F01 | 1.04 [1.01-1.08] | 1.03 [1.00-1.07] | 1.03 [1.00-1.07] | | 1.04 [1.00-1.07] | 1.04 [1.00-1.07] | 1.00 [0.97-1.04] | 1.00 [0.97-1.04] | | 1.04 [1.00-1.07] | 10,012 | 10,023 | 10,004 | NA | 9,758 |
| F02 | 0.97 [0.89-1.06] | 0.95 [0.87-1.03] | 0.95 [0.87-1.03] | | 0.92 [0.85-1.00] | 0.99 [0.91-1.08] | 0.95 [0.87-1.03] | 0.95 [0.87-1.04] | | 0.94 [0.87-1.03] | 1,536 | 1,537 | 1,535 | NA | 1,471 |
| F03 | 1.03 [1.01-1.05] | 1.02 [1.00-1.04] | 1.03 [1.00-1.05] | | 1.03 [1.00-1.05] | 1.03 [1.00-1.05] | 1.01 [0.98-1.03] | 1.01 [0.99-1.04] | | 1.03 [1.01-1.06] | 23,316 | 23,290 | 23,245 | NA | 22,287 |
| F05 | 1.11 [1.09-1.14] | 1.11 [1.09-1.14] | 1.10 [1.08-1.13] | | 1.11 [1.08-1.13] | 1.10 [1.07-1.12] | 1.07 [1.04-1.10] | 1.06 [1.04-1.09] | | 1.10 [1.08-1.13] | 22,934 | 22,916 | 22,832 | NA | 22,664 |
| F06 | 1.11 [1.05-1.18] | 1.09 [1.03-1.16] | 1.11 [1.04-1.18] | | 1.06 [1.00-1.13] | 1.09 [1.03-1.16] | 1.05 [0.99-1.12] | 1.07 [1.00-1.13] | | 1.06 [1.00-1.13] | 3,455 | 3,439 | 3,408 | NA | 3,371 |
| F07 | 1.16 [0.91-1.49] | 1.24 [0.96-1.59] | 1.37 [1.04-1.81] | | 1.07 [0.83-1.38] | 1.07 [0.83-1.39] | 1.16 [0.89-1.50] | 1.30 [0.98-1.73] | | 1.05 [0.81-1.36] | 190 | 184 | 149 | NA | 173 |
| F10 | 1.18 [1.14-1.21] | 1.17 [1.14-1.21] | 1.20 [1.16-1.24] | 0.92 [0.80-1.05] | 1.16 [1.13-1.20] | 1.16 [1.12-1.19] | 1.10 [1.06-1.13] | 1.13 [1.09-1.17] | 0.93 [0.80-1.07] | 1.17 [1.14-1.21] | 13,285 | 13,280 | 11,000 | 701 | 11,863 |
| F11 | 1.19 [1.06-1.34] | 1.12 [1.00-1.26] | 1.18 [1.02-1.37] | | 1.13 [1.01-1.27] | 1.11 [0.99-1.26] | 0.94 [0.83-1.06] | 1.02 [0.88-1.19] | | 1.11 [0.98-1.25] | 878 | 901 | 518 | NA | 830 |
| F12 | 1.09 [0.99-1.19] | 1.06 [0.97-1.16] | 1.14 [0.99-1.30] | 0.93 [0.77-1.13] | 0.98 [0.90-1.08] | 1.02 [0.92-1.12] | 0.92 [0.84-1.01] | 1.00 [0.87-1.16] | 0.96 [0.78-1.18] | 0.98 [0.89-1.08] | 1,468 | 1,526 | 635 | 369 | 1,346 |
| F13 | 1.13 [0.91-1.42] | 1.09 [0.87-1.36] | 1.31 [1.01-1.68] | | 1.02 [0.82-1.28] | 1.06 [0.84-1.34] | 0.91 [0.72-1.16] | 1.10 [0.84-1.44] | | 0.99 [0.79-1.24] | 241 | 239 | 187 | NA | 230 |
| F14 | 1.07 [0.93-1.23] | 1.06 [0.92-1.22] | 1.17 [0.93-1.46] | 0.82 [0.60-1.14] | 0.98 [0.85-1.13] | 1.01 [0.87-1.18] | 0.91 [0.78-1.05] | 1.00 [0.79-1.27] | 0.93 [0.66-1.31] | 0.99 [0.85-1.14] | 621 | 642 | 234 | 127 | 574 |
| F15 | 1.07 [0.84-1.37] | 0.92 [0.72-1.17] | | | 0.96 [0.75-1.22] | 1.01 [0.79-1.30] | 0.86 [0.67-1.10] | | | 0.96 [0.75-1.23] | 207 | 195 | NA | NA | 188 |
| F17 | 1.08 [1.07-1.10] | 1.08 [1.07-1.10] | 1.09 [1.08-1.11] | 0.92 [0.87-0.96] | 1.05 [1.04-1.07] | 1.08 [1.06-1.09] | 0.99 [0.98-1.00] | 1.02 [1.00-1.04] | 0.98 [0.93-1.04] | 1.08 [1.07-1.10] | 63,060 | 65,557 | 43,722 | 5,535 | 57,665 |
| F19 | 0.98 [0.86-1.12] | 0.98 [0.86-1.11] | 1.01 [0.84-1.21] | 0.70 [0.49-1.00] | 0.93 [0.82-1.05] | 0.90 [0.79-1.03] | 0.83 [0.72-0.95] | 0.86 [0.71-1.05] | 0.71 [0.49-1.04] | 0.93 [0.82-1.07] | 693 | 713 | 325 | 104 | 679 |
| F20 | 1.14 [1.07-1.23] | 1.15 [1.07-1.24] | 1.21 [1.12-1.31] | | 1.14 [1.06-1.23] | 1.16 [1.07-1.24] | 1.12 [1.05-1.21] | 1.21 [1.12-1.31] | | 1.18 [1.09-1.27] | 2,229 | 2,264 | 1,868 | NA | 1,949 |
| F22 | 1.27 [1.14-1.40] | 1.23 [1.11-1.36] | 1.24 [1.11-1.38] | | 1.20 [1.08-1.33] | 1.24 [1.11-1.37] | 1.15 [1.04-1.28] | 1.16 [1.04-1.30] | | 1.19 [1.07-1.32] | 1,115 | 1,104 | 983 | NA | 1,044 |
| F23 | 1.17 [1.01-1.36] | 1.15 [0.99-1.34] | 1.25 [1.04-1.49] | | 1.10 [0.94-1.29] | 1.13 [0.97-1.32] | 1.09 [0.94-1.28] | 1.21 [1.01-1.45] | | 1.11 [0.94-1.30] | 525 | 522 | 360 | NA | 451 |
| F25 | 1.24 [1.07-1.45] | 1.24 [1.07-1.44] | 1.26 [1.06-1.49] | | 1.10 [0.95-1.27] | 1.26 [1.08-1.48] | 1.18 [1.01-1.38] | 1.25 [1.05-1.48] | | 1.16 [1.00-1.35] | 530 | 532 | 384 | NA | 507 |
| F29 | 1.27 [1.14-1.40] | 1.26 [1.14-1.40] | 1.30 [1.15-1.46] | | 1.12 [1.01-1.24] | 1.21 [1.09-1.35] | 1.15 [1.03-1.28] | 1.21 [1.07-1.36] | | 1.11 [0.99-1.23] | 1,133 | 1,157 | 847 | NA | 1,059 |
| F30 | 1.41 [1.17-1.72] | 1.31 [1.08-1.59] | 1.45 [1.18-1.79] | | 1.34 [1.10-1.63] | 1.38 [1.14-1.68] | 1.23 [1.01-1.50] | 1.39 [1.13-1.72] | | 1.34 [1.10-1.64] | 327 | 325 | 271 | NA | 294 |
| F31 | 1.16 [1.09-1.24] | 1.16 [1.09-1.23] | 1.17 [1.09-1.26] | 0.87 [0.68-1.12] | 1.09 [1.03-1.16] | 1.08 [1.01-1.16] | 1.00 [0.94-1.07] | 1.06 [0.98-1.14] | 0.87 [0.67-1.13] | 1.07 [1.00-1.14] | 2,932 | 3,013 | 2,046 | 201 | 2,779 |
| F32 | 1.24 [1.22-1.26] | 1.23 [1.22-1.25] | 1.24 [1.22-1.26] | 0.97 [0.91-1.03] | 1.20 [1.18-1.22] | 1.14 [1.12-1.16] | 1.06 [1.04-1.08] | 1.08 [1.06-1.10] | 1.00 [0.93-1.06] | 1.15 [1.13-1.16] | 56,319 | 57,470 | 43,384 | 3,400 | 53,486 |
| F33 | 1.15 [1.06-1.25] | 1.19 [1.09-1.29] | 1.09 [1.00-1.19] | | 1.12 [1.03-1.22] | 1.07 [0.98-1.16] | 1.03 [0.95-1.13] | 0.96 [0.87-1.05] | | 1.09 [1.00-1.19] | 1,658 | 1,709 | 1,335 | NA | 1,580 |
| F34 | 1.22 [0.99-1.50] | 1.25 [1.01-1.55] | 1.31 [1.04-1.67] | | 1.16 [0.94-1.42] | 1.09 [0.87-1.35] | 1.06 [0.85-1.32] | 1.12 [0.88-1.44] | | 1.06 [0.86-1.31] | 277 | 282 | 214 | NA | 260 |
| F38 | 1.15 [0.84-1.58] | 1.16 [0.86-1.58] | | | 1.17 [0.87-1.58] | 1.05 [0.76-1.46] | 1.07 [0.78-1.47] | | | 1.14 [0.84-1.55] | 123 | 125 | NA | NA | 127 |
| F39 | 1.17 [0.93-1.49] | 1.09 [0.87-1.38] | 1.27 [0.98-1.64] | | 1.16 [0.93-1.46] | 1.09 [0.86-1.39] | 0.97 [0.76-1.23] | 1.13 [0.86-1.47] | | 1.11 [0.88-1.39] | 224 | 221 | 178 | NA | 227 |
| F40 | 1.19 [1.13-1.25] | 1.18 [1.12-1.25] | 1.22 [1.15-1.30] | 0.98 [0.83-1.15] | 1.16 [1.10-1.22] | 1.11 [1.06-1.17] | 1.07 [1.01-1.13] | 1.11 [1.05-1.18] | 0.96 [0.81-1.13] | 1.11 [1.05-1.17] | 4,691 | 4,579 | 3,298 | 557 | 4,352 |
| F41 | 1.22 [1.20-1.24] | 1.21 [1.19-1.23] | 1.23 [1.21-1.26] | 0.97 [0.91-1.03] | 1.18 [1.16-1.20] | 1.11 [1.10-1.13] | 1.05 [1.03-1.06] | 1.08 [1.06-1.10] | 0.98 [0.92-1.05] | 1.12 [1.10-1.14] | 46,858 | 47,954 | 34,383 | 3,904 | 44,519 |
| F42 | 1.53 [1.38-1.70] | 1.51 [1.36-1.67] | 1.68 [1.45-1.94] | 0.92 [0.69-1.21] | 1.43 [1.29-1.58] | 1.36 [1.22-1.52] | 1.25 [1.12-1.40] | 1.47 [1.26-1.70] | 0.83 [0.61-1.11] | 1.32 [1.18-1.47] | 1,227 | 1,234 | 650 | 177 | 1,147 |
| F43 | 1.17 [1.09-1.26] | 1.15 [1.08-1.24] | 1.19 [1.08-1.30] | 0.85 [0.69-1.04] | 1.10 [1.02-1.18] | 1.04 [0.96-1.12] | 0.94 [0.87-1.01] | 0.99 [0.90-1.10] | 0.87 [0.70-1.09] | 1.04 [0.97-1.12] | 2,340 | 2,405 | 1,384 | 282 | 2,271 |
| F44 | 1.23 [1.07-1.41] | 1.23 [1.07-1.42] | 1.28 [1.07-1.52] | | 1.13 [0.98-1.29] | 1.10 [0.95-1.27] | 1.04 [0.90-1.20] | 1.06 [0.89-1.28] | | 1.05 [0.92-1.21] | 632 | 642 | 402 | NA | 627 |
| F45 | 1.30 [1.15-1.47] | 1.28 [1.13-1.44] | 1.20 [1.05-1.38] | | 1.22 [1.08-1.38] | 1.16 [1.02-1.32] | 1.10 [0.97-1.26] | 1.06 [0.92-1.22] | | 1.12 [0.99-1.27] | 808 | 780 | 608 | NA | 756 |
| F48 | 1.45 [1.14-1.85] | 1.26 [0.98-1.60] | 1.19 [0.89-1.58] | | 1.08 [0.85-1.38] | 1.32 [1.03-1.69] | 1.07 [0.82-1.38] | 1.00 [0.74-1.36] | | 0.98 [0.76-1.27] | 215 | 204 | 144 | NA | 188 |
| F50 | 1.20 [1.07-1.35] | 1.19 [1.06-1.34] | 1.28 [1.06-1.55] | 0.92 [0.74-1.14] | 1.06 [0.95-1.20] | 1.10 [0.97-1.24] | 1.03 [0.92-1.17] | 1.14 [0.94-1.39] | 0.95 [0.76-1.19] | 1.03 [0.91-1.16] | 943 | 912 | 327 | 293 | 834 |
| F51 | 1.29 [1.03-1.63] | 1.36 [1.07-1.74] | 1.26 [0.95-1.67] | | 1.32 [1.05-1.66] | 1.18 [0.93-1.50] | 1.22 [0.95-1.57] | 1.10 [0.82-1.48] | | 1.25 [0.99-1.58] | 240 | 210 | 153 | NA | 237 |
| F52 | 1.18 [1.00-1.38] | 1.14 [0.97-1.34] | 1.21 [1.03-1.43] | | 1.17 [1.00-1.38] | 1.11 [0.94-1.31] | 1.04 [0.88-1.22] | 1.09 [0.92-1.29] | | 1.13 [0.96-1.33] | 445 | 454 | 430 | NA | 435 |
| F53 | 1.29 [1.10-1.52] | 1.27 [1.08-1.49] | | | 1.23 [1.06-1.43] | 1.23 [1.04-1.45] | 1.12 [0.95-1.32] | | | 1.18 [1.01-1.39] | 468 | 499 | NA | NA | 480 |
| F60 | 1.29 [1.19-1.39] | 1.23 [1.14-1.33] | 1.38 [1.24-1.53] | 0.87 [0.72-1.06] | 1.16 [1.08-1.25] | 1.14 [1.05-1.23] | 0.98 [0.90-1.06] | 1.16 [1.04-1.30] | 0.93 [0.76-1.13] | 1.11 [1.02-1.19] | 2,140 | 2,237 | 1,035 | 348 | 2,106 |
| F69 | 1.21 [0.91-1.60] | 1.39 [1.04-1.84] | 1.57 [1.12-2.21] | | 1.24 [0.94-1.62] | 1.10 [0.82-1.47] | 1.15 [0.85-1.55] | 1.32 [0.92-1.89] | | 1.18 [0.89-1.56] | 157 | 150 | 104 | NA | 155 |
| F70 | 1.59 [1.31-1.94] | 1.78 [1.45-2.18] | 2.06 [1.63-2.61] | | 1.55 [1.27-1.89] | 1.47 [1.20-1.81] | 1.63 [1.32-2.02] | 1.83 [1.43-2.33] | | 1.44 [1.17-1.78] | 363 | 335 | 246 | NA | 325 |
| F71 | 2.01 [1.45-2.77] | 2.05 [1.48-2.85] | | | 2.06 [1.48-2.87] | 1.90 [1.36-2.64] | 2.02 [1.44-2.84] | | | 2.08 [1.47-2.94] | 149 | 140 | NA | NA | 130 |
| F72 | 1.78 [1.37-2.29] | 1.81 [1.38-2.37] | 2.37 [1.71-3.28] | | 1.46 [1.14-1.88] | 1.69 [1.30-2.21] | 1.93 [1.45-2.56] | 2.24 [1.60-3.14] | | 1.44 [1.11-1.86] | 220 | 195 | 143 | NA | 213 |
| F79 | 1.55 [1.40-1.71] | 1.72 [1.55-1.91] | 1.82 [1.62-2.05] | 0.86 [0.68-1.08] | 1.50 [1.36-1.65] | 1.49 [1.34-1.64] | 1.66 [1.49-1.85] | 1.76 [1.56-1.99] | 0.86 [0.68-1.08] | 1.44 [1.30-1.60] | 1,442 | 1,274 | 985 | 264 | 1,353 |
| F80 | 1.19 [1.08-1.33] | 1.31 [1.12-1.52] | 1.28 [1.10-1.50] | 1.11 [0.96-1.28] | 1.16 [1.04-1.29] | 1.16 [1.04-1.29] | 1.24 [1.07-1.45] | 1.22 [1.04-1.44] | 1.08 [0.93-1.25] | 1.15 [1.03-1.28] | 1,149 | 511 | 488 | 651 | 1,097 |
| F81 | 1.54 [1.45-1.64] | 1.66 [1.56-1.78] | 1.87 [1.74-2.02] | 0.97 [0.85-1.10] | 1.45 [1.36-1.55] | 1.47 [1.38-1.57] | 1.63 [1.53-1.75] | 1.82 [1.69-1.97] | 0.96 [0.84-1.09] | 1.41 [1.32-1.51] | 3,760 | 3,214 | 2,399 | 879 | 3,253 |
| F82 | 0.96 [0.69-1.33] | | | | 1.06 [0.76-1.49] | 0.92 [0.66-1.28] | | | | 1.01 [0.72-1.44] | 117 | NA | NA | NA | 113 |
| F84 | 1.36 [1.26-1.48] | 1.52 [1.36-1.70] | 1.92 [1.62-2.27] | 1.14 [1.03-1.26] | 1.29 [1.18-1.40] | 1.24 [1.13-1.35] | 1.37 [1.22-1.54] | 1.70 [1.43-2.03] | 1.10 [0.99-1.22] | 1.21 [1.11-1.32] | 2,261 | 1,118 | 492 | 1,500 | 2,041 |
| F89 | 0.98 [0.84-1.14] | | | 0.91 [0.77-1.07] | 0.93 [0.80-1.08] | 0.92 [0.78-1.07] | | | 0.87 [0.74-1.03] | 0.88 [0.75-1.03] | 545 | NA | NA | 486 | 542 |
| F90 | 1.10 [0.98-1.24] | 1.17 [0.99-1.37] | 1.58 [1.10-2.26] | 0.98 [0.85-1.12] | 1.11 [0.98-1.25] | 1.04 [0.91-1.18] | 1.03 [0.87-1.23] | 1.20 [0.81-1.79] | 0.98 [0.85-1.14] | 1.10 [0.97-1.25] | 990 | 516 | 103 | 727 | 916 |
| F91 | 1.10 [0.86-1.40] | | | 1.21 [0.87-1.69] | 1.14 [0.89-1.46] | 1.06 [0.83-1.36] | | | 1.18 [0.84-1.66] | 1.14 [0.89-1.46] | 219 | NA | NA | 136 | 209 |
| F95 | 1.18 [0.85-1.63] | | | | 1.23 [0.88-1.71] | 1.04 [0.74-1.46] | | | | 1.14 [0.81-1.61] | 138 | NA | NA | NA | 131 |
| F98 | 1.05 [0.78-1.41] | | | 1.13 [0.78-1.64] | 1.03 [0.76-1.40] | 0.95 [0.69-1.30] | | | 1.08 [0.73-1.59] | 1.00 [0.73-1.37] | 151 | NA | NA | 107 | 148 |
| F99 | 1.11 [0.98-1.26] | 1.15 [1.02-1.30] | 1.17 [1.01-1.36] | | 1.11 [0.98-1.25] | 1.05 [0.92-1.20] | 1.01 [0.89-1.16] | 1.07 [0.92-1.24] | | 1.08 [0.95-1.23] | 752 | 766 | 516 | NA | 734 |
| Diseases of the nervous system (G00-G99) | | | | | | | | | | | | | | | |
| G00 | 1.06 [0.83-1.35] | 1.07 [0.83-1.39] | 1.05 [0.79-1.41] | | 0.99 [0.76-1.28] | 1.01 [0.78-1.30] | 1.01 [0.77-1.32] | 1.00 [0.74-1.34] | | 0.96 [0.74-1.26] | 186 | 169 | 125 | NA | 160 |
| G03 | 0.99 [0.79-1.24] | 1.01 [0.80-1.28] | 0.99 [0.76-1.30] | | 0.91 [0.72-1.14] | 0.94 [0.74-1.18] | 0.92 [0.72-1.17] | 0.91 [0.69-1.21] | | 0.86 [0.68-1.08] | 233 | 204 | 145 | NA | 200 |

### Supplementary Table 4: Hazard ratios and events from all cohorts (excluding non-consulters)

| | Hazard ratio (99% confidence interval) | | | | | | | | | | Events (in exposed) | | | | |
|---|---|---|---|---|---|---|---|---|---|---|---|---|---|---|---|
| | crude | | | | | adjusted | | | | | | | | | |
| Outcome | any age | 18+ | 40+ | <18 | hosp. | any age | 18+ | 40+ | <18 | hosp. | any age | 18+ | 40+ | <18 | hosp. |
| G04 | 1.17 [1.02-1.34] | 1.16 [1.01-1.34] | 1.19 [1.03-1.38] | | 1.09 [0.95-1.25] | 1.15 [1.00-1.32] | 1.09 [0.94-1.26] | 1.13 [0.97-1.31] | | 1.07 [0.93-1.23] | 628 | 586 | 514 | NA | 583 |
| G05 | 1.39 [1.07-1.81] | 1.48 [1.13-1.94] | 1.39 [1.05-1.84] | | 1.52 [1.16-1.99] | 1.37 [1.05-1.78] | 1.44 [1.10-1.90] | 1.35 [1.02-1.80] | | 1.45 [1.10-1.91] | 177 | 164 | 150 | NA | 162 |
| G06 | 1.04 [0.84-1.29] | 1.04 [0.83-1.29] | 1.01 [0.81-1.27] | | 1.08 [0.87-1.34] | 1.00 [0.81-1.25] | 1.01 [0.80-1.26] | 0.99 [0.79-1.24] | | 1.08 [0.87-1.34] | 243 | 227 | 211 | NA | 235 |
| G08 | 1.12 [0.83-1.51] | 1.35 [1.00-1.82] | | | 1.19 [0.88-1.63] | 1.05 [0.77-1.43] | 1.31 [0.96-1.78] | | | 1.19 [0.87-1.62] | 130 | 135 | NA | NA | 115 |
| G09 | 1.19 [0.91-1.56] | 1.18 [0.90-1.56] | 1.10 [0.82-1.48] | | 1.15 [0.89-1.50] | 1.15 [0.87-1.52] | 1.13 [0.85-1.50] | 1.06 [0.78-1.43] | | 1.10 [0.84-1.43] | 160 | 154 | 127 | NA | 163 |
| G11 | 1.06 [0.86-1.29] | 1.16 [0.95-1.43] | 1.16 [0.94-1.44] | | 1.15 [0.94-1.40] | 1.04 [0.85-1.28] | 1.10 [0.90-1.36] | 1.08 [0.87-1.35] | | 1.15 [0.94-1.40] | 287 | 268 | 241 | NA | 278 |
| G12 | 1.08 [0.95-1.24] | 1.12 [0.98-1.28] | 1.05 [0.92-1.20] | | 1.07 [0.93-1.22] | 1.06 [0.93-1.21] | 1.09 [0.96-1.25] | 1.03 [0.90-1.18] | | 1.06 [0.92-1.21] | 627 | 620 | 598 | NA | 554 |
| G20 | 1.01 [0.97-1.06] | 1.00 [0.96-1.04] | 1.02 [0.97-1.06] | | 1.01 [0.96-1.05] | 1.02 [0.97-1.06] | 0.98 [0.94-1.03] | 1.01 [0.96-1.05] | | 1.01 [0.97-1.06] | 5,956 | 5,962 | 5,945 | NA | 5,377 |
| G21 | 1.26 [1.09-1.44] | 1.21 [1.05-1.38] | 1.24 [1.08-1.43] | | 1.19 [1.04-1.37] | 1.26 [1.09-1.44] | 1.17 [1.02-1.34] | 1.22 [1.06-1.40] | | 1.21 [1.06-1.39] | 646 | 648 | 634 | NA | 632 |
| G23 | 1.08 [0.91-1.30] | 1.05 [0.88-1.26] | 1.04 [0.87-1.25] | | 1.03 [0.86-1.23] | 1.08 [0.90-1.29] | 1.05 [0.88-1.26] | 1.02 [0.85-1.23] | | 1.02 [0.85-1.23] | 350 | 343 | 337 | NA | 334 |
| G24 | 1.12 [0.98-1.28] | 1.15 [1.00-1.31] | 1.11 [0.96-1.28] | | 1.11 [0.97-1.26] | 1.09 [0.95-1.25] | 1.07 [0.93-1.23] | 1.06 [0.91-1.22] | | 1.09 [0.96-1.25] | 657 | 613 | 538 | NA | 621 |
| G25 | 1.17 [1.11-1.24] | 1.18 [1.11-1.25] | 1.16 [1.10-1.23] | | 1.15 [1.09-1.21] | 1.08 [1.02-1.15] | 1.05 [0.99-1.12] | 1.05 [0.99-1.12] | | 1.09 [1.03-1.15] | 3,742 | 3,708 | 3,508 | NA | 3,700 |
| G30 | 1.02 [0.99-1.06] | 1.03 [0.99-1.06] | 1.03 [1.00-1.07] | | 1.04 [1.01-1.07] | 1.02 [0.99-1.06] | 1.01 [0.98-1.05] | 1.02 [0.99-1.06] | | 1.04 [1.00-1.07] | 11,206 | 11,223 | 11,212 | NA | 10,627 |
| G31 | 1.11 [1.06-1.15] | 1.11 [1.07-1.15] | 1.09 [1.05-1.14] | | 1.09 [1.05-1.13] | 1.09 [1.05-1.13] | 1.07 [1.03-1.11] | 1.06 [1.02-1.10] | | 1.09 [1.04-1.13] | 8,278 | 8,257 | 8,155 | NA | 7,918 |
| G32 | 1.27 [0.93-1.74] | 1.13 [0.83-1.55] | 1.42 [1.03-1.97] | | 1.27 [0.93-1.74] | 1.22 [0.88-1.69] | 1.05 [0.77-1.45] | 1.33 [0.95-1.85] | | 1.30 [0.94-1.78] | 120 | 116 | 116 | NA | 114 |
| G35 | 1.13 [1.03-1.23] | 1.12 [1.02-1.22] | 1.15 [1.04-1.27] | | 1.10 [1.00-1.22] | 1.08 [0.98-1.18] | 1.01 [0.92-1.11] | 1.05 [0.95-1.17] | | 1.10 [1.00-1.21] | 1,393 | 1,413 | 1,119 | NA | 1,181 |
| G37 | 1.18 [0.99-1.40] | 1.13 [0.94-1.34] | 1.30 [1.06-1.61] | | 1.18 [0.99-1.41] | 1.13 [0.95-1.35] | 1.01 [0.84-1.22] | 1.23 [0.99-1.53] | | 1.18 [0.98-1.41] | 384 | 380 | 265 | NA | 355 |
| G40 | 1.21 [1.17-1.25] | 1.22 [1.18-1.26] | 1.24 [1.19-1.28] | 1.03 [0.91-1.15] | 1.16 [1.12-1.20] | 1.14 [1.10-1.18] | 1.11 [1.07-1.15] | 1.12 [1.08-1.16] | 1.04 [0.92-1.17] | 1.13 [1.09-1.17] | 10,243 | 9,698 | 8,141 | 990 | 9,088 |
| G41 | 1.17 [1.03-1.33] | 1.25 [1.09-1.44] | 1.30 [1.12-1.51] | 0.92 [0.67-1.27] | 1.10 [0.97-1.25] | 1.13 [0.99-1.29] | 1.17 [1.01-1.35] | 1.22 [1.05-1.42] | 0.94 [0.68-1.29] | 1.10 [0.97-1.25] | 711 | 610 | 545 | 124 | 713 |
| G43 | 1.18 [1.14-1.22] | 1.17 [1.14-1.22] | 1.19 [1.14-1.24] | 1.01 [0.90-1.13] | 1.13 [1.09-1.17] | 1.06 [1.02-1.10] | 1.00 [0.96-1.04] | 1.02 [0.97-1.07] | 1.00 [0.89-1.12] | 1.05 [1.01-1.08] | 10,276 | 10,449 | 6,311 | 1,111 | 9,772 |
| G44 | 1.18 [1.07-1.29] | 1.14 [1.04-1.25] | 1.25 [1.12-1.39] | 0.89 [0.64-1.23] | 1.17 [1.07-1.28] | 1.08 [0.98-1.19] | 1.02 [0.93-1.12] | 1.11 [0.99-1.24] | 0.90 [0.64-1.26] | 1.10 [1.01-1.21] | 1,442 | 1,449 | 992 | 124 | 1,402 |
| G45 | 1.10 [1.06-1.14] | 1.11 [1.06-1.15] | 1.09 [1.05-1.14] | | 1.07 [1.03-1.11] | 1.07 [1.03-1.11] | 1.06 [1.02-1.10] | 1.04 [1.00-1.08] | | 1.06 [1.02-1.10] | 7,704 | 7,732 | 7,625 | NA | 7,223 |
| G46 | 1.12 [0.96-1.30] | 1.14 [0.98-1.33] | 1.09 [0.94-1.27] | | 1.07 [0.93-1.24] | 1.09 [0.94-1.27] | 1.10 [0.95-1.29] | 1.04 [0.89-1.21] | | 1.06 [0.91-1.23] | 518 | 519 | 506 | NA | 511 |
| G47 | 1.31 [1.27-1.35] | 1.34 [1.30-1.39] | 1.34 [1.29-1.39] | 1.21 [1.12-1.31] | 1.29 [1.26-1.33] | 1.18 [1.15-1.22] | 1.14 [1.11-1.18] | 1.15 [1.11-1.19] | 1.16 [1.07-1.26] | 1.20 [1.16-1.23] | 13,363 | 11,232 | 10,088 | 2,412 | 12,881 |
| G50 | 1.32 [1.20-1.45] | 1.33 [1.21-1.46] | 1.30 [1.18-1.43] | | 1.26 [1.15-1.39] | 1.22 [1.11-1.34] | 1.18 [1.07-1.30] | 1.15 [1.04-1.27] | | 1.19 [1.08-1.30] | 1,397 | 1,402 | 1,282 | NA | 1,315 |
| G51 | 1.15 [1.08-1.23] | 1.18 [1.10-1.27] | 1.14 [1.06-1.22] | 1.23 [0.90-1.67] | 1.13 [1.05-1.21] | 1.10 [1.03-1.18] | 1.10 [1.03-1.19] | 1.07 [0.99-1.15] | 1.20 [0.87-1.66] | 1.10 [1.02-1.18] | 2,495 | 2,419 | 2,162 | 146 | 2,334 |
| G52 | 1.11 [0.86-1.41] | 1.15 [0.90-1.48] | 1.16 [0.89-1.51] | | 1.17 [0.91-1.49] | 1.07 [0.84-1.38] | 1.08 [0.84-1.39] | 1.08 [0.83-1.42] | | 1.13 [0.88-1.45] | 184 | 187 | 167 | NA | 180 |
| G53 | 1.25 [1.07-1.47] | 1.21 [1.03-1.42] | 1.23 [1.04-1.44] | | 1.21 [1.03-1.42] | 1.18 [1.00-1.39] | 1.14 [0.96-1.34] | 1.17 [0.99-1.38] | | 1.16 [0.99-1.36] | 462 | 461 | 440 | NA | 444 |
| G54 | 1.24 [1.09-1.40] | 1.16 [1.03-1.32] | 1.23 [1.08-1.40] | | 1.18 [1.05-1.33] | 1.15 [1.02-1.31] | 1.03 [0.91-1.17] | 1.10 [0.96-1.25] | | 1.13 [1.00-1.28] | 772 | 768 | 660 | NA | 753 |
| G55 | 1.16 [1.11-1.20] | 1.15 [1.11-1.19] | 1.17 [1.13-1.22] | 0.86 [0.63-1.18] | 1.12 [1.08-1.17] | 1.08 [1.04-1.12] | 1.04 [1.00-1.08] | 1.06 [1.01-1.10] | 0.77 [0.55-1.09] | 1.07 [1.03-1.11] | 8,112 | 8,175 | 7,096 | 123 | 7,942 |
| G56 | 1.22 [1.18-1.26] | 1.21 [1.18-1.25] | 1.20 [1.16-1.23] | 1.07 [0.77-1.48] | 1.18 [1.14-1.21] | 1.14 [1.10-1.17] | 1.10 [1.07-1.14] | 1.10 [1.06-1.13] | 1.02 [0.71-1.46] | 1.11 [1.08-1.15] | 13,152 | 13,206 | 11,717 | 125 | 12,002 |
| G57 | 1.25 [1.15-1.37] | 1.26 [1.15-1.37] | 1.25 [1.14-1.37] | | 1.16 [1.06-1.27] | 1.18 [1.08-1.29] | 1.15 [1.05-1.26] | 1.15 [1.04-1.26] | | 1.11 [1.02-1.22] | 1,441 | 1,459 | 1,257 | NA | 1,371 |
| G58 | 1.23 [1.09-1.39] | 1.26 [1.12-1.43] | 1.31 [1.15-1.48] | | 1.19 [1.06-1.34] | 1.14 [1.01-1.29] | 1.13 [1.00-1.28] | 1.17 [1.03-1.34] | | 1.14 [1.01-1.29] | 802 | 802 | 700 | NA | 782 |
| G59 | 1.24 [1.10-1.40] | 1.21 [1.07-1.36] | 1.22 [1.08-1.37] | | 1.17 [1.04-1.31] | 1.14 [1.01-1.29] | 1.04 [0.92-1.18] | 1.05 [0.93-1.19] | | 1.12 [1.00-1.27] | 787 | 788 | 762 | NA | 767 |
| G60 | 1.21 [1.01-1.44] | 1.12 [0.94-1.34] | 1.20 [1.00-1.45] | | 1.18 [0.99-1.41] | 1.11 [0.92-1.33] | 1.02 [0.85-1.23] | 1.10 [0.91-1.33] | | 1.09 [0.91-1.32] | 367 | 355 | 325 | NA | 345 |
| G61 | 1.22 [1.03-1.44] | 1.23 [1.05-1.46] | 1.25 [1.05-1.48] | | 1.13 [0.95-1.35] | 1.20 [1.01-1.41] | 1.19 [1.01-1.41] | 1.19 [1.00-1.42] | | 1.10 [0.92-1.31] | 409 | 406 | 372 | NA | 363 |
| G62 | 1.19 [1.13-1.24] | 1.20 [1.14-1.25] | 1.19 [1.14-1.25] | | 1.14 [1.09-1.19] | 1.10 [1.05-1.16] | 1.07 [1.02-1.12] | 1.06 [1.01-1.11] | | 1.09 [1.04-1.14] | 5,343 | 5,356 | 5,129 | NA | 5,257 |
| G63 | 1.22 [1.15-1.30] | 1.20 [1.13-1.27] | 1.23 [1.16-1.31] | | 1.18 [1.11-1.25] | 1.14 [1.07-1.21] | 1.03 [0.97-1.10] | 1.06 [1.00-1.13] | | 1.13 [1.06-1.19] | 3,330 | 3,329 | 3,271 | NA | 3,363 |
| G70 | 1.06 [0.90-1.25] | 1.07 [0.90-1.26] | 1.14 [0.96-1.36] | | 1.15 [0.97-1.36] | 1.03 [0.87-1.21] | 1.02 [0.86-1.21] | 1.09 [0.91-1.29] | | 1.11 [0.94-1.32] | 424 | 395 | 376 | NA | 382 |
| G71 | 1.03 [0.84-1.26] | 1.15 [0.93-1.43] | 1.08 [0.85-1.37] | | 1.02 [0.83-1.26] | 1.01 [0.82-1.24] | 1.10 [0.88-1.38] | 1.04 [0.81-1.33] | | 1.02 [0.82-1.26] | 275 | 253 | 188 | NA | 252 |
| G72 | 1.36 [1.18-1.57] | 1.33 [1.15-1.53] | 1.35 [1.17-1.57] | | 1.29 [1.12-1.48] | 1.29 [1.11-1.49] | 1.22 [1.06-1.42] | 1.27 [1.09-1.48] | | 1.25 [1.08-1.44] | 594 | 575 | 536 | NA | 566 |
| G80 | 1.07 [0.94-1.22] | 1.30 [1.10-1.52] | 1.44 [1.19-1.73] | 0.78 [0.63-0.96] | 1.02 [0.89-1.18] | 1.05 [0.92-1.20] | 1.31 [1.11-1.54] | 1.46 [1.21-1.77] | 0.77 [0.62-0.95] | 1.02 [0.88-1.17] | 676 | 458 | 356 | 253 | 591 |
| G81 | 1.10 [1.06-1.14] | 1.11 [1.07-1.15] | 1.11 [1.07-1.15] | 0.98 [0.72-1.33] | 1.08 [1.04-1.12] | 1.09 [1.05-1.13] | 1.07 [1.03-1.11] | 1.06 [1.03-1.10] | 0.99 [0.72-1.36] | 1.08 [1.04-1.12] | 9,002 | 8,870 | 8,701 | 138 | 8,612 |
| G82 | 1.16 [1.03-1.31] | 1.13 [1.00-1.28] | 1.10 [0.97-1.25] | | 1.11 [0.98-1.25] | 1.13 [1.00-1.27] | 1.10 [0.97-1.24] | 1.06 [0.93-1.20] | | 1.10 [0.97-1.25] | 793 | 752 | 683 | NA | 737 |
| G83 | 1.09 [0.99-1.20] | 1.07 [0.97-1.18] | 1.11 [1.00-1.23] | | 1.05 [0.95-1.15] | 1.05 [0.95-1.16] | 1.00 [0.91-1.11] | 1.05 [0.94-1.16] | | 1.03 [0.93-1.13] | 1,227 | 1,220 | 1,083 | NA | 1,205 |
| G90 | 1.14 [1.00-1.29] | 1.09 [0.96-1.24] | 1.11 [0.97-1.28] | | 1.01 [0.89-1.15] | 1.07 [0.94-1.22] | 0.99 [0.87-1.12] | 1.01 [0.88-1.17] | | 0.96 [0.84-1.09] | 706 | 711 | 566 | NA | 663 |
| G91 | 1.09 [1.00-1.19] | 1.11 [1.02-1.22] | 1.10 [1.01-1.21] | | 1.11 [1.01-1.21] | 1.08 [0.99-1.18] | 1.09 [0.99-1.19] | 1.07 [0.98-1.18] | | 1.10 [1.00-1.20] | 1,510 | 1,436 | 1,307 | NA | 1,376 |
| G93 | 1.15 [1.10-1.20] | 1.15 [1.11-1.20] | 1.15 [1.10-1.20] | 1.02 [0.88-1.19] | 1.12 [1.07-1.17] | 1.09 [1.04-1.14] | 1.06 [1.01-1.11] | 1.06 [1.01-1.12] | 0.99 [0.85-1.16] | 1.08 [1.03-1.13] | 6,539 | 6,403 | 5,103 | 553 | 6,271 |
| G94 | 1.09 [0.86-1.38] | 1.16 [0.90-1.50] | 1.17 [0.91-1.52] | | 1.06 [0.83-1.36] | 1.08 [0.85-1.37] | 1.14 [0.88-1.48] | 1.17 [0.90-1.52] | | 1.08 [0.84-1.38] | 195 | 179 | 166 | NA | 181 |
| G95 | 1.09 [1.01-1.19] | 1.12 [1.03-1.21] | 1.14 [1.05-1.24] | | 1.07 [0.99-1.16] | 1.06 [0.97-1.15] | 1.07 [0.98-1.16] | 1.08 [1.00-1.18] | | 1.05 [0.97-1.14] | 1,733 | 1,708 | 1,606 | NA | 1,636 |
| G96 | 1.22 [1.05-1.41] | 1.23 [1.06-1.43] | 1.31 [1.11-1.53] | | 1.22 [1.06-1.41] | 1.13 [0.97-1.31] | 1.11 [0.95-1.29] | 1.19 [1.01-1.41] | | 1.18 [1.02-1.37] | 566 | 548 | 461 | NA | 540 |
| G97 | 1.10 [0.94-1.28] | 1.14 [0.97-1.33] | 1.21 [0.99-1.48] | | 1.10 [0.94-1.28] | 1.01 [0.86-1.18] | 1.02 [0.86-1.20] | 1.11 [0.90-1.36] | | 1.05 [0.90-1.23] | 497 | 481 | 279 | NA | 474 |
| G98 | 1.23 [0.97-1.55] | 1.18 [0.93-1.49] | 1.28 [1.00-1.65] | | 1.23 [0.98-1.54] | 1.13 [0.89-1.43] | 1.03 [0.80-1.31] | 1.10 [0.84-1.43] | | 1.12 [0.89-1.42] | 222 | 216 | 194 | NA | 221 |
| G99 | 1.17 [1.10-1.24] | 1.16 [1.09-1.24] | 1.15 [1.08-1.22] | | 1.13 [1.07-1.20] | 1.11 [1.04-1.19] | 1.07 [1.00-1.14] | 1.06 [0.99-1.13] | | 1.09 [1.03-1.17] | 2,947 | 2,941 | 2,737 | NA | 2,861 |
| Diseases of the eye and adnexa (H00-H59) | | | | | | | | | | | | | | | |
| H00 | 1.18 [1.08-1.28] | 1.21 [1.11-1.33] | 1.18 [1.06-1.31] | 1.06 [0.87-1.28] | 1.19 [1.09-1.30] | 1.14 [1.04-1.24] | 1.18 [1.08-1.30] | 1.16 [1.04-1.29] | 1.05 [0.86-1.28] | 1.17 [1.06-1.28] | 1,657 | 1,403 | 1,018 | 359 | 1,410 |
| H01 | 1.30 [1.21-1.41] | 1.29 [1.19-1.40] | 1.31 [1.21-1.42] | | 1.30 [1.20-1.40] | 1.24 [1.14-1.34] | 1.22 [1.13-1.32] | 1.24 [1.14-1.35] | | 1.24 [1.14-1.34] | 1,952 | 1,889 | 1,824 | NA | 1,855 |
| H02 | 1.21 [1.17-1.25] | 1.21 [1.17-1.26] | 1.22 [1.17-1.26] | 1.14 [0.89-1.47] | 1.18 [1.13-1.22] | 1.16 [1.12-1.20] | 1.16 [1.11-1.20] | 1.16 [1.12-1.20] | 1.10 [0.85-1.43] | 1.14 [1.09-1.18] | 9,523 | 9,381 | 9,001 | 209 | 8,646 |
| H03 | 1.30 [0.94-1.79] | 1.23 [0.88-1.73] | | | 1.29 [0.92-1.80] | 1.24 [0.89-1.72] | 1.14 [0.81-1.62] | | | 1.19 [0.84-1.69] | 118 | 101 | NA | NA | 107 |
| H04 | 1.29 [1.23-1.35] | 1.31 [1.25-1.38] | 1.31 [1.25-1.38] | 1.13 [0.93-1.39] | 1.26 [1.20-1.32] | 1.21 [1.15-1.27] | 1.23 [1.17-1.29] | 1.23 [1.16-1.29] | 1.09 [0.89-1.33] | 1.19 [1.13-1.25] | 5,073 | 4,820 | 4,603 | 318 | 4,659 |
| H05 | 1.31 [1.18-1.46] | 1.32 [1.17-1.50] | 1.33 [1.17-1.52] | 1.23 [1.02-1.48] | 1.31 [1.17-1.46] | 1.27 [1.14-1.41] | 1.25 [1.10-1.42] | 1.25 [1.10-1.43] | 1.18 [0.97-1.42] | 1.27 [1.14-1.42] | 1,164 | 799 | 693 | 403 | 1,031 |
| H06 | 1.51 [1.19-1.92] | 1.55 [1.23-1.96] | 1.42 [1.10-1.83] | | 1.42 [1.12-1.81] | 1.45 [1.14-1.85] | 1.41 [1.11-1.79] | 1.34 [1.03-1.74] | | 1.38 [1.08-1.76] | 219 | 229 | 187 | NA | 195 |
| H10 | 1.25 [1.17-1.34] | 1.32 [1.22-1.43] | 1.23 [1.14-1.34] | 1.26 [1.06-1.48] | 1.27 [1.18-1.36] | 1.19 [1.11-1.28] | 1.24 [1.14-1.34] | 1.16 [1.07-1.26] | 1.15 [0.97-1.36] | 1.22 [1.13-1.31] | 2,558 | 2,121 | 1,957 | 490 | 2,465 |
| H11 | 1.14 [1.06-1.24] | 1.15 [1.06-1.24] | 1.16 [1.07-1.26] | | 1.13 [1.05-1.23] | 1.11 [1.03-1.21] | 1.11 [1.02-1.20] | 1.12 [1.03-1.22] | | 1.11 [1.02-1.20] | 1,836 | 1,797 | 1,678 | NA | 1,698 |
| H13 | 1.38 [1.02-1.85] | 1.67 [1.21-2.29] | 1.65 [1.18-2.29] | | 1.31 [0.97-1.77] | 1.35 [1.00-1.84] | 1.62 [1.17-2.24] | 1.56 [1.12-2.19] | | 1.28 [0.94-1.73] | 144 | 126 | 113 | NA | 129 |
| H15 | 1.44 [1.11-1.87] | 1.39 [1.07-1.79] | 1.38 [1.05-1.81] | | 1.36 [1.05-1.77] | 1.37 [1.05-1.79] | 1.31 [1.01-1.69] | 1.29 [0.97-1.70] | | 1.28 [0.98-1.68] | 191 | 189 | 163 | NA | 169 |
| H16 | 1.43 [1.27-1.60] | 1.34 [1.19-1.51] | 1.40 [1.23-1.58] | | 1.41 [1.25-1.59] | 1.36 [1.21-1.54] | 1.29 [1.14-1.45] | 1.34 [1.18-1.52] | | 1.37 [1.21-1.54] | 915 | 881 | 800 | NA | 875 |
| H17 | 1.26 [1.08-1.46] | 1.20 [1.03-1.40] | 1.27 [1.08-1.48] | | 1.27 [1.09-1.48] | 1.21 [1.03-1.41] | 1.16 [0.99-1.35] | 1.22 [1.05-1.43] | | 1.22 [1.04-1.42] | 537 | 520 | 495 | NA | 506 |
| H18 | 1.17 [1.11-1.25] | 1.17 [1.10-1.24] | 1.13 [1.07-1.21] | 1.76 [1.24-2.50] | 1.18 [1.11-1.25] | 1.14 [1.07-1.21] | 1.13 [1.06-1.20] | 1.10 [1.03-1.17] | 1.53 [1.04-2.25] | 1.14 [1.08-1.22] | 3,409 | 3,382 | 3,032 | 140 | 3,216 |
| H19 | 1.40 [1.11-1.76] | 1.44 [1.15-1.82] | 1.49 [1.17-1.90] | | 1.48 [1.17-1.86] | 1.27 [1.00-1.61] | 1.33 [1.05-1.68] | 1.32 [1.03-1.70] | | 1.33 [1.05-1.69] | 240 | 239 | 213 | NA | 230 |
| H20 | 1.30 [1.16-1.47] | 1.19 [1.06-1.34] | 1.19 [1.06-1.35] | | 1.25 [1.12-1.41] | 1.28 [1.14-1.44] | 1.14 [1.01-1.29] | 1.16 [1.02-1.31] | | 1.23 [1.10-1.39] | 900 | 855 | 764 | NA | 847 |
| H21 | 1.13 [1.04-1.24] | 1.12 [1.03-1.23] | 1.08 [0.99-1.18] | | 1.09 [1.00-1.19] | 1.11 [1.01-1.21] | 1.09 [1.00-1.19] | 1.05 [0.96-1.15] | | 1.07 [0.98-1.17] | 1,471 | 1,462 | 1,428 | NA | 1,401 |
| H25 | 1.10 [1.08-1.12] | 1.10 [1.08-1.12] | 1.11 [1.09-1.13] | | 1.09 [1.07-1.11] | 1.07 [1.05-1.09] | 1.06 [1.04-1.08] | 1.08 [1.06-1.10] | | 1.06 [1.04-1.08] | 35,929 | 36,002 | 35,913 | NA | 32,562 |
| H26 | 1.12 [1.11-1.14] | 1.12 [1.10-1.14] | 1.12 [1.10-1.14] | 1.67 [1.14-2.46] | 1.11 [1.09-1.13] | 1.09 [1.08-1.11] | 1.08 [1.06-1.09] | 1.08 [1.06-1.10] | 1.66 [1.11-2.48] | 1.08 [1.07-1.10] | 52,608 | 52,595 | 52,245 | 113 | 47,168 |
| H27 | 1.10 [0.97-1.26] | 1.16 [1.02-1.33] | 1.11 [0.97-1.27] | | 1.17 [1.03-1.34] | 1.09 [0.95-1.25] | 1.13 [0.99-1.29] | 1.08 [0.94-1.24] | | 1.15 [1.01-1.32] | 663 | 652 | 622 | NA | 628 |
| H28 | 1.17 [0.97-1.41] | 1.23 [1.03-1.49] | 1.17 [0.97-1.41] | | 1.21 [1.00-1.46] | 1.12 [0.93-1.36] | 1.15 [0.95-1.39] | 1.09 [0.90-1.32] | | 1.18 [0.97-1.43] | 321 | 323 | 308 | NA | 302 |
| H31 | 1.06 [0.94-1.20] | 1.05 [0.93-1.18] | 1.02 [0.91-1.16] | | 1.05 [0.93-1.18] | 1.05 [0.94-1.19] | 1.03 [0.92-1.17] | 1.00 [0.88-1.13] | | 1.04 [0.93-1.18] | 819 | 817 | 796 | NA | 785 |
| H33 | 0.98 [0.92-1.05] | 1.02 [0.96-1.09] | 0.99 [0.92-1.06] | | 1.01 [0.94-1.08] | 0.97 [0.91-1.04] | 1.01 [0.94-1.08] | 0.97 [0.91-1.04] | | 1.00 [0.93-1.07] | 2,569 | 2,550 | 2,359 | NA | 2,221 |
| H34 | 1.09 [1.01-1.17] | 1.09 [1.02-1.18] | 1.09 [1.01-1.17] | | 1.06 [0.98-1.14] | 1.07 [1.00-1.16] | 1.06 [0.98-1.14] | 1.04 [0.97-1.13] | | 1.05 [0.97-1.13] | 2,072 | 2,081 | 2,052 | NA | 1,955 |
| H35 | 1.10 [1.07-1.13] | 1.11 [1.09-1.14] | 1.10 [1.07-1.13] | | 1.09 [1.06-1.11] | 1.08 [1.05-1.11] | 1.08 [1.05-1.11] | 1.07 [1.04-1.10] | | 1.07 [1.04-1.10] | 19,118 | 19,098 | 18,853 | NA | 17,748 |
| H36 | 1.05 [1.02-1.09] | 1.06 [1.02-1.09] | 1.06 [1.02-1.09] | | 1.05 [1.01-1.09] | 1.03 [0.99-1.06] | 0.98 [0.94-1.01] | 0.97 [0.94-1.01] | | 1.04 [1.00-1.08] | 8,639 | 8,638 | 8,365 | NA | 8,222 |
| H40 | 1.11 [1.08-1.13] | 1.11 [1.08-1.14] | 1.11 [1.08-1.14] | | 1.10 [1.07-1.13] | 1.08 [1.06-1.11] | 1.08 [1.06-1.11] | 1.09 [1.06-1.12] | | 1.08 [1.05-1.11] | 17,114 | 17,086 | 16,991 | NA | 15,494 |
| H43 | 1.06 [1.00-1.12] | 1.07 [1.01-1.13] | 1.08 [1.02-1.15] | | 1.04 [0.98-1.10] | 1.03 [0.97-1.10] | 1.03 [0.97-1.09] | 1.04 [0.98-1.11] | | 1.02 [0.96-1.08] | 3,277 | 3,291 | 3,156 | NA | 3,071 |
| H44 | 1.23 [1.11-1.37] | 1.21 [1.09-1.34] | 1.20 [1.08-1.34] | | 1.18 [1.06-1.31] | 1.19 [1.07-1.32] | 1.16 [1.05-1.29] | 1.15 [1.03-1.28] | | 1.14 [1.02-1.26] | 1,097 | 1,065 | 1,005 | NA | 1,020 |
| H46 | 1.41 [1.16-1.70] | 1.40 [1.15-1.70] | 1.38 [1.11-1.71] | | 1.38 [1.14-1.68] | 1.37 [1.13-1.67] | 1.30 [1.06-1.58] | 1.28 [1.02-1.60] | | 1.34 [1.10-1.64] | 348 | 345 | 257 | NA | 323 |
| H47 | 1.12 [1.01-1.24] | 1.15 [1.04-1.27] | 1.16 [1.03-1.30] | 1.21 [0.93-1.57] | 1.10 [0.99-1.21] | 1.10 [0.99-1.22] | 1.10 [0.99-1.22] | 1.14 [1.01-1.28] | 1.20 [0.91-1.59] | 1.10 [0.99-1.21] | 1,212 | 1,132 | 842 | 204 | 1,123 |
| H49 | 0.96 [0.85-1.09] | 1.01 [0.89-1.15] | 1.02 [0.89-1.16] | | 1.01 [0.89-1.14] | 0.94 [0.83-1.06] | 0.97 [0.85-1.10] | 0.97 [0.85-1.11] | | 1.00 [0.88-1.13] | 744 | 706 | 639 | NA | 681 |
| H50 | 1.11 [1.03-1.19] | 1.22 [1.11-1.35] | 1.21 [1.08-1.35] | 0.97 [0.86-1.08] | 1.08 [1.00-1.17] | 1.07 [0.99-1.16] | 1.16 [1.05-1.28] | 1.15 [1.02-1.28] | 0.96 [0.85-1.08] | 1.06 [0.98-1.15] | 2,170 | 1,246 | 914 | 989 | 1,877 |
| H51 | 1.22 [0.90-1.65] | 1.14 [0.83-1.56] | | | 1.13 [0.84-1.51] | 1.15 [0.84-1.56] | 1.06 [0.77-1.47] | | | 1.09 [0.81-1.48] | 134 | 115 | NA | NA | 131 |
| H52 | 1.06 [1.01-1.11] | 1.06 [1.01-1.11] | 1.06 [1.01-1.12] | 1.02 [0.80-1.29] | 1.05 [1.00-1.10] | 1.04 [0.99-1.09] | 1.04 [0.99-1.09] | 1.04 [0.99-1.09] | 0.98 [0.77-1.25] | 1.03 [0.98-1.08] | 5,215 | 5,023 | 4,887 | 251 | 4,884 |
| H53 | 1.14 [1.10-1.18] | 1.14 [1.11-1.18] | 1.15 [1.11-1.19] | 1.02 [0.91-1.16] | 1.10 [1.07-1.14] | 1.07 [1.04-1.11] | 1.05 [1.01-1.08] | 1.07 [1.03-1.11] | 1.01 [0.89-1.14] | 1.06 [1.03-1.10] | 10,579 | 10,452 | 7,983 | 962 | 10,026 |
| H54 | 1.14 [1.10-1.19] | 1.14 [1.09-1.18] | 1.14 [1.09-1.18] | 0.97 [0.75-1.25] | 1.13 [1.09-1.17] | 1.12 [1.08-1.16] | 1.10 [1.05-1.14] | 1.09 [1.05-1.13] | 0.93 [0.72-1.21] | 1.12 [1.07-1.16] | 8,810 | 8,649 | 8,343 | 204 | 8,476 |
| H55 | 1.08 [0.95-1.22] | 1.11 [0.98-1.27] | 1.05 [0.92-1.21] | 0.88 [0.62-1.25] | 1.09 [0.96-1.23] | 1.04 [0.91-1.18] | 1.04 [0.91-1.20] | 0.98 [0.85-1.14] | 0.87 [0.61-1.24] | 1.06 [0.93-1.20] | 741 | 667 | 580 | 108 | 716 |
| H57 | 1.19 [1.10-1.29] | 1.17 [1.08-1.28] | 1.19 [1.10-1.30] | 1.28 [0.96-1.71] | 1.13 [1.05-1.23] | 1.13 [1.04-1.23] | 1.09 [1.00-1.19] | 1.12 [1.03-1.23] | 1.24 [0.92-1.67] | 1.09 [1.00-1.18] | 1,849 | 1,714 | 1,554 | 171 | 1,753 |
| H58 | 1.05 [0.89-1.25] | 1.09 [0.92-1.30] | 1.06 [0.89-1.26] | | 1.08 [0.91-1.28] | 1.03 [0.87-1.22] | 1.05 [0.88-1.25] | 0.99 [0.83-1.18] | | 1.07 [0.90-1.27] | 395 | 389 | 374 | NA | 367 |
| H59 | 1.07 [0.94-1.23] | 1.08 [0.95-1.24] | 1.03 [0.90-1.18] | | 1.10 [0.96-1.26] | 1.07 [0.94-1.23] | 1.08 [0.94-1.24] | 1.02 [0.89-1.17] | | 1.10 [0.96-1.26] | 617 | 610 | 597 | NA | 582 |
| Diseases of the ear and mastoid process (H60-H95) | | | | | | | | | | | | | | | |
| H60 | 1.53 [1.41-1.67] | 1.55 [1.42-1.70] | 1.69 [1.52-1.87] | 1.35 [1.09-1.69] | 1.48 [1.36-1.61] | 1.44 [1.32-1.57] | 1.42 [1.30-1.56] | 1.58 [1.42-1.75] | 1.30 [1.03-1.64] | 1.43 [1.31-1.56] | 1,834 | 1,648 | 1,264 | 312 | 1,706 |
| H61 | 1.32 [1.23-1.41] | 1.30 [1.20-1.40] | 1.33 [1.23-1.44] | 1.26 [1.10-1.44] | 1.30 [1.22-1.39] | 1.28 [1.19-1.37] | 1.24 [1.14-1.34] | 1.26 [1.16-1.37] | 1.22 [1.06-1.40] | 1.26 [1.18-1.35] | 2,833 | 2,130 | 1,902 | 764 | 2,654 |
| H65 | 1.11 [1.05-1.17] | 1.21 [1.09-1.34] | 1.19 [1.06-1.34] | 1.06 [1.00-1.13] | 1.08 [1.02-1.14] | 1.07 [1.01-1.13] | 1.12 [1.01-1.25] | 1.12 [1.00-1.26] | 1.03 [0.97-1.10] | 1.05 [0.99-1.11] | 4,278 | 1,007 | 810 | 3,320 | 3,717 |
| H66 | 1.21 [1.13-1.29] | 1.38 [1.26-1.52] | 1.38 [1.24-1.54] | 1.12 [1.03-1.22] | 1.20 [1.13-1.29] | 1.16 [1.09-1.24] | 1.29 [1.17-1.42] | 1.29 [1.15-1.44] | 1.10 [1.01-1.20] | 1.17 [1.09-1.25] | 2,960 | 1,419 | 1,051 | 1,672 | 2,718 |
| H69 | 1.34 [1.08-1.66] | 1.24 [0.99-1.55] | 1.29 [0.99-1.69] | | 1.21 [0.98-1.49] | 1.25 [1.00-1.56] | 1.10 [0.87-1.39] | 1.18 [0.89-1.55] | | 1.13 [0.91-1.41] | 283 | 250 | 158 | NA | 261 |
| H70 | 1.15 [0.95-1.40] | 1.27 [1.03-1.58] | 1.29 [1.02-1.63] | | 1.09 [0.90-1.32] | 1.11 [0.91-1.35] | 1.17 [0.94-1.46] | 1.22 [0.96-1.55] | | 1.07 [0.88-1.30] | 325 | 260 | 221 | NA | 308 |
| H71 | 0.97 [0.84-1.11] | 1.03 [0.89-1.20] | 1.08 [0.90-1.29] | 0.95 [0.71-1.29] | 1.00 [0.86-1.15] | 0.96 [0.83-1.10] | 1.00 [0.86-1.16] | 1.04 [0.87-1.25] | 1.00 [0.73-1.36] | 0.99 [0.86-1.15] | 590 | 498 | 334 | 151 | 518 |
| H72 | 1.16 [1.06-1.28] | 1.20 [1.08-1.33] | 1.17 [1.03-1.32] | 1.16 [0.96-1.40] | 1.09 [0.99-1.21] | 1.11 [1.01-1.22] | 1.13 [1.02-1.26] | 1.12 [0.99-1.27] | 1.13 [0.93-1.38] | 1.05 [0.95-1.16] | 1,371 | 1,072 | 742 | 393 | 1,206 |
| H73 | 1.12 [0.99-1.28] | 1.25 [1.06-1.47] | 1.29 [1.06-1.57] | 1.10 [0.90-1.34] | 1.08 [0.94-1.23] | 1.09 [0.95-1.25] | 1.20 [1.01-1.42] | 1.23 [1.01-1.50] | 1.08 [0.88-1.32] | 1.05 [0.92-1.21] | 737 | 450 | 304 | 363 | 678 |
| H74 | 1.10 [0.96-1.25] | 1.10 [0.95-1.27] | 1.22 [1.04-1.44] | 0.95 [0.72-1.27] | 1.08 [0.94-1.23] | 1.08 [0.95-1.24] | 1.05 [0.91-1.22] | 1.17 [0.99-1.39] | 0.96 [0.72-1.29] | 1.06 [0.92-1.21] | 705 | 566 | 432 | 177 | 626 |
| H80 | 0.98 [0.76-1.26] | 0.98 [0.76-1.27] | 0.93 [0.70-1.24] | | 1.09 [0.84-1.41] | 0.98 [0.76-1.27] | 0.98 [0.75-1.27] | 0.91 [0.68-1.23] | | 1.09 [0.84-1.42] | 178 | 176 | 129 | NA | 160 |
| H81 | 1.17 [1.10-1.23] | 1.17 [1.11-1.24] | 1.17 [1.11-1.24] | | 1.12 [1.06-1.18] | 1.11 [1.04-1.17] | 1.09 [1.03-1.16] | 1.10 [1.04-1.16] | | 1.07 [1.01-1.13] | 3,769 | 3,771 | 3,621 | NA | 3,536 |
| H83 | 1.16 [1.06-1.27] | 1.20 [1.09-1.31] | 1.16 [1.05-1.27] | | 1.14 [1.04-1.25] | 1.09 [0.99-1.19] | 1.12 [1.02-1.23] | 1.07 [0.97-1.18] | | 1.09 [0.99-1.19] | 1,414 | 1,390 | 1,253 | NA | 1,326 |
| H90 | 1.19 [1.12-1.26] | 1.23 [1.16-1.31] | 1.24 [1.17-1.33] | 1.00 [0.86-1.17] | 1.16 [1.10-1.23] | 1.13 [1.07-1.20] | 1.15 [1.08-1.22] | 1.17 [1.09-1.25] | 0.98 [0.84-1.15] | 1.12 [1.05-1.18] | 3,866 | 3,387 | 3,030 | 592 | 3,712 |
| H91 | 1.16 [1.13-1.19] | 1.16 [1.13-1.19] | 1.16 [1.13-1.19] | 0.99 [0.87-1.13] | 1.15 [1.12-1.18] | 1.12 [1.10-1.15] | 1.11 [1.08-1.14] | 1.11 [1.09-1.14] | 0.96 [0.84-1.11] | 1.12 [1.10-1.15] | 22,799 | 22,237 | 21,579 | 739 | 21,877 |
| H92 | 1.29 [1.17-1.42] | 1.40 [1.26-1.56] | 1.37 [1.22-1.55] | 1.00 [0.81-1.24] | 1.28 [1.16-1.41] | 1.21 [1.10-1.34] | 1.28 [1.15-1.43] | 1.25 [1.11-1.42] | 1.00 [0.81-1.25] | 1.22 [1.11-1.35] | 1,323 | 1,107 | 823 | 306 | 1,266 |
| H93 | 1.22 [1.13-1.32] | 1.21 [1.12-1.30] | 1.22 [1.13-1.33] | 1.35 [0.92-1.98] | 1.23 [1.14-1.32] | 1.16 [1.08-1.26] | 1.13 [1.04-1.22] | 1.15 [1.06-1.24] | 1.41 [0.94-2.10] | 1.18 [1.09-1.27] | 2,067 | 2,043 | 1,819 | 109 | 1,971 |
| H95 | 1.14 [0.84-1.55] | 1.23 [0.90-1.70] | | | 1.06 [0.78-1.43] | 1.14 [0.84-1.56] | 1.19 [0.86-1.65] | | | 1.04 [0.76-1.42] | 128 | 117 | NA | NA | 125 |
| Diseases of the circulatory system (I00-I99) | | | | | | | | | | | | | | | |
| I05 | 1.19 [1.08-1.31] | 1.20 [1.09-1.32] | 1.23 [1.12-1.36] | | 1.17 [1.07-1.29] | 1.14 [1.04-1.25] | 1.14 [1.04-1.26] | 1.17 [1.06-1.29] | | 1.15 [1.04-1.26] | 1,292 | 1,296 | 1,262 | NA | 1,208 |
| I06 | 1.22 [0.90-1.64] | 1.24 [0.93-1.66] | 1.36 [1.01-1.82] | | 1.11 [0.82-1.49] | 1.15 [0.85-1.56] | 1.17 [0.87-1.58] | 1.28 [0.95-1.72] | | 1.05 [0.78-1.42] | 126 | 127 | 126 | NA | 119 |
| I07 | 1.21 [1.14-1.29] | 1.21 [1.14-1.28] | 1.21 [1.14-1.28] | | 1.18 [1.11-1.25] | 1.17 [1.10-1.25] | 1.14 [1.07-1.21] | 1.14 [1.07-1.21] | | 1.15 [1.08-1.22] | 3,304 | 3,293 | 3,227 | NA | 3,229 |
| I08 | 1.14 [1.11-1.17] | 1.14 [1.11-1.17] | 1.14 [1.11-1.17] | | 1.10 [1.07-1.13] | 1.11 [1.08-1.14] | 1.08 [1.05-1.11] | 1.08 [1.05-1.11] | | 1.08 [1.05-1.11] | 16,519 | 16,490 | 16,300 | NA | 16,138 |
| I09 | 1.21 [0.95-1.54] | 1.25 [0.99-1.58] | 1.36 [1.07-1.72] | | 1.26 [0.99-1.60] | 1.18 [0.93-1.51] | 1.21 [0.95-1.53] | 1.30 [1.02-1.66] | | 1.23 [0.96-1.57] | 199 | 200 | 197 | NA | 190 |
| I10 | 1.08 [1.07-1.09] | 1.08 [1.07-1.09] | 1.09 [1.08-1.10] | 0.92 [0.78-1.09] | 1.08 [1.07-1.09] | 1.06 [1.05-1.07] | 1.02 [1.01-1.03] | 1.02 [1.01-1.03] | 0.93 [0.78-1.10] | 1.07 [1.05-1.08] | 157,034 | 157,068 | 153,830 | 506 | 119,978 |
| I11 | 1.07 [0.96-1.20] | 1.07 [0.96-1.20] | 1.09 [0.98-1.22] | | 1.00 [0.89-1.12] | 1.03 [0.92-1.15] | 1.00 [0.90-1.12] | 1.01 [0.91-1.13] | | 0.98 [0.88-1.10] | 921 | 928 | 919 | NA | 850 |
| I12 | 1.16 [1.12-1.20] | 1.16 [1.12-1.20] | 1.17 [1.13-1.21] | | 1.12 [1.09-1.16] | 1.13 [1.10-1.17] | 1.09 [1.06-1.13] | 1.10 [1.06-1.14] | | 1.12 [1.08-1.16] | 9,880 | 9,878 | 9,802 | NA | 9,300 |
| I13 | 1.23 [1.06-1.43] | 1.19 [1.02-1.38] | 1.17 [1.00-1.36] | | 1.13 [0.98-1.32] | 1.17 [1.00-1.37] | 1.09 [0.93-1.28] | 1.07 [0.92-1.26] | | 1.10 [0.95-1.28] | 518 | 518 | 518 | NA | 518 |
| I15 | 1.17 [1.03-1.34] | 1.19 [1.04-1.36] | 1.15 [1.00-1.32] | | 1.06 [0.93-1.21] | 1.14 [1.00-1.31] | 1.11 [0.97-1.27] | 1.07 [0.93-1.23] | | 1.06 [0.93-1.21] | 663 | 629 | 589 | NA | 635 |
| I20 | 1.13 [1.11-1.15] | 1.12 [1.10-1.15] | 1.13 [1.11-1.15] | | 1.10 [1.08-1.12] | 1.08 [1.06-1.10] | 1.04 [1.02-1.06] | 1.04 [1.03-1.06] | | 1.06 [1.04-1.08] | 35,262 | 35,320 | 35,085 | NA | 31,682 |
| I21 | 1.10 [1.07-1.12] | 1.08 [1.06-1.11] | 1.09 [1.07-1.12] | | 1.08 [1.05-1.11] | 1.07 [1.05-1.10] | 1.04 [1.01-1.06] | 1.04 [1.02-1.07] | | 1.07 [1.04-1.09] | 21,458 | 21,437 | 21,330 | NA | 19,587 |
| I22 | 1.15 [1.07-1.24] | 1.16 [1.09-1.25] | 1.19 [1.11-1.27] | | 1.15 [1.08-1.24] | 1.12 [1.04-1.20] | 1.09 [1.01-1.17] | 1.11 [1.03-1.19] | | 1.14 [1.06-1.22] | 2,257 | 2,265 | 2,268 | NA | 2,176 |
| I23 | 1.25 [0.92-1.69] | 1.22 [0.91-1.65] | 1.35 [0.99-1.83] | | 1.36 [1.00-1.85] | 1.23 [0.91-1.67] | 1.14 [0.84-1.55] | 1.32 [0.96-1.80] | | 1.33 [0.98-1.82] | 123 | 124 | 123 | NA | 115 |
| I24 | 1.13 [1.09-1.18] | 1.13 [1.08-1.18] | 1.15 [1.11-1.20] | | 1.12 [1.08-1.17] | 1.09 [1.05-1.14] | 1.06 [1.01-1.10] | 1.07 [1.02-1.11] | | 1.10 [1.05-1.14] | 6,864 | 6,833 | 6,812 | NA | 6,691 |
| I25 | 1.10 [1.09-1.12] | 1.10 [1.09-1.12] | 1.10 [1.09-1.12] | | 1.08 [1.07-1.10] | 1.06 [1.05-1.08] | 1.03 [1.02-1.05] | 1.03 [1.02-1.05] | | 1.06 [1.04-1.07] | 59,154 | 59,160 | 58,817 | NA | 53,341 |
| I26 | 1.09 [1.05-1.12] | 1.07 [1.04-1.11] | 1.07 [1.04-1.11] | | 1.06 [1.02-1.09] | 1.05 [1.02-1.09] | 1.02 [0.98-1.05] | 1.02 [0.99-1.06] | | 1.03 [0.99-1.06] | 10,547 | 10,591 | 10,054 | NA | 9,785 |
| I27 | 1.21 [1.16-1.26] | 1.20 [1.16-1.25] | 1.20 [1.15-1.25] | | 1.19 [1.14-1.24] | 1.15 [1.11-1.20] | 1.12 [1.08-1.17] | 1.11 [1.07-1.15] | | 1.15 [1.11-1.20] | 7,864 | 7,862 | 7,772 | NA | 7,790 |
| I28 | 1.07 [0.83-1.39] | 1.01 [0.78-1.30] | 1.03 [0.78-1.34] | | 0.98 [0.76-1.25] | 1.01 [0.77-1.31] | 0.91 [0.70-1.19] | 0.93 [0.71-1.23] | | 0.90 [0.69-1.16] | 174 | 165 | 158 | NA | 176 |
| I30 | 1.05 [0.85-1.29] | 1.07 [0.87-1.30] | 1.06 [0.85-1.32] | | 1.09 [0.88-1.34] | 1.03 [0.84-1.27] | 1.02 [0.83-1.25] | 1.02 [0.81-1.28] | | 1.07 [0.86-1.32] | 272 | 270 | 218 | NA | 241 |
| I31 | 1.15 [1.09-1.21] | 1.13 [1.07-1.19] | 1.15 [1.09-1.22] | 0.90 [0.64-1.27] | 1.08 [1.02-1.14] | 1.11 [1.05-1.17] | 1.07 [1.01-1.13] | 1.08 [1.02-1.15] | 0.93 [0.65-1.34] | 1.05 [0.99-1.11] | 4,133 | 4,095 | 3,840 | 104 | 3,884 |
| I33 | 1.28 [1.13-1.45] | 1.20 [1.06-1.37] | 1.20 [1.05-1.36] | | 1.18 [1.04-1.34] | 1.22 [1.08-1.39] | 1.14 [1.00-1.30] | 1.13 [0.99-1.29] | | 1.14 [1.00-1.30] | 730 | 727 | 696 | NA | 694 |
| I34 | 1.09 [1.06-1.13] | 1.10 [1.07-1.14] | 1.10 [1.06-1.14] | | 1.08 [1.04-1.11] | 1.06 [1.02-1.09] | 1.05 [1.02-1.09] | 1.04 [1.01-1.08] | | 1.05 [1.02-1.09] | 10,125 | 10,126 | 9,857 | NA | 9,734 |
| I35 | 1.15 [1.12-1.18] | 1.14 [1.11-1.17] | 1.15 [1.12-1.19] | | 1.12 [1.09-1.15] | 1.11 [1.08-1.14] | 1.08 [1.05-1.11] | 1.09 [1.06-1.12] | | 1.09 [1.06-1.12] | 15,731 | 15,697 | 15,580 | NA | 14,969 |
| I36 | 1.14 [1.03-1.26] | 1.11 [1.00-1.23] | 1.12 [1.01-1.24] | | 1.10 [0.99-1.21] | 1.08 [0.97-1.20] | 1.05 [0.95-1.17] | 1.06 [0.95-1.18] | | 1.07 [0.97-1.19] | 1,131 | 1,117 | 1,089 | NA | 1,122 |
| I37 | 1.16 [1.05-1.29] | 1.18 [1.07-1.31] | 1.19 [1.07-1.32] | | 1.10 [1.00-1.22] | 1.12 [1.00-1.24] | 1.13 [1.02-1.26] | 1.11 [0.99-1.24] | | 1.07 [0.97-1.19] | 1,164 | 1,156 | 1,075 | NA | 1,184 |
| I38 | 1.21 [1.11-1.31] | 1.21 [1.11-1.32] | 1.24 [1.14-1.35] | | 1.15 [1.06-1.25] | 1.16 [1.07-1.26] | 1.15 [1.05-1.25] | 1.17 [1.07-1.27] | | 1.11 [1.02-1.21] | 1,735 | 1,703 | 1,663 | NA | 1,639 |
| I40 | 1.19 [0.83-1.69] | | | | 1.46 [1.03-2.05] | 1.15 [0.81-1.65] | | | | 1.39 [0.97-1.99] | 105 | NA | NA | NA | 109 |
| I42 | 1.11 [1.05-1.18] | 1.14 [1.07-1.20] | 1.09 [1.03-1.16] | | 1.11 [1.05-1.18] | 1.06 [1.00-1.13] | 1.06 [1.00-1.12] | 1.02 [0.96-1.08] | | 1.07 [1.01-1.13] | 3,509 | 3,494 | 3,301 | NA | 3,416 |
| I43 | 0.96 [0.72-1.28] | 0.98 [0.73-1.32] | 0.91 [0.68-1.22] | | 0.99 [0.75-1.30] | 0.93 [0.69-1.25] | 0.92 [0.68-1.25] | 0.85 [0.63-1.15] | | 0.95 [0.72-1.26] | 132 | 131 | 126 | NA | 138 |

### Supplementary Table 4: Hazard ratios and events from all cohorts (excluding non-consulters)

| | Hazard ratio (99% confidence interval) | | | | | | | | | | | | | | |
|---|---|---|---|---|---|---|---|---|---|---|---|---|---|---|---|
| | crude | | | | | adjusted | | | | | Events (in exposed) | | | | |
| Outcome | any age | 18+ | 40+ | <18 | hosp. | any age | 18+ | 40+ | <18 | hosp. | any age | 18+ | 40+ | <18 | hosp. |
| I44 | 1.12 [1.10-1.15] | 1.12 [1.10-1.15] | 1.12 [1.10-1.15] | | 1.11 [1.09-1.14] | 1.09 [1.07-1.12] | 1.07 [1.05-1.10] | 1.07 [1.05-1.10] | | 1.09 [1.07-1.12] | 24,310 | 24,301 | 24,127 | NA | 23,421 |
| I45 | 1.13 [1.10-1.16] | 1.12 [1.09-1.15] | 1.13 [1.09-1.16] | 0.95 [0.72-1.25] | 1.11 [1.08-1.14] | 1.09 [1.06-1.12] | 1.06 [1.03-1.09] | 1.07 [1.03-1.10] | 0.92 [0.69-1.24] | 1.09 [1.05-1.12] | 14,712 | 14,713 | 14,244 | 171 | 14,182 |
| I46 | 1.15 [1.11-1.20] | 1.16 [1.12-1.21] | 1.15 [1.11-1.20] | | 1.14 [1.10-1.19] | 1.13 [1.08-1.17] | 1.11 [1.06-1.15] | 1.10 [1.05-1.14] | | 1.13 [1.08-1.18] | 7,450 | 7,442 | 7,219 | NA | 7,099 |
| I47 | 1.13 [1.08-1.17] | 1.12 [1.08-1.16] | 1.13 [1.08-1.17] | 0.89 [0.68-1.17] | 1.09 [1.04-1.13] | 1.08 [1.03-1.12] | 1.05 [1.01-1.09] | 1.06 [1.01-1.10] | 0.90 [0.68-1.21] | 1.05 [1.01-1.09] | 7,921 | 7,881 | 7,256 | 176 | 7,406 |
| I48 | 1.09 [1.07-1.10] | 1.09 [1.07-1.10] | 1.09 [1.07-1.10] | | 1.08 [1.06-1.09] | 1.06 [1.05-1.08] | 1.05 [1.03-1.06] | 1.05 [1.03-1.06] | | 1.06 [1.05-1.08] | 65,261 | 65,329 | 64,987 | NA | 59,478 |
| I49 | 1.09 [1.05-1.13] | 1.09 [1.06-1.13] | 1.09 [1.05-1.13] | 0.93 [0.71-1.22] | 1.06 [1.03-1.10] | 1.05 [1.01-1.08] | 1.03 [0.99-1.06] | 1.03 [0.99-1.07] | 0.84 [0.63-1.12] | 1.03 [1.00-1.07] | 9,678 | 9,684 | 9,055 | 187 | 9,306 |
| I50 | 1.17 [1.15-1.19] | 1.17 [1.15-1.19] | 1.18 [1.16-1.19] | | 1.16 [1.14-1.18] | 1.13 [1.11-1.15] | 1.10 [1.08-1.11] | 1.10 [1.08-1.12] | | 1.13 [1.11-1.15] | 53,392 | 53,464 | 53,159 | NA | 51,208 |
| I51 | 1.15 [1.13-1.18] | 1.15 [1.13-1.18] | 1.15 [1.13-1.17] | 0.86 [0.66-1.12] | 1.12 [1.10-1.15] | 1.11 [1.08-1.13] | 1.08 [1.06-1.10] | 1.07 [1.05-1.10] | 0.85 [0.65-1.12] | 1.09 [1.07-1.12] | 28,705 | 28,639 | 28,163 | 175 | 28,071 |
| I60 | 1.15 [1.05-1.26] | 1.16 [1.05-1.27] | 1.11 [1.01-1.22] | | 1.08 [0.99-1.19] | 1.14 [1.04-1.26] | 1.13 [1.03-1.24] | 1.08 [0.98-1.19] | | 1.09 [0.99-1.20] | 1,315 | 1,323 | 1,242 | NA | 1,165 |
| I61 | 1.15 [1.09-1.22] | 1.14 [1.07-1.20] | 1.13 [1.07-1.20] | | 1.13 [1.07-1.19] | 1.15 [1.09-1.21] | 1.12 [1.06-1.18] | 1.10 [1.04-1.17] | | 1.13 [1.07-1.19] | 3,764 | 3,734 | 3,682 | NA | 3,472 |
| I62 | 1.10 [1.03-1.19] | 1.13 [1.05-1.22] | 1.11 [1.03-1.20] | | 1.05 [0.98-1.13] | 1.09 [1.02-1.18] | 1.11 [1.03-1.19] | 1.08 [1.00-1.16] | | 1.06 [0.98-1.14] | 2,240 | 2,234 | 2,208 | NA | 2,095 |
| I63 | 1.04 [1.02-1.07] | 1.04 [1.02-1.07] | 1.04 [1.01-1.07] | | 1.04 [1.01-1.06] | 1.04 [1.01-1.06] | 1.02 [0.99-1.04] | 1.01 [0.99-1.04] | | 1.04 [1.01-1.07] | 17,592 | 17,525 | 17,374 | NA | 16,278 |
| I64 | 1.09 [1.04-1.14] | 1.10 [1.04-1.15] | 1.09 [1.04-1.14] | | 1.10 [1.04-1.16] | 1.08 [1.02-1.13] | 1.06 [1.01-1.12] | 1.05 [1.00-1.10] | | 1.10 [1.04-1.16] | 4,490 | 4,502 | 4,471 | NA | 4,030 |
| I65 | 1.10 [1.05-1.15] | 1.08 [1.03-1.14] | 1.10 [1.04-1.15] | | 1.08 [1.02-1.13] | 1.08 [1.02-1.13] | 1.03 [0.98-1.08] | 1.03 [0.98-1.09] | | 1.07 [1.02-1.12] | 4,569 | 4,530 | 4,546 | NA | 4,288 |
| I66 | 1.12 [0.90-1.37] | 1.06 [0.86-1.31] | 1.05 [0.85-1.30] | | 1.03 [0.84-1.26] | 1.09 [0.89-1.35] | 1.02 [0.82-1.27] | 0.99 [0.79-1.23] | | 1.01 [0.82-1.24] | 262 | 254 | 244 | NA | 258 |
| I67 | 1.11 [1.09-1.13] | 1.11 [1.09-1.14] | 1.11 [1.09-1.14] | | 1.10 [1.08-1.12] | 1.09 [1.07-1.11] | 1.07 [1.05-1.09] | 1.06 [1.04-1.09] | | 1.09 [1.07-1.11] | 29,676 | 29,648 | 29,521 | NA | 28,813 |
| I68 | 1.17 [0.91-1.52] | 1.12 [0.87-1.45] | 1.18 [0.92-1.53] | | 1.14 [0.88-1.48] | 1.15 [0.88-1.49] | 1.08 [0.83-1.41] | 1.11 [0.86-1.45] | | 1.10 [0.85-1.43] | 173 | 170 | 170 | NA | 165 |
| I69 | 1.10 [1.06-1.14] | 1.10 [1.06-1.14] | 1.09 [1.05-1.13] | | 1.04 [1.01-1.08] | 1.08 [1.04-1.12] | 1.06 [1.02-1.10] | 1.04 [1.00-1.08] | | 1.05 [1.01-1.08] | 8,442 | 8,401 | 8,317 | NA | 8,146 |
| I70 | 1.16 [1.11-1.21] | 1.15 [1.10-1.19] | 1.17 [1.12-1.21] | | 1.12 [1.08-1.16] | 1.13 [1.09-1.18] | 1.08 [1.04-1.12] | 1.09 [1.05-1.13] | | 1.12 [1.07-1.16] | 7,561 | 7,587 | 7,572 | NA | 7,291 |
| I71 | 1.11 [1.07-1.15] | 1.10 [1.06-1.14] | 1.11 [1.07-1.15] | | 1.08 [1.04-1.12] | 1.10 [1.06-1.15] | 1.06 [1.03-1.10] | 1.06 [1.03-1.11] | | 1.08 [1.04-1.12] | 8,528 | 8,510 | 8,457 | NA | 8,059 |
| I72 | 1.05 [0.97-1.13] | 1.05 [0.98-1.14] | 1.08 [1.00-1.17] | | 1.06 [0.98-1.15] | 1.03 [0.95-1.11] | 1.00 [0.93-1.08] | 1.02 [0.94-1.10] | | 1.05 [0.98-1.14] | 1,971 | 1,971 | 1,912 | NA | 1,891 |
| I73 | 1.27 [1.24-1.31] | 1.27 [1.24-1.30] | 1.26 [1.23-1.30] | 1.13 [0.84-1.53] | 1.21 [1.18-1.25] | 1.22 [1.19-1.25] | 1.17 [1.13-1.20] | 1.16 [1.13-1.19] | 1.09 [0.79-1.50] | 1.18 [1.15-1.21] | 17,224 | 17,298 | 16,664 | 146 | 16,427 |
| I74 | 1.08 [1.03-1.15] | 1.07 [1.01-1.13] | 1.10 [1.05-1.17] | | 1.06 [1.00-1.12] | 1.06 [1.00-1.12] | 1.01 [0.95-1.06] | 1.03 [0.98-1.09] | | 1.05 [1.00-1.11] | 3,879 | 3,877 | 3,819 | NA | 3,675 |
| I77 | 1.20 [1.15-1.26] | 1.19 [1.13-1.24] | 1.21 [1.16-1.27] | | 1.16 [1.11-1.21] | 1.17 [1.12-1.22] | 1.11 [1.06-1.16] | 1.13 [1.08-1.18] | | 1.15 [1.10-1.21] | 5,583 | 5,581 | 5,454 | NA | 5,331 |
| I78 | 1.24 [1.13-1.36] | 1.26 [1.15-1.39] | 1.21 [1.10-1.34] | | 1.18 [1.07-1.29] | 1.19 [1.09-1.31] | 1.19 [1.08-1.30] | 1.15 [1.04-1.27] | | 1.14 [1.04-1.25] | 1,335 | 1,335 | 1,267 | NA | 1,285 |
| I79 | 1.14 [1.00-1.31] | 1.15 [1.00-1.31] | 1.13 [0.99-1.29] | | 1.08 [0.94-1.24] | 1.09 [0.95-1.25] | 1.02 [0.88-1.17] | 1.01 [0.87-1.16] | | 1.05 [0.91-1.20] | 613 | 605 | 600 | NA | 586 |
| I80 | 1.19 [1.15-1.23] | 1.19 [1.15-1.23] | 1.19 [1.15-1.23] | | 1.17 [1.13-1.21] | 1.15 [1.11-1.19] | 1.13 [1.09-1.17] | 1.13 [1.09-1.17] | | 1.13 [1.09-1.17] | 9,973 | 9,998 | 9,379 | NA | 9,315 |
| I81 | 1.07 [0.92-1.23] | 1.11 [0.96-1.28] | 1.06 [0.92-1.23] | | 0.99 [0.86-1.15] | 1.03 [0.89-1.19] | 1.07 [0.92-1.23] | 1.01 [0.87-1.18] | | 1.00 [0.86-1.15] | 526 | 541 | 510 | NA | 506 |
| I82 | 1.12 [1.01-1.23] | 1.14 [1.03-1.26] | 1.15 [1.03-1.27] | | 1.07 [0.97-1.18] | 1.08 [0.98-1.19] | 1.08 [0.98-1.19] | 1.07 [0.96-1.19] | | 1.04 [0.94-1.15] | 1,206 | 1,215 | 1,069 | NA | 1,156 |
| I83 | 1.53 [1.49-1.58] | 1.55 [1.51-1.60] | 1.57 [1.52-1.62] | | 1.50 [1.46-1.55] | 1.50 [1.45-1.54] | 1.50 [1.46-1.55] | 1.51 [1.46-1.56] | | 1.47 [1.42-1.52] | 13,662 | 13,715 | 12,449 | NA | 12,335 |
| I84 | 1.24 [1.21-1.27] | 1.24 [1.20-1.27] | 1.22 [1.19-1.25] | 1.08 [0.86-1.37] | 1.20 [1.17-1.23] | 1.18 [1.15-1.21] | 1.16 [1.13-1.19] | 1.15 [1.12-1.18] | 1.04 [0.81-1.33] | 1.15 [1.12-1.18] | 17,611 | 17,688 | 15,413 | 230 | 15,824 |
| I85 | 1.34 [1.23-1.45] | 1.31 [1.21-1.43] | 1.34 [1.23-1.45] | | 1.32 [1.22-1.43] | 1.29 [1.18-1.40] | 1.22 [1.12-1.33] | 1.23 [1.12-1.34] | | 1.30 [1.19-1.41] | 1,692 | 1,691 | 1,617 | NA | 1,657 |
| I86 | 1.21 [1.09-1.35] | 1.25 [1.12-1.39] | 1.27 [1.13-1.43] | | 1.17 [1.05-1.30] | 1.15 [1.03-1.28] | 1.17 [1.05-1.31] | 1.18 [1.05-1.33] | | 1.15 [1.03-1.28] | 1,037 | 1,024 | 862 | NA | 986 |
| I87 | 1.50 [1.41-1.59] | 1.49 [1.40-1.58] | 1.54 [1.44-1.64] | | 1.46 [1.37-1.55] | 1.44 [1.35-1.53] | 1.40 [1.31-1.49] | 1.45 [1.36-1.55] | | 1.43 [1.34-1.52] | 3,287 | 3,285 | 3,104 | NA | 3,171 |
| I88 | 1.18 [1.04-1.33] | 1.26 [1.05-1.52] | 1.38 [1.09-1.74] | 1.18 [1.02-1.38] | 1.14 [1.01-1.29] | 1.16 [1.02-1.31] | 1.17 [0.97-1.41] | 1.35 [1.06-1.71] | 1.17 [1.00-1.36] | 1.12 [0.98-1.27] | 941 | 367 | 222 | 630 | 815 |
| I89 | 1.40 [1.33-1.48] | 1.39 [1.31-1.46] | 1.41 [1.33-1.49] | | 1.36 [1.29-1.44] | 1.34 [1.26-1.41] | 1.29 [1.22-1.36] | 1.30 [1.23-1.38] | | 1.32 [1.25-1.40] | 4,178 | 4,197 | 4,037 | NA | 4,095 |
| I95 | 1.12 [1.10-1.14] | 1.12 [1.10-1.14] | 1.12 [1.10-1.14] | 0.92 [0.76-1.11] | 1.10 [1.08-1.12] | 1.09 [1.07-1.11] | 1.07 [1.05-1.09] | 1.06 [1.04-1.08] | 0.91 [0.74-1.10] | 1.09 [1.07-1.11] | 35,050 | 35,052 | 33,770 | 379 | 34,239 |
| I97 | 1.22 [1.04-1.43] | 1.22 [1.04-1.43] | 1.19 [1.01-1.41] | | 1.15 [0.98-1.35] | 1.19 [1.01-1.40] | 1.16 [0.99-1.37] | 1.15 [0.97-1.36] | | 1.15 [0.98-1.36] | 428 | 427 | 410 | NA | 407 |
| I98 | 1.30 [1.17-1.44] | 1.34 [1.21-1.48] | 1.32 [1.19-1.46] | | 1.27 [1.15-1.41] | 1.23 [1.11-1.37] | 1.25 [1.12-1.38] | 1.21 [1.09-1.34] | | 1.25 [1.13-1.38] | 1,123 | 1,146 | 1,088 | NA | 1,127 |
| I99 | 1.12 [0.98-1.29] | 1.12 [0.98-1.28] | 1.16 [1.02-1.33] | | 1.11 [0.97-1.27] | 1.10 [0.96-1.26] | 1.06 [0.92-1.21] | 1.10 [0.96-1.27] | | 1.10 [0.96-1.26] | 624 | 624 | 603 | NA | 594 |
| Diseases of the respiratory system (J00-J99) | | | | | | | | | | | | | | | |
| J00 | 1.31 [1.19-1.43] | 1.24 [1.09-1.40] | 1.33 [1.15-1.54] | 1.35 [1.18-1.54] | 1.29 [1.17-1.42] | 1.25 [1.14-1.38] | 1.15 [1.01-1.31] | 1.25 [1.07-1.45] | 1.30 [1.14-1.49] | 1.26 [1.15-1.39] | 1,449 | 739 | 527 | 764 | 1,355 |
| J01 | 1.24 [1.04-1.46] | 1.25 [1.05-1.48] | 1.17 [0.95-1.44] | | 1.15 [0.97-1.35] | 1.11 [0.93-1.33] | 1.11 [0.93-1.33] | 1.02 [0.83-1.27] | | 1.07 [0.90-1.27] | 434 | 413 | 272 | NA | 409 |
| J02 | 1.27 [1.19-1.36] | 1.27 [1.19-1.37] | 1.31 [1.20-1.43] | 1.04 [0.91-1.19] | 1.21 [1.14-1.30] | 1.20 [1.12-1.28] | 1.16 [1.08-1.25] | 1.19 [1.08-1.30] | 1.01 [0.88-1.16] | 1.17 [1.09-1.25] | 2,827 | 2,344 | 1,499 | 783 | 2,640 |
| J03 | 1.19 [1.14-1.23] | 1.18 [1.12-1.24] | 1.16 [1.03-1.31] | 1.14 [1.09-1.19] | 1.14 [1.10-1.19] | 1.14 [1.09-1.18] | 1.08 [1.03-1.14] | 1.02 [0.90-1.16] | 1.11 [1.06-1.16] | 1.11 [1.06-1.15] | 9,686 | 4,572 | 755 | 6,950 | 8,569 |
| J04 | 1.20 [1.01-1.42] | 1.37 [1.15-1.64] | 1.25 [1.02-1.52] | | 1.29 [1.08-1.53] | 1.07 [0.90-1.28] | 1.20 [0.99-1.44] | 1.07 [0.87-1.32] | | 1.20 [1.00-1.43] | 420 | 382 | 293 | NA | 386 |
| J05 | 1.00 [0.92-1.09] | 0.97 [0.73-1.30] | 1.15 [0.84-1.56] | 1.01 [0.93-1.11] | 1.01 [0.92-1.10] | 0.98 [0.90-1.07] | 0.91 [0.68-1.22] | 1.02 [0.74-1.41] | 0.99 [0.90-1.08] | 0.99 [0.90-1.08] | 1,540 | 135 | 118 | 1,417 | 1,401 |
| J06 | 1.28 [1.23-1.33] | 1.26 [1.18-1.35] | 1.26 [1.17-1.36] | 1.28 [1.22-1.34] | 1.25 [1.20-1.30] | 1.22 [1.17-1.27] | 1.12 [1.05-1.20] | 1.14 [1.05-1.23] | 1.25 [1.19-1.31] | 1.20 [1.15-1.25] | 7,951 | 2,718 | 1,916 | 5,508 | 7,331 |
| J10 | 1.22 [1.15-1.30] | 1.25 [1.17-1.34] | 1.24 [1.16-1.33] | 1.17 [0.96-1.43] | 1.22 [1.15-1.30] | 1.13 [1.06-1.20] | 1.11 [1.04-1.19] | 1.11 [1.03-1.19] | 1.11 [0.90-1.36] | 1.14 [1.07-1.22] | 3,180 | 2,962 | 2,556 | 344 | 3,138 |
| J11 | 1.17 [1.01-1.35] | 1.19 [1.02-1.37] | 1.24 [1.05-1.46] | | 1.15 [1.00-1.33] | 1.07 [0.92-1.24] | 1.06 [0.91-1.23] | 1.11 [0.94-1.32] | | 1.09 [0.94-1.26] | 588 | 556 | 437 | NA | 576 |
| J12 | 1.16 [1.11-1.22] | 1.16 [1.10-1.21] | 1.17 [1.12-1.23] | 1.06 [0.81-1.37] | 1.16 [1.11-1.22] | 1.10 [1.04-1.15] | 1.07 [1.02-1.13] | 1.08 [1.03-1.14] | 1.00 [0.76-1.31] | 1.12 [1.06-1.17] | 5,383 | 5,253 | 5,042 | 188 | 5,318 |
| J13 | 1.23 [1.10-1.36] | 1.30 [1.17-1.44] | 1.26 [1.13-1.40] | | 1.29 [1.16-1.43] | 1.14 [1.03-1.27] | 1.18 [1.06-1.31] | 1.13 [1.01-1.26] | | 1.21 [1.09-1.35] | 1,106 | 1,094 | 1,028 | NA | 1,034 |
| J14 | 1.23 [1.05-1.44] | 1.30 [1.11-1.53] | 1.24 [1.05-1.45] | | 1.25 [1.07-1.46] | 1.10 [0.94-1.30] | 1.11 [0.94-1.31] | 1.04 [0.88-1.23] | | 1.12 [0.96-1.31] | 482 | 469 | 452 | NA | 477 |
| J15 | 1.28 [1.19-1.37] | 1.30 [1.21-1.40] | 1.30 [1.21-1.40] | 1.28 [0.91-1.79] | 1.30 [1.21-1.39] | 1.18 [1.10-1.27] | 1.18 [1.10-1.27] | 1.16 [1.08-1.25] | 1.27 [0.90-1.79] | 1.22 [1.14-1.31] | 2,530 | 2,435 | 2,344 | 123 | 2,443 |
| J17 | 1.34 [1.15-1.56] | 1.32 [1.13-1.53] | 1.23 [1.05-1.44] | | 1.27 [1.09-1.48] | 1.26 [1.08-1.47] | 1.22 [1.04-1.43] | 1.13 [0.96-1.33] | | 1.19 [1.02-1.40] | 498 | 495 | 469 | NA | 477 |
| J18 | 1.18 [1.17-1.20] | 1.18 [1.16-1.19] | 1.18 [1.16-1.19] | 1.20 [1.11-1.30] | 1.16 [1.15-1.18] | 1.14 [1.12-1.15] | 1.11 [1.09-1.12] | 1.11 [1.09-1.12] | 1.18 [1.09-1.27] | 1.13 [1.12-1.15] | 72,453 | 70,790 | 68,632 | 2,192 | 69,584 |
| J20 | 1.21 [1.08-1.35] | 1.21 [1.08-1.36] | 1.22 [1.08-1.38] | | 1.22 [1.09-1.36] | 1.11 [0.99-1.25] | 1.09 [0.97-1.23] | 1.11 [0.98-1.27] | | 1.13 [1.00-1.27] | 926 | 855 | 777 | NA | 894 |
| J21 | 1.17 [1.01-1.35] | 1.28 [1.02-1.59] | 1.22 [0.97-1.54] | 1.05 [0.87-1.26] | 1.13 [0.98-1.31] | 1.09 [0.94-1.27] | 1.11 [0.88-1.40] | 1.09 [0.86-1.39] | 0.99 [0.82-1.20] | 1.04 [0.89-1.20] | 564 | 240 | 219 | 330 | 523 |
| J22 | 1.24 [1.22-1.26] | 1.23 [1.21-1.25] | 1.21 [1.19-1.23] | 1.35 [1.28-1.42] | 1.20 [1.18-1.22] | 1.18 [1.16-1.20] | 1.14 [1.12-1.16] | 1.13 [1.11-1.15] | 1.29 [1.22-1.36] | 1.16 [1.14-1.18] | 44,787 | 40,875 | 37,563 | 4,816 | 42,632 |
| J30 | 1.77 [1.69-1.85] | 1.58 [1.51-1.66] | 1.50 [1.42-1.58] | 2.91 [2.64-3.21] | 1.73 [1.66-1.80] | 1.45 [1.38-1.51] | 1.26 [1.20-1.32] | 1.23 [1.15-1.30] | 2.45 [2.22-2.72] | 1.41 [1.35-1.48] | 8,255 | 6,685 | 4,111 | 2,475 | 7,484 |
| J31 | 1.50 [1.37-1.63] | 1.54 [1.41-1.69] | 1.46 [1.31-1.62] | 1.72 [1.34-2.20] | 1.46 [1.34-1.60] | 1.25 [1.14-1.37] | 1.28 [1.16-1.41] | 1.19 [1.07-1.33] | 1.45 [1.12-1.89] | 1.23 [1.12-1.35] | 1,726 | 1,577 | 1,128 | 280 | 1,598 |
| J32 | 1.26 [1.19-1.33] | 1.29 [1.22-1.37] | 1.27 [1.19-1.36] | 1.23 [1.00-1.52] | 1.24 [1.17-1.31] | 1.10 [1.04-1.17] | 1.11 [1.04-1.18] | 1.12 [1.05-1.19] | 1.15 [0.92-1.43] | 1.10 [1.04-1.17] | 3,916 | 3,815 | 2,877 | 326 | 3,615 |
| J33 | 1.40 [1.30-1.50] | 1.37 [1.28-1.47] | 1.37 [1.27-1.48] | 1.87 [1.28-2.73] | 1.33 [1.24-1.43] | 1.20 [1.12-1.29] | 1.16 [1.08-1.25] | 1.18 [1.09-1.28] | 1.71 [1.15-2.55] | 1.15 [1.07-1.24] | 2,619 | 2,550 | 2,082 | 117 | 2,194 |
| J34 | 1.32 [1.27-1.38] | 1.30 [1.25-1.36] | 1.30 [1.23-1.37] | 1.37 [1.23-1.52] | 1.28 [1.23-1.34] | 1.20 [1.15-1.25] | 1.15 [1.10-1.21] | 1.17 [1.11-1.23] | 1.28 [1.15-1.43] | 1.17 [1.12-1.22] | 7,296 | 6,696 | 4,250 | 1,370 | 6,322 |
| J35 | 1.18 [1.13-1.24] | 1.19 [1.11-1.28] | 1.14 [1.01-1.29] | 1.19 [1.13-1.26] | 1.17 [1.11-1.22] | 1.14 [1.09-1.19] | 1.12 [1.04-1.20] | 1.05 [0.93-1.19] | 1.16 [1.10-1.22] | 1.13 [1.08-1.19] | 6,855 | 2,312 | 720 | 5,158 | 5,798 |
| J36 | 1.05 [0.93-1.17] | 1.06 [0.95-1.18] | 1.07 [0.88-1.30] | 1.10 [0.87-1.39] | 1.05 [0.93-1.18] | 1.05 [0.93-1.18] | 1.06 [0.95-1.19] | 1.05 [0.87-1.28] | 1.13 [0.88-1.45] | 1.05 [0.93-1.19] | 939 | 951 | 301 | 244 | 811 |
| J38 | 1.20 [1.13-1.28] | 1.21 [1.14-1.29] | 1.23 [1.15-1.31] | 1.13 [0.87-1.47] | 1.15 [1.08-1.23] | 1.14 [1.07-1.22] | 1.09 [1.02-1.17] | 1.13 [1.06-1.21] | 1.05 [0.80-1.38] | 1.10 [1.03-1.17] | 3,002 | 2,897 | 2,632 | 187 | 2,757 |
| J39 | 1.20 [1.10-1.31] | 1.21 [1.11-1.32] | 1.19 [1.08-1.31] | 1.09 [0.81-1.46] | 1.14 [1.05-1.24] | 1.12 [1.03-1.23] | 1.10 [1.00-1.20] | 1.09 [0.99-1.20] | 1.03 [0.76-1.39] | 1.08 [0.99-1.18] | 1,589 | 1,484 | 1,308 | 157 | 1,519 |
| J40 | 1.31 [1.21-1.41] | 1.30 [1.21-1.40] | 1.30 [1.20-1.40] | | 1.23 [1.14-1.32] | 1.19 [1.10-1.28] | 1.15 [1.07-1.25] | 1.15 [1.06-1.24] | | 1.12 [1.04-1.21] | 2,142 | 2,125 | 1,985 | NA | 1,999 |
| J42 | 1.37 [1.22-1.54] | 1.35 [1.20-1.52] | 1.31 [1.16-1.47] | | 1.31 [1.17-1.47] | 1.23 [1.10-1.39] | 1.19 [1.06-1.34] | 1.13 [1.00-1.28] | | 1.18 [1.04-1.33] | 897 | 880 | 853 | NA | 839 |
| J43 | 1.27 [1.23-1.31] | 1.26 [1.22-1.30] | 1.27 [1.23-1.31] | | 1.24 [1.21-1.28] | 1.19 [1.15-1.23] | 1.13 [1.10-1.17] | 1.15 [1.11-1.18] | | 1.18 [1.14-1.22] | 11,572 | 11,588 | 11,449 | NA | 11,218 |
| J44 | 1.32 [1.30-1.34] | 1.32 [1.30-1.34] | 1.31 [1.29-1.34] | | 1.29 [1.27-1.31] | 1.19 [1.17-1.21] | 1.14 [1.12-1.16] | 1.14 [1.12-1.16] | | 1.17 [1.15-1.19] | 49,128 | 49,132 | 48,741 | NA | 45,504 |
| J45 | 1.72 [1.69-1.74] | 1.65 [1.63-1.68] | 1.57 [1.54-1.59] | 2.22 [2.14-2.29] | 1.65 [1.63-1.67] | 1.32 [1.30-1.34] | 1.21 [1.20-1.23] | 1.20 [1.18-1.22] | 1.86 [1.79-1.93] | 1.29 [1.27-1.31] | 85,315 | 77,417 | 53,422 | 15,741 | 68,712 |
| J46 | 2.24 [2.01-2.48] | 1.81 [1.61-2.05] | 1.67 [1.44-1.94] | 3.34 [2.75-4.04] | 2.05 [1.84-2.28] | 1.74 [1.54-1.95] | 1.29 [1.12-1.50] | 1.17 [0.98-1.39] | 2.85 [2.33-3.48] | 1.57 [1.40-1.77] | 1,451 | 952 | 574 | 616 | 1,323 |
| J47 | 1.34 [1.30-1.39] | 1.35 [1.30-1.40] | 1.33 [1.29-1.38] | | 1.32 [1.27-1.37] | 1.19 [1.14-1.23] | 1.17 [1.13-1.22] | 1.17 [1.12-1.21] | | 1.17 [1.12-1.21] | 9,197 | 9,196 | 8,977 | NA | 8,996 |
| J61 | 1.28 [1.16-1.41] | 1.30 [1.18-1.44] | 1.29 [1.16-1.42] | | 1.29 [1.17-1.42] | 1.22 [1.10-1.35] | 1.23 [1.11-1.36] | 1.20 [1.09-1.33] | | 1.23 [1.12-1.36] | 1,223 | 1,228 | 1,215 | NA | 1,212 |
| J67 | 1.39 [1.16-1.66] | 1.45 [1.21-1.73] | 1.38 [1.15-1.66] | | 1.33 [1.11-1.59] | 1.27 [1.05-1.53] | 1.26 [1.04-1.52] | 1.26 [1.04-1.52] | | 1.22 [1.01-1.46] | 375 | 380 | 350 | NA | 351 |
| J69 | 1.09 [1.06-1.13] | 1.10 [1.06-1.14] | 1.11 [1.07-1.14] | | 1.10 [1.06-1.14] | 1.09 [1.05-1.13] | 1.08 [1.04-1.11] | 1.08 [1.04-1.11] | | 1.11 [1.07-1.15] | 10,736 | 10,688 | 10,511 | NA | 10,551 |
| J70 | 1.40 [1.13-1.74] | 1.37 [1.11-1.70] | 1.44 [1.15-1.79] | | 1.29 [1.04-1.60] | 1.31 [1.05-1.63] | 1.25 [1.00-1.56] | 1.33 [1.06-1.67] | | 1.22 [0.98-1.52] | 247 | 249 | 243 | NA | 243 |
| J80 | 1.27 [1.13-1.43] | 1.29 [1.15-1.46] | 1.30 [1.15-1.47] | | 1.26 [1.12-1.42] | 1.22 [1.08-1.38] | 1.23 [1.09-1.39] | 1.21 [1.07-1.38] | | 1.26 [1.12-1.42] | 844 | 832 | 779 | NA | 803 |
| J81 | 1.20 [1.14-1.27] | 1.20 [1.14-1.27] | 1.20 [1.14-1.27] | | 1.15 [1.09-1.21] | 1.17 [1.11-1.24] | 1.13 [1.07-1.20] | 1.14 [1.08-1.21] | | 1.13 [1.07-1.20] | 3,873 | 3,907 | 3,819 | NA | 3,671 |
| J82 | 1.86 [1.54-2.25] | 1.85 [1.53-2.23] | 1.68 [1.37-2.05] | | 1.83 [1.52-2.21] | 1.35 [1.09-1.68] | 1.35 [1.09-1.68] | 1.23 [0.98-1.54] | | 1.46 [1.19-1.79] | 404 | 418 | 332 | NA | 399 |
| J84 | 1.30 [1.25-1.35] | 1.33 [1.28-1.38] | 1.31 [1.26-1.36] | | 1.32 [1.27-1.37] | 1.25 [1.20-1.30] | 1.25 [1.20-1.30] | 1.24 [1.19-1.29] | | 1.28 [1.23-1.33] | 7,855 | 7,858 | 7,775 | NA | 7,625 |
| J85 | 1.30 [1.08-1.56] | 1.33 [1.10-1.61] | 1.26 [1.04-1.52] | | 1.27 [1.06-1.53] | 1.21 [1.00-1.46] | 1.24 [1.02-1.51] | 1.17 [0.96-1.43] | | 1.23 [1.02-1.48] | 344 | 331 | 314 | NA | 323 |
| J86 | 1.18 [1.07-1.31] | 1.22 [1.10-1.36] | 1.21 [1.09-1.35] | | 1.13 [1.02-1.25] | 1.16 [1.04-1.28] | 1.17 [1.05-1.30] | 1.14 [1.02-1.28] | | 1.12 [1.01-1.24] | 1,118 | 1,066 | 1,004 | NA | 1,021 |
| J90 | 1.13 [1.11-1.15] | 1.12 [1.10-1.15] | 1.13 [1.11-1.15] | 0.99 [0.81-1.20] | 1.11 [1.09-1.13] | 1.11 [1.08-1.13] | 1.08 [1.06-1.10] | 1.08 [1.06-1.10] | 1.04 [0.84-1.28] | 1.10 [1.08-1.12] | 34,393 | 34,273 | 33,495 | 323 | 33,010 |
| J91 | 1.19 [1.05-1.34] | 1.15 [1.02-1.30] | 1.17 [1.03-1.32] | | 1.19 [1.05-1.34] | 1.16 [1.03-1.32] | 1.09 [0.96-1.23] | 1.10 [0.97-1.24] | | 1.18 [1.05-1.33] | 822 | 822 | 802 | NA | 813 |
| J92 | 1.20 [1.14-1.27] | 1.23 [1.16-1.30] | 1.22 [1.15-1.28] | | 1.19 [1.13-1.25] | 1.17 [1.11-1.24] | 1.18 [1.12-1.25] | 1.17 [1.11-1.24] | | 1.17 [1.10-1.23] | 4,095 | 4,094 | 4,070 | NA | 4,001 |
| J93 | 1.08 [1.01-1.16] | 1.07 [1.00-1.14] | 1.08 [1.01-1.16] | 0.97 [0.69-1.35] | 1.06 [0.99-1.14] | 1.06 [0.99-1.13] | 1.03 [0.96-1.10] | 1.04 [0.97-1.12] | 1.00 [0.71-1.41] | 1.05 [0.98-1.13] | 2,609 | 2,543 | 2,317 | 121 | 2,414 |
| J94 | 1.18 [1.08-1.30] | 1.16 [1.05-1.28] | 1.16 [1.05-1.28] | | 1.10 [1.00-1.21] | 1.16 [1.06-1.28] | 1.11 [1.00-1.22] | 1.10 [1.00-1.21] | | 1.09 [0.99-1.20] | 1,258 | 1,254 | 1,219 | NA | 1,197 |
| J95 | 1.14 [1.02-1.27] | 1.08 [0.97-1.20] | 1.12 [1.00-1.25] | | 1.11 [0.99-1.24] | 1.12 [1.00-1.24] | 1.01 [0.91-1.13] | 1.07 [0.95-1.20] | | 1.10 [0.99-1.23] | 924 | 894 | 868 | NA | 847 |
| J96 | 1.22 [1.19-1.24] | 1.20 [1.18-1.23] | 1.20 [1.18-1.23] | 1.01 [0.78-1.32] | 1.19 [1.17-1.22] | 1.15 [1.13-1.18] | 1.10 [1.08-1.13] | 1.10 [1.08-1.13] | 0.95 [0.72-1.26] | 1.15 [1.12-1.17] | 23,696 | 23,699 | 23,196 | 183 | 23,167 |
| J98 | 1.16 [1.14-1.19] | 1.15 [1.12-1.18] | 1.15 [1.13-1.18] | 1.19 [1.03-1.37] | 1.14 [1.11-1.16] | 1.10 [1.08-1.13] | 1.06 [1.04-1.09] | 1.07 [1.04-1.10] | 1.13 [0.97-1.30] | 1.10 [1.07-1.12] | 20,600 | 20,132 | 19,337 | 685 | 20,025 |
| J99 | 1.44 [1.26-1.64] | 1.44 [1.27-1.64] | 1.46 [1.27-1.66] | | 1.39 [1.22-1.57] | 1.27 [1.11-1.45] | 1.29 [1.13-1.48] | 1.32 [1.15-1.51] | | 1.24 [1.09-1.42] | 722 | 724 | 667 | NA | 729 |
| Diseases of the digestive system (K00-K95) | | | | | | | | | | | | | | | |
| K00 | 1.15 [1.04-1.26] | 1.13 [1.00-1.28] | 1.17 [0.91-1.50] | 1.11 [0.99-1.26] | 1.06 [0.96-1.18] | 1.11 [1.01-1.22] | 1.08 [0.94-1.23] | 1.09 [0.84-1.41] | 1.09 [0.96-1.23] | 1.05 [0.95-1.17] | 1,578 | 766 | 178 | 1,019 | 1,265 |
| K01 | 1.15 [1.10-1.21] | 1.15 [1.10-1.21] | 1.09 [0.97-1.22] | 1.15 [1.05-1.26] | 1.09 [1.03-1.15] | 1.10 [1.05-1.16] | 1.08 [1.03-1.13] | 1.05 [0.94-1.18] | 1.11 [1.01-1.22] | 1.05 [0.99-1.10] | 5,622 | 5,317 | 854 | 1,757 | 4,342 |
| K02 | 1.11 [1.08-1.14] | 1.16 [1.13-1.20] | 1.19 [1.14-1.25] | 0.95 [0.91-1.00] | 1.07 [1.04-1.10] | 1.06 [1.03-1.10] | 1.07 [1.03-1.11] | 1.10 [1.05-1.15] | 0.95 [0.90-1.00] | 1.04 [1.01-1.08] | 14,861 | 10,641 | 5,775 | 5,478 | 13,078 |
| K03 | 1.21 [1.01-1.45] | 1.38 [1.12-1.70] | 1.38 [1.08-1.77] | 0.96 [0.72-1.29] | 1.15 [0.96-1.38] | 1.15 [0.95-1.38] | 1.30 [1.05-1.62] | 1.26 [0.98-1.63] | 0.93 [0.68-1.26] | 1.12 [0.93-1.34] | 411 | 287 | 192 | 162 | 376 |
| K04 | 1.11 [1.05-1.17] | 1.13 [1.06-1.19] | 1.20 [1.12-1.29] | 0.87 [0.76-0.99] | 1.08 [1.02-1.14] | 1.05 [0.99-1.11] | 1.03 [0.97-1.10] | 1.10 [1.03-1.19] | 0.88 [0.77-1.00] | 1.04 [0.99-1.10] | 4,067 | 3,592 | 2,350 | 797 | 3,673 |
| K05 | 1.20 [1.14-1.26] | 1.21 [1.15-1.27] | 1.24 [1.14-1.36] | 1.14 [1.01-1.29] | 1.14 [1.08-1.21] | 1.14 [1.08-1.20] | 1.11 [1.06-1.17] | 1.14 [1.04-1.25] | 1.13 [0.99-1.29] | 1.11 [1.05-1.17] | 4,660 | 5,027 | 1,456 | 913 | 3,958 |
| K06 | 1.29 [1.13-1.47] | 1.26 [1.10-1.44] | 1.31 [1.13-1.52] | | 1.16 [1.02-1.33] | 1.24 [1.08-1.42] | 1.21 [1.05-1.39] | 1.21 [1.04-1.41] | | 1.12 [0.97-1.29] | 662 | 644 | 524 | NA | 605 |
| K07 | 1.13 [1.05-1.21] | 1.19 [1.10-1.28] | 1.17 [1.02-1.34] | 1.08 [0.97-1.20] | 1.14 [1.06-1.23] | 1.07 [1.00-1.15] | 1.08 [1.00-1.17] | 1.05 [0.92-1.21] | 1.05 [0.94-1.18] | 1.10 [1.02-1.19] | 2,700 | 2,147 | 622 | 1,229 | 2,266 |
| K08 | 1.16 [1.11-1.21] | 1.19 [1.14-1.24] | 1.22 [1.15-1.28] | 0.94 [0.83-1.06] | 1.13 [1.08-1.18] | 1.11 [1.06-1.16] | 1.10 [1.05-1.16] | 1.12 [1.07-1.19] | 0.93 [0.82-1.05] | 1.09 [1.04-1.14] | 6,461 | 6,008 | 4,167 | 965 | 5,913 |
| K09 | 1.20 [1.03-1.41] | 1.13 [0.96-1.33] | 1.12 [0.93-1.35] | | 1.18 [0.99-1.39] | 1.16 [0.98-1.37] | 1.06 [0.90-1.25] | 1.10 [0.91-1.33] | | 1.12 [0.94-1.33] | 471 | 431 | 314 | NA | 383 |
| K10 | 1.17 [1.04-1.32] | 1.16 [1.03-1.31] | 1.19 [1.04-1.36] | | 1.13 [1.00-1.28] | 1.09 [0.96-1.23] | 1.04 [0.92-1.18] | 1.09 [0.95-1.25] | | 1.09 [0.97-1.23] | 785 | 785 | 651 | NA | 766 |
| K11 | 1.29 [1.19-1.40] | 1.27 [1.17-1.38] | 1.28 [1.17-1.40] | 1.02 [0.77-1.35] | 1.22 [1.12-1.32] | 1.24 [1.14-1.35] | 1.20 [1.11-1.31] | 1.20 [1.09-1.32] | 1.00 [0.74-1.35] | 1.17 [1.07-1.27] | 1,749 | 1,665 | 1,405 | 162 | 1,616 |
| K12 | 1.25 [1.17-1.34] | 1.24 [1.16-1.33] | 1.28 [1.18-1.38] | 1.16 [0.97-1.40] | 1.19 [1.11-1.27] | 1.20 [1.12-1.29] | 1.17 [1.09-1.26] | 1.20 [1.11-1.30] | 1.19 [0.99-1.44] | 1.16 [1.08-1.24] | 2,730 | 2,442 | 2,066 | 406 | 2,549 |
| K13 | 1.28 [1.21-1.36] | 1.27 [1.20-1.35] | 1.29 [1.21-1.37] | 1.22 [0.99-1.50] | 1.26 [1.19-1.34] | 1.22 [1.15-1.29] | 1.19 [1.12-1.26] | 1.20 [1.12-1.28] | 1.16 [0.94-1.43] | 1.22 [1.15-1.29] | 3,653 | 3,476 | 2,943 | 328 | 3,297 |
| K14 | 1.30 [1.21-1.40] | 1.29 [1.19-1.39] | 1.29 [1.19-1.39] | 1.23 [0.88-1.71] | 1.26 [1.17-1.36] | 1.23 [1.13-1.33] | 1.17 [1.08-1.27] | 1.19 [1.10-1.29] | 1.12 [0.79-1.58] | 1.20 [1.11-1.30] | 2,026 | 1,946 | 1,744 | 126 | 1,896 |
| K20 | 1.20 [1.16-1.23] | 1.19 [1.16-1.23] | 1.18 [1.15-1.22] | 1.32 [1.06-1.66] | 1.15 [1.12-1.19] | 1.12 [1.09-1.16] | 1.10 [1.06-1.13] | 1.09 [1.06-1.13] | 1.20 [0.95-1.52] | 1.10 [1.06-1.13] | 12,640 | 12,544 | 11,699 | 300 | 11,728 |
| K21 | 1.22 [1.20-1.24] | 1.22 [1.20-1.24] | 1.22 [1.20-1.24] | 1.03 [0.94-1.13] | 1.18 [1.16-1.20] | 1.11 [1.09-1.13] | 1.10 [1.08-1.11] | 1.10 [1.08-1.12] | 0.93 [0.84-1.03] | 1.09 [1.08-1.11] | 51,515 | 51,270 | 46,144 | 1,507 | 48,545 |
| K22 | 1.19 [1.16-1.22] | 1.19 [1.16-1.22] | 1.18 [1.15-1.21] | 1.05 [0.85-1.30] | 1.16 [1.13-1.19] | 1.14 [1.11-1.16] | 1.11 [1.09-1.14] | 1.11 [1.08-1.14] | 0.96 [0.77-1.21] | 1.11 [1.08-1.14] | 19,006 | 18,956 | 18,032 | 289 | 17,638 |
| K23 | 1.36 [1.09-1.69] | 1.29 [1.04-1.61] | 1.38 [1.10-1.72] | | 1.36 [1.10-1.69] | 1.26 [1.01-1.57] | 1.14 [0.91-1.43] | 1.21 [0.96-1.52] | | 1.24 [1.00-1.54] | 263 | 262 | 259 | NA | 254 |
| K25 | 1.19 [1.15-1.23] | 1.19 [1.15-1.23] | 1.17 [1.13-1.21] | | 1.16 [1.12-1.20] | 1.14 [1.10-1.18] | 1.11 [1.07-1.15] | 1.10 [1.06-1.14] | | 1.13 [1.09-1.17] | 9,750 | 9,770 | 9,274 | NA | 9,017 |
| K26 | 1.11 [1.07-1.16] | 1.12 [1.07-1.17] | 1.11 [1.07-1.16] | | 1.11 [1.06-1.16] | 1.10 [1.05-1.14] | 1.08 [1.03-1.12] | 1.08 [1.03-1.12] | | 1.10 [1.06-1.15] | 6,495 | 6,483 | 6,249 | NA | 5,953 |
| K27 | 1.13 [1.03-1.25] | 1.13 [1.03-1.25] | 1.14 [1.03-1.26] | | 1.13 [1.03-1.25] | 1.06 [0.96-1.18] | 1.03 [0.94-1.14] | 1.05 [0.95-1.16] | | 1.09 [0.99-1.21] | 1,180 | 1,186 | 1,113 | NA | 1,141 |
| K28 | 1.29 [1.00-1.66] | 1.34 [1.04-1.73] | 1.29 [0.99-1.69] | | 1.39 [1.08-1.77] | 1.17 [0.91-1.52] | 1.15 [0.88-1.50] | 1.08 [0.81-1.43] | | 1.29 [1.00-1.66] | 190 | 187 | 161 | NA | 198 |
| K29 | 1.20 [1.18-1.22] | 1.20 [1.18-1.22] | 1.20 [1.18-1.22] | 1.01 [0.92-1.12] | 1.16 [1.14-1.18] | 1.13 [1.12-1.15] | 1.11 [1.09-1.13] | 1.11 [1.09-1.13] | 0.95 [0.86-1.06] | 1.11 [1.09-1.13] | 48,814 | 48,504 | 44,100 | 1,375 | 45,096 |
| K30 | 1.22 [1.18-1.25] | 1.21 [1.18-1.25] | 1.21 [1.17-1.25] | 1.22 [0.95-1.56] | 1.19 [1.15-1.23] | 1.13 [1.09-1.17] | 1.11 [1.07-1.14] | 1.12 [1.08-1.16] | 1.13 [0.87-1.47] | 1.12 [1.08-1.16] | 11,654 | 11,723 | 10,147 | 221 | 10,613 |
| K31 | 1.18 [1.15-1.21] | 1.18 [1.15-1.21] | 1.20 [1.17-1.23] | 0.93 [0.70-1.23] | 1.15 [1.12-1.18] | 1.10 [1.07-1.13] | 1.07 [1.05-1.10] | 1.09 [1.06-1.12] | 0.91 [0.67-1.22] | 1.08 [1.05-1.11] | 17,145 | 17,143 | 16,337 | 175 | 16,558 |
| K35 | 1.03 [0.98-1.09] | 1.05 [1.00-1.11] | 1.01 [0.94-1.09] | 0.95 [0.86-1.05] | 1.00 [0.95-1.05] | 1.00 [0.95-1.06] | 1.01 [0.95-1.06] | 0.99 [0.92-1.06] | 0.93 [0.83-1.03] | 0.99 [0.93-1.04] | 4,838 | 4,229 | 2,120 | 1,411 | 4,024 |
| K36 | 0.98 [0.73-1.33] | 1.08 [0.80-1.47] | | | 1.00 [0.73-1.37] | 0.96 [0.70-1.30] | 0.98 [0.71-1.35] | | | 0.92 [0.66-1.28] | 118 | 120 | NA | NA | 109 |
| K37 | 1.04 [0.93-1.16] | 1.06 [0.95-1.19] | 1.05 [0.88-1.26] | 0.99 [0.80-1.22] | 1.01 [0.90-1.13] | 0.99 [0.88-1.11] | 0.98 [0.87-1.10] | 0.99 [0.83-1.20] | 0.93 [0.75-1.17] | 0.96 [0.85-1.08] | 965 | 888 | 327 | 309 | 848 |
| K38 | 1.02 [0.90-1.16] | 1.03 [0.90-1.16] | 1.02 [0.86-1.21] | 0.87 [0.66-1.15] | 0.98 [0.87-1.12] | 0.98 [0.86-1.12] | 0.94 [0.82-1.07] | 0.97 [0.82-1.15] | 0.85 [0.63-1.14] | 0.96 [0.84-1.10] | 747 | 715 | 395 | 172 | 675 |
| K40 | 1.04 [1.01-1.07] | 1.05 [1.02-1.08] | 1.05 [1.02-1.08] | 0.94 [0.82-1.09] | 1.04 [1.01-1.07] | 1.01 [0.99-1.04] | 1.03 [1.00-1.06] | 1.04 [1.01-1.07] | 0.91 [0.79-1.05] | 1.02 [0.99-1.05] | 14,245 | 13,659 | 13,188 | 626 | 12,360 |
| K41 | 1.12 [1.01-1.23] | 1.12 [1.01-1.23] | 1.14 [1.03-1.26] | | 1.08 [0.98-1.20] | 1.06 [0.96-1.17] | 1.08 [0.97-1.19] | 1.09 [0.98-1.21] | | 1.04 [0.94-1.15] | 1,186 | 1,186 | 1,106 | NA | 1,093 |
| K42 | 1.17 [1.12-1.22] | 1.15 [1.11-1.20] | 1.17 [1.12-1.22] | 1.03 [0.86-1.23] | 1.16 [1.11-1.21] | 1.08 [1.04-1.13] | 1.05 [1.01-1.10] | 1.06 [1.01-1.11] | 0.98 [0.81-1.18] | 1.09 [1.05-1.14] | 7,649 | 7,387 | 6,396 | 406 | 7,200 |
| K43 | 1.16 [1.11-1.21] | 1.17 [1.12-1.22] | 1.18 [1.13-1.23] | 1.07 [0.82-1.41] | 1.12 [1.08-1.17] | 1.09 [1.04-1.14] | 1.07 [1.03-1.12] | 1.09 [1.04-1.14] | 1.04 [0.78-1.39] | 1.08 [1.04-1.13] | 6,341 | 6,260 | 5,692 | 180 | 6,234 |
| K44 | 1.19 [1.17-1.21] | 1.19 [1.17-1.21] | 1.18 [1.16-1.20] | 1.07 [0.90-1.26] | 1.15 [1.14-1.17] | 1.11 [1.10-1.13] | 1.10 [1.08-1.12] | 1.09 [1.07-1.11] | 0.97 [0.81-1.16] | 1.09 [1.07-1.11] | 48,147 | 48,297 | 45,224 | 491 | 44,437 |
| K45 | 1.14 [0.94-1.38] | 1.07 [0.88-1.30] | 0.99 [0.81-1.21] | | 1.11 [0.92-1.34] | 1.06 [0.87-1.29] | 0.97 [0.79-1.18] | 0.91 [0.74-1.12] | | 1.02 [0.84-1.24] | 299 | 297 | 270 | NA | 301 |
| K46 | 1.18 [1.08-1.30] | 1.19 [1.08-1.31] | 1.20 [1.09-1.32] | | 1.16 [1.06-1.27] | 1.09 [0.99-1.20] | 1.07 [0.97-1.17] | 1.09 [0.98-1.20] | | 1.10 [1.01-1.21] | 1,370 | 1,359 | 1,295 | NA | 1,374 |
| K50 | 1.44 [1.35-1.52] | 1.43 [1.35-1.52] | 1.34 [1.25-1.44] | 1.44 [1.18-1.75] | 1.40 [1.32-1.49] | 1.27 [1.20-1.36] | 1.24 [1.16-1.32] | 1.16 [1.08-1.25] | 1.38 [1.12-1.71] | 1.27 [1.20-1.35] | 3,492 | 3,412 | 2,308 | 415 | 3,235 |
| K51 | 1.35 [1.29-1.42] | 1.32 [1.26-1.39] | 1.29 [1.22-1.36] | 1.41 [1.14-1.75] | 1.28 [1.22-1.34] | 1.24 [1.17-1.30] | 1.17 [1.11-1.24] | 1.15 [1.09-1.22] | 1.30 [1.03-1.65] | 1.20 [1.14-1.26] | 4,939 | 4,961 | 3,728 | 332 | 4,485 |
| K52 | 1.25 [1.22-1.27] | 1.25 [1.22-1.28] | 1.24 [1.21-1.27] | 1.15 [1.06-1.24] | 1.21 [1.18-1.23] | 1.18 [1.16-1.21] | 1.16 [1.13-1.18] | 1.15 [1.13-1.18] | 1.13 [1.04-1.23] | 1.16 [1.14-1.19] | 26,610 | 25,640 | 22,004 | 1,885 | 24,352 |
| K55 | 1.16 [1.10-1.22] | 1.15 [1.10-1.21] | 1.16 [1.10-1.22] | | 1.10 [1.05-1.16] | 1.12 [1.06-1.18] | 1.09 [1.03-1.14] | 1.09 [1.03-1.15] | | 1.08 [1.03-1.14] | 4,398 | 4,404 | 4,282 | NA | 4,230 |
| K56 | 1.12 [1.09-1.16] | 1.12 [1.08-1.15] | 1.11 [1.08-1.15] | 1.07 [0.88-1.31] | 1.10 [1.06-1.13] | 1.08 [1.05-1.11] | 1.06 [1.02-1.09] | 1.06 [1.02-1.09] | 1.05 [0.85-1.28] | 1.07 [1.04-1.10] | 12,145 | 12,020 | 11,246 | 358 | 11,630 |
| K57 | 1.16 [1.14-1.17] | 1.15 [1.13-1.16] | 1.16 [1.14-1.17] | | 1.12 [1.10-1.14] | 1.10 [1.08-1.12] | 1.07 [1.06-1.09] | 1.08 [1.07-1.10] | | 1.08 [1.06-1.09] | 54,908 | 54,918 | 54,410 | NA | 51,008 |
| K58 | 1.33 [1.30-1.37] | 1.34 [1.30-1.38] | 1.35 [1.31-1.39] | 1.03 [0.92-1.15] | 1.29 [1.25-1.32] | 1.16 [1.13-1.19] | 1.10 [1.07-1.13] | 1.12 [1.08-1.15] | 0.98 [0.87-1.10] | 1.15 [1.12-1.18] | 16,813 | 17,367 | 12,036 | 1,034 | 16,145 |
| K59 | 1.19 [1.17-1.21] | 1.19 [1.17-1.21] | 1.19 [1.17-1.20] | 1.13 [1.06-1.20] | 1.16 [1.14-1.18] | 1.13 [1.12-1.15] | 1.11 [1.10-1.13] | 1.11 [1.09-1.13] | 1.11 [1.03-1.18] | 1.12 [1.10-1.14] | 52,797 | 51,061 | 46,256 | 3,194 | 50,573 |
| K60 | 1.34 [1.27-1.41] | 1.35 [1.28-1.43] | 1.37 [1.29-1.47] | 1.33 [1.09-1.62] | 1.32 [1.25-1.40] | 1.26 [1.19-1.33] | 1.25 [1.18-1.32] | 1.28 [1.19-1.37] | 1.33 [1.08-1.63] | 1.27 [1.20-1.35] | 4,155 | 4,122 | 2,784 | 388 | 3,780 |
| K61 | 1.29 [1.20-1.40] | 1.30 [1.20-1.40] | 1.28 [1.17-1.40] | 1.18 [0.92-1.52] | 1.24 [1.15-1.34] | 1.25 [1.16-1.35] | 1.20 [1.11-1.30] | 1.19 [1.09-1.31] | 1.23 [0.94-1.60] | 1.22 [1.13-1.32] | 2,219 | 2,206 | 1,434 | 240 | 1,976 |
| K62 | 1.20 [1.18-1.23] | 1.20 [1.18-1.23] | 1.19 [1.16-1.22] | 1.23 [1.08-1.41] | 1.16 [1.14-1.19] | 1.14 [1.12-1.17] | 1.13 [1.10-1.15] | 1.12 [1.09-1.14] | 1.17 [1.02-1.35] | 1.12 [1.10-1.15] | 26,704 | 26,653 | 23,349 | 792 | 24,350 |
| K63 | 1.14 [1.12-1.17] | 1.15 [1.13-1.17] | 1.14 [1.12-1.17] | 1.21 [1.00-1.47] | 1.13 [1.11-1.15] | 1.09 [1.06-1.11] | 1.07 [1.04-1.09] | 1.06 [1.04-1.09] | 1.12 [0.92-1.37] | 1.09 [1.07-1.11] | 27,517 | 27,533 | 25,686 | 407 | 26,081 |
| K64 | 1.19 [1.16-1.22] | 1.18 [1.15-1.21] | 1.19 [1.16-1.22] | 1.05 [0.89-1.24] | 1.16 [1.13-1.19] | 1.12 [1.09-1.15] | 1.09 [1.06-1.12] | 1.10 [1.07-1.13] | 1.02 [0.85-1.22] | 1.10 [1.08-1.13] | 19,851 | 20,018 | 17,641 | 520 | 19,125 |
| K65 | 1.14 [1.07-1.21] | 1.11 [1.04-1.18] | 1.10 [1.03-1.17] | | 1.10 [1.03-1.17] | 1.11 [1.04-1.19] | 1.06 [0.99-1.13] | 1.06 [0.99-1.13] | | 1.09 [1.03-1.17] | 2,819 | 2,795 | 2,538 | NA | 2,658 |
| K66 | 1.11 [1.07-1.14] | 1.10 [1.06-1.14] | 1.10 [1.06-1.14] | 1.00 [0.84-1.18] | 1.06 [1.03-1.10] | 1.05 [1.02-1.09] | 1.02 [0.99-1.06] | 1.03 [0.99-1.07] | 0.97 [0.81-1.16] | 1.04 [1.00-1.07] | 9,791 | 9,940 | 7,610 | 466 | 9,381 |

### Supplementary Table 4: Hazard ratios and events from all cohorts (excluding non-consulters)

| Outcome | Hazard ratio (99% confidence interval) | | | | | | | | | | Events (in exposed) | | | | |
|---|---|---|---|---|---|---|---|---|---|---|---|---|---|---|---|
| | crude | | | | | adjusted | | | | | | | | | |
| | any age | 18+ | 40+ | <18 | hosp. | any age | 18+ | 40+ | <18 | hosp. | any age | 18+ | 40+ | <18 | hosp. |
| K70 | 1.36 [1.28-1.44] | 1.35 [1.27-1.43] | 1.35 [1.27-1.43] | | 1.30 [1.22-1.37] | 1.31 [1.24-1.39] | 1.25 [1.18-1.32] | 1.23 [1.15-1.31] | | 1.29 [1.22-1.37] | 3,519 | 3,533 | 3,298 | NA | 3,270 |
| K71 | 1.27 [1.00-1.60] | 1.29 [1.02-1.62] | 1.42 [1.11-1.83] | | 1.18 [0.94-1.48] | 1.24 [0.98-1.57] | 1.21 [0.95-1.53] | 1.33 [1.03-1.72] | | 1.16 [0.92-1.47] | 219 | 221 | 194 | NA | 217 |
| K72 | 1.25 [1.16-1.34] | 1.27 [1.18-1.36] | 1.28 [1.19-1.38] | | 1.28 [1.19-1.37] | 1.20 [1.12-1.29] | 1.18 [1.10-1.26] | 1.18 [1.10-1.27] | | 1.26 [1.17-1.35] | 2,388 | 2,401 | 2,273 | NA | 2,314 |
| K73 | 1.36 [1.08-1.69] | 1.32 [1.06-1.66] | 1.38 [1.09-1.74] | | 1.43 [1.15-1.79] | 1.27 [1.01-1.59] | 1.24 [0.99-1.57] | 1.26 [0.99-1.61] | | 1.36 [1.09-1.71] | 231 | 224 | 211 | NA | 222 |
| K74 | 1.30 [1.22-1.37] | 1.31 [1.24-1.39] | 1.30 [1.23-1.38] | | 1.29 [1.22-1.36] | 1.23 [1.16-1.30] | 1.20 [1.14-1.28] | 1.18 [1.12-1.26] | | 1.25 [1.19-1.33] | 3,771 | 3,756 | 3,630 | NA | 3,691 |
| K75 | 1.24 [1.16-1.33] | 1.25 [1.17-1.34] | 1.25 [1.17-1.35] | | 1.24 [1.15-1.32] | 1.17 [1.09-1.26] | 1.15 [1.07-1.23] | 1.15 [1.07-1.24] | | 1.20 [1.12-1.29] | 2,409 | 2,394 | 2,238 | NA | 2,346 |
| K76 | 1.20 [1.17-1.23] | 1.19 [1.16-1.23] | 1.20 [1.17-1.23] | 0.79 [0.64-0.99] | 1.17 [1.14-1.20] | 1.13 [1.10-1.16] | 1.09 [1.06-1.12] | 1.09 [1.06-1.12] | 0.81 [0.64-1.03] | 1.12 [1.09-1.15] | 16,807 | 16,847 | 15,484 | 271 | 16,350 |
| K80 | 1.09 [1.06-1.11] | 1.08 [1.06-1.10] | 1.08 [1.05-1.10] | 0.87 [0.76-0.99] | 1.05 [1.03-1.07] | 1.04 [1.02-1.07] | 1.02 [1.00-1.04] | 1.02 [1.00-1.04] | 0.89 [0.77-1.03] | 1.02 [1.00-1.04] | 28,674 | 28,917 | 24,159 | 673 | 26,681 |
| K81 | 1.09 [1.04-1.15] | 1.08 [1.03-1.13] | 1.08 [1.03-1.13] | 0.81 [0.58-1.13] | 1.10 [1.05-1.15] | 1.05 [1.00-1.10] | 1.01 [0.97-1.06] | 1.01 [0.96-1.07] | 0.79 [0.55-1.12] | 1.06 [1.02-1.12] | 5,284 | 5,376 | 4,543 | 111 | 5,055 |
| K82 | 1.07 [1.02-1.13] | 1.07 [1.01-1.13] | 1.07 [1.01-1.14] | 0.92 [0.66-1.29] | 1.07 [1.02-1.13] | 1.03 [0.97-1.09] | 1.00 [0.95-1.06] | 1.01 [0.96-1.08] | 0.89 [0.62-1.26] | 1.04 [0.99-1.10] | 3,865 | 3,904 | 3,123 | 118 | 3,675 |
| K83 | 1.08 [1.04-1.13] | 1.09 [1.05-1.13] | 1.08 [1.04-1.13] | 0.77 [0.55-1.08] | 1.08 [1.04-1.12] | 1.05 [1.01-1.09] | 1.04 [1.00-1.08] | 1.03 [0.99-1.07] | 0.76 [0.54-1.07] | 1.06 [1.02-1.10] | 7,814 | 7,851 | 7,057 | 108 | 7,473 |
| K85 | 1.11 [1.06-1.16] | 1.11 [1.06-1.16] | 1.10 [1.04-1.16] | 0.98 [0.72-1.32] | 1.09 [1.04-1.14] | 1.07 [1.02-1.12] | 1.04 [0.99-1.10] | 1.03 [0.98-1.09] | 1.09 [0.79-1.50] | 1.07 [1.02-1.12] | 4,845 | 4,900 | 4,188 | 136 | 4,546 |
| K86 | 1.12 [1.06-1.18] | 1.14 [1.08-1.20] | 1.12 [1.06-1.18] | | 1.12 [1.06-1.17] | 1.07 [1.02-1.13] | 1.06 [1.00-1.11] | 1.04 [0.99-1.10] | | 1.09 [1.04-1.15] | 4,443 | 4,453 | 4,212 | NA | 4,367 |
| K90 | 1.53 [1.45-1.61] | 1.41 [1.33-1.50] | 1.46 [1.37-1.56] | 2.03 [1.81-2.28] | 1.51 [1.43-1.59] | 1.37 [1.30-1.45] | 1.24 [1.17-1.32] | 1.29 [1.20-1.38] | 1.71 [1.51-1.93] | 1.37 [1.30-1.45] | 4,645 | 3,769 | 2,721 | 1,258 | 4,293 |
| K91 | 1.19 [1.13-1.25] | 1.17 [1.11-1.24] | 1.18 [1.12-1.25] | 0.90 [0.67-1.20] | 1.14 [1.08-1.20] | 1.13 [1.07-1.19] | 1.08 [1.03-1.15] | 1.11 [1.04-1.17] | 0.84 [0.62-1.14] | 1.10 [1.04-1.16] | 3,911 | 3,857 | 3,413 | 157 | 3,759 |
| K92 | 1.18 [1.16-1.20] | 1.18 [1.16-1.21] | 1.17 [1.15-1.20] | 1.02 [0.92-1.13] | 1.14 [1.12-1.16] | 1.13 [1.11-1.15] | 1.11 [1.09-1.13] | 1.10 [1.08-1.12] | 0.99 [0.89-1.11] | 1.11 [1.09-1.13] | 34,874 | 34,921 | 30,925 | 1,261 | 33,035 |
| Diseases of the skin and subcutaneous tissue (L00-L99) | | | | | | | | | | | | | | | |
| L01 | 2.19 [1.83-2.62] | 2.14 [1.68-2.73] | 2.16 [1.65-2.84] | 2.38 [1.84-3.06] | 2.10 [1.74-2.52] | 2.04 [1.69-2.45] | 1.85 [1.43-2.38] | 1.96 [1.48-2.59] | 2.23 [1.71-2.90] | 1.96 [1.62-2.38] | 499 | 251 | 192 | 282 | 456 |
| L02 | 1.25 [1.20-1.31] | 1.27 [1.21-1.32] | 1.26 [1.19-1.32] | 1.17 [1.03-1.34] | 1.20 [1.15-1.25] | 1.19 [1.14-1.25] | 1.16 [1.11-1.22] | 1.16 [1.10-1.22] | 1.14 [0.99-1.30] | 1.17 [1.12-1.23] | 6,363 | 6,163 | 4,282 | 796 | 5,804 |
| L03 | 1.50 [1.47-1.54] | 1.52 [1.49-1.55] | 1.51 [1.48-1.54] | 1.30 [1.16-1.45] | 1.45 [1.42-1.48] | 1.45 [1.42-1.48] | 1.42 [1.39-1.45] | 1.42 [1.39-1.45] | 1.26 [1.13-1.42] | 1.41 [1.38-1.44] | 32,062 | 31,439 | 29,524 | 1,201 | 30,055 |
| L04 | 1.26 [1.06-1.49] | 1.23 [0.96-1.57] | 1.31 [0.99-1.75] | 1.20 [0.95-1.52] | 1.19 [0.99-1.42] | 1.24 [1.04-1.48] | 1.18 [0.92-1.52] | 1.30 [0.97-1.73] | 1.19 [0.94-1.51] | 1.17 [0.98-1.40] | 426 | 199 | 144 | 237 | 383 |
| L05 | 1.04 [0.94-1.15] | 1.06 [0.96-1.17] | 1.27 [1.05-1.52] | 0.93 [0.77-1.11] | 1.05 [0.95-1.18] | 1.02 [0.92-1.14] | 1.01 [0.91-1.11] | 1.18 [0.98-1.43] | 0.96 [0.79-1.16] | 1.05 [0.94-1.18] | 1,261 | 1,297 | 349 | 427 | 987 |
| L08 | 1.39 [1.34-1.45] | 1.39 [1.33-1.44] | 1.41 [1.35-1.46] | 1.61 [1.34-1.93] | 1.38 [1.33-1.44] | 1.35 [1.29-1.40] | 1.30 [1.25-1.35] | 1.32 [1.26-1.37] | 1.63 [1.34-1.97] | 1.36 [1.31-1.41] | 8,468 | 8,107 | 7,598 | 460 | 8,123 |
| L10 | 1.80 [1.29-2.51] | 2.10 [1.50-2.93] | 2.01 [1.44-2.82] | | 1.85 [1.33-2.56] | 1.68 [1.20-2.35] | 1.93 [1.37-2.72] | 1.77 [1.24-2.50] | | 1.72 [1.23-2.40] | 127 | 126 | 123 | NA | 121 |
| L12 | 2.66 [2.36-3.00] | 2.66 [2.36-3.00] | 2.65 [2.36-2.99] | | 2.54 [2.25-2.87] | 2.58 [2.29-2.91] | 2.54 [2.25-2.87] | 2.54 [2.25-2.86] | | 2.47 [2.19-2.80] | 1,160 | 1,159 | 1,151 | NA | 1,060 |
| L13 | 2.61 [1.94-3.51] | 3.28 [2.40-4.49] | 2.95 [2.14-4.06] | | 2.40 [1.78-3.24] | 2.50 [1.85-3.38] | 3.12 [2.27-4.28] | 2.79 [2.02-3.87] | | 2.31 [1.70-3.13] | 183 | 181 | 163 | NA | 167 |
| L20 | 24.14 [20.01-29.12] | 29.51 [22.97-37.90] | 29.60 [22.34-39.24] | 17.64 [13.61-22.86] | 24.80 [20.43-30.10] | 21.95 [18.16-26.53] | 26.47 [20.56-34.06] | 27.25 [20.53-36.19] | 14.94 [11.46-19.48] | 22.36 [18.38-27.20] | 2,663 | 1,677 | 1,233 | 1,165 | 2,423 |
| L21 | 2.74 [2.28-3.30] | 2.57 [2.14-3.10] | 2.72 [2.24-3.30] | | 2.65 [2.20-3.18] | 2.62 [2.17-3.17] | 2.47 [2.04-2.98] | 2.57 [2.11-3.14] | | 2.57 [2.13-3.10] | 516 | 477 | 451 | NA | 485 |
| L22 | 1.01 [0.81-1.27] | 0.91 [0.67-1.24] | 0.91 [0.67-1.24] | | 1.03 [0.82-1.31] | 1.01 [0.80-1.27] | 0.90 [0.66-1.23] | 0.89 [0.65-1.22] | | 1.03 [0.81-1.31] | 225 | 132 | 131 | NA | 217 |
| L23 | 3.18 [2.64-3.82] | 2.92 [2.39-3.56] | 3.21 [2.56-4.02] | 4.71 [2.94-7.55] | 3.08 [2.57-3.71] | 2.94 [2.44-3.55] | 2.69 [2.20-3.30] | 2.96 [2.36-3.73] | 3.71 [2.25-6.12] | 2.90 [2.41-3.50] | 536 | 434 | 338 | 126 | 494 |
| L24 | 1.35 [1.25-1.46] | 1.36 [1.25-1.47] | 1.34 [1.24-1.45] | | 1.32 [1.22-1.43] | 1.32 [1.21-1.43] | 1.29 [1.19-1.40] | 1.26 [1.16-1.37] | | 1.32 [1.22-1.43] | 2,175 | 2,163 | 2,136 | NA | 2,159 |
| L25 | 2.52 [2.09-3.03] | 2.83 [2.34-3.41] | 2.54 [2.07-3.12] | | 2.37 [1.97-2.85] | 2.41 [2.00-2.91] | 2.66 [2.20-3.22] | 2.42 [1.96-2.97] | | 2.31 [1.92-2.79] | 492 | 490 | 394 | NA | 449 |
| L27 | 1.66 [1.53-1.80] | 1.48 [1.36-1.61] | 1.52 [1.39-1.66] | 2.66 [2.04-3.46] | 1.58 [1.45-1.71] | 1.55 [1.43-1.69] | 1.38 [1.26-1.50] | 1.42 [1.30-1.56] | 2.33 [1.76-3.08] | 1.51 [1.39-1.64] | 1,969 | 1,766 | 1,532 | 265 | 1,883 |
| L28 | 3.96 [3.48-4.51] | 3.92 [3.44-4.46] | 3.66 [3.21-4.19] | | 3.41 [3.00-3.87] | 3.72 [3.26-4.25] | 3.64 [3.19-4.15] | 3.37 [2.95-3.86] | | 3.21 [2.82-3.65] | 1,192 | 1,155 | 1,047 | NA | 1,081 |
| L29 | 1.61 [1.51-1.72] | 1.56 [1.46-1.66] | 1.86 [1.72-2.01] | 1.32 [1.07-1.63] | 1.54 [1.44-1.64] | 1.53 [1.44-1.64] | 1.42 [1.33-1.52] | 1.71 [1.58-1.86] | 1.25 [1.00-1.57] | 1.46 [1.36-1.56] | 3,273 | 3,208 | 2,128 | 340 | 3,070 |
| L30 | 8.55 [8.28-8.83] | 8.44 [8.14-8.76] | 7.93 [7.62-8.26] | 9.02 [8.48-9.59] | 8.16 [7.90-8.42] | 8.01 [7.76-8.28] | 7.81 [7.52-8.10] | 7.46 [7.16-7.77] | 8.32 [7.81-8.85] | 7.65 [7.40-7.90] | 36,133 | 26,652 | 19,541 | 11,825 | 32,561 |
| L40 | 1.99 [1.91-2.07] | 1.97 [1.89-2.05] | 1.95 [1.87-2.03] | 1.59 [1.28-1.97] | 1.95 [1.87-2.03] | 1.93 [1.85-2.01] | 1.86 [1.78-1.93] | 1.85 [1.77-1.93] | 1.63 [1.30-2.05] | 1.90 [1.83-1.98] | 9,255 | 9,284 | 7,897 | 327 | 8,542 |
| L42 | 1.44 [1.05-1.98] | 1.58 [1.15-2.19] | | | 1.23 [0.88-1.70] | 1.39 [1.00-1.93] | 1.37 [0.97-1.93] | | | 1.15 [0.83-1.61] | 127 | 123 | NA | NA | 111 |
| L43 | 1.98 [1.77-2.22] | 1.92 [1.71-2.15] | 1.91 [1.70-2.15] | | 1.82 [1.62-2.05] | 1.89 [1.68-2.12] | 1.80 [1.61-2.03] | 1.82 [1.62-2.05] | | 1.76 [1.56-1.98] | 1,043 | 1,047 | 982 | NA | 932 |
| L50 | 1.79 [1.64-1.95] | 1.61 [1.46-1.78] | 1.71 [1.52-1.92] | 2.20 [1.89-2.57] | 1.77 [1.62-1.93] | 1.54 [1.41-1.68] | 1.37 [1.23-1.52] | 1.44 [1.27-1.63] | 1.86 [1.59-2.19] | 1.51 [1.38-1.65] | 1,944 | 1,290 | 933 | 772 | 1,816 |
| L51 | 1.31 [1.08-1.58] | 1.77 [1.41-2.21] | 1.75 [1.37-2.24] | | 1.42 [1.17-1.73] | 1.23 [1.02-1.50] | 1.63 [1.29-2.05] | 1.61 [1.26-2.07] | | 1.34 [1.10-1.63] | 344 | 266 | 220 | NA | 328 |
| L52 | 1.37 [1.03-1.82] | 1.24 [0.94-1.65] | | | 1.28 [0.95-1.74] | 1.28 [0.96-1.71] | 1.18 [0.88-1.57] | | | 1.18 [0.86-1.62] | 162 | 158 | NA | NA | 145 |
| L53 | 1.59 [1.49-1.70] | 1.60 [1.49-1.71] | 1.67 [1.56-1.80] | 1.19 [0.92-1.54] | 1.54 [1.44-1.64] | 1.54 [1.44-1.65] | 1.50 [1.40-1.61] | 1.58 [1.47-1.70] | 1.11 [0.85-1.46] | 1.50 [1.40-1.60] | 2,947 | 2,799 | 2,546 | 210 | 2,790 |
| L56 | 2.54 [1.83-3.51] | 2.41 [1.75-3.34] | 2.49 [1.74-3.55] | | 2.27 [1.64-3.15] | 2.48 [1.79-3.45] | 2.28 [1.64-3.17] | 2.38 [1.66-3.43] | | 2.20 [1.57-3.07] | 145 | 144 | 127 | NA | 131 |
| L57 | 1.24 [1.18-1.29] | 1.21 [1.16-1.27] | 1.24 [1.19-1.30] | | 1.21 [1.16-1.26] | 1.19 [1.14-1.25] | 1.16 [1.11-1.22] | 1.19 [1.13-1.24] | | 1.16 [1.11-1.22] | 5,879 | 5,881 | 5,843 | NA | 5,558 |
| L60 | 1.38 [1.26-1.50] | 1.38 [1.26-1.51] | 1.41 [1.28-1.57] | 1.13 [0.91-1.41] | 1.25 [1.15-1.37] | 1.28 [1.17-1.40] | 1.27 [1.16-1.40] | 1.29 [1.16-1.44] | 1.07 [0.85-1.35] | 1.17 [1.06-1.28] | 1,640 | 1,494 | 1,130 | 301 | 1,459 |
| L63 | 2.57 [1.86-3.56] | 2.26 [1.66-3.08] | 2.75 [1.89-4.01] | | 2.62 [1.90-3.61] | 2.07 [1.46-2.93] | 1.73 [1.23-2.41] | 2.29 [1.54-3.41] | | 2.15 [1.52-3.02] | 166 | 162 | 118 | NA | 155 |
| L65 | 1.72 [1.44-2.05] | 1.77 [1.48-2.12] | 1.94 [1.59-2.38] | | 1.81 [1.51-2.16] | 1.55 [1.29-1.86] | 1.54 [1.27-1.86] | 1.73 [1.41-2.14] | | 1.60 [1.33-1.93] | 449 | 423 | 329 | NA | 418 |
| L68 | 1.14 [0.82-1.59] | 1.18 [0.85-1.65] | | | 1.23 [0.88-1.72] | 1.10 [0.78-1.56] | 1.03 [0.73-1.46] | | | 1.19 [0.85-1.68] | 104 | 108 | NA | NA | 105 |
| L70 | 1.25 [1.03-1.52] | 1.18 [0.97-1.42] | 1.22 [0.88-1.69] | 1.22 [0.85-1.75] | 1.18 [0.97-1.44] | 1.16 [0.95-1.42] | 1.06 [0.87-1.29] | 1.06 [0.76-1.49] | 1.13 [0.77-1.65] | 1.13 [0.92-1.38] | 350 | 348 | 111 | 117 | 324 |
| L71 | 1.39 [1.21-1.61] | 1.46 [1.27-1.69] | 1.38 [1.19-1.59] | | 1.42 [1.23-1.64] | 1.31 [1.13-1.52] | 1.41 [1.22-1.62] | 1.30 [1.12-1.51] | | 1.35 [1.16-1.56] | 613 | 603 | 566 | NA | 581 |
| L72 | 1.07 [1.03-1.12] | 1.07 [1.03-1.12] | 1.10 [1.04-1.15] | 0.97 [0.80-1.18] | 1.05 [1.01-1.10] | 1.03 [0.99-1.08] | 1.01 [0.97-1.06] | 1.04 [0.99-1.09] | 0.93 [0.76-1.13] | 1.03 [0.98-1.07] | 6,167 | 6,011 | 4,738 | 372 | 5,322 |
| L73 | 1.39 [1.24-1.55] | 1.41 [1.26-1.58] | 1.37 [1.19-1.58] | 1.08 [0.75-1.56] | 1.28 [1.14-1.43] | 1.29 [1.15-1.45] | 1.26 [1.13-1.42] | 1.23 [1.06-1.42] | 1.02 [0.70-1.49] | 1.23 [1.10-1.38] | 1,006 | 1,026 | 580 | 104 | 928 |
| L80 | 1.62 [1.31-2.01] | 1.54 [1.25-1.91] | 1.57 [1.25-1.97] | | 1.67 [1.35-2.08] | 1.47 [1.18-1.84] | 1.46 [1.17-1.82] | 1.42 [1.12-1.80] | | 1.61 [1.29-2.01] | 291 | 287 | 247 | NA | 279 |
| L81 | 1.21 [1.09-1.33] | 1.18 [1.07-1.31] | 1.21 [1.09-1.35] | 1.66 [1.13-2.45] | 1.22 [1.10-1.35] | 1.16 [1.05-1.28] | 1.13 [1.02-1.25] | 1.16 [1.04-1.30] | 1.67 [1.12-2.48] | 1.17 [1.06-1.30] | 1,189 | 1,109 | 972 | 106 | 1,113 |
| L82 | 1.15 [1.09-1.22] | 1.19 [1.12-1.25] | 1.19 [1.12-1.26] | | 1.16 [1.10-1.23] | 1.10 [1.04-1.17] | 1.13 [1.06-1.19] | 1.13 [1.07-1.20] | | 1.11 [1.05-1.18] | 3,562 | 3,569 | 3,440 | NA | 3,267 |
| L83 | 1.66 [1.32-2.09] | 1.67 [1.31-2.12] | 1.61 [1.25-2.06] | | 1.60 [1.27-2.01] | 1.60 [1.26-2.02] | 1.53 [1.20-1.96] | 1.52 [1.18-1.95] | | 1.53 [1.21-1.94] | 252 | 232 | 210 | NA | 230 |
| L84 | 1.43 [1.22-1.69] | 1.40 [1.19-1.65] | 1.35 [1.13-1.59] | | 1.30 [1.10-1.53] | 1.36 [1.15-1.61] | 1.29 [1.09-1.53] | 1.24 [1.04-1.47] | | 1.26 [1.06-1.49] | 453 | 447 | 411 | NA | 416 |
| L85 | 1.51 [1.39-1.65] | 1.49 [1.37-1.62] | 1.46 [1.34-1.59] | | 1.52 [1.40-1.65] | 1.46 [1.35-1.59] | 1.41 [1.30-1.54] | 1.39 [1.27-1.51] | | 1.48 [1.36-1.61] | 1,816 | 1,793 | 1,721 | NA | 1,731 |
| L88 | 1.61 [1.15-2.26] | 1.64 [1.17-2.28] | | | 1.28 [0.92-1.77] | 1.49 [1.05-2.10] | 1.46 [1.03-2.06] | | | 1.19 [0.85-1.66] | 108 | 109 | NA | NA | 112 |
| L89 | 1.17 [1.14-1.19] | 1.15 [1.13-1.18] | 1.16 [1.13-1.18] | | 1.16 [1.14-1.19] | 1.15 [1.12-1.18] | 1.11 [1.09-1.14] | 1.11 [1.09-1.14] | | 1.16 [1.14-1.19] | 21,530 | 21,508 | 21,367 | NA | 21,128 |
| L90 | 1.24 [1.17-1.31] | 1.27 [1.20-1.34] | 1.25 [1.17-1.33] | 0.93 [0.71-1.22] | 1.22 [1.16-1.30] | 1.19 [1.12-1.26] | 1.20 [1.13-1.27] | 1.18 [1.10-1.26] | 0.91 [0.69-1.21] | 1.18 [1.12-1.26] | 3,508 | 3,505 | 2,806 | 182 | 3,338 |
| L91 | 1.29 [1.16-1.43] | 1.22 [1.09-1.35] | 1.25 [1.10-1.42] | 1.46 [1.11-1.93] | 1.19 [1.07-1.33] | 1.23 [1.10-1.37] | 1.16 [1.04-1.29] | 1.18 [1.04-1.35] | 1.45 [1.09-1.92] | 1.16 [1.04-1.29] | 1,110 | 1,020 | 677 | 199 | 974 |
| L92 | 1.24 [1.10-1.39] | 1.21 [1.07-1.36] | 1.19 [1.04-1.36] | 1.04 [0.76-1.43] | 1.15 [1.03-1.29] | 1.17 [1.04-1.31] | 1.11 [0.99-1.26] | 1.11 [0.97-1.27] | 1.03 [0.74-1.43] | 1.11 [0.99-1.24] | 917 | 850 | 657 | 134 | 857 |
| L93 | 1.71 [1.47-1.98] | 1.65 [1.41-1.92] | 1.64 [1.39-1.93] | | 1.69 [1.46-1.97] | 1.58 [1.35-1.84] | 1.47 [1.26-1.72] | 1.47 [1.24-1.74] | | 1.57 [1.35-1.83] | 565 | 569 | 467 | NA | 542 |
| L94 | 1.33 [1.10-1.62] | 1.32 [1.08-1.62] | 1.54 [1.24-1.91] | | 1.31 [1.08-1.59] | 1.27 [1.05-1.55] | 1.25 [1.01-1.53] | 1.39 [1.11-1.74] | | 1.28 [1.05-1.55] | 317 | 294 | 266 | NA | 309 |
| L95 | 1.61 [1.36-1.90] | 1.78 [1.51-2.11] | 1.64 [1.37-1.96] | | 1.55 [1.31-1.83] | 1.50 [1.27-1.78] | 1.67 [1.41-1.99] | 1.50 [1.25-1.81] | | 1.45 [1.22-1.72] | 472 | 463 | 409 | NA | 439 |
| L97 | 1.44 [1.41-1.48] | 1.42 [1.38-1.46] | 1.44 [1.40-1.48] | | 1.40 [1.37-1.44] | 1.41 [1.37-1.45] | 1.35 [1.31-1.39] | 1.36 [1.32-1.40] | | 1.39 [1.35-1.43] | 17,841 | 17,848 | 17,693 | NA | 17,055 |
| L98 | 1.31 [1.27-1.35] | 1.31 [1.27-1.36] | 1.31 [1.27-1.35] | 1.27 [1.06-1.52] | 1.27 [1.23-1.31] | 1.26 [1.22-1.30] | 1.25 [1.21-1.29] | 1.24 [1.20-1.28] | 1.28 [1.06-1.54] | 1.23 [1.19-1.27] | 11,909 | 11,662 | 10,675 | 437 | 11,106 |
| Diseases of the musculoskeletal system and connective tissue (M00-M99) | | | | | | | | | | | | | | | |
| M00 | 1.34 [1.22-1.47] | 1.31 [1.19-1.43] | 1.37 [1.24-1.51] | 1.45 [1.00-2.09] | 1.30 [1.18-1.42] | 1.29 [1.17-1.42] | 1.23 [1.12-1.35] | 1.29 [1.17-1.42] | 1.44 [0.98-2.10] | 1.27 [1.16-1.39] | 1,458 | 1,378 | 1,323 | 105 | 1,421 |
| M02 | 1.16 [0.93-1.45] | 1.30 [1.00-1.68] | 1.14 [0.86-1.53] | | 1.15 [0.91-1.46] | 1.12 [0.89-1.41] | 1.21 [0.93-1.58] | 1.10 [0.82-1.48] | | 1.08 [0.85-1.38] | 246 | 178 | 133 | NA | 216 |
| M05 | 1.21 [1.12-1.32] | 1.20 [1.11-1.31] | 1.19 [1.10-1.30] | | 1.16 [1.07-1.26] | 1.19 [1.10-1.29] | 1.16 [1.07-1.26] | 1.15 [1.06-1.25] | | 1.14 [1.05-1.24] | 1,718 | 1,725 | 1,632 | NA | 1,666 |
| M06 | 1.24 [1.20-1.28] | 1.25 [1.21-1.29] | 1.24 [1.21-1.28] | | 1.22 [1.19-1.26] | 1.18 [1.14-1.21] | 1.16 [1.13-1.20] | 1.16 [1.12-1.19] | | 1.17 [1.13-1.20] | 13,260 | 13,321 | 12,536 | NA | 12,362 |
| M07 | 1.48 [1.35-1.62] | 1.45 [1.33-1.58] | 1.40 [1.27-1.54] | | 1.44 [1.32-1.57] | 1.40 [1.28-1.53] | 1.34 [1.22-1.46] | 1.29 [1.17-1.43] | | 1.37 [1.26-1.50] | 1,635 | 1,643 | 1,351 | NA | 1,561 |
| M08 | 1.10 [0.86-1.40] | 1.06 [0.75-1.49] | | 1.18 [0.85-1.65] | 1.08 [0.84-1.39] | 1.04 [0.81-1.34] | 1.04 [0.73-1.47] | | 1.17 [0.83-1.65] | 1.09 [0.84-1.41] | 212 | 105 | NA | 130 | 196 |
| M10 | 1.20 [1.17-1.24] | 1.21 [1.18-1.24] | 1.20 [1.17-1.23] | | 1.19 [1.16-1.22] | 1.16 [1.13-1.19] | 1.14 [1.11-1.17] | 1.12 [1.09-1.16] | | 1.16 [1.13-1.19] | 15,577 | 15,584 | 15,495 | NA | 14,894 |
| M11 | 1.20 [1.10-1.31] | 1.17 [1.08-1.28] | 1.18 [1.08-1.29] | | 1.23 [1.13-1.34] | 1.14 [1.05-1.25] | 1.10 [1.00-1.20] | 1.10 [1.01-1.21] | | 1.18 [1.09-1.29] | 1,717 | 1,713 | 1,693 | NA | 1,692 |
| M12 | 1.36 [1.12-1.66] | 1.27 [1.04-1.54] | 1.31 [1.07-1.62] | | 1.11 [0.92-1.35] | 1.25 [1.02-1.53] | 1.15 [0.94-1.41] | 1.14 [0.92-1.42] | | 1.09 [0.89-1.32] | 307 | 308 | 260 | NA | 291 |
| M13 | 1.21 [1.19-1.23] | 1.20 [1.18-1.22] | 1.21 [1.18-1.23] | 1.12 [0.85-1.49] | 1.19 [1.17-1.21] | 1.14 [1.12-1.16] | 1.11 [1.09-1.13] | 1.11 [1.09-1.13] | 1.07 [0.80-1.43] | 1.13 [1.11-1.15] | 37,875 | 37,923 | 37,011 | 188 | 36,172 |
| M14 | 1.30 [1.10-1.52] | 1.37 [1.17-1.61] | 1.42 [1.20-1.68] | | 1.21 [1.03-1.41] | 1.21 [1.03-1.43] | 1.20 [1.01-1.42] | 1.23 [1.04-1.47] | | 1.16 [0.99-1.36] | 460 | 460 | 444 | NA | 450 |
| M15 | 1.25 [1.21-1.29] | 1.26 [1.22-1.30] | 1.23 [1.20-1.27] | | 1.20 [1.17-1.24] | 1.17 [1.13-1.20] | 1.16 [1.12-1.19] | 1.13 [1.10-1.17] | | 1.14 [1.11-1.17] | 14,249 | 14,280 | 14,162 | NA | 13,843 |
| M16 | 1.06 [1.04-1.09] | 1.06 [1.03-1.08] | 1.06 [1.04-1.09] | | 1.04 [1.01-1.06] | 1.04 [1.02-1.07] | 1.03 [1.00-1.05] | 1.03 [1.00-1.05] | | 1.02 [0.99-1.04] | 19,401 | 19,372 | 19,104 | NA | 17,634 |
| M17 | 1.15 [1.13-1.17] | 1.15 [1.13-1.17] | 1.14 [1.12-1.16] | | 1.12 [1.10-1.14] | 1.10 [1.08-1.12] | 1.09 [1.07-1.11] | 1.08 [1.06-1.10] | | 1.08 [1.06-1.10] | 33,741 | 33,770 | 33,007 | NA | 30,919 |
| M18 | 1.29 [1.19-1.39] | 1.25 [1.16-1.35] | 1.27 [1.18-1.37] | | 1.20 [1.11-1.29] | 1.17 [1.08-1.26] | 1.11 [1.02-1.20] | 1.14 [1.05-1.23] | | 1.11 [1.03-1.20] | 2,057 | 2,056 | 2,034 | NA | 1,969 |
| M19 | 1.23 [1.21-1.25] | 1.23 [1.21-1.25] | 1.22 [1.21-1.24] | | 1.19 [1.17-1.21] | 1.16 [1.14-1.17] | 1.13 [1.12-1.15] | 1.12 [1.11-1.14] | | 1.13 [1.12-1.15] | 59,981 | 60,071 | 59,234 | NA | 57,853 |
| M20 | 1.21 [1.16-1.26] | 1.19 [1.15-1.24] | 1.18 [1.13-1.23] | 1.24 [0.98-1.58] | 1.15 [1.11-1.20] | 1.15 [1.11-1.20] | 1.13 [1.08-1.18] | 1.12 [1.08-1.17] | 1.22 [0.95-1.57] | 1.11 [1.06-1.15] | 7,141 | 7,127 | 6,234 | 260 | 6,336 |
| M21 | 1.12 [1.06-1.18] | 1.13 [1.07-1.20] | 1.12 [1.06-1.19] | 0.97 [0.78-1.20] | 1.10 [1.05-1.16] | 1.06 [1.01-1.12] | 1.06 [1.01-1.13] | 1.06 [1.00-1.12] | 0.97 [0.77-1.21] | 1.07 [1.01-1.13] | 4,036 | 3,820 | 3,497 | 304 | 3,919 |
| M22 | 1.22 [1.10-1.35] | 1.19 [1.07-1.31] | 1.15 [1.02-1.31] | 1.22 [0.89-1.68] | 1.23 [1.11-1.36] | 1.13 [1.02-1.25] | 1.08 [0.97-1.20] | 1.06 [0.93-1.21] | 1.15 [0.82-1.61] | 1.15 [1.03-1.28] | 1,172 | 1,146 | 671 | 151 | 1,057 |
| M23 | 1.11 [1.07-1.15] | 1.12 [1.08-1.15] | 1.11 [1.07-1.15] | 1.06 [0.90-1.24] | 1.08 [1.04-1.12] | 1.05 [1.02-1.09] | 1.05 [1.02-1.09] | 1.04 [1.00-1.08] | 1.02 [0.86-1.21] | 1.03 [1.00-1.07] | 10,814 | 10,779 | 8,874 | 553 | 9,402 |
| M24 | 1.18 [1.12-1.24] | 1.17 [1.11-1.23] | 1.15 [1.08-1.22] | 1.10 [0.94-1.29] | 1.14 [1.08-1.19] | 1.11 [1.06-1.17] | 1.08 [1.03-1.14] | 1.07 [1.01-1.14] | 1.07 [0.90-1.26] | 1.09 [1.04-1.15] | 4,616 | 4,470 | 3,246 | 549 | 4,331 |
| M25 | 1.19 [1.16-1.21] | 1.18 [1.16-1.21] | 1.17 [1.15-1.20] | 1.10 [1.00-1.20] | 1.16 [1.13-1.18] | 1.12 [1.10-1.14] | 1.10 [1.07-1.12] | 1.09 [1.07-1.11] | 1.05 [0.96-1.16] | 1.11 [1.09-1.13] | 34,297 | 33,572 | 30,035 | 1,682 | 32,837 |
| M30 | 1.17 [0.92-1.48] | 1.30 [0.98-1.72] | 1.25 [0.94-1.66] | | 1.14 [0.90-1.45] | 1.08 [0.85-1.38] | 1.10 [0.82-1.49] | 1.09 [0.81-1.47] | | 1.02 [0.80-1.31] | 222 | 147 | 137 | NA | 200 |
| M31 | 1.26 [1.17-1.36] | 1.25 [1.16-1.35] | 1.24 [1.15-1.33] | | 1.19 [1.11-1.29] | 1.20 [1.11-1.29] | 1.18 [1.09-1.27] | 1.15 [1.07-1.24] | | 1.14 [1.05-1.23] | 2,102 | 2,113 | 2,038 | NA | 1,942 |
| M32 | 1.52 [1.36-1.69] | 1.54 [1.38-1.71] | 1.50 [1.33-1.69] | | 1.48 [1.33-1.64] | 1.43 [1.28-1.59] | 1.40 [1.25-1.56] | 1.38 [1.22-1.56] | | 1.39 [1.25-1.55] | 1,094 | 1,102 | 841 | NA | 1,030 |
| M33 | 1.93 [1.53-2.43] | 2.01 [1.59-2.53] | 1.94 [1.52-2.47] | | 1.74 [1.37-2.22] | 1.83 [1.44-2.32] | 1.92 [1.51-2.43] | 1.89 [1.48-2.42] | | 1.68 [1.31-2.15] | 248 | 246 | 217 | NA | 207 |
| M34 | 1.45 [1.23-1.72] | 1.44 [1.22-1.70] | 1.44 [1.21-1.71] | | 1.43 [1.21-1.69] | 1.38 [1.16-1.64] | 1.34 [1.13-1.60] | 1.33 [1.12-1.60] | | 1.36 [1.14-1.61] | 423 | 418 | 389 | NA | 398 |
| M35 | 1.28 [1.24-1.33] | 1.29 [1.25-1.33] | 1.29 [1.24-1.33] | 1.19 [0.99-1.43] | 1.26 [1.22-1.31] | 1.21 [1.17-1.25] | 1.20 [1.16-1.24] | 1.20 [1.16-1.25] | 1.12 [0.92-1.36] | 1.20 [1.15-1.24] | 10,397 | 10,287 | 9,374 | 430 | 9,715 |
| M40 | 1.20 [1.11-1.29] | 1.15 [1.07-1.23] | 1.18 [1.09-1.27] | | 1.15 [1.07-1.24] | 1.14 [1.06-1.23] | 1.08 [1.00-1.16] | 1.11 [1.03-1.20] | | 1.11 [1.03-1.20] | 2,299 | 2,286 | 2,196 | NA | 2,274 |
| M41 | 1.19 [1.14-1.25] | 1.20 [1.15-1.26] | 1.18 [1.12-1.24] | 0.96 [0.80-1.16] | 1.13 [1.08-1.18] | 1.13 [1.08-1.18] | 1.13 [1.08-1.18] | 1.10 [1.05-1.16] | 0.95 [0.78-1.15] | 1.08 [1.03-1.13] | 5,972 | 5,807 | 5,191 | 404 | 5,718 |
| M43 | 1.21 [1.15-1.27] | 1.21 [1.15-1.26] | 1.20 [1.14-1.26] | 0.95 [0.70-1.29] | 1.15 [1.10-1.21] | 1.13 [1.08-1.19] | 1.10 [1.04-1.15] | 1.10 [1.04-1.15] | 0.88 [0.64-1.20] | 1.09 [1.04-1.14] | 5,217 | 5,181 | 4,835 | 145 | 5,034 |
| M45 | 1.30 [1.18-1.43] | 1.38 [1.25-1.52] | 1.32 [1.19-1.46] | | 1.35 [1.22-1.49] | 1.22 [1.10-1.35] | 1.26 [1.14-1.40] | 1.21 [1.09-1.34] | | 1.26 [1.14-1.40] | 1,245 | 1,261 | 1,131 | NA | 1,197 |
| M46 | 1.29 [1.23-1.36] | 1.29 [1.23-1.36] | 1.28 [1.22-1.35] | | 1.24 [1.18-1.30] | 1.19 [1.13-1.25] | 1.16 [1.10-1.22] | 1.15 [1.09-1.21] | | 1.17 [1.11-1.23] | 4,513 | 4,531 | 4,319 | NA | 4,386 |
| M47 | 1.22 [1.20-1.25] | 1.24 [1.21-1.27] | 1.22 [1.19-1.25] | | 1.19 [1.17-1.22] | 1.14 [1.11-1.17] | 1.12 [1.10-1.15] | 1.11 [1.08-1.13] | | 1.13 [1.10-1.15] | 25,023 | 25,080 | 24,283 | NA | 24,218 |
| M48 | 1.18 [1.15-1.22] | 1.18 [1.15-1.22] | 1.19 [1.15-1.22] | | 1.16 [1.12-1.19] | 1.11 [1.08-1.15] | 1.09 [1.05-1.12] | 1.08 [1.05-1.12] | | 1.10 [1.07-1.14] | 14,272 | 14,276 | 14,003 | NA | 13,875 |
| M49 | 1.01 [0.86-1.18] | 1.03 [0.88-1.22] | 1.11 [0.94-1.31] | | 1.02 [0.87-1.20] | 0.99 [0.84-1.16] | 1.01 [0.86-1.19] | 1.09 [0.92-1.29] | | 1.03 [0.88-1.22] | 420 | 417 | 403 | NA | 397 |
| M50 | 1.22 [1.15-1.29] | 1.19 [1.13-1.26] | 1.20 [1.13-1.27] | | 1.13 [1.07-1.20] | 1.15 [1.08-1.21] | 1.08 [1.02-1.14] | 1.08 [1.02-1.15] | | 1.08 [1.02-1.14] | 3,530 | 3,530 | 3,277 | NA | 3,384 |
| M51 | 1.19 [1.16-1.22] | 1.18 [1.15-1.21] | 1.19 [1.16-1.22] | 0.98 [0.79-1.22] | 1.15 [1.12-1.18] | 1.11 [1.08-1.14] | 1.06 [1.04-1.09] | 1.07 [1.04-1.11] | 0.88 [0.70-1.11] | 1.09 [1.06-1.12] | 16,572 | 16,674 | 14,545 | 277 | 15,952 |
| M53 | 1.12 [1.01-1.25] | 1.11 [1.00-1.23] | 1.11 [0.99-1.25] | | 1.11 [1.00-1.23] | 1.03 [0.93-1.15] | 0.98 [0.88-1.10] | 0.99 [0.88-1.12] | | 1.03 [0.92-1.14] | 1,024 | 1,019 | 808 | NA | 1,002 |
| M54 | 1.19 [1.17-1.21] | 1.18 [1.16-1.20] | 1.19 [1.17-1.21] | 1.00 [0.91-1.09] | 1.14 [1.12-1.16] | 1.11 [1.09-1.13] | 1.07 [1.05-1.09] | 1.08 [1.06-1.10] | 0.98 [0.89-1.07] | 1.08 [1.07-1.10] | 42,416 | 42,685 | 35,152 | 1,735 | 40,355 |
| M60 | 1.33 [1.18-1.51] | 1.32 [1.16-1.50] | 1.36 [1.18-1.55] | | 1.31 [1.15-1.48] | 1.29 [1.14-1.47] | 1.24 [1.09-1.42] | 1.29 [1.12-1.48] | | 1.29 [1.13-1.47] | 785 | 716 | 637 | NA | 744 |
| M62 | 1.18 [1.11-1.25] | 1.18 [1.11-1.25] | 1.18 [1.11-1.26] | 0.88 [0.67-1.15] | 1.12 [1.06-1.19] | 1.14 [1.07-1.21] | 1.11 [1.05-1.18] | 1.13 [1.06-1.20] | 0.81 [0.61-1.08] | 1.11 [1.05-1.18] | 3,455 | 3,385 | 3,050 | 174 | 3,341 |
| M65 | 1.25 [1.20-1.30] | 1.24 [1.19-1.29] | 1.23 [1.17-1.28] | 1.24 [1.03-1.48] | 1.21 [1.17-1.27] | 1.16 [1.11-1.21] | 1.14 [1.09-1.19] | 1.13 [1.08-1.18] | 1.18 [0.98-1.42] | 1.15 [1.10-1.20] | 6,878 | 6,642 | 5,811 | 408 | 6,406 |
| M66 | 1.20 [1.05-1.37] | 1.19 [1.05-1.36] | 1.12 [0.99-1.28] | | 1.10 [0.97-1.26] | 1.14 [1.00-1.30] | 1.13 [0.99-1.29] | 1.06 [0.92-1.21] | | 1.06 [0.92-1.21] | 692 | 685 | 647 | NA | 620 |
| M67 | 1.18 [1.12-1.25] | 1.19 [1.12-1.25] | 1.25 [1.18-1.33] | 1.02 [0.87-1.20] | 1.14 [1.08-1.20] | 1.12 [1.06-1.18] | 1.11 [1.05-1.17] | 1.17 [1.09-1.24] | 0.98 [0.83-1.16] | 1.09 [1.03-1.15] | 4,100 | 3,879 | 2,927 | 502 | 3,701 |
| M70 | 1.28 [1.20-1.36] | 1.27 [1.20-1.36] | 1.27 [1.19-1.35] | | 1.22 [1.15-1.30] | 1.19 [1.11-1.27] | 1.15 [1.08-1.23] | 1.15 [1.08-1.23] | | 1.14 [1.07-1.22] | 2,910 | 2,902 | 2,712 | NA | 2,800 |
| M71 | 1.23 [1.14-1.34] | 1.25 [1.15-1.35] | 1.22 [1.12-1.32] | | 1.25 [1.15-1.35] | 1.16 [1.07-1.26] | 1.16 [1.07-1.26] | 1.12 [1.03-1.22] | | 1.18 [1.09-1.28] | 1,909 | 1,906 | 1,787 | NA | 1,819 |
| M72 | 1.02 [0.96-1.08] | 1.05 [0.99-1.11] | 1.04 [0.98-1.10] | | 1.04 [0.99-1.11] | 1.00 [0.94-1.06] | 1.02 [0.96-1.08] | 1.02 [0.96-1.08] | | 1.03 [0.97-1.09] | 3,398 | 3,408 | 3,255 | NA | 3,060 |
| M75 | 1.18 [1.14-1.21] | 1.19 [1.15-1.22] | 1.17 [1.13-1.21] | | 1.13 [1.10-1.17] | 1.10 [1.06-1.13] | 1.09 [1.05-1.12] | 1.07 [1.03-1.10] | | 1.07 [1.04-1.11] | 11,012 | 11,050 | 10,280 | NA | 10,333 |
| M76 | 1.26 [1.13-1.41] | 1.29 [1.15-1.44] | 1.32 [1.17-1.49] | | 1.28 [1.15-1.43] | 1.16 [1.04-1.31] | 1.16 [1.03-1.30] | 1.18 [1.04-1.34] | | 1.19 [1.06-1.33] | 945 | 938 | 795 | NA | 918 |
| M77 | 1.34 [1.25-1.45] | 1.31 [1.22-1.41] | 1.30 [1.20-1.40] | | 1.26 [1.17-1.35] | 1.23 [1.14-1.33] | 1.17 [1.09-1.27] | 1.17 [1.08-1.26] | | 1.18 [1.09-1.27] | 2,210 | 2,213 | 1,934 | NA | 2,089 |
| M79 | 1.27 [1.25-1.29] | 1.27 [1.25-1.30] | 1.28 [1.26-1.31] | 1.04 [0.95-1.14] | 1.23 [1.20-1.25] | 1.18 [1.16-1.20] | 1.15 [1.13-1.17] | 1.16 [1.14-1.19] | 1.01 [0.91-1.11] | 1.16 [1.14-1.18] | 37,345 | 36,967 | 31,656 | 1,573 | 35,587 |
| M80 | 1.19 [1.13-1.25] | 1.21 [1.14-1.27] | 1.20 [1.14-1.27] | | 1.18 [1.13-1.25] | 1.14 [1.08-1.20] | 1.13 [1.07-1.19] | 1.12 [1.07-1.18] | | 1.14 [1.08-1.20] | 4,601 | 4,591 | 4,563 | NA | 4,381 |
| M81 | 1.21 [1.18-1.23] | 1.20 [1.17-1.22] | 1.20 [1.18-1.23] | | 1.19 [1.16-1.21] | 1.15 [1.12-1.17] | 1.11 [1.09-1.13] | 1.12 [1.09-1.14] | | 1.13 [1.11-1.15] | 28,513 | 28,498 | 28,165 | NA | 27,378 |
| M83 | 1.30 [1.01-1.69] | 1.10 [0.85-1.42] | 1.35 [1.03-1.78] | | 1.24 [0.96-1.60] | 1.20 [0.92-1.57] | 1.05 [0.81-1.37] | 1.25 [0.95-1.66] | | 1.18 [0.91-1.53] | 173 | 174 | 162 | NA | 171 |
| M84 | 1.15 [1.08-1.22] | 1.16 [1.09-1.24] | 1.16 [1.09-1.24] | 0.88 [0.65-1.20] | 1.14 [1.07-1.21] | 1.10 [1.03-1.17] | 1.08 [1.02-1.15] | 1.09 [1.02-1.16] | 0.87 [0.63-1.20] | 1.12 [1.05-1.19] | 3,150 | 3,100 | 2,746 | 144 | 2,964 |
| M85 | 1.20 [1.15-1.26] | 1.20 [1.14-1.25] | 1.17 [1.12-1.23] | 1.01 [0.70-1.44] | 1.18 [1.13-1.24] | 1.12 [1.07-1.17] | 1.09 [1.04-1.14] | 1.08 [1.03-1.13] | 0.94 [0.65-1.37] | 1.12 [1.07-1.17] | 6,142 | 6,124 | 5,801 | 115 | 6,060 |
| M86 | 1.22 [1.14-1.30] | 1.22 [1.14-1.30] | 1.23 [1.15-1.32] | 1.41 [1.02-1.96] | 1.22 [1.14-1.30] | 1.18 [1.11-1.27] | 1.13 [1.06-1.21] | 1.15 [1.07-1.24] | 1.36 [0.97-1.91] | 1.22 [1.14-1.30] | 2,723 | 2,618 | 2,525 | 139 | 2,667 |
| M87 | 1.14 [1.03-1.27] | 1.11 [1.00-1.24] | 1.16 [1.04-1.30] | | 1.11 [1.00-1.23] | 1.09 [0.98-1.22] | 1.05 [0.94-1.17] | 1.10 [0.98-1.23] | | 1.09 [0.98-1.21] | 1,018 | 1,002 | 926 | NA | 984 |
| M88 | 1.11 [0.97-1.27] | 1.05 [0.92-1.20] | 1.11 [0.97-1.27] | | 1.02 [0.89-1.17] | 1.10 [0.96-1.25] | 1.03 [0.90-1.18] | 1.08 [0.94-1.24] | | 1.00 [0.87-1.15] | 667 | 678 | 656 | NA | 598 |
| M89 | 1.22 [1.14-1.30] | 1.19 [1.11-1.27] | 1.21 [1.12-1.30] | 1.03 [0.77-1.37] | 1.14 [1.07-1.22] | 1.15 [1.07-1.23] | 1.10 [1.02-1.18] | 1.11 [1.03-1.20] | 1.00 [0.74-1.35] | 1.10 [1.03-1.18] | 2,554 | 2,501 | 2,102 | 157 | 2,432 |
| M90 | 1.06 [0.95-1.18] | 1.03 [0.93-1.15] | 1.02 [0.91-1.13] | | 0.97 [0.87-1.08] | 1.06 [0.95-1.18] | 1.03 [0.92-1.14] | 1.00 [0.90-1.12] | | 0.99 [0.88-1.10] | 982 | 978 | 944 | NA | 895 |
| M91 | 1.17 [0.87-1.58] | | | | 1.17 [0.86-1.59] | 1.12 [0.83-1.53] | | | | 1.12 [0.82-1.53] | 137 | NA | NA | NA | 122 |
| M92 | 1.38 [1.10-1.72] | 1.36 [1.08-1.71] | 1.33 [1.00-1.77] | | 1.27 [1.01-1.59] | 1.27 [1.01-1.60] | 1.24 [0.98-1.57] | 1.20 [0.89-1.62] | | 1.16 [0.91-1.47] | 266 | 244 | 142 | NA | 242 |
| M93 | 1.31 [1.11-1.55] | 1.21 [1.02-1.43] | 1.25 [1.02-1.53] | | 1.13 [0.95-1.33] | 1.22 [1.03-1.45] | 1.10 [0.92-1.31] | 1.16 [0.94-1.43] | | 1.05 [0.88-1.25] | 462 | 406 | 276 | NA | 408 |
| M94 | 1.25 [1.16-1.35] | 1.28 [1.18-1.38] | 1.33 [1.21-1.45] | 0.99 [0.75-1.32] | 1.25 [1.15-1.35] | 1.12 [1.03-1.22] | 1.12 [1.03-1.22] | 1.18 [1.07-1.29] | 0.98 [0.73-1.32] | 1.17 [1.08-1.27] | 1,962 | 1,982 | 1,444 | 170 | 1,932 |
| M95 | 1.27 [1.06-1.52] | 1.23 [1.02-1.47] | 1.29 [1.02-1.63] | | 1.22 [1.02-1.47] | 1.22 [1.01-1.46] | 1.14 [0.94-1.37] | 1.18 [0.93-1.51] | | 1.16 [0.96-1.40] | 391 | 374 | 208 | NA | 346 |
| M96 | 1.24 [1.14-1.36] | 1.23 [1.12-1.34] | 1.25 [1.15-1.37] | | 1.25 [1.14-1.36] | 1.19 [1.08-1.30] | 1.15 [1.05-1.26] | 1.17 [1.07-1.29] | | 1.23 [1.12-1.34] | 1,544 | 1,537 | 1,475 | NA | 1,548 |
| M99 | 1.25 [1.06-1.47] | 1.18 [1.01-1.40] | 1.24 [1.05-1.47] | | 1.18 [1.00-1.39] | 1.13 [0.96-1.34] | 1.07 [0.90-1.26] | 1.09 [0.92-1.30] | | 1.10 [0.94-1.30] | 438 | 433 | 415 | NA | 420 |
| Diseases of the genitourinary system (N00-N99) | | | | | | | | | | | | | | | |
| N02 | 1.17 [1.05-1.29] | 1.19 [1.07-1.32] | 1.20 [1.07-1.33] | | 1.12 [1.01-1.24] | 1.14 [1.03-1.27] | 1.14 [1.03-1.27] | 1.16 [1.04-1.29] | | 1.11 [1.00-1.23] | 1,082 | 1,092 | 959 | NA | 1,042 |
| N03 | 1.20 [1.14-1.25] | 1.21 [1.15-1.27] | 1.21 [1.16-1.27] | | 1.20 [1.15-1.26] | 1.15 [1.10-1.21] | 1.13 [1.08-1.19] | 1.14 [1.08-1.19] | | 1.17 [1.12-1.23] | 5,200 | 5,207 | 5,138 | NA | 5,039 |
| N04 | 1.28 [1.12-1.46] | 1.30 [1.14-1.49] | 1.22 [1.06-1.40] | | 1.14 [1.00-1.30] | 1.21 [1.06-1.38] | 1.23 [1.07-1.41] | 1.14 [0.99-1.32] | | 1.12 [0.98-1.28] | 698 | 661 | 586 | NA | 655 |
| N05 | 1.37 [1.20-1.56] | 1.30 [1.14-1.49] | 1.31 [1.14-1.51] | | 1.26 [1.11-1.44] | 1.33 [1.16-1.52] | 1.23 [1.07-1.41] | 1.24 [1.07-1.43] | | 1.24 [1.09-1.41] | 722 | 674 | 591 | NA | 697 |
| N08 | 1.23 [1.15-1.32] | 1.22 [1.14-1.30] | 1.22 [1.14-1.31] | | 1.16 [1.09-1.24] | 1.19 [1.11-1.27] | 1.11 [1.03-1.19] | 1.10 [1.03-1.19] | | 1.16 [1.08-1.24] | 2,513 | 2,519 | 2,385 | NA | 2,486 |

### Supplementary Table 4: Hazard ratios and events from all cohorts (excluding non-consulters)

| | Hazard ratio (99% confidence interval) | | | | | | | | | | Events (in exposed) | | | | |
|---|---|---|---|---|---|---|---|---|---|---|---|---|---|---|---|
| | crude | | | | | adjusted | | | | | | | | | |
| Outcome | any age | 18+ | 40+ | <18 | hosp. | any age | 18+ | 40+ | <18 | hosp. | any age | 18+ | 40+ | <18 | hosp. |
| N10 | 1.10 [0.98-1.25] | 1.10 [0.97-1.24] | 1.16 [1.00-1.36] | 0.92 [0.66-1.30] | 1.06 [0.94-1.20] | 1.05 [0.92-1.19] | 1.01 [0.89-1.15] | 1.08 [0.92-1.26] | 0.95 [0.66-1.37] | 1.03 [0.90-1.17] | 776 | 778 | 472 | 111 | 728 |
| N11 | 1.18 [1.00-1.40] | 1.15 [0.97-1.36] | 1.14 [0.95-1.38] | | 1.05 [0.89-1.25] | 1.14 [0.96-1.35] | 1.09 [0.92-1.29] | 1.07 [0.89-1.30] | | 1.05 [0.88-1.24] | 409 | 408 | 319 | NA | 383 |
| N12 | 1.11 [1.05-1.17] | 1.13 [1.08-1.19] | 1.15 [1.08-1.23] | 1.01 [0.87-1.16] | 1.06 [1.01-1.12] | 1.04 [0.99-1.10] | 1.03 [0.98-1.09] | 1.06 [0.99-1.13] | 1.00 [0.86-1.16] | 1.04 [0.98-1.10] | 4,313 | 4,485 | 2,601 | 647 | 4,050 |
| N13 | 1.01 [0.98-1.05] | 1.02 [0.98-1.05] | 1.03 [0.99-1.06] | 0.83 [0.69-1.00] | 1.00 [0.97-1.04] | 1.00 [0.97-1.04] | 1.00 [0.96-1.04] | 1.01 [0.98-1.05] | 0.81 [0.67-0.97] | 1.01 [0.98-1.05] | 9,107 | 9,032 | 8,165 | 372 | 8,586 |
| N14 | 1.13 [0.87-1.46] | 1.16 [0.89-1.51] | 1.24 [0.95-1.61] | | 1.17 [0.91-1.51] | 1.12 [0.86-1.45] | 1.09 [0.83-1.42] | 1.17 [0.90-1.54] | | 1.14 [0.88-1.48] | 178 | 178 | 170 | NA | 177 |
| N15 | 1.12 [0.91-1.38] | 1.11 [0.90-1.36] | 1.07 [0.86-1.34] | | 1.03 [0.84-1.26] | 1.08 [0.87-1.34] | 1.04 [0.84-1.28] | 1.02 [0.81-1.28] | | 1.01 [0.82-1.24] | 263 | 269 | 220 | NA | 264 |
| N16 | 1.31 [0.98-1.75] | 1.28 [0.96-1.71] | 1.27 [0.93-1.74] | | 1.15 [0.87-1.52] | 1.29 [0.96-1.74] | 1.19 [0.88-1.60] | 1.17 [0.85-1.62] | | 1.18 [0.89-1.56] | 147 | 149 | 118 | NA | 149 |
| N17 | 1.14 [1.13-1.16] | 1.14 [1.12-1.15] | 1.15 [1.13-1.16] | 0.80 [0.66-0.98] | 1.13 [1.11-1.14] | 1.12 [1.10-1.14] | 1.08 [1.07-1.10] | 1.09 [1.07-1.10] | 0.82 [0.66-1.01] | 1.12 [1.10-1.14] | 63,036 | 63,147 | 61,984 | 322 | 61,515 |
| N18 | 1.15 [1.13-1.16] | 1.15 [1.13-1.17] | 1.15 [1.13-1.17] | 1.00 [0.70-1.42] | 1.12 [1.11-1.14] | 1.11 [1.10-1.13] | 1.08 [1.07-1.10] | 1.08 [1.06-1.10] | 0.98 [0.68-1.41] | 1.11 [1.09-1.12] | 55,729 | 55,773 | 55,339 | 109 | 54,713 |
| N19 | 1.20 [1.15-1.25] | 1.20 [1.16-1.25] | 1.19 [1.14-1.23] | | 1.19 [1.14-1.23] | 1.16 [1.12-1.21] | 1.14 [1.09-1.18] | 1.12 [1.08-1.17] | | 1.16 [1.12-1.21] | 7,585 | 7,557 | 7,382 | NA | 7,161 |
| N20 | 1.04 [1.00-1.08] | 1.04 [1.00-1.08] | 1.04 [1.00-1.08] | 0.78 [0.61-0.99] | 1.04 [1.00-1.08] | 1.01 [0.97-1.05] | 0.99 [0.95-1.03] | 0.99 [0.95-1.03] | 0.82 [0.63-1.06] | 1.03 [0.98-1.07] | 7,234 | 7,316 | 6,153 | 196 | 6,744 |
| N21 | 0.94 [0.87-1.02] | 0.92 [0.85-0.99] | 0.94 [0.87-1.02] | | 0.91 [0.84-0.99] | 0.94 [0.87-1.02] | 0.91 [0.84-0.99] | 0.93 [0.86-1.01] | | 0.92 [0.85-1.00] | 1,683 | 1,664 | 1,588 | NA | 1,555 |
| N23 | 1.06 [0.98-1.16] | 1.04 [0.95-1.13] | 1.01 [0.92-1.12] | | 0.99 [0.91-1.08] | 1.00 [0.91-1.09] | 0.96 [0.88-1.05] | 0.93 [0.84-1.03] | | 0.95 [0.87-1.04] | 1,495 | 1,565 | 1,034 | NA | 1,354 |
| N25 | 1.20 [1.02-1.40] | 1.24 [1.05-1.45] | 1.24 [1.05-1.46] | | 1.07 [0.91-1.25] | 1.17 [1.00-1.37] | 1.19 [1.01-1.40] | 1.19 [1.01-1.40] | | 1.07 [0.92-1.26] | 467 | 457 | 431 | NA | 439 |
| N26 | 1.08 [0.96-1.21] | 1.08 [0.96-1.21] | 1.05 [0.93-1.18] | | 1.03 [0.92-1.15] | 1.04 [0.92-1.17] | 1.00 [0.89-1.13] | 0.97 [0.86-1.10] | | 1.02 [0.91-1.15] | 857 | 858 | 791 | NA | 858 |
| N27 | 1.24 [1.06-1.45] | 1.22 [1.04-1.44] | 1.28 [1.08-1.51] | | 1.12 [0.95-1.30] | 1.23 [1.05-1.44] | 1.17 [1.00-1.38] | 1.23 [1.03-1.45] | | 1.10 [0.94-1.29] | 487 | 460 | 417 | NA | 459 |
| N28 | 1.10 [1.06-1.13] | 1.09 [1.06-1.13] | 1.09 [1.06-1.13] | 0.89 [0.70-1.13] | 1.08 [1.04-1.11] | 1.07 [1.03-1.10] | 1.03 [1.00-1.07] | 1.04 [1.00-1.07] | 0.93 [0.73-1.20] | 1.06 [1.03-1.10] | 11,855 | 11,792 | 11,234 | 220 | 11,417 |
| N30 | 1.16 [1.11-1.21] | 1.16 [1.11-1.21] | 1.12 [1.06-1.17] | 1.16 [0.86-1.57] | 1.12 [1.07-1.17] | 1.12 [1.07-1.17] | 1.10 [1.05-1.15] | 1.06 [1.01-1.11] | 1.11 [0.82-1.52] | 1.10 [1.05-1.15] | 5,348 | 5,414 | 4,754 | 155 | 5,061 |
| N31 | 1.23 [1.13-1.34] | 1.28 [1.18-1.40] | 1.26 [1.15-1.38] | | 1.15 [1.06-1.25] | 1.15 [1.05-1.25] | 1.16 [1.06-1.27] | 1.15 [1.05-1.26] | | 1.11 [1.02-1.20] | 1,598 | 1,581 | 1,374 | NA | 1,545 |
| N32 | 1.13 [1.10-1.16] | 1.12 [1.09-1.16] | 1.12 [1.09-1.15] | 0.94 [0.76-1.15] | 1.10 [1.07-1.13] | 1.09 [1.06-1.12] | 1.07 [1.04-1.10] | 1.07 [1.04-1.10] | 0.92 [0.74-1.15] | 1.08 [1.05-1.11] | 15,374 | 15,340 | 14,355 | 299 | 14,514 |
| N34 | 1.23 [0.99-1.54] | 1.17 [0.94-1.47] | 1.14 [0.89-1.46] | | 1.11 [0.88-1.40] | 1.21 [0.97-1.52] | 1.14 [0.91-1.44] | 1.11 [0.87-1.43] | | 1.10 [0.87-1.39] | 242 | 233 | 190 | NA | 206 |
| N35 | 1.20 [1.15-1.26] | 1.18 [1.13-1.24] | 1.19 [1.14-1.25] | 1.14 [0.88-1.48] | 1.14 [1.09-1.20] | 1.16 [1.11-1.22] | 1.12 [1.07-1.18] | 1.14 [1.08-1.20] | 1.09 [0.83-1.42] | 1.12 [1.07-1.18] | 5,195 | 5,138 | 4,621 | 212 | 4,820 |
| N36 | 1.15 [1.05-1.26] | 1.16 [1.06-1.27] | 1.13 [1.03-1.24] | | 1.07 [0.97-1.17] | 1.11 [1.02-1.22] | 1.12 [1.03-1.23] | 1.07 [0.98-1.18] | | 1.05 [0.95-1.15] | 1,420 | 1,392 | 1,235 | NA | 1,284 |
| N39 | 1.12 [1.11-1.14] | 1.12 [1.11-1.14] | 1.12 [1.11-1.14] | 1.05 [0.97-1.13] | 1.10 [1.09-1.12] | 1.09 [1.08-1.11] | 1.07 [1.06-1.08] | 1.07 [1.06-1.09] | 1.02 [0.95-1.11] | 1.09 [1.07-1.10] | 71,936 | 71,311 | 66,045 | 2,343 | 67,612 |
| N40 | 1.08 [1.06-1.10] | 1.08 [1.05-1.10] | 1.07 [1.05-1.10] | | 1.06 [1.04-1.09] | 1.05 [1.02-1.07] | 1.04 [1.02-1.07] | 1.03 [1.01-1.06] | | 1.04 [1.01-1.06] | 23,087 | 23,094 | 23,034 | NA | 21,241 |
| N41 | 1.03 [0.94-1.12] | 1.01 [0.93-1.11] | 1.00 [0.91-1.09] | | 1.00 [0.92-1.09] | 1.00 [0.91-1.09] | 0.98 [0.90-1.08] | 0.97 [0.89-1.06] | | 0.98 [0.89-1.07] | 1,382 | 1,385 | 1,366 | NA | 1,291 |
| N42 | 1.04 [0.97-1.12] | 1.02 [0.95-1.10] | 1.04 [0.97-1.12] | | 1.00 [0.93-1.08] | 1.02 [0.94-1.10] | 1.00 [0.93-1.08] | 1.01 [0.94-1.09] | | 0.99 [0.92-1.07] | 1,963 | 1,959 | 1,954 | NA | 1,837 |
| N43 | 1.05 [0.98-1.13] | 1.11 [1.02-1.20] | 1.08 [1.00-1.18] | 1.02 [0.86-1.20] | 1.11 [1.02-1.19] | 1.04 [0.96-1.12] | 1.08 [1.00-1.18] | 1.06 [0.97-1.15] | 0.98 [0.83-1.17] | 1.10 [1.01-1.18] | 2,105 | 1,693 | 1,601 | 461 | 1,909 |
| N44 | 1.19 [0.96-1.47] | | | 1.12 [0.89-1.40] | 1.20 [0.94-1.53] | 1.16 [0.93-1.45] | | | 1.11 [0.88-1.40] | 1.19 [0.93-1.52] | 335 | NA | NA | 310 | 269 |
| N45 | 1.08 [0.99-1.17] | 1.12 [1.02-1.23] | 1.10 [1.00-1.22] | 1.05 [0.82-1.34] | 1.06 [0.97-1.15] | 1.04 [0.95-1.14] | 1.05 [0.96-1.16] | 1.04 [0.94-1.15] | 1.00 [0.78-1.30] | 1.04 [0.95-1.14] | 1,561 | 1,400 | 1,224 | 233 | 1,425 |
| N47 | 1.26 [1.19-1.32] | 1.22 [1.15-1.29] | 1.23 [1.16-1.31] | 1.34 [1.22-1.46] | 1.22 [1.16-1.29] | 1.22 [1.16-1.29] | 1.17 [1.10-1.24] | 1.18 [1.11-1.26] | 1.32 [1.20-1.45] | 1.21 [1.15-1.28] | 5,234 | 3,611 | 3,141 | 1,811 | 4,483 |
| N48 | 1.25 [1.18-1.33] | 1.29 [1.21-1.38] | 1.24 [1.15-1.32] | 1.25 [1.09-1.45] | 1.22 [1.15-1.30] | 1.21 [1.14-1.29] | 1.22 [1.14-1.31] | 1.17 [1.09-1.26] | 1.23 [1.06-1.42] | 1.20 [1.13-1.28] | 3,468 | 2,865 | 2,645 | 697 | 3,136 |
| N49 | 1.15 [0.98-1.35] | 1.32 [1.12-1.55] | 1.24 [1.05-1.47] | | 1.15 [0.98-1.35] | 1.13 [0.96-1.33] | 1.25 [1.06-1.48] | 1.18 [0.99-1.40] | | 1.15 [0.97-1.36] | 455 | 442 | 414 | NA | 427 |
| N50 | 1.11 [1.04-1.18] | 1.14 [1.07-1.21] | 1.14 [1.07-1.22] | 1.00 [0.85-1.17] | 1.10 [1.04-1.17] | 1.07 [1.00-1.14] | 1.09 [1.02-1.16] | 1.09 [1.02-1.17] | 0.96 [0.81-1.14] | 1.08 [1.01-1.15] | 3,220 | 2,832 | 2,528 | 536 | 2,959 |
| N60 | 1.14 [1.04-1.24] | 1.09 [1.00-1.19] | 1.11 [1.00-1.22] | | 1.06 [0.97-1.16] | 1.10 [1.00-1.20] | 1.04 [0.95-1.13] | 1.07 [0.97-1.18] | | 1.05 [0.95-1.15] | 1,436 | 1,450 | 1,115 | NA | 1,265 |
| N61 | 1.27 [1.15-1.41] | 1.29 [1.17-1.43] | 1.41 [1.23-1.61] | | 1.25 [1.12-1.39] | 1.23 [1.10-1.37] | 1.18 [1.06-1.32] | 1.29 [1.12-1.48] | | 1.22 [1.10-1.36] | 1,109 | 1,127 | 649 | NA | 1,004 |
| N62 | 1.16 [1.03-1.32] | 1.20 [1.07-1.36] | 1.24 [1.07-1.44] | | 1.15 [1.01-1.31] | 1.10 [0.97-1.26] | 1.12 [0.99-1.27] | 1.15 [0.99-1.34] | | 1.12 [0.98-1.27] | 754 | 784 | 536 | NA | 661 |
| N63 | 1.11 [1.02-1.21] | 1.10 [1.01-1.19] | 1.16 [1.06-1.27] | | 1.06 [0.98-1.16] | 1.07 [0.98-1.16] | 1.04 [0.96-1.14] | 1.09 [0.99-1.20] | | 1.05 [0.96-1.14] | 1,637 | 1,646 | 1,346 | NA | 1,444 |
| N64 | 1.19 [1.10-1.30] | 1.17 [1.08-1.27] | 1.20 [1.09-1.31] | | 1.19 [1.09-1.29] | 1.15 [1.06-1.26] | 1.09 [1.01-1.19] | 1.13 [1.02-1.24] | | 1.16 [1.06-1.26] | 1,640 | 1,676 | 1,263 | NA | 1,517 |
| N70 | 1.04 [0.93-1.16] | 1.09 [0.98-1.21] | 0.97 [0.84-1.12] | | 1.04 [0.93-1.16] | 1.01 [0.90-1.13] | 1.03 [0.92-1.15] | 0.92 [0.79-1.07] | | 1.01 [0.90-1.14] | 963 | 997 | 495 | NA | 845 |
| N71 | 1.12 [0.97-1.30] | 1.21 [1.05-1.39] | 1.25 [1.05-1.49] | | 1.11 [0.96-1.29] | 1.06 [0.91-1.23] | 1.14 [0.99-1.32] | 1.17 [0.97-1.40] | | 1.05 [0.90-1.22] | 550 | 577 | 354 | NA | 518 |
| N72 | 1.08 [0.97-1.20] | 1.09 [0.98-1.22] | 1.10 [0.95-1.27] | | 1.01 [0.90-1.13] | 1.04 [0.93-1.16] | 1.06 [0.95-1.19] | 1.06 [0.92-1.23] | | 0.98 [0.88-1.11] | 946 | 995 | 506 | NA | 811 |
| N73 | 1.07 [1.02-1.11] | 1.08 [1.03-1.12] | 1.07 [1.01-1.14] | 1.02 [0.87-1.19] | 1.04 [1.00-1.09] | 1.02 [0.97-1.06] | 1.00 [0.95-1.05] | 1.01 [0.95-1.08] | 1.04 [0.88-1.22] | 1.01 [0.97-1.06] | 5,763 | 6,012 | 2,721 | 547 | 5,310 |
| N75 | 1.03 [0.92-1.16] | 1.03 [0.92-1.15] | 1.03 [0.86-1.22] | | 1.01 [0.90-1.14] | 1.01 [0.90-1.13] | 0.98 [0.87-1.10] | 1.00 [0.84-1.20] | | 1.01 [0.89-1.14] | 858 | 904 | 333 | NA | 743 |
| N76 | 1.32 [1.21-1.44] | 1.32 [1.21-1.44] | 1.37 [1.23-1.52] | 1.10 [0.85-1.43] | 1.33 [1.22-1.45] | 1.25 [1.14-1.36] | 1.21 [1.10-1.32] | 1.28 [1.15-1.43] | 1.13 [0.86-1.47] | 1.28 [1.18-1.40] | 1,675 | 1,653 | 1,035 | 208 | 1,517 |
| N77 | 1.18 [1.10-1.28] | 1.21 [1.12-1.30] | 1.36 [1.21-1.53] | 0.90 [0.74-1.10] | 1.14 [1.06-1.23] | 1.10 [1.01-1.19] | 1.07 [1.00-1.16] | 1.19 [1.05-1.35] | 0.88 [0.71-1.09] | 1.08 [1.00-1.17] | 2,075 | 2,268 | 875 | 308 | 2,002 |
| N80 | 1.07 [1.02-1.12] | 1.09 [1.04-1.14] | 1.03 [0.97-1.11] | 0.96 [0.83-1.11] | 1.05 [1.01-1.11] | 1.01 [0.96-1.06] | 1.01 [0.96-1.05] | 0.98 [0.92-1.05] | 0.93 [0.80-1.09] | 1.01 [0.96-1.06] | 5,599 | 5,944 | 2,392 | 578 | 4,982 |
| N81 | 1.14 [1.11-1.18] | 1.14 [1.10-1.18] | 1.13 [1.09-1.17] | | 1.11 [1.07-1.15] | 1.10 [1.06-1.13] | 1.09 [1.05-1.12] | 1.08 [1.04-1.12] | | 1.07 [1.03-1.11] | 10,329 | 10,342 | 9,347 | NA | 9,208 |
| N82 | 1.25 [1.08-1.45] | 1.20 [1.04-1.39] | 1.18 [1.01-1.39] | | 1.22 [1.06-1.42] | 1.19 [1.03-1.39] | 1.13 [0.97-1.31] | 1.10 [0.93-1.29] | | 1.19 [1.03-1.38] | 528 | 523 | 427 | NA | 510 |
| N83 | 1.08 [1.04-1.12] | 1.08 [1.05-1.12] | 1.06 [1.01-1.11] | 0.96 [0.86-1.08] | 1.05 [1.01-1.09] | 1.03 [0.99-1.07] | 1.00 [0.97-1.04] | 1.01 [0.96-1.06] | 0.94 [0.84-1.06] | 1.01 [0.98-1.05] | 9,134 | 9,544 | 4,765 | 968 | 8,230 |
| N84 | 1.09 [1.05-1.13] | 1.09 [1.05-1.13] | 1.09 [1.05-1.14] | | 1.06 [1.01-1.10] | 1.08 [1.04-1.12] | 1.07 [1.03-1.12] | 1.08 [1.04-1.13] | | 1.05 [1.01-1.09] | 7,674 | 7,738 | 6,706 | NA | 6,387 |
| N85 | 1.11 [1.07-1.16] | 1.12 [1.07-1.16] | 1.10 [1.05-1.15] | 1.06 [0.82-1.35] | 1.08 [1.04-1.13] | 1.08 [1.04-1.13] | 1.07 [1.02-1.11] | 1.06 [1.01-1.11] | 1.00 [0.77-1.31] | 1.07 [1.02-1.11] | 6,475 | 6,591 | 5,058 | 214 | 5,716 |
| N86 | 1.09 [1.01-1.18] | 1.10 [1.02-1.19] | 1.14 [0.98-1.33] | 0.88 [0.70-1.09] | 1.06 [0.98-1.16] | 1.05 [0.97-1.14] | 1.05 [0.97-1.14] | 1.11 [0.95-1.30] | 0.88 [0.70-1.11] | 1.02 [0.94-1.11] | 1,855 | 2,036 | 466 | 256 | 1,621 |
| N87 | 1.03 [0.96-1.11] | 1.06 [0.98-1.14] | 1.00 [0.87-1.13] | 1.19 [0.94-1.51] | 1.04 [0.96-1.13] | 1.00 [0.93-1.08] | 1.01 [0.93-1.08] | 0.96 [0.84-1.10] | 1.23 [0.95-1.58] | 1.04 [0.96-1.12] | 2,147 | 2,231 | 608 | 245 | 1,742 |
| N88 | 1.15 [1.06-1.24] | 1.14 [1.06-1.23] | 1.11 [1.02-1.21] | | 1.11 [1.02-1.20] | 1.12 [1.04-1.21] | 1.11 [1.03-1.20] | 1.07 [0.98-1.16] | | 1.09 [1.01-1.18] | 1,943 | 1,984 | 1,583 | NA | 1,724 |
| N89 | 1.19 [1.12-1.26] | 1.20 [1.14-1.27] | 1.24 [1.13-1.35] | 0.92 [0.77-1.09] | 1.12 [1.05-1.19] | 1.13 [1.07-1.21] | 1.11 [1.05-1.18] | 1.18 [1.08-1.29] | 0.89 [0.74-1.07] | 1.08 [1.01-1.15] | 3,319 | 3,497 | 1,512 | 389 | 3,009 |
| N90 | 1.29 [1.22-1.37] | 1.26 [1.19-1.34] | 1.31 [1.22-1.40] | 1.01 [0.77-1.33] | 1.21 [1.14-1.29] | 1.23 [1.16-1.31] | 1.19 [1.12-1.27] | 1.23 [1.15-1.32] | 1.04 [0.78-1.38] | 1.17 [1.10-1.24] | 3,385 | 3,431 | 2,367 | 178 | 3,050 |
| N91 | 1.22 [1.01-1.47] | 1.23 [1.02-1.48] | 1.65 [1.20-2.27] | | 1.16 [0.95-1.40] | 1.16 [0.95-1.41] | 1.09 [0.90-1.33] | 1.61 [1.16-2.24] | | 1.09 [0.89-1.32] | 343 | 349 | 121 | NA | 305 |
| N92 | 1.13 [1.09-1.16] | 1.14 [1.11-1.18] | 1.13 [1.09-1.18] | 1.01 [0.88-1.16] | 1.10 [1.07-1.14] | 1.07 [1.04-1.11] | 1.06 [1.02-1.09] | 1.07 [1.03-1.11] | 1.01 [0.87-1.16] | 1.06 [1.03-1.10] | 12,290 | 12,542 | 7,436 | 711 | 10,871 |
| N93 | 1.11 [1.07-1.16] | 1.12 [1.07-1.17] | 1.11 [1.04-1.18] | 0.99 [0.86-1.14] | 1.04 [0.99-1.08] | 1.07 [1.02-1.12] | 1.04 [0.99-1.08] | 1.05 [0.99-1.12] | 0.99 [0.86-1.15] | 1.01 [0.97-1.06] | 5,970 | 6,258 | 2,939 | 622 | 5,380 |
| N94 | 1.17 [1.11-1.23] | 1.18 [1.12-1.24] | 1.17 [1.08-1.26] | 1.13 [0.98-1.31] | 1.14 [1.09-1.20] | 1.09 [1.04-1.15] | 1.06 [1.01-1.12] | 1.09 [1.00-1.17] | 1.10 [0.94-1.28] | 1.08 [1.03-1.14] | 4,819 | 5,103 | 1,880 | 626 | 4,356 |
| N95 | 1.18 [1.14-1.23] | 1.15 [1.11-1.20] | 1.18 [1.13-1.22] | | 1.14 [1.10-1.19] | 1.16 [1.11-1.20] | 1.11 [1.07-1.15] | 1.14 [1.09-1.18] | | 1.12 [1.08-1.17] | 7,543 | 7,524 | 7,504 | NA | 6,451 |
| N97 | 0.89 [0.79-1.00] | 0.93 [0.83-1.05] | | | 1.01 [0.89-1.16] | 0.88 [0.78-0.99] | 0.93 [0.82-1.04] | | | 1.01 [0.88-1.16] | 781 | 805 | NA | NA | 534 |
| N99 | 1.13 [1.04-1.22] | 1.13 [1.05-1.22] | 1.11 [1.02-1.21] | | 1.11 [1.02-1.20] | 1.10 [1.01-1.19] | 1.08 [1.00-1.17] | 1.07 [0.98-1.16] | | 1.09 [1.00-1.18] | 1,838 | 1,819 | 1,505 | NA | 1,733 |
| Pregnancy, childbirth and the puerperium (O00-O9A) | | | | | | | | | | | | | | | |
| O00 | 0.96 [0.89-1.05] | 1.01 [0.93-1.09] | | 1.02 [0.81-1.28] | 0.96 [0.88-1.05] | 0.95 [0.87-1.04] | 0.97 [0.89-1.06] | | 1.03 [0.81-1.31] | 0.96 [0.88-1.05] | 1,619 | 1,737 | NA | 247 | 1,479 |
| O02 | 1.01 [0.95-1.06] | 1.02 [0.97-1.07] | 0.91 [0.77-1.07] | 0.91 [0.77-1.07] | 1.01 [0.95-1.06] | 0.99 [0.93-1.04] | 1.00 [0.95-1.06] | 0.94 [0.80-1.12] | 0.93 [0.78-1.11] | 1.00 [0.95-1.06] | 3,955 | 4,214 | 375 | 441 | 3,411 |
| O03 | 1.00 [0.95-1.04] | 0.99 [0.95-1.03] | 0.93 [0.81-1.06] | 0.92 [0.81-1.04] | 0.98 [0.93-1.02] | 0.99 [0.95-1.04] | 0.97 [0.92-1.01] | 0.95 [0.82-1.08] | 0.93 [0.82-1.06] | 0.98 [0.94-1.03] | 5,746 | 6,180 | 560 | 794 | 5,036 |
| O04 | 0.99 [0.94-1.03] | 0.98 [0.94-1.02] | 0.90 [0.75-1.07] | 0.97 [0.88-1.06] | 0.94 [0.90-0.98] | 0.98 [0.93-1.03] | 0.95 [0.91-0.99] | 0.92 [0.76-1.11] | 1.02 [0.92-1.12] | 0.95 [0.90-0.99] | 5,800 | 6,654 | 311 | 1,416 | 5,051 |
| O08 | 0.99 [0.86-1.14] | 0.98 [0.86-1.12] | | 0.99 [0.74-1.34] | 0.98 [0.86-1.13] | 0.96 [0.83-1.11] | 0.93 [0.81-1.06] | | 1.10 [0.79-1.53] | 0.99 [0.86-1.14] | 578 | 644 | NA | 131 | 554 |
| O10 | 0.96 [0.85-1.09] | 1.00 [0.88-1.12] | 1.05 [0.80-1.37] | | 0.95 [0.84-1.07] | 0.94 [0.83-1.07] | 0.94 [0.83-1.07] | 0.94 [0.71-1.25] | | 0.93 [0.82-1.06] | 744 | 787 | 142 | NA | 692 |
| O11 | 0.98 [0.72-1.35] | 1.04 [0.76-1.41] | | | 1.07 [0.77-1.49] | 0.93 [0.67-1.30] | 0.94 [0.68-1.30] | | | 1.04 [0.74-1.45] | 115 | 121 | NA | NA | 108 |
| O12 | 1.05 [0.96-1.15] | 1.07 [0.98-1.17] | | 0.91 [0.72-1.15] | 1.04 [0.94-1.14] | 1.01 [0.92-1.12] | 1.01 [0.92-1.11] | | 0.90 [0.70-1.14] | 1.00 [0.90-1.10] | 1,328 | 1,473 | NA | 234 | 1,175 |
| O13 | 1.00 [0.94-1.06] | 1.02 [0.97-1.08] | 0.91 [0.73-1.13] | 0.94 [0.80-1.09] | 1.02 [0.96-1.08] | 0.99 [0.93-1.04] | 1.01 [0.95-1.07] | 0.92 [0.73-1.15] | 0.94 [0.80-1.10] | 1.01 [0.95-1.08] | 3,626 | 4,006 | 201 | 519 | 2,911 |
| O14 | 1.02 [0.95-1.09] | 1.01 [0.95-1.08] | 0.92 [0.70-1.21] | 0.99 [0.83-1.17] | 1.00 [0.93-1.08] | 1.01 [0.94-1.08] | 1.00 [0.93-1.07] | 0.92 [0.69-1.22] | 1.02 [0.85-1.22] | 0.98 [0.91-1.06] | 2,545 | 2,861 | 136 | 448 | 1,996 |
| O16 | 1.00 [0.94-1.07] | 1.00 [0.94-1.06] | 0.81 [0.63-1.04] | 0.92 [0.77-1.11] | 0.99 [0.92-1.06] | 1.00 [0.93-1.06] | 0.99 [0.93-1.05] | 0.82 [0.63-1.06] | 0.92 [0.76-1.11] | 0.98 [0.91-1.05] | 2,949 | 3,170 | 152 | 376 | 2,303 |
| O20 | 1.00 [0.95-1.05] | 1.03 [0.98-1.08] | 0.89 [0.72-1.10] | 0.93 [0.83-1.04] | 0.98 [0.94-1.03] | 0.98 [0.93-1.03] | 0.99 [0.94-1.04] | 0.96 [0.77-1.20] | 0.94 [0.84-1.06] | 0.98 [0.93-1.03] | 4,983 | 5,510 | 220 | 945 | 4,460 |
| O21 | 1.03 [0.97-1.09] | 1.06 [1.01-1.12] | | 0.88 [0.78-0.99] | 0.98 [0.93-1.04] | 0.98 [0.92-1.04] | 0.98 [0.93-1.04] | | 0.88 [0.77-1.00] | 0.96 [0.91-1.02] | 3,415 | 3,979 | NA | 821 | 3,147 |
| O22 | 1.03 [0.90-1.18] | 1.00 [0.87-1.13] | | | 0.99 [0.87-1.13] | 1.04 [0.91-1.20] | 1.01 [0.88-1.15] | | | 1.01 [0.88-1.15] | 654 | 664 | NA | NA | 603 |
| O23 | 1.07 [1.02-1.13] | 1.10 [1.04-1.16] | | 0.94 [0.84-1.06] | 1.05 [1.00-1.11] | 1.04 [0.99-1.10] | 1.03 [0.98-1.09] | | 0.95 [0.84-1.08] | 1.04 [0.98-1.10] | 4,038 | 4,631 | NA | 895 | 3,752 |
| O24 | 0.97 [0.93-1.02] | 0.99 [0.94-1.03] | 0.92 [0.80-1.07] | 0.86 [0.75-0.99] | 1.00 [0.95-1.04] | 0.97 [0.93-1.02] | 0.99 [0.94-1.03] | 0.93 [0.80-1.08] | 0.92 [0.79-1.06] | 0.99 [0.94-1.04] | 5,280 | 5,620 | 470 | 646 | 4,797 |
| O26 | 1.06 [1.03-1.09] | 1.08 [1.05-1.11] | 0.83 [0.72-0.96] | 0.89 [0.83-0.95] | 1.03 [1.00-1.06] | 1.03 [1.00-1.06] | 1.02 [1.00-1.05] | 0.83 [0.72-0.97] | 0.89 [0.83-0.95] | 1.01 [0.98-1.04] | 15,473 | 17,384 | 467 | 2,941 | 13,341 |
| O28 | 1.00 [0.91-1.09] | 0.99 [0.91-1.07] | | 0.94 [0.78-1.14] | 0.96 [0.88-1.05] | 0.96 [0.87-1.05] | 0.93 [0.85-1.02] | | 0.94 [0.76-1.15] | 0.94 [0.86-1.03] | 1,479 | 1,654 | NA | 327 | 1,361 |
| O30 | 0.93 [0.86-1.01] | 0.94 [0.87-1.02] | | 1.20 [0.95-1.51] | 0.95 [0.88-1.03] | 0.92 [0.85-1.00] | 0.93 [0.86-1.01] | | 1.23 [0.96-1.57] | 0.94 [0.86-1.02] | 1,737 | 1,884 | NA | 242 | 1,571 |
| O31 | 1.01 [0.72-1.40] | 1.12 [0.80-1.55] | | | 1.06 [0.77-1.47] | 0.97 [0.69-1.38] | 1.03 [0.73-1.46] | | | 1.05 [0.75-1.47] | 106 | 111 | NA | NA | 105 |
| O32 | 0.94 [0.91-0.98] | 0.98 [0.94-1.01] | 0.90 [0.77-1.05] | 0.86 [0.78-0.96] | 0.96 [0.92-1.00] | 0.94 [0.90-0.98] | 0.98 [0.94-1.02] | 0.95 [0.80-1.12] | 0.87 [0.78-0.98] | 0.96 [0.92-1.00] | 7,303 | 7,977 | 390 | 1,013 | 6,152 |
| O33 | 0.94 [0.73-1.21] | 0.97 [0.76-1.23] | | | 0.91 [0.71-1.18] | 0.90 [0.70-1.17] | 0.93 [0.72-1.19] | | | 0.90 [0.69-1.17] | 173 | 180 | NA | NA | 152 |
| O34 | 1.00 [0.97-1.03] | 1.01 [0.98-1.04] | 0.90 [0.81-1.01] | 0.84 [0.76-0.92] | 0.98 [0.95-1.01] | 1.01 [0.97-1.04] | 1.01 [0.98-1.04] | 0.96 [0.86-1.07] | 0.86 [0.78-0.95] | 0.98 [0.95-1.01] | 13,334 | 13,998 | 906 | 1,369 | 13,034 |
| O35 | 0.95 [0.87-1.03] | 0.97 [0.90-1.05] | 0.88 [0.68-1.13] | 0.91 [0.73-1.13] | 0.97 [0.90-1.06] | 0.94 [0.86-1.02] | 0.97 [0.89-1.05] | 0.93 [0.71-1.22] | 0.93 [0.74-1.18] | 0.98 [0.90-1.06] | 1,742 | 1,903 | 149 | 253 | 1,577 |
| O36 | 1.02 [1.00-1.05] | 1.04 [1.02-1.07] | 0.91 [0.81-1.02] | 0.93 [0.88-0.98] | 1.02 [0.99-1.04] | 1.00 [0.98-1.03] | 1.01 [0.99-1.04] | 0.93 [0.82-1.05] | 0.95 [0.89-1.00] | 1.01 [0.98-1.04] | 20,280 | 23,035 | 702 | 4,313 | 17,222 |
| O40 | 1.04 [0.96-1.13] | 1.05 [0.97-1.14] | 0.84 [0.62-1.13] | 0.86 [0.70-1.06] | 1.05 [0.97-1.14] | 1.02 [0.94-1.11] | 1.02 [0.94-1.11] | 0.90 [0.66-1.22] | 0.93 [0.74-1.16] | 1.04 [0.96-1.13] | 1,777 | 1,934 | 104 | 274 | 1,727 |
| O41 | 1.04 [0.97-1.12] | 1.07 [1.00-1.14] | | 0.92 [0.77-1.10] | 1.03 [0.96-1.12] | 1.03 [0.96-1.11] | 1.05 [0.98-1.13] | | 1.00 [0.82-1.21] | 1.03 [0.95-1.11] | 2,213 | 2,433 | NA | 359 | 1,866 |
| O42 | 0.96 [0.93-0.99] | 0.98 [0.95-1.01] | 0.77 [0.65-0.91] | 0.92 [0.85-1.00] | 0.99 [0.96-1.02] | 0.98 [0.94-1.01] | 0.99 [0.96-1.02] | 0.85 [0.71-1.01] | 0.95 [0.87-1.03] | 1.00 [0.97-1.04] | 11,697 | 12,869 | 345 | 1,995 | 9,405 |
| O43 | 0.95 [0.88-1.04] | 0.97 [0.90-1.06] | | 0.89 [0.71-1.11] | 0.98 [0.90-1.07] | 0.95 [0.87-1.04] | 0.98 [0.90-1.06] | | 0.97 [0.76-1.23] | 0.98 [0.90-1.07] | 1,629 | 1,778 | NA | 250 | 1,426 |
| O44 | 0.94 [0.85-1.04] | 0.94 [0.85-1.04] | | 0.80 [0.59-1.10] | 0.97 [0.87-1.07] | 0.92 [0.83-1.02] | 0.91 [0.83-1.01] | | 0.80 [0.58-1.12] | 0.96 [0.86-1.06] | 1,135 | 1,198 | NA | 115 | 1,029 |
| O45 | 0.93 [0.79-1.08] | 0.93 [0.81-1.09] | | | 0.98 [0.84-1.15] | 0.90 [0.76-1.05] | 0.93 [0.79-1.08] | | | 1.00 [0.85-1.17] | 468 | 524 | NA | NA | 437 |
| O46 | 1.04 [0.99-1.09] | 1.07 [1.02-1.11] | 0.97 [0.79-1.20] | 0.90 [0.80-1.01] | 1.02 [0.97-1.06] | 1.02 [0.98-1.07] | 1.03 [0.99-1.08] | 1.01 [0.81-1.26] | 0.91 [0.81-1.03] | 1.01 [0.96-1.06] | 5,711 | 6,317 | 225 | 943 | 4,948 |
| O47 | 1.06 [1.00-1.12] | 1.06 [1.01-1.12] | 0.94 [0.69-1.28] | 0.90 [0.78-1.04] | 1.02 [0.97-1.08] | 1.04 [0.98-1.10] | 1.02 [0.97-1.08] | 0.94 [0.68-1.31] | 0.88 [0.76-1.03] | 1.01 [0.95-1.07] | 3,995 | 4,324 | 102 | 564 | 3,297 |
| O48 | 0.97 [0.93-1.01] | 0.95 [0.92-0.99] | 0.75 [0.60-0.95] | 0.93 [0.84-1.04] | 0.96 [0.92-1.00] | 0.99 [0.95-1.03] | 0.98 [0.94-1.02] | 0.84 [0.66-1.07] | 0.95 [0.85-1.07] | 0.96 [0.92-1.01] | 6,866 | 7,465 | 173 | 1,051 | 5,232 |
| O60 | 1.00 [0.95-1.04] | 1.00 [0.96-1.04] | 0.92 [0.77-1.09] | 0.96 [0.87-1.08] | 1.00 [0.96-1.05] | 0.99 [0.94-1.03] | 0.97 [0.93-1.02] | 0.93 [0.77-1.11] | 1.00 [0.89-1.12] | 1.00 [0.95-1.05] | 6,252 | 6,849 | 313 | 1,041 | 5,501 |
| O61 | 1.04 [0.95-1.15] | 1.05 [0.96-1.16] | | 1.12 [0.86-1.45] | 1.05 [0.94-1.17] | 1.01 [0.91-1.12] | 1.03 [0.93-1.13] | | 1.08 [0.81-1.43] | 1.00 [0.90-1.12] | 1,157 | 1,260 | NA | 195 | 898 |
| O62 | 0.98 [0.93-1.03] | 0.98 [0.93-1.02] | 0.81 [0.64-1.02] | 0.88 [0.78-0.99] | 1.00 [0.95-1.05] | 0.99 [0.94-1.04] | 0.99 [0.94-1.04] | 0.86 [0.67-1.10] | 0.91 [0.80-1.04] | 1.00 [0.95-1.06] | 4,615 | 5,070 | 175 | 805 | 3,840 |
| O63 | 0.94 [0.91-0.97] | 0.95 [0.92-0.98] | 0.84 [0.70-1.00] | 0.93 [0.85-1.01] | 0.98 [0.95-1.02] | 0.95 [0.91-0.98] | 0.96 [0.93-0.99] | 0.93 [0.77-1.12] | 0.92 [0.84-1.01] | 0.98 [0.94-1.02] | 9,989 | 11,007 | 303 | 1,674 | 6,834 |
| O64 | 0.95 [0.88-1.02] | 0.95 [0.88-1.02] | | 0.90 [0.71-1.13] | 0.97 [0.89-1.06] | 0.95 [0.87-1.02] | 0.97 [0.90-1.04] | | 0.84 [0.66-1.08] | 0.98 [0.90-1.07] | 1,892 | 2,061 | NA | 227 | 1,397 |
| O65 | 0.78 [0.58-1.04] | 0.85 [0.64-1.12] | | | | 0.79 [0.59-1.07] | 0.89 [0.67-1.18] | | | | 123 | 135 | NA | NA | NA |
| O66 | 0.93 [0.87-1.01] | 0.95 [0.88-1.02] | | 0.79 [0.64-0.97] | 0.95 [0.87-1.03] | 0.94 [0.87-1.02] | 0.97 [0.89-1.04] | | 0.80 [0.64-0.99] | 0.95 [0.88-1.04] | 1,929 | 2,082 | NA | 288 | 1,536 |
| O67 | 0.97 [0.83-1.12] | 0.98 [0.85-1.13] | | | 1.00 [0.86-1.17] | 0.98 [0.84-1.14] | 1.01 [0.87-1.17] | | | 0.99 [0.84-1.17] | 524 | 546 | NA | NA | 429 |
| O68 | 0.96 [0.94-0.98] | 0.98 [0.96-1.01] | 0.83 [0.74-0.94] | 0.87 [0.82-0.92] | 0.97 [0.94-0.99] | 0.97 [0.95-0.99] | 0.99 [0.97-1.02] | 0.89 [0.79-1.01] | 0.89 [0.84-0.94] | 0.97 [0.94-1.00] | 21,677 | 24,275 | 734 | 3,819 | 16,077 |
| O69 | 0.97 [0.90-1.05] | 1.00 [0.93-1.07] | | 0.88 [0.71-1.08] | 0.99 [0.92-1.07] | 0.98 [0.90-1.05] | 1.01 [0.94-1.09] | | 0.91 [0.73-1.14] | 1.00 [0.92-1.08] | 1,991 | 2,174 | NA | 277 | 1,718 |
| O70 | 0.94 [0.92-0.96] | 0.96 [0.94-0.97] | 0.83 [0.73-0.94] | 0.87 [0.83-0.92] | 0.94 [0.92-0.97] | 0.96 [0.94-0.98] | 0.98 [0.96-1.00] | 0.95 [0.83-1.08] | 0.91 [0.86-0.96] | 0.96 [0.94-0.99] | 29,008 | 32,532 | 669 | 5,391 | 21,905 |
| O71 | 0.94 [0.87-1.02] | 0.97 [0.90-1.04] | | 0.97 [0.80-1.19] | 0.96 [0.88-1.05] | 0.95 [0.88-1.03] | 0.99 [0.92-1.07] | | 1.08 [0.87-1.34] | 0.98 [0.89-1.07] | 1,739 | 1,921 | NA | 305 | 1,447 |
| O72 | 0.98 [0.95-1.00] | 0.99 [0.96-1.02] | 0.89 [0.79-1.00] | 0.94 [0.87-1.00] | 1.00 [0.97-1.03] | 0.99 [0.96-1.02] | 1.01 [0.98-1.03] | 0.97 [0.85-1.10] | 0.96 [0.89-1.03] | 1.00 [0.97-1.03] | 15,399 | 16,864 | 701 | 2,538 | 12,591 |
| O73 | 0.99 [0.90-1.10] | 0.98 [0.89-1.08] | | 0.99 [0.74-1.33] | 1.02 [0.92-1.14] | 1.00 [0.90-1.11] | 0.98 [0.88-1.08] | | 0.94 [0.69-1.29] | 1.02 [0.92-1.14] | 1,084 | 1,137 | NA | 141 | 919 |
| O74 | 1.15 [0.94-1.40] | 1.06 [0.88-1.28] | | | 1.05 [0.86-1.29] | 1.08 [0.88-1.32] | 1.01 [0.83-1.23] | | | 1.03 [0.83-1.28] | 304 | 325 | NA | NA | 257 |
| O75 | 0.95 [0.91-0.99] | 0.96 [0.92-0.99] | 0.86 [0.71-1.04] | 0.76 [0.68-0.84] | 0.98 [0.94-1.02] | 0.96 [0.93-1.00] | 0.97 [0.94-1.01] | 0.94 [0.77-1.14] | 0.76 [0.68-0.85] | 0.99 [0.95-1.04] | 7,301 | 8,033 | 277 | 1,099 | 6,158 |
| O80 | 0.98 [0.94-1.02] | 0.98 [0.95-1.02] | 0.89 [0.71-1.12] | 0.81 [0.73-0.90] | 0.94 [0.91-0.98] | 1.01 [0.97-1.05] | 1.01 [0.97-1.05] | 0.96 [0.76-1.22] | 0.86 [0.77-0.96] | 0.99 [0.95-1.03] | 7,943 | 8,617 | 197 | 1,153 | 7,142 |
| O81 | 1.02 [0.75-1.38] | 0.95 [0.70-1.29] | | | | 1.01 [0.74-1.39] | 0.98 [0.71-1.35] | | | | 111 | 111 | NA | NA | NA |
| O82 | 1.05 [0.96-1.16] | 1.02 [0.93-1.11] | | 1.25 [0.88-1.76] | 0.99 [0.90-1.10] | 1.04 [0.95-1.15] | 1.01 [0.92-1.11] | | 1.31 [0.91-1.90] | 0.98 [0.89-1.09] | 1,228 | 1,269 | NA | 106 | 1,051 |
| O85 | 0.95 [0.83-1.09] | 0.96 [0.84-1.09] | | 0.73 [0.54-1.00] | 1.00 [0.88-1.15] | 0.92 [0.80-1.07] | 0.94 [0.82-1.08] | | 0.77 [0.56-1.07] | 0.98 [0.85-1.14] | 627 | 700 | NA | 122 | 572 |
| O86 | 1.08 [1.01-1.16] | 1.07 [1.00-1.14] | 1.16 [0.88-1.51] | 0.93 [0.78-1.11] | 1.11 [1.03-1.19] | 1.06 [0.98-1.14] | 1.04 [0.97-1.11] | 1.11 [0.84-1.48] | 0.95 [0.79-1.15] | 1.09 [1.01-1.17] | 2,427 | 2,666 | 149 | 411 | 2,090 |
| O87 | 1.16 [0.98-1.39] | 1.24 [1.04-1.47] | | | 1.17 [0.97-1.40] | 1.18 [0.98-1.41] | 1.26 [1.05-1.50] | | | 1.17 [0.96-1.41] | 377 | 390 | NA | NA | 331 |
| O88 | 1.09 [0.81-1.47] | 1.18 [0.89-1.57] | | | 1.11 [0.83-1.48] | 1.05 [0.77-1.43] | 1.14 [0.85-1.53] | | | 1.06 [0.79-1.42] | 137 | 152 | NA | NA | 134 |
| O89 | 1.08 [0.78-1.50] | 1.27 [0.93-1.73] | | | 1.13 [0.81-1.56] | 1.04 [0.74-1.46] | 1.23 [0.89-1.71] | | | 1.10 [0.79-1.54] | 115 | 126 | NA | NA | 101 |
| O90 | 1.07 [1.00-1.14] | 1.07 [1.00-1.13] | 0.96 [0.74-1.24] | 0.90 [0.77-1.06] | 1.05 [0.99-1.12] | 1.05 [0.98-1.12] | 1.03 [0.97-1.10] | 0.97 [0.74-1.27] | 0.88 [0.74-1.05] | 1.03 [0.96-1.10] | 2,926 | 3,204 | 155 | 466 | 2,551 |
| O91 | 0.84 [0.69-1.03] | 0.93 [0.77-1.14] | | | 0.89 [0.72-1.11] | 0.82 [0.66-1.02] | 0.93 [0.76-1.14] | | | 0.86 [0.68-1.07] | 264 | 293 | NA | NA | 220 |
| O92 | 1.00 [0.85-1.17] | 1.01 [0.87-1.18] | | | 0.98 [0.82-1.16] | 0.98 [0.82-1.16] | 1.01 [0.86-1.19] | | | 1.01 [0.84-1.21] | 413 | 454 | NA | NA | 345 |
| O94 | 1.21 [0.87-1.67] | 1.13 [0.83-1.54] | | | 1.18 [0.86-1.61] | 1.12 [0.80-1.58] | 1.02 [0.74-1.43] | | | 1.14 [0.82-1.58] | 117 | 122 | NA | NA | 113 |
| O98 | 1.06 [0.95-1.17] | 1.05 [0.95-1.16] | | 0.82 [0.64-1.06] | 1.05 [0.95-1.17] | 1.04 [0.93-1.16] | 1.03 [0.92-1.14] | | 0.83 [0.63-1.08] | 1.03 [0.93-1.15] | 1,067 | 1,182 | NA | 191 | 988 |
| O99 | 1.08 [1.05-1.10] | 1.08 [1.05-1.11] | 0.95 [0.85-1.07] | 0.93 [0.88-0.99] | 1.06 [1.03-1.09] | 1.05 [1.02-1.07] | 1.03 [1.00-1.05] | 0.94 [0.84-1.07] | 0.95 [0.89-1.02] | 1.04 [1.01-1.07] | 17,744 | 19,624 | 770 | 3,254 | 15,517 |
| O01 | | 0.91 [0.65-1.26] | | | | | 0.89 [0.63-1.26] | | | | NA | 104 | NA | NA | NA |
| O15 | | 1.07 [0.77-1.49] | | | | | 1.02 [0.72-1.43] | | | | NA | 104 | NA | NA | NA |
| Congenital malformations, deformations and chromosomal abnormalities (Q00-Q99) | | | | | | | | | | | | | | | |
| Q02 | 0.97 [0.73-1.28] | | | 0.96 [0.71-1.31] | 0.90 [0.67-1.21] | 0.93 [0.69-1.24] | | | 0.96 [0.70-1.30] | 0.88 [0.65-1.19] | 151 | NA | NA | 135 | 146 |
| Q04 | 1.03 [0.80-1.32] | 1.13 [0.82-1.55] | | | 1.12 [0.87-1.44] | 0.96 [0.74-1.25] | 1.02 [0.74-1.43] | | | 1.11 [0.86-1.44] | 196 | 123 | NA | NA | 190 |
| Q05 | 1.13 [0.91-1.39] | 1.19 [0.97-1.46] | 1.08 [0.84-1.37] | | 1.16 [0.94-1.43] | 1.07 [0.87-1.33] | 1.11 [0.90-1.37] | 0.99 [0.77-1.26] | | 1.07 [0.86-1.33] | 271 | 275 | 181 | NA | 239 |
| Q07 | 1.04 [0.84-1.30] | 1.15 [0.92-1.45] | | | 1.07 [0.87-1.31] | 0.97 [0.78-1.21] | 0.99 [0.78-1.26] | | | 0.99 [0.80-1.22] | 265 | 232 | NA | NA | 267 |
| Q10 | 1.30 [1.02-1.66] | | | 1.25 [0.94-1.67] | 1.20 [0.94-1.53] | 1.31 [1.02-1.66] | | | 1.25 [0.94-1.68] | 1.19 [0.93-1.52] | 210 | NA | NA | 152 | 199 |
| Q12 | 1.29 [0.99-1.68] | 1.15 [0.84-1.57] | | | 1.21 [0.92-1.61] | 1.27 [0.97-1.67] | 1.09 [0.79-1.51] | | | 1.18 [0.89-1.58] | 180 | 124 | NA | NA | 150 |
| Q17 | 0.97 [0.77-1.22] | | | 0.94 [0.74-1.20] | 1.08 [0.83-1.39] | 0.95 [0.75-1.19] | | | 0.93 [0.73-1.18] | 1.07 [0.82-1.39] | 240 | NA | NA | 222 | 195 |
| Q18 | 1.13 [0.93-1.38] | 1.18 [0.91-1.52] | 1.19 [0.86-1.67] | 1.18 [0.86-1.62] | 1.10 [0.88-1.37] | 1.10 [0.89-1.35] | 1.08 [0.83-1.40] | 1.18 [0.85-1.66] | 1.19 [0.86-1.65] | 1.09 [0.87-1.36] | 296 | 188 | 101 | 131 | 244 |
| Q21 | 1.10 [1.00-1.21] | 1.12 [1.01-1.24] | 1.16 [1.03-1.30] | 0.86 [0.68-1.08] | 1.08 [0.98-1.19] | 1.05 [0.95-1.16] | 1.06 [0.95-1.18] | 1.09 [0.96-1.23] | 0.85 [0.67-1.07] | 1.04 [0.94-1.15] | 1,219 | 1,049 | 814 | 223 | 1,111 |
| Q22 | 0.99 [0.71-1.38] | | | | | 1.00 [0.71-1.40] | | | | | 105 | NA | NA | NA | NA |
| Q23 | 1.04 [0.89-1.20] | 1.04 [0.90-1.22] | 0.99 [0.83-1.17] | | 0.91 [0.79-1.06] | 1.01 [0.87-1.18] | 1.01 [0.87-1.18] | 0.97 [0.82-1.15] | | 0.89 [0.77-1.04] | 523 | 476 | 384 | NA | 462 |
| Q24 | 1.09 [0.92-1.29] | 1.21 [1.01-1.45] | 1.16 [0.94-1.43] | | 1.11 [0.94-1.31] | 1.02 [0.85-1.21] | 1.11 [0.92-1.33] | 1.03 [0.82-1.28] | | 1.06 [0.89-1.26] | 413 | 376 | 250 | NA | 398 |
| Q25 | 0.96 [0.80-1.16] | 1.07 [0.87-1.33] | 1.20 [0.93-1.55] | 0.79 [0.58-1.06] | 0.94 [0.78-1.13] | 0.93 [0.77-1.12] | 1.01 [0.81-1.25] | 1.10 [0.85-1.43] | 0.71 [0.51-0.97] | 0.91 [0.75-1.10] | 345 | 252 | 173 | 125 | 328 |
| Q27 | 1.03 [0.85-1.25] | 1.09 [0.88-1.34] | 1.07 [0.84-1.36] | | 0.91 [0.75-1.11] | 0.99 [0.81-1.20] | 1.03 [0.83-1.27] | 0.99 [0.77-1.27] | | 0.90 [0.74-1.09] | 314 | 258 | 192 | NA | 303 |
| Q28 | 1.27 [0.99-1.63] | 1.36 [1.06-1.74] | 1.33 [1.01-1.75] | | 1.27 [0.99-1.62] | 1.24 [0.96-1.59] | 1.29 [1.00-1.67] | 1.28 [0.96-1.69] | | 1.24 [0.96-1.60] | 198 | 197 | 154 | NA | 181 |

### Supplementary Table 4: Hazard ratios and events from all cohorts (excluding non-consulters)

| | Hazard ratio (99% confidence interval) | | | | | | | | | | Events (in exposed) | | | | |
|---|---|---|---|---|---|---|---|---|---|---|---|---|---|---|---|
| | crude | | | | | adjusted | | | | | | | | | |
| Outcome | any age | 18+ | 40+ | <18 | hosp. | any age | 18+ | 40+ | <18 | hosp. | any age | 18+ | 40+ | <18 | hosp. |
| Q31 | 0.96 [0.71-1.31] | | | | 0.84 [0.62-1.14] | 0.91 [0.67-1.24] | | | | 0.80 [0.59-1.09] | 115 | NA | NA | NA | 116 |
| Q35 | 1.11 [0.78-1.58] | | | | | 1.07 [0.74-1.53] | | | | | 101 | NA | NA | NA | NA |
| Q38 | 1.12 [0.99-1.26] | 1.11 [0.95-1.30] | 1.11 [0.95-1.30] | 1.13 [0.95-1.35] | 1.11 [0.98-1.25] | 1.08 [0.96-1.22] | 1.05 [0.90-1.23] | 1.04 [0.88-1.23] | 1.12 [0.93-1.33] | 1.07 [0.95-1.22] | 889 | 492 | 454 | 402 | 802 |
| Q39 | 1.21 [0.98-1.49] | 1.23 [1.00-1.51] | 1.23 [1.00-1.52] | | 1.00 [0.81-1.23] | 1.14 [0.92-1.41] | 1.15 [0.93-1.42] | 1.15 [0.93-1.43] | | 0.92 [0.74-1.14] | 269 | 267 | 254 | NA | 246 |
| Q40 | 1.30 [1.00-1.69] | 1.22 [0.94-1.60] | 1.13 [0.86-1.49] | | 1.24 [0.96-1.61] | 1.19 [0.90-1.56] | 1.07 [0.81-1.41] | 1.02 [0.77-1.35] | | 1.17 [0.90-1.53] | 160 | 160 | 143 | NA | 161 |
| Q43 | 1.19 [1.03-1.38] | 1.15 [0.99-1.34] | 1.20 [1.02-1.42] | | 1.28 [1.11-1.49] | 1.13 [0.98-1.32] | 1.08 [0.92-1.25] | 1.13 [0.95-1.34] | | 1.22 [1.05-1.42] | 550 | 501 | 390 | NA | 522 |
| Q44 | 0.95 [0.74-1.21] | 0.95 [0.75-1.21] | 1.00 [0.76-1.31] | | 0.99 [0.78-1.26] | 0.92 [0.72-1.18] | 0.89 [0.69-1.14] | 0.93 [0.71-1.23] | | 0.96 [0.76-1.23] | 184 | 184 | 148 | NA | 186 |
| Q50 | 1.22 [1.03-1.46] | 1.26 [1.07-1.49] | 1.03 [0.78-1.36] | | 1.12 [0.94-1.34] | 1.15 [0.96-1.38] | 1.16 [0.97-1.38] | 0.94 [0.70-1.26] | | 1.10 [0.92-1.32] | 406 | 425 | 139 | NA | 357 |
| Q51 | 0.98 [0.82-1.17] | 0.97 [0.82-1.15] | 0.99 [0.77-1.27] | | 1.00 [0.84-1.20] | 0.94 [0.78-1.12] | 0.92 [0.77-1.10] | 0.90 [0.69-1.17] | | 0.98 [0.81-1.18] | 366 | 387 | 169 | NA | 328 |
| Q52 | 1.14 [0.86-1.51] | 1.12 [0.83-1.53] | | | 1.06 [0.79-1.42] | 1.06 [0.79-1.42] | 1.11 [0.81-1.53] | | | 1.05 [0.77-1.42] | 155 | 119 | NA | NA | 129 |
| Q53 | 0.87 [0.77-0.99] | | | 0.91 [0.79-1.04] | 0.85 [0.74-0.98] | 0.87 [0.76-0.99] | | | 0.90 [0.79-1.03] | 0.84 [0.73-0.97] | 711 | NA | NA | 680 | 608 |
| Q54 | 1.11 [0.93-1.32] | 1.14 [0.90-1.44] | 1.16 [0.91-1.47] | 1.10 [0.84-1.44] | 0.96 [0.79-1.15] | 1.11 [0.93-1.33] | 1.10 [0.86-1.39] | 1.13 [0.89-1.44] | 1.12 [0.86-1.47] | 0.97 [0.80-1.17] | 363 | 207 | 202 | 168 | 323 |
| Q55 | 1.14 [0.97-1.33] | | | 1.16 [0.98-1.37] | 1.15 [0.97-1.37] | 1.13 [0.96-1.33] | | | 1.15 [0.97-1.36] | 1.13 [0.95-1.35] | 543 | NA | NA | 478 | 453 |
| Q60 | 1.03 [0.86-1.23] | 1.08 [0.91-1.29] | 1.02 [0.84-1.24] | | 1.10 [0.93-1.31] | 0.99 [0.83-1.19] | 1.01 [0.85-1.22] | 0.96 [0.79-1.17] | | 1.10 [0.92-1.31] | 373 | 372 | 304 | NA | 379 |
| Q61 | 1.00 [0.90-1.11] | 1.01 [0.91-1.12] | 1.05 [0.94-1.17] | | 1.01 [0.91-1.12] | 0.97 [0.88-1.08] | 0.96 [0.86-1.06] | 1.01 [0.90-1.13] | | 1.00 [0.90-1.11] | 1,056 | 1,039 | 915 | NA | 987 |
| Q62 | 0.92 [0.69-1.22] | | | | 0.93 [0.70-1.24] | 0.91 [0.68-1.22] | | | | 0.94 [0.70-1.25] | 143 | NA | NA | NA | 139 |
| Q63 | 1.00 [0.88-1.14] | 1.05 [0.92-1.20] | 0.98 [0.83-1.14] | 0.93 [0.68-1.27] | 0.97 [0.85-1.10] | 0.94 [0.82-1.07] | 0.98 [0.85-1.12] | 0.90 [0.76-1.06] | 0.90 [0.65-1.24] | 0.92 [0.80-1.05] | 677 | 628 | 432 | 132 | 642 |
| Q64 | 1.12 [0.81-1.55] | | | | 1.07 [0.76-1.49] | 1.07 [0.77-1.49] | | | | 1.07 [0.76-1.50] | 117 | NA | NA | NA | 105 |
| Q65 | 1.13 [0.96-1.34] | 1.05 [0.87-1.27] | 1.17 [0.90-1.52] | 1.00 [0.76-1.31] | 1.07 [0.91-1.26] | 1.07 [0.90-1.26] | 0.98 [0.81-1.19] | 1.09 [0.83-1.43] | 0.96 [0.73-1.28] | 1.04 [0.88-1.23] | 445 | 330 | 163 | 161 | 420 |
| Q66 | 1.25 [1.06-1.48] | 1.23 [1.02-1.49] | 1.41 [1.14-1.76] | 1.06 [0.79-1.44] | 1.23 [1.03-1.45] | 1.15 [0.97-1.37] | 1.13 [0.93-1.38] | 1.31 [1.04-1.64] | 1.01 [0.75-1.37] | 1.16 [0.98-1.38] | 464 | 336 | 251 | 153 | 432 |
| Q67 | 1.22 [1.00-1.49] | 1.14 [0.87-1.49] | 1.35 [0.98-1.84] | 1.01 [0.76-1.34] | 1.13 [0.92-1.39] | 1.19 [0.97-1.46] | 1.06 [0.81-1.40] | 1.30 [0.95-1.79] | 0.97 [0.72-1.29] | 1.12 [0.91-1.38] | 315 | 169 | 121 | 157 | 293 |
| Q68 | 1.35 [0.96-1.90] | | | | | 1.25 [0.88-1.78] | | | | | 108 | NA | NA | NA | NA |
| Q74 | 1.13 [0.92-1.39] | 1.25 [0.97-1.61] | 1.26 [0.90-1.75] | 1.13 [0.83-1.54] | 1.08 [0.88-1.33] | 1.08 [0.88-1.34] | 1.15 [0.88-1.50] | 1.18 [0.83-1.66] | 1.10 [0.80-1.51] | 1.06 [0.85-1.31] | 295 | 193 | 109 | 138 | 262 |
| Q75 | 0.92 [0.69-1.21] | | | 0.82 [0.61-1.11] | 0.85 [0.64-1.13] | 0.89 [0.67-1.19] | | | 0.81 [0.60-1.09] | 0.84 [0.63-1.12] | 144 | NA | NA | 129 | 138 |
| Q76 | 1.19 [0.99-1.42] | 1.14 [0.94-1.38] | 1.12 [0.90-1.40] | | 1.14 [0.95-1.37] | 1.08 [0.90-1.31] | 1.00 [0.82-1.22] | 0.97 [0.77-1.22] | | 1.07 [0.89-1.30] | 360 | 315 | 223 | NA | 328 |
| Q78 | 1.16 [0.92-1.47] | 1.25 [0.95-1.63] | 1.19 [0.89-1.58] | | 1.18 [0.93-1.50] | 1.09 [0.85-1.39] | 1.11 [0.84-1.47] | 1.06 [0.79-1.42] | | 1.12 [0.88-1.43] | 211 | 165 | 137 | NA | 198 |
| Q79 | 1.26 [1.07-1.48] | 1.23 [1.05-1.46] | 1.04 [0.81-1.32] | 0.97 [0.70-1.35] | 1.14 [0.97-1.34] | 1.05 [0.88-1.25] | 0.97 [0.81-1.16] | 0.83 [0.64-1.08] | 0.90 [0.64-1.27] | 0.98 [0.83-1.16] | 481 | 470 | 181 | 121 | 453 |
| Q82 | 1.45 [1.20-1.75] | 1.64 [1.30-2.07] | 1.65 [1.28-2.13] | 1.06 [0.77-1.46] | 1.39 [1.14-1.69] | 1.36 [1.12-1.65] | 1.49 [1.17-1.90] | 1.51 [1.16-1.96] | 1.03 [0.74-1.43] | 1.32 [1.09-1.61] | 358 | 242 | 196 | 120 | 314 |
| Q83 | 0.96 [0.74-1.25] | 0.99 [0.78-1.26] | | | 0.98 [0.74-1.29] | 0.93 [0.71-1.21] | 0.95 [0.74-1.22] | | | 0.96 [0.72-1.27] | 165 | 190 | NA | NA | 140 |
| Q85 | 0.91 [0.73-1.12] | 0.99 [0.79-1.25] | 1.06 [0.81-1.38] | | 0.98 [0.80-1.21] | 0.88 [0.70-1.09] | 0.93 [0.74-1.18] | 1.01 [0.77-1.33] | | 0.95 [0.77-1.18] | 248 | 207 | 150 | NA | 243 |
| Q87 | 1.16 [0.96-1.40] | 1.31 [1.05-1.64] | 1.36 [1.03-1.80] | 1.07 [0.79-1.44] | 1.12 [0.93-1.35] | 1.07 [0.88-1.30] | 1.19 [0.95-1.51] | 1.25 [0.93-1.67] | 1.05 [0.76-1.43] | 1.08 [0.89-1.31] | 363 | 246 | 152 | 139 | 332 |
| Q89 | 1.06 [0.87-1.29] | 1.18 [0.93-1.49] | 0.98 [0.73-1.31] | 0.92 [0.66-1.27] | 1.09 [0.89-1.33] | 1.02 [0.83-1.25] | 1.08 [0.85-1.38] | 0.94 [0.70-1.26] | 0.89 [0.64-1.25] | 1.06 [0.86-1.31] | 312 | 203 | 127 | 124 | 275 |
| Q90 | 3.11 [2.52-3.84] | 3.08 [2.48-3.82] | 3.71 [2.89-4.75] | | 2.01 [1.54-2.63] | 3.04 [2.45-3.77] | 3.34 [2.66-4.19] | 3.78 [2.93-4.87] | | 1.93 [1.46-2.55] | 412 | 385 | 312 | NA | 190 |
| Q93 | 1.43 [1.07-1.91] | | | 1.20 [0.85-1.68] | 1.44 [1.07-1.93] | 1.31 [0.97-1.76] | | | 1.13 [0.80-1.60] | 1.40 [1.03-1.89] | 161 | NA | NA | 114 | 159 |
| Q99 | 1.02 [0.78-1.32] | | | | 1.14 [0.88-1.49] | 0.94 [0.72-1.24] | | | | 1.10 [0.84-1.45] | 173 | NA | NA | NA | 177 |
| Symptoms, signs and abnormal clinical and laboratory findings, not elsewhere classified (R00-R99) | | | | | | | | | | | | | | | |
| R00 | 1.11 [1.09-1.13] | 1.10 [1.08-1.13] | 1.11 [1.09-1.14] | 0.99 [0.91-1.08] | 1.09 [1.07-1.11] | 1.06 [1.04-1.08] | 1.03 [1.01-1.06] | 1.05 [1.02-1.07] | 0.98 [0.90-1.06] | 1.06 [1.04-1.08] | 30,681 | 30,445 | 25,366 | 2,001 | 29,090 |
| R01 | 1.14 [1.08-1.20] | 1.15 [1.09-1.21] | 1.17 [1.10-1.24] | 1.24 [1.07-1.44] | 1.14 [1.08-1.20] | 1.09 [1.04-1.15] | 1.09 [1.03-1.15] | 1.11 [1.04-1.17] | 1.18 [1.02-1.37] | 1.11 [1.05-1.17] | 4,686 | 4,248 | 3,696 | 640 | 4,483 |
| R02 | 1.14 [1.09-1.20] | 1.14 [1.09-1.20] | 1.16 [1.10-1.21] | | 1.13 [1.08-1.19] | 1.12 [1.07-1.18] | 1.08 [1.02-1.13] | 1.08 [1.03-1.14] | | 1.13 [1.08-1.19] | 4,934 | 4,916 | 4,712 | NA | 4,767 |
| R03 | 1.12 [1.08-1.15] | 1.10 [1.06-1.14] | 1.13 [1.09-1.17] | 1.06 [0.91-1.23] | 1.08 [1.05-1.12] | 1.08 [1.05-1.12] | 1.05 [1.02-1.09] | 1.08 [1.04-1.12] | 1.02 [0.87-1.20] | 1.06 [1.03-1.10] | 10,725 | 10,827 | 8,673 | 547 | 10,140 |
| R04 | 1.23 [1.19-1.27] | 1.23 [1.20-1.27] | 1.24 [1.20-1.28] | 1.13 [0.98-1.30] | 1.20 [1.16-1.23] | 1.17 [1.13-1.20] | 1.15 [1.11-1.19] | 1.15 [1.11-1.19] | 1.12 [0.97-1.29] | 1.15 [1.12-1.19] | 12,489 | 12,004 | 11,284 | 703 | 11,680 |
| R05 | 1.24 [1.20-1.28] | 1.22 [1.18-1.27] | 1.24 [1.19-1.28] | 1.34 [1.23-1.45] | 1.21 [1.17-1.25] | 1.16 [1.12-1.19] | 1.11 [1.07-1.15] | 1.12 [1.08-1.16] | 1.28 [1.18-1.40] | 1.14 [1.11-1.18] | 12,434 | 10,823 | 9,545 | 1,997 | 11,902 |
| R06 | 1.32 [1.30-1.34] | 1.21 [1.19-1.23] | 1.21 [1.19-1.24] | 1.77 [1.71-1.84] | 1.28 [1.26-1.30] | 1.25 [1.23-1.27] | 1.10 [1.08-1.13] | 1.11 [1.08-1.13] | 1.73 [1.66-1.80] | 1.22 [1.20-1.24] | 40,702 | 30,863 | 26,850 | 10,957 | 38,707 |
| R07 | 1.16 [1.15-1.18] | 1.17 [1.15-1.19] | 1.18 [1.16-1.20] | 0.92 [0.85-1.00] | 1.13 [1.11-1.15] | 1.09 [1.08-1.11] | 1.08 [1.06-1.09] | 1.08 [1.07-1.10] | 0.92 [0.84-1.00] | 1.08 [1.06-1.10] | 52,023 | 52,322 | 44,229 | 1,976 | 47,918 |
| R09 | 1.20 [1.14-1.26] | 1.19 [1.13-1.25] | 1.19 [1.13-1.26] | 1.08 [0.86-1.36] | 1.15 [1.09-1.20] | 1.13 [1.07-1.19] | 1.10 [1.04-1.15] | 1.10 [1.04-1.16] | 1.09 [0.85-1.38] | 1.11 [1.05-1.16] | 4,818 | 4,733 | 4,154 | 248 | 4,634 |
| R10 | 1.16 [1.15-1.18] | 1.16 [1.15-1.18] | 1.19 [1.17-1.21] | 1.03 [0.99-1.08] | 1.13 [1.12-1.15] | 1.10 [1.08-1.11] | 1.07 [1.05-1.08] | 1.10 [1.08-1.12] | 1.01 [0.97-1.05] | 1.08 [1.07-1.10] | 64,379 | 63,596 | 39,642 | 8,806 | 57,549 |
| R11 | 1.15 [1.13-1.17] | 1.15 [1.13-1.17] | 1.15 [1.13-1.18] | 1.07 [1.02-1.12] | 1.11 [1.09-1.13] | 1.09 [1.07-1.11] | 1.06 [1.04-1.08] | 1.08 [1.05-1.10] | 1.05 [1.00-1.10] | 1.08 [1.06-1.10] | 38,199 | 35,420 | 28,135 | 5,415 | 35,888 |
| R12 | 1.21 [1.14-1.29] | 1.22 [1.15-1.30] | 1.25 [1.16-1.34] | 0.96 [0.69-1.35] | 1.18 [1.11-1.26] | 1.12 [1.04-1.19] | 1.10 [1.03-1.18] | 1.14 [1.06-1.22] | 0.88 [0.61-1.25] | 1.11 [1.03-1.18] | 2,857 | 2,875 | 2,333 | 116 | 2,654 |
| R13 | 1.20 [1.17-1.23] | 1.20 [1.18-1.23] | 1.20 [1.17-1.23] | 1.14 [0.95-1.37] | 1.17 [1.15-1.20] | 1.13 [1.11-1.16] | 1.11 [1.09-1.14] | 1.12 [1.09-1.14] | 1.05 [0.87-1.28] | 1.13 [1.10-1.16] | 22,523 | 22,408 | 21,197 | 423 | 21,803 |
| R14 | 1.18 [1.13-1.24] | 1.18 [1.12-1.24] | 1.19 [1.13-1.26] | 0.99 [0.80-1.23] | 1.17 [1.12-1.23] | 1.10 [1.05-1.16] | 1.07 [1.01-1.12] | 1.09 [1.03-1.15] | 0.97 [0.78-1.22] | 1.11 [1.06-1.17] | 4,885 | 4,849 | 3,924 | 275 | 4,641 |
| R15 | 1.11 [1.07-1.15] | 1.11 [1.07-1.15] | 1.11 [1.07-1.15] | 0.99 [0.71-1.36] | 1.10 [1.06-1.14] | 1.09 [1.05-1.13] | 1.07 [1.03-1.11] | 1.08 [1.04-1.12] | 0.98 [0.70-1.37] | 1.10 [1.06-1.14] | 9,490 | 9,401 | 9,092 | 131 | 9,238 |
| R16 | 1.30 [1.22-1.38] | 1.35 [1.27-1.43] | 1.32 [1.24-1.41] | 0.95 [0.74-1.21] | 1.22 [1.15-1.29] | 1.23 [1.16-1.31] | 1.24 [1.17-1.32] | 1.22 [1.15-1.31] | 0.91 [0.71-1.16] | 1.19 [1.12-1.26] | 3,350 | 3,290 | 2,897 | 211 | 3,195 |
| R17 | 1.17 [1.09-1.25] | 1.19 [1.11-1.27] | 1.18 [1.10-1.26] | | 1.17 [1.09-1.25] | 1.15 [1.07-1.23] | 1.14 [1.07-1.22] | 1.14 [1.06-1.22] | | 1.17 [1.09-1.25] | 2,578 | 2,562 | 2,356 | NA | 2,370 |
| R18 | 1.16 [1.12-1.21] | 1.17 [1.12-1.22] | 1.17 [1.13-1.23] | 0.85 [0.61-1.19] | 1.15 [1.11-1.20] | 1.14 [1.10-1.19] | 1.13 [1.08-1.18] | 1.14 [1.09-1.18] | 0.87 [0.62-1.22] | 1.16 [1.11-1.21] | 6,807 | 6,790 | 6,356 | 121 | 6,439 |
| R19 | 1.22 [1.19-1.24] | 1.21 [1.19-1.24] | 1.20 [1.18-1.23] | 1.02 [0.91-1.14] | 1.19 [1.16-1.21] | 1.14 [1.12-1.17] | 1.12 [1.09-1.14] | 1.12 [1.09-1.14] | 0.99 [0.88-1.11] | 1.13 [1.11-1.16] | 27,081 | 27,030 | 23,324 | 1,097 | 25,303 |
| R20 | 1.16 [1.11-1.21] | 1.16 [1.11-1.20] | 1.14 [1.09-1.20] | 1.03 [0.87-1.23] | 1.12 [1.07-1.17] | 1.07 [1.03-1.12] | 1.04 [1.00-1.09] | 1.03 [0.98-1.08] | 1.03 [0.86-1.24] | 1.06 [1.02-1.11] | 6,862 | 6,943 | 5,153 | 450 | 6,589 |
| R21 | 1.53 [1.48-1.58] | 1.60 [1.54-1.66] | 1.70 [1.63-1.77] | 1.16 [1.08-1.25] | 1.46 [1.41-1.51] | 1.47 [1.41-1.52] | 1.50 [1.44-1.56] | 1.59 [1.53-1.66] | 1.14 [1.06-1.23] | 1.42 [1.37-1.47] | 10,581 | 8,610 | 7,270 | 2,391 | 9,807 |
| R22 | 1.29 [1.23-1.35] | 1.25 [1.19-1.31] | 1.29 [1.23-1.37] | 1.43 [1.23-1.67] | 1.25 [1.19-1.31] | 1.22 [1.16-1.28] | 1.15 [1.09-1.21] | 1.20 [1.14-1.27] | 1.40 [1.20-1.63] | 1.19 [1.13-1.25] | 5,293 | 4,889 | 4,066 | 625 | 4,884 |
| R23 | 1.20 [1.14-1.27] | 1.21 [1.14-1.30] | 1.23 [1.15-1.31] | 1.17 [1.05-1.32] | 1.20 [1.14-1.28] | 1.16 [1.09-1.23] | 1.14 [1.06-1.22] | 1.15 [1.07-1.23] | 1.13 [1.00-1.27] | 1.17 [1.10-1.24] | 3,748 | 2,862 | 2,563 | 996 | 3,607 |
| R25 | 1.14 [1.08-1.20] | 1.16 [1.10-1.22] | 1.15 [1.09-1.22] | 1.01 [0.83-1.22] | 1.12 [1.06-1.18] | 1.07 [1.02-1.13] | 1.06 [1.00-1.12] | 1.06 [1.00-1.12] | 0.98 [0.80-1.19] | 1.08 [1.02-1.14] | 4,131 | 3,892 | 3,457 | 341 | 4,004 |
| R26 | 1.14 [1.12-1.16] | 1.13 [1.11-1.15] | 1.13 [1.11-1.16] | 1.01 [0.88-1.16] | 1.12 [1.10-1.14] | 1.11 [1.09-1.13] | 1.08 [1.06-1.10] | 1.08 [1.06-1.10] | 1.00 [0.86-1.15] | 1.11 [1.08-1.13] | 36,004 | 35,529 | 35,015 | 644 | 35,245 |
| R27 | 1.09 [0.98-1.21] | 1.09 [0.98-1.21] | 1.10 [0.98-1.23] | 1.01 [0.74-1.39] | 1.05 [0.94-1.16] | 1.05 [0.95-1.17] | 1.02 [0.92-1.14] | 1.03 [0.92-1.16] | 0.99 [0.71-1.37] | 1.03 [0.93-1.15] | 1,100 | 985 | 894 | 138 | 1,029 |
| R29 | 1.12 [1.10-1.14] | 1.12 [1.10-1.14] | 1.12 [1.11-1.14] | 0.86 [0.72-1.03] | 1.11 [1.09-1.13] | 1.09 [1.07-1.11] | 1.06 [1.05-1.08] | 1.07 [1.05-1.09] | 0.86 [0.72-1.03] | 1.10 [1.08-1.11] | 47,875 | 47,735 | 46,731 | 428 | 47,120 |
| R30 | 1.26 [1.19-1.34] | 1.20 [1.13-1.28] | 1.23 [1.14-1.31] | 1.09 [0.90-1.32] | 1.20 [1.13-1.27] | 1.18 [1.11-1.25] | 1.09 [1.02-1.16] | 1.10 [1.02-1.18] | 1.08 [0.89-1.32] | 1.14 [1.07-1.21] | 3,306 | 3,194 | 2,419 | 366 | 3,154 |
| R31 | 1.10 [1.07-1.12] | 1.10 [1.07-1.12] | 1.09 [1.06-1.11] | 1.09 [0.93-1.28] | 1.06 [1.03-1.08] | 1.07 [1.05-1.10] | 1.06 [1.03-1.08] | 1.05 [1.02-1.07] | 1.07 [0.90-1.25] | 1.04 [1.02-1.07] | 22,024 | 21,931 | 20,276 | 540 | 19,990 |
| R32 | 1.10 [1.08-1.13] | 1.10 [1.07-1.13] | 1.09 [1.06-1.12] | 1.10 [0.91-1.33] | 1.08 [1.05-1.11] | 1.08 [1.05-1.11] | 1.05 [1.03-1.08] | 1.05 [1.02-1.08] | 1.09 [0.89-1.33] | 1.07 [1.05-1.10] | 17,893 | 17,683 | 16,966 | 401 | 17,449 |
| R33 | 1.10 [1.08-1.12] | 1.09 [1.07-1.11] | 1.10 [1.07-1.12] | 1.00 [0.85-1.17] | 1.09 [1.07-1.11] | 1.07 [1.05-1.10] | 1.05 [1.03-1.07] | 1.06 [1.04-1.08] | 0.97 [0.82-1.15] | 1.08 [1.06-1.10] | 33,293 | 33,293 | 31,650 | 526 | 31,859 |
| R34 | 1.24 [1.11-1.39] | 1.23 [1.09-1.38] | 1.22 [1.09-1.38] | | 1.25 [1.11-1.40] | 1.19 [1.06-1.33] | 1.14 [1.01-1.29] | 1.14 [1.01-1.29] | | 1.23 [1.10-1.38] | 932 | 910 | 864 | NA | 886 |
| R35 | 1.25 [1.20-1.31] | 1.22 [1.17-1.28] | 1.21 [1.16-1.28] | 1.01 [0.81-1.27] | 1.18 [1.13-1.24] | 1.18 [1.13-1.24] | 1.13 [1.08-1.19] | 1.13 [1.08-1.19] | 0.99 [0.78-1.25] | 1.13 [1.08-1.18] | 5,488 | 5,424 | 4,643 | 250 | 5,141 |
| R36 | 1.30 [0.94-1.80] | 1.42 [1.01-1.99] | 1.41 [1.00-1.99] | | 1.34 [0.97-1.85] | 1.31 [0.94-1.83] | 1.36 [0.97-1.92] | 1.26 [0.88-1.80] | | 1.32 [0.95-1.83] | 121 | 114 | 109 | NA | 114 |
| R39 | 1.20 [1.15-1.26] | 1.21 [1.16-1.26] | 1.20 [1.15-1.26] | 1.02 [0.81-1.27] | 1.15 [1.10-1.20] | 1.13 [1.08-1.18] | 1.12 [1.07-1.17] | 1.12 [1.06-1.17] | 0.98 [0.78-1.24] | 1.10 [1.05-1.15] | 6,208 | 6,194 | 5,460 | 255 | 5,844 |
| R40 | 1.13 [1.07-1.18] | 1.13 [1.08-1.19] | 1.14 [1.08-1.20] | 1.00 [0.81-1.24] | 1.11 [1.06-1.17] | 1.09 [1.04-1.15] | 1.06 [1.01-1.12] | 1.07 [1.02-1.13] | 1.00 [0.80-1.25] | 1.09 [1.04-1.15] | 5,149 | 4,996 | 4,648 | 281 | 5,070 |
| R41 | 1.12 [1.10-1.14] | 1.12 [1.10-1.15] | 1.11 [1.09-1.13] | 0.94 [0.78-1.13] | 1.11 [1.09-1.13] | 1.10 [1.08-1.12] | 1.08 [1.06-1.10] | 1.07 [1.05-1.09] | 0.94 [0.78-1.15] | 1.10 [1.08-1.13] | 32,986 | 32,816 | 32,286 | 373 | 31,660 |
| R42 | 1.14 [1.11-1.17] | 1.14 [1.11-1.17] | 1.13 [1.10-1.16] | 0.95 [0.83-1.09] | 1.11 [1.08-1.14] | 1.08 [1.06-1.11] | 1.06 [1.03-1.08] | 1.06 [1.03-1.09] | 0.95 [0.82-1.10] | 1.07 [1.04-1.10] | 17,602 | 17,551 | 15,276 | 708 | 16,910 |
| R43 | 1.11 [0.88-1.39] | 1.05 [0.83-1.32] | 1.19 [0.92-1.54] | | 1.13 [0.89-1.42] | 1.03 [0.81-1.31] | 0.96 [0.75-1.22] | 1.06 [0.81-1.38] | | 1.08 [0.85-1.37] | 220 | 215 | 177 | NA | 208 |
| R44 | 1.14 [1.09-1.21] | 1.13 [1.08-1.20] | 1.15 [1.09-1.22] | 0.96 [0.75-1.22] | 1.10 [1.05-1.16] | 1.11 [1.05-1.17] | 1.07 [1.01-1.13] | 1.09 [1.03-1.15] | 0.97 [0.76-1.25] | 1.09 [1.03-1.15] | 4,448 | 4,331 | 3,971 | 219 | 4,241 |
| R45 | 1.13 [1.09-1.18] | 1.12 [1.08-1.16] | 1.15 [1.10-1.20] | 0.96 [0.86-1.08] | 1.10 [1.06-1.15] | 1.09 [1.05-1.13] | 1.03 [0.99-1.07] | 1.07 [1.02-1.11] | 1.00 [0.88-1.13] | 1.09 [1.05-1.14] | 8,788 | 8,371 | 6,945 | 1,012 | 8,519 |
| R46 | 1.14 [1.06-1.21] | 1.10 [1.03-1.18] | 1.09 [1.02-1.17] | 1.07 [0.81-1.41] | 1.13 [1.05-1.20] | 1.13 [1.05-1.20] | 1.06 [0.99-1.14] | 1.06 [0.98-1.13] | 1.03 [0.77-1.37] | 1.15 [1.07-1.23] | 2,727 | 2,567 | 2,411 | 188 | 2,636 |
| R47 | 1.09 [1.05-1.13] | 1.09 [1.05-1.13] | 1.08 [1.04-1.12] | 1.11 [0.85-1.45] | 1.07 [1.04-1.11] | 1.07 [1.03-1.11] | 1.04 [1.00-1.08] | 1.03 [0.99-1.07] | 1.15 [0.87-1.52] | 1.07 [1.03-1.11] | 8,917 | 8,804 | 8,409 | 185 | 8,525 |
| R48 | 1.28 [1.03-1.59] | 1.18 [0.94-1.47] | 1.28 [0.95-1.71] | | 1.29 [1.04-1.60] | 1.15 [0.92-1.45] | 1.05 [0.83-1.32] | 1.17 [0.86-1.58] | | 1.22 [0.98-1.53] | 281 | 256 | 138 | NA | 260 |
| R49 | 1.19 [1.10-1.30] | 1.23 [1.13-1.34] | 1.18 [1.08-1.29] | | 1.19 [1.10-1.30] | 1.11 [1.01-1.20] | 1.09 [1.00-1.19] | 1.05 [0.96-1.15] | | 1.11 [1.02-1.21] | 1,663 | 1,626 | 1,483 | NA | 1,584 |
| R50 | 1.18 [1.14-1.21] | 1.19 [1.15-1.23] | 1.19 [1.15-1.23] | 1.13 [1.07-1.20] | 1.15 [1.12-1.19] | 1.13 [1.10-1.16] | 1.12 [1.08-1.15] | 1.13 [1.09-1.17] | 1.11 [1.04-1.18] | 1.13 [1.09-1.16] | 14,839 | 11,749 | 9,530 | 3,766 | 13,835 |
| R51 | 1.17 [1.14-1.20] | 1.17 [1.14-1.20] | 1.19 [1.16-1.22] | 1.06 [0.99-1.13] | 1.13 [1.10-1.16] | 1.09 [1.06-1.12] | 1.05 [1.03-1.08] | 1.08 [1.05-1.11] | 1.04 [0.97-1.12] | 1.08 [1.05-1.10] | 22,028 | 21,466 | 14,228 | 2,813 | 20,771 |
| R52 | 1.25 [1.19-1.32] | 1.24 [1.17-1.30] | 1.24 [1.17-1.31] | 0.85 [0.64-1.13] | 1.21 [1.15-1.27] | 1.15 [1.09-1.21] | 1.09 [1.03-1.15] | 1.11 [1.04-1.17] | 0.83 [0.62-1.13] | 1.14 [1.08-1.20] | 4,190 | 4,190 | 3,532 | 160 | 4,153 |
| R53 | 1.13 [1.10-1.16] | 1.13 [1.10-1.16] | 1.13 [1.10-1.16] | 1.08 [0.95-1.21] | 1.11 [1.08-1.14] | 1.09 [1.06-1.12] | 1.06 [1.03-1.09] | 1.06 [1.03-1.09] | 1.05 [0.93-1.19] | 1.09 [1.06-1.12] | 16,722 | 16,192 | 14,781 | 916 | 16,261 |
| R54 | 1.11 [1.09-1.14] | 1.12 [1.09-1.14] | 1.11 [1.09-1.14] | | 1.10 [1.08-1.13] | 1.10 [1.07-1.12] | 1.08 [1.05-1.10] | 1.07 [1.05-1.10] | | 1.10 [1.07-1.13] | 22,174 | 22,166 | 22,113 | NA | 21,393 |
| R55 | 1.12 [1.10-1.14] | 1.12 [1.10-1.14] | 1.13 [1.11-1.15] | 0.93 [0.84-1.04] | 1.11 [1.09-1.13] | 1.09 [1.07-1.11] | 1.07 [1.05-1.09] | 1.08 [1.06-1.10] | 0.91 [0.82-1.02] | 1.09 [1.07-1.11] | 31,010 | 30,710 | 28,043 | 1,171 | 29,017 |
| R56 | 1.12 [1.09-1.16] | 1.16 [1.12-1.21] | 1.17 [1.12-1.21] | 1.01 [0.95-1.08] | 1.09 [1.05-1.12] | 1.10 [1.06-1.13] | 1.10 [1.06-1.14] | 1.11 [1.06-1.16] | 1.00 [0.93-1.07] | 1.08 [1.05-1.12] | 10,570 | 8,105 | 6,985 | 2,794 | 9,840 |
| R57 | 1.10 [1.03-1.17] | 1.11 [1.04-1.19] | 1.11 [1.04-1.19] | | 1.12 [1.05-1.20] | 1.08 [1.01-1.16] | 1.07 [1.00-1.14] | 1.06 [1.00-1.14] | | 1.13 [1.06-1.20] | 2,785 | 2,763 | 2,680 | NA | 2,712 |
| R58 | 1.29 [1.10-1.53] | 1.22 [1.03-1.44] | 1.22 [1.03-1.45] | | 1.17 [1.00-1.38] | 1.23 [1.04-1.46] | 1.14 [0.96-1.35] | 1.15 [0.97-1.37] | | 1.12 [0.95-1.32] | 450 | 442 | 419 | NA | 433 |
| R59 | 1.26 [1.21-1.31] | 1.25 [1.21-1.30] | 1.26 [1.21-1.31] | 1.14 [1.00-1.29] | 1.22 [1.18-1.27] | 1.24 [1.19-1.28] | 1.20 [1.15-1.25] | 1.21 [1.16-1.26] | 1.13 [0.99-1.28] | 1.21 [1.17-1.26] | 8,395 | 7,815 | 6,886 | 863 | 7,786 |
| R60 | 1.24 [1.21-1.28] | 1.24 [1.20-1.27] | 1.25 [1.21-1.28] | 0.97 [0.71-1.33] | 1.23 [1.19-1.26] | 1.19 [1.16-1.22] | 1.16 [1.13-1.19] | 1.17 [1.14-1.20] | 0.96 [0.69-1.33] | 1.19 [1.16-1.22] | 16,685 | 16,642 | 16,239 | 132 | 16,254 |
| R61 | 1.25 [1.14-1.36] | 1.25 [1.14-1.36] | 1.18 [1.07-1.30] | 1.16 [0.82-1.64] | 1.19 [1.09-1.29] | 1.15 [1.05-1.26] | 1.14 [1.04-1.25] | 1.07 [0.97-1.19] | 1.06 [0.74-1.52] | 1.13 [1.03-1.24] | 1,558 | 1,567 | 1,177 | 116 | 1,457 |
| R62 | 1.10 [0.99-1.21] | 1.06 [0.86-1.29] | 1.18 [0.91-1.52] | 1.09 [0.97-1.21] | 1.05 [0.95-1.17] | 1.06 [0.96-1.18] | 1.02 [0.83-1.25] | 1.12 [0.86-1.45] | 1.04 [0.93-1.17] | 1.03 [0.93-1.14] | 1,316 | 298 | 179 | 1,034 | 1,224 |
| R63 | 1.16 [1.14-1.18] | 1.15 [1.13-1.18] | 1.16 [1.14-1.19] | 1.03 [0.94-1.12] | 1.15 [1.13-1.17] | 1.12 [1.09-1.14] | 1.08 [1.06-1.11] | 1.10 [1.07-1.12] | 0.99 [0.91-1.09] | 1.12 [1.10-1.14] | 30,767 | 29,783 | 27,511 | 1,713 | 29,257 |
| R64 | 1.14 [1.01-1.29] | 1.11 [0.99-1.26] | 1.08 [0.96-1.22] | | 1.06 [0.94-1.20] | 1.14 [1.01-1.29] | 1.09 [0.96-1.24] | 1.08 [0.95-1.22] | | 1.09 [0.96-1.23] | 794 | 795 | 786 | NA | 767 |
| R65 | 1.13 [1.05-1.23] | 1.11 [1.02-1.20] | 1.14 [1.05-1.23] | | 1.14 [1.06-1.23] | 1.09 [1.01-1.18] | 1.05 [0.97-1.14] | 1.06 [0.98-1.15] | | 1.12 [1.03-1.21] | 1,988 | 1,968 | 1,911 | NA | 1,966 |
| R68 | 1.19 [1.12-1.27] | 1.16 [1.09-1.24] | 1.17 [1.09-1.25] | 1.20 [0.93-1.55] | 1.18 [1.11-1.26] | 1.18 [1.11-1.26] | 1.13 [1.05-1.20] | 1.13 [1.05-1.21] | 1.14 [0.88-1.49] | 1.18 [1.11-1.26] | 3,069 | 2,919 | 2,766 | 189 | 2,985 |
| R69 | 1.15 [1.13-1.18] | 1.14 [1.12-1.17] | 1.15 [1.12-1.18] | 1.01 [0.91-1.13] | 1.13 [1.11-1.16] | 1.12 [1.10-1.15] | 1.09 [1.06-1.11] | 1.10 [1.07-1.13] | 1.00 [0.89-1.12] | 1.11 [1.09-1.14] | 24,225 | 23,904 | 20,373 | 1,101 | 21,557 |
| R70 | 1.33 [1.07-1.65] | 1.31 [1.04-1.63] | 1.29 [1.03-1.61] | | 1.22 [0.97-1.54] | 1.28 [1.03-1.60] | 1.22 [0.97-1.54] | 1.21 [0.96-1.53] | | 1.18 [0.93-1.49] | 239 | 236 | 221 | NA | 218 |
| R71 | 1.10 [0.85-1.42] | 1.01 [0.78-1.31] | 1.13 [0.86-1.49] | | 0.99 [0.77-1.29] | 1.08 [0.83-1.41] | 0.98 [0.75-1.28] | 1.10 [0.83-1.45] | | 1.00 [0.77-1.31] | 162 | 162 | 149 | NA | 152 |
| R72 | 1.12 [1.01-1.26] | 1.10 [0.99-1.23] | 1.10 [0.98-1.24] | | 1.09 [0.98-1.22] | 1.10 [0.98-1.23] | 1.04 [0.93-1.17] | 1.07 [0.94-1.20] | | 1.07 [0.96-1.20] | 954 | 949 | 803 | NA | 912 |
| R73 | 1.16 [1.10-1.23] | 1.16 [1.10-1.23] | 1.17 [1.11-1.24] | 0.86 [0.60-1.23] | 1.17 [1.10-1.23] | 1.10 [1.04-1.16] | 1.08 [1.02-1.14] | 1.07 [1.01-1.14] | 0.84 [0.58-1.22] | 1.12 [1.06-1.19] | 4,007 | 3,975 | 3,673 | 102 | 3,905 |
| R74 | 1.12 [1.02-1.22] | 1.08 [0.99-1.18] | 1.12 [1.03-1.23] | 1.15 [0.81-1.63] | 1.08 [0.99-1.18] | 1.07 [0.98-1.17] | 1.00 [0.92-1.10] | 1.05 [0.96-1.15] | 1.09 [0.75-1.58] | 1.06 [0.98-1.16] | 1,650 | 1,608 | 1,431 | 111 | 1,580 |
| R76 | 1.28 [1.09-1.51] | 1.28 [1.09-1.50] | 1.30 [1.08-1.56] | | 1.27 [1.08-1.50] | 1.25 [1.06-1.47] | 1.23 [1.04-1.45] | 1.25 [1.04-1.50] | | 1.23 [1.04-1.45] | 472 | 450 | 366 | NA | 439 |
| R77 | 1.22 [1.10-1.36] | 1.21 [1.09-1.34] | 1.25 [1.12-1.40] | | 1.21 [1.09-1.34] | 1.16 [1.05-1.30] | 1.13 [1.01-1.26] | 1.18 [1.05-1.32] | | 1.17 [1.05-1.30] | 1,135 | 1,102 | 997 | NA | 1,085 |
| R78 | 1.29 [0.97-1.71] | 1.19 [0.90-1.59] | 1.04 [0.78-1.40] | | 1.05 [0.79-1.39] | 1.28 [0.96-1.71] | 1.15 [0.86-1.54] | 1.00 [0.74-1.35] | | 1.06 [0.79-1.43] | 145 | 141 | 128 | NA | 125 |
| R79 | 1.13 [1.11-1.16] | 1.12 [1.10-1.15] | 1.14 [1.11-1.16] | 1.07 [0.89-1.28] | 1.12 [1.09-1.15] | 1.09 [1.07-1.12] | 1.06 [1.04-1.09] | 1.07 [1.05-1.10] | 1.07 [0.89-1.29] | 1.09 [1.07-1.12] | 21,522 | 21,380 | 20,454 | 415 | 20,755 |
| R80 | 1.19 [1.06-1.35] | 1.22 [1.08-1.38] | 1.24 [1.08-1.42] | | 1.18 [1.05-1.34] | 1.17 [1.03-1.32] | 1.17 [1.03-1.32] | 1.18 [1.02-1.35] | | 1.17 [1.03-1.32] | 800 | 758 | 622 | NA | 750 |
| R81 | 1.01 [0.79-1.29] | 1.04 [0.81-1.33] | | | 1.18 [0.91-1.54] | 0.98 [0.76-1.26] | 1.01 [0.79-1.30] | | | 1.19 [0.91-1.56] | 182 | 176 | NA | NA | 154 |
| R82 | 1.09 [1.00-1.19] | 1.09 [1.00-1.19] | 1.06 [0.96-1.16] | 1.00 [0.75-1.34] | 1.06 [0.97-1.15] | 1.05 [0.96-1.15] | 1.02 [0.93-1.11] | 0.99 [0.89-1.09] | 1.01 [0.74-1.38] | 1.04 [0.96-1.14] | 1,597 | 1,574 | 1,223 | 150 | 1,580 |
| R84 | 1.23 [1.06-1.43] | 1.22 [1.05-1.43] | 1.31 [1.12-1.54] | | 1.14 [0.99-1.32] | 1.13 [0.97-1.32] | 1.08 [0.92-1.26] | 1.18 [1.00-1.39] | | 1.05 [0.90-1.22] | 544 | 490 | 467 | NA | 530 |
| R85 | 1.25 [1.02-1.53] | 1.25 [1.02-1.54] | 1.18 [0.94-1.47] | | 1.23 [1.00-1.51] | 1.17 [0.95-1.44] | 1.17 [0.95-1.45] | 1.08 [0.86-1.36] | | 1.15 [0.94-1.42] | 290 | 281 | 231 | NA | 276 |
| R87 | 1.06 [0.96-1.16] | 1.08 [0.98-1.18] | 0.98 [0.85-1.14] | | 0.97 [0.88-1.08] | 1.02 [0.93-1.13] | 1.03 [0.94-1.14] | 0.95 [0.81-1.10] | | 0.97 [0.87-1.08] | 1,293 | 1,332 | 464 | NA | 982 |
| R89 | 1.31 [1.07-1.61] | 1.39 [1.13-1.72] | 1.41 [1.13-1.75] | | 1.31 [1.07-1.61] | 1.28 [1.04-1.58] | 1.31 [1.06-1.63] | 1.33 [1.07-1.67] | | 1.29 [1.05-1.59] | 295 | 276 | 256 | NA | 289 |
| R90 | 1.14 [1.01-1.28] | 1.10 [0.98-1.24] | 1.15 [1.01-1.30] | | 1.19 [1.06-1.34] | 1.10 [0.98-1.24] | 1.05 [0.93-1.18] | 1.10 [0.96-1.24] | | 1.17 [1.04-1.32] | 832 | 809 | 712 | NA | 793 |
| R91 | 1.18 [1.14-1.22] | 1.17 [1.14-1.21] | 1.19 [1.15-1.23] | 1.17 [0.82-1.66] | 1.17 [1.13-1.20] | 1.14 [1.10-1.18] | 1.10 [1.06-1.14] | 1.12 [1.08-1.16] | 1.19 [0.82-1.72] | 1.14 [1.10-1.18] | 10,382 | 10,345 | 10,063 | 108 | 9,895 |
| R93 | 1.15 [1.11-1.19] | 1.13 [1.09-1.16] | 1.14 [1.10-1.18] | 1.07 [0.85-1.34] | 1.11 [1.08-1.15] | 1.11 [1.07-1.15] | 1.06 [1.03-1.10] | 1.08 [1.04-1.12] | 1.05 [0.83-1.33] | 1.09 [1.06-1.13] | 9,957 | 9,914 | 9,169 | 257 | 9,482 |
| R94 | 1.18 [1.15-1.21] | 1.18 [1.15-1.21] | 1.17 [1.14-1.21] | 0.98 [0.84-1.14] | 1.15 [1.12-1.18] | 1.14 [1.12-1.18] | 1.11 [1.08-1.14] | 1.11 [1.09-1.15] | 0.96 [0.82-1.13] | 1.13 [1.10-1.16] | 18,296 | 18,265 | 16,589 | 564 | 17,461 |
| Injury, poisoning and certain other consequences of external causes (S00-T88) | | | | | | | | | | | | | | | |
| S00 | 1.12 [1.09-1.15] | 1.13 [1.10-1.17] | 1.14 [1.11-1.18] | 0.95 [0.86-1.04] | 1.11 [1.08-1.14] | 1.09 [1.06-1.12] | 1.09 [1.06-1.12] | 1.09 [1.06-1.12] | 0.95 [0.86-1.04] | 1.09 [1.06-1.12] | 17,330 | 16,214 | 15,353 | 1,393 | 16,648 |
| S01 | 1.13 [1.10-1.17] | 1.13 [1.10-1.17] | 1.13 [1.10-1.17] | 0.98 [0.90-1.06] | 1.12 [1.09-1.15] | 1.11 [1.08-1.14] | 1.09 [1.06-1.12] | 1.09 [1.06-1.12] | 0.97 [0.90-1.06] | 1.11 [1.08-1.14] | 17,034 | 15,476 | 14,575 | 1,875 | 16,101 |
| S02 | 1.09 [1.04-1.15] | 1.11 [1.05-1.18] | 1.16 [1.09-1.24] | 0.97 [0.85-1.10] | 1.11 [1.05-1.17] | 1.07 [1.01-1.13] | 1.07 [1.01-1.13] | 1.12 [1.05-1.19] | 0.97 [0.85-1.11] | 1.09 [1.03-1.15] | 4,481 | 4,079 | 3,073 | 826 | 4,016 |
| S03 | 1.08 [0.83-1.41] | 1.16 [0.83-1.62] | | | 1.07 [0.81-1.41] | 1.01 [0.77-1.33] | 1.08 [0.77-1.52] | | | 1.06 [0.80-1.40] | 178 | 112 | NA | NA | 156 |
| S05 | 1.10 [1.01-1.21] | 1.11 [1.01-1.22] | 1.15 [1.05-1.27] | | 1.15 [1.05-1.26] | 1.08 [0.99-1.19] | 1.06 [0.96-1.17] | 1.11 [1.00-1.22] | | 1.15 [1.05-1.26] | 1,394 | 1,339 | 1,256 | NA | 1,329 |
| S06 | 1.06 [1.01-1.11] | 1.08 [1.02-1.13] | 1.08 [1.02-1.13] | 1.06 [0.86-1.32] | 1.04 [0.99-1.10] | 1.04 [0.99-1.10] | 1.04 [0.99-1.10] | 1.04 [0.99-1.10] | 1.07 [0.86-1.34] | 1.04 [0.99-1.09] | 4,809 | 4,639 | 4,322 | 291 | 4,469 |
| S09 | 1.13 [1.09-1.17] | 1.15 [1.10-1.19] | 1.18 [1.13-1.23] | 0.96 [0.87-1.06] | 1.12 [1.08-1.16] | 1.09 [1.05-1.13] | 1.07 [1.03-1.12] | 1.11 [1.06-1.16] | 0.94 [0.84-1.04] | 1.10 [1.05-1.14] | 8,595 | 7,571 | 6,909 | 1,248 | 8,073 |
| S10 | 1.06 [0.89-1.25] | 1.15 [0.96-1.37] | 1.07 [0.88-1.31] | | 0.92 [0.77-1.09] | 1.01 [0.85-1.20] | 1.06 [0.89-1.27] | 1.00 [0.82-1.23] | | 0.91 [0.76-1.08] | 404 | 380 | 299 | NA | 379 |
| S11 | 1.17 [0.92-1.49] | 1.17 [0.91-1.49] | 1.26 [0.93-1.70] | | 1.16 [0.91-1.48] | 1.08 [0.85-1.39] | 1.10 [0.85-1.41] | 1.20 [0.88-1.63] | | 1.18 [0.92-1.51] | 207 | 202 | 147 | NA | 204 |
| S12 | 1.15 [1.04-1.26] | 1.18 [1.07-1.30] | 1.12 [1.01-1.23] | | 1.15 [1.04-1.26] | 1.12 [1.01-1.24] | 1.12 [1.02-1.24] | 1.06 [0.96-1.18] | | 1.13 [1.03-1.25] | 1,299 | 1,294 | 1,234 | NA | 1,242 |
| S13 | 1.19 [0.96-1.47] | 1.13 [0.91-1.40] | 1.08 [0.84-1.39] | | 1.05 [0.85-1.30] | 1.12 [0.90-1.40] | 1.05 [0.84-1.30] | 1.02 [0.79-1.32] | | 1.02 [0.82-1.27] | 260 | 258 | 185 | NA | 236 |
| S14 | 0.96 [0.76-1.23] | 0.92 [0.72-1.17] | 1.02 [0.80-1.31] | | 0.93 [0.72-1.19] | 0.98 [0.76-1.25] | 0.91 [0.71-1.16] | 1.03 [0.80-1.32] | | 0.92 [0.71-1.19] | 187 | 185 | 171 | NA | 165 |
| S19 | 1.14 [0.95-1.37] | 1.27 [1.05-1.53] | 1.18 [0.95-1.46] | | 1.20 [1.00-1.45] | 1.06 [0.88-1.28] | 1.13 [0.93-1.38] | 1.06 [0.85-1.32] | | 1.14 [0.94-1.38] | 356 | 335 | 252 | NA | 339 |
| S20 | 1.18 [1.09-1.27] | 1.16 [1.07-1.25] | 1.14 [1.05-1.24] | | 1.14 [1.05-1.24] | 1.13 [1.04-1.22] | 1.07 [0.99-1.16] | 1.06 [0.97-1.15] | | 1.11 [1.02-1.20] | 1,930 | 1,889 | 1,767 | NA | 1,858 |
| S21 | 1.24 [0.99-1.55] | 1.19 [0.95-1.49] | 1.36 [1.05-1.76] | | 1.20 [0.95-1.50] | 1.22 [0.97-1.54] | 1.10 [0.87-1.39] | 1.29 [0.99-1.68] | | 1.21 [0.96-1.53] | 261 | 249 | 191 | NA | 236 |
| S22 | 1.17 [1.12-1.22] | 1.15 [1.10-1.20] | 1.16 [1.11-1.21] | 1.01 [0.72-1.42] | 1.13 [1.08-1.18] | 1.13 [1.08-1.18] | 1.07 [1.03-1.12] | 1.08 [1.03-1.13] | 1.03 [0.72-1.47] | 1.10 [1.05-1.15] | 6,461 | 6,410 | 6,121 | 111 | 6,132 |
| S27 | 1.11 [1.02-1.22] | 1.09 [1.00-1.19] | 1.13 [1.03-1.24] | | 1.05 [0.96-1.15] | 1.09 [1.00-1.19] | 1.05 [0.96-1.15] | 1.09 [0.99-1.19] | | 1.05 [0.96-1.15] | 1,547 | 1,499 | 1,347 | NA | 1,387 |
| S29 | 1.37 [1.18-1.59] | 1.40 [1.21-1.63] | 1.38 [1.18-1.62] | | 1.29 [1.11-1.50] | 1.29 [1.10-1.50] | 1.29 [1.11-1.51] | 1.26 [1.08-1.49] | | 1.22 [1.05-1.43] | 557 | 551 | 514 | NA | 534 |
| S30 | 1.19 [1.12-1.26] | 1.17 [1.10-1.25] | 1.22 [1.14-1.30] | 0.93 [0.74-1.16] | 1.15 [1.08-1.22] | 1.14 [1.07-1.21] | 1.09 [1.02-1.16] | 1.14 [1.07-1.22] | 0.94 [0.75-1.17] | 1.12 [1.05-1.19] | 3,322 | 3,159 | 2,858 | 267 | 3,236 |
| S31 | 1.11 [0.97-1.27] | 1.15 [0.99-1.33] | 1.31 [1.10-1.57] | 1.02 [0.78-1.34] | 1.16 [1.01-1.33] | 1.08 [0.94-1.24] | 1.05 [0.90-1.22] | 1.20 [1.00-1.44] | 1.05 [0.79-1.40] | 1.14 [0.99-1.32] | 684 | 565 | 400 | 183 | 619 |
| S32 | 1.10 [1.06-1.15] | 1.10 [1.05-1.14] | 1.12 [1.08-1.17] | 0.88 [0.64-1.22] | 1.08 [1.04-1.13] | 1.07 [1.03-1.12] | 1.05 [1.01-1.10] | 1.08 [1.03-1.13] | 0.97 [0.69-1.36] | 1.06 [1.02-1.11] | 7,154 | 7,141 | 6,881 | 123 | 6,811 |
| S36 | 1.09 [0.94-1.26] | 1.14 [0.98-1.32] | 1.08 [0.92-1.28] | | 1.04 [0.90-1.21] | 1.08 [0.94-1.25] | 1.10 [0.95-1.28] | 1.05 [0.89-1.25] | | 1.05 [0.90-1.22] | 564 | 534 | 406 | NA | 494 |
| S37 | 0.94 [0.81-1.10] | 1.01 [0.86-1.19] | 0.97 [0.81-1.15] | | 0.95 [0.81-1.12] | 0.93 [0.79-1.09] | 0.99 [0.84-1.17] | 0.94 [0.79-1.13] | | 0.95 [0.81-1.12] | 467 | 449 | 383 | NA | 438 |
| S39 | 1.08 [0.98-1.20] | 1.09 [0.99-1.21] | 1.11 [0.99-1.24] | 1.09 [0.79-1.50] | 1.13 [1.02-1.25] | 1.02 [0.92-1.13] | 1.01 [0.91-1.12] | 1.03 [0.92-1.15] | 1.08 [0.77-1.51] | 1.08 [0.98-1.20] | 1,191 | 1,128 | 941 | 133 | 1,161 |
| S40 | 1.11 [1.04-1.19] | 1.13 [1.05-1.20] | 1.13 [1.05-1.21] | 0.84 [0.60-1.16] | 1.12 [1.05-1.20] | 1.07 [1.00-1.15] | 1.06 [0.99-1.13] | 1.07 [0.99-1.14] | 0.86 [0.61-1.21] | 1.09 [1.02-1.16] | 2,849 | 2,771 | 2,646 | 114 | 2,777 |
| S41 | 1.22 [1.09-1.37] | 1.24 [1.10-1.39] | 1.23 [1.09-1.39] | | 1.22 [1.09-1.36] | 1.16 [1.04-1.30] | 1.15 [1.02-1.29] | 1.13 [1.00-1.28] | | 1.17 [1.05-1.31] | 1,080 | 1,037 | 930 | NA | 1,040 |
| S42 | 1.09 [1.05-1.13] | 1.09 [1.05-1.14] | 1.10 [1.06-1.15] | 1.02 [0.90-1.15] | 1.07 [1.03-1.12] | 1.07 [1.03-1.11] | 1.05 [1.01-1.10] | 1.06 [1.02-1.11] | 1.02 [0.90-1.16] | 1.06 [1.02-1.11] | 8,104 | 7,361 | 6,993 | 886 | 7,380 |

### Supplementary Table 4: Hazard ratios and events from all cohorts (excluding non-consulters)

| Outcome | Hazard ratio (99% confidence interval): crude: any age | 18+ | 40+ | <18 | hosp. | adjusted: any age | 18+ | 40+ | <18 | hosp. | Events (in exposed): any age | 18+ | 40+ | <18 | hosp. |
|---|---|---|---|---|---|---|---|---|---|---|---|---|---|---|---|
| S43 | 1.04 [0.95-1.15] | 1.06 [0.96-1.16] | 1.04 [0.94-1.15] | | 1.08 [0.98-1.19] | 1.02 [0.93-1.12] | 1.03 [0.93-1.13] | 1.01 [0.91-1.12] | | 1.07 [0.97-1.18] | 1,282 | 1,285 | 1,185 | NA | 1,199 |
| S46 | 1.05 [0.89-1.23] | 1.05 [0.89-1.23] | 1.01 [0.86-1.20] | | 1.02 [0.87-1.19] | 1.01 [0.86-1.19] | 1.01 [0.85-1.19] | 0.94 [0.79-1.12] | | 0.98 [0.84-1.15] | 483 | 465 | 423 | NA | 463 |
| S49 | 1.10 [0.98-1.24] | 1.19 [1.06-1.34] | 1.13 [1.01-1.28] | | 1.13 [1.01-1.27] | 1.04 [0.92-1.17] | 1.10 [0.98-1.24] | 1.06 [0.94-1.20] | | 1.10 [0.97-1.23] | 891 | 874 | 826 | NA | 858 |
| S50 | 1.15 [1.08-1.23] | 1.12 [1.05-1.20] | 1.14 [1.07-1.22] | 0.88 [0.67-1.15] | 1.11 [1.04-1.18] | 1.11 [1.04-1.19] | 1.06 [0.99-1.13] | 1.08 [1.01-1.16] | 0.89 [0.68-1.18] | 1.08 [1.01-1.16] | 3,024 | 2,893 | 2,715 | 174 | 2,898 |
| S51 | 1.15 [1.09-1.22] | 1.19 [1.12-1.26] | 1.17 [1.11-1.25] | 0.93 [0.74-1.17] | 1.18 [1.12-1.25] | 1.09 [1.03-1.15] | 1.10 [1.04-1.17] | 1.09 [1.03-1.16] | 0.98 [0.77-1.25] | 1.13 [1.07-1.20] | 4,342 | 4,218 | 3,853 | 250 | 4,175 |
| S52 | 1.08 [1.04-1.11] | 1.09 [1.05-1.13] | 1.09 [1.05-1.13] | 0.97 [0.89-1.05] | 1.06 [1.02-1.09] | 1.05 [1.02-1.09] | 1.05 [1.02-1.09] | 1.06 [1.02-1.10] | 0.95 [0.87-1.04] | 1.04 [1.00-1.08] | 10,338 | 8,549 | 7,791 | 2,009 | 8,934 |
| S53 | 1.04 [0.84-1.29] | 1.01 [0.79-1.28] | 0.93 [0.71-1.22] | | 1.16 [0.92-1.46] | 0.98 [0.78-1.22] | 0.93 [0.73-1.20] | 0.87 [0.66-1.15] | | 1.13 [0.89-1.43] | 260 | 189 | 148 | NA | 224 |
| S54 | 1.10 [0.78-1.53] | 1.14 [0.80-1.61] | | | 1.30 [0.92-1.83] | 1.06 [0.75-1.51] | 1.02 [0.71-1.46] | | | 1.23 [0.86-1.75] | 103 | 101 | NA | NA | 104 |
| S56 | 0.97 [0.80-1.18] | 0.99 [0.81-1.21] | 1.04 [0.81-1.33] | | 0.94 [0.77-1.16] | 0.95 [0.78-1.16] | 0.94 [0.77-1.16] | 1.00 [0.78-1.30] | | 0.94 [0.76-1.17] | 309 | 287 | 172 | NA | 254 |
| S59 | 1.26 [1.05-1.52] | 1.18 [0.97-1.43] | 1.25 [1.03-1.53] | | 1.19 [0.99-1.44] | 1.20 [0.99-1.46] | 1.09 [0.89-1.33] | 1.15 [0.94-1.42] | | 1.16 [0.96-1.40] | 362 | 332 | 314 | NA | 345 |
| S60 | 1.15 [1.06-1.23] | 1.15 [1.07-1.25] | 1.18 [1.09-1.28] | 0.88 [0.68-1.14] | 1.11 [1.03-1.20] | 1.11 [1.03-1.20] | 1.10 [1.01-1.18] | 1.12 [1.03-1.21] | 0.90 [0.69-1.18] | 1.08 [1.00-1.17] | 2,231 | 2,105 | 1,883 | 195 | 2,148 |
| S61 | 1.07 [1.02-1.12] | 1.07 [1.02-1.12] | 1.09 [1.04-1.15] | 0.94 [0.85-1.04] | 1.05 [1.01-1.10] | 1.03 [0.98-1.07] | 1.01 [0.96-1.05] | 1.04 [0.99-1.10] | 0.94 [0.85-1.05] | 1.03 [0.98-1.08] | 6,425 | 5,633 | 4,340 | 1,254 | 5,766 |
| S62 | 1.05 [1.00-1.11] | 1.06 [1.00-1.12] | 1.07 [1.00-1.14] | 1.01 [0.89-1.15] | 1.01 [0.95-1.07] | 1.03 [0.97-1.09] | 1.01 [0.95-1.07] | 1.02 [0.96-1.09] | 1.00 [0.87-1.14] | 1.00 [0.94-1.06] | 4,095 | 3,643 | 2,795 | 831 | 3,577 |
| S63 | 1.19 [1.05-1.35] | 1.19 [1.05-1.35] | 1.20 [1.04-1.38] | | 1.15 [1.01-1.31] | 1.16 [1.02-1.32] | 1.14 [1.00-1.30] | 1.17 [1.01-1.35] | | 1.11 [0.98-1.27] | 771 | 750 | 581 | NA | 673 |
| S64 | 0.96 [0.85-1.10] | 0.94 [0.83-1.07] | 0.95 [0.81-1.12] | 0.86 [0.64-1.16] | 0.94 [0.82-1.07] | 0.95 [0.84-1.09] | 0.90 [0.79-1.03] | 0.93 [0.78-1.10] | 0.89 [0.65-1.21] | 0.93 [0.81-1.06] | 711 | 662 | 390 | 153 | 590 |
| S65 | 0.89 [0.70-1.14] | 0.97 [0.76-1.26] | 0.90 [0.66-1.23] | | 0.96 [0.74-1.24] | 0.88 [0.69-1.13] | 0.94 [0.73-1.22] | 0.87 [0.63-1.19] | | 0.96 [0.74-1.24] | 193 | 171 | 111 | NA | 163 |
| S66 | 1.07 [0.97-1.18] | 0.98 [0.89-1.08] | 0.95 [0.84-1.08] | 1.02 [0.81-1.28] | 0.97 [0.88-1.08] | 1.05 [0.95-1.16] | 0.94 [0.85-1.04] | 0.93 [0.82-1.06] | 1.03 [0.81-1.31] | 0.96 [0.87-1.07] | 1,297 | 1,228 | 703 | 260 | 1,045 |
| S67 | 0.78 [0.59-1.04] | | | | 0.73 [0.53-0.99] | 0.77 [0.57-1.03] | | | | 0.74 [0.54-1.02] | 137 | NA | NA | NA | 108 |
| S68 | 0.98 [0.84-1.15] | 0.94 [0.78-1.13] | 1.02 [0.83-1.25] | 0.94 [0.70-1.26] | 0.92 [0.77-1.09] | 0.97 [0.83-1.14] | 0.96 [0.80-1.16] | 1.03 [0.84-1.26] | 0.96 [0.71-1.29] | 0.92 [0.77-1.10] | 447 | 324 | 264 | 147 | 359 |
| S69 | 1.23 [1.09-1.39] | 1.22 [1.07-1.38] | 1.20 [1.05-1.37] | 1.04 [0.74-1.48] | 1.20 [1.07-1.36] | 1.17 [1.03-1.32] | 1.11 [0.97-1.26] | 1.10 [0.96-1.26] | 1.16 [0.80-1.67] | 1.15 [1.02-1.30] | 868 | 783 | 665 | 108 | 816 |
| S70 | 1.13 [1.06-1.21] | 1.11 [1.04-1.19] | 1.13 [1.06-1.21] | 1.02 [0.74-1.42] | 1.10 [1.03-1.18] | 1.09 [1.02-1.16] | 1.04 [0.97-1.11] | 1.07 [1.00-1.15] | 1.05 [0.75-1.48] | 1.08 [1.01-1.15] | 2,873 | 2,763 | 2,668 | 126 | 2,791 |
| S71 | 1.07 [0.89-1.28] | 1.15 [0.95-1.40] | 1.23 [0.98-1.54] | | 1.13 [0.94-1.35] | 1.01 [0.84-1.21] | 1.07 [0.88-1.30] | 1.15 [0.91-1.46] | | 1.08 [0.89-1.30] | 391 | 342 | 231 | NA | 352 |
| S72 | 1.07 [1.05-1.10] | 1.08 [1.05-1.11] | 1.08 [1.06-1.11] | 0.95 [0.76-1.19] | 1.09 [1.06-1.11] | 1.07 [1.04-1.10] | 1.06 [1.04-1.09] | 1.06 [1.04-1.09] | 0.98 [0.78-1.24] | 1.09 [1.06-1.12] | 19,778 | 19,593 | 19,431 | 244 | 18,303 |
| S73 | 1.07 [0.86-1.34] | 1.15 [0.92-1.44] | 1.17 [0.92-1.47] | | 1.12 [0.90-1.40] | 1.03 [0.82-1.29] | 1.08 [0.86-1.36] | 1.10 [0.86-1.39] | | 1.11 [0.89-1.39] | 237 | 221 | 209 | NA | 225 |
| S76 | 0.93 [0.79-1.10] | 0.93 [0.79-1.10] | 0.97 [0.82-1.15] | | 1.06 [0.90-1.24] | 0.88 [0.75-1.04] | 0.90 [0.76-1.06] | 0.92 [0.77-1.10] | | 1.01 [0.85-1.19] | 426 | 408 | 371 | NA | 398 |
| S79 | 1.16 [1.06-1.27] | 1.14 [1.04-1.24] | 1.19 [1.08-1.30] | | 1.16 [1.06-1.26] | 1.12 [1.02-1.23] | 1.08 [0.98-1.18] | 1.13 [1.03-1.24] | | 1.14 [1.04-1.24] | 1,565 | 1,541 | 1,495 | NA | 1,499 |
| S80 | 1.11 [1.06-1.17] | 1.12 [1.07-1.17] | 1.11 [1.06-1.17] | 0.90 [0.71-1.13] | 1.08 [1.03-1.13] | 1.06 [1.01-1.11] | 1.04 [0.99-1.10] | 1.04 [0.98-1.09] | 0.89 [0.70-1.13] | 1.04 [0.99-1.09] | 5,411 | 5,250 | 5,013 | 225 | 5,227 |
| S81 | 1.19 [1.13-1.26] | 1.19 [1.12-1.26] | 1.22 [1.15-1.29] | 1.07 [0.84-1.37] | 1.17 [1.10-1.24] | 1.11 [1.05-1.18] | 1.09 [1.03-1.15] | 1.12 [1.06-1.19] | 1.09 [0.84-1.40] | 1.10 [1.04-1.17] | 4,074 | 3,930 | 3,716 | 224 | 3,896 |
| S82 | 1.07 [1.03-1.11] | 1.06 [1.02-1.10] | 1.05 [1.01-1.09] | 1.09 [0.96-1.24] | 1.05 [1.01-1.09] | 1.04 [1.00-1.07] | 1.01 [0.97-1.05] | 1.00 [0.96-1.05] | 1.09 [0.96-1.24] | 1.03 [0.99-1.07] | 8,511 | 8,003 | 6,755 | 899 | 7,470 |
| S83 | 1.14 [1.02-1.27] | 1.17 [1.05-1.31] | 1.12 [0.98-1.29] | 1.39 [1.04-1.86] | 1.11 [0.99-1.25] | 1.08 [0.96-1.21] | 1.10 [0.97-1.23] | 1.05 [0.91-1.20] | 1.35 [0.99-1.84] | 1.06 [0.94-1.19] | 984 | 931 | 636 | 183 | 866 |
| S86 | 1.10 [0.95-1.27] | 1.15 [0.99-1.33] | 1.15 [0.98-1.35] | | 1.08 [0.93-1.26] | 1.06 [0.91-1.23] | 1.09 [0.94-1.27] | 1.07 [0.91-1.27] | | 1.03 [0.88-1.20] | 559 | 549 | 440 | NA | 473 |
| S89 | 1.16 [1.05-1.29] | 1.18 [1.06-1.31] | 1.19 [1.07-1.32] | | 1.13 [1.02-1.25] | 1.11 [1.00-1.23] | 1.10 [0.99-1.22] | 1.11 [1.00-1.24] | | 1.10 [0.99-1.22] | 1,187 | 1,155 | 1,076 | NA | 1,133 |
| S90 | 1.08 [0.98-1.20] | 1.08 [0.97-1.19] | 1.05 [0.95-1.17] | 0.97 [0.68-1.38] | 1.05 [0.95-1.15] | 1.04 [0.94-1.15] | 0.99 [0.89-1.10] | 0.98 [0.88-1.09] | 1.00 [0.70-1.44] | 1.01 [0.92-1.12] | 1,242 | 1,161 | 1,059 | 106 | 1,185 |
| S91 | 1.13 [1.01-1.25] | 1.19 [1.06-1.33] | 1.23 [1.09-1.40] | 1.03 [0.80-1.33] | 1.10 [0.99-1.23] | 1.07 [0.96-1.19] | 1.08 [0.97-1.22] | 1.13 [0.99-1.28] | 0.98 [0.76-1.28] | 1.07 [0.96-1.19] | 1,101 | 981 | 818 | 207 | 1,021 |
| S92 | 1.11 [1.03-1.20] | 1.11 [1.03-1.20] | 1.16 [1.06-1.26] | 1.09 [0.81-1.47] | 1.12 [1.04-1.22] | 1.06 [0.98-1.15] | 1.03 [0.95-1.12] | 1.07 [0.98-1.16] | 1.12 [0.82-1.53] | 1.09 [1.00-1.18] | 1,920 | 1,849 | 1,594 | 166 | 1,747 |
| S93 | 1.10 [0.96-1.26] | 1.06 [0.92-1.21] | 1.10 [0.95-1.28] | | 1.06 [0.92-1.21] | 1.04 [0.91-1.20] | 0.98 [0.85-1.13] | 1.03 [0.89-1.20] | | 1.02 [0.89-1.18] | 631 | 615 | 515 | NA | 599 |
| S96 | 1.11 [0.81-1.53] | 1.07 [0.77-1.47] | | | 1.06 [0.77-1.46] | 1.02 [0.73-1.41] | 0.90 [0.64-1.26] | | | 0.93 [0.66-1.30] | 118 | 112 | NA | NA | 108 |
| S99 | 1.08 [0.93-1.26] | 1.07 [0.92-1.24] | 1.06 [0.90-1.25] | | 1.03 [0.89-1.20] | 0.97 [0.83-1.14] | 0.93 [0.79-1.09] | 0.95 [0.81-1.13] | | 0.99 [0.85-1.15] | 531 | 504 | 448 | NA | 498 |
| T00 | 1.10 [0.99-1.22] | 1.11 [1.00-1.24] | 1.13 [1.01-1.26] | | 1.12 [1.00-1.24] | 1.06 [0.95-1.18] | 1.03 [0.92-1.15] | 1.06 [0.95-1.19] | | 1.09 [0.98-1.21] | 1,147 | 1,089 | 1,021 | NA | 1,114 |
| T01 | 1.04 [0.88-1.22] | 1.03 [0.88-1.22] | 1.09 [0.92-1.30] | | 1.08 [0.93-1.26] | 0.98 [0.83-1.16] | 0.93 [0.78-1.10] | 0.99 [0.83-1.19] | | 1.04 [0.89-1.22] | 527 | 490 | 424 | NA | 518 |
| T02 | 1.20 [1.00-1.45] | 1.21 [1.00-1.47] | 1.26 [1.03-1.55] | | 1.07 [0.88-1.30] | 1.19 [0.98-1.44] | 1.17 [0.96-1.42] | 1.22 [1.00-1.50] | | 1.06 [0.87-1.29] | 328 | 303 | 282 | NA | 291 |
| T09 | 1.32 [1.03-1.70] | 1.22 [0.95-1.58] | 1.33 [1.02-1.75] | | 1.19 [0.93-1.53] | 1.25 [0.96-1.62] | 1.16 [0.89-1.50] | 1.22 [0.92-1.61] | | 1.15 [0.89-1.48] | 193 | 176 | 162 | NA | 178 |
| T11 | 1.21 [1.07-1.37] | 1.18 [1.04-1.34] | 1.21 [1.06-1.37] | | 1.23 [1.09-1.39] | 1.15 [1.01-1.30] | 1.08 [0.95-1.23] | 1.11 [0.97-1.27] | | 1.19 [1.05-1.35] | 852 | 822 | 748 | NA | 837 |
| T13 | 1.19 [1.02-1.39] | 1.24 [1.06-1.45] | 1.20 [1.02-1.41] | | 1.14 [0.98-1.33] | 1.13 [0.97-1.32] | 1.16 [0.99-1.36] | 1.09 [0.93-1.29] | | 1.10 [0.94-1.28] | 521 | 503 | 469 | NA | 495 |
| T14 | 1.21 [1.03-1.42] | 1.08 [0.92-1.28] | 1.17 [0.99-1.39] | | 1.15 [0.98-1.35] | 1.14 [0.97-1.35] | 1.01 [0.85-1.19] | 1.07 [0.89-1.27] | | 1.11 [0.94-1.31] | 454 | 428 | 376 | NA | 412 |
| T16 | 0.97 [0.82-1.14] | | | 0.96 [0.80-1.14] | 1.04 [0.87-1.24] | 0.95 [0.81-1.12] | | | 0.98 [0.81-1.17] | 1.03 [0.86-1.23] | 472 | NA | NA | 403 | 434 |
| T17 | 1.16 [1.06-1.27] | 1.24 [1.11-1.37] | 1.18 [1.06-1.32] | 1.04 [0.87-1.25] | 1.13 [1.03-1.24] | 1.10 [1.00-1.21] | 1.15 [1.03-1.28] | 1.11 [0.99-1.24] | 1.03 [0.86-1.24] | 1.09 [0.99-1.19] | 1,432 | 1,080 | 979 | 381 | 1,305 |
| T18 | 1.15 [1.03-1.27] | 1.24 [1.11-1.39] | 1.18 [1.04-1.34] | 0.93 [0.74-1.16] | 1.06 [0.95-1.18] | 1.09 [0.98-1.21] | 1.16 [1.03-1.30] | 1.10 [0.97-1.25] | 0.88 [0.69-1.11] | 1.02 [0.91-1.13] | 1,102 | 907 | 732 | 240 | 961 |
| T19 | 1.21 [0.88-1.65] | 1.38 [1.01-1.90] | | | 1.20 [0.87-1.65] | 1.11 [0.80-1.54] | 1.33 [0.96-1.84] | | | 1.18 [0.85-1.64] | 121 | 114 | NA | NA | 110 |
| T20 | 1.11 [0.94-1.32] | 1.18 [0.95-1.47] | 1.18 [0.93-1.51] | 0.99 [0.76-1.28] | 1.06 [0.88-1.26] | 1.09 [0.92-1.30] | 1.12 [0.90-1.40] | 1.14 [0.89-1.46] | 0.99 [0.76-1.30] | 1.06 [0.89-1.28] | 402 | 245 | 188 | 171 | 346 |
| T21 | 1.05 [0.92-1.21] | 1.18 [0.99-1.39] | 1.20 [1.00-1.44] | 0.93 [0.74-1.17] | 1.08 [0.93-1.24] | 1.05 [0.91-1.20] | 1.12 [0.95-1.33] | 1.14 [0.95-1.37] | 0.95 [0.75-1.20] | 1.07 [0.93-1.23] | 632 | 437 | 369 | 218 | 577 |
| T22 | 1.11 [0.95-1.29] | 1.06 [0.88-1.28] | 1.09 [0.88-1.36] | 1.11 [0.87-1.42] | 1.06 [0.90-1.23] | 1.10 [0.94-1.28] | 1.01 [0.84-1.23] | 1.07 [0.86-1.33] | 1.13 [0.88-1.46] | 1.05 [0.90-1.23] | 496 | 316 | 240 | 199 | 462 |
| T23 | 1.11 [0.95-1.31] | 1.03 [0.84-1.26] | 1.12 [0.89-1.42] | 1.15 [0.91-1.47] | 0.98 [0.83-1.16] | 1.10 [0.94-1.30] | 0.97 [0.79-1.20] | 1.07 [0.84-1.36] | 1.16 [0.90-1.49] | 0.97 [0.81-1.15] | 467 | 283 | 208 | 203 | 394 |
| T24 | 0.85 [0.72-1.00] | 0.91 [0.75-1.10] | 0.93 [0.76-1.13] | | 0.87 [0.73-1.04] | 0.85 [0.71-1.01] | 0.87 [0.72-1.06] | 0.91 [0.74-1.12] | | 0.88 [0.74-1.05] | 400 | 318 | 278 | NA | 357 |
| T25 | 0.80 [0.63-1.02] | 0.88 [0.65-1.18] | 0.84 [0.61-1.17] | | 0.90 [0.70-1.15] | 0.77 [0.60-0.99] | 0.84 [0.62-1.13] | 0.79 [0.56-1.10] | | 0.88 [0.69-1.14] | 198 | 127 | 106 | NA | 177 |
| T31 | 1.00 [0.90-1.10] | 1.00 [0.88-1.13] | 1.07 [0.93-1.24] | 0.99 [0.85-1.15] | 0.99 [0.90-1.10] | 0.98 [0.89-1.08] | 0.95 [0.84-1.08] | 1.03 [0.89-1.19] | 1.00 [0.86-1.16] | 0.99 [0.89-1.10] | 1,231 | 730 | 569 | 544 | 1,109 |
| T36 | 1.26 [1.04-1.52] | 1.20 [0.99-1.46] | 1.37 [1.04-1.81] | | 1.22 [1.01-1.48] | 1.15 [0.94-1.40] | 1.05 [0.86-1.29] | 1.24 [0.93-1.65] | | 1.19 [0.98-1.45] | 341 | 316 | 158 | NA | 317 |
| T37 | 1.33 [1.01-1.76] | 1.37 [1.02-1.84] | | | 1.30 [0.99-1.71] | 1.20 [0.90-1.61] | 1.14 [0.84-1.56] | | | 1.25 [0.95-1.66] | 162 | 149 | NA | NA | 160 |
| T38 | 1.15 [1.01-1.30] | 1.13 [0.99-1.28] | 1.14 [0.98-1.32] | | 1.09 [0.97-1.24] | 1.06 [0.93-1.20] | 0.96 [0.84-1.10] | 0.98 [0.84-1.14] | | 1.04 [0.92-1.18] | 742 | 709 | 538 | NA | 752 |
| T39 | 1.09 [1.04-1.14] | 1.07 [1.02-1.12] | 1.20 [1.12-1.28] | 0.95 [0.87-1.04] | 1.04 [0.99-1.09] | 1.04 [0.99-1.09] | 0.94 [0.90-0.99] | 1.06 [0.99-1.14] | 0.96 [0.87-1.05] | 1.03 [0.98-1.08] | 5,432 | 5,003 | 2,411 | 1,548 | 4,851 |
| T40 | 1.17 [1.10-1.25] | 1.16 [1.09-1.23] | 1.21 [1.12-1.30] | 0.98 [0.83-1.16] | 1.08 [1.02-1.15] | 1.08 [1.01-1.15] | 0.98 [0.92-1.05] | 1.03 [0.95-1.12] | 1.00 [0.84-1.19] | 1.05 [0.99-1.12] | 3,171 | 3,202 | 1,882 | 465 | 3,091 |
| T42 | 1.11 [1.03-1.19] | 1.14 [1.07-1.23] | 1.18 [1.08-1.29] | 0.75 [0.60-0.93] | 1.04 [0.97-1.12] | 1.03 [0.96-1.11] | 0.98 [0.91-1.05] | 1.02 [0.93-1.12] | 0.80 [0.63-1.00] | 1.03 [0.96-1.11] | 2,336 | 2,354 | 1,432 | 251 | 2,213 |
| T43 | 1.14 [1.07-1.21] | 1.14 [1.07-1.20] | 1.21 [1.11-1.31] | 0.91 [0.80-1.05] | 1.06 [1.00-1.13] | 1.07 [1.00-1.13] | 0.98 [0.92-1.04] | 1.05 [0.96-1.15] | 0.95 [0.83-1.10] | 1.06 [1.00-1.13] | 3,376 | 3,399 | 1,555 | 672 | 3,099 |
| T44 | 1.09 [0.96-1.23] | 1.10 [0.97-1.24] | 1.11 [0.95-1.30] | 0.92 [0.68-1.24] | 1.08 [0.96-1.22] | 1.05 [0.92-1.19] | 0.98 [0.86-1.11] | 0.98 [0.83-1.16] | 0.94 [0.69-1.28] | 1.08 [0.96-1.23] | 772 | 755 | 455 | 136 | 752 |
| T45 | 1.21 [1.08-1.35] | 1.19 [1.06-1.33] | 1.21 [1.06-1.39] | 1.14 [0.90-1.45] | 1.14 [1.03-1.27] | 1.11 [0.99-1.24] | 1.06 [0.94-1.19] | 1.06 [0.92-1.23] | 1.13 [0.88-1.45] | 1.10 [0.99-1.23] | 1,026 | 946 | 621 | 238 | 1,002 |
| T46 | 1.09 [0.98-1.22] | 1.10 [0.98-1.23] | 1.12 [1.00-1.26] | | 1.06 [0.95-1.18] | 1.04 [0.93-1.16] | 0.99 [0.88-1.12] | 1.00 [0.89-1.13] | | 1.03 [0.92-1.16] | 920 | 888 | 795 | NA | 900 |
| T47 | 1.20 [1.01-1.42] | 1.21 [1.02-1.44] | 1.17 [0.95-1.44] | | 1.13 [0.96-1.34] | 1.08 [0.91-1.29] | 1.00 [0.83-1.20] | 0.99 [0.79-1.23] | | 1.09 [0.92-1.29] | 421 | 406 | 269 | NA | 416 |
| T48 | 1.26 [0.96-1.65] | 1.42 [1.04-1.96] | | | 1.22 [0.93-1.60] | 1.03 [0.77-1.38] | 1.07 [0.76-1.52] | | | 1.06 [0.80-1.41] | 178 | 122 | NA | NA | 165 |
| T49 | 1.05 [0.78-1.40] | | | | 1.05 [0.77-1.42] | 0.99 [0.74-1.33] | | | | 1.03 [0.76-1.40] | 138 | NA | NA | NA | 120 |
| T50 | 1.11 [0.99-1.24] | 1.11 [0.99-1.25] | 1.17 [1.02-1.34] | 1.02 [0.75-1.38] | 1.10 [0.98-1.23] | 1.03 [0.92-1.16] | 0.97 [0.86-1.09] | 1.03 [0.90-1.19] | 1.03 [0.75-1.41] | 1.06 [0.95-1.19] | 957 | 881 | 636 | 133 | 880 |
| T51 | 1.12 [1.05-1.20] | 1.07 [1.00-1.14] | 1.17 [1.07-1.29] | 0.96 [0.81-1.13] | 1.04 [0.97-1.11] | 1.05 [0.98-1.13] | 0.92 [0.86-0.98] | 1.00 [0.91-1.10] | 1.07 [0.89-1.28] | 1.04 [0.97-1.11] | 2,733 | 2,806 | 1,315 | 451 | 2,525 |
| T52 | 1.02 [0.74-1.40] | | | | 1.03 [0.74-1.43] | 0.96 [0.69-1.34] | | | | 1.02 [0.73-1.42] | 111 | NA | NA | NA | 106 |
| T54 | 0.90 [0.66-1.24] | | | | 0.97 [0.71-1.32] | 0.89 [0.64-1.23] | | | | 0.98 [0.72-1.35] | 122 | NA | NA | NA | 116 |
| T59 | 0.94 [0.75-1.18] | 0.93 [0.74-1.17] | 1.04 [0.82-1.33] | | 0.85 [0.68-1.07] | 0.94 [0.74-1.18] | 0.86 [0.68-1.09] | 0.99 [0.78-1.27] | | 0.84 [0.67-1.06] | 217 | 209 | 186 | NA | 206 |
| T63 | 0.95 [0.69-1.33] | | | | | 0.85 [0.60-1.20] | | | | | 103 | NA | NA | NA | NA |
| T65 | 1.01 [0.76-1.34] | | | | 0.95 [0.71-1.27] | 0.97 [0.73-1.30] | | | | 0.93 [0.69-1.25] | 142 | NA | NA | NA | 135 |
| T68 | 1.07 [0.98-1.18] | 1.08 [0.98-1.18] | 1.08 [0.99-1.19] | | 1.06 [0.96-1.16] | 1.09 [0.99-1.20] | 1.08 [0.98-1.19] | 1.09 [0.99-1.20] | | 1.10 [1.00-1.20] | 1,443 | 1,438 | 1,411 | NA | 1,420 |
| T74 | 1.03 [0.75-1.40] | | | | 1.04 [0.77-1.41] | 0.96 [0.70-1.32] | | | | 1.04 [0.76-1.42] | 128 | NA | NA | NA | 123 |
| T78 | 2.27 [2.13-2.40] | 1.67 [1.55-1.79] | 1.55 [1.43-1.68] | 5.30 [4.70-5.99] | 2.18 [2.05-2.31] | 1.96 [1.84-2.09] | 1.41 [1.31-1.52] | 1.37 [1.26-1.49] | 4.23 [3.72-4.80] | 1.90 [1.78-2.02] | 4,543 | 2,758 | 1,866 | 2,136 | 4,068 |
| T79 | 1.08 [0.99-1.17] | 1.09 [1.00-1.19] | 1.09 [0.99-1.19] | | 1.04 [0.95-1.14] | 1.06 [0.97-1.15] | 1.05 [0.96-1.15] | 1.05 [0.96-1.15] | | 1.05 [0.96-1.14] | 1,685 | 1,643 | 1,504 | NA | 1,516 |
| T80 | 1.25 [1.11-1.40] | 1.24 [1.10-1.40] | 1.30 [1.14-1.47] | | 1.22 [1.09-1.37] | 1.19 [1.06-1.34] | 1.17 [1.04-1.32] | 1.21 [1.07-1.38] | | 1.21 [1.07-1.36] | 873 | 832 | 746 | NA | 824 |
| T81 | 1.18 [1.16-1.21] | 1.19 [1.16-1.21] | 1.18 [1.15-1.20] | 1.16 [1.06-1.28] | 1.15 [1.13-1.18] | 1.13 [1.11-1.16] | 1.11 [1.09-1.14] | 1.11 [1.08-1.13] | 1.15 [1.05-1.27] | 1.12 [1.10-1.15] | 26,204 | 25,714 | 22,140 | 1,560 | 24,446 |
| T82 | 1.21 [1.16-1.26] | 1.21 [1.16-1.26] | 1.20 [1.15-1.25] | 1.03 [0.82-1.30] | 1.14 [1.10-1.19] | 1.17 [1.12-1.22] | 1.13 [1.09-1.18] | 1.11 [1.06-1.16] | 1.02 [0.81-1.29] | 1.13 [1.09-1.18] | 7,103 | 6,922 | 6,570 | 258 | 6,949 |
| T83 | 1.06 [1.01-1.10] | 1.07 [1.03-1.12] | 1.06 [1.01-1.11] | 0.88 [0.64-1.21] | 1.04 [1.00-1.08] | 1.04 [0.99-1.08] | 1.04 [0.99-1.08] | 1.02 [0.98-1.07] | 0.91 [0.65-1.27] | 1.04 [1.00-1.09] | 6,466 | 6,498 | 5,893 | 129 | 6,378 |
| T84 | 1.21 [1.17-1.26] | 1.22 [1.17-1.26] | 1.21 [1.17-1.26] | 1.14 [0.90-1.44] | 1.17 [1.13-1.21] | 1.16 [1.12-1.21] | 1.15 [1.11-1.20] | 1.15 [1.10-1.19] | 1.13 [0.88-1.44] | 1.14 [1.10-1.18] | 8,565 | 8,481 | 7,868 | 274 | 8,290 |
| T85 | 1.16 [1.11-1.22] | 1.20 [1.15-1.26] | 1.19 [1.14-1.25] | 0.88 [0.73-1.07] | 1.13 [1.08-1.18] | 1.12 [1.07-1.17] | 1.13 [1.08-1.19] | 1.12 [1.07-1.18] | 0.86 [0.71-1.05] | 1.11 [1.06-1.16] | 5,497 | 5,315 | 4,722 | 348 | 5,290 |
| T86 | 1.17 [1.03-1.32] | 1.16 [1.02-1.31] | 1.17 [1.02-1.34] | | 1.13 [1.00-1.28] | 1.14 [1.00-1.30] | 1.12 [0.98-1.27] | 1.09 [0.95-1.26] | | 1.15 [1.01-1.30] | 735 | 709 | 601 | NA | 729 |
| T87 | 1.17 [1.01-1.36] | 1.11 [0.96-1.29] | 1.15 [0.99-1.34] | | 1.12 [0.97-1.30] | 1.11 [0.95-1.29] | 1.03 [0.88-1.20] | 1.02 [0.88-1.20] | | 1.10 [0.94-1.28] | 517 | 510 | 492 | NA | 501 |
| T88 | 1.31 [1.21-1.42] | 1.32 [1.21-1.43] | 1.27 [1.16-1.39] | 1.54 [1.15-2.07] | 1.28 [1.18-1.39] | 1.20 [1.10-1.30] | 1.18 [1.08-1.28] | 1.15 [1.05-1.26] | 1.49 [1.09-2.02] | 1.20 [1.11-1.31] | 1,807 | 1,713 | 1,432 | 179 | 1,718 |
| T90 | 1.20 [1.08-1.33] | 1.15 [1.04-1.29] | 1.27 [1.12-1.44] | 1.33 [0.99-1.80] | 1.11 [0.99-1.23] | 1.15 [1.03-1.28] | 1.09 [0.97-1.21] | 1.18 [1.03-1.34] | 1.34 [0.98-1.83] | 1.09 [0.98-1.22] | 1,087 | 1,021 | 722 | 164 | 982 |
| T91 | 1.19 [1.04-1.38] | 1.12 [0.97-1.29] | 1.12 [0.96-1.31] | | 1.11 [0.97-1.28] | 1.12 [0.97-1.30] | 1.03 [0.89-1.19] | 1.03 [0.88-1.21] | | 1.06 [0.92-1.22] | 580 | 563 | 458 | NA | 557 |
| T92 | 1.09 [0.97-1.22] | 1.11 [0.98-1.25] | 1.14 [1.00-1.30] | | 1.06 [0.94-1.20] | 1.05 [0.94-1.19] | 1.04 [0.92-1.18] | 1.09 [0.95-1.24] | | 1.04 [0.92-1.18] | 829 | 805 | 636 | NA | 736 |
| T93 | 1.19 [1.08-1.32] | 1.19 [1.08-1.32] | 1.18 [1.05-1.32] | 1.36 [0.93-1.98] | 1.15 [1.04-1.28] | 1.13 [1.02-1.26] | 1.10 [0.99-1.23] | 1.09 [0.96-1.22] | 1.34 [0.91-1.97] | 1.11 [1.00-1.24] | 1,116 | 1,088 | 839 | 104 | 1,020 |
| T94 | 1.02 [0.75-1.39] | 1.10 [0.81-1.50] | 1.06 [0.76-1.48] | | 1.18 [0.86-1.62] | 1.02 [0.74-1.39] | 1.00 [0.73-1.38] | 0.97 [0.69-1.37] | | 1.12 [0.81-1.55] | 122 | 119 | 105 | NA | 116 |
| T95 | 1.13 [0.81-1.56] | | | | | 1.10 [0.78-1.53] | | | | | 106 | NA | NA | NA | NA |
| T98 | 1.16 [0.93-1.45] | 1.33 [1.06-1.67] | 1.29 [1.02-1.63] | | 1.08 [0.87-1.35] | 1.13 [0.89-1.41] | 1.27 [1.01-1.60] | 1.23 [0.97-1.56] | | 1.04 [0.83-1.30] | 226 | 232 | 210 | NA | 213 |
| T56 | | | | | 1.10 [0.80-1.53] | | | | | 1.12 [0.81-1.56] | NA | NA | NA | NA | 102 |
| Codes for special purposes (U00-U89) | | | | | | | | | | | | | | | |
| U07 | 1.15 [1.11-1.20] | 1.13 [1.09-1.18] | 1.16 [1.12-1.21] | 0.90 [0.71-1.14] | 1.15 [1.11-1.20] | 1.09 [1.05-1.14] | 1.06 [1.02-1.10] | 1.08 [1.04-1.12] | 0.85 [0.66-1.09] | 1.11 [1.07-1.16] | 9,064 | 9,073 | 8,310 | 248 | 8,991 |
| U80 | 1.38 [1.30-1.47] | 1.39 [1.31-1.47] | 1.36 [1.28-1.45] | 1.47 [1.04-2.10] | 1.37 [1.30-1.46] | 1.32 [1.25-1.41] | 1.29 [1.22-1.37] | 1.26 [1.18-1.34] | 1.43 [0.99-2.08] | 1.33 [1.26-1.41] | 3,571 | 3,519 | 3,276 | 126 | 3,550 |
| U81 | 1.13 [0.87-1.47] | 1.05 [0.81-1.36] | 0.99 [0.76-1.29] | | 1.08 [0.83-1.39] | 1.10 [0.84-1.44] | 0.99 [0.76-1.29] | 0.95 [0.72-1.24] | | 1.04 [0.80-1.35] | 173 | 177 | 166 | NA | 171 |
| U82 | 1.26 [1.21-1.32] | 1.26 [1.20-1.32] | 1.24 [1.19-1.30] | 0.99 [0.79-1.24] | 1.26 [1.20-1.31] | 1.21 [1.16-1.26] | 1.16 [1.11-1.22] | 1.15 [1.10-1.21] | 1.02 [0.80-1.29] | 1.23 [1.18-1.29] | 6,721 | 6,637 | 6,117 | 265 | 6,688 |
| U83 | 1.20 [1.13-1.27] | 1.22 [1.15-1.29] | 1.22 [1.15-1.29] | 1.01 [0.75-1.37] | 1.23 [1.17-1.30] | 1.16 [1.09-1.23] | 1.13 [1.06-1.19] | 1.13 [1.06-1.20] | 1.01 [0.74-1.38] | 1.21 [1.15-1.28] | 3,966 | 3,917 | 3,637 | 158 | 3,971 |
| U88 | 1.20 [1.06-1.35] | 1.16 [1.03-1.31] | 1.23 [1.09-1.40] | | 1.12 [0.99-1.26] | 1.15 [1.01-1.30] | 1.09 [0.96-1.24] | 1.16 [1.02-1.32] | | 1.08 [0.96-1.22] | 802 | 792 | 762 | NA | 798 |
| U89 | 1.24 [1.11-1.39] | 1.21 [1.08-1.36] | 1.21 [1.08-1.36] | | 1.24 [1.11-1.39] | 1.19 [1.06-1.33] | 1.12 [0.99-1.26] | 1.12 [0.99-1.26] | | 1.20 [1.07-1.35] | 925 | 895 | 846 | NA | 919 |
| External causes of morbidity (V00-Y99) | | | | | | | | | | | | | | | |
| V03 | 1.16 [1.03-1.30] | 1.10 [0.97-1.25] | 1.20 [1.05-1.37] | 1.07 [0.84-1.38] | 1.06 [0.94-1.20] | 1.13 [1.01-1.27] | 1.06 [0.93-1.20] | 1.16 [1.01-1.33] | 1.09 [0.84-1.41] | 1.05 [0.93-1.19] | 913 | 746 | 632 | 206 | 768 |
| V09 | 1.19 [0.97-1.46] | 1.28 [1.03-1.58] | 1.21 [0.98-1.51] | | 1.18 [0.97-1.45] | 1.10 [0.90-1.36] | 1.13 [0.91-1.41] | 1.10 [0.88-1.38] | | 1.13 [0.92-1.38] | 303 | 271 | 254 | NA | 290 |
| V13 | 0.82 [0.64-1.06] | 0.84 [0.64-1.10] | | | 0.76 [0.58-1.01] | 0.82 [0.64-1.06] | 0.87 [0.66-1.15] | | | 0.77 [0.58-1.03] | 178 | 154 | NA | NA | 131 |
| V18 | 1.02 [0.91-1.14] | 1.02 [0.90-1.17] | 1.07 [0.93-1.25] | 1.03 [0.87-1.24] | 1.03 [0.92-1.16] | 1.00 [0.89-1.12] | 1.02 [0.89-1.17] | 1.10 [0.94-1.28] | 0.99 [0.83-1.19] | 1.02 [0.91-1.15] | 1,051 | 653 | 498 | 451 | 849 |
| V19 | 1.09 [0.83-1.43] | 0.96 [0.68-1.34] | | | 1.06 [0.79-1.42] | 1.07 [0.82-1.41] | 0.98 [0.69-1.38] | | | 1.05 [0.78-1.42] | 174 | 103 | NA | NA | 141 |
| V23 | 0.93 [0.73-1.19] | 0.86 [0.67-1.10] | 0.80 [0.58-1.11] | | 0.80 [0.61-1.05] | 0.91 [0.71-1.17] | 0.81 [0.62-1.05] | 0.80 [0.58-1.11] | | 0.82 [0.62-1.08] | 194 | 168 | 101 | NA | 150 |
| V28 | 0.99 [0.81-1.21] | 0.98 [0.79-1.21] | 0.95 [0.73-1.23] | | 0.88 [0.71-1.10] | 0.94 [0.76-1.16] | 0.94 [0.75-1.18] | 0.88 [0.67-1.16] | | 0.87 [0.69-1.09] | 286 | 253 | 163 | NA | 230 |
| V43 | 1.11 [1.00-1.24] | 1.08 [0.97-1.20] | 1.12 [0.99-1.27] | 0.99 [0.71-1.38] | 1.04 [0.93-1.16] | 1.05 [0.95-1.18] | 1.02 [0.92-1.14] | 1.04 [0.92-1.19] | 0.99 [0.70-1.40] | 1.02 [0.91-1.14] | 1,016 | 1,011 | 687 | 116 | 883 |
| V47 | 1.14 [0.96-1.35] | 1.29 [1.09-1.52] | 1.30 [1.05-1.60] | | 1.28 [1.08-1.53] | 1.11 [0.93-1.32] | 1.25 [1.05-1.48] | 1.20 [0.97-1.49] | | 1.25 [1.04-1.49] | 424 | 415 | 282 | NA | 390 |
| V48 | 1.07 [0.93-1.22] | 1.03 [0.90-1.18] | 1.10 [0.95-1.28] | | 1.03 [0.89-1.18] | 1.03 [0.90-1.19] | 0.99 [0.86-1.13] | 1.05 [0.90-1.22] | | 1.01 [0.87-1.16] | 623 | 617 | 513 | NA | 560 |
| V49 | 1.06 [0.89-1.25] | 1.07 [0.91-1.27] | 1.05 [0.86-1.28] | | 1.11 [0.93-1.32] | 1.02 [0.86-1.21] | 1.01 [0.85-1.19] | 0.99 [0.81-1.22] | | 1.10 [0.92-1.31] | 420 | 424 | 280 | NA | 362 |
| V78 | 1.11 [0.92-1.35] | 1.13 [0.94-1.37] | 1.15 [0.95-1.40] | | 1.14 [0.94-1.38] | 1.07 [0.89-1.30] | 1.08 [0.89-1.31] | 1.09 [0.90-1.33] | | 1.12 [0.92-1.36] | 317 | 313 | 300 | NA | 302 |
| V80 | 0.93 [0.80-1.10] | 0.88 [0.75-1.04] | 0.84 [0.66-1.07] | 0.90 [0.65-1.24] | 0.89 [0.75-1.07] | 0.92 [0.78-1.08] | 0.87 [0.74-1.03] | 0.86 [0.67-1.10] | 0.89 [0.63-1.25] | 0.89 [0.75-1.07] | 428 | 412 | 165 | 123 | 341 |
| V89 | 1.24 [0.97-1.59] | 1.25 [0.97-1.60] | 1.54 [1.11-2.15] | | 1.07 [0.83-1.39] | 1.15 [0.89-1.48] | 1.13 [0.87-1.47] | 1.37 [0.97-1.94] | | 1.02 [0.78-1.34] | 188 | 187 | 107 | NA | 163 |
| W00 | 1.05 [0.95-1.17] | 1.04 [0.93-1.16] | 1.06 [0.95-1.19] | | 1.07 [0.95-1.20] | 1.04 [0.93-1.16] | 1.03 [0.92-1.15] | 1.05 [0.94-1.18] | | 1.05 [0.94-1.18] | 938 | 942 | 832 | NA | 806 |
| W01 | 1.11 [1.09-1.14] | 1.12 [1.09-1.14] | 1.11 [1.09-1.14] | 1.02 [0.91-1.13] | 1.10 [1.08-1.13] | 1.08 [1.05-1.10] | 1.07 [1.05-1.10] | 1.07 [1.04-1.09] | 1.01 [0.91-1.13] | 1.09 [1.06-1.11] | 23,340 | 22,519 | 21,652 | 1,105 | 21,582 |
| W02 | 1.00 [0.85-1.18] | 1.09 [0.88-1.35] | 1.10 [0.81-1.48] | 0.97 [0.76-1.23] | 0.94 [0.78-1.13] | 0.98 [0.82-1.16] | 1.07 [0.86-1.33] | 1.07 [0.78-1.47] | 0.98 [0.77-1.25] | 0.92 [0.76-1.10] | 445 | 250 | 126 | 236 | 354 |
| W03 | 1.02 [0.89-1.17] | 0.99 [0.84-1.16] | 1.04 [0.85-1.28] | 0.95 [0.78-1.17] | 0.95 [0.82-1.11] | 1.01 [0.88-1.16] | 0.95 [0.80-1.12] | 1.00 [0.81-1.23] | 0.95 [0.77-1.17] | 0.95 [0.81-1.10] | 659 | 429 | 257 | 327 | 523 |
| W04 | 0.92 [0.69-1.23] | | | | 1.07 [0.80-1.44] | 0.93 [0.69-1.25] | | | | 1.13 [0.84-1.53] | 134 | NA | NA | NA | 133 |
| W05 | 1.15 [1.00-1.34] | 1.18 [1.02-1.37] | 1.19 [1.03-1.38] | | 1.13 [0.98-1.30] | 1.12 [0.96-1.30] | 1.14 [0.98-1.32] | 1.11 [0.96-1.29] | | 1.12 [0.97-1.29] | 551 | 549 | 537 | NA | 550 |
| W06 | 1.10 [1.06-1.16] | 1.11 [1.06-1.16] | 1.11 [1.06-1.16] | 1.05 [0.85-1.30] | 1.11 [1.06-1.16] | 1.08 [1.03-1.13] | 1.06 [1.01-1.11] | 1.06 [1.01-1.11] | 1.04 [0.84-1.29] | 1.09 [1.05-1.15] | 6,095 | 5,838 | 5,790 | 270 | 5,961 |
| W07 | 1.13 [1.06-1.20] | 1.13 [1.06-1.20] | 1.14 [1.07-1.21] | 1.01 [0.82-1.25] | 1.10 [1.03-1.17] | 1.10 [1.04-1.17] | 1.08 [1.02-1.16] | 1.09 [1.02-1.16] | 1.04 [0.84-1.28] | 1.09 [1.02-1.16] | 3,396 | 3,132 | 3,091 | 278 | 3,259 |
| W08 | 1.20 [1.07-1.34] | 1.17 [1.02-1.33] | 1.15 [1.01-1.31] | 0.98 [0.77-1.24] | 1.14 [1.02-1.28] | 1.16 [1.03-1.30] | 1.09 [0.96-1.25] | 1.08 [0.94-1.23] | 0.97 [0.76-1.23] | 1.12 [1.00-1.26] | 956 | 753 | 726 | 210 | 914 |
| W09 | 0.98 [0.88-1.09] | | | 0.97 [0.86-1.09] | 1.00 [0.88-1.13] | 0.96 [0.86-1.08] | | | 0.96 [0.85-1.08] | 0.99 [0.87-1.12] | 1,045 | NA | NA | 979 | 903 |
| W10 | 1.10 [1.07-1.14] | 1.11 [1.07-1.15] | 1.12 [1.08-1.17] | 0.97 [0.84-1.12] | 1.09 [1.05-1.13] | 1.07 [1.03-1.11] | 1.05 [1.02-1.09] | 1.07 [1.03-1.11] | 0.94 [0.81-1.09] | 1.07 [1.03-1.11] | 9,070 | 8,726 | 8,027 | 569 | 8,461 |
| W11 | 1.02 [0.92-1.12] | 1.04 [0.94-1.15] | 1.00 [0.90-1.11] | | 1.03 [0.92-1.14] | 0.98 [0.89-1.09] | 1.01 [0.92-1.12] | 0.96 [0.87-1.07] | | 1.01 [0.90-1.12] | 1,096 | 1,061 | 1,007 | NA | 935 |
| W13 | 1.10 [0.94-1.28] | 1.01 [0.84-1.20] | 1.04 [0.85-1.28] | 1.03 [0.80-1.33] | 1.06 [0.90-1.24] | 1.07 [0.91-1.25] | 0.96 [0.80-1.15] | 1.00 [0.81-1.23] | 1.03 [0.79-1.34] | 1.06 [0.90-1.25] | 514 | 363 | 263 | 193 | 437 |
| W14 | 1.13 [0.82-1.57] | | | | | 1.13 [0.81-1.57] | | | | | 124 | NA | NA | NA | NA |
| W17 | 1.08 [0.97-1.19] | 1.13 [1.01-1.27] | 1.14 [1.01-1.29] | 1.00 [0.83-1.20] | 1.06 [0.96-1.18] | 1.06 [0.96-1.17] | 1.09 [0.97-1.22] | 1.09 [0.96-1.24] | 0.99 [0.82-1.20] | 1.04 [0.93-1.15] | 1,223 | 909 | 770 | 377 | 1,085 |
| W18 | 1.15 [1.12-1.18] | 1.16 [1.13-1.19] | 1.16 [1.13-1.19] | 0.95 [0.82-1.10] | 1.13 [1.10-1.16] | 1.12 [1.09-1.15] | 1.10 [1.07-1.13] | 1.10 [1.07-1.13] | 0.94 [0.81-1.09] | 1.11 [1.08-1.14] | 16,837 | 16,373 | 15,857 | 597 | 16,089 |
| W19 | 1.13 [1.11-1.15] | 1.13 [1.11-1.15] | 1.13 [1.11-1.15] | 0.97 [0.86-1.08] | 1.12 [1.10-1.14] | 1.10 [1.08-1.12] | 1.09 [1.07-1.11] | 1.08 [1.06-1.11] | 0.96 [0.85-1.07] | 1.11 [1.09-1.13] | 38,290 | 37,557 | 36,306 | 1,031 | 36,033 |
| W20 | 1.05 [0.91-1.20] | 1.02 [0.88-1.19] | 1.16 [0.98-1.37] | 0.90 [0.66-1.23] | 1.04 [0.89-1.20] | 1.01 [0.87-1.16] | 0.96 [0.82-1.12] | 1.09 [0.91-1.29] | 0.87 [0.63-1.20] | 0.97 [0.84-1.13] | 593 | 490 | 388 | 133 | 512 |
| W21 | 1.04 [0.84-1.28] | 0.94 [0.72-1.23] | | 0.94 [0.71-1.24] | 0.91 [0.73-1.15] | 0.97 [0.78-1.21] | 0.87 [0.66-1.15] | | 0.94 [0.71-1.26] | 0.87 [0.68-1.10] | 301 | 168 | NA | 185 | 242 |
| W22 | 1.13 [1.06-1.20] | 1.12 [1.05-1.19] | 1.13 [1.05-1.21] | 1.09 [0.96-1.23] | 1.10 [1.04-1.17] | 1.08 [1.02-1.15] | 1.04 [0.98-1.11] | 1.05 [0.98-1.13] | 1.07 [0.94-1.22] | 1.06 [1.00-1.13] | 3,559 | 2,946 | 2,430 | 836 | 3,332 |
| W23 | 1.08 [0.99-1.17] | 1.07 [0.96-1.19] | 1.09 [0.97-1.23] | 1.00 [0.88-1.15] | 1.02 [0.93-1.11] | 1.05 [0.97-1.15] | 1.03 [0.92-1.14] | 1.07 [0.94-1.20] | 1.00 [0.87-1.14] | 1.00 [0.92-1.10] | 1,671 | 1,042 | 813 | 717 | 1,445 |
| W25 | 1.06 [0.93-1.20] | 1.07 [0.94-1.21] | 1.14 [0.95-1.37] | 0.98 [0.78-1.24] | 1.09 [0.96-1.24] | 1.03 [0.90-1.17] | 0.99 [0.86-1.14] | 1.08 [0.89-1.30] | 0.98 [0.77-1.24] | 1.07 [0.93-1.22] | 784 | 683 | 341 | 251 | 687 |
| W26 | 1.00 [0.88-1.13] | 0.99 [0.87-1.13] | 1.04 [0.88-1.22] | 0.87 [0.66-1.14] | 0.94 [0.83-1.07] | 0.95 [0.84-1.08] | 0.93 [0.82-1.06] | 0.96 [0.81-1.13] | 0.87 [0.65-1.16] | 0.92 [0.81-1.05] | 808 | 755 | 427 | 169 | 694 |
| W27 | 0.98 [0.82-1.18] | 1.02 [0.85-1.22] | 1.04 [0.84-1.29] | | 0.98 [0.82-1.18] | 0.95 [0.79-1.14] | 0.95 [0.78-1.15] | 0.95 [0.76-1.19] | | 0.93 [0.77-1.13] | 358 | 326 | 230 | NA | 315 |
| W29 | 0.92 [0.78-1.10] | 0.87 [0.74-1.04] | 0.93 [0.77-1.13] | | 0.88 [0.74-1.05] | 0.91 [0.76-1.08] | 0.85 [0.72-1.02] | 0.92 [0.76-1.11] | | 0.87 [0.73-1.05] | 369 | 370 | 297 | NA | 330 |
| W31 | 0.90 [0.75-1.08] | 0.92 [0.76-1.12] | 0.94 [0.76-1.15] | | 0.94 [0.77-1.15] | 0.90 [0.75-1.09] | 0.95 [0.78-1.15] | 0.93 [0.75-1.14] | | 0.94 [0.76-1.15] | 328 | 301 | 249 | NA | 262 |
| W44 | 1.11 [1.03-1.20] | 1.23 [1.12-1.35] | 1.22 [1.10-1.36] | 0.97 [0.86-1.09] | 1.09 [1.01-1.18] | 1.06 [0.98-1.15] | 1.15 [1.04-1.27] | 1.15 [1.03-1.29] | 0.96 [0.85-1.08] | 1.05 [0.97-1.14] | 2,046 | 1,286 | 1,042 | 853 | 1,818 |

### Supplementary Table 4: Hazard ratios and events from all cohorts (excluding non-consulters)

| | Hazard ratio (99% confidence interval) | | | | | | | | | | Events (in exposed) | | | | |
|---|---|---|---|---|---|---|---|---|---|---|---|---|---|---|---|
| | crude | | | | | adjusted | | | | | | | | | |
| Outcome | any age | 18+ | 40+ | <18 | hosp. | any age | 18+ | 40+ | <18 | hosp. | any age | 18+ | 40+ | <18 | hosp. |
| W45 | 1.03 [0.89-1.18] | 1.07 [0.92-1.25] | 0.97 [0.81-1.17] | 1.28 [0.98-1.66] | 1.03 [0.89-1.19] | 1.00 [0.86-1.15] | 1.02 [0.87-1.19] | 0.93 [0.77-1.12] | 1.21 [0.92-1.59] | 1.00 [0.86-1.16] | 632 | 489 | 319 | 218 | 530 |
| W46 | 0.97 [0.69-1.36] | 1.12 [0.80-1.57] | | | 1.09 [0.78-1.52] | 0.93 [0.65-1.31] | 1.00 [0.71-1.43] | | | 1.10 [0.77-1.56] | 105 | 105 | NA | NA | 103 |
| W49 | 1.36 [1.04-1.77] | 1.28 [0.97-1.68] | 1.34 [0.98-1.84] | | 1.35 [1.03-1.76] | 1.34 [1.02-1.76] | 1.21 [0.91-1.61] | 1.29 [0.94-1.77] | | 1.29 [0.98-1.70] | 179 | 158 | 121 | NA | 166 |
| W50 | 1.11 [0.96-1.28] | 1.13 [0.96-1.33] | 1.15 [0.90-1.47] | 1.15 [0.93-1.42] | 1.09 [0.94-1.26] | 1.08 [0.94-1.25] | 1.03 [0.87-1.22] | 1.08 [0.84-1.39] | 1.13 [0.90-1.40] | 1.08 [0.92-1.25] | 685 | 494 | 190 | 326 | 558 |
| W51 | 0.97 [0.82-1.14] | 0.96 [0.79-1.18] | 1.01 [0.76-1.34] | 1.07 [0.85-1.35] | 1.08 [0.91-1.28] | 0.95 [0.80-1.12] | 0.90 [0.73-1.11] | 0.98 [0.73-1.30] | 1.03 [0.81-1.32] | 1.05 [0.88-1.26] | 476 | 274 | 142 | 259 | 415 |
| W54 | 1.03 [0.93-1.15] | 1.05 [0.94-1.17] | 1.11 [0.98-1.26] | 0.95 [0.75-1.20] | 1.04 [0.93-1.16] | 0.99 [0.89-1.10] | 0.98 [0.87-1.10] | 1.03 [0.91-1.18] | 0.94 [0.73-1.19] | 1.00 [0.89-1.12] | 1,020 | 882 | 683 | 219 | 914 |
| W55 | 0.93 [0.80-1.07] | 0.95 [0.82-1.10] | 0.99 [0.84-1.16] | | 0.95 [0.82-1.10] | 0.86 [0.74-0.99] | 0.87 [0.75-1.01] | 0.90 [0.76-1.06] | | 0.91 [0.78-1.05] | 530 | 526 | 411 | NA | 485 |
| W57 | 1.38 [1.22-1.56] | 1.34 [1.18-1.52] | 1.33 [1.14-1.54] | 1.25 [0.88-1.77] | 1.33 [1.17-1.51] | 1.31 [1.15-1.49] | 1.24 [1.09-1.41] | 1.24 [1.06-1.44] | 1.22 [0.85-1.75] | 1.27 [1.12-1.45] | 798 | 765 | 525 | 113 | 723 |
| W64 | 0.91 [0.69-1.20] | 1.05 [0.80-1.38] | 1.01 [0.75-1.36] | | 1.03 [0.79-1.36] | 0.90 [0.68-1.19] | 0.99 [0.75-1.31] | 0.98 [0.72-1.33] | | 1.04 [0.79-1.38] | 156 | 156 | 130 | NA | 154 |
| W78 | 1.03 [0.90-1.19] | 1.04 [0.90-1.19] | 1.03 [0.89-1.19] | | 1.04 [0.90-1.20] | 1.03 [0.89-1.18] | 1.02 [0.88-1.18] | 1.01 [0.87-1.17] | | 1.06 [0.92-1.22] | 602 | 592 | 570 | NA | 587 |
| W79 | 1.10 [0.98-1.23] | 1.15 [1.02-1.29] | 1.09 [0.97-1.23] | | 1.12 [1.00-1.26] | 1.08 [0.96-1.21] | 1.11 [0.98-1.25] | 1.06 [0.93-1.20] | | 1.11 [0.99-1.25] | 892 | 847 | 785 | NA | 864 |
| W80 | 1.19 [0.99-1.42] | 1.15 [0.95-1.38] | 1.21 [1.01-1.46] | | 1.14 [0.95-1.36] | 1.14 [0.95-1.37] | 1.09 [0.91-1.32] | 1.13 [0.94-1.37] | | 1.10 [0.92-1.32] | 370 | 364 | 354 | NA | 359 |
| W84 | 1.01 [0.84-1.21] | 0.98 [0.82-1.18] | 1.00 [0.83-1.20] | | 0.97 [0.81-1.16] | 0.99 [0.82-1.19] | 0.93 [0.77-1.13] | 0.96 [0.80-1.16] | | 0.97 [0.81-1.17] | 339 | 335 | 334 | NA | 340 |
| X10 | 1.07 [0.90-1.28] | 1.15 [0.90-1.48] | 1.26 [0.95-1.68] | 1.06 [0.84-1.34] | 1.01 [0.84-1.22] | 1.06 [0.89-1.27] | 1.08 [0.84-1.40] | 1.19 [0.89-1.59] | 1.06 [0.83-1.34] | 0.99 [0.83-1.20] | 391 | 192 | 144 | 224 | 354 |
| X11 | 0.93 [0.67-1.28] | | | | | 0.91 [0.65-1.26] | | | | | 111 | NA | NA | NA | NA |
| X12 | 0.94 [0.75-1.18] | 1.03 [0.78-1.35] | 1.07 [0.79-1.46] | | 0.97 [0.76-1.23] | 0.92 [0.73-1.16] | 0.97 [0.73-1.29] | 1.00 [0.73-1.37] | | 0.93 [0.73-1.19] | 236 | 157 | 123 | NA | 202 |
| X15 | 1.06 [0.80-1.40] | | | | 1.01 [0.76-1.35] | 1.04 [0.78-1.38] | | | | 1.02 [0.76-1.37] | 146 | NA | NA | NA | 144 |
| X16 | 0.95 [0.72-1.24] | 0.95 [0.69-1.29] | 0.96 [0.71-1.31] | | 0.93 [0.70-1.23] | 0.93 [0.71-1.22] | 0.90 [0.66-1.24] | 0.92 [0.68-1.26] | | 0.91 [0.68-1.20] | 156 | 127 | 120 | NA | 140 |
| X23 | 1.02 [0.77-1.36] | 1.06 [0.79-1.42] | 1.01 [0.74-1.39] | | 1.07 [0.79-1.44] | 0.97 [0.72-1.30] | 1.00 [0.74-1.34] | 0.96 [0.69-1.33] | | 1.00 [0.74-1.36] | 148 | 136 | 108 | NA | 122 |
| X31 | 1.09 [0.83-1.42] | 0.92 [0.71-1.20] | 0.98 [0.75-1.27] | | 0.94 [0.72-1.23] | 1.10 [0.84-1.44] | 0.91 [0.70-1.19] | 0.99 [0.76-1.29] | | 0.98 [0.75-1.28] | 171 | 170 | 169 | NA | 161 |
| X40 | 1.07 [0.98-1.17] | 1.06 [0.96-1.17] | 1.18 [1.03-1.33] | 1.01 [0.85-1.19] | 1.01 [0.92-1.10] | 1.02 [0.93-1.12] | 0.94 [0.85-1.05] | 1.02 [0.90-1.17] | 1.02 [0.86-1.22] | 0.99 [0.90-1.08] | 1,430 | 1,164 | 705 | 453 | 1,340 |
| X41 | 1.11 [0.99-1.23] | 1.15 [1.03-1.29] | 1.14 [1.00-1.30] | 0.95 [0.73-1.23] | 1.03 [0.92-1.14] | 1.04 [0.94-1.17] | 1.00 [0.89-1.12] | 1.01 [0.88-1.16] | 0.97 [0.74-1.26] | 1.02 [0.91-1.14] | 987 | 922 | 613 | 184 | 955 |
| X42 | 1.17 [1.05-1.30] | 1.17 [1.05-1.30] | 1.23 [1.09-1.39] | 0.98 [0.70-1.37] | 1.10 [0.99-1.22] | 1.07 [0.96-1.19] | 0.99 [0.89-1.11] | 1.07 [0.94-1.21] | 0.99 [0.70-1.40] | 1.06 [0.95-1.18] | 1,058 | 1,045 | 793 | 116 | 1,065 |
| X43 | 1.00 [0.80-1.26] | 1.04 [0.81-1.32] | 0.99 [0.77-1.29] | | 0.98 [0.78-1.23] | 0.98 [0.77-1.23] | 0.97 [0.76-1.25] | 0.91 [0.70-1.19] | | 0.97 [0.77-1.22] | 222 | 194 | 169 | NA | 212 |
| X44 | 1.09 [1.00-1.19] | 1.11 [1.01-1.23] | 1.14 [1.03-1.27] | 0.96 [0.79-1.16] | 1.09 [1.00-1.19] | 1.03 [0.94-1.13] | 0.99 [0.89-1.09] | 1.02 [0.91-1.13] | 0.97 [0.80-1.18] | 1.06 [0.97-1.16] | 1,565 | 1,291 | 1,118 | 330 | 1,524 |
| X45 | 1.16 [0.96-1.40] | 1.03 [0.86-1.25] | 1.04 [0.80-1.36] | | 0.97 [0.80-1.17] | 1.08 [0.89-1.32] | 0.88 [0.72-1.07] | 0.90 [0.68-1.19] | | 0.96 [0.79-1.16] | 335 | 325 | 155 | NA | 306 |
| X47 | 1.03 [0.77-1.38] | 0.99 [0.74-1.34] | 1.14 [0.82-1.56] | | 1.01 [0.75-1.35] | 1.03 [0.77-1.38] | 0.91 [0.67-1.24] | 1.11 [0.80-1.53] | | 0.98 [0.72-1.33] | 136 | 131 | 117 | NA | 128 |
| X49 | 1.24 [1.06-1.45] | 1.20 [0.98-1.47] | 1.13 [0.90-1.42] | 1.41 [1.11-1.78] | 1.28 [1.09-1.49] | 1.17 [1.00-1.37] | 1.07 [0.87-1.32] | 1.06 [0.84-1.34] | 1.34 [1.05-1.70] | 1.20 [1.02-1.41] | 527 | 304 | 217 | 248 | 496 |
| X50 | 1.07 [0.99-1.16] | 1.11 [1.02-1.20] | 1.07 [0.98-1.17] | 1.08 [0.84-1.37] | 1.05 [0.97-1.14] | 1.01 [0.93-1.10] | 1.03 [0.95-1.12] | 1.00 [0.91-1.10] | 1.02 [0.79-1.32] | 1.01 [0.93-1.09] | 1,878 | 1,798 | 1,374 | 227 | 1,687 |
| X51 | 1.02 [0.77-1.33] | 1.01 [0.77-1.33] | 1.07 [0.79-1.44] | | 1.04 [0.78-1.38] | 1.00 [0.76-1.32] | 1.00 [0.76-1.31] | 1.01 [0.75-1.37] | | 1.04 [0.78-1.38] | 142 | 146 | 118 | NA | 130 |
| X58 | 1.36 [1.20-1.55] | 1.40 [1.22-1.61] | 1.25 [1.07-1.45] | 2.16 [1.53-3.05] | 1.29 [1.13-1.47] | 1.26 [1.10-1.44] | 1.29 [1.12-1.49] | 1.17 [1.00-1.36] | 1.91 [1.32-2.76] | 1.23 [1.08-1.41] | 773 | 679 | 542 | 152 | 736 |
| X59 | 1.18 [1.14-1.22] | 1.18 [1.14-1.23] | 1.20 [1.15-1.25] | 1.06 [0.95-1.18] | 1.16 [1.12-1.20] | 1.13 [1.09-1.18] | 1.11 [1.07-1.15] | 1.13 [1.09-1.18] | 1.04 [0.93-1.17] | 1.13 [1.09-1.18] | 9,634 | 8,773 | 7,825 | 1,161 | 8,935 |
| X60 | 1.10 [1.05-1.16] | 1.07 [1.01-1.13] | 1.22 [1.13-1.32] | 0.93 [0.84-1.04] | 1.05 [0.99-1.11] | 1.05 [0.99-1.11] | 0.94 [0.89-1.00] | 1.08 [1.00-1.17] | 0.94 [0.84-1.06] | 1.04 [0.98-1.10] | 4,252 | 4,065 | 1,812 | 1,141 | 3,773 |
| X61 | 1.15 [1.08-1.21] | 1.14 [1.08-1.20] | 1.23 [1.13-1.33] | 0.91 [0.79-1.04] | 1.07 [1.01-1.13] | 1.07 [1.01-1.14] | 0.98 [0.93-1.04] | 1.06 [0.98-1.15] | 0.95 [0.82-1.11] | 1.06 [1.00-1.13] | 3,720 | 3,786 | 1,811 | 657 | 3,405 |
| X62 | 1.17 [1.08-1.26] | 1.14 [1.06-1.23] | 1.19 [1.07-1.32] | 0.99 [0.82-1.20] | 1.07 [1.00-1.16] | 1.06 [0.98-1.15] | 0.96 [0.89-1.04] | 1.00 [0.89-1.12] | 1.01 [0.83-1.23] | 1.04 [0.97-1.13] | 2,099 | 2,140 | 1,029 | 360 | 2,013 |
| X63 | 1.12 [0.97-1.31] | 1.12 [0.97-1.30] | 1.16 [0.94-1.43] | | 1.07 [0.93-1.24] | 1.07 [0.92-1.25] | 0.97 [0.84-1.14] | 1.01 [0.81-1.25] | | 1.08 [0.93-1.26] | 530 | 542 | 265 | NA | 517 |
| X64 | 1.18 [1.08-1.28] | 1.11 [1.03-1.21] | 1.26 [1.13-1.41] | 1.05 [0.87-1.26] | 1.10 [1.01-1.19] | 1.09 [1.00-1.19] | 0.96 [0.88-1.04] | 1.09 [0.97-1.22] | 1.01 [0.83-1.23] | 1.08 [1.00-1.18] | 1,809 | 1,700 | 930 | 400 | 1,675 |
| X65 | 1.12 [1.05-1.21] | 1.08 [1.01-1.16] | 1.21 [1.10-1.33] | 0.95 [0.79-1.13] | 1.07 [1.00-1.15] | 1.05 [0.98-1.13] | 0.93 [0.87-1.00] | 1.02 [0.92-1.13] | 1.06 [0.88-1.29] | 1.07 [0.99-1.15] | 2,511 | 2,604 | 1,232 | 382 | 2,353 |
| X69 | 0.93 [0.71-1.23] | 0.78 [0.58-1.05] | | | 0.92 [0.69-1.22] | 0.88 [0.67-1.18] | 0.68 [0.50-0.93] | | | 0.94 [0.70-1.25] | 155 | 118 | NA | NA | 140 |
| X70 | 1.01 [0.72-1.40] | | | | 0.92 [0.67-1.28] | 1.01 [0.71-1.42] | | | | 0.95 [0.67-1.33] | 106 | NA | NA | NA | 104 |
| X78 | 1.08 [0.97-1.21] | 1.08 [0.96-1.21] | 1.18 [0.99-1.41] | 0.98 [0.80-1.20] | 1.06 [0.95-1.19] | 0.97 [0.86-1.10] | 0.90 [0.80-1.02] | 0.99 [0.83-1.20] | 0.99 [0.80-1.23] | 1.02 [0.91-1.15] | 966 | 850 | 367 | 317 | 881 |
| X84 | 1.06 [0.79-1.42] | 0.98 [0.73-1.32] | | | 1.10 [0.83-1.47] | 0.95 [0.69-1.29] | 0.83 [0.60-1.13] | | | 1.03 [0.76-1.40] | 147 | 127 | NA | NA | 143 |
| X99 | 1.13 [0.91-1.40] | 1.02 [0.81-1.29] | | | 0.97 [0.76-1.22] | 1.11 [0.89-1.39] | 0.97 [0.76-1.23] | | | 0.97 [0.76-1.24] | 281 | 228 | NA | NA | 210 |
| Y00 | 1.00 [0.77-1.30] | 0.99 [0.76-1.30] | | | 0.92 [0.70-1.20] | 0.92 [0.70-1.21] | 0.90 [0.68-1.19] | | | 0.89 [0.67-1.18] | 169 | 159 | NA | NA | 152 |
| Y04 | 1.06 [0.96-1.16] | 1.05 [0.96-1.16] | 1.15 [1.00-1.32] | 0.91 [0.75-1.10] | 1.02 [0.93-1.13] | 1.01 [0.92-1.12] | 0.96 [0.86-1.06] | 1.04 [0.90-1.20] | 0.98 [0.80-1.21] | 1.02 [0.92-1.13] | 1,401 | 1,331 | 584 | 345 | 1,207 |
| Y07 | 0.94 [0.69-1.28] | | | | 0.88 [0.66-1.18] | 0.88 [0.64-1.20] | | | | 0.87 [0.64-1.17] | 131 | NA | NA | NA | 120 |
| Y09 | 1.09 [0.86-1.38] | 1.01 [0.80-1.29] | 1.17 [0.84-1.63] | | 1.09 [0.85-1.39] | 1.04 [0.82-1.32] | 0.92 [0.72-1.18] | 1.05 [0.74-1.47] | | 1.07 [0.83-1.36] | 219 | 201 | 109 | NA | 190 |
| Y40 | 1.43 [1.35-1.52] | 1.41 [1.33-1.50] | 1.44 [1.35-1.54] | 1.09 [0.84-1.42] | 1.36 [1.28-1.44] | 1.33 [1.25-1.42] | 1.28 [1.21-1.37] | 1.32 [1.23-1.41] | 1.09 [0.83-1.43] | 1.29 [1.22-1.37] | 3,502 | 3,418 | 3,069 | 191 | 3,384 |
| Y41 | 1.30 [1.14-1.48] | 1.27 [1.11-1.45] | 1.32 [1.14-1.51] | | 1.21 [1.06-1.38] | 1.20 [1.05-1.37] | 1.16 [1.01-1.33] | 1.19 [1.03-1.37] | | 1.13 [0.99-1.30] | 700 | 692 | 604 | NA | 662 |
| Y42 | 1.27 [1.19-1.36] | 1.23 [1.15-1.31] | 1.24 [1.16-1.33] | | 1.22 [1.14-1.30] | 1.15 [1.07-1.23] | 1.08 [1.01-1.15] | 1.09 [1.02-1.17] | | 1.13 [1.06-1.21] | 2,822 | 2,793 | 2,555 | NA | 2,773 |
| Y43 | 1.13 [1.08-1.19] | 1.14 [1.08-1.19] | 1.14 [1.09-1.20] | 1.13 [0.85-1.49] | 1.09 [1.04-1.15] | 1.11 [1.06-1.17] | 1.11 [1.05-1.17] | 1.12 [1.06-1.17] | 1.14 [0.86-1.53] | 1.09 [1.04-1.15] | 4,678 | 4,587 | 4,253 | 173 | 4,330 |
| Y44 | 1.17 [1.08-1.27] | 1.19 [1.10-1.28] | 1.16 [1.07-1.26] | | 1.13 [1.05-1.23] | 1.13 [1.05-1.22] | 1.12 [1.03-1.21] | 1.09 [1.01-1.18] | | 1.10 [1.02-1.19] | 1,923 | 1,922 | 1,854 | NA | 1,879 |
| Y45 | 1.24 [1.18-1.31] | 1.24 [1.18-1.31] | 1.24 [1.17-1.31] | 1.15 [0.80-1.67] | 1.19 [1.13-1.25] | 1.18 [1.11-1.24] | 1.14 [1.08-1.20] | 1.15 [1.08-1.21] | 1.08 [0.72-1.61] | 1.15 [1.09-1.21] | 4,335 | 4,354 | 3,954 | 104 | 4,160 |
| Y46 | 1.27 [1.11-1.46] | 1.24 [1.07-1.42] | 1.27 [1.10-1.46] | | 1.20 [1.05-1.38] | 1.20 [1.04-1.38] | 1.14 [0.99-1.32] | 1.17 [1.01-1.35] | | 1.17 [1.02-1.35] | 617 | 589 | 572 | NA | 608 |
| Y47 | 1.16 [0.94-1.41] | 1.34 [1.09-1.65] | 1.28 [1.03-1.58] | | 1.18 [0.96-1.44] | 1.08 [0.88-1.33] | 1.25 [1.01-1.54] | 1.21 [0.97-1.50] | | 1.15 [0.94-1.41] | 296 | 287 | 256 | NA | 289 |
| Y48 | 1.07 [0.88-1.30] | 1.15 [0.94-1.41] | 1.11 [0.89-1.39] | | 1.06 [0.87-1.28] | 1.00 [0.82-1.22] | 1.06 [0.86-1.30] | 1.02 [0.82-1.28] | | 1.03 [0.84-1.25] | 294 | 289 | 230 | NA | 287 |
| Y49 | 1.25 [1.13-1.39] | 1.30 [1.18-1.45] | 1.25 [1.12-1.39] | | 1.27 [1.14-1.40] | 1.23 [1.10-1.36] | 1.23 [1.11-1.37] | 1.19 [1.07-1.33] | | 1.26 [1.13-1.40] | 1,081 | 1,086 | 990 | NA | 1,047 |
| Y51 | 1.11 [1.02-1.21] | 1.15 [1.06-1.25] | 1.13 [1.04-1.23] | | 1.11 [1.02-1.21] | 1.10 [1.01-1.19] | 1.11 [1.02-1.21] | 1.09 [1.00-1.19] | | 1.12 [1.03-1.21] | 1,663 | 1,665 | 1,630 | NA | 1,611 |
| Y52 | 1.20 [1.14-1.27] | 1.24 [1.17-1.30] | 1.19 [1.13-1.25] | | 1.18 [1.12-1.25] | 1.16 [1.10-1.22] | 1.16 [1.10-1.22] | 1.12 [1.06-1.18] | | 1.16 [1.10-1.22] | 4,300 | 4,307 | 4,266 | NA | 4,218 |
| Y53 | 1.32 [1.17-1.49] | 1.28 [1.13-1.44] | 1.32 [1.16-1.49] | | 1.20 [1.06-1.36] | 1.23 [1.08-1.39] | 1.14 [1.01-1.30] | 1.20 [1.06-1.37] | | 1.14 [1.00-1.29] | 816 | 818 | 782 | NA | 794 |
| Y54 | 1.23 [1.16-1.30] | 1.26 [1.18-1.33] | 1.25 [1.18-1.33] | | 1.22 [1.15-1.29] | 1.18 [1.11-1.25] | 1.17 [1.10-1.24] | 1.17 [1.10-1.24] | | 1.18 [1.11-1.25] | 3,479 | 3,486 | 3,452 | NA | 3,355 |
| Y55 | 1.26 [1.05-1.50] | 1.20 [1.01-1.44] | 1.43 [1.12-1.83] | | 1.17 [0.98-1.40] | 1.12 [0.94-1.35] | 1.05 [0.87-1.26] | 1.21 [0.93-1.57] | | 1.03 [0.86-1.25] | 396 | 394 | 190 | NA | 356 |
| Y56 | 1.67 [1.38-2.01] | 1.63 [1.35-1.96] | 1.70 [1.39-2.09] | | 1.58 [1.31-1.90] | 1.53 [1.26-1.85] | 1.47 [1.21-1.78] | 1.51 [1.22-1.86] | | 1.49 [1.23-1.80] | 382 | 372 | 320 | NA | 355 |
| Y57 | 1.24 [1.15-1.35] | 1.23 [1.13-1.33] | 1.25 [1.15-1.36] | | 1.23 [1.14-1.34] | 1.16 [1.07-1.26] | 1.12 [1.03-1.22] | 1.14 [1.05-1.24] | | 1.18 [1.09-1.28] | 1,862 | 1,861 | 1,749 | NA | 1,827 |
| Y59 | 1.28 [0.98-1.66] | 1.18 [0.89-1.57] | 1.25 [0.92-1.70] | | 1.37 [1.05-1.79] | 1.20 [0.92-1.58] | 1.09 [0.81-1.46] | 1.14 [0.83-1.57] | | 1.27 [0.96-1.66] | 185 | 142 | 121 | NA | 172 |
| Y60 | 1.15 [1.06-1.24] | 1.16 [1.07-1.25] | 1.20 [1.11-1.30] | | 1.15 [1.07-1.24] | 1.11 [1.02-1.19] | 1.09 [1.01-1.18] | 1.14 [1.05-1.24] | | 1.12 [1.04-1.21] | 2,050 | 2,048 | 1,848 | NA | 1,931 |
| Y65 | 1.06 [0.86-1.31] | 1.08 [0.87-1.34] | 1.13 [0.91-1.41] | | 1.04 [0.84-1.29] | 1.02 [0.82-1.26] | 1.01 [0.81-1.26] | 1.06 [0.85-1.33] | | 1.03 [0.83-1.28] | 256 | 251 | 229 | NA | 246 |
| Y71 | 1.32 [0.99-1.77] | 1.31 [0.98-1.75] | 1.37 [1.02-1.85] | | 1.28 [0.96-1.71] | 1.29 [0.96-1.73] | 1.23 [0.92-1.65] | 1.31 [0.97-1.77] | | 1.25 [0.93-1.68] | 139 | 140 | 130 | NA | 136 |
| Y77 | 1.00 [0.74-1.36] | 0.96 [0.71-1.30] | 0.94 [0.70-1.26] | | 1.07 [0.79-1.44] | 0.99 [0.73-1.34] | 0.91 [0.67-1.23] | 0.91 [0.68-1.23] | | 1.04 [0.77-1.41] | 121 | 118 | 118 | NA | 117 |
| Y79 | 1.03 [0.84-1.26] | 1.07 [0.87-1.31] | 1.04 [0.85-1.28] | | 1.06 [0.86-1.30] | 0.99 [0.80-1.21] | 1.01 [0.82-1.24] | 1.00 [0.81-1.23] | | 1.01 [0.82-1.25] | 260 | 258 | 254 | NA | 254 |
| Y82 | 1.27 [0.91-1.78] | | | | 1.34 [0.97-1.84] | 1.23 [0.88-1.72] | | | | 1.32 [0.96-1.83] | 101 | NA | NA | NA | 106 |
| Y83 | 1.17 [1.15-1.19] | 1.17 [1.15-1.19] | 1.16 [1.13-1.18] | 1.12 [1.03-1.22] | 1.13 [1.11-1.15] | 1.12 [1.10-1.14] | 1.10 [1.07-1.12] | 1.09 [1.07-1.11] | 1.11 [1.02-1.22] | 1.10 [1.08-1.12] | 34,875 | 34,170 | 29,806 | 1,936 | 33,031 |
| Y84 | 1.14 [1.09-1.18] | 1.15 [1.11-1.19] | 1.13 [1.08-1.17] | 0.98 [0.75-1.27] | 1.10 [1.06-1.15] | 1.10 [1.06-1.15] | 1.09 [1.05-1.13] | 1.06 [1.02-1.11] | 0.95 [0.72-1.25] | 1.09 [1.05-1.13] | 8,348 | 8,265 | 7,747 | 205 | 8,111 |
| Y85 | 1.17 [1.03-1.34] | 1.22 [1.07-1.39] | 1.26 [1.08-1.46] | | 1.08 [0.95-1.24] | 1.11 [0.97-1.27] | 1.11 [0.97-1.27] | 1.17 [1.00-1.37] | | 1.04 [0.91-1.19] | 682 | 677 | 520 | NA | 623 |
| Y86 | 1.12 [1.01-1.24] | 1.12 [1.01-1.24] | 1.13 [1.01-1.27] | 1.20 [0.88-1.65] | 1.09 [0.99-1.21] | 1.08 [0.98-1.20] | 1.04 [0.94-1.16] | 1.07 [0.95-1.20] | 1.19 [0.85-1.65] | 1.06 [0.95-1.17] | 1,172 | 1,110 | 841 | 141 | 1,073 |
| Y87 | 1.27 [0.97-1.65] | 1.16 [0.89-1.52] | | | 1.12 [0.85-1.46] | 1.19 [0.90-1.56] | 1.01 [0.76-1.34] | | | 1.07 [0.80-1.42] | 178 | 175 | NA | NA | 162 |
| Y88 | 1.01 [0.82-1.23] | 1.12 [0.91-1.38] | 1.09 [0.88-1.35] | | 0.92 [0.76-1.12] | 0.94 [0.76-1.16] | 1.01 [0.82-1.25] | 0.95 [0.76-1.19] | | 0.86 [0.70-1.06] | 265 | 265 | 232 | NA | 259 |
| Y90 | 1.25 [0.92-1.70] | 1.29 [0.96-1.73] | | | 1.01 [0.76-1.34] | 1.28 [0.93-1.74] | 1.24 [0.91-1.67] | | | 1.07 [0.80-1.43] | 130 | 137 | NA | NA | 129 |
| Y91 | 1.11 [0.88-1.39] | 0.97 [0.77-1.21] | 1.12 [0.87-1.45] | | 1.08 [0.86-1.35] | 1.06 [0.84-1.34] | 0.90 [0.71-1.13] | 1.01 [0.77-1.31] | | 1.08 [0.86-1.36] | 219 | 219 | 164 | NA | 204 |
| Y95 | 1.17 [1.14-1.20] | 1.16 [1.13-1.19] | 1.17 [1.14-1.20] | | 1.15 [1.13-1.18] | 1.12 [1.10-1.15] | 1.09 [1.06-1.12] | 1.09 [1.06-1.12] | | 1.13 [1.10-1.15] | 21,092 | 21,111 | 20,890 | NA | 21,025 |
| Factors influencing health status and contact with health services (Z00-Z99) | | | | | | | | | | | | | | | |
| Z00 | 1.12 [0.99-1.28] | 1.11 [0.97-1.27] | 1.18 [1.01-1.37] | | 1.06 [0.93-1.22] | 1.06 [0.93-1.22] | 1.05 [0.91-1.20] | 1.11 [0.95-1.30] | | 1.04 [0.90-1.19] | 653 | 636 | 488 | NA | 572 |
| Z01 | 1.97 [1.86-2.09] | 1.23 [1.14-1.33] | 1.28 [1.17-1.40] | 3.98 [3.62-4.38] | 1.92 [1.81-2.04] | 1.72 [1.62-1.83] | 1.11 [1.02-1.20] | 1.18 [1.08-1.30] | 3.14 [2.83-3.47] | 1.67 [1.57-1.77] | 4,661 | 2,018 | 1,434 | 2,782 | 4,087 |
| Z02 | 1.09 [1.02-1.17] | 1.09 [1.02-1.17] | 1.12 [1.04-1.20] | | 1.08 [1.00-1.15] | 1.05 [0.98-1.13] | 1.03 [0.96-1.10] | 1.06 [0.99-1.15] | | 1.05 [0.98-1.13] | 2,679 | 2,719 | 2,137 | NA | 2,456 |
| Z03 | 1.23 [1.20-1.25] | 1.22 [1.19-1.25] | 1.22 [1.19-1.25] | 1.35 [1.25-1.45] | 1.20 [1.17-1.22] | 1.16 [1.13-1.19] | 1.13 [1.10-1.15] | 1.13 [1.10-1.16] | 1.27 [1.17-1.37] | 1.15 [1.12-1.17] | 22,897 | 21,255 | 18,261 | 2,474 | 21,583 |
| Z04 | 1.14 [1.07-1.21] | 1.16 [1.09-1.23] | 1.21 [1.12-1.30] | 1.02 [0.88-1.18] | 1.12 [1.05-1.19] | 1.10 [1.04-1.17] | 1.08 [1.02-1.15] | 1.15 [1.07-1.24] | 1.02 [0.88-1.19] | 1.09 [1.03-1.16] | 3,339 | 3,015 | 2,076 | 561 | 2,990 |
| Z08 | 1.01 [0.97-1.06] | 1.01 [0.96-1.05] | 1.01 [0.97-1.05] | | 1.01 [0.97-1.06] | 1.01 [0.96-1.05] | 0.99 [0.95-1.03] | 0.99 [0.95-1.04] | | 1.01 [0.97-1.06] | 6,074 | 5,996 | 5,867 | NA | 5,865 |
| Z09 | 1.17 [1.13-1.20] | 1.16 [1.13-1.20] | 1.17 [1.13-1.21] | 1.09 [0.95-1.26] | 1.13 [1.10-1.16] | 1.11 [1.07-1.14] | 1.08 [1.05-1.11] | 1.09 [1.05-1.13] | 1.05 [0.90-1.22] | 1.09 [1.06-1.12] | 12,760 | 12,375 | 11,061 | 645 | 12,274 |
| Z11 | 1.12 [1.10-1.14] | 1.11 [1.09-1.13] | 1.13 [1.11-1.16] | 1.05 [1.00-1.11] | 1.10 [1.08-1.12] | 1.07 [1.05-1.09] | 1.03 [1.02-1.05] | 1.05 [1.03-1.07] | 1.05 [0.99-1.11] | 1.08 [1.06-1.10] | 40,378 | 39,851 | 30,858 | 4,427 | 39,570 |
| Z12 | 1.08 [1.04-1.12] | 1.07 [1.03-1.12] | 1.07 [1.03-1.11] | | 1.06 [1.02-1.11] | 1.06 [1.02-1.10] | 1.06 [1.02-1.10] | 1.05 [1.01-1.09] | | 1.04 [1.00-1.09] | 7,780 | 7,718 | 7,479 | NA | 6,871 |
| Z13 | 1.16 [1.14-1.19] | 1.16 [1.13-1.18] | 1.16 [1.13-1.19] | 1.31 [1.21-1.43] | 1.13 [1.10-1.15] | 1.11 [1.09-1.14] | 1.09 [1.06-1.11] | 1.09 [1.06-1.11] | 1.26 [1.15-1.37] | 1.09 [1.07-1.12] | 24,529 | 23,895 | 18,218 | 2,161 | 22,043 |
| Z20 | 1.15 [1.05-1.26] | 1.13 [1.03-1.25] | 1.12 [1.01-1.24] | 1.20 [0.89-1.63] | 1.12 [1.02-1.23] | 1.10 [1.00-1.21] | 1.05 [0.96-1.16] | 1.03 [0.93-1.15] | 1.19 [0.87-1.64] | 1.08 [0.99-1.19] | 1,470 | 1,397 | 1,211 | 144 | 1,420 |
| Z22 | 1.27 [1.24-1.31] | 1.28 [1.24-1.32] | 1.33 [1.29-1.39] | 0.93 [0.84-1.04] | 1.25 [1.21-1.28] | 1.23 [1.19-1.26] | 1.21 [1.17-1.24] | 1.24 [1.20-1.29] | 0.94 [0.83-1.06] | 1.22 [1.18-1.25] | 14,010 | 14,384 | 8,745 | 1,011 | 13,522 |
| Z24 | 0.99 [0.77-1.27] | 1.09 [0.87-1.38] | | | 0.95 [0.73-1.23] | 1.03 [0.80-1.34] | 1.10 [0.87-1.39] | | | 0.95 [0.73-1.25] | 183 | 216 | NA | NA | 161 |
| Z25 | 2.61 [1.80-3.78] | | | 3.84 [2.29-6.41] | 1.94 [1.36-2.76] | 2.29 [1.56-3.37] | | | 2.95 [1.70-5.12] | 1.72 [1.19-2.49] | 141 | NA | NA | 103 | 128 |
| Z27 | 1.85 [1.61-2.12] | 0.99 [0.85-1.17] | | 3.29 [2.68-4.03] | 1.66 [1.44-1.92] | 1.73 [1.50-2.00] | 0.99 [0.84-1.17] | | 2.78 [2.24-3.44] | 1.56 [1.34-1.81] | 725 | 439 | NA | 470 | 626 |
| Z29 | 1.15 [1.11-1.20] | 1.15 [1.11-1.20] | 1.18 [1.12-1.24] | 1.00 [0.87-1.16] | 1.15 [1.10-1.20] | 1.11 [1.07-1.16] | 1.09 [1.04-1.14] | 1.10 [1.05-1.16] | 1.02 [0.87-1.18] | 1.13 [1.08-1.17] | 6,989 | 6,965 | 4,751 | 618 | 6,741 |
| Z30 | 1.08 [1.03-1.13] | 1.10 [1.05-1.14] | 1.13 [1.05-1.22] | 0.86 [0.72-1.04] | 1.01 [0.97-1.06] | 1.05 [1.01-1.10] | 1.04 [1.00-1.09] | 1.07 [0.99-1.16] | 0.89 [0.73-1.08] | 1.01 [0.96-1.06] | 6,248 | 6,431 | 1,870 | 366 | 5,440 |
| Z32 | 1.16 [1.00-1.35] | 1.14 [0.99-1.32] | | | 1.06 [0.91-1.23] | 1.13 [0.97-1.32] | 1.10 [0.95-1.28] | | | 1.06 [0.90-1.23] | 516 | 574 | NA | NA | 470 |
| Z33 | 1.15 [1.10-1.21] | 1.17 [1.12-1.23] | 1.13 [0.88-1.43] | 0.96 [0.86-1.08] | 1.11 [1.06-1.17] | 1.09 [1.04-1.15] | 1.07 [1.02-1.13] | 1.09 [0.85-1.40] | 0.97 [0.86-1.09] | 1.08 [1.02-1.13] | 4,967 | 5,590 | 185 | 944 | 4,635 |
| Z34 | 1.00 [0.97-1.03] | 1.03 [1.00-1.06] | 0.90 [0.76-1.06] | 0.91 [0.84-0.98] | 0.99 [0.96-1.03] | 0.99 [0.96-1.03] | 1.01 [0.98-1.04] | 0.94 [0.79-1.12] | 0.92 [0.85-0.99] | 0.99 [0.96-1.03] | 12,482 | 13,959 | 362 | 2,327 | 10,399 |
| Z35 | 0.98 [0.95-1.02] | 0.99 [0.96-1.03] | 0.86 [0.78-0.95] | 0.85 [0.78-0.92] | 0.98 [0.95-1.01] | 0.98 [0.95-1.02] | 0.99 [0.96-1.02] | 0.91 [0.82-1.00] | 0.86 [0.79-0.95] | 0.99 [0.95-1.02] | 11,651 | 12,347 | 1,065 | 1,587 | 10,964 |
| Z36 | 1.03 [0.96-1.11] | 1.02 [0.95-1.09] | 0.87 [0.64-1.18] | 0.82 [0.65-1.04] | 1.02 [0.94-1.11] | 1.03 [0.96-1.11] | 1.02 [0.95-1.09] | 0.93 [0.68-1.28] | 0.84 [0.66-1.07] | 1.02 [0.94-1.11] | 2,276 | 2,369 | 102 | 220 | 1,690 |
| Z37 | 0.96 [0.94-0.98] | 0.99 [0.97-1.00] | 0.93 [0.84-1.03] | 0.86 [0.83-0.90] | 0.95 [0.93-0.98] | 0.98 [0.96-1.00] | 0.99 [0.97-1.01] | 1.04 [0.94-1.16] | 0.89 [0.86-0.93] | 0.97 [0.95-1.00] | 41,764 | 48,042 | 1,140 | 8,899 | 22,630 |
| Z38 | 0.91 [0.68-1.22] | 1.17 [0.88-1.55] | | | 1.06 [0.78-1.44] | 0.91 [0.68-1.24] | 1.22 [0.91-1.64] | | | 1.03 [0.75-1.41] | 124 | 140 | NA | NA | 110 |
| Z39 | 0.99 [0.93-1.04] | 0.99 [0.94-1.04] | 1.00 [0.78-1.27] | 1.00 [0.86-1.16] | 1.00 [0.95-1.07] | 1.00 [0.94-1.05] | 1.00 [0.94-1.05] | 1.06 [0.82-1.37] | 1.02 [0.87-1.19] | 1.00 [0.94-1.07] | 3,837 | 4,159 | 173 | 583 | 3,048 |
| Z40 | 1.03 [0.90-1.18] | 0.98 [0.86-1.12] | 1.05 [0.91-1.22] | | 0.93 [0.82-1.06] | 1.03 [0.90-1.18] | 0.94 [0.82-1.08] | 1.01 [0.87-1.18] | | 0.94 [0.82-1.08] | 644 | 633 | 491 | NA | 604 |
| Z41 | 1.16 [1.02-1.32] | 1.14 [0.99-1.31] | 1.17 [0.98-1.40] | 1.23 [0.90-1.67] | 1.14 [1.00-1.29] | 1.11 [0.97-1.27] | 1.07 [0.93-1.23] | 1.08 [0.90-1.30] | 1.22 [0.89-1.67] | 1.14 [0.99-1.30] | 691 | 605 | 345 | 150 | 643 |
| Z42 | 1.09 [1.01-1.18] | 1.11 [1.03-1.20] | 1.09 [1.00-1.18] | | 1.08 [1.00-1.17] | 1.08 [0.99-1.17] | 1.06 [0.98-1.15] | 1.05 [0.96-1.14] | | 1.09 [1.01-1.18] | 1,845 | 1,833 | 1,503 | NA | 1,767 |
| Z43 | 1.08 [1.02-1.15] | 1.14 [1.07-1.22] | 1.12 [1.05-1.19] | 0.79 [0.61-1.01] | 1.08 [1.01-1.14] | 1.07 [1.01-1.14] | 1.11 [1.04-1.19] | 1.09 [1.02-1.16] | 0.75 [0.58-0.97] | 1.08 [1.02-1.15] | 2,884 | 2,747 | 2,509 | 191 | 2,782 |
| Z45 | 1.13 [1.09-1.18] | 1.14 [1.10-1.19] | 1.12 [1.07-1.16] | 1.01 [0.82-1.25] | 1.08 [1.04-1.12] | 1.10 [1.05-1.14] | 1.08 [1.04-1.13] | 1.05 [1.01-1.10] | 1.00 [0.81-1.24] | 1.06 [1.02-1.11] | 6,871 | 6,695 | 6,183 | 301 | 6,645 |
| Z46 | 1.06 [1.02-1.10] | 1.06 [1.02-1.10] | 1.06 [1.01-1.10] | 1.05 [0.83-1.35] | 1.03 [0.99-1.07] | 1.04 [1.00-1.08] | 1.03 [0.99-1.07] | 1.03 [0.99-1.07] | 1.00 [0.78-1.29] | 1.02 [0.98-1.06] | 7,883 | 7,782 | 7,237 | 238 | 7,350 |
| Z47 | 1.12 [1.06-1.18] | 1.15 [1.08-1.22] | 1.15 [1.07-1.23] | 1.03 [0.91-1.16] | 1.12 [1.06-1.19] | 1.08 [1.02-1.14] | 1.08 [1.01-1.15] | 1.10 [1.03-1.18] | 1.02 [0.90-1.15] | 1.09 [1.03-1.16] | 3,808 | 3,024 | 2,258 | 1,008 | 3,426 |
| Z48 | 1.13 [1.05-1.22] | 1.15 [1.06-1.24] | 1.11 [1.02-1.21] | 1.11 [0.91-1.36] | 1.08 [1.00-1.17] | 1.10 [1.02-1.19] | 1.11 [1.02-1.20] | 1.07 [0.98-1.17] | 1.09 [0.89-1.34] | 1.07 [0.99-1.15] | 1,972 | 1,715 | 1,445 | 322 | 1,763 |
| Z49 | 1.27 [1.17-1.38] | 1.27 [1.17-1.38] | 1.23 [1.13-1.33] | | 1.17 [1.08-1.27] | 1.26 [1.16-1.37] | 1.21 [1.12-1.32] | 1.16 [1.06-1.26] | | 1.19 [1.09-1.29] | 1,674 | 1,681 | 1,583 | NA | 1,635 |
| Z50 | 1.11 [1.09-1.13] | 1.12 [1.10-1.13] | 1.11 [1.10-1.13] | 0.98 [0.87-1.10] | 1.10 [1.09-1.12] | 1.08 [1.07-1.10] | 1.07 [1.05-1.08] | 1.06 [1.05-1.08] | 0.95 [0.84-1.08] | 1.09 [1.07-1.10] | 53,539 | 53,191 | 51,299 | 958 | 51,059 |
| Z51 | 1.14 [1.12-1.15] | 1.13 [1.12-1.15] | 1.13 [1.11-1.14] | 1.14 [1.03-1.26] | 1.11 [1.10-1.13] | 1.11 [1.09-1.12] | 1.08 [1.07-1.10] | 1.08 [1.06-1.09] | 1.11 [1.01-1.23] | 1.09 [1.08-1.11] | 61,301 | 60,872 | 56,491 | 1,396 | 58,085 |
| Z52 | 0.90 [0.71-1.15] | 0.92 [0.72-1.17] | 0.89 [0.67-1.18] | | 0.88 [0.68-1.14] | 0.90 [0.70-1.15] | 0.92 [0.72-1.18] | 0.91 [0.68-1.22] | | 0.89 [0.68-1.16] | 188 | 186 | 127 | NA | 151 |
| Z53 | 1.21 [1.19-1.23] | 1.20 [1.18-1.22] | 1.20 [1.18-1.22] | 1.19 [1.12-1.27] | 1.16 [1.14-1.18] | 1.15 [1.13-1.17] | 1.12 [1.10-1.13] | 1.12 [1.11-1.14] | 1.15 [1.08-1.23] | 1.12 [1.11-1.14] | 50,321 | 48,556 | 42,356 | 3,302 | 46,665 |
| Z54 | 1.32 [1.13-1.55] | 1.24 [1.06-1.45] | 1.24 [1.06-1.45] | | 1.27 [1.07-1.50] | 1.29 [1.10-1.51] | 1.20 [1.03-1.41] | 1.19 [1.01-1.40] | | 1.26 [1.06-1.49] | 465 | 467 | 457 | NA | 398 |
| Z56 | 1.14 [0.97-1.33] | 1.13 [0.97-1.32] | 1.08 [0.89-1.32] | | 1.06 [0.91-1.25] | 1.11 [0.94-1.31] | 1.00 [0.85-1.18] | 1.01 [0.83-1.24] | | 1.02 [0.86-1.20] | 486 | 497 | 291 | NA | 449 |
| Z57 | 1.20 [1.12-1.30] | 1.21 [1.12-1.31] | 1.23 [1.14-1.32] | | 1.18 [1.10-1.28] | 1.16 [1.08-1.25] | 1.15 [1.06-1.24] | 1.16 [1.07-1.25] | | 1.14 [1.05-1.23] | 1,984 | 1,989 | 1,956 | NA | 1,884 |
| Z58 | 1.17 [1.06-1.29] | 1.12 [1.01-1.24] | 1.16 [1.04-1.29] | 1.52 [1.17-1.97] | 1.16 [1.06-1.28] | 1.10 [1.00-1.21] | 1.02 [0.92-1.14] | 1.05 [0.93-1.17] | 1.53 [1.16-2.00] | 1.09 [0.99-1.20] | 1,301 | 1,149 | 987 | 214 | 1,258 |
| Z59 | 1.08 [0.97-1.19] | 1.04 [0.94-1.15] | 1.06 [0.94-1.19] | 1.18 [0.85-1.66] | 1.03 [0.93-1.14] | 1.05 [0.94-1.16] | 0.98 [0.88-1.09] | 0.99 [0.88-1.12] | 1.27 [0.88-1.82] | 1.05 [0.95-1.17] | 1,145 | 1,128 | 824 | 118 | 1,092 |
| Z60 | 1.12 [1.09-1.15] | 1.12 [1.09-1.15] | 1.12 [1.09-1.15] | 0.62 [0.44-0.86] | 1.10 [1.07-1.13] | 1.10 [1.07-1.13] | 1.07 [1.04-1.10] | 1.07 [1.04-1.10] | 0.63 [0.44-0.91] | 1.08 [1.05-1.11] | 16,880 | 16,832 | 16,418 | 103 | 15,460 |
| Z61 | 1.11 [0.93-1.33] | 1.02 [0.84-1.23] | 1.36 [1.02-1.81] | 0.86 [0.64-1.16] | 0.94 [0.79-1.13] | 1.07 [0.89-1.28] | 0.88 [0.72-1.08] | 1.17 [0.87-1.58] | 0.87 [0.64-1.19] | 0.95 [0.79-1.14] | 385 | 312 | 145 | 146 | 354 |
| Z63 | 1.11 [1.05-1.17] | 1.09 [1.04-1.16] | 1.13 [1.06-1.21] | 0.85 [0.73-0.98] | 1.07 [1.01-1.12] | 1.04 [0.99-1.10] | 0.99 [0.93-1.04] | 1.03 [0.96-1.10] | 0.88 [0.76-1.02] | 1.04 [0.99-1.10] | 4,178 | 3,937 | 2,867 | 628 | 3,969 |
| Z65 | 1.03 [0.85-1.26] | 1.12 [0.91-1.37] | 1.06 [0.81-1.39] | | 0.98 [0.81-1.19] | 1.01 [0.82-1.24] | 0.96 [0.77-1.19] | 0.96 [0.72-1.28] | | 0.99 [0.81-1.21] | 303 | 282 | 155 | NA | 296 |
| Z71 | 1.29 [1.23-1.36] | 1.09 [1.03-1.15] | 1.06 [1.00-1.12] | 3.28 [2.89-3.72] | 1.24 [1.18-1.31] | 1.22 [1.16-1.29] | 1.03 [0.97-1.09] | 1.01 [0.95-1.07] | 2.65 [2.32-3.03] | 1.19 [1.13-1.26] | 5,302 | 3,984 | 3,453 | 1,481 | 4,947 |
| Z72 | 1.13 [1.11-1.15] | 1.13 [1.11-1.15] | 1.14 [1.12-1.17] | 0.98 [0.89-1.08] | 1.09 [1.07-1.12] | 1.11 [1.09-1.13] | 1.05 [1.03-1.07] | 1.08 [1.05-1.10] | 1.02 [0.93-1.13] | 1.10 [1.08-1.13] | 28,055 | 28,553 | 21,098 | 1,459 | 23,916 |
| Z73 | 1.11 [1.07-1.16] | 1.11 [1.07-1.15] | 1.11 [1.07-1.16] | | 1.08 [1.04-1.12] | 1.08 [1.04-1.13] | 1.05 [1.01-1.09] | 1.06 [1.02-1.10] | | 1.07 [1.03-1.11] | 9,071 | 9,107 | 8,747 | NA | 8,793 |
| Z74 | 1.13 [1.08-1.17] | 1.11 [1.06-1.15] | 1.11 [1.07-1.16] | | 1.10 [1.06-1.15] | 1.11 [1.06-1.16] | 1.08 [1.03-1.12] | 1.08 [1.03-1.12] | | 1.10 [1.06-1.15] | 6,919 | 6,929 | 6,848 | NA | 6,546 |
| Z75 | 1.09 [1.06-1.12] | 1.08 [1.05-1.11] | 1.09 [1.05-1.12] | | 1.08 [1.05-1.11] | 1.08 [1.04-1.11] | 1.05 [1.02-1.08] | 1.05 [1.02-1.09] | | 1.08 [1.05-1.12] | 13,694 | 13,705 | 13,520 | NA | 13,300 |
| Z76 | 1.16 [1.04-1.30] | 1.09 [0.97-1.22] | 1.29 [1.04-1.59] | 1.04 [0.76-1.43] | 1.13 [1.00-1.27] | 1.17 [1.04-1.32] | 1.09 [0.97-1.23] | 1.25 [1.01-1.55] | 1.00 [0.71-1.41] | 1.13 [1.00-1.28] | 897 | 896 | 283 | 127 | 775 |
| Z80 | 1.09 [1.06-1.12] | 1.09 [1.06-1.13] | 1.08 [1.05-1.12] | 0.96 [0.79-1.16] | 1.05 [1.02-1.09] | 1.05 [1.02-1.08] | 1.04 [1.01-1.07] | 1.03 [1.00-1.07] | 0.94 [0.76-1.15] | 1.02 [0.99-1.06] | 12,119 | 12,203 | 10,395 | 341 | 10,864 |
| Z81 | 1.11 [1.01-1.22] | 1.06 [0.96-1.16] | 1.32 [1.12-1.57] | 1.10 [0.95-1.28] | 1.04 [0.95-1.14] | 1.05 [0.96-1.16] | 0.93 [0.84-1.03] | 1.19 [1.00-1.42] | 1.08 [0.92-1.26] | 1.02 [0.93-1.12] | 1,492 | 1,220 | 391 | 614 | 1,416 |
| Z82 | 1.16 [1.14-1.18] | 1.11 [1.09-1.13] | 1.13 [1.10-1.15] | 1.60 [1.51-1.71] | 1.12 [1.10-1.15] | 1.11 [1.09-1.14] | 1.04 [1.02-1.06] | 1.05 [1.03-1.08] | 1.57 [1.47-1.68] | 1.09 [1.07-1.12] | 27,494 | 25,100 | 19,859 | 3,863 | 25,152 |
| Z83 | 1.12 [1.08-1.16] | 1.09 [1.05-1.13] | 1.12 [1.07-1.18] | 1.15 [1.06-1.26] | 1.09 [1.05-1.13] | 1.08 [1.04-1.12] | 1.04 [1.00-1.08] | 1.05 [1.00-1.11] | 1.13 [1.03-1.24] | 1.06 [1.02-1.10] | 9,665 | 9,145 | 4,517 | 1,816 | 8,861 |
| Z84 | 1.53 [1.38-1.69] | 1.12 [0.99-1.27] | 1.22 [1.00-1.49] | 1.86 [1.61-2.14] | 1.41 [1.28-1.56] | 1.52 [1.37-1.69] | 1.09 [0.95-1.24] | 1.17 [0.95-1.43] | 1.86 [1.60-2.16] | 1.40 [1.26-1.55] | 1,369 | 755 | 300 | 758 | 1,296 |
| Z85 | 1.10 [1.08-1.12] | 1.10 [1.08-1.12] | 1.11 [1.09-1.12] | 0.97 [0.79-1.19] | 1.07 [1.05-1.09] | 1.08 [1.06-1.09] | 1.06 [1.04-1.07] | 1.06 [1.05-1.08] | 1.00 [0.80-1.23] | 1.07 [1.05-1.08] | 49,392 | 49,283 | 47,881 | 296 | 47,097 |
| Z86 | 1.16 [1.15-1.17] | 1.16 [1.15-1.17] | 1.17 [1.16-1.18] | 0.98 [0.94-1.03] | 1.12 [1.11-1.13] | 1.12 [1.11-1.13] | 1.08 [1.07-1.09] | 1.09 [1.08-1.10] | 1.02 [0.97-1.07] | 1.10 [1.09-1.11] | 160,334 | 161,425 | 136,870 | 6,238 | 149,123 |
| Z87 | 1.17 [1.16-1.19] | 1.17 [1.16-1.19] | 1.18 [1.16-1.19] | 1.16 [1.11-1.21] | 1.13 [1.12-1.15] | 1.11 [1.09-1.12] | 1.08 [1.07-1.10] | 1.09 [1.07-1.11] | 1.12 [1.07-1.17] | 1.09 [1.08-1.10] | 79,896 | 76,878 | 61,747 | 7,339 | 77,310 |
| Z88 | 1.33 [1.31-1.34] | 1.32 [1.30-1.34] | 1.32 [1.31-1.34] | 1.30 [1.23-1.37] | 1.30 [1.28-1.32] | 1.22 [1.21-1.24] | 1.19 [1.18-1.21] | 1.20 [1.19-1.22] | 1.22 [1.15-1.28] | 1.21 [1.20-1.23] | 89,234 | 88,383 | 72,465 | 5,464 | 81,876 |
| Z89 | 1.12 [1.05-1.20] | 1.13 [1.06-1.21] | 1.14 [1.07-1.22] | | 1.08 [1.01-1.15] | 1.09 [1.01-1.16] | 1.06 [0.99-1.13] | 1.06 [0.99-1.14] | | 1.08 [1.01-1.15] | 2,548 | 2,522 | 2,480 | NA | 2,463 |
| Z90 | 1.13 [1.11-1.15] | 1.12 [1.11-1.14] | 1.12 [1.10-1.14] | 1.03 [0.92-1.16] | 1.09 [1.07-1.11] | 1.08 [1.06-1.09] | 1.05 [1.03-1.07] | 1.05 [1.04-1.07] | 1.02 [0.90-1.15] | 1.06 [1.04-1.08] | 43,872 | 43,913 | 39,685 | 991 | 42,789 |
| Z91 | 1.52 [1.49-1.54] | 1.38 [1.35-1.41] | 1.38 [1.35-1.41] | 2.28 [2.17-2.39] | 1.44 [1.42-1.47] | 1.36 [1.33-1.39] | 1.21 [1.18-1.23] | 1.23 [1.20-1.26] | 2.01 [1.91-2.12] | 1.33 [1.30-1.35] | 36,843 | 32,108 | 22,361 | 8,172 | 34,862 |
| Z92 | 1.12 [1.11-1.13] | 1.12 [1.11-1.13] | 1.12 [1.11-1.13] | 1.04 [0.94-1.15] | 1.08 [1.07-1.10] | 1.08 [1.07-1.09] | 1.05 [1.04-1.07] | 1.05 [1.04-1.06] | 1.00 [0.90-1.11] | 1.06 [1.05-1.08] | 110,285 | 110,141 | 105,614 | 1,366 | 104,999 |
| Z93 | 1.11 [1.07-1.15] | 1.13 [1.09-1.17] | 1.11 [1.07-1.15] | 0.82 [0.68-1.00] | 1.09 [1.06-1.13] | 1.08 [1.05-1.12] | 1.09 [1.05-1.13] | 1.08 [1.04-1.12] | 0.78 [0.64-0.96] | 1.09 [1.05-1.13] | 9,435 | 9,293 | 8,700 | 319 | 9,097 |

Supplementary Table 4: Hazard ratios and events from all cohorts (excluding non-consulters)

| | Hazard ratio (99% confidence interval) | | | | | | | | | | Events (in exposed) | | | | |
|---|---|---|---|---|---|---|---|---|---|---|---|---|---|---|---|
| | crude | | | | | adjusted | | | | | | | | | |
| Outcome | any age | 18+ | 40+ | <18 | hosp. | any age | 18+ | 40+ | <18 | hosp. | any age | 18+ | 40+ | <18 | hosp. |
| Z94 | 1.19 [1.10-1.29] | 1.19 [1.10-1.29] | 1.21 [1.11-1.32] | 1.06 [0.73-1.52] | 1.07 [0.99-1.15] | 1.16 [1.07-1.26] | 1.15 [1.06-1.24] | 1.17 [1.07-1.27] | 1.01 [0.69-1.49] | 1.07 [0.99-1.16] | 1,846 | 1,784 | 1,541 | 106 | 1,801 |
| Z95 | 1.10 [1.08-1.12] | 1.11 [1.09-1.13] | 1.11 [1.09-1.13] | 0.72 [0.58-0.90] | 1.06 [1.05-1.08] | 1.07 [1.05-1.09] | 1.05 [1.03-1.07] | 1.04 [1.02-1.06] | 0.71 [0.56-0.89] | 1.05 [1.04-1.07] | 36,396 | 36,311 | 35,702 | 248 | 35,794 |
| Z96 | 1.13 [1.11-1.14] | 1.13 [1.11-1.14] | 1.12 [1.11-1.14] | 0.94 [0.80-1.10] | 1.10 [1.09-1.11] | 1.09 [1.08-1.11] | 1.08 [1.06-1.09] | 1.08 [1.06-1.09] | 0.95 [0.80-1.12] | 1.08 [1.07-1.09] | 68,729 | 68,441 | 67,217 | 531 | 66,196 |
| Z97 | 1.18 [1.13-1.23] | 1.20 [1.15-1.25] | 1.16 [1.11-1.22] | 1.03 [0.85-1.24] | 1.16 [1.11-1.21] | 1.12 [1.07-1.17] | 1.12 [1.07-1.17] | 1.09 [1.04-1.15] | 1.05 [0.86-1.28] | 1.13 [1.08-1.18] | 6,590 | 6,639 | 5,281 | 374 | 6,440 |
| Z98 | 1.18 [1.13-1.22] | 1.19 [1.15-1.24] | 1.17 [1.13-1.22] | 0.98 [0.79-1.22] | 1.14 [1.10-1.18] | 1.10 [1.06-1.14] | 1.08 [1.04-1.13] | 1.07 [1.03-1.12] | 0.96 [0.77-1.21] | 1.08 [1.04-1.12] | 8,583 | 8,554 | 7,525 | 293 | 8,577 |
| Z99 | 1.24 [1.20-1.28] | 1.23 [1.20-1.27] | 1.23 [1.19-1.27] | 0.85 [0.68-1.06] | 1.20 [1.16-1.23] | 1.17 [1.13-1.20] | 1.12 [1.08-1.16] | 1.12 [1.08-1.15] | 0.83 [0.66-1.05] | 1.15 [1.11-1.18] | 12,321 | 12,205 | 11,685 | 270 | 12,359 |
