## Supplementary Table 5 for "Mapping risks of hospital-recorded health conditions in people with eczema"

### Supplementary Table 5: GBD-mapped results (hazard ratios and events from all cohorts)

| | Hazard ratio (99% confidence interval) | | | | | | | | | | | | | | |
|---|---|---|---|---|---|---|---|---|---|---|---|---|---|---|---|
| | crude | | | | | adjusted | | | | | Events (in exposed) | | | | |
| Outcome | any age | 18+ | 40+ | <18 | hosp. | any age | 18+ | 40+ | <18 | hosp. | any age | 18+ | 40+ | <18 | hosp. |
| Infectious and parasitic diseases | | | | | | | | | | | | | | | |
| 20 | 1.32 [1.31-1.33] | 1.29 [1.28-1.31] | 1.29 [1.27-1.30] | 1.38 [1.37-1.40] | 1.25 [1.24-1.26] | 1.26 [1.25-1.27] | 1.19 [1.18-1.20] | 1.18 [1.17-1.19] | 1.35 [1.33-1.36] | 1.22 [1.21-1.23] | 220,610 | 162,198 | 132,012 | 69,009 | 190,523 |
| 30 | 1.26 [1.15-1.37] | 1.32 [1.21-1.45] | 1.36 [1.22-1.51] | 0.89 [0.70-1.13] | 1.28 [1.16-1.41] | 1.20 [1.09-1.31] | 1.25 [1.13-1.37] | 1.24 [1.12-1.38] | 0.95 [0.74-1.21] | 1.22 [1.10-1.35] | 1,239 | 1,145 | 900 | 165 | 1,020 |
| 40 | 1.13 [1.10-1.17] | 1.16 [1.13-1.20] | 1.14 [1.09-1.19] | 1.09 [1.00-1.17] | 1.07 [1.04-1.11] | 1.07 [1.04-1.11] | 1.07 [1.04-1.10] | 1.06 [1.01-1.10] | 1.06 [0.98-1.15] | 1.04 [1.01-1.07] | 11,723 | 11,485 | 5,586 | 1,693 | 9,824 |
| 90 | 1.13 [1.10-1.17] | 1.16 [1.13-1.19] | 1.14 [1.09-1.18] | 1.09 [1.01-1.18] | 1.07 [1.04-1.10] | 1.07 [1.04-1.10] | 1.07 [1.03-1.10] | 1.06 [1.01-1.10] | 1.06 [0.98-1.15] | 1.04 [1.00-1.07] | 11,559 | 11,337 | 5,549 | 1,649 | 9,710 |
| 100 | 1.84 [1.39-2.43] | 1.90 [1.42-2.52] | | | | 1.69 [1.26-2.26] | 1.70 [1.26-2.29] | | | | 125 | 119 | NA | NA | NA |
| 102 | 1.84 [1.39-2.43] | 1.90 [1.42-2.52] | | | | 1.69 [1.26-2.26] | 1.70 [1.26-2.29] | | | | 125 | 119 | NA | NA | NA |
| 110 | 1.27 [1.25-1.28] | 1.31 [1.29-1.33] | 1.29 [1.27-1.31] | 1.17 [1.14-1.19] | 1.20 [1.18-1.21] | 1.19 [1.18-1.21] | 1.17 [1.15-1.19] | 1.16 [1.14-1.17] | 1.13 [1.10-1.16] | 1.15 [1.14-1.17] | 72,317 | 56,748 | 46,663 | 19,725 | 65,055 |
| 120 | 1.21 [0.96-1.53] | | | 1.10 [0.82-1.46] | 1.15 [0.89-1.48] | 1.14 [0.90-1.45] | | | 1.05 [0.79-1.41] | 1.09 [0.84-1.42] | 181 | NA | NA | 121 | 150 |
| 170 | 1.13 [1.04-1.23] | 1.16 [1.03-1.31] | 1.17 [1.02-1.35] | 1.10 [0.98-1.23] | 1.04 [0.94-1.14] | 1.08 [0.99-1.18] | 1.07 [0.95-1.21] | 1.08 [0.93-1.25] | 1.07 [0.95-1.20] | 1.01 [0.92-1.11] | 1,275 | 671 | 457 | 706 | 1,047 |
| 180 | 1.20 [1.10-1.30] | 1.23 [1.12-1.36] | 1.21 [1.09-1.35] | 1.01 [0.85-1.20] | 1.12 [1.03-1.23] | 1.15 [1.05-1.26] | 1.13 [1.02-1.25] | 1.11 [0.99-1.24] | 0.99 [0.83-1.18] | 1.10 [1.00-1.20] | 1,317 | 1,037 | 855 | 342 | 1,129 |
| 185 | 1.19 [1.12-1.27] | 1.24 [1.16-1.32] | 1.29 [1.20-1.39] | 1.04 [0.84-1.29] | 1.11 [1.04-1.19] | 1.17 [1.09-1.24] | 1.15 [1.07-1.22] | 1.19 [1.10-1.28] | 1.12 [0.90-1.41] | 1.12 [1.04-1.20] | 2,473 | 2,366 | 1,765 | 218 | 2,050 |
| 186 | 1.10 [0.86-1.39] | 1.24 [0.95-1.60] | 1.35 [1.00-1.82] | | 1.06 [0.82-1.39] | 1.06 [0.83-1.36] | 1.17 [0.89-1.53] | 1.26 [0.93-1.72] | | 1.03 [0.79-1.35] | 171 | 142 | 108 | NA | 139 |
| 190 | 1.20 [1.13-1.28] | 1.23 [1.16-1.32] | 1.29 [1.19-1.38] | 1.07 [0.84-1.37] | 1.11 [1.04-1.19] | 1.17 [1.09-1.25] | 1.14 [1.07-1.22] | 1.18 [1.09-1.27] | 1.18 [0.92-1.52] | 1.12 [1.04-1.20] | 2,324 | 2,246 | 1,675 | 179 | 1,931 |
| 200 | 1.17 [1.07-1.29] | 1.22 [1.11-1.34] | 1.26 [1.14-1.40] | | 1.06 [0.96-1.17] | 1.12 [1.02-1.23] | 1.06 [0.96-1.16] | 1.12 [1.00-1.25] | | 1.07 [0.97-1.19] | 1,058 | 1,048 | 825 | NA | 888 |
| 210 | 1.00 [0.85-1.17] | 1.10 [0.91-1.32] | 1.05 [0.84-1.32] | 0.86 [0.65-1.15] | 0.97 [0.79-1.18] | 0.99 [0.84-1.17] | 1.14 [0.94-1.38] | 1.07 [0.85-1.35] | 0.81 [0.60-1.10] | 0.98 [0.80-1.19] | 368 | 276 | 177 | 115 | 243 |
| 220 | 0.97 [0.80-1.16] | 1.04 [0.84-1.28] | 1.03 [0.80-1.33] | | 0.92 [0.73-1.15] | 0.97 [0.80-1.17] | 1.09 [0.88-1.36] | 1.06 [0.81-1.38] | | 0.93 [0.74-1.18] | 284 | 209 | 133 | NA | 175 |
| 330 | 1.21 [1.06-1.37] | 1.24 [1.04-1.49] | 1.18 [0.94-1.49] | 1.17 [1.00-1.37] | 1.17 [1.02-1.33] | 1.14 [1.00-1.29] | 1.14 [0.94-1.37] | 1.09 [0.86-1.39] | 1.11 [0.95-1.31] | 1.11 [0.97-1.27] | 650 | 291 | 168 | 414 | 562 |
| 362 | 1.20 [1.06-1.37] | 1.23 [1.02-1.48] | 1.19 [0.94-1.51] | 1.18 [1.00-1.38] | 1.15 [1.01-1.32] | 1.14 [1.00-1.29] | 1.12 [0.93-1.36] | 1.11 [0.87-1.43] | 1.12 [0.95-1.32] | 1.10 [0.96-1.27] | 626 | 272 | 159 | 406 | 542 |
| 370 | 1.36 [1.35-1.37] | 1.30 [1.29-1.32] | 1.29 [1.28-1.31] | 1.47 [1.45-1.49] | 1.29 [1.28-1.30] | 1.30 [1.29-1.31] | 1.20 [1.19-1.21] | 1.19 [1.18-1.20] | 1.43 [1.41-1.45] | 1.26 [1.25-1.27] | 170,698 | 122,439 | 104,438 | 54,657 | 149,347 |
| Respiratory infectious | | | | | | | | | | | | | | | |
| 380 | 1.28 [1.27-1.29] | 1.29 [1.28-1.30] | 1.27 [1.26-1.29] | 1.25 [1.24-1.27] | 1.22 [1.21-1.23] | 1.22 [1.21-1.23] | 1.18 [1.16-1.19] | 1.17 [1.15-1.18] | 1.21 [1.20-1.23] | 1.18 [1.17-1.19] | 207,228 | 140,428 | 122,072 | 73,961 | 178,108 |
| 390 | 1.30 [1.29-1.31] | 1.29 [1.28-1.30] | 1.27 [1.26-1.28] | 1.39 [1.36-1.42] | 1.24 [1.22-1.25] | 1.23 [1.22-1.24] | 1.17 [1.16-1.19] | 1.16 [1.15-1.18] | 1.33 [1.30-1.36] | 1.19 [1.18-1.20] | 147,648 | 127,381 | 117,393 | 23,454 | 131,283 |
| 400 | 1.25 [1.23-1.27] | 1.34 [1.31-1.37] | 1.36 [1.31-1.41] | 1.22 [1.20-1.24] | 1.20 [1.18-1.21] | 1.20 [1.18-1.21] | 1.19 [1.16-1.22] | 1.19 [1.15-1.24] | 1.18 [1.16-1.20] | 1.16 [1.14-1.17] | 60,682 | 16,586 | 6,996 | 48,823 | 50,826 |
| 410 | 1.22 [1.19-1.24] | 1.45 [1.37-1.53] | 1.43 [1.35-1.53] | 1.19 [1.16-1.22] | 1.18 [1.15-1.20] | 1.18 [1.15-1.20] | 1.31 [1.24-1.38] | 1.31 [1.22-1.39] | 1.16 [1.13-1.19] | 1.15 [1.12-1.17] | 22,933 | 3,432 | 2,487 | 19,831 | 19,093 |
| Maternal conditions | | | | | | | | | | | | | | | |
| 420 | 1.07 [1.05-1.08] | 1.12 [1.10-1.13] | 1.05 [0.99-1.13] | 0.99 [0.97-1.01] | 0.97 [0.95-0.99] | 1.05 [1.04-1.07] | 1.08 [1.06-1.09] | 1.09 [1.01-1.17] | 0.98 [0.97-1.00] | 0.97 [0.95-0.99] | 81,715 | 77,703 | 2,347 | 31,825 | 37,492 |
| Neonatal conditions | | | | | | | | | | | | | | | |
| 490 | 1.06 [0.91-1.23] | | | 1.02 [0.86-1.20] | 0.98 [0.82-1.17] | 1.02 [0.87-1.19] | | | 0.99 [0.84-1.17] | 0.98 [0.82-1.17] | 448 | NA | NA | 371 | 336 |
| 530 | 1.01 [0.86-1.18] | | | 1.02 [0.87-1.20] | 0.94 [0.79-1.13] | 0.99 [0.84-1.16] | | | 1.01 [0.86-1.19] | 0.94 [0.79-1.13] | 399 | NA | NA | 379 | 318 |
| Nutritional deficiencies | | | | | | | | | | | | | | | |
| 540 | 1.32 [1.31-1.34] | 1.32 [1.31-1.34] | 1.32 [1.30-1.33] | 1.28 [1.23-1.33] | 1.25 [1.24-1.27] | 1.25 [1.23-1.26] | 1.20 [1.19-1.21] | 1.19 [1.18-1.21] | 1.22 [1.17-1.28] | 1.20 [1.19-1.22] | 81,934 | 77,944 | 69,277 | 6,889 | 74,156 |
| 550 | 1.20 [1.12-1.28] | 1.17 [1.09-1.25] | 1.16 [1.08-1.24] | 1.28 [0.98-1.67] | 1.15 [1.07-1.23] | 1.18 [1.10-1.26] | 1.09 [1.02-1.17] | 1.10 [1.02-1.18] | 1.24 [0.94-1.64] | 1.16 [1.08-1.24] | 2,309 | 2,231 | 2,032 | 156 | 2,146 |
| 560 | 1.16 [0.99-1.36] | 1.21 [1.03-1.41] | 1.15 [0.98-1.36] | | 1.12 [0.95-1.31] | 1.14 [0.98-1.34] | 1.16 [0.99-1.36] | 1.11 [0.94-1.32] | | 1.11 [0.95-1.31] | 439 | 434 | 386 | NA | 402 |
| 580 | 1.32 [1.31-1.34] | 1.32 [1.30-1.34] | 1.32 [1.30-1.34] | 1.28 [1.21-1.35] | 1.25 [1.23-1.27] | 1.25 [1.23-1.27] | 1.20 [1.18-1.22] | 1.20 [1.18-1.22] | 1.24 [1.18-1.31] | 1.20 [1.19-1.22] | 53,162 | 50,841 | 45,714 | 4,007 | 47,985 |
| 590 | 1.35 [1.33-1.37] | 1.35 [1.33-1.38] | 1.33 [1.31-1.36] | 1.30 [1.22-1.38] | 1.28 [1.25-1.30] | 1.26 [1.24-1.28] | 1.21 [1.19-1.23] | 1.20 [1.18-1.22] | 1.22 [1.15-1.30] | 1.22 [1.20-1.24] | 37,096 | 35,187 | 30,940 | 3,365 | 34,254 |
| Malignant neoplasms | | | | | | | | | | | | | | | |
| 610 | 1.09 [1.08-1.10] | 1.09 [1.08-1.10] | 1.09 [1.08-1.10] | 1.08 [1.00-1.16] | 1.06 [1.05-1.07] | 1.08 [1.07-1.09] | 1.07 [1.06-1.08] | 1.06 [1.05-1.07] | 1.07 [0.99-1.16] | 1.05 [1.04-1.07] | 108,133 | 107,036 | 103,666 | 1,800 | 86,707 |
| 620 | 1.16 [1.09-1.24] | 1.18 [1.11-1.26] | 1.19 [1.12-1.27] | | 1.13 [1.06-1.21] | 1.16 [1.09-1.23] | 1.14 [1.07-1.21] | 1.15 [1.07-1.22] | | 1.15 [1.08-1.24] | 2,437 | 2,425 | 2,348 | NA | 2,035 |
| 621 | 1.18 [1.10-1.28] | 1.19 [1.11-1.29] | 1.19 [1.10-1.28] | | 1.16 [1.07-1.26] | 1.18 [1.09-1.27] | 1.15 [1.06-1.24] | 1.15 [1.06-1.24] | | 1.18 [1.08-1.28] | 1,693 | 1,685 | 1,613 | NA | 1,427 |
| 622 | 1.19 [0.90-1.58] | 1.20 [0.90-1.60] | 1.36 [1.01-1.83] | | | 1.19 [0.89-1.58] | 1.17 [0.87-1.56] | 1.31 [0.97-1.77] | | | 115 | 111 | 108 | NA | NA |
| 623 | 1.12 [1.01-1.24] | 1.17 [1.05-1.29] | 1.17 [1.05-1.29] | | 1.08 [0.97-1.21] | 1.11 [1.00-1.23] | 1.12 [1.01-1.24] | 1.12 [1.01-1.24] | | 1.11 [0.99-1.25] | 875 | 875 | 873 | NA | 729 |
| 630 | 1.18 [1.12-1.25] | 1.15 [1.09-1.22] | 1.17 [1.10-1.23] | | 1.13 [1.07-1.20] | 1.17 [1.11-1.24] | 1.12 [1.06-1.18] | 1.13 [1.07-1.19] | | 1.14 [1.07-1.21] | 3,259 | 3,259 | 3,242 | NA | 2,722 |
| 640 | 1.08 [1.01-1.15] | 1.07 [1.00-1.13] | 1.09 [1.02-1.16] | | 1.06 [0.99-1.14] | 1.07 [1.01-1.14] | 1.04 [0.98-1.11] | 1.06 [0.99-1.13] | | 1.06 [0.99-1.14] | 2,508 | 2,508 | 2,476 | NA | 2,102 |
| 650 | 0.99 [0.97-1.02] | 0.99 [0.97-1.02] | 0.99 [0.97-1.02] | | 0.98 [0.95-1.01] | 0.99 [0.96-1.02] | 0.99 [0.96-1.02] | 0.98 [0.96-1.01] | | 0.98 [0.95-1.02] | 11,626 | 11,618 | 11,444 | NA | 9,393 |
| 660 | 1.25 [1.16-1.34] | 1.25 [1.16-1.34] | 1.23 [1.14-1.32] | | 1.18 [1.09-1.27] | 1.21 [1.13-1.31] | 1.18 [1.10-1.27] | 1.17 [1.08-1.26] | | 1.17 [1.08-1.26] | 1,925 | 1,919 | 1,894 | NA | 1,708 |
| 670 | 1.16 [1.10-1.23] | 1.14 [1.08-1.21] | 1.15 [1.09-1.22] | | 1.13 [1.07-1.20] | 1.15 [1.09-1.22] | 1.10 [1.04-1.17] | 1.11 [1.05-1.18] | | 1.13 [1.07-1.21] | 3,016 | 3,008 | 2,998 | NA | 2,573 |
| 680 | 1.15 [1.12-1.18] | 1.15 [1.12-1.18] | 1.15 [1.12-1.18] | | 1.11 [1.08-1.14] | 1.14 [1.11-1.17] | 1.09 [1.06-1.12] | 1.09 [1.06-1.12] | | 1.12 [1.09-1.15] | 13,085 | 13,068 | 13,034 | NA | 11,190 |
| 690 | 1.08 [1.06-1.10] | 1.09 [1.07-1.11] | 1.09 [1.07-1.11] | 1.11 [0.86-1.44] | 1.04 [1.02-1.07] | 1.06 [1.04-1.08] | 1.05 [1.03-1.07] | 1.05 [1.03-1.07] | 1.01 [0.77-1.32] | 1.03 [1.00-1.05] | 27,302 | 27,276 | 26,659 | 150 | 23,012 |
| 691 | 0.95 [0.90-1.01] | 0.97 [0.92-1.02] | 0.96 [0.90-1.01] | | 0.97 [0.91-1.02] | 0.94 [0.89-1.00] | 0.95 [0.90-1.01] | 0.94 [0.89-0.99] | | 0.96 [0.90-1.02] | 3,170 | 3,159 | 2,886 | NA | 2,682 |
| 692 | 1.10 [1.08-1.12] | 1.11 [1.08-1.13] | 1.10 [1.08-1.13] | | 1.06 [1.03-1.08] | 1.07 [1.05-1.09] | 1.07 [1.04-1.09] | 1.06 [1.04-1.08] | | 1.03 [1.01-1.06] | 25,094 | 25,077 | 24,709 | NA | 21,264 |
| 700 | 1.04 [1.02-1.07] | 1.04 [1.01-1.06] | 1.03 [1.01-1.06] | | 1.02 [0.99-1.05] | 1.04 [1.01-1.07] | 1.03 [1.00-1.05] | 1.02 [0.99-1.05] | | 1.03 [1.00-1.06] | 14,359 | 14,360 | 13,755 | NA | 11,473 |
| 710 | 0.96 [0.86-1.07] | 0.96 [0.86-1.07] | 0.96 [0.84-1.09] | | 1.02 [0.90-1.15] | 0.97 [0.87-1.09] | 0.97 [0.87-1.08] | 0.98 [0.86-1.12] | | 1.05 [0.93-1.19] | 755 | 752 | 523 | NA | 615 |
| 720 | 1.04 [0.98-1.11] | 1.04 [0.98-1.11] | 1.03 [0.97-1.09] | | 1.03 [0.96-1.10] | 1.06 [0.99-1.12] | 1.06 [1.00-1.13] | 1.05 [0.98-1.11] | | 1.04 [0.97-1.12] | 2,358 | 2,355 | 2,327 | NA | 1,822 |
| 730 | 1.03 [0.97-1.10] | 1.01 [0.95-1.08] | 1.02 [0.96-1.09] | | 1.00 [0.93-1.08] | 1.04 [0.97-1.11] | 1.02 [0.95-1.09] | 1.02 [0.96-1.10] | | 1.01 [0.94-1.09] | 2,094 | 2,078 | 1,979 | NA | 1,630 |
| 740 | 1.06 [1.03-1.09] | 1.06 [1.03-1.09] | 1.05 [1.03-1.08] | | 1.01 [0.98-1.04] | 1.04 [1.01-1.06] | 1.03 [1.00-1.06] | 1.02 [1.00-1.05] | | 1.00 [0.97-1.03] | 13,721 | 13,725 | 13,723 | NA | 11,486 |
| 742 | 1.04 [0.89-1.22] | 1.02 [0.87-1.20] | 0.99 [0.78-1.26] | | 1.02 [0.84-1.24] | 1.02 [0.87-1.20] | 0.99 [0.84-1.17] | 0.95 [0.74-1.22] | | 1.03 [0.84-1.25] | 362 | 345 | 146 | NA | 242 |
| 745 | 1.11 [1.06-1.17] | 1.10 [1.05-1.16] | 1.11 [1.06-1.18] | | 1.05 [0.99-1.11] | 1.09 [1.04-1.15] | 1.05 [1.00-1.11] | 1.06 [1.01-1.12] | | 1.05 [0.99-1.11] | 3,343 | 3,296 | 3,251 | NA | 2,871 |
| 750 | 1.08 [1.04-1.12] | 1.09 [1.05-1.13] | 1.08 [1.04-1.13] | | 1.03 [0.99-1.07] | 1.07 [1.03-1.12] | 1.06 [1.02-1.10] | 1.06 [1.02-1.10] | | 1.03 [0.99-1.08] | 6,425 | 6,415 | 6,366 | NA | 5,376 |
| 751 | 1.12 [1.04-1.21] | 1.14 [1.06-1.23] | 1.13 [1.04-1.22] | 1.01 [0.84-1.22] | 1.09 [1.01-1.18] | 1.10 [1.03-1.19] | 1.11 [1.03-1.20] | 1.10 [1.02-1.20] | 1.02 [0.84-1.24] | 1.08 [1.00-1.17] | 1,820 | 1,586 | 1,437 | 278 | 1,454 |
| 752 | 1.08 [0.96-1.21] | 1.07 [0.95-1.20] | 1.06 [0.94-1.19] | | 1.03 [0.91-1.17] | 1.07 [0.95-1.21] | 1.04 [0.92-1.17] | 1.03 [0.91-1.16] | | 1.03 [0.90-1.17] | 688 | 689 | 682 | NA | 578 |
| 753 | 1.14 [1.02-1.28] | 1.17 [1.04-1.32] | 1.16 [1.03-1.30] | | 1.08 [0.95-1.22] | 1.16 [1.03-1.30] | 1.14 [1.01-1.28] | 1.12 [0.99-1.26] | | 1.11 [0.98-1.26] | 690 | 689 | 680 | NA | 586 |
| 754 | 1.06 [0.96-1.18] | 1.06 [0.95-1.18] | 1.06 [0.94-1.20] | | 1.02 [0.91-1.15] | 1.04 [0.93-1.16] | 1.02 [0.92-1.14] | 1.03 [0.92-1.17] | | 1.01 [0.90-1.14] | 819 | 798 | 618 | NA | 675 |
| 755 | 0.98 [0.88-1.10] | 1.00 [0.89-1.12] | 1.01 [0.90-1.13] | | 1.02 [0.91-1.16] | 0.99 [0.89-1.11] | 1.01 [0.90-1.13] | 1.02 [0.91-1.14] | | 1.04 [0.92-1.18] | 725 | 723 | 725 | NA | 614 |
| 760 | 1.29 [1.25-1.34] | 1.29 [1.25-1.34] | 1.26 [1.22-1.31] | 1.34 [1.13-1.59] | 1.22 [1.17-1.26] | 1.26 [1.22-1.31] | 1.26 [1.22-1.31] | 1.22 [1.18-1.27] | 1.31 [1.09-1.56] | 1.20 [1.15-1.25] | 7,749 | 7,541 | 7,011 | 353 | 6,306 |
| 761 | 1.71 [1.54-1.90] | 1.72 [1.54-1.91] | 1.62 [1.42-1.85] | 1.41 [1.11-1.79] | 1.46 [1.30-1.65] | 1.64 [1.48-1.83] | 1.65 [1.48-1.85] | 1.52 [1.33-1.74] | 1.33 [1.04-1.71] | 1.43 [1.26-1.61] | 969 | 870 | 588 | 185 | 701 |
| 762 | 1.32 [1.26-1.38] | 1.32 [1.26-1.38] | 1.29 [1.23-1.35] | 1.28 [1.02-1.61] | 1.24 [1.18-1.30] | 1.28 [1.23-1.34] | 1.28 [1.22-1.34] | 1.25 [1.19-1.31] | 1.28 [1.01-1.62] | 1.22 [1.17-1.29] | 5,070 | 4,947 | 4,641 | 198 | 4,166 |
| 763 | 1.16 [1.09-1.24] | 1.17 [1.10-1.25] | 1.16 [1.09-1.24] | | 1.11 [1.04-1.19] | 1.13 [1.06-1.21] | 1.14 [1.07-1.22] | 1.13 [1.06-1.21] | | 1.10 [1.02-1.17] | 2,394 | 2,385 | 2,354 | NA | 2,040 |
| 770 | 1.15 [1.09-1.21] | 1.18 [1.12-1.25] | 1.16 [1.10-1.22] | 0.99 [0.85-1.15] | 1.11 [1.05-1.17] | 1.13 [1.07-1.18] | 1.15 [1.09-1.21] | 1.12 [1.06-1.19] | 0.99 [0.84-1.16] | 1.10 [1.05-1.17] | 3,956 | 3,611 | 3,460 | 400 | 3,325 |
| 780 | 1.19 [1.18-1.20] | 1.19 [1.18-1.21] | 1.19 [1.18-1.20] | 1.01 [0.96-1.07] | 1.14 [1.13-1.16] | 1.15 [1.14-1.16] | 1.10 [1.09-1.11] | 1.09 [1.08-1.10] | 1.00 [0.95-1.06] | 1.12 [1.11-1.14] | 108,970 | 106,556 | 101,983 | 3,358 | 85,808 |
| Other neoplasms | | | | | | | | | | | | | | | |
| 790 | 1.16 [1.15-1.17] | 1.16 [1.15-1.18] | 1.16 [1.15-1.17] | 1.12 [1.09-1.16] | 1.10 [1.09-1.11] | 1.11 [1.10-1.12] | 1.09 [1.08-1.10] | 1.09 [1.07-1.10] | 1.09 [1.05-1.13] | 1.07 [1.06-1.08] | 93,227 | 87,836 | 73,590 | 8,908 | 75,311 |
| Diabetes mellitus | | | | | | | | | | | | | | | |
| 800 | 1.24 [1.23-1.26] | 1.25 [1.24-1.27] | 1.25 [1.23-1.26] | 1.01 [0.95-1.07] | 1.19 [1.18-1.21] | 1.18 [1.17-1.19] | 1.12 [1.11-1.14] | 1.11 [1.10-1.13] | 0.99 [0.94-1.06] | 1.15 [1.14-1.17] | 78,753 | 76,641 | 72,970 | 2,904 | 60,805 |
| Endocrine, blood, immune disorders | | | | | | | | | | | | | | | |
| 810 | 1.27 [1.26-1.27] | 1.28 [1.27-1.28] | 1.27 [1.26-1.28] | 1.17 [1.15-1.19] | 1.19 [1.18-1.20] | 1.20 [1.20-1.21] | 1.17 [1.16-1.17] | 1.16 [1.15-1.17] | 1.13 [1.11-1.16] | 1.15 [1.15-1.16] | 290,840 | 273,746 | 227,123 | 32,523 | 238,894 |
| 811 | 1.40 [1.31-1.50] | 1.42 [1.32-1.53] | 1.54 [1.40-1.70] | 1.34 [1.16-1.56] | 1.27 [1.18-1.37] | 1.37 [1.27-1.47] | 1.37 [1.27-1.48] | 1.42 [1.28-1.57] | 1.31 [1.12-1.53] | 1.25 [1.16-1.35] | 2,201 | 1,898 | 1,101 | 518 | 1,811 |
| 812 | 1.31 [1.23-1.40] | 1.37 [1.27-1.47] | 1.30 [1.17-1.44] | 1.36 [1.23-1.51] | 1.22 [1.13-1.31] | 1.28 [1.20-1.37] | 1.32 [1.22-1.43] | 1.23 [1.10-1.37] | 1.36 [1.22-1.51] | 1.21 [1.12-1.30] | 2,519 | 1,759 | 859 | 1,054 | 1,979 |
| 813 | 1.34 [1.23-1.46] | 1.37 [1.24-1.51] | 1.34 [1.20-1.49] | 1.24 [1.04-1.48] | 1.28 [1.17-1.41] | 1.32 [1.20-1.44] | 1.32 [1.20-1.46] | 1.28 [1.15-1.43] | 1.24 [1.03-1.48] | 1.29 [1.17-1.42] | 1,318 | 1,053 | 844 | 333 | 1,142 |
| 814 | 1.27 [1.26-1.27] | 1.28 [1.27-1.28] | 1.27 [1.26-1.28] | 1.16 [1.14-1.19] | 1.19 [1.18-1.20] | 1.20 [1.20-1.21] | 1.17 [1.16-1.17] | 1.16 [1.15-1.17] | 1.13 [1.11-1.15] | 1.15 [1.15-1.16] | 288,595 | 272,399 | 226,681 | 31,345 | 237,480 |
| Mental and substance use disorders | | | | | | | | | | | | | | | |
| 820 | 1.22 [1.21-1.23] | 1.24 [1.23-1.25] | 1.25 [1.24-1.26] | 1.08 [1.06-1.09] | 1.15 [1.14-1.16] | 1.16 [1.16-1.17] | 1.10 [1.09-1.11] | 1.12 [1.11-1.13] | 1.06 [1.04-1.07] | 1.13 [1.12-1.14] | 240,813 | 217,026 | 151,120 | 52,334 | 198,517 |
| 830 | 1.37 [1.35-1.38] | 1.38 [1.36-1.39] | 1.39 [1.37-1.41] | 1.14 [1.10-1.17] | 1.26 [1.25-1.28] | 1.23 [1.22-1.25] | 1.13 [1.12-1.14] | 1.15 [1.13-1.17] | 1.13 [1.09-1.17] | 1.20 [1.18-1.21] | 83,481 | 80,770 | 58,389 | 11,116 | 73,730 |
| 831 | 1.37 [1.35-1.38] | 1.38 [1.36-1.39] | 1.39 [1.37-1.41] | 1.14 [1.10-1.17] | 1.26 [1.25-1.28] | 1.23 [1.22-1.25] | 1.13 [1.12-1.14] | 1.15 [1.13-1.17] | 1.13 [1.09-1.17] | 1.20 [1.18-1.21] | 83,481 | 80,770 | 58,389 | 11,116 | 73,730 |
| 840 | 1.30 [1.24-1.36] | 1.32 [1.26-1.38] | 1.33 [1.26-1.41] | 1.08 [0.94-1.23] | 1.16 [1.11-1.22] | 1.19 [1.13-1.24] | 1.09 [1.03-1.14] | 1.13 [1.07-1.20] | 1.09 [0.95-1.25] | 1.12 [1.07-1.18] | 4,470 | 4,378 | 2,830 | 601 | 3,878 |
| 850 | 1.25 [1.20-1.30] | 1.24 [1.20-1.29] | 1.31 [1.25-1.37] | 0.97 [0.87-1.08] | 1.14 [1.09-1.19] | 1.22 [1.17-1.27] | 1.14 [1.10-1.19] | 1.23 [1.17-1.29] | 0.99 [0.88-1.11] | 1.15 [1.10-1.20] | 6,436 | 6,180 | 4,492 | 860 | 5,088 |
| 860 | 1.20 [1.18-1.23] | 1.23 [1.20-1.25] | 1.26 [1.23-1.29] | 0.98 [0.93-1.03] | 1.13 [1.11-1.16] | 1.17 [1.14-1.19] | 1.11 [1.09-1.14] | 1.15 [1.12-1.18] | 0.97 [0.92-1.02] | 1.15 [1.12-1.17] | 23,745 | 22,275 | 16,448 | 3,835 | 18,898 |
| 870 | 1.14 [1.10-1.17] | 1.18 [1.14-1.22] | 1.31 [1.25-1.38] | 0.97 [0.92-1.03] | 1.03 [1.00-1.07] | 1.06 [1.03-1.10] | 1.00 [0.97-1.04] | 1.08 [1.03-1.14] | 0.98 [0.93-1.03] | 1.02 [0.99-1.06] | 9,576 | 8,354 | 3,688 | 3,466 | 7,900 |
| 871 | 1.19 [1.13-1.25] | 1.23 [1.16-1.30] | 1.34 [1.24-1.44] | 0.98 [0.86-1.11] | 1.11 [1.04-1.17] | 1.09 [1.03-1.16] | 1.00 [0.94-1.06] | 1.08 [1.00-1.17] | 0.99 [0.87-1.13] | 1.08 [1.02-1.14] | 3,240 | 3,030 | 1,739 | 616 | 2,801 |
| 872 | 1.12 [1.03-1.21] | 1.16 [1.07-1.26] | 1.42 [1.22-1.65] | 1.02 [0.89-1.16] | 0.98 [0.90-1.07] | 1.04 [0.95-1.13] | 0.97 [0.89-1.05] | 1.13 [0.97-1.33] | 1.03 [0.89-1.18] | 0.99 [0.91-1.08] | 1,455 | 1,412 | 404 | 530 | 1,208 |
| 873 | 1.03 [0.90-1.19] | 1.04 [0.90-1.20] | 1.16 [0.91-1.48] | 0.95 [0.76-1.18] | 0.93 [0.80-1.08] | 0.97 [0.85-1.12] | 0.93 [0.80-1.08] | 0.98 [0.76-1.27] | 0.92 [0.73-1.16] | 0.94 [0.81-1.10] | 503 | 438 | 148 | 200 | 403 |
| 874 | 1.08 [1.03-1.14] | 1.12 [1.06-1.18] | 1.28 [1.17-1.41] | 0.96 [0.89-1.04] | 0.95 [0.90-1.01] | 1.01 [0.96-1.07] | 0.95 [0.90-1.00] | 1.06 [0.96-1.17] | 0.97 [0.89-1.05] | 0.96 [0.91-1.02] | 3,794 | 3,351 | 1,033 | 1,749 | 3,051 |
| 875 | 1.16 [1.09-1.23] | 1.23 [1.15-1.32] | 1.31 [1.19-1.43] | 0.99 [0.90-1.10] | 1.06 [0.99-1.13] | 1.08 [1.02-1.16] | 1.03 [0.96-1.11] | 1.08 [0.98-1.19] | 1.00 [0.90-1.11] | 1.05 [0.98-1.12] | 2,491 | 1,933 | 1,086 | 935 | 2,148 |
| 880 | 1.33 [1.31-1.34] | 1.34 [1.33-1.36] | 1.35 [1.33-1.37] | 1.16 [1.13-1.19] | 1.24 [1.22-1.25] | 1.20 [1.18-1.21] | 1.12 [1.10-1.13] | 1.13 [1.12-1.15] | 1.13 [1.10-1.16] | 1.16 [1.15-1.18] | 78,552 | 73,087 | 49,431 | 15,339 | 69,390 |
| 890 | 1.28 [1.19-1.37] | 1.29 [1.18-1.41] | 1.42 [1.22-1.64] | 1.13 [1.02-1.24] | 1.19 [1.10-1.28] | 1.17 [1.08-1.26] | 1.08 [0.99-1.18] | 1.19 [1.02-1.39] | 1.09 [0.99-1.20] | 1.13 [1.04-1.22] | 2,058 | 1,258 | 443 | 1,189 | 1,711 |
| 900 | 1.32 [1.28-1.38] | 1.54 [1.44-1.65] | 2.15 [1.90-2.44] | 1.22 [1.17-1.27] | 1.23 [1.18-1.28] | 1.18 [1.14-1.23] | 1.34 [1.24-1.44] | 1.73 [1.51-1.97] | 1.15 [1.11-1.20] | 1.14 [1.09-1.19] | 7,896 | 2,095 | 695 | 6,653 | 6,860 |
| 910 | 1.22 [1.16-1.28] | 1.22 [1.11-1.33] | 1.39 [1.14-1.70] | 1.17 [1.11-1.24] | 1.14 [1.08-1.20] | 1.13 [1.08-1.19] | 1.08 [0.99-1.19] | 1.14 [0.93-1.40] | 1.14 [1.08-1.20] | 1.11 [1.05-1.17] | 4,568 | 1,169 | 251 | 3,997 | 3,979 |
| 911 | 1.22 [1.15-1.28] | 1.23 [1.12-1.35] | 1.75 [1.34-2.29] | 1.16 [1.09-1.22] | 1.14 [1.08-1.21] | 1.12 [1.06-1.19] | 1.09 [0.99-1.21] | 1.26 [0.94-1.68] | 1.12 [1.06-1.19] | 1.11 [1.04-1.17] | 3,883 | 1,020 | 139 | 3,444 | 3,393 |
| 912 | 1.24 [1.11-1.39] | 1.14 [0.90-1.44] | 1.10 [0.82-1.46] | 1.27 [1.12-1.43] | 1.15 [1.02-1.30] | 1.17 [1.05-1.31] | 1.04 [0.81-1.32] | 1.02 [0.76-1.37] | 1.22 [1.07-1.38] | 1.11 [0.98-1.25] | 867 | 174 | 113 | 730 | 754 |
| 920 | 1.58 [1.49-1.67] | 1.89 [1.77-2.02] | 2.15 [1.98-2.33] | 1.09 [1.00-1.18] | 1.37 [1.29-1.45] | 1.45 [1.37-1.53] | 1.74 [1.63-1.86] | 1.94 [1.78-2.11] | 1.05 [0.97-1.15] | 1.30 [1.22-1.38] | 3,632 | 2,534 | 1,643 | 1,483 | 3,148 |
| 930 | 1.16 [1.15-1.17] | 1.18 [1.17-1.19] | 1.18 [1.17-1.20] | 1.03 [1.01-1.05] | 1.09 [1.08-1.10] | 1.13 [1.12-1.14] | 1.06 [1.05-1.07] | 1.08 [1.07-1.09] | 1.03 [1.01-1.05] | 1.11 [1.10-1.12] | 149,446 | 139,440 | 95,176 | 29,738 | 126,704 |
| Neurological conditions | | | | | | | | | | | | | | | |
| 940 | 1.24 [1.23-1.25] | 1.24 [1.23-1.25] | 1.24 [1.22-1.25] | 1.18 [1.15-1.21] | 1.17 [1.16-1.18] | 1.17 [1.16-1.18] | 1.13 [1.12-1.14] | 1.13 [1.12-1.14] | 1.14 [1.11-1.16] | 1.13 [1.12-1.14] | 161,265 | 145,227 | 126,193 | 21,702 | 138,222 |
| 950 | 1.10 [1.08-1.11] | 1.09 [1.08-1.11] | 1.09 [1.08-1.11] | 1.02 [0.78-1.33] | 1.07 [1.05-1.08] | 1.08 [1.07-1.10] | 1.06 [1.04-1.08] | 1.06 [1.04-1.07] | 0.99 [0.75-1.30] | 1.07 [1.05-1.08] | 47,325 | 47,210 | 47,009 | 139 | 42,954 |
| 960 | 1.07 [1.03-1.11] | 1.06 [1.03-1.10] | 1.07 [1.04-1.11] | | 1.04 [1.00-1.08] | 1.06 [1.02-1.10] | 1.03 [1.00-1.07] | 1.04 [1.01-1.08] | | 1.04 [1.00-1.08] | 7,838 | 7,840 | 7,793 | NA | 6,778 |
| 970 | 1.30 [1.27-1.33] | 1.35 [1.32-1.39] | 1.37 [1.33-1.41] | 1.07 [1.02-1.12] | 1.18 [1.15-1.21] | 1.20 [1.17-1.23] | 1.18 [1.15-1.21] | 1.19 [1.16-1.23] | 1.05 [1.00-1.10] | 1.13 [1.10-1.16] | 17,589 | 14,393 | 11,345 | 4,197 | 14,416 |
| 980 | 1.23 [1.15-1.31] | 1.22 [1.15-1.30] | 1.23 [1.14-1.32] | 1.14 [0.85-1.53] | 1.14 [1.06-1.22] | 1.16 [1.08-1.24] | 1.08 [1.01-1.15] | 1.09 [1.01-1.18] | 1.15 [0.85-1.55] | 1.12 [1.04-1.21] | 2,212 | 2,199 | 1,684 | 125 | 1,677 |
| 990 | 1.32 [1.28-1.35] | 1.32 [1.29-1.36] | 1.33 [1.29-1.38] | 1.20 [1.14-1.26] | 1.21 [1.18-1.25] | 1.16 [1.13-1.19] | 1.08 [1.05-1.11] | 1.10 [1.06-1.14] | 1.13 [1.07-1.20] | 1.12 [1.09-1.15] | 16,417 | 14,623 | 8,600 | 3,948 | 14,478 |
| 1,000 | 1.33 [1.24-1.42] | 1.31 [1.23-1.40] | 1.37 [1.26-1.49] | 1.22 [1.05-1.42] | 1.23 [1.15-1.32] | 1.20 [1.12-1.28] | 1.12 [1.04-1.20] | 1.15 [1.06-1.26] | 1.19 [1.02-1.39] | 1.15 [1.07-1.23] | 2,317 | 2,109 | 1,353 | 470 | 2,084 |
| 1,010 | 1.29 [1.28-1.30] | 1.30 [1.28-1.31] | 1.29 [1.28-1.30] | 1.19 [1.16-1.23] | 1.20 [1.19-1.22] | 1.20 [1.19-1.22] | 1.15 [1.14-1.17] | 1.15 [1.14-1.16] | 1.15 [1.12-1.18] | 1.15 [1.14-1.16] | 102,753 | 90,649 | 79,225 | 15,059 | 89,516 |
| Sense organ diseases | | | | | | | | | | | | | | | |
| 1,020 | 1.24 [1.24-1.25] | 1.26 [1.25-1.27] | 1.25 [1.24-1.26] | 1.18 [1.16-1.20] | 1.18 [1.17-1.19] | 1.19 [1.18-1.20] | 1.17 [1.16-1.18] | 1.17 [1.16-1.18] | 1.14 [1.12-1.17] | 1.14 [1.13-1.15] | 179,196 | 154,467 | 141,161 | 29,279 | 145,632 |
| 1,030 | 1.18 [1.15-1.20] | 1.18 [1.15-1.20] | 1.18 [1.15-1.20] | 1.29 [1.00-1.67] | 1.13 [1.11-1.16] | 1.14 [1.12-1.17] | 1.13 [1.11-1.16] | 1.13 [1.11-1.16] | 1.26 [0.97-1.64] | 1.11 [1.08-1.14] | 21,887 | 21,771 | 21,555 | 166 | 18,784 |
| 1,040 | 1.21 [1.20-1.22] | 1.21 [1.19-1.22] | 1.21 [1.20-1.22] | 1.59 [1.36-1.86] | 1.15 [1.14-1.17] | 1.16 [1.15-1.18] | 1.14 [1.13-1.15] | 1.14 [1.13-1.16] | 1.56 [1.33-1.83] | 1.12 [1.11-1.14] | 93,063 | 92,792 | 92,062 | 471 | 77,516 |
| 1,050 | 1.10 [1.07-1.13] | 1.15 [1.12-1.19] | 1.15 [1.11-1.18] | 1.02 [0.97-1.06] | 1.06 [1.03-1.09] | 1.07 [1.04-1.10] | 1.10 [1.07-1.14] | 1.09 [1.06-1.13] | 0.99 [0.95-1.04] | 1.04 [1.01-1.07] | 14,365 | 9,472 | 8,504 | 5,228 | 12,293 |
| 1,070 | 1.18 [1.17-1.20] | 1.19 [1.17-1.21] | 1.18 [1.16-1.20] | 1.15 [1.10-1.21] | 1.13 [1.11-1.15] | 1.14 [1.12-1.15] | 1.12 [1.10-1.13] | 1.11 [1.09-1.13] | 1.11 [1.05-1.16] | 1.10 [1.09-1.12] | 49,522 | 46,488 | 41,633 | 4,784 | 43,251 |
| 1,080 | 1.23 [1.21-1.26] | 1.25 [1.22-1.27] | 1.24 [1.22-1.27] | 1.16 [1.11-1.21] | 1.18 [1.16-1.20] | 1.18 [1.16-1.20] | 1.17 [1.14-1.19] | 1.17 [1.14-1.19] | 1.12 [1.07-1.17] | 1.14 [1.12-1.16] | 35,538 | 30,845 | 29,334 | 5,219 | 32,606 |
| 1,090 | 1.29 [1.28-1.31] | 1.31 [1.30-1.33] | 1.30 [1.28-1.32] | 1.24 [1.21-1.27] | 1.22 [1.20-1.24] | 1.23 [1.22-1.25] | 1.22 [1.20-1.23] | 1.21 [1.19-1.23] | 1.20 [1.17-1.23] | 1.18 [1.16-1.19] | 73,023 | 56,768 | 48,813 | 19,041 | 61,598 |
| Cardiovascular diseases | | | | | | | | | | | | | | | |
| 1,100 | 1.26 [1.25-1.27] | 1.26 [1.25-1.27] | 1.26 [1.25-1.27] | 1.14 [1.11-1.18] | 1.18 [1.18-1.19] | 1.20 [1.20-1.21] | 1.15 [1.14-1.16] | 1.15 [1.14-1.16] | 1.11 [1.07-1.14] | 1.15 [1.14-1.16] | 264,656 | 257,075 | 239,279 | 12,155 | 181,766 |
| 1,110 | 1.23 [1.20-1.25] | 1.22 [1.20-1.25] | 1.23 [1.20-1.25] | 0.97 [0.81-1.16] | 1.16 [1.13-1.18] | 1.18 [1.15-1.20] | 1.14 [1.11-1.16] | 1.14 [1.11-1.16] | 0.96 [0.80-1.16] | 1.13 [1.11-1.16] | 23,896 | 23,737 | 23,254 | 315 | 22,215 |
| 1,120 | 1.28 [1.24-1.31] | 1.28 [1.25-1.32] | 1.29 [1.26-1.33] | 1.08 [0.87-1.33] | 1.17 [1.14-1.21] | 1.23 [1.20-1.27] | 1.17 [1.14-1.20] | 1.17 [1.14-1.21] | 1.08 [0.87-1.34] | 1.16 [1.12-1.19] | 13,243 | 13,058 | 12,889 | 224 | 11,938 |
| 1,130 | 1.22 [1.21-1.23] | 1.22 [1.21-1.24] | 1.22 [1.21-1.24] | 1.06 [0.87-1.31] | 1.15 [1.13-1.16] | 1.16 [1.15-1.18] | 1.12 [1.11-1.13] | 1.11 [1.10-1.13] | 1.06 [0.86-1.31] | 1.11 [1.10-1.13] | 84,291 | 84,251 | 83,224 | 233 | 68,531 |
| 1,140 | 1.16 [1.14-1.17] | 1.16 [1.14-1.17] | 1.16 [1.14-1.17] | 0.99 [0.86-1.13] | 1.12 [1.11-1.14] | 1.13 [1.12-1.15] | 1.10 [1.08-1.11] | 1.10 [1.08-1.11] | 0.99 [0.86-1.14] | 1.12 [1.10-1.13] | 60,756 | 60,404 | 59,576 | 523 | 53,772 |
| 1,150 | 1.23 [1.20-1.27] | 1.24 [1.21-1.28] | 1.24 [1.21-1.28] | 1.07 [0.97-1.19] | 1.16 [1.13-1.19] | 1.18 [1.14-1.21] | 1.14 [1.11-1.18] | 1.14 [1.11-1.18] | 1.06 [0.95-1.18] | 1.13 [1.09-1.16] | 13,408 | 12,890 | 11,745 | 917 | 11,840 |
| 1,160 | 1.24 [1.23-1.25] | 1.24 [1.23-1.25] | 1.24 [1.23-1.25] | 1.15 [1.11-1.18] | 1.18 [1.17-1.19] | 1.20 [1.19-1.21] | 1.16 [1.15-1.17] | 1.16 [1.15-1.16] | 1.11 [1.07-1.15] | 1.15 [1.14-1.16] | 193,587 | 187,338 | 175,285 | 9,860 | 159,624 |
| Respiratory diseases | | | | | | | | | | | | | | | |
| 1,170 | 1.62 [1.61-1.63] | 1.55 [1.54-1.56] | 1.46 [1.45-1.47] | 1.88 [1.85-1.90] | 1.49 [1.48-1.50] | 1.39 [1.38-1.40] | 1.24 [1.23-1.25] | 1.22 [1.21-1.23] | 1.65 [1.63-1.67] | 1.32 [1.31-1.33] | 296,188 | 223,550 | 171,643 | 91,824 | 226,829 |
| 1,180 | 1.44 [1.42-1.46] | 1.44 [1.42-1.45] | 1.43 [1.42-1.45] | 1.47 [1.29-1.69] | 1.34 [1.32-1.36] | 1.28 [1.27-1.30] | 1.20 [1.18-1.22] | 1.20 [1.19-1.22] | 1.34 [1.16-1.54] | 1.22 [1.20-1.24] | 68,734 | 68,403 | 67,240 | 607 | 59,581 |
| 1,190 | 2.12 [2.11-2.14] | 1.98 [1.96-1.99] | 1.84 [1.82-1.86] | 2.50 [2.46-2.53] | 1.93 [1.91-1.95] | 1.61 [1.60-1.63] | 1.34 [1.33-1.36] | 1.31 [1.29-1.33] | 2.06 [2.03-2.09] | 1.51 [1.50-1.53] | 165,315 | 117,360 | 75,382 | 63,614 | 122,153 |
| 1,200 | 1.33 [1.32-1.34] | 1.31 [1.30-1.32] | 1.29 [1.28-1.30] | 1.41 [1.39-1.44] | 1.26 [1.25-1.27] | 1.25 [1.24-1.26] | 1.17 [1.16-1.18] | 1.17 [1.16-1.18] | 1.33 [1.31-1.35] | 1.20 [1.19-1.21] | 165,151 | 130,657 | 112,436 | 41,511 | 141,040 |
| Digestive diseases | | | | | | | | | | | | | | | |

### Supplementary Table 5: GBD-mapped results (hazard ratios and events from all cohorts)

| Outcome | Hazard ratio (99% confidence interval) | | | | | | | | | | Events (in exposed) | | | | |
|---|---|---|---|---|---|---|---|---|---|---|---|---|---|---|---|
| | crude | | | | | adjusted | | | | | | | | | |
| | any age | 18+ | 40+ | <18 | hosp. | any age | 18+ | 40+ | <18 | hosp. | any age | 18+ | 40+ | <18 | hosp. |
| 1,210 | 1.28 [1.27-1.29] | 1.29 [1.28-1.30] | 1.29 [1.28-1.30] | 1.19 [1.17-1.20] | 1.19 [1.19-1.20] | 1.21 [1.20-1.21] | 1.18 [1.17-1.19] | 1.18 [1.17-1.19] | 1.14 [1.12-1.16] | 1.14 [1.14-1.15] | 315,904 | 282,825 | 227,352 | 50,932 | 238,006 |
| 1,220 | 1.26 [1.23-1.29] | 1.27 [1.24-1.29] | 1.26 [1.23-1.29] | 1.26 [1.11-1.44] | 1.19 [1.16-1.21] | 1.20 [1.17-1.23] | 1.16 [1.14-1.19] | 1.16 [1.13-1.18] | 1.23 [1.08-1.42] | 1.16 [1.13-1.18] | 20,957 | 20,678 | 19,339 | 624 | 18,027 |
| 1,230 | 1.43 [1.38-1.48] | 1.45 [1.40-1.50] | 1.45 [1.40-1.50] | 1.09 [0.84-1.42] | 1.34 [1.29-1.38] | 1.34 [1.30-1.39] | 1.28 [1.24-1.33] | 1.28 [1.24-1.33] | 1.03 [0.79-1.35] | 1.31 [1.26-1.36] | 8,739 | 8,692 | 8,048 | 147 | 7,676 |
| 1,240 | 1.09 [1.06-1.11] | 1.10 [1.07-1.13] | 1.06 [1.02-1.12] | 1.05 [1.02-1.09] | 1.02 [0.99-1.05] | 1.05 [1.02-1.08] | 1.03 [1.00-1.07] | 1.01 [0.97-1.06] | 1.03 [0.99-1.06] | 1.00 [0.97-1.03] | 16,027 | 9,242 | 3,848 | 9,326 | 11,863 |
| 1,241 | 1.32 [1.31-1.34] | 1.33 [1.31-1.35] | 1.33 [1.31-1.35] | 1.23 [1.17-1.28] | 1.22 [1.21-1.24] | 1.22 [1.21-1.24] | 1.19 [1.17-1.20] | 1.19 [1.17-1.20] | 1.16 [1.11-1.22] | 1.16 [1.15-1.18] | 69,990 | 66,962 | 59,204 | 5,411 | 60,277 |
| 1,242 | 1.21 [1.18-1.24] | 1.21 [1.18-1.24] | 1.20 [1.17-1.23] | 1.08 [0.99-1.18] | 1.14 [1.11-1.17] | 1.15 [1.12-1.17] | 1.11 [1.09-1.14] | 1.11 [1.08-1.14] | 1.04 [0.95-1.14] | 1.10 [1.07-1.13] | 17,069 | 16,153 | 14,775 | 1,354 | 15,433 |
| 1,244 | 1.38 [1.36-1.40] | 1.40 [1.38-1.42] | 1.37 [1.35-1.39] | 1.28 [1.24-1.32] | 1.26 [1.24-1.28] | 1.28 [1.26-1.30] | 1.25 [1.23-1.27] | 1.23 [1.21-1.25] | 1.23 [1.19-1.27] | 1.21 [1.19-1.22] | 49,842 | 43,247 | 34,394 | 9,371 | 41,117 |
| 1,246 | 1.17 [1.16-1.19] | 1.18 [1.16-1.19] | 1.17 [1.15-1.18] | 1.05 [0.99-1.12] | 1.11 [1.09-1.13] | 1.11 [1.10-1.13] | 1.08 [1.07-1.10] | 1.08 [1.06-1.09] | 1.02 [0.96-1.09] | 1.08 [1.06-1.09] | 49,054 | 48,597 | 40,227 | 2,480 | 42,828 |
| 1,248 | 1.20 [1.16-1.23] | 1.22 [1.18-1.25] | 1.20 [1.16-1.24] | 0.95 [0.84-1.07] | 1.13 [1.10-1.17] | 1.14 [1.10-1.17] | 1.11 [1.08-1.14] | 1.09 [1.06-1.13] | 0.93 [0.82-1.06] | 1.10 [1.07-1.14] | 11,248 | 11,026 | 9,607 | 635 | 9,978 |
| 1,250 | 1.30 [1.29-1.31] | 1.31 [1.30-1.31] | 1.30 [1.29-1.30] | 1.23 [1.21-1.25] | 1.21 [1.20-1.22] | 1.21 [1.21-1.22] | 1.18 [1.17-1.19] | 1.18 [1.17-1.19] | 1.17 [1.15-1.19] | 1.15 [1.15-1.16] | 283,770 | 261,648 | 216,773 | 36,299 | 224,276 |
| Genitourinary diseases | | | | | | | | | | | | | | | |
| 1,260 | 1.22 [1.21-1.23] | 1.22 [1.21-1.23] | 1.22 [1.21-1.23] | 1.19 [1.17-1.20] | 1.15 [1.14-1.16] | 1.17 [1.16-1.18] | 1.14 [1.13-1.14] | 1.14 [1.13-1.15] | 1.15 [1.13-1.17] | 1.12 [1.12-1.13] | 263,963 | 236,324 | 191,230 | 42,723 | 211,374 |
| 1,270 | 1.22 [1.21-1.23] | 1.22 [1.21-1.23] | 1.22 [1.21-1.23] | 1.07 [1.02-1.11] | 1.16 [1.15-1.17] | 1.17 [1.16-1.18] | 1.13 [1.12-1.14] | 1.13 [1.12-1.14] | 1.04 [1.00-1.09] | 1.14 [1.13-1.15] | 124,174 | 121,174 | 114,202 | 5,781 | 112,699 |
| 1,271 | 1.35 [1.06-1.73] | 1.31 [0.98-1.76] | 1.34 [0.98-1.83] | | 1.20 [0.93-1.54] | 1.34 [1.04-1.73] | 1.26 [0.93-1.70] | 1.24 [0.90-1.71] | | 1.18 [0.91-1.53] | 174 | 121 | 101 | NA | 160 |
| 1,272 | 1.45 [1.37-1.55] | 1.45 [1.36-1.54] | 1.45 [1.36-1.55] | | 1.29 [1.21-1.37] | 1.35 [1.27-1.44] | 1.24 [1.16-1.32] | 1.22 [1.14-1.31] | | 1.27 [1.19-1.35] | 2,642 | 2,638 | 2,538 | NA | 2,542 |
| 1,273 | 1.22 [1.21-1.23] | 1.22 [1.21-1.23] | 1.22 [1.21-1.23] | 1.07 [1.02-1.11] | 1.16 [1.15-1.17] | 1.17 [1.16-1.18] | 1.13 [1.12-1.14] | 1.13 [1.12-1.14] | 1.04 [1.00-1.09] | 1.14 [1.13-1.15] | 124,119 | 121,149 | 114,183 | 5,750 | 112,653 |
| 1,280 | 1.16 [1.14-1.19] | 1.16 [1.14-1.18] | 1.16 [1.14-1.18] | | 1.10 [1.08-1.12] | 1.12 [1.10-1.14] | 1.10 [1.08-1.12] | 1.10 [1.08-1.12] | | 1.07 [1.05-1.09] | 30,069 | 30,057 | 29,975 | NA | 26,000 |
| 1,290 | 1.14 [1.11-1.17] | 1.14 [1.11-1.17] | 1.13 [1.10-1.17] | 0.97 [0.88-1.07] | 1.06 [1.03-1.09] | 1.09 [1.06-1.12] | 1.06 [1.04-1.09] | 1.06 [1.03-1.09] | 0.94 [0.85-1.04] | 1.04 [1.01-1.07] | 14,150 | 13,909 | 11,059 | 1,052 | 11,806 |
| 1,300 | 1.23 [1.22-1.24] | 1.22 [1.21-1.24] | 1.22 [1.20-1.23] | 1.22 [1.19-1.24] | 1.15 [1.14-1.16] | 1.18 [1.17-1.19] | 1.14 [1.13-1.15] | 1.13 [1.12-1.15] | 1.18 [1.16-1.21] | 1.13 [1.12-1.14] | 147,268 | 124,802 | 109,348 | 28,564 | 124,457 |
| 1,310 | 0.95 [0.87-1.03] | 0.97 [0.89-1.05] | 1.00 [0.78-1.27] | | 0.98 [0.89-1.09] | 0.94 [0.86-1.02] | 0.95 [0.87-1.03] | 1.00 [0.78-1.29] | | 0.97 [0.87-1.07] | 1,353 | 1,353 | 137 | NA | 823 |
| 1,320 | 1.19 [1.18-1.21] | 1.20 [1.18-1.21] | 1.19 [1.18-1.21] | 1.15 [1.12-1.19] | 1.13 [1.11-1.14] | 1.14 [1.13-1.15] | 1.11 [1.10-1.13] | 1.12 [1.11-1.14] | 1.11 [1.08-1.15] | 1.10 [1.08-1.11] | 80,226 | 76,332 | 47,501 | 12,194 | 61,923 |
| Skin diseases | | | | | | | | | | | | | | | |
| 1,330 | 1.96 [1.94-1.97] | 1.74 [1.72-1.75] | 1.67 [1.66-1.69] | 2.81 [2.77-2.86] | 1.83 [1.82-1.85] | 1.86 [1.85-1.88] | 1.60 [1.59-1.62] | 1.55 [1.54-1.57] | 2.69 [2.65-2.73] | 1.77 [1.75-1.78] | 212,545 | 160,139 | 127,239 | 64,263 | 175,421 |
| Musculoskeletal diseases | | | | | | | | | | | | | | | |
| 1,340 | 1.29 [1.28-1.30] | 1.29 [1.29-1.30] | 1.30 [1.29-1.31] | 1.17 [1.15-1.19] | 1.20 [1.19-1.21] | 1.21 [1.20-1.22] | 1.18 [1.17-1.18] | 1.18 [1.17-1.19] | 1.12 [1.10-1.15] | 1.15 [1.14-1.15] | 267,151 | 245,258 | 205,775 | 34,599 | 208,501 |
| 1,350 | 1.41 [1.38-1.44] | 1.40 [1.37-1.43] | 1.39 [1.36-1.42] | 1.27 [1.05-1.54] | 1.31 [1.28-1.35] | 1.32 [1.28-1.35] | 1.27 [1.24-1.30] | 1.26 [1.23-1.29] | 1.20 [0.98-1.47] | 1.24 [1.21-1.28] | 17,774 | 17,725 | 16,515 | 291 | 15,460 |
| 1,360 | 1.28 [1.27-1.29] | 1.28 [1.27-1.29] | 1.28 [1.27-1.29] | 1.18 [1.05-1.32] | 1.20 [1.19-1.21] | 1.20 [1.19-1.21] | 1.17 [1.16-1.18] | 1.16 [1.15-1.17] | 1.10 [0.98-1.24] | 1.14 [1.13-1.15] | 117,124 | 117,031 | 113,542 | 802 | 100,636 |
| 1,370 | 1.36 [1.33-1.39] | 1.36 [1.33-1.39] | 1.36 [1.33-1.39] | | 1.27 [1.24-1.30] | 1.29 [1.26-1.32] | 1.24 [1.21-1.27] | 1.23 [1.20-1.26] | | 1.22 [1.19-1.25] | 19,181 | 19,179 | 18,861 | NA | 17,324 |
| 1,380 | 1.30 [1.28-1.31] | 1.30 [1.29-1.31] | 1.30 [1.29-1.32] | 1.15 [1.11-1.20] | 1.20 [1.19-1.21] | 1.20 [1.19-1.21] | 1.15 [1.14-1.16] | 1.15 [1.14-1.17] | 1.10 [1.05-1.14] | 1.14 [1.13-1.15] | 96,513 | 93,934 | 79,147 | 6,828 | 84,881 |
| 1,390 | 1.30 [1.29-1.30] | 1.30 [1.29-1.31] | 1.31 [1.30-1.32] | 1.18 [1.16-1.20] | 1.20 [1.19-1.21] | 1.21 [1.20-1.22] | 1.18 [1.17-1.19] | 1.19 [1.18-1.20] | 1.13 [1.11-1.15] | 1.15 [1.14-1.16] | 209,238 | 189,096 | 159,953 | 29,805 | 171,806 |
| Congenital anomalies | | | | | | | | | | | | | | | |
| 1,400 | 1.19 [1.17-1.21] | 1.26 [1.23-1.29] | 1.29 [1.25-1.33] | 1.09 [1.06-1.12] | 1.11 [1.09-1.13] | 1.13 [1.11-1.15] | 1.15 [1.12-1.18] | 1.17 [1.14-1.21] | 1.06 [1.04-1.09] | 1.08 [1.05-1.10] | 32,251 | 17,025 | 11,591 | 17,158 | 25,999 |
| 1,410 | 1.28 [1.11-1.48] | 1.32 [1.13-1.54] | 1.32 [1.10-1.59] | 1.08 [0.81-1.45] | 1.17 [1.01-1.37] | 1.17 [1.01-1.36] | 1.19 [1.01-1.39] | 1.16 [0.96-1.41] | 1.02 [0.76-1.38] | 1.09 [0.93-1.28] | 476 | 404 | 268 | 116 | 395 |
| 1,420 | 1.03 [0.87-1.21] | | | 1.02 [0.85-1.22] | 0.98 [0.82-1.18] | 0.98 [0.83-1.16] | | | 0.96 [0.80-1.16] | 0.96 [0.79-1.15] | 348 | NA | NA | 286 | 292 |
| 1,430 | 2.77 [2.46-3.12] | 3.76 [3.28-4.32] | 4.16 [3.54-4.89] | 1.03 [0.81-1.31] | 2.00 [1.68-2.37] | 2.68 [2.37-3.02] | 3.86 [3.35-4.45] | 4.05 [3.44-4.78] | 1.00 [0.79-1.29] | 1.90 [1.59-2.27] | 855 | 725 | 526 | 161 | 367 |
| 1,440 | 1.17 [1.12-1.22] | 1.22 [1.15-1.28] | 1.23 [1.16-1.31] | 1.04 [0.96-1.11] | 1.10 [1.05-1.16] | 1.12 [1.07-1.17] | 1.13 [1.07-1.19] | 1.14 [1.07-1.21] | 1.02 [0.94-1.09] | 1.07 [1.02-1.12] | 5,055 | 3,427 | 2,545 | 1,913 | 4,275 |
| 1,450 | 1.29 [1.18-1.41] | 1.54 [1.33-1.78] | 1.71 [1.40-2.08] | 1.14 [1.02-1.27] | 1.22 [1.11-1.34] | 1.21 [1.11-1.33] | 1.38 [1.19-1.61] | 1.55 [1.26-1.89] | 1.09 [0.97-1.21] | 1.16 [1.05-1.28] | 1,285 | 485 | 262 | 873 | 1,126 |
| 1,460 | 1.17 [1.14-1.19] | 1.23 [1.20-1.26] | 1.26 [1.21-1.30] | 1.09 [1.07-1.12] | 1.10 [1.08-1.13] | 1.11 [1.09-1.13] | 1.11 [1.08-1.14] | 1.14 [1.10-1.18] | 1.06 [1.04-1.09] | 1.07 [1.04-1.09] | 26,608 | 12,766 | 8,409 | 15,480 | 21,781 |
| Oral conditions | | | | | | | | | | | | | | | |
| 1,470 | 1.18 [1.17-1.19] | 1.27 [1.25-1.29] | 1.29 [1.26-1.31] | 1.08 [1.07-1.10] | 1.11 [1.10-1.12] | 1.13 [1.12-1.14] | 1.15 [1.14-1.17] | 1.17 [1.15-1.19] | 1.06 [1.04-1.08] | 1.08 [1.07-1.09] | 91,561 | 50,669 | 26,576 | 49,756 | 70,760 |
| 1,480 | 1.11 [1.09-1.13] | 1.25 [1.22-1.28] | 1.28 [1.24-1.32] | 1.02 [1.00-1.04] | 1.05 [1.03-1.07] | 1.07 [1.05-1.09] | 1.12 [1.10-1.15] | 1.15 [1.11-1.19] | 1.01 [0.99-1.03] | 1.03 [1.01-1.05] | 42,689 | 17,603 | 8,465 | 28,068 | 34,584 |
| 1,490 | 1.29 [1.25-1.34] | 1.33 [1.28-1.38] | 1.33 [1.24-1.42] | 1.21 [1.14-1.29] | 1.19 [1.15-1.24] | 1.22 [1.17-1.26] | 1.20 [1.15-1.24] | 1.21 [1.13-1.29] | 1.16 [1.09-1.24] | 1.15 [1.10-1.19] | 8,466 | 7,996 | 2,186 | 2,842 | 6,400 |
| 1,502 | 1.22 [1.20-1.24] | 1.27 [1.25-1.29] | 1.30 [1.27-1.32] | 1.13 [1.11-1.15] | 1.15 [1.13-1.16] | 1.16 [1.15-1.18] | 1.16 [1.14-1.18] | 1.18 [1.15-1.21] | 1.10 [1.08-1.12] | 1.11 [1.10-1.13] | 61,180 | 38,767 | 21,493 | 29,069 | 47,317 |
| Unintentional injuries | | | | | | | | | | | | | | | |
| 1,520 | 1.16 [1.16-1.17] | 1.21 [1.20-1.22] | 1.21 [1.20-1.22] | 1.06 [1.04-1.07] | 1.11 [1.10-1.12] | 1.12 [1.11-1.13] | 1.13 [1.12-1.14] | 1.13 [1.12-1.14] | 1.04 [1.02-1.05] | 1.09 [1.08-1.10] | 246,179 | 184,392 | 152,605 | 74,802 | 198,640 |
| 1,530 | 1.16 [1.16-1.17] | 1.21 [1.20-1.22] | 1.21 [1.20-1.22] | 1.06 [1.04-1.07] | 1.11 [1.10-1.12] | 1.12 [1.11-1.13] | 1.13 [1.12-1.14] | 1.13 [1.12-1.14] | 1.04 [1.02-1.05] | 1.09 [1.08-1.10] | 246,179 | 184,392 | 152,605 | 74,802 | 198,640 |
| 1,540 | 1.13 [1.08-1.17] | 1.19 [1.13-1.27] | 1.25 [1.16-1.35] | 1.07 [1.02-1.13] | 1.07 [1.03-1.12] | 1.08 [1.03-1.12] | 1.05 [0.99-1.12] | 1.09 [1.01-1.18] | 1.05 [0.99-1.11] | 1.05 [1.00-1.10] | 5,432 | 2,840 | 1,623 | 3,151 | 4,586 |
| 1,550 | 1.14 [1.13-1.15] | 1.17 [1.16-1.19] | 1.18 [1.17-1.19] | 1.02 [1.00-1.04] | 1.10 [1.09-1.11] | 1.10 [1.09-1.12] | 1.11 [1.09-1.12] | 1.11 [1.10-1.13] | 1.01 [0.99-1.03] | 1.08 [1.07-1.10] | 123,750 | 97,427 | 89,267 | 29,486 | 104,225 |
| 1,560 | 1.01 [0.96-1.06] | 1.11 [1.02-1.20] | 1.12 [1.02-1.23] | 0.96 [0.90-1.03] | 0.98 [0.92-1.04] | 0.99 [0.93-1.04] | 1.03 [0.95-1.12] | 1.04 [0.94-1.14] | 0.95 [0.89-1.02] | 0.97 [0.91-1.03] | 3,270 | 1,460 | 1,057 | 2,004 | 2,799 |
| 1,570 | 0.88 [0.67-1.14] | | | | 0.83 [0.62-1.11] | 0.84 [0.64-1.10] | | | | 0.81 [0.60-1.09] | 128 | NA | NA | NA | 104 |
| 1,575 | 1.06 [1.04-1.08] | 1.11 [1.08-1.13] | 1.12 [1.09-1.16] | 1.02 [1.00-1.05] | 1.01 [0.99-1.04] | 1.03 [1.01-1.05] | 1.04 [1.01-1.06] | 1.05 [1.02-1.09] | 1.01 [0.98-1.04] | 1.00 [0.98-1.02] | 28,689 | 16,072 | 8,886 | 16,270 | 22,319 |
| 1,580 | 0.98 [0.72-1.33] | | | | | 0.92 [0.67-1.27] | | | | | 105 | NA | NA | NA | NA |
| 1,590 | 1.25 [1.24-1.26] | 1.28 [1.27-1.29] | 1.27 [1.26-1.28] | 1.14 [1.12-1.16] | 1.17 [1.16-1.19] | 1.19 [1.18-1.20] | 1.17 [1.16-1.18] | 1.17 [1.15-1.18] | 1.10 [1.08-1.13] | 1.14 [1.13-1.15] | 123,759 | 105,231 | 88,344 | 24,731 | 106,478 |
| Intentional injuries | | | | | | | | | | | | | | | |
| 1,600 | 1.10 [1.08-1.12] | 1.14 [1.11-1.16] | 1.27 [1.23-1.32] | 1.00 [0.98-1.03] | 1.01 [0.99-1.04] | 1.05 [1.03-1.07] | 1.00 [0.98-1.03] | 1.09 [1.05-1.13] | 0.99 [0.96-1.02] | 1.00 [0.98-1.03] | 23,875 | 16,855 | 6,452 | 12,388 | 17,833 |
| 1,610 | 1.11 [1.09-1.14] | 1.14 [1.11-1.17] | 1.28 [1.23-1.34] | 1.00 [0.97-1.04] | 1.02 [1.00-1.05] | 1.05 [1.03-1.08] | 0.99 [0.96-1.02] | 1.09 [1.04-1.14] | 0.99 [0.96-1.03] | 1.01 [0.98-1.04] | 17,707 | 12,089 | 5,089 | 9,073 | 13,698 |
| 1,620 | 1.07 [1.03-1.11] | 1.13 [1.08-1.18] | 1.24 [1.15-1.34] | 1.01 [0.96-1.07] | 0.98 [0.94-1.03] | 1.03 [0.99-1.07] | 1.03 [0.98-1.07] | 1.08 [1.00-1.17] | 1.00 [0.95-1.05] | 0.98 [0.94-1.02] | 6,784 | 5,326 | 1,554 | 3,482 | 4,828 |
