## Supplementary Table 6 for "Mapping risks of hospital-recorded health conditions in people with eczema"

### Supplementary Table 6: phecode-mapped results (hazard ratios and events from all cohorts)

| | Hazard ratio (99% confidence interval) | | | | | | | | | | | | | | |
|---|---|---|---|---|---|---|---|---|---|---|---|---|---|---|---|
| | crude | | | | | adjusted | | | | | Events (in exposed) | | | | |
| Outcome | any age | 18+ | 40+ | <18 | hosp. | any age | 18+ | 40+ | <18 | hosp. | any age | 18+ | 40+ | <18 | hosp. |
| infectious diseases | | | | | | | | | | | | | | | |
| 8 | 1.29 [1.27-1.31] | 1.32 [1.30-1.34] | 1.29 [1.27-1.32] | 1.19 [1.15-1.23] | 1.21 [1.19-1.23] | 1.20 [1.18-1.22] | 1.17 [1.15-1.19] | 1.15 [1.13-1.17] | 1.14 [1.10-1.18] | 1.16 [1.14-1.18] | 51,853 | 44,673 | 36,068 | 10,855 | 47,318 |
| 8.5 | 1.25 [1.20-1.31] | 1.26 [1.20-1.32] | 1.28 [1.21-1.34] | 1.11 [0.99-1.25] | 1.15 [1.10-1.21] | 1.17 [1.11-1.22] | 1.14 [1.09-1.20] | 1.15 [1.09-1.21] | 1.07 [0.95-1.21] | 1.09 [1.04-1.15] | 4,846 | 4,328 | 3,579 | 751 | 4,083 |
| 8.51 | 1.20 [0.95-1.50] | 1.29 [0.96-1.74] | | | 1.18 [0.92-1.51] | 1.13 [0.90-1.43] | 1.22 [0.90-1.65] | | | 1.15 [0.90-1.48] | 187 | 110 | NA | NA | 162 |
| 8.52 | 1.28 [1.23-1.33] | 1.28 [1.23-1.33] | 1.27 [1.22-1.32] | 1.16 [0.94-1.43] | 1.21 [1.17-1.26] | 1.23 [1.19-1.28] | 1.19 [1.15-1.24] | 1.18 [1.14-1.23] | 1.10 [0.88-1.37] | 1.19 [1.14-1.24] | 6,908 | 6,779 | 6,507 | 226 | 6,244 |
| 8.6 | 1.20 [1.17-1.23] | 1.31 [1.26-1.37] | 1.31 [1.25-1.38] | 1.14 [1.10-1.17] | 1.15 [1.12-1.18] | 1.15 [1.12-1.18] | 1.17 [1.12-1.22] | 1.17 [1.12-1.23] | 1.11 [1.08-1.15] | 1.12 [1.09-1.16] | 14,673 | 5,645 | 4,555 | 9,469 | 13,130 |
| 8.7 | 1.19 [0.97-1.47] | 1.21 [0.93-1.57] | | 1.48 [1.07-2.04] | 1.14 [0.90-1.43] | 1.12 [0.90-1.39] | 1.06 [0.81-1.39] | | 1.40 [1.01-1.95] | 1.10 [0.88-1.39] | 221 | 139 | NA | 101 | 185 |
| 10 | 1.26 [1.15-1.38] | 1.33 [1.21-1.45] | 1.36 [1.23-1.51] | 0.89 [0.70-1.14] | 1.28 [1.16-1.41] | 1.20 [1.10-1.32] | 1.25 [1.14-1.37] | 1.25 [1.12-1.39] | 0.95 [0.74-1.22] | 1.22 [1.11-1.35] | 1,244 | 1,150 | 905 | 165 | 1,025 |
| 31 | 1.39 [1.04-1.85] | | | | 1.28 [0.95-1.72] | 1.26 [0.94-1.70] | | | | 1.13 [0.83-1.53] | 117 | NA | NA | NA | 110 |
| 38 | 1.24 [1.22-1.26] | 1.25 [1.23-1.27] | 1.25 [1.22-1.27] | 1.07 [1.00-1.16] | 1.18 [1.16-1.21] | 1.20 [1.18-1.22] | 1.17 [1.15-1.19] | 1.16 [1.14-1.18] | 1.05 [0.97-1.13] | 1.17 [1.15-1.19] | 35,491 | 34,297 | 32,322 | 1,893 | 32,907 |
| 38.1 | 1.21 [1.17-1.25] | 1.21 [1.17-1.26] | 1.23 [1.19-1.28] | 1.03 [0.92-1.16] | 1.15 [1.12-1.20] | 1.17 [1.13-1.21] | 1.13 [1.09-1.17] | 1.14 [1.10-1.19] | 1.02 [0.90-1.15] | 1.14 [1.10-1.18] | 9,006 | 8,421 | 8,036 | 710 | 8,335 |
| 38.2 | 1.51 [1.44-1.59] | 1.52 [1.44-1.60] | 1.52 [1.44-1.60] | 1.27 [1.08-1.50] | 1.40 [1.33-1.47] | 1.45 [1.38-1.53] | 1.41 [1.34-1.48] | 1.41 [1.33-1.49] | 1.23 [1.04-1.45] | 1.38 [1.31-1.45] | 4,414 | 4,102 | 3,811 | 391 | 3,973 |
| 41 | 1.29 [1.27-1.32] | 1.31 [1.29-1.34] | 1.31 [1.29-1.34] | 1.13 [1.07-1.19] | 1.22 [1.20-1.25] | 1.23 [1.21-1.25] | 1.20 [1.18-1.23] | 1.20 [1.18-1.22] | 1.09 [1.04-1.15] | 1.19 [1.17-1.21] | 34,181 | 31,214 | 28,566 | 3,812 | 30,330 |
| 41.1 | 1.60 [1.57-1.64] | 1.59 [1.56-1.63] | 1.57 [1.54-1.61] | 1.70 [1.61-1.80] | 1.50 [1.46-1.53] | 1.53 [1.50-1.57] | 1.46 [1.43-1.50] | 1.45 [1.41-1.48] | 1.65 [1.56-1.75] | 1.46 [1.43-1.50] | 23,132 | 20,137 | 17,620 | 3,862 | 20,430 |
| 41.2 | 1.38 [1.34-1.42] | 1.40 [1.36-1.44] | 1.40 [1.35-1.45] | 1.25 [1.18-1.32] | 1.29 [1.25-1.33] | 1.32 [1.29-1.36] | 1.29 [1.26-1.34] | 1.29 [1.24-1.33] | 1.22 [1.15-1.29] | 1.27 [1.23-1.31] | 13,748 | 11,189 | 8,920 | 3,252 | 12,204 |
| 41.21 | 1.06 [0.81-1.39] | 1.00 [0.74-1.34] | 1.10 [0.82-1.49] | | 1.03 [0.78-1.37] | 1.00 [0.76-1.32] | 0.90 [0.67-1.22] | 1.03 [0.76-1.40] | | 1.00 [0.75-1.33] | 130 | 107 | 105 | NA | 117 |
| 41.4 | 1.17 [1.15-1.19] | 1.18 [1.15-1.20] | 1.19 [1.16-1.21] | 1.04 [0.98-1.11] | 1.11 [1.09-1.14] | 1.13 [1.10-1.15] | 1.09 [1.07-1.11] | 1.10 [1.08-1.13] | 1.03 [0.96-1.09] | 1.10 [1.07-1.12] | 26,594 | 24,751 | 22,561 | 2,705 | 24,487 |
| 53 | 1.42 [1.34-1.50] | 1.41 [1.33-1.49] | 1.40 [1.32-1.49] | 1.41 [1.20-1.67] | 1.32 [1.24-1.40] | 1.34 [1.26-1.42] | 1.29 [1.21-1.37] | 1.29 [1.21-1.37] | 1.35 [1.13-1.60] | 1.26 [1.19-1.33] | 3,475 | 3,158 | 2,946 | 394 | 3,139 |
| 53.1 | 1.29 [1.13-1.47] | 1.32 [1.16-1.51] | 1.33 [1.16-1.52] | | 1.24 [1.08-1.42] | 1.22 [1.07-1.40] | 1.21 [1.06-1.39] | 1.24 [1.08-1.42] | | 1.19 [1.03-1.36] | 621 | 608 | 584 | NA | 571 |
| 54 | 2.46 [2.33-2.60] | 2.02 [1.87-2.19] | 1.73 [1.57-1.90] | 3.00 [2.78-3.24] | 2.28 [2.14-2.42] | 2.29 [2.16-2.42] | 1.77 [1.63-1.92] | 1.54 [1.39-1.69] | 2.85 [2.63-3.08] | 2.15 [2.02-2.29] | 3,948 | 1,866 | 1,186 | 2,363 | 3,327 |
| 70 | 1.20 [1.12-1.28] | 1.23 [1.15-1.32] | 1.29 [1.19-1.39] | 1.06 [0.79-1.41] | 1.10 [1.02-1.18] | 1.16 [1.08-1.25] | 1.14 [1.06-1.22] | 1.17 [1.08-1.27] | 1.18 [0.87-1.59] | 1.11 [1.03-1.19] | 2,021 | 1,972 | 1,495 | 123 | 1,714 |
| 70.1 | 1.10 [0.86-1.39] | 1.24 [0.95-1.60] | 1.35 [1.00-1.82] | | 1.06 [0.82-1.39] | 1.06 [0.83-1.36] | 1.17 [0.89-1.53] | 1.26 [0.93-1.72] | | 1.03 [0.79-1.35] | 171 | 142 | 108 | NA | 139 |
| 70.2 | 1.21 [1.10-1.34] | 1.24 [1.13-1.37] | 1.38 [1.23-1.55] | | 1.11 [1.00-1.23] | 1.21 [1.10-1.35] | 1.23 [1.11-1.36] | 1.30 [1.16-1.47] | | 1.12 [1.00-1.25] | 973 | 963 | 720 | NA | 804 |
| 70.3 | 1.17 [1.07-1.29] | 1.22 [1.11-1.34] | 1.26 [1.14-1.40] | | 1.06 [0.96-1.17] | 1.12 [1.02-1.23] | 1.06 [0.96-1.16] | 1.12 [1.00-1.25] | | 1.07 [0.97-1.19] | 1,058 | 1,048 | 825 | NA | 888 |
| 70.4 | 1.42 [1.27-1.59] | 1.40 [1.24-1.56] | 1.42 [1.25-1.60] | | 1.34 [1.19-1.50] | 1.33 [1.18-1.49] | 1.26 [1.12-1.42] | 1.28 [1.13-1.46] | | 1.27 [1.13-1.44] | 799 | 736 | 640 | NA | 703 |
| 70.9 | 1.48 [1.39-1.58] | 1.52 [1.42-1.63] | 1.52 [1.41-1.63] | 1.28 [1.00-1.63] | 1.39 [1.30-1.49] | 1.36 [1.27-1.45] | 1.32 [1.23-1.42] | 1.31 [1.22-1.41] | 1.22 [0.95-1.57] | 1.33 [1.24-1.42] | 2,397 | 2,317 | 2,020 | 190 | 2,198 |
| 71.1 | 1.83 [1.38-2.43] | 1.90 [1.42-2.54] | | | | 1.65 [1.22-2.22] | 1.69 [1.25-2.30] | | | | 120 | 115 | NA | NA | NA |
| 78 | 1.41 [1.31-1.52] | 1.40 [1.29-1.52] | 1.42 [1.28-1.56] | 1.44 [1.22-1.69] | 1.33 [1.22-1.45] | 1.30 [1.20-1.41] | 1.25 [1.15-1.36] | 1.29 [1.17-1.43] | 1.32 [1.12-1.56] | 1.26 [1.15-1.37] | 1,723 | 1,411 | 1,005 | 409 | 1,392 |
| 79 | 1.50 [1.48-1.52] | 1.34 [1.30-1.38] | 1.33 [1.28-1.39] | 1.53 [1.51-1.56] | 1.44 [1.42-1.46] | 1.43 [1.41-1.45] | 1.17 [1.13-1.21] | 1.16 [1.11-1.21] | 1.49 [1.46-1.51] | 1.39 [1.37-1.41] | 48,807 | 10,855 | 6,395 | 39,583 | 43,067 |
| 79.1 | 1.61 [1.52-1.72] | 1.39 [1.14-1.68] | 1.43 [1.10-1.87] | 1.63 [1.53-1.74] | 1.53 [1.44-1.64] | 1.56 [1.46-1.66] | 1.27 [1.04-1.55] | 1.29 [0.99-1.70] | 1.58 [1.48-1.69] | 1.49 [1.40-1.60] | 2,710 | 260 | 139 | 2,482 | 2,395 |
| 79.2 | 1.15 [1.06-1.24] | 1.11 [1.00-1.24] | | 1.16 [1.06-1.26] | 1.08 [0.98-1.19] | 1.10 [1.01-1.19] | 1.02 [0.91-1.14] | | 1.10 [1.01-1.21] | 1.04 [0.94-1.15] | 1,551 | 787 | NA | 1,319 | 1,099 |
| 80 | 1.38 [1.35-1.42] | 1.39 [1.35-1.43] | 1.39 [1.35-1.43] | 1.25 [1.17-1.35] | 1.27 [1.24-1.31] | 1.30 [1.26-1.33] | 1.25 [1.22-1.29] | 1.26 [1.22-1.30] | 1.19 [1.11-1.28] | 1.23 [1.19-1.26] | 14,249 | 12,851 | 10,760 | 2,107 | 12,312 |
| 81 | 1.32 [1.27-1.38] | 1.36 [1.30-1.42] | 1.33 [1.27-1.39] | 1.19 [1.07-1.33] | 1.20 [1.14-1.25] | 1.27 [1.21-1.33] | 1.25 [1.20-1.31] | 1.22 [1.16-1.29] | 1.14 [1.02-1.27] | 1.19 [1.13-1.24] | 5,621 | 4,971 | 4,303 | 899 | 5,162 |
| 110.1 | 1.73 [1.61-1.86] | 1.78 [1.65-1.91] | 1.72 [1.59-1.86] | 1.48 [1.22-1.79] | 1.64 [1.53-1.77] | 1.59 [1.48-1.71] | 1.57 [1.45-1.70] | 1.53 [1.41-1.66] | 1.38 [1.13-1.69] | 1.55 [1.44-1.67] | 2,264 | 2,009 | 1,827 | 302 | 2,033 |
| 110.11 | 1.33 [1.12-1.57] | 1.45 [1.22-1.72] | 1.39 [1.16-1.65] | | 1.38 [1.16-1.64] | 1.18 [0.99-1.40] | 1.26 [1.05-1.50] | 1.22 [1.02-1.47] | | 1.25 [1.05-1.50] | 371 | 359 | 335 | NA | 340 |
| 110.12 | 1.89 [1.66-2.16] | 1.95 [1.70-2.23] | 1.87 [1.62-2.16] | | 1.71 [1.49-1.96] | 1.75 [1.53-2.01] | 1.73 [1.51-1.99] | 1.69 [1.45-1.96] | | 1.63 [1.41-1.88] | 648 | 617 | 535 | NA | 560 |
| 110.13 | 1.75 [1.50-2.04] | 1.76 [1.49-2.06] | 1.77 [1.50-2.09] | | 1.72 [1.47-2.01] | 1.61 [1.37-1.88] | 1.54 [1.30-1.81] | 1.54 [1.30-1.83] | | 1.63 [1.38-1.91] | 493 | 453 | 431 | NA | 459 |
| 110.2 | 1.87 [1.63-2.14] | 1.86 [1.62-2.14] | 1.77 [1.53-2.05] | | 1.68 [1.46-1.92] | 1.72 [1.50-1.97] | 1.66 [1.44-1.92] | 1.58 [1.36-1.84] | | 1.60 [1.39-1.84] | 651 | 592 | 538 | NA | 596 |
| 112 | 1.36 [1.33-1.39] | 1.37 [1.34-1.40] | 1.38 [1.34-1.41] | 1.11 [1.04-1.20] | 1.26 [1.23-1.29] | 1.25 [1.22-1.28] | 1.20 [1.17-1.23] | 1.21 [1.18-1.24] | 1.06 [0.99-1.14] | 1.19 [1.16-1.22] | 18,913 | 17,961 | 15,081 | 1,965 | 17,283 |
| 112.3 | 1.56 [1.37-1.78] | 1.61 [1.41-1.85] | 1.61 [1.40-1.86] | | 1.47 [1.28-1.68] | 1.46 [1.28-1.67] | 1.43 [1.24-1.65] | 1.44 [1.25-1.67] | | 1.42 [1.24-1.63] | 649 | 594 | 562 | NA | 604 |
| 117 | 1.57 [1.40-1.76] | 1.52 [1.35-1.71] | 1.52 [1.34-1.72] | 1.48 [1.10-2.00] | 1.45 [1.29-1.62] | 1.46 [1.30-1.64] | 1.36 [1.20-1.54] | 1.36 [1.20-1.55] | 1.46 [1.07-1.98] | 1.36 [1.21-1.53] | 829 | 722 | 658 | 120 | 761 |
| 117.4 | 1.68 [1.51-1.88] | 1.84 [1.64-2.06] | 1.89 [1.68-2.13] | | 1.51 [1.35-1.69] | 1.40 [1.24-1.57] | 1.46 [1.30-1.65] | 1.50 [1.32-1.70] | | 1.27 [1.13-1.43] | 857 | 811 | 745 | NA | 784 |
| 130 | 1.20 [0.97-1.49] | 1.18 [0.92-1.50] | 1.22 [0.92-1.62] | | 1.11 [0.88-1.40] | 1.13 [0.90-1.41] | 1.04 [0.81-1.34] | 1.10 [0.82-1.48] | | 1.05 [0.83-1.33] | 202 | 155 | 112 | NA | 175 |
| 130.1 | 1.20 [0.89-1.62] | | | | | 1.08 [0.79-1.47] | | | | | 109 | NA | NA | NA | NA |
| 132 | 2.28 [1.99-2.61] | 2.33 [2.01-2.70] | 2.36 [2.03-2.75] | | 2.00 [1.73-2.31] | 2.23 [1.95-2.56] | 2.18 [1.88-2.53] | 2.22 [1.90-2.59] | | 2.00 [1.72-2.31] | 669 | 574 | 548 | NA | 572 |
| 132.1 | 1.12 [0.91-1.37] | 1.19 [0.89-1.59] | | 0.95 [0.71-1.26] | 1.04 [0.83-1.30] | 1.11 [0.90-1.36] | 1.09 [0.81-1.47] | | 0.95 [0.71-1.26] | 1.03 [0.82-1.29] | 228 | 117 | NA | 115 | 189 |
| 133 | 0.97 [0.81-1.16] | 1.03 [0.85-1.27] | 1.00 [0.78-1.29] | | 0.92 [0.74-1.15] | 0.97 [0.80-1.16] | 1.08 [0.87-1.33] | 1.02 [0.79-1.32] | | 0.93 [0.74-1.17] | 302 | 223 | 141 | NA | 189 |
| 134 | 1.33 [1.05-1.68] | 1.54 [1.14-2.08] | | | 1.35 [1.06-1.74] | 1.29 [1.01-1.65] | 1.48 [1.08-2.02] | | | 1.35 [1.05-1.74] | 187 | 114 | NA | NA | 165 |
| 134.1 | 1.18 [1.06-1.32] | 1.20 [1.02-1.41] | 1.16 [0.95-1.42] | 1.14 [0.98-1.31] | 1.13 [1.00-1.27] | 1.12 [1.00-1.26] | 1.12 [0.95-1.32] | 1.08 [0.87-1.33] | 1.08 [0.93-1.26] | 1.08 [0.96-1.22] | 787 | 358 | 218 | 490 | 683 |
| 136 | 1.28 [1.19-1.37] | 1.29 [1.20-1.39] | 1.24 [1.15-1.35] | 1.23 [1.01-1.48] | 1.22 [1.14-1.32] | 1.22 [1.13-1.31] | 1.18 [1.09-1.27] | 1.15 [1.06-1.24] | 1.20 [0.99-1.45] | 1.20 [1.11-1.29] | 2,133 | 1,902 | 1,709 | 291 | 1,987 |
| neoplasms | | | | | | | | | | | | | | | |
| 145 | 1.20 [1.04-1.38] | 1.21 [1.05-1.40] | 1.20 [1.04-1.38] | | 1.13 [0.97-1.31] | 1.19 [1.03-1.37] | 1.15 [1.00-1.33] | 1.12 [0.97-1.30] | | 1.15 [0.98-1.34] | 472 | 475 | 452 | NA | 410 |
| 145.1 | 1.04 [0.81-1.35] | 1.08 [0.84-1.40] | 1.10 [0.85-1.42] | | 1.04 [0.80-1.36] | 1.03 [0.80-1.33] | 1.05 [0.81-1.36] | 1.05 [0.81-1.37] | | 1.03 [0.79-1.35] | 146 | 146 | 144 | NA | 128 |
| 145.2 | 1.23 [1.10-1.37] | 1.24 [1.11-1.39] | 1.23 [1.10-1.38] | | 1.23 [1.09-1.39] | 1.22 [1.09-1.36] | 1.19 [1.06-1.33] | 1.19 [1.06-1.34] | | 1.26 [1.11-1.42] | 754 | 754 | 735 | NA | 644 |
| 145.3 | 1.06 [0.89-1.27] | 1.07 [0.89-1.28] | 1.04 [0.86-1.26] | | 1.06 [0.87-1.29] | 1.05 [0.88-1.25] | 1.01 [0.84-1.22] | 0.99 [0.82-1.20] | | 1.04 [0.86-1.27] | 297 | 288 | 258 | NA | 246 |
| 145.4 | 1.47 [1.12-1.92] | 1.38 [1.06-1.81] | 1.44 [1.09-1.91] | | 1.32 [0.99-1.76] | 1.46 [1.11-1.91] | 1.38 [1.05-1.82] | 1.42 [1.07-1.88] | | 1.32 [0.98-1.77] | 139 | 139 | 132 | NA | 118 |
| 145.5 | 1.13 [0.88-1.45] | 1.15 [0.90-1.48] | 1.12 [0.87-1.44] | | 1.10 [0.84-1.43] | 1.14 [0.89-1.47] | 1.08 [0.84-1.39] | 1.06 [0.82-1.37] | | 1.13 [0.86-1.48] | 151 | 150 | 145 | NA | 128 |
| 149 | 1.10 [0.91-1.34] | 1.12 [0.92-1.36] | 1.07 [0.87-1.30] | | 0.95 [0.77-1.18] | 1.09 [0.89-1.33] | 1.07 [0.87-1.31] | 1.00 [0.82-1.23] | | 0.97 [0.78-1.20] | 235 | 235 | 234 | NA | 202 |
| 149.1 | 1.08 [0.95-1.24] | 1.12 [0.99-1.28] | 1.14 [1.00-1.30] | | 1.06 [0.92-1.22] | 1.07 [0.94-1.22] | 1.06 [0.93-1.22] | 1.09 [0.95-1.24] | | 1.08 [0.93-1.25] | 531 | 531 | 530 | NA | 435 |
| 149.2 | 1.19 [0.90-1.58] | 1.20 [0.90-1.60] | 1.36 [1.01-1.83] | | | 1.19 [0.89-1.58] | 1.17 [0.87-1.56] | 1.31 [0.97-1.77] | | | 115 | 111 | 108 | NA | NA |
| 149.3 | 1.21 [0.98-1.48] | 1.25 [1.02-1.54] | 1.28 [1.04-1.57] | | 1.22 [0.98-1.52] | 1.23 [1.00-1.52] | 1.23 [0.99-1.51] | 1.26 [1.02-1.55] | | 1.31 [1.05-1.64] | 221 | 221 | 221 | NA | 191 |
| 149.4 | 1.14 [1.02-1.28] | 1.17 [1.05-1.31] | 1.15 [1.03-1.29] | | 1.07 [0.95-1.21] | 1.16 [1.03-1.30] | 1.13 [1.01-1.27] | 1.12 [1.00-1.26] | | 1.11 [0.98-1.26] | 737 | 736 | 726 | NA | 624 |
| 149.9 | 1.21 [0.96-1.51] | 1.17 [0.94-1.47] | 1.28 [1.01-1.61] | | 1.23 [0.96-1.57] | 1.20 [0.95-1.51] | 1.13 [0.90-1.42] | 1.22 [0.96-1.54] | | 1.21 [0.94-1.55] | 183 | 180 | 177 | NA | 156 |
| 150 | 1.18 [1.11-1.24] | 1.15 [1.09-1.22] | 1.16 [1.10-1.23] | | 1.13 [1.06-1.20] | 1.17 [1.10-1.23] | 1.12 [1.06-1.18] | 1.13 [1.07-1.19] | | 1.13 [1.07-1.20] | 3,271 | 3,271 | 3,254 | NA | 2,734 |
| 151 | 1.08 [1.01-1.15] | 1.07 [1.00-1.13] | 1.09 [1.02-1.16] | | 1.06 [0.99-1.14] | 1.07 [1.01-1.14] | 1.04 [0.98-1.11] | 1.06 [0.99-1.13] | | 1.06 [0.99-1.14] | 2,515 | 2,515 | 2,483 | NA | 2,109 |
| 153.2 | 1.02 [0.98-1.05] | 1.01 [0.98-1.05] | 1.01 [0.98-1.05] | | 1.00 [0.96-1.03] | 1.00 [0.97-1.04] | 1.00 [0.96-1.03] | 1.00 [0.96-1.03] | | 0.99 [0.96-1.03] | 8,495 | 8,486 | 8,349 | NA | 7,040 |
| 153.3 | 0.97 [0.93-1.02] | 0.98 [0.94-1.02] | 0.98 [0.94-1.02] | | 0.96 [0.91-1.01] | 0.98 [0.94-1.02] | 0.98 [0.94-1.03] | 0.98 [0.94-1.03] | | 0.97 [0.92-1.02] | 4,787 | 4,786 | 4,707 | NA | 3,800 |
| 155 | 1.24 [1.16-1.33] | 1.25 [1.16-1.34] | 1.23 [1.14-1.32] | | 1.18 [1.09-1.27] | 1.21 [1.13-1.30] | 1.18 [1.10-1.27] | 1.16 [1.08-1.25] | | 1.17 [1.08-1.26] | 1,928 | 1,922 | 1,897 | NA | 1,711 |
| 155.1 | 1.31 [1.18-1.45] | 1.34 [1.21-1.49] | 1.32 [1.19-1.47] | | 1.26 [1.13-1.40] | 1.25 [1.13-1.39] | 1.24 [1.11-1.38] | 1.22 [1.10-1.36] | | 1.24 [1.12-1.39] | 915 | 909 | 900 | NA | 833 |
| 157 | 1.16 [1.10-1.23] | 1.14 [1.08-1.21] | 1.15 [1.09-1.22] | | 1.13 [1.07-1.20] | 1.15 [1.09-1.22] | 1.10 [1.04-1.17] | 1.11 [1.05-1.18] | | 1.13 [1.07-1.21] | 3,016 | 3,008 | 2,998 | NA | 2,573 |
| 158 | 1.10 [1.04-1.16] | 1.10 [1.04-1.17] | 1.11 [1.05-1.17] | | 1.09 [1.03-1.16] | 1.07 [1.01-1.13] | 1.06 [1.00-1.12] | 1.05 [1.00-1.12] | | 1.09 [1.02-1.15] | 3,061 | 3,036 | 2,947 | NA | 2,623 |
| 159 | 1.11 [1.08-1.14] | 1.11 [1.08-1.14] | 1.11 [1.08-1.14] | | 1.06 [1.03-1.09] | 1.09 [1.06-1.12] | 1.06 [1.03-1.09] | 1.07 [1.04-1.10] | | 1.05 [1.03-1.09] | 13,238 | 13,203 | 12,938 | NA | 12,018 |
| 159.2 | 1.11 [0.97-1.27] | 1.15 [1.00-1.31] | 1.12 [0.98-1.29] | | 1.07 [0.93-1.23] | 1.09 [0.95-1.25] | 1.08 [0.95-1.24] | 1.09 [0.95-1.25] | | 1.06 [0.92-1.22] | 530 | 529 | 521 | NA | 469 |
| 159.3 | 1.08 [0.96-1.21] | 1.07 [0.95-1.20] | 1.06 [0.94-1.19] | | 1.03 [0.91-1.17] | 1.07 [0.95-1.21] | 1.04 [0.92-1.17] | 1.03 [0.91-1.16] | | 1.03 [0.90-1.17] | 688 | 689 | 682 | NA | 578 |
| 159.4 | 1.04 [0.90-1.20] | 1.07 [0.93-1.23] | 1.06 [0.92-1.23] | | 1.04 [0.89-1.21] | 1.03 [0.89-1.19] | 1.06 [0.92-1.23] | 1.05 [0.91-1.21] | | 1.03 [0.88-1.21] | 451 | 444 | 439 | NA | 373 |
| 164 | 1.17 [0.94-1.46] | 1.12 [0.89-1.40] | 1.11 [0.87-1.40] | | 1.16 [0.91-1.47] | 1.16 [0.92-1.45] | 1.09 [0.87-1.38] | 1.09 [0.86-1.39] | | 1.16 [0.91-1.48] | 193 | 179 | 163 | NA | 161 |
| 165 | 1.18 [1.12-1.23] | 1.18 [1.13-1.24] | 1.17 [1.12-1.23] | | 1.10 [1.05-1.16] | 1.13 [1.08-1.19] | 1.09 [1.04-1.15] | 1.08 [1.03-1.13] | | 1.08 [1.03-1.14] | 4,438 | 4,415 | 4,368 | NA | 4,129 |
| 165.1 | 1.15 [1.12-1.18] | 1.15 [1.12-1.18] | 1.15 [1.12-1.18] | | 1.11 [1.08-1.14] | 1.14 [1.11-1.17] | 1.09 [1.06-1.12] | 1.09 [1.06-1.12] | | 1.12 [1.09-1.15] | 13,101 | 13,084 | 13,050 | NA | 11,204 |
| 170.1 | 1.20 [1.04-1.37] | 1.17 [1.01-1.36] | 1.15 [0.98-1.35] | 1.25 [0.92-1.70] | 1.15 [0.98-1.33] | 1.19 [1.03-1.37] | 1.14 [0.98-1.32] | 1.12 [0.95-1.32] | 1.26 [0.92-1.73] | 1.15 [0.99-1.34] | 507 | 424 | 364 | 109 | 416 |
| 170.2 | 1.09 [0.98-1.21] | 1.13 [1.01-1.26] | 1.13 [1.01-1.27] | | 1.10 [0.98-1.24] | 1.06 [0.96-1.18] | 1.10 [0.99-1.23] | 1.08 [0.97-1.22] | | 1.10 [0.98-1.23] | 871 | 796 | 711 | NA | 743 |
| 172.11 | 0.99 [0.94-1.04] | 1.00 [0.96-1.05] | 0.99 [0.95-1.04] | | 0.98 [0.94-1.04] | 0.97 [0.93-1.02] | 0.98 [0.94-1.03] | 0.97 [0.93-1.02] | | 0.97 [0.93-1.03] | 4,204 | 4,190 | 3,863 | NA | 3,528 |
| 172.2 | 1.10 [1.08-1.12] | 1.11 [1.08-1.13] | 1.10 [1.08-1.13] | | 1.06 [1.03-1.08] | 1.07 [1.05-1.09] | 1.07 [1.04-1.09] | 1.06 [1.04-1.08] | | 1.03 [1.01-1.06] | 25,094 | 25,077 | 24,709 | NA | 21,264 |
| 172.3 | 1.25 [1.18-1.33] | 1.26 [1.19-1.34] | 1.26 [1.19-1.34] | | 1.20 [1.13-1.28] | 1.19 [1.12-1.27] | 1.18 [1.11-1.25] | 1.17 [1.10-1.24] | | 1.16 [1.09-1.23] | 3,098 | 3,093 | 3,069 | NA | 2,827 |
| 173 | 1.04 [0.88-1.23] | 1.06 [0.89-1.26] | 1.04 [0.86-1.25] | | 0.96 [0.79-1.15] | 0.98 [0.83-1.17] | 1.01 [0.84-1.20] | 0.99 [0.82-1.19] | | 0.92 [0.76-1.12] | 331 | 303 | 276 | NA | 259 |
| 174 | 0.96 [0.74-1.25] | 0.96 [0.74-1.25] | 0.93 [0.72-1.21] | | 0.92 [0.70-1.22] | 0.95 [0.73-1.24] | 0.94 [0.72-1.22] | 0.93 [0.71-1.21] | | 0.92 [0.70-1.22] | 126 | 126 | 123 | NA | 108 |
| 174.1 | 1.01 [0.95-1.07] | 1.01 [0.95-1.07] | 1.00 [0.94-1.06] | | 0.96 [0.90-1.03] | 0.99 [0.93-1.06] | 1.00 [0.93-1.06] | 0.98 [0.92-1.05] | | 0.96 [0.89-1.03] | 2,263 | 2,265 | 2,184 | NA | 1,812 |
| 174.11 | 1.06 [1.04-1.09] | 1.06 [1.04-1.08] | 1.05 [1.03-1.08] | | 1.04 [1.01-1.07] | 1.06 [1.03-1.08] | 1.04 [1.02-1.07] | 1.03 [1.01-1.06] | | 1.05 [1.02-1.07] | 18,072 | 18,066 | 17,357 | NA | 14,324 |
| 174.3 | 0.99 [0.74-1.34] | 0.96 [0.71-1.29] | | | | 0.99 [0.73-1.33] | 0.93 [0.69-1.26] | | | | 104 | 104 | NA | NA | NA |
| 175 | 1.22 [1.21-1.24] | 1.22 [1.21-1.24] | 1.21 [1.20-1.23] | 1.21 [1.14-1.28] | 1.13 [1.12-1.15] | 1.15 [1.13-1.16] | 1.11 [1.09-1.12] | 1.10 [1.09-1.12] | 1.16 [1.09-1.23] | 1.10 [1.08-1.11] | 59,573 | 58,287 | 52,051 | 3,107 | 55,520 |
| 180.1 | 1.14 [1.12-1.18] | 1.14 [1.11-1.17] | 1.14 [1.11-1.17] | 1.06 [0.79-1.42] | 1.07 [1.04-1.10] | 1.11 [1.08-1.14] | 1.08 [1.06-1.11] | 1.08 [1.05-1.11] | 1.09 [0.81-1.48] | 1.07 [1.04-1.10] | 13,997 | 13,964 | 13,435 | 117 | 12,876 |
| 180.3 | 1.08 [1.04-1.13] | 1.10 [1.06-1.15] | 1.06 [0.97-1.14] | 1.04 [0.93-1.17] | 1.03 [0.98-1.08] | 1.05 [1.00-1.10] | 1.03 [0.99-1.08] | 1.01 [0.93-1.09] | 1.05 [0.93-1.19] | 1.03 [0.98-1.08] | 4,977 | 4,983 | 1,335 | 736 | 3,779 |
| 182 | 1.04 [0.98-1.11] | 1.04 [0.98-1.11] | 1.03 [0.97-1.10] | | 1.03 [0.96-1.10] | 1.06 [0.99-1.12] | 1.06 [1.00-1.13] | 1.05 [0.98-1.11] | | 1.04 [0.98-1.12] | 2,510 | 2,506 | 2,472 | NA | 1,945 |
| 184 | 1.06 [0.89-1.25] | 1.06 [0.89-1.25] | 1.00 [0.82-1.21] | | 1.09 [0.91-1.32] | 1.05 [0.89-1.25] | 1.05 [0.88-1.24] | 0.96 [0.79-1.16] | | 1.11 [0.92-1.35] | 330 | 313 | 243 | NA | 266 |
| 184.1 | 1.17 [0.98-1.41] | 1.20 [1.00-1.45] | 1.20 [1.00-1.45] | | 1.16 [0.95-1.41] | 1.19 [0.99-1.44] | 1.22 [1.01-1.47] | 1.21 [1.00-1.46] | | 1.17 [0.95-1.43] | 276 | 276 | 270 | NA | 228 |
| 184.11 | 1.14 [1.11-1.17] | 1.13 [1.10-1.16] | 1.13 [1.10-1.16] | 1.00 [0.75-1.33] | 1.06 [1.04-1.09] | 1.11 [1.08-1.14] | 1.08 [1.05-1.11] | 1.08 [1.05-1.11] | 1.01 [0.75-1.36] | 1.06 [1.03-1.09] | 14,907 | 14,868 | 14,399 | 123 | 13,494 |
| 184.2 | 1.14 [1.05-1.24] | 1.14 [1.05-1.24] | 1.14 [1.04-1.25] | | 1.15 [1.05-1.26] | 1.12 [1.03-1.22] | 1.10 [1.01-1.20] | 1.09 [0.99-1.20] | | 1.14 [1.04-1.26] | 1,277 | 1,254 | 1,078 | NA | 1,074 |
| 185 | 1.08 [1.06-1.10] | 1.08 [1.06-1.10] | 1.08 [1.05-1.10] | | 1.02 [1.00-1.05] | 1.06 [1.03-1.08] | 1.05 [1.02-1.07] | 1.04 [1.02-1.07] | | 1.02 [0.99-1.04] | 20,634 | 20,611 | 20,204 | NA | 17,639 |
| 187 | 1.25 [1.01-1.53] | 1.26 [1.03-1.55] | 1.18 [0.96-1.46] | | 1.18 [0.95-1.47] | 1.22 [0.99-1.51] | 1.20 [0.98-1.49] | 1.16 [0.93-1.44] | | 1.17 [0.94-1.47] | 217 | 216 | 203 | NA | 184 |
| 187.1 | 1.15 [1.12-1.19] | 1.15 [1.12-1.18] | 1.15 [1.12-1.18] | 1.10 [0.80-1.51] | 1.07 [1.04-1.10] | 1.12 [1.09-1.15] | 1.09 [1.06-1.12] | 1.09 [1.06-1.12] | 1.14 [0.82-1.58] | 1.07 [1.04-1.10] | 13,776 | 13,744 | 13,331 | 101 | 12,748 |
| 187.2 | 1.15 [1.12-1.18] | 1.15 [1.12-1.18] | 1.14 [1.11-1.18] | 1.07 [0.84-1.38] | 1.07 [1.04-1.10] | 1.12 [1.09-1.15] | 1.09 [1.06-1.12] | 1.09 [1.06-1.12] | 1.13 [0.87-1.46] | 1.07 [1.04-1.10] | 13,848 | 13,807 | 13,264 | 160 | 12,744 |
| 187.8 | 1.12 [0.88-1.43] | 1.13 [0.88-1.45] | 1.21 [0.92-1.59] | | 1.13 [0.86-1.47] | 1.11 [0.87-1.43] | 1.11 [0.86-1.43] | 1.17 [0.88-1.54] | | 1.11 [0.85-1.46] | 158 | 151 | 122 | NA | 130 |
| 189 | 1.14 [1.10-1.18] | 1.14 [1.10-1.18] | 1.14 [1.10-1.18] | | 1.08 [1.04-1.12] | 1.11 [1.07-1.15] | 1.09 [1.05-1.13] | 1.08 [1.05-1.12] | | 1.07 [1.03-1.11] | 8,077 | 8,029 | 7,939 | NA | 7,248 |
| 189.11 | 1.11 [1.05-1.18] | 1.10 [1.04-1.17] | 1.11 [1.05-1.18] | | 1.06 [0.99-1.12] | 1.09 [1.03-1.15] | 1.05 [0.99-1.11] | 1.06 [1.00-1.12] | | 1.05 [0.99-1.12] | 2,914 | 2,867 | 2,824 | NA | 2,501 |
| 189.12 | 1.22 [1.01-1.47] | 1.22 [1.01-1.47] | 1.22 [1.01-1.48] | | 1.11 [0.91-1.36] | 1.24 [1.02-1.50] | 1.18 [0.98-1.43] | 1.21 [1.00-1.47] | | 1.15 [0.94-1.41] | 282 | 280 | 277 | NA | 241 |
| 189.2 | 1.09 [1.02-1.16] | 1.08 [1.01-1.16] | 1.09 [1.02-1.17] | | 1.01 [0.95-1.09] | 1.07 [1.00-1.15] | 1.05 [0.98-1.12] | 1.07 [1.00-1.14] | | 1.02 [0.95-1.09] | 2,176 | 2,175 | 2,143 | NA | 1,863 |
| 189.21 | 1.08 [1.04-1.12] | 1.09 [1.05-1.13] | 1.08 [1.04-1.13] | | 1.03 [0.99-1.07] | 1.07 [1.03-1.12] | 1.06 [1.02-1.10] | 1.06 [1.02-1.10] | | 1.03 [0.99-1.08] | 6,425 | 6,415 | 6,366 | NA | 5,376 |
| 189.4 | 1.08 [0.94-1.25] | 1.08 [0.94-1.24] | 1.09 [0.95-1.26] | | 0.98 [0.84-1.13] | 1.08 [0.93-1.24] | 1.06 [0.92-1.22] | 1.06 [0.92-1.22] | | 0.99 [0.85-1.15] | 473 | 474 | 471 | NA | 416 |
| 190 | 1.02 [0.85-1.22] | 0.99 [0.82-1.20] | 1.08 [0.89-1.31] | | 1.00 [0.83-1.22] | 0.99 [0.82-1.19] | 0.97 [0.80-1.18] | 1.04 [0.85-1.27] | | 1.00 [0.82-1.22] | 280 | 257 | 241 | NA | 239 |
| 191 | 1.13 [1.01-1.25] | 1.10 [0.98-1.23] | 1.05 [0.93-1.18] | 1.24 [0.96-1.59] | 1.08 [0.96-1.20] | 1.10 [0.99-1.22] | 1.06 [0.94-1.18] | 0.99 [0.88-1.12] | 1.25 [0.97-1.61] | 1.07 [0.95-1.20] | 881 | 747 | 651 | 166 | 748 |
| 191.1 | 1.12 [0.89-1.41] | 1.09 [0.84-1.42] | 1.12 [0.84-1.49] | | 1.06 [0.83-1.35] | 1.08 [0.86-1.36] | 1.05 [0.80-1.37] | 1.12 [0.84-1.49] | | 1.07 [0.84-1.36] | 186 | 135 | 112 | NA | 161 |
| 191.11 | 1.12 [1.04-1.21] | 1.15 [1.06-1.24] | 1.13 [1.04-1.23] | 0.99 [0.81-1.21] | 1.09 [1.00-1.19] | 1.11 [1.03-1.19] | 1.12 [1.03-1.21] | 1.10 [1.01-1.20] | 1.02 [0.83-1.24] | 1.09 [1.00-1.18] | 1,672 | 1,469 | 1,338 | 243 | 1,326 |
| 193 | 1.06 [0.96-1.18] | 1.06 [0.95-1.18] | 1.06 [0.94-1.20] | | 1.02 [0.91-1.15] | 1.04 [0.93-1.16] | 1.02 [0.92-1.14] | 1.03 [0.92-1.17] | | 1.01 [0.90-1.14] | 819 | 798 | 618 | NA | 675 |
| 194 | 1.15 [0.95-1.38] | 1.06 [0.87-1.30] | 1.14 [0.93-1.41] | | 1.05 [0.87-1.27] | 1.13 [0.93-1.35] | 1.01 [0.82-1.25] | 1.08 [0.87-1.34] | | 1.05 [0.87-1.28] | 281 | 228 | 209 | NA | 249 |
| 195 | 1.11 [1.03-1.19] | 1.12 [1.04-1.20] | 1.12 [1.04-1.20] | | 1.07 [0.99-1.16] | 1.08 [1.00-1.16] | 1.07 [0.99-1.15] | 1.07 [0.99-1.15] | | 1.06 [0.98-1.14] | 1,820 | 1,800 | 1,741 | NA | 1,597 |
| 195.1 | 1.16 [1.15-1.18] | 1.16 [1.15-1.18] | 1.16 [1.15-1.18] | 1.09 [1.01-1.18] | 1.10 [1.08-1.11] | 1.13 [1.11-1.14] | 1.10 [1.09-1.11] | 1.10 [1.09-1.11] | 1.09 [1.01-1.18] | 1.09 [1.08-1.10] | 80,474 | 79,523 | 75,993 | 1,693 | 72,445 |
| 195.3 | 1.28 [1.01-1.62] | 1.28 [1.00-1.64] | 1.21 [0.95-1.55] | | 1.30 [1.00-1.68] | 1.29 [1.02-1.64] | 1.26 [0.98-1.62] | 1.19 [0.92-1.53] | | 1.34 [1.03-1.73] | 172 | 161 | 158 | NA | 150 |
| 197 | 1.27 [1.25-1.29] | 1.28 [1.26-1.30] | 1.27 [1.24-1.29] | 1.16 [1.10-1.22] | 1.19 [1.17-1.21] | 1.20 [1.18-1.22] | 1.17 [1.15-1.19] | 1.16 [1.14-1.18] | 1.12 [1.06-1.18] | 1.15 [1.13-1.17] | 40,144 | 37,469 | 31,884 | 4,078 | 34,553 |
| 198 | 1.07 [1.04-1.09] | 1.06 [1.04-1.08] | 1.06 [1.04-1.09] | 1.01 [0.78-1.30] | 1.05 [1.03-1.07] | 1.07 [1.05-1.09] | 1.04 [1.02-1.06] | 1.05 [1.02-1.07] | 1.01 [0.78-1.31] | 1.07 [1.05-1.09] | 22,768 | 22,653 | 22,271 | 164 | 19,792 |
| 198.1 | 1.06 [1.03-1.09] | 1.05 [1.03-1.08] | 1.05 [1.03-1.08] | 1.14 [0.86-1.51] | 1.04 [1.01-1.07] | 1.06 [1.04-1.09] | 1.04 [1.01-1.07] | 1.04 [1.01-1.07] | 1.14 [0.86-1.53] | 1.05 [1.02-1.08] | 14,180 | 14,120 | 13,563 | 140 | 11,924 |
| 198.2 | 1.06 [1.03-1.09] | 1.05 [1.02-1.08] | 1.05 [1.02-1.09] | 1.03 [0.76-1.40] | 1.05 [1.01-1.08] | 1.07 [1.04-1.10] | 1.03 [1.00-1.07] | 1.04 [1.01-1.07] | 1.05 [0.77-1.44] | 1.07 [1.04-1.11] | 10,415 | 10,342 | 10,137 | 109 | 9,342 |
| 198.3 | 1.05 [1.01-1.10] | 1.04 [1.00-1.09] | 1.05 [1.01-1.10] | | 1.04 [1.00-1.09] | 1.06 [1.02-1.10] | 1.04 [0.99-1.08] | 1.04 [1.00-1.09] | | 1.06 [1.01-1.10] | 5,766 | 5,739 | 5,623 | NA | 4,965 |
| 198.4 | 1.05 [1.03-1.08] | 1.04 [1.02-1.07] | 1.06 [1.03-1.09] | | 1.05 [1.01-1.08] | 1.06 [1.03-1.09] | 1.03 [1.00-1.06] | 1.04 [1.01-1.07] | | 1.06 [1.03-1.10] | 12,201 | 12,178 | 11,983 | NA | 10,663 |
| 198.5 | 1.12 [1.06-1.18] | 1.11 [1.06-1.17] | 1.13 [1.08-1.19] | | 1.10 [1.04-1.16] | 1.13 [1.07-1.19] | 1.08 [1.03-1.14] | 1.11 [1.05-1.17] | | 1.13 [1.07-1.19] | 3,672 | 3,626 | 3,549 | NA | 3,245 |
| 198.6 | 1.07 [1.04-1.10] | 1.07 [1.04-1.10] | 1.08 [1.05-1.11] | 1.06 [0.77-1.45] | 1.05 [1.02-1.08] | 1.07 [1.04-1.11] | 1.05 [1.02-1.08] | 1.05 [1.02-1.09] | 1.10 [0.79-1.51] | 1.08 [1.04-1.11] | 11,056 | 10,974 | 10,800 | 107 | 9,843 |
| 198.7 | 1.01 [0.90-1.13] | 1.00 [0.89-1.13] | 0.99 [0.88-1.12] | | 0.96 [0.86-1.09] | 1.02 [0.91-1.15] | 0.99 [0.88-1.11] | 0.98 [0.87-1.11] | | 0.99 [0.88-1.12] | 694 | 688 | 665 | NA | 638 |
| 199 | 1.10 [1.06-1.15] | 1.11 [1.06-1.15] | 1.12 [1.07-1.16] | 0.96 [0.80-1.14] | 1.05 [1.00-1.09] | 1.08 [1.04-1.13] | 1.06 [1.02-1.11] | 1.07 [1.03-1.12] | 0.94 [0.78-1.13] | 1.05 [1.00-1.09] | 6,264 | 6,052 | 5,681 | 319 | 5,352 |
| 199.4 | 0.94 [0.78-1.12] | 0.91 [0.72-1.15] | 0.89 [0.67-1.19] | 0.89 [0.69-1.15] | 0.87 [0.72-1.06] | 0.90 [0.75-1.08] | 0.83 [0.65-1.05] | 0.83 [0.62-1.11] | 0.89 [0.69-1.16] | 0.85 [0.70-1.03] | 291 | 163 | 106 | 148 | 255 |
| 200 | 1.27 [1.21-1.34] | 1.29 [1.22-1.35] | 1.26 [1.20-1.33] | 1.30 [0.99-1.71] | 1.20 [1.14-1.27] | 1.23 [1.16-1.29] | 1.21 [1.15-1.28] | 1.20 [1.13-1.26] | 1.27 [0.96-1.68] | 1.18 [1.12-1.25] | 3,886 | 3,804 | 3,634 | 147 | 3,493 |
| 200.1 | 1.10 [0.99-1.23] | 1.13 [1.01-1.25] | 1.06 [0.96-1.19] | | 1.06 [0.95-1.19] | 1.09 [0.98-1.21] | 1.06 [0.95-1.18] | 1.00 [0.89-1.11] | | 1.04 [0.93-1.17] | 839 | 837 | 776 | NA | 704 |
| 201 | 1.71 [1.54-1.90] | 1.72 [1.54-1.91] | 1.62 [1.42-1.85] | 1.41 [1.11-1.79] | 1.46 [1.30-1.65] | 1.64 [1.48-1.83] | 1.65 [1.48-1.85] | 1.52 [1.33-1.74] | 1.33 [1.04-1.71] | 1.43 [1.26-1.61] | 969 | 870 | 588 | 185 | 701 |
| 202 | 1.33 [1.26-1.40] | 1.32 [1.26-1.40] | 1.31 [1.24-1.38] | 1.19 [0.94-1.49] | 1.22 [1.16-1.28] | 1.26 [1.20-1.33] | 1.22 [1.16-1.28] | 1.19 [1.12-1.25] | 1.19 [0.94-1.51] | 1.18 [1.12-1.24] | 3,821 | 3,718 | 3,407 | 197 | 3,596 |
| 202.2 | 1.32 [1.27-1.38] | 1.33 [1.27-1.39] | 1.30 [1.24-1.36] | 1.29 [1.01-1.66] | 1.25 [1.19-1.31] | 1.29 [1.23-1.35] | 1.29 [1.23-1.35] | 1.25 [1.19-1.31] | 1.30 [1.00-1.67] | 1.23 [1.17-1.30] | 4,824 | 4,724 | 4,442 | 168 | 3,988 |
| 202.21 | 1.16 [1.04-1.29] | 1.10 [0.99-1.23] | 1.10 [0.98-1.23] | | 1.04 [0.93-1.17] | 1.10 [0.99-1.23] | 1.04 [0.93-1.16] | 1.05 [0.94-1.17] | | 1.02 [0.90-1.15] | 790 | 782 | 744 | NA | 658 |
| 202.24 | 1.31 [1.22-1.42] | 1.32 [1.22-1.43] | 1.29 [1.19-1.39] | | 1.24 [1.14-1.35] | 1.27 [1.18-1.37] | 1.28 [1.19-1.39] | 1.24 [1.15-1.34] | | 1.22 [1.12-1.33] | 1,690 | 1,672 | 1,575 | NA | 1,437 |
| 204 | 1.19 [1.10-1.28] | 1.30 [1.20-1.41] | 1.29 [1.19-1.41] | 0.90 [0.74-1.10] | 1.11 [1.03-1.20] | 1.15 [1.07-1.25] | 1.23 [1.13-1.34] | 1.20 [1.10-1.31] | 0.93 [0.76-1.14] | 1.09 [1.01-1.18] | 1,708 | 1,506 | 1,380 | 250 | 1,559 |
| 204.1 | 1.07 [1.00-1.15] | 1.12 [1.04-1.21] | 1.09 [1.02-1.18] | 0.97 [0.81-1.15] | 1.04 [0.97-1.12] | 1.05 [0.99-1.13] | 1.08 [1.01-1.17] | 1.05 [0.98-1.13] | 0.97 [0.81-1.15] | 1.03 [0.95-1.11] | 2,123 | 1,828 | 1,763 | 321 | 1,756 |
| 204.11 | 1.00 [0.87-1.16] | 1.23 [0.98-1.54] | 1.22 [0.95-1.58] | 0.94 [0.79-1.12] | 0.97 [0.82-1.13] | 0.99 [0.86-1.14] | 1.18 [0.94-1.49] | 1.15 [0.89-1.50] | 0.94 [0.79-1.13] | 0.96 [0.82-1.13] | 470 | 184 | 142 | 307 | 369 |
| 204.12 | 1.04 [0.96-1.13] | 1.07 [0.99-1.16] | 1.04 [0.96-1.12] | | 1.01 [0.93-1.10] | 1.02 [0.94-1.11] | 1.03 [0.95-1.12] | 1.00 [0.92-1.08] | | 1.00 [0.91-1.09] | 1,512 | 1,480 | 1,464 | NA | 1,269 |
| 204.2 | 1.28 [1.18-1.38] | 1.29 [1.20-1.39] | 1.28 [1.19-1.39] | 1.09 [0.81-1.49] | 1.24 [1.15-1.35] | 1.25 [1.16-1.35] | 1.26 [1.16-1.36] | 1.25 [1.15-1.35] | 1.10 [0.80-1.51] | 1.24 [1.14-1.35] | 1,752 | 1,680 | 1,574 | 107 | 1,488 |
| 204.21 | 1.32 [1.21-1.44] | 1.33 [1.22-1.45] | 1.33 [1.22-1.46] | | 1.29 [1.17-1.41] | 1.30 [1.19-1.42] | 1.30 [1.19-1.42] | 1.30 [1.19-1.43] | | 1.29 [1.17-1.41] | 1,349 | 1,282 | 1,203 | NA | 1,151 |
| 204.22 | 1.17 [1.00-1.37] | 1.21 [1.03-1.41] | 1.16 [0.98-1.36] | | 1.16 [0.98-1.37] | 1.14 [0.97-1.33] | 1.16 [0.99-1.36] | 1.11 [0.94-1.31] | | 1.16 [0.98-1.38] | 413 | 406 | 374 | NA | 353 |
| 204.3 | 1.17 [0.94-1.46] | 1.18 [0.95-1.47] | 1.10 [0.88-1.37] | | 1.11 [0.89-1.40] | 1.19 [0.95-1.48] | 1.14 [0.91-1.42] | 1.07 [0.85-1.34] | | 1.15 [0.91-1.44] | 216 | 213 | 211 | NA | 199 |
| 204.4 | 1.16 [1.09-1.24] | 1.17 [1.10-1.25] | 1.17 [1.09-1.24] | | 1.11 [1.04-1.19] | 1.13 [1.06-1.21] | 1.14 [1.07-1.22] | 1.13 [1.06-1.21] | | 1.09 [1.02-1.17] | 2,358 | 2,351 | 2,322 | NA | 2,008 |
| 208 | 1.23 [1.22-1.25] | 1.23 [1.22-1.25] | 1.22 [1.20-1.24] | 1.38 [1.20-1.60] | 1.15 [1.13-1.17] | 1.16 [1.14-1.17] | 1.12 [1.10-1.14] | 1.11 [1.09-1.13] | 1.30 [1.12-1.51] | 1.11 [1.09-1.12] | 43,766 | 43,635 | 41,594 | 536 | 37,979 |

### Supplementary Table 6: phecode-mapped results (hazard ratios and events from all cohorts)

| | Hazard ratio (99% confidence interval) | | | | | | | | | | Events (in exposed) | | | | |
|---|---|---|---|---|---|---|---|---|---|---|---|---|---|---|---|
| | crude | | | | | adjusted | | | | | | | | | |
| Outcome | any age | 18+ | 40+ | <18 | hosp. | any age | 18+ | 40+ | <18 | hosp. | any age | 18+ | 40+ | <18 | hosp. |
| 210 | 1.15 [1.07-1.24] | 1.17 [1.08-1.26] | 1.16 [1.07-1.26] | 1.09 [0.88-1.35] | 1.13 [1.04-1.23] | 1.12 [1.04-1.21] | 1.08 [1.00-1.17] | 1.08 [0.99-1.17] | 1.09 [0.88-1.36] | 1.11 [1.02-1.21] | 1,714 | 1,585 | 1,262 | 221 | 1,388 |
| 211 | 1.28 [1.24-1.31] | 1.28 [1.24-1.31] | 1.27 [1.24-1.31] | 1.06 [0.81-1.39] | 1.18 [1.15-1.21] | 1.15 [1.11-1.18] | 1.12 [1.09-1.15] | 1.11 [1.08-1.14] | 1.00 [0.76-1.33] | 1.09 [1.06-1.12] | 13,365 | 13,319 | 12,827 | 139 | 12,195 |
| 212 | 1.25 [1.13-1.40] | 1.25 [1.12-1.40] | 1.26 [1.11-1.42] | 1.31 [0.97-1.79] | 1.13 [1.00-1.27] | 1.20 [1.08-1.34] | 1.15 [1.02-1.29] | 1.18 [1.04-1.34] | 1.25 [0.91-1.72] | 1.10 [0.98-1.24] | 835 | 772 | 632 | 108 | 677 |
| 213 | 1.18 [1.07-1.29] | 1.27 [1.13-1.43] | 1.20 [1.03-1.40] | 1.07 [0.94-1.22] | 1.08 [0.98-1.21] | 1.14 [1.03-1.25] | 1.18 [1.04-1.33] | 1.09 [0.93-1.28] | 1.06 [0.92-1.21] | 1.05 [0.94-1.17] | 1,123 | 647 | 390 | 604 | 878 |
| 214 | 1.16 [1.12-1.20] | 1.16 [1.12-1.20] | 1.13 [1.09-1.18] | 1.13 [0.98-1.30] | 1.07 [1.03-1.11] | 1.10 [1.07-1.14] | 1.08 [1.05-1.12] | 1.06 [1.02-1.10] | 1.07 [0.93-1.23] | 1.03 [0.99-1.07] | 7,797 | 7,547 | 6,147 | 516 | 6,248 |
| 214.1 | 1.16 [1.12-1.22] | 1.16 [1.11-1.21] | 1.13 [1.07-1.18] | 1.20 [1.02-1.41] | 1.06 [1.01-1.11] | 1.11 [1.06-1.16] | 1.09 [1.04-1.14] | 1.06 [1.01-1.11] | 1.13 [0.95-1.33] | 1.03 [0.98-1.08] | 5,078 | 4,904 | 3,769 | 379 | 3,941 |
| 215 | 1.23 [1.14-1.32] | 1.25 [1.16-1.34] | 1.22 [1.13-1.33] | 1.07 [0.87-1.32] | 1.14 [1.05-1.23] | 1.16 [1.07-1.25] | 1.15 [1.07-1.24] | 1.12 [1.03-1.22] | 1.04 [0.84-1.29] | 1.09 [1.01-1.18] | 1,812 | 1,661 | 1,316 | 235 | 1,475 |
| 216 | 1.18 [1.15-1.21] | 1.20 [1.16-1.23] | 1.18 [1.14-1.23] | 1.14 [1.09-1.20] | 1.10 [1.07-1.13] | 1.13 [1.10-1.16] | 1.12 [1.09-1.16] | 1.11 [1.07-1.15] | 1.11 [1.05-1.16] | 1.07 [1.03-1.10] | 13,503 | 10,708 | 6,986 | 3,861 | 10,387 |
| 217 | 1.19 [1.00-1.41] | | | 1.16 [0.96-1.40] | 1.16 [0.96-1.41] | 1.17 [0.98-1.39] | | | 1.15 [0.95-1.39] | 1.14 [0.94-1.39] | 338 | NA | NA | 287 | 271 |
| 217.1 | 1.34 [1.24-1.45] | 1.36 [1.25-1.47] | 1.33 [1.23-1.45] | | 1.28 [1.18-1.39] | 1.27 [1.17-1.38] | 1.24 [1.14-1.35] | 1.23 [1.13-1.34] | | 1.25 [1.15-1.36] | 1,597 | 1,510 | 1,419 | NA | 1,467 |
| 218.1 | 1.07 [1.04-1.10] | 1.08 [1.05-1.11] | 1.09 [1.06-1.12] | 1.14 [0.89-1.44] | 1.06 [1.03-1.09] | 1.04 [1.01-1.07] | 1.05 [1.02-1.08] | 1.05 [1.02-1.08] | 1.12 [0.87-1.44] | 1.03 [1.00-1.06] | 12,552 | 12,549 | 10,372 | 190 | 9,894 |
| 218.2 | 1.12 [0.96-1.29] | 1.10 [0.95-1.27] | 1.08 [0.92-1.27] | | 1.09 [0.93-1.29] | 1.09 [0.94-1.27] | 1.08 [0.93-1.25] | 1.04 [0.88-1.23] | | 1.07 [0.90-1.26] | 424 | 420 | 330 | NA | 342 |
| 220 | 1.04 [0.98-1.10] | 1.06 [1.00-1.12] | 1.04 [0.97-1.11] | 1.07 [0.90-1.28] | 1.03 [0.96-1.09] | 1.02 [0.96-1.08] | 1.01 [0.96-1.07] | 0.99 [0.93-1.07] | 1.04 [0.87-1.25] | 1.01 [0.95-1.07] | 2,900 | 2,818 | 1,877 | 349 | 2,332 |
| 221 | 1.25 [1.06-1.47] | 1.26 [1.06-1.49] | 1.14 [0.93-1.40] | | 1.20 [1.00-1.44] | 1.19 [1.00-1.41] | 1.19 [1.00-1.41] | 1.11 [0.90-1.37] | | 1.17 [0.97-1.40] | 347 | 332 | 218 | NA | 287 |
| 222 | 1.23 [1.02-1.49] | 1.21 [0.99-1.49] | 1.14 [0.92-1.42] | | 1.17 [0.95-1.44] | 1.19 [0.98-1.45] | 1.16 [0.95-1.43] | 1.10 [0.89-1.38] | | 1.14 [0.92-1.40] | 263 | 232 | 208 | NA | 225 |
| 223 | 1.11 [0.96-1.28] | 1.08 [0.94-1.25] | 1.08 [0.94-1.26] | | 1.03 [0.88-1.20] | 1.08 [0.93-1.25] | 1.02 [0.88-1.18] | 1.03 [0.89-1.20] | | 1.01 [0.87-1.17] | 450 | 443 | 422 | NA | 400 |
| 224 | 1.38 [1.15-1.64] | 1.39 [1.12-1.72] | 1.34 [1.04-1.72] | 1.31 [0.99-1.73] | 1.41 [1.16-1.71] | 1.34 [1.12-1.61] | 1.28 [1.03-1.60] | 1.26 [0.98-1.63] | 1.27 [0.95-1.70] | 1.39 [1.14-1.69] | 319 | 214 | 154 | 132 | 270 |
| 224.1 | 1.17 [0.97-1.43] | 1.21 [0.99-1.48] | 1.21 [0.99-1.48] | | 1.16 [0.94-1.43] | 1.16 [0.95-1.41] | 1.18 [0.96-1.44] | 1.15 [0.94-1.42] | | 1.14 [0.92-1.40] | 258 | 249 | 244 | NA | 232 |
| 225.1 | 1.13 [1.06-1.21] | 1.09 [1.02-1.17] | 1.11 [1.04-1.19] | 1.39 [1.02-1.89] | 1.08 [1.00-1.15] | 1.08 [1.01-1.15] | 1.02 [0.95-1.09] | 1.04 [0.97-1.12] | 1.33 [0.97-1.83] | 1.05 [0.98-1.13] | 2,253 | 2,182 | 2,041 | 112 | 2,001 |
| 225.2 | 1.12 [0.85-1.49] | 1.13 [0.84-1.50] | | | | 1.05 [0.79-1.40] | 1.06 [0.78-1.42] | | | | 121 | 112 | NA | NA | NA |
| 226 | 1.07 [0.93-1.24] | 1.04 [0.90-1.21] | 1.04 [0.89-1.23] | | 1.06 [0.91-1.24] | 1.03 [0.88-1.19] | 0.98 [0.84-1.14] | 1.00 [0.85-1.19] | | 1.01 [0.86-1.19] | 432 | 419 | 333 | NA | 381 |
| 227.1 | 1.11 [0.99-1.23] | 1.11 [1.00-1.23] | 1.11 [1.00-1.24] | | 1.04 [0.93-1.16] | 1.05 [0.94-1.17] | 0.99 [0.89-1.10] | 0.99 [0.88-1.10] | | 1.02 [0.91-1.14] | 848 | 845 | 808 | NA | 769 |
| 227.2 | 1.08 [0.97-1.21] | 1.07 [0.96-1.19] | 1.04 [0.93-1.16] | | 1.00 [0.89-1.12] | 1.05 [0.94-1.17] | 1.02 [0.91-1.14] | 1.00 [0.90-1.13] | | 0.98 [0.87-1.10] | 776 | 774 | 722 | NA | 665 |
| 227.3 | 1.21 [1.09-1.35] | 1.22 [1.10-1.35] | 1.24 [1.10-1.40] | | 1.05 [0.94-1.18] | 1.16 [1.04-1.29] | 1.15 [1.04-1.28] | 1.17 [1.03-1.32] | | 1.03 [0.91-1.15] | 891 | 868 | 670 | NA | 734 |
| 228 | 1.17 [1.11-1.23] | 1.16 [1.10-1.23] | 1.19 [1.12-1.26] | 1.14 [1.02-1.28] | 1.11 [1.05-1.17] | 1.11 [1.05-1.17] | 1.07 [1.01-1.13] | 1.09 [1.02-1.16] | 1.13 [1.01-1.27] | 1.07 [1.01-1.13] | 3,729 | 3,083 | 2,567 | 790 | 3,237 |
| 229 | 1.21 [1.15-1.27] | 1.20 [1.14-1.27] | 1.18 [1.11-1.26] | 1.11 [0.99-1.24] | 1.11 [1.05-1.17] | 1.15 [1.09-1.20] | 1.12 [1.06-1.19] | 1.10 [1.03-1.17] | 1.07 [0.95-1.20] | 1.07 [1.01-1.14] | 3,836 | 3,264 | 2,316 | 811 | 3,041 |
| 229.1 | 1.66 [1.22-2.26] | | | | | 1.59 [1.16-2.17] | | | | | 103 | NA | NA | NA | NA |
| 860 | 1.24 [1.13-1.37] | 1.30 [1.17-1.44] | 1.27 [1.13-1.42] | 1.07 [0.86-1.33] | 1.15 [1.04-1.27] | 1.21 [1.10-1.34] | 1.25 [1.12-1.39] | 1.22 [1.09-1.37] | 1.08 [0.86-1.35] | 1.16 [1.05-1.28] | 1,036 | 882 | 709 | 215 | 945 |
| endocrine/metabolic | | | | | | | | | | | | | | | |
| 240 | 1.14 [1.06-1.23] | 1.12 [1.03-1.21] | 1.07 [0.99-1.16] | | 1.11 [1.02-1.20] | 1.08 [1.00-1.17] | 1.02 [0.95-1.11] | 0.99 [0.91-1.08] | | 1.07 [0.99-1.16] | 1,654 | 1,603 | 1,419 | NA | 1,455 |
| 241.1 | 1.15 [1.06-1.24] | 1.13 [1.05-1.22] | 1.14 [1.05-1.24] | | 1.07 [0.99-1.16] | 1.10 [1.02-1.19] | 1.06 [0.98-1.15] | 1.07 [0.98-1.16] | | 1.04 [0.96-1.14] | 1,583 | 1,550 | 1,318 | NA | 1,388 |
| 241.2 | 1.15 [1.07-1.24] | 1.15 [1.07-1.24] | 1.15 [1.06-1.24] | | 1.10 [1.02-1.19] | 1.09 [1.01-1.17] | 1.07 [1.00-1.16] | 1.07 [0.99-1.15] | | 1.07 [0.99-1.15] | 1,836 | 1,827 | 1,669 | NA | 1,617 |
| 242 | 1.29 [1.25-1.34] | 1.29 [1.24-1.33] | 1.27 [1.22-1.32] | 1.28 [1.09-1.50] | 1.21 [1.17-1.25] | 1.23 [1.18-1.27] | 1.18 [1.14-1.22] | 1.17 [1.12-1.21] | 1.24 [1.05-1.46] | 1.16 [1.12-1.21] | 8,314 | 8,168 | 6,931 | 419 | 7,289 |
| 242.1 | 1.37 [1.27-1.47] | 1.36 [1.26-1.46] | 1.36 [1.25-1.48] | 1.36 [1.07-1.74] | 1.28 [1.18-1.38] | 1.30 [1.21-1.40] | 1.24 [1.15-1.34] | 1.25 [1.14-1.36] | 1.38 [1.07-1.78] | 1.23 [1.14-1.33] | 1,916 | 1,856 | 1,320 | 182 | 1,654 |
| 242.2 | 1.11 [0.95-1.29] | 1.10 [0.94-1.28] | 1.07 [0.92-1.25] | | 1.02 [0.87-1.19] | 1.07 [0.92-1.25] | 1.03 [0.88-1.21] | 1.00 [0.85-1.18] | | 0.98 [0.84-1.16] | 424 | 424 | 389 | NA | 369 |
| 242.3 | 1.47 [1.22-1.78] | 1.42 [1.18-1.72] | 1.46 [1.19-1.80] | | 1.35 [1.11-1.65] | 1.43 [1.18-1.73] | 1.25 [1.03-1.53] | 1.35 [1.09-1.67] | | 1.32 [1.07-1.62] | 277 | 274 | 229 | NA | 237 |
| 244.1 | 1.24 [1.18-1.30] | 1.25 [1.19-1.32] | 1.24 [1.18-1.31] | 1.13 [0.82-1.56] | 1.16 [1.10-1.22] | 1.16 [1.11-1.22] | 1.14 [1.08-1.20] | 1.12 [1.06-1.18] | 1.10 [0.79-1.55] | 1.11 [1.06-1.17] | 4,029 | 3,999 | 3,602 | 103 | 3,734 |
| 244.2 | 1.29 [1.16-1.44] | 1.31 [1.17-1.47] | 1.27 [1.13-1.43] | 1.54 [1.15-2.07] | 1.22 [1.09-1.36] | 1.23 [1.10-1.37] | 1.23 [1.09-1.37] | 1.19 [1.05-1.34] | 1.44 [1.06-1.95] | 1.18 [1.05-1.32] | 923 | 831 | 723 | 132 | 841 |
| 244.4 | 1.34 [1.32-1.36] | 1.34 [1.32-1.36] | 1.33 [1.31-1.35] | 1.20 [1.11-1.31] | 1.26 [1.24-1.28] | 1.28 [1.26-1.30] | 1.25 [1.23-1.26] | 1.24 [1.22-1.26] | 1.18 [1.08-1.28] | 1.22 [1.20-1.24] | 50,313 | 49,752 | 44,667 | 1,561 | 41,359 |
| 244.5 | 1.17 [0.98-1.41] | 1.42 [1.14-1.77] | 1.38 [1.08-1.75] | | 1.11 [0.91-1.35] | 1.10 [0.91-1.33] | 1.28 [1.02-1.60] | 1.23 [0.96-1.57] | | 1.04 [0.85-1.27] | 286 | 221 | 178 | NA | 253 |
| 245 | 1.22 [1.10-1.35] | 1.22 [1.10-1.36] | 1.24 [1.10-1.40] | | 1.20 [1.07-1.34] | 1.17 [1.05-1.30] | 1.13 [1.01-1.26] | 1.16 [1.02-1.31] | | 1.14 [1.02-1.28] | 883 | 841 | 636 | NA | 787 |
| 245.21 | 1.25 [1.11-1.41] | 1.26 [1.11-1.43] | 1.27 [1.10-1.46] | | 1.24 [1.09-1.41] | 1.21 [1.07-1.37] | 1.17 [1.03-1.33] | 1.19 [1.03-1.38] | | 1.18 [1.04-1.35] | 654 | 621 | 460 | NA | 579 |
| 246 | 1.26 [1.16-1.37] | 1.30 [1.19-1.41] | 1.28 [1.17-1.40] | | 1.18 [1.08-1.29] | 1.17 [1.07-1.27] | 1.18 [1.08-1.29] | 1.16 [1.06-1.27] | | 1.11 [1.02-1.22] | 1,422 | 1,394 | 1,233 | NA | 1,293 |
| 246.7 | 1.14 [0.96-1.35] | 1.16 [0.97-1.40] | 1.22 [1.00-1.49] | | 1.10 [0.92-1.31] | 1.07 [0.90-1.28] | 1.07 [0.89-1.29] | 1.11 [0.91-1.36] | | 1.06 [0.89-1.27] | 360 | 310 | 269 | NA | 329 |
| 250 | 1.36 [1.33-1.40] | 1.37 [1.34-1.41] | 1.36 [1.33-1.40] | 0.90 [0.74-1.11] | 1.27 [1.23-1.30] | 1.26 [1.23-1.30] | 1.18 [1.15-1.22] | 1.17 [1.13-1.20] | 0.91 [0.74-1.12] | 1.22 [1.18-1.25] | 14,114 | 14,014 | 13,328 | 236 | 12,768 |
| 250.1 | 1.23 [1.19-1.26] | 1.30 [1.25-1.34] | 1.32 [1.27-1.36] | 0.97 [0.91-1.04] | 1.14 [1.11-1.18] | 1.15 [1.12-1.19] | 1.12 [1.09-1.16] | 1.13 [1.09-1.17] | 0.96 [0.90-1.03] | 1.11 [1.07-1.15] | 10,587 | 8,646 | 7,458 | 2,324 | 8,645 |
| 250.11 | 1.09 [1.02-1.17] | 1.21 [1.11-1.31] | 1.26 [1.12-1.41] | 0.91 [0.83-1.00] | 0.99 [0.92-1.06] | 1.05 [0.98-1.12] | 1.08 [0.98-1.17] | 1.06 [0.95-1.20] | 0.94 [0.85-1.03] | 0.99 [0.91-1.06] | 2,084 | 1,291 | 718 | 1,077 | 1,809 |
| 250.12 | 1.51 [1.32-1.73] | 1.61 [1.40-1.85] | 1.71 [1.47-1.98] | | 1.32 [1.15-1.51] | 1.39 [1.21-1.60] | 1.34 [1.16-1.54] | 1.40 [1.19-1.63] | | 1.29 [1.12-1.49] | 527 | 525 | 450 | NA | 501 |
| 250.13 | 1.30 [1.20-1.40] | 1.33 [1.24-1.43] | 1.32 [1.22-1.43] | | 1.17 [1.09-1.27] | 1.21 [1.13-1.31] | 1.13 [1.05-1.22] | 1.11 [1.02-1.21] | | 1.16 [1.08-1.26] | 1,764 | 1,755 | 1,438 | NA | 1,616 |
| 250.14 | 1.52 [1.36-1.71] | 1.52 [1.35-1.70] | 1.67 [1.47-1.88] | | 1.30 [1.16-1.45] | 1.38 [1.23-1.55] | 1.22 [1.08-1.37] | 1.30 [1.14-1.48] | | 1.25 [1.11-1.40] | 762 | 763 | 667 | NA | 739 |
| 250.2 | 1.26 [1.24-1.27] | 1.26 [1.25-1.28] | 1.25 [1.24-1.26] | 1.12 [1.00-1.26] | 1.20 [1.18-1.22] | 1.19 [1.18-1.21] | 1.13 [1.11-1.14] | 1.11 [1.10-1.13] | 1.10 [0.98-1.23] | 1.16 [1.15-1.18] | 76,947 | 76,654 | 73,593 | 821 | 60,531 |
| 250.21 | 1.19 [1.09-1.29] | 1.18 [1.08-1.28] | 1.16 [1.06-1.26] | | 1.09 [1.00-1.19] | 1.12 [1.03-1.22] | 1.03 [0.94-1.12] | 1.00 [0.91-1.09] | | 1.08 [0.99-1.18] | 1,328 | 1,311 | 1,230 | NA | 1,202 |
| 250.22 | 1.45 [1.36-1.55] | 1.43 [1.34-1.53] | 1.43 [1.34-1.53] | | 1.28 [1.20-1.37] | 1.35 [1.26-1.44] | 1.22 [1.14-1.31] | 1.20 [1.12-1.29] | | 1.26 [1.18-1.35] | 2,350 | 2,348 | 2,312 | NA | 2,274 |
| 250.23 | 1.24 [1.20-1.28] | 1.25 [1.21-1.28] | 1.24 [1.20-1.28] | 1.09 [0.79-1.49] | 1.16 [1.13-1.20] | 1.17 [1.14-1.21] | 1.09 [1.06-1.13] | 1.08 [1.04-1.11] | 1.14 [0.82-1.58] | 1.14 [1.11-1.18] | 10,348 | 10,339 | 9,952 | 102 | 9,448 |
| 250.24 | 1.41 [1.34-1.48] | 1.41 [1.34-1.48] | 1.43 [1.36-1.50] | | 1.28 [1.22-1.35] | 1.27 [1.21-1.34] | 1.14 [1.08-1.20] | 1.16 [1.10-1.22] | | 1.21 [1.15-1.27] | 4,276 | 4,270 | 4,152 | NA | 4,098 |
| 250.3 | 1.21 [1.03-1.43] | 1.30 [1.09-1.56] | 1.43 [1.17-1.76] | | 1.14 [0.96-1.34] | 1.10 [0.93-1.30] | 1.11 [0.92-1.34] | 1.19 [0.96-1.48] | | 1.11 [0.93-1.31] | 380 | 304 | 247 | NA | 363 |
| 250.41 | 1.34 [1.27-1.42] | 1.34 [1.27-1.42] | 1.34 [1.26-1.43] | 1.33 [1.01-1.77] | 1.29 [1.21-1.37] | 1.24 [1.17-1.32] | 1.21 [1.13-1.28] | 1.20 [1.12-1.28] | 1.29 [0.96-1.72] | 1.22 [1.15-1.30] | 3,035 | 2,933 | 2,683 | 150 | 2,778 |
| 250.42 | 1.28 [1.20-1.36] | 1.29 [1.20-1.38] | 1.28 [1.19-1.37] | 1.15 [0.97-1.37] | 1.21 [1.14-1.29] | 1.18 [1.11-1.26] | 1.13 [1.06-1.21] | 1.12 [1.04-1.21] | 1.08 [0.90-1.29] | 1.16 [1.08-1.24] | 2,477 | 2,209 | 1,978 | 337 | 2,265 |
| 250.5 | 1.19 [1.06-1.34] | 1.16 [1.02-1.33] | 1.10 [0.82-1.47] | 1.16 [0.96-1.41] | 1.14 [0.99-1.30] | 1.16 [1.02-1.31] | 1.09 [0.95-1.25] | 1.01 [0.75-1.36] | 1.16 [0.95-1.42] | 1.12 [0.98-1.29] | 674 | 512 | 106 | 271 | 537 |
| 250.6 | 1.46 [1.38-1.54] | 1.45 [1.37-1.53] | 1.47 [1.39-1.55] | | 1.33 [1.26-1.40] | 1.31 [1.24-1.38] | 1.16 [1.10-1.23] | 1.17 [1.11-1.24] | | 1.24 [1.18-1.31] | 3,551 | 3,548 | 3,456 | NA | 3,443 |
| 250.7 | 1.25 [1.21-1.29] | 1.25 [1.21-1.29] | 1.24 [1.20-1.28] | 1.07 [0.78-1.46] | 1.17 [1.13-1.21] | 1.18 [1.14-1.22] | 1.09 [1.06-1.13] | 1.08 [1.05-1.12] | 1.11 [0.81-1.53] | 1.15 [1.11-1.19] | 9,859 | 9,848 | 9,450 | 104 | 8,998 |
| 251 | 1.25 [1.22-1.29] | 1.27 [1.24-1.31] | 1.27 [1.23-1.31] | 1.07 [0.99-1.15] | 1.16 [1.13-1.20] | 1.18 [1.14-1.21] | 1.11 [1.08-1.14] | 1.10 [1.07-1.14] | 1.05 [0.97-1.14] | 1.14 [1.10-1.17] | 12,636 | 11,294 | 10,504 | 1,633 | 11,955 |
| 251.1 | 1.25 [1.22-1.29] | 1.27 [1.24-1.31] | 1.27 [1.23-1.31] | 1.06 [0.98-1.15] | 1.16 [1.13-1.20] | 1.18 [1.14-1.21] | 1.11 [1.08-1.14] | 1.10 [1.07-1.14] | 1.05 [0.97-1.14] | 1.14 [1.10-1.17] | 12,590 | 11,257 | 10,471 | 1,622 | 11,910 |
| 252.1 | 1.17 [1.11-1.23] | 1.17 [1.11-1.23] | 1.16 [1.10-1.22] | | 1.09 [1.03-1.15] | 1.12 [1.06-1.18] | 1.08 [1.02-1.14] | 1.07 [1.02-1.14] | | 1.06 [1.00-1.12] | 3,538 | 3,509 | 3,366 | NA | 3,236 |
| 252.2 | 1.40 [1.25-1.57] | 1.42 [1.27-1.60] | 1.42 [1.26-1.60] | | 1.26 [1.13-1.41] | 1.27 [1.13-1.43] | 1.26 [1.12-1.41] | 1.26 [1.12-1.43] | | 1.20 [1.07-1.35] | 818 | 784 | 706 | NA | 772 |
| 253 | 1.39 [1.20-1.62] | 1.39 [1.19-1.64] | 1.47 [1.22-1.76] | | 1.35 [1.15-1.57] | 1.29 [1.11-1.50] | 1.23 [1.04-1.46] | 1.31 [1.09-1.58] | | 1.30 [1.10-1.52] | 460 | 388 | 304 | NA | 418 |
| 253.1 | 1.47 [1.39-1.56] | 1.46 [1.38-1.55] | 1.50 [1.41-1.60] | 1.33 [1.06-1.68] | 1.39 [1.30-1.47] | 1.40 [1.32-1.49] | 1.36 [1.28-1.44] | 1.39 [1.31-1.49] | 1.27 [1.00-1.61] | 1.34 [1.26-1.43] | 3,085 | 2,955 | 2,737 | 205 | 2,799 |
| 253.11 | 1.27 [1.00-1.60] | 1.23 [0.93-1.63] | | | 1.01 [0.78-1.30] | 1.17 [0.92-1.49] | 1.13 [0.84-1.50] | | | 0.98 [0.75-1.27] | 181 | 118 | NA | NA | 145 |
| 253.2 | 1.44 [1.30-1.58] | 1.54 [1.38-1.73] | 1.57 [1.38-1.79] | 1.31 [1.11-1.54] | 1.24 [1.12-1.37] | 1.34 [1.22-1.48] | 1.36 [1.21-1.53] | 1.41 [1.24-1.61] | 1.24 [1.05-1.47] | 1.19 [1.08-1.32] | 1,120 | 777 | 605 | 415 | 1,010 |
| 253.3 | 1.44 [1.22-1.70] | 1.43 [1.19-1.72] | 1.61 [1.30-1.99] | | 1.30 [1.09-1.55] | 1.35 [1.14-1.61] | 1.29 [1.06-1.57] | 1.43 [1.15-1.77] | | 1.24 [1.03-1.48] | 360 | 282 | 227 | NA | 313 |
| 253.7 | 1.52 [1.42-1.63] | 1.52 [1.41-1.62] | 1.52 [1.42-1.64] | | 1.44 [1.34-1.55] | 1.46 [1.36-1.57] | 1.42 [1.33-1.53] | 1.43 [1.33-1.54] | | 1.40 [1.30-1.51] | 2,343 | 2,311 | 2,288 | NA | 2,147 |
| 255 | 1.23 [1.13-1.34] | 1.22 [1.11-1.34] | 1.17 [1.06-1.28] | | 1.17 [1.06-1.28] | 1.18 [1.08-1.29] | 1.12 [1.02-1.23] | 1.06 [0.96-1.17] | | 1.15 [1.05-1.26] | 1,257 | 1,192 | 1,101 | NA | 1,161 |
| 255.11 | 1.60 [1.35-1.90] | 1.46 [1.22-1.74] | 1.61 [1.33-1.96] | | 1.40 [1.17-1.67] | 1.32 [1.10-1.58] | 1.12 [0.93-1.36] | 1.24 [1.01-1.52] | | 1.21 [1.01-1.45] | 357 | 319 | 263 | NA | 327 |
| 255.12 | 1.21 [0.95-1.54] | 1.16 [0.91-1.49] | 1.14 [0.87-1.50] | | 1.16 [0.90-1.50] | 1.13 [0.88-1.44] | 1.04 [0.80-1.34] | 0.97 [0.73-1.29] | | 1.14 [0.88-1.48] | 163 | 150 | 124 | NA | 142 |
| 255.21 | 1.72 [1.60-1.85] | 1.73 [1.60-1.87] | 1.75 [1.61-1.90] | 1.45 [1.21-1.74] | 1.55 [1.44-1.67] | 1.47 [1.36-1.58] | 1.43 [1.32-1.55] | 1.45 [1.33-1.59] | 1.31 [1.08-1.58] | 1.38 [1.28-1.49] | 2,070 | 1,833 | 1,510 | 338 | 1,932 |
| 256 | 1.20 [1.16-1.25] | 1.24 [1.19-1.28] | 1.25 [1.14-1.38] | 1.13 [1.06-1.21] | 1.17 [1.12-1.22] | 1.10 [1.06-1.14] | 1.07 [1.03-1.12] | 1.04 [0.94-1.15] | 1.10 [1.03-1.18] | 1.10 [1.06-1.15] | 7,246 | 6,975 | 990 | 2,350 | 5,906 |
| 256.4 | 1.21 [1.16-1.26] | 1.24 [1.19-1.29] | 1.27 [1.15-1.41] | 1.14 [1.06-1.22] | 1.17 [1.12-1.22] | 1.10 [1.06-1.15] | 1.07 [1.03-1.12] | 1.05 [0.94-1.16] | 1.10 [1.03-1.18] | 1.10 [1.06-1.15] | 6,957 | 6,700 | 902 | 2,307 | 5,676 |
| 257 | 1.71 [1.47-1.98] | 1.76 [1.51-2.05] | 1.76 [1.50-2.08] | | 1.45 [1.25-1.69] | 1.50 [1.28-1.75] | 1.42 [1.21-1.67] | 1.40 [1.18-1.66] | | 1.32 [1.13-1.55] | 466 | 445 | 385 | NA | 430 |
| 257.1 | 1.71 [1.47-2.00] | 1.76 [1.51-2.06] | 1.77 [1.50-2.09] | | 1.47 [1.26-1.71] | 1.50 [1.28-1.75] | 1.43 [1.21-1.68] | 1.40 [1.18-1.67] | | 1.34 [1.14-1.57] | 449 | 430 | 373 | NA | 417 |
| 258 | 1.47 [1.22-1.79] | 1.45 [1.19-1.77] | 1.45 [1.17-1.79] | | 1.23 [1.01-1.49] | 1.33 [1.09-1.63] | 1.24 [1.01-1.53] | 1.24 [0.99-1.54] | | 1.18 [0.97-1.44] | 272 | 253 | 221 | NA | 255 |
| 259 | 1.25 [1.15-1.35] | 1.35 [1.16-1.57] | 1.46 [1.21-1.77] | 1.20 [1.09-1.32] | 1.17 [1.08-1.28] | 1.16 [1.06-1.26] | 1.17 [1.00-1.37] | 1.28 [1.05-1.56] | 1.12 [1.02-1.23] | 1.12 [1.03-1.23] | 1,544 | 426 | 270 | 1,169 | 1,378 |
| 259.1 | 1.43 [1.16-1.77] | 1.40 [1.11-1.76] | 1.27 [0.97-1.67] | | 1.35 [1.08-1.69] | 1.25 [1.00-1.56] | 1.21 [0.95-1.54] | 1.10 [0.83-1.46] | | 1.24 [0.98-1.57] | 227 | 188 | 129 | NA | 208 |
| 259.3 | 1.18 [0.98-1.43] | | | 1.16 [0.95-1.40] | 1.00 [0.81-1.23] | 1.16 [0.95-1.41] | | | 1.12 [0.92-1.37] | 1.00 [0.81-1.23] | 297 | NA | NA | 295 | 257 |
| 259.4 | 1.22 [1.06-1.41] | | | 1.22 [1.05-1.40] | 1.20 [1.03-1.40] | 1.17 [1.01-1.35] | | | 1.16 [1.00-1.34] | 1.17 [1.00-1.36] | 521 | NA | NA | 516 | 449 |
| 260 | 1.21 [1.13-1.30] | 1.18 [1.10-1.27] | 1.16 [1.08-1.25] | 1.31 [0.99-1.74] | 1.16 [1.08-1.24] | 1.19 [1.11-1.28] | 1.10 [1.03-1.19] | 1.09 [1.01-1.18] | 1.27 [0.95-1.71] | 1.16 [1.08-1.25] | 2,081 | 2,012 | 1,832 | 139 | 1,930 |
| 260.1 | 1.16 [1.05-1.28] | 1.12 [1.01-1.24] | 1.10 [1.00-1.22] | | 1.09 [0.98-1.21] | 1.17 [1.06-1.30] | 1.09 [0.99-1.21] | 1.08 [0.97-1.20] | | 1.12 [1.01-1.25] | 1,044 | 1,034 | 1,014 | NA | 970 |
| 260.22 | 1.07 [0.80-1.42] | 1.05 [0.78-1.41] | 1.10 [0.82-1.48] | | 1.15 [0.86-1.55] | 1.10 [0.82-1.47] | 1.06 [0.78-1.42] | 1.09 [0.81-1.48] | | 1.20 [0.89-1.61] | 123 | 121 | 113 | NA | 118 |
| 260.6 | 1.21 [1.17-1.26] | 1.21 [1.16-1.26] | 1.23 [1.18-1.28] | 1.07 [0.98-1.17] | 1.15 [1.11-1.20] | 1.16 [1.12-1.21] | 1.12 [1.07-1.16] | 1.15 [1.10-1.19] | 1.03 [0.94-1.13] | 1.13 [1.09-1.18] | 7,715 | 6,704 | 5,932 | 1,361 | 6,978 |
| 261 | 1.27 [1.00-1.62] | 1.29 [1.00-1.67] | 1.21 [0.92-1.60] | | 1.21 [0.94-1.54] | 1.19 [0.93-1.53] | 1.14 [0.87-1.48] | 1.10 [0.83-1.46] | | 1.18 [0.92-1.52] | 176 | 155 | 132 | NA | 165 |
| 261.2 | 1.31 [1.27-1.35] | 1.31 [1.27-1.35] | 1.29 [1.25-1.33] | 1.26 [1.11-1.44] | 1.24 [1.20-1.28] | 1.23 [1.20-1.27] | 1.18 [1.15-1.22] | 1.17 [1.13-1.21] | 1.19 [1.03-1.36] | 1.20 [1.16-1.23] | 12,428 | 12,315 | 10,879 | 652 | 11,582 |
| 261.4 | 1.37 [1.34-1.40] | 1.38 [1.34-1.42] | 1.35 [1.32-1.39] | 1.34 [1.25-1.45] | 1.30 [1.26-1.33] | 1.26 [1.23-1.29] | 1.22 [1.19-1.25] | 1.20 [1.17-1.24] | 1.26 [1.17-1.35] | 1.23 [1.19-1.26] | 18,051 | 16,575 | 14,208 | 2,279 | 16,868 |
| 261.41 | 1.33 [1.11-1.60] | 1.24 [1.01-1.52] | 1.29 [1.04-1.60] | | 1.31 [1.08-1.58] | 1.22 [1.02-1.47] | 1.13 [0.91-1.39] | 1.17 [0.93-1.46] | | 1.23 [1.02-1.50] | 306 | 242 | 217 | NA | 285 |
| 262 | 1.40 [1.32-1.49] | 1.40 [1.31-1.49] | 1.40 [1.30-1.50] | 1.39 [1.20-1.61] | 1.29 [1.22-1.38] | 1.28 [1.20-1.36] | 1.23 [1.15-1.31] | 1.23 [1.15-1.32] | 1.31 [1.12-1.52] | 1.22 [1.14-1.30] | 2,987 | 2,637 | 2,192 | 540 | 2,778 |
| 263 | 1.21 [1.02-1.44] | 1.10 [0.92-1.31] | 1.28 [1.06-1.55] | | 1.13 [0.95-1.35] | 1.17 [0.98-1.40] | 1.01 [0.84-1.21] | 1.20 [0.99-1.46] | | 1.12 [0.94-1.34] | 345 | 308 | 278 | NA | 325 |
| 264.3 | 0.98 [0.85-1.14] | | | 1.00 [0.86-1.16] | 0.90 [0.77-1.05] | 0.93 [0.80-1.08] | | | 0.96 [0.83-1.11] | 0.88 [0.75-1.03] | 434 | NA | NA | 435 | 401 |
| 264.9 | 1.08 [1.03-1.14] | 1.10 [0.94-1.28] | 1.25 [1.01-1.55] | 1.07 [1.01-1.13] | 1.01 [0.95-1.07] | 1.03 [0.98-1.09] | 1.00 [0.86-1.18] | 1.17 [0.94-1.46] | 1.03 [0.97-1.09] | 0.98 [0.93-1.04] | 3,528 | 408 | 227 | 3,186 | 3,157 |
| 269 | 1.33 [1.23-1.45] | 1.38 [1.26-1.52] | 1.44 [1.29-1.60] | 1.17 [0.99-1.39] | 1.24 [1.13-1.36] | 1.28 [1.18-1.40] | 1.26 [1.14-1.39] | 1.30 [1.16-1.44] | 1.16 [0.98-1.38] | 1.21 [1.10-1.32] | 1,442 | 1,128 | 889 | 396 | 1,232 |
| 270.1 | 1.25 [0.99-1.57] | 1.33 [0.99-1.80] | | | 1.20 [0.93-1.54] | 1.20 [0.95-1.52] | 1.22 [0.90-1.67] | | | 1.17 [0.90-1.51] | 188 | 108 | NA | NA | 158 |
| 270.32 | 1.35 [1.25-1.46] | 1.36 [1.26-1.47] | 1.36 [1.26-1.47] | | 1.28 [1.18-1.38] | 1.27 [1.17-1.37] | 1.25 [1.16-1.35] | 1.23 [1.14-1.33] | | 1.23 [1.14-1.33] | 1,905 | 1,906 | 1,896 | NA | 1,811 |
| 270.33 | 1.25 [1.10-1.41] | 1.22 [1.07-1.38] | 1.18 [1.04-1.35] | | 1.18 [1.04-1.34] | 1.20 [1.06-1.36] | 1.14 [1.00-1.29] | 1.10 [0.96-1.25] | | 1.14 [1.00-1.30] | 655 | 623 | 590 | NA | 596 |
| 270.35 | 1.35 [1.11-1.64] | 1.28 [1.06-1.56] | 1.36 [1.12-1.66] | | 1.24 [1.01-1.52] | 1.32 [1.08-1.60] | 1.26 [1.03-1.53] | 1.32 [1.08-1.62] | | 1.22 [0.99-1.50] | 267 | 266 | 265 | NA | 240 |
| 271.3 | 1.39 [1.27-1.52] | 1.37 [1.22-1.54] | 1.33 [1.16-1.53] | 1.35 [1.19-1.53] | 1.31 [1.19-1.44] | 1.22 [1.11-1.34] | 1.13 [1.00-1.28] | 1.11 [0.96-1.29] | 1.22 [1.07-1.39] | 1.17 [1.06-1.28] | 1,313 | 728 | 486 | 679 | 1,157 |
| 271.9 | 1.36 [1.09-1.69] | 1.40 [1.05-1.87] | | 1.26 [0.94-1.70] | 1.31 [1.05-1.63] | 1.28 [1.02-1.60] | 1.28 [0.95-1.73] | | 1.23 [0.91-1.68] | 1.27 [1.01-1.60] | 227 | 118 | NA | 123 | 206 |
| 272.1 | 1.27 [1.23-1.30] | 1.26 [1.23-1.30] | 1.26 [1.23-1.29] | | 1.17 [1.14-1.20] | 1.19 [1.16-1.22] | 1.12 [1.10-1.15] | 1.12 [1.09-1.15] | | 1.13 [1.10-1.16] | 16,512 | 16,499 | 16,297 | NA | 14,971 |
| 272.11 | 1.23 [1.22-1.24] | 1.23 [1.22-1.24] | 1.23 [1.22-1.24] | 1.19 [0.96-1.48] | 1.15 [1.14-1.17] | 1.17 [1.16-1.18] | 1.11 [1.10-1.13] | 1.11 [1.10-1.12] | 1.17 [0.94-1.47] | 1.12 [1.11-1.14] | 90,425 | 90,380 | 89,053 | 219 | 77,085 |
| 272.12 | 1.58 [1.34-1.86] | 1.69 [1.43-2.00] | 1.54 [1.30-1.84] | | 1.46 [1.23-1.73] | 1.41 [1.19-1.68] | 1.46 [1.23-1.74] | 1.33 [1.11-1.60] | | 1.40 [1.17-1.66] | 388 | 381 | 336 | NA | 352 |
| 272.13 | 1.40 [1.23-1.59] | 1.44 [1.26-1.63] | 1.42 [1.25-1.61] | | 1.30 [1.14-1.48] | 1.26 [1.11-1.44] | 1.16 [1.02-1.33] | 1.18 [1.03-1.35] | | 1.21 [1.06-1.38] | 639 | 636 | 622 | NA | 582 |
| 272.9 | 1.26 [1.16-1.36] | 1.26 [1.17-1.37] | 1.31 [1.21-1.42] | | 1.17 [1.08-1.28] | 1.16 [1.07-1.26] | 1.09 [1.01-1.19] | 1.13 [1.04-1.23] | | 1.12 [1.03-1.22] | 1,554 | 1,553 | 1,527 | NA | 1,455 |
| 274.1 | 1.36 [1.33-1.39] | 1.36 [1.33-1.39] | 1.36 [1.33-1.39] | | 1.27 [1.24-1.30] | 1.29 [1.26-1.32] | 1.24 [1.21-1.27] | 1.23 [1.20-1.26] | | 1.22 [1.19-1.25] | 19,181 | 19,179 | 18,861 | NA | 17,324 |
| 274.11 | 1.31 [1.15-1.48] | 1.33 [1.17-1.51] | 1.26 [1.11-1.44] | | 1.21 [1.06-1.38] | 1.26 [1.11-1.44] | 1.24 [1.09-1.41] | 1.16 [1.02-1.33] | | 1.19 [1.04-1.36] | 640 | 643 | 625 | NA | 591 |
| 274.2 | 1.33 [1.17-1.50] | 1.31 [1.16-1.48] | 1.37 [1.21-1.55] | | 1.30 [1.15-1.47] | 1.25 [1.10-1.41] | 1.19 [1.05-1.35] | 1.25 [1.11-1.42] | | 1.25 [1.10-1.42] | 781 | 781 | 774 | NA | 744 |
| 274.21 | 1.31 [1.22-1.42] | 1.32 [1.23-1.43] | 1.29 [1.20-1.39] | | 1.25 [1.16-1.35] | 1.22 [1.13-1.32] | 1.20 [1.11-1.29] | 1.17 [1.08-1.26] | | 1.18 [1.10-1.28] | 1,911 | 1,885 | 1,775 | NA | 1,754 |
| 275 | 1.21 [1.18-1.24] | 1.22 [1.19-1.25] | 1.20 [1.17-1.23] | 1.08 [0.96-1.20] | 1.15 [1.12-1.18] | 1.16 [1.13-1.19] | 1.12 [1.09-1.15] | 1.10 [1.08-1.13] | 1.06 [0.95-1.19] | 1.13 [1.10-1.16] | 17,831 | 17,352 | 16,193 | 875 | 16,376 |
| 275.3 | 1.25 [1.20-1.30] | 1.26 [1.21-1.32] | 1.25 [1.20-1.30] | 1.02 [0.83-1.26] | 1.19 [1.14-1.24] | 1.17 [1.13-1.22] | 1.12 [1.07-1.17] | 1.11 [1.06-1.16] | 1.01 [0.82-1.25] | 1.16 [1.11-1.21] | 6,400 | 6,267 | 5,950 | 249 | 6,060 |
| 275.5 | 1.18 [1.14-1.22] | 1.18 [1.15-1.22] | 1.17 [1.13-1.21] | 0.96 [0.81-1.15] | 1.13 [1.09-1.16] | 1.14 [1.10-1.18] | 1.10 [1.07-1.14] | 1.09 [1.05-1.12] | 0.95 [0.79-1.13] | 1.12 [1.08-1.16] | 10,042 | 9,830 | 9,362 | 334 | 9,210 |
| 275.53 | 1.25 [1.18-1.32] | 1.27 [1.20-1.34] | 1.23 [1.16-1.30] | 1.14 [0.95-1.36] | 1.16 [1.09-1.22] | 1.19 [1.12-1.26] | 1.16 [1.09-1.22] | 1.12 [1.05-1.19] | 1.13 [0.94-1.35] | 1.13 [1.07-1.20] | 3,624 | 3,453 | 3,070 | 337 | 3,359 |
| 276.1 | 1.20 [1.11-1.30] | 1.23 [1.14-1.33] | 1.22 [1.13-1.33] | 1.18 [0.88-1.57] | 1.18 [1.09-1.28] | 1.13 [1.04-1.22] | 1.11 [1.02-1.20] | 1.10 [1.01-1.20] | 1.11 [0.82-1.49] | 1.16 [1.07-1.26] | 1,686 | 1,622 | 1,478 | 125 | 1,596 |
| 276.11 | 1.15 [1.10-1.20] | 1.16 [1.11-1.21] | 1.17 [1.12-1.22] | 0.87 [0.69-1.09] | 1.12 [1.07-1.17] | 1.14 [1.09-1.19] | 1.11 [1.06-1.16] | 1.12 [1.07-1.17] | 0.84 [0.67-1.06] | 1.13 [1.07-1.18] | 5,363 | 5,226 | 5,096 | 190 | 5,020 |
| 276.12 | 1.38 [1.36-1.41] | 1.38 [1.36-1.41] | 1.38 [1.36-1.41] | 0.99 [0.86-1.14] | 1.32 [1.29-1.35] | 1.33 [1.30-1.35] | 1.28 [1.25-1.30] | 1.28 [1.25-1.30] | 0.99 [0.86-1.14] | 1.29 [1.26-1.31] | 29,451 | 29,107 | 28,537 | 500 | 26,962 |
| 276.13 | 1.29 [1.25-1.33] | 1.29 [1.25-1.32] | 1.28 [1.25-1.32] | 0.97 [0.79-1.20] | 1.21 [1.17-1.24] | 1.24 [1.21-1.28] | 1.18 [1.15-1.22] | 1.17 [1.14-1.21] | 0.96 [0.78-1.19] | 1.20 [1.16-1.23] | 13,239 | 13,106 | 12,753 | 237 | 12,419 |
| 276.14 | 1.22 [1.20-1.25] | 1.23 [1.20-1.26] | 1.22 [1.19-1.25] | 1.12 [1.02-1.24] | 1.17 [1.15-1.20] | 1.17 [1.14-1.20] | 1.13 [1.10-1.16] | 1.12 [1.09-1.15] | 1.10 [0.99-1.22] | 1.15 [1.12-1.18] | 21,009 | 20,373 | 19,152 | 1,090 | 19,482 |
| 276.41 | 1.24 [1.21-1.27] | 1.24 [1.21-1.27] | 1.24 [1.21-1.28] | 1.04 [0.94-1.15] | 1.18 [1.15-1.21] | 1.18 [1.15-1.21] | 1.12 [1.09-1.15] | 1.12 [1.09-1.15] | 1.02 [0.92-1.13] | 1.15 [1.12-1.18] | 16,781 | 16,079 | 15,262 | 999 | 15,629 |
| 276.42 | 1.27 [1.17-1.37] | 1.29 [1.19-1.39] | 1.27 [1.17-1.37] | 0.93 [0.71-1.22] | 1.18 [1.10-1.28] | 1.18 [1.10-1.28] | 1.14 [1.05-1.24] | 1.13 [1.04-1.23] | 0.86 [0.65-1.14] | 1.13 [1.04-1.22] | 1,779 | 1,711 | 1,536 | 138 | 1,654 |
| 276.5 | 1.20 [1.18-1.21] | 1.19 [1.17-1.21] | 1.19 [1.17-1.21] | 1.13 [1.08-1.18] | 1.14 [1.12-1.16] | 1.16 [1.14-1.18] | 1.12 [1.10-1.14] | 1.12 [1.10-1.14] | 1.10 [1.05-1.15] | 1.13 [1.11-1.15] | 47,668 | 43,859 | 41,005 | 4,893 | 43,955 |
| 276.6 | 1.32 [1.28-1.37] | 1.33 [1.29-1.37] | 1.32 [1.28-1.37] | 1.02 [0.80-1.29] | 1.26 [1.22-1.30] | 1.26 [1.22-1.30] | 1.20 [1.17-1.24] | 1.19 [1.15-1.23] | 1.02 [0.79-1.30] | 1.22 [1.19-1.27] | 10,985 | 10,880 | 10,645 | 184 | 10,552 |
| 276.8 | 1.34 [1.20-1.50] | 1.53 [1.32-1.79] | 1.59 [1.34-1.89] | 1.14 [0.97-1.33] | 1.24 [1.10-1.39] | 1.26 [1.13-1.41] | 1.37 [1.17-1.60] | 1.38 [1.15-1.65] | 1.09 [0.93-1.28] | 1.17 [1.04-1.32] | 808 | 442 | 348 | 400 | 694 |
| 277 | 1.22 [1.19-1.26] | 1.24 [1.20-1.28] | 1.22 [1.18-1.26] | 1.16 [1.09-1.24] | 1.16 [1.12-1.19] | 1.17 [1.14-1.21] | 1.15 [1.11-1.19] | 1.14 [1.10-1.18] | 1.13 [1.06-1.21] | 1.14 [1.10-1.17] | 12,880 | 10,907 | 9,674 | 2,445 | 11,980 |
| 277.2 | 1.37 [1.05-1.78] | 1.38 [1.06-1.82] | 1.44 [1.07-1.93] | | 1.23 [0.93-1.62] | 1.31 [1.00-1.70] | 1.24 [0.94-1.64] | 1.25 [0.92-1.70] | | 1.17 [0.88-1.54] | 149 | 137 | 116 | NA | 131 |
| 277.4 | 1.16 [1.06-1.27] | 1.20 [1.10-1.32] | 1.20 [1.07-1.34] | 1.11 [0.91-1.36] | 1.07 [0.97-1.18] | 1.09 [0.99-1.19] | 1.11 [1.00-1.22] | 1.12 [1.00-1.25] | 1.04 [0.84-1.28] | 1.02 [0.92-1.12] | 1,218 | 1,090 | 769 | 256 | 1,055 |
| 277.5 | 1.30 [1.18-1.43] | 1.37 [1.23-1.51] | 1.37 [1.23-1.53] | 1.00 [0.78-1.27] | 1.22 [1.10-1.34] | 1.21 [1.10-1.34] | 1.19 [1.07-1.32] | 1.19 [1.06-1.33] | 0.97 [0.76-1.25] | 1.17 [1.06-1.30] | 1,100 | 974 | 865 | 169 | 1,035 |
| 277.51 | 1.57 [1.31-1.88] | 1.59 [1.31-1.92] | 1.70 [1.39-2.08] | | 1.39 [1.15-1.68] | 1.48 [1.23-1.78] | 1.39 [1.14-1.70] | 1.50 [1.22-1.85] | | 1.35 [1.11-1.64] | 305 | 284 | 257 | NA | 280 |
| 278.1 | 1.30 [1.29-1.32] | 1.32 [1.30-1.33] | 1.32 [1.31-1.34] | 1.18 [1.14-1.22] | 1.22 [1.21-1.24] | 1.20 [1.19-1.21] | 1.15 [1.14-1.17] | 1.15 [1.13-1.16] | 1.13 [1.10-1.17] | 1.15 [1.14-1.17] | 88,230 | 86,031 | 60,487 | 11,250 | 77,698 |
| 278.3 | 1.45 [1.08-1.95] | 1.54 [1.14-2.08] | | | | 1.33 [0.98-1.80] | 1.39 [1.02-1.90] | | | | 113 | 107 | NA | NA | NA |
| 278.4 | 1.51 [1.30-1.76] | 1.51 [1.26-1.81] | 1.53 [1.25-1.89] | 1.48 [1.16-1.88] | 1.39 [1.19-1.62] | 1.36 [1.17-1.59] | 1.26 [1.04-1.52] | 1.28 [1.03-1.59] | 1.37 [1.06-1.76] | 1.28 [1.09-1.50] | 482 | 318 | 233 | 203 | 448 |
| 279 | 1.66 [1.31-2.09] | 1.62 [1.23-2.13] | 1.55 [1.13-2.14] | | 1.50 [1.18-1.90] | 1.47 [1.15-1.87] | 1.33 [0.99-1.78] | 1.30 [0.93-1.82] | | 1.39 [1.09-1.77] | 204 | 141 | 105 | NA | 190 |
| 279.1 | 1.52 [1.38-1.67] | 1.57 [1.41-1.76] | 1.49 [1.32-1.68] | 1.45 [1.22-1.72] | 1.44 [1.30-1.59] | 1.34 [1.21-1.48] | 1.33 [1.18-1.49] | 1.23 [1.09-1.40] | 1.31 [1.10-1.56] | 1.30 [1.18-1.44] | 1,161 | 828 | 681 | 384 | 1,085 |
| 279.11 | 1.51 [1.34-1.70] | 1.54 [1.35-1.77] | 1.48 [1.28-1.71] | 1.46 [1.16-1.82] | 1.44 [1.27-1.62] | 1.33 [1.18-1.51] | 1.31 [1.14-1.51] | 1.23 [1.05-1.44] | 1.32 [1.05-1.66] | 1.30 [1.15-1.47] | 744 | 565 | 463 | 216 | 696 |
| 279.7 | 1.37 [1.21-1.55] | 1.32 [1.15-1.52] | 1.37 [1.17-1.60] | 1.32 [1.02-1.70] | 1.28 [1.12-1.47] | 1.29 [1.14-1.47] | 1.22 [1.06-1.41] | 1.26 [1.08-1.48] | 1.27 [0.98-1.65] | 1.23 [1.07-1.41] | 648 | 521 | 418 | 168 | 549 |
| 279.8 | 1.64 [1.31-2.06] | 1.66 [1.28-2.16] | 1.49 [1.10-2.02] | | 1.67 [1.32-2.11] | 1.49 [1.18-1.89] | 1.48 [1.13-1.95] | 1.32 [0.97-1.81] | | 1.53 [1.20-1.95] | 207 | 151 | 109 | NA | 193 |
| hematopoietic | | | | | | | | | | | | | | | |
| 275.1 | 1.30 [1.20-1.42] | 1.32 [1.21-1.43] | 1.29 [1.18-1.42] | | 1.22 [1.12-1.34] | 1.26 [1.16-1.38] | 1.25 [1.15-1.37] | 1.23 [1.12-1.35] | | 1.20 [1.10-1.32] | 1,332 | 1,290 | 1,110 | NA | 1,125 |
| 280.1 | 1.32 [1.31-1.34] | 1.32 [1.30-1.34] | 1.32 [1.30-1.34] | 1.28 [1.21-1.35] | 1.25 [1.23-1.27] | 1.25 [1.23-1.27] | 1.20 [1.18-1.22] | 1.20 [1.18-1.22] | 1.24 [1.18-1.31] | 1.20 [1.19-1.22] | 53,162 | 50,841 | 45,714 | 4,007 | 47,985 |
| 280.2 | 1.22 [1.15-1.30] | 1.21 [1.14-1.28] | 1.23 [1.16-1.31] | 1.11 [0.90-1.36] | 1.15 [1.08-1.22] | 1.18 [1.12-1.26] | 1.14 [1.08-1.21] | 1.16 [1.09-1.24] | 1.10 [0.89-1.37] | 1.13 [1.06-1.20] | 3,120 | 3,000 | 2,489 | 254 | 2,730 |
| 281.1 | 1.16 [0.87-1.53] | 1.18 [0.88-1.57] | 1.29 [0.95-1.74] | | 0.94 [0.68-1.28] | 1.14 [0.85-1.52] | 1.09 [0.81-1.46] | 1.23 [0.90-1.67] | | 0.93 [0.67-1.28] | 125 | 122 | 114 | NA | 104 |
| 281.11 | 1.41 [1.33-1.48] | 1.39 [1.32-1.47] | 1.40 [1.32-1.48] | | 1.33 [1.25-1.40] | 1.31 [1.23-1.38] | 1.25 [1.18-1.32] | 1.26 [1.19-1.33] | | 1.26 [1.18-1.33] | 3,472 | 3,466 | 3,227 | NA | 3,117 |
| 281.12 | 1.38 [1.32-1.44] | 1.35 [1.29-1.42] | 1.36 [1.30-1.43] | 1.43 [1.07-1.91] | 1.29 [1.23-1.35] | 1.29 [1.23-1.35] | 1.23 [1.17-1.29] | 1.24 [1.18-1.31] | 1.33 [0.99-1.80] | 1.23 [1.17-1.29] | 4,681 | 4,667 | 4,277 | 132 | 4,210 |
| 281.13 | 1.28 [1.21-1.35] | 1.31 [1.24-1.38] | 1.30 [1.23-1.38] | | 1.25 [1.18-1.32] | 1.23 [1.17-1.31] | 1.22 [1.15-1.29] | 1.20 [1.14-1.28] | | 1.24 [1.17-1.31] | 3,437 | 3,417 | 3,254 | NA | 3,181 |

### Supplementary Table 6: phecode-mapped results (hazard ratios and events from all cohorts)

| | Hazard ratio (99% confidence interval) | | | | | | | | | | Events (in exposed) | | | | |
|---|---|---|---|---|---|---|---|---|---|---|---|---|---|---|---|
| | crude | | | | | adjusted | | | | | | | | | |
| Outcome | any age | 18+ | 40+ | <18 | hosp. | any age | 18+ | 40+ | <18 | hosp. | any age | 18+ | 40+ | <18 | hosp. |
| 281.9 | 1.48 [1.32-1.66] | 1.46 [1.30-1.64] | 1.56 [1.38-1.76] | | 1.42 [1.26-1.60] | 1.39 [1.23-1.56] | 1.32 [1.17-1.49] | 1.38 [1.22-1.57] | | 1.37 [1.22-1.55] | 835 | 811 | 786 | NA | 785 |
| 282.5 | 1.31 [1.23-1.40] | 1.37 [1.27-1.47] | 1.30 [1.17-1.44] | 1.36 [1.23-1.51] | 1.22 [1.13-1.31] | 1.28 [1.20-1.37] | 1.32 [1.22-1.43] | 1.23 [1.10-1.37] | 1.36 [1.22-1.51] | 1.21 [1.12-1.30] | 2,519 | 1,759 | 859 | 1,054 | 1,979 |
| 282.8 | 1.38 [1.29-1.48] | 1.41 [1.32-1.52] | 1.51 [1.38-1.66] | 1.31 [1.14-1.51] | 1.26 [1.17-1.35] | 1.35 [1.26-1.44] | 1.36 [1.27-1.47] | 1.40 [1.27-1.54] | 1.29 [1.12-1.49] | 1.25 [1.16-1.34] | 2,393 | 2,059 | 1,188 | 572 | 1,987 |
| 282.9 | 1.27 [1.13-1.43] | 1.30 [1.14-1.49] | 1.28 [1.09-1.50] | 1.22 [0.98-1.52] | 1.22 [1.08-1.39] | 1.25 [1.11-1.41] | 1.24 [1.08-1.43] | 1.21 [1.03-1.43] | 1.23 [0.98-1.54] | 1.22 [1.07-1.39] | 710 | 538 | 390 | 216 | 618 |
| 283 | 1.44 [1.28-1.62] | 1.47 [1.30-1.67] | 1.43 [1.25-1.64] | 1.27 [0.96-1.67] | 1.36 [1.20-1.54] | 1.42 [1.26-1.59] | 1.43 [1.25-1.63] | 1.38 [1.20-1.58] | 1.25 [0.94-1.67] | 1.36 [1.20-1.55] | 745 | 639 | 566 | 133 | 654 |
| 283.1 | 1.57 [1.36-1.81] | 1.58 [1.36-1.83] | 1.50 [1.29-1.75] | | 1.48 [1.27-1.72] | 1.53 [1.33-1.77] | 1.55 [1.33-1.80] | 1.44 [1.24-1.69] | | 1.47 [1.26-1.72] | 517 | 490 | 444 | NA | 455 |
| 284 | 1.31 [1.24-1.39] | 1.33 [1.25-1.41] | 1.30 [1.22-1.38] | 1.08 [0.89-1.31] | 1.25 [1.18-1.32] | 1.27 [1.20-1.35] | 1.26 [1.19-1.33] | 1.22 [1.15-1.30] | 1.05 [0.86-1.28] | 1.24 [1.17-1.31] | 3,302 | 3,131 | 2,874 | 259 | 3,032 |
| 285 | 1.30 [1.28-1.31] | 1.30 [1.29-1.32] | 1.30 [1.29-1.32] | 1.17 [1.11-1.23] | 1.23 [1.21-1.24] | 1.24 [1.23-1.26] | 1.21 [1.19-1.22] | 1.21 [1.19-1.22] | 1.14 [1.08-1.21] | 1.20 [1.18-1.21] | 69,443 | 67,794 | 61,959 | 3,476 | 62,148 |
| 285.1 | 1.13 [1.04-1.23] | 1.13 [1.04-1.23] | 1.18 [1.05-1.33] | 0.99 [0.79-1.23] | 1.07 [0.98-1.18] | 1.12 [1.03-1.22] | 1.10 [1.01-1.20] | 1.12 [0.99-1.26] | 0.98 [0.78-1.23] | 1.08 [0.99-1.19] | 1,430 | 1,401 | 686 | 211 | 1,170 |
| 285.2 | 1.43 [1.34-1.52] | 1.43 [1.34-1.52] | 1.41 [1.32-1.50] | | 1.30 [1.22-1.38] | 1.35 [1.27-1.44] | 1.28 [1.20-1.37] | 1.25 [1.17-1.34] | | 1.27 [1.19-1.36] | 2,731 | 2,710 | 2,583 | NA | 2,620 |
| 285.22 | 1.14 [1.07-1.22] | 1.15 [1.07-1.23] | 1.13 [1.06-1.21] | | 1.10 [1.02-1.18] | 1.14 [1.06-1.22] | 1.13 [1.06-1.21] | 1.11 [1.04-1.19] | | 1.11 [1.03-1.20] | 2,146 | 2,101 | 2,044 | NA | 1,845 |
| 286.1 | 1.85 [1.41-2.43] | 1.82 [1.32-2.51] | | | 1.46 [1.09-1.97] | 1.65 [1.25-2.19] | 1.53 [1.09-2.14] | | | 1.41 [1.04-1.91] | 148 | 104 | NA | NA | 117 |
| 286.11 | 1.26 [1.06-1.50] | 1.38 [1.13-1.69] | 1.32 [1.00-1.74] | 1.17 [0.91-1.52] | 1.18 [0.98-1.43] | 1.15 [0.96-1.37] | 1.19 [0.96-1.47] | 1.14 [0.85-1.52] | 1.11 [0.85-1.44] | 1.11 [0.91-1.35] | 334 | 231 | 116 | 158 | 269 |
| 286.12 | 1.22 [1.09-1.36] | 1.27 [1.13-1.43] | 1.35 [1.16-1.57] | 1.06 [0.81-1.38] | 1.11 [0.99-1.25] | 1.11 [0.99-1.25] | 1.12 [0.99-1.26] | 1.16 [0.99-1.36] | 0.98 [0.75-1.28] | 1.03 [0.92-1.17] | 744 | 663 | 399 | 152 | 652 |
| 286.13 | 1.15 [0.94-1.40] | 1.13 [0.90-1.43] | 1.18 [0.90-1.57] | | 1.20 [0.97-1.50] | 1.11 [0.90-1.36] | 1.03 [0.81-1.31] | 1.09 [0.82-1.45] | | 1.17 [0.94-1.47] | 242 | 177 | 121 | NA | 204 |
| 286.3 | 1.05 [0.94-1.17] | 1.04 [0.94-1.16] | | 0.96 [0.76-1.22] | 0.98 [0.88-1.10] | 1.03 [0.92-1.15] | 1.00 [0.90-1.12] | | 0.94 [0.74-1.20] | 0.98 [0.87-1.10] | 820 | 811 | NA | 176 | 673 |
| 286.5 | 1.31 [1.11-1.54] | 1.35 [1.15-1.59] | 1.34 [1.13-1.58] | | 1.11 [0.93-1.32] | 1.25 [1.06-1.47] | 1.24 [1.05-1.46] | 1.24 [1.04-1.46] | | 1.07 [0.90-1.27] | 370 | 371 | 361 | NA | 327 |
| 286.6 | 1.28 [1.22-1.35] | 1.32 [1.25-1.39] | 1.33 [1.25-1.41] | 1.09 [0.97-1.24] | 1.18 [1.13-1.25] | 1.19 [1.13-1.25] | 1.18 [1.12-1.24] | 1.18 [1.11-1.26] | 1.03 [0.91-1.17] | 1.13 [1.07-1.19] | 4,176 | 3,783 | 2,765 | 675 | 3,675 |
| 286.7 | 1.29 [1.22-1.35] | 1.32 [1.25-1.39] | 1.34 [1.26-1.42] | 1.09 [0.96-1.23] | 1.19 [1.13-1.25] | 1.19 [1.13-1.25] | 1.18 [1.11-1.24] | 1.18 [1.11-1.26] | 1.03 [0.90-1.17] | 1.13 [1.07-1.19] | 3,974 | 3,599 | 2,602 | 650 | 3,500 |
| 287 | 1.23 [1.20-1.27] | 1.27 [1.23-1.31] | 1.28 [1.24-1.33] | 1.06 [1.00-1.13] | 1.18 [1.14-1.21] | 1.20 [1.16-1.23] | 1.20 [1.16-1.24] | 1.20 [1.16-1.25] | 1.06 [1.00-1.13] | 1.17 [1.13-1.20] | 11,780 | 9,406 | 8,184 | 2,747 | 10,458 |
| 287.1 | 1.18 [1.11-1.24] | 1.32 [1.17-1.49] | 1.42 [1.24-1.61] | 1.14 [1.07-1.21] | 1.12 [1.06-1.19] | 1.13 [1.07-1.20] | 1.20 [1.07-1.36] | 1.30 [1.14-1.48] | 1.11 [1.04-1.18] | 1.09 [1.02-1.16] | 3,222 | 720 | 613 | 2,545 | 2,877 |
| 287.2 | 1.10 [1.01-1.19] | 1.32 [1.07-1.63] | 1.44 [1.10-1.89] | 1.05 [0.97-1.15] | 1.11 [1.02-1.21] | 1.08 [1.00-1.17] | 1.22 [0.98-1.51] | 1.29 [0.98-1.70] | 1.05 [0.97-1.15] | 1.10 [1.01-1.20] | 1,587 | 218 | 136 | 1,406 | 1,341 |
| 287.3 | 1.26 [1.22-1.30] | 1.27 [1.22-1.31] | 1.29 [1.24-1.34] | 1.07 [0.96-1.20] | 1.20 [1.15-1.24] | 1.22 [1.18-1.27] | 1.20 [1.16-1.25] | 1.21 [1.16-1.26] | 1.06 [0.95-1.19] | 1.19 [1.15-1.23] | 8,323 | 7,736 | 6,795 | 855 | 7,552 |
| 287.31 | 1.25 [1.16-1.34] | 1.30 [1.20-1.41] | 1.31 [1.20-1.43] | 1.12 [0.97-1.30] | 1.18 [1.09-1.27] | 1.21 [1.12-1.30] | 1.23 [1.13-1.34] | 1.23 [1.12-1.34] | 1.10 [0.95-1.28] | 1.16 [1.07-1.26] | 1,872 | 1,487 | 1,252 | 453 | 1,617 |
| 287.32 | 1.39 [1.20-1.62] | 1.34 [1.15-1.57] | 1.34 [1.14-1.58] | | 1.27 [1.08-1.48] | 1.35 [1.16-1.58] | 1.25 [1.07-1.47] | 1.24 [1.05-1.46] | | 1.28 [1.09-1.50] | 452 | 420 | 376 | NA | 404 |
| 288 | 1.26 [1.20-1.34] | 1.26 [1.19-1.34] | 1.25 [1.18-1.33] | 1.22 [1.06-1.40] | 1.20 [1.13-1.27] | 1.20 [1.13-1.26] | 1.15 [1.08-1.22] | 1.15 [1.08-1.23] | 1.18 [1.02-1.36] | 1.15 [1.08-1.22] | 3,404 | 3,053 | 2,603 | 521 | 3,043 |
| 288.11 | 1.23 [1.19-1.27] | 1.24 [1.20-1.28] | 1.23 [1.19-1.28] | 1.14 [1.04-1.24] | 1.18 [1.14-1.22] | 1.20 [1.16-1.24] | 1.19 [1.15-1.23] | 1.19 [1.15-1.23] | 1.11 [1.02-1.22] | 1.17 [1.13-1.21] | 9,777 | 8,724 | 7,840 | 1,334 | 8,508 |
| 288.2 | 1.43 [1.34-1.53] | 1.44 [1.35-1.55] | 1.40 [1.30-1.51] | 1.41 [1.18-1.68] | 1.35 [1.26-1.45] | 1.31 [1.22-1.40] | 1.27 [1.18-1.37] | 1.25 [1.16-1.35] | 1.31 [1.10-1.58] | 1.26 [1.17-1.35] | 2,421 | 2,176 | 1,866 | 358 | 2,199 |
| 288.3 | 2.09 [1.82-2.40] | 2.13 [1.84-2.46] | 2.08 [1.77-2.45] | 2.02 [1.49-2.76] | 1.95 [1.69-2.25] | 1.82 [1.58-2.10] | 1.75 [1.50-2.05] | 1.77 [1.49-2.09] | 1.71 [1.23-2.36] | 1.72 [1.49-2.00] | 636 | 546 | 440 | 138 | 573 |
| 289 | 1.25 [1.18-1.31] | 1.25 [1.18-1.31] | 1.25 [1.18-1.32] | 1.19 [0.91-1.56] | 1.18 [1.12-1.25] | 1.19 [1.13-1.26] | 1.14 [1.08-1.21] | 1.15 [1.09-1.22] | 1.13 [0.85-1.51] | 1.16 [1.09-1.22] | 3,575 | 3,494 | 3,277 | 140 | 3,174 |
| 289.3 | 1.34 [1.26-1.43] | 1.35 [1.26-1.44] | 1.37 [1.26-1.48] | 1.22 [1.06-1.41] | 1.22 [1.14-1.30] | 1.25 [1.17-1.33] | 1.22 [1.14-1.31] | 1.22 [1.13-1.32] | 1.19 [1.03-1.38] | 1.16 [1.09-1.24] | 2,606 | 2,270 | 1,648 | 551 | 2,423 |
| 289.4 | 1.33 [1.30-1.36] | 1.35 [1.32-1.39] | 1.33 [1.28-1.37] | 1.25 [1.21-1.30] | 1.25 [1.22-1.29] | 1.29 [1.26-1.32] | 1.26 [1.23-1.30] | 1.25 [1.21-1.29] | 1.22 [1.17-1.26] | 1.23 [1.20-1.27] | 18,440 | 11,712 | 9,577 | 7,528 | 15,745 |
| 289.5 | 1.25 [1.17-1.34] | 1.24 [1.16-1.32] | 1.26 [1.17-1.35] | 0.92 [0.72-1.18] | 1.16 [1.08-1.24] | 1.20 [1.12-1.28] | 1.15 [1.07-1.23] | 1.16 [1.08-1.25] | 0.95 [0.74-1.22] | 1.14 [1.07-1.23] | 2,222 | 2,116 | 1,898 | 169 | 2,030 |
| 289.8 | 1.16 [1.06-1.27] | 1.13 [1.04-1.23] | 1.13 [1.03-1.24] | | 1.10 [1.00-1.21] | 1.12 [1.02-1.22] | 1.03 [0.94-1.12] | 1.03 [0.94-1.13] | | 1.09 [1.00-1.20] | 1,244 | 1,232 | 1,141 | NA | 1,125 |
| 289.9 | 1.27 [1.07-1.52] | 1.17 [0.98-1.41] | 1.16 [0.95-1.41] | | 1.14 [0.94-1.37] | 1.21 [1.01-1.45] | 1.11 [0.92-1.34] | 1.08 [0.88-1.32] | | 1.12 [0.92-1.35] | 313 | 282 | 246 | NA | 273 |
| mental disorders | | | | | | | | | | | | | | | |
| 290 | 1.17 [1.14-1.19] | 1.17 [1.15-1.19] | 1.17 [1.14-1.19] | | 1.13 [1.10-1.15] | 1.14 [1.12-1.17] | 1.11 [1.09-1.14] | 1.11 [1.08-1.13] | | 1.12 [1.10-1.14] | 26,809 | 26,795 | 26,750 | NA | 24,968 |
| 290.1 | 1.07 [1.05-1.09] | 1.06 [1.04-1.08] | 1.06 [1.05-1.08] | | 1.04 [1.02-1.06] | 1.06 [1.04-1.08] | 1.04 [1.02-1.06] | 1.04 [1.02-1.06] | | 1.04 [1.02-1.06] | 34,205 | 34,185 | 34,155 | NA | 31,222 |
| 290.11 | 1.07 [1.04-1.10] | 1.07 [1.04-1.10] | 1.08 [1.05-1.11] | | 1.06 [1.03-1.09] | 1.06 [1.03-1.09] | 1.05 [1.02-1.08] | 1.06 [1.03-1.09] | | 1.06 [1.02-1.09] | 13,779 | 13,770 | 13,766 | NA | 12,505 |
| 290.12 | 1.01 [0.91-1.11] | 1.01 [0.91-1.11] | 1.00 [0.91-1.10] | | 0.95 [0.86-1.05] | 1.03 [0.93-1.13] | 1.00 [0.91-1.10] | 0.99 [0.90-1.09] | | 0.98 [0.89-1.08] | 1,100 | 1,101 | 1,101 | NA | 1,046 |
| 290.16 | 1.10 [1.07-1.13] | 1.10 [1.06-1.13] | 1.09 [1.06-1.12] | | 1.06 [1.03-1.10] | 1.08 [1.05-1.12] | 1.05 [1.02-1.08] | 1.04 [1.00-1.07] | | 1.06 [1.03-1.09] | 11,780 | 11,779 | 11,773 | NA | 11,049 |
| 290.2 | 1.18 [1.15-1.20] | 1.18 [1.15-1.20] | 1.18 [1.15-1.20] | 0.98 [0.76-1.27] | 1.14 [1.12-1.17] | 1.15 [1.13-1.18] | 1.11 [1.09-1.13] | 1.11 [1.08-1.13] | 0.98 [0.75-1.27] | 1.14 [1.11-1.16] | 25,766 | 25,664 | 25,415 | 166 | 24,422 |
| 290.3 | 1.08 [1.00-1.17] | 1.09 [1.00-1.18] | 1.26 [1.11-1.42] | 0.93 [0.81-1.06] | 0.97 [0.90-1.06] | 1.01 [0.93-1.09] | 0.90 [0.82-0.97] | 0.99 [0.86-1.13] | 0.96 [0.83-1.11] | 0.99 [0.91-1.08] | 1,605 | 1,475 | 563 | 539 | 1,395 |
| 291.1 | 1.16 [1.05-1.29] | 1.12 [1.00-1.24] | 1.28 [1.11-1.47] | 0.94 [0.75-1.18] | 1.07 [0.95-1.20] | 1.13 [1.02-1.26] | 1.04 [0.94-1.16] | 1.19 [1.04-1.38] | 0.93 [0.74-1.18] | 1.08 [0.96-1.21] | 923 | 842 | 497 | 197 | 725 |
| 291.4 | 1.17 [1.11-1.23] | 1.17 [1.11-1.23] | 1.18 [1.12-1.24] | 1.03 [0.77-1.38] | 1.10 [1.04-1.15] | 1.13 [1.08-1.19] | 1.10 [1.05-1.16] | 1.11 [1.06-1.17] | 1.11 [0.82-1.51] | 1.09 [1.04-1.15] | 4,526 | 4,455 | 4,306 | 120 | 4,215 |
| 291.8 | 1.25 [1.20-1.31] | 1.27 [1.21-1.33] | 1.28 [1.22-1.35] | 1.08 [0.96-1.21] | 1.18 [1.13-1.24] | 1.18 [1.12-1.23] | 1.13 [1.07-1.19] | 1.15 [1.09-1.21] | 1.03 [0.92-1.17] | 1.14 [1.09-1.20] | 5,157 | 4,582 | 4,131 | 734 | 4,836 |
| 292 | 1.17 [1.16-1.19] | 1.18 [1.16-1.20] | 1.17 [1.15-1.19] | 1.09 [1.01-1.18] | 1.14 [1.12-1.16] | 1.14 [1.12-1.16] | 1.11 [1.09-1.13] | 1.11 [1.09-1.13] | 1.04 [0.96-1.13] | 1.13 [1.11-1.15] | 41,977 | 40,673 | 39,528 | 1,764 | 38,419 |
| 292.1 | 1.17 [1.13-1.20] | 1.17 [1.13-1.20] | 1.16 [1.13-1.20] | 1.09 [0.97-1.22] | 1.11 [1.08-1.15] | 1.13 [1.09-1.16] | 1.08 [1.05-1.12] | 1.08 [1.04-1.11] | 1.04 [0.92-1.18] | 1.10 [1.06-1.13] | 11,628 | 11,115 | 10,439 | 719 | 10,584 |
| 292.12 | 1.29 [1.12-1.48] | 1.32 [1.11-1.56] | 1.31 [1.03-1.66] | 1.16 [0.96-1.40] | 1.23 [1.06-1.42] | 1.15 [1.00-1.33] | 1.10 [0.92-1.32] | 1.13 [0.88-1.45] | 1.09 [0.90-1.32] | 1.16 [1.00-1.35] | 570 | 352 | 175 | 309 | 512 |
| 292.2 | 1.15 [1.09-1.21] | 1.14 [1.08-1.21] | 1.16 [1.10-1.23] | | 1.10 [1.04-1.16] | 1.12 [1.06-1.18] | 1.09 [1.03-1.15] | 1.10 [1.04-1.17] | | 1.09 [1.03-1.15] | 3,771 | 3,754 | 3,715 | NA | 3,562 |
| 292.3 | 1.17 [1.12-1.23] | 1.18 [1.12-1.24] | 1.19 [1.13-1.26] | 1.01 [0.88-1.15] | 1.12 [1.07-1.18] | 1.12 [1.07-1.18] | 1.10 [1.04-1.16] | 1.11 [1.05-1.17] | 0.96 [0.83-1.10] | 1.10 [1.04-1.15] | 4,779 | 4,370 | 3,995 | 582 | 4,269 |
| 292.4 | 1.17 [1.15-1.19] | 1.17 [1.15-1.19] | 1.17 [1.14-1.19] | 1.14 [1.01-1.29] | 1.14 [1.12-1.16] | 1.14 [1.12-1.16] | 1.11 [1.09-1.13] | 1.10 [1.08-1.12] | 1.10 [0.97-1.25] | 1.13 [1.11-1.15] | 32,842 | 32,350 | 31,720 | 712 | 30,123 |
| 292.6 | 1.20 [1.14-1.27] | 1.22 [1.16-1.29] | 1.25 [1.17-1.32] | 1.10 [0.99-1.23] | 1.14 [1.08-1.21] | 1.14 [1.08-1.20] | 1.11 [1.05-1.17] | 1.14 [1.07-1.21] | 1.07 [0.96-1.20] | 1.12 [1.06-1.18] | 4,107 | 3,484 | 2,954 | 864 | 3,714 |
| 293 | 1.37 [1.32-1.42] | 1.46 [1.37-1.55] | 1.46 [1.36-1.57] | 1.33 [1.27-1.39] | 1.31 [1.25-1.36] | 1.28 [1.23-1.33] | 1.24 [1.16-1.32] | 1.21 [1.12-1.31] | 1.26 [1.20-1.32] | 1.23 [1.18-1.28] | 7,400 | 2,382 | 1,762 | 5,150 | 6,239 |
| 293.1 | 1.39 [1.33-1.46] | 1.33 [1.26-1.41] | 1.29 [1.21-1.37] | 1.48 [1.37-1.60] | 1.32 [1.26-1.39] | 1.30 [1.24-1.36] | 1.20 [1.13-1.26] | 1.18 [1.10-1.25] | 1.41 [1.31-1.53] | 1.26 [1.20-1.32] | 5,012 | 3,433 | 2,592 | 1,903 | 4,347 |
| 295 | 1.30 [1.02-1.65] | 1.37 [1.08-1.75] | 1.40 [1.06-1.85] | | 1.18 [0.92-1.51] | 1.18 [0.92-1.51] | 1.18 [0.92-1.52] | 1.23 [0.93-1.64] | | 1.12 [0.87-1.44] | 180 | 173 | 130 | NA | 166 |
| 295.1 | 1.27 [1.21-1.34] | 1.26 [1.20-1.33] | 1.31 [1.23-1.39] | 1.01 [0.86-1.20] | 1.14 [1.07-1.20] | 1.25 [1.19-1.32] | 1.16 [1.10-1.23] | 1.25 [1.17-1.33] | 1.05 [0.88-1.25] | 1.16 [1.09-1.23] | 3,632 | 3,578 | 2,596 | 355 | 2,819 |
| 295.2 | 1.28 [1.18-1.38] | 1.27 [1.17-1.37] | 1.31 [1.20-1.43] | 0.96 [0.71-1.30] | 1.18 [1.09-1.29] | 1.22 [1.13-1.33] | 1.16 [1.07-1.26] | 1.20 [1.10-1.31] | 0.95 [0.69-1.29] | 1.18 [1.08-1.28] | 1,605 | 1,558 | 1,321 | 117 | 1,411 |
| 295.3 | 1.23 [1.15-1.32] | 1.28 [1.19-1.38] | 1.40 [1.28-1.53] | 0.91 [0.77-1.07] | 1.12 [1.04-1.21] | 1.17 [1.09-1.26] | 1.14 [1.06-1.23] | 1.25 [1.14-1.37] | 0.92 [0.77-1.09] | 1.11 [1.03-1.20] | 1,942 | 1,826 | 1,197 | 386 | 1,665 |
| 296 | 1.36 [1.23-1.50] | 1.37 [1.24-1.52] | 1.38 [1.22-1.55] | 1.18 [0.93-1.48] | 1.21 [1.09-1.34] | 1.21 [1.09-1.34] | 1.13 [1.01-1.26] | 1.16 [1.02-1.31] | 1.09 [0.85-1.39] | 1.13 [1.01-1.25] | 1,042 | 934 | 691 | 202 | 923 |
| 296.1 | 1.30 [1.24-1.36] | 1.32 [1.26-1.38] | 1.34 [1.26-1.42] | 1.07 [0.94-1.22] | 1.16 [1.11-1.22] | 1.18 [1.13-1.24] | 1.08 [1.03-1.14] | 1.14 [1.07-1.21] | 1.07 [0.94-1.23] | 1.12 [1.07-1.18] | 4,635 | 4,532 | 2,958 | 616 | 4,028 |
| 296.2 | 1.37 [1.35-1.38] | 1.38 [1.36-1.40] | 1.39 [1.37-1.41] | 1.14 [1.10-1.17] | 1.26 [1.25-1.28] | 1.23 [1.22-1.25] | 1.13 [1.12-1.14] | 1.15 [1.14-1.17] | 1.13 [1.09-1.17] | 1.20 [1.18-1.21] | 83,464 | 80,758 | 58,420 | 11,105 | 73,790 |
| 296.22 | 1.24 [1.13-1.35] | 1.24 [1.13-1.36] | 1.22 [1.10-1.35] | | 1.13 [1.02-1.24] | 1.14 [1.03-1.25] | 1.04 [0.95-1.15] | 1.05 [0.95-1.17] | | 1.11 [1.01-1.22] | 1,145 | 1,119 | 882 | NA | 1,000 |
| 297 | 0.98 [0.83-1.16] | 1.08 [0.89-1.31] | 1.07 [0.81-1.41] | 0.89 [0.69-1.14] | 0.91 [0.75-1.09] | 0.92 [0.77-1.09] | 0.95 [0.77-1.16] | 0.96 [0.72-1.27] | 0.85 [0.66-1.10] | 0.86 [0.71-1.04] | 346 | 239 | 123 | 155 | 264 |
| 297.2 | 1.08 [1.02-1.14] | 1.09 [1.02-1.17] | 1.27 [1.13-1.41] | 1.00 [0.93-1.07] | 0.99 [0.93-1.05] | 1.01 [0.96-1.07] | 0.92 [0.85-0.99] | 1.05 [0.94-1.18] | 0.98 [0.91-1.06] | 0.97 [0.91-1.03] | 3,237 | 1,886 | 767 | 1,924 | 2,653 |
| 300 | 1.27 [1.20-1.34] | 1.28 [1.20-1.36] | 1.29 [1.20-1.38] | 1.20 [1.07-1.35] | 1.16 [1.10-1.23] | 1.18 [1.11-1.25] | 1.11 [1.05-1.19] | 1.12 [1.04-1.21] | 1.18 [1.05-1.33] | 1.13 [1.06-1.20] | 3,259 | 2,730 | 1,951 | 846 | 2,914 |
| 300.1 | 1.34 [1.33-1.36] | 1.36 [1.34-1.37] | 1.36 [1.34-1.39] | 1.16 [1.13-1.20] | 1.25 [1.23-1.27] | 1.21 [1.19-1.22] | 1.12 [1.10-1.13] | 1.14 [1.12-1.15] | 1.14 [1.10-1.17] | 1.17 [1.16-1.19] | 66,705 | 63,188 | 43,294 | 11,799 | 59,643 |
| 300.11 | 1.37 [1.26-1.49] | 1.37 [1.26-1.49] | 1.32 [1.18-1.47] | 1.31 [1.08-1.59] | 1.22 [1.12-1.33] | 1.22 [1.12-1.34] | 1.12 [1.02-1.23] | 1.09 [0.97-1.22] | 1.32 [1.08-1.62] | 1.14 [1.04-1.25] | 1,455 | 1,347 | 823 | 315 | 1,256 |
| 300.12 | 1.29 [1.25-1.34] | 1.30 [1.25-1.34] | 1.30 [1.24-1.36] | 1.13 [1.03-1.23] | 1.22 [1.17-1.26] | 1.17 [1.13-1.21] | 1.08 [1.04-1.12] | 1.08 [1.03-1.13] | 1.11 [1.01-1.22] | 1.14 [1.10-1.19] | 7,923 | 7,372 | 5,281 | 1,355 | 7,048 |
| 300.13 | 1.26 [1.21-1.30] | 1.30 [1.25-1.35] | 1.30 [1.24-1.36] | 1.13 [1.05-1.21] | 1.19 [1.14-1.24] | 1.16 [1.12-1.21] | 1.13 [1.09-1.18] | 1.14 [1.08-1.19] | 1.08 [1.01-1.16] | 1.13 [1.09-1.18] | 7,880 | 6,570 | 4,367 | 2,286 | 6,833 |
| 300.3 | 1.64 [1.52-1.76] | 1.67 [1.54-1.80] | 1.84 [1.65-2.06] | 1.25 [1.09-1.43] | 1.47 [1.36-1.58] | 1.43 [1.32-1.54] | 1.33 [1.22-1.44] | 1.53 [1.37-1.72] | 1.15 [1.00-1.32] | 1.34 [1.24-1.46] | 2,087 | 1,795 | 888 | 635 | 1,802 |
| 300.4 | 1.52 [1.26-1.83] | 1.50 [1.25-1.82] | 1.56 [1.27-1.91] | | 1.28 [1.06-1.55] | 1.32 [1.09-1.60] | 1.21 [0.99-1.48] | 1.29 [1.04-1.59] | | 1.16 [0.95-1.42] | 301 | 290 | 242 | NA | 262 |
| 300.8 | 1.22 [1.06-1.40] | 1.21 [1.05-1.40] | 1.36 [1.13-1.64] | 1.11 [0.82-1.50] | 1.15 [0.99-1.34] | 1.13 [0.98-1.31] | 1.05 [0.90-1.22] | 1.17 [0.96-1.42] | 1.07 [0.78-1.48] | 1.11 [0.95-1.30] | 495 | 450 | 278 | 118 | 412 |
| 300.9 | 1.29 [1.21-1.37] | 1.31 [1.23-1.40] | 1.43 [1.31-1.57] | 0.98 [0.87-1.12] | 1.18 [1.10-1.26] | 1.09 [1.02-1.17] | 0.98 [0.92-1.05] | 1.08 [0.98-1.19] | 0.96 [0.84-1.09] | 1.09 [1.02-1.17] | 2,542 | 2,350 | 1,193 | 641 | 2,345 |
| 301 | 1.38 [1.29-1.48] | 1.41 [1.32-1.52] | 1.57 [1.43-1.72] | 1.03 [0.88-1.19] | 1.24 [1.15-1.33] | 1.19 [1.11-1.28] | 1.09 [1.01-1.17] | 1.24 [1.12-1.37] | 1.03 [0.88-1.20] | 1.16 [1.08-1.25] | 2,122 | 2,002 | 1,095 | 450 | 1,961 |
| 301.1 | 1.52 [1.18-1.97] | 1.51 [1.16-1.95] | | | 1.38 [1.06-1.80] | 1.38 [1.06-1.80] | 1.28 [0.97-1.67] | | | 1.32 [1.01-1.74] | 157 | 147 | NA | NA | 138 |
| 301.2 | 1.30 [1.22-1.39] | 1.36 [1.27-1.45] | 1.66 [1.48-1.86] | 1.00 [0.89-1.13] | 1.17 [1.09-1.25] | 1.09 [1.01-1.17] | 0.98 [0.91-1.06] | 1.20 [1.06-1.36] | 0.99 [0.87-1.12] | 1.08 [1.01-1.16] | 2,267 | 2,190 | 734 | 715 | 2,100 |
| 302 | 1.49 [1.33-1.68] | 1.48 [1.32-1.67] | 1.48 [1.31-1.68] | | 1.33 [1.17-1.51] | 1.36 [1.21-1.54] | 1.26 [1.12-1.43] | 1.27 [1.12-1.45] | | 1.26 [1.11-1.43] | 707 | 698 | 616 | NA | 598 |
| 303.1 | 1.40 [1.26-1.55] | 1.42 [1.28-1.58] | 1.45 [1.26-1.66] | 1.26 [1.03-1.54] | 1.22 [1.10-1.36] | 1.21 [1.08-1.34] | 1.11 [0.99-1.24] | 1.14 [0.99-1.32] | 1.19 [0.96-1.46] | 1.11 [1.00-1.24] | 1,022 | 914 | 529 | 282 | 934 |
| 303.3 | 1.41 [1.27-1.58] | 1.34 [1.20-1.50] | 1.32 [1.17-1.50] | 1.16 [0.86-1.56] | 1.26 [1.12-1.41] | 1.26 [1.12-1.41] | 1.13 [1.01-1.27] | 1.14 [1.00-1.30] | 1.02 [0.75-1.39] | 1.15 [1.02-1.29] | 848 | 773 | 600 | 119 | 748 |
| 303.4 | 1.44 [1.34-1.55] | 1.41 [1.30-1.53] | 1.40 [1.27-1.54] | 1.28 [1.08-1.51] | 1.28 [1.18-1.38] | 1.26 [1.16-1.36] | 1.13 [1.04-1.23] | 1.15 [1.05-1.28] | 1.19 [1.00-1.41] | 1.16 [1.07-1.26] | 1,793 | 1,582 | 1,064 | 401 | 1,624 |
| 304 | 1.21 [1.10-1.33] | 1.23 [1.11-1.35] | 1.26 [1.11-1.42] | 0.99 [0.80-1.22] | 1.11 [1.01-1.23] | 1.12 [1.02-1.24] | 1.04 [0.94-1.15] | 1.06 [0.93-1.20] | 1.04 [0.83-1.29] | 1.09 [0.99-1.21] | 1,141 | 1,048 | 647 | 224 | 965 |
| 305.2 | 1.29 [1.20-1.38] | 1.29 [1.18-1.41] | 1.41 [1.22-1.64] | 1.15 [1.04-1.26] | 1.20 [1.11-1.29] | 1.17 [1.09-1.26] | 1.08 [0.98-1.18] | 1.19 [1.02-1.39] | 1.11 [1.01-1.22] | 1.14 [1.05-1.23] | 2,118 | 1,258 | 443 | 1,249 | 1,770 |
| 305.21 | 1.22 [1.09-1.36] | 1.25 [1.08-1.45] | 1.40 [1.10-1.79] | 1.09 [0.95-1.26] | 1.13 [1.00-1.27] | 1.14 [1.02-1.28] | 1.08 [0.93-1.26] | 1.21 [0.94-1.56] | 1.06 [0.92-1.23] | 1.09 [0.96-1.24] | 847 | 460 | 159 | 533 | 682 |
| 306 | 1.26 [1.25-1.28] | 1.27 [1.26-1.28] | 1.28 [1.27-1.29] | 1.04 [1.01-1.07] | 1.18 [1.17-1.19] | 1.20 [1.19-1.21] | 1.14 [1.13-1.15] | 1.16 [1.15-1.17] | 1.04 [1.01-1.08] | 1.16 [1.15-1.17] | 150,754 | 149,919 | 121,961 | 10,438 | 132,415 |
| 306.9 | 1.30 [1.19-1.41] | 1.28 [1.17-1.39] | 1.40 [1.26-1.57] | 1.13 [0.94-1.37] | 1.20 [1.10-1.31] | 1.19 [1.09-1.29] | 1.11 [1.02-1.22] | 1.21 [1.08-1.36] | 1.10 [0.91-1.34] | 1.12 [1.03-1.23] | 1,410 | 1,263 | 803 | 305 | 1,255 |
| 312 | 1.24 [1.11-1.39] | 1.14 [0.90-1.44] | 1.10 [0.82-1.46] | 1.27 [1.12-1.43] | 1.15 [1.02-1.30] | 1.17 [1.05-1.31] | 1.04 [0.81-1.32] | 1.02 [0.76-1.37] | 1.22 [1.07-1.38] | 1.11 [0.98-1.25] | 867 | 174 | 113 | 730 | 754 |
| 313 | 1.25 [1.09-1.44] | 1.33 [1.00-1.77] | | 1.21 [1.05-1.41] | 1.13 [0.97-1.32] | 1.14 [0.99-1.32] | 1.11 [0.82-1.50] | | 1.16 [1.00-1.35] | 1.06 [0.91-1.24] | 559 | 115 | NA | 511 | 481 |
| 313.1 | 1.22 [1.15-1.28] | 1.23 [1.12-1.35] | 1.75 [1.34-2.29] | 1.16 [1.09-1.22] | 1.14 [1.08-1.21] | 1.12 [1.06-1.19] | 1.09 [0.99-1.21] | 1.26 [0.94-1.68] | 1.12 [1.06-1.19] | 1.11 [1.04-1.17] | 3,883 | 1,020 | 139 | 3,444 | 3,393 |
| 313.2 | 1.36 [1.19-1.57] | 1.40 [1.11-1.76] | | 1.29 [1.10-1.52] | 1.24 [1.07-1.44] | 1.23 [1.06-1.42] | 1.16 [0.91-1.48] | | 1.21 [1.03-1.42] | 1.17 [1.00-1.36] | 581 | 187 | NA | 447 | 513 |
| 313.3 | 1.32 [1.28-1.38] | 1.54 [1.44-1.65] | 2.15 [1.90-2.44] | 1.22 [1.17-1.27] | 1.23 [1.18-1.28] | 1.18 [1.14-1.23] | 1.34 [1.24-1.44] | 1.73 [1.51-1.97] | 1.15 [1.11-1.20] | 1.14 [1.09-1.19] | 7,896 | 2,095 | 695 | 6,653 | 6,860 |
| 315 | 1.14 [1.10-1.19] | 1.22 [1.16-1.28] | 1.23 [1.17-1.30] | 1.01 [0.95-1.08] | 1.08 [1.03-1.12] | 1.10 [1.06-1.15] | 1.12 [1.07-1.18] | 1.14 [1.08-1.21] | 0.97 [0.91-1.04] | 1.06 [1.01-1.10] | 6,293 | 4,238 | 3,699 | 2,319 | 5,799 |
| 315.1 | 1.56 [1.50-1.62] | 1.92 [1.83-2.00] | 2.15 [2.03-2.27] | 1.13 [1.07-1.19] | 1.36 [1.31-1.42] | 1.44 [1.39-1.50] | 1.79 [1.71-1.88] | 1.99 [1.88-2.11] | 1.09 [1.03-1.15] | 1.29 [1.24-1.35] | 8,341 | 5,297 | 3,441 | 3,817 | 6,832 |
| 315.2 | 1.22 [1.15-1.29] | 1.38 [1.22-1.56] | 1.41 [1.24-1.61] | 1.16 [1.09-1.24] | 1.14 [1.08-1.22] | 1.16 [1.10-1.24] | 1.29 [1.13-1.46] | 1.31 [1.14-1.49] | 1.11 [1.04-1.19] | 1.12 [1.05-1.19] | 2,868 | 666 | 603 | 2,234 | 2,631 |
| 315.3 | 1.58 [1.49-1.67] | 1.90 [1.78-2.02] | 2.15 [1.98-2.33] | 1.09 [1.00-1.18] | 1.37 [1.29-1.45] | 1.45 [1.37-1.53] | 1.74 [1.63-1.87] | 1.94 [1.78-2.11] | 1.05 [0.97-1.15] | 1.30 [1.22-1.38] | 3,630 | 2,534 | 1,641 | 1,482 | 3,146 |
| 316 | 1.19 [1.17-1.21] | 1.21 [1.19-1.24] | 1.27 [1.24-1.29] | 0.98 [0.94-1.03] | 1.11 [1.09-1.14] | 1.15 [1.13-1.17] | 1.09 [1.07-1.11] | 1.15 [1.12-1.18] | 0.98 [0.94-1.02] | 1.12 [1.10-1.15] | 27,927 | 26,131 | 17,706 | 5,543 | 22,075 |
| 316.1 | 1.24 [1.05-1.47] | 1.22 [1.02-1.45] | 1.24 [1.03-1.50] | | 1.15 [0.96-1.37] | 1.21 [1.01-1.44] | 1.18 [0.98-1.42] | 1.22 [1.01-1.47] | | 1.15 [0.96-1.37] | 330 | 303 | 285 | NA | 304 |
| 317 | 1.20 [1.17-1.22] | 1.22 [1.19-1.24] | 1.26 [1.23-1.29] | 0.99 [0.95-1.04] | 1.13 [1.11-1.15] | 1.16 [1.14-1.18] | 1.10 [1.08-1.12] | 1.15 [1.12-1.18] | 0.99 [0.94-1.04] | 1.14 [1.12-1.17] | 25,955 | 24,196 | 16,882 | 4,875 | 20,728 |
| 317.1 | 1.22 [1.19-1.24] | 1.22 [1.20-1.25] | 1.23 [1.20-1.26] | 1.02 [0.96-1.09] | 1.13 [1.11-1.16] | 1.17 [1.15-1.19] | 1.11 [1.09-1.13] | 1.12 [1.10-1.15] | 1.01 [0.95-1.08] | 1.13 [1.10-1.15] | 24,191 | 23,744 | 18,465 | 2,336 | 20,394 |
| 317.11 | 1.44 [1.38-1.51] | 1.46 [1.40-1.52] | 1.46 [1.40-1.53] | | 1.35 [1.29-1.41] | 1.37 [1.31-1.43] | 1.29 [1.23-1.35] | 1.28 [1.23-1.34] | | 1.34 [1.28-1.41] | 5,266 | 5,262 | 4,799 | NA | 4,536 |
| 318 | 1.13 [1.12-1.14] | 1.15 [1.14-1.16] | 1.15 [1.14-1.16] | 0.99 [0.97-1.02] | 1.06 [1.05-1.07] | 1.12 [1.11-1.13] | 1.03 [1.02-1.04] | 1.06 [1.04-1.07] | 1.00 [0.98-1.02] | 1.09 [1.08-1.10] | 112,963 | 109,604 | 65,828 | 22,575 | 86,891 |
| neurological | | | | | | | | | | | | | | | |
| 320 | 1.24 [1.15-1.33] | 1.28 [1.18-1.39] | 1.25 [1.12-1.41] | 1.21 [1.08-1.35] | 1.11 [1.03-1.20] | 1.16 [1.08-1.25] | 1.14 [1.05-1.25] | 1.13 [1.00-1.27] | 1.18 [1.05-1.33] | 1.06 [0.98-1.15] | 1,921 | 1,447 | 691 | 782 | 1,575 |
| 323 | 1.23 [1.14-1.33] | 1.27 [1.17-1.39] | 1.26 [1.15-1.39] | 1.02 [0.87-1.20] | 1.16 [1.07-1.26] | 1.18 [1.09-1.28] | 1.17 [1.07-1.29] | 1.16 [1.05-1.28] | 1.00 [0.85-1.18] | 1.13 [1.04-1.23] | 1,586 | 1,255 | 1,049 | 404 | 1,359 |
| 323.2 | 1.16 [0.92-1.46] | 1.23 [0.96-1.58] | 1.32 [0.99-1.76] | | 1.13 [0.88-1.46] | 1.13 [0.89-1.43] | 1.15 [0.89-1.49] | 1.25 [0.93-1.67] | | 1.12 [0.86-1.45] | 168 | 147 | 109 | NA | 139 |
| 323.8 | 1.11 [0.85-1.46] | | | | 0.94 [0.71-1.26] | 1.07 [0.81-1.41] | | | | 0.94 [0.70-1.26] | 134 | NA | NA | NA | 106 |
| 324 | 1.26 [1.24-1.28] | 1.27 [1.25-1.29] | 1.26 [1.24-1.28] | 1.18 [1.12-1.25] | 1.18 [1.16-1.20] | 1.18 [1.16-1.20] | 1.14 [1.13-1.16] | 1.14 [1.12-1.16] | 1.14 [1.07-1.20] | 1.13 [1.11-1.15] | 44,811 | 42,291 | 39,029 | 3,658 | 41,695 |
| 325 | 1.16 [0.95-1.42] | 1.32 [1.06-1.65] | 1.14 [0.86-1.51] | | 1.29 [1.04-1.60] | 1.12 [0.92-1.38] | 1.26 [1.00-1.58] | 1.06 [0.79-1.42] | | 1.29 [1.03-1.61] | 239 | 199 | 116 | NA | 206 |
| 327 | 1.48 [1.45-1.51] | 1.58 [1.54-1.62] | 1.58 [1.54-1.62] | 1.31 [1.27-1.36] | 1.38 [1.35-1.41] | 1.32 [1.29-1.35] | 1.27 [1.24-1.31] | 1.27 [1.24-1.31] | 1.25 [1.21-1.30] | 1.27 [1.24-1.29] | 23,995 | 15,545 | 13,302 | 9,075 | 21,559 |
| 327.3 | 1.51 [1.48-1.55] | 1.63 [1.59-1.68] | 1.63 [1.58-1.67] | 1.32 [1.27-1.37] | 1.40 [1.37-1.44] | 1.35 [1.32-1.38] | 1.31 [1.27-1.35] | 1.30 [1.26-1.34] | 1.26 [1.22-1.31] | 1.29 [1.26-1.32] | 20,679 | 13,191 | 11,662 | 7,845 | 18,668 |
| 327.4 | 1.25 [0.97-1.61] | 1.24 [0.93-1.65] | | | 1.21 [0.93-1.58] | 1.12 [0.86-1.45] | 1.01 [0.75-1.36] | | | 1.13 [0.86-1.48] | 153 | 123 | NA | NA | 143 |
| 327.41 | 1.34 [1.23-1.46] | 1.32 [1.21-1.44] | 1.36 [1.23-1.51] | 1.26 [1.03-1.55] | 1.24 [1.14-1.36] | 1.18 [1.08-1.29] | 1.09 [0.99-1.20] | 1.14 [1.03-1.27] | 1.19 [0.96-1.47] | 1.16 [1.05-1.26] | 1,414 | 1,254 | 999 | 268 | 1,302 |
| 327.5 | 1.50 [1.22-1.84] | 1.90 [1.45-2.51] | | 1.25 [0.93-1.66] | 1.37 [1.11-1.71] | 1.37 [1.11-1.69] | 1.61 [1.21-2.15] | | 1.22 [0.91-1.64] | 1.28 [1.02-1.59] | 254 | 144 | NA | 130 | 222 |
| 327.6 | 1.33 [1.02-1.72] | 1.52 [1.11-2.06] | | | 1.28 [0.98-1.67] | 1.12 [0.85-1.47] | 1.14 [0.81-1.59] | | | 1.19 [0.90-1.56] | 157 | 106 | NA | NA | 144 |
| 331 | 1.20 [1.13-1.27] | 1.21 [1.14-1.29] | 1.22 [1.14-1.30] | 1.03 [0.83-1.28] | 1.14 [1.07-1.21] | 1.16 [1.09-1.23] | 1.13 [1.06-1.20] | 1.13 [1.06-1.21] | 1.03 [0.82-1.28] | 1.13 [1.06-1.21] | 2,687 | 2,498 | 2,370 | 212 | 2,445 |
| 331.1 | 1.17 [1.10-1.25] | 1.21 [1.14-1.30] | 1.18 [1.10-1.26] | 0.96 [0.83-1.13] | 1.14 [1.07-1.22] | 1.15 [1.08-1.23] | 1.17 [1.09-1.25] | 1.13 [1.05-1.22] | 0.97 [0.83-1.13] | 1.14 [1.06-1.22] | 2,544 | 2,204 | 1,937 | 412 | 2,170 |
| 331.9 | 1.16 [1.12-1.21] | 1.18 [1.14-1.22] | 1.16 [1.12-1.20] | | 1.13 [1.09-1.17] | 1.14 [1.10-1.18] | 1.12 [1.08-1.16] | 1.10 [1.06-1.14] | | 1.13 [1.09-1.17] | 8,113 | 8,032 | 7,940 | NA | 7,506 |
| 332 | 1.06 [1.02-1.09] | 1.05 [1.01-1.09] | 1.06 [1.02-1.10] | | 1.03 [0.99-1.07] | 1.05 [1.01-1.09] | 1.02 [0.99-1.06] | 1.03 [0.99-1.07] | | 1.03 [0.99-1.07] | 7,432 | 7,435 | 7,409 | NA | 6,407 |
| 333 | 1.28 [1.22-1.34] | 1.30 [1.24-1.36] | 1.28 [1.22-1.35] | 1.07 [0.91-1.27] | 1.20 [1.15-1.26] | 1.16 [1.11-1.22] | 1.12 [1.07-1.17] | 1.11 [1.06-1.17] | 1.04 [0.87-1.24] | 1.13 [1.07-1.18] | 4,924 | 4,668 | 4,350 | 353 | 4,614 |
| 333.1 | 1.18 [1.09-1.27] | 1.17 [1.09-1.26] | 1.15 [1.06-1.24] | | 1.12 [1.04-1.20] | 1.08 [1.00-1.16] | 1.03 [0.95-1.11] | 1.02 [0.94-1.10] | | 1.05 [0.97-1.14] | 1,851 | 1,814 | 1,743 | NA | 1,735 |
| 333.2 | 1.28 [1.12-1.45] | 1.32 [1.15-1.52] | 1.38 [1.19-1.61] | 1.09 [0.82-1.44] | 1.21 [1.06-1.38] | 1.19 [1.04-1.35] | 1.18 [1.02-1.36] | 1.23 [1.06-1.44] | 1.06 [0.80-1.41] | 1.16 [1.02-1.33] | 616 | 517 | 453 | 126 | 571 |
| 333.3 | 1.33 [1.09-1.61] | 1.32 [1.08-1.63] | 1.25 [1.01-1.55] | | 1.22 [0.99-1.52] | 1.31 [1.08-1.60] | 1.31 [1.06-1.61] | 1.24 [1.00-1.55] | | 1.23 [0.99-1.53] | 266 | 239 | 216 | NA | 218 |
| 333.4 | 1.12 [1.02-1.23] | 1.21 [1.09-1.35] | 1.21 [1.08-1.37] | 0.81 [0.67-0.97] | 1.02 [0.93-1.13] | 1.06 [0.96-1.17] | 1.10 [0.99-1.23] | 1.11 [0.99-1.25] | 0.77 [0.64-0.93] | 1.00 [0.90-1.10] | 1,090 | 844 | 708 | 296 | 994 |
| 333.8 | 1.17 [1.00-1.36] | 1.12 [0.96-1.30] | 1.08 [0.93-1.27] | | 1.02 [0.87-1.19] | 1.14 [0.97-1.33] | 1.07 [0.92-1.25] | 1.03 [0.88-1.21] | | 1.01 [0.86-1.18] | 428 | 418 | 405 | NA | 393 |
| 334 | 1.22 [1.17-1.28] | 1.24 [1.19-1.30] | 1.23 [1.18-1.29] | 1.10 [0.91-1.34] | 1.15 [1.10-1.20] | 1.16 [1.11-1.21] | 1.14 [1.09-1.19] | 1.13 [1.08-1.18] | 1.06 [0.87-1.30] | 1.12 [1.07-1.17] | 5,259 | 5,092 | 4,666 | 266 | 4,683 |
| 334.1 | 1.17 [1.01-1.36] | 1.25 [1.07-1.46] | 1.20 [1.02-1.43] | | 1.15 [0.99-1.34] | 1.13 [0.97-1.31] | 1.16 [0.99-1.36] | 1.10 [0.93-1.31] | | 1.14 [0.97-1.33] | 454 | 380 | 328 | NA | 400 |
| 334.2 | 1.08 [0.97-1.20] | 1.12 [1.00-1.24] | 1.10 [0.99-1.22] | | 1.06 [0.95-1.19] | 1.04 [0.93-1.15] | 1.07 [0.96-1.20] | 1.06 [0.95-1.18] | | 1.05 [0.94-1.18] | 857 | 824 | 796 | NA | 732 |
| 335 | 1.23 [1.15-1.31] | 1.22 [1.15-1.30] | 1.23 [1.14-1.32] | 1.14 [0.85-1.53] | 1.14 [1.06-1.22] | 1.16 [1.08-1.24] | 1.08 [1.01-1.15] | 1.09 [1.01-1.18] | 1.15 [0.85-1.55] | 1.12 [1.04-1.21] | 2,212 | 2,199 | 1,684 | 125 | 1,677 |
| 337 | 1.26 [1.15-1.40] | 1.23 [1.11-1.36] | 1.22 [1.08-1.36] | 1.01 [0.77-1.33] | 1.13 [1.02-1.25] | 1.17 [1.05-1.29] | 1.07 [0.97-1.19] | 1.08 [0.96-1.22] | 0.94 [0.71-1.25] | 1.06 [0.95-1.18] | 1,018 | 945 | 747 | 143 | 926 |
| 337.1 | 1.36 [1.11-1.67] | 1.28 [1.05-1.57] | 1.28 [1.04-1.57] | | 1.26 [1.02-1.56] | 1.26 [1.03-1.56] | 1.15 [0.93-1.42] | 1.13 [0.91-1.40] | | 1.20 [0.97-1.49] | 246 | 243 | 237 | NA | 222 |
| 338.1 | 1.40 [1.30-1.51] | 1.46 [1.35-1.58] | 1.46 [1.34-1.60] | 1.08 [0.89-1.31] | 1.28 [1.18-1.38] | 1.29 [1.19-1.39] | 1.27 [1.17-1.38] | 1.29 [1.18-1.42] | 1.02 [0.84-1.25] | 1.20 [1.11-1.30] | 1,800 | 1,608 | 1,278 | 284 | 1,630 |
| 338.2 | 1.46 [1.37-1.56] | 1.48 [1.39-1.58] | 1.46 [1.36-1.57] | 1.20 [0.97-1.47] | 1.32 [1.24-1.41] | 1.25 [1.16-1.33] | 1.15 [1.08-1.24] | 1.15 [1.07-1.25] | 1.10 [0.89-1.36] | 1.20 [1.12-1.28] | 2,496 | 2,367 | 1,855 | 261 | 2,412 |
| 339 | 1.28 [1.26-1.31] | 1.29 [1.27-1.32] | 1.32 [1.29-1.35] | 1.20 [1.17-1.24] | 1.19 [1.17-1.21] | 1.18 [1.16-1.20] | 1.13 [1.11-1.15] | 1.15 [1.13-1.18] | 1.15 [1.12-1.19] | 1.13 [1.11-1.14] | 39,639 | 32,105 | 20,115 | 11,841 | 34,211 |
| 340 | 1.32 [1.29-1.35] | 1.32 [1.29-1.36] | 1.33 [1.29-1.38] | 1.20 [1.14-1.27] | 1.22 [1.19-1.25] | 1.16 [1.13-1.19] | 1.08 [1.05-1.11] | 1.10 [1.06-1.13] | 1.14 [1.08-1.20] | 1.12 [1.09-1.15] | 16,716 | 14,898 | 8,783 | 4,008 | 14,746 |
| 340.1 | 1.29 [1.18-1.40] | 1.31 [1.19-1.43] | 1.22 [1.08-1.37] | 1.15 [0.98-1.37] | 1.19 [1.09-1.30] | 1.13 [1.04-1.24] | 1.06 [0.96-1.16] | 0.98 [0.86-1.11] | 1.11 [0.94-1.33] | 1.09 [0.99-1.19] | 1,393 | 1,201 | 647 | 386 | 1,256 |
| 341 | 1.27 [1.13-1.43] | 1.29 [1.14-1.46] | 1.36 [1.17-1.58] | 1.00 [0.75-1.34] | 1.20 [1.06-1.37] | 1.20 [1.07-1.36] | 1.17 [1.03-1.32] | 1.24 [1.06-1.45] | 0.98 [0.73-1.32] | 1.17 [1.02-1.33] | 691 | 624 | 413 | 120 | 575 |
| 342 | 1.17 [1.14-1.20] | 1.19 [1.15-1.23] | 1.17 [1.14-1.21] | 0.98 [0.86-1.13] | 1.12 [1.08-1.15] | 1.14 [1.11-1.18] | 1.12 [1.08-1.15] | 1.10 [1.07-1.13] | 0.98 [0.85-1.12] | 1.11 [1.08-1.15] | 11,632 | 11,192 | 10,789 | 541 | 10,542 |
| 343 | 1.01 [0.94-1.09] | 1.39 [1.25-1.55] | 1.66 [1.46-1.90] | 0.79 [0.72-0.87] | 0.92 [0.85-1.00] | 0.96 [0.89-1.04] | 1.33 [1.19-1.49] | 1.59 [1.38-1.82] | 0.77 [0.70-0.85] | 0.90 [0.83-0.97] | 1,750 | 801 | 552 | 1,019 | 1,431 |
| 344 | 1.19 [1.13-1.26] | 1.22 [1.15-1.29] | 1.22 [1.15-1.30] | 0.86 [0.73-1.01] | 1.09 [1.02-1.15] | 1.13 [1.07-1.20] | 1.12 [1.05-1.19] | 1.13 [1.06-1.21] | 0.84 [0.71-0.99] | 1.07 [1.00-1.13] | 2,968 | 2,727 | 2,329 | 349 | 2,621 |
| 345 | 1.31 [1.28-1.34] | 1.36 [1.33-1.40] | 1.38 [1.34-1.42] | 1.08 [1.02-1.13] | 1.19 [1.16-1.22] | 1.21 [1.18-1.24] | 1.19 [1.15-1.22] | 1.20 [1.16-1.23] | 1.06 [1.00-1.11] | 1.14 [1.11-1.17] | 17,304 | 14,194 | 11,163 | 4,102 | 14,182 |
| 345.1 | 1.26 [1.19-1.34] | 1.43 [1.33-1.54] | 1.39 [1.27-1.51] | 0.99 [0.90-1.09] | 1.13 [1.06-1.20] | 1.17 [1.10-1.24] | 1.24 [1.15-1.34] | 1.18 [1.09-1.29] | 0.98 [0.89-1.08] | 1.09 [1.03-1.17] | 2,752 | 1,824 | 1,416 | 1,074 | 2,522 |
| 345.11 | 1.25 [1.19-1.32] | 1.32 [1.24-1.41] | 1.40 [1.30-1.51] | 1.09 [1.00-1.18] | 1.14 [1.08-1.21] | 1.15 [1.09-1.21] | 1.13 [1.06-1.21] | 1.18 [1.09-1.28] | 1.07 [0.99-1.16] | 1.10 [1.04-1.16] | 3,745 | 2,481 | 1,757 | 1,554 | 3,343 |
| 345.12 | 1.46 [1.33-1.59] | 1.54 [1.39-1.70] | 1.45 [1.30-1.62] | 1.28 [1.09-1.50] | 1.30 [1.19-1.42] | 1.29 [1.18-1.41] | 1.25 [1.12-1.38] | 1.15 [1.02-1.30] | 1.27 [1.08-1.49] | 1.20 [1.09-1.31] | 1,394 | 1,060 | 796 | 422 | 1,261 |
| 345.3 | 1.14 [1.12-1.17] | 1.26 [1.23-1.30] | 1.27 [1.23-1.31] | 1.02 [0.99-1.05] | 1.08 [1.05-1.10] | 1.11 [1.08-1.13] | 1.15 [1.12-1.19] | 1.16 [1.13-1.20] | 1.01 [0.98-1.04] | 1.07 [1.04-1.09] | 21,453 | 12,161 | 9,715 | 10,228 | 18,506 |
| 346.1 | 1.10 [1.00-1.22] | 1.12 [1.01-1.25] | 1.12 [1.00-1.26] | 0.84 [0.66-1.07] | 1.05 [0.94-1.17] | 1.07 [0.96-1.19] | 1.05 [0.93-1.17] | 1.05 [0.93-1.19] | 0.86 [0.67-1.10] | 1.04 [0.93-1.16] | 930 | 802 | 685 | 168 | 831 |
| 346.2 | 1.19 [0.93-1.52] | | | | 1.14 [0.88-1.47] | 1.14 [0.89-1.47] | | | | 1.11 [0.86-1.44] | 164 | NA | NA | NA | 151 |
| 346.3 | 1.20 [0.89-1.62] | | | | | 1.14 [0.84-1.56] | | | | | 101 | NA | NA | NA | NA |

### Supplementary Table 6: phecode-mapped results (hazard ratios and events from all cohorts)

| | Hazard ratio (99% confidence interval) | | | | | | | | | | | | | | |
|---|---|---|---|---|---|---|---|---|---|---|---|---|---|---|---|
| | crude | | | | | adjusted | | | | | Events (in exposed) | | | | |
| Outcome | any age | 18+ | 40+ | <18 | hosp. | any age | 18+ | 40+ | <18 | hosp. | any age | 18+ | 40+ | <18 | hosp. |
| 347 | 1.43 [1.14-1.79] | 1.45 [1.13-1.86] | 1.47 [1.09-1.99] | | 1.42 [1.12-1.81] | 1.27 [1.00-1.61] | 1.20 [0.93-1.56] | 1.26 [0.92-1.72] | | 1.31 [1.02-1.67] | 209 | 161 | 106 | NA | 184 |
| 348 | 1.21 [1.10-1.32] | 1.29 [1.17-1.42] | 1.25 [1.13-1.39] | 0.94 [0.73-1.21] | 1.14 [1.04-1.26] | 1.16 [1.05-1.27] | 1.18 [1.07-1.30] | 1.15 [1.03-1.27] | 0.92 [0.71-1.19] | 1.14 [1.03-1.26] | 1,144 | 1,046 | 915 | 157 | 1,020 |
| 348.2 | 1.17 [1.09-1.25] | 1.16 [1.08-1.24] | 1.10 [1.01-1.20] | 1.16 [1.02-1.32] | 1.10 [1.02-1.18] | 1.10 [1.02-1.17] | 1.04 [0.97-1.12] | 1.02 [0.94-1.12] | 1.10 [0.96-1.26] | 1.05 [0.98-1.13] | 2,306 | 1,973 | 1,237 | 644 | 2,012 |
| 348.4 | 1.20 [1.08-1.33] | 1.20 [1.06-1.36] | 1.14 [0.98-1.32] | 1.20 [1.01-1.42] | 1.15 [1.03-1.28] | 1.15 [1.03-1.28] | 1.08 [0.95-1.23] | 1.03 [0.88-1.20] | 1.15 [0.96-1.37] | 1.11 [0.99-1.24] | 884 | 617 | 439 | 340 | 787 |
| 348.7 | 1.17 [1.10-1.24] | 1.19 [1.11-1.27] | 1.18 [1.10-1.27] | 1.05 [0.90-1.22] | 1.12 [1.05-1.20] | 1.13 [1.06-1.20] | 1.10 [1.03-1.18] | 1.09 [1.01-1.18] | 1.02 [0.88-1.20] | 1.10 [1.03-1.18] | 2,569 | 2,282 | 1,947 | 443 | 2,292 |
| 348.8 | 1.40 [1.28-1.54] | 1.43 [1.29-1.58] | 1.45 [1.30-1.61] | 0.93 [0.73-1.17] | 1.32 [1.20-1.46] | 1.30 [1.18-1.43] | 1.26 [1.14-1.40] | 1.26 [1.13-1.40] | 0.92 [0.73-1.17] | 1.30 [1.18-1.43] | 1,143 | 988 | 901 | 178 | 1,078 |
| 348.9 | 1.14 [1.08-1.21] | 1.16 [1.09-1.23] | 1.18 [1.11-1.26] | 1.01 [0.87-1.17] | 1.07 [1.00-1.13] | 1.10 [1.03-1.16] | 1.08 [1.01-1.14] | 1.09 [1.02-1.16] | 0.97 [0.83-1.12] | 1.05 [0.98-1.11] | 3,027 | 2,691 | 2,402 | 461 | 2,737 |
| 349 | 1.32 [1.24-1.41] | 1.34 [1.25-1.43] | 1.40 [1.30-1.50] | 1.03 [0.85-1.25] | 1.24 [1.16-1.32] | 1.25 [1.17-1.33] | 1.20 [1.12-1.28] | 1.26 [1.17-1.35] | 1.00 [0.82-1.22] | 1.21 [1.13-1.29] | 2,442 | 2,267 | 1,993 | 271 | 2,248 |
| 350.1 | 1.24 [1.19-1.29] | 1.29 [1.23-1.35] | 1.29 [1.23-1.35] | 1.10 [1.01-1.20] | 1.17 [1.12-1.22] | 1.15 [1.11-1.20] | 1.15 [1.10-1.20] | 1.15 [1.10-1.21] | 1.06 [0.96-1.16] | 1.12 [1.07-1.17] | 6,355 | 5,322 | 4,542 | 1,312 | 5,761 |
| 350.2 | 1.21 [1.19-1.23] | 1.20 [1.19-1.22] | 1.20 [1.18-1.22] | 1.10 [1.03-1.17] | 1.15 [1.13-1.17] | 1.16 [1.15-1.18] | 1.12 [1.10-1.14] | 1.12 [1.10-1.14] | 1.07 [1.00-1.15] | 1.13 [1.11-1.15] | 45,075 | 43,012 | 42,038 | 2,374 | 42,416 |
| 350.3 | 1.18 [1.10-1.27] | 1.18 [1.08-1.29] | 1.20 [1.09-1.31] | 1.17 [1.03-1.33] | 1.12 [1.04-1.21] | 1.12 [1.04-1.21] | 1.07 [0.98-1.16] | 1.09 [0.99-1.20] | 1.12 [0.98-1.28] | 1.09 [1.01-1.18] | 1,904 | 1,344 | 1,171 | 629 | 1,698 |
| 350.6 | 1.35 [1.14-1.61] | 1.28 [1.07-1.52] | 1.36 [1.11-1.66] | | 1.24 [1.04-1.49] | 1.23 [1.03-1.47] | 1.12 [0.93-1.34] | 1.14 [0.92-1.41] | | 1.17 [0.98-1.41] | 348 | 318 | 243 | NA | 305 |
| 351 | 1.35 [1.32-1.38] | 1.35 [1.32-1.38] | 1.34 [1.31-1.37] | 1.14 [1.00-1.31] | 1.24 [1.21-1.27] | 1.24 [1.21-1.27] | 1.19 [1.16-1.21] | 1.17 [1.15-1.20] | 1.08 [0.94-1.24] | 1.16 [1.14-1.19] | 22,138 | 21,937 | 19,241 | 596 | 19,088 |
| 352 | 1.30 [1.17-1.45] | 1.32 [1.18-1.47] | 1.33 [1.19-1.49] | | 1.26 [1.13-1.41] | 1.23 [1.10-1.37] | 1.20 [1.07-1.35] | 1.22 [1.09-1.37] | | 1.20 [1.07-1.35] | 886 | 849 | 795 | NA | 813 |
| 352.1 | 1.42 [1.32-1.53] | 1.43 [1.33-1.55] | 1.44 [1.33-1.56] | | 1.32 [1.22-1.42] | 1.29 [1.20-1.39] | 1.22 [1.13-1.32] | 1.24 [1.15-1.35] | | 1.22 [1.13-1.32] | 1,837 | 1,821 | 1,646 | NA | 1,651 |
| 352.2 | 1.21 [1.15-1.28] | 1.24 [1.17-1.31] | 1.21 [1.14-1.29] | 1.09 [0.95-1.24] | 1.15 [1.09-1.21] | 1.15 [1.09-1.21] | 1.13 [1.06-1.19] | 1.10 [1.04-1.17] | 1.09 [0.95-1.24] | 1.11 [1.05-1.17] | 3,683 | 3,203 | 2,781 | 595 | 3,191 |
| 353 | 1.28 [1.25-1.32] | 1.27 [1.24-1.31] | 1.29 [1.25-1.32] | 1.14 [0.99-1.31] | 1.18 [1.15-1.22] | 1.18 [1.14-1.21] | 1.11 [1.08-1.14] | 1.12 [1.08-1.15] | 1.07 [0.93-1.24] | 1.12 [1.09-1.15] | 12,810 | 12,750 | 10,597 | 528 | 11,564 |
| 353.1 | 1.31 [1.09-1.58] | 1.31 [1.09-1.57] | 1.32 [1.05-1.66] | | 1.22 [1.01-1.48] | 1.19 [0.98-1.44] | 1.15 [0.95-1.39] | 1.15 [0.91-1.46] | | 1.16 [0.95-1.41] | 292 | 278 | 183 | NA | 258 |
| 353.2 | 1.27 [1.00-1.61] | 1.20 [0.95-1.53] | 1.23 [0.96-1.59] | | 1.18 [0.92-1.51] | 1.14 [0.89-1.45] | 0.98 [0.77-1.27] | 1.03 [0.79-1.35] | | 1.09 [0.85-1.41] | 169 | 168 | 144 | NA | 154 |
| 356 | 1.28 [1.11-1.46] | 1.26 [1.10-1.46] | 1.26 [1.09-1.47] | | 1.17 [1.01-1.35] | 1.17 [1.02-1.34] | 1.12 [0.97-1.30] | 1.11 [0.95-1.30] | | 1.07 [0.93-1.24] | 529 | 483 | 419 | NA | 473 |
| 357 | 1.36 [1.32-1.40] | 1.36 [1.32-1.40] | 1.36 [1.32-1.41] | 1.02 [0.84-1.23] | 1.24 [1.20-1.28] | 1.24 [1.20-1.28] | 1.15 [1.11-1.19] | 1.15 [1.11-1.19] | 0.99 [0.81-1.20] | 1.17 [1.14-1.21] | 9,917 | 9,762 | 9,264 | 284 | 9,311 |
| 358 | 1.27 [1.00-1.61] | 1.34 [1.02-1.75] | 1.21 [0.90-1.62] | | 1.11 [0.86-1.42] | 1.21 [0.95-1.55] | 1.25 [0.95-1.65] | 1.13 [0.83-1.52] | | 1.09 [0.84-1.40] | 174 | 136 | 113 | NA | 157 |
| 358.1 | 1.09 [0.95-1.26] | 1.11 [0.96-1.28] | 1.16 [0.99-1.35] | | 1.13 [0.97-1.32] | 1.05 [0.90-1.21] | 1.04 [0.90-1.21] | 1.07 [0.92-1.25] | | 1.09 [0.93-1.27] | 459 | 446 | 408 | NA | 400 |
| 359.1 | 1.20 [1.04-1.38] | 1.22 [1.03-1.45] | 1.13 [0.93-1.38] | 1.00 [0.79-1.27] | 1.04 [0.89-1.21] | 1.17 [1.01-1.35] | 1.18 [0.99-1.40] | 1.08 [0.89-1.32] | 1.00 [0.79-1.28] | 1.03 [0.88-1.21] | 488 | 341 | 243 | 172 | 400 |
| 359.2 | 1.35 [1.27-1.43] | 1.37 [1.28-1.47] | 1.35 [1.26-1.46] | 1.21 [1.07-1.36] | 1.26 [1.18-1.34] | 1.29 [1.21-1.37] | 1.26 [1.17-1.35] | 1.26 [1.17-1.35] | 1.18 [1.04-1.33] | 1.23 [1.15-1.31] | 2,818 | 2,185 | 1,838 | 728 | 2,470 |
| sense organs | | | | | | | | | | | | | | | |
| 360 | 1.30 [1.20-1.41] | 1.33 [1.23-1.44] | 1.30 [1.19-1.41] | 1.29 [0.99-1.67] | 1.25 [1.15-1.36] | 1.24 [1.15-1.34] | 1.26 [1.16-1.37] | 1.23 [1.12-1.34] | 1.17 [0.89-1.53] | 1.20 [1.10-1.31] | 1,642 | 1,518 | 1,390 | 159 | 1,453 |
| 360.2 | 1.17 [0.99-1.38] | 1.13 [0.96-1.34] | 1.13 [0.96-1.34] | | 1.09 [0.91-1.30] | 1.15 [0.97-1.35] | 1.12 [0.94-1.32] | 1.11 [0.93-1.32] | | 1.04 [0.86-1.24] | 351 | 339 | 325 | NA | 286 |
| 361 | 1.08 [1.02-1.15] | 1.11 [1.04-1.18] | 1.09 [1.01-1.16] | 1.01 [0.76-1.35] | 1.06 [0.99-1.14] | 1.05 [0.98-1.12] | 1.07 [1.00-1.14] | 1.05 [0.98-1.12] | 1.01 [0.75-1.34] | 1.05 [0.98-1.13] | 2,275 | 2,204 | 2,000 | 128 | 1,834 |
| 361.1 | 1.05 [1.00-1.10] | 1.08 [1.02-1.13] | 1.04 [0.99-1.10] | 1.10 [0.89-1.35] | 1.03 [0.98-1.09] | 1.03 [0.98-1.08] | 1.05 [1.00-1.11] | 1.02 [0.97-1.08] | 1.08 [0.87-1.34] | 1.03 [0.97-1.09] | 3,916 | 3,790 | 3,415 | 243 | 3,116 |
| 361.2 | 1.19 [0.96-1.46] | 1.18 [0.96-1.46] | 1.16 [0.94-1.44] | | 1.15 [0.92-1.44] | 1.17 [0.95-1.45] | 1.14 [0.92-1.42] | 1.15 [0.92-1.43] | | 1.14 [0.91-1.43] | 219 | 216 | 206 | NA | 197 |
| 362 | 1.17 [1.14-1.19] | 1.17 [1.15-1.20] | 1.16 [1.14-1.19] | 1.16 [0.96-1.39] | 1.12 [1.10-1.15] | 1.13 [1.11-1.16] | 1.12 [1.10-1.15] | 1.11 [1.09-1.13] | 1.14 [0.94-1.38] | 1.10 [1.08-1.13] | 23,935 | 23,741 | 23,343 | 308 | 21,152 |
| 362.2 | 1.15 [1.13-1.18] | 1.16 [1.13-1.19] | 1.15 [1.13-1.18] | | 1.12 [1.09-1.14] | 1.12 [1.09-1.15] | 1.11 [1.09-1.14] | 1.10 [1.08-1.13] | | 1.09 [1.07-1.12] | 19,942 | 19,927 | 19,823 | NA | 17,772 |
| 362.29 | 1.15 [1.13-1.18] | 1.16 [1.13-1.19] | 1.15 [1.13-1.18] | | 1.12 [1.09-1.14] | 1.12 [1.09-1.15] | 1.11 [1.09-1.14] | 1.10 [1.08-1.13] | | 1.09 [1.07-1.12] | 19,942 | 19,927 | 19,823 | NA | 17,772 |
| 362.3 | 1.24 [1.01-1.53] | 1.30 [1.05-1.60] | 1.24 [0.99-1.54] | | 1.11 [0.90-1.38] | 1.19 [0.97-1.47] | 1.17 [0.94-1.45] | 1.09 [0.87-1.38] | | 1.12 [0.90-1.39] | 233 | 218 | 194 | NA | 207 |
| 362.31 | 1.23 [1.07-1.42] | 1.24 [1.08-1.43] | 1.20 [1.04-1.38] | | 1.17 [1.00-1.35] | 1.19 [1.04-1.38] | 1.18 [1.03-1.36] | 1.15 [1.00-1.33] | | 1.14 [0.98-1.33] | 521 | 516 | 501 | NA | 457 |
| 362.4 | 1.19 [1.14-1.26] | 1.20 [1.15-1.27] | 1.18 [1.12-1.24] | | 1.13 [1.07-1.19] | 1.16 [1.10-1.22] | 1.13 [1.08-1.19] | 1.10 [1.05-1.16] | | 1.11 [1.05-1.17] | 4,086 | 4,030 | 3,898 | NA | 3,633 |
| 362.6 | 1.09 [0.89-1.33] | 1.09 [0.89-1.34] | 1.02 [0.82-1.26] | | 1.02 [0.81-1.27] | 1.07 [0.87-1.31] | 1.07 [0.87-1.32] | 0.98 [0.78-1.22] | | 1.00 [0.80-1.25] | 234 | 230 | 199 | NA | 191 |
| 362.7 | 1.25 [1.22-1.29] | 1.26 [1.22-1.30] | 1.25 [1.21-1.29] | 1.16 [0.90-1.49] | 1.18 [1.14-1.22] | 1.19 [1.15-1.23] | 1.10 [1.07-1.14] | 1.09 [1.05-1.13] | 1.19 [0.91-1.54] | 1.16 [1.12-1.20] | 10,332 | 10,279 | 9,810 | 164 | 9,418 |
| 362.8 | 1.23 [1.07-1.42] | 1.20 [1.04-1.38] | 1.22 [1.06-1.42] | | 1.20 [1.04-1.40] | 1.19 [1.03-1.37] | 1.13 [0.97-1.30] | 1.15 [0.99-1.34] | | 1.18 [1.01-1.38] | 513 | 482 | 450 | NA | 439 |
| 363 | 1.12 [1.02-1.23] | 1.13 [1.03-1.25] | 1.11 [1.00-1.22] | | 1.05 [0.95-1.17] | 1.10 [0.99-1.21] | 1.09 [0.99-1.21] | 1.06 [0.96-1.17] | | 1.04 [0.94-1.16] | 1,077 | 1,054 | 1,007 | NA | 966 |
| 363.3 | 1.21 [1.05-1.39] | 1.20 [1.04-1.38] | 1.16 [1.00-1.34] | | 1.16 [1.00-1.34] | 1.17 [1.02-1.36] | 1.15 [0.99-1.33] | 1.11 [0.95-1.28] | | 1.15 [0.99-1.34] | 510 | 498 | 476 | NA | 477 |
| 363.4 | 1.05 [0.78-1.42] | | | | | 1.03 [0.76-1.40] | | | | | 104 | NA | NA | NA | NA |
| 364 | 1.38 [1.32-1.44] | 1.36 [1.30-1.43] | 1.24 [1.18-1.31] | 1.91 [1.67-2.18] | 1.29 [1.23-1.36] | 1.31 [1.25-1.37] | 1.27 [1.22-1.34] | 1.19 [1.13-1.25] | 1.70 [1.48-1.95] | 1.24 [1.18-1.30] | 5,208 | 4,964 | 3,982 | 735 | 4,460 |
| 364.1 | 1.41 [1.26-1.59] | 1.40 [1.24-1.57] | 1.37 [1.21-1.56] | | 1.39 [1.23-1.57] | 1.34 [1.19-1.51] | 1.30 [1.15-1.48] | 1.28 [1.12-1.46] | | 1.33 [1.18-1.50] | 774 | 722 | 635 | NA | 694 |
| 364.2 | 1.18 [1.02-1.37] | 1.19 [1.02-1.38] | 1.24 [1.06-1.44] | | 1.16 [1.00-1.36] | 1.12 [0.96-1.30] | 1.11 [0.96-1.29] | 1.17 [1.00-1.37] | | 1.10 [0.94-1.29] | 451 | 447 | 426 | NA | 410 |
| 364.4 | 1.21 [1.12-1.30] | 1.21 [1.12-1.30] | 1.20 [1.11-1.29] | | 1.18 [1.09-1.27] | 1.18 [1.09-1.27] | 1.16 [1.08-1.25] | 1.15 [1.07-1.24] | | 1.15 [1.06-1.24] | 1,875 | 1,859 | 1,825 | NA | 1,731 |
| 364.41 | 2.36 [2.13-2.62] | 2.37 [2.13-2.63] | 2.17 [1.74-2.71] | 2.00 [1.73-2.32] | 2.24 [1.98-2.52] | 2.01 [1.80-2.25] | 1.98 [1.77-2.21] | 1.80 [1.42-2.28] | 1.77 [1.51-2.06] | 1.94 [1.71-2.21] | 1,203 | 1,051 | 218 | 602 | 862 |
| 364.5 | 1.26 [1.14-1.40] | 1.21 [1.09-1.34] | 1.20 [1.08-1.33] | | 1.16 [1.04-1.29] | 1.22 [1.10-1.35] | 1.15 [1.04-1.27] | 1.14 [1.03-1.27] | | 1.13 [1.02-1.26] | 978 | 969 | 946 | NA | 860 |
| 364.9 | 1.73 [1.50-2.00] | 1.70 [1.47-1.96] | 1.65 [1.39-1.94] | | 1.66 [1.43-1.93] | 1.57 [1.35-1.83] | 1.50 [1.29-1.74] | 1.48 [1.25-1.75] | | 1.51 [1.29-1.76] | 507 | 497 | 381 | NA | 466 |
| 365 | 1.18 [1.15-1.20] | 1.18 [1.15-1.20] | 1.18 [1.15-1.20] | 1.30 [1.01-1.67] | 1.13 [1.11-1.16] | 1.14 [1.12-1.17] | 1.13 [1.11-1.16] | 1.13 [1.11-1.16] | 1.26 [0.97-1.64] | 1.11 [1.08-1.14] | 21,888 | 21,771 | 21,556 | 166 | 18,785 |
| 365.11 | 1.16 [1.11-1.20] | 1.15 [1.10-1.20] | 1.16 [1.11-1.20] | | 1.11 [1.06-1.16] | 1.13 [1.09-1.18] | 1.12 [1.08-1.17] | 1.13 [1.08-1.18] | | 1.09 [1.04-1.14] | 5,891 | 5,870 | 5,851 | NA | 5,142 |
| 365.2 | 1.14 [1.06-1.22] | 1.13 [1.06-1.22] | 1.15 [1.07-1.24] | | 1.09 [1.01-1.18] | 1.10 [1.03-1.19] | 1.10 [1.02-1.18] | 1.11 [1.04-1.20] | | 1.07 [0.99-1.16] | 1,881 | 1,879 | 1,858 | NA | 1,581 |
| 366 | 1.21 [1.20-1.22] | 1.21 [1.20-1.23] | 1.21 [1.20-1.23] | 1.49 [1.28-1.73] | 1.16 [1.14-1.17] | 1.17 [1.15-1.18] | 1.14 [1.13-1.16] | 1.14 [1.13-1.16] | 1.44 [1.23-1.67] | 1.13 [1.11-1.14] | 77,912 | 77,601 | 76,878 | 503 | 66,285 |
| 366.1 | 1.47 [1.15-1.86] | 1.49 [1.15-1.93] | 1.40 [1.05-1.86] | | 1.32 [1.00-1.73] | 1.43 [1.12-1.83] | 1.38 [1.05-1.80] | 1.26 [0.94-1.70] | | 1.28 [0.97-1.69] | 171 | 143 | 113 | NA | 130 |
| 366.2 | 1.17 [1.15-1.19] | 1.17 [1.16-1.19] | 1.18 [1.17-1.20] | | 1.12 [1.11-1.14] | 1.13 [1.12-1.15] | 1.12 [1.10-1.13] | 1.13 [1.11-1.14] | | 1.10 [1.08-1.12] | 46,074 | 46,065 | 45,897 | NA | 39,385 |
| 366.3 | 1.08 [0.85-1.36] | 1.12 [0.87-1.45] | 1.08 [0.82-1.43] | | 1.00 [0.76-1.32] | 1.07 [0.84-1.37] | 1.11 [0.85-1.45] | 1.05 [0.78-1.40] | | 1.00 [0.75-1.32] | 164 | 141 | 116 | NA | 122 |
| 367 | 1.12 [1.09-1.16] | 1.13 [1.09-1.17] | 1.12 [1.08-1.17] | 1.12 [1.01-1.23] | 1.09 [1.05-1.14] | 1.09 [1.05-1.13] | 1.09 [1.05-1.13] | 1.08 [1.04-1.12] | 1.09 [0.99-1.20] | 1.07 [1.03-1.11] | 8,017 | 7,057 | 6,729 | 1,094 | 7,038 |
| 367.1 | 1.08 [1.02-1.15] | 1.08 [1.02-1.15] | 1.08 [1.01-1.15] | 1.10 [0.91-1.33] | 1.05 [0.99-1.12] | 1.06 [1.00-1.12] | 1.06 [0.99-1.12] | 1.06 [0.99-1.13] | 1.07 [0.88-1.30] | 1.03 [0.97-1.10] | 2,804 | 2,561 | 2,398 | 308 | 2,335 |
| 367.2 | 1.17 [1.11-1.23] | 1.14 [1.08-1.21] | 1.13 [1.07-1.20] | 1.35 [1.13-1.61] | 1.13 [1.06-1.19] | 1.13 [1.07-1.20] | 1.09 [1.03-1.15] | 1.07 [1.01-1.14] | 1.31 [1.09-1.57] | 1.09 [1.03-1.16] | 3,421 | 3,124 | 3,017 | 338 | 3,137 |
| 367.8 | 1.12 [1.02-1.22] | 1.25 [1.13-1.39] | 1.20 [1.08-1.34] | 0.98 [0.84-1.14] | 1.04 [0.94-1.14] | 1.08 [0.99-1.18] | 1.20 [1.08-1.34] | 1.15 [1.03-1.28] | 0.95 [0.81-1.11] | 1.01 [0.92-1.12] | 1,287 | 887 | 845 | 424 | 1,107 |
| 367.9 | 1.20 [1.16-1.24] | 1.22 [1.18-1.25] | 1.21 [1.17-1.25] | 1.00 [0.89-1.12] | 1.14 [1.11-1.18] | 1.16 [1.12-1.20] | 1.14 [1.11-1.18] | 1.14 [1.10-1.17] | 0.96 [0.86-1.08] | 1.12 [1.09-1.16] | 11,421 | 10,807 | 10,273 | 773 | 10,552 |
| 368 | 1.24 [1.21-1.27] | 1.25 [1.22-1.28] | 1.25 [1.21-1.29] | 1.18 [1.12-1.24] | 1.16 [1.13-1.19] | 1.16 [1.13-1.18] | 1.11 [1.08-1.14] | 1.13 [1.09-1.16] | 1.13 [1.07-1.19] | 1.11 [1.08-1.14] | 16,638 | 14,327 | 10,510 | 3,753 | 14,631 |
| 368.1 | 1.19 [1.12-1.27] | 1.23 [1.15-1.31] | 1.21 [1.13-1.30] | 1.15 [0.99-1.33] | 1.17 [1.09-1.25] | 1.15 [1.08-1.23] | 1.16 [1.08-1.24] | 1.16 [1.08-1.24] | 1.14 [0.98-1.33] | 1.14 [1.07-1.22] | 2,540 | 2,123 | 2,005 | 465 | 2,205 |
| 368.2 | 1.23 [1.16-1.31] | 1.18 [1.11-1.26] | 1.21 [1.13-1.30] | 1.27 [1.11-1.45] | 1.15 [1.09-1.23] | 1.15 [1.08-1.22] | 1.05 [0.99-1.13] | 1.08 [1.00-1.16] | 1.24 [1.08-1.42] | 1.10 [1.04-1.18] | 2,891 | 2,440 | 2,009 | 609 | 2,537 |
| 368.3 | 1.15 [1.00-1.32] | 1.14 [0.98-1.32] | 1.09 [0.93-1.27] | | 1.15 [0.99-1.33] | 1.12 [0.97-1.29] | 1.08 [0.93-1.27] | 1.04 [0.89-1.22] | | 1.11 [0.95-1.29] | 499 | 421 | 411 | NA | 441 |
| 368.4 | 1.17 [1.08-1.25] | 1.16 [1.08-1.25] | 1.13 [1.05-1.22] | 1.28 [0.99-1.67] | 1.10 [1.02-1.19] | 1.13 [1.05-1.22] | 1.10 [1.02-1.18] | 1.07 [0.99-1.15] | 1.30 [0.99-1.70] | 1.09 [1.01-1.18] | 1,907 | 1,794 | 1,661 | 157 | 1,685 |
| 368.9 | 1.25 [1.20-1.31] | 1.26 [1.21-1.32] | 1.24 [1.18-1.31] | 1.18 [1.09-1.29] | 1.16 [1.11-1.21] | 1.16 [1.11-1.21] | 1.11 [1.06-1.16] | 1.11 [1.05-1.17] | 1.13 [1.03-1.23] | 1.12 [1.07-1.16] | 6,221 | 5,337 | 3,782 | 1,474 | 5,632 |
| 369 | 1.42 [1.33-1.51] | 1.47 [1.33-1.63] | 1.45 [1.30-1.63] | 1.35 [1.25-1.45] | 1.33 [1.25-1.43] | 1.36 [1.28-1.45] | 1.35 [1.22-1.50] | 1.35 [1.20-1.51] | 1.30 [1.21-1.41] | 1.30 [1.21-1.39] | 2,692 | 1,009 | 790 | 1,757 | 2,239 |
| 369.2 | 1.54 [1.39-1.71] | 1.56 [1.39-1.76] | 1.42 [1.25-1.60] | 1.67 [1.34-2.09] | 1.40 [1.25-1.56] | 1.44 [1.29-1.60] | 1.42 [1.26-1.60] | 1.28 [1.13-1.46] | 1.55 [1.24-1.95] | 1.30 [1.16-1.46] | 985 | 792 | 694 | 229 | 858 |
| 369.5 | 1.43 [1.36-1.50] | 1.43 [1.34-1.52] | 1.35 [1.26-1.45] | 1.42 [1.32-1.54] | 1.35 [1.28-1.42] | 1.34 [1.27-1.41] | 1.29 [1.21-1.38] | 1.24 [1.16-1.33] | 1.32 [1.22-1.43] | 1.28 [1.22-1.35] | 4,406 | 2,770 | 2,435 | 1,760 | 3,999 |
| 370 | 1.72 [1.58-1.87] | 1.66 [1.52-1.81] | 1.61 [1.46-1.77] | 1.90 [1.52-2.38] | 1.61 [1.47-1.76] | 1.61 [1.48-1.75] | 1.53 [1.40-1.67] | 1.50 [1.36-1.66] | 1.73 [1.37-2.18] | 1.54 [1.41-1.69] | 1,569 | 1,397 | 1,162 | 243 | 1,348 |
| 370.1 | 1.69 [1.51-1.90] | 1.64 [1.46-1.86] | 1.53 [1.34-1.75] | 1.99 [1.44-2.75] | 1.58 [1.40-1.79] | 1.58 [1.41-1.79] | 1.52 [1.34-1.73] | 1.42 [1.24-1.63] | 1.85 [1.32-2.59] | 1.52 [1.34-1.73] | 802 | 725 | 595 | 111 | 686 |
| 370.3 | 2.82 [2.23-3.57] | 2.47 [1.85-3.28] | 2.15 [1.57-2.93] | 4.44 [2.90-6.81] | 2.39 [1.85-3.07] | 2.53 [1.97-3.24] | 2.06 [1.53-2.79] | 1.98 [1.43-2.72] | 3.71 [2.36-5.83] | 2.14 [1.65-2.78] | 243 | 153 | 117 | 105 | 206 |
| 371 | 1.44 [1.28-1.63] | 1.47 [1.29-1.67] | 1.40 [1.22-1.59] | | 1.37 [1.21-1.56] | 1.36 [1.20-1.54] | 1.37 [1.20-1.56] | 1.31 [1.15-1.50] | | 1.32 [1.16-1.50] | 717 | 645 | 599 | NA | 637 |
| 371.1 | 1.38 [1.28-1.48] | 1.32 [1.23-1.43] | 1.31 [1.20-1.42] | 1.40 [1.14-1.72] | 1.32 [1.22-1.42] | 1.33 [1.23-1.43] | 1.25 [1.15-1.35] | 1.24 [1.14-1.35] | 1.36 [1.10-1.68] | 1.29 [1.19-1.39] | 1,890 | 1,678 | 1,459 | 272 | 1,670 |
| 371.2 | 1.89 [1.38-2.58] | | | | | 1.58 [1.14-2.19] | | | | | 117 | NA | NA | NA | NA |
| 371.3 | 1.31 [1.26-1.36] | 1.33 [1.27-1.39] | 1.33 [1.27-1.40] | 1.23 [1.14-1.32] | 1.24 [1.18-1.29] | 1.25 [1.20-1.30] | 1.25 [1.19-1.31] | 1.25 [1.19-1.32] | 1.20 [1.11-1.29] | 1.19 [1.14-1.24] | 6,529 | 4,781 | 3,800 | 2,103 | 5,206 |
| 371.33 | 3.77 [2.55-5.57] | | | | | 3.42 [2.29-5.11] | | | | | 103 | NA | NA | NA | NA |
| 372 | 1.24 [1.16-1.31] | 1.24 [1.16-1.32] | 1.28 [1.19-1.36] | 1.11 [0.92-1.33] | 1.17 [1.10-1.25] | 1.19 [1.12-1.26] | 1.18 [1.11-1.26] | 1.21 [1.13-1.29] | 1.08 [0.89-1.30] | 1.15 [1.08-1.23] | 2,749 | 2,548 | 2,268 | 293 | 2,312 |
| 374 | 1.31 [1.27-1.34] | 1.31 [1.28-1.35] | 1.31 [1.27-1.35] | 1.23 [1.11-1.35] | 1.22 [1.19-1.26] | 1.24 [1.21-1.28] | 1.22 [1.19-1.26] | 1.22 [1.19-1.26] | 1.19 [1.07-1.31] | 1.17 [1.14-1.21] | 13,550 | 12,700 | 11,939 | 1,055 | 11,510 |
| 374.1 | 1.36 [1.31-1.42] | 1.35 [1.29-1.41] | 1.34 [1.28-1.40] | 1.55 [1.11-2.16] | 1.26 [1.21-1.32] | 1.31 [1.26-1.37] | 1.27 [1.22-1.33] | 1.27 [1.21-1.32] | 1.61 [1.14-2.28] | 1.23 [1.17-1.28] | 5,689 | 5,602 | 5,528 | 105 | 4,912 |
| 374.2 | 1.24 [0.96-1.61] | 1.32 [1.00-1.74] | 1.16 [0.87-1.54] | | 1.18 [0.90-1.54] | 1.20 [0.92-1.56] | 1.23 [0.92-1.63] | 1.11 [0.83-1.49] | | 1.14 [0.86-1.50] | 144 | 129 | 117 | NA | 131 |
| 374.3 | 1.25 [1.19-1.32] | 1.27 [1.20-1.33] | 1.28 [1.21-1.35] | 1.12 [0.95-1.32] | 1.17 [1.11-1.23] | 1.17 [1.12-1.23] | 1.15 [1.09-1.22] | 1.16 [1.10-1.23] | 1.09 [0.92-1.29] | 1.11 [1.05-1.17] | 4,020 | 3,713 | 3,520 | 367 | 3,570 |
| 375 | 1.40 [1.35-1.45] | 1.44 [1.38-1.50] | 1.44 [1.39-1.51] | 1.20 [1.09-1.32] | 1.31 [1.26-1.37] | 1.30 [1.25-1.35] | 1.32 [1.27-1.37] | 1.32 [1.26-1.37] | 1.15 [1.04-1.26] | 1.24 [1.19-1.29] | 7,312 | 6,384 | 6,029 | 1,027 | 6,318 |
| 375.2 | 1.44 [1.35-1.53] | 1.54 [1.44-1.65] | 1.51 [1.41-1.62] | 1.24 [1.07-1.43] | 1.36 [1.28-1.46] | 1.35 [1.27-1.44] | 1.43 [1.33-1.53] | 1.39 [1.29-1.49] | 1.20 [1.03-1.38] | 1.30 [1.22-1.39] | 2,649 | 2,219 | 2,093 | 459 | 2,286 |
| 376 | 1.40 [1.32-1.49] | 1.43 [1.30-1.57] | 1.47 [1.33-1.64] | 1.33 [1.23-1.43] | 1.32 [1.23-1.41] | 1.35 [1.27-1.44] | 1.32 [1.20-1.45] | 1.35 [1.21-1.50] | 1.29 [1.19-1.39] | 1.29 [1.21-1.38] | 2,911 | 1,174 | 928 | 1,822 | 2,424 |
| 377 | 1.12 [1.03-1.22] | 1.19 [1.07-1.31] | 1.19 [1.05-1.36] | 1.09 [0.96-1.25] | 1.11 [1.01-1.22] | 1.08 [0.99-1.18] | 1.09 [0.98-1.20] | 1.16 [1.02-1.32] | 1.04 [0.91-1.20] | 1.07 [0.98-1.18] | 1,372 | 969 | 557 | 607 | 1,184 |
| 377.1 | 1.31 [1.10-1.55] | 1.29 [1.07-1.56] | 1.36 [1.11-1.66] | | 1.23 [1.03-1.47] | 1.25 [1.05-1.49] | 1.20 [0.99-1.46] | 1.26 [1.03-1.55] | | 1.21 [1.01-1.45] | 353 | 289 | 255 | NA | 332 |
| 377.3 | 1.31 [1.18-1.45] | 1.34 [1.20-1.50] | 1.28 [1.14-1.45] | 1.28 [1.00-1.64] | 1.21 [1.08-1.35] | 1.28 [1.15-1.42] | 1.24 [1.11-1.39] | 1.19 [1.05-1.35] | 1.29 [1.00-1.66] | 1.19 [1.06-1.33] | 927 | 819 | 660 | 176 | 798 |
| 378 | 1.19 [1.02-1.38] | 1.29 [1.06-1.57] | 1.21 [0.95-1.52] | 0.99 [0.79-1.24] | 1.06 [0.90-1.25] | 1.11 [0.95-1.30] | 1.16 [0.94-1.42] | 1.11 [0.87-1.41] | 0.94 [0.75-1.19] | 1.02 [0.87-1.20] | 416 | 247 | 179 | 192 | 367 |
| 378.1 | 1.07 [1.03-1.11] | 1.26 [1.18-1.36] | 1.28 [1.17-1.39] | 1.00 [0.96-1.05] | 1.03 [0.98-1.08] | 1.04 [1.00-1.08] | 1.18 [1.10-1.28] | 1.19 [1.09-1.30] | 0.99 [0.94-1.03] | 1.01 [0.97-1.06] | 6,070 | 1,823 | 1,238 | 4,451 | 5,057 |
| 378.2 | 1.12 [1.03-1.23] | 1.19 [1.07-1.32] | 1.15 [1.03-1.29] | 0.95 [0.80-1.11] | 1.06 [0.97-1.17] | 1.07 [0.98-1.17] | 1.09 [0.98-1.21] | 1.04 [0.93-1.17] | 0.92 [0.78-1.09] | 1.04 [0.94-1.14] | 1,222 | 903 | 763 | 368 | 1,120 |
| 378.5 | 1.06 [0.97-1.16] | 1.10 [1.00-1.21] | 1.13 [1.02-1.26] | 0.97 [0.80-1.18] | 1.02 [0.93-1.13] | 1.01 [0.92-1.11] | 1.02 [0.92-1.13] | 1.05 [0.94-1.17] | 0.92 [0.75-1.12] | 1.00 [0.91-1.10] | 1,185 | 965 | 849 | 258 | 1,024 |
| 379 | 1.32 [1.21-1.43] | 1.34 [1.22-1.46] | 1.34 [1.22-1.47] | 1.20 [1.00-1.45] | 1.26 [1.16-1.37] | 1.24 [1.14-1.35] | 1.23 [1.12-1.34] | 1.24 [1.13-1.36] | 1.15 [0.95-1.39] | 1.19 [1.09-1.30] | 1,573 | 1,315 | 1,159 | 302 | 1,384 |
| 379.1 | 1.62 [1.29-2.04] | 1.65 [1.31-2.07] | 1.46 [1.13-1.90] | | 1.56 [1.24-1.98] | 1.50 [1.19-1.90] | 1.43 [1.12-1.81] | 1.28 [0.98-1.68] | | 1.42 [1.11-1.81] | 207 | 197 | 151 | NA | 182 |
| 379.2 | 1.12 [1.07-1.18] | 1.13 [1.07-1.18] | 1.14 [1.09-1.20] | | 1.08 [1.03-1.13] | 1.08 [1.03-1.14] | 1.06 [1.01-1.12] | 1.08 [1.03-1.13] | | 1.06 [1.01-1.11] | 4,314 | 4,273 | 4,086 | NA | 3,817 |
| 379.3 | 1.15 [1.11-1.18] | 1.15 [1.11-1.19] | 1.16 [1.12-1.20] | | 1.10 [1.06-1.14] | 1.11 [1.07-1.15] | 1.09 [1.06-1.13] | 1.10 [1.06-1.14] | | 1.07 [1.03-1.11] | 9,161 | 9,105 | 9,014 | NA | 7,958 |
| 379.4 | 1.08 [0.95-1.22] | 1.08 [0.94-1.23] | 1.05 [0.91-1.21] | 1.08 [0.79-1.47] | 1.01 [0.88-1.15] | 1.03 [0.90-1.17] | 0.99 [0.86-1.14] | 0.98 [0.84-1.13] | 1.00 [0.72-1.38] | 0.98 [0.85-1.12] | 610 | 533 | 469 | 106 | 548 |
| 379.5 | 1.19 [1.11-1.28] | 1.18 [1.10-1.27] | 1.15 [1.07-1.24] | | 1.15 [1.07-1.24] | 1.15 [1.07-1.24] | 1.12 [1.04-1.21] | 1.09 [1.01-1.17] | | 1.13 [1.05-1.22] | 2,010 | 1,955 | 1,871 | NA | 1,834 |
| 379.9 | 1.41 [1.28-1.56] | 1.40 [1.25-1.56] | 1.41 [1.25-1.59] | 1.45 [1.19-1.78] | 1.29 [1.17-1.43] | 1.29 [1.16-1.43] | 1.22 [1.09-1.37] | 1.23 [1.09-1.40] | 1.41 [1.15-1.74] | 1.22 [1.10-1.35] | 1,057 | 841 | 684 | 275 | 952 |
| 380 | 1.35 [1.29-1.40] | 1.45 [1.37-1.54] | 1.45 [1.36-1.55] | 1.26 [1.19-1.33] | 1.27 [1.22-1.33] | 1.30 [1.24-1.35] | 1.35 [1.27-1.43] | 1.34 [1.26-1.43] | 1.22 [1.15-1.29] | 1.24 [1.19-1.30] | 5,997 | 3,053 | 2,561 | 3,129 | 5,249 |
| 380.1 | 1.60 [1.52-1.70] | 1.72 [1.61-1.83] | 1.85 [1.71-2.00] | 1.33 [1.21-1.46] | 1.48 [1.39-1.57] | 1.51 [1.42-1.60] | 1.54 [1.44-1.65] | 1.67 [1.54-1.81] | 1.28 [1.16-1.41] | 1.43 [1.34-1.52] | 3,397 | 2,476 | 1,794 | 1,182 | 2,917 |
| 380.4 | 1.28 [1.22-1.35] | 1.32 [1.21-1.43] | 1.35 [1.23-1.48] | 1.25 [1.18-1.33] | 1.22 [1.16-1.29] | 1.24 [1.18-1.30] | 1.22 [1.12-1.34] | 1.26 [1.14-1.38] | 1.21 [1.14-1.29] | 1.19 [1.13-1.26] | 4,195 | 1,476 | 1,265 | 2,803 | 3,729 |
| 381.1 | 1.21 [1.18-1.24] | 1.36 [1.26-1.47] | 1.35 [1.24-1.48] | 1.20 [1.17-1.23] | 1.17 [1.14-1.20] | 1.17 [1.14-1.20] | 1.22 [1.13-1.32] | 1.24 [1.13-1.35] | 1.16 [1.13-1.19] | 1.14 [1.11-1.17] | 16,747 | 1,588 | 1,181 | 15,302 | 13,800 |
| 381.11 | 1.26 [1.22-1.31] | 1.53 [1.43-1.64] | 1.52 [1.40-1.65] | 1.21 [1.16-1.25] | 1.21 [1.16-1.25] | 1.21 [1.17-1.26] | 1.39 [1.29-1.49] | 1.37 [1.26-1.50] | 1.17 [1.12-1.22] | 1.18 [1.13-1.22] | 8,185 | 2,130 | 1,463 | 6,314 | 7,188 |
| 381.2 | 1.43 [1.27-1.62] | 1.47 [1.26-1.71] | 1.46 [1.21-1.76] | 1.41 [1.17-1.70] | 1.32 [1.15-1.50] | 1.33 [1.17-1.51] | 1.26 [1.07-1.48] | 1.25 [1.03-1.52] | 1.41 [1.17-1.71] | 1.24 [1.08-1.42] | 669 | 422 | 266 | 315 | 577 |
| 381.3 | 1.15 [1.03-1.29] | 1.33 [1.14-1.55] | 1.20 [1.01-1.44] | 1.05 [0.89-1.22] | 1.06 [0.94-1.20] | 1.10 [0.98-1.24] | 1.19 [1.02-1.40] | 1.10 [0.91-1.32] | 1.02 [0.87-1.19] | 1.04 [0.92-1.18] | 766 | 422 | 301 | 386 | 673 |
| 381.9 | 1.33 [1.21-1.46] | 1.55 [1.34-1.78] | 1.41 [1.19-1.67] | 1.28 [1.13-1.45] | 1.23 [1.11-1.36] | 1.27 [1.15-1.41] | 1.41 [1.22-1.63] | 1.28 [1.07-1.52] | 1.25 [1.10-1.41] | 1.19 [1.07-1.32] | 1,112 | 505 | 339 | 676 | 979 |
| 382 | 1.35 [1.25-1.46] | 1.47 [1.34-1.61] | 1.44 [1.29-1.61] | 1.19 [1.07-1.33] | 1.28 [1.18-1.38] | 1.25 [1.16-1.35] | 1.27 [1.15-1.40] | 1.25 [1.11-1.41] | 1.15 [1.03-1.29] | 1.21 [1.11-1.31] | 1,858 | 1,132 | 791 | 841 | 1,639 |
| 383 | 1.09 [0.91-1.30] | 1.12 [0.93-1.34] | 1.14 [0.93-1.40] | | 1.11 [0.92-1.35] | 1.07 [0.90-1.29] | 1.09 [0.91-1.32] | 1.12 [0.90-1.38] | | 1.09 [0.89-1.33] | 291 | 286 | 209 | NA | 238 |
| 384 | 1.22 [1.14-1.30] | 1.32 [1.18-1.48] | 1.34 [1.16-1.55] | 1.19 [1.10-1.28] | 1.14 [1.06-1.22] | 1.17 [1.10-1.25] | 1.23 [1.09-1.38] | 1.23 [1.06-1.42] | 1.16 [1.07-1.25] | 1.12 [1.05-1.20] | 2,421 | 755 | 449 | 1,806 | 2,153 |
| 384.4 | 1.24 [1.18-1.31] | 1.25 [1.16-1.35] | 1.31 [1.19-1.43] | 1.17 [1.09-1.26] | 1.13 [1.07-1.20] | 1.19 [1.13-1.26] | 1.18 [1.09-1.27] | 1.20 [1.10-1.32] | 1.14 [1.06-1.23] | 1.11 [1.05-1.18] | 3,386 | 1,686 | 1,115 | 1,886 | 2,890 |
| 385 | 1.19 [1.10-1.29] | 1.24 [1.12-1.37] | 1.33 [1.18-1.51] | 1.10 [0.99-1.23] | 1.12 [1.03-1.21] | 1.15 [1.06-1.24] | 1.16 [1.04-1.29] | 1.24 [1.09-1.41] | 1.06 [0.95-1.19] | 1.09 [1.00-1.18] | 1,687 | 906 | 622 | 880 | 1,471 |
| 385.3 | 1.16 [1.09-1.25] | 1.19 [1.09-1.30] | 1.23 [1.10-1.38] | 1.11 [1.01-1.23] | 1.09 [1.01-1.18] | 1.14 [1.06-1.22] | 1.14 [1.04-1.25] | 1.17 [1.04-1.31] | 1.10 [0.99-1.22] | 1.10 [1.01-1.18] | 2,024 | 1,168 | 765 | 1,002 | 1,691 |
| 385.5 | 1.20 [1.07-1.33] | 1.17 [1.01-1.36] | 1.22 [1.01-1.48] | 1.20 [1.03-1.39] | 1.12 [1.00-1.25] | 1.15 [1.02-1.28] | 1.09 [0.93-1.27] | 1.13 [0.92-1.37] | 1.16 [1.00-1.35] | 1.08 [0.96-1.22] | 838 | 412 | 244 | 488 | 748 |
| 386 | 1.25 [1.20-1.31] | 1.26 [1.20-1.32] | 1.25 [1.20-1.31] | 1.27 [0.94-1.71] | 1.16 [1.11-1.22] | 1.17 [1.11-1.22] | 1.14 [1.09-1.20] | 1.14 [1.09-1.20] | 1.20 [0.88-1.64] | 1.10 [1.05-1.16] | 4,860 | 4,798 | 4,556 | 122 | 4,363 |
| 386.1 | 1.25 [1.17-1.34] | 1.26 [1.17-1.35] | 1.25 [1.17-1.35] | | 1.16 [1.07-1.25] | 1.17 [1.09-1.26] | 1.15 [1.07-1.24] | 1.15 [1.07-1.23] | | 1.10 [1.02-1.19] | 2,001 | 2,001 | 1,918 | NA | 1,736 |
| 386.2 | 1.25 [1.17-1.33] | 1.25 [1.18-1.34] | 1.26 [1.18-1.34] | | 1.18 [1.10-1.26] | 1.15 [1.08-1.23] | 1.13 [1.05-1.20] | 1.14 [1.07-1.22] | | 1.12 [1.04-1.19] | 2,583 | 2,554 | 2,427 | NA | 2,389 |
| 386.3 | 1.26 [1.17-1.35] | 1.27 [1.18-1.37] | 1.24 [1.15-1.34] | 1.23 [0.96-1.58] | 1.17 [1.09-1.27] | 1.16 [1.08-1.25] | 1.15 [1.07-1.24] | 1.12 [1.03-1.21] | 1.15 [0.89-1.49] | 1.10 [1.02-1.19] | 1,901 | 1,784 | 1,557 | 168 | 1,633 |
| 386.9 | 1.24 [1.21-1.26] | 1.25 [1.22-1.27] | 1.24 [1.22-1.27] | 1.11 [1.05-1.18] | 1.16 [1.13-1.18] | 1.15 [1.13-1.18] | 1.12 [1.09-1.14] | 1.12 [1.09-1.15] | 1.07 [1.01-1.14] | 1.10 [1.08-1.13] | 24,441 | 22,855 | 19,322 | 2,819 | 22,003 |
| 388 | 1.37 [1.19-1.58] | 1.39 [1.16-1.67] | 1.43 [1.16-1.77] | 1.24 [0.99-1.54] | 1.29 [1.11-1.51] | 1.30 [1.13-1.51] | 1.25 [1.03-1.51] | 1.31 [1.05-1.62] | 1.19 [0.95-1.49] | 1.24 [1.06-1.44] | 494 | 300 | 217 | 221 | 438 |
| 389 | 1.23 [1.21-1.25] | 1.24 [1.22-1.26] | 1.24 [1.22-1.26] | 1.16 [1.11-1.21] | 1.17 [1.16-1.19] | 1.18 [1.16-1.19] | 1.15 [1.13-1.17] | 1.15 [1.14-1.17] | 1.12 [1.07-1.17] | 1.14 [1.12-1.16] | 46,430 | 41,503 | 39,398 | 5,674 | 42,616 |
| 389.1 | 1.33 [1.26-1.40] | 1.39 [1.31-1.47] | 1.37 [1.29-1.46] | 1.03 [0.91-1.18] | 1.22 [1.16-1.29] | 1.23 [1.17-1.30] | 1.24 [1.17-1.32] | 1.23 [1.16-1.31] | 0.99 [0.87-1.13] | 1.16 [1.10-1.23] | 3,710 | 3,237 | 2,952 | 573 | 3,432 |
| 389.2 | 1.23 [1.16-1.31] | 1.28 [1.16-1.41] | 1.28 [1.14-1.44] | 1.21 [1.13-1.30] | 1.17 [1.09-1.24] | 1.18 [1.11-1.26] | 1.19 [1.07-1.31] | 1.18 [1.05-1.33] | 1.17 [1.09-1.26] | 1.14 [1.07-1.21] | 2,886 | 1,019 | 692 | 1,992 | 2,626 |
| 389.3 | 1.07 [0.92-1.23] | 1.08 [0.94-1.25] | 1.11 [0.96-1.28] | | 1.08 [0.94-1.25] | 1.01 [0.87-1.17] | 1.02 [0.88-1.18] | 1.03 [0.89-1.19] | | 1.04 [0.90-1.21] | 524 | 520 | 519 | NA | 501 |
| 389.4 | 1.37 [1.29-1.46] | 1.38 [1.29-1.47] | 1.35 [1.27-1.45] | 1.35 [1.04-1.75] | 1.30 [1.21-1.38] | 1.28 [1.20-1.37] | 1.24 [1.16-1.32] | 1.22 [1.14-1.31] | 1.32 [1.01-1.73] | 1.23 [1.15-1.31] | 2,572 | 2,496 | 2,204 | 176 | 2,306 |
| 389.5 | 1.39 [1.06-1.82] | | | | 1.27 [0.96-1.68] | 1.18 [0.89-1.56] | | | | 1.15 [0.86-1.54] | 144 | NA | NA | NA | 125 |
| circulatory system | | | | | | | | | | | | | | | |
| 394 | 1.21 [1.17-1.24] | 1.22 [1.18-1.25] | 1.22 [1.18-1.26] | | 1.14 [1.11-1.18] | 1.16 [1.12-1.20] | 1.14 [1.10-1.17] | 1.14 [1.10-1.17] | | 1.12 [1.08-1.15] | 12,028 | 11,979 | 11,829 | NA | 11,236 |
| 394.2 | 1.20 [1.17-1.23] | 1.20 [1.17-1.23] | 1.20 [1.18-1.23] | 0.99 [0.83-1.19] | 1.14 [1.11-1.17] | 1.15 [1.12-1.18] | 1.12 [1.09-1.14] | 1.11 [1.09-1.14] | 0.94 [0.78-1.13] | 1.11 [1.09-1.14] | 19,411 | 19,234 | 18,706 | 330 | 17,719 |
| 394.3 | 1.22 [1.18-1.26] | 1.22 [1.18-1.27] | 1.23 [1.19-1.28] | 0.99 [0.79-1.25] | 1.15 [1.11-1.19] | 1.16 [1.12-1.21] | 1.13 [1.09-1.17] | 1.13 [1.09-1.17] | 0.99 [0.78-1.26] | 1.12 [1.08-1.16] | 8,616 | 8,497 | 8,289 | 188 | 7,873 |
| 394.4 | 1.41 [1.16-1.70] | 1.37 [1.13-1.66] | 1.46 [1.20-1.78] | | 1.31 [1.08-1.60] | 1.34 [1.11-1.63] | 1.28 [1.05-1.56] | 1.34 [1.10-1.64] | | 1.27 [1.04-1.55] | 271 | 264 | 257 | NA | 250 |
| 394.7 | 1.25 [1.21-1.28] | 1.24 [1.20-1.27] | 1.24 [1.20-1.27] | 1.01 [0.81-1.25] | 1.18 [1.15-1.21] | 1.20 [1.16-1.23] | 1.14 [1.11-1.18] | 1.14 [1.11-1.17] | 0.99 [0.80-1.24] | 1.15 [1.12-1.19] | 12,847 | 12,740 | 12,413 | 219 | 12,071 |
| 395.1 | 1.19 [1.17-1.22] | 1.20 [1.17-1.23] | 1.20 [1.17-1.23] | 1.02 [0.85-1.21] | 1.14 [1.11-1.17] | 1.15 [1.12-1.17] | 1.12 [1.09-1.14] | 1.11 [1.09-1.14] | 0.96 [0.80-1.16] | 1.11 [1.09-1.14] | 18,964 | 18,789 | 18,269 | 326 | 17,378 |
| 395.2 | 1.22 [1.20-1.25] | 1.23 [1.20-1.26] | 1.23 [1.20-1.26] | 0.97 [0.79-1.18] | 1.17 [1.14-1.19] | 1.17 [1.15-1.20] | 1.14 [1.11-1.16] | 1.14 [1.11-1.16] | 0.97 [0.79-1.19] | 1.13 [1.10-1.16] | 19,913 | 19,749 | 19,498 | 249 | 18,136 |
| 395.3 | 1.23 [1.12-1.34] | 1.21 [1.11-1.32] | 1.24 [1.13-1.35] | | 1.17 [1.07-1.28] | 1.16 [1.06-1.26] | 1.12 [1.02-1.22] | 1.13 [1.04-1.24] | | 1.13 [1.04-1.24] | 1,417 | 1,373 | 1,313 | NA | 1,334 |
| 395.4 | 1.22 [1.12-1.32] | 1.26 [1.15-1.37] | 1.29 [1.18-1.41] | 0.95 [0.75-1.20] | 1.13 [1.04-1.23] | 1.15 [1.06-1.25] | 1.17 [1.07-1.27] | 1.17 [1.07-1.28] | 0.94 [0.74-1.20] | 1.10 [1.01-1.20] | 1,558 | 1,448 | 1,318 | 167 | 1,463 |
| 395.6 | 1.21 [1.17-1.26] | 1.23 [1.19-1.28] | 1.22 [1.17-1.27] | 0.85 [0.63-1.15] | 1.11 [1.06-1.15] | 1.17 [1.12-1.21] | 1.15 [1.10-1.20] | 1.13 [1.08-1.18] | 0.85 [0.63-1.16] | 1.08 [1.04-1.13] | 6,087 | 6,033 | 5,856 | 108 | 5,680 |
| 396 | 1.21 [1.19-1.23] | 1.22 [1.20-1.24] | 1.22 [1.20-1.24] | 1.16 [1.12-1.20] | 1.14 [1.13-1.16] | 1.14 [1.13-1.16] | 1.11 [1.09-1.13] | 1.12 [1.10-1.13] | 1.12 [1.08-1.16] | 1.10 [1.09-1.12] | 51,043 | 45,453 | 36,501 | 9,356 | 45,151 |
| 401.1 | 1.24 [1.23-1.25] | 1.24 [1.23-1.25] | 1.24 [1.23-1.25] | 1.21 [1.12-1.31] | 1.17 [1.16-1.18] | 1.19 [1.18-1.20] | 1.13 [1.12-1.14] | 1.12 [1.11-1.13] | 1.17 [1.08-1.27] | 1.14 [1.13-1.15] | 207,382 | 206,548 | 200,764 | 1,786 | 153,264 |
| 401.2 | 1.30 [1.13-1.49] | 1.29 [1.12-1.48] | 1.28 [1.11-1.47] | | 1.17 [1.02-1.35] | 1.22 [1.06-1.40] | 1.15 [1.00-1.33] | 1.14 [0.99-1.31] | | 1.14 [0.99-1.31] | 579 | 577 | 573 | NA | 561 |
| 401.21 | 1.21 [1.10-1.33] | 1.21 [1.10-1.33] | 1.22 [1.11-1.35] | | 1.06 [0.96-1.17] | 1.15 [1.04-1.27] | 1.08 [0.98-1.19] | 1.10 [0.99-1.21] | | 1.04 [0.94-1.15] | 1,103 | 1,092 | 1,084 | NA | 972 |
| 401.22 | 1.29 [1.26-1.33] | 1.30 [1.27-1.34] | 1.31 [1.27-1.35] | | 1.19 [1.16-1.23] | 1.25 [1.21-1.29] | 1.19 [1.15-1.23] | 1.19 [1.16-1.23] | | 1.18 [1.14-1.22] | 11,437 | 11,390 | 11,255 | NA | 10,333 |
| 401.3 | 1.26 [1.15-1.39] | 1.29 [1.17-1.42] | 1.26 [1.14-1.40] | 1.03 [0.81-1.32] | 1.12 [1.02-1.24] | 1.22 [1.11-1.34] | 1.17 [1.05-1.30] | 1.13 [1.02-1.25] | 1.05 [0.81-1.35] | 1.12 [1.01-1.23] | 1,158 | 1,009 | 950 | 166 | 1,068 |
| 402 | 1.16 [1.11-1.20] | 1.16 [1.12-1.21] | 1.20 [1.14-1.25] | 1.13 [1.03-1.24] | 1.10 [1.06-1.15] | 1.11 [1.07-1.16] | 1.10 [1.06-1.15] | 1.13 [1.08-1.19] | 1.11 [1.01-1.22] | 1.07 [1.03-1.12] | 7,371 | 6,868 | 4,612 | 1,282 | 6,126 |
| 411.1 | 1.29 [1.26-1.33] | 1.29 [1.25-1.33] | 1.29 [1.25-1.32] | | 1.17 [1.13-1.20] | 1.21 [1.17-1.24] | 1.15 [1.11-1.18] | 1.14 [1.10-1.17] | | 1.12 [1.08-1.15] | 12,002 | 12,009 | 11,890 | NA | 10,376 |

### Supplementary Table 6: phecode-mapped results (hazard ratios and events from all cohorts)

| | Hazard ratio (99% confidence interval) | | | | | | | | | | Events (in exposed) | | | | |
|---|---|---|---|---|---|---|---|---|---|---|---|---|---|---|---|
| | crude | | | | | adjusted | | | | | | | | | |
| Outcome | any age | 18+ | 40+ | <18 | hosp. | any age | 18+ | 40+ | <18 | hosp. | any age | 18+ | 40+ | <18 | hosp. |
| 411.2 | 1.19 [1.17-1.20] | 1.18 [1.17-1.20] | 1.19 [1.17-1.20] | 1.07 [0.79-1.45] | 1.12 [1.10-1.14] | 1.14 [1.12-1.16] | 1.09 [1.07-1.10] | 1.09 [1.07-1.10] | 1.11 [0.81-1.51] | 1.10 [1.08-1.11] | 45,699 | 45,672 | 45,192 | 109 | 39,874 |
| 411.3 | 1.28 [1.26-1.29] | 1.27 [1.26-1.29] | 1.28 [1.26-1.30] | | 1.17 [1.15-1.19] | 1.20 [1.18-1.21] | 1.14 [1.12-1.15] | 1.14 [1.12-1.15] | | 1.12 [1.10-1.14] | 44,789 | 44,808 | 44,361 | NA | 37,819 |
| 411.4 | 1.21 [1.19-1.23] | 1.21 [1.19-1.23] | 1.21 [1.19-1.23] | 1.00 [0.74-1.34] | 1.14 [1.12-1.16] | 1.15 [1.14-1.17] | 1.11 [1.09-1.12] | 1.11 [1.09-1.13] | 0.99 [0.73-1.34] | 1.11 [1.09-1.13] | 45,156 | 45,122 | 44,671 | 115 | 37,893 |
| 411.41 | 1.11 [1.03-1.19] | 1.10 [1.02-1.18] | 1.13 [1.05-1.21] | 1.20 [0.88-1.63] | 1.06 [0.98-1.14] | 1.06 [0.99-1.14] | 1.04 [0.97-1.12] | 1.07 [0.99-1.15] | 1.15 [0.84-1.58] | 1.02 [0.95-1.10] | 1,925 | 1,855 | 1,737 | 112 | 1,686 |
| 411.8 | 1.22 [1.21-1.23] | 1.22 [1.21-1.24] | 1.22 [1.21-1.24] | 1.06 [0.81-1.37] | 1.14 [1.13-1.16] | 1.16 [1.15-1.18] | 1.11 [1.10-1.13] | 1.11 [1.10-1.12] | 1.08 [0.83-1.41] | 1.11 [1.10-1.12] | 75,749 | 75,716 | 75,062 | 144 | 64,357 |
| 411.9 | 1.24 [1.20-1.28] | 1.25 [1.21-1.29] | 1.24 [1.20-1.28] | | 1.17 [1.13-1.21] | 1.18 [1.13-1.22] | 1.13 [1.09-1.17] | 1.11 [1.07-1.15] | | 1.14 [1.10-1.18] | 8,471 | 8,469 | 8,361 | NA | 7,812 |
| 414 | 1.22 [1.19-1.26] | 1.24 [1.20-1.27] | 1.23 [1.20-1.27] | 1.09 [0.87-1.37] | 1.16 [1.13-1.19] | 1.16 [1.13-1.19] | 1.12 [1.09-1.15] | 1.11 [1.08-1.14] | 1.06 [0.84-1.34] | 1.12 [1.09-1.15] | 14,411 | 14,312 | 13,974 | 209 | 13,480 |
| 414.2 | 1.35 [1.15-1.59] | 1.39 [1.18-1.64] | 1.36 [1.16-1.60] | | 1.24 [1.05-1.46] | 1.27 [1.08-1.50] | 1.26 [1.07-1.49] | 1.19 [1.01-1.41] | | 1.19 [1.01-1.42] | 408 | 407 | 396 | NA | 370 |
| 415 | 1.13 [1.10-1.16] | 1.12 [1.09-1.15] | 1.11 [1.08-1.14] | 1.01 [0.84-1.20] | 1.07 [1.04-1.10] | 1.08 [1.05-1.11] | 1.05 [1.02-1.08] | 1.04 [1.01-1.07] | 0.97 [0.81-1.16] | 1.04 [1.01-1.07] | 14,034 | 13,958 | 13,043 | 322 | 12,255 |
| 415.11 | 1.13 [1.10-1.16] | 1.12 [1.09-1.15] | 1.12 [1.09-1.15] | 1.07 [0.88-1.29] | 1.07 [1.04-1.10] | 1.08 [1.05-1.11] | 1.05 [1.02-1.08] | 1.04 [1.01-1.07] | 1.03 [0.84-1.26] | 1.04 [1.01-1.07] | 14,114 | 14,073 | 13,190 | 277 | 12,322 |
| 415.2 | 1.44 [1.34-1.55] | 1.44 [1.34-1.55] | 1.42 [1.33-1.53] | | 1.35 [1.26-1.46] | 1.30 [1.21-1.40] | 1.25 [1.16-1.34] | 1.21 [1.12-1.30] | | 1.24 [1.15-1.34] | 2,022 | 2,013 | 1,981 | NA | 1,894 |
| 415.21 | 1.36 [1.28-1.44] | 1.33 [1.25-1.41] | 1.32 [1.24-1.40] | | 1.25 [1.18-1.33] | 1.28 [1.21-1.36] | 1.21 [1.14-1.28] | 1.20 [1.13-1.27] | | 1.20 [1.13-1.28] | 2,975 | 2,949 | 2,858 | NA | 2,740 |
| 416 | 1.27 [1.24-1.29] | 1.26 [1.24-1.29] | 1.26 [1.24-1.29] | 1.11 [0.96-1.28] | 1.20 [1.17-1.22] | 1.20 [1.18-1.23] | 1.15 [1.13-1.18] | 1.15 [1.13-1.18] | 1.07 [0.92-1.25] | 1.16 [1.14-1.19] | 25,068 | 24,757 | 24,144 | 504 | 23,402 |
| 418 | 1.30 [1.28-1.32] | 1.31 [1.30-1.33] | 1.32 [1.30-1.33] | 1.13 [1.08-1.17] | 1.19 [1.17-1.20] | 1.20 [1.18-1.21] | 1.16 [1.15-1.18] | 1.16 [1.15-1.18] | 1.09 [1.04-1.13] | 1.13 [1.11-1.14] | 68,204 | 65,789 | 53,445 | 6,513 | 57,864 |
| 418.1 | 1.32 [1.28-1.35] | 1.31 [1.28-1.34] | 1.32 [1.28-1.36] | 1.13 [1.02-1.26] | 1.21 [1.18-1.24] | 1.21 [1.17-1.24] | 1.14 [1.11-1.17] | 1.14 [1.11-1.18] | 1.12 [1.00-1.24] | 1.14 [1.11-1.17] | 14,587 | 14,318 | 12,440 | 977 | 13,330 |
| 420.1 | 1.30 [1.14-1.49] | 1.25 [1.09-1.44] | 1.41 [1.19-1.67] | 1.05 [0.76-1.44] | 1.22 [1.05-1.41] | 1.23 [1.07-1.42] | 1.14 [0.98-1.31] | 1.29 [1.09-1.54] | 1.08 [0.78-1.50] | 1.17 [1.01-1.36] | 530 | 496 | 351 | 102 | 453 |
| 420.2 | 1.22 [1.17-1.28] | 1.23 [1.18-1.28] | 1.22 [1.17-1.28] | 1.02 [0.88-1.18] | 1.13 [1.08-1.18] | 1.16 [1.12-1.22] | 1.13 [1.08-1.18] | 1.13 [1.08-1.18] | 0.99 [0.85-1.16] | 1.10 [1.05-1.15] | 5,741 | 5,523 | 4,918 | 442 | 5,058 |
| 420.21 | 1.24 [1.09-1.43] | 1.20 [1.04-1.38] | 1.28 [1.09-1.50] | | 1.18 [1.01-1.37] | 1.20 [1.04-1.38] | 1.13 [0.98-1.31] | 1.20 [1.02-1.41] | | 1.15 [0.99-1.34] | 508 | 477 | 360 | NA | 415 |
| 420.22 | 1.27 [1.07-1.50] | 1.32 [1.11-1.57] | 1.33 [1.11-1.59] | | 1.22 [1.02-1.45] | 1.22 [1.03-1.45] | 1.21 [1.01-1.44] | 1.22 [1.01-1.46] | | 1.17 [0.98-1.39] | 341 | 318 | 289 | NA | 314 |
| 420.3 | 1.30 [1.24-1.37] | 1.32 [1.25-1.38] | 1.32 [1.25-1.39] | 1.12 [0.91-1.38] | 1.21 [1.15-1.28] | 1.25 [1.19-1.31] | 1.22 [1.16-1.28] | 1.23 [1.16-1.29] | 1.10 [0.89-1.36] | 1.18 [1.12-1.24] | 4,356 | 4,164 | 3,977 | 235 | 3,937 |
| 425.1 | 1.21 [1.16-1.27] | 1.23 [1.18-1.29] | 1.22 [1.16-1.28] | 1.17 [0.96-1.43] | 1.15 [1.10-1.20] | 1.15 [1.10-1.20] | 1.12 [1.07-1.17] | 1.10 [1.05-1.16] | 1.18 [0.95-1.45] | 1.11 [1.06-1.16] | 4,772 | 4,639 | 4,297 | 242 | 4,273 |
| 425.11 | 1.26 [1.10-1.46] | 1.31 [1.14-1.52] | 1.27 [1.09-1.48] | | 1.13 [0.97-1.31] | 1.21 [1.05-1.40] | 1.20 [1.03-1.39] | 1.17 [1.00-1.36] | | 1.11 [0.95-1.29] | 473 | 459 | 422 | NA | 420 |
| 425.12 | 1.16 [1.03-1.32] | 1.21 [1.06-1.37] | 1.19 [1.04-1.36] | | 1.08 [0.95-1.23] | 1.11 [0.98-1.26] | 1.10 [0.97-1.26] | 1.09 [0.95-1.25] | | 1.05 [0.93-1.20] | 640 | 607 | 554 | NA | 586 |
| 425.2 | 0.98 [0.78-1.25] | 1.07 [0.85-1.36] | 0.98 [0.77-1.25] | | 0.97 [0.76-1.23] | 0.94 [0.74-1.20] | 0.99 [0.78-1.27] | 0.92 [0.72-1.18] | | 0.93 [0.73-1.18] | 178 | 172 | 160 | NA | 167 |
| 425.8 | 1.53 [1.23-1.90] | 1.50 [1.21-1.86] | 1.45 [1.17-1.81] | | 1.34 [1.07-1.69] | 1.47 [1.18-1.84] | 1.34 [1.08-1.68] | 1.30 [1.04-1.63] | | 1.34 [1.06-1.69] | 210 | 210 | 199 | NA | 181 |
| 426 | 1.18 [1.07-1.30] | 1.20 [1.09-1.33] | 1.20 [1.08-1.33] | | 1.17 [1.06-1.30] | 1.12 [1.01-1.24] | 1.11 [1.00-1.23] | 1.10 [1.00-1.23] | | 1.15 [1.04-1.27] | 1,037 | 1,020 | 995 | NA | 981 |
| 426.2 | 1.15 [1.03-1.29] | 1.17 [1.04-1.31] | 1.17 [1.05-1.31] | | 1.12 [1.00-1.26] | 1.10 [0.98-1.24] | 1.10 [0.98-1.23] | 1.09 [0.97-1.22] | | 1.10 [0.98-1.24] | 837 | 828 | 811 | NA | 795 |
| 426.21 | 1.27 [1.23-1.31] | 1.25 [1.21-1.30] | 1.27 [1.23-1.31] | | 1.20 [1.17-1.24] | 1.22 [1.18-1.26] | 1.17 [1.14-1.21] | 1.18 [1.14-1.22] | | 1.18 [1.14-1.22] | 10,840 | 10,780 | 10,661 | NA | 10,115 |
| 426.23 | 1.18 [1.11-1.25] | 1.20 [1.13-1.28] | 1.20 [1.13-1.27] | | 1.15 [1.08-1.23] | 1.13 [1.07-1.20] | 1.12 [1.06-1.19] | 1.12 [1.06-1.20] | | 1.13 [1.06-1.20] | 2,963 | 2,939 | 2,877 | NA | 2,714 |
| 426.24 | 1.13 [1.08-1.19] | 1.15 [1.09-1.20] | 1.15 [1.09-1.20] | | 1.08 [1.03-1.13] | 1.10 [1.05-1.16] | 1.09 [1.04-1.14] | 1.09 [1.04-1.14] | | 1.07 [1.01-1.12] | 4,628 | 4,597 | 4,547 | NA | 4,207 |
| 426.25 | 1.23 [1.08-1.40] | 1.24 [1.09-1.41] | 1.21 [1.06-1.39] | | 1.13 [0.99-1.30] | 1.19 [1.04-1.36] | 1.18 [1.03-1.35] | 1.15 [1.01-1.32] | | 1.10 [0.96-1.27] | 606 | 599 | 585 | NA | 546 |
| 426.3 | 1.22 [1.15-1.29] | 1.22 [1.15-1.30] | 1.21 [1.14-1.29] | | 1.17 [1.11-1.25] | 1.17 [1.10-1.24] | 1.14 [1.08-1.21] | 1.13 [1.06-1.20] | | 1.14 [1.07-1.21] | 3,251 | 3,247 | 3,221 | NA | 3,027 |
| 426.31 | 1.21 [1.18-1.24] | 1.21 [1.18-1.24] | 1.20 [1.17-1.24] | 1.03 [0.88-1.20] | 1.16 [1.13-1.19] | 1.15 [1.12-1.18] | 1.12 [1.09-1.15] | 1.11 [1.08-1.14] | 0.99 [0.84-1.17] | 1.12 [1.09-1.15] | 14,848 | 14,703 | 14,078 | 420 | 13,663 |
| 426.32 | 1.19 [1.16-1.22] | 1.19 [1.16-1.22] | 1.19 [1.16-1.22] | | 1.14 [1.11-1.17] | 1.14 [1.11-1.17] | 1.10 [1.07-1.13] | 1.10 [1.07-1.13] | | 1.11 [1.08-1.14] | 15,083 | 15,061 | 14,924 | NA | 13,937 |
| 426.4 | 1.16 [1.02-1.33] | 1.16 [1.01-1.35] | 1.21 [1.01-1.46] | 1.06 [0.85-1.33] | 1.07 [0.92-1.24] | 1.11 [0.97-1.27] | 1.09 [0.94-1.27] | 1.15 [0.95-1.39] | 1.01 [0.80-1.27] | 1.03 [0.88-1.20] | 550 | 434 | 257 | 207 | 425 |
| 426.8 | 1.11 [0.94-1.32] | 1.16 [0.97-1.39] | 1.21 [1.00-1.46] | | 1.02 [0.86-1.22] | 1.04 [0.88-1.24] | 1.01 [0.84-1.22] | 1.08 [0.88-1.31] | | 0.98 [0.82-1.17] | 347 | 311 | 282 | NA | 323 |
| 426.9 | 1.39 [1.24-1.56] | 1.45 [1.29-1.62] | 1.40 [1.25-1.57] | | 1.23 [1.10-1.38] | 1.28 [1.14-1.43] | 1.25 [1.11-1.40] | 1.20 [1.06-1.35] | | 1.18 [1.05-1.32] | 779 | 769 | 753 | NA | 751 |
| 426.91 | 1.20 [1.17-1.23] | 1.22 [1.19-1.25] | 1.21 [1.18-1.24] | 1.10 [0.86-1.41] | 1.13 [1.10-1.16] | 1.15 [1.12-1.18] | 1.12 [1.09-1.15] | 1.11 [1.08-1.14] | 1.08 [0.83-1.39] | 1.10 [1.07-1.13] | 16,208 | 16,143 | 15,877 | 166 | 15,540 |
| 427 | 1.18 [1.07-1.30] | 1.21 [1.09-1.33] | 1.18 [1.07-1.31] | | 1.11 [1.01-1.23] | 1.12 [1.01-1.23] | 1.11 [1.00-1.22] | 1.09 [0.98-1.21] | | 1.07 [0.97-1.19] | 1,087 | 1,035 | 940 | NA | 986 |
| 427.1 | 1.22 [1.19-1.26] | 1.22 [1.18-1.26] | 1.22 [1.18-1.26] | 1.16 [1.03-1.31] | 1.14 [1.10-1.18] | 1.15 [1.12-1.19] | 1.11 [1.08-1.15] | 1.11 [1.07-1.15] | 1.11 [0.98-1.25] | 1.10 [1.06-1.13] | 10,826 | 10,396 | 9,373 | 764 | 9,433 |
| 427.11 | 1.25 [1.21-1.30] | 1.25 [1.20-1.29] | 1.25 [1.20-1.30] | 1.12 [0.99-1.27] | 1.17 [1.13-1.22] | 1.17 [1.13-1.22] | 1.14 [1.09-1.18] | 1.14 [1.10-1.19] | 1.08 [0.95-1.23] | 1.12 [1.08-1.16] | 7,882 | 7,502 | 6,648 | 652 | 6,834 |
| 427.12 | 1.14 [1.08-1.20] | 1.13 [1.07-1.19] | 1.13 [1.07-1.20] | 1.26 [0.95-1.68] | 1.04 [0.98-1.10] | 1.08 [1.02-1.14] | 1.03 [0.97-1.09] | 1.03 [0.97-1.09] | 1.17 [0.87-1.58] | 1.01 [0.95-1.07] | 3,264 | 3,198 | 3,007 | 134 | 2,923 |
| 427.2 | 1.16 [1.15-1.18] | 1.16 [1.15-1.18] | 1.16 [1.15-1.18] | 1.06 [0.85-1.32] | 1.12 [1.11-1.13] | 1.13 [1.11-1.14] | 1.10 [1.09-1.11] | 1.09 [1.08-1.11] | 1.01 [0.80-1.27] | 1.10 [1.09-1.11] | 81,212 | 81,194 | 80,479 | 207 | 70,255 |
| 427.3 | 1.19 [1.16-1.21] | 1.20 [1.17-1.23] | 1.20 [1.17-1.23] | 1.08 [0.99-1.19] | 1.12 [1.09-1.15] | 1.14 [1.12-1.17] | 1.12 [1.09-1.15] | 1.12 [1.09-1.15] | 1.04 [0.95-1.15] | 1.10 [1.07-1.13] | 16,985 | 16,114 | 15,079 | 1,293 | 15,400 |
| 427.4 | 1.07 [0.98-1.17] | 1.07 [0.98-1.18] | 1.07 [0.97-1.17] | | 1.03 [0.94-1.14] | 1.04 [0.95-1.15] | 1.01 [0.92-1.11] | 0.99 [0.90-1.09] | | 1.03 [0.93-1.13] | 1,133 | 1,113 | 1,059 | NA | 1,006 |
| 427.41 | 1.07 [0.98-1.17] | 1.07 [0.98-1.18] | 1.07 [0.97-1.17] | | 1.03 [0.94-1.14] | 1.04 [0.95-1.15] | 1.01 [0.92-1.11] | 0.99 [0.90-1.09] | | 1.03 [0.93-1.13] | 1,133 | 1,113 | 1,059 | NA | 1,006 |
| 427.42 | 1.23 [1.19-1.27] | 1.24 [1.20-1.28] | 1.23 [1.19-1.27] | 1.08 [0.89-1.30] | 1.18 [1.14-1.22] | 1.19 [1.15-1.23] | 1.15 [1.11-1.19] | 1.14 [1.10-1.18] | 1.04 [0.85-1.26] | 1.17 [1.13-1.21] | 9,596 | 9,458 | 9,070 | 275 | 8,594 |
| 427.5 | 1.25 [1.19-1.32] | 1.26 [1.19-1.32] | 1.27 [1.20-1.34] | 1.28 [1.03-1.58] | 1.19 [1.13-1.25] | 1.17 [1.12-1.24] | 1.14 [1.09-1.21] | 1.16 [1.10-1.23] | 1.21 [0.97-1.51] | 1.14 [1.08-1.20] | 4,023 | 3,903 | 3,607 | 234 | 3,683 |
| 427.6 | 1.20 [1.14-1.27] | 1.20 [1.14-1.27] | 1.20 [1.13-1.27] | 1.08 [0.83-1.40] | 1.13 [1.07-1.20] | 1.13 [1.07-1.20] | 1.10 [1.04-1.16] | 1.09 [1.03-1.16] | 1.05 [0.80-1.37] | 1.08 [1.02-1.15] | 3,507 | 3,421 | 3,165 | 167 | 3,181 |
| 427.61 | 1.17 [1.03-1.32] | 1.12 [0.99-1.28] | 1.15 [1.01-1.32] | | 1.07 [0.94-1.23] | 1.07 [0.94-1.22] | 1.03 [0.90-1.17] | 1.05 [0.91-1.20] | | 1.02 [0.89-1.17] | 662 | 641 | 602 | NA | 581 |
| 427.7 | 1.22 [1.19-1.25] | 1.22 [1.20-1.25] | 1.22 [1.19-1.26] | 1.15 [1.10-1.20] | 1.15 [1.13-1.18] | 1.14 [1.12-1.17] | 1.10 [1.07-1.12] | 1.10 [1.07-1.13] | 1.11 [1.06-1.17] | 1.10 [1.08-1.13] | 20,616 | 18,181 | 12,895 | 4,809 | 18,176 |
| 427.8 | 1.12 [1.04-1.19] | 1.13 [1.06-1.21] | 1.12 [1.05-1.20] | | 1.05 [0.98-1.12] | 1.07 [1.00-1.14] | 1.05 [0.98-1.13] | 1.04 [0.97-1.11] | | 1.02 [0.95-1.09] | 2,271 | 2,274 | 2,214 | NA | 2,127 |
| 427.9 | 1.24 [1.20-1.27] | 1.25 [1.21-1.28] | 1.24 [1.20-1.28] | 1.12 [1.04-1.21] | 1.16 [1.12-1.19] | 1.14 [1.11-1.17] | 1.10 [1.07-1.14] | 1.09 [1.06-1.13] | 1.07 [0.99-1.16] | 1.09 [1.05-1.12] | 11,971 | 11,314 | 8,436 | 1,752 | 10,457 |
| 428.1 | 1.29 [1.27-1.32] | 1.29 [1.27-1.31] | 1.29 [1.27-1.31] | | 1.23 [1.21-1.25] | 1.23 [1.21-1.25] | 1.17 [1.15-1.19] | 1.17 [1.15-1.19] | | 1.18 [1.16-1.20] | 41,385 | 41,364 | 41,173 | NA | 38,454 |
| 428.2 | 1.26 [1.25-1.28] | 1.26 [1.24-1.27] | 1.26 [1.25-1.28] | 1.09 [0.90-1.33] | 1.20 [1.19-1.22] | 1.20 [1.19-1.22] | 1.15 [1.14-1.17] | 1.15 [1.14-1.17] | 1.10 [0.90-1.35] | 1.17 [1.15-1.19] | 64,824 | 64,744 | 64,197 | 270 | 58,996 |
| 429.1 | 1.17 [0.91-1.50] | 1.30 [0.99-1.71] | 1.46 [1.09-1.97] | | 1.00 [0.77-1.30] | 1.12 [0.87-1.44] | 1.19 [0.89-1.57] | 1.30 [0.96-1.77] | | 0.95 [0.72-1.24] | 147 | 126 | 108 | NA | 127 |
| 429.2 | 1.28 [1.21-1.36] | 1.28 [1.20-1.36] | 1.31 [1.23-1.40] | 1.26 [1.02-1.55] | 1.23 [1.15-1.31] | 1.21 [1.14-1.29] | 1.16 [1.09-1.23] | 1.18 [1.10-1.26] | 1.19 [0.96-1.48] | 1.20 [1.12-1.28] | 2,811 | 2,689 | 2,383 | 252 | 2,514 |
| 429.3 | 1.28 [1.17-1.41] | 1.33 [1.21-1.47] | 1.27 [1.14-1.40] | 1.29 [1.06-1.58] | 1.22 [1.11-1.33] | 1.19 [1.09-1.31] | 1.18 [1.07-1.30] | 1.14 [1.02-1.27] | 1.19 [0.96-1.46] | 1.16 [1.06-1.28] | 1,266 | 1,070 | 925 | 263 | 1,157 |
| 430 | 1.17 [1.06-1.29] | 1.19 [1.07-1.32] | 1.22 [1.09-1.37] | 0.94 [0.75-1.18] | 1.10 [0.99-1.23] | 1.15 [1.04-1.28] | 1.15 [1.03-1.28] | 1.17 [1.04-1.32] | 0.91 [0.72-1.15] | 1.11 [0.99-1.24] | 1,004 | 879 | 764 | 190 | 854 |
| 430.1 | 1.16 [1.08-1.24] | 1.16 [1.08-1.25] | 1.14 [1.06-1.22] | | 1.10 [1.02-1.18] | 1.15 [1.07-1.24] | 1.13 [1.05-1.21] | 1.11 [1.03-1.19] | | 1.10 [1.02-1.20] | 1,910 | 1,880 | 1,738 | NA | 1,556 |
| 430.2 | 1.17 [1.12-1.23] | 1.17 [1.12-1.23] | 1.17 [1.12-1.22] | 0.88 [0.65-1.19] | 1.14 [1.09-1.20] | 1.16 [1.11-1.22] | 1.14 [1.09-1.20] | 1.13 [1.08-1.19] | 0.88 [0.64-1.20] | 1.14 [1.09-1.20] | 4,912 | 4,837 | 4,715 | 104 | 4,230 |
| 430.3 | 1.14 [1.07-1.22] | 1.16 [1.09-1.24] | 1.15 [1.08-1.23] | | 1.08 [1.01-1.16] | 1.12 [1.05-1.20] | 1.12 [1.04-1.20] | 1.10 [1.03-1.18] | | 1.09 [1.01-1.17] | 2,297 | 2,267 | 2,229 | NA | 2,071 |
| 433 | 1.21 [1.18-1.25] | 1.20 [1.17-1.24] | 1.22 [1.18-1.25] | | 1.15 [1.12-1.19] | 1.17 [1.13-1.20] | 1.12 [1.09-1.15] | 1.12 [1.09-1.16] | | 1.13 [1.10-1.17] | 12,642 | 12,611 | 12,487 | NA | 11,809 |
| 433.1 | 1.17 [1.13-1.22] | 1.17 [1.12-1.22] | 1.17 [1.12-1.22] | | 1.11 [1.06-1.16] | 1.14 [1.10-1.19] | 1.09 [1.04-1.14] | 1.08 [1.04-1.13] | | 1.10 [1.05-1.15] | 5,593 | 5,582 | 5,554 | NA | 4,973 |
| 433.11 | 1.10 [0.98-1.23] | 1.11 [0.99-1.24] | 1.12 [1.00-1.26] | | 1.10 [0.97-1.24] | 1.10 [0.98-1.23] | 1.08 [0.96-1.21] | 1.08 [0.96-1.22] | | 1.11 [0.98-1.25] | 775 | 772 | 759 | NA | 681 |
| 433.12 | 1.13 [1.05-1.21] | 1.16 [1.08-1.25] | 1.14 [1.06-1.23] | | 1.08 [1.00-1.16] | 1.10 [1.03-1.18] | 1.11 [1.03-1.19] | 1.07 [1.00-1.15] | | 1.07 [1.00-1.15] | 2,085 | 2,078 | 2,071 | NA | 1,934 |
| 433.2 | 1.09 [1.07-1.11] | 1.09 [1.07-1.12] | 1.09 [1.07-1.11] | 1.17 [0.93-1.47] | 1.07 [1.04-1.09] | 1.08 [1.06-1.10] | 1.05 [1.03-1.07] | 1.04 [1.02-1.07] | 1.22 [0.96-1.55] | 1.07 [1.05-1.09] | 22,286 | 22,175 | 21,851 | 191 | 19,622 |
| 433.21 | 1.11 [1.09-1.14] | 1.12 [1.09-1.15] | 1.11 [1.08-1.14] | | 1.09 [1.06-1.12] | 1.11 [1.08-1.14] | 1.07 [1.04-1.10] | 1.07 [1.04-1.10] | | 1.10 [1.07-1.13] | 14,370 | 14,329 | 14,170 | NA | 12,566 |
| 433.3 | 1.18 [1.16-1.20] | 1.18 [1.16-1.20] | 1.18 [1.16-1.20] | 1.08 [0.82-1.41] | 1.14 [1.12-1.16] | 1.15 [1.13-1.17] | 1.11 [1.09-1.13] | 1.11 [1.09-1.13] | 1.08 [0.82-1.43] | 1.13 [1.11-1.15] | 35,644 | 35,555 | 35,303 | 135 | 33,238 |
| 433.31 | 1.19 [1.15-1.23] | 1.19 [1.15-1.23] | 1.19 [1.15-1.23] | | 1.12 [1.08-1.16] | 1.14 [1.10-1.18] | 1.11 [1.07-1.15] | 1.10 [1.07-1.14] | | 1.10 [1.06-1.14] | 9,101 | 9,072 | 8,944 | NA | 8,049 |
| 433.5 | 1.24 [1.13-1.36] | 1.23 [1.12-1.35] | 1.24 [1.13-1.37] | | 1.17 [1.06-1.29] | 1.21 [1.10-1.33] | 1.14 [1.04-1.25] | 1.15 [1.05-1.27] | | 1.18 [1.07-1.30] | 1,115 | 1,101 | 1,043 | NA | 999 |
| 433.8 | 1.18 [1.15-1.22] | 1.19 [1.15-1.23] | 1.18 [1.15-1.22] | 0.94 [0.73-1.21] | 1.10 [1.06-1.13] | 1.15 [1.12-1.19] | 1.11 [1.07-1.14] | 1.10 [1.06-1.13] | 0.91 [0.70-1.17] | 1.09 [1.06-1.13] | 10,401 | 10,290 | 10,123 | 153 | 9,596 |
| 440 | 1.26 [1.22-1.30] | 1.26 [1.22-1.30] | 1.27 [1.23-1.31] | | 1.18 [1.14-1.22] | 1.22 [1.18-1.26] | 1.14 [1.10-1.18] | 1.15 [1.11-1.19] | | 1.17 [1.13-1.21] | 9,463 | 9,462 | 9,410 | NA | 8,712 |
| 440.1 | 1.38 [1.23-1.55] | 1.39 [1.24-1.56] | 1.41 [1.25-1.59] | | 1.26 [1.12-1.43] | 1.35 [1.20-1.52] | 1.29 [1.14-1.45] | 1.29 [1.14-1.46] | | 1.26 [1.12-1.43] | 741 | 736 | 717 | NA | 670 |
| 440.2 | 1.28 [1.23-1.34] | 1.28 [1.22-1.34] | 1.29 [1.23-1.35] | | 1.19 [1.14-1.25] | 1.24 [1.19-1.30] | 1.16 [1.11-1.21] | 1.16 [1.10-1.21] | | 1.19 [1.14-1.25] | 5,105 | 5,107 | 5,091 | NA | 4,665 |
| 440.9 | 1.21 [1.14-1.28] | 1.20 [1.13-1.27] | 1.23 [1.16-1.30] | | 1.13 [1.07-1.20] | 1.16 [1.10-1.23] | 1.10 [1.04-1.16] | 1.12 [1.05-1.19] | | 1.12 [1.05-1.18] | 3,247 | 3,246 | 3,234 | NA | 3,045 |
| 441 | 1.25 [1.20-1.30] | 1.25 [1.20-1.30] | 1.25 [1.20-1.31] | | 1.15 [1.10-1.21] | 1.19 [1.14-1.24] | 1.14 [1.10-1.20] | 1.14 [1.09-1.19] | | 1.13 [1.08-1.19] | 5,570 | 5,532 | 5,358 | NA | 5,112 |
| 441.1 | 1.23 [1.14-1.33] | 1.21 [1.12-1.31] | 1.21 [1.12-1.31] | | 1.13 [1.04-1.22] | 1.17 [1.08-1.26] | 1.12 [1.03-1.21] | 1.11 [1.02-1.20] | | 1.11 [1.02-1.20] | 1,671 | 1,643 | 1,568 | NA | 1,502 |
| 441.2 | 1.24 [1.05-1.45] | 1.24 [1.05-1.46] | 1.22 [1.03-1.44] | | 1.11 [0.94-1.31] | 1.17 [0.99-1.37] | 1.08 [0.91-1.28] | 1.09 [0.92-1.29] | | 1.10 [0.93-1.30] | 399 | 390 | 379 | NA | 380 |
| 442 | 1.17 [1.10-1.25] | 1.18 [1.11-1.25] | 1.18 [1.10-1.25] | | 1.11 [1.04-1.18] | 1.14 [1.07-1.21] | 1.09 [1.02-1.16] | 1.09 [1.02-1.16] | | 1.10 [1.03-1.17] | 2,555 | 2,547 | 2,443 | NA | 2,312 |
| 442.1 | 1.15 [1.09-1.22] | 1.15 [1.09-1.22] | 1.17 [1.11-1.23] | | 1.07 [1.01-1.13] | 1.12 [1.06-1.18] | 1.09 [1.03-1.15] | 1.09 [1.03-1.15] | | 1.06 [1.00-1.12] | 3,563 | 3,514 | 3,430 | NA | 3,251 |
| 442.11 | 1.19 [1.15-1.23] | 1.19 [1.15-1.24] | 1.20 [1.16-1.24] | | 1.14 [1.10-1.18] | 1.18 [1.14-1.22] | 1.13 [1.09-1.17] | 1.13 [1.09-1.17] | | 1.15 [1.11-1.19] | 8,355 | 8,345 | 8,343 | NA | 7,485 |
| 442.2 | 1.12 [0.99-1.26] | 1.11 [0.98-1.24] | 1.13 [1.01-1.28] | | 1.02 [0.90-1.15] | 1.10 [0.97-1.24] | 1.04 [0.92-1.17] | 1.06 [0.94-1.20] | | 1.02 [0.90-1.15] | 708 | 708 | 705 | NA | 633 |
| 442.3 | 1.22 [1.09-1.36] | 1.22 [1.09-1.37] | 1.21 [1.08-1.35] | | 1.14 [1.02-1.29] | 1.18 [1.05-1.32] | 1.11 [0.99-1.24] | 1.09 [0.97-1.23] | | 1.13 [1.01-1.27] | 790 | 789 | 770 | NA | 715 |
| 442.8 | 1.17 [1.06-1.30] | 1.18 [1.07-1.30] | 1.18 [1.06-1.31] | | 1.14 [1.02-1.26] | 1.14 [1.03-1.27] | 1.10 [0.99-1.22] | 1.10 [0.99-1.23] | | 1.13 [1.02-1.25] | 976 | 970 | 903 | NA | 896 |
| 443 | 1.40 [1.37-1.43] | 1.40 [1.37-1.43] | 1.39 [1.36-1.42] | 1.44 [1.24-1.68] | 1.29 [1.26-1.32] | 1.32 [1.29-1.35] | 1.24 [1.21-1.27] | 1.23 [1.20-1.26] | 1.35 [1.15-1.58] | 1.25 [1.22-1.28] | 21,801 | 21,629 | 20,652 | 481 | 19,729 |
| 443.1 | 1.59 [1.52-1.66] | 1.56 [1.49-1.63] | 1.59 [1.51-1.67] | 1.51 [1.29-1.78] | 1.46 [1.40-1.53] | 1.43 [1.37-1.50] | 1.35 [1.29-1.42] | 1.38 [1.32-1.46] | 1.41 [1.19-1.68] | 1.33 [1.27-1.40] | 5,376 | 5,213 | 4,374 | 438 | 4,803 |
| 443.7 | 1.39 [1.33-1.47] | 1.39 [1.32-1.46] | 1.38 [1.31-1.45] | | 1.30 [1.23-1.37] | 1.32 [1.25-1.39] | 1.20 [1.14-1.26] | 1.18 [1.12-1.24] | | 1.27 [1.21-1.34] | 3,937 | 3,934 | 3,870 | NA | 3,675 |
| 443.8 | 1.40 [1.37-1.43] | 1.40 [1.37-1.43] | 1.39 [1.36-1.42] | 1.44 [1.24-1.68] | 1.29 [1.26-1.32] | 1.32 [1.29-1.35] | 1.24 [1.21-1.27] | 1.23 [1.20-1.26] | 1.35 [1.15-1.58] | 1.25 [1.22-1.28] | 21,801 | 21,629 | 20,652 | 481 | 19,729 |
| 443.9 | 1.35 [1.32-1.39] | 1.36 [1.32-1.39] | 1.35 [1.32-1.38] | | 1.25 [1.22-1.28] | 1.29 [1.26-1.32] | 1.21 [1.18-1.24] | 1.20 [1.17-1.23] | | 1.22 [1.19-1.26] | 16,835 | 16,835 | 16,695 | NA | 15,327 |
| 444 | 1.18 [1.13-1.23] | 1.17 [1.12-1.22] | 1.19 [1.14-1.25] | | 1.11 [1.06-1.17] | 1.14 [1.09-1.19] | 1.07 [1.02-1.12] | 1.09 [1.04-1.14] | | 1.11 [1.06-1.17] | 4,919 | 4,896 | 4,775 | NA | 4,434 |
| 444.1 | 1.19 [1.12-1.26] | 1.18 [1.12-1.25] | 1.21 [1.14-1.28] | | 1.12 [1.05-1.18] | 1.15 [1.08-1.21] | 1.08 [1.02-1.14] | 1.09 [1.03-1.16] | | 1.12 [1.05-1.18] | 3,172 | 3,161 | 3,123 | NA | 2,865 |
| 444.2 | 1.12 [0.92-1.37] | 1.10 [0.90-1.34] | 1.21 [0.99-1.48] | | 1.03 [0.83-1.26] | 1.12 [0.91-1.37] | 1.01 [0.83-1.24] | 1.12 [0.91-1.38] | | 1.06 [0.86-1.31] | 249 | 249 | 248 | NA | 228 |
| 446 | 1.39 [1.21-1.59] | 1.47 [1.19-1.81] | 1.41 [1.12-1.76] | 1.28 [1.07-1.53] | 1.31 [1.13-1.52] | 1.28 [1.12-1.48] | 1.23 [0.99-1.53] | 1.17 [0.92-1.48] | 1.29 [1.07-1.55] | 1.23 [1.06-1.43] | 525 | 229 | 193 | 309 | 456 |
| 446.2 | 1.26 [1.05-1.51] | | | 1.24 [1.03-1.49] | 1.24 [1.02-1.51] | 1.27 [1.05-1.53] | | | 1.26 [1.04-1.52] | 1.26 [1.03-1.54] | 294 | NA | NA | 288 | 261 |
| 446.3 | 1.52 [1.16-1.98] | 1.52 [1.15-2.00] | 1.48 [1.11-1.97] | | 1.16 [0.87-1.55] | 1.51 [1.16-1.98] | 1.47 [1.11-1.94] | 1.42 [1.06-1.89] | | 1.15 [0.86-1.54] | 140 | 134 | 120 | NA | 116 |
| 446.4 | 1.53 [1.29-1.80] | 1.50 [1.27-1.77] | 1.56 [1.31-1.86] | | 1.44 [1.21-1.72] | 1.37 [1.16-1.63] | 1.32 [1.11-1.57] | 1.36 [1.13-1.64] | | 1.30 [1.08-1.56] | 373 | 365 | 318 | NA | 308 |
| 446.5 | 1.31 [1.22-1.40] | 1.31 [1.23-1.41] | 1.30 [1.22-1.40] | | 1.23 [1.15-1.33] | 1.22 [1.14-1.31] | 1.21 [1.12-1.30] | 1.19 [1.11-1.28] | | 1.16 [1.07-1.25] | 2,049 | 2,051 | 2,043 | NA | 1,841 |
| 446.6 | 1.49 [1.14-1.94] | 1.47 [1.12-1.93] | 1.44 [1.07-1.92] | | 1.30 [0.97-1.74] | 1.40 [1.07-1.83] | 1.37 [1.04-1.81] | 1.30 [0.97-1.76] | | 1.23 [0.92-1.66] | 141 | 131 | 115 | NA | 118 |
| 446.8 | 1.30 [0.96-1.75] | 1.44 [1.05-1.98] | | | | 1.24 [0.91-1.68] | 1.29 [0.93-1.78] | | | | 113 | 102 | NA | NA | NA |
| 446.9 | 1.59 [1.45-1.74] | 1.55 [1.41-1.70] | 1.52 [1.38-1.67] | | 1.42 [1.30-1.56] | 1.47 [1.34-1.61] | 1.41 [1.28-1.54] | 1.37 [1.24-1.51] | | 1.34 [1.22-1.48] | 1,278 | 1,236 | 1,120 | NA | 1,148 |
| 447 | 1.30 [1.26-1.35] | 1.32 [1.27-1.37] | 1.33 [1.28-1.38] | 1.06 [0.87-1.29] | 1.21 [1.16-1.26] | 1.25 [1.20-1.30] | 1.19 [1.15-1.24] | 1.20 [1.15-1.25] | 1.03 [0.84-1.27] | 1.20 [1.15-1.24] | 7,470 | 7,317 | 7,026 | 253 | 6,714 |
| 447.1 | 1.30 [1.24-1.36] | 1.30 [1.24-1.36] | 1.32 [1.26-1.38] | | 1.19 [1.14-1.25] | 1.25 [1.19-1.31] | 1.16 [1.11-1.22] | 1.18 [1.13-1.24] | | 1.20 [1.14-1.26] | 4,377 | 4,368 | 4,339 | NA | 3,969 |
| 448 | 1.30 [1.21-1.40] | 1.35 [1.25-1.45] | 1.31 [1.21-1.42] | 1.03 [0.77-1.36] | 1.24 [1.15-1.34] | 1.24 [1.15-1.34] | 1.24 [1.14-1.34] | 1.21 [1.12-1.31] | 1.01 [0.76-1.35] | 1.21 [1.12-1.31] | 1,829 | 1,727 | 1,615 | 121 | 1,655 |
| 450 | 1.52 [1.46-1.59] | 1.52 [1.46-1.59] | 1.50 [1.44-1.57] | 1.15 [0.91-1.47] | 1.41 [1.35-1.48] | 1.43 [1.37-1.49] | 1.37 [1.31-1.44] | 1.36 [1.30-1.42] | 1.08 [0.84-1.38] | 1.37 [1.31-1.43] | 5,602 | 5,490 | 5,228 | 177 | 5,193 |
| 451 | 1.25 [1.22-1.29] | 1.24 [1.21-1.28] | 1.25 [1.21-1.28] | 1.15 [0.98-1.36] | 1.19 [1.15-1.22] | 1.20 [1.17-1.23] | 1.16 [1.12-1.19] | 1.16 [1.13-1.20] | 1.11 [0.93-1.32] | 1.15 [1.12-1.19] | 13,733 | 13,610 | 12,516 | 371 | 11,862 |
| 451.2 | 1.24 [1.21-1.27] | 1.23 [1.20-1.26] | 1.24 [1.21-1.28] | 1.05 [0.87-1.27] | 1.18 [1.14-1.21] | 1.19 [1.15-1.22] | 1.15 [1.12-1.18] | 1.16 [1.13-1.20] | 1.01 [0.82-1.23] | 1.14 [1.11-1.18] | 12,447 | 12,365 | 11,446 | 279 | 10,705 |
| 452 | 1.18 [1.11-1.26] | 1.19 [1.12-1.27] | 1.17 [1.09-1.25] | 0.95 [0.74-1.21] | 1.09 [1.02-1.17] | 1.13 [1.06-1.20] | 1.11 [1.04-1.18] | 1.08 [1.01-1.16] | 0.94 [0.73-1.21] | 1.07 [1.00-1.15] | 2,397 | 2,320 | 2,043 | 157 | 2,130 |
| 454 | 1.31 [1.22-1.41] | 1.36 [1.26-1.47] | 1.36 [1.24-1.49] | 1.03 [0.88-1.21] | 1.24 [1.15-1.34] | 1.22 [1.14-1.32] | 1.22 [1.13-1.32] | 1.21 [1.10-1.33] | 0.99 [0.84-1.17] | 1.21 [1.12-1.31] | 1,918 | 1,691 | 1,175 | 412 | 1,611 |
| 454.1 | 1.63 [1.59-1.67] | 1.63 [1.59-1.67] | 1.68 [1.64-1.72] | 1.05 [0.86-1.27] | 1.52 [1.48-1.56] | 1.57 [1.54-1.61] | 1.55 [1.52-1.59] | 1.59 [1.55-1.63] | 1.01 [0.82-1.24] | 1.48 [1.45-1.52] | 19,256 | 19,220 | 17,136 | 254 | 15,892 |
| 454.11 | 2.21 [2.12-2.30] | 2.19 [2.11-2.28] | 2.19 [2.10-2.27] | | 2.05 [1.97-2.14] | 2.14 [2.06-2.23] | 2.07 [1.99-2.15] | 2.05 [1.97-2.14] | | 2.02 [1.93-2.10] | 8,205 | 8,205 | 7,948 | NA | 7,356 |
| 456 | 1.88 [1.76-2.01] | 1.85 [1.73-1.98] | 1.86 [1.74-1.99] | | 1.75 [1.63-1.87] | 1.78 [1.66-1.90] | 1.69 [1.58-1.81] | 1.71 [1.59-1.83] | | 1.69 [1.58-1.81] | 2,673 | 2,670 | 2,587 | NA | 2,497 |
| 458 | 1.21 [1.17-1.25] | 1.22 [1.17-1.26] | 1.21 [1.16-1.26] | 1.10 [0.94-1.29] | 1.13 [1.09-1.18] | 1.17 [1.12-1.21] | 1.13 [1.09-1.18] | 1.13 [1.08-1.17] | 1.08 [0.92-1.27] | 1.12 [1.07-1.16] | 7,942 | 7,770 | 6,977 | 443 | 7,319 |
| 458.1 | 1.21 [1.18-1.24] | 1.21 [1.18-1.24] | 1.21 [1.18-1.24] | 1.12 [0.90-1.38] | 1.15 [1.12-1.18] | 1.16 [1.13-1.19] | 1.11 [1.09-1.14] | 1.12 [1.09-1.15] | 1.04 [0.83-1.30] | 1.12 [1.10-1.15] | 17,005 | 16,905 | 16,578 | 233 | 16,016 |
| 458.2 | 1.23 [1.14-1.32] | 1.27 [1.18-1.37] | 1.25 [1.16-1.34] | | 1.20 [1.11-1.29] | 1.18 [1.10-1.27] | 1.17 [1.08-1.26] | 1.14 [1.05-1.23] | | 1.17 [1.09-1.26] | 1,976 | 1,972 | 1,937 | NA | 1,851 |
| 458.9 | 1.20 [1.18-1.22] | 1.21 [1.19-1.23] | 1.20 [1.19-1.22] | 1.06 [0.97-1.16] | 1.15 [1.13-1.16] | 1.16 [1.14-1.17] | 1.12 [1.11-1.14] | 1.12 [1.10-1.14] | 1.04 [0.95-1.14] | 1.13 [1.11-1.15] | 44,151 | 43,521 | 41,490 | 1,343 | 41,026 |
| 459 | 1.20 [1.07-1.34] | 1.22 [1.09-1.37] | 1.25 [1.10-1.41] | | 1.11 [0.99-1.25] | 1.17 [1.04-1.31] | 1.14 [1.02-1.29] | 1.17 [1.03-1.32] | | 1.11 [0.98-1.26] | 745 | 707 | 634 | NA | 661 |
| 459.1 | 1.35 [1.19-1.53] | 1.34 [1.17-1.52] | 1.34 [1.17-1.54] | | 1.30 [1.14-1.48] | 1.30 [1.14-1.48] | 1.22 [1.07-1.40] | 1.22 [1.06-1.41] | | 1.29 [1.13-1.48] | 644 | 582 | 533 | NA | 597 |
| 459.9 | 1.25 [1.23-1.26] | 1.25 [1.24-1.26] | 1.25 [1.24-1.27] | 1.03 [0.96-1.11] | 1.15 [1.14-1.17] | 1.18 [1.17-1.20] | 1.14 [1.13-1.15] | 1.14 [1.13-1.15] | 0.99 [0.92-1.07] | 1.12 [1.11-1.14] | 82,816 | 81,957 | 77,655 | 1,925 | 75,883 |
| respiratory | | | | | | | | | | | | | | | |
| 464 | 1.46 [1.31-1.63] | 1.38 [1.21-1.56] | 1.38 [1.18-1.62] | 1.44 [1.19-1.74] | 1.28 [1.14-1.44] | 1.31 [1.17-1.47] | 1.18 [1.03-1.35] | 1.16 [0.99-1.37] | 1.36 [1.12-1.66] | 1.19 [1.05-1.34] | 851 | 617 | 388 | 306 | 714 |
| 465 | 1.30 [1.28-1.33] | 1.36 [1.30-1.42] | 1.36 [1.29-1.43] | 1.29 [1.27-1.32] | 1.25 [1.22-1.27] | 1.25 [1.22-1.27] | 1.18 [1.13-1.24] | 1.19 [1.13-1.26] | 1.25 [1.22-1.28] | 1.20 [1.18-1.23] | 25,551 | 5,142 | 3,385 | 21,050 | 22,206 |
| 465.2 | 1.32 [1.27-1.38] | 1.39 [1.31-1.46] | 1.44 [1.34-1.54] | 1.19 [1.13-1.26] | 1.23 [1.18-1.28] | 1.24 [1.19-1.29] | 1.24 [1.17-1.31] | 1.26 [1.18-1.36] | 1.15 [1.09-1.22] | 1.18 [1.13-1.23] | 6,206 | 3,525 | 2,082 | 3,263 | 5,339 |
| 465.4 | 1.34 [1.19-1.51] | 1.47 [1.28-1.68] | 1.41 [1.21-1.65] | 1.32 [1.07-1.63] | 1.33 [1.17-1.50] | 1.23 [1.09-1.39] | 1.29 [1.12-1.48] | 1.19 [1.01-1.40] | 1.29 [1.05-1.60] | 1.23 [1.08-1.40] | 724 | 538 | 396 | 242 | 633 |
| 470 | 1.55 [1.50-1.59] | 1.53 [1.48-1.58] | 1.52 [1.45-1.59] | 1.56 [1.48-1.64] | 1.42 [1.37-1.47] | 1.38 [1.33-1.42] | 1.29 [1.25-1.34] | 1.29 [1.23-1.35] | 1.44 [1.36-1.52] | 1.28 [1.24-1.33] | 11,262 | 8,889 | 4,277 | 4,190 | 8,388 |
| 471 | 1.56 [1.49-1.64] | 1.54 [1.47-1.62] | 1.52 [1.44-1.61] | 1.41 [1.21-1.64] | 1.42 [1.35-1.50] | 1.29 [1.23-1.36] | 1.25 [1.19-1.32] | 1.25 [1.18-1.32] | 1.21 [1.03-1.42] | 1.20 [1.14-1.27] | 4,446 | 4,224 | 3,253 | 471 | 3,366 |
| 472 | 1.75 [1.66-1.86] | 1.70 [1.59-1.81] | 1.59 [1.47-1.72] | 1.96 [1.78-2.17] | 1.62 [1.52-1.72] | 1.44 [1.35-1.53] | 1.33 [1.24-1.43] | 1.24 [1.14-1.34] | 1.68 [1.52-1.87] | 1.34 [1.26-1.43] | 3,383 | 2,421 | 1,625 | 1,259 | 2,875 |
| 473 | 1.35 [1.28-1.42] | 1.37 [1.30-1.45] | 1.35 [1.27-1.43] | 1.23 [1.06-1.41] | 1.24 [1.18-1.31] | 1.24 [1.18-1.31] | 1.18 [1.12-1.25] | 1.18 [1.11-1.25] | 1.12 [0.96-1.29] | 1.16 [1.10-1.23] | 3,700 | 3,315 | 2,863 | 499 | 3,171 |
| 473.1 | 1.39 [1.07-1.81] | 1.40 [1.07-1.84] | 1.31 [0.98-1.75] | | 1.19 [0.90-1.58] | 1.24 [0.95-1.63] | 1.15 [0.86-1.53] | 1.10 [0.81-1.49] | | 1.09 [0.82-1.45] | 143 | 129 | 108 | NA | 118 |
| 473.3 | 1.19 [1.09-1.30] | 1.19 [1.08-1.31] | 1.22 [1.10-1.35] | 1.17 [0.94-1.47] | 1.14 [1.03-1.25] | 1.13 [1.03-1.24] | 1.10 [0.99-1.22] | 1.14 [1.03-1.27] | 1.10 [0.88-1.39] | 1.12 [1.02-1.23] | 1,165 | 1,007 | 912 | 198 | 1,063 |
| 473.4 | 1.35 [1.27-1.44] | 1.35 [1.27-1.44] | 1.32 [1.24-1.42] | 1.25 [1.05-1.48] | 1.23 [1.15-1.31] | 1.23 [1.16-1.32] | 1.17 [1.10-1.26] | 1.14 [1.07-1.23] | 1.20 [1.01-1.42] | 1.15 [1.08-1.23] | 2,591 | 2,269 | 2,050 | 364 | 2,279 |
| 474 | 1.14 [1.07-1.22] | 1.16 [1.08-1.24] | 1.14 [1.00-1.30] | 1.11 [1.01-1.22] | 1.07 [1.00-1.15] | 1.11 [1.04-1.18] | 1.11 [1.03-1.19] | 1.09 [0.95-1.25] | 1.10 [1.00-1.21] | 1.06 [0.99-1.15] | 2,461 | 1,982 | 525 | 1,193 | 1,763 |
| 474.1 | 1.22 [1.20-1.24] | 1.31 [1.27-1.36] | 1.25 [1.14-1.36] | 1.20 [1.17-1.22] | 1.17 [1.15-1.19] | 1.17 [1.15-1.19] | 1.17 [1.13-1.21] | 1.08 [0.98-1.19] | 1.16 [1.14-1.18] | 1.14 [1.11-1.16] | 32,053 | 8,122 | 1,121 | 27,861 | 26,714 |
| 474.2 | 1.28 [1.26-1.31] | 1.30 [1.24-1.36] | 1.23 [1.13-1.35] | 1.28 [1.25-1.30] | 1.24 [1.21-1.27] | 1.23 [1.21-1.26] | 1.17 [1.11-1.23] | 1.10 [1.01-1.21] | 1.23 [1.20-1.26] | 1.20 [1.17-1.23] | 25,871 | 4,000 | 1,120 | 23,193 | 20,766 |
| 475 | 1.42 [1.36-1.47] | 1.46 [1.40-1.52] | 1.42 [1.35-1.49] | 1.31 [1.20-1.44] | 1.32 [1.27-1.38] | 1.22 [1.18-1.28] | 1.20 [1.15-1.26] | 1.18 [1.13-1.25] | 1.21 [1.10-1.33] | 1.17 [1.12-1.22] | 6,587 | 5,843 | 4,146 | 1,286 | 5,596 |
| 476 | 2.14 [2.08-2.20] | 1.76 [1.71-1.82] | 1.67 [1.60-1.74] | 2.83 [2.72-2.95] | 2.03 [1.98-2.09] | 1.73 [1.68-1.78] | 1.32 [1.27-1.36] | 1.29 [1.23-1.35] | 2.38 [2.28-2.48] | 1.66 [1.61-1.71] | 17,750 | 10,536 | 6,066 | 9,124 | 15,193 |
| 477 | 1.29 [1.26-1.33] | 1.30 [1.26-1.34] | 1.29 [1.25-1.34] | 1.26 [1.18-1.34] | 1.20 [1.16-1.24] | 1.24 [1.20-1.27] | 1.21 [1.17-1.25] | 1.20 [1.16-1.24] | 1.21 [1.14-1.29] | 1.17 [1.14-1.21] | 11,914 | 9,611 | 8,860 | 2,635 | 10,369 |
| 478 | 1.30 [1.17-1.44] | 1.29 [1.15-1.45] | 1.22 [1.06-1.40] | 1.29 [1.07-1.56] | 1.18 [1.06-1.32] | 1.18 [1.06-1.31] | 1.12 [1.00-1.27] | 1.06 [0.92-1.22] | 1.24 [1.02-1.50] | 1.11 [0.99-1.24] | 939 | 717 | 506 | 306 | 821 |
| 479 | 1.43 [1.39-1.47] | 1.44 [1.40-1.49] | 1.39 [1.34-1.45] | 1.37 [1.31-1.44] | 1.31 [1.27-1.35] | 1.29 [1.26-1.33] | 1.24 [1.20-1.28] | 1.21 [1.17-1.26] | 1.28 [1.22-1.34] | 1.20 [1.17-1.24] | 13,200 | 9,664 | 7,027 | 4,500 | 11,197 |
| 480 | 1.27 [1.26-1.29] | 1.27 [1.26-1.28] | 1.26 [1.25-1.28] | 1.30 [1.26-1.35] | 1.21 [1.20-1.22] | 1.21 [1.20-1.22] | 1.16 [1.15-1.18] | 1.16 [1.14-1.17] | 1.25 [1.21-1.30] | 1.17 [1.16-1.19] | 98,311 | 90,942 | 86,748 | 8,658 | 89,044 |
| 480.1 | 1.30 [1.25-1.35] | 1.32 [1.27-1.38] | 1.31 [1.25-1.36] | 1.13 [1.00-1.28] | 1.24 [1.19-1.29] | 1.22 [1.17-1.27] | 1.19 [1.14-1.24] | 1.17 [1.12-1.22] | 1.11 [0.98-1.25] | 1.19 [1.14-1.23] | 7,315 | 6,757 | 6,320 | 696 | 6,770 |
| 480.11 | 1.28 [1.26-1.29] | 1.28 [1.26-1.29] | 1.27 [1.25-1.29] | 1.29 [1.24-1.34] | 1.21 [1.19-1.23] | 1.21 [1.19-1.22] | 1.16 [1.15-1.18] | 1.15 [1.14-1.17] | 1.24 [1.19-1.30] | 1.17 [1.15-1.18] | 62,603 | 57,581 | 54,753 | 5,925 | 57,060 |
| 480.12 | 1.43 [1.28-1.60] | 1.55 [1.38-1.74] | 1.58 [1.40-1.77] | | 1.35 [1.20-1.51] | 1.27 [1.13-1.42] | 1.30 [1.15-1.47] | 1.30 [1.15-1.47] | | 1.20 [1.07-1.36] | 800 | 775 | 743 | NA | 747 |
| 480.2 | 1.28 [1.23-1.33] | 1.29 [1.24-1.34] | 1.31 [1.25-1.36] | 1.14 [1.00-1.29] | 1.23 [1.18-1.28] | 1.19 [1.14-1.24] | 1.16 [1.11-1.20] | 1.16 [1.11-1.21] | 1.08 [0.95-1.23] | 1.17 [1.12-1.22] | 7,159 | 6,672 | 6,277 | 619 | 6,720 |
| 480.3 | 1.47 [1.33-1.62] | 1.49 [1.35-1.65] | 1.49 [1.34-1.65] | | 1.34 [1.21-1.48] | 1.36 [1.23-1.50] | 1.33 [1.19-1.47] | 1.33 [1.19-1.48] | | 1.26 [1.14-1.40] | 1,055 | 989 | 895 | NA | 950 |
| 480.5 | 1.24 [1.19-1.29] | 1.26 [1.21-1.31] | 1.24 [1.19-1.29] | 1.25 [1.06-1.47] | 1.18 [1.14-1.23] | 1.19 [1.14-1.24] | 1.17 [1.12-1.22] | 1.16 [1.11-1.21] | 1.21 [1.02-1.43] | 1.15 [1.10-1.20] | 6,951 | 6,624 | 6,456 | 366 | 6,113 |
| 481 | 1.35 [1.29-1.41] | 1.39 [1.32-1.45] | 1.36 [1.29-1.43] | 1.23 [1.12-1.34] | 1.28 [1.22-1.33] | 1.22 [1.17-1.28] | 1.19 [1.13-1.25] | 1.18 [1.11-1.24] | 1.15 [1.05-1.25] | 1.19 [1.13-1.24] | 5,744 | 4,672 | 3,759 | 1,414 | 5,332 |
| 483 | 1.08 [1.04-1.12] | 1.32 [1.21-1.43] | 1.31 [1.19-1.43] | 1.03 [0.98-1.07] | 1.04 [1.00-1.09] | 1.04 [1.00-1.08] | 1.14 [1.05-1.24] | 1.15 [1.04-1.26] | 1.00 [0.96-1.04] | 1.01 [0.97-1.05] | 6,647 | 1,420 | 1,248 | 5,295 | 5,902 |

Supplementary Table 6: phecode-mapped results (hazard ratios and events from all cohorts)

| | Hazard ratio (99% confidence interval) | | | | | | | | | | | | | | |
|---|---|---|---|---|---|---|---|---|---|---|---|---|---|---|---|
| | crude | | | | | adjusted | | | | | Events (in exposed) | | | | |
| Outcome | any age | 18+ | 40+ | <18 | hosp. | any age | 18+ | 40+ | <18 | hosp. | any age | 18+ | 40+ | <18 | hosp. |
| 495 | 2.12 [2.11-2.14] | 1.98 [1.96-2.00] | 1.84 [1.82-1.87] | 2.50 [2.46-2.53] | 1.93 [1.91-1.95] | 1.61 [1.59-1.62] | 1.34 [1.33-1.36] | 1.31 [1.29-1.33] | 2.06 [2.02-2.09] | 1.51 [1.50-1.53] | 165,066 | 117,603 | 75,502 | 63,223 | 122,304 |
| 495.2 | 2.95 [2.79-3.13] | 2.46 [2.25-2.68] | 2.08 [1.85-2.33] | 3.34 [3.10-3.60] | 2.59 [2.43-2.76] | 2.29 [2.15-2.44] | 1.55 [1.40-1.72] | 1.35 [1.18-1.54] | 2.76 [2.55-2.99] | 2.01 [1.88-2.15] | 3,968 | 1,559 | 809 | 2,673 | 3,324 |
| 496 | 1.48 [1.46-1.50] | 1.48 [1.46-1.50] | 1.48 [1.46-1.50] | 1.60 [1.27-2.02] | 1.37 [1.35-1.39] | 1.30 [1.28-1.31] | 1.21 [1.19-1.23] | 1.21 [1.20-1.23] | 1.39 [1.09-1.76] | 1.23 [1.21-1.24] | 56,567 | 56,445 | 55,924 | 218 | 50,540 |
| 496.1 | 1.35 [1.32-1.39] | 1.35 [1.32-1.39] | 1.36 [1.32-1.39] | 1.21 [0.93-1.57] | 1.28 [1.25-1.32] | 1.25 [1.22-1.29] | 1.18 [1.15-1.21] | 1.18 [1.15-1.22] | 1.17 [0.89-1.53] | 1.21 [1.18-1.25] | 15,001 | 14,949 | 14,688 | 151 | 13,692 |
| 496.2 | 1.45 [1.33-1.58] | 1.41 [1.29-1.54] | 1.41 [1.29-1.55] | | 1.29 [1.18-1.42] | 1.28 [1.17-1.40] | 1.18 [1.08-1.30] | 1.18 [1.07-1.30] | | 1.16 [1.05-1.28] | 1,286 | 1,246 | 1,191 | NA | 1,145 |
| 496.21 | 1.46 [1.44-1.48] | 1.45 [1.44-1.47] | 1.45 [1.43-1.47] | 1.81 [1.49-2.20] | 1.36 [1.34-1.38] | 1.29 [1.27-1.31] | 1.20 [1.19-1.22] | 1.21 [1.19-1.22] | 1.54 [1.26-1.89] | 1.22 [1.21-1.24] | 63,230 | 63,040 | 62,368 | 310 | 55,206 |
| 496.3 | 1.49 [1.45-1.54] | 1.51 [1.47-1.56] | 1.50 [1.45-1.55] | 1.29 [1.06-1.58] | 1.39 [1.35-1.44] | 1.29 [1.25-1.33] | 1.27 [1.23-1.31] | 1.26 [1.22-1.30] | 1.15 [0.93-1.42] | 1.22 [1.18-1.26] | 11,900 | 11,734 | 11,381 | 269 | 11,013 |
| 497 | 1.44 [1.36-1.53] | 1.44 [1.36-1.53] | 1.45 [1.36-1.54] | 1.32 [0.96-1.82] | 1.30 [1.23-1.39] | 1.29 [1.21-1.37] | 1.23 [1.16-1.31] | 1.25 [1.17-1.33] | 1.24 [0.89-1.73] | 1.19 [1.12-1.27] | 2,869 | 2,824 | 2,580 | 108 | 2,545 |
| 499 | 1.23 [0.96-1.58] | | | | 1.14 [0.87-1.50] | 1.11 [0.86-1.43] | | | | 1.03 [0.78-1.36] | 160 | NA | NA | NA | 134 |
| 500 | 1.37 [1.22-1.53] | 1.34 [1.19-1.51] | 1.44 [1.28-1.63] | | 1.22 [1.08-1.37] | 1.30 [1.16-1.47] | 1.22 [1.08-1.38] | 1.32 [1.17-1.50] | | 1.18 [1.05-1.34] | 763 | 735 | 709 | NA | 688 |
| 500.1 | 1.58 [1.37-1.82] | 1.57 [1.36-1.81] | 1.56 [1.35-1.81] | | 1.42 [1.22-1.65] | 1.41 [1.22-1.63] | 1.36 [1.17-1.58] | 1.37 [1.18-1.60] | | 1.28 [1.10-1.49] | 511 | 506 | 478 | NA | 453 |
| 500.2 | 1.35 [1.25-1.46] | 1.35 [1.24-1.46] | 1.35 [1.25-1.46] | | 1.28 [1.18-1.39] | 1.28 [1.18-1.39] | 1.24 [1.14-1.34] | 1.24 [1.14-1.34] | | 1.22 [1.12-1.32] | 1,684 | 1,684 | 1,678 | NA | 1,572 |
| 501 | 1.17 [1.14-1.20] | 1.18 [1.14-1.21] | 1.17 [1.14-1.21] | 0.94 [0.79-1.12] | 1.13 [1.09-1.16] | 1.15 [1.11-1.18] | 1.12 [1.09-1.16] | 1.12 [1.08-1.15] | 0.92 [0.77-1.10] | 1.13 [1.10-1.17] | 13,257 | 13,076 | 12,692 | 313 | 12,447 |
| 502 | 1.45 [1.40-1.50] | 1.47 [1.42-1.52] | 1.45 [1.40-1.50] | | 1.38 [1.33-1.44] | 1.37 [1.32-1.42] | 1.35 [1.30-1.40] | 1.34 [1.29-1.39] | | 1.33 [1.28-1.38] | 8,675 | 8,663 | 8,561 | NA | 7,999 |
| 503 | 1.30 [1.24-1.36] | 1.28 [1.23-1.34] | 1.31 [1.25-1.37] | 0.98 [0.73-1.32] | 1.19 [1.13-1.25] | 1.25 [1.19-1.31] | 1.18 [1.13-1.24] | 1.21 [1.15-1.26] | 0.97 [0.71-1.31] | 1.17 [1.11-1.22] | 4,997 | 4,933 | 4,786 | 110 | 4,488 |
| 504 | 1.54 [1.45-1.64] | 1.52 [1.43-1.62] | 1.55 [1.45-1.65] | | 1.45 [1.35-1.54] | 1.45 [1.36-1.54] | 1.38 [1.29-1.47] | 1.40 [1.31-1.50] | | 1.39 [1.30-1.48] | 2,740 | 2,710 | 2,635 | NA | 2,545 |
| 505 | 2.40 [2.08-2.78] | 2.31 [2.00-2.68] | 2.11 [1.80-2.47] | | 2.24 [1.93-2.59] | 1.72 [1.47-2.02] | 1.55 [1.31-1.82] | 1.49 [1.25-1.77] | | 1.69 [1.44-1.98] | 600 | 574 | 451 | NA | 570 |
| 506 | 1.16 [1.11-1.21] | 1.16 [1.11-1.21] | 1.19 [1.13-1.24] | 1.06 [0.95-1.18] | 1.09 [1.04-1.14] | 1.12 [1.08-1.17] | 1.09 [1.04-1.14] | 1.11 [1.06-1.17] | 1.07 [0.96-1.19] | 1.08 [1.03-1.13] | 5,655 | 5,112 | 4,419 | 862 | 4,745 |
| 507 | 1.20 [1.19-1.22] | 1.20 [1.19-1.22] | 1.21 [1.19-1.22] | 1.10 [1.02-1.20] | 1.15 [1.13-1.17] | 1.17 [1.15-1.18] | 1.13 [1.11-1.15] | 1.13 [1.11-1.15] | 1.11 [1.02-1.20] | 1.13 [1.12-1.15] | 50,126 | 49,149 | 47,087 | 1,605 | 45,275 |
| 508 | 1.22 [1.19-1.25] | 1.21 [1.18-1.24] | 1.21 [1.18-1.24] | 1.19 [1.09-1.31] | 1.16 [1.13-1.19] | 1.15 [1.12-1.18] | 1.10 [1.07-1.13] | 1.10 [1.07-1.12] | 1.13 [1.03-1.24] | 1.12 [1.09-1.15] | 18,974 | 18,039 | 17,062 | 1,293 | 17,468 |
| 509.1 | 1.32 [1.30-1.34] | 1.31 [1.29-1.34] | 1.30 [1.28-1.33] | 1.21 [1.07-1.36] | 1.24 [1.22-1.26] | 1.23 [1.21-1.25] | 1.17 [1.14-1.19] | 1.16 [1.13-1.18] | 1.12 [0.99-1.26] | 1.18 [1.16-1.21] | 30,130 | 29,782 | 28,727 | 709 | 27,859 |
| 509.2 | 1.31 [1.29-1.34] | 1.31 [1.29-1.34] | 1.30 [1.28-1.33] | 1.17 [1.02-1.33] | 1.24 [1.21-1.26] | 1.22 [1.20-1.25] | 1.16 [1.14-1.19] | 1.16 [1.13-1.18] | 1.09 [0.95-1.25] | 1.18 [1.15-1.20] | 25,270 | 24,998 | 24,069 | 587 | 23,419 |
| 509.5 | 1.28 [1.17-1.40] | 1.29 [1.17-1.42] | 1.27 [1.15-1.41] | 1.16 [0.92-1.48] | 1.17 [1.06-1.29] | 1.21 [1.10-1.32] | 1.16 [1.05-1.28] | 1.16 [1.05-1.29] | 1.12 [0.88-1.43] | 1.13 [1.03-1.25] | 1,209 | 1,081 | 969 | 172 | 1,062 |
| 509.8 | 1.51 [1.45-1.57] | 1.53 [1.47-1.59] | 1.53 [1.46-1.59] | 0.89 [0.74-1.07] | 1.37 [1.31-1.42] | 1.32 [1.27-1.38] | 1.25 [1.20-1.31] | 1.24 [1.19-1.30] | 0.82 [0.68-0.99] | 1.22 [1.17-1.28] | 6,613 | 6,428 | 6,141 | 280 | 6,314 |
| 510 | 1.27 [1.22-1.32] | 1.29 [1.24-1.34] | 1.30 [1.25-1.35] | 0.95 [0.82-1.10] | 1.20 [1.16-1.25] | 1.18 [1.13-1.22] | 1.14 [1.10-1.19] | 1.15 [1.11-1.20] | 0.87 [0.75-1.01] | 1.14 [1.10-1.19] | 7,653 | 7,307 | 6,975 | 453 | 7,123 |
| 510.2 | 1.02 [0.79-1.30] | 1.06 [0.82-1.38] | 1.08 [0.81-1.43] | | 0.94 [0.73-1.22] | 0.95 [0.74-1.22] | 0.96 [0.73-1.25] | 0.95 [0.71-1.27] | | 0.86 [0.66-1.13] | 150 | 138 | 113 | NA | 132 |
| 512 | 1.25 [1.23-1.28] | 1.24 [1.22-1.26] | 1.23 [1.21-1.26] | 1.25 [1.17-1.33] | 1.18 [1.16-1.21] | 1.17 [1.15-1.20] | 1.12 [1.09-1.14] | 1.11 [1.09-1.13] | 1.17 [1.10-1.25] | 1.14 [1.11-1.16] | 28,140 | 26,101 | 24,543 | 2,583 | 25,833 |
| 512.1 | 1.96 [1.92-2.00] | 1.49 [1.38-1.60] | 1.41 [1.30-1.53] | 2.00 [1.96-2.04] | 1.87 [1.83-1.91] | 1.88 [1.84-1.92] | 1.24 [1.15-1.34] | 1.20 [1.11-1.31] | 1.92 [1.88-1.97] | 1.80 [1.76-1.84] | 29,127 | 1,915 | 1,583 | 27,345 | 26,362 |
| 512.2 | 1.25 [1.13-1.37] | 1.27 [1.15-1.40] | 1.26 [1.12-1.42] | 1.16 [0.89-1.52] | 1.12 [1.01-1.24] | 1.13 [1.02-1.25] | 1.09 [0.99-1.21] | 1.06 [0.94-1.20] | 1.15 [0.87-1.52] | 1.04 [0.94-1.16] | 1,041 | 993 | 700 | 145 | 921 |
| 512.7 | 1.35 [1.33-1.38] | 1.33 [1.31-1.36] | 1.33 [1.31-1.36] | 1.39 [1.33-1.46] | 1.26 [1.24-1.28] | 1.25 [1.23-1.27] | 1.18 [1.16-1.20] | 1.18 [1.16-1.20] | 1.33 [1.27-1.40] | 1.19 [1.17-1.21] | 37,295 | 34,544 | 29,430 | 4,423 | 33,881 |
| 512.8 | 1.38 [1.35-1.41] | 1.35 [1.32-1.39] | 1.36 [1.32-1.39] | 1.37 [1.32-1.43] | 1.29 [1.26-1.32] | 1.27 [1.24-1.30] | 1.19 [1.15-1.22] | 1.19 [1.15-1.22] | 1.30 [1.25-1.35] | 1.21 [1.18-1.24] | 20,701 | 14,452 | 12,383 | 6,900 | 18,750 |
| 512.9 | 1.35 [1.30-1.41] | 1.35 [1.28-1.42] | 1.34 [1.27-1.42] | 1.33 [1.25-1.42] | 1.26 [1.21-1.32] | 1.25 [1.20-1.31] | 1.17 [1.11-1.24] | 1.16 [1.10-1.24] | 1.26 [1.18-1.35] | 1.19 [1.14-1.25] | 6,008 | 3,624 | 3,038 | 2,578 | 5,501 |
| 513 | 1.52 [1.50-1.54] | 1.34 [1.32-1.36] | 1.33 [1.31-1.35] | 1.76 [1.72-1.79] | 1.43 [1.41-1.44] | 1.43 [1.41-1.44] | 1.18 [1.16-1.20] | 1.18 [1.16-1.20] | 1.70 [1.67-1.73] | 1.36 [1.35-1.38] | 74,870 | 41,003 | 34,545 | 35,908 | 67,136 |
| 513.3 | 1.46 [1.35-1.58] | 1.53 [1.40-1.67] | 1.58 [1.45-1.73] | 1.05 [0.89-1.24] | 1.34 [1.24-1.45] | 1.33 [1.23-1.44] | 1.30 [1.18-1.42] | 1.31 [1.19-1.44] | 0.99 [0.83-1.17] | 1.25 [1.15-1.36] | 1,670 | 1,361 | 1,267 | 342 | 1,554 |
| 513.31 | 1.34 [0.97-1.84] | | | | | 1.30 [0.94-1.80] | | | | | 104 | NA | NA | NA | NA |
| 513.4 | 1.31 [1.19-1.45] | 1.28 [1.15-1.43] | 1.19 [1.04-1.37] | 1.34 [1.11-1.63] | 1.17 [1.05-1.31] | 1.15 [1.04-1.28] | 1.05 [0.94-1.19] | 0.99 [0.85-1.14] | 1.21 [0.99-1.48] | 1.07 [0.95-1.19] | 972 | 785 | 495 | 292 | 827 |
| 513.8 | 1.29 [1.15-1.44] | 1.34 [1.19-1.51] | 1.30 [1.15-1.47] | | 1.26 [1.11-1.42] | 1.22 [1.08-1.37] | 1.22 [1.08-1.38] | 1.19 [1.05-1.36] | | 1.22 [1.08-1.37] | 734 | 715 | 651 | NA | 679 |
| 514 | 1.26 [1.22-1.29] | 1.25 [1.22-1.29] | 1.25 [1.22-1.29] | 1.29 [1.09-1.53] | 1.20 [1.17-1.24] | 1.20 [1.17-1.24] | 1.15 [1.12-1.18] | 1.15 [1.12-1.18] | 1.26 [1.06-1.50] | 1.17 [1.14-1.21] | 13,726 | 13,469 | 13,019 | 377 | 12,284 |
| 514.1 | 1.41 [1.13-1.76] | 1.46 [1.15-1.86] | 1.41 [1.09-1.82] | | 1.25 [0.99-1.58] | 1.29 [1.02-1.62] | 1.28 [1.00-1.64] | 1.19 [0.92-1.56] | | 1.15 [0.91-1.46] | 210 | 177 | 157 | NA | 181 |
| 516 | 1.31 [1.20-1.44] | 1.37 [1.24-1.51] | 1.45 [1.31-1.61] | 1.04 [0.86-1.26] | 1.20 [1.09-1.31] | 1.17 [1.07-1.29] | 1.15 [1.04-1.27] | 1.23 [1.10-1.37] | 0.97 [0.80-1.19] | 1.10 [1.00-1.20] | 1,289 | 1,065 | 969 | 267 | 1,185 |
| 516.1 | 1.33 [1.30-1.36] | 1.35 [1.31-1.38] | 1.34 [1.31-1.38] | 1.24 [1.17-1.31] | 1.23 [1.20-1.26] | 1.25 [1.22-1.28] | 1.22 [1.18-1.25] | 1.21 [1.18-1.24] | 1.19 [1.12-1.26] | 1.19 [1.16-1.22] | 18,561 | 15,989 | 14,425 | 3,187 | 16,162 |
| 519 | 1.61 [1.40-1.85] | 1.68 [1.45-1.96] | 1.59 [1.36-1.86] | | 1.50 [1.30-1.73] | 1.45 [1.26-1.67] | 1.40 [1.20-1.64] | 1.38 [1.17-1.63] | | 1.37 [1.18-1.58] | 556 | 475 | 423 | NA | 500 |
| 519.2 | 1.21 [1.10-1.34] | 1.21 [1.10-1.34] | 1.25 [1.12-1.38] | | 1.13 [1.01-1.25] | 1.17 [1.06-1.29] | 1.12 [1.01-1.24] | 1.17 [1.05-1.30] | | 1.11 [1.00-1.24] | 999 | 921 | 863 | NA | 843 |
| 519.8 | 1.37 [1.35-1.39] | 1.35 [1.34-1.37] | 1.32 [1.30-1.34] | 1.43 [1.40-1.46] | 1.29 [1.27-1.30] | 1.27 [1.26-1.29] | 1.20 [1.19-1.22] | 1.19 [1.17-1.21] | 1.35 [1.32-1.38] | 1.22 [1.20-1.23] | 82,314 | 62,929 | 55,037 | 22,170 | 74,222 |
| 519.9 | 1.27 [1.16-1.38] | 1.23 [1.12-1.36] | 1.28 [1.15-1.43] | 1.16 [0.95-1.42] | 1.16 [1.05-1.27] | 1.20 [1.10-1.32] | 1.13 [1.03-1.25] | 1.19 [1.06-1.33] | 1.13 [0.92-1.38] | 1.12 [1.01-1.23] | 1,228 | 1,032 | 850 | 253 | 1,056 |
| digestive | | | | | | | | | | | | | | | |
| 520 | 1.16 [1.12-1.21] | 1.15 [1.06-1.25] | 1.22 [1.02-1.47] | 1.14 [1.09-1.19] | 1.09 [1.05-1.15] | 1.12 [1.08-1.17] | 1.05 [0.96-1.14] | 1.12 [0.93-1.35] | 1.11 [1.06-1.16] | 1.07 [1.02-1.13] | 6,500 | 1,359 | 276 | 5,703 | 4,997 |
| 520.1 | 1.15 [1.07-1.22] | 1.14 [0.95-1.36] | | 1.13 [1.06-1.21] | 1.06 [0.98-1.14] | 1.12 [1.05-1.20] | 1.10 [0.91-1.32] | | 1.11 [1.04-1.19] | 1.05 [0.97-1.13] | 2,644 | 273 | NA | 2,518 | 2,085 |
| 520.2 | 1.21 [1.18-1.24] | 1.23 [1.19-1.27] | 1.15 [1.07-1.24] | 1.18 [1.14-1.22] | 1.13 [1.10-1.17] | 1.15 [1.13-1.18] | 1.13 [1.09-1.16] | 1.10 [1.02-1.18] | 1.14 [1.10-1.17] | 1.10 [1.07-1.13] | 17,520 | 9,951 | 1,617 | 10,828 | 12,168 |
| 521 | 1.21 [1.01-1.46] | 1.45 [1.09-1.94] | | 1.14 [0.91-1.43] | 1.07 [0.88-1.31] | 1.15 [0.96-1.40] | 1.34 [0.99-1.80] | | 1.11 [0.88-1.40] | 1.05 [0.86-1.29] | 303 | 113 | NA | 208 | 256 |
| 521.1 | 1.11 [1.09-1.13] | 1.25 [1.22-1.28] | 1.28 [1.24-1.32] | 1.02 [1.00-1.04] | 1.05 [1.03-1.07] | 1.07 [1.05-1.09] | 1.12 [1.10-1.15] | 1.15 [1.11-1.19] | 1.01 [0.99-1.03] | 1.03 [1.01-1.05] | 42,689 | 17,603 | 8,465 | 28,068 | 34,584 |
| 521.2 | 1.11 [0.83-1.48] | | | | 1.10 [0.81-1.48] | 1.00 [0.74-1.36] | | | | 1.01 [0.74-1.38] | 121 | NA | NA | NA | 114 |
| 521.4 | 1.10 [0.91-1.34] | 1.19 [0.88-1.60] | | 1.05 [0.84-1.32] | 1.11 [0.90-1.37] | 1.08 [0.88-1.32] | 1.08 [0.79-1.47] | | 1.05 [0.83-1.33] | 1.10 [0.89-1.37] | 285 | 104 | NA | 216 | 240 |
| 522 | 1.14 [1.10-1.18] | 1.22 [1.17-1.27] | 1.25 [1.19-1.32] | 1.00 [0.95-1.05] | 1.07 [1.03-1.11] | 1.09 [1.05-1.13] | 1.10 [1.05-1.15] | 1.13 [1.07-1.19] | 0.99 [0.94-1.05] | 1.04 [1.00-1.08] | 8,999 | 5,838 | 3,471 | 3,950 | 7,404 |
| 522.1 | 1.28 [1.09-1.51] | 1.35 [1.11-1.65] | 1.32 [1.01-1.73] | 1.17 [0.91-1.50] | 1.22 [1.02-1.47] | 1.16 [0.98-1.38] | 1.12 [0.91-1.39] | 1.08 [0.81-1.44] | 1.13 [0.88-1.46] | 1.13 [0.94-1.36] | 370 | 243 | 126 | 169 | 312 |
| 522.5 | 1.12 [1.08-1.16] | 1.20 [1.14-1.26] | 1.25 [1.17-1.33] | 1.00 [0.94-1.05] | 1.05 [1.00-1.09] | 1.07 [1.03-1.11] | 1.08 [1.03-1.14] | 1.12 [1.05-1.20] | 0.99 [0.94-1.05] | 1.02 [0.98-1.07] | 6,595 | 3,939 | 2,296 | 3,226 | 5,462 |
| 523 | 1.33 [1.24-1.42] | 1.36 [1.27-1.47] | 1.40 [1.29-1.53] | 1.02 [0.89-1.16] | 1.20 [1.12-1.29] | 1.26 [1.18-1.35] | 1.25 [1.16-1.35] | 1.27 [1.16-1.39] | 0.96 [0.84-1.10] | 1.16 [1.08-1.24] | 2,347 | 1,883 | 1,363 | 637 | 2,033 |
| 523.1 | 1.33 [1.19-1.50] | 1.46 [1.26-1.68] | 1.47 [1.23-1.76] | 1.21 [1.01-1.43] | 1.32 [1.16-1.49] | 1.27 [1.12-1.43] | 1.32 [1.14-1.53] | 1.33 [1.10-1.60] | 1.19 [0.99-1.42] | 1.27 [1.12-1.44] | 783 | 499 | 306 | 361 | 685 |
| 523.31 | 1.34 [1.19-1.52] | 1.38 [1.21-1.57] | 1.52 [1.23-1.89] | 1.24 [1.00-1.55] | 1.21 [1.05-1.39] | 1.30 [1.14-1.47] | 1.26 [1.10-1.44] | 1.37 [1.10-1.72] | 1.27 [1.01-1.59] | 1.19 [1.04-1.37] | 661 | 587 | 207 | 214 | 508 |
| 523.32 | 1.26 [1.22-1.31] | 1.32 [1.28-1.37] | 1.28 [1.19-1.37] | 1.18 [1.11-1.26] | 1.17 [1.12-1.22] | 1.19 [1.14-1.23] | 1.19 [1.14-1.23] | 1.16 [1.08-1.25] | 1.12 [1.05-1.20] | 1.12 [1.07-1.17] | 7,692 | 7,339 | 1,866 | 2,575 | 5,756 |
| 524 | 1.22 [0.95-1.57] | | | 1.08 [0.80-1.47] | 1.09 [0.81-1.45] | 1.19 [0.92-1.54] | | | 1.05 [0.77-1.45] | 1.03 [0.76-1.39] | 159 | NA | NA | 106 | 120 |
| 524.3 | 1.16 [1.12-1.21] | 1.13 [1.07-1.20] | 1.07 [0.92-1.25] | 1.14 [1.09-1.19] | 1.09 [1.04-1.14] | 1.12 [1.08-1.17] | 1.06 [1.00-1.13] | 1.00 [0.85-1.16] | 1.11 [1.07-1.16] | 1.07 [1.03-1.12] | 7,826 | 2,563 | 373 | 6,447 | 5,587 |
| 525 | 1.18 [1.15-1.22] | 1.25 [1.21-1.29] | 1.29 [1.24-1.34] | 1.04 [0.99-1.08] | 1.12 [1.08-1.15] | 1.13 [1.10-1.16] | 1.14 [1.10-1.17] | 1.15 [1.11-1.20] | 1.02 [0.98-1.07] | 1.08 [1.05-1.12] | 13,088 | 9,194 | 5,859 | 5,019 | 10,876 |
| 526.1 | 1.21 [1.08-1.36] | 1.19 [1.05-1.36] | 1.11 [0.95-1.29] | 1.16 [0.93-1.44] | 1.16 [1.01-1.32] | 1.20 [1.06-1.35] | 1.13 [0.99-1.29] | 1.07 [0.91-1.25] | 1.13 [0.90-1.42] | 1.14 [0.99-1.30] | 721 | 553 | 374 | 224 | 544 |
| 526.3 | 1.21 [1.06-1.38] | 1.31 [1.13-1.52] | | 1.19 [1.02-1.39] | 1.08 [0.93-1.25] | 1.16 [1.01-1.32] | 1.23 [1.06-1.44] | | 1.15 [0.98-1.35] | 1.05 [0.90-1.22] | 601 | 427 | NA | 439 | 449 |
| 526.41 | 1.44 [1.30-1.59] | 1.47 [1.33-1.63] | 1.51 [1.31-1.73] | 1.38 [1.13-1.69] | 1.36 [1.22-1.51] | 1.27 [1.14-1.41] | 1.22 [1.10-1.36] | 1.28 [1.11-1.47] | 1.24 [1.00-1.54] | 1.23 [1.10-1.37] | 1,041 | 967 | 522 | 270 | 907 |
| 526.5 | 1.16 [0.99-1.37] | 1.17 [0.99-1.38] | 1.12 [0.94-1.35] | | 1.06 [0.90-1.26] | 1.09 [0.92-1.29] | 1.03 [0.87-1.23] | 1.01 [0.83-1.22] | | 1.03 [0.87-1.23] | 364 | 344 | 278 | NA | 336 |
| 526.9 | 1.29 [1.16-1.43] | 1.31 [1.17-1.47] | 1.36 [1.20-1.54] | 1.31 [1.02-1.68] | 1.18 [1.06-1.32] | 1.18 [1.05-1.31] | 1.14 [1.01-1.28] | 1.19 [1.04-1.36] | 1.26 [0.98-1.63] | 1.12 [1.00-1.26] | 863 | 771 | 601 | 176 | 762 |
| 527 | 1.31 [1.24-1.39] | 1.36 [1.28-1.45] | 1.36 [1.27-1.46] | 1.16 [1.04-1.30] | 1.24 [1.16-1.32] | 1.24 [1.17-1.32] | 1.25 [1.17-1.33] | 1.23 [1.15-1.33] | 1.12 [1.00-1.25] | 1.19 [1.12-1.27] | 3,061 | 2,405 | 1,914 | 848 | 2,583 |
| 527.1 | 1.28 [0.99-1.65] | 1.51 [1.15-1.99] | | | 1.25 [0.95-1.66] | 1.18 [0.91-1.53] | 1.37 [1.03-1.82] | | | 1.19 [0.89-1.58] | 146 | 126 | NA | NA | 121 |
| 527.2 | 1.37 [1.25-1.50] | 1.36 [1.23-1.49] | 1.35 [1.21-1.50] | 1.31 [1.08-1.59] | 1.27 [1.16-1.40] | 1.28 [1.17-1.40] | 1.22 [1.11-1.35] | 1.22 [1.09-1.36] | 1.26 [1.03-1.54] | 1.21 [1.10-1.34] | 1,278 | 1,081 | 890 | 286 | 1,110 |
| 527.7 | 1.21 [1.05-1.41] | 1.29 [1.07-1.56] | 1.34 [1.10-1.64] | 1.02 [0.81-1.28] | 1.08 [0.93-1.26] | 1.11 [0.95-1.29] | 1.13 [0.93-1.38] | 1.19 [0.97-1.47] | 0.94 [0.75-1.19] | 1.02 [0.87-1.19] | 448 | 267 | 240 | 193 | 421 |
| 527.8 | 1.39 [1.18-1.65] | 1.40 [1.18-1.66] | 1.37 [1.14-1.65] | | 1.28 [1.07-1.53] | 1.32 [1.11-1.56] | 1.26 [1.06-1.51] | 1.22 [1.00-1.48] | | 1.23 [1.02-1.47] | 358 | 327 | 272 | NA | 303 |
| 528 | 1.37 [1.31-1.44] | 1.38 [1.31-1.46] | 1.39 [1.31-1.48] | 1.26 [1.13-1.42] | 1.29 [1.22-1.36] | 1.30 [1.24-1.37] | 1.27 [1.20-1.34] | 1.27 [1.20-1.35] | 1.23 [1.09-1.38] | 1.25 [1.18-1.32] | 4,011 | 3,375 | 2,729 | 821 | 3,297 |
| 528.11 | 1.41 [1.33-1.51] | 1.45 [1.35-1.57] | 1.43 [1.32-1.56] | 1.33 [1.18-1.48] | 1.32 [1.23-1.41] | 1.34 [1.25-1.43] | 1.32 [1.22-1.42] | 1.30 [1.19-1.42] | 1.28 [1.14-1.43] | 1.27 [1.18-1.36] | 2,454 | 1,776 | 1,423 | 831 | 2,177 |
| 528.12 | 1.38 [1.19-1.62] | 1.42 [1.16-1.73] | 1.46 [1.15-1.85] | 1.34 [1.07-1.67] | 1.29 [1.09-1.53] | 1.31 [1.12-1.53] | 1.22 [0.99-1.51] | 1.25 [0.98-1.61] | 1.28 [1.02-1.61] | 1.24 [1.05-1.47] | 419 | 246 | 180 | 203 | 362 |
| 528.3 | 1.06 [0.96-1.17] | 1.11 [0.99-1.24] | 1.16 [1.01-1.35] | 0.95 [0.80-1.11] | 1.04 [0.93-1.15] | 1.04 [0.94-1.15] | 1.02 [0.91-1.15] | 1.09 [0.94-1.26] | 0.97 [0.82-1.15] | 1.05 [0.94-1.17] | 979 | 716 | 438 | 367 | 843 |
| 528.41 | 1.17 [0.99-1.39] | 1.25 [1.00-1.55] | 1.38 [1.04-1.82] | 1.10 [0.87-1.39] | 1.18 [0.98-1.43] | 1.13 [0.95-1.35] | 1.20 [0.96-1.50] | 1.25 [0.93-1.66] | 1.07 [0.84-1.35] | 1.14 [0.93-1.38] | 356 | 197 | 119 | 193 | 273 |
| 528.5 | 1.37 [1.28-1.47] | 1.40 [1.29-1.52] | 1.36 [1.24-1.49] | 1.37 [1.22-1.53] | 1.34 [1.24-1.44] | 1.30 [1.22-1.40] | 1.26 [1.16-1.37] | 1.23 [1.12-1.36] | 1.33 [1.19-1.50] | 1.27 [1.18-1.37] | 2,191 | 1,468 | 1,105 | 863 | 1,819 |
| 528.6 | 1.40 [1.23-1.59] | 1.36 [1.20-1.55] | 1.43 [1.25-1.64] | | 1.34 [1.16-1.53] | 1.33 [1.17-1.52] | 1.24 [1.09-1.42] | 1.29 [1.12-1.49] | | 1.30 [1.13-1.49] | 584 | 568 | 519 | NA | 499 |
| 528.7 | 1.32 [1.15-1.52] | 1.33 [1.15-1.53] | 1.28 [1.08-1.52] | | 1.25 [1.08-1.46] | 1.27 [1.10-1.46] | 1.25 [1.07-1.45] | 1.18 [0.99-1.40] | | 1.21 [1.04-1.42] | 503 | 448 | 327 | NA | 407 |
| 529 | 1.39 [1.31-1.47] | 1.40 [1.32-1.49] | 1.37 [1.29-1.46] | 1.32 [1.14-1.53] | 1.29 [1.21-1.37] | 1.28 [1.21-1.36] | 1.24 [1.17-1.32] | 1.23 [1.15-1.31] | 1.26 [1.09-1.47] | 1.21 [1.14-1.29] | 3,114 | 2,746 | 2,364 | 495 | 2,705 |
| 529.1 | 1.42 [1.28-1.56] | 1.49 [1.34-1.65] | 1.48 [1.33-1.66] | 1.29 [1.02-1.64] | 1.34 [1.21-1.49] | 1.31 [1.18-1.44] | 1.30 [1.16-1.45] | 1.28 [1.14-1.44] | 1.25 [0.97-1.60] | 1.26 [1.14-1.41] | 1,040 | 899 | 793 | 182 | 922 |
| 530 | 1.39 [1.29-1.49] | 1.37 [1.27-1.48] | 1.35 [1.25-1.46] | | 1.26 [1.17-1.36] | 1.27 [1.18-1.37] | 1.21 [1.12-1.31] | 1.21 [1.12-1.31] | | 1.19 [1.10-1.28] | 1,815 | 1,780 | 1,646 | NA | 1,634 |
| 530.1 | 1.33 [1.30-1.36] | 1.32 [1.29-1.36] | 1.30 [1.27-1.34] | 1.49 [1.35-1.64] | 1.23 [1.20-1.26] | 1.22 [1.19-1.25] | 1.18 [1.15-1.20] | 1.16 [1.13-1.19] | 1.34 [1.21-1.49] | 1.15 [1.12-1.18] | 18,438 | 17,762 | 16,047 | 1,158 | 15,910 |
| 530.11 | 1.36 [1.34-1.37] | 1.36 [1.35-1.38] | 1.36 [1.34-1.37] | 1.23 [1.18-1.29] | 1.25 [1.23-1.26] | 1.21 [1.20-1.23] | 1.18 [1.17-1.19] | 1.17 [1.16-1.19] | 1.11 [1.06-1.17] | 1.15 [1.13-1.16] | 73,030 | 70,858 | 62,008 | 4,889 | 64,783 |
| 530.12 | 1.30 [1.26-1.34] | 1.29 [1.25-1.34] | 1.28 [1.24-1.32] | 1.30 [1.03-1.64] | 1.18 [1.14-1.22] | 1.22 [1.18-1.26] | 1.18 [1.14-1.22] | 1.18 [1.14-1.22] | 1.27 [1.00-1.61] | 1.13 [1.09-1.17] | 9,007 | 8,920 | 8,459 | 197 | 7,512 |
| 530.14 | 1.32 [1.29-1.35] | 1.33 [1.30-1.36] | 1.32 [1.29-1.35] | 1.20 [1.08-1.33] | 1.21 [1.19-1.24] | 1.20 [1.18-1.23] | 1.18 [1.15-1.20] | 1.17 [1.14-1.20] | 1.08 [0.97-1.20] | 1.14 [1.11-1.16] | 21,183 | 20,879 | 18,546 | 980 | 18,033 |
| 530.2 | 1.36 [1.31-1.41] | 1.38 [1.33-1.43] | 1.36 [1.31-1.42] | 1.13 [0.93-1.37] | 1.27 [1.23-1.33] | 1.25 [1.20-1.30] | 1.22 [1.17-1.27] | 1.21 [1.16-1.26] | 1.00 [0.82-1.23] | 1.20 [1.16-1.25] | 7,168 | 7,034 | 6,487 | 274 | 6,482 |
| 530.3 | 1.30 [1.25-1.36] | 1.31 [1.26-1.36] | 1.30 [1.24-1.35] | 1.29 [0.99-1.67] | 1.25 [1.20-1.30] | 1.22 [1.18-1.28] | 1.21 [1.16-1.26] | 1.20 [1.15-1.25] | 1.18 [0.90-1.55] | 1.20 [1.15-1.25] | 6,260 | 6,186 | 5,883 | 150 | 5,507 |
| 530.5 | 1.38 [1.29-1.47] | 1.36 [1.28-1.46] | 1.39 [1.30-1.49] | 1.22 [0.89-1.68] | 1.30 [1.22-1.39] | 1.25 [1.17-1.33] | 1.20 [1.12-1.28] | 1.21 [1.13-1.30] | 1.06 [0.76-1.48] | 1.20 [1.13-1.29] | 2,436 | 2,396 | 2,214 | 104 | 2,267 |
| 530.6 | 1.22 [1.08-1.38] | 1.22 [1.08-1.37] | 1.22 [1.08-1.38] | | 1.14 [1.01-1.29] | 1.13 [1.00-1.28] | 1.13 [1.00-1.27] | 1.12 [0.99-1.27] | | 1.08 [0.95-1.22] | 721 | 717 | 710 | NA | 661 |
| 530.7 | 1.20 [1.10-1.30] | 1.25 [1.14-1.37] | 1.32 [1.18-1.47] | 1.11 [0.94-1.30] | 1.12 [1.02-1.22] | 1.11 [1.02-1.21] | 1.09 [0.99-1.19] | 1.18 [1.05-1.32] | 1.05 [0.89-1.24] | 1.07 [0.98-1.17] | 1,404 | 1,173 | 826 | 389 | 1,217 |
| 530.9 | 1.35 [1.29-1.42] | 1.35 [1.28-1.42] | 1.34 [1.27-1.42] | 1.08 [0.90-1.28] | 1.23 [1.17-1.30] | 1.22 [1.16-1.28] | 1.18 [1.12-1.24] | 1.17 [1.11-1.24] | 1.00 [0.83-1.20] | 1.14 [1.08-1.20] | 4,130 | 4,046 | 3,156 | 336 | 3,611 |
| 531 | 1.21 [1.18-1.25] | 1.22 [1.19-1.26] | 1.21 [1.18-1.25] | 1.26 [1.05-1.51] | 1.16 [1.12-1.20] | 1.17 [1.13-1.21] | 1.14 [1.10-1.17] | 1.13 [1.09-1.17] | 1.21 [1.01-1.47] | 1.15 [1.11-1.19] | 10,355 | 10,202 | 9,563 | 325 | 8,880 |
| 531.1 | 1.21 [1.15-1.28] | 1.21 [1.14-1.28] | 1.21 [1.14-1.28] | | 1.16 [1.10-1.24] | 1.21 [1.14-1.28] | 1.16 [1.10-1.23] | 1.16 [1.09-1.23] | | 1.18 [1.11-1.25] | 3,261 | 3,239 | 3,113 | NA | 2,785 |
| 531.2 | 1.29 [1.26-1.33] | 1.30 [1.26-1.33] | 1.29 [1.25-1.32] | 1.30 [1.10-1.54] | 1.21 [1.17-1.24] | 1.22 [1.18-1.25] | 1.17 [1.14-1.20] | 1.17 [1.13-1.20] | 1.27 [1.07-1.52] | 1.16 [1.13-1.20] | 13,006 | 12,851 | 12,008 | 368 | 11,348 |
| 531.3 | 1.19 [1.15-1.23] | 1.21 [1.17-1.25] | 1.20 [1.16-1.24] | 1.21 [0.99-1.48] | 1.15 [1.11-1.19] | 1.16 [1.12-1.20] | 1.14 [1.10-1.18] | 1.13 [1.09-1.17] | 1.18 [0.95-1.46] | 1.14 [1.10-1.18] | 8,838 | 8,712 | 8,214 | 259 | 7,495 |
| 531.4 | 1.29 [1.19-1.40] | 1.28 [1.18-1.39] | 1.29 [1.18-1.40] | | 1.18 [1.09-1.28] | 1.19 [1.10-1.29] | 1.12 [1.03-1.22] | 1.14 [1.04-1.24] | | 1.12 [1.03-1.22] | 1,511 | 1,492 | 1,381 | NA | 1,373 |
| 531.5 | 1.53 [1.24-1.89] | 1.55 [1.25-1.92] | 1.45 [1.15-1.82] | | 1.48 [1.19-1.83] | 1.29 [1.04-1.61] | 1.25 [0.99-1.56] | 1.09 [0.85-1.40] | | 1.32 [1.06-1.64] | 232 | 226 | 183 | NA | 225 |
| 532 | 1.31 [1.29-1.34] | 1.32 [1.29-1.34] | 1.31 [1.29-1.34] | 1.21 [1.11-1.32] | 1.23 [1.21-1.25] | 1.22 [1.19-1.24] | 1.18 [1.16-1.21] | 1.18 [1.16-1.21] | 1.12 [1.02-1.23] | 1.17 [1.15-1.19] | 30,132 | 29,322 | 27,273 | 1,407 | 27,584 |
| 535 | 1.32 [1.31-1.34] | 1.33 [1.31-1.35] | 1.33 [1.31-1.35] | 1.23 [1.17-1.28] | 1.22 [1.21-1.24] | 1.22 [1.21-1.24] | 1.19 [1.17-1.20] | 1.19 [1.17-1.20] | 1.16 [1.11-1.22] | 1.16 [1.15-1.18] | 69,990 | 66,962 | 59,204 | 5,411 | 60,277 |
| 535.1 | 1.41 [1.34-1.48] | 1.40 [1.33-1.47] | 1.40 [1.32-1.47] | 1.14 [0.93-1.38] | 1.27 [1.21-1.34] | 1.30 [1.24-1.37] | 1.24 [1.18-1.31] | 1.24 [1.17-1.31] | 1.11 [0.91-1.36] | 1.22 [1.15-1.29] | 3,754 | 3,588 | 3,256 | 257 | 3,299 |
| 535.2 | 1.43 [1.31-1.55] | 1.39 [1.28-1.51] | 1.40 [1.28-1.52] | | 1.32 [1.21-1.44] | 1.34 [1.23-1.45] | 1.28 [1.18-1.39] | 1.26 [1.16-1.38] | | 1.27 [1.16-1.38] | 1,556 | 1,553 | 1,492 | NA | 1,404 |
| 535.6 | 1.31 [1.28-1.34] | 1.31 [1.28-1.34] | 1.30 [1.27-1.33] | 1.30 [1.14-1.48] | 1.20 [1.16-1.23] | 1.23 [1.20-1.26] | 1.19 [1.16-1.22] | 1.18 [1.15-1.22] | 1.21 [1.05-1.38] | 1.16 [1.13-1.19] | 15,226 | 14,943 | 13,538 | 637 | 12,984 |
| 535.8 | 1.33 [1.31-1.36] | 1.34 [1.31-1.37] | 1.33 [1.30-1.36] | 1.35 [1.25-1.47] | 1.24 [1.21-1.27] | 1.23 [1.20-1.25] | 1.18 [1.16-1.21] | 1.18 [1.15-1.20] | 1.27 [1.17-1.38] | 1.17 [1.15-1.20] | 23,457 | 22,460 | 20,228 | 1,673 | 20,800 |
| 537 | 1.30 [1.27-1.33] | 1.31 [1.28-1.33] | 1.30 [1.27-1.33] | 1.11 [0.97-1.26] | 1.21 [1.18-1.23] | 1.18 [1.16-1.21] | 1.15 [1.12-1.17] | 1.14 [1.12-1.17] | 1.04 [0.91-1.19] | 1.13 [1.10-1.15] | 22,728 | 22,438 | 21,097 | 633 | 20,728 |
| 539 | 1.32 [0.99-1.75] | 1.41 [1.05-1.90] | | | 1.20 [0.90-1.59] | 1.11 [0.83-1.50] | 1.20 [0.88-1.63] | | | 1.11 [0.83-1.48] | 119 | 112 | NA | NA | 117 |
| 540 | 1.15 [1.07-1.24] | 1.16 [1.06-1.27] | 1.10 [0.97-1.25] | 1.08 [0.97-1.21] | 1.06 [0.98-1.15] | 1.10 [1.01-1.18] | 1.06 [0.97-1.16] | 1.05 [0.92-1.19] | 1.05 [0.94-1.18] | 1.04 [0.95-1.13] | 1,735 | 1,171 | 581 | 837 | 1,394 |
| 540.1 | 1.14 [1.08-1.21] | 1.14 [1.06-1.23] | 1.14 [1.01-1.29] | 1.10 [1.01-1.18] | 1.07 [1.00-1.14] | 1.08 [1.02-1.15] | 1.02 [0.95-1.10] | 1.06 [0.94-1.21] | 1.06 [0.98-1.15] | 1.03 [0.96-1.10] | 2,929 | 1,691 | 600 | 1,758 | 2,275 |
| 540.11 | 1.08 [1.05-1.11] | 1.09 [1.05-1.13] | 1.05 [1.00-1.11] | 1.04 [1.00-1.08] | 1.01 [0.98-1.04] | 1.04 [1.01-1.07] | 1.03 [1.00-1.07] | 1.01 [0.95-1.06] | 1.02 [0.98-1.06] | 0.99 [0.96-1.03] | 13,415 | 7,735 | 3,340 | 7,734 | 9,842 |
| 550 | 1.27 [1.23-1.31] | 1.29 [1.25-1.34] | 1.30 [1.25-1.34] | 1.13 [1.00-1.28] | 1.18 [1.14-1.22] | 1.17 [1.13-1.21] | 1.15 [1.11-1.19] | 1.15 [1.11-1.19] | 1.06 [0.93-1.21] | 1.12 [1.09-1.17] | 8,593 | 8,112 | 7,263 | 632 | 7,848 |
| 550.1 | 1.10 [1.07-1.12] | 1.11 [1.09-1.14] | 1.11 [1.08-1.13] | 1.02 [0.96-1.08] | 1.06 [1.04-1.09] | 1.06 [1.04-1.08] | 1.08 [1.06-1.10] | 1.07 [1.05-1.10] | 0.98 [0.93-1.04] | 1.04 [1.02-1.06] | 23,888 | 21,407 | 19,508 | 3,081 | 18,644 |
| 550.2 | 1.31 [1.29-1.32] | 1.32 [1.30-1.33] | 1.30 [1.29-1.32] | 1.27 [1.17-1.38] | 1.22 [1.20-1.23] | 1.20 [1.18-1.21] | 1.17 [1.16-1.19] | 1.16 [1.15-1.18] | 1.17 [1.08-1.28] | 1.14 [1.12-1.15] | 66,300 | 66,090 | 60,673 | 1,682 | 57,225 |
| 550.3 | 1.19 [1.10-1.28] | 1.18 [1.09-1.27] | 1.16 [1.08-1.26] | | 1.12 [1.03-1.21] | 1.13 [1.05-1.22] | 1.12 [1.04-1.21] | 1.10 [1.02-1.19] | | 1.07 [0.98-1.16] | 1,699 | 1,670 | 1,555 | NA | 1,454 |
| 550.4 | 1.29 [1.25-1.32] | 1.29 [1.26-1.33] | 1.29 [1.25-1.34] | 1.19 [1.10-1.28] | 1.20 [1.17-1.24] | 1.17 [1.14-1.21] | 1.14 [1.11-1.18] | 1.14 [1.10-1.17] | 1.09 [1.00-1.18] | 1.13 [1.09-1.16] | 11,993 | 10,583 | 8,923 | 1,665 | 10,301 |
| 550.5 | 1.27 [1.23-1.31] | 1.28 [1.24-1.32] | 1.29 [1.25-1.34] | 1.13 [1.00-1.27] | 1.18 [1.14-1.22] | 1.17 [1.13-1.20] | 1.14 [1.10-1.18] | 1.15 [1.11-1.19] | 1.05 [0.93-1.19] | 1.13 [1.09-1.16] | 10,222 | 9,728 | 8,749 | 664 | 9,462 |
| 555.1 | 1.70 [1.63-1.77] | 1.65 [1.58-1.72] | 1.56 [1.47-1.64] | 1.80 [1.65-1.96] | 1.55 [1.48-1.62] | 1.47 [1.41-1.54] | 1.35 [1.29-1.42] | 1.29 [1.22-1.36] | 1.67 [1.53-1.82] | 1.38 [1.32-1.45] | 6,324 | 5,403 | 3,344 | 1,728 | 5,217 |
| 555.2 | 1.51 [1.46-1.56] | 1.51 [1.46-1.57] | 1.45 [1.39-1.51] | 1.44 [1.31-1.58] | 1.36 [1.31-1.42] | 1.35 [1.30-1.40] | 1.29 [1.24-1.34] | 1.24 [1.19-1.30] | 1.36 [1.23-1.50] | 1.25 [1.20-1.30] | 8,057 | 7,573 | 5,375 | 1,254 | 6,659 |
| 555.21 | 1.62 [1.52-1.73] | 1.60 [1.50-1.71] | 1.57 [1.45-1.69] | 1.51 [1.30-1.77] | 1.44 [1.35-1.54] | 1.39 [1.30-1.49] | 1.30 [1.22-1.39] | 1.27 [1.17-1.38] | 1.43 [1.21-1.68] | 1.29 [1.21-1.38] | 2,605 | 2,415 | 1,624 | 490 | 2,356 |
| 556 | 1.49 [1.37-1.62] | 1.50 [1.38-1.64] | 1.45 [1.32-1.59] | 1.52 [1.16-1.98] | 1.36 [1.25-1.48] | 1.38 [1.26-1.50] | 1.35 [1.23-1.47] | 1.31 [1.19-1.45] | 1.44 [1.09-1.90] | 1.31 [1.20-1.43] | 1,455 | 1,391 | 1,105 | 173 | 1,293 |
| 556.1 | 1.54 [1.45-1.63] | 1.53 [1.44-1.63] | 1.46 [1.37-1.57] | 1.60 [1.36-1.88] | 1.40 [1.32-1.49] | 1.37 [1.29-1.46] | 1.30 [1.22-1.39] | 1.28 [1.19-1.37] | 1.48 [1.25-1.74] | 1.30 [1.22-1.38] | 2,973 | 2,814 | 2,066 | 458 | 2,642 |
| 557 | 1.78 [1.72-1.85] | 1.60 [1.53-1.67] | 1.61 [1.53-1.70] | 2.04 [1.93-2.17] | 1.67 [1.61-1.73] | 1.56 [1.50-1.62] | 1.36 [1.30-1.42] | 1.37 [1.30-1.44] | 1.76 [1.65-1.87] | 1.49 [1.43-1.55] | 8,443 | 5,443 | 3,833 | 3,599 | 7,356 |
| 557.1 | 1.57 [1.50-1.64] | 1.59 [1.52-1.67] | 1.62 [1.53-1.71] | 1.38 [1.26-1.50] | 1.45 [1.38-1.52] | 1.41 [1.35-1.47] | 1.38 [1.31-1.45] | 1.40 [1.32-1.48] | 1.30 [1.18-1.42] | 1.33 [1.27-1.40] | 5,403 | 4,409 | 3,104 | 1,479 | 4,508 |
| 558 | 1.35 [1.33-1.37] | 1.39 [1.37-1.41] | 1.37 [1.34-1.39] | 1.21 [1.16-1.25] | 1.25 [1.22-1.27] | 1.26 [1.24-1.29] | 1.24 [1.22-1.27] | 1.23 [1.21-1.25] | 1.17 [1.13-1.22] | 1.19 [1.17-1.21] | 37,902 | 32,476 | 26,597 | 7,035 | 31,731 |
| 559 | 1.29 [1.23-1.35] | 1.29 [1.23-1.36] | 1.22 [1.16-1.29] | 1.43 [1.17-1.74] | 1.21 [1.16-1.28] | 1.21 [1.15-1.27] | 1.17 [1.11-1.23] | 1.13 [1.07-1.19] | 1.32 [1.07-1.62] | 1.16 [1.11-1.23] | 4,140 | 4,029 | 3,460 | 279 | 3,698 |
| 560.1 | 1.18 [1.11-1.25] | 1.20 [1.13-1.27] | 1.20 [1.12-1.28] | 0.87 [0.70-1.08] | 1.14 [1.07-1.21] | 1.12 [1.06-1.19] | 1.11 [1.04-1.18] | 1.12 [1.05-1.19] | 0.84 [0.67-1.05] | 1.11 [1.04-1.18] | 2,870 | 2,729 | 2,425 | 226 | 2,599 |
| 560.2 | 1.23 [1.16-1.31] | 1.26 [1.18-1.35] | 1.28 [1.19-1.37] | 1.11 [0.96-1.28] | 1.16 [1.09-1.23] | 1.17 [1.10-1.24] | 1.13 [1.06-1.21] | 1.16 [1.08-1.24] | 1.07 [0.92-1.24] | 1.11 [1.04-1.18] | 2,820 | 2,405 | 2,190 | 498 | 2,602 |
| 560.3 | 1.19 [1.12-1.26] | 1.19 [1.12-1.27] | 1.17 [1.10-1.24] | 0.90 [0.69-1.17] | 1.09 [1.03-1.16] | 1.11 [1.05-1.18] | 1.08 [1.02-1.15] | 1.07 [1.01-1.14] | 0.85 [0.65-1.11] | 1.06 [1.00-1.12] | 2,843 | 2,772 | 2,543 | 141 | 2,633 |
| 560.4 | 1.21 [1.18-1.25] | 1.21 [1.18-1.25] | 1.19 [1.16-1.23] | 1.16 [1.03-1.31] | 1.14 [1.11-1.17] | 1.15 [1.12-1.19] | 1.12 [1.09-1.15] | 1.10 [1.07-1.14] | 1.11 [0.98-1.26] | 1.10 [1.07-1.14] | 12,494 | 12,100 | 11,131 | 705 | 11,306 |
| 561 | 1.32 [1.30-1.34] | 1.33 [1.31-1.35] | 1.32 [1.29-1.34] | 1.22 [1.15-1.30] | 1.23 [1.21-1.25] | 1.23 [1.21-1.25] | 1.20 [1.18-1.22] | 1.19 [1.17-1.21] | 1.16 [1.08-1.23] | 1.17 [1.15-1.19] | 40,513 | 39,486 | 34,220 | 2,734 | 35,935 |
| 561.1 | 1.62 [1.51-1.74] | 1.58 [1.46-1.70] | 1.56 [1.44-1.69] | 1.75 [1.42-2.14] | 1.51 [1.40-1.62] | 1.50 [1.39-1.61] | 1.40 [1.29-1.51] | 1.40 [1.29-1.52] | 1.53 [1.24-1.89] | 1.42 [1.31-1.53] | 2,038 | 1,820 | 1,646 | 276 | 1,878 |
| 561.2 | 1.31 [1.27-1.36] | 1.32 [1.27-1.37] | 1.30 [1.25-1.36] | 1.16 [1.04-1.30] | 1.22 [1.17-1.26] | 1.20 [1.16-1.25] | 1.15 [1.11-1.20] | 1.15 [1.10-1.20] | 1.11 [0.99-1.24] | 1.14 [1.10-1.19] | 7,064 | 6,608 | 5,225 | 903 | 6,356 |
| 562.1 | 1.25 [1.24-1.27] | 1.25 [1.23-1.26] | 1.25 [1.23-1.26] | 1.15 [0.93-1.43] | 1.17 [1.15-1.18] | 1.17 [1.16-1.19] | 1.14 [1.12-1.15] | 1.14 [1.12-1.15] | 1.06 [0.84-1.32] | 1.11 [1.10-1.13] | 72,988 | 72,950 | 71,810 | 235 | 63,834 |
| 563 | 1.27 [1.26-1.29] | 1.29 [1.27-1.30] | 1.27 [1.25-1.29] | 1.20 [1.16-1.23] | 1.20 [1.18-1.21] | 1.20 [1.18-1.21] | 1.17 [1.15-1.18] | 1.16 [1.14-1.18] | 1.15 [1.12-1.19] | 1.15 [1.14-1.17] | 73,619 | 64,164 | 56,852 | 12,054 | 66,813 |
| 564 | 1.27 [1.26-1.29] | 1.29 [1.27-1.30] | 1.27 [1.26-1.29] | 1.19 [1.16-1.23] | 1.20 [1.18-1.21] | 1.20 [1.18-1.21] | 1.17 [1.15-1.18] | 1.16 [1.14-1.18] | 1.15 [1.11-1.18] | 1.15 [1.14-1.17] | 75,465 | 65,892 | 58,307 | 12,257 | 68,411 |
| 564.1 | 1.50 [1.47-1.53] | 1.52 [1.49-1.55] | 1.53 [1.49-1.56] | 1.29 [1.21-1.37] | 1.39 [1.36-1.42] | 1.27 [1.24-1.30] | 1.19 [1.17-1.22] | 1.20 [1.17-1.23] | 1.21 [1.14-1.29] | 1.22 [1.19-1.25] | 24,237 | 23,632 | 16,259 | 3,011 | 21,882 |
| 564.8 | 1.28 [1.23-1.32] | 1.27 [1.23-1.32] | 1.26 [1.21-1.31] | 1.32 [1.11-1.58] | 1.20 [1.15-1.24] | 1.21 [1.16-1.25] | 1.17 [1.12-1.21] | 1.15 [1.11-1.20] | 1.26 [1.05-1.51] | 1.16 [1.11-1.20] | 7,607 | 7,483 | 6,803 | 353 | 6,879 |
| 564.9 | 1.31 [1.29-1.32] | 1.32 [1.30-1.34] | 1.30 [1.29-1.32] | 1.17 [1.12-1.23] | 1.20 [1.18-1.21] | 1.20 [1.18-1.21] | 1.16 [1.15-1.18] | 1.16 [1.14-1.17] | 1.09 [1.03-1.14] | 1.14 [1.12-1.15] | 57,424 | 54,908 | 47,972 | 4,667 | 53,093 |
| 565 | 1.40 [1.37-1.43] | 1.41 [1.38-1.44] | 1.38 [1.35-1.42] | 1.30 [1.22-1.38] | 1.29 [1.26-1.32] | 1.30 [1.28-1.33] | 1.27 [1.24-1.30] | 1.25 [1.22-1.28] | 1.25 [1.18-1.33] | 1.23 [1.20-1.26] | 21,100 | 19,711 | 14,491 | 2,957 | 17,782 |
| 565.1 | 1.24 [1.21-1.27] | 1.24 [1.21-1.28] | 1.23 [1.20-1.26] | 1.35 [1.13-1.61] | 1.15 [1.12-1.18] | 1.17 [1.14-1.20] | 1.13 [1.10-1.16] | 1.12 [1.09-1.15] | 1.29 [1.07-1.55] | 1.11 [1.08-1.14] | 15,436 | 15,327 | 14,143 | 342 | 13,376 |
| 567 | 1.19 [1.14-1.25] | 1.18 [1.13-1.24] | 1.17 [1.11-1.23] | 1.04 [0.87-1.25] | 1.13 [1.07-1.19] | 1.16 [1.10-1.21] | 1.10 [1.05-1.16] | 1.10 [1.04-1.16] | 1.05 [0.87-1.26] | 1.12 [1.06-1.18] | 4,231 | 4,044 | 3,566 | 335 | 3,710 |
| 568 | 1.19 [1.16-1.22] | 1.20 [1.17-1.23] | 1.18 [1.14-1.21] | 1.11 [1.02-1.21] | 1.11 [1.08-1.14] | 1.12 [1.09-1.15] | 1.09 [1.06-1.12] | 1.07 [1.04-1.11] | 1.07 [0.99-1.17] | 1.08 [1.05-1.11] | 14,367 | 13,909 | 10,436 | 1,499 | 12,929 |
| 568.1 | 1.20 [1.17-1.23] | 1.20 [1.17-1.24] | 1.18 [1.15-1.22] | 1.12 [1.03-1.22] | 1.12 [1.09-1.15] | 1.12 [1.09-1.15] | 1.09 [1.06-1.12] | 1.08 [1.04-1.11] | 1.08 [0.99-1.19] | 1.08 [1.05-1.11] | 13,670 | 13,248 | 9,849 | 1,391 | 12,346 |
| 569 | 1.28 [1.26-1.30] | 1.28 [1.26-1.30] | 1.26 [1.24-1.28] | 1.38 [1.27-1.50] | 1.19 [1.17-1.21] | 1.19 [1.17-1.21] | 1.15 [1.13-1.17] | 1.14 [1.12-1.15] | 1.28 [1.17-1.39] | 1.14 [1.12-1.16] | 40,631 | 39,999 | 36,652 | 1,586 | 36,108 |
| 569.1 | 1.24 [1.14-1.35] | 1.26 [1.15-1.37] | 1.24 [1.13-1.35] | | 1.19 [1.09-1.30] | 1.17 [1.07-1.27] | 1.14 [1.05-1.25] | 1.13 [1.03-1.24] | | 1.15 [1.06-1.26] | 1,433 | 1,394 | 1,310 | NA | 1,340 |
| 569.2 | 1.24 [1.16-1.34] | 1.25 [1.16-1.35] | 1.22 [1.12-1.32] | 1.16 [0.92-1.47] | 1.17 [1.08-1.26] | 1.16 [1.08-1.25] | 1.13 [1.04-1.22] | 1.11 [1.02-1.20] | 1.08 [0.85-1.38] | 1.12 [1.03-1.20] | 1,834 | 1,732 | 1,472 | 184 | 1,640 |
| 571 | 1.48 [1.41-1.55] | 1.51 [1.44-1.58] | 1.50 [1.43-1.58] | | 1.39 [1.32-1.46] | 1.37 [1.30-1.44] | 1.33 [1.26-1.40] | 1.31 [1.25-1.38] | | 1.34 [1.27-1.41] | 4,213 | 4,197 | 4,021 | NA | 3,916 |

### Supplementary Table 6: phecode-mapped results (hazard ratios and events from all cohorts)

| | Hazard ratio (99% confidence interval) | | | | | | | | | | | | | | |
|---|---|---|---|---|---|---|---|---|---|---|---|---|---|---|---|
| | crude | | | | | adjusted | | | | | Events (in exposed) | | | | |
| Outcome | any age | 18+ | 40+ | <18 | hosp. | any age | 18+ | 40+ | <18 | hosp. | any age | 18+ | 40+ | <18 | hosp. |
| 571.5 | 1.45 [1.41-1.49] | 1.45 [1.41-1.50] | 1.46 [1.42-1.50] | 1.19 [1.04-1.36] | 1.34 [1.30-1.38] | 1.29 [1.25-1.33] | 1.21 [1.17-1.25] | 1.20 [1.17-1.24] | 1.14 [0.99-1.31] | 1.23 [1.20-1.27] | 12,671 | 12,461 | 10,860 | 655 | 11,737 |
| 571.51 | 1.47 [1.40-1.54] | 1.50 [1.43-1.57] | 1.50 [1.43-1.57] | | 1.38 [1.32-1.45] | 1.36 [1.30-1.43] | 1.32 [1.26-1.39] | 1.31 [1.24-1.37] | | 1.33 [1.27-1.40] | 4,611 | 4,572 | 4,331 | NA | 4,277 |
| 571.6 | 1.44 [1.28-1.62] | 1.41 [1.25-1.59] | 1.46 [1.29-1.65] | | 1.34 [1.18-1.51] | 1.31 [1.16-1.48] | 1.25 [1.10-1.41] | 1.28 [1.13-1.45] | | 1.26 [1.11-1.43] | 697 | 687 | 654 | NA | 630 |
| 571.8 | 1.34 [1.28-1.40] | 1.35 [1.29-1.41] | 1.37 [1.31-1.44] | 0.85 [0.66-1.09] | 1.29 [1.23-1.35] | 1.27 [1.21-1.33] | 1.22 [1.17-1.28] | 1.24 [1.18-1.30] | 0.85 [0.65-1.09] | 1.27 [1.21-1.33] | 5,162 | 5,076 | 4,753 | 152 | 4,657 |
| 571.81 | 1.46 [1.38-1.54] | 1.47 [1.39-1.55] | 1.46 [1.38-1.54] | | 1.37 [1.29-1.44] | 1.36 [1.29-1.44] | 1.30 [1.23-1.37] | 1.28 [1.21-1.35] | | 1.33 [1.26-1.41] | 3,513 | 3,470 | 3,245 | NA | 3,220 |
| 572 | 1.20 [1.17-1.24] | 1.22 [1.18-1.26] | 1.22 [1.18-1.26] | 0.94 [0.81-1.10] | 1.17 [1.13-1.21] | 1.17 [1.14-1.21] | 1.15 [1.11-1.19] | 1.15 [1.11-1.19] | 0.93 [0.80-1.09] | 1.17 [1.13-1.21] | 9,491 | 9,236 | 8,539 | 441 | 8,376 |
| 573 | 1.33 [1.30-1.36] | 1.33 [1.31-1.36] | 1.33 [1.30-1.36] | 1.11 [1.00-1.23] | 1.25 [1.22-1.27] | 1.23 [1.20-1.25] | 1.17 [1.14-1.19] | 1.16 [1.14-1.19] | 1.06 [0.95-1.19] | 1.18 [1.16-1.21] | 22,960 | 22,590 | 20,335 | 975 | 20,969 |
| 573.1 | 1.28 [1.08-1.52] | 1.34 [1.13-1.59] | 1.46 [1.22-1.74] | | 1.29 [1.09-1.54] | 1.24 [1.04-1.48] | 1.24 [1.04-1.48] | 1.35 [1.12-1.62] | | 1.28 [1.07-1.53] | 360 | 355 | 347 | NA | 347 |
| 573.2 | 1.57 [1.31-1.89] | 1.68 [1.38-2.04] | 1.78 [1.43-2.20] | | 1.39 [1.16-1.68] | 1.43 [1.18-1.72] | 1.46 [1.19-1.79] | 1.56 [1.25-1.95] | | 1.32 [1.09-1.61] | 299 | 259 | 213 | NA | 275 |
| 573.3 | 1.38 [1.32-1.45] | 1.44 [1.38-1.51] | 1.41 [1.34-1.49] | 1.11 [0.99-1.24] | 1.27 [1.21-1.33] | 1.30 [1.24-1.36] | 1.28 [1.22-1.35] | 1.27 [1.20-1.34] | 1.08 [0.96-1.21] | 1.23 [1.17-1.29] | 5,156 | 4,624 | 3,839 | 829 | 4,592 |
| 573.5 | 1.22 [1.16-1.28] | 1.24 [1.18-1.31] | 1.23 [1.17-1.30] | 1.12 [0.94-1.33] | 1.19 [1.12-1.25] | 1.19 [1.13-1.25] | 1.18 [1.12-1.24] | 1.17 [1.11-1.24] | 1.13 [0.95-1.34] | 1.18 [1.12-1.25] | 3,810 | 3,566 | 3,200 | 362 | 3,246 |
| 573.7 | 1.26 [1.23-1.29] | 1.26 [1.23-1.29] | 1.26 [1.23-1.29] | 1.12 [1.03-1.22] | 1.19 [1.16-1.21] | 1.21 [1.18-1.23] | 1.17 [1.14-1.19] | 1.17 [1.14-1.19] | 1.09 [1.00-1.19] | 1.16 [1.14-1.19] | 20,802 | 20,176 | 17,720 | 1,533 | 18,675 |
| 573.9 | 1.21 [1.14-1.30] | 1.17 [1.09-1.26] | 1.21 [1.12-1.30] | 1.24 [1.05-1.46] | 1.15 [1.08-1.23] | 1.15 [1.07-1.23] | 1.06 [0.98-1.14] | 1.10 [1.02-1.19] | 1.14 [0.96-1.36] | 1.12 [1.04-1.20] | 2,353 | 2,104 | 1,804 | 390 | 2,149 |
| 574.1 | 1.16 [1.14-1.18] | 1.17 [1.15-1.19] | 1.15 [1.13-1.17] | 1.05 [0.97-1.12] | 1.10 [1.08-1.12] | 1.10 [1.09-1.12] | 1.08 [1.06-1.09] | 1.07 [1.05-1.08] | 1.02 [0.95-1.10] | 1.06 [1.04-1.08] | 39,352 | 39,031 | 32,181 | 1,957 | 34,401 |
| 574.11 | 1.14 [1.09-1.19] | 1.15 [1.11-1.20] | 1.13 [1.08-1.19] | 1.11 [0.90-1.38] | 1.06 [1.01-1.11] | 1.09 [1.04-1.14] | 1.07 [1.02-1.11] | 1.05 [1.00-1.10] | 1.12 [0.89-1.39] | 1.03 [0.98-1.08] | 5,154 | 5,128 | 4,363 | 220 | 4,513 |
| 574.12 | 1.15 [1.12-1.18] | 1.16 [1.13-1.19] | 1.14 [1.10-1.17] | 1.07 [0.96-1.19] | 1.09 [1.06-1.12] | 1.09 [1.06-1.12] | 1.07 [1.04-1.10] | 1.05 [1.02-1.08] | 1.04 [0.93-1.16] | 1.05 [1.02-1.08] | 13,627 | 13,549 | 10,395 | 881 | 11,886 |
| 574.2 | 1.12 [1.08-1.16] | 1.13 [1.09-1.17] | 1.11 [1.07-1.15] | 1.00 [0.86-1.17] | 1.07 [1.03-1.10] | 1.07 [1.03-1.11] | 1.04 [1.01-1.08] | 1.03 [0.99-1.07] | 0.98 [0.84-1.15] | 1.04 [1.00-1.08] | 8,555 | 8,498 | 6,989 | 445 | 7,591 |
| 574.3 | 1.19 [1.15-1.23] | 1.19 [1.15-1.24] | 1.19 [1.14-1.24] | 1.06 [0.89-1.26] | 1.13 [1.09-1.18] | 1.12 [1.08-1.17] | 1.09 [1.05-1.13] | 1.09 [1.04-1.13] | 1.03 [0.86-1.24] | 1.10 [1.05-1.14] | 7,288 | 7,250 | 6,041 | 326 | 6,430 |
| 575 | 1.18 [1.13-1.23] | 1.19 [1.14-1.24] | 1.18 [1.13-1.24] | 0.99 [0.84-1.17] | 1.11 [1.06-1.16] | 1.11 [1.07-1.16] | 1.09 [1.04-1.14] | 1.08 [1.03-1.14] | 0.96 [0.81-1.15] | 1.07 [1.02-1.12] | 5,528 | 5,479 | 4,323 | 344 | 4,861 |
| 575.1 | 1.19 [1.13-1.26] | 1.21 [1.15-1.28] | 1.18 [1.12-1.25] | | 1.14 [1.08-1.21] | 1.15 [1.08-1.21] | 1.12 [1.06-1.19] | 1.10 [1.04-1.17] | | 1.12 [1.06-1.18] | 3,424 | 3,395 | 3,235 | NA | 3,222 |
| 575.2 | 1.12 [1.06-1.17] | 1.12 [1.06-1.17] | 1.12 [1.06-1.18] | 0.99 [0.78-1.25] | 1.09 [1.04-1.15] | 1.10 [1.05-1.16] | 1.07 [1.02-1.13] | 1.07 [1.01-1.13] | 0.98 [0.77-1.25] | 1.09 [1.03-1.15] | 3,876 | 3,819 | 3,208 | 174 | 3,365 |
| 575.6 | 1.12 [1.02-1.23] | 1.12 [1.02-1.23] | 1.11 [0.99-1.24] | | 1.07 [0.97-1.17] | 1.04 [0.94-1.14] | 1.01 [0.92-1.11] | 1.00 [0.89-1.12] | | 1.00 [0.91-1.11] | 1,095 | 1,093 | 688 | NA | 958 |
| 575.7 | 1.19 [1.14-1.25] | 1.20 [1.15-1.26] | 1.19 [1.14-1.26] | 1.04 [0.86-1.27] | 1.11 [1.06-1.17] | 1.13 [1.08-1.18] | 1.11 [1.06-1.16] | 1.10 [1.04-1.16] | 1.01 [0.82-1.24] | 1.08 [1.03-1.14] | 4,563 | 4,517 | 3,721 | 260 | 4,022 |
| 575.8 | 1.16 [1.12-1.19] | 1.17 [1.13-1.20] | 1.16 [1.12-1.20] | 1.03 [0.88-1.21] | 1.12 [1.09-1.16] | 1.11 [1.08-1.15] | 1.08 [1.05-1.12] | 1.08 [1.04-1.11] | 1.00 [0.85-1.17] | 1.10 [1.06-1.14] | 10,320 | 10,204 | 9,066 | 394 | 9,357 |
| 575.9 | 1.14 [1.05-1.23] | 1.12 [1.03-1.21] | 1.14 [1.05-1.23] | | 1.09 [1.00-1.18] | 1.11 [1.02-1.20] | 1.04 [0.96-1.13] | 1.07 [0.98-1.16] | | 1.08 [0.99-1.17] | 1,554 | 1,523 | 1,424 | NA | 1,382 |
| 577 | 1.23 [1.16-1.30] | 1.25 [1.18-1.32] | 1.26 [1.19-1.33] | 0.85 [0.67-1.09] | 1.16 [1.09-1.22] | 1.16 [1.09-1.22] | 1.12 [1.05-1.18] | 1.13 [1.07-1.20] | 0.84 [0.66-1.08] | 1.12 [1.06-1.19] | 3,389 | 3,298 | 3,114 | 158 | 3,154 |
| 577.1 | 1.20 [1.16-1.25] | 1.21 [1.17-1.26] | 1.19 [1.14-1.24] | 1.03 [0.89-1.18] | 1.13 [1.09-1.18] | 1.14 [1.10-1.18] | 1.11 [1.07-1.15] | 1.08 [1.04-1.13] | 1.02 [0.88-1.18] | 1.10 [1.06-1.15] | 7,005 | 6,863 | 5,652 | 497 | 6,079 |
| 577.2 | 1.30 [1.22-1.39] | 1.35 [1.26-1.44] | 1.31 [1.21-1.41] | | 1.20 [1.12-1.29] | 1.22 [1.13-1.30] | 1.17 [1.09-1.26] | 1.13 [1.05-1.22] | | 1.18 [1.10-1.26] | 2,132 | 2,104 | 1,822 | NA | 1,943 |
| 577.3 | 1.16 [1.07-1.25] | 1.16 [1.08-1.26] | 1.15 [1.06-1.24] | | 1.06 [0.98-1.15] | 1.10 [1.02-1.19] | 1.08 [0.99-1.16] | 1.06 [0.97-1.15] | | 1.04 [0.96-1.13] | 1,616 | 1,602 | 1,492 | NA | 1,478 |
| 578.1 | 1.20 [1.17-1.24] | 1.23 [1.20-1.27] | 1.21 [1.17-1.25] | 1.10 [1.01-1.21] | 1.12 [1.08-1.15] | 1.14 [1.11-1.18] | 1.12 [1.09-1.16] | 1.11 [1.08-1.15] | 1.07 [0.97-1.17] | 1.09 [1.06-1.13] | 11,643 | 10,993 | 9,429 | 1,234 | 10,327 |
| 578.2 | 1.25 [1.22-1.29] | 1.25 [1.22-1.29] | 1.25 [1.22-1.29] | 1.15 [1.02-1.29] | 1.17 [1.14-1.21] | 1.20 [1.16-1.23] | 1.15 [1.12-1.19] | 1.15 [1.11-1.19] | 1.07 [0.95-1.21] | 1.15 [1.11-1.18] | 11,586 | 11,080 | 10,263 | 766 | 10,329 |
| 578.8 | 1.39 [1.36-1.43] | 1.39 [1.36-1.43] | 1.37 [1.34-1.41] | 1.34 [1.23-1.47] | 1.26 [1.23-1.30] | 1.30 [1.26-1.33] | 1.25 [1.22-1.29] | 1.24 [1.21-1.28] | 1.26 [1.15-1.38] | 1.20 [1.17-1.24] | 16,108 | 15,501 | 12,518 | 1,407 | 13,033 |
| 578.9 | 1.32 [1.30-1.34] | 1.33 [1.31-1.35] | 1.30 [1.28-1.33] | 1.24 [1.17-1.32] | 1.23 [1.21-1.25] | 1.23 [1.21-1.25] | 1.19 [1.17-1.22] | 1.17 [1.15-1.20] | 1.17 [1.10-1.25] | 1.18 [1.16-1.20] | 32,769 | 31,805 | 26,516 | 2,905 | 29,328 |
| 579 | 1.11 [1.05-1.16] | 1.11 [1.06-1.17] | 1.09 [1.03-1.15] | 1.02 [0.86-1.22] | 1.07 [1.01-1.13] | 1.09 [1.04-1.15] | 1.06 [1.01-1.12] | 1.05 [0.99-1.11] | 1.02 [0.86-1.23] | 1.07 [1.01-1.13] | 3,647 | 3,445 | 3,048 | 318 | 3,157 |
| 579.2 | 1.38 [1.32-1.45] | 1.44 [1.38-1.51] | 1.41 [1.34-1.49] | 1.11 [0.99-1.24] | 1.27 [1.21-1.33] | 1.30 [1.24-1.36] | 1.28 [1.22-1.35] | 1.27 [1.20-1.34] | 1.08 [0.96-1.21] | 1.23 [1.17-1.29] | 5,156 | 4,624 | 3,839 | 829 | 4,592 |
| 579.8 | 1.32 [1.27-1.37] | 1.35 [1.31-1.41] | 1.32 [1.26-1.37] | 1.18 [1.08-1.29] | 1.25 [1.20-1.30] | 1.22 [1.18-1.27] | 1.20 [1.16-1.25] | 1.18 [1.13-1.23] | 1.13 [1.02-1.24] | 1.19 [1.14-1.23] | 7,913 | 7,159 | 5,915 | 1,249 | 7,191 |
| genitourinary | | | | | | | | | | | | | | | |
| 580 | 1.34 [1.23-1.47] | 1.48 [1.33-1.63] | 1.46 [1.30-1.62] | 1.01 [0.85-1.21] | 1.23 [1.12-1.36] | 1.29 [1.17-1.41] | 1.35 [1.22-1.50] | 1.33 [1.19-1.49] | 1.01 [0.84-1.21] | 1.20 [1.09-1.33] | 1,254 | 973 | 825 | 316 | 1,065 |
| 580.11 | 1.38 [1.13-1.69] | 1.38 [1.10-1.72] | 1.39 [1.10-1.76] | | 1.24 [1.00-1.54] | 1.35 [1.10-1.66] | 1.29 [1.03-1.62] | 1.33 [1.04-1.68] | | 1.24 [0.99-1.54] | 242 | 206 | 176 | NA | 212 |
| 580.12 | 1.54 [1.35-1.75] | 1.56 [1.36-1.78] | 1.47 [1.27-1.70] | | 1.36 [1.19-1.56] | 1.45 [1.27-1.66] | 1.44 [1.25-1.66] | 1.34 [1.15-1.56] | | 1.31 [1.14-1.51] | 595 | 551 | 470 | NA | 522 |
| 580.14 | 1.34 [1.28-1.39] | 1.34 [1.29-1.40] | 1.35 [1.29-1.41] | | 1.26 [1.21-1.32] | 1.27 [1.22-1.32] | 1.22 [1.17-1.27] | 1.22 [1.17-1.28] | | 1.22 [1.17-1.28] | 6,043 | 6,015 | 5,937 | NA | 5,629 |
| 580.2 | 1.42 [1.35-1.49] | 1.47 [1.39-1.55] | 1.45 [1.37-1.53] | 1.02 [0.86-1.21] | 1.28 [1.21-1.35] | 1.34 [1.27-1.41] | 1.29 [1.22-1.36] | 1.26 [1.19-1.33] | 1.02 [0.86-1.21] | 1.26 [1.19-1.33] | 3,904 | 3,623 | 3,315 | 349 | 3,595 |
| 580.31 | 1.43 [1.35-1.52] | 1.45 [1.36-1.53] | 1.43 [1.35-1.52] | | 1.28 [1.21-1.36] | 1.35 [1.27-1.43] | 1.26 [1.18-1.33] | 1.22 [1.15-1.31] | | 1.26 [1.19-1.34] | 2,958 | 2,908 | 2,705 | NA | 2,814 |
| 580.32 | 1.47 [1.36-1.59] | 1.48 [1.36-1.62] | 1.48 [1.35-1.63] | 1.31 [1.10-1.56] | 1.34 [1.23-1.46] | 1.40 [1.29-1.53] | 1.35 [1.23-1.48] | 1.34 [1.22-1.48] | 1.26 [1.05-1.51] | 1.30 [1.20-1.42] | 1,569 | 1,272 | 1,077 | 350 | 1,408 |
| 585.1 | 1.23 [1.21-1.24] | 1.22 [1.21-1.23] | 1.23 [1.21-1.24] | 1.07 [0.98-1.17] | 1.17 [1.16-1.19] | 1.19 [1.17-1.20] | 1.13 [1.12-1.15] | 1.14 [1.12-1.15] | 1.07 [0.97-1.17] | 1.16 [1.15-1.18] | 76,985 | 76,432 | 74,248 | 1,369 | 71,764 |
| 585.2 | 1.30 [1.26-1.34] | 1.31 [1.27-1.36] | 1.29 [1.25-1.34] | 1.08 [0.91-1.29] | 1.23 [1.19-1.27] | 1.24 [1.20-1.28] | 1.21 [1.17-1.25] | 1.19 [1.15-1.23] | 1.05 [0.87-1.26] | 1.20 [1.16-1.24] | 9,785 | 9,554 | 9,235 | 316 | 8,723 |
| 585.3 | 1.26 [1.24-1.28] | 1.26 [1.24-1.28] | 1.25 [1.24-1.27] | 1.14 [0.97-1.34] | 1.19 [1.17-1.20] | 1.21 [1.19-1.22] | 1.15 [1.14-1.17] | 1.15 [1.13-1.16] | 1.12 [0.95-1.33] | 1.16 [1.15-1.18] | 65,241 | 65,046 | 64,301 | 393 | 61,464 |
| 585.31 | 1.42 [1.34-1.50] | 1.42 [1.34-1.50] | 1.39 [1.31-1.48] | 1.11 [0.85-1.45] | 1.24 [1.17-1.31] | 1.38 [1.30-1.46] | 1.30 [1.22-1.38] | 1.26 [1.18-1.34] | 1.11 [0.84-1.46] | 1.24 [1.17-1.32] | 3,115 | 3,038 | 2,793 | 143 | 2,877 |
| 586 | 1.16 [1.11-1.21] | 1.18 [1.13-1.23] | 1.17 [1.12-1.22] | 1.03 [0.91-1.16] | 1.08 [1.03-1.12] | 1.11 [1.06-1.16] | 1.08 [1.04-1.13] | 1.07 [1.02-1.12] | 1.02 [0.90-1.15] | 1.06 [1.01-1.10] | 5,806 | 5,314 | 4,760 | 665 | 5,263 |
| 586.11 | 1.23 [1.14-1.33] | 1.23 [1.14-1.33] | 1.24 [1.14-1.35] | 0.98 [0.78-1.22] | 1.11 [1.03-1.20] | 1.19 [1.10-1.28] | 1.13 [1.04-1.23] | 1.13 [1.04-1.24] | 0.96 [0.76-1.21] | 1.10 [1.02-1.20] | 1,692 | 1,553 | 1,408 | 194 | 1,574 |
| 586.12 | 1.08 [0.93-1.24] | 1.17 [0.89-1.52] | | 1.04 [0.88-1.22] | 0.98 [0.85-1.14] | 1.04 [0.90-1.20] | 1.06 [0.81-1.40] | | 1.01 [0.85-1.19] | 0.97 [0.83-1.12] | 472 | 133 | NA | 357 | 438 |
| 586.2 | 1.19 [1.16-1.23] | 1.19 [1.15-1.23] | 1.20 [1.16-1.24] | 0.88 [0.70-1.11] | 1.13 [1.09-1.17] | 1.14 [1.11-1.18] | 1.10 [1.06-1.14] | 1.11 [1.07-1.14] | 0.86 [0.68-1.10] | 1.11 [1.07-1.15] | 9,727 | 9,633 | 9,308 | 183 | 8,957 |
| 586.3 | 1.25 [1.10-1.43] | 1.25 [1.09-1.42] | 1.21 [1.06-1.39] | | 1.13 [0.98-1.29] | 1.22 [1.07-1.40] | 1.14 [0.99-1.31] | 1.11 [0.97-1.28] | | 1.12 [0.97-1.29] | 563 | 554 | 529 | NA | 511 |
| 586.4 | 1.07 [1.01-1.13] | 1.08 [1.02-1.14] | 1.07 [1.01-1.13] | 0.99 [0.84-1.16] | 1.01 [0.95-1.07] | 1.05 [1.00-1.11] | 1.04 [0.98-1.10] | 1.04 [0.98-1.10] | 0.98 [0.83-1.15] | 1.02 [0.96-1.08] | 3,404 | 3,160 | 2,732 | 390 | 3,021 |
| 587 | 1.39 [1.26-1.54] | 1.45 [1.31-1.61] | 1.40 [1.24-1.57] | 1.07 [0.80-1.43] | 1.15 [1.04-1.27] | 1.36 [1.23-1.51] | 1.35 [1.22-1.51] | 1.26 [1.12-1.42] | 1.04 [0.77-1.40] | 1.18 [1.07-1.31] | 980 | 905 | 712 | 117 | 909 |
| 588 | 1.36 [1.20-1.53] | 1.42 [1.25-1.61] | 1.47 [1.29-1.68] | | 1.19 [1.05-1.35] | 1.30 [1.15-1.48] | 1.31 [1.15-1.49] | 1.34 [1.17-1.54] | | 1.18 [1.04-1.34] | 667 | 632 | 584 | NA | 618 |
| 589 | 1.25 [1.13-1.38] | 1.22 [1.10-1.35] | 1.20 [1.08-1.34] | 1.09 [0.80-1.48] | 1.13 [1.02-1.25] | 1.20 [1.08-1.33] | 1.12 [1.01-1.25] | 1.08 [0.97-1.21] | 1.12 [0.82-1.54] | 1.11 [1.00-1.23] | 1,044 | 956 | 864 | 118 | 953 |
| 590 | 1.17 [1.13-1.20] | 1.19 [1.15-1.23] | 1.23 [1.18-1.28] | 1.05 [0.99-1.12] | 1.08 [1.04-1.12] | 1.10 [1.07-1.14] | 1.07 [1.03-1.11] | 1.11 [1.06-1.16] | 1.03 [0.96-1.10] | 1.05 [1.02-1.09] | 9,587 | 8,432 | 5,143 | 2,655 | 8,346 |
| 591 | 1.19 [1.17-1.20] | 1.19 [1.17-1.20] | 1.19 [1.17-1.20] | 1.11 [1.07-1.15] | 1.13 [1.11-1.14] | 1.15 [1.13-1.16] | 1.11 [1.09-1.12] | 1.11 [1.10-1.13] | 1.08 [1.04-1.12] | 1.11 [1.09-1.12] | 90,056 | 84,342 | 77,587 | 8,358 | 80,678 |
| 592.1 | 1.27 [1.22-1.32] | 1.27 [1.22-1.31] | 1.23 [1.18-1.28] | 1.40 [1.21-1.62] | 1.16 [1.12-1.21] | 1.20 [1.16-1.25] | 1.17 [1.12-1.21] | 1.14 [1.09-1.18] | 1.33 [1.15-1.55] | 1.13 [1.09-1.17] | 7,370 | 7,189 | 6,222 | 517 | 6,559 |
| 592.11 | 1.28 [1.06-1.53] | 1.26 [1.05-1.52] | 1.26 [1.04-1.52] | | 1.17 [0.97-1.41] | 1.25 [1.04-1.50] | 1.17 [0.97-1.42] | 1.20 [0.99-1.46] | | 1.17 [0.96-1.42] | 293 | 284 | 260 | NA | 266 |
| 592.12 | 1.31 [1.23-1.40] | 1.30 [1.22-1.39] | 1.29 [1.20-1.38] | | 1.19 [1.11-1.28] | 1.24 [1.16-1.33] | 1.20 [1.12-1.29] | 1.19 [1.10-1.28] | | 1.16 [1.08-1.24] | 2,115 | 2,088 | 1,855 | NA | 1,906 |
| 592.13 | 1.45 [1.28-1.64] | 1.43 [1.26-1.62] | 1.31 [1.12-1.53] | | 1.30 [1.14-1.48] | 1.31 [1.15-1.48] | 1.24 [1.09-1.41] | 1.15 [0.98-1.35] | | 1.20 [1.05-1.37] | 657 | 639 | 393 | NA | 594 |
| 592.2 | 1.35 [1.16-1.58] | 1.34 [1.13-1.59] | 1.25 [1.03-1.52] | | 1.12 [0.94-1.33] | 1.30 [1.11-1.53] | 1.25 [1.05-1.49] | 1.19 [0.97-1.45] | | 1.09 [0.91-1.29] | 404 | 334 | 251 | NA | 327 |
| 593 | 1.20 [1.18-1.22] | 1.20 [1.18-1.22] | 1.19 [1.17-1.21] | 1.12 [1.05-1.20] | 1.12 [1.10-1.14] | 1.15 [1.13-1.17] | 1.12 [1.10-1.14] | 1.12 [1.10-1.14] | 1.08 [1.01-1.15] | 1.09 [1.07-1.12] | 32,057 | 30,628 | 27,600 | 2,331 | 27,062 |
| 594 | 1.15 [1.11-1.18] | 1.15 [1.12-1.19] | 1.15 [1.11-1.19] | 0.94 [0.83-1.05] | 1.07 [1.04-1.10] | 1.10 [1.07-1.13] | 1.07 [1.04-1.10] | 1.07 [1.03-1.10] | 0.92 [0.81-1.03] | 1.05 [1.02-1.08] | 10,973 | 10,813 | 8,693 | 738 | 9,287 |
| 594.1 | 1.16 [1.12-1.20] | 1.17 [1.13-1.21] | 1.16 [1.12-1.21] | 0.97 [0.84-1.11] | 1.07 [1.03-1.11] | 1.11 [1.07-1.15] | 1.07 [1.04-1.11] | 1.07 [1.03-1.11] | 0.94 [0.81-1.08] | 1.05 [1.01-1.08] | 8,334 | 8,207 | 6,743 | 513 | 7,256 |
| 594.2 | 1.02 [0.96-1.08] | 1.01 [0.95-1.08] | 1.03 [0.96-1.10] | | 0.95 [0.89-1.02] | 1.00 [0.94-1.07] | 0.98 [0.92-1.05] | 1.00 [0.94-1.07] | | 0.96 [0.90-1.02] | 2,439 | 2,414 | 2,229 | NA | 2,124 |
| 594.3 | 1.11 [1.06-1.16] | 1.11 [1.06-1.16] | 1.10 [1.05-1.17] | 0.84 [0.70-1.02] | 1.02 [0.97-1.07] | 1.06 [1.01-1.11] | 1.03 [0.98-1.09] | 1.02 [0.97-1.08] | 0.82 [0.67-1.00] | 1.00 [0.95-1.06] | 3,964 | 3,923 | 3,048 | 261 | 3,273 |
| 594.8 | 1.17 [1.11-1.25] | 1.19 [1.12-1.26] | 1.14 [1.05-1.23] | 1.03 [0.87-1.22] | 1.05 [0.98-1.12] | 1.08 [1.02-1.15] | 1.06 [1.00-1.13] | 1.01 [0.94-1.09] | 0.98 [0.83-1.18] | 1.01 [0.94-1.08] | 2,585 | 2,517 | 1,569 | 334 | 2,068 |
| 595 | 1.05 [1.02-1.08] | 1.06 [1.02-1.09] | 1.06 [1.02-1.09] | 1.03 [0.92-1.15] | 1.00 [0.97-1.04] | 1.04 [1.01-1.07] | 1.02 [0.99-1.06] | 1.03 [1.00-1.07] | 1.00 [0.90-1.12] | 1.01 [0.98-1.05] | 10,444 | 10,029 | 8,820 | 863 | 9,209 |
| 596 | 1.23 [1.20-1.26] | 1.22 [1.20-1.25] | 1.22 [1.19-1.25] | 1.15 [1.05-1.27] | 1.14 [1.12-1.17] | 1.17 [1.14-1.19] | 1.13 [1.11-1.16] | 1.14 [1.11-1.16] | 1.10 [0.99-1.21] | 1.11 [1.09-1.14] | 21,205 | 20,503 | 18,963 | 1,124 | 18,799 |
| 596.1 | 1.21 [1.13-1.29] | 1.20 [1.13-1.28] | 1.22 [1.14-1.30] | | 1.13 [1.06-1.21] | 1.15 [1.08-1.23] | 1.12 [1.05-1.19] | 1.13 [1.06-1.21] | | 1.10 [1.02-1.17] | 2,419 | 2,403 | 2,326 | NA | 2,096 |
| 596.5 | 1.35 [1.26-1.43] | 1.39 [1.30-1.48] | 1.40 [1.30-1.51] | 0.98 [0.82-1.16] | 1.19 [1.12-1.27] | 1.24 [1.16-1.32] | 1.20 [1.12-1.28] | 1.22 [1.13-1.31] | 0.95 [0.79-1.13] | 1.13 [1.06-1.21] | 2,392 | 2,148 | 1,849 | 319 | 2,190 |
| 597 | 1.23 [1.15-1.32] | 1.29 [1.20-1.38] | 1.23 [1.14-1.32] | 1.04 [0.86-1.25] | 1.15 [1.07-1.23] | 1.18 [1.10-1.26] | 1.21 [1.12-1.29] | 1.15 [1.07-1.24] | 1.00 [0.83-1.22] | 1.12 [1.04-1.20] | 2,182 | 1,971 | 1,700 | 293 | 1,908 |
| 597.1 | 1.31 [1.26-1.36] | 1.30 [1.26-1.35] | 1.30 [1.25-1.35] | 1.30 [1.17-1.45] | 1.20 [1.15-1.25] | 1.24 [1.20-1.29] | 1.21 [1.16-1.25] | 1.20 [1.16-1.25] | 1.26 [1.13-1.41] | 1.17 [1.12-1.21] | 7,751 | 7,187 | 6,246 | 879 | 6,658 |
| 597.2 | 0.99 [0.83-1.18] | 1.02 [0.86-1.21] | 1.02 [0.85-1.22] | | 0.96 [0.81-1.15] | 0.97 [0.81-1.16] | 0.97 [0.82-1.16] | 0.98 [0.81-1.17] | | 0.97 [0.81-1.16] | 321 | 310 | 281 | NA | 303 |
| 598 | 1.33 [1.28-1.38] | 1.35 [1.30-1.41] | 1.33 [1.27-1.38] | 1.24 [1.09-1.41] | 1.21 [1.16-1.26] | 1.23 [1.18-1.28] | 1.20 [1.15-1.25] | 1.18 [1.13-1.23] | 1.17 [1.03-1.34] | 1.15 [1.10-1.19] | 6,555 | 6,226 | 5,317 | 651 | 5,794 |
| 598.9 | 1.20 [1.13-1.28] | 1.21 [1.13-1.30] | 1.16 [1.07-1.25] | 1.18 [1.03-1.35] | 1.12 [1.05-1.20] | 1.15 [1.07-1.22] | 1.10 [1.03-1.18] | 1.06 [0.98-1.15] | 1.15 [1.00-1.32] | 1.10 [1.02-1.17] | 2,432 | 2,073 | 1,563 | 562 | 2,206 |
| 599 | 1.21 [1.18-1.23] | 1.21 [1.19-1.24] | 1.19 [1.17-1.22] | 1.14 [1.05-1.25] | 1.13 [1.10-1.15] | 1.16 [1.14-1.19] | 1.13 [1.11-1.15] | 1.11 [1.09-1.14] | 1.10 [1.00-1.21] | 1.11 [1.09-1.13] | 24,847 | 24,175 | 22,258 | 1,266 | 22,715 |
| 599.1 | 1.06 [1.03-1.09] | 1.07 [1.04-1.10] | 1.07 [1.04-1.10] | 1.05 [0.97-1.15] | 1.01 [0.99-1.04] | 1.05 [1.02-1.08] | 1.03 [1.01-1.06] | 1.04 [1.01-1.07] | 1.02 [0.94-1.12] | 1.02 [0.99-1.05] | 13,110 | 12,270 | 10,762 | 1,400 | 11,521 |
| 599.2 | 1.17 [1.15-1.19] | 1.16 [1.14-1.18] | 1.16 [1.14-1.18] | 1.16 [1.08-1.25] | 1.12 [1.10-1.14] | 1.13 [1.11-1.15] | 1.10 [1.08-1.11] | 1.10 [1.08-1.12] | 1.10 [1.02-1.19] | 1.10 [1.09-1.12] | 43,140 | 42,266 | 39,647 | 1,817 | 39,109 |
| 599.3 | 1.35 [1.29-1.41] | 1.34 [1.28-1.41] | 1.36 [1.29-1.44] | 1.18 [1.08-1.29] | 1.21 [1.16-1.27] | 1.24 [1.19-1.30] | 1.17 [1.11-1.23] | 1.18 [1.11-1.25] | 1.14 [1.05-1.25] | 1.14 [1.09-1.20] | 5,315 | 4,379 | 3,136 | 1,412 | 4,639 |
| 599.4 | 1.23 [1.21-1.26] | 1.23 [1.21-1.26] | 1.22 [1.19-1.24] | 1.20 [1.12-1.29] | 1.16 [1.14-1.18] | 1.18 [1.15-1.20] | 1.14 [1.11-1.16] | 1.13 [1.10-1.15] | 1.14 [1.06-1.23] | 1.13 [1.11-1.16] | 31,011 | 29,430 | 27,205 | 2,064 | 28,402 |
| 599.5 | 1.35 [1.30-1.40] | 1.33 [1.29-1.38] | 1.32 [1.27-1.38] | 1.22 [1.10-1.35] | 1.23 [1.19-1.28] | 1.26 [1.21-1.30] | 1.20 [1.15-1.25] | 1.19 [1.14-1.24] | 1.19 [1.07-1.32] | 1.17 [1.12-1.21] | 7,985 | 7,287 | 6,145 | 1,046 | 6,959 |
| 599.6 | 1.21 [1.11-1.33] | 1.30 [1.18-1.43] | 1.26 [1.14-1.39] | 0.95 [0.73-1.23] | 1.23 [1.11-1.35] | 1.15 [1.05-1.27] | 1.17 [1.06-1.30] | 1.15 [1.03-1.27] | 0.92 [0.71-1.21] | 1.20 [1.09-1.32] | 1,225 | 1,113 | 1,027 | 145 | 1,131 |
| 599.7 | 1.28 [1.00-1.63] | 1.35 [1.04-1.75] | 1.39 [1.05-1.85] | | 1.19 [0.92-1.54] | 1.25 [0.97-1.60] | 1.23 [0.94-1.61] | 1.24 [0.92-1.66] | | 1.15 [0.88-1.48] | 176 | 156 | 132 | NA | 155 |
| 599.9 | 1.39 [1.31-1.47] | 1.35 [1.27-1.43] | 1.33 [1.25-1.42] | 1.47 [1.25-1.74] | 1.24 [1.17-1.32] | 1.28 [1.21-1.36] | 1.20 [1.13-1.28] | 1.19 [1.11-1.27] | 1.44 [1.21-1.70] | 1.18 [1.11-1.26] | 2,972 | 2,692 | 2,310 | 412 | 2,602 |
| 600 | 1.16 [1.14-1.19] | 1.16 [1.14-1.18] | 1.16 [1.14-1.18] | | 1.10 [1.08-1.12] | 1.12 [1.10-1.14] | 1.10 [1.08-1.12] | 1.10 [1.08-1.12] | | 1.07 [1.05-1.09] | 30,069 | 30,057 | 29,975 | NA | 26,000 |
| 601.1 | 1.15 [1.08-1.23] | 1.14 [1.07-1.22] | 1.11 [1.03-1.18] | | 1.04 [0.97-1.12] | 1.10 [1.02-1.17] | 1.08 [1.00-1.15] | 1.05 [0.98-1.12] | | 1.01 [0.93-1.08] | 2,047 | 2,045 | 1,963 | NA | 1,726 |
| 601.11 | 1.09 [0.93-1.28] | 1.10 [0.94-1.29] | 1.07 [0.91-1.25] | | 0.95 [0.80-1.13] | 1.05 [0.90-1.24] | 1.04 [0.89-1.22] | 1.01 [0.86-1.19] | | 0.92 [0.77-1.10] | 362 | 362 | 356 | NA | 300 |
| 601.12 | 1.08 [0.98-1.19] | 1.07 [0.97-1.17] | 1.03 [0.94-1.14] | | 0.97 [0.88-1.08] | 1.03 [0.94-1.14] | 1.02 [0.93-1.13] | 1.00 [0.91-1.10] | | 0.94 [0.85-1.04] | 984 | 985 | 957 | NA | 840 |
| 601.3 | 1.21 [1.15-1.28] | 1.26 [1.18-1.33] | 1.25 [1.16-1.35] | 1.12 [1.02-1.23] | 1.11 [1.04-1.18] | 1.15 [1.09-1.22] | 1.15 [1.08-1.22] | 1.15 [1.07-1.24] | 1.09 [0.99-1.20] | 1.09 [1.02-1.15] | 3,388 | 2,598 | 1,768 | 1,211 | 2,758 |
| 601.4 | 1.45 [1.35-1.56] | 1.55 [1.37-1.75] | 1.54 [1.35-1.76] | 1.37 [1.25-1.50] | 1.31 [1.21-1.43] | 1.40 [1.30-1.51] | 1.40 [1.24-1.58] | 1.41 [1.23-1.61] | 1.33 [1.21-1.46] | 1.29 [1.18-1.40] | 1,874 | 694 | 572 | 1,233 | 1,539 |
| 601.8 | 1.36 [1.25-1.49] | 1.39 [1.26-1.53] | 1.37 [1.23-1.52] | 1.28 [1.08-1.52] | 1.22 [1.11-1.34] | 1.31 [1.20-1.43] | 1.28 [1.16-1.41] | 1.25 [1.12-1.39] | 1.25 [1.05-1.48] | 1.21 [1.10-1.33] | 1,355 | 1,073 | 883 | 362 | 1,125 |
| 602 | 1.11 [1.05-1.18] | 1.11 [1.05-1.18] | 1.13 [1.06-1.20] | | 1.03 [0.97-1.10] | 1.07 [1.00-1.13] | 1.06 [1.00-1.13] | 1.08 [1.01-1.14] | | 1.01 [0.95-1.08] | 2,640 | 2,638 | 2,612 | NA | 2,322 |
| 603 | 1.20 [1.14-1.25] | 1.23 [1.16-1.30] | 1.24 [1.15-1.33] | 1.12 [1.06-1.19] | 1.11 [1.06-1.17] | 1.15 [1.10-1.20] | 1.12 [1.06-1.19] | 1.14 [1.06-1.23] | 1.10 [1.03-1.17] | 1.10 [1.04-1.15] | 5,311 | 2,922 | 1,852 | 2,965 | 4,266 |
| 603.1 | 1.12 [1.07-1.18] | 1.21 [1.14-1.28] | 1.19 [1.12-1.27] | 1.04 [0.97-1.12] | 1.10 [1.04-1.16] | 1.10 [1.05-1.15] | 1.16 [1.09-1.23] | 1.14 [1.07-1.22] | 1.02 [0.95-1.10] | 1.09 [1.03-1.14] | 4,538 | 2,690 | 2,253 | 2,075 | 3,697 |
| 604 | 1.38 [1.31-1.45] | 1.42 [1.34-1.51] | 1.40 [1.31-1.50] | 1.30 [1.20-1.41] | 1.27 [1.20-1.34] | 1.33 [1.26-1.40] | 1.32 [1.24-1.41] | 1.30 [1.21-1.40] | 1.27 [1.17-1.39] | 1.25 [1.18-1.33] | 4,085 | 2,747 | 2,122 | 1,637 | 3,445 |
| 604.1 | 1.46 [1.42-1.50] | 1.43 [1.38-1.48] | 1.38 [1.32-1.44] | 1.46 [1.41-1.50] | 1.37 [1.33-1.41] | 1.40 [1.37-1.44] | 1.32 [1.28-1.37] | 1.28 [1.23-1.34] | 1.41 [1.36-1.46] | 1.34 [1.30-1.38] | 16,264 | 7,471 | 4,922 | 10,132 | 12,324 |
| 605 | 1.44 [1.30-1.61] | 1.44 [1.29-1.60] | 1.44 [1.29-1.61] | | 1.29 [1.16-1.45] | 1.31 [1.17-1.46] | 1.24 [1.11-1.38] | 1.24 [1.10-1.39] | | 1.22 [1.08-1.37] | 853 | 849 | 789 | NA | 760 |
| 608 | 1.22 [1.18-1.27] | 1.25 [1.20-1.31] | 1.26 [1.20-1.33] | 1.12 [1.06-1.19] | 1.15 [1.10-1.20] | 1.17 [1.12-1.21] | 1.16 [1.11-1.21] | 1.17 [1.11-1.23] | 1.08 [1.02-1.15] | 1.12 [1.08-1.17] | 7,430 | 5,100 | 3,792 | 2,982 | 6,096 |
| 610.1 | 1.21 [1.10-1.34] | 1.17 [1.06-1.30] | 1.15 [1.03-1.29] | | 1.14 [1.02-1.27] | 1.16 [1.05-1.29] | 1.13 [1.02-1.25] | 1.09 [0.97-1.23] | | 1.13 [1.02-1.27] | 906 | 891 | 698 | NA | 762 |
| 610.2 | 1.06 [0.89-1.27] | 1.01 [0.84-1.20] | 0.91 [0.74-1.13] | | 0.99 [0.81-1.21] | 1.03 [0.86-1.24] | 0.97 [0.81-1.17] | 0.88 [0.71-1.09] | | 0.99 [0.81-1.22] | 285 | 273 | 193 | NA | 214 |
| 610.3 | 1.15 [0.96-1.36] | 1.10 [0.92-1.30] | 1.08 [0.90-1.31] | | 1.03 [0.85-1.25] | 1.12 [0.94-1.34] | 1.06 [0.89-1.27] | 1.07 [0.88-1.30] | | 1.04 [0.86-1.27] | 304 | 302 | 243 | NA | 241 |
| 610.4 | 1.05 [0.99-1.11] | 1.04 [0.98-1.11] | 1.02 [0.94-1.11] | 1.13 [1.00-1.28] | 0.99 [0.93-1.06] | 1.01 [0.95-1.07] | 0.99 [0.93-1.06] | 0.98 [0.90-1.07] | 1.12 [0.98-1.27] | 0.97 [0.90-1.04] | 2,632 | 2,432 | 1,299 | 675 | 1,975 |
| 610.8 | 1.17 [1.10-1.25] | 1.14 [1.07-1.22] | 1.11 [1.03-1.19] | 1.37 [1.01-1.85] | 1.10 [1.02-1.18] | 1.13 [1.06-1.21] | 1.09 [1.02-1.16] | 1.05 [0.97-1.13] | 1.34 [0.98-1.84] | 1.09 [1.01-1.17] | 2,183 | 2,148 | 1,645 | 118 | 1,789 |
| 611.1 | 1.09 [0.81-1.46] | 1.09 [0.81-1.46] | 1.07 [0.80-1.44] | | | 1.06 [0.78-1.43] | 0.97 [0.72-1.32] | 1.01 [0.75-1.37] | | | 109 | 110 | 108 | NA | NA |
| 611.3 | 1.16 [1.09-1.23] | 1.17 [1.09-1.24] | 1.16 [1.08-1.25] | 1.19 [0.93-1.53] | 1.10 [1.02-1.18] | 1.11 [1.04-1.18] | 1.09 [1.02-1.16] | 1.08 [1.00-1.16] | 1.16 [0.90-1.50] | 1.08 [1.00-1.16] | 2,320 | 2,264 | 1,839 | 163 | 1,932 |
| 612.2 | 1.28 [1.18-1.39] | 1.28 [1.17-1.39] | 1.27 [1.13-1.42] | 1.30 [1.09-1.55] | 1.16 [1.05-1.27] | 1.21 [1.11-1.32] | 1.15 [1.05-1.26] | 1.11 [0.99-1.25] | 1.26 [1.05-1.51] | 1.12 [1.01-1.23] | 1,388 | 1,242 | 739 | 339 | 1,060 |
| 612.3 | 1.13 [0.96-1.33] | 1.15 [0.96-1.38] | | 0.97 [0.76-1.25] | 1.04 [0.86-1.26] | 1.06 [0.90-1.26] | 1.05 [0.87-1.26] | | 0.93 [0.73-1.20] | 0.99 [0.82-1.21] | 352 | 278 | NA | 154 | 250 |
| 613 | 1.25 [1.18-1.34] | 1.25 [1.17-1.33] | 1.21 [1.12-1.30] | 1.37 [1.09-1.71] | 1.18 [1.11-1.27] | 1.19 [1.12-1.27] | 1.15 [1.07-1.23] | 1.11 [1.03-1.20] | 1.35 [1.07-1.70] | 1.15 [1.08-1.24] | 2,374 | 2,310 | 1,728 | 217 | 2,070 |
| 613.1 | 1.31 [1.22-1.40] | 1.33 [1.24-1.43] | 1.32 [1.20-1.45] | 1.25 [1.02-1.52] | 1.26 [1.17-1.36] | 1.25 [1.16-1.34] | 1.21 [1.12-1.30] | 1.20 [1.09-1.32] | 1.22 [0.99-1.50] | 1.24 [1.15-1.34] | 1,921 | 1,833 | 1,124 | 267 | 1,658 |
| 613.5 | 1.47 [1.25-1.73] | 1.41 [1.20-1.66] | 1.35 [1.10-1.66] | | 1.32 [1.12-1.55] | 1.36 [1.15-1.60] | 1.22 [1.03-1.44] | 1.19 [0.96-1.47] | | 1.24 [1.05-1.47] | 395 | 379 | 231 | NA | 365 |
| 613.7 | 1.23 [1.10-1.37] | 1.22 [1.09-1.37] | 1.22 [1.08-1.39] | | 1.13 [1.00-1.27] | 1.16 [1.04-1.31] | 1.13 [1.00-1.27] | 1.12 [0.99-1.28] | | 1.09 [0.96-1.23] | 738 | 719 | 578 | NA | 617 |
| 613.8 | 1.27 [1.13-1.42] | 1.29 [1.15-1.45] | 1.26 [1.11-1.44] | | 1.24 [1.09-1.40] | 1.23 [1.09-1.38] | 1.23 [1.09-1.38] | 1.19 [1.04-1.36] | | 1.21 [1.07-1.37] | 733 | 715 | 539 | NA | 639 |
| 613.9 | 1.23 [1.01-1.49] | 1.18 [0.97-1.43] | 1.16 [0.93-1.45] | | 1.17 [0.95-1.43] | 1.14 [0.93-1.39] | 1.07 [0.88-1.31] | 1.08 [0.86-1.35] | | 1.15 [0.93-1.42] | 262 | 249 | 198 | NA | 228 |
| 614 | 1.05 [0.78-1.41] | | | | | 0.95 [0.70-1.29] | | | | | 103 | NA | NA | NA | NA |
| 614.1 | 1.14 [1.10-1.18] | 1.16 [1.12-1.20] | 1.13 [1.07-1.19] | 1.12 [1.00-1.26] | 1.08 [1.04-1.12] | 1.08 [1.04-1.12] | 1.07 [1.03-1.11] | 1.04 [0.99-1.10] | 1.11 [0.98-1.25] | 1.05 [1.01-1.09] | 7,098 | 7,047 | 3,522 | 775 | 6,138 |
| 614.3 | 1.07 [0.99-1.15] | 1.08 [1.00-1.17] | 1.06 [0.95-1.18] | 1.08 [0.85-1.37] | 1.00 [0.92-1.09] | 1.03 [0.95-1.11] | 1.01 [0.93-1.09] | 1.00 [0.89-1.11] | 1.06 [0.83-1.35] | 0.98 [0.90-1.07] | 1,521 | 1,491 | 779 | 181 | 1,282 |
| 614.31 | 1.10 [0.86-1.40] | 1.18 [0.93-1.51] | | | 1.02 [0.78-1.32] | 1.05 [0.82-1.35] | 1.08 [0.84-1.40] | | | 1.00 [0.76-1.30] | 163 | 159 | NA | NA | 136 |
| 614.32 | 1.08 [0.98-1.18] | 1.10 [1.00-1.21] | 1.10 [0.97-1.25] | 1.23 [0.91-1.67] | 1.01 [0.91-1.12] | 1.04 [0.95-1.15] | 1.03 [0.93-1.13] | 1.04 [0.91-1.18] | 1.22 [0.89-1.67] | 0.99 [0.89-1.10] | 1,042 | 1,034 | 523 | 112 | 875 |
| 614.33 | 1.13 [1.05-1.21] | 1.16 [1.08-1.25] | 1.14 [1.01-1.29] | 1.08 [0.95-1.23] | 1.03 [0.95-1.11] | 1.04 [0.96-1.12] | 1.00 [0.92-1.08] | 1.05 [0.92-1.19] | 1.03 [0.90-1.18] | 0.97 [0.90-1.05] | 1,898 | 1,761 | 568 | 611 | 1,609 |
| 614.4 | 1.17 [1.05-1.31] | 1.23 [1.10-1.37] | 1.25 [1.09-1.42] | | 1.14 [1.02-1.28] | 1.08 [0.97-1.21] | 1.12 [1.01-1.26] | 1.13 [0.99-1.30] | | 1.08 [0.96-1.22] | 792 | 791 | 507 | NA | 699 |
| 614.5 | 1.15 [1.08-1.22] | 1.15 [1.08-1.22] | 1.21 [1.11-1.31] | 1.00 [0.86-1.16] | 1.11 [1.04-1.18] | 1.11 [1.04-1.18] | 1.07 [1.01-1.14] | 1.13 [1.04-1.23] | 1.00 [0.85-1.16] | 1.08 [1.01-1.15] | 2,641 | 2,545 | 1,310 | 441 | 2,182 |
| 614.51 | 1.09 [1.01-1.18] | 1.12 [1.04-1.21] | 1.12 [1.01-1.24] | 0.95 [0.72-1.26] | 1.04 [0.96-1.14] | 1.04 [0.96-1.13] | 1.06 [0.98-1.15] | 1.07 [0.96-1.19] | 0.96 [0.72-1.28] | 1.02 [0.93-1.12] | 1,444 | 1,441 | 782 | 126 | 1,136 |
| 614.52 | 1.36 [1.25-1.48] | 1.44 [1.31-1.59] | 1.57 [1.40-1.78] | 1.26 [1.09-1.47] | 1.32 [1.21-1.45] | 1.28 [1.17-1.39] | 1.29 [1.17-1.42] | 1.42 [1.25-1.61] | 1.21 [1.03-1.41] | 1.26 [1.15-1.39] | 1,382 | 1,081 | 680 | 452 | 1,188 |
| 614.53 | 1.06 [0.97-1.15] | 1.04 [0.96-1.13] | 1.06 [0.93-1.20] | 0.92 [0.77-1.11] | 1.01 [0.92-1.10] | 1.04 [0.95-1.12] | 0.98 [0.91-1.07] | 1.02 [0.89-1.16] | 0.92 [0.76-1.11] | 1.00 [0.91-1.10] | 1,427 | 1,374 | 515 | 297 | 1,115 |
| 614.54 | 1.40 [1.27-1.54] | 1.43 [1.30-1.59] | 1.49 [1.30-1.70] | 1.12 [0.91-1.37] | 1.32 [1.19-1.46] | 1.33 [1.20-1.46] | 1.28 [1.15-1.42] | 1.32 [1.15-1.52] | 1.07 [0.87-1.32] | 1.28 [1.15-1.42] | 1,088 | 976 | 536 | 262 | 910 |
| 615 | 1.13 [1.09-1.17] | 1.15 [1.11-1.18] | 1.09 [1.03-1.14] | 1.16 [1.06-1.27] | 1.08 [1.04-1.12] | 1.06 [1.02-1.10] | 1.04 [1.00-1.07] | 1.00 [0.95-1.05] | 1.12 [1.02-1.22] | 1.03 [0.99-1.07] | 8,651 | 8,539 | 3,717 | 1,438 | 7,115 |
| 617 | 1.34 [1.21-1.47] | 1.35 [1.22-1.48] | 1.31 [1.18-1.46] | | 1.25 [1.13-1.38] | 1.26 [1.14-1.39] | 1.24 [1.12-1.37] | 1.19 [1.07-1.33] | | 1.21 [1.10-1.34] | 1,059 | 1,046 | 826 | NA | 966 |
| 618 | 1.24 [1.10-1.40] | 1.24 [1.11-1.40] | 1.18 [1.04-1.34] | | 1.12 [0.99-1.26] | 1.16 [1.03-1.31] | 1.15 [1.02-1.30] | 1.08 [0.95-1.23] | | 1.07 [0.94-1.21] | 678 | 678 | 569 | NA | 603 |
| 618.1 | 1.21 [1.18-1.24] | 1.20 [1.17-1.24] | 1.20 [1.17-1.23] | 1.28 [0.97-1.69] | 1.15 [1.12-1.18] | 1.15 [1.12-1.18] | 1.13 [1.10-1.16] | 1.12 [1.09-1.15] | 1.28 [0.96-1.69] | 1.10 [1.07-1.13] | 14,347 | 14,319 | 13,052 | 141 | 12,136 |
| 618.2 | 1.14 [1.10-1.18] | 1.13 [1.09-1.17] | 1.13 [1.09-1.17] | | 1.08 [1.04-1.13] | 1.11 [1.06-1.15] | 1.09 [1.05-1.13] | 1.09 [1.05-1.13] | | 1.06 [1.02-1.11] | 6,596 | 6,598 | 6,086 | NA | 5,466 |
| 618.5 | 1.24 [1.08-1.42] | 1.24 [1.08-1.43] | 1.23 [1.07-1.42] | | 1.08 [0.94-1.25] | 1.18 [1.03-1.36] | 1.16 [1.01-1.34] | 1.15 [1.00-1.32] | | 1.04 [0.90-1.20] | 480 | 479 | 470 | NA | 425 |
| 618.6 | 1.20 [1.08-1.33] | 1.22 [1.10-1.36] | 1.21 [1.08-1.34] | | 1.14 [1.02-1.27] | 1.12 [1.00-1.25] | 1.12 [1.00-1.25] | 1.09 [0.98-1.22] | | 1.07 [0.96-1.20] | 818 | 816 | 777 | NA | 750 |
| 619 | 1.37 [1.12-1.69] | 1.45 [1.17-1.79] | 1.52 [1.14-2.04] | | 1.17 [0.93-1.48] | 1.27 [1.02-1.56] | 1.28 [1.03-1.60] | 1.37 [1.01-1.86] | | 1.08 [0.85-1.37] | 225 | 209 | 111 | NA | 166 |
| 619.1 | 1.13 [1.10-1.16] | 1.15 [1.12-1.18] | 1.12 [1.08-1.16] | 1.11 [1.05-1.17] | 1.08 [1.05-1.11] | 1.07 [1.04-1.10] | 1.04 [1.01-1.07] | 1.04 [1.00-1.07] | 1.06 [1.00-1.13] | 1.04 [1.01-1.07] | 14,639 | 13,619 | 6,998 | 3,120 | 12,225 |
| 619.2 | 1.16 [1.12-1.21] | 1.18 [1.14-1.23] | 1.19 [1.14-1.25] | 1.10 [0.96-1.27] | 1.12 [1.08-1.17] | 1.11 [1.07-1.15] | 1.10 [1.06-1.14] | 1.12 [1.07-1.17] | 1.08 [0.93-1.25] | 1.08 [1.04-1.13] | 6,636 | 6,594 | 4,757 | 535 | 5,644 |
| 619.3 | 1.18 [1.14-1.23] | 1.19 [1.14-1.24] | 1.19 [1.12-1.26] | 1.09 [0.97-1.21] | 1.11 [1.07-1.16] | 1.13 [1.09-1.18] | 1.12 [1.07-1.16] | 1.13 [1.06-1.19] | 1.05 [0.94-1.18] | 1.08 [1.03-1.13] | 6,068 | 6,016 | 2,852 | 856 | 4,976 |
| 619.4 | 1.28 [1.22-1.33] | 1.29 [1.23-1.35] | 1.30 [1.22-1.39] | 1.08 [0.98-1.18] | 1.16 [1.11-1.21] | 1.20 [1.15-1.26] | 1.17 [1.12-1.22] | 1.21 [1.13-1.29] | 1.04 [0.95-1.15] | 1.11 [1.06-1.17] | 5,278 | 4,969 | 2,047 | 1,166 | 4,498 |
| 619.5 | 1.21 [1.16-1.27] | 1.20 [1.15-1.26] | 1.38 [1.29-1.49] | 1.09 [0.97-1.22] | 1.14 [1.08-1.20] | 1.17 [1.12-1.23] | 1.14 [1.09-1.20] | 1.27 [1.18-1.37] | 1.08 [0.95-1.21] | 1.10 [1.05-1.16] | 4,461 | 4,302 | 1,748 | 737 | 3,762 |
| 620 | 1.48 [1.22-1.81] | 1.48 [1.21-1.80] | 1.44 [1.15-1.80] | | 1.41 [1.14-1.73] | 1.41 [1.15-1.73] | 1.33 [1.09-1.63] | 1.29 [1.03-1.62] | | 1.33 [1.07-1.64] | 254 | 255 | 195 | NA | 220 |
| 621 | 1.17 [1.09-1.27] | 1.17 [1.08-1.26] | 1.14 [1.05-1.23] | | 1.07 [0.98-1.16] | 1.15 [1.06-1.24] | 1.13 [1.04-1.22] | 1.10 [1.02-1.20] | | 1.06 [0.98-1.16] | 1,550 | 1,551 | 1,362 | NA | 1,255 |
| 622 | 1.21 [1.02-1.44] | 1.28 [1.07-1.52] | 1.22 [1.00-1.50] | | 1.27 [1.05-1.53] | 1.15 [0.96-1.37] | 1.20 [1.01-1.44] | 1.15 [0.94-1.42] | | 1.22 [1.01-1.47] | 316 | 304 | 218 | NA | 268 |
| 622.1 | 1.12 [1.09-1.16] | 1.13 [1.09-1.16] | 1.12 [1.09-1.16] | 1.17 [0.93-1.48] | 1.08 [1.05-1.12] | 1.10 [1.07-1.13] | 1.09 [1.06-1.12] | 1.09 [1.06-1.13] | 1.09 [0.86-1.39] | 1.07 [1.03-1.10] | 11,263 | 11,250 | 9,786 | 199 | 8,858 |
| 622.2 | 1.10 [1.03-1.16] | 1.10 [1.04-1.17] | 1.09 [1.02-1.16] | | 1.06 [0.99-1.13] | 1.08 [1.01-1.15] | 1.10 [1.03-1.17] | 1.08 [1.01-1.15] | | 1.05 [0.98-1.12] | 2,477 | 2,478 | 2,127 | NA | 1,887 |
| 623 | 1.13 [1.06-1.20] | 1.14 [1.07-1.21] | 1.14 [1.06-1.22] | 1.05 [0.83-1.33] | 1.09 [1.02-1.17] | 1.09 [1.02-1.16] | 1.06 [1.00-1.13] | 1.07 [0.99-1.15] | 1.01 [0.79-1.29] | 1.06 [0.99-1.13] | 2,484 | 2,432 | 1,760 | 179 | 2,138 |

### Supplementary Table 6: phecode-mapped results (hazard ratios and events from all cohorts)

| | Hazard ratio (99% confidence interval) | | | | | | | | | | Events (in exposed) | | | | |
|---|---|---|---|---|---|---|---|---|---|---|---|---|---|---|---|
| | crude | | | | | adjusted | | | | | | | | | |
| Outcome | any age | 18+ | 40+ | <18 | hosp. | any age | 18+ | 40+ | <18 | hosp. | any age | 18+ | 40+ | <18 | hosp. |
| 624.1 | 1.64 [1.48-1.83] | 1.63 [1.46-1.81] | 1.64 [1.47-1.83] | | 1.50 [1.34-1.68] | 1.53 [1.37-1.70] | 1.48 [1.32-1.65] | 1.50 [1.34-1.69] | | 1.43 [1.28-1.60] | 927 | 915 | 837 | NA | 823 |
| 624.2 | 1.30 [1.18-1.43] | 1.28 [1.16-1.41] | 1.28 [1.16-1.41] | | 1.22 [1.10-1.35] | 1.25 [1.13-1.38] | 1.22 [1.10-1.34] | 1.20 [1.09-1.33] | | 1.19 [1.07-1.32] | 999 | 996 | 979 | NA | 857 |
| 624.9 | 1.38 [1.33-1.43] | 1.38 [1.33-1.43] | 1.37 [1.32-1.42] | 1.48 [1.12-1.94] | 1.27 [1.23-1.33] | 1.26 [1.21-1.31] | 1.22 [1.18-1.27] | 1.21 [1.16-1.26] | 1.38 [1.04-1.84] | 1.20 [1.16-1.25] | 6,882 | 6,834 | 5,957 | 148 | 6,068 |
| 625 | 1.23 [1.18-1.30] | 1.27 [1.21-1.34] | 1.22 [1.14-1.32] | 1.14 [1.02-1.28] | 1.13 [1.07-1.19] | 1.16 [1.10-1.22] | 1.14 [1.08-1.20] | 1.11 [1.03-1.20] | 1.12 [0.99-1.26] | 1.08 [1.03-1.14] | 4,109 | 3,978 | 1,652 | 764 | 3,414 |
| 625.1 | 1.33 [1.25-1.41] | 1.36 [1.28-1.44] | 1.37 [1.23-1.52] | 1.32 [1.16-1.49] | 1.24 [1.16-1.32] | 1.21 [1.14-1.29] | 1.18 [1.11-1.26] | 1.22 [1.09-1.36] | 1.25 [1.10-1.42] | 1.16 [1.09-1.24] | 2,946 | 2,923 | 828 | 678 | 2,459 |
| 626 | 1.21 [1.16-1.26] | 1.21 [1.16-1.26] | 1.19 [1.12-1.26] | 1.18 [1.07-1.29] | 1.11 [1.07-1.16] | 1.14 [1.10-1.19] | 1.09 [1.04-1.14] | 1.10 [1.03-1.17] | 1.14 [1.04-1.25] | 1.09 [1.04-1.14] | 5,712 | 5,363 | 2,693 | 1,214 | 4,937 |
| 626.1 | 1.17 [1.10-1.26] | 1.19 [1.11-1.27] | 1.14 [1.04-1.25] | 1.19 [0.99-1.42] | 1.09 [1.02-1.18] | 1.11 [1.03-1.19] | 1.08 [1.00-1.16] | 1.05 [0.96-1.15] | 1.19 [0.99-1.43] | 1.05 [0.98-1.14] | 1,996 | 1,872 | 1,047 | 334 | 1,662 |
| 626.11 | 1.29 [1.13-1.47] | 1.30 [1.13-1.49] | 1.52 [1.19-1.93] | 1.14 [0.92-1.41] | 1.15 [1.00-1.32] | 1.22 [1.06-1.39] | 1.14 [0.99-1.32] | 1.41 [1.10-1.81] | 1.11 [0.89-1.38] | 1.10 [0.95-1.27] | 609 | 491 | 160 | 239 | 485 |
| 626.12 | 1.20 [1.17-1.23] | 1.20 [1.17-1.23] | 1.17 [1.14-1.21] | 1.23 [1.13-1.35] | 1.14 [1.11-1.18] | 1.13 [1.10-1.16] | 1.10 [1.07-1.13] | 1.08 [1.05-1.11] | 1.19 [1.09-1.31] | 1.10 [1.07-1.13] | 14,375 | 13,834 | 9,103 | 1,385 | 12,061 |
| 626.13 | 1.21 [1.14-1.30] | 1.23 [1.15-1.31] | 1.23 [1.12-1.34] | 1.18 [1.00-1.39] | 1.15 [1.07-1.24] | 1.12 [1.05-1.20] | 1.10 [1.02-1.18] | 1.12 [1.03-1.23] | 1.13 [0.95-1.34] | 1.10 [1.03-1.19] | 2,162 | 2,046 | 1,125 | 396 | 1,793 |
| 626.14 | 1.22 [1.17-1.27] | 1.23 [1.18-1.28] | 1.19 [1.13-1.25] | 1.15 [0.99-1.34] | 1.16 [1.11-1.22] | 1.14 [1.09-1.19] | 1.12 [1.07-1.17] | 1.10 [1.05-1.17] | 1.08 [0.93-1.27] | 1.11 [1.06-1.16] | 5,245 | 5,150 | 3,165 | 464 | 4,438 |
| 626.2 | 1.22 [1.16-1.29] | 1.23 [1.17-1.30] | 1.18 [1.08-1.28] | 1.21 [1.07-1.37] | 1.16 [1.09-1.23] | 1.12 [1.06-1.19] | 1.08 [1.02-1.15] | 1.05 [0.96-1.15] | 1.13 [1.00-1.28] | 1.11 [1.04-1.18] | 3,238 | 2,983 | 1,225 | 753 | 2,703 |
| 626.21 | 1.12 [0.92-1.37] | 1.04 [0.82-1.32] | | 1.12 [0.87-1.44] | 1.01 [0.81-1.25] | 1.04 [0.85-1.27] | 0.94 [0.73-1.20] | | 1.09 [0.84-1.41] | 0.97 [0.77-1.22] | 267 | 164 | NA | 170 | 203 |
| 626.4 | 1.18 [0.94-1.48] | 1.21 [0.96-1.52] | 1.34 [1.00-1.78] | | 1.11 [0.88-1.40] | 1.02 [0.81-1.29] | 0.99 [0.78-1.26] | 1.16 [0.86-1.57] | | 1.05 [0.82-1.33] | 184 | 176 | 107 | NA | 169 |
| 626.8 | 0.93 [0.86-1.01] | 0.95 [0.87-1.03] | 0.97 [0.75-1.26] | | 0.97 [0.88-1.08] | 0.92 [0.85-1.00] | 0.93 [0.86-1.01] | 0.98 [0.76-1.27] | | 0.96 [0.86-1.07] | 1,306 | 1,306 | 125 | NA | 800 |
| 627 | 1.19 [1.02-1.40] | 1.22 [1.04-1.43] | 1.22 [1.04-1.43] | | 1.11 [0.93-1.32] | 1.12 [0.95-1.32] | 1.14 [0.97-1.34] | 1.13 [0.96-1.33] | | 1.05 [0.88-1.26] | 360 | 361 | 353 | NA | 283 |
| 627.1 | 1.24 [1.20-1.28] | 1.23 [1.19-1.27] | 1.23 [1.19-1.27] | | 1.18 [1.14-1.23] | 1.19 [1.15-1.23] | 1.17 [1.13-1.21] | 1.16 [1.12-1.20] | | 1.15 [1.11-1.20] | 8,737 | 8,737 | 8,697 | NA | 6,993 |
| 627.2 | 1.39 [1.10-1.76] | 1.49 [1.17-1.89] | 1.49 [1.17-1.90] | | 1.38 [1.07-1.78] | 1.26 [0.99-1.60] | 1.30 [1.02-1.66] | 1.33 [1.04-1.71] | | 1.30 [1.01-1.68] | 175 | 174 | 163 | NA | 148 |
| 627.3 | 1.30 [1.21-1.39] | 1.32 [1.23-1.42] | 1.30 [1.22-1.40] | | 1.22 [1.14-1.32] | 1.23 [1.14-1.32] | 1.23 [1.15-1.32] | 1.22 [1.13-1.30] | | 1.18 [1.10-1.28] | 2,050 | 2,051 | 2,045 | NA | 1,791 |
| 627.4 | 1.46 [1.22-1.74] | 1.45 [1.21-1.74] | 1.40 [1.16-1.68] | | 1.49 [1.22-1.81] | 1.39 [1.16-1.67] | 1.36 [1.13-1.64] | 1.31 [1.08-1.58] | | 1.41 [1.16-1.73] | 292 | 290 | 270 | NA | 249 |
| 627.5 | 1.39 [1.09-1.76] | 1.61 [1.26-2.06] | | | 1.48 [1.15-1.89] | 1.25 [0.98-1.60] | 1.33 [1.03-1.72] | | | 1.38 [1.07-1.78] | 187 | 175 | NA | NA | 163 |
| 628 | 1.14 [1.11-1.17] | 1.15 [1.12-1.19] | 1.13 [1.08-1.17] | 1.10 [1.03-1.16] | 1.08 [1.05-1.12] | 1.07 [1.04-1.10] | 1.04 [1.01-1.07] | 1.04 [1.00-1.08] | 1.05 [0.98-1.12] | 1.04 [1.01-1.07] | 13,108 | 12,192 | 6,131 | 2,839 | 10,981 |
| pregnancy complications | | | | | | | | | | | | | | | |
| 634 | 1.07 [1.06-1.08] | 1.12 [1.10-1.13] | 1.06 [0.99-1.14] | 0.98 [0.96-1.00] | 0.98 [0.96-1.00] | 1.06 [1.05-1.07] | 1.08 [1.07-1.09] | 1.10 [1.02-1.18] | 0.98 [0.96-1.00] | 0.98 [0.96-1.00] | 80,817 | 77,228 | 2,350 | 30,496 | 38,506 |
| 634.1 | 1.10 [1.05-1.14] | 1.11 [1.07-1.16] | 1.03 [0.92-1.15] | 1.07 [0.98-1.18] | 1.05 [1.00-1.09] | 1.07 [1.02-1.11] | 1.07 [1.03-1.12] | 1.01 [0.89-1.14] | 1.07 [0.97-1.17] | 1.03 [0.99-1.08] | 5,887 | 5,819 | 624 | 1,182 | 4,714 |
| 634.3 | 1.04 [0.98-1.11] | 1.09 [1.03-1.16] | 1.02 [0.81-1.29] | 1.02 [0.90-1.16] | 0.99 [0.92-1.05] | 1.02 [0.95-1.08] | 1.03 [0.97-1.10] | 1.01 [0.79-1.29] | 0.99 [0.87-1.13] | 0.98 [0.92-1.05] | 2,641 | 2,620 | 150 | 632 | 2,156 |
| 635 | 1.01 [0.76-1.33] | 1.08 [0.82-1.43] | | | 0.94 [0.71-1.26] | 0.99 [0.75-1.32] | 1.09 [0.82-1.45] | | | 0.97 [0.72-1.30] | 120 | 120 | NA | NA | 106 |
| 635.2 | 1.09 [1.06-1.13] | 1.13 [1.09-1.16] | 1.02 [0.90-1.17] | 1.03 [0.97-1.09] | 1.05 [1.01-1.08] | 1.07 [1.04-1.10] | 1.07 [1.04-1.11] | 1.01 [0.88-1.16] | 1.01 [0.96-1.08] | 1.03 [1.00-1.07] | 11,241 | 11,068 | 479 | 3,014 | 8,809 |
| 635.3 | 1.05 [1.03-1.08] | 1.08 [1.06-1.10] | 0.97 [0.89-1.06] | 0.98 [0.94-1.02] | 1.02 [1.00-1.04] | 1.05 [1.02-1.07] | 1.07 [1.05-1.09] | 1.00 [0.92-1.10] | 0.97 [0.93-1.02] | 1.02 [0.99-1.04] | 23,598 | 23,373 | 1,079 | 6,369 | 17,714 |
| 636 | 1.16 [1.12-1.21] | 1.17 [1.13-1.22] | 0.95 [0.75-1.20] | 1.05 [0.97-1.14] | 1.07 [1.02-1.12] | 1.12 [1.07-1.16] | 1.10 [1.06-1.14] | 0.91 [0.71-1.16] | 1.04 [0.95-1.13] | 1.05 [1.00-1.10] | 6,347 | 6,178 | 146 | 1,471 | 4,703 |
| 636.2 | 1.08 [1.04-1.11] | 1.11 [1.07-1.14] | 1.06 [0.93-1.21] | 1.02 [0.96-1.08] | 1.03 [0.99-1.06] | 1.05 [1.01-1.08] | 1.05 [1.02-1.09] | 1.03 [0.90-1.18] | 1.02 [0.96-1.09] | 1.02 [0.98-1.06] | 9,664 | 9,541 | 479 | 2,675 | 7,666 |
| 636.3 | 1.06 [1.03-1.10] | 1.10 [1.06-1.14] | 0.97 [0.83-1.13] | 1.00 [0.94-1.07] | 1.00 [0.96-1.04] | 1.03 [0.99-1.07] | 1.03 [1.00-1.07] | 0.98 [0.84-1.15] | 1.00 [0.93-1.06] | 0.99 [0.95-1.03] | 8,064 | 7,869 | 356 | 2,475 | 6,494 |
| 636.8 | 1.06 [0.92-1.22] | 1.06 [0.92-1.22] | | | 0.99 [0.86-1.15] | 1.05 [0.91-1.21] | 1.04 [0.90-1.20] | | | 1.00 [0.87-1.17] | 483 | 482 | NA | NA | 431 |
| 638 | 1.10 [1.04-1.16] | 1.14 [1.07-1.20] | 0.94 [0.83-1.06] | 1.03 [0.90-1.18] | 1.03 [0.97-1.09] | 1.07 [1.00-1.13] | 1.09 [1.03-1.16] | 0.96 [0.85-1.09] | 0.99 [0.86-1.13] | 1.02 [0.96-1.09] | 2,961 | 2,932 | 563 | 561 | 2,662 |
| 639 | 1.05 [0.95-1.16] | 1.09 [0.98-1.20] | | 0.99 [0.83-1.18] | 0.97 [0.87-1.09] | 1.01 [0.91-1.12] | 1.01 [0.91-1.12] | | 0.99 [0.83-1.18] | 0.97 [0.87-1.09] | 942 | 916 | NA | 336 | 792 |
| 642 | 1.09 [1.06-1.12] | 1.12 [1.08-1.15] | 1.05 [0.93-1.19] | 1.05 [0.98-1.12] | 1.06 [1.02-1.10] | 1.06 [1.03-1.10] | 1.08 [1.05-1.12] | 1.03 [0.91-1.17] | 1.03 [0.96-1.10] | 1.04 [1.00-1.08] | 9,573 | 9,424 | 560 | 2,369 | 6,712 |
| 642.1 | 1.10 [1.05-1.16] | 1.13 [1.07-1.18] | 0.98 [0.80-1.20] | 1.11 [1.02-1.22] | 1.06 [1.00-1.12] | 1.07 [1.02-1.13] | 1.09 [1.03-1.14] | 0.96 [0.78-1.18] | 1.11 [1.01-1.22] | 1.03 [0.97-1.09] | 4,171 | 4,093 | 207 | 1,292 | 2,917 |
| 643 | 1.11 [1.06-1.15] | 1.16 [1.11-1.21] | | 1.03 [0.97-1.10] | 1.02 [0.97-1.06] | 1.04 [1.00-1.09] | 1.05 [1.00-1.09] | | 1.01 [0.94-1.08] | 0.99 [0.94-1.03] | 5,975 | 5,800 | NA | 2,422 | 4,869 |
| 643.1 | 1.08 [1.03-1.14] | 1.14 [1.09-1.20] | | 1.01 [0.94-1.09] | 1.01 [0.96-1.07] | 1.02 [0.97-1.08] | 1.04 [0.98-1.09] | | 0.99 [0.91-1.07] | 0.98 [0.93-1.04] | 4,069 | 3,948 | NA | 1,725 | 3,330 |
| 644 | 1.07 [1.03-1.11] | 1.10 [1.06-1.13] | 1.00 [0.86-1.15] | 1.01 [0.95-1.08] | 1.00 [0.97-1.04] | 1.06 [1.02-1.09] | 1.08 [1.04-1.12] | 0.98 [0.85-1.14] | 1.01 [0.94-1.08] | 1.01 [0.97-1.04] | 8,673 | 8,547 | 393 | 2,495 | 6,718 |
| 645 | 1.03 [1.00-1.06] | 1.05 [1.02-1.07] | 0.87 [0.75-1.01] | 0.99 [0.93-1.04] | 0.98 [0.95-1.01] | 1.02 [1.00-1.05] | 1.04 [1.01-1.07] | 0.89 [0.77-1.04] | 0.98 [0.92-1.03] | 0.98 [0.94-1.01] | 12,342 | 12,175 | 376 | 3,275 | 8,546 |
| 646 | 1.18 [1.16-1.20] | 1.21 [1.18-1.23] | 1.01 [0.93-1.11] | 1.07 [1.03-1.10] | 1.09 [1.07-1.12] | 1.12 [1.10-1.14] | 1.10 [1.08-1.13] | 0.98 [0.89-1.08] | 1.04 [1.01-1.08] | 1.06 [1.04-1.08] | 31,167 | 30,354 | 1,005 | 9,895 | 23,736 |
| 647 | 1.11 [1.04-1.18] | 1.16 [1.09-1.23] | | 0.99 [0.88-1.10] | 1.07 [1.00-1.14] | 1.08 [1.01-1.15] | 1.11 [1.04-1.18] | | 0.97 [0.86-1.08] | 1.04 [0.97-1.12] | 2,723 | 2,693 | NA | 851 | 2,184 |
| 647.1 | 1.16 [1.12-1.20] | 1.19 [1.15-1.24] | 1.02 [0.82-1.27] | 1.04 [0.97-1.10] | 1.08 [1.04-1.13] | 1.11 [1.07-1.15] | 1.09 [1.05-1.14] | 0.97 [0.78-1.21] | 1.03 [0.96-1.09] | 1.06 [1.02-1.11] | 7,294 | 7,072 | 177 | 2,648 | 5,969 |
| 647.3 | 1.09 [0.99-1.20] | 1.10 [1.00-1.21] | | 0.95 [0.80-1.13] | 1.04 [0.94-1.15] | 1.05 [0.95-1.15] | 1.03 [0.93-1.14] | | 0.92 [0.77-1.10] | 1.01 [0.91-1.12] | 1,088 | 1,078 | NA | 356 | 897 |
| 649 | 1.21 [1.17-1.25] | 1.26 [1.22-1.30] | 1.11 [0.97-1.27] | 1.09 [1.02-1.15] | 1.13 [1.09-1.17] | 1.15 [1.11-1.19] | 1.16 [1.12-1.20] | 1.06 [0.92-1.21] | 1.06 [0.99-1.13] | 1.10 [1.06-1.14] | 9,473 | 9,451 | 495 | 2,958 | 8,033 |
| 649.1 | 1.07 [1.03-1.11] | 1.09 [1.06-1.13] | 1.04 [0.93-1.16] | 1.00 [0.92-1.09] | 1.02 [0.98-1.06] | 1.05 [1.01-1.09] | 1.07 [1.03-1.11] | 1.03 [0.92-1.16] | 1.02 [0.94-1.11] | 1.01 [0.97-1.05] | 7,581 | 7,552 | 683 | 1,507 | 6,382 |
| 650 | 1.08 [1.05-1.11] | 1.09 [1.06-1.12] | 1.05 [0.89-1.24] | 0.94 [0.89-1.00] | 0.97 [0.94-1.00] | 1.09 [1.06-1.12] | 1.09 [1.06-1.12] | 1.11 [0.94-1.32] | 0.96 [0.91-1.02] | 1.00 [0.97-1.03] | 12,482 | 12,266 | 312 | 2,989 | 10,079 |
| 651 | 1.07 [1.06-1.09] | 1.12 [1.10-1.13] | 1.06 [0.99-1.13] | 0.98 [0.96-1.00] | 1.00 [0.98-1.01] | 1.06 [1.05-1.08] | 1.08 [1.07-1.10] | 1.12 [1.04-1.20] | 0.98 [0.96-1.00] | 1.00 [0.99-1.02] | 78,231 | 76,189 | 2,196 | 27,276 | 41,408 |
| 652 | 1.03 [1.01-1.06] | 1.06 [1.03-1.08] | 0.95 [0.86-1.06] | 0.98 [0.93-1.03] | 1.00 [0.97-1.02] | 1.02 [1.00-1.05] | 1.04 [1.01-1.07] | 0.96 [0.86-1.07] | 0.97 [0.92-1.02] | 0.99 [0.96-1.02] | 16,378 | 16,241 | 755 | 3,926 | 12,111 |
| 653 | 1.05 [1.03-1.08] | 1.08 [1.06-1.10] | 0.89 [0.81-0.99] | 1.00 [0.96-1.04] | 1.02 [1.00-1.05] | 1.05 [1.02-1.07] | 1.06 [1.03-1.08] | 0.94 [0.84-1.04] | 1.00 [0.96-1.04] | 1.03 [1.00-1.05] | 23,548 | 23,281 | 790 | 6,589 | 17,376 |
| 654 | 1.04 [0.97-1.12] | 1.07 [1.00-1.15] | | 0.97 [0.85-1.11] | 1.01 [0.93-1.10] | 1.03 [0.95-1.11] | 1.06 [0.98-1.14] | | 0.97 [0.85-1.11] | 1.00 [0.92-1.09] | 1,807 | 1,769 | NA | 573 | 1,361 |
| 654.1 | 1.10 [1.08-1.13] | 1.12 [1.09-1.15] | 1.03 [0.94-1.12] | 0.98 [0.92-1.04] | 1.02 [0.99-1.04] | 1.08 [1.06-1.11] | 1.10 [1.07-1.13] | 1.05 [0.96-1.15] | 0.97 [0.91-1.04] | 1.02 [0.99-1.04] | 15,511 | 15,518 | 1,145 | 2,277 | 14,592 |
| 654.2 | 1.09 [1.01-1.17] | 1.11 [1.02-1.19] | | 1.09 [0.93-1.27] | 1.01 [0.92-1.10] | 1.07 [0.99-1.16] | 1.08 [1.00-1.17] | | 1.07 [0.91-1.26] | 1.01 [0.93-1.10] | 1,621 | 1,598 | NA | 416 | 1,254 |
| 655 | 1.08 [1.06-1.09] | 1.11 [1.10-1.13] | 0.97 [0.90-1.04] | 1.01 [0.98-1.04] | 1.02 [1.01-1.04] | 1.06 [1.04-1.07] | 1.07 [1.06-1.09] | 0.98 [0.91-1.05] | 1.00 [0.98-1.03] | 1.02 [1.00-1.03] | 52,442 | 51,477 | 1,728 | 17,094 | 35,811 |
| 656 | 1.18 [1.14-1.22] | 1.22 [1.18-1.25] | 1.14 [0.99-1.32] | 1.08 [1.02-1.14] | 1.09 [1.05-1.13] | 1.11 [1.08-1.15] | 1.09 [1.05-1.12] | 1.06 [0.91-1.23] | 1.07 [1.01-1.13] | 1.05 [1.01-1.09] | 9,997 | 9,657 | 395 | 3,308 | 8,356 |
| 656.2 | 1.08 [0.96-1.21] | 1.11 [0.98-1.25] | 1.10 [0.97-1.25] | | 1.03 [0.92-1.16] | 1.04 [0.93-1.17] | 1.06 [0.94-1.20] | 1.04 [0.92-1.19] | | 1.04 [0.92-1.17] | 782 | 726 | 692 | NA | 740 |
| 656.7 | 1.02 [0.87-1.20] | | | 1.04 [0.89-1.22] | 1.00 [0.84-1.20] | 1.02 [0.87-1.20] | | | 1.02 [0.87-1.20] | 1.01 [0.84-1.21] | 366 | NA | NA | 365 | 298 |
| 661 | 1.03 [1.01-1.05] | 1.05 [1.03-1.07] | 0.89 [0.80-0.99] | 0.99 [0.96-1.04] | 1.02 [0.99-1.04] | 1.02 [1.00-1.04] | 1.04 [1.02-1.06] | 0.93 [0.83-1.05] | 0.98 [0.94-1.03] | 1.01 [0.98-1.04] | 21,601 | 21,334 | 668 | 6,145 | 14,205 |
| 663 | 1.09 [1.03-1.15] | 1.10 [1.04-1.16] | 1.18 [0.93-1.49] | 0.99 [0.88-1.11] | 1.03 [0.97-1.10] | 1.08 [1.02-1.14] | 1.10 [1.04-1.16] | 1.22 [0.95-1.56] | 0.99 [0.88-1.12] | 1.04 [0.98-1.11] | 3,144 | 3,121 | 152 | 714 | 2,453 |
| 665 | 1.04 [1.02-1.05] | 1.06 [1.05-1.08] | 0.94 [0.86-1.02] | 0.97 [0.95-1.00] | 0.98 [0.96-1.00] | 1.04 [1.02-1.05] | 1.06 [1.04-1.07] | 1.01 [0.93-1.11] | 0.98 [0.95-1.00] | 0.99 [0.97-1.01] | 49,808 | 48,741 | 1,227 | 15,768 | 34,052 |
| 668 | 1.20 [1.06-1.37] | 1.21 [1.06-1.37] | | 0.91 [0.71-1.17] | 1.14 [0.99-1.31] | 1.16 [1.02-1.32] | 1.16 [1.02-1.32] | | 0.89 [0.69-1.15] | 1.12 [0.97-1.28] | 626 | 615 | NA | 157 | 500 |
| 669 | 1.05 [1.02-1.07] | 1.07 [1.05-1.09] | 0.96 [0.87-1.06] | 1.01 [0.97-1.05] | 1.01 [0.99-1.04] | 1.04 [1.02-1.06] | 1.05 [1.03-1.08] | 1.00 [0.90-1.11] | 1.01 [0.97-1.05] | 1.01 [0.99-1.04] | 24,245 | 23,891 | 905 | 6,935 | 16,759 |
| 671 | 1.12 [1.05-1.20] | 1.15 [1.08-1.23] | 1.10 [0.87-1.39] | 0.97 [0.83-1.13] | 1.04 [0.97-1.12] | 1.09 [1.02-1.17] | 1.10 [1.03-1.18] | 1.05 [0.83-1.34] | 0.93 [0.80-1.09] | 1.03 [0.96-1.11] | 2,196 | 2,187 | 153 | 441 | 1,872 |
| 674 | 1.19 [1.14-1.25] | 1.22 [1.17-1.27] | 1.11 [0.94-1.31] | 1.07 [0.98-1.17] | 1.13 [1.07-1.18] | 1.15 [1.10-1.20] | 1.15 [1.10-1.21] | 1.08 [0.91-1.29] | 1.05 [0.96-1.15] | 1.09 [1.04-1.15] | 5,402 | 5,356 | 301 | 1,445 | 4,289 |
| 676 | 1.10 [0.97-1.24] | 1.11 [0.99-1.26] | | 0.95 [0.73-1.24] | 1.04 [0.90-1.19] | 1.06 [0.93-1.21] | 1.07 [0.94-1.22] | | 0.90 [0.69-1.19] | 1.02 [0.89-1.18] | 632 | 626 | NA | 145 | 487 |
| dermatologic | | | | | | | | | | | | | | | |
| 681 | 1.61 [1.58-1.63] | 1.65 [1.62-1.67] | 1.64 [1.61-1.66] | 1.38 [1.32-1.44] | 1.49 [1.47-1.52] | 1.53 [1.50-1.55] | 1.50 [1.48-1.53] | 1.49 [1.47-1.52] | 1.34 [1.28-1.40] | 1.44 [1.42-1.46] | 45,803 | 41,863 | 37,950 | 5,186 | 40,017 |
| 681.1 | 1.31 [1.23-1.39] | 1.40 [1.31-1.50] | 1.37 [1.27-1.47] | 1.17 [1.05-1.30] | 1.24 [1.16-1.32] | 1.25 [1.18-1.33] | 1.27 [1.19-1.37] | 1.23 [1.14-1.33] | 1.15 [1.03-1.28] | 1.20 [1.12-1.28] | 2,930 | 2,174 | 1,782 | 913 | 2,545 |
| 681.2 | 1.36 [1.28-1.44] | 1.48 [1.38-1.59] | 1.55 [1.43-1.69] | 1.16 [1.06-1.28] | 1.30 [1.22-1.38] | 1.32 [1.24-1.40] | 1.37 [1.28-1.48] | 1.45 [1.33-1.58] | 1.16 [1.05-1.28] | 1.29 [1.20-1.37] | 2,886 | 1,939 | 1,415 | 1,128 | 2,437 |
| 681.3 | 1.64 [1.61-1.67] | 1.66 [1.63-1.69] | 1.66 [1.63-1.69] | 1.42 [1.34-1.52] | 1.52 [1.49-1.54] | 1.56 [1.53-1.58] | 1.52 [1.49-1.55] | 1.51 [1.48-1.54] | 1.38 [1.29-1.47] | 1.46 [1.44-1.49] | 37,614 | 35,648 | 32,976 | 2,784 | 33,044 |
| 681.5 | 1.64 [1.61-1.67] | 1.66 [1.63-1.69] | 1.66 [1.63-1.69] | 1.42 [1.34-1.52] | 1.52 [1.49-1.54] | 1.56 [1.53-1.58] | 1.52 [1.49-1.55] | 1.51 [1.48-1.54] | 1.38 [1.29-1.47] | 1.46 [1.44-1.49] | 37,614 | 35,648 | 32,976 | 2,784 | 33,044 |
| 681.6 | 1.64 [1.61-1.67] | 1.66 [1.63-1.69] | 1.66 [1.63-1.69] | 1.42 [1.34-1.52] | 1.52 [1.49-1.54] | 1.56 [1.53-1.58] | 1.52 [1.49-1.55] | 1.51 [1.48-1.54] | 1.38 [1.29-1.47] | 1.46 [1.44-1.49] | 37,614 | 35,648 | 32,976 | 2,784 | 33,044 |
| 681.7 | 1.53 [1.45-1.62] | 1.55 [1.47-1.65] | 1.54 [1.45-1.65] | 1.31 [1.13-1.53] | 1.41 [1.33-1.50] | 1.43 [1.35-1.51] | 1.37 [1.29-1.46] | 1.37 [1.28-1.46] | 1.25 [1.07-1.46] | 1.36 [1.28-1.44] | 3,265 | 2,986 | 2,431 | 454 | 2,937 |
| 686 | 1.55 [1.50-1.60] | 1.52 [1.47-1.57] | 1.52 [1.47-1.57] | 1.71 [1.58-1.84] | 1.46 [1.42-1.51] | 1.48 [1.44-1.53] | 1.39 [1.35-1.44] | 1.38 [1.33-1.43] | 1.67 [1.54-1.80] | 1.43 [1.38-1.48] | 12,334 | 10,798 | 9,706 | 1,884 | 11,127 |
| 686.1 | 1.37 [1.34-1.41] | 1.38 [1.34-1.42] | 1.37 [1.32-1.43] | 1.29 [1.22-1.36] | 1.26 [1.22-1.30] | 1.30 [1.26-1.34] | 1.23 [1.19-1.27] | 1.23 [1.18-1.28] | 1.26 [1.19-1.33] | 1.23 [1.19-1.27] | 12,261 | 9,906 | 6,135 | 3,705 | 9,961 |
| 686.2 | 2.28 [2.08-2.51] | 2.65 [2.21-3.17] | 2.27 [1.83-2.82] | 2.22 [2.00-2.48] | 2.16 [1.95-2.39] | 2.10 [1.91-2.32] | 2.19 [1.81-2.65] | 1.96 [1.57-2.46] | 2.09 [1.87-2.33] | 2.01 [1.81-2.23] | 1,345 | 383 | 245 | 1,019 | 1,124 |
| 686.3 | 1.15 [1.09-1.21] | 1.17 [1.11-1.24] | 1.30 [1.15-1.48] | 1.09 [1.02-1.17] | 1.09 [1.02-1.16] | 1.11 [1.05-1.17] | 1.08 [1.02-1.14] | 1.17 [1.03-1.34] | 1.06 [0.98-1.14] | 1.09 [1.02-1.16] | 3,813 | 3,135 | 557 | 2,154 | 2,607 |
| 686.4 | 1.30 [1.18-1.43] | 1.32 [1.18-1.49] | 1.29 [1.12-1.48] | 1.26 [1.08-1.47] | 1.25 [1.13-1.39] | 1.24 [1.12-1.36] | 1.21 [1.08-1.37] | 1.20 [1.04-1.39] | 1.22 [1.05-1.42] | 1.19 [1.07-1.33] | 1,099 | 710 | 487 | 470 | 912 |
| 686.5 | 1.84 [1.54-2.21] | 1.90 [1.56-2.30] | 1.85 [1.49-2.30] | | 1.58 [1.31-1.91] | 1.71 [1.42-2.06] | 1.66 [1.36-2.03] | 1.65 [1.32-2.07] | | 1.52 [1.26-1.84] | 344 | 293 | 228 | NA | 306 |
| 687 | 1.25 [1.20-1.29] | 1.33 [1.26-1.40] | 1.33 [1.26-1.41] | 1.16 [1.10-1.22] | 1.18 [1.14-1.23] | 1.19 [1.14-1.24] | 1.21 [1.15-1.28] | 1.21 [1.14-1.28] | 1.12 [1.06-1.18] | 1.14 [1.10-1.19] | 7,127 | 3,782 | 3,302 | 3,510 | 6,405 |
| 687.1 | 1.47 [1.43-1.50] | 1.68 [1.64-1.73] | 1.76 [1.70-1.81] | 1.18 [1.14-1.22] | 1.38 [1.35-1.41] | 1.39 [1.36-1.42] | 1.53 [1.49-1.58] | 1.61 [1.56-1.66] | 1.14 [1.10-1.18] | 1.33 [1.30-1.36] | 21,832 | 13,595 | 10,995 | 9,151 | 19,112 |
| 687.2 | 1.38 [1.34-1.43] | 1.37 [1.31-1.42] | 1.36 [1.31-1.42] | 1.42 [1.33-1.52] | 1.31 [1.27-1.36] | 1.29 [1.25-1.34] | 1.22 [1.17-1.27] | 1.23 [1.18-1.29] | 1.36 [1.27-1.45] | 1.25 [1.20-1.29] | 8,990 | 6,898 | 5,433 | 2,598 | 7,743 |
| 687.3 | 1.60 [1.38-1.85] | 1.59 [1.35-1.87] | 1.46 [1.22-1.74] | 1.44 [1.04-2.00] | 1.48 [1.26-1.73] | 1.53 [1.31-1.78] | 1.44 [1.21-1.70] | 1.31 [1.09-1.58] | 1.42 [1.02-1.99] | 1.42 [1.21-1.66] | 485 | 395 | 330 | 101 | 424 |
| 687.4 | 1.27 [1.23-1.30] | 1.27 [1.23-1.31] | 1.27 [1.23-1.32] | 1.15 [1.06-1.24] | 1.16 [1.13-1.20] | 1.16 [1.12-1.20] | 1.11 [1.07-1.14] | 1.11 [1.07-1.15] | 1.09 [1.00-1.18] | 1.10 [1.06-1.13] | 10,595 | 9,813 | 7,028 | 1,678 | 9,380 |
| 689 | 1.54 [1.51-1.58] | 1.49 [1.46-1.53] | 1.47 [1.43-1.51] | 1.88 [1.78-2.00] | 1.44 [1.40-1.47] | 1.46 [1.43-1.49] | 1.38 [1.34-1.41] | 1.36 [1.32-1.39] | 1.79 [1.68-1.90] | 1.38 [1.35-1.42] | 21,581 | 18,840 | 16,230 | 3,546 | 18,853 |
| 690 | 27.42 [23.35-32.19] | 19.22 [15.25-24.22] | 13.15 [10.11-17.10] | 32.12 [26.19-39.39] | 24.24 [20.43-28.76] | 25.91 [22.01-30.51] | 16.97 [13.43-21.46] | 12.33 [9.45-16.10] | 29.95 [24.37-36.80] | 22.62 [19.02-26.91] | 2,347 | 781 | 453 | 1,711 | 1,917 |
| 690.1 | 3.29 [2.87-3.77] | 3.13 [2.71-3.62] | 3.22 [2.77-3.75] | 3.22 [2.24-4.64] | 2.86 [2.48-3.29] | 3.09 [2.69-3.56] | 2.85 [2.45-3.30] | 2.91 [2.49-3.40] | 3.12 [2.12-4.57] | 2.72 [2.36-3.15] | 776 | 682 | 616 | 114 | 678 |
| 691 | 1.49 [1.27-1.76] | 1.85 [1.48-2.30] | 1.99 [1.56-2.54] | 1.24 [0.97-1.58] | 1.44 [1.20-1.71] | 1.39 [1.17-1.64] | 1.71 [1.36-2.13] | 1.74 [1.36-2.24] | 1.18 [0.92-1.51] | 1.34 [1.12-1.60] | 380 | 223 | 183 | 168 | 325 |
| 691.1 | 2.79 [1.97-3.95] | | | | 2.74 [1.90-3.95] | 2.58 [1.81-3.68] | | | | 2.50 [1.71-3.64] | 119 | NA | NA | NA | 105 |
| 694.1 | 1.97 [1.69-2.29] | 1.85 [1.57-2.18] | 1.90 [1.59-2.27] | 2.46 [1.72-3.53] | 1.83 [1.56-2.15] | 1.80 [1.54-2.11] | 1.65 [1.40-1.96] | 1.67 [1.39-2.01] | 2.34 [1.60-3.42] | 1.73 [1.46-2.04] | 492 | 410 | 343 | 106 | 427 |
| 694.2 | 1.29 [1.20-1.39] | 1.29 [1.19-1.39] | 1.27 [1.17-1.38] | 1.50 [1.26-1.78] | 1.24 [1.15-1.34] | 1.23 [1.14-1.32] | 1.20 [1.11-1.30] | 1.19 [1.10-1.30] | 1.45 [1.21-1.72] | 1.19 [1.10-1.29] | 1,910 | 1,583 | 1,347 | 395 | 1,633 |
| 694.3 | 1.62 [1.43-1.84] | 1.79 [1.56-2.04] | 1.70 [1.47-1.96] | | 1.51 [1.32-1.73] | 1.51 [1.32-1.72] | 1.64 [1.43-1.88] | 1.53 [1.32-1.77] | | 1.42 [1.24-1.63] | 660 | 603 | 524 | NA | 569 |
| 695.1 | 1.32 [1.19-1.47] | 1.81 [1.56-2.10] | 2.03 [1.71-2.39] | 0.99 [0.86-1.15] | 1.27 [1.13-1.42] | 1.26 [1.13-1.40] | 1.62 [1.39-1.89] | 1.81 [1.52-2.15] | 0.98 [0.84-1.13] | 1.21 [1.08-1.36] | 896 | 483 | 390 | 442 | 760 |
| 695.21 | 3.51 [2.89-4.26] | 3.96 [3.22-4.86] | 3.58 [2.87-4.48] | | 3.07 [2.49-3.78] | 3.29 [2.70-4.02] | 3.61 [2.93-4.46] | 3.32 [2.64-4.17] | | 2.92 [2.36-3.61] | 375 | 341 | 282 | NA | 311 |
| 695.22 | 2.81 [2.55-3.09] | 2.76 [2.50-3.03] | 2.76 [2.51-3.04] | | 2.57 [2.32-2.84] | 2.69 [2.44-2.96] | 2.59 [2.35-2.85] | 2.58 [2.34-2.84] | | 2.48 [2.24-2.75] | 1,545 | 1,526 | 1,504 | NA | 1,375 |
| 695.3 | 1.55 [1.39-1.72] | 1.59 [1.43-1.78] | 1.52 [1.36-1.70] | | 1.50 [1.34-1.68] | 1.44 [1.29-1.61] | 1.49 [1.33-1.66] | 1.40 [1.25-1.57] | | 1.43 [1.27-1.60] | 890 | 873 | 798 | NA | 784 |
| 695.41 | 1.86 [1.66-2.10] | 1.86 [1.66-2.10] | 1.84 [1.62-2.10] | | 1.68 [1.49-1.90] | 1.67 [1.48-1.88] | 1.61 [1.42-1.82] | 1.58 [1.38-1.81] | | 1.55 [1.36-1.75] | 786 | 768 | 620 | NA | 700 |
| 695.42 | 1.79 [1.65-1.95] | 1.75 [1.61-1.90] | 1.69 [1.54-1.86] | 1.73 [1.30-2.30] | 1.62 [1.49-1.76] | 1.63 [1.49-1.77] | 1.52 [1.39-1.65] | 1.48 [1.34-1.63] | 1.73 [1.29-2.32] | 1.49 [1.36-1.63] | 1,578 | 1,517 | 1,106 | 150 | 1,379 |
| 695.7 | 3.00 [2.81-3.19] | 2.88 [2.70-3.07] | 2.79 [2.60-2.98] | 5.26 [3.96-7.00] | 2.65 [2.48-2.84] | 2.79 [2.61-2.98] | 2.64 [2.47-2.82] | 2.55 [2.38-2.73] | 4.75 [3.54-6.38] | 2.51 [2.34-2.69] | 3,243 | 3,039 | 2,718 | 259 | 2,749 |
| 695.8 | 1.82 [1.66-1.98] | 1.88 [1.70-2.08] | 1.74 [1.57-1.93] | 1.63 [1.36-1.94] | 1.67 [1.52-1.83] | 1.72 [1.57-1.88] | 1.67 [1.51-1.85] | 1.56 [1.40-1.74] | 1.59 [1.33-1.90] | 1.59 [1.45-1.75] | 1,463 | 1,166 | 1,027 | 344 | 1,272 |
| 695.81 | 1.49 [1.25-1.79] | 1.50 [1.22-1.84] | 1.49 [1.12-1.96] | 1.47 [1.11-1.96] | 1.36 [1.12-1.66] | 1.38 [1.15-1.66] | 1.33 [1.07-1.64] | 1.28 [0.96-1.71] | 1.36 [1.01-1.82] | 1.24 [1.01-1.52] | 329 | 233 | 127 | 139 | 264 |
| 695.9 | 1.78 [1.69-1.87] | 1.85 [1.75-1.96] | 1.88 [1.77-1.99] | 1.45 [1.29-1.64] | 1.66 [1.57-1.75] | 1.68 [1.60-1.78] | 1.70 [1.60-1.80] | 1.71 [1.61-1.82] | 1.37 [1.20-1.55] | 1.61 [1.52-1.70] | 4,146 | 3,600 | 3,135 | 714 | 3,688 |
| 696.3 | 1.80 [1.46-2.23] | 1.94 [1.54-2.44] | 2.09 [1.59-2.74] | | 1.69 [1.34-2.13] | 1.70 [1.37-2.11] | 1.74 [1.37-2.21] | 1.92 [1.45-2.54] | | 1.60 [1.27-2.03] | 250 | 211 | 159 | NA | 208 |
| 696.4 | 2.23 [2.17-2.30] | 2.19 [2.13-2.26] | 2.21 [2.14-2.28] | 2.04 [1.84-2.26] | 2.11 [2.04-2.17] | 2.13 [2.07-2.20] | 2.02 [1.96-2.08] | 2.03 [1.97-2.10] | 2.01 [1.80-2.23] | 2.04 [1.98-2.11] | 14,006 | 13,506 | 11,116 | 1,195 | 11,883 |
| 696.41 | 2.30 [2.23-2.37] | 2.27 [2.20-2.34] | 2.28 [2.20-2.36] | 2.11 [1.90-2.35] | 2.17 [2.10-2.24] | 2.20 [2.13-2.27] | 2.08 [2.02-2.15] | 2.09 [2.02-2.17] | 2.08 [1.86-2.33] | 2.11 [2.04-2.18] | 12,400 | 11,939 | 9,842 | 1,114 | 10,571 |
| 696.42 | 1.73 [1.62-1.85] | 1.69 [1.58-1.81] | 1.70 [1.58-1.83] | 1.37 [1.00-1.86] | 1.59 [1.48-1.71] | 1.62 [1.51-1.74] | 1.52 [1.42-1.63] | 1.53 [1.42-1.65] | 1.33 [0.97-1.82] | 1.52 [1.41-1.63] | 2,322 | 2,269 | 1,879 | 116 | 1,993 |
| 697 | 1.57 [1.46-1.70] | 1.55 [1.43-1.67] | 1.51 [1.39-1.64] | 1.78 [1.27-2.51] | 1.43 [1.31-1.55] | 1.44 [1.33-1.56] | 1.39 [1.29-1.51] | 1.34 [1.23-1.46] | 1.67 [1.17-2.37] | 1.32 [1.22-1.44] | 1,717 | 1,649 | 1,425 | 102 | 1,485 |
| 698 | 1.72 [1.64-1.80] | 1.68 [1.60-1.76] | 1.99 [1.87-2.12] | 1.46 [1.31-1.62] | 1.59 [1.51-1.67] | 1.60 [1.52-1.68] | 1.50 [1.42-1.58] | 1.78 [1.67-1.90] | 1.36 [1.21-1.51] | 1.50 [1.42-1.58] | 4,866 | 4,324 | 2,764 | 1,002 | 4,307 |
| 700 | 1.49 [1.31-1.69] | 1.47 [1.29-1.67] | 1.50 [1.31-1.72] | | 1.32 [1.16-1.51] | 1.39 [1.22-1.58] | 1.33 [1.16-1.51] | 1.35 [1.18-1.55] | | 1.26 [1.10-1.44] | 634 | 609 | 550 | NA | 566 |
| 701 | 1.36 [1.31-1.40] | 1.38 [1.34-1.43] | 1.38 [1.33-1.43] | 1.18 [1.10-1.28] | 1.27 [1.22-1.31] | 1.28 [1.24-1.33] | 1.28 [1.24-1.33] | 1.27 [1.23-1.32] | 1.17 [1.09-1.27] | 1.22 [1.18-1.26] | 10,113 | 8,842 | 6,975 | 1,829 | 8,668 |
| 701.1 | 2.28 [1.79-2.90] | 2.04 [1.60-2.62] | 2.21 [1.71-2.87] | | 2.22 [1.73-2.86] | 2.14 [1.68-2.74] | 1.87 [1.45-2.41] | 2.07 [1.58-2.70] | | 2.13 [1.65-2.75] | 204 | 183 | 169 | NA | 184 |
| 701.2 | 1.19 [1.14-1.25] | 1.24 [1.18-1.31] | 1.20 [1.13-1.27] | 1.00 [0.90-1.12] | 1.12 [1.07-1.18] | 1.13 [1.08-1.18] | 1.15 [1.10-1.21] | 1.11 [1.05-1.18] | 0.99 [0.88-1.11] | 1.09 [1.04-1.15] | 4,378 | 3,873 | 2,861 | 789 | 3,863 |
| 701.3 | 1.46 [1.18-1.81] | 1.50 [1.19-1.89] | 1.69 [1.31-2.16] | | 1.33 [1.06-1.67] | 1.37 [1.10-1.70] | 1.38 [1.08-1.75] | 1.50 [1.16-1.94] | | 1.27 [1.00-1.60] | 221 | 182 | 166 | NA | 186 |
| 701.4 | 1.29 [1.17-1.42] | 1.31 [1.16-1.47] | 1.33 [1.11-1.58] | 1.27 [1.11-1.46] | 1.19 [1.06-1.32] | 1.22 [1.10-1.35] | 1.22 [1.08-1.38] | 1.22 [1.02-1.47] | 1.26 [1.10-1.45] | 1.14 [1.02-1.27] | 1,088 | 689 | 306 | 579 | 854 |
| 701.5 | 1.29 [1.20-1.39] | 1.39 [1.27-1.51] | 1.37 [1.24-1.51] | 1.10 [0.96-1.27] | 1.20 [1.11-1.30] | 1.21 [1.12-1.30] | 1.25 [1.15-1.36] | 1.24 [1.12-1.37] | 1.04 [0.90-1.19] | 1.15 [1.06-1.25] | 1,791 | 1,376 | 1,000 | 545 | 1,521 |
| 701.6 | 1.67 [1.43-1.96] | 1.82 [1.51-2.18] | 1.71 [1.41-2.08] | 1.58 [1.18-2.12] | 1.55 [1.31-1.83] | 1.56 [1.33-1.84] | 1.60 [1.32-1.93] | 1.56 [1.27-1.90] | 1.51 [1.11-2.04] | 1.46 [1.23-1.74] | 456 | 322 | 278 | 150 | 387 |
| 702.1 | 1.37 [1.32-1.42] | 1.35 [1.30-1.40] | 1.37 [1.32-1.41] | | 1.29 [1.24-1.34] | 1.30 [1.25-1.34] | 1.26 [1.22-1.31] | 1.27 [1.22-1.32] | | 1.23 [1.18-1.28] | 8,337 | 8,281 | 8,115 | NA | 7,420 |
| 702.2 | 1.22 [1.17-1.28] | 1.25 [1.19-1.31] | 1.26 [1.20-1.31] | | 1.18 [1.13-1.24] | 1.16 [1.11-1.21] | 1.17 [1.12-1.22] | 1.17 [1.12-1.23] | | 1.13 [1.07-1.18] | 4,915 | 4,889 | 4,688 | NA | 4,220 |
| 703 | 1.42 [1.34-1.49] | 1.48 [1.39-1.58] | 1.51 [1.39-1.63] | 1.32 [1.22-1.43] | 1.33 [1.25-1.41] | 1.30 [1.23-1.38] | 1.33 [1.24-1.42] | 1.32 [1.22-1.43] | 1.24 [1.14-1.34] | 1.24 [1.17-1.32] | 3,705 | 2,360 | 1,554 | 1,741 | 2,910 |
| 703.1 | 1.42 [1.34-1.51] | 1.52 [1.41-1.64] | 1.58 [1.43-1.75] | 1.34 [1.24-1.46] | 1.36 [1.27-1.46] | 1.31 [1.23-1.39] | 1.36 [1.25-1.47] | 1.37 [1.23-1.52] | 1.26 [1.16-1.37] | 1.27 [1.19-1.37] | 2,927 | 1,683 | 991 | 1,606 | 2,260 |
| 704 | 1.17 [1.12-1.21] | 1.17 [1.13-1.22] | 1.16 [1.11-1.21] | 1.13 [1.02-1.25] | 1.10 [1.05-1.14] | 1.12 [1.08-1.17] | 1.10 [1.06-1.14] | 1.09 [1.04-1.14] | 1.08 [0.97-1.20] | 1.07 [1.03-1.12] | 6,656 | 6,255 | 4,344 | 951 | 5,126 |
| 704.1 | 1.96 [1.76-2.19] | 2.02 [1.79-2.27] | 2.00 [1.74-2.30] | 1.84 [1.49-2.28] | 1.86 [1.66-2.08] | 1.75 [1.56-1.95] | 1.73 [1.53-1.96] | 1.72 [1.49-1.98] | 1.72 [1.38-2.15] | 1.66 [1.47-1.87] | 1,010 | 811 | 577 | 271 | 853 |
| 704.11 | 2.87 [2.32-3.56] | 2.84 [2.24-3.59] | 2.62 [1.97-3.48] | | 2.70 [2.15-3.40] | 2.27 [1.80-2.85] | 2.08 [1.61-2.68] | 2.06 [1.53-2.79] | | 2.19 [1.71-2.79] | 290 | 234 | 151 | NA | 243 |
| 704.2 | 1.25 [1.00-1.56] | 1.28 [1.00-1.64] | | | 1.10 [0.86-1.41] | 1.19 [0.95-1.49] | 1.12 [0.87-1.45] | | | 1.05 [0.82-1.35] | 211 | 157 | NA | NA | 167 |
| 704.8 | 1.40 [1.10-1.79] | 1.35 [1.05-1.74] | 1.41 [1.06-1.88] | | 1.29 [1.00-1.67] | 1.31 [1.02-1.68] | 1.20 [0.92-1.56] | 1.30 [0.97-1.74] | | 1.22 [0.94-1.59] | 169 | 152 | 114 | NA | 148 |
| 705.1 | 5.39 [4.03-7.21] | 4.31 [3.17-5.86] | 3.59 [2.55-5.05] | | 5.00 [3.69-6.77] | 5.05 [3.73-6.83] | 3.75 [2.73-5.14] | 3.11 [2.18-4.44] | | 4.73 [3.45-6.47] | 228 | 171 | 132 | NA | 191 |
| 705.3 | 1.56 [1.40-1.72] | 1.54 [1.39-1.71] | 1.51 [1.28-1.77] | 1.32 [1.07-1.63] | 1.32 [1.19-1.47] | 1.40 [1.26-1.56] | 1.28 [1.14-1.42] | 1.23 [1.04-1.46] | 1.25 [1.01-1.56] | 1.26 [1.12-1.40] | 991 | 944 | 362 | 240 | 847 |
| 705.8 | 1.36 [1.27-1.45] | 1.38 [1.29-1.48] | 1.34 [1.24-1.45] | 1.24 [1.07-1.45] | 1.23 [1.15-1.32] | 1.24 [1.16-1.32] | 1.21 [1.13-1.30] | 1.17 [1.08-1.27] | 1.18 [1.01-1.37] | 1.16 [1.09-1.25] | 2,502 | 2,240 | 1,590 | 476 | 2,119 |
| 706.1 | 1.46 [1.31-1.63] | 1.43 [1.26-1.62] | 1.45 [1.15-1.84] | 1.36 [1.17-1.58] | 1.30 [1.16-1.47] | 1.34 [1.19-1.50] | 1.24 [1.09-1.41] | 1.26 [0.99-1.62] | 1.27 [1.09-1.49] | 1.23 [1.09-1.40] | 911 | 642 | 175 | 518 | 738 |
| 706.2 | 1.16 [1.13-1.20] | 1.18 [1.14-1.21] | 1.17 [1.13-1.22] | 1.11 [1.04-1.20] | 1.09 [1.06-1.13] | 1.12 [1.08-1.15] | 1.09 [1.06-1.13] | 1.10 [1.06-1.14] | 1.08 [1.00-1.17] | 1.07 [1.03-1.10] | 11,104 | 10,136 | 7,216 | 1,829 | 8,562 |
| 706.8 | 2.65 [1.99-3.52] | 2.55 [1.85-3.52] | 2.85 [2.04-3.98] | | 2.47 [1.86-3.29] | 2.49 [1.86-3.34] | 2.30 [1.65-3.19] | 2.56 [1.81-3.60] | | 2.42 [1.81-3.24] | 171 | 139 | 127 | NA | 165 |
| 707 | 1.54 [1.50-1.57] | 1.53 [1.49-1.56] | 1.52 [1.49-1.56] | 1.28 [0.99-1.64] | 1.45 [1.41-1.48] | 1.48 [1.45-1.52] | 1.42 [1.39-1.45] | 1.41 [1.38-1.44] | 1.28 [0.99-1.66] | 1.42 [1.39-1.46] | 22,548 | 22,452 | 22,090 | 165 | 20,699 |
| 707.1 | 1.22 [1.20-1.25] | 1.21 [1.19-1.24] | 1.22 [1.19-1.24] | 0.99 [0.82-1.20] | 1.18 [1.16-1.21] | 1.20 [1.17-1.22] | 1.15 [1.13-1.18] | 1.15 [1.13-1.17] | 0.97 [0.80-1.18] | 1.18 [1.16-1.21] | 26,249 | 26,064 | 25,754 | 284 | 24,650 |
| 709.2 | 1.66 [1.54-1.80] | 1.69 [1.56-1.84] | 1.67 [1.54-1.82] | | 1.51 [1.39-1.64] | 1.48 [1.36-1.61] | 1.46 [1.34-1.59] | 1.43 [1.31-1.56] | | 1.35 [1.24-1.47] | 1,608 | 1,600 | 1,483 | NA | 1,444 |
| 709.3 | 1.73 [1.52-1.98] | 1.64 [1.44-1.87] | 1.65 [1.44-1.89] | | 1.54 [1.34-1.77] | 1.61 [1.40-1.84] | 1.48 [1.30-1.70] | 1.49 [1.29-1.71] | | 1.45 [1.26-1.67] | 611 | 591 | 535 | NA | 530 |
| 709.4 | 1.53 [1.19-1.96] | 1.52 [1.18-1.94] | 1.44 [1.11-1.88] | | 1.46 [1.11-1.91] | 1.47 [1.14-1.90] | 1.41 [1.09-1.82] | 1.36 [1.04-1.79] | | 1.39 [1.05-1.83] | 164 | 162 | 140 | NA | 135 |
| 709.5 | 2.10 [1.78-2.47] | 2.12 [1.78-2.52] | 2.04 [1.69-2.45] | | 1.92 [1.60-2.31] | 2.03 [1.71-2.40] | 2.00 [1.67-2.39] | 1.94 [1.61-2.35] | | 1.84 [1.52-2.22] | 412 | 363 | 315 | NA | 322 |
| 709.6 | 1.79 [1.47-2.18] | 1.77 [1.45-2.17] | 1.78 [1.42-2.22] | | 1.51 [1.24-1.85] | 1.60 [1.31-1.96] | 1.56 [1.26-1.92] | 1.58 [1.25-1.98] | | 1.38 [1.12-1.69] | 290 | 267 | 211 | NA | 259 |
| 709.7 | 1.40 [1.34-1.47] | 1.39 [1.33-1.46] | 1.41 [1.34-1.49] | 1.29 [1.14-1.45] | 1.29 [1.23-1.35] | 1.28 [1.23-1.35] | 1.23 [1.17-1.30] | 1.24 [1.18-1.31] | 1.26 [1.11-1.43] | 1.21 [1.15-1.27] | 4,677 | 4,045 | 3,723 | 689 | 4,076 |
| 938 | 1.97 [1.65-2.36] | 1.97 [1.63-2.37] | 1.97 [1.62-2.41] | | 1.64 [1.35-1.98] | 1.87 [1.55-2.24] | 1.84 [1.52-2.22] | 1.82 [1.49-2.24] | | 1.61 [1.32-1.97] | 350 | 320 | 279 | NA | 290 |
| 938.1 | 1.48 [1.15-1.90] | 1.69 [1.25-2.30] | | | 1.50 [1.15-1.97] | 1.43 [1.11-1.84] | 1.60 [1.17-2.20] | | | 1.45 [1.10-1.91] | 171 | 111 | NA | NA | 150 |
| 938.2 | 1.32 [1.27-1.37] | 1.31 [1.26-1.35] | 1.32 [1.28-1.37] | | 1.25 [1.20-1.30] | 1.25 [1.21-1.30] | 1.22 [1.18-1.27] | 1.24 [1.19-1.28] | | 1.20 [1.15-1.24] | 7,794 | 7,772 | 7,653 | NA | 6,959 |
| 939 | 7.81 [7.68-7.95] | 6.82 [6.67-6.96] | 6.01 [5.86-6.15] | 9.08 [8.86-9.31] | 6.95 [6.83-7.07] | 7.23 [7.10-7.35] | 6.12 [5.98-6.25] | 5.50 [5.36-5.64] | 8.34 [8.13-8.56] | 6.48 [6.36-6.60] | 84,723 | 46,492 | 32,628 | 43,598 | 71,715 |
| 947 | 1.90 [1.81-1.99] | 1.77 [1.66-1.88] | 1.81 [1.69-1.95] | 2.04 [1.91-2.18] | 1.83 [1.74-1.92] | 1.64 [1.56-1.72] | 1.46 [1.37-1.55] | 1.50 [1.39-1.61] | 1.77 [1.65-1.89] | 1.58 [1.50-1.66] | 5,321 | 2,835 | 2,084 | 2,772 | 4,679 |

Supplementary Table 6: phecode-mapped results (hazard ratios and events from all cohorts)

Hazard ratio (99% confidence interval)

| | crude | | | | | adjusted | | | | | Events (in exposed) | | | | |
|---|---|---|---|---|---|---|---|---|---|---|---|---|---|---|---|
| Outcome | any age | 18+ | 40+ | <18 | hosp. | any age | 18+ | 40+ | <18 | hosp. | any age | 18+ | 40+ | <18 | hosp. |
| musculoskeletal | | | | | | | | | | | | | | | |
| 710 | 1.49 [1.15-1.92] | 1.61 [1.24-2.08] | 1.48 [1.14-1.93] | | 1.57 [1.19-2.06] | 1.50 [1.16-1.94] | 1.56 [1.20-2.03] | 1.41 [1.08-1.84] | | 1.61 [1.22-2.12] | 154 | 151 | 143 | NA | 135 |
| 710.11 | 1.35 [1.12-1.62] | 1.35 [1.10-1.66] | 1.39 [1.12-1.73] | | 1.34 [1.10-1.62] | 1.31 [1.09-1.58] | 1.24 [1.00-1.53] | 1.27 [1.02-1.58] | | 1.33 [1.10-1.62] | 309 | 231 | 214 | NA | 278 |
| 710.12 | 1.39 [1.25-1.54] | 1.43 [1.28-1.59] | 1.45 [1.29-1.64] | 1.27 [0.98-1.65] | 1.26 [1.13-1.40] | 1.32 [1.19-1.46] | 1.29 [1.15-1.44] | 1.32 [1.17-1.49] | 1.24 [0.95-1.62] | 1.22 [1.10-1.36] | 953 | 825 | 735 | 160 | 855 |
| 710.19 | 1.37 [1.31-1.45] | 1.39 [1.32-1.47] | 1.38 [1.30-1.46] | 1.37 [1.20-1.56] | 1.28 [1.22-1.35] | 1.32 [1.25-1.39] | 1.25 [1.19-1.33] | 1.23 [1.16-1.31] | 1.34 [1.17-1.54] | 1.26 [1.20-1.33] | 4,040 | 3,496 | 3,260 | 622 | 3,674 |
| 711 | 1.36 [1.11-1.66] | 1.30 [1.04-1.63] | 1.36 [1.07-1.73] | | 1.29 [1.05-1.59] | 1.23 [1.00-1.51] | 1.12 [0.88-1.41] | 1.16 [0.90-1.49] | | 1.22 [0.98-1.51] | 254 | 196 | 169 | NA | 227 |
| 711.1 | 1.48 [1.39-1.59] | 1.50 [1.39-1.61] | 1.54 [1.42-1.66] | 1.40 [1.20-1.64] | 1.35 [1.26-1.45] | 1.41 [1.32-1.51] | 1.37 [1.27-1.48] | 1.41 [1.30-1.52] | 1.37 [1.16-1.61] | 1.31 [1.22-1.41] | 2,261 | 1,904 | 1,742 | 421 | 1,990 |
| 711.2 | 1.24 [0.92-1.67] | | | | | 1.14 [0.84-1.54] | | | | | 111 | NA | NA | NA | NA |
| 711.3 | 1.31 [0.98-1.76] | 1.29 [0.96-1.74] | | | | 1.22 [0.90-1.65] | 1.14 [0.83-1.55] | | | | 119 | 108 | NA | NA | NA |
| 712 | 1.38 [1.21-1.57] | 1.36 [1.18-1.56] | 1.31 [1.13-1.53] | 1.47 [1.08-1.98] | 1.29 [1.12-1.48] | 1.34 [1.17-1.53] | 1.28 [1.10-1.48] | 1.25 [1.07-1.46] | 1.42 [1.03-1.96] | 1.29 [1.12-1.49] | 597 | 491 | 420 | 123 | 525 |
| 713 | 1.25 [1.11-1.40] | 1.40 [1.21-1.64] | 1.31 [1.10-1.56] | 1.13 [0.95-1.34] | 1.18 [1.04-1.34] | 1.20 [1.07-1.35] | 1.29 [1.10-1.51] | 1.21 [1.01-1.45] | 1.11 [0.94-1.32] | 1.15 [1.02-1.31] | 735 | 431 | 309 | 347 | 629 |
| 713.5 | 1.69 [1.46-1.96] | 1.70 [1.46-1.97] | 1.67 [1.44-1.94] | | 1.42 [1.22-1.64] | 1.52 [1.31-1.77] | 1.39 [1.19-1.63] | 1.34 [1.14-1.57] | | 1.34 [1.15-1.55] | 468 | 468 | 452 | NA | 443 |
| 714 | 1.46 [1.36-1.56] | 1.43 [1.34-1.53] | 1.45 [1.35-1.56] | | 1.32 [1.23-1.42] | 1.32 [1.23-1.41] | 1.25 [1.16-1.34] | 1.27 [1.18-1.36] | | 1.23 [1.15-1.32] | 2,226 | 2,188 | 2,056 | NA | 2,038 |
| 714.1 | 1.41 [1.37-1.44] | 1.40 [1.37-1.43] | 1.39 [1.36-1.43] | 1.28 [1.05-1.57] | 1.31 [1.28-1.34] | 1.32 [1.28-1.35] | 1.27 [1.24-1.30] | 1.26 [1.23-1.29] | 1.22 [1.00-1.50] | 1.24 [1.21-1.27] | 17,617 | 17,574 | 16,389 | 279 | 15,331 |
| 714.2 | 1.33 [1.19-1.48] | 1.38 [1.09-1.73] | | 1.34 [1.19-1.51] | 1.25 [1.10-1.41] | 1.28 [1.14-1.43] | 1.26 [1.00-1.60] | | 1.30 [1.15-1.47] | 1.23 [1.08-1.39] | 852 | 183 | NA | 723 | 706 |
| 715 | 1.38 [1.33-1.43] | 1.37 [1.32-1.43] | 1.38 [1.32-1.44] | 1.08 [0.83-1.41] | 1.27 [1.22-1.32] | 1.25 [1.20-1.31] | 1.20 [1.15-1.25] | 1.20 [1.15-1.25] | 1.09 [0.83-1.42] | 1.19 [1.15-1.24] | 6,405 | 6,314 | 5,957 | 153 | 5,911 |
| 715.1 | 1.43 [1.24-1.64] | 1.44 [1.25-1.66] | 1.43 [1.22-1.67] | | 1.28 [1.12-1.48] | 1.26 [1.09-1.45] | 1.21 [1.04-1.40] | 1.17 [0.99-1.37] | | 1.19 [1.03-1.37] | 537 | 510 | 400 | NA | 502 |
| 715.2 | 1.50 [1.39-1.62] | 1.56 [1.45-1.68] | 1.48 [1.37-1.61] | | 1.42 [1.32-1.54] | 1.39 [1.29-1.50] | 1.39 [1.28-1.50] | 1.31 [1.20-1.42] | | 1.33 [1.23-1.45] | 1,810 | 1,796 | 1,543 | NA | 1,588 |
| 716 | 1.48 [1.31-1.68] | 1.49 [1.31-1.70] | 1.54 [1.34-1.78] | | 1.34 [1.18-1.53] | 1.33 [1.17-1.51] | 1.25 [1.08-1.43] | 1.31 [1.13-1.52] | | 1.26 [1.10-1.43] | 648 | 570 | 502 | NA | 583 |
| 716.1 | 1.37 [1.34-1.41] | 1.37 [1.34-1.41] | 1.36 [1.33-1.40] | | 1.27 [1.24-1.30] | 1.26 [1.23-1.29] | 1.22 [1.19-1.25] | 1.21 [1.18-1.24] | | 1.19 [1.16-1.22] | 18,108 | 18,032 | 17,859 | NA | 16,872 |
| 716.2 | 1.25 [1.24-1.27] | 1.25 [1.24-1.27] | 1.25 [1.23-1.27] | 1.17 [1.00-1.37] | 1.17 [1.15-1.19] | 1.18 [1.16-1.20] | 1.16 [1.14-1.18] | 1.15 [1.14-1.17] | 1.12 [0.95-1.33] | 1.12 [1.10-1.14] | 45,851 | 45,760 | 44,156 | 393 | 39,506 |
| 716.8 | 1.65 [1.26-2.15] | 1.56 [1.19-2.05] | 1.48 [1.11-1.97] | | 1.43 [1.09-1.88] | 1.52 [1.16-2.01] | 1.37 [1.03-1.81] | 1.27 [0.94-1.72] | | 1.37 [1.04-1.81] | 143 | 140 | 115 | NA | 127 |
| 716.9 | 1.32 [1.31-1.33] | 1.32 [1.30-1.33] | 1.31 [1.30-1.33] | 1.24 [1.14-1.35] | 1.23 [1.22-1.24] | 1.23 [1.21-1.24] | 1.19 [1.18-1.20] | 1.18 [1.17-1.19] | 1.16 [1.07-1.27] | 1.16 [1.15-1.18] | 121,759 | 121,279 | 116,846 | 1,514 | 106,193 |
| 717 | 1.38 [1.34-1.43] | 1.39 [1.34-1.44] | 1.38 [1.34-1.43] | | 1.32 [1.28-1.37] | 1.30 [1.26-1.35] | 1.29 [1.25-1.33] | 1.28 [1.24-1.32] | | 1.26 [1.22-1.31] | 9,733 | 9,731 | 9,658 | NA | 8,595 |
| 720 | 1.30 [1.27-1.34] | 1.30 [1.27-1.34] | 1.30 [1.26-1.33] | 1.10 [0.82-1.48] | 1.20 [1.17-1.24] | 1.19 [1.16-1.22] | 1.15 [1.11-1.18] | 1.13 [1.10-1.16] | 1.02 [0.75-1.38] | 1.14 [1.11-1.17] | 14,146 | 14,135 | 13,686 | 120 | 13,127 |
| 721 | 1.35 [1.33-1.38] | 1.36 [1.34-1.38] | 1.35 [1.32-1.37] | 1.06 [0.84-1.34] | 1.26 [1.23-1.28] | 1.23 [1.21-1.26] | 1.19 [1.17-1.21] | 1.18 [1.16-1.20] | 0.99 [0.78-1.27] | 1.18 [1.16-1.20] | 32,132 | 32,112 | 30,898 | 193 | 29,839 |
| 721.1 | 1.35 [1.32-1.38] | 1.36 [1.33-1.39] | 1.36 [1.33-1.39] | 0.96 [0.72-1.29] | 1.25 [1.23-1.28] | 1.23 [1.20-1.26] | 1.19 [1.16-1.22] | 1.19 [1.16-1.22] | 0.93 [0.69-1.27] | 1.18 [1.15-1.20] | 20,283 | 20,277 | 19,516 | 117 | 18,927 |
| 721.2 | 1.32 [1.18-1.49] | 1.32 [1.17-1.49] | 1.26 [1.12-1.42] | | 1.23 [1.09-1.39] | 1.24 [1.10-1.39] | 1.19 [1.05-1.34] | 1.11 [0.98-1.25] | | 1.18 [1.04-1.33] | 704 | 702 | 682 | NA | 661 |
| 721.8 | 1.27 [1.21-1.35] | 1.26 [1.19-1.33] | 1.26 [1.19-1.34] | 1.12 [0.87-1.45] | 1.19 [1.12-1.26] | 1.19 [1.13-1.26] | 1.14 [1.08-1.21] | 1.15 [1.08-1.22] | 1.03 [0.79-1.34] | 1.13 [1.07-1.20] | 3,522 | 3,420 | 3,251 | 165 | 3,312 |
| 722 | 1.30 [1.27-1.33] | 1.29 [1.26-1.32] | 1.31 [1.28-1.35] | 1.18 [1.05-1.32] | 1.19 [1.16-1.22] | 1.18 [1.15-1.21] | 1.12 [1.09-1.14] | 1.13 [1.10-1.16] | 1.11 [0.98-1.25] | 1.12 [1.09-1.15] | 16,846 | 16,747 | 13,505 | 801 | 15,012 |
| 722.1 | 1.36 [1.23-1.49] | 1.33 [1.21-1.47] | 1.36 [1.22-1.50] | | 1.22 [1.10-1.34] | 1.23 [1.11-1.36] | 1.13 [1.02-1.25] | 1.14 [1.02-1.27] | | 1.13 [1.02-1.25] | 1,051 | 1,049 | 910 | NA | 969 |
| 722.3 | 1.26 [0.99-1.59] | 1.27 [1.00-1.61] | 1.22 [0.93-1.61] | | 1.17 [0.92-1.49] | 1.14 [0.89-1.46] | 1.13 [0.88-1.45] | 1.08 [0.81-1.43] | | 1.14 [0.89-1.46] | 179 | 169 | 127 | NA | 164 |
| 722.6 | 1.35 [1.31-1.39] | 1.34 [1.31-1.39] | 1.34 [1.30-1.38] | 1.48 [1.17-1.86] | 1.24 [1.20-1.28] | 1.22 [1.19-1.26] | 1.17 [1.13-1.20] | 1.16 [1.13-1.20] | 1.40 [1.10-1.78] | 1.16 [1.13-1.20] | 11,334 | 11,306 | 10,383 | 207 | 10,584 |
| 722.7 | 1.33 [1.21-1.45] | 1.35 [1.23-1.47] | 1.29 [1.17-1.42] | | 1.25 [1.14-1.38] | 1.24 [1.13-1.36] | 1.19 [1.08-1.31] | 1.14 [1.04-1.26] | | 1.21 [1.10-1.33] | 1,179 | 1,175 | 1,087 | NA | 1,058 |
| 722.8 | 1.40 [1.09-1.80] | 1.46 [1.13-1.88] | 1.46 [1.12-1.90] | | 1.27 [0.99-1.63] | 1.21 [0.93-1.58] | 1.18 [0.90-1.54] | 1.18 [0.89-1.55] | | 1.18 [0.91-1.52] | 155 | 155 | 139 | NA | 150 |
| 722.9 | 1.30 [1.26-1.33] | 1.28 [1.25-1.32] | 1.30 [1.26-1.34] | 1.13 [0.98-1.31] | 1.19 [1.16-1.22] | 1.19 [1.16-1.23] | 1.12 [1.09-1.15] | 1.13 [1.10-1.17] | 1.07 [0.92-1.23] | 1.13 [1.10-1.16] | 13,980 | 13,913 | 11,719 | 508 | 12,608 |
| 723 | 1.29 [1.18-1.41] | 1.31 [1.20-1.43] | 1.27 [1.16-1.39] | | 1.21 [1.11-1.33] | 1.21 [1.10-1.32] | 1.16 [1.06-1.27] | 1.12 [1.02-1.23] | | 1.17 [1.07-1.29] | 1,259 | 1,248 | 1,137 | NA | 1,127 |
| 723.1 | 1.09 [0.98-1.21] | 1.14 [1.00-1.30] | 1.18 [1.01-1.39] | 1.05 [0.89-1.23] | 1.03 [0.92-1.15] | 1.00 [0.90-1.12] | 0.99 [0.87-1.14] | 1.04 [0.88-1.23] | 1.01 [0.86-1.20] | 0.97 [0.86-1.09] | 862 | 542 | 356 | 391 | 759 |
| 724.8 | 1.25 [1.04-1.49] | 1.22 [1.02-1.47] | 1.18 [0.97-1.42] | | 1.18 [0.98-1.42] | 1.15 [0.95-1.39] | 1.08 [0.89-1.31] | 1.04 [0.86-1.27] | | 1.14 [0.95-1.38] | 300 | 296 | 275 | NA | 282 |
| 724.9 | 1.26 [1.20-1.33] | 1.26 [1.20-1.33] | 1.26 [1.19-1.32] | 1.02 [0.78-1.33] | 1.19 [1.13-1.25] | 1.18 [1.12-1.25] | 1.14 [1.08-1.20] | 1.14 [1.08-1.21] | 0.94 [0.71-1.23] | 1.14 [1.08-1.20] | 4,121 | 4,031 | 3,820 | 148 | 3,835 |
| 726 | 1.33 [1.30-1.36] | 1.33 [1.30-1.36] | 1.33 [1.30-1.37] | 1.17 [1.01-1.36] | 1.21 [1.18-1.25] | 1.21 [1.18-1.24] | 1.18 [1.15-1.21] | 1.18 [1.15-1.21] | 1.12 [0.96-1.30] | 1.14 [1.11-1.17] | 17,418 | 17,298 | 15,635 | 462 | 15,207 |
| 726.1 | 1.29 [1.27-1.32] | 1.30 [1.27-1.33] | 1.29 [1.26-1.32] | 1.14 [0.98-1.31] | 1.19 [1.16-1.22] | 1.19 [1.16-1.22] | 1.17 [1.14-1.19] | 1.16 [1.13-1.18] | 1.05 [0.90-1.22] | 1.12 [1.09-1.15] | 19,439 | 19,293 | 17,857 | 489 | 17,313 |
| 726.2 | 1.42 [1.26-1.60] | 1.40 [1.25-1.58] | 1.43 [1.26-1.62] | | 1.26 [1.11-1.42] | 1.27 [1.12-1.43] | 1.22 [1.07-1.38] | 1.22 [1.07-1.39] | | 1.16 [1.02-1.32] | 685 | 685 | 612 | NA | 609 |
| 726.3 | 1.51 [1.41-1.62] | 1.52 [1.41-1.63] | 1.49 [1.38-1.60] | 1.43 [1.09-1.88] | 1.36 [1.26-1.46] | 1.40 [1.30-1.50] | 1.36 [1.26-1.46] | 1.32 [1.22-1.43] | 1.38 [1.04-1.84] | 1.28 [1.19-1.38] | 2,124 | 2,041 | 1,733 | 154 | 1,784 |
| 726.4 | 1.40 [1.14-1.72] | 1.32 [1.08-1.62] | 1.35 [1.09-1.67] | | 1.26 [1.02-1.55] | 1.30 [1.06-1.60] | 1.16 [0.94-1.42] | 1.16 [0.93-1.45] | | 1.19 [0.96-1.48] | 242 | 239 | 207 | NA | 220 |
| 727 | 1.32 [1.24-1.40] | 1.33 [1.25-1.42] | 1.32 [1.23-1.42] | 1.19 [1.00-1.43] | 1.19 [1.12-1.27] | 1.21 [1.13-1.28] | 1.19 [1.11-1.27] | 1.17 [1.08-1.25] | 1.09 [0.91-1.31] | 1.12 [1.05-1.20] | 2,592 | 2,437 | 1,881 | 326 | 2,241 |
| 727.1 | 1.35 [1.31-1.38] | 1.35 [1.31-1.40] | 1.35 [1.30-1.40] | 1.24 [1.16-1.32] | 1.25 [1.21-1.29] | 1.24 [1.21-1.28] | 1.21 [1.17-1.25] | 1.20 [1.16-1.24] | 1.21 [1.13-1.29] | 1.18 [1.14-1.22] | 11,691 | 9,598 | 8,155 | 2,518 | 10,204 |
| 727.2 | 1.51 [1.22-1.87] | 1.47 [1.19-1.82] | 1.46 [1.15-1.85] | | 1.28 [1.02-1.61] | 1.42 [1.14-1.76] | 1.31 [1.05-1.64] | 1.32 [1.03-1.69] | | 1.23 [0.98-1.55] | 224 | 216 | 171 | NA | 188 |
| 727.4 | 1.30 [1.25-1.36] | 1.30 [1.24-1.36] | 1.33 [1.26-1.40] | 1.15 [1.01-1.30] | 1.21 [1.15-1.27] | 1.22 [1.16-1.28] | 1.20 [1.14-1.26] | 1.22 [1.15-1.29] | 1.10 [0.97-1.26] | 1.15 [1.09-1.21] | 4,579 | 4,263 | 3,032 | 650 | 3,641 |
| 727.5 | 1.36 [1.27-1.46] | 1.35 [1.26-1.45] | 1.36 [1.26-1.46] | | 1.30 [1.21-1.40] | 1.27 [1.18-1.36] | 1.24 [1.15-1.33] | 1.23 [1.14-1.32] | | 1.23 [1.14-1.32] | 2,135 | 2,099 | 2,008 | NA | 1,917 |
| 727.6 | 1.23 [1.08-1.39] | 1.24 [1.09-1.40] | 1.21 [1.06-1.38] | | 1.10 [0.96-1.25] | 1.16 [1.02-1.32] | 1.16 [1.02-1.31] | 1.11 [0.96-1.27] | | 1.05 [0.91-1.20] | 605 | 596 | 521 | NA | 508 |
| 727.7 | 1.04 [0.93-1.17] | 1.27 [1.10-1.48] | 1.37 [1.15-1.62] | 0.83 [0.71-0.98] | 0.94 [0.84-1.06] | 0.97 [0.86-1.09] | 1.13 [0.97-1.32] | 1.19 [1.00-1.42] | 0.79 [0.67-0.93] | 0.90 [0.80-1.02] | 768 | 436 | 329 | 373 | 684 |
| 728 | 1.28 [1.16-1.41] | 1.32 [1.19-1.46] | 1.35 [1.22-1.51] | | 1.17 [1.06-1.30] | 1.19 [1.08-1.32] | 1.20 [1.08-1.33] | 1.23 [1.10-1.38] | | 1.14 [1.03-1.26] | 1,065 | 1,014 | 878 | NA | 978 |
| 728.2 | 1.42 [1.35-1.50] | 1.44 [1.35-1.53] | 1.43 [1.30-1.58] | 1.38 [1.27-1.50] | 1.30 [1.23-1.38] | 1.26 [1.19-1.34] | 1.20 [1.12-1.28] | 1.20 [1.09-1.33] | 1.28 [1.18-1.40] | 1.19 [1.12-1.27] | 3,422 | 2,351 | 986 | 1,653 | 2,994 |
| 728.7 | 1.12 [1.07-1.17] | 1.12 [1.07-1.17] | 1.12 [1.07-1.17] | 1.23 [0.91-1.67] | 1.06 [1.01-1.11] | 1.08 [1.03-1.13] | 1.07 [1.02-1.11] | 1.07 [1.02-1.12] | 1.18 [0.85-1.62] | 1.04 [0.99-1.09] | 5,021 | 4,955 | 4,699 | 114 | 4,137 |
| 728.71 | 1.03 [0.98-1.08] | 1.03 [0.98-1.08] | 1.04 [0.99-1.09] | | 0.98 [0.92-1.03] | 1.01 [0.96-1.07] | 1.01 [0.96-1.07] | 1.02 [0.97-1.08] | | 0.97 [0.92-1.03] | 3,475 | 3,470 | 3,427 | NA | 2,770 |
| 729 | 1.40 [1.38-1.43] | 1.41 [1.38-1.44] | 1.42 [1.39-1.45] | 1.22 [1.13-1.32] | 1.30 [1.28-1.33] | 1.31 [1.28-1.33] | 1.26 [1.24-1.29] | 1.27 [1.25-1.30] | 1.19 [1.10-1.28] | 1.23 [1.21-1.26] | 26,684 | 25,680 | 22,682 | 1,788 | 23,758 |
| 729.1 | 1.68 [1.54-1.83] | 1.66 [1.53-1.81] | 1.66 [1.51-1.82] | | 1.48 [1.35-1.62] | 1.38 [1.26-1.51] | 1.29 [1.18-1.42] | 1.30 [1.18-1.44] | | 1.27 [1.16-1.39] | 1,338 | 1,334 | 1,127 | NA | 1,213 |
| 729.3 | 1.38 [1.21-1.57] | 1.37 [1.20-1.57] | 1.42 [1.21-1.67] | 1.21 [0.88-1.67] | 1.28 [1.11-1.47] | 1.26 [1.10-1.44] | 1.22 [1.06-1.40] | 1.25 [1.05-1.48] | 1.11 [0.80-1.55] | 1.20 [1.04-1.38] | 584 | 547 | 360 | 104 | 503 |
| 731 | 1.07 [0.98-1.16] | 1.04 [0.96-1.13] | 1.05 [0.96-1.14] | | 1.02 [0.94-1.11] | 1.06 [0.98-1.16] | 1.03 [0.94-1.12] | 1.03 [0.94-1.12] | | 1.04 [0.96-1.14] | 1,375 | 1,340 | 1,287 | NA | 1,220 |
| 731.1 | 1.16 [1.04-1.29] | 1.14 [1.03-1.28] | 1.17 [1.05-1.31] | | 1.07 [0.95-1.20] | 1.13 [1.01-1.26] | 1.10 [0.99-1.23] | 1.13 [1.01-1.26] | | 1.04 [0.93-1.18] | 890 | 887 | 851 | NA | 738 |
| 732 | 1.27 [1.15-1.40] | 1.30 [1.16-1.46] | 1.31 [1.13-1.51] | 1.17 [1.01-1.36] | 1.17 [1.05-1.30] | 1.17 [1.06-1.29] | 1.13 [1.01-1.28] | 1.16 [1.00-1.34] | 1.10 [0.94-1.28] | 1.10 [0.99-1.23] | 1,066 | 703 | 437 | 473 | 860 |
| 732.1 | 1.31 [1.19-1.43] | 1.36 [1.20-1.53] | 1.38 [1.17-1.63] | 1.22 [1.08-1.38] | 1.22 [1.10-1.35] | 1.21 [1.10-1.33] | 1.20 [1.06-1.36] | 1.24 [1.05-1.47] | 1.14 [1.00-1.29] | 1.13 [1.02-1.25] | 1,218 | 665 | 355 | 690 | 981 |
| 732.7 | 1.37 [1.15-1.64] | 1.25 [1.01-1.55] | | 1.50 [1.16-1.94] | 1.20 [0.98-1.47] | 1.25 [1.04-1.50] | 1.07 [0.86-1.34] | | 1.39 [1.07-1.81] | 1.12 [0.91-1.38] | 319 | 206 | NA | 172 | 247 |
| 733 | 1.31 [1.27-1.34] | 1.32 [1.28-1.35] | 1.32 [1.29-1.36] | 1.18 [1.10-1.27] | 1.22 [1.19-1.26] | 1.20 [1.17-1.23] | 1.17 [1.14-1.20] | 1.18 [1.15-1.22] | 1.13 [1.05-1.22] | 1.16 [1.13-1.19] | 16,105 | 14,991 | 12,630 | 2,032 | 14,549 |
| 733.2 | 1.18 [1.05-1.32] | 1.20 [1.06-1.36] | 1.21 [1.05-1.38] | 0.98 [0.76-1.27] | 1.09 [0.96-1.24] | 1.12 [0.99-1.26] | 1.11 [0.98-1.27] | 1.11 [0.97-1.28] | 0.97 [0.74-1.26] | 1.06 [0.93-1.21] | 699 | 580 | 488 | 150 | 588 |
| 733.4 | 1.28 [1.18-1.39] | 1.27 [1.17-1.38] | 1.28 [1.17-1.39] | 1.22 [0.93-1.59] | 1.16 [1.07-1.26] | 1.21 [1.11-1.31] | 1.16 [1.07-1.26] | 1.16 [1.06-1.27] | 1.16 [0.88-1.53] | 1.13 [1.04-1.23] | 1,542 | 1,451 | 1,285 | 146 | 1,373 |
| 733.6 | 1.41 [1.31-1.52] | 1.42 [1.32-1.54] | 1.50 [1.37-1.64] | 1.20 [1.04-1.40] | 1.31 [1.22-1.42] | 1.26 [1.16-1.36] | 1.19 [1.10-1.29] | 1.24 [1.12-1.37] | 1.17 [1.00-1.37] | 1.22 [1.13-1.31] | 1,967 | 1,708 | 1,170 | 495 | 1,812 |
| 733.8 | 1.24 [1.17-1.30] | 1.25 [1.19-1.32] | 1.28 [1.20-1.37] | 1.15 [1.02-1.29] | 1.15 [1.08-1.21] | 1.16 [1.10-1.22] | 1.13 [1.07-1.20] | 1.16 [1.08-1.24] | 1.11 [0.98-1.25] | 1.12 [1.06-1.19] | 3,449 | 3,079 | 2,264 | 732 | 2,846 |
| 733.9 | 1.33 [1.11-1.60] | 1.37 [1.14-1.64] | 1.44 [1.17-1.77] | | 1.22 [1.00-1.48] | 1.20 [0.99-1.45] | 1.20 [0.99-1.45] | 1.31 [1.06-1.62] | | 1.14 [0.94-1.40] | 288 | 278 | 215 | NA | 243 |
| 735 | 1.29 [1.13-1.48] | 1.42 [1.22-1.66] | 1.43 [1.21-1.70] | 1.01 [0.78-1.32] | 1.17 [1.02-1.35] | 1.21 [1.05-1.39] | 1.29 [1.10-1.51] | 1.31 [1.10-1.56] | 0.98 [0.75-1.29] | 1.13 [0.98-1.30] | 539 | 419 | 334 | 150 | 485 |
| 735.1 | 1.33 [1.17-1.50] | 1.41 [1.22-1.63] | 1.39 [1.19-1.63] | 1.18 [0.94-1.48] | 1.28 [1.12-1.46] | 1.20 [1.05-1.36] | 1.25 [1.08-1.45] | 1.22 [1.03-1.43] | 1.09 [0.86-1.38] | 1.19 [1.04-1.36] | 658 | 457 | 379 | 228 | 598 |
| 735.2 | 1.42 [1.34-1.50] | 1.42 [1.34-1.50] | 1.42 [1.33-1.51] | 1.21 [1.04-1.40] | 1.27 [1.20-1.35] | 1.32 [1.24-1.39] | 1.29 [1.22-1.37] | 1.28 [1.21-1.37] | 1.15 [0.99-1.34] | 1.20 [1.13-1.28] | 3,176 | 2,826 | 2,467 | 467 | 2,731 |
| 735.21 | 1.37 [1.29-1.46] | 1.36 [1.28-1.45] | 1.35 [1.26-1.44] | 1.34 [1.00-1.79] | 1.26 [1.18-1.34] | 1.27 [1.19-1.35] | 1.23 [1.16-1.31] | 1.23 [1.15-1.31] | 1.31 [0.97-1.77] | 1.17 [1.10-1.26] | 2,492 | 2,411 | 2,256 | 127 | 2,165 |
| 735.23 | 1.22 [1.12-1.32] | 1.26 [1.17-1.37] | 1.26 [1.16-1.37] | | 1.17 [1.08-1.28] | 1.13 [1.05-1.23] | 1.15 [1.06-1.25] | 1.16 [1.06-1.26] | | 1.11 [1.01-1.21] | 1,423 | 1,419 | 1,339 | NA | 1,217 |
| 735.3 | 1.28 [1.23-1.33] | 1.27 [1.22-1.32] | 1.27 [1.22-1.32] | 1.20 [1.02-1.40] | 1.18 [1.13-1.23] | 1.21 [1.16-1.25] | 1.18 [1.13-1.23] | 1.18 [1.14-1.23] | 1.16 [0.99-1.36] | 1.13 [1.08-1.18] | 6,813 | 6,626 | 5,717 | 433 | 5,659 |
| 736 | 1.20 [1.15-1.24] | 1.24 [1.19-1.30] | 1.23 [1.18-1.29] | 1.03 [0.95-1.12] | 1.12 [1.07-1.16] | 1.12 [1.08-1.17] | 1.14 [1.09-1.19] | 1.13 [1.08-1.19] | 0.99 [0.91-1.08] | 1.08 [1.03-1.12] | 6,788 | 5,505 | 4,784 | 1,559 | 6,095 |
| 736.2 | 1.17 [1.02-1.34] | 1.23 [1.06-1.42] | 1.19 [1.00-1.40] | 1.05 [0.77-1.43] | 1.07 [0.92-1.24] | 1.12 [0.97-1.29] | 1.17 [1.01-1.35] | 1.14 [0.96-1.36] | 0.98 [0.72-1.35] | 1.03 [0.89-1.20] | 510 | 442 | 326 | 107 | 419 |
| 737 | 1.24 [1.07-1.45] | 1.27 [1.08-1.50] | 1.36 [1.13-1.63] | | 1.16 [0.99-1.36] | 1.16 [0.99-1.35] | 1.14 [0.96-1.35] | 1.25 [1.03-1.51] | | 1.11 [0.95-1.30] | 444 | 370 | 302 | NA | 408 |
| 737.1 | 1.24 [1.16-1.32] | 1.22 [1.15-1.31] | 1.23 [1.15-1.31] | 1.10 [0.83-1.45] | 1.18 [1.10-1.25] | 1.17 [1.10-1.25] | 1.13 [1.05-1.20] | 1.14 [1.07-1.22] | 1.02 [0.77-1.35] | 1.13 [1.06-1.21] | 2,707 | 2,616 | 2,513 | 136 | 2,564 |
| 737.2 | 1.41 [1.18-1.69] | 1.41 [1.17-1.70] | 1.41 [1.15-1.73] | | 1.23 [1.02-1.47] | 1.31 [1.09-1.57] | 1.24 [1.02-1.51] | 1.27 [1.03-1.57] | | 1.15 [0.95-1.38] | 332 | 303 | 256 | NA | 303 |
| 737.3 | 1.24 [1.20-1.28] | 1.27 [1.23-1.32] | 1.27 [1.22-1.32] | 1.07 [0.99-1.15] | 1.14 [1.10-1.18] | 1.16 [1.12-1.20] | 1.16 [1.12-1.21] | 1.16 [1.11-1.21] | 1.03 [0.96-1.12] | 1.09 [1.05-1.13] | 8,868 | 7,537 | 6,538 | 1,747 | 8,022 |
| 738 | 1.34 [1.22-1.48] | 1.32 [1.20-1.46] | 1.29 [1.13-1.47] | 1.21 [1.00-1.47] | 1.18 [1.06-1.31] | 1.24 [1.12-1.37] | 1.16 [1.04-1.29] | 1.09 [0.95-1.25] | 1.19 [0.98-1.45] | 1.12 [1.01-1.24] | 1,092 | 944 | 573 | 293 | 924 |
| 738.4 | 1.32 [1.26-1.37] | 1.31 [1.25-1.37] | 1.30 [1.24-1.36] | 1.06 [0.79-1.42] | 1.20 [1.15-1.25] | 1.21 [1.16-1.27] | 1.15 [1.10-1.21] | 1.14 [1.09-1.19] | 0.99 [0.73-1.34] | 1.12 [1.07-1.17] | 5,500 | 5,432 | 5,196 | 123 | 5,084 |
| 739 | 1.16 [1.07-1.26] | 1.27 [1.16-1.39] | 1.29 [1.18-1.43] | 0.89 [0.74-1.06] | 1.09 [1.00-1.19] | 1.10 [1.01-1.20] | 1.18 [1.08-1.29] | 1.20 [1.09-1.33] | 0.88 [0.74-1.06] | 1.06 [0.98-1.16] | 1,464 | 1,226 | 1,038 | 308 | 1,323 |
| 740.1 | 1.14 [1.12-1.17] | 1.14 [1.12-1.16] | 1.13 [1.11-1.16] | 1.11 [0.84-1.48] | 1.09 [1.07-1.11] | 1.10 [1.08-1.12] | 1.08 [1.06-1.10] | 1.07 [1.05-1.09] | 1.07 [0.80-1.43] | 1.06 [1.04-1.08] | 27,965 | 27,940 | 27,430 | 132 | 24,100 |
| 740.11 | 1.19 [1.15-1.23] | 1.18 [1.15-1.22] | 1.18 [1.15-1.22] | | 1.11 [1.07-1.15] | 1.14 [1.10-1.17] | 1.12 [1.08-1.15] | 1.11 [1.08-1.15] | | 1.08 [1.04-1.11] | 9,407 | 9,401 | 9,174 | NA | 7,603 |
| 740.12 | 1.31 [1.17-1.47] | 1.29 [1.15-1.45] | 1.33 [1.18-1.51] | | 1.12 [0.99-1.26] | 1.23 [1.09-1.38] | 1.20 [1.07-1.35] | 1.25 [1.11-1.42] | | 1.08 [0.96-1.22] | 727 | 717 | 616 | NA | 637 |
| 740.2 | 1.34 [1.23-1.46] | 1.35 [1.24-1.48] | 1.32 [1.21-1.44] | | 1.22 [1.12-1.34] | 1.23 [1.13-1.35] | 1.22 [1.11-1.33] | 1.19 [1.08-1.30] | | 1.15 [1.05-1.26] | 1,275 | 1,275 | 1,262 | NA | 1,159 |
| 740.9 | 1.34 [1.32-1.36] | 1.33 [1.31-1.35] | 1.33 [1.31-1.35] | 1.30 [1.12-1.51] | 1.25 [1.23-1.27] | 1.24 [1.22-1.25] | 1.19 [1.17-1.21] | 1.19 [1.17-1.21] | 1.21 [1.04-1.41] | 1.18 [1.16-1.20] | 48,612 | 48,412 | 47,100 | 490 | 44,277 |
| 741 | 1.37 [1.30-1.44] | 1.39 [1.32-1.47] | 1.35 [1.26-1.44] | 1.21 [1.06-1.37] | 1.25 [1.18-1.32] | 1.25 [1.19-1.32] | 1.24 [1.17-1.31] | 1.18 [1.11-1.27] | 1.15 [1.00-1.31] | 1.18 [1.11-1.25] | 3,589 | 3,312 | 2,302 | 621 | 3,109 |
| 741.1 | 1.15 [0.95-1.40] | 1.20 [0.98-1.46] | 1.10 [0.88-1.37] | | 1.07 [0.88-1.31] | 1.06 [0.87-1.30] | 1.11 [0.90-1.36] | 0.98 [0.78-1.24] | | 1.02 [0.84-1.25] | 260 | 240 | 187 | NA | 237 |
| 741.2 | 1.33 [1.22-1.44] | 1.35 [1.23-1.47] | 1.35 [1.22-1.48] | 1.16 [0.95-1.41] | 1.26 [1.15-1.37] | 1.24 [1.14-1.35] | 1.23 [1.12-1.34] | 1.23 [1.12-1.36] | 1.11 [0.90-1.36] | 1.20 [1.10-1.31] | 1,473 | 1,291 | 1,032 | 273 | 1,313 |
| 741.3 | 1.23 [1.17-1.29] | 1.27 [1.20-1.33] | 1.25 [1.18-1.31] | 0.98 [0.84-1.14] | 1.17 [1.12-1.23] | 1.17 [1.11-1.22] | 1.15 [1.09-1.21] | 1.13 [1.08-1.19] | 0.95 [0.82-1.12] | 1.14 [1.08-1.20] | 4,840 | 4,451 | 4,353 | 415 | 4,521 |
| 741.4 | 1.29 [1.25-1.34] | 1.31 [1.26-1.36] | 1.30 [1.25-1.35] | 1.17 [1.07-1.28] | 1.20 [1.16-1.24] | 1.21 [1.16-1.25] | 1.18 [1.14-1.23] | 1.18 [1.13-1.23] | 1.12 [1.03-1.23] | 1.15 [1.11-1.20] | 8,354 | 7,184 | 6,528 | 1,383 | 7,485 |
| 741.5 | 1.29 [1.14-1.45] | 1.37 [1.21-1.55] | 1.43 [1.25-1.62] | | 1.26 [1.11-1.43] | 1.22 [1.08-1.38] | 1.25 [1.10-1.43] | 1.28 [1.12-1.47] | | 1.21 [1.06-1.37] | 712 | 683 | 641 | NA | 659 |
| 741.6 | 1.19 [0.88-1.59] | | | | | 1.10 [0.81-1.49] | | | | | 112 | NA | NA | NA | NA |
| 742 | 1.06 [0.82-1.35] | 1.08 [0.84-1.40] | 1.14 [0.88-1.47] | | 0.99 [0.76-1.29] | 1.03 [0.80-1.33] | 1.03 [0.79-1.33] | 1.11 [0.85-1.43] | | 0.98 [0.75-1.27] | 150 | 149 | 143 | NA | 135 |
| 742.1 | 1.38 [1.22-1.57] | 1.32 [1.16-1.50] | 1.40 [1.20-1.64] | 1.39 [1.04-1.85] | 1.19 [1.04-1.37] | 1.31 [1.15-1.49] | 1.20 [1.05-1.37] | 1.26 [1.08-1.49] | 1.39 [1.03-1.87] | 1.15 [1.00-1.32] | 607 | 559 | 382 | 118 | 490 |
| 742.2 | 1.39 [1.21-1.58] | 1.45 [1.25-1.69] | 1.52 [1.13-2.04] | 1.28 [1.07-1.53] | 1.28 [1.11-1.49] | 1.25 [1.09-1.43] | 1.26 [1.07-1.47] | 1.33 [0.98-1.81] | 1.18 [0.98-1.42] | 1.19 [1.02-1.38] | 599 | 429 | 111 | 333 | 467 |
| 742.8 | 1.33 [1.25-1.40] | 1.30 [1.23-1.38] | 1.31 [1.22-1.41] | 1.27 [1.11-1.46] | 1.19 [1.12-1.26] | 1.21 [1.14-1.29] | 1.15 [1.08-1.22] | 1.15 [1.07-1.24] | 1.20 [1.04-1.38] | 1.12 [1.06-1.19] | 3,081 | 2,901 | 1,818 | 588 | 2,634 |
| 742.9 | 1.26 [1.22-1.31] | 1.29 [1.24-1.33] | 1.30 [1.24-1.36] | 1.13 [1.05-1.21] | 1.15 [1.11-1.20] | 1.16 [1.12-1.20] | 1.14 [1.10-1.18] | 1.14 [1.09-1.20] | 1.07 [0.99-1.15] | 1.09 [1.05-1.13] | 8,330 | 7,532 | 4,477 | 2,078 | 6,999 |
| 743.1 | 1.28 [1.23-1.34] | 1.29 [1.24-1.36] | 1.30 [1.24-1.36] | | 1.22 [1.17-1.28] | 1.21 [1.16-1.27] | 1.18 [1.13-1.24] | 1.19 [1.13-1.25] | | 1.17 [1.12-1.23] | 4,996 | 4,983 | 4,950 | NA | 4,693 |
| 743.11 | 1.30 [1.28-1.33] | 1.30 [1.28-1.32] | 1.30 [1.28-1.32] | 1.38 [1.07-1.78] | 1.23 [1.20-1.25] | 1.22 [1.20-1.24] | 1.17 [1.15-1.19] | 1.17 [1.15-1.20] | 1.30 [1.00-1.69] | 1.16 [1.14-1.18] | 35,604 | 35,547 | 35,063 | 173 | 32,807 |
| 743.12 | 1.33 [1.07-1.66] | 1.37 [1.10-1.71] | 1.39 [1.11-1.73] | | 1.38 [1.10-1.73] | 1.27 [1.02-1.59] | 1.25 [0.99-1.56] | 1.27 [1.01-1.60] | | 1.29 [1.02-1.62] | 213 | 212 | 212 | NA | 204 |
| 743.13 | 1.40 [1.27-1.56] | 1.42 [1.27-1.57] | 1.38 [1.24-1.53] | | 1.30 [1.17-1.45] | 1.28 [1.15-1.43] | 1.24 [1.11-1.38] | 1.23 [1.10-1.37] | | 1.20 [1.07-1.34] | 934 | 908 | 848 | NA | 812 |
| 743.2 | 1.22 [1.12-1.32] | 1.26 [1.15-1.37] | 1.24 [1.14-1.35] | | 1.16 [1.07-1.26] | 1.17 [1.08-1.27] | 1.16 [1.07-1.27] | 1.16 [1.07-1.27] | | 1.14 [1.05-1.24] | 1,509 | 1,434 | 1,396 | NA | 1,370 |
| 743.21 | 1.22 [1.16-1.29] | 1.24 [1.17-1.30] | 1.24 [1.17-1.31] | | 1.16 [1.10-1.22] | 1.15 [1.09-1.22] | 1.13 [1.07-1.19] | 1.13 [1.07-1.19] | | 1.11 [1.05-1.18] | 3,940 | 3,922 | 3,867 | NA | 3,706 |
| 743.4 | 1.39 [1.13-1.71] | 1.37 [1.11-1.69] | 1.34 [1.09-1.65] | | 1.18 [0.95-1.45] | 1.32 [1.07-1.62] | 1.25 [1.01-1.55] | 1.26 [1.02-1.56] | | 1.14 [0.92-1.40] | 262 | 252 | 246 | NA | 238 |
| 743.9 | 1.27 [1.19-1.36] | 1.29 [1.20-1.39] | 1.28 [1.18-1.38] | 1.14 [0.98-1.32] | 1.18 [1.10-1.27] | 1.19 [1.11-1.28] | 1.16 [1.07-1.25] | 1.15 [1.06-1.24] | 1.10 [0.94-1.28] | 1.13 [1.05-1.22] | 2,210 | 1,849 | 1,526 | 459 | 1,919 |
| 745 | 1.30 [1.28-1.33] | 1.31 [1.28-1.33] | 1.30 [1.27-1.32] | 1.22 [1.17-1.28] | 1.21 [1.19-1.23] | 1.21 [1.19-1.23] | 1.16 [1.14-1.18] | 1.15 [1.13-1.18] | 1.17 [1.11-1.22] | 1.15 [1.13-1.17] | 36,720 | 32,923 | 28,365 | 5,197 | 33,207 |
| congenital anomalies | | | | | | | | | | | | | | | |
| 747 | 1.31 [1.12-1.54] | 1.46 [1.23-1.74] | 1.36 [1.11-1.66] | 0.96 [0.70-1.31] | 1.17 [0.98-1.39] | 1.26 [1.07-1.48] | 1.36 [1.13-1.62] | 1.25 [1.01-1.54] | 0.95 [0.69-1.32] | 1.16 [0.97-1.38] | 397 | 328 | 237 | 102 | 319 |
| 747.1 | 1.20 [1.12-1.28] | 1.25 [1.16-1.36] | 1.24 [1.13-1.37] | 1.04 [0.93-1.16] | 1.10 [1.03-1.18] | 1.14 [1.06-1.22] | 1.14 [1.05-1.24] | 1.14 [1.04-1.26] | 1.00 [0.89-1.12] | 1.06 [0.98-1.14] | 2,218 | 1,555 | 1,027 | 854 | 1,912 |
| 747.11 | 1.15 [1.08-1.23] | 1.23 [1.14-1.33] | 1.27 [1.16-1.39] | 1.00 [0.90-1.10] | 1.11 [1.04-1.19] | 1.10 [1.03-1.17] | 1.14 [1.05-1.24] | 1.16 [1.05-1.27] | 0.97 [0.87-1.07] | 1.06 [0.99-1.14] | 2,389 | 1,558 | 1,130 | 961 | 2,019 |
| 747.12 | 1.11 [1.02-1.22] | 1.16 [1.04-1.30] | 1.14 [1.00-1.29] | 1.01 [0.88-1.17] | 1.02 [0.93-1.13] | 1.09 [1.00-1.20] | 1.10 [0.98-1.23] | 1.09 [0.96-1.24] | 1.00 [0.86-1.15] | 1.01 [0.91-1.12] | 1,159 | 764 | 576 | 465 | 981 |
| 747.13 | 1.23 [1.20-1.26] | 1.24 [1.20-1.27] | 1.24 [1.21-1.28] | 0.98 [0.87-1.12] | 1.17 [1.14-1.21] | 1.18 [1.15-1.21] | 1.14 [1.11-1.17] | 1.15 [1.12-1.18] | 0.95 [0.84-1.08] | 1.14 [1.10-1.17] | 15,224 | 14,686 | 14,457 | 616 | 13,912 |
| 747.2 | 1.18 [1.06-1.30] | 1.31 [1.15-1.48] | 1.28 [1.10-1.48] | 1.02 [0.87-1.21] | 1.10 [0.99-1.23] | 1.13 [1.02-1.26] | 1.23 [1.08-1.39] | 1.18 [1.01-1.37] | 1.01 [0.86-1.19] | 1.09 [0.98-1.22] | 942 | 624 | 441 | 391 | 823 |
| 748 | 1.05 [0.92-1.21] | 1.30 [1.02-1.64] | 1.46 [1.12-1.90] | 0.98 [0.83-1.16] | 0.98 [0.85-1.13] | 0.94 [0.81-1.08] | 1.09 [0.85-1.40] | 1.27 [0.97-1.67] | 0.86 [0.73-1.02] | 0.88 [0.76-1.02] | 491 | 168 | 143 | 335 | 454 |
| 749 | 1.13 [1.07-1.19] | 1.18 [1.04-1.34] | 1.35 [1.13-1.61] | 1.09 [1.03-1.16] | 1.09 [1.02-1.16] | 1.10 [1.04-1.17] | 1.08 [0.95-1.23] | 1.25 [1.04-1.50] | 1.07 [1.01-1.14] | 1.08 [1.01-1.15] | 3,107 | 593 | 284 | 2,653 | 2,401 |
| 749.1 | 1.03 [0.87-1.21] | | | 1.02 [0.85-1.22] | 0.98 [0.82-1.18] | 0.98 [0.83-1.16] | | | 0.96 [0.80-1.16] | 0.96 [0.79-1.15] | 348 | NA | NA | 286 | 292 |
| 749.2 | 1.06 [0.93-1.21] | | | 1.00 [0.87-1.15] | 1.00 [0.87-1.14] | 1.01 [0.89-1.16] | | | 0.97 [0.84-1.12] | 0.98 [0.85-1.12] | 560 | NA | NA | 490 | 501 |
| 750 | 1.34 [1.02-1.76] | 1.36 [1.01-1.83] | | | 1.37 [1.03-1.81] | 1.24 [0.93-1.64] | 1.13 [0.82-1.54] | | | 1.29 [0.97-1.72] | 135 | 111 | NA | NA | 126 |
| 750.1 | 1.19 [0.99-1.42] | 1.18 [0.96-1.45] | 1.22 [0.98-1.51] | | 1.13 [0.94-1.36] | 1.09 [0.90-1.31] | 1.02 [0.82-1.26] | 1.07 [0.85-1.34] | | 1.06 [0.88-1.29] | 294 | 224 | 200 | NA | 267 |
| 750.11 | 1.42 [1.08-1.87] | 1.53 [1.14-2.07] | 1.54 [1.13-2.10] | | 1.45 [1.09-1.94] | 1.31 [0.99-1.73] | 1.40 [1.03-1.90] | 1.39 [1.02-1.91] | | 1.38 [1.03-1.85] | 134 | 114 | 108 | NA | 121 |
| 750.13 | 1.14 [1.07-1.22] | 1.15 [1.02-1.31] | 1.18 [1.04-1.35] | 1.11 [1.03-1.20] | 1.10 [1.02-1.18] | 1.11 [1.04-1.18] | 1.08 [0.95-1.23] | 1.09 [0.95-1.25] | 1.10 [1.02-1.18] | 1.07 [1.00-1.16] | 2,319 | 657 | 591 | 1,694 | 1,931 |
| 750.14 | 1.27 [1.08-1.49] | 1.30 [1.09-1.53] | 1.29 [1.08-1.53] | | 1.11 [0.93-1.31] | 1.17 [0.99-1.39] | 1.16 [0.98-1.38] | 1.16 [0.97-1.39] | | 1.03 [0.87-1.23] | 376 | 352 | 326 | NA | 334 |
| 750.15 | 1.26 [1.00-1.59] | 1.18 [0.93-1.50] | 1.17 [0.91-1.49] | | 1.22 [0.96-1.55] | 1.12 [0.88-1.42] | 1.00 [0.78-1.28] | 1.01 [0.78-1.30] | | 1.13 [0.88-1.44] | 178 | 168 | 152 | NA | 168 |
| 750.21 | 1.18 [1.07-1.31] | 1.22 [1.09-1.37] | 1.21 [1.06-1.39] | 1.01 [0.85-1.21] | 1.14 [1.02-1.26] | 1.11 [1.00-1.23] | 1.09 [0.97-1.23] | 1.10 [0.96-1.26] | 0.99 [0.82-1.19] | 1.07 [0.96-1.20] | 970 | 742 | 530 | 303 | 832 |
| 750.22 | 1.20 [1.02-1.41] | 1.23 [1.03-1.45] | 1.14 [0.94-1.38] | | 1.15 [0.97-1.36] | 1.13 [0.96-1.34] | 1.09 [0.92-1.30] | 1.02 [0.84-1.25] | | 1.10 [0.93-1.31] | 381 | 328 | 256 | NA | 338 |
| 751.1 | 1.13 [1.06-1.21] | 1.22 [0.96-1.54] | | 1.11 [1.03-1.19] | 1.13 [1.04-1.21] | 1.10 [1.03-1.19] | 1.11 [0.87-1.42] | | 1.08 [1.00-1.16] | 1.10 [1.02-1.19] | 2,131 | 164 | NA | 2,016 | 1,782 |
| 751.11 | 1.17 [1.08-1.27] | 1.20 [1.10-1.30] | 1.21 [1.06-1.37] | 1.23 [1.07-1.41] | 1.13 [1.03-1.23] | 1.11 [1.02-1.20] | 1.10 [1.01-1.20] | 1.10 [0.97-1.26] | 1.16 [1.00-1.34] | 1.09 [1.00-1.19] | 1,604 | 1,319 | 564 | 541 | 1,301 |
| 751.12 | 1.03 [0.98-1.09] | 1.22 [1.06-1.41] | 1.17 [0.99-1.39] | 1.01 [0.96-1.07] | 0.99 [0.93-1.04] | 1.02 [0.97-1.07] | 1.14 [0.98-1.33] | 1.12 [0.94-1.33] | 0.99 [0.94-1.05] | 0.98 [0.93-1.04] | 3,929 | 455 | 327 | 3,525 | 3,175 |
| 751.2 | 1.12 [0.97-1.28] | 1.22 [0.99-1.52] | 1.22 [0.94-1.59] | 1.03 [0.87-1.23] | 1.04 [0.89-1.20] | 1.08 [0.93-1.24] | 1.16 [0.93-1.44] | 1.15 [0.88-1.51] | 1.01 [0.84-1.21] | 1.02 [0.88-1.19] | 485 | 199 | 133 | 316 | 427 |
| 751.21 | 1.16 [1.07-1.26] | 1.14 [1.05-1.24] | 1.19 [1.08-1.30] | 1.05 [0.86-1.29] | 1.07 [0.99-1.17] | 1.11 [1.03-1.21] | 1.04 [0.95-1.14] | 1.10 [1.01-1.21] | 1.00 [0.82-1.23] | 1.05 [0.96-1.15] | 1,502 | 1,317 | 1,141 | 239 | 1,307 |
| 751.22 | 1.16 [1.08-1.25] | 1.18 [1.08-1.28] | 1.14 [1.03-1.26] | 1.09 [0.96-1.24] | 1.06 [0.98-1.15] | 1.10 [1.02-1.19] | 1.06 [0.97-1.15] | 1.03 [0.93-1.14] | 1.07 [0.94-1.21] | 1.02 [0.94-1.11] | 1,806 | 1,332 | 944 | 630 | 1,582 |
| 751.3 | 1.00 [0.85-1.18] | 1.01 [0.77-1.31] | | 0.99 [0.81-1.21] | 0.96 [0.81-1.15] | 0.98 [0.83-1.16] | 0.98 [0.75-1.29] | | 0.97 [0.79-1.19] | 0.96 [0.80-1.15] | 338 | 130 | NA | 228 | 300 |
| 752.1 | 1.29 [1.06-1.56] | 1.28 [1.03-1.59] | 1.46 [1.11-1.92] | | 1.17 [0.96-1.43] | 1.18 [0.97-1.44] | 1.08 [0.86-1.36] | 1.18 [0.88-1.57] | | 1.11 [0.91-1.36] | 263 | 198 | 127 | NA | 234 |
| 752.11 | 1.30 [1.17-1.44] | 1.35 [1.20-1.52] | 1.33 [1.14-1.55] | 1.12 [0.95-1.32] | 1.19 [1.07-1.33] | 1.18 [1.06-1.31] | 1.18 [1.04-1.33] | 1.15 [0.98-1.35] | 1.04 [0.88-1.24] | 1.11 [0.99-1.23] | 964 | 722 | 395 | 361 | 829 |
| 752.2 | 1.14 [1.05-1.23] | 1.35 [1.20-1.52] | 1.38 [1.17-1.63] | 1.04 [0.94-1.14] | 1.07 [0.99-1.16] | 1.06 [0.98-1.15] | 1.17 [1.04-1.33] | 1.18 [0.99-1.40] | 1.00 [0.91-1.10] | 1.02 [0.94-1.11] | 1,632 | 710 | 345 | 1,073 | 1,471 |
| 753 | 1.18 [0.96-1.45] | 1.28 [0.96-1.70] | | 1.05 [0.79-1.40] | 1.11 [0.89-1.39] | 1.11 [0.90-1.37] | 1.22 [0.91-1.63] | | 0.98 [0.73-1.31] | 1.08 [0.86-1.35] | 223 | 119 | NA | 117 | 193 |
| 753.1 | 1.22 [1.02-1.46] | 1.26 [1.00-1.58] | 1.23 [0.93-1.62] | 1.25 [0.97-1.63] | 1.16 [0.95-1.40] | 1.17 [0.98-1.41] | 1.19 [0.94-1.51] | 1.18 [0.88-1.56] | 1.21 [0.93-1.57] | 1.13 [0.93-1.38] | 313 | 185 | 123 | 150 | 261 |
| 753.2 | 1.11 [0.86-1.44] | | | | 1.08 [0.82-1.42] | 1.07 [0.82-1.39] | | | | 1.08 [0.82-1.43] | 142 | NA | NA | NA | 126 |
| 754 | 1.22 [1.07-1.40] | 1.33 [1.13-1.57] | 1.41 [1.16-1.72] | 1.04 [0.84-1.28] | 1.12 [0.97-1.29] | 1.14 [0.99-1.31] | 1.16 [0.98-1.38] | 1.22 [0.99-1.50] | 0.97 [0.78-1.21] | 1.07 [0.93-1.24] | 544 | 351 | 240 | 233 | 480 |
| 755.1 | 1.26 [1.15-1.37] | 1.37 [1.20-1.57] | 1.60 [1.36-1.89] | 1.12 [1.01-1.25] | 1.12 [1.02-1.23] | 1.19 [1.09-1.30] | 1.25 [1.09-1.44] | 1.44 [1.21-1.71] | 1.08 [0.97-1.21] | 1.09 [0.99-1.20] | 1,329 | 528 | 363 | 872 | 1,112 |
| 755.3 | 1.02 [0.87-1.20] | | | 0.95 [0.80-1.13] | 0.94 [0.78-1.12] | 0.98 [0.83-1.16] | | | 0.94 [0.78-1.12] | 0.94 [0.78-1.12] | 366 | NA | NA | 297 | 286 |
| 755.4 | 1.22 [1.08-1.38] | 1.16 [0.91-1.46] | 1.32 [0.98-1.78] | 1.21 [1.05-1.40] | 1.06 [0.92-1.22] | 1.19 [1.05-1.36] | 1.08 [0.85-1.38] | 1.20 [0.88-1.63] | 1.20 [1.04-1.38] | 1.05 [0.91-1.21] | 638 | 174 | 109 | 495 | 523 |

### Supplementary Table 6: phecode-mapped results (hazard ratios and events from all cohorts)

| | Hazard ratio (99% confidence interval) | | | | | | | | | | | | | | |
|---|---|---|---|---|---|---|---|---|---|---|---|---|---|---|---|
| | crude | | | | | adjusted | | | | | Events (in exposed) | | | | |
| Outcome | any age | 18+ | 40+ | <18 | hosp. | any age | 18+ | 40+ | <18 | hosp. | any age | 18+ | 40+ | <18 | hosp. |
| 755.6 | 1.21 [1.05-1.38] | 1.26 [1.03-1.54] | 1.33 [1.01-1.75] | 1.10 [0.93-1.31] | 1.08 [0.93-1.25] | 1.15 [1.00-1.33] | 1.14 [0.93-1.40] | 1.18 [0.89-1.58] | 1.07 [0.90-1.27] | 1.04 [0.89-1.21] | 566 | 241 | 124 | 381 | 476 |
| 755.61 | 1.04 [0.94-1.15] | 1.17 [1.02-1.34] | 1.18 [0.97-1.43] | 0.89 [0.78-1.02] | 0.93 [0.84-1.04] | 0.99 [0.89-1.09] | 1.07 [0.93-1.23] | 1.08 [0.88-1.32] | 0.86 [0.75-0.99] | 0.91 [0.82-1.02] | 947 | 502 | 238 | 525 | 815 |
| 756 | 1.20 [1.05-1.36] | 1.20 [0.98-1.47] | 1.31 [1.00-1.72] | 1.12 [0.96-1.32] | 1.03 [0.89-1.19] | 1.12 [0.98-1.29] | 1.06 [0.86-1.31] | 1.16 [0.87-1.54] | 1.08 [0.92-1.28] | 0.99 [0.85-1.14] | 569 | 228 | 125 | 386 | 476 |
| 756.1 | 1.00 [0.75-1.33] | | | | | 0.90 [0.67-1.21] | | | | | 114 | NA | NA | NA | NA |
| 756.21 | 1.26 [1.04-1.54] | 1.27 [0.96-1.68] | | 1.19 [0.94-1.52] | 1.24 [1.00-1.54] | 1.24 [1.02-1.52] | 1.26 [0.94-1.69] | | 1.15 [0.90-1.46] | 1.23 [0.99-1.53] | 267 | 119 | NA | 183 | 223 |
| 756.3 | 1.43 [1.28-1.61] | 1.39 [1.21-1.59] | 1.24 [0.98-1.56] | 1.32 [1.12-1.56] | 1.25 [1.10-1.41] | 1.18 [1.04-1.33] | 1.01 [0.87-1.17] | 0.89 [0.69-1.14] | 1.19 [1.00-1.42] | 1.07 [0.94-1.22] | 797 | 540 | 173 | 411 | 697 |
| 756.5 | 1.27 [1.18-1.37] | 1.31 [1.21-1.42] | 1.32 [1.20-1.44] | 1.07 [0.91-1.25] | 1.15 [1.06-1.24] | 1.19 [1.10-1.28] | 1.20 [1.10-1.30] | 1.17 [1.07-1.29] | 1.03 [0.88-1.21] | 1.11 [1.02-1.20] | 1,865 | 1,550 | 1,251 | 417 | 1,651 |
| 757 | 1.36 [1.22-1.51] | 1.77 [1.50-2.10] | 1.81 [1.49-2.20] | 1.20 [1.04-1.38] | 1.32 [1.18-1.49] | 1.28 [1.15-1.43] | 1.59 [1.34-1.89] | 1.58 [1.29-1.93] | 1.16 [1.00-1.33] | 1.27 [1.12-1.43] | 857 | 373 | 279 | 519 | 718 |
| 758 | 1.28 [1.10-1.49] | 1.42 [1.16-1.73] | 1.50 [1.15-1.96] | 1.05 [0.85-1.29] | 1.12 [0.95-1.31] | 1.21 [1.04-1.42] | 1.26 [1.02-1.54] | 1.36 [1.03-1.79] | 1.04 [0.84-1.28] | 1.09 [0.92-1.29] | 444 | 247 | 138 | 235 | 374 |
| 758.1 | 1.71 [1.59-1.84] | 2.48 [2.24-2.74] | 2.96 [2.61-3.35] | 1.13 [1.02-1.25] | 1.38 [1.27-1.51] | 1.62 [1.50-1.75] | 2.39 [2.15-2.65] | 2.81 [2.48-3.20] | 1.08 [0.97-1.20] | 1.31 [1.20-1.43] | 2,051 | 1,166 | 762 | 983 | 1,424 |
| 759 | 1.13 [1.01-1.26] | 1.26 [1.08-1.47] | 1.36 [1.13-1.64] | 1.05 [0.91-1.22] | 1.08 [0.96-1.21] | 1.06 [0.95-1.19] | 1.14 [0.97-1.34] | 1.25 [1.03-1.52] | 1.00 [0.86-1.17] | 1.04 [0.92-1.17] | 797 | 413 | 265 | 454 | 724 |
| 759.1 | 1.06 [0.92-1.22] | 1.11 [0.88-1.38] | 1.02 [0.77-1.36] | 1.04 [0.87-1.24] | 1.04 [0.89-1.21] | 1.04 [0.90-1.20] | 1.03 [0.82-1.29] | 0.99 [0.74-1.32] | 1.03 [0.86-1.23] | 1.03 [0.88-1.21] | 481 | 185 | 108 | 325 | 399 |
| symptoms | | | | | | | | | | | | | | | |
| 760 | 1.30 [1.28-1.32] | 1.30 [1.28-1.32] | 1.30 [1.28-1.32] | 1.15 [1.10-1.19] | 1.19 [1.17-1.20] | 1.19 [1.17-1.21] | 1.13 [1.12-1.15] | 1.14 [1.12-1.16] | 1.09 [1.04-1.14] | 1.12 [1.11-1.14] | 60,233 | 57,895 | 46,321 | 5,857 | 53,676 |
| 761 | 1.34 [1.30-1.39] | 1.33 [1.28-1.38] | 1.36 [1.30-1.41] | 1.19 [1.07-1.31] | 1.22 [1.17-1.26] | 1.21 [1.16-1.25] | 1.13 [1.09-1.18] | 1.15 [1.11-1.20] | 1.12 [1.01-1.25] | 1.13 [1.09-1.17] | 7,994 | 7,406 | 6,088 | 1,030 | 7,247 |
| 764 | 1.29 [1.25-1.33] | 1.30 [1.26-1.34] | 1.29 [1.25-1.33] | 1.16 [0.98-1.38] | 1.19 [1.16-1.23] | 1.17 [1.13-1.21] | 1.11 [1.08-1.15] | 1.10 [1.06-1.14] | 1.12 [0.94-1.34] | 1.12 [1.08-1.15] | 10,148 | 10,113 | 8,600 | 361 | 9,225 |
| 765 | 1.32 [1.22-1.44] | 1.31 [1.20-1.42] | 1.33 [1.22-1.46] | | 1.21 [1.11-1.31] | 1.19 [1.09-1.30] | 1.10 [1.00-1.20] | 1.11 [1.02-1.22] | | 1.12 [1.03-1.23] | 1,339 | 1,340 | 1,189 | NA | 1,245 |
| 766 | 1.33 [1.27-1.39] | 1.33 [1.27-1.39] | 1.33 [1.27-1.40] | 1.22 [0.94-1.56] | 1.21 [1.15-1.27] | 1.18 [1.13-1.24] | 1.12 [1.07-1.17] | 1.11 [1.06-1.17] | 1.16 [0.89-1.51] | 1.12 [1.07-1.18] | 4,757 | 4,713 | 4,155 | 169 | 4,450 |
| 770 | 1.37 [1.30-1.44] | 1.37 [1.30-1.45] | 1.42 [1.34-1.51] | 1.24 [1.12-1.37] | 1.27 [1.21-1.34] | 1.25 [1.19-1.32] | 1.20 [1.13-1.27] | 1.24 [1.17-1.32] | 1.17 [1.06-1.30] | 1.20 [1.14-1.27] | 4,383 | 3,601 | 2,770 | 1,035 | 3,877 |
| 771 | 1.36 [1.28-1.44] | 1.39 [1.30-1.47] | 1.40 [1.31-1.50] | 1.25 [1.06-1.47] | 1.30 [1.22-1.38] | 1.26 [1.19-1.34] | 1.24 [1.16-1.32] | 1.25 [1.17-1.34] | 1.20 [1.01-1.43] | 1.24 [1.16-1.32] | 2,936 | 2,660 | 2,254 | 385 | 2,520 |
| 771.1 | 1.40 [1.37-1.43] | 1.41 [1.38-1.44] | 1.42 [1.39-1.45] | 1.21 [1.10-1.32] | 1.31 [1.28-1.34] | 1.31 [1.28-1.34] | 1.26 [1.24-1.29] | 1.27 [1.24-1.30] | 1.17 [1.07-1.28] | 1.24 [1.21-1.27] | 21,683 | 20,952 | 18,627 | 1,339 | 19,449 |
| 772 | 1.22 [1.16-1.29] | 1.26 [1.19-1.33] | 1.24 [1.16-1.31] | 1.05 [0.91-1.22] | 1.16 [1.09-1.22] | 1.17 [1.11-1.24] | 1.16 [1.10-1.23] | 1.16 [1.09-1.24] | 1.00 [0.86-1.16] | 1.13 [1.07-1.20] | 3,592 | 3,276 | 2,820 | 484 | 3,259 |
| 772.1 | 1.25 [1.10-1.42] | 1.28 [1.12-1.46] | 1.36 [1.19-1.56] | | 1.16 [1.02-1.32] | 1.16 [1.02-1.33] | 1.18 [1.03-1.35] | 1.26 [1.09-1.44] | | 1.13 [0.99-1.29] | 639 | 603 | 562 | NA | 586 |
| 772.4 | 1.07 [0.88-1.29] | 1.34 [1.07-1.68] | 1.39 [1.08-1.80] | | 0.93 [0.76-1.13] | 0.97 [0.80-1.18] | 1.20 [0.95-1.53] | 1.24 [0.96-1.62] | | 0.88 [0.72-1.08] | 273 | 188 | 151 | NA | 244 |
| 773 | 1.37 [1.34-1.40] | 1.38 [1.35-1.41] | 1.39 [1.36-1.42] | 1.21 [1.15-1.28] | 1.25 [1.23-1.28] | 1.26 [1.23-1.29] | 1.21 [1.19-1.24] | 1.23 [1.20-1.26] | 1.17 [1.10-1.24] | 1.19 [1.16-1.21] | 25,521 | 23,243 | 19,458 | 3,343 | 22,908 |
| 780 | 1.16 [1.10-1.23] | 1.17 [1.11-1.24] | 1.16 [1.09-1.22] | 1.04 [0.84-1.29] | 1.14 [1.08-1.21] | 1.18 [1.12-1.25] | 1.16 [1.10-1.23] | 1.15 [1.08-1.21] | 1.04 [0.84-1.30] | 1.17 [1.11-1.24] | 3,844 | 3,690 | 3,544 | 221 | 3,561 |
| 781 | 1.18 [1.16-1.19] | 1.19 [1.17-1.21] | 1.19 [1.17-1.20] | 1.06 [1.02-1.10] | 1.13 [1.12-1.15] | 1.14 [1.12-1.15] | 1.11 [1.10-1.13] | 1.11 [1.10-1.13] | 1.04 [1.00-1.08] | 1.11 [1.10-1.13] | 61,354 | 56,317 | 52,697 | 6,297 | 54,633 |
| 782.3 | 1.33 [1.30-1.36] | 1.33 [1.30-1.36] | 1.34 [1.31-1.37] | 1.21 [1.05-1.40] | 1.27 [1.24-1.30] | 1.26 [1.23-1.29] | 1.22 [1.19-1.24] | 1.22 [1.19-1.25] | 1.16 [1.00-1.35] | 1.22 [1.19-1.25] | 20,865 | 20,570 | 19,881 | 481 | 19,325 |
| 782.6 | 1.26 [1.13-1.39] | 1.26 [1.12-1.41] | 1.15 [1.01-1.31] | 1.39 [1.14-1.70] | 1.17 [1.05-1.30] | 1.17 [1.05-1.30] | 1.13 [1.00-1.27] | 1.04 [0.91-1.18] | 1.30 [1.06-1.60] | 1.11 [0.99-1.24] | 935 | 718 | 596 | 259 | 848 |
| 783 | 1.22 [1.20-1.25] | 1.27 [1.24-1.30] | 1.27 [1.24-1.31] | 1.15 [1.12-1.18] | 1.16 [1.14-1.18] | 1.17 [1.15-1.19] | 1.17 [1.14-1.20] | 1.17 [1.14-1.21] | 1.12 [1.09-1.15] | 1.13 [1.11-1.15] | 28,770 | 16,423 | 12,706 | 13,760 | 25,222 |
| 785 | 1.27 [1.26-1.28] | 1.28 [1.27-1.30] | 1.31 [1.29-1.32] | 1.19 [1.17-1.21] | 1.18 [1.17-1.19] | 1.19 [1.17-1.20] | 1.15 [1.13-1.16] | 1.17 [1.15-1.18] | 1.14 [1.12-1.16] | 1.12 [1.11-1.14] | 125,464 | 102,812 | 62,876 | 37,604 | 101,217 |
| 788 | 1.20 [1.18-1.21] | 1.20 [1.18-1.22] | 1.20 [1.18-1.22] | 1.10 [1.05-1.14] | 1.14 [1.12-1.16] | 1.15 [1.13-1.17] | 1.12 [1.10-1.14] | 1.12 [1.10-1.14] | 1.05 [1.01-1.10] | 1.11 [1.09-1.13] | 43,614 | 40,258 | 35,478 | 5,290 | 38,136 |
| 789 | 1.23 [1.22-1.25] | 1.25 [1.23-1.27] | 1.25 [1.23-1.27] | 1.17 [1.14-1.19] | 1.15 [1.13-1.17] | 1.16 [1.14-1.17] | 1.12 [1.10-1.14] | 1.13 [1.11-1.15] | 1.13 [1.10-1.15] | 1.11 [1.09-1.12] | 66,001 | 49,305 | 37,038 | 21,941 | 57,542 |
| 790 | 1.25 [1.15-1.36] | 1.27 [1.17-1.39] | 1.26 [1.15-1.38] | 1.13 [0.88-1.45] | 1.22 [1.13-1.33] | 1.18 [1.08-1.28] | 1.16 [1.06-1.26] | 1.16 [1.05-1.27] | 1.12 [0.87-1.46] | 1.17 [1.08-1.28] | 1,549 | 1,443 | 1,261 | 174 | 1,429 |
| 790.1 | 1.52 [1.28-1.82] | 1.49 [1.24-1.79] | 1.48 [1.22-1.79] | | 1.33 [1.10-1.61] | 1.44 [1.20-1.72] | 1.37 [1.13-1.66] | 1.37 [1.12-1.66] | | 1.25 [1.03-1.52] | 331 | 291 | 268 | NA | 274 |
| 790.6 | 1.24 [1.22-1.26] | 1.24 [1.21-1.26] | 1.24 [1.22-1.27] | 1.18 [1.09-1.29] | 1.17 [1.15-1.19] | 1.18 [1.15-1.20] | 1.14 [1.11-1.16] | 1.14 [1.11-1.16] | 1.15 [1.06-1.25] | 1.14 [1.11-1.16] | 28,223 | 27,256 | 25,657 | 1,540 | 25,681 |
| 791 | 1.22 [1.17-1.27] | 1.23 [1.18-1.28] | 1.22 [1.17-1.28] | 1.08 [0.92-1.28] | 1.16 [1.11-1.21] | 1.19 [1.14-1.24] | 1.13 [1.08-1.18] | 1.12 [1.07-1.17] | 1.07 [0.91-1.27] | 1.16 [1.11-1.21] | 6,432 | 6,203 | 5,777 | 376 | 5,857 |
| 793 | 1.18 [1.06-1.32] | 1.11 [0.99-1.24] | 1.14 [1.01-1.29] | 1.06 [0.77-1.44] | 1.13 [1.01-1.26] | 1.16 [1.04-1.29] | 1.04 [0.92-1.17] | 1.08 [0.96-1.22] | 1.05 [0.76-1.45] | 1.14 [1.02-1.28] | 837 | 744 | 692 | 108 | 772 |
| 793.2 | 1.17 [1.09-1.25] | 1.17 [1.09-1.25] | 1.18 [1.09-1.27] | 1.14 [0.90-1.44] | 1.10 [1.02-1.18] | 1.12 [1.04-1.20] | 1.09 [1.01-1.17] | 1.10 [1.02-1.18] | 1.08 [0.84-1.38] | 1.07 [0.99-1.15] | 2,110 | 1,999 | 1,786 | 189 | 1,876 |
| 794 | 1.06 [0.95-1.19] | 1.05 [0.94-1.18] | 1.16 [0.87-1.54] | 1.12 [0.87-1.44] | 1.09 [0.96-1.23] | 1.03 [0.92-1.16] | 1.02 [0.90-1.15] | 1.04 [0.77-1.40] | 1.13 [0.87-1.46] | 1.08 [0.95-1.23] | 710 | 692 | 111 | 160 | 577 |
| 797 | 1.18 [1.02-1.38] | 1.31 [1.12-1.54] | 1.21 [1.03-1.43] | | 1.19 [1.02-1.39] | 1.16 [0.99-1.35] | 1.25 [1.06-1.47] | 1.14 [0.96-1.35] | | 1.20 [1.03-1.41] | 439 | 404 | 368 | NA | 401 |
| 797.1 | 1.09 [0.98-1.20] | 1.08 [0.98-1.20] | 1.06 [0.96-1.17] | | 1.07 [0.96-1.19] | 1.10 [0.99-1.22] | 1.05 [0.95-1.17] | 1.02 [0.92-1.13] | | 1.10 [0.98-1.22] | 948 | 939 | 911 | NA | 834 |
| 798 | 1.24 [1.21-1.27] | 1.23 [1.20-1.26] | 1.22 [1.19-1.25] | 1.22 [1.16-1.29] | 1.17 [1.14-1.19] | 1.18 [1.15-1.20] | 1.12 [1.10-1.15] | 1.12 [1.09-1.14] | 1.18 [1.11-1.25] | 1.13 [1.10-1.16] | 23,449 | 20,786 | 18,510 | 3,424 | 21,508 |
| 798.1 | 1.49 [1.40-1.60] | 1.49 [1.39-1.60] | 1.46 [1.34-1.59] | 1.31 [1.12-1.54] | 1.34 [1.25-1.44] | 1.27 [1.18-1.36] | 1.17 [1.09-1.26] | 1.15 [1.05-1.26] | 1.23 [1.04-1.44] | 1.16 [1.08-1.25] | 2,283 | 2,076 | 1,346 | 448 | 2,004 |
| injuries & poisonings | | | | | | | | | | | | | | | |
| 800 | 1.09 [1.06-1.13] | 1.10 [1.06-1.15] | 1.12 [1.08-1.17] | 1.02 [0.94-1.09] | 1.05 [1.01-1.09] | 1.06 [1.03-1.10] | 1.04 [1.00-1.08] | 1.07 [1.02-1.11] | 1.00 [0.93-1.08] | 1.04 [1.00-1.08] | 8,111 | 6,863 | 5,613 | 1,795 | 6,526 |
| 800.1 | 1.11 [1.08-1.13] | 1.11 [1.09-1.14] | 1.12 [1.09-1.14] | 1.07 [0.85-1.36] | 1.09 [1.07-1.12] | 1.10 [1.08-1.12] | 1.08 [1.06-1.11] | 1.08 [1.06-1.11] | 1.07 [0.83-1.37] | 1.09 [1.07-1.12] | 22,544 | 22,421 | 22,271 | 173 | 19,750 |
| 800.2 | 1.10 [1.08-1.12] | 1.11 [1.09-1.14] | 1.12 [1.10-1.14] | 0.98 [0.90-1.07] | 1.08 [1.06-1.11] | 1.09 [1.07-1.11] | 1.08 [1.06-1.11] | 1.08 [1.06-1.11] | 0.97 [0.89-1.06] | 1.09 [1.06-1.11] | 25,942 | 24,948 | 24,537 | 1,190 | 22,614 |
| 800.3 | 1.08 [1.05-1.12] | 1.11 [1.07-1.16] | 1.11 [1.06-1.16] | 1.04 [0.99-1.10] | 1.05 [1.01-1.09] | 1.05 [1.02-1.09] | 1.04 [1.00-1.09] | 1.04 [0.99-1.10] | 1.03 [0.97-1.08] | 1.03 [0.99-1.08] | 8,131 | 5,262 | 3,886 | 3,466 | 6,389 |
| 800.4 | 1.10 [1.02-1.18] | 1.13 [1.05-1.21] | 1.10 [1.02-1.20] | 0.93 [0.77-1.12] | 1.02 [0.94-1.10] | 1.06 [0.99-1.14] | 1.07 [0.99-1.15] | 1.05 [0.96-1.14] | 0.90 [0.74-1.09] | 1.01 [0.93-1.09] | 1,880 | 1,726 | 1,395 | 278 | 1,491 |
| 801 | 1.10 [1.06-1.15] | 1.12 [1.07-1.17] | 1.15 [1.09-1.21] | 0.96 [0.88-1.05] | 1.07 [1.03-1.12] | 1.06 [1.01-1.10] | 1.04 [0.99-1.08] | 1.06 [1.01-1.12] | 0.95 [0.87-1.04] | 1.05 [1.00-1.10] | 5,848 | 5,124 | 3,794 | 1,304 | 4,681 |
| 801.1 | 1.10 [0.94-1.30] | 1.18 [0.99-1.40] | 1.30 [1.04-1.63] | 0.99 [0.73-1.33] | 1.10 [0.92-1.33] | 1.06 [0.90-1.25] | 1.11 [0.92-1.32] | 1.16 [0.92-1.47] | 0.99 [0.73-1.35] | 1.08 [0.89-1.30] | 362 | 309 | 186 | 111 | 287 |
| 802 | 1.11 [1.07-1.16] | 1.12 [1.08-1.17] | 1.14 [1.09-1.19] | 0.88 [0.74-1.05] | 1.07 [1.02-1.11] | 1.08 [1.04-1.13] | 1.07 [1.03-1.12] | 1.09 [1.04-1.13] | 0.89 [0.74-1.07] | 1.05 [1.01-1.10] | 6,509 | 6,340 | 6,052 | 313 | 5,836 |
| 803 | 1.09 [1.02-1.16] | 1.16 [1.06-1.26] | 1.19 [1.08-1.31] | 1.01 [0.92-1.11] | 1.03 [0.96-1.11] | 1.06 [0.99-1.13] | 1.09 [1.00-1.19] | 1.14 [1.03-1.26] | 0.99 [0.89-1.09] | 1.02 [0.95-1.10] | 2,203 | 1,257 | 1,027 | 1,058 | 1,715 |
| 803.1 | 1.11 [1.08-1.14] | 1.14 [1.10-1.18] | 1.14 [1.09-1.18] | 1.06 [1.01-1.11] | 1.08 [1.05-1.12] | 1.09 [1.05-1.12] | 1.08 [1.04-1.12] | 1.08 [1.04-1.12] | 1.06 [1.00-1.11] | 1.08 [1.04-1.11] | 10,942 | 7,254 | 6,760 | 3,901 | 9,330 |
| 803.2 | 1.06 [1.04-1.08] | 1.10 [1.07-1.13] | 1.11 [1.07-1.14] | 1.01 [0.98-1.04] | 1.02 [1.00-1.05] | 1.04 [1.02-1.06] | 1.06 [1.03-1.09] | 1.07 [1.03-1.10] | 1.00 [0.97-1.03] | 1.01 [0.99-1.04] | 23,610 | 12,452 | 10,625 | 11,920 | 18,431 |
| 803.3 | 1.15 [1.11-1.20] | 1.16 [1.12-1.21] | 1.19 [1.14-1.24] | 1.01 [0.90-1.12] | 1.10 [1.05-1.14] | 1.11 [1.07-1.15] | 1.08 [1.04-1.13] | 1.10 [1.06-1.15] | 0.99 [0.89-1.11] | 1.08 [1.03-1.12] | 7,012 | 6,527 | 5,667 | 901 | 5,997 |
| 804 | 1.05 [1.02-1.08] | 1.09 [1.05-1.12] | 1.10 [1.06-1.15] | 1.01 [0.98-1.05] | 1.00 [0.97-1.03] | 1.02 [0.99-1.05] | 1.03 [0.99-1.06] | 1.04 [1.00-1.09] | 1.00 [0.96-1.03] | 0.99 [0.96-1.02] | 15,344 | 9,028 | 5,771 | 8,096 | 11,805 |
| 805 | 1.18 [1.14-1.23] | 1.18 [1.14-1.22] | 1.21 [1.17-1.26] | 1.01 [0.89-1.15] | 1.14 [1.10-1.19] | 1.13 [1.09-1.18] | 1.10 [1.06-1.14] | 1.13 [1.08-1.17] | 1.01 [0.89-1.15] | 1.12 [1.08-1.16] | 8,082 | 7,844 | 7,027 | 627 | 7,155 |
| 807 | 1.22 [1.17-1.27] | 1.21 [1.16-1.26] | 1.24 [1.19-1.29] | 1.09 [0.91-1.29] | 1.14 [1.10-1.19] | 1.16 [1.12-1.21] | 1.11 [1.07-1.16] | 1.13 [1.08-1.18] | 1.11 [0.93-1.33] | 1.12 [1.07-1.17] | 6,514 | 6,407 | 5,894 | 328 | 5,765 |
| 809 | 1.19 [1.10-1.28] | 1.19 [1.10-1.28] | 1.24 [1.14-1.35] | 1.05 [0.83-1.34] | 1.11 [1.03-1.21] | 1.14 [1.06-1.24] | 1.10 [1.02-1.20] | 1.14 [1.05-1.25] | 1.03 [0.81-1.33] | 1.10 [1.01-1.20] | 1,690 | 1,602 | 1,443 | 177 | 1,430 |
| 817 | 1.10 [1.00-1.21] | 1.14 [1.00-1.30] | 1.12 [0.94-1.33] | 1.08 [0.95-1.24] | 1.06 [0.95-1.18] | 1.05 [0.95-1.16] | 1.02 [0.89-1.17] | 1.03 [0.86-1.22] | 1.05 [0.92-1.20] | 1.03 [0.92-1.15] | 1,015 | 544 | 319 | 574 | 821 |
| 818 | 1.11 [1.06-1.16] | 1.13 [1.07-1.18] | 1.15 [1.09-1.21] | 1.00 [0.87-1.14] | 1.07 [1.01-1.12] | 1.09 [1.04-1.14] | 1.07 [1.02-1.13] | 1.09 [1.04-1.15] | 0.99 [0.86-1.14] | 1.06 [1.01-1.11] | 4,885 | 4,583 | 4,148 | 536 | 4,312 |
| 819 | 1.12 [1.10-1.15] | 1.16 [1.13-1.20] | 1.19 [1.16-1.23] | 1.04 [1.00-1.08] | 1.06 [1.03-1.09] | 1.09 [1.07-1.12] | 1.09 [1.06-1.12] | 1.12 [1.08-1.16] | 1.02 [0.98-1.06] | 1.05 [1.02-1.07] | 18,084 | 14,150 | 9,555 | 6,304 | 14,386 |
| 830 | 1.15 [1.11-1.19] | 1.17 [1.13-1.22] | 1.13 [1.07-1.19] | 1.08 [1.02-1.15] | 1.09 [1.05-1.13] | 1.09 [1.05-1.13] | 1.07 [1.03-1.11] | 1.07 [1.01-1.13] | 1.06 [0.99-1.13] | 1.06 [1.02-1.10] | 8,227 | 6,917 | 3,358 | 2,574 | 6,858 |
| 835 | 1.23 [1.20-1.25] | 1.23 [1.21-1.26] | 1.24 [1.21-1.27] | 1.16 [1.10-1.22] | 1.13 [1.10-1.16] | 1.15 [1.12-1.17] | 1.13 [1.11-1.16] | 1.13 [1.10-1.16] | 1.11 [1.05-1.17] | 1.07 [1.05-1.10] | 21,687 | 20,302 | 14,578 | 3,603 | 16,501 |
| 836 | 1.30 [1.18-1.42] | 1.30 [1.18-1.43] | 1.34 [1.21-1.48] | | 1.19 [1.08-1.31] | 1.21 [1.10-1.34] | 1.17 [1.06-1.29] | 1.21 [1.09-1.34] | | 1.15 [1.04-1.27] | 1,071 | 1,065 | 908 | NA | 967 |
| 840 | 1.22 [1.09-1.35] | 1.19 [1.06-1.33] | 1.16 [0.99-1.35] | 1.21 [1.01-1.46] | 1.14 [1.02-1.29] | 1.15 [1.03-1.28] | 1.09 [0.97-1.23] | 1.07 [0.92-1.25] | 1.16 [0.96-1.40] | 1.10 [0.98-1.25] | 860 | 722 | 399 | 299 | 664 |
| 840.3 | 1.03 [0.87-1.23] | 1.01 [0.83-1.22] | 0.98 [0.77-1.26] | | 1.02 [0.83-1.25] | 1.00 [0.83-1.20] | 0.94 [0.77-1.15] | 0.95 [0.74-1.22] | | 1.01 [0.82-1.24] | 291 | 246 | 145 | NA | 218 |
| 850 | 1.27 [1.24-1.30] | 1.27 [1.24-1.31] | 1.26 [1.22-1.30] | 1.22 [1.15-1.30] | 1.19 [1.16-1.22] | 1.20 [1.17-1.23] | 1.16 [1.13-1.19] | 1.15 [1.12-1.19] | 1.17 [1.10-1.25] | 1.15 [1.12-1.18] | 16,761 | 14,625 | 12,023 | 3,111 | 14,552 |
| 851 | 1.28 [1.26-1.30] | 1.29 [1.27-1.31] | 1.27 [1.25-1.29] | 1.23 [1.18-1.27] | 1.18 [1.17-1.20] | 1.21 [1.19-1.22] | 1.17 [1.16-1.19] | 1.17 [1.15-1.18] | 1.17 [1.13-1.22] | 1.15 [1.13-1.16] | 53,275 | 47,864 | 40,200 | 8,014 | 46,944 |
| 853 | 1.33 [1.24-1.42] | 1.36 [1.26-1.46] | 1.30 [1.20-1.40] | 1.15 [0.90-1.48] | 1.20 [1.11-1.29] | 1.22 [1.14-1.32] | 1.19 [1.10-1.28] | 1.17 [1.08-1.27] | 1.09 [0.84-1.40] | 1.14 [1.06-1.23] | 1,957 | 1,860 | 1,631 | 163 | 1,807 |
| 854 | 1.31 [1.27-1.35] | 1.34 [1.30-1.38] | 1.34 [1.29-1.38] | 1.01 [0.92-1.12] | 1.19 [1.16-1.23] | 1.25 [1.21-1.29] | 1.21 [1.17-1.25] | 1.20 [1.16-1.25] | 0.99 [0.90-1.10] | 1.17 [1.13-1.21] | 9,866 | 9,093 | 8,384 | 996 | 9,084 |
| 857 | 1.14 [1.10-1.18] | 1.16 [1.12-1.20] | 1.14 [1.09-1.18] | 1.03 [0.87-1.21] | 1.08 [1.04-1.12] | 1.11 [1.07-1.15] | 1.10 [1.06-1.14] | 1.08 [1.04-1.12] | 0.97 [0.82-1.15] | 1.07 [1.03-1.11] | 8,538 | 8,392 | 7,525 | 387 | 7,974 |
| 858 | 1.31 [1.27-1.34] | 1.32 [1.28-1.35] | 1.34 [1.30-1.38] | 1.13 [1.03-1.24] | 1.20 [1.17-1.24] | 1.24 [1.20-1.27] | 1.21 [1.18-1.24] | 1.23 [1.20-1.27] | 1.10 [1.00-1.21] | 1.17 [1.14-1.20] | 13,678 | 13,074 | 11,614 | 1,217 | 12,548 |
| 859 | 1.31 [1.27-1.34] | 1.33 [1.29-1.37] | 1.33 [1.29-1.38] | 1.16 [1.08-1.25] | 1.19 [1.16-1.23] | 1.23 [1.19-1.27] | 1.21 [1.17-1.25] | 1.21 [1.17-1.25] | 1.11 [1.03-1.20] | 1.16 [1.12-1.19] | 12,458 | 11,132 | 9,626 | 1,821 | 11,466 |
| 870 | 1.09 [1.01-1.18] | 1.12 [1.02-1.23] | 1.40 [1.24-1.58] | 0.92 [0.81-1.04] | 1.05 [0.96-1.14] | 1.05 [0.96-1.14] | 1.01 [0.92-1.12] | 1.27 [1.11-1.44] | 0.91 [0.80-1.03] | 1.04 [0.95-1.14] | 1,537 | 1,102 | 675 | 668 | 1,249 |
| 870.1 | 1.13 [1.08-1.19] | 1.15 [1.08-1.22] | 1.18 [1.10-1.25] | 1.02 [0.93-1.12] | 1.09 [1.03-1.15] | 1.10 [1.05-1.16] | 1.08 [1.02-1.15] | 1.11 [1.04-1.18] | 1.02 [0.93-1.12] | 1.08 [1.02-1.14] | 4,027 | 3,061 | 2,530 | 1,212 | 3,469 |
| 870.2 | 1.08 [0.99-1.19] | 1.08 [0.96-1.21] | 1.17 [1.02-1.34] | 1.05 [0.93-1.20] | 1.04 [0.94-1.15] | 1.07 [0.98-1.17] | 1.02 [0.91-1.15] | 1.13 [0.98-1.29] | 1.03 [0.90-1.18] | 1.04 [0.94-1.15] | 1,219 | 764 | 555 | 574 | 1,011 |
| 870.3 | 1.13 [1.11-1.15] | 1.18 [1.16-1.21] | 1.20 [1.18-1.23] | 0.97 [0.94-1.01] | 1.08 [1.06-1.11] | 1.10 [1.08-1.12] | 1.12 [1.09-1.14] | 1.14 [1.11-1.17] | 0.96 [0.93-1.00] | 1.07 [1.05-1.09] | 28,524 | 21,364 | 18,584 | 8,473 | 24,852 |
| 870.4 | 1.12 [1.05-1.20] | 1.20 [1.11-1.29] | 1.21 [1.11-1.31] | 0.89 [0.77-1.03] | 1.08 [1.00-1.16] | 1.08 [1.01-1.16] | 1.12 [1.03-1.21] | 1.12 [1.03-1.22] | 0.89 [0.77-1.03] | 1.05 [0.97-1.14] | 2,051 | 1,705 | 1,504 | 441 | 1,809 |
| 870.5 | 1.02 [0.98-1.07] | 1.12 [1.05-1.21] | 1.19 [1.09-1.31] | 0.95 [0.90-1.00] | 0.98 [0.94-1.03] | 1.00 [0.96-1.05] | 1.08 [1.00-1.16] | 1.12 [1.02-1.23] | 0.94 [0.89-1.00] | 0.98 [0.93-1.03] | 4,608 | 1,848 | 1,227 | 3,072 | 3,837 |
| 870.6 | 1.15 [1.05-1.27] | 1.17 [1.05-1.29] | 1.24 [1.10-1.40] | 1.04 [0.88-1.24] | 1.12 [1.02-1.24] | 1.11 [1.01-1.22] | 1.09 [0.98-1.21] | 1.14 [1.00-1.29] | 1.03 [0.86-1.23] | 1.12 [1.01-1.24] | 1,141 | 951 | 686 | 333 | 981 |
| 870.8 | 1.13 [1.00-1.28] | 1.08 [0.89-1.31] | 1.32 [1.00-1.74] | 1.10 [0.94-1.27] | 1.06 [0.92-1.22] | 1.10 [0.97-1.24] | 1.02 [0.83-1.24] | 1.19 [0.90-1.58] | 1.09 [0.93-1.27] | 1.05 [0.91-1.21] | 652 | 254 | 132 | 452 | 516 |
| 871 | 1.10 [1.08-1.12] | 1.15 [1.12-1.18] | 1.20 [1.17-1.24] | 0.98 [0.95-1.01] | 1.05 [1.03-1.08] | 1.05 [1.03-1.07] | 1.06 [1.04-1.08] | 1.11 [1.08-1.14] | 0.97 [0.94-1.01] | 1.03 [1.01-1.05] | 27,381 | 20,760 | 15,409 | 9,328 | 22,587 |
| 871.3 | 1.11 [1.02-1.21] | 1.24 [1.11-1.40] | 1.33 [1.15-1.53] | 1.01 [0.90-1.14] | 1.09 [0.99-1.20] | 1.06 [0.97-1.16] | 1.10 [0.98-1.24] | 1.19 [1.03-1.38] | 0.99 [0.88-1.12] | 1.05 [0.95-1.16] | 1,299 | 727 | 477 | 700 | 1,081 |
| 871.4 | 1.12 [0.99-1.27] | 1.24 [1.06-1.45] | 1.33 [1.12-1.58] | 0.94 [0.77-1.14] | 1.07 [0.93-1.22] | 1.09 [0.96-1.24] | 1.13 [0.96-1.32] | 1.19 [0.99-1.42] | 0.95 [0.78-1.17] | 1.05 [0.92-1.21] | 640 | 424 | 351 | 253 | 555 |
| 872 | 0.99 [0.91-1.07] | 0.98 [0.88-1.09] | 1.01 [0.88-1.15] | 0.98 [0.87-1.11] | 0.94 [0.85-1.03] | 0.99 [0.91-1.08] | 0.97 [0.87-1.09] | 1.01 [0.88-1.16] | 0.97 [0.86-1.10] | 0.94 [0.85-1.04] | 1,306 | 732 | 497 | 685 | 995 |
| 873 | 1.13 [1.05-1.23] | 1.26 [1.14-1.39] | 1.36 [1.21-1.53] | 1.00 [0.89-1.12] | 1.05 [0.97-1.15] | 1.11 [1.02-1.20] | 1.15 [1.04-1.28] | 1.25 [1.10-1.41] | 1.00 [0.89-1.13] | 1.04 [0.95-1.13] | 1,602 | 1,037 | 686 | 720 | 1,327 |
| 874 | 1.35 [1.20-1.52] | 1.30 [1.16-1.47] | 1.28 [1.13-1.45] | | 1.21 [1.07-1.36] | 1.25 [1.11-1.41] | 1.14 [1.01-1.29] | 1.08 [0.95-1.23] | | 1.17 [1.04-1.32] | 716 | 681 | 622 | NA | 663 |
| 905 | 1.28 [1.10-1.49] | 1.29 [1.11-1.51] | 1.31 [1.08-1.59] | | 1.14 [0.96-1.34] | 1.20 [1.03-1.40] | 1.19 [1.02-1.40] | 1.17 [0.96-1.44] | | 1.09 [0.92-1.29] | 419 | 389 | 250 | NA | 337 |
| 907 | 1.10 [1.05-1.16] | 1.13 [1.07-1.19] | 1.16 [1.09-1.24] | 0.99 [0.91-1.09] | 1.02 [0.97-1.08] | 1.07 [1.01-1.12] | 1.04 [0.99-1.10] | 1.07 [1.01-1.15] | 1.00 [0.90-1.10] | 1.00 [0.95-1.06] | 4,011 | 3,421 | 2,275 | 1,144 | 3,175 |
| 913 | 1.10 [0.88-1.36] | 0.98 [0.77-1.26] | 0.95 [0.72-1.25] | | 1.00 [0.78-1.28] | 1.02 [0.81-1.27] | 0.88 [0.68-1.13] | 0.88 [0.66-1.16] | | 0.94 [0.73-1.21] | 199 | 145 | 111 | NA | 151 |
| 915 | 1.15 [1.13-1.16] | 1.19 [1.17-1.21] | 1.21 [1.19-1.23] | 0.99 [0.96-1.02] | 1.09 [1.07-1.11] | 1.10 [1.09-1.12] | 1.10 [1.08-1.11] | 1.12 [1.10-1.14] | 0.97 [0.95-1.00] | 1.07 [1.05-1.08] | 47,549 | 38,385 | 33,586 | 11,393 | 42,090 |
| 916 | 1.16 [1.13-1.19] | 1.18 [1.15-1.22] | 1.20 [1.17-1.24] | 0.97 [0.91-1.04] | 1.10 [1.07-1.13] | 1.11 [1.08-1.14] | 1.09 [1.06-1.13] | 1.12 [1.08-1.15] | 0.96 [0.90-1.03] | 1.08 [1.05-1.11] | 13,528 | 11,824 | 10,631 | 2,224 | 12,240 |
| 930 | 5.58 [5.28-5.90] | 2.80 [2.54-3.08] | 2.02 [1.73-2.34] | 7.34 [6.88-7.84] | 5.48 [5.15-5.82] | 4.37 [4.11-4.63] | 1.81 [1.62-2.02] | 1.57 [1.34-1.85] | 5.82 [5.42-6.25] | 4.27 [4.01-4.56] | 6,437 | 1,302 | 460 | 5,577 | 5,437 |
| 941 | 1.53 [1.40-1.66] | 1.45 [1.33-1.60] | 1.40 [1.27-1.54] | 1.85 [1.52-2.25] | 1.43 [1.31-1.56] | 1.39 [1.27-1.51] | 1.27 [1.16-1.40] | 1.24 [1.12-1.38] | 1.71 [1.40-2.10] | 1.32 [1.21-1.45] | 1,423 | 1,192 | 1,037 | 298 | 1,306 |
| 942 | 1.70 [1.28-2.25] | | | | 1.66 [1.25-2.21] | 1.70 [1.28-2.27] | | | | 1.70 [1.27-2.29] | 137 | NA | NA | NA | 130 |
| 946 | 1.96 [1.84-2.08] | 1.76 [1.64-1.89] | 1.66 [1.54-1.80] | 2.76 [2.44-3.11] | 1.81 [1.69-1.93] | 1.68 [1.58-1.79] | 1.44 [1.34-1.54] | 1.40 [1.29-1.53] | 2.33 [2.05-2.64] | 1.60 [1.49-1.71] | 2,962 | 2,249 | 1,619 | 968 | 2,562 |
| 949 | 2.22 [2.08-2.37] | 1.91 [1.76-2.07] | 1.71 [1.55-1.90] | 2.71 [2.46-2.98] | 2.07 [1.93-2.22] | 1.91 [1.79-2.05] | 1.58 [1.45-1.73] | 1.47 [1.32-1.64] | 2.32 [2.10-2.57] | 1.81 [1.68-1.95] | 2,834 | 1,569 | 948 | 1,480 | 2,350 |
| 949.1 | 1.05 [0.90-1.22] | 1.08 [0.83-1.40] | 1.00 [0.77-1.31] | 1.01 [0.84-1.21] | 1.01 [0.86-1.18] | 1.01 [0.87-1.18] | 1.02 [0.79-1.33] | 0.96 [0.73-1.25] | 0.95 [0.78-1.15] | 0.99 [0.84-1.16] | 424 | 164 | 160 | 264 | 405 |
| 952 | 1.03 [0.88-1.20] | 1.04 [0.89-1.22] | 1.10 [0.93-1.32] | | 1.00 [0.85-1.19] | 1.02 [0.88-1.19] | 1.00 [0.85-1.17] | 1.06 [0.89-1.27] | | 1.00 [0.84-1.18] | 422 | 382 | 303 | NA | 341 |
| 957 | 1.31 [1.19-1.45] | 1.35 [1.21-1.49] | 1.33 [1.20-1.47] | | 1.20 [1.08-1.34] | 1.22 [1.10-1.36] | 1.20 [1.08-1.33] | 1.17 [1.05-1.30] | | 1.16 [1.04-1.29] | 939 | 936 | 905 | NA | 872 |
| 958 | 1.09 [1.03-1.17] | 1.11 [1.04-1.18] | 1.12 [1.04-1.21] | 1.05 [0.90-1.22] | 1.04 [0.98-1.12] | 1.07 [1.00-1.14] | 1.04 [0.97-1.11] | 1.07 [0.99-1.15] | 1.03 [0.89-1.21] | 1.04 [0.97-1.12] | 2,569 | 2,323 | 1,932 | 438 | 2,157 |
| 958.1 | 1.18 [0.92-1.50] | 1.16 [0.91-1.49] | 1.12 [0.87-1.45] | | 1.17 [0.91-1.51] | 1.13 [0.88-1.45] | 1.10 [0.85-1.41] | 1.07 [0.82-1.39] | | 1.17 [0.90-1.52] | 160 | 150 | 139 | NA | 140 |
| 958.2 | 1.05 [0.91-1.20] | 1.09 [0.95-1.26] | 1.09 [0.93-1.29] | 1.12 [0.85-1.49] | 1.04 [0.90-1.21] | 1.00 [0.87-1.16] | 1.01 [0.87-1.17] | 1.02 [0.86-1.22] | 1.08 [0.81-1.44] | 1.03 [0.89-1.20] | 515 | 463 | 342 | 131 | 430 |
| 960 | 1.62 [1.58-1.65] | 1.63 [1.59-1.66] | 1.64 [1.60-1.68] | 1.42 [1.33-1.51] | 1.49 [1.46-1.53] | 1.40 [1.37-1.43] | 1.36 [1.33-1.39] | 1.37 [1.33-1.40] | 1.31 [1.23-1.40] | 1.33 [1.30-1.36] | 24,400 | 22,914 | 18,975 | 2,787 | 22,398 |
| 960.1 | 1.52 [1.32-1.76] | 1.64 [1.40-1.93] | 1.76 [1.44-2.14] | 1.30 [1.04-1.64] | 1.39 [1.19-1.61] | 1.38 [1.19-1.59] | 1.38 [1.16-1.63] | 1.44 [1.17-1.78] | 1.24 [0.98-1.57] | 1.29 [1.11-1.51] | 521 | 392 | 267 | 193 | 450 |
| 960.2 | 1.44 [1.42-1.46] | 1.45 [1.43-1.47] | 1.46 [1.44-1.48] | 1.37 [1.33-1.41] | 1.36 [1.35-1.38] | 1.32 [1.31-1.34] | 1.29 [1.27-1.30] | 1.30 [1.28-1.32] | 1.28 [1.24-1.32] | 1.27 [1.25-1.29] | 75,992 | 68,701 | 52,815 | 12,829 | 62,561 |
| 961 | 1.52 [1.42-1.62] | 1.54 [1.44-1.65] | 1.52 [1.41-1.64] | 1.27 [1.05-1.53] | 1.41 [1.32-1.50] | 1.33 [1.24-1.42] | 1.28 [1.19-1.37] | 1.26 [1.16-1.36] | 1.21 [0.99-1.47] | 1.28 [1.19-1.37] | 2,474 | 2,315 | 1,859 | 312 | 2,307 |
| 961.1 | 1.59 [1.49-1.69] | 1.56 [1.47-1.67] | 1.58 [1.47-1.69] | | 1.46 [1.36-1.56] | 1.39 [1.30-1.49] | 1.32 [1.23-1.41] | 1.33 [1.24-1.43] | | 1.32 [1.23-1.41] | 2,515 | 2,482 | 2,241 | NA | 2,273 |
| 962 | 1.26 [1.16-1.38] | 1.34 [1.22-1.48] | 1.35 [1.20-1.52] | 1.04 [0.88-1.23] | 1.15 [1.05-1.26] | 1.15 [1.05-1.26] | 1.10 [0.99-1.22] | 1.10 [0.97-1.25] | 1.02 [0.86-1.21] | 1.10 [1.00-1.21] | 1,223 | 986 | 696 | 370 | 1,122 |
| 962.3 | 1.60 [1.49-1.71] | 1.60 [1.49-1.72] | 1.55 [1.43-1.67] | 1.37 [1.13-1.67] | 1.44 [1.34-1.55] | 1.38 [1.28-1.48] | 1.30 [1.21-1.40] | 1.27 [1.17-1.37] | 1.29 [1.05-1.58] | 1.28 [1.19-1.37] | 2,226 | 2,047 | 1,753 | 290 | 2,061 |
| 963 | 1.23 [1.19-1.27] | 1.24 [1.20-1.28] | 1.22 [1.17-1.27] | 1.17 [1.08-1.26] | 1.15 [1.11-1.19] | 1.18 [1.14-1.22] | 1.16 [1.12-1.21] | 1.16 [1.11-1.20] | 1.12 [1.04-1.22] | 1.14 [1.10-1.18] | 8,518 | 7,403 | 6,255 | 1,601 | 7,406 |
| 963.1 | 1.33 [1.05-1.70] | 1.56 [1.18-2.04] | | | 1.15 [0.89-1.48] | 1.28 [1.00-1.64] | 1.45 [1.10-1.91] | | | 1.11 [0.86-1.45] | 167 | 132 | NA | NA | 144 |
| 964 | 1.24 [1.17-1.32] | 1.25 [1.18-1.34] | 1.25 [1.17-1.34] | 1.07 [0.89-1.28] | 1.17 [1.10-1.24] | 1.18 [1.11-1.26] | 1.15 [1.08-1.23] | 1.14 [1.07-1.22] | 1.07 [0.89-1.29] | 1.13 [1.06-1.21] | 2,723 | 2,514 | 2,302 | 305 | 2,523 |
| 964.1 | 1.27 [1.18-1.36] | 1.28 [1.19-1.37] | 1.27 [1.18-1.37] | | 1.19 [1.11-1.28] | 1.21 [1.13-1.30] | 1.17 [1.09-1.26] | 1.16 [1.08-1.25] | | 1.15 [1.07-1.24] | 2,103 | 2,073 | 2,008 | NA | 1,996 |
| 965 | 1.31 [1.29-1.33] | 1.35 [1.33-1.37] | 1.41 [1.38-1.43] | 1.08 [1.05-1.12] | 1.22 [1.20-1.24] | 1.18 [1.16-1.20] | 1.14 [1.12-1.16] | 1.19 [1.16-1.21] | 1.05 [1.02-1.09] | 1.13 [1.11-1.15] | 39,157 | 33,202 | 24,181 | 9,715 | 34,065 |
| 965.1 | 1.38 [1.34-1.42] | 1.39 [1.35-1.44] | 1.37 [1.32-1.42] | 1.26 [1.12-1.42] | 1.27 [1.23-1.31] | 1.22 [1.18-1.26] | 1.17 [1.13-1.21] | 1.16 [1.12-1.21] | 1.18 [1.04-1.34] | 1.16 [1.12-1.20] | 10,675 | 10,555 | 8,596 | 759 | 10,260 |
| 965.3 | 1.17 [1.08-1.26] | 1.24 [1.14-1.34] | 1.25 [1.13-1.38] | 0.93 [0.81-1.07] | 1.07 [0.99-1.17] | 1.12 [1.03-1.21] | 1.10 [1.01-1.19] | 1.12 [1.01-1.24] | 0.93 [0.81-1.08] | 1.06 [0.97-1.15] | 1,698 | 1,436 | 969 | 467 | 1,405 |
| 966 | 1.25 [1.18-1.32] | 1.29 [1.22-1.36] | 1.33 [1.24-1.43] | 1.00 [0.89-1.13] | 1.11 [1.05-1.17] | 1.14 [1.08-1.20] | 1.07 [1.00-1.13] | 1.10 [1.03-1.19] | 1.02 [0.90-1.15] | 1.08 [1.02-1.14] | 3,375 | 3,007 | 1,949 | 724 | 3,019 |
| 967 | 1.38 [1.29-1.47] | 1.35 [1.26-1.45] | 1.34 [1.23-1.46] | 1.26 [1.12-1.43] | 1.24 [1.16-1.32] | 1.23 [1.15-1.31] | 1.15 [1.07-1.23] | 1.13 [1.03-1.23] | 1.19 [1.05-1.35] | 1.15 [1.07-1.23] | 2,483 | 1,986 | 1,289 | 744 | 2,243 |
| 969 | 1.16 [1.12-1.19] | 1.20 [1.16-1.24] | 1.31 [1.25-1.37] | 0.97 [0.92-1.01] | 1.06 [1.03-1.09] | 1.08 [1.05-1.11] | 1.01 [0.98-1.04] | 1.08 [1.03-1.13] | 0.97 [0.92-1.02] | 1.04 [1.01-1.08] | 11,596 | 9,490 | 4,536 | 4,368 | 9,695 |
| 971 | 1.19 [1.13-1.26] | 1.25 [1.18-1.32] | 1.25 [1.17-1.33] | 1.01 [0.89-1.15] | 1.14 [1.08-1.20] | 1.15 [1.08-1.21] | 1.13 [1.07-1.20] | 1.13 [1.06-1.21] | 0.99 [0.87-1.13] | 1.13 [1.07-1.20] | 3,453 | 3,086 | 2,498 | 615 | 3,113 |
| 972 | 1.30 [1.24-1.35] | 1.32 [1.27-1.38] | 1.30 [1.24-1.35] | 0.98 [0.81-1.18] | 1.22 [1.17-1.27] | 1.23 [1.18-1.28] | 1.19 [1.14-1.24] | 1.17 [1.12-1.22] | 0.95 [0.79-1.15] | 1.18 [1.13-1.23] | 6,146 | 5,946 | 5,731 | 285 | 5,717 |
| 972.2 | 1.39 [1.12-1.72] | 1.34 [1.08-1.66] | 1.38 [1.11-1.71] | | 1.27 [1.01-1.58] | 1.34 [1.08-1.66] | 1.23 [0.99-1.54] | 1.29 [1.04-1.61] | | 1.26 [1.00-1.58] | 216 | 215 | 214 | NA | 198 |
| 973 | 1.27 [1.17-1.37] | 1.33 [1.23-1.45] | 1.34 [1.22-1.46] | 1.02 [0.86-1.22] | 1.20 [1.11-1.30] | 1.15 [1.06-1.24] | 1.11 [1.02-1.21] | 1.13 [1.03-1.25] | 1.00 [0.83-1.19] | 1.13 [1.05-1.23] | 1,780 | 1,555 | 1,266 | 347 | 1,643 |
| 974 | 1.43 [1.32-1.54] | 1.44 [1.33-1.55] | 1.41 [1.30-1.52] | | 1.30 [1.20-1.41] | 1.33 [1.23-1.44] | 1.28 [1.18-1.38] | 1.25 [1.15-1.36] | | 1.25 [1.15-1.35] | 1,860 | 1,795 | 1,733 | NA | 1,708 |
| 975 | 1.33 [1.21-1.47] | 1.41 [1.25-1.59] | 1.56 [1.31-1.86] | 1.16 [1.00-1.34] | 1.23 [1.11-1.37] | 1.19 [1.08-1.32] | 1.18 [1.04-1.33] | 1.26 [1.05-1.52] | 1.10 [0.95-1.28] | 1.11 [0.99-1.24] | 1,052 | 729 | 331 | 488 | 889 |
| 976 | 1.25 [1.09-1.43] | 1.75 [1.43-2.14] | 1.84 [1.44-2.37] | 1.01 [0.84-1.21] | 1.24 [1.07-1.43] | 1.17 [1.02-1.35] | 1.48 [1.20-1.82] | 1.56 [1.21-2.03] | 0.97 [0.81-1.16] | 1.19 [1.02-1.38] | 527 | 265 | 178 | 290 | 465 |
| 979 | 1.31 [1.22-1.40] | 1.38 [1.28-1.49] | 1.46 [1.33-1.61] | 1.04 [0.92-1.17] | 1.19 [1.10-1.28] | 1.19 [1.11-1.28] | 1.14 [1.06-1.24] | 1.20 [1.08-1.32] | 1.01 [0.89-1.15] | 1.13 [1.05-1.22] | 2,180 | 1,741 | 1,134 | 656 | 1,859 |
| 981 | 1.13 [1.09-1.18] | 1.16 [1.11-1.21] | 1.26 [1.18-1.35] | 0.96 [0.90-1.03] | 1.05 [1.00-1.10] | 1.06 [1.02-1.11] | 0.98 [0.93-1.02] | 1.04 [0.97-1.11] | 0.98 [0.91-1.05] | 1.05 [1.00-1.09] | 5,797 | 5,029 | 2,102 | 2,038 | 4,832 |
| 983 | 0.98 [0.82-1.16] | 1.01 [0.80-1.29] | | 0.86 [0.69-1.06] | 0.95 [0.79-1.15] | 0.96 [0.80-1.14] | 0.94 [0.73-1.20] | | 0.85 [0.68-1.06] | 0.94 [0.78-1.14] | 338 | 162 | NA | 215 | 271 |
| 985 | 1.16 [0.91-1.49] | 1.17 [0.89-1.53] | 1.17 [0.87-1.58] | | 1.10 [0.85-1.42] | 1.10 [0.85-1.41] | 1.05 [0.80-1.39] | 1.06 [0.78-1.44] | | 1.11 [0.86-1.44] | 153 | 131 | 102 | NA | 138 |
| 986 | 1.08 [0.81-1.43] | | | | | 1.07 [0.80-1.43] | | | | | 115 | NA | NA | NA | NA |
| 987 | 1.05 [0.90-1.23] | 1.08 [0.91-1.27] | 1.12 [0.93-1.36] | 1.04 [0.77-1.41] | 1.04 [0.89-1.22] | 1.02 [0.87-1.19] | 0.97 [0.82-1.16] | 1.02 [0.84-1.25] | 1.05 [0.77-1.43] | 1.03 [0.87-1.21] | 405 | 330 | 256 | 102 | 360 |
| 988 | 1.31 [1.05-1.63] | | | 1.12 [0.84-1.48] | 1.22 [0.95-1.56] | 1.23 [0.98-1.53] | | | 1.08 [0.81-1.44] | 1.15 [0.90-1.48] | 204 | NA | NA | 117 | 165 |
| 989 | 0.99 [0.87-1.13] | 1.11 [0.89-1.38] | 1.21 [0.94-1.58] | 0.92 [0.78-1.08] | 0.92 [0.80-1.06] | 0.95 [0.82-1.09] | 0.95 [0.76-1.19] | 1.05 [0.80-1.38] | 0.89 [0.76-1.05] | 0.90 [0.78-1.05] | 503 | 192 | 139 | 337 | 438 |
| 990 | 1.17 [1.14-1.20] | 1.17 [1.14-1.20] | 1.16 [1.13-1.19] | 1.09 [0.91-1.30] | 1.09 [1.07-1.12] | 1.14 [1.11-1.17] | 1.10 [1.08-1.13] | 1.10 [1.07-1.13] | 1.13 [0.94-1.36] | 1.10 [1.07-1.13] | 15,940 | 15,749 | 15,211 | 329 | 15,016 |
| 994.2 | 1.24 [1.22-1.26] | 1.25 [1.23-1.27] | 1.25 [1.22-1.27] | 1.09 [1.01-1.17] | 1.19 [1.16-1.21] | 1.20 [1.18-1.22] | 1.16 [1.14-1.19] | 1.16 [1.14-1.18] | 1.06 [0.98-1.14] | 1.17 [1.15-1.19] | 34,356 | 33,202 | 31,291 | 1,833 | 31,894 |
| other | | | | | | | | | | | | | | | |
| 1000 | 1.00 [0.95-1.04] | 1.11 [1.04-1.19] | 1.13 [1.05-1.23] | 0.93 [0.87-0.98] | 0.96 [0.92-1.01] | 0.98 [0.93-1.02] | 1.04 [0.97-1.12] | 1.05 [0.97-1.14] | 0.92 [0.86-0.98] | 0.96 [0.91-1.01] | 4,449 | 2,160 | 1,498 | 2,585 | 3,769 |
| 1001 | 1.12 [1.09-1.16] | 1.31 [1.25-1.38] | 1.30 [1.22-1.37] | 1.02 [0.98-1.07] | 1.06 [1.02-1.10] | 1.07 [1.04-1.11] | 1.18 [1.13-1.25] | 1.18 [1.11-1.25] | 1.00 [0.96-1.05] | 1.03 [0.99-1.07] | 8,527 | 3,990 | 3,011 | 4,947 | 7,126 |
| 1002 | 1.27 [1.24-1.29] | 1.27 [1.25-1.29] | 1.27 [1.24-1.29] | 1.15 [1.09-1.22] | 1.20 [1.18-1.22] | 1.20 [1.17-1.22] | 1.16 [1.13-1.18] | 1.15 [1.13-1.18] | 1.11 [1.05-1.18] | 1.16 [1.14-1.18] | 32,744 | 30,266 | 27,215 | 3,638 | 29,116 |
| 1004 | 1.19 [1.15-1.23] | 1.18 [1.13-1.22] | 1.27 [1.21-1.33] | 1.06 [1.01-1.12] | 1.11 [1.07-1.15] | 1.11 [1.07-1.15] | 1.01 [0.97-1.05] | 1.09 [1.03-1.14] | 1.04 [0.99-1.10] | 1.09 [1.05-1.12] | 9,786 | 6,775 | 4,516 | 4,166 | 8,760 |
| 1005 | 1.28 [1.24-1.33] | 1.31 [1.26-1.36] | 1.31 [1.25-1.36] | 1.05 [0.94-1.16] | 1.21 [1.17-1.26] | 1.22 [1.18-1.27] | 1.20 [1.16-1.25] | 1.20 [1.15-1.25] | 1.02 [0.92-1.14] | 1.18 [1.14-1.23] | 7,534 | 6,857 | 6,110 | 890 | 6,933 |

Supplementary Table 6: phecode-mapped results (hazard ratios and events from all cohorts)

| | Hazard ratio (99% confidence interval) | | | | | | | | | | Events (in exposed) | | | | |
|---|---|---|---|---|---|---|---|---|---|---|---|---|---|---|---|
| | crude | | | | | adjusted | | | | | | | | | |
| Outcome | any age | 18+ | 40+ | <18 | hosp. | any age | 18+ | 40+ | <18 | hosp. | any age | 18+ | 40+ | <18 | hosp. |
| 1006 | 0.92 [0.82-1.03] | 0.89 [0.75-1.05] | 0.90 [0.72-1.11] | 0.94 [0.81-1.08] | 0.91 [0.80-1.03] | 0.91 [0.81-1.02] | 0.87 [0.74-1.04] | 0.88 [0.70-1.09] | 0.93 [0.80-1.09] | 0.91 [0.80-1.04] | 684 | 304 | 174 | 431 | 542 |
| 1007 | 1.09 [0.99-1.19] | 1.12 [1.01-1.24] | 1.16 [1.02-1.32] | 1.00 [0.85-1.17] | 1.02 [0.92-1.13] | 1.08 [0.98-1.19] | 1.07 [0.97-1.19] | 1.11 [0.98-1.27] | 1.02 [0.87-1.21] | 1.04 [0.93-1.16] | 1,122 | 929 | 553 | 400 | 858 |
| 1008 | 1.11 [1.06-1.16] | 1.13 [1.07-1.18] | 1.16 [1.10-1.23] | 1.01 [0.92-1.10] | 1.05 [1.00-1.11] | 1.08 [1.03-1.13] | 1.06 [1.01-1.11] | 1.09 [1.03-1.15] | 1.01 [0.93-1.11] | 1.05 [1.00-1.10] | 5,009 | 4,318 | 3,179 | 1,306 | 4,128 |
| 1009 | 1.16 [1.15-1.18] | 1.20 [1.18-1.22] | 1.23 [1.21-1.26] | 1.06 [1.03-1.09] | 1.10 [1.09-1.12] | 1.11 [1.09-1.13] | 1.10 [1.08-1.12] | 1.13 [1.10-1.15] | 1.04 [1.01-1.07] | 1.07 [1.06-1.09] | 45,692 | 35,468 | 26,227 | 14,115 | 38,103 |
| 1010 | 1.44 [1.42-1.46] | 1.33 [1.31-1.35] | 1.33 [1.31-1.35] | 1.97 [1.91-2.03] | 1.35 [1.34-1.37] | 1.35 [1.33-1.37] | 1.22 [1.20-1.23] | 1.22 [1.20-1.24] | 1.74 [1.68-1.79] | 1.29 [1.28-1.31] | 60,388 | 50,680 | 37,629 | 13,628 | 53,513 |
| 1011 | 1.29 [1.26-1.33] | 1.30 [1.27-1.34] | 1.29 [1.25-1.33] | 1.20 [1.12-1.28] | 1.21 [1.17-1.24] | 1.21 [1.18-1.24] | 1.17 [1.14-1.21] | 1.17 [1.13-1.21] | 1.15 [1.07-1.24] | 1.16 [1.12-1.19] | 13,419 | 11,896 | 9,589 | 2,259 | 11,695 |
| 1012 | 0.97 [0.79-1.19] | 1.32 [1.00-1.73] | | 0.79 [0.59-1.06] | 1.06 [0.86-1.31] | 0.96 [0.78-1.18] | 1.27 [0.96-1.68] | | 0.81 [0.60-1.09] | 1.08 [0.87-1.34] | 222 | 125 | NA | 106 | 204 |
| 1013 | 1.25 [1.17-1.33] | 1.25 [1.17-1.33] | 1.27 [1.18-1.36] | 1.14 [0.95-1.38] | 1.18 [1.11-1.26] | 1.20 [1.12-1.28] | 1.14 [1.07-1.22] | 1.16 [1.08-1.25] | 1.15 [0.95-1.39] | 1.16 [1.08-1.24] | 2,516 | 2,300 | 2,119 | 293 | 2,317 |
| 1014 | 1.00 [0.95-1.06] | 1.12 [1.02-1.22] | 1.09 [0.99-1.21] | 0.96 [0.90-1.04] | 0.98 [0.92-1.04] | 0.99 [0.93-1.05] | 1.05 [0.96-1.15] | 1.03 [0.93-1.14] | 0.96 [0.89-1.03] | 0.97 [0.91-1.04] | 2,854 | 1,270 | 983 | 1,724 | 2,470 |
| 1015 | 1.47 [1.45-1.50] | 1.33 [1.31-1.36] | 1.35 [1.32-1.38] | 1.66 [1.62-1.71] | 1.35 [1.33-1.38] | 1.34 [1.32-1.37] | 1.15 [1.12-1.17] | 1.18 [1.15-1.21] | 1.54 [1.50-1.59] | 1.28 [1.26-1.31] | 36,043 | 24,828 | 15,848 | 15,319 | 30,824 |
| 1019 | 1.24 [1.22-1.25] | 1.24 [1.22-1.25] | 1.23 [1.21-1.25] | 1.18 [1.14-1.23] | 1.16 [1.14-1.17] | 1.19 [1.17-1.20] | 1.14 [1.13-1.16] | 1.15 [1.13-1.17] | 1.14 [1.10-1.19] | 1.13 [1.11-1.15] | 45,241 | 40,851 | 33,837 | 6,336 | 37,400 |
| 1100 | 1.23 [1.21-1.27] | 1.22 [1.19-1.26] | 1.20 [1.16-1.25] | 1.24 [1.19-1.29] | 1.17 [1.14-1.20] | 1.17 [1.14-1.20] | 1.13 [1.10-1.16] | 1.11 [1.07-1.15] | 1.19 [1.14-1.24] | 1.13 [1.10-1.16] | 17,209 | 13,223 | 6,277 | 6,352 | 14,733 |
