## Supplementary Table 7 for "Mapping risks of hospital-recorded health conditions in people with eczema"

Supplementary Table 7: Largest differences in crude hazard ratios between any-age and <18 cohorts

| | Hazard ratio (99% confidence interval) | | Events (in exposed) | | |
|---|---|---|---|---|---|
| outcome | any age | <18 | any age | <18 | change[†] |
| Q90 Down syndrome | 2.77 [2.46-3.12] | 1.03 [0.81-1.31] | 855 | 161 | −62.70% |
| F72 Severe mental retardation | 1.67 [1.43-1.94] | 1.03 [0.82-1.29] | 485 | 197 | −38.42% |
| I83 Varicose veins of lower extremities | 1.63 [1.59-1.67] | 1.05 [0.86-1.27] | 19,256 | 254 | −35.76% |
| I27 Other pulmonary heart diseases | 1.32 [1.28-1.37] | 0.85 [0.63-1.15] | 9,635 | 106 | −35.58% |
| M77 Other enthesopathies | 1.47 [1.40-1.56] | 0.95 [0.73-1.26] | 3,405 | 136 | −35.23% |
| N02 Recurrent and persistent haematuria | 1.32 [1.22-1.43] | 0.86 [0.66-1.12] | 1,604 | 139 | −35.12% |
| L25 Unspecified contact dermatitis | 2.99 [2.63-3.39] | 1.95 [1.49-2.56] | 847 | 164 | −34.65% |
| G82 Paraplegia and tetraplegia | 1.15 [1.05-1.25] | 0.75 [0.61-0.93] | 1,261 | 198 | −34.51% |
| K72 Hepatic failure, NEC | 1.36 [1.28-1.43] | 0.89 [0.67-1.18] | 3,232 | 125 | −34.37% |
| I87 Other disorders of veins | 1.60 [1.52-1.69] | 1.06 [0.79-1.42] | 4,228 | 125 | −33.92% |
| E14 Unspecified diabetes mellitus | 1.37 [1.33-1.41] | 0.92 [0.75-1.14] | 11,113 | 228 | −32.67% |
| F79 Unspecified mental retardation | 1.56 [1.47-1.66] | 1.06 [0.96-1.16] | 2,891 | 1,151 | −32.39% |
| Z99 Dependence on enabling machines and devices, NEC | 1.39 [1.36-1.43] | 0.94 [0.86-1.04] | 16,202 | 1,037 | −32.01% |
| I07 Rheumatic tricuspid valve diseases | 1.31 [1.24-1.38] | 0.89 [0.67-1.18] | 4,088 | 118 | −31.99% |
| F70 Mild mental retardation | 1.72 [1.51-1.97] | 1.19 [0.94-1.51] | 625 | 183 | −30.95% |
| K86 Other diseases of pancreas | 1.21 [1.17-1.27] | 0.86 [0.70-1.05] | 5,937 | 244 | −29.38% |
| F53 Mental and behavioural disorders associated with the puerperium, NEC | 1.35 [1.19-1.53] | 0.96 [0.73-1.26] | 619 | 135 | −28.98% |
| T13 Other injuries of lower limb, level unspecified | 1.17 [1.04-1.31] | 0.83 [0.63-1.10] | 755 | 121 | −28.80% |
| G24 Dystonia | 1.14 [1.03-1.25] | 0.82 [0.67-1.00] | 1,028 | 250 | −27.96% |
| G62 Other polyneuropathies | 1.35 [1.30-1.40] | 0.97 [0.78-1.22] | 6,954 | 198 | −27.76% |
| N31 Neuromuscular dysfunction of bladder, NEC | 1.35 [1.26-1.43] | 0.98 [0.82-1.16] | 2,392 | 319 | −27.36% |
| F81 Specific developmental disorders of scholastic skills | 1.56 [1.50-1.61] | 1.13 [1.07-1.19] | 8,353 | 3,825 | −27.33% |
| N04 Nephrotic syndrome | 1.34 [1.22-1.48] | 0.99 [0.82-1.20] | 1,140 | 273 | −26.10% |
| D73 Diseases of spleen | 1.25 [1.17-1.34] | 0.92 [0.72-1.18] | 2,222 | 169 | −26.10% |
| N27 Small kidney of unknown cause | 1.30 [1.15-1.47] | 0.96 [0.73-1.27] | 659 | 129 | −26.07% |
| F29 Unspecified nonorganic psychosis | 1.22 [1.13-1.32] | 0.91 [0.77-1.07] | 1,809 | 375 | −25.65% |
| Q28 Other congenital malformations of circulatory system | 1.32 [1.12-1.54] | 0.98 [0.71-1.35] | 391 | 101 | −25.59% |
| J81 Pulmonary oedema | 1.30 [1.24-1.36] | 0.97 [0.72-1.31] | 4,916 | 107 | −25.39% |
| I61 Intracerebral haemorrhage | 1.17 [1.12-1.23] | 0.88 [0.65-1.19] | 4,912 | 104 | −25.11% |
| V89 Motor or nonmotor vehicle accident, type of vehicle unspecified | 1.20 [1.02-1.40] | 0.90 [0.67-1.20] | 370 | 111 | −24.90% |
| I89 Other noninfective disorders of lymphatic vessels and lymph nodes | 1.53 [1.46-1.59] | 1.16 [0.91-1.47] | 5,461 | 177 | −24.13% |
| O74 Complications of anaesthesia during labour and delivery | 1.16 [1.00-1.34] | 0.88 [0.66-1.16] | 464 | 122 | −24.07% |
| L51 Erythema multiforme | 1.28 [1.14-1.43] | 0.97 [0.83-1.13] | 767 | 416 | −24.05% |
| S20 Superficial injury of thorax | 1.18 [1.11-1.25] | 0.90 [0.77-1.05] | 2,809 | 409 | −23.95% |
| F42 Obsessive compulsive disorder | 1.64 [1.52-1.76] | 1.25 [1.09-1.43] | 2,087 | 635 | −23.80% |
| L20 Atopic dermatitis | 24.72 [22.44-27.23] | 18.85 [16.88-21.04] | 6,205 | 3,853 | −23.76% |
| F60 Specific personality disorders | 1.34 [1.27-1.42] | 1.02 [0.92-1.14] | 3,390 | 915 | −23.63% |
| T82 Complications of cardiac and vascular prosthetic devices, implants and grafts | 1.31 [1.27-1.35] | 1.01 [0.91-1.11] | 9,620 | 979 | −23.30% |
| T86 Failure and rejection of transplanted organs and tissues | 1.33 [1.22-1.46] | 1.02 [0.82-1.28] | 1,200 | 200 | −23.26% |
| S21 Open wound of thorax | 1.11 [0.97-1.26] | 0.85 [0.70-1.04] | 604 | 247 | −23.24% |
| T14 Injury of unspecified body region | 1.16 [1.03-1.29] | 0.89 [0.71-1.13] | 756 | 169 | −22.77% |
| M46 Other inflammatory spondylopathies | 1.42 [1.36-1.48] | 1.10 [0.83-1.45] | 5,856 | 135 | −22.71% |
| Z95 Presence of cardiac and vascular implants and grafts | 1.22 [1.21-1.24] | 0.95 [0.86-1.05] | 45,729 | 1,021 | −22.54% |
| I37 Pulmonary valve disorders | 1.22 [1.12-1.32] | 0.95 [0.75-1.20] | 1,558 | 167 | −22.20% |
| F22 Persistent delusional disorders | 1.29 [1.19-1.40] | 1.00 [0.74-1.36] | 1,560 | 113 | −22.18% |
| I35 Nonrheumatic aortic valve disorders | 1.23 [1.20-1.26] | 0.95 [0.78-1.17] | 19,023 | 238 | −22.16% |
| R34 Anuria and oliguria | 1.21 [1.11-1.33] | 0.95 [0.73-1.23] | 1,225 | 145 | −22.05% |
| R18 Ascites | 1.20 [1.17-1.24] | 0.94 [0.81-1.10] | 9,491 | 441 | −21.92% |
| Z60 Problems related to social environment | 1.18 [1.15-1.21] | 0.92 [0.80-1.06] | 21,873 | 524 | −21.81% |
| G80 Cerebral palsy | 1.01 [0.94-1.09] | 0.79 [0.72-0.87] | 1,750 | 1,019 | −21.48% |
| I78 Diseases of capillaries | 1.30 [1.21-1.40] | 1.03 [0.77-1.36] | 1,829 | 121 | −21.45% |
| F20 Schizophrenia | 1.28 [1.21-1.34] | 1.00 [0.84-1.20] | 3,454 | 313 | −21.31% |
| I86 Varicose veins of other sites | 1.31 [1.22-1.41] | 1.03 [0.88-1.21] | 1,918 | 412 | −21.15% |
| Q60 Renal agenesis and other reduction defects of kidney | 1.17 [1.03-1.33] | 0.92 [0.72-1.18] | 628 | 166 | −21.15% |
| M47 Spondylosis | 1.35 [1.33-1.38] | 1.07 [0.85-1.35] | 32,121 | 193 | −21.02% |
| E10 Type 1 diabetes mellitus | 1.23 [1.19-1.26] | 0.97 [0.91-1.04] | 10,587 | 2,324 | −20.90% |
| Q76 Congenital malformations of spine and bony thorax | 1.15 [1.02-1.31] | 0.91 [0.75-1.12] | 639 | 242 | −20.85% |
| G96 Other disorders of central nervous system | 1.30 [1.17-1.44] | 1.03 [0.79-1.33] | 875 | 145 | −20.75% |
| J69 Pneumonitis due to solids and liquids | 1.17 [1.14-1.20] | 0.93 [0.78-1.10] | 13,535 | 323 | −20.74% |
| I69 Sequelae of cerebrovascular disease | 1.18 [1.15-1.22] | 0.94 [0.73-1.21] | 10,401 | 153 | −20.43% |
| F43 Reaction to severe stress, and adjustment disorders | 1.26 [1.20-1.33] | 1.00 [0.90-1.12] | 3,877 | 901 | −20.36% |
| R12 Heartburn | 1.35 [1.29-1.42] | 1.08 [0.90-1.28] | 4,130 | 336 | −20.33% |
| M43 Other deforming dorsopathies | 1.29 [1.24-1.34] | 1.02 [0.90-1.17] | 7,046 | 575 | −20.32% |
| T11 Other injuries of upper limb, level unspecified | 1.19 [1.08-1.31] | 0.95 [0.78-1.16] | 1,227 | 253 | −20.12% |
| Z94 Transplanted organ and tissue status | 1.36 [1.28-1.44] | 1.09 [0.94-1.26] | 2,996 | 449 | −20.04% |
| R52 Pain, NEC | 1.40 [1.34-1.46] | 1.12 [0.98-1.28] | 6,030 | 610 | −20.02% |
| R16 Hepatomegaly and splenomegaly, NEC | 1.38 [1.32-1.45] | 1.11 [0.99-1.24] | 5,156 | 829 | −19.92% |
| G90 Disorders of autonomic nervous system | 1.26 [1.15-1.40] | 1.01 [0.77-1.33] | 1,018 | 143 | −19.92% |
| Z93 Artificial opening status | 1.19 [1.16-1.23] | 0.96 [0.87-1.05] | 13,568 | 1,158 | −19.86% |
| G41 Status epilepticus | 1.20 [1.10-1.31] | 0.96 [0.84-1.11] | 1,276 | 479 | −19.80% |

[†] Percent change of crude hazard ratio from any-age cohort to <18 cohort.
