## Supplementary Table 8 for "Mapping risks of hospital-recorded health conditions in people with eczema"

Supplementary Table 8: Largest differences in crude hazard ratios between any-age and 40+ cohorts

| | Hazard ratio (99% confidence interval) | | Events (in exposed) | | |
|---|---|---|---|---|---|
| outcome | any age | 40+ | any age | 40+ | change[1] |
| T78 Adverse effects, NEC | 3.07 [2.96-3.18] | 1.72 [1.61-1.83] | 10,510 | 2,499 | −44.05% |
| Z01 Other special examinations and investigations of persons withou... | 2.34 [2.27-2.42] | 1.43 [1.34-1.53] | 11,656 | 2,154 | −38.88% |
| B00 Herpesviral [herpes simplex] infections | 2.52 [2.38-2.67] | 1.73 [1.58-1.91] | 3,846 | 1,174 | −31.16% |
| J46 Status asthmaticus | 2.95 [2.79-3.13] | 2.08 [1.85-2.33] | 3,968 | 809 | −29.63% |
| Z71 Persons encountering health services for other counselling and ... | 1.65 [1.59-1.70] | 1.18 [1.12-1.23] | 9,814 | 4,554 | −28.54% |
| J30 Vasomotor and allergic rhinitis | 2.14 [2.08-2.20] | 1.67 [1.60-1.74] | 17,750 | 6,066 | −22.09% |
| O82 Single delivery by caesarean section | 1.10 [1.03-1.19] | 0.86 [0.67-1.11] | 1,727 | 120 | −21.99% |
| O23 Infections of genitourinary tract in pregnancy | 1.16 [1.12-1.20] | 0.93 [0.73-1.19] | 6,682 | 135 | −19.91% |
| Z84 Family history of other conditions | 1.76 [1.66-1.87] | 1.41 [1.22-1.64] | 3,330 | 433 | −19.90% |
| V80 Animal rider or occupant of animal drawn vehicle injured in tra... | 0.95 [0.86-1.04] | 0.76 [0.64-0.90] | 1,094 | 288 | −19.87% |
| H03 Disorders of eyelid in diseases CE | 1.61 [1.30-1.98] | 1.30 [0.96-1.76] | 247 | 107 | −19.28% |
| O47 False labour | 1.16 [1.12-1.21] | 0.95 [0.75-1.20] | 6,347 | 146 | −18.54% |
| O42 Premature rupture of membranes | 1.03 [1.01-1.06] | 0.84 [0.75-0.95] | 18,440 | 557 | −18.31% |
| O26 Maternal care for other conditions predominantly related to pre... | 1.16 [1.13-1.18] | 0.95 [0.85-1.05] | 25,422 | 756 | −17.89% |
| O48 Prolonged pregnancy | 1.02 [0.99-1.05] | 0.84 [0.71-1.00] | 10,930 | 279 | −17.48% |
| W21 Striking against or struck by sports equipment | 1.11 [1.03-1.21] | 0.94 [0.73-1.22] | 1,513 | 131 | −15.21% |
| F91 Conduct disorders | 1.27 [1.13-1.42] | 1.09 [0.81-1.47] | 820 | 106 | −14.35% |
| T63 Toxic effect of contact with venomous animals | 1.10 [0.89-1.37] | 0.95 [0.72-1.25] | 200 | 111 | −13.87% |
| B34 Viral infection of unspecified site | 1.54 [1.52-1.57] | 1.33 [1.27-1.40] | 40,130 | 3,879 | −13.70% |
| O30 Multiple gestation | 1.00 [0.94-1.06] | 0.86 [0.69-1.07] | 2,655 | 170 | −13.61% |
| X58 Exposure to other specified factors | 1.60 [1.47-1.75] | 1.39 [1.23-1.56] | 1,466 | 729 | −13.41% |
| J45 Asthma | 2.12 [2.11-2.14] | 1.84 [1.82-1.87] | 165,066 | 75,502 | −13.16% |
| O40 Polyhydramnios | 1.09 [1.03-1.16] | 0.95 [0.76-1.19] | 2,620 | 158 | −12.81% |
| B90 Sequelae of tuberculosis | 1.60 [1.25-2.03] | 1.39 [1.08-1.79] | 184 | 167 | −12.71% |
| J82 Pulmonary eosinophilia, NEC | 2.40 [2.08-2.78] | 2.11 [1.80-2.47] | 600 | 451 | −12.34% |
| R06 Abnormalities of breathing | 1.52 [1.50-1.54] | 1.33 [1.31-1.35] | 74,870 | 34,545 | −12.29% |
| T94 Sequelae of injuries involving multiple and unspecified body re... | 1.26 [1.01-1.57] | 1.11 [0.85-1.44] | 196 | 139 | −12.26% |
| O75 Other CO labour and delivery, NEC | 1.04 [1.01-1.07] | 0.91 [0.79-1.05] | 11,471 | 410 | −12.25% |
| Q79 Congenital malformations of the musculoskeletal system, NEC | 1.35 [1.22-1.51] | 1.19 [0.98-1.45] | 920 | 242 | −11.73% |
| Z34 Supervision of normal pregnancy | 1.09 [1.07-1.11] | 0.97 [0.85-1.09] | 20,140 | 555 | −11.39% |
| Z91 PH of risk factors, NEC | 1.76 [1.73-1.78] | 1.56 [1.53-1.59] | 66,982 | 30,367 | −11.21% |
| O14 Pre eclampsia | 1.10 [1.05-1.16] | 0.98 [0.80-1.20] | 4,133 | 205 | −11.17% |
| B01 Varicella [chickenpox] | 1.61 [1.52-1.72] | 1.43 [1.10-1.87] | 2,710 | 139 | −11.12% |
| O13 Gestational [pregnancy induced] hypertension | 1.08 [1.04-1.13] | 0.96 [0.81-1.14] | 5,577 | 288 | −10.98% |
| O68 Labour and delivery complicated by fetal stress [distress] | 1.04 [1.03-1.06] | 0.93 [0.85-1.01] | 35,814 | 1,175 | −10.95% |
| Q50 Congenital malformations of ovaries, fallopian tubes and broad ... | 1.25 [1.11-1.41] | 1.12 [0.91-1.37] | 682 | 206 | −10.74% |
| L28 Lichen simplex chronicus and prurigo | 4.46 [4.06-4.91] | 3.99 [3.59-4.42] | 1,757 | 1,387 | −10.69% |
| O63 Long labour | 1.02 [0.99-1.04] | 0.91 [0.80-1.03] | 15,999 | 487 | −10.63% |
| G10 Huntington disease | 1.40 [1.07-1.84] | 1.26 [0.94-1.67] | 135 | 113 | −10.60% |
| O70 Perineal laceration during delivery | 1.03 [1.02-1.05] | 0.93 [0.85-1.01] | 48,711 | 1,170 | −10.51% |
| O04 Medical abortion | 1.02 [1.00-1.05] | 0.92 [0.81-1.04] | 11,751 | 521 | −10.44% |
| K09 Cysts of oral region, NEC | 1.26 [1.14-1.40] | 1.13 [0.99-1.30] | 971 | 489 | −10.38% |
| Z35 Supervision of high risk pregnancy | 1.07 [1.04-1.09] | 0.96 [0.89-1.03] | 17,420 | 1,680 | −10.33% |
| O41 Other disorders of amniotic fluid and membranes | 1.10 [1.04-1.16] | 0.99 [0.77-1.27] | 3,493 | 132 | −10.12% |
| O61 Failed induction of labour | 1.08 [1.01-1.17] | 0.98 [0.75-1.27] | 1,800 | 120 | −10.00% |
| H18 Other disorders of cornea | 1.38 [1.32-1.44] | 1.24 [1.18-1.31] | 5,208 | 3,982 | −9.68% |
| K90 Intestinal malabsorption | 1.78 [1.72-1.85] | 1.61 [1.53-1.70] | 8,443 | 3,833 | −9.59% |
| J31 Chronic rhinitis, nasopharyngitis and pharyngitis | 1.75 [1.66-1.86] | 1.59 [1.47-1.72] | 3,383 | 1,625 | −9.32% |
| G09 Sequelae of inflammatory diseases of central nervous system | 1.36 [1.14-1.63] | 1.24 [0.98-1.56] | 311 | 176 | −9.15% |
| O20 Haemorrhage in early pregnancy | 1.06 [1.03-1.10] | 0.97 [0.83-1.13] | 8,064 | 356 | −9.14% |
| O99 Other maternal diseases classifiable elsewhere but complicating... | 1.16 [1.14-1.19] | 1.06 [0.97-1.15] | 27,987 | 1,210 | −9.12% |
| O36 Maternal care for other known or suspected fetal problems | 1.12 [1.10-1.14] | 1.02 [0.93-1.11] | 33,543 | 1,131 | −9.10% |
| L63 Alopecia areata | 2.87 [2.32-3.56] | 2.62 [1.97-3.48] | 290 | 151 | −8.96% |
| O28 Abnormal findings on antenatal screening of mother | 1.06 [0.99-1.13] | 0.97 [0.75-1.25] | 2,337 | 122 | −8.78% |
| B59 Pneumocystosis | 1.53 [1.24-1.88] | 1.39 [1.12-1.73] | 236 | 205 | −8.73% |
| O43 Placental disorders | 1.03 [0.97-1.10] | 0.94 [0.71-1.25] | 2,479 | 103 | −8.65% |
| O62 Abnormalities of forces of labour | 1.05 [1.01-1.09] | 0.96 [0.81-1.15] | 7,213 | 271 | −8.58% |
| K50 Crohn disease [regional enteritis] | 1.70 [1.63-1.77] | 1.56 [1.47-1.64] | 6,324 | 3,344 | −8.52% |
| L27 Dermatitis due to substances taken internally | 1.75 [1.65-1.86] | 1.60 [1.49-1.72] | 3,074 | 2,008 | −8.50% |
| G03 Meningitis DTOAUC | 1.24 [1.07-1.43] | 1.13 [0.92-1.40] | 464 | 200 | −8.42% |
| L30 Other dermatitis | 10.30 [10.10-10.50] | 9.44 [9.16-9.73] | 77,273 | 27,658 | −8.36% |
| D28 Benign neoplasm of OAU female genital organs | 1.25 [1.06-1.47] | 1.14 [0.93-1.40] | 347 | 218 | −8.33% |
| H15 Disorders of sclera | 1.41 [1.17-1.71] | 1.30 [1.04-1.62] | 277 | 208 | −8.26% |
| O16 Unspecified maternal hypertension | 1.12 [1.07-1.17] | 1.03 [0.86-1.24] | 4,610 | 243 | −8.06% |
| O72 Postpartum haemorrhage | 1.05 [1.03-1.08] | 0.97 [0.89-1.06] | 23,598 | 1,079 | −8.06% |
| D39 Neoplasm of uncertain or unknown behaviour of female genital or... | 1.06 [0.92-1.22] | 0.97 [0.83-1.14] | 459 | 329 | −8.01% |
| O34 Maternal care for known or suspected abnormality of pelvic organs | 1.09 [1.06-1.11] | 1.00 [0.93-1.08] | 18,898 | 1,365 | −7.96% |
| R71 Abnormality of red blood cells | 1.22 [1.00-1.49] | 1.13 [0.91-1.40] | 249 | 200 | −7.95% |
| N34 Urethritis and urethral syndrome | 1.35 [1.16-1.58] | 1.25 [1.03-1.52] | 404 | 251 | −7.56% |
| D29 Benign neoplasm of male genital organs | 1.23 [1.02-1.49] | 1.14 [0.92-1.42] | 263 | 208 | −7.26% |

[1] Percent change of crude hazard ratio from any-age cohort to 40+ cohort.
